# Comparing the Phenotypic and Genetic Differences Between Autistic Regression and Autism Without Regression Using the SPARK Cohort

**DOI:** 10.64898/2026.10.26.26366210

**Authors:** Ashraf Yahia, Yali Zhang, Antonia Gkerdi, Shyam Sundar Rajagopalan, Kristiina Tammimies

**Author notes:** Correspondence: Kristiina Tammimies; postal address: K6 Kvinnors och barns hälsa, K6 Neuropsykiatri Tammimies, 171 77 Stockholm.

## Abstract

The biology of autistic regression (Autreg) remains poorly understood. We investigated Autreg’s phenotypic and genetic landscape in 51 633 individuals from the SPARK cohort. We calculated the frequencies of parental-reported language regression and regression affecting social, play, toileting, and/or other developmental domains, and ages at skill regression and regain within SPARK. We compared developmental milestones, quantitative developmental measures, and selected other phenotypes in Autreg and autism without regression (Autnreg). Rare damaging coding variants and polygenic scores (PGS) in Autreg and Autnreg were compared using genome sequencing data. Rare variants were analyzed using the Transmission And De novo Association (TADA) framework in 1 107 Autreg and 974 Autnreg individuals, followed by pathway enrichment analysis. PGS for autism, attention-deficit/hyperactivity disorder, epilepsy, and schizophrenia were initially compared. PGS validation was conducted in 14 001 Autnreg and 11 638 Autreg individuals using larger SPARK genotyping data. We identified regression in 43% of autistic individuals. Other regression occurred later than language regression, and 60% of skill regain occurred within two years. Autreg was associated with lower Vineland-3 Adaptive Behavior Composite scores (p<0.001) and >twofold increased odds of epilepsy and intellectual disability (p<0.001). Developmental milestones and quantitative measures had modest differences. TADA analysis highlighted genes involved in nucleotide catabolism, transcription, and focal adhesion, with *PTEN* emerging as a strong candidate for Autreg. Schizophrenia PGS explained 0.087% of variance in liability to regression among autistic individuals. In conclusion, Autreg was associated with increased neurodevelopmental morbidity. Schizophrenia PGS and rare variants explained a minimal fraction of its etiology.

## Introduction

Autistic regression (Autreg) signifies a developmental trajectory in a subset of autistic children characterized by loss of previously established language, social, and/or other skills(1–3). The exact prevalence of Autreg is hampered by the lack of a consensus definition; however, two meta-analyses performed in 2013 and 2021 indicated that approximately 30% of autistic children had Autreg, with the age-at-regression estimated to be 21.35 and 19.8 months, respectively(1, 3).

Regression is not restricted to autism and can occur in other neurodevelopmental and non-developmental phenotypes. Regression can also occur in neurodegenerative and nutritional disorders, and following brain injury(2). Developmental phenotypes that can encompass regression include developmental epileptic encephalopathy (DEE) and multiple monogenic neurodevelopmental disorders(2, 4). Previous studies have detected no or minimal association between Autreg and epilepsy(5–7). Data on the association between Autreg and other neurodevelopmental and neuropsychiatric disorders, such as attention deficit hyperactivity disorder (ADHD) and schizophrenia, is limited.

A growing body of evidence suggests that Autreg is associated with more prominent core autism features, such as impaired social communication and restricted and repetitive behaviors, interests, and activities, and reduced cognitive and language developmental functioning compared to autism without regression (Autnreg)(7–12). However, some studies have suggested no differences between the two groups(13, 14). Whether Autreg is a distinct autism subtype is more than an issue of prognosis or nosology. A randomized, single-blinded, placebo-controlled trial involving 26 participants suggested that prednisolone as an adjuvant to risperidone could lead to a favorable behavioral outcome in Autreg(15). Another study found a favorable outcome of behavioral intervention in Autreg before 4 years old, an effect lost if started after 4 years of age(10).

The etiology of Autreg is poorly understood(2, 16). Previous research suggested roles for fever, febrile seizures, family history of neuropsychiatric and autoimmune disorders(17, 18), autoimmunity(19), genetic(4, 16) and environmental determinants(20), and alterations in subcellular processes such as synaptic pruning(21) and mitochondrial function(22).

In this study, we aimed to enhance the understanding of Autreg and its biological underpinnings. We leveraged clinical and genetic data in a large autism cohort, the Simons Foundation Powering Autism Research for Knowledge (SPARK)(23, 24), to explore the clinical differences between individuals with Autreg and Autnreg and within Autreg subtypes, including language regression, other regression (regression affecting social, play, toileting, and other developmental aspects), and both language and other regression. Furthermore, we investigated the possible contributions of rare protein-coding variants and common genetic variants in Autreg.

## Subjects and methods

### Ethics

Ethical approval for the data collection and written informed consents were obtained from the participants within the SPARK project. The Swedish Ethical Committee approved this study and data analysis in Sweden (dnr 2020–00400).

### Study cohorts and demographic and clinical comparisons

We leveraged demographic and phenotype data from the SPARK collection version 14 with a release date of 2024-12-17. We utilized the child background history, basic medical screening, and clinical lab results datasets. Out of 221 969 autistic individuals with available basic medical screening, we proceeded with 51 633 autistic individuals with available information on their regression status. We compared the ages at regression between those with language, other, and both language and other regression using two-sample t-test and summarized the age at skill regain, if any. To minimize including those who could develop Autreg later in life in our Autnreg group, we set the age threshold for the Autnreg group to be > the age at which 95% of those with Autreg manifested regression. Similarly, we set the age for those who did not regain their lost skill to be > the age at which 95% of those with Autreg regained their skills.

We subsequently compared the ages at reaching selected developmental milestones, the values of selected child development quantitative measures, and the occurrence of selected phenotypes and conditions in those with Autreg vs Autnreg, language vs other regression, single vs double regression (both language and other regression), and those who regained vs did not regain their lost skills using logistic regression after adjusting for sex and age at evaluation. The developmental milestones that we assessed were smiling, sitting unsupported, crawling, walking, eating with a spoon, using words, combining words, combining phrases, bladder control, and bowel control. The assessed child development quantitative measures were the Vineland-3 adaptive behavior scales standard Adaptive Behavior Composite (ABC) score(25), Child Behavior Checklist 6-18 years old total problems T score(26), Developmental Coordination Disorder Questionnaire final score(27), Non-Verbal Intelligence Quotient overall full-scale IQ score(28), Repetitive Behavior Scale-Revised(29), and Social-Communication Questionnaire - Lifetime total final score(30).

We included the following clinical conditions: audiovisual abnormalities, sleep problems, neurological and developmental phenotypes (besides autism), mood disturbances, growth and behavioral abnormalities, eating problems, and birth defects. Furthermore, we analyzed the occurrences of ADHD, epilepsy, intellectual disability, schizophrenia, and genomic conditions (monogenic conditions and copy number or structural variants).

For the genetic part, the variant calling dataset SPARK genome sequencing (GS) version1.1 was used. After applying our age cutoffs, we had groups of 1 107 and 974 individuals from the Autreg and Autnreg groups, respectively. We also used a dataset of annotated de novo coding variants in autism discovery cohorts published by Zhou et al., 2022(24), for gene-level validation. The dataset contained 16 779 individuals from multiple ASD cohorts, including the SPARK cohort, with exome sequencing. We included only individuals from SPARK and selected only those who do not overlap with the SPARK GS1.1 (4 884 individuals).

### Variants calling, annotation, and filtering after genome sequencing

De novo variants (DNVs) and transmitted and non-transmitted variants were called from GS using Slivar (version0.2.7) and bcftools (version1.17). Details are in Supplementary Information (SI).

### De Novo rates analysis

We compared DNV rates in coding-only and both coding and non-coding sequences in Autreg and Autnreg using negative binomial regression in MASS R (4.4.2) package.

### Transmission And De novo Association (TADA) Analysis

We applied the Transmission And De novo Association (TADA) framework to assess the contribution of LoF and damaging missense variants (misD, defined as missense variants with REVEL score ≥ 0.75) to Autreg risk. Analyses were performed separately in 1 107 Autreg trios and 974 Autnreg trios. For each gene and variant class, we incorporated counts of DNV, transmitted, and non-transmitted variants in the two groups together with gene-specific mutation rates. Mutation rates for each gene and variant class were obtained from published estimates(31). As misD was not included in the mutation rates estimates by Samocha et al., 2014(31), we estimated misD rate for each gene as 32% of its missense rate based on a recent study(32). We only included genes with mutation rates in Samocha et al., 2014(31) (18,271 genes) and used HGNChelper R package(33) to align the gene names in our dataset and in the mutation rates table. We excluded genes with extreme variant counts (transmitted LoF > 12, transmitted synonymous > 57, or transmitted LoF > 12 with LoF mutation rate < 1e^-06^) prior to analysis to reduce bias from potential technical or annotation artifacts. Those cutoffs corresponded to 99.5% of variants in the Autnreg dataset (Supplementary Figure 1).

TADA integrates DNV and inherited variant evidence within a hierarchical Bayesian framework and calculates gene-level Bayes factors (BFs) comparing a phenotype-risk model with a neutral model(34–36). Herein, we used the TADA-R TADA version(36). We used GPT-5.6 (accessed on August 2026) to refine the TADA analysis pipeline (code is provided in SI). We reviewed, tested, and revised the refined code and confirmed its accuracy and results reproducibility. LoF and misD evidence were modeled separately and combined multiplicatively to obtain an overall gene-level BF. To assess empirical significance, independent null distributions were generated for the Autreg and Autnreg cohorts using 20 000 simulations. Under the null, DNV counts were simulated according to gene-specific mutation rates, while transmitted and non-transmitted counts were generated under the 50:50 transmission model. To improve computational efficiency, possible transmitted/non-transmitted BF values were precomputed for each observed transmission total and used as lookup tables during null simulations. Bayesian false discovery rates were calculated assuming a prior null probability of 0.95, and empirical p-values were derived by comparison with the corresponding cohort-specific null distributions. Finally, Autreg and Autnreg results were integrated to identify possible Autreg-specific risk genes, defined by strong evidence of association in Autreg together with comparatively weak evidence in Autnreg. Details on the TADA analysis are in SI.

### Pathway enrichment analysis

We tested the pathway enrichment of the candidate Autreg-specific genes using Enrichr-KG(37) (https://maayanlab.cloud/enrichr-kg accessed on 12/08/2026). We analyzed the enrichment with the Gene Ontology (GO) biological process, KEGG, and Reactome. We set the minimum libraries per gene and minimum links per gene at 1 and the minimum links per term at 2.

### Gene-level validation

We evaluated whether the candidate Autreg-specific genes were significantly enriched for ultra-rare protein-altering DNVs in Autreg relative to Autnreg in the autism discovery cohort published by Zhou et al., 2022(24). Out of 4 884 individuals in the Zhou et al.,2022, discovery cohort, 1 449 individuals remained after selecting only LoF and misD variants, and 820 remained after applying our age cutoffs for Autreg (386 individuals) and Autnreg (434 individuals).

### Polygenic scores

PGS analysis was performed in two phases, an exploratory and a validation analysis. In the exploratory analysis, we utilized the SPARK GS dataset version1.1 to compare autism(38), ADHD(39), epilepsy(40), and schizophrenia(41) standardized PGS in Autreg (n=857) and Autnreg (n=836). In the validation analysis, we utilized single-nucleotide polymorphisms (SNPs) array data from the SPARK genomic data iWES version3 to compare schizophrenia standardized PGS in Autreg (n=14 001) and Autnreg (n=11 638). We excluded all individuals in the exploratory PGS analysis from the validation PGS analysis.

Following quality control (QC) of the GS, we included only individuals of predicted European descent (9 879 individuals). Details for QC and population stratification are provided in SI and Supplementary Figures 2-6. We selected individuals with available regression information who fulfilled our Autreg (895 individuals) and Autnreg (869 individuals) age cutoffs. We combined the two lists and excluded related individuals using KING (version2.3.2) with a kinship threshold of 0.125. We ended up with 857 individuals with Autreg and 836 individuals with Autnreg.

PGS was calculated using PRS-CS(42). Details for base genome-wide association studies (GWAS) and PGS calculation are in SI. We tested the association between standardized PGS of ASD(38), ADHD(39), epilepsy(40), and schizophrenia(41) and the presence of Autreg using logistic regression adjusting for age at evaluation and sex (glm(group∼ Standardized_PGS + age_at_eval_years + sex)). Similarly, we tested the association between PGS strata and regression status. For each phenotype, individuals were categorized into three PGS strata based on percentile cutoffs: bottom 10% (0–10), middle 80% (10–90), and top 10% (90–100). Multinomial logistic regression models were used to estimate ORs and 95% CIs for the association between PGS strata and regression status, with the 10-90 stratum as the reference. P-values were adjusted for multiple comparisons using the Benjamini-Hochberg FDR method.

Furthermore, we compared the schizophrenia PGS in Autreg (n=14 001) and Autnreg (n=11 638) after excluding individuals included in the exploratory PGS analysis and their relatives. Genotype information was obtained from the SNPs microarray data, the SPARK genomic data iWES version3. Phasing and imputation are described in SI.

To quantify the association between schizophrenia PGS and regression status, we fit a logistic regression model with regression status as the binary outcome and standardized schizophrenia PGS as the primary predictor, adjusting for age at evaluation and sex. The effect of schizophrenia PGS was expressed as ORs per standard deviation (SD) increase in PGS. Model improvement attributable to the schizophrenia PGS was evaluated using a likelihood ratio test comparing the full model to a reduced model that excludes the schizophrenia PGS. The incremental variance explained by the schizophrenia PGS on the observed scale was estimated as the difference in model-based R2 between the linear predictors of the full and reduced logistic regression models.

Observed R^2^ estimates were subsequently transformed onto the liability scale using the method described by Lee et al.,2012(43). We assumed an underlying normally distributed liability to regression and incorporated both the sample case proportion (Pr) and the assumed population prevalence of regression among autistic individuals (K). In our sample, the proportion of individuals with regression was Pr = 0.546 (14 001/25 639). We assumed an Autreg prevalence of K = 0.30 among autistic individuals. The liability-scale R2 was calculated as:

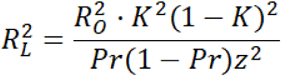

where R^2^_O_ is the incremental observed-scale R² attributable to schizophrenia PGS and z is the standard normal density evaluated at the liability threshold corresponding to K.

## Results

### Approximately 43% of autistic individuals in SPARK reported with regression

The mean age at evaluation of the SPARK individuals used for this study was 9.4 years (SD 6.1, median 8, range 1-77). Supplementary Figure 7. Within the 51 633 (39 644 males and 11 989 females) autistic individuals in SPARK with available information on regression, 43% had Autreg and 57% had Autnreg. The occurrence of regression differed slightly between males (43.8%) and females (42.4%) (two-proportion z-test, χ² = 7.36, p-value = 0.0067), although the effect size was negligible (Cohen’s h 0.03). Isolated language regression was slightly more prevalent than other types of regression (38% and 34%, respectively), the remaining 28% had both language and other regression (double regression) (Figure1).

### Other regressions started later than language regression

The mean age at language regression was 23.1 months (range = 3-85 months, median = 18, 95^th^ centile = 54) while the mean age at other regression was 36.2 months (range = 3-85 months, median = 27, 95^th^ centile = 85). The difference between the two means was statistically significant using a two-sample t-test (t = -54.999, df = 21297, p-value < 2.2e^-16^). Cohen’s d indicated a moderate effect size (0.67). The mean age at other regressions increased to 45.7 months in individuals with single regression and dropped to 28.6 months in those with double regression. The mean ages at language regression were comparable in single and double regression, 21.8 and 24.4 months, respectively (Figure1 and Supplementary Figures 8-10).

### Most regain occurred within 2 years of regression

Approximately 60% of skill regain occurred within 2 years of regression onset. A greater proportion of those with single regression regained their skills within 2 years of regression than those with double regression. For instance, 67.9% and 69.4% of males and 71.1% and 64.2% of females with isolated language and other regression, respectively, regained their lost skill within 2 years. This was in contrast to 57.7% and 52.7% among males, and 63.2% and 60.2% among females, with double regression (Supplementary Figure 11).

### Small to moderate differences in developmental milestones and quantitative measures were detected between the groups

Differences of moderate effect sizes were detected when comparing the Vineland-3 standard Adaptive Behavior Composite (ABC) score in Autreg (mean 68.4, SD 12.6) and Autnreg (mean 74.2, SD 11.8) groups (Cohen’s d 0.49), and skills regain (mean 71.4, SD 11.6) and no regain (mean 61.9, SD 13.8) groups (Cohen’s d 0.62). Also, we detected moderate differences in the ages of combining words (Cohen’s d 0.51) and phrases (Cohen’s d 0.59) in those with isolated language and other regressions. Other studied clinical scores and ages at reaching developmental milestones were of small effect sizes and/or not statistically significant. Table1 and Supplementary Figures 12-19.

### The occurrence of developmental phenotypes, epilepsy, and ID were elevated in AutReg group compared to Autnreg group

Between-group differences in the odds of the studied conditions and phenotypes ranged from non-significant to approximately two-fold increases (Table2, Figure2, and Supplementary Figures 20-25). There was no statistically significant difference in the odds of receiving a genetic/genomic diagnosis between the tested group pairs. Also, no statistically significant difference in the odds of having visual symptoms was detected, except when comparing other to language regression (odds ratio (OR) 1.54, 95% CI 1.28-1.86, p-value 5.18e^-06^).

### No significant difference in de novo variant counts between Autreg and Autnreg

Autreg and Autnreg had comparable DNV counts in both coding-only (incidence rate ratio = 1.075, 95% CI 0.979-1.181, p = 0.130) and coding and non-coding (incidence rate ratio = 0.999, 95% CI 0.975-1.024, p = 0.942) sequences. Mean DNV counts and their distributions are shown in Supplementary Figure 26.

### TADA followed by pathway enrichment analysis suggested a possible role for ectonucleoside triphosphate diphosphohydrolases (NTPDase)-mediated breakdown of nucleotides in Autreg

We analyzed 15 641 genes that harbored de novo, transmitted, and/or non-transmitted variants in the Autreg (n=1 107) and/or Autnreg (n=974) groups using TADA framework (full results in Supplementary Table 1). The analysis revealed 53 significant candidate Autreg-specific genes (genes with adjusted p-value < 0.05 in Autreg TADA and > 0.05 in Autnreg TADA; Supplementary Table 2). We further estimated effect sizes as log₂ rate ratios to characterize the magnitude of enrichment of those 53 genes, and we decomposed signals by inheritance class to distinguish de novo and inherited contributions (Figure 3). Enrichment analysis of those 53 genes showed significant enrichment in the phosphate bond hydrolysis by NTPDase proteins pathway (R-HSA-8850843; q-value 0.04) and nonsignificant enrichment trends in pathways related to transcription and focal adhesion (Supplementary Table 5).

**Figure 1.**
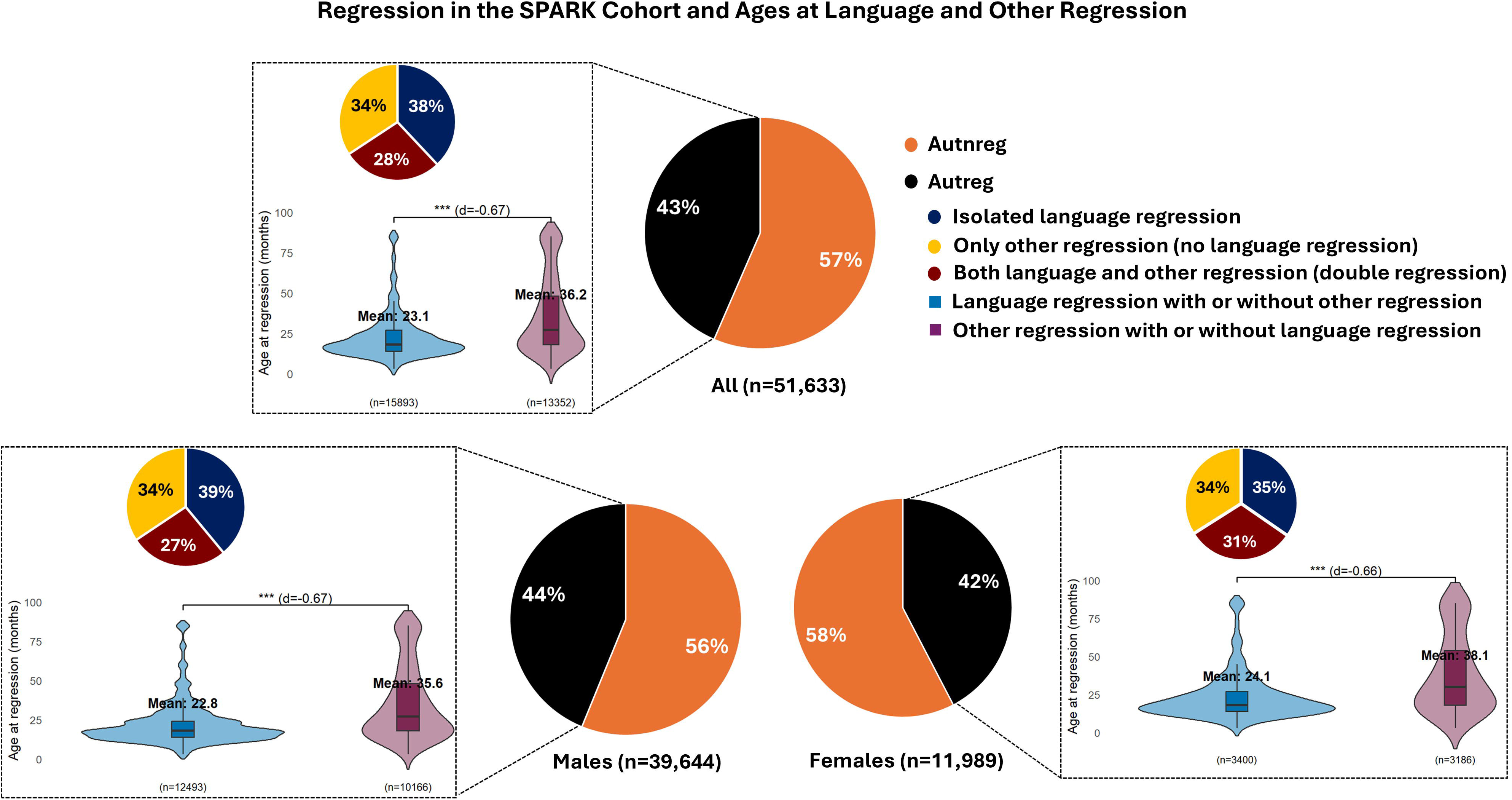
Regression in SPARK cohort and ages at regression. In SPARK cohort 29,183 individuals had autism without regression (Autnreg), 22,450 individuals had autistic regression (Autreg). Among those with regression 8,532 had only language regression, 6,224 had only other regression, and 7,694 had both language and other regression. Other regression includes social, play, toileting, and/or other skills. The ratio of those with regression to those without regression can be inaccurate because we excluded individuals with no available data on regression status. The differences between the mean ages at language and other regression were statistically significant using two sample t-test and Cohen’s d indicated moderate effect sizes (further details are in Supplementary Figure8).

**Figure 2.**
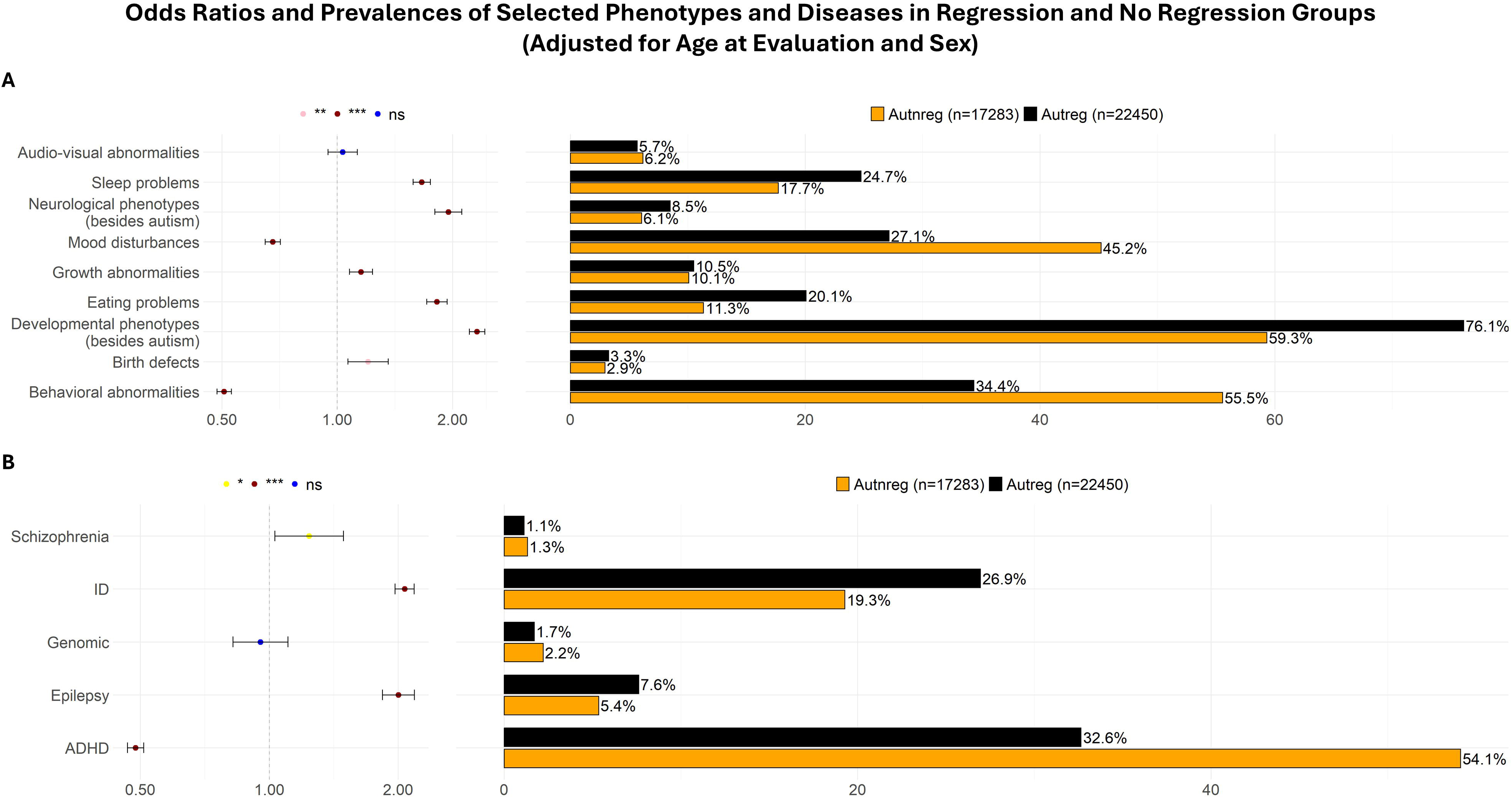
Comparing the occurrence of selected phenotypes and conditions in the autistic regression (Autreg) and autism without regression (Autnreg) groups. In the Autnreg group, we included individuals without regression who were >7.08 years old at evaluation, which was the age at which 95% of individuals in the SPARK v14 phenotype data with autistic regression developed regression. In the Autreg category we included all individuals with autistic regression (language regression, other regression, or both). Age at evaluation and sex were adjusted for in a regression model. A: selected phenotypes. B: selected diseases. ID, intellectual disability; ADHD, attention deficit hyperactivity disorder; Genomic, individuals with available genetic diagnosis of a single-gene disease or a genomic disease due to copy number variation or chromosomal abnormality; * Benjamini-Hochberg false discovery rate (FDR) < 0.05; ** < 0.01; *** < 0.001; ns, statistically non-significant.

**Figure 3.**
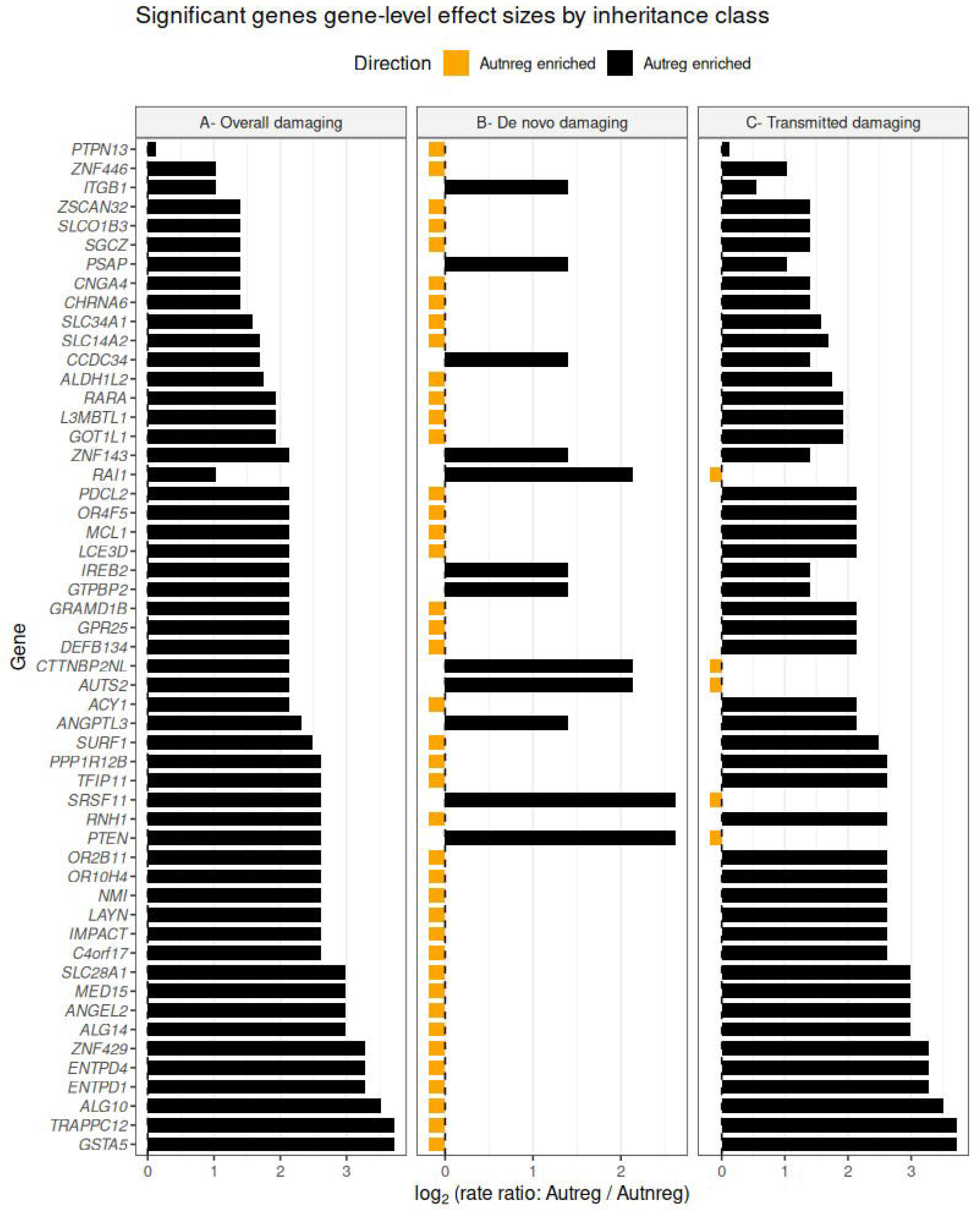
Significant autistic regression candidate genes gene-level effect sizes by inheritance class. Gene-level effect sizes were compared in autistic individuals with regression (Autreg) to those without regression (Autnreg). Bars represent log₂ rate ratios of damaging variant burden (de novo and transmitted loss-of-function and damaging missense variants combined). Effect sizes were computed using per-sample variant rates with a pseudocount to stabilize estimates for low counts. Genes are ordered by absolute effect size incrementally. Only genes surpassing false discovery rate-corrected significance in the Autreg but not the Autnreg TADA analysis (Autreg specific) are shown.

### The rare variants validation dataset supported the association between *PTEN* and Autreg

Analyzing the presence of the 53 significant candidate Autreg-specific genes in the dataset published by Zhou et al., 2022(24), we identified 6 genes (*PTEN*, *ACY1*, *AUTS2*, *RAI1*, *IREB2*, and *ITGB1*) in 12 individuals (9 with Autreg and 3 with Autnreg). Of those 6 genes, 3 were found exclusively in the Autreg group (*PTEN* in 6 and *ACY1* and *IREB2* each in one individual). *AUTS2* was detected in a single individual in each group while *RAI1* and *ITGB1* were each detected in a single individual from the Autnreg and were absent in the Autreg group (Supplementary Table 3).

Furthermore, we revisited data from individuals with a confirmed genetic diagnosis of *PTEN*-related disorder from the genetic diagnosis dataset within SPARK and found 15 additional individuals with a clinical genetic diagnosis based on *PTEN* variants, of whom 67% (10 individuals) had reported Autreg and 33% (five individuals) Autnreg (Supplementary Table 4).

### Schizophrenia PGS slightly contributed to the variance in liability to regression among autistic individuals

Comparing the means of standardized autism, ADHD, epilepsy, and schizophrenia PGS in the exploratory PGS analysis showed no statistically significant differences between Autreg and Autnreg after correcting for multiple testing (Figure 4A). Multinominal logistic regression, after adjusting for sex and age at evaluation, showed no significant difference in the occurrence of Autreg in the different PGS strata (Supplementary Table 6). However, a statistically non-significant overrepresentation of Autreg was observed in the upper decile schizophrenia PGS stratum compared with the lower strata (Figure 4B).

**Figure 4.**
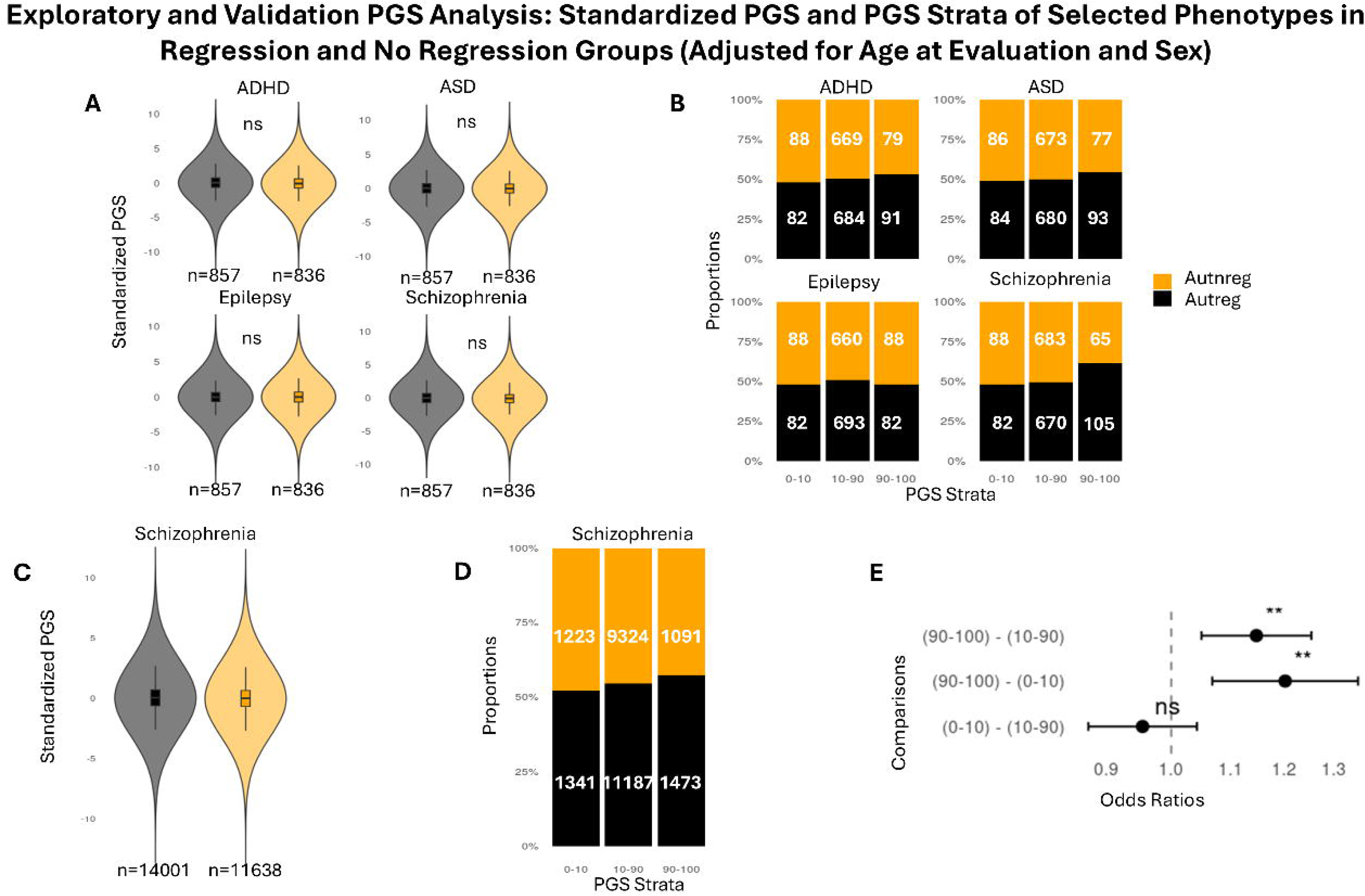
Comparing ASD, ADHD, epilepsy, and schizophrenia PGS in the regression (Autreg) and no regression (Autnreg) groups. Analysis was done using exploratory (A and B) and validation PGS datasets (C-E). A: No statistically significant differences were detected between the means of standardized ASD (nominal p-value 0.39, adjusted p-value 0.52), ADHDH (nominal p-value 0.16, adjusted p-value 0.31), epilepsy (nominal p-value 0.64, adjusted p-value 0.64), and schizophrenia PGS (nominal p-value 0.06, adjusted p-value 0.25) in the Autreg and Autnreg groups using logistic regression (equation: glm(group ∼ Standardized PGS + age at evaluation + sex)) after correcting for multiple testing using Benjamini-Hochberg false discovery rate (FDR). B: Proportions of individuals with and without regression in different PGS strata in the exploratory PGS dataset (sample sizes are inside the bars). C: Comparing schizophrenia PGS in the Autreg andAutnreg groups using the PGS validation dataset. Genomic data was obtained from the SPARK genomic data set v3. A statistically significant difference was detected between the means of standardized schizophrenia PGS (p-value 0.0002) in the regression and no regression groups using linear regression (equation: lm(Standardized PGS ∼ group + age at evaluation + sex)). D: Proportions of individuals with and without regression in different schizophrenia PGS strata (sample sizes are inside the bars) in the validation PGS dataset. E: A forest plot showing pairwise autistic regression odds ratios by schizophrenia PGS strata. The 90-100 schizophrenia PGS stratum had statistically significantly higher odds of regression compared to the middle stratum (OR 1.15, 95% CI 1.05-1.25, adjusted p-value 0.0036) and the lower stratum (OR 1.20, 95% CI 1.07-1.35, adjusted p-value 0.0036). Odds ratios were generated from logistic regression and estimated marginal means (see text). Odds ratios were plotted on a log₁₀-transformed axis. ASD, autism; ADHD, attention-deficit hyperactivity disorder.

To further explore the trend in schizophrenia PGS differences between Autreg and Autnreg, we used a dataset containing genomic data from 14 001 and 11 638 individuals with Autreg and Autnreg, respectively, and excluded individuals from the exploratory PGS analysis and their families. Logistic regression showed that higher schizophrenia PGS was significantly associated with increased odds of regression status after adjusting for age at evaluation and sex (OR = 1.056, 95% CI 1.029-1.084, p-value = 4.44 × 10^-5^) (Figure 4C).

Model comparison using a likelihood ratio test indicated that inclusion of schizophrenia PGS significantly improved model fit compared to the covariate-only model (Δdeviance = 16.68, p-value = 4.4 × 10^-5^). The incremental variance in regression status explained by schizophrenia PGS on the observed scale was small (R^2^O = 5.89 x 10^-4^). After transformation to the liability scale, schizophrenia PGS accounted for approximately 0.087% of variance in liability to regression.

Similarly, the odds of Autreg were significantly higher in the upper schizophrenia PGS decile stratum compared to the middle 10-90% stratum (OR 1.15, 95% CI 1.05-1.25, adjusted p-value 0.0036) and to the lower decile stratum (OR 1.20, 95% CI 1.07-1.35, adjusted p-value 0.0036) (Figure 4D,E).

## Discussion

Here, we analyzed the clinical and genotypic differences between autistic individuals with and without regression using one of the largest available autism research cohorts. We found that 43% of autistic individuals in the SPARK cohort, composed mainly of multiplex families, had Autreg. This is higher than the 30% pooled prevalence estimated in the two most recent meta-analyses(1, 3). However, we believe it is likely driven by differences in Autreg definition. We defined Autreg as language and/or other regression. Other regressions included regression in social, play, toileting, and other nonlinguistic developmental skills. A similar approach was undertaken before in simplex families, and Autreg prevalence was found to be 37%(8). In the two most recent meta-analysis, Autreg pooled prevalence was found to be 40% and 38.1% when Autreg was defined as language and/or social regression, and 20% and 24.9% when only language regression was considered(1, 3). However, we detected a prevalence of language regression of 31.4%.

We found that language regression started earlier than other regression, with an average age of 23.1 and 36.2 months, respectively. In simplex families, Goin-Kochel, et al., 2014, found that the age at language and other regression was 20.2 and 20.7 months, respectively(8). One possibility for this difference was that we had a wide range of ages at evaluation (1-77 years) (Supplementary Figure 7) while Goin-Kochel, et al., 2014, had a narrower age range of 4-18 years. However, the mean ages at evaluation in the two studies were comparable, 9.4 years in our study and 8.9 years in Goin-Kochel, et al., 2014(8). Other studies also showed no differences between age at language and other regression, but they considered only children(44, 45). Another factor could be the difference in the ability to retrospectively specify the age at onset of language and other regressions. One study showed significant forward telescoping in age at social but not at language regression(44). This telescoping difference could be, as put by Goin-Kochel et al., 2014 in a different context, due to the ease of detecting words loss compared to losses in less discrete skills such as social engagement(8). In line with this, and unlike language regression, we detected notable differences in age at other regression between individuals with isolated and double regression (45.7 and 28.6 months, respectively). This may indicate greater attention to other regression in individuals with language regression. However, the SPARK cohort we used mainly consisted of multiplex families. Thus, if it was an issue of increased attention, the age at other regression in our study would be expected to be lower than in simplex families(46). Also, previous studies found no differences in reporting regression onset between simplex and multiplex families(46). The reason for the late age at other regression in our study was unclear and may have been due to one or more of the above factors. The late age at other regression should be validated in future studies.

We found significant differences between Autreg and Autnreg and within the Autreg group in developmental milestones and quantitative developmental measures. All those differences had a small/moderate effect size, and we postulate that many were driven by the large sample size. This was comparable to previous studies that used large datasets(8, 9, 47). We also found that the OR of epilepsy and ID were approximately doubled in Autreg. A meta-analysis reported an epilepsy OR of 1.59 (95% CI 1.31-1.95) in Autreg(48). Other studies did not find an association between Autreg and epilepsy(49, 50). A higher prevalence of ID in Autreg has been noted before(8, 51). We expected differences in obtaining a genetic/genomic diagnosis between those with Autreg and Autnreg but none was found. However, differences can exist in specific genetic diagnoses such as *PTEN*-related disorders.

We highlighted 53 possible Autreg associated genes. Links between autism in general and some of those genes have been established/suggested. Some of the remaining have functions unrelated to brain development and are likely false positives (Supplementary Table 3). Six of the 53 genes were identified to harbor ultra-rare protein-altering DNVs in the discovery cohort published by Zhou et al.(24) with *ACY1*, *IREB2*, and *PTEN* exclusively identified in the Autreg group. This low overlap of only 6 genes could be due to the discovery cohort published by Zhou et al. only contained ultra-rare (< 1 × 10^-5^) DNVs(24). In our study, we had a more relaxed frequency filter (< 1 × 10^-3^) and we investigated both DNV and inherited variants. We included only individuals from the discovery cohort who were within our specified age thresholds. *PTEN* stood out because it was identified in 6 individuals from the discovery cohort. It is known to cause a dominant NDD with macrocephaly(52, 53). However, *PTEN* has not been linked specifically to Autreg or any regressive NDD(2, 54, 55). Similarly, while pathogenic variants in *ACY1*(56) and *IREB2*(57) cause recessive neurometabolic/neurodegenerative phenotypes, they were not linked exclusively to Autreg either in recessive or dominant states. More natural history studies and closer phenotyping in *PTEN*-related disorders and *ACY1, and IREB2* carriers are needed to understand their possible association with Autreg.

We detected a statistically significant association between Autreg and NTPDase-mediated breakdown of nucleotides pathway driven by *ENTPD1* and *ENTPD4,* a pathway implicated in synapse formation during development(58) and the regulation of extracellular purinergic signaling(59–61). Interestingly, dysregulation of extracellular ATP signaling has been implicated in autism and has been proposed to contribute to altered neurodevelopmental homeostasis through persistent activation of stress-response pathways, consistent with the cell danger response framework(62). Although the role of NTPDase signaling in Autreg remains unclear, altered regulation of extracellular nucleotide metabolism could represent a biologically plausible mechanism contributing to the destabilization of developing neural networks following an initial period of typical development(59, 63).

Previous studies have suggested other pathways and biological mechanisms such as postsynaptic density and mitochondrial functions to be implicated in Autreg(4, 22, 64). We did not detect statistically significant associations between Autreg and genes involved in those specific pathways and biological mechanisms, possibly due to our hypothesis-free approach. However, we noted trends of associations, though not statistically significant, between Autreg and transcription-related processes (Supplementary Table 5). The association between transcription genes and neuroregression has been pointed out before(4, 65). The biological significance of many of the genes we identified in Autreg is unclear and only 14/53 genes contributed to the outcome of our enrichment analysis. A recent study showed dysregulation of sphingolipid metabolism pathways in Autreg using untargeted metabolomic approach(66). Probably, such approaches that consider other omics layers and multi-omic analysis can give better resolution of the pathways involved in Autreg.

We compared autism, ADHD, epilepsy, and schizophrenia PGS in the Autreg and Autnreg groups. Autism was the primary phenotype, and it was intuitive to measure its PGS. ADHD PGS was selected due to its frequent co-occurrence with autism (67). We also measured epilepsy and schizophrenia PGS as both disorders can cause, though rarely, developmental regression in children(68–70), were included in the SPARK phenotypic dataset, and had available large GWAS. Despite epileptic encephalopathy being the epilepsy subtype associated with regression, we utilized an “all epilepsy” GWAS as a base GWAS due to its higher sample size. A previous study showed that “all epilepsy” PGS was comparable to epileptic encephalopathy PGS in patients with epileptic encephalopathy(71). A recent study compared PGS derived from 18 phenotypes, including the phenotypes we measured in this study, in Autreg and Autnreg and did not identify statistically significant differences between the two groups(47). We reached a similar result except for a previously unreported weak association between schizophrenia PGS and Autreg. One study reported a family history of neuropsychiatric disorders, including schizophrenia, as a risk factor for Autreg(18). However, a family history of schizophrenia in that study was reported only in 2/34 Autreg individuals compared to 3/136 Autnreg individuals(18). On the other hand, a recent study compared schizophrenia PGS in 211 Autreg and 719 Autnreg individuals and did not detect statistically significant differences(47). The discordance between our results and those of Tonekaboni et al. is likely due to our larger sample size and the low effect size of schizophrenia PGS in Autreg. We had also used stricter age cutoffs to define Autreg and Autnreg groups. Further studies are required to validate our findings and investigate the biological significance, if any, of this small difference in schizophrenia PGS between Autreg and Autnreg.

Although we could not detect any major polygenic or monogenic differences between Autreg and Autnreg, we highlighted some possible candidates, especially indicating that more studies should focus on investigating possible regression in *PTEN* cases. Also, further investigation of how the schizophrenia PGS is associated with regressive phenotype needs further investigation. Although we found very modest association, it is consistent with cross-trait polygenic associations observed in psychiatric genetics(72, 73) and suggested the presence of a subtle shared common variant architecture between schizophrenia genetic liability and Autreg. Environmental factors could contribute to a proportion of the remaining Autreg risk(4). Further studies are needed to explore the phenotypic and genetic differences we found between Autreg and Autnreg in depth as well as the role of environmental factors in Autreg.

Our study had several limitations. The SPARK cohort is mainly composed of multiplex families, and this could limit the generalizability of our findings. Also, the information was collected retrospectively which could lead to recall bias that could afflict the accuracy of the results. We restricted our PGS analysis to individuals of European ancestry, and this also limits the generalizability of our results. However, we were limited by the scarcity of data from other populations.

In conclusion, we highlight that regression is an important phenotype within autism, and is associated with epilepsy and ID. However, for genetic discoveries, much larger sample sizes are needed. Our study highlighted *PTEN*, NTPDase-mediated breakdown of nucleotides pathway, and schizophrenia PGS as possible contributors to Autreg.

Supplementary information is available at MP’s website.

## Data Availability

Phenotypic and genetic data for the SPARK cohort can be accessed by applying at SFARI ([https://base.sfari.org] (https:/base.sfari.org)).

## Acknowledgements

We thank the SPARK cohort’s participants and the Simons Foundation for access for the data. Computations resources were provided by the National Academic Infrastructure for Supercomputing in Sweden (NAISS) and the Swedish National Infrastructure for Computing (SNIC) at Uppsala University partially funded by the Swedish Research Council through grants no. 2022-06725 and no. 2018-05973. Open access funding provided by Karolinska Institutet. This work was supported by grants from the Swedish Foundation for Strategic Research (FFL18-0104), The Swedish Brain Foundation (FO2021-0073 and F02023-0186), the Irstadska stiftelsen, Strategic Research Area in Neuroscience at Karolinska Institutet (StratNeuro), and to Y.Z the China Scholarship Council and the Sällskapet Barnavård.

