## Supplementary Table 1 for "Comparing the Phenotypic and Genetic Differences Between Autistic Regression and Autism Without Regression Using the SPARK Cohort"

**Supplementary Table 1. TADA results. Gene-level Bayes factors (BFs), Bayesian false discovery rates (q-values), empirical P-values, and empirical null-distribution characteristics are shown separately for the regression (Autreg) and no-regression (Autnreg) cohorts. The ratio of the Autreg to Autnreg BF and the corresponding log10-transformed BF contrast were used to assess case-specificity. Boolean indicators identify genes with strong Autreg evidence, neutral or weak Autnreg evidence, and those meeting the predefined criteria for a specific candidate.**

| Gene | Autreg_BF | Autreg_qval | Autreg_pval | Autreg_null_median | Autreg_null_q95 | Autreg_null_q99 | Autnreg_BF | Autnreg_qval | Autnreg_pval | Autnreg_null_median | Autnreg_null_q95 | Autnreg_null_q99 | Autnreg_Control_BF_ratio | Autreg_Autnreg_log10BF | Autreg_strong_signal | Autreg_neutral_or_weak | Autreg_specific_candidate |
|---|---|---|---|---|---|---|---|---|---|---|---|---|---|---|---|---|---|
| CD34 | 2.35E+291 | 0 | 0.0032 | 2.5465E+249 | 1.2275E+275 | 2.9416E+286 | 1.429E+223 | 0 | 0.33175 | 4.4576E+216 | 1.6384E+241 | 1.463E+251 | 1.6461E+68 | 68.21645582 | TRUE | FALSE | FALSE |
| COMT | 9.22E+251 | 0 | 0.3844 | 6.0165E+248 | 2.9796E+274 | 7.5691E+285 | 4.692E+181 | 0 | 0.6584 | 9.4775E+186 | 3.9671E+208 | 1.8032E+218 | 1.96409E+70 | 70.2931613 | TRUE | FALSE | FALSE |
| POLR1B | 6.87E+218 | 0 | 0.1604 | 9.063E+204 | 1.5209E+228 | 3.9724E+237 | 3.666E+142 | 0 | 0.66705 | 1.2144E+147 | 8.8637E+166 | 2.0123E+175 | 1.87307E+76 | 76.27255402 | TRUE | FALSE | FALSE |
| SULT1C3 | 2.68E+202 | 0 | 0.46075 | 0.588067007 | 1.0616E+306 | 4.7669E+307 | 3.151E+169 | 0 | 1 | 1.0189E+232 | 7.4724E+256 | 4.3866E+268 | 8.51306E+32 | 32.93008571 | TRUE | FALSE | FALSE |
| ST3GAL3 | 9.6E+190 | 0 | 0.000025 | 2.5353E+130 | 1.3523E+148 | 4.3211E+155 | 4.076E+132 | 0 | 0.01235 | 3.6206E+110 | 4.4014E+126 | 1.2922E+134 | 2.35628E+58 | 58.3722267 | TRUE | FALSE | FALSE |
| OR11G2 | 6.07E+186 | 0 | 0.00015 | 1.3269E+147 | 1.6332E+165 | 2.5016E+174 | 4.14E+143 | 0 | 0.0003 | 3.1423E+109 | 3.649E+125 | 1.0506E+133 | 1.46633E+43 | 43.16623278 | TRUE | FALSE | FALSE |
| CHRNE | 5.18E+173 | 0 | 0.6527 | 2.5193E+178 | 3.5977E+200 | 1.5283E+210 | 6.487E+150 | 0 | 0.25325 | 1.8402E+143 | 1.0611E+163 | 1.9653E+172 | 7.9917E+22 | 22.90263934 | TRUE | FALSE | FALSE |
| USP48 | 1.31E+166 | 0 | 0.98175 | 1.1725E+194 | 2.1043E+216 | 4.0558E+227 | 1.512E+180 | 0 | 0.69975 | 2.7316E+186 | 4.249E+208 | 1.8745E+218 | 8.69403E-15 | -14.06077897 | TRUE | FALSE | FALSE |
| UBE2D3 | 7.56E+150 | 0 | 1 | 5.7553E+206 | 8.1214E+230 | 4.9989E+240 | 1.565E+130 | 0 | 0.99975 | 8.9448E+173 | 7.064E+195 | 2.3658E+205 | 4.83248E+20 | 20.68417024 | TRUE | FALSE | FALSE |
| PCDHGA10 | 3.03E+150 | 0 | 0.754 | 4.3286E+159 | 2.392E+180 | 1.6107E+189 | 2.058E+135 | 0 | 0.45175 | 6.7481E+133 | 6.7007E+151 | 2.3383E+160 | 1.47264E+15 | 15.16809681 | TRUE | FALSE | FALSE |
| OR51B5 | 3.113E+96 | 0 | 0.94355 | 2.1243E+113 | 1.4738E+129 | 3.0912E+136 | 7.492E+120 | 0 | 0.0078 | 1.39242E+98 | 4.7898E+113 | 7.4917E+120 | 4.15462E-25 | -24.38146877 | TRUE | FALSE | FALSE |
| EIF3E | 2.208E+95 | 0 | 1 | 1.2043E+150 | 5.8067E+169 | 1.1261E+178 | 2.2043E+95 | 0 | 1 | 2.4175E+147 | 3.7711E+165 | 6.5154E+174 | 1.001546783 | 0.00067124 | TRUE | FALSE | FALSE |
| OR2T35 | 6.148E+94 | 0 | 0.000025 | 1.02998E+40 | 1.50184E+48 | 9.15871E+52 | 3.8004E+70 | 0 | 0.000025 | 5.69509E+31 | 1.24436E+40 | 5.64769E+43 | 1.61777E+24 | 24.20891555 | TRUE | FALSE | FALSE |
| PAX6 | 2.851E+91 | 0 | 0.99055 | 1.87E+115 | 1.5166E+131 | 3.402E+138 | 2.8231E+83 | 0 | 0.9504 | 3.04755E+97 | 2.5268E+113 | 2.0083E+119 | 101002706.6 | 8.004333012 | TRUE | FALSE | FALSE |
| KATNAL2 | 9.304E+74 | 0 | 0.0316 | 7.8854E+62 | 4.07701E+73 | 1.17001E+79 | 5.1985E+64 | 0 | 0.1002 | 3.38182E+55 | 1.11342E+66 | 2.59342E+71 | 17897271765 | 10.25278683 | TRUE | FALSE | FALSE |
| ADAMTSL1 | 1.645E+67 | 0 | 0.041 | 6.76391E+55 | 9.4164E+66 | 4.29011E+71 | 5.4036E+56 | 0 | 0.0399 | 3.0029E+46 | 1.09945E+56 | 1.09646E+60 | 30445660665 | 10.4835254 | TRUE | FALSE | FALSE |
| TNRC18 | 7.154E+66 | 0 | 0.6073 | 3.3455E+69 | 5.6003E+81 | 1.93094E+87 | 2.602E+58 | 0 | 0.84585 | 1.0017E+65 | 1.36276E+77 | 4.32504E+82 | 274944741.5 | 8.439245418 | TRUE | FALSE | FALSE |
| CAMKK2 | 1.946E+66 | 0 | 0.961 | 7.07836E+79 | 6.42654E+93 | 3.1243E+99 | 3.6082E+64 | 0 | 0.9608 | 3.02211E+79 | 2.94404E+93 | 1.4752E+99 | 53.92287543 | 1.731773043 | TRUE | FALSE | FALSE |
| CWC27 | 2.537E+62 | 0 | 0.99025 | 5.32424E+80 | 1.95742E+94 | 6.6151E+99 | 1.7207E+46 | 0 | 0.99325 | 1.04267E+64 | 9.11865E+75 | 4.25386E+80 | 1.47461E+16 | 16.16867684 | TRUE | FALSE | FALSE |
| ADPRHL1 | 2.994E+48 | 0 | 0.08925 | 1.72987E+42 | 1.02165E+51 | 7.04382E+54 | 4.645E+37 | 0 | 0.5933 | 7.90423E+38 | 4.77373E+47 | 3.36162E+51 | 64462991176 | 10.80931045 | TRUE | FALSE | FALSE |
| OLFM1 | 1.023E+46 | 0 | 0.67015 | 3.6008E+48 | 4.28852E+57 | 3.94377E+61 | 5.0868E+49 | 0 | 0.3324 | 1.41744E+47 | 3.13079E+57 | 2.86611E+61 | 0.000201048 | -3.696701026 | TRUE | FALSE | FALSE |
| NLRP13 | 4.041E+39 | 0 | 0.37145 | 1.69264E+37 | 4.4242E+45 | 4.55856E+49 | 1.3534E+20 | 0 | 0.6109 | 1.85269E+21 | 1.12677E+27 | 3.98375E+30 | 2.98599E+19 | 19.47508872 | TRUE | FALSE | FALSE |
| ZNF749 | 2.701E+35 | 0 | 0.2691 | 7.35162E+31 | 1.85549E+40 | 5.26401E+42 | 2.7793E+28 | 0 | 0.1365 | 9.40137E+24 | 6.17451E+30 | 2.28004E+34 | 9717265.855 | 6.987544085 | TRUE | FALSE | FALSE |
| ABHD16B | 8.359E+32 | 0 | 0.3083 | 3.37132E+29 | 5.15239E+37 | 2.32245E+41 | 1.6503E+16 | 6.2628E-17 | 0.68315 | 2.84143E+18 | 1.49206E+24 | 3.24673E+26 | 5.06512E+16 | 16.70458957 | TRUE | FALSE | FALSE |
| CHST15 | 1.943E+32 | 0 | 1 | 3.59006E+98 | 3.0098E+114 | 2.331E+120 | 2.9012E+35 | 0 | 1 | 4.97296E+76 | 3.38327E+90 | 5.68461E+94 | 0.000669647 | -3.174153874 | TRUE | FALSE | FALSE |
| FBP2 | 9.659E+26 | 0 | 0.26275 | 3.34462E+25 | 1.04555E+30 | 1.58064E+32 | 1.3135E+28 | 0 | 0.6627 | 5.31338E+28 | 1.55815E+34 | 1.0398E+36 | 0.073538239 | -1.133486775 | TRUE | FALSE | FALSE |
| OR6K2 | 1.156E+26 | 0 | 0.05965 | 2.87745E+21 | 1.70329E+27 | 3.87346E+29 | 555824940 | 3.72547E-09 | 0.96905 | 2.03388E+14 | 5.23442E+18 | 9.35051E+20 | 2.07966E+17 | 17.31799206 | TRUE | FALSE | FALSE |
| PTCHD3 | 1.563E+25 | 0 | 0.02595 | 2.49296E+18 | 1.56298E+24 | 3.26388E+27 | 6.1762E+11 | 1.17279E-12 | 0.64075 | 1.06825E+12 | 3.25418E+17 | 5.42921E+19 | 2.53002E+13 | 13.40312344 | TRUE | FALSE | FALSE |
| MEOX2 | 3.638E+24 | 0 | 0.78895 | 1.26953E+28 | 2.32704E+35 | 6.88198E+37 | 4.5702E+21 | 0 | 0.80125 | 2.13623E+26 | 2.09006E+32 | 9.80424E+35 | 796.0552314 | 2.900943201 | TRUE | FALSE | FALSE |
| ZNF506 | 1.24E+23 | 0 | 0.01405 | 2.14316E+15 | 6.8991E+20 | 1.24015E+23 | 21876616 | 1.10474E-07 | 0.2151 | 200024.8094 | 236366124.3 | 2608333714 | 5.66886E+15 | 15.75349554 | TRUE | FALSE | FALSE |
| TMEM134 | 9.789E+21 | 0 | 0.1226 | 3.18905E+17 | 1.35223E+23 | 2.71101E+25 | 517375003 | 4.00994E-09 | 0.59065 | 517375002.7 | 9.29532E+12 | 1.13122E+14 | 1.89197E+13 | 13.27691488 | TRUE | FALSE | FALSE |
| FOXN4 | 2.667E+21 | 0 | 0.2082 | 4.05827E+18 | 2.20804E+24 | 4.88159E+26 | 2922171489 | 5.17236E-10 | 0.83735 | 4.18111E+11 | 1.08318E+16 | 1.94984E+18 | 9.12689E+11 | 11.96032299 | TRUE | FALSE | FALSE |
| C1orf116 | 5.453E+20 | 0 | 0.38465 | 5.45267E+20 | 2.97561E+26 | 6.5619E+28 | 5.2106E+18 | 0 | 0.3122 | 4.00275E+17 | 1.78062E+23 | 3.63902E+25 | 104.645426 | 2.01972025 | TRUE | FALSE | FALSE |
| SPHK1 | 1.723E+19 | 0 | 0.3841 | 1.72314E+19 | 8.865E+24 | 1.91469E+27 | 2.4524E+11 | 3.51181E-12 | 0.41165 | 20917034963 | 4.45451E+14 | 5.64895E+15 | 70264402.9 | 7.84673536 | TRUE | FALSE | FALSE |
| ZNF229 | 1.524E+19 | 0 | 0.19255 | 6.48047E+15 | 2.94537E+21 | 6.09771E+23 | 4.6075E+12 | 2.15854E-13 | 0.7699 | 9.27771E+15 | 3.06132E+20 | 6.18118E+22 | 3306834.95 | 6.519412519 | TRUE | FALSE | FALSE |
| UBE2I | 1.435E+18 | 0 | 0.30915 | 1.08079E+17 | 5.32697E+22 | 1.13694E+25 | 62544153.3 | 2.37996E-08 | 0.3942 | 5643743.812 | 8201119972 | 96947861441 | 22944337374 | 10.36067552 | TRUE | FALSE | FALSE |
| DCHS2 | 1.428E+18 | 0 | 0.68915 | 1.88041E+20 | 8.8862E+25 | 7.01996E+28 | 6.6097E+18 | 0 | 0.10225 | 4.53389E+15 | 1.05414E+20 | 4.40542E+22 | 0.215999971 | -0.665546307 | TRUE | FALSE | FALSE |
| MYL6 | 1.312E+18 | 0 | 0.31285 | 9.94444E+16 | 4.73361E+22 | 9.95442E+24 | 2.1847E+13 | 4.91486E-14 | 0.56945 | 2.18473E+13 | 2.34176E+18 | 4.27742E+20 | 60034.68036 | 4.778402203 | TRUE | FALSE | FALSE |
| SPATA33 | 3.163E+17 | 0 | 0.0284 | 1.17093E+12 | 3.1632E+17 | 5.3529E+19 | 777.39351 | 0.002901362 | 0.99875 | 1007769533 | 1.31885E+13 | 1.596E+14 | 4.06898E+14 | 14.60948521 | TRUE | FALSE | FALSE |
| CLEC11A | 2.253E+17 | 0 | 0.50305 | 2.90435E+18 | 9.55139E+22 | 2.65929E+25 | 8.3256E+10 | 2.70018E-11 | 0.26465 | 7302524731 | 1.39085E+14 | 1.71537E+15 | 2705606.57 | 6.432264645 | TRUE | FALSE | FALSE |
| OSCP1 | 1.811E+17 | 0 | 0.5034 | 1.81114E+17 | 5.55912E+21 | 1.07856E+24 | 2.1672E+19 | 0 | 0.5591 | 2.92174E+20 | 1.67228E+26 | 5.76645E+29 | 0.008357171 | -2.077940713 | TRUE | FALSE | FALSE |
| FAM111A | 1.104E+17 | 5.2868E-18 | 0.0439 | 3.72656E+11 | 8.56684E+15 | 1.44924E+18 | 1.3139E+11 | 1.06365E-11 | 0.4287 | 16740192875 | 2.31927E+14 | 3.72166E+16 | 840568.9494 | 5.924573343 | TRUE | FALSE | FALSE |
| GCKR | 6.138E+16 | 1.291E-17 | 0.0127 | 2.01863E+11 | 4.73417E+15 | 8.10089E+17 | 633148529 | 2.7001E-09 | 0.49945 | 633148528.9 | 1.6089E+14 | 2.05217E+15 | 96936550.05 | 7.986487559 | TRUE | FALSE | FALSE |
| APOBEC1 | 1.322E+16 | 4.7941E-17 | 0.68695 | 1.67601E+17 | 7.14638E+22 | 1.43631E+25 | 1.8147E+17 | 0 | 0.13895 | 8.97971E+13 | 3.17849E+19 | 5.97234E+21 | 0.072860271 | -1.137509215 | TRUE | FALSE | FALSE |
| MMP10 | 1.047E+16 | 8.6351E-17 | 0.093 | 4.55984E+11 | 1.04695E+16 | 1.77019E+18 | 23460.3534 | 7.63892E-05 | 0.96795 | 26056918.87 | 4.09333E+11 | 4.85252E+12 | 4.46263E+11 | 11.64959068 | TRUE | FALSE | FALSE |
| NTN3 | 6.362E+15 | 1.4964E-16 | 0.96125 | 1.51405E+23 | 9.70185E+29 | 4.03363E+32 | 582359936 | 3.4583E-09 | 0.96395 | 1.87243E+14 | 9.41241E+19 | 2.01832E+22 | 10924778.4 | 7.038412636 | TRUE | FALSE | FALSE |
| ORAI1 | 4.082E+15 | 2.433E-16 | 0.5689 | 4.08185E+15 | 1.70863E+21 | 3.42202E+23 | 2407241051 | 7.19752E-10 | 0.58825 | 2407241051 | 4.79289E+13 | 5.98303E+14 | 1695655.782 | 6.229337695 | TRUE | FALSE | FALSE |
| SNAPC1 | 9.877E+14 | 6.4069E-16 | 0.01875 | 49869152347 | 8.08781E+13 | 1.22892E+16 | 9.9441E+12 | 1.08381E-13 | 0.06965 | 6320956035 | 1.20301E+14 | 1.4834E+15 | 99.32249126 | 1.997047604 | TRUE | FALSE | FALSE |
| FAM221A | 8.222E+14 | 1.1012E-15 | 0.49665 | 6.65084E+13 | 2.31291E+19 | 4.31823E+21 | 2.861E+14 | 4.75842E-15 | 0.04715 | 13481274341 | 2.86097E+14 | 4.68022E+16 | 2.873775806 | 0.458452884 | TRUE | FALSE | FALSE |
| PRDM2 | 6.14E+14 | 1.6986E-15 | 1 | 5.34839E+36 | 2.53037E+45 | 1.61306E+49 | 5.4815E+16 | 9.00181E-18 | 1 | 8.85459E+56 | 2.97897E+67 | 1.21776E+72 | 0.011200604 | -1.950758559 | TRUE | FALSE | FALSE |
| SVOPL | 4.02E+14 | 2.5905E-15 | 0.00635 | 89029829.18 | 2.75142E+12 | 3.29415E+13 | 170516.065 | 1.18613E-05 | 0.29415 | 170516.0646 | 19380033.8 | 2156445778 | 2357416503 | 9.37243632 | TRUE | FALSE | FALSE |
| HDDC3 | 3.975E+14 | 3.4588E-15 | 0.0065 | 1671979276 | 3.21273E+13 | 3.97464E+14 | 2.3419E+10 | 7.15368E-11 | 0.41745 | 23418909738 | 4.98766E+14 | 6.32516E+15 | 16971.94685 | 4.229731663 | TRUE | FALSE | FALSE |
| SSH2 | 1.137E+14 | 6.5461E-15 | 0.87355 | 7.31514E+17 | 3.56031E+23 | 2.79436E+25 | 2.7411E+15 | 4.71845E-16 | 0.05705 | 1.42439E+12 | 3.5411E+16 | 4.65933E+17 | 0.041493203 | -1.382023039 | TRUE | FALSE | FALSE |
| SYT15 | 3.233E+13 | 1.7305E-14 | 0.3492 | 2.70075E+12 | 6.21193E+16 | 1.0511E+19 | 3005431482 | 4.52184E-10 | 0.7261 | 4.45419E+11 | 9.8695E+15 | 1.64243E+18 | 10758.65026 | 4.03175779 | TRUE | FALSE | FALSE |
| AUNIP | 1.076E+13 | 4.9086E-14 | 0.1523 | 41218909002 | 1.5725E+15 | 5.52651E+16 | 1.7131E+21 | 0 | 0.2135 | 6.99435E+17 | 3.38732E+23 | 7.14804E+25 | 6.28311E-09 | -8.201825582 | TRUE | FALSE | FALSE |
| CCDC66 | 1.011E+13 | 8.1762E-14 | 0.87365 | 5.08602E+16 | 2.64942E+21 | 1.76123E+24 | 1.1132E+15 | 1.00885E-15 | 0.05525 | 21068766379 | 1.11318E+15 | 1.4261E+17 | 0.009084265 | -2.041710225 | TRUE | FALSE | FALSE |
| TRAF3IP2 | 1.001E+13 | 1.1364E-13 | 0.50235 | 1.25595E+14 | 3.75022E+18 | 7.20718E+20 | 2.8887E+10 | 6.25151E-11 | 0.20635 | 2464027313 | 5.24585E+13 | 8.59426E+15 | 346.3892058 | 2.53956435 | TRUE | FALSE | FALSE |
| SLC13A1 | 9.183E+12 | 1.4736E-13 | 0.1642 | 70430971981 | 5.39235E+14 | 7.84643E+16 | 149.690274 | 0.018933279 | 0.9829 | 138332.1181 | 1336641252 | 1703933782 | 61348824752 | 10.78780625 | TRUE | FALSE | FALSE |
| PPP1R3D | 8.692E+12 | 1.8191E-13 | 0.0058 | 102719091.6 | 1.82565E+11 | 8.69194E+12 | 1761358010 | 1.19557E-09 | 0.6599 | 80909079379 | 4.309E+14 | 2.18557E+16 | 4934.796047 | 3.693269208 | TRUE | FALSE | FALSE |
| GPR142 | 8.094E+12 | 2.18E-13 | 0.1602 | 40320344059 | 1.18269E+15 | 1.86245E+17 | 2.2187E+15 | 6.51331E-16 | 0.4325 | 1.80091E+14 | 7.27285E+18 | 8.27858E+20 | 0.003647979 | -2.437947724 | TRUE | FALSE | FALSE |
| NEURL2 | 7.666E+12 | 2.5506E-13 | 0.9861 | 4.50313E+19 | 2.17836E+26 | 4.80561E+28 | 3.0384E+14 | 3.5006E-15 | 0.1003 | 1.30192E+11 | 1.21583E+15 | 1.54637E+17 | 0.02523192 | -1.598049705 | TRUE | FALSE | FALSE |
| ZNF100 | 3.944E+12 | 3.2864E-13 | 0.02235 | 24879412.52 | 3.45631E+11 | 3.944E+12 | 70.9056852 | 0.040413329 | 0.92805 | 6417.075415 | 6560678.578 | 69053267.26 | 55623151796 | 10.74525559 | TRUE | FALSE | FALSE |
| ZC3H8 | 3.145E+12 | 4.1932E-13 | 0.82295 | 4.76071E+14 | 1.86395E+20 | 2.58392E+21 | 5.2731E+13 | 1.50094E-14 | 0.64115 | 6.57654E+14 | 2.624E+20 | 5.16772E+22 | 0.059642477 | -1.224444327 | TRUE | FALSE | FALSE |
| NAT9 | 3.096E+12 | 5.0867E-13 | 0.1458 | 22323993385 | 4.64673E+14 | 5.85917E+15 | 869143.65 | 1.84408E-06 | 0.03665 | 8210.745926 | 869143.6503 | 9258872.117 | 3561738.065 | 6.551661978 | TRUE | FALSE | FALSE |
| RNF212 | 3.049E+12 | 5.9671E-13 | 0.27325 | 22522103440 | 4.46254E+14 | 7.03861E+16 | 2.9437E+10 | 5.51666E-11 | 0.32775 | 397285991.2 | 4.07692E+12 | 6.10416E+14 | 103.5865273 | 2.015303274 | TRUE | FALSE | FALSE |
| ABCC6 | 1.7E+12 | 7.5705E-13 | 0.4976 | 1.28732E+12 | 3.40823E+16 | 2.64084E+18 | 41226.2026 | 4.73968E-05 | 0.81795 | 5859820.848 | 24220306892 | 7.38199E+11 | 41225890.24 | 7.615170043 | TRUE | FALSE | FALSE |

| | | | | | | | | | | | | | | | | | |
|---|---|---|---|---|---|---|---|---|---|---|---|---|---|---|---|---|---|
| GMPPB | 1.407E+12 | 9.4736E-13 | 0.43555 | 1.40658E+12 | 3.63817E+16 | 6.53177E+18 | 1.501E+12 | 6.95517E-13 | 0.81225 | 2.89032E+15 | 3.58356E+20 | 7.00926E+22 | 0.937095664 | -0.028216072 | TRUE | FALSE | FALSE |
| CLCA4 | 7.15E+11 | 1.3242E-12 | 0.0041 | 2029153.645 | 5862500350 | 93875451327 | 3289.94097 | 0.000647031 | 0.3107 | 344.9068495 | 128532.6153 | 1086461.001 | 217340592.2 | 8.337140846 | TRUE | FALSE | FALSE |
| SLC6A13 | 7.095E+11 | 1.6931E-12 | 0.4934 | 5.0445E+11 | 1.44687E+16 | 9.28409E+17 | 4.8984E+10 | 3.16886E-11 | 0.27205 | 1634211606 | 2.65516E+13 | 1.2032E+15 | 14.48411604 | 1.160891995 | TRUE | FALSE | FALSE |
| KDM1A | 6.999E+11 | 2.0567E-12 | 0.91415 | 2.22296E+17 | 7.76906E+21 | 1.60847E+24 | 975832550 | 1.87903E-09 | 0.4985 | 153228268.1 | 1.6072E+12 | 5.42981E+13 | 717.2179773 | 2.855651167 | TRUE | FALSE | FALSE |
| MLKL | 6.411E+11 | 2.4452E-12 | 0.6607 | 1.6205E+13 | 1.72268E+18 | 9.19558E+19 | 4.4911E+10 | 4.13589E-11 | 0.223 | 359819095.8 | 2.27353E+12 | 2.28601E+14 | 14.27516411 | 1.15458111 | TRUE | FALSE | FALSE |
| CWH43 | 5.227E+11 | 2.9161E-12 | 0.4247 | 1.32644E+11 | 9.48765E+14 | 1.54851E+17 | 3.7814E+15 | 3.20157E-16 | 0.3664 | 2.98609E+14 | 8.56464E+18 | 1.61263E+21 | 0.000138216 | -3.859441528 | TRUE | FALSE | FALSE |
| GSG1 | 4.968E+11 | 3.4001E-12 | 0.65045 | 9.7237E+12 | 2.23655E+17 | 2.88366E+18 | 6245175006 | 1.28827E-10 | 0.6596 | 11129604124 | 2.31075E+14 | 3.57329E+16 | 79.55037948 | 1.900642256 | TRUE | FALSE | FALSE |
| TNFRSF10C | 4.144E+11 | 3.9737E-12 | 0.34155 | 35239559248 | 6.08437E+13 | 9.6492E+15 | 1058161.29 | 1.41078E-06 | 0.6987 | 1058161.292 | 15809716422 | 1.85304E+11 | 391644.4307 | 5.592891955 | TRUE | FALSE | FALSE |
| PYGM | 3.379E+11 | 4.6703E-12 | 1 | 7.21896E+29 | 1.79392E+37 | 6.24577E+39 | 125217563 | 1.15433E-08 | 1 | 4.69939E+30 | 1.88186E+38 | 1.8691E+41 | 2698.894596 | 3.431185924 | TRUE | FALSE | FALSE |
| COBL | 3.11E+11 | 5.4128E-12 | 0.1371 | 249823319.9 | 3.65438E+12 | 4.37855E+13 | 3034964228 | 3.87682E-10 | 0.169 | 41348157.74 | 34647131040 | 4.03628E+11 | 102.4676442 | 2.010586751 | TRUE | FALSE | FALSE |
| DNAH12 | 2.629E+11 | 6.281E-12 | 0.65165 | 1.05182E+12 | 9.01399E+16 | 4.57843E+18 | 1.2708E+11 | 1.44873E-11 | 0.0257 | 18034512.03 | 19300774435 | 1.27076E+11 | 2.068891927 | 0.315737805 | TRUE | FALSE | FALSE |
| ADARB2 | 2.158E+11 | 7.3295E-12 | 0.0848 | 831004442 | 2.15766E+11 | 3.48194E+12 | 1379370820 | 1.71308E-09 | 0.5036 | 1379370820 | 3.58332E+11 | 5.78412E+12 | 156.4235573 | 2.194302158 | TRUE | FALSE | FALSE |
| GPR162 | 2.025E+11 | 8.4242E-12 | 0.052 | 135459954.9 | 3.41361E+11 | 2.93201E+13 | 166681440 | 9.49993E-09 | 0.0228 | 163224.5149 | 166681439.9 | 1892368191 | 1215.041712 | 3.084591187 | TRUE | FALSE | FALSE |
| ART5 | 1.992E+11 | 9.511E-12 | 0.1345 | 1493170451 | 2.88245E+13 | 3.57033E+14 | 3.9094E+13 | 2.38949E-14 | 0.0105 | 1976085219 | 3.1985E+12 | 4.87223E+14 | 0.005096232 | -2.292750793 | TRUE | FALSE | FALSE |
| TEX15 | 1.729E+11 | 1.075E-11 | 0.0475 | 17501196.14 | 1.72922E+11 | 1.92384E+13 | 874670.811 | 1.75184E-06 | 0.7965 | 81861626.71 | 9.08416E+11 | 1.74861E+13 | 197699.9765 | 5.296006618 | TRUE | FALSE | FALSE |
| TRIM6 | 1.572E+11 | 1.2093E-11 | 0.12175 | 104945390.2 | 1.87368E+12 | 2.27831E+13 | 153132.55 | 1.30626E-05 | 0.256 | 1443.673007 | 1632970.551 | 16732272.91 | 1026428.485 | 6.011328696 | TRUE | FALSE | FALSE |
| FBXW8 | 7.108E+10 | 1.5168E-11 | 0.6434 | 9.83427E+12 | 2.37282E+17 | 4.11613E+19 | 181249.999 | 1.11186E-05 | 0.86955 | 211617880.6 | 1.32289E+12 | 1.59877E+13 | 392169.3294 | 5.593473626 | TRUE | FALSE | FALSE |
| RHBDL1 | 6.493E+10 | 1.8471E-11 | 0.4218 | 5538490509 | 1.17936E+14 | 1.49583E+15 | 1.3089E+11 | 1.25583E-11 | 0.01595 | 7394215.658 | 10996630441 | 1.30888E+11 | 0.496043692 | -0.304480069 | TRUE | FALSE | FALSE |
| M1AP | 4.702E+10 | 2.3008E-11 | 0.0516 | 50983827.44 | 2.11529E+11 | 6.37814E+12 | 80892333.7 | 2.17707E-08 | 0.4034 | 7473489.102 | 5346903160 | 7.12475E+11 | 581.2432937 | 2.764357955 | TRUE | FALSE | FALSE |
| LENG9 | 4.605E+10 | 2.7538E-11 | 0.00515 | 3396992.476 | 4380891534 | 46049882179 | 1.2535E+11 | 1.63913E-11 | 0.027 | 51369064.04 | 1.2535E+11 | 1.45688E+12 | 0.367371187 | -0.434894908 | TRUE | FALSE | FALSE |
| TBP | 3.774E+10 | 3.3009E-11 | 0.331 | 37738305896 | 6.73945E+13 | 1.09399E+16 | 8.1717E+15 | 2.08828E-16 | 0.1742 | 5.04306E+13 | 1.42719E+18 | 2.67933E+20 | 4.61815E-06 | -5.335532336 | TRUE | FALSE | FALSE |
| MOK | 3.743E+10 | 3.8402E-11 | 0.14875 | 311222938.8 | 4.87457E+12 | 2.70955E+13 | 809044526 | 2.25726E-09 | 0.10855 | 679235.8641 | 809044526 | 1.00898E+11 | 46.26335649 | 1.665237138 | TRUE | FALSE | FALSE |
| DUXA | 3.215E+10 | 4.4611E-11 | 0.42055 | 32147682622 | 7.19792E+14 | 1.20693E+17 | 604036840 | 3.20018E-09 | 0.5017 | 6935150070 | 1.18596E+13 | 1.87019E+15 | 53.22139395 | 1.726086245 | TRUE | FALSE | FALSE |
| ZNF528 | 1.971E+10 | 5.4828E-11 | 0.6541 | 2.20677E+11 | 6.79718E+15 | 1.3449E+17 | 44739006.1 | 3.53863E-08 | 0.58745 | 44739006.15 | 6.74936E+11 | 6.3856E+12 | 440.4728378 | 2.643919132 | TRUE | FALSE | FALSE |
| MICU2 | 1.719E+10 | 6.6375E-11 | 0.342 | 17185948606 | 4.2151E+13 | 6.39489E+15 | 3.419E+10 | 4.77883E-11 | 0.1341 | 271206374.7 | 8.35245E+12 | 1.00958E+14 | 0.502654185 | -0.298730697 | TRUE | FALSE | FALSE |
| WDR36 | 1.683E+10 | 7.7921E-11 | 0.8836 | 4.38391E+12 | 4.61196E+15 | 7.47223E+16 | 1.6441E+11 | 6.71667E-12 | 0.1276 | 632253727.3 | 6.6063E+11 | 1.06743E+13 | 0.102389853 | -0.989743079 | TRUE | FALSE | FALSE |
| HKDC1 | 1.681E+10 | 8.9237E-11 | 0.08635 | 16238996.79 | 44480330103 | 1.08606E+12 | 56085293.6 | 3.00804E-08 | 0.1083 | 871385.9114 | 901878173.5 | 14517051844 | 299.7168739 | 2.476711194 | TRUE | FALSE | FALSE |
| TRIM31 | 9.508E+09 | 1.0955E-10 | 0.66175 | 1.10786E+11 | 1.97515E+14 | 3.19651E+16 | 124502109 | 1.26202E-08 | 0.59565 | 124502108.9 | 3.34333E+13 | 4.15953E+14 | 76.37199384 | 1.882934129 | TRUE | FALSE | FALSE |
| CORO7 | 7.45E+09 | 1.3524E-10 | 0.02175 | 2778453.375 | 1858888317 | 19810020122 | 1849345095 | 9.05576E-10 | 0.37175 | 461558745.2 | 8.56799E+11 | 1.37722E+13 | 4.028400544 | 0.605132646 | TRUE | FALSE | FALSE |
| LPO | 6.878E+09 | 1.626E-10 | 0.0142 | 36848.67137 | 269686501.5 | 8357368355 | 17.3594406 | 0.146668007 | 0.366 | 4.335315677 | 2667.298537 | 32174.82383 | 396227212.4 | 8.597944299 | TRUE | FALSE | FALSE |
| TRIM61 | 5.203E+09 | 1.9857E-10 | 0.1524 | 39733737.21 | 61471218390 | 7.41345E+11 | 40.2801497 | 0.067923171 | 0.9792 | 4059.310174 | 4913657.763 | 54813154.3 | 129179666.4 | 8.111194159 | TRUE | FALSE | FALSE |
| THAP8 | 4.706E+09 | 2.3775E-10 | 0.1714 | 2737373.245 | 54894053309 | 6.53442E+11 | 3923.07808 | 0.000478718 | 0.9394 | 4638450.599 | 7134889296 | 83781634927 | 1199491.076 | 6.078997021 | TRUE | FALSE | FALSE |
| TKTL2 | 3.944E+09 | 2.8401E-10 | 0.3823 | 986079889.1 | 1.85549E+13 | 8.88707E+14 | 28729249.3 | 7.98724E-08 | 0.44165 | 18998560.19 | 36349721218 | 6.84587E+11 | 137.2725662 | 2.137583753 | TRUE | FALSE | FALSE |
| HS3ST6 | 3.171E+09 | 3.4109E-10 | 0.52775 | 6291957311 | 4.21733E+13 | 8.367E+14 | 1543310.86 | 1.03152E-06 | 0.65765 | 12928454.5 | 19344655062 | 3.10198E+11 | 2054.724088 | 3.312753512 | TRUE | FALSE | FALSE |
| CPA4 | 2.575E+09 | 4.1075E-10 | 0.3659 | 2575349691 | 3.43192E+11 | 4.9518E+13 | 712918.313 | 2.4469E-06 | 0.1364 | 69250.68687 | 14147105.15 | 886200586.3 | 3612.405019 | 3.557796437 | TRUE | FALSE | FALSE |
| TTC39A | 2.499E+09 | 4.8127E-10 | 1 | 7.10395E+29 | 2.00081E+38 | 5.88653E+40 | 1.0434E+14 | 8.23742E-15 | 0.99325 | 1.30991E+24 | 1.49318E+31 | 3.79352E+33 | 2.39496E-05 | -4.620702221 | TRUE | FALSE | FALSE |
| ZNF660 | 2.429E+09 | 5.5253E-10 | 0.827 | 3.94737E+12 | 9.91417E+16 | 1.7554E+19 | 109314590 | 1.59493E-08 | 0.5933 | 109314589.9 | 2.35276E+12 | 2.87079E+13 | 22.22266891 | 1.346796216 | TRUE | FALSE | FALSE |
| TTLL10 | 2.413E+09 | 6.2292E-10 | 0.4217 | 2413267820 | 5.57144E+13 | 6.79861E+14 | 4.6999E+10 | 3.64652E-11 | 0.3358 | 4156715131 | 8.64441E+12 | 1.28723E+15 | 0.051347143 | -1.289483718 | TRUE | FALSE | FALSE |
| HBZ | 1.799E+09 | 7.1757E-10 | 0.9942 | 9.32638E+16 | 3.7155E+22 | 7.90998E+24 | 32438055.5 | 5.36793E-08 | 0.97085 | 9.27244E+12 | 2.28982E+17 | 4.03167E+19 | 55.46103946 | 1.743988005 | TRUE | FALSE | FALSE |
| RBM43 | 1.79E+09 | 8.1094E-10 | 0.0718 | 98429.89773 | 20862580251 | 2.48119E+11 | 163.824622 | 0.016369934 | 0.37615 | 163.824622 | 17070.65324 | 180427.1577 | 10926267.13 | 7.038471814 | TRUE | FALSE | FALSE |
| SDCBP2 | 1.651E+09 | 9.1089E-10 | 0.5897 | 18880906372 | 3.94935E+14 | 4.98618E+15 | 61127487.7 | 2.58498E-08 | 0.6747 | 303721854 | 1.03933E+12 | 3.83293E+13 | 27.01648924 | 1.431628912 | TRUE | FALSE | FALSE |
| SPNS3 | 1.63E+09 | 1.0103E-09 | 0.2438 | 53384140.74 | 1.28676E+11 | 2.183E+12 | 247004213 | 7.36938E-09 | 0.50435 | 247004213.3 | 3.89199E+11 | 1.37372E+13 | 6.601075071 | 0.819614672 | TRUE | FALSE | FALSE |
| RORC | 1.491E+09 | 1.118E-09 | 0.02935 | 611924.5028 | 301647153.5 | 10140185858 | 96276.7766 | 2.11866E-05 | 0.25825 | 9111.581122 | 1040614.552 | 11503356.78 | 15485.72691 | 4.189931596 | TRUE | FALSE | FALSE |
| ZNF677 | 1.315E+09 | 1.2392E-09 | 0.78115 | 1.55707E+11 | 2.75324E+15 | 9.08951E+17 | 340856.489 | 5.38021E-06 | 0.81865 | 35774709.15 | 45031858760 | 5.06235E+11 | 3857.156755 | 3.586267289 | TRUE | FALSE | FALSE |
| NME3 | 1.219E+09 | 1.3684E-09 | 0.1684 | 7846239.476 | 1.62503E+11 | 1.93407E+12 | 5454.86435 | 0.00031755 | 0.2267 | 549.2389885 | 55498.97029 | 144558.9122 | 223549.4367 | 5.34937358 | TRUE | FALSE | FALSE |
| KRT82 | 1.158E+09 | 1.5027E-09 | 0.5392 | 1639024778 | 5.88581E+13 | 2.87099E+15 | 5008358583 | 1.70952E-10 | 0.09325 | 3788082.197 | 20022857251 | 1.03183E+12 | 0.231275914 | -0.635869594 | TRUE | FALSE | FALSE |
| ZNF543 | 1.054E+09 | 1.6489E-09 | 0.91965 | 1.89388E+14 | 1.11097E+19 | 1.85771E+21 | 8.2655E+15 | 1.57056E-16 | 0.7412 | 1.28632E+18 | 5.2425E+23 | 1.03056E+26 | 1.27482E-07 | -6.894552612 | TRUE | FALSE | FALSE |
| CTSW | 1.031E+09 | 1.7961E-09 | 0.7851 | 1.34774E+11 | 2.91473E+15 | 3.70477E+16 | 1609701340 | 1.49079E-09 | 0.3224 | 32466278.07 | 2.2149E+11 | 4.39856E+12 | 0.640580876 | -0.193426031 | TRUE | FALSE | FALSE |
| PCDHGA9 | 947844367 | 1.9548E-09 | 0.66345 | 10462461319 | 3.37096E+12 | 3.10119E+14 | 22991.472 | 8.09475E-05 | 0.15195 | 239.2818691 | 22991.47202 | 233362.9005 | 41225.91048 | 4.615170256 | TRUE | FALSE | FALSE |
| NEIL1 | 943587016 | 2.1115E-09 | 0.04125 | 483578.8149 | 517618076.6 | 15093276241 | 22672420.4 | 1.05823E-07 | 0.1483 | 137081.4531 | 239638897.5 | 2588132053 | 41.61827452 | 1.619284071 | TRUE | FALSE | FALSE |
| DCAF13 | 711850379 | 2.3216E-09 | 0.1871 | 64831390.33 | 91284218431 | 3.08463E+12 | 11767262.3 | 1.72868E-07 | 0.59145 | 11767262.3 | 3.02893E+11 | 3.59071E+12 | 60.49413712 | 1.781713286 | TRUE | FALSE | FALSE |
| MLH1 | 708682350 | 2.5291E-09 | 0.92095 | 3.43382E+12 | 3.6054E+15 | 2.34938E+17 | 1.0454E+16 | 1.05471E-16 | 0.0916 | 3.99681E+13 | 1.69321E+17 | 2.74518E+18 | 6.77909E-08 | -7.1688288 | TRUE | FALSE | FALSE |
| OCM | 701237218 | 2.7356E-09 | 0.63325 | 3898411643 | 1.075E+13 | 3.39341E+14 | 8.7003E+14 | 1.45274E-15 | 0.1711 | 2.08159E+12 | 4.30479E+16 | 2.33884E+18 | 8.05996E-07 | -6.093667193 | TRUE | FALSE | FALSE |
| HRCT1 | 642179286 | 2.9593E-09 | 0.99835 | 1.45727E+18 | 8.61238E+23 | 1.88773E+27 | 247595.367 | 7.63804E-06 | 0.9994 | 4.85988E+16 | 2.16808E+22 | 3.56671E+24 | 2593.664391 | 3.41391378 | TRUE | FALSE | FALSE |
| USP45 | 634327856 | 3.1824E-09 | 0.266 | 23654306.42 | 2.95126E+11 | 9.28331E+12 | 2652315.8 | 6.35644E-07 | 0.43855 | 663026.8685 | 2860068399 | 45802607275 | 239.1600034 | 2.378688551 | TRUE | FALSE | FALSE |
| ZNF417 | 581612124 | 3.4241E-09 | 0.66675 | 11506089279 | 7.70292E+13 | 1.2415E+16 | 827154.858 | 2.03234E-06 | 0.83375 | 97156514.25 | 1.67318E+11 | 2.00147E+12 | 703.1478058 | 2.847046626 | TRUE | FALSE | FALSE |
| CABP1 | 578295832 | 3.6634E-09 | 0.07345 | 427485.4684 | 578295832.1 | 6681349262 | 623889348 | 2.9479E-09 | 0.04525 | 459732.3475 | 623889348.5 | 85230548587 | 0.926920508 | -0.032957509 | TRUE | FALSE | FALSE |
| UBXN10 | 566036716 | 3.9045E-09 | 0.7404 | 7584362462 | 1.57694E+14 | 1.98787E+15 | 701438.566 | 2.55641E-06 | 0.7638 | 81727335.99 | 1.73555E+11 | 2.53397E+13 | 806.9654895 | 2.906854962 | TRUE | FALSE | FALSE |
| MYO1A | 561258569 | 4.1441E-09 | 0.80045 | 1.33154E+11 | 9.64449E+15 | 8.79108E+17 | 1.2149E+13 | 7.6197E-14 | 0.19265 | 28186280134 | 1.41387E+15 | 1.77181E+17 | 4.61989E-05 | -4.335368348 | TRUE | FALSE | FALSE |
| PPP1R15A | 424946144 | 4.4661E-09 | 0.87035 | 59762314103 | 1.01937E+14 | 1.59102E+16 | 17219944.7 | 1.37086E-07 | 0.09315 | 17641.06749 | 17219944.65 | 178312081.1 | 24.67755572 | 1.392302141 | TRUE | FALSE | FALSE |
| LCNL1 | 393611608 | 4.811E-09 | 0.4035 | 35292561.81 | 52201170676 | 7.51875E+12 | 364673.964 | 4.96266E-06 | 0.7089 | 364673.9643 | 501252814.3 | 67189404588 | 1079.352097 | 3.03316314 | TRUE | FALSE | FALSE |
| PCK2 | 383861335 | 5.1601E-09 | 0.7225 | 43211366178 | 6.93454E+14 | 9.73524E+16 | 8.3262E+10 | 2.4319E-11 | 0.9931 | 8.55513E+18 | 4.65195E+24 | 3.43683E+27 | 0.004610288 | -2.336271937 | TRUE | FALSE | FALSE |
| HSD3B1 | 374743306 | 5.5131E-09 | 0.00695 | 52869.65478 | 35203615.74 | 374743305.5 | 1477204.5 | 1.0884E-06 | 0.12295 | 14414.18398 | 5910012.513 | 61949006.81 | 253.6841072 | 2.40429326 | TRUE | FALSE | FALSE |
| SCN10A | 355633204 | 5.8817E-09 | 0.039 | 21400.99132 | 88662696.47 | 5725953532 | 4.89236286 | 0.311381242 | 0.9986 | 561643099.3 | 9.38135E+12 | 3.44706E+14 | 72691501.87 | 7.861483642 | TRUE | FALSE | FALSE |
| ENDOU | 347745813 | 6.2539E-09 | 0.09215 | 286171.8772 | 347745813.2 | 3873337892 | 453023.549 | 4.07436E-06 | 0.29295 | 453023.5487 | 51414485.15 | 5711340651 | 767.6108983 | 2.885141132 | TRUE | FALSE | FALSE |
| G6PC2 | 298452775 | 6.6888E-09 | 0.23405 | 6844800.728 | 13294506984 | 7.25487E+11 | 7920542.4 | 2.24353E-07 | 0.2557 | 767395.7229 | 902318909 | 17508191734 | 37.68085062 | 1.576120698 | TRUE | FALSE | FALSE |
| ADIPOQ | 280545701 | 7.1477E-09 | 0.28755 | 17428606.58 | 4520403357 | 72909339134 | 1232.33122 | 0.002038637 | 0.6031 | 4935.794441 | 80515.84559 | 1294306.523 | 227654.4629 | 5.357276169 | TRUE | FALSE | FALSE |
| MC1R | 211740143 | 7.764E-09 | 0.7405 | 9464033517 | 3.34256E+13 | 1.63287E+15 | 400581.615 | 4.58411E-06 | 0.2909 | 153798.7293 | 46468125.94 | 955358824.3 | 528.5817797 | 2.723112189 | TRUE | FALSE | FALSE |
| MMACHC | 202805438 | 8.4005E-09 | 0.01205 | 15824.16414 | 12655923.08 | 202805438 | 325.246031 | 0.007576342 | 0.0906 | 9.703553706 | 698.2577927 | 5215.998621 | 623544.6972 | 5.79486759 | TRUE | FALSE | FALSE |
| RBM44 | 152827759 | 9.2528E-09 | 0.00495 | 12181.41386 | 13973808.75 | 152827759 | 2.28536725 | 0.466014945 | 0.85485 | 207.5102316 | 20753.22276 | 214899.553 | 66872297.66 | 7.825246245 | TRUE | FALSE | FALSE |
| PHYKPL | 145093590 | 1.0141E-08 | 0.50255 | 145093589.9 | 37710328416 | 6.08856E+11 | 34308.0684 | 5.64865E-05 | 0.7191 | 865172.8473 | 224083892.4 | 3611712988 | 4229.13899 | 3.626251958 | TRUE | FALSE | FALSE |
| CCDC7 | 141158314 | 1.1043E-08 | 0.5783 | 141158313.8 | 1.94469E+12 | 2.2124E+13 | 38529565.6 | 4.70776E-08 | 0.1531 | 370569.2506 | 405634068.1 | 4359859913 | 3.663636267 | 0.56391235 | TRUE | FALSE | FALSE |
| PTGER3 | 130337722 | 1.2012E-08 | 0.93905 | 3.69443E+13 | 9.70152E+17 | 1.29925E+19 | 4.6652E+14 | 2.27133E-15 | 0.6413 | 7.64323E+16 | 2.55045E+21 | 5.16855E+23 | 2.79383E-07 | -6.55380039 | TRUE | FALSE | FALSE |

| | | | | | | | | | | | | | | | | | |
|---|---|---|---|---|---|---|---|---|---|---|---|---|---|---|---|---|---|
| TNFRSF13B | 125075004 | 1.3012E-08 | 0.59645 | 501759075.4 | 5.23287E+11 | 1.86409E+13 | 29508512.5 | 7.24403E-08 | 0.5718 | 118393633.1 | 66914589283 | 1.07958E+12 | 4.238607565 | 0.627223209 | TRUE | FALSE | FALSE |
| KCNK16 | 113346777 | 1.4108E-08 | 0.81195 | 18619791638 | 3.13134E+13 | 4.88308E+15 | 17405442.5 | 1.31335E-07 | 0.30515 | 1613296.254 | 32553575928 | 3.81824E+11 | 6.512145686 | 0.813724108 | TRUE | FALSE | FALSE |
| OR6V1 | 109226157 | 1.5234E-08 | 0.1345 | 837653.4862 | 109226157.4 | 14472911045 | 10079256.1 | 2.00526E-07 | 0.3049 | 10079256.13 | 15005620074 | 1.78666E+11 | 10.83672803 | 1.034898174 | TRUE | FALSE | FALSE |
| DPYD | 103658678 | 1.6409E-08 | 0.19845 | 2584091.91 | 6704548885 | 1.96928E+11 | 5790.46314 | 0.00029306 | 0.2772 | 1446.481145 | 371998.949 | 1490622.213 | 17901.6213 | 4.252892366 | TRUE | FALSE | FALSE |
| SYNM | 99718159 | 1.7618E-08 | 0.03 | 77837.18514 | 35909366.93 | 398942541.9 | 97895.4371 | 1.93055E-05 | 0.3825 | 24475.47247 | 7636387.825 | 55289288 | 1018.619067 | 3.008011801 | TRUE | FALSE | FALSE |
| CAPN12 | 99252826 | 1.8817E-08 | 0.6429 | 801163609.1 | 2.29141E+12 | 8.89076E+13 | 63502.966 | 3.22025E-05 | 0.42155 | 23091.91811 | 19250277.22 | 211961277.1 | 1562.963629 | 3.193948872 | TRUE | FALSE | FALSE |
| OMA1 | 98863546 | 2.0004E-08 | 0.1313 | 6137223.055 | 1594164305 | 25731338985 | 1252814.58 | 1.33274E-06 | 0.19285 | 312401.1023 | 20163227.17 | 324837471.1 | 78.9131507 | 1.897149383 | TRUE | FALSE | FALSE |
| USP54 | 85677162 | 2.1377E-08 | 0.18285 | 8012242.397 | 10433718659 | 1.18722E+11 | 190.508707 | 0.0140196 | 0.5108 | 190.5087069 | 18485.3752 | 188546.8205 | 449728.3264 | 5.652950243 | TRUE | FALSE | FALSE |
| PAH | 84044152 | 2.276E-08 | 0.0916 | 325835.9955 | 158286521.1 | 5244619156 | 3904785.64 | 4.08319E-07 | 0.79915 | 251616368.8 | 7.30165E+11 | 1.17795E+13 | 21.52337153 | 1.332910302 | TRUE | FALSE | FALSE |
| HABP2 | 77494322 | 2.4253E-08 | 0.6695 | 990207552 | 1.00316E+12 | 6.5546E+13 | 1487950511 | 1.59821E-09 | 0.18555 | 13235935.49 | 16287962951 | 2.90948E+11 | 0.052081249 | -1.283318607 | TRUE | FALSE | FALSE |
| IFIH1 | 75891997 | 2.576E-08 | 0.33385 | 7584250.694 | 8074470552 | 9.29056E+11 | 2.1924E+16 | 3.2138E-17 | 0.6759 | 3.16126E+18 | 2.13502E+24 | 3.32195E+27 | 3.46165E-09 | -8.460716783 | TRUE | FALSE | FALSE |
| PRSS57 | 66721819 | 2.7475E-08 | 0.49985 | 47462390.42 | 1.09711E+11 | 3.76018E+12 | 1964.56593 | 0.001222705 | 0.68285 | 7857.893819 | 2100930.152 | 22489811.17 | 33962.62678 | 4.531001273 | TRUE | FALSE | FALSE |
| PPP1R12B | 61258324 | 2.9335E-08 | 0.01495 | 52196.8407 | 18154181.42 | 674463880.8 | 0.14433006 | 0.865812454 | 0.96705 | 115.9488122 | 11318.30981 | 115790.2382 | 424432179 | 8.627808303 | TRUE | TRUE | TRUE |
| CD5L | 56252025 | 3.1351E-08 | 0.0746 | 47489.03524 | 56252025.27 | 621813542.1 | 7395.38391 | 0.00022961 | 0.50735 | 7395.383912 | 788494.7228 | 8423612.947 | 7606.369857 | 3.881177439 | TRUE | FALSE | FALSE |
| PELP1 | 52152492 | 3.3513E-08 | 0.15925 | 5654384.12 | 6736086230 | 78985697546 | 83.6382812 | 0.034794318 | 0.7553 | 8453.451339 | 938855.2302 | 9428689.848 | 623548.1026 | 5.794869962 | TRUE | FALSE | FALSE |
| ECM2 | 44969104 | 3.6023E-08 | 0.28195 | 4275278.888 | 21223781325 | 6.80065E+11 | 233.614696 | 0.010831254 | 0.74675 | 2278.847215 | 232619.8083 | 2431664.872 | 192492.6142 | 5.284414071 | TRUE | FALSE | FALSE |
| ATXN2 | 44290794 | 3.8542E-08 | 0.91665 | 7.9755E+11 | 1.97411E+16 | 3.47356E+18 | 733.284912 | 0.003111316 | 0.9854 | 99254151 | 1.57313E+11 | 2.36776E+13 | 60400.52525 | 4.781040715 | TRUE | FALSE | FALSE |
| IQCE | 43441479 | 4.1082E-08 | 0.89445 | 9013593968 | 9.82746E+14 | 1.48558E+17 | 1.364E+11 | 8.66634E-12 | 0.013 | 1079093.751 | 2193813611 | 1.36402E+11 | 0.000318481 | -3.496916286 | TRUE | FALSE | FALSE |
| MYBPHL | 41989919 | 4.3686E-08 | 0.0584 | 280015.0413 | 72340094.22 | 675243589.3 | 803011305 | 2.45184E-09 | 0.13065 | 9382759.627 | 3221511223 | 38201345036 | 0.05229057 | -1.281576623 | TRUE | FALSE | FALSE |
| SLC3A1 | 39824771 | 4.6412E-08 | 0.6161 | 159586728.4 | 5.96983E+11 | 5.10966E+12 | 568376.203 | 3.18671E-06 | 0.50675 | 568376.2035 | 585774161 | 9412918637 | 70.06762604 | 1.845517403 | TRUE | FALSE | FALSE |
| ATP4A | 33977562 | 4.9617E-08 | 0.62475 | 547888303.7 | 6.90385E+11 | 3.74115E+13 | 29680881.6 | 6.49733E-08 | 0.6238 | 478592360.2 | 5.01853E+11 | 9.78188E+12 | 1.14476255 | 0.058715413 | TRUE | FALSE | FALSE |
| IQCA1 | 33829094 | 5.2797E-08 | 0.74385 | 1372555625 | 6.53887E+12 | 2.23688E+14 | 40915700.5 | 4.10769E-08 | 0.1148 | 770034.85 | 274524132.7 | 4420289529 | 0.826799825 | -0.082599624 | TRUE | FALSE | FALSE |
| MPO | 32555320 | 5.6074E-08 | 0.1224 | 403513.4965 | 303585686 | 3203949444 | 176497.542 | 1.14842E-05 | 0.632 | 706520.0623 | 728918085.6 | 7031167056 | 184.4519763 | 2.265883313 | TRUE | FALSE | FALSE |
| SIX6 | 31254987 | 5.9459E-08 | 0.64625 | 503106904.1 | 1.30749E+11 | 2.11095E+12 | 969939.634 | 1.57572E-06 | 0.6616 | 969939.6339 | 309641218.8 | 4041161947 | 32.22364182 | 1.508174621 | TRUE | FALSE | FALSE |
| SLC6A18 | 29306104 | 6.305E-08 | 0.54275 | 55459892.81 | 1.28509E+11 | 2.07074E+12 | 24451613.6 | 8.80732E-08 | 0.61535 | 86651771.25 | 1.02075E+11 | 1.64684E+12 | 1.198534555 | 0.07865056 | TRUE | FALSE | FALSE |
| TRIM49 | 28654307 | 6.6686E-08 | 0.5088 | 28654306.84 | 43770189255 | 5.2564E+11 | 2888.8048 | 0.000802654 | 0.7335 | 317402.0073 | 38178554.77 | 432870375.9 | 9919.087255 | 3.996471711 | TRUE | FALSE | FALSE |
| VWA7 | 27965758 | 7.0377E-08 | 0.0206 | 3772.157253 | 2916065.501 | 68476265.57 | 189064.973 | 1.04055E-05 | 0.13835 | 2396.127712 | 1021592.389 | 16363644.29 | 147.9161241 | 2.170015518 | TRUE | FALSE | FALSE |
| IQCC | 27205969 | 7.4138E-08 | 0.0072 | 24653.24539 | 2574840.664 | 27205968.97 | 5.32019029 | 0.293349312 | 0.81505 | 49.57921528 | 473.6105342 | 4634.830007 | 5113721.028 | 6.708737032 | TRUE | FALSE | FALSE |
| DLEC1 | 24937016 | 7.8232E-08 | 0.78805 | 2539907040 | 3.32346E+13 | 4.24577E+15 | 9.7376E+10 | 1.88396E-11 | 0.2361 | 9150005402 | 1.31499E+14 | 1.76024E+16 | 0.00025609 | -3.591607187 | TRUE | FALSE | FALSE |
| PKM | 22568223 | 8.275E-08 | 0.54395 | 22568223.19 | 1.75714E+12 | 9.31534E+13 | 120042.08 | 1.64025E-05 | 0.65465 | 759404.3709 | 9418256219 | 4.68445E+11 | 188.0025996 | 2.274163854 | TRUE | FALSE | FALSE |
| BCO2 | 21702730 | 8.7413E-08 | 0.77865 | 349185049.4 | 8.47028E+11 | 2.36343E+13 | 439738.895 | 4.23947E-06 | 0.0734 | 7999.688885 | 439738.895 | 3712294.428 | 49.35367471 | 1.693319494 | TRUE | FALSE | FALSE |
| TMC7 | 19956057 | 9.247E-08 | 0.0355 | 17277.90398 | 1860237.692 | 19956056.89 | 2881320160 | 6.44246E-10 | 0.2582 | 257358832.9 | 7.84292E+12 | 9.48006E+13 | 0.006926012 | -2.159516785 | TRUE | FALSE | FALSE |
| RAPGEF3 | 19422805 | 9.762E-08 | 0.81405 | 5778108435 | 9.66468E+13 | 3.68176E+15 | 142957622 | 1.04566E-08 | 0.24475 | 3384914.888 | 16983367548 | 1.9087E+11 | 0.135864075 | -0.866895364 | TRUE | FALSE | FALSE |
| GPR150 | 17424790 | 1.0336E-07 | 0.39865 | 1608235.049 | 1828441474 | 21156876732 | 232.11258 | 0.011038907 | 0.7779 | 2315.808342 | 247681.0355 | 247681.0355 | 75070.42336 | 4.875468865 | TRUE | FALSE | FALSE |
| GPRC5D | 16968372 | 1.092E-07 | 0.99995 | 6.15609E+17 | 3.00312E+24 | 6.67872E+26 | 25260.6435 | 6.75614E-05 | 1 | 2.65629E+17 | 4.76984E+23 | 1.05244E+26 | 671.731575 | 2.827195763 | TRUE | FALSE | FALSE |
| C1S | 16730062 | 1.1507E-07 | 0.64935 | 269301331.8 | 69987107330 | 1.12994E+12 | 29213665.4 | 7.6145E-08 | 0.28145 | 7285422.452 | 1886380456 | 30399440982 | 0.572679317 | -0.242088501 | TRUE | FALSE | FALSE |
| ZRANB3 | 15739341 | 1.2127E-07 | 0.26175 | 614479.9001 | 1760692677 | 42151646653 | 2.4706E+11 | 2.35612E-12 | 0.0276 | 19080376.66 | 89715985642 | 2.77837E+12 | 6.37076E-05 | -4.195808493 | TRUE | FALSE | FALSE |
| CLIC5 | 15728277 | 1.2741E-07 | 0.06095 | 89198.01442 | 63063937.06 | 592217802.4 | 15266.9492 | 0.000135733 | 0.58965 | 15266.94918 | 2691053.659 | 43280589.62 | 1030.217398 | 3.01292888 | TRUE | FALSE | FALSE |
| ATP8B3 | 13718231 | 1.3448E-07 | 0.3019 | 858439.9978 | 6734714482 | 2.98918E+11 | 0.23527927 | 0.843270518 | 0.96015 | 323.0973585 | 306400.7855 | 8254778.325 | 58306160.83 | 7.765714446 | TRUE | TRUE | TRUE |
| MYH11 | 13019645 | 1.4188E-07 | 0.92295 | 13483560720 | 2.0401E+14 | 3.89278E+15 | 18701784.6 | 1.25585E-07 | 0.6836 | 300489010.9 | 5.10095E+11 | 8.67409E+13 | 0.696171243 | -0.15728392 | TRUE | FALSE | FALSE |
| HSPA6 | 11848001 | 1.5E-07 | 0.68045 | 1491620191 | 2.466E+12 | 3.81046E+14 | 16708029.6 | 1.48616E-07 | 0.76835 | 186416189.9 | 3.56742E+12 | 4.41372E+13 | 0.709120175 | -0.149280158 | TRUE | FALSE | FALSE |
| ATG4C | 11104926 | 1.5862E-07 | 0.049 | 11399.03184 | 7196599.936 | 115130937.6 | 39821381.7 | 4.40433E-08 | 0.5936 | 105981465.9 | 45620834535 | 5.18547E+11 | 0.27886842 | -0.554600665 | TRUE | FALSE | FALSE |
| ZNF273 | 10050284 | 1.6814E-07 | 0.97025 | 6.53938E+12 | 1.61848E+17 | 2.83539E+19 | 117.292333 | 0.025032965 | 1 | 6.34759E+18 | 2.99097E+24 | 6.24154E+26 | 85685.77389 | 4.932908723 | TRUE | FALSE | FALSE |
| TOMM20L | 9816360.6 | 1.778E-07 | 0.40015 | 9816360.56 | 11170044205 | 1.54641E+12 | 13212740.1 | 1.56113E-07 | 0.40355 | 13212740.05 | 14673927998 | 2.05514E+12 | 0.74294662 | -0.129042389 | TRUE | FALSE | FALSE |
| FGFRL1 | 9675862.6 | 1.875E-07 | 0.05505 | 7935.296312 | 9675862.607 | 108779771.9 | 1154.51423 | 0.00220434 | 0.2312 | 230.0558357 | 125973.6327 | 234920.6401 | 8380.895051 | 3.923290402 | TRUE | FALSE | FALSE |
| PCTP | 8666059.2 | 1.9834E-07 | 0.0756 | 76659.41475 | 96687974.89 | 1211422152 | 96886.6847 | 1.99345E-05 | 0.60935 | 1022012.183 | 1795406702 | 20628567681 | 89.44530671 | 1.951557558 | TRUE | FALSE | FALSE |
| MRPL32 | 8658538.5 | 2.0907E-07 | 0.5034 | 8658538.47 | 16463017252 | 1.91953E+11 | 4353572.59 | 3.84735E-07 | 0.59565 | 4353572.588 | 22844737492 | 1.07146E+12 | 1.988835214 | 0.298598801 | TRUE | FALSE | FALSE |
| ZNF701 | 7494941.9 | 2.2151E-07 | 0.05105 | 71753.53151 | 14377489.95 | 853545410 | 1586.48092 | 0.001523953 | 0.22995 | 165.3912459 | 15581.45338 | 31482.36143 | 4724.255942 | 3.674333418 | TRUE | FALSE | FALSE |
| FTCD | 6420862 | 2.3607E-07 | 0.7033 | 67012963.48 | 3.47216E+11 | 1.42402E+13 | 323041634 | 6.80847E-09 | 0.69 | 9903450326 | 1.50477E+14 | 1.19446E+16 | 0.019876268 | -1.701665164 | TRUE | FALSE | FALSE |
| BEST4 | 5467759.4 | 2.5321E-07 | 0.98135 | 5.84127E+11 | 9.75046E+15 | 6.34515E+17 | 5397.91243 | 0.000325879 | 1 | 4.50357E+18 | 1.12117E+24 | 1.22612E+26 | 1012.939618 | 3.005583558 | TRUE | FALSE | FALSE |
| KIAA0586 | 5448306 | 2.7023E-07 | 0.8787 | 69232222804 | 1.20307E+15 | 2.35114E+16 | 1794119931 | 1.0034E-09 | 0.27215 | 167202009.5 | 2.49975E+12 | 2.90241E+13 | 0.003036757 | -2.517589977 | TRUE | FALSE | FALSE |
| ACVRL1 | 5412328.7 | 2.8719E-07 | 0.14825 | 336738.3021 | 87078060.99 | 349408389.9 | 540.244926 | 0.004091211 | 0.8388 | 14364.59838 | 924779.4492 | 3708255.291 | 10018.28693 | 4.000793466 | TRUE | FALSE | FALSE |
| AQP7 | 4661688.1 | 3.0693E-07 | 0.50055 | 8543590.619 | 79735216397 | 9.61199E+11 | 52112.8153 | 3.61934E-05 | 0.72305 | 554270.9438 | 67145679.11 | 764007345.1 | 89.45377525 | 1.951598674 | TRUE | FALSE | FALSE |
| CITED4 | 4555696.4 | 3.2694E-07 | 0.70945 | 32009715.19 | 19016809597 | 3.06806E+11 | 16020.1382 | 0.000119799 | 0.67835 | 64198.47911 | 16597467.9 | 66595022.34 | 284.3731025 | 2.453888516 | TRUE | FALSE | FALSE |
| S100A3 | 4383072.7 | 3.476E-07 | 0.496 | 411828.4527 | 543139182.4 | 73358774824 | 43454936.5 | 3.81577E-08 | 0.9038 | 5568570021 | 1.17351E+14 | 1.91727E+16 | 0.100864783 | -0.996260441 | TRUE | FALSE | FALSE |
| CARF | 3944273.3 | 3.7052E-07 | 0.50905 | 3944273.266 | 6589682216 | 9.26817E+11 | 437578.453 | 4.40409E-06 | 0.9434 | 12629676244 | 2.94349E+14 | 3.76315E+15 | 9.013865367 | 0.954911067 | TRUE | FALSE | FALSE |
| ABTB1 | 3673597.1 | 3.9502E-07 | 0.6705 | 38630407.09 | 39973033931 | 5.36531E+12 | 353338656 | 5.98209E-09 | 0.2462 | 8273017.602 | 3854361599 | 33043361906 | 0.010396816 | -1.983099624 | TRUE | FALSE | FALSE |
| ZNF684 | 3614204.6 | 4.197E-07 | 0.05175 | 56071.38406 | 14498390.23 | 58174822.67 | 64078.0901 | 3.13008E-05 | 0.5895 | 256860.006 | 66485524.88 | 266842710.7 | 56.40312652 | 1.751303178 | TRUE | FALSE | FALSE |
| CHIT1 | 3224980.4 | 4.4734E-07 | 0.14005 | 309557.1446 | 63861849.2 | 4154622131 | 479826075 | 4.58898E-09 | 0.2484 | 43903401.53 | 67069873401 | 1.64337E+13 | 0.006721144 | -2.17255677 | TRUE | FALSE | FALSE |
| ECHDC2 | 3082404.8 | 4.7606E-07 | 0.75285 | 226188535.9 | 1.28412E+12 | 1.01849E+14 | 2.0196E+10 | 8.18846E-11 | 0.15055 | 113539765 | 9.17579E+11 | 2.00246E+13 | 0.000152621 | -3.816386082 | TRUE | FALSE | FALSE |
| INSRR | 3036131.8 | 5.0497E-07 | 0.276 | 389997.8443 | 404543606 | 28215052389 | 195.818151 | 0.013670519 | 0.6681 | 784.1304234 | 202121.3194 | 810058.287 | 15504.85404 | 4.190467682 | TRUE | FALSE | FALSE |
| ZMYND15 | 3026038.7 | 5.337E-07 | 0.6038 | 3026038.7 | 3909111736 | 7.10767E+11 | 5190004.27 | 3.2409E-07 | 0.1386 | 492249.6216 | 102492343.7 | 6221491369 | 0.583051293 | -0.234293237 | TRUE | FALSE | FALSE |
| CELA1 | 2693583.4 | 5.6598E-07 | 0.8977 | 1602954266 | 2.39307E+12 | 3.85784E+13 | 123221654 | 1.36927E-08 | 0.83745 | 7956649016 | 8.31058E+12 | 1.34261E+14 | 0.02185966 | -1.660356607 | TRUE | FALSE | FALSE |
| CEPT1 | 2672879 | 5.982E-07 | 0.9322 | 46285612387 | 1.04359E+15 | 1.34249E+16 | 1.7288E+18 | 0 | 0.068 | 7.69862E+14 | 2.34825E+19 | 4.56773E+21 | 1.54606E-12 | -11.81077383 | TRUE | FALSE | FALSE |
| IGFLR1 | 2671109.3 | 6.3014E-07 | 0.0357 | 9670.070053 | 2671109.275 | 28874604.51 | 3888.06006 | 0.000489679 | 0.5043 | 3888.060058 | 412536.2092 | 4396513.978 | 687.0030902 | 2.836958691 | TRUE | FALSE | FALSE |
| ACTBL2 | 2588941.5 | 6.6287E-07 | 0.82995 | 372386666 | 1.57669E+12 | 5.80373E+13 | 46922.3795 | 3.9757E-05 | 0.87765 | 12141018.31 | 29742919323 | 4.80103E+11 | 55.17498419 | 1.741742217 | TRUE | FALSE | FALSE |
| DCLRE1A | 2586159.2 | 6.9531E-07 | 0.505 | 2586159.225 | 4632150666 | 1.40968E+11 | 1652.52226 | 0.001423452 | 0.85145 | 167098.3471 | 200248219.6 | 2218198656 | 1564.976933 | 3.194507941 | TRUE | FALSE | FALSE |
| SLC5A9 | 2540035.1 | 7.2809E-07 | 0.05045 | 8122.727697 | 3119264.059 | 40767030.44 | 4792.22893 | 0.000388136 | 0.25485 | 414.9699724 | 132477.9037 | 1228296.789 | 530.0320903 | 2.724302164 | TRUE | FALSE | FALSE |
| ESF1 | 2507249.9 | 7.6102E-07 | 0.3955 | 239407.9104 | 293652339.5 | 3281014317 | 38100942.3 | 5.01085E-08 | 0.3292 | 38100942.35 | 67709506850 | 7.87044E+11 | 0.065805458 | -1.181738086 | TRUE | FALSE | FALSE |
| ANGPTL3 | 2376300.9 | 7.9564E-07 | 0.000025 | 31.93346589 | 11942.0718 | 29922.81348 | 0.5566129 | 0.730133177 | 0.9783 | 81.63381385 | 7397.298856 | 26010.66255 | 4269216.317 | 6.630348161 | TRUE | TRUE | TRUE |
| ABCA12 | 2309896.3 | 8.3102E-07 | 0.72155 | 130228183.9 | 1.19582E+13 | 1.14773E+15 | 1252.8048 | 0.001957616 | 0.91635 | 5714690.306 | 1.24954E+11 | 1.63275E+13 | 1843.779897 | 3.265709075 | TRUE | FALSE | FALSE |
| DNAH14 | 2291698 | 8.6637E-07 | 0.7459 | 386174260.1 | 2.80701E+13 | 2.77874E+15 | 4838803259 | 2.1363E-10 | 0.68265 | 45714774022 | 4.45433E+15 | 4.94724E+17 | 0.000473608 | -3.324580581 | TRUE | FALSE | FALSE |
| LIME1 | 2273407.3 | 9.017E-07 | 0.0218 | 13807.09422 | 2273407.305 | 23131046.37 | 253.262713 | 0.009910835 | 0.858 | 2518.07552 | 267453.8388 | 2853975.039 | 8976.478536 | 3.953105997 | TRUE | FALSE | FALSE |

| | | | | | | | | | | | | | | | | | |
|---|---|---|---|---|---|---|---|---|---|---|---|---|---|---|---|---|---|
| PPP1R3A | 2216312.4 | 9.3772E-07 | 0.939 | 3.217E+11 | 8.09205E+15 | 1.2201E+18 | 6101326.04 | 3.06053E-07 | 0.50425 | 64375276.76 | 85691299273 | 1.6868E+12 | 0.363250933 | -0.439793262 | TRUE | FALSE | FALSE |
| PCDHB11 | 2118732 | 9.7524E-07 | 0.6836 | 22396608.3 | 2768546639 | 3.8415E+11 | 57.677467 | 0.04869461 | 0.7344 | 5462.732385 | 567393.0534 | 5978500.492 | 36734.13725 | 4.565069844 | TRUE | FALSE | FALSE |
| DDX10 | 2048628.6 | 1.0138E-06 | 0.0353 | 6709.429073 | 2048628.627 | 21841130.05 | 3476.5513 | 0.000547849 | 0.07445 | 353.0045065 | 35055.76391 | 361724.2703 | 589.2703576 | 2.770314595 | TRUE | FALSE | FALSE |
| HPSE | 2005087.7 | 1.053E-06 | 0.29325 | 193986.3017 | 84741633.36 | 2522789703 | 121829337 | 1.47623E-08 | 0.35875 | 11450782.58 | 41677697237 | 6.66903E+11 | 0.016458168 | -1.783618508 | TRUE | FALSE | FALSE |
| ZNF211 | 1818940.1 | 1.0963E-06 | 0.60185 | 1818940.091 | 2271669106 | 2.92398E+11 | 3.3501E+13 | 3.39543E-14 | 0.2138 | 2.33808E+11 | 5.18029E+15 | 6.62264E+16 | 5.42951E-08 | -7.265239593 | TRUE | FALSE | FALSE |
| EMID1 | 1710430.9 | 1.1422E-06 | 0.1933 | 15231.98565 | 18738540.76 | 287432315.7 | 3.08356062 | 0.407031757 | 0.8907 | 61.25501258 | 11839.81806 | 30655.62085 | 554693.4608 | 5.744053046 | TRUE | FALSE | FALSE |
| SLC5A10 | 1661833.8 | 1.1892E-06 | 0.86895 | 241559363.7 | 8.3814E+11 | 2.83649E+13 | 1631406414 | 1.39067E-09 | 0.0223 | 683455.3877 | 406525393.6 | 3549336084 | 0.001018651 | -2.991974581 | TRUE | FALSE | FALSE |
| PTPRH | 1656641.7 | 1.2359E-06 | 0.58295 | 1656641.726 | 17535667757 | 3.4223E+11 | 0.1569194 | 0.862767415 | 0.9902 | 934.6990816 | 805145.6241 | 7997270.681 | 10557277.96 | 7.023551956 | TRUE | TRUE | TRUE |
| MCCC2 | 1651252.9 | 1.2824E-06 | 0.08465 | 14568.1072 | 6616897.851 | 60093133.23 | 0.00922816 | 0.896951948 | 0.98845 | 1.315336541 | 79.7441355 | 336.3358652 | 178936261.2 | 8.252698359 | TRUE | TRUE | TRUE |
| LY75 | 1633501.1 | 1.329E-06 | 0.699 | 17982744.02 | 27326594991 | 3.27601E+11 | 102419740 | 1.72053E-08 | 0.08455 | 855609.1348 | 1158436119 | 13387718153 | 0.015949085 | -1.797264235 | TRUE | FALSE | FALSE |
| TIGD4 | 1586142 | 1.3768E-06 | 0.4063 | 1586142.018 | 1930813201 | 2.44321E+11 | 31.6700001 | 0.08846399 | 0.9285 | 28653.9785 | 31908304.53 | 345534328.8 | 50083.4232 | 4.699694005 | TRUE | FALSE | FALSE |
| FGG | 1452708.9 | 1.429E-06 | 0.19225 | 263794.5086 | 23360746.64 | 376034480.2 | 21656.1172 | 9.03192E-05 | 0.8182 | 811575.66 | 526014771.3 | 3444621502 | 67.08076562 | 1.826598011 | TRUE | FALSE | FALSE |
| TP53I3 | 1414247.4 | 1.4824E-06 | 0.12035 | 134517.8388 | 15207217.05 | 169651516.5 | 183489.873 | 1.0761E-05 | 0.2512 | 1755.427354 | 1942415.806 | 21030353.3 | 7.707495487 | 0.886913279 | TRUE | FALSE | FALSE |
| TRPM4 | 1076897.6 | 1.5539E-06 | 0.29435 | 67237.29751 | 152110719.3 | 2440148330 | 2688652.33 | 6.02508E-07 | 0.45715 | 1075471.729 | 4584440269 | 1.3273E+11 | 0.400534337 | -0.397360246 | TRUE | FALSE | FALSE |
| ZNF626 | 1066577.2 | 1.6255E-06 | 0.07175 | 1056.213279 | 1066577.158 | 11180207.73 | 1887.91461 | 0.001279442 | 0.27475 | 1887.914605 | 190897.736 | 1984665.523 | 564.9498948 | 2.752009932 | TRUE | FALSE | FALSE |
| FAN1 | 1052660 | 1.6976E-06 | 0.55295 | 1896228.174 | 3252936235 | 1.19098E+11 | 10.7573564 | 0.190467279 | 0.32065 | 5.658481978 | 362.8238223 | 1452.929449 | 97854.89566 | 4.990582558 | TRUE | FALSE | FALSE |
| ALPK2 | 1048854.4 | 1.7692E-06 | 0.12125 | 11085.39728 | 10534248.59 | 1130444944 | 3389355.04 | 5.09869E-07 | 0.017 | 3568.504062 | 3389355.04 | 34778588.51 | 0.30945546 | -0.50940185 | TRUE | FALSE | FALSE |
| PON3 | 941747.71 | 1.8493E-06 | 0.83735 | 18714037.66 | 2.2074E+11 | 2.62549E+12 | 2584658.24 | 6.6939E-07 | 0.0545 | 6180.345456 | 2584658.244 | 27361565.37 | 0.36436063 | -0.438468555 | TRUE | FALSE | FALSE |
| CROT | 904307.71 | 1.9322E-06 | 0.13285 | 5661.210425 | 6047417.558 | 96592825.46 | 42791.1862 | 4.60594E-05 | 0.4338 | 18094.49634 | 16439200.96 | 352177937.6 | 21.13303677 | 1.324961909 | TRUE | FALSE | FALSE |
| CATSPER2 | 864798.82 | 2.0186E-06 | 0.9887 | 1.3498E+11 | 1.51946E+15 | 8.20742E+16 | 7663420.82 | 2.3688E-07 | 0.9989 | 1.71025E+15 | 1.95628E+20 | 2.74898E+22 | 0.112847623 | -0.947507585 | TRUE | FALSE | FALSE |
| FOXS1 | 744168.7 | 2.1195E-06 | 0.07955 | 26798.20642 | 1726158.645 | 27771878.05 | 1053.24526 | 0.00232569 | 0.4031 | 1053.245265 | 16893.72111 | 83656.77061 | 706.5483454 | 2.849141884 | TRUE | FALSE | FALSE |
| VAV1 | 718043.29 | 2.2235E-06 | 0.3001 | 718043.2899 | 1007410112 | 11770697994 | 109.329973 | 0.026484168 | 0.66005 | 109.3299731 | 11373.36116 | 120614.3706 | 6567.670963 | 3.817411387 | TRUE | FALSE | FALSE |
| ATP13A5 | 670626.88 | 2.3346E-06 | 0.25655 | 17880.46156 | 62900776.66 | 1601328274 | 4742516.71 | 3.43781E-07 | 0.26335 | 193287.2438 | 1486124049 | 26083032309 | 0.141407384 | -0.849527912 | TRUE | FALSE | FALSE |
| PIWIL3 | 618480.16 | 2.4549E-06 | 0.24465 | 63414.57596 | 62747668.91 | 6917182641 | 219.216588 | 0.011674843 | 0.15745 | 24.04025019 | 2046.787845 | 19556.99949 | 2821.32008 | 3.45045236 | TRUE | FALSE | FALSE |
| SFXN2 | 587147.93 | 2.5811E-06 | 0.0567 | 5555.800709 | 6245033.386 | 11655478.86 | 7295401.83 | 2.49961E-07 | 0.506 | 7295401.834 | 4981653150 | 2.32668E+11 | 0.080481918 | -1.094301684 | TRUE | FALSE | FALSE |
| GJB4 | 584224.33 | 2.7069E-06 | 0.3922 | 240894.7619 | 246558262 | 3960491897 | 1752791144 | 1.29012E-09 | 0.11185 | 7692233.904 | 10066764809 | 1.66901E+11 | 0.000333311 | -3.477150532 | TRUE | FALSE | FALSE |
| CYP4F3 | 539753.64 | 2.8428E-06 | 0.1171 | 7570.406634 | 1269372.591 | 5086738.18 | 169.099249 | 0.015854511 | 0.49655 | 169.0992493 | 29705.88958 | 209120.8412 | 3191.933992 | 3.504053902 | TRUE | FALSE | FALSE |
| CHIA | 528877.6 | 2.9807E-06 | 0.0566 | 5095.916367 | 2115445.842 | 22297707.86 | 9.08445265 | 0.209774571 | 0.81455 | 86.01561526 | 834.8333211 | 8300.566392 | 58217.88336 | 4.765056412 | TRUE | FALSE | FALSE |
| CHEK1 | 458541.12 | 3.1402E-06 | 0.17535 | 17676.47515 | 4832519.313 | 52064393.97 | 8.2727906 | 0.222903232 | 0.50845 | 78.38871589 | 761.3752778 | 7575.812754 | 55427.62315 | 4.743726256 | TRUE | FALSE | FALSE |
| OTOS | 442980.19 | 3.3045E-06 | 0.1466 | 4075.286192 | 442980.1885 | 4777904.621 | 511.302141 | 0.004374518 | 0.06375 | 51.84923586 | 5168.234057 | 5168.234057 | 866.3765562 | 2.937706692 | TRUE | FALSE | FALSE |
| ALKBH3 | 430615.71 | 3.4724E-06 | 0.19585 | 42077.14227 | 48231386.12 | 527358649.6 | 72.5121456 | 0.03957986 | 0.8559 | 699.3964241 | 69914.18216 | 723791.0718 | 5938.532154 | 3.773679112 | TRUE | FALSE | FALSE |
| MTMR9 | 424835.68 | 3.6415E-06 | 0.14215 | 39317.36253 | 6830060.722 | 109915879.5 | 20804244.5 | 1.15339E-07 | 0.78775 | 334793297.6 | 3.49325E+11 | 5.64112E+12 | 0.020420625 | -1.689930964 | TRUE | FALSE | FALSE |
| PCSK7 | 414822.53 | 3.8136E-06 | 0.06445 | 2148.110119 | 550409.0793 | 8806719.533 | 0.70854683 | 0.662139647 | 0.9543 | 64.52190337 | 5072.316488 | 25932.72932 | 585455.345 | 5.767493775 | TRUE | TRUE | TRUE |
| ZNF99 | 407120.41 | 3.9878E-06 | 0.1416 | 40375.38602 | 44207092.84 | 475611981.9 | 0.14040802 | 0.866717971 | 0.96885 | 11.40919377 | 106.771629 | 1023.656024 | 2899552.275 | 6.462330943 | TRUE | TRUE | TRUE |
| CYP3A5 | 380579.37 | 4.1737E-06 | 0.07515 | 3690.694346 | 3995700.312 | 42866340.37 | 70.4632434 | 0.040624328 | 0.9549 | 26985.5027 | 30038571.86 | 899639942.8 | 5401.104964 | 3.732482617 | TRUE | FALSE | FALSE |
| TOP3A | 321147.85 | 4.3954E-06 | 0.0964 | 9308.087079 | 1287681.931 | 20717651.7 | 1739531.48 | 9.23032E-07 | 0.15275 | 108217.5269 | 27989815 | 207992537.6 | 0.184617441 | -0.733727273 | TRUE | FALSE | FALSE |
| C20orf144 | 314314.32 | 4.6205E-06 | 0.2542 | 30080.11513 | 3359008.834 | 38281690.51 | 35872.1568 | 5.49087E-05 | 0.3637 | 3500.197499 | 376401.594 | 4136934.216 | 8.762069124 | 0.942606675 | TRUE | FALSE | FALSE |
| KIR2DL1 | 305508.83 | 4.8508E-06 | 0.9991 | 1.54583E+15 | 6.47403E+20 | 1.28507E+22 | 47556414 | 3.26639E-08 | 0.99745 | 2.1145E+15 | 9.1174E+20 | 1.28709E+22 | 0.006424135 | -2.19218533 | TRUE | FALSE | FALSE |
| SERGEF | 295216.01 | 5.0795E-06 | 0.19875 | 7410.421858 | 3216569.137 | 34653764.29 | 141915.914 | 1.35117E-05 | 0.1187 | 5379.260385 | 1507886.768 | 16378440.13 | 2.150682044 | 0.332576209 | TRUE | FALSE | FALSE |
| DNLZ | 292699.89 | 5.3169E-06 | 0.50445 | 292699.8881 | 33722913.27 | 378225178.5 | 328277.146 | 5.59535E-06 | 0.3852 | 328277.1463 | 432599195.7 | 4966065236 | 0.891624322 | -0.049818093 | TRUE | FALSE | FALSE |
| ARMC9 | 267305.93 | 5.5768E-06 | 0.39935 | 26255.47996 | 29630849.68 | 322308190.6 | 0.07829878 | 0.880772242 | 0.51365 | 0.700831619 | 0.700831619 | 2.65107205 | 3413922.042 | 6.5332536 | TRUE | TRUE | TRUE |
| MBP | 266947.9 | 5.8351E-06 | 0.02175 | 224.5121868 | 266947.8968 | 2960608.091 | 21.7611321 | 0.124929882 | 0.98025 | 24129.30285 | 32793069.32 | 379457185.9 | 12267.1879 | 4.088745018 | TRUE | FALSE | FALSE |
| RAD1 | 253291.34 | 6.1063E-06 | 0.00875 | 234.3865607 | 24116.21493 | 46763.8476 | 31.9132376 | 0.087316351 | 0.51145 | 31.91323759 | 314.4853329 | 3174.754568 | 7936.873923 | 3.899649482 | TRUE | FALSE | FALSE |
| TFAP2E | 245765.63 | 6.3844E-06 | 0.353 | 245765.6344 | 58255291.69 | 233758934.4 | 108.446218 | 0.026977592 | 0.8921 | 1462.624583 | 6960.669992 | 94113.69497 | 2266.244399 | 3.355306744 | TRUE | FALSE | FALSE |
| SERPINI2 | 244144.64 | 6.6624E-06 | 0.83295 | 2568785.855 | 16370909161 | 1.89985E+11 | 0.06235638 | 0.884221657 | 0.99785 | 4775.604233 | 508356.11 | 5426488.776 | 3915311.465 | 6.592766316 | TRUE | TRUE | TRUE |
| TMEM25 | 241282.52 | 6.9417E-06 | 1 | 1.23266E+18 | 3.65556E+23 | 5.42089E+25 | 1.59313054 | 0.52666905 | 1 | 1.39788E+21 | 1.085E+27 | 2.50904E+29 | 151451.8226 | 5.180274504 | TRUE | FALSE | FALSE |
| TMPRSS4 | 240386.69 | 7.2201E-06 | 0.66205 | 1716563.838 | 444776498.8 | 4012059065 | 3474832.31 | 4.83465E-07 | 0.5806 | 3474832.313 | 900618976.8 | 14520901789 | 0.069179364 | -1.160023433 | TRUE | FALSE | FALSE |
| ITIH1 | 236740.18 | 7.501E-06 | 0.0101 | 157.5538332 | 59195.6255 | 340333.5558 | 1.97988468 | 0.490684401 | 0.29205 | 0.495158402 | 7.918523318 | 31.6779477 | 119572.7092 | 5.077632069 | TRUE | FALSE | FALSE |
| LIMS1 | 236509.48 | 7.78E-06 | 0.8454 | 15160674.26 | 36796716749 | 5.89224E+11 | 77480.7965 | 2.53208E-05 | 0.83125 | 3260662.757 | 4250680501 | 68069993966 | 3.05249157 | 0.484654473 | TRUE | FALSE | FALSE |
| HK3 | 229990.22 | 8.0656E-06 | 0.1783 | 6650.999086 | 8159829.893 | 130780403.2 | 0.10576372 | 0.874442328 | 0.85565 | 1.688404627 | 431.5742123 | 2115.369352 | 2174566.248 | 6.337372643 | TRUE | TRUE | TRUE |
| MS4A4A | 226959.17 | 8.3532E-06 | 0.38645 | 226959.1746 | 275046909 | 3126750203 | 32.324278 | 0.086166483 | 0.8964 | 314.9802561 | 32191.05767 | 337231.3392 | 7021.322329 | 3.846418911 | TRUE | FALSE | FALSE |
| IGLL1 | 224584.85 | 8.642E-06 | 0.87905 | 26200492.91 | 53343877463 | 7.69765E+12 | 3647994.69 | 4.57505E-07 | 0.2196 | 338315.6409 | 609707877 | 7004599884 | 0.06156392 | -1.210673735 | TRUE | FALSE | FALSE |
| AFM | 221588.52 | 8.9329E-06 | 0.44075 | 88123.21453 | 96058107.25 | 1033830756 | 430.256992 | 0.005509561 | 0.6848 | 1721.22637 | 421341.2784 | 4384590.355 | 515.0143487 | 2.711819329 | TRUE | FALSE | FALSE |
| TCHP | 220259.34 | 9.2236E-06 | 0.09465 | 2189.003104 | 220259.3445 | 2286012.891 | 42.5565444 | 0.06441313 | 0.07095 | 4.553426812 | 407.7018279 | 407.7018279 | 5175.686782 | 3.713967986 | TRUE | FALSE | FALSE |
| SYCP1 | 213367.08 | 9.5226E-06 | 1 | 4.82861E+34 | 2.00717E+43 | 6.55637E+45 | 579.605876 | 0.003760496 | 1 | 1.71971E+25 | 2.19598E+32 | 5.78771E+34 | 368.1244264 | 2.565994635 | TRUE | FALSE | FALSE |
| CYB561D2 | 207192.99 | 9.8292E-06 | 0.50225 | 2198268.82 | 236005013.7 | 31085860431 | 2981066.04 | 5.69527E-07 | 0.0548 | 26735.51378 | 33604303.19 | 33604303.19 | 0.069502987 | -1.157996532 | TRUE | FALSE | FALSE |
| TMEM54 | 202588.97 | 1.0141E-05 | 0.6084 | 2147514.61 | 2485194963 | 28789894651 | 23909219.5 | 9.68484E-08 | 0.21635 | 2613471.674 | 3033677298 | 35279019133 | 0.008473257 | -2.07194961 | TRUE | FALSE | FALSE |
| RMND5B | 201384.38 | 1.0453E-05 | 0.05055 | 837.3983177 | 201384.381 | 805668.2884 | 1327.11389 | 0.001851086 | 0.0342 | 15.07874825 | 331.7211943 | 1327.113889 | 151.7461182 | 2.18111759 | TRUE | FALSE | FALSE |
| CASP5 | 193638.45 | 1.0777E-05 | 0.9313 | 2454191310 | 5.78436E+13 | 7.05844E+14 | 7.8067E+12 | 1.48475E-13 | 0.0733 | 4060333853 | 9.43606E+13 | 1.16249E+15 | 2.48042E-08 | -7.605474475 | TRUE | FALSE | FALSE |
| POR | 192656.51 | 1.1099E-05 | 0.47085 | 103488.5348 | 51710723.84 | 1750601452 | 1109.39396 | 0.002234023 | 0.11155 | 31.74993772 | 1367.266864 | 17790.92197 | 173.659243 | 2.239697904 | TRUE | FALSE | FALSE |
| OR2A12 | 176051.89 | 1.1454E-05 | 0.5067 | 176051.8949 | 20156519.94 | 177058869.6 | 0.33887641 | 0.81235784 | 1 | 3.137566308 | 3.137566308 | 3.137566308 | 519516.5247 | 5.715599366 | TRUE | TRUE | TRUE |
| SHQ1 | 175166.99 | 1.1808E-05 | 0.2768 | 17334.96432 | 1810402.481 | 19128326.73 | 3.73976006 | 0.368934412 | 0.89325 | 139.2678366 | 12996.3232 | 64383.14876 | 46839.09857 | 4.670608528 | TRUE | FALSE | FALSE |
| GPD2 | 174863.29 | 1.216E-05 | 0.17385 | 7649.63039 | 2800471.776 | 27756645.75 | 240.142131 | 0.010316849 | 0.89055 | 22329.48835 | 14289887.8 | 146128682.5 | 728.1657926 | 2.862230273 | TRUE | FALSE | FALSE |
| ZNF479 | 172219.79 | 1.2516E-05 | 0.06405 | 1637.869894 | 172219.7861 | 172219.7861 | 23.0075246 | 0.117612673 | 0.3153 | 23.00752463 | 2226.219694 | 2226.219694 | 7485.36789 | 3.87421315 | TRUE | FALSE | FALSE |
| OR5M3 | 161419.41 | 1.2896E-05 | 0.61485 | 161419.4104 | 173795579.4 | 1971025796 | 22248.9295 | 8.56226E-05 | 0.1466 | 2182.128163 | 232256.7626 | 2480978.719 | 7.255154039 | 0.860646638 | TRUE | FALSE | FALSE |
| PNPLA7 | 156364.56 | 1.3286E-05 | 0.0956 | 609.6672199 | 346422.9664 | 5553448.182 | 2.09350421 | 0.480499753 | 0.96735 | 2146.764205 | 652814.6623 | 5901177.804 | 74690.34952 | 4.873264492 | TRUE | FALSE | FALSE |
| ZNF662 | 154737 | 1.3679E-05 | 0.2959 | 15395.2917 | 53881096.5 | 1969122576 | 261631.277 | 6.8089E-06 | 0.19635 | 25749.25389 | 28886099.91 | 313576312.8 | 0.591431562 | -0.228095503 | TRUE | FALSE | FALSE |
| RIPK3 | 151000.55 | 1.4079E-05 | 0.2944 | 29791.04954 | 30915817.98 | 1979545306 | 2973.87279 | 0.000775596 | 0.0688 | 303.3217971 | 29852.79469 | 30142.97059 | 50.77572694 | 1.70565615 | TRUE | FALSE | FALSE |
| GCAT | 148331.25 | 1.4485E-05 | 0.0953 | 348.2053049 | 711924.2483 | 14091732.76 | 4871.04082 | 0.000378982 | 0.051 | 15.30332633 | 7191.48925 | 78686.14655 | 30.45165349 | 1.483610879 | TRUE | FALSE | FALSE |
| DENND2C | 144618.8 | 1.4899E-05 | 0.26615 | 36068.09136 | 9336395.058 | 37472599.7 | 213.625142 | 0.012437787 | 0.8699 | 13740.98297 | 1965831.245 | 9662399.797 | 676.9746463 | 2.830572404 | TRUE | FALSE | FALSE |
| OXR1 | 143191.5 | 1.5315E-05 | 0.89475 | 15887120.9 | 23347672297 | 3.34869E+12 | 3.246E+12 | 3.09813E-13 | 0.43515 | 3.246E+12 | 8.11785E+16 | 1.43539E+19 | 4.41132E-08 | -7.355431216 | TRUE | FALSE | FALSE |
| DLX4 | 137731.42 | 1.5747E-05 | 0.1858 | 8585.209741 | 589351.6053 | 4674977.98 | 12487.7943 | 0.00017142 | 0.139 | 1351.413922 | 50015.52722 | 605451.7784 | 11.02928302 | 1.042547281 | TRUE | FALSE | FALSE |
| SIGLEC1 | 136091.58 | 1.6182E-05 | 0.4427 | 71925.30703 | 137231361.9 | 2207892546 | 36059.3244 | 5.33977E-05 | 0.072 | 117.1094415 | 81889.31838 | 1045276.705 | 3.774102299 | 0.576813668 | TRUE | FALSE | FALSE |

| | | | | | | | | | | | | | | | | | |
|---|---|---|---|---|---|---|---|---|---|---|---|---|---|---|---|---|---|
| PDE1A | 133617.02 | 1.6622E-05 | 0.5115 | 1420175.312 | 1805354671 | 21033695572 | 23211415.4 | 1.01303E-07 | 0.50875 | 92796947.18 | 2.43537E+11 | 2.96563E+12 | 0.005756522 | -2.239839839 | TRUE | FALSE | FALSE |
| CCDC17 | 133137.86 | 1.7061E-05 | 0.6002 | 1311265.913 | 4594331431 | 1.66632E+11 | 0.58977928 | 0.716758323 | 0.9799 | 427.8887884 | 38919.52323 | 384063.0966 | 225741.8345 | 5.35361205 | TRUE | TRUE | TRUE |
| NCL | 132920.76 | 1.7499E-05 | 0.6154 | 132920.7642 | 171674907.6 | 1968516668 | 752.965112 | 0.003026354 | 0.00145 | 2.312211267 | 22.26123492 | 45.71247378 | 176.5297781 | 2.246817975 | TRUE | FALSE | FALSE |
| PDPR | 128400.09 | 1.795E-05 | 0.9998 | 9.73379E+12 | 5.20852E+16 | 1.48996E+18 | 6841310.91 | 2.90792E-07 | 0.999 | 5.3717E+14 | 9.10066E+18 | 3.71106E+20 | 0.018768346 | -1.726573997 | TRUE | FALSE | FALSE |
| IDUA | 126796.04 | 1.8405E-05 | 0.1082 | 1982.675509 | 794987.6312 | 12716616.51 | 765.153033 | 0.002942938 | 0.43895 | 455.7389564 | 177660.9945 | 1816409.567 | 165.7133109 | 2.219357395 | TRUE | FALSE | FALSE |
| CYP39A1 | 125464.66 | 1.8862E-05 | 0.4995 | 125464.6606 | 71792608.91 | 288077088.5 | 1.77605659 | 0.510430461 | 0.9532 | 113.7622039 | 1823.481013 | 14359.54306 | 70642.2651 | 4.849064616 | TRUE | FALSE | FALSE |
| CLCN1 | 122462.83 | 1.9328E-05 | 0.87745 | 66816327.14 | 3.32446E+11 | 9.7935E+12 | 750610.929 | 2.23477E-06 | 0.81615 | 56547063.56 | 2.37424E+11 | 8.60078E+12 | 0.16315088 | -0.78741058 | TRUE | FALSE | FALSE |
| SALL1 | 122307.13 | 1.9793E-05 | 0.7936 | 1299879.629 | 1825609518 | 21338013107 | 185.151779 | 0.01473236 | 0.7782 | 1866.307689 | 204539.08 | 650528.7325 | 660.5776426 | 2.819923871 | TRUE | FALSE | FALSE |
| GDNF | 116939.98 | 2.0278E-05 | 0.5008 | 116939.9803 | 17938281.05 | 288606019.3 | 419453256 | 5.22443E-09 | 0.01395 | 1621593.523 | 104552840.9 | 1683214574 | 0.000278791 | -3.554720553 | TRUE | FALSE | FALSE |
| ALDH1L1 | 114414.45 | 2.0772E-05 | 0.73195 | 1840865.816 | 1919848946 | 68433379600 | 27671710.1 | 8.37126E-08 | 0.12575 | 353323.2489 | 199487739.4 | 1788559720 | 0.004134708 | -2.383555131 | TRUE | FALSE | FALSE |
| ATP8B4 | 113229.34 | 2.1268E-05 | 0.3418 | 28193.89686 | 118739862.3 | 1921348746 | 93243.2858 | 2.18293E-05 | 0.5391 | 93243.28582 | 429989892.6 | 13116682432 | 1.214343122 | 0.084341417 | TRUE | FALSE | FALSE |
| RERGL | 113103.64 | 2.1762E-05 | 0.2541 | 10823.25869 | 1208459.283 | 13807440.71 | 2454.03603 | 0.0009811 | 0.22695 | 250.7513707 | 24617.88463 | 63248.5616 | 46.08882421 | 1.663595629 | TRUE | FALSE | FALSE |
| OR5J2 | 111996.8 | 2.2259E-05 | 0.5037 | 1173396.331 | 137736471.2 | 24226131417 | 0.24557829 | 0.840525182 | 0.75165 | 2.255692458 | 21.26491259 | 21.26491259 | 456053.3277 | 5.659015629 | TRUE | TRUE | TRUE |
| TLR1 | 103539.28 | 2.2798E-05 | 0.60625 | 103539.2782 | 111566930.6 | 13016915184 | 0.44815329 | 0.774911277 | 0.6609 | 3.999377607 | 342.888334 | 3290.352153 | 231035.4073 | 5.363678543 | TRUE | TRUE | TRUE |
| GSTA5 | 100643.27 | 2.3351E-05 | 0.02195 | 955.4055004 | 100643.2717 | 1068737.28 | 0.02078594 | 0.893630367 | 1 | 0.188868865 | 0.188868865 | 0.188868865 | 4841891.278 | 6.685015033 | TRUE | TRUE | TRUE |
| GMPR | 99402.175 | 2.3908E-05 | 0.5116 | 99402.1747 | 38042714.36 | 385455547.1 | 160.051438 | 0.0166367 | 0.98655 | 1627873.071 | 1669093697 | 37931890158 | 621.0639279 | 2.793136306 | TRUE | FALSE | FALSE |
| DOHH | 97737.646 | 2.4473E-05 | 0.146 | 953.7443396 | 97737.64631 | 1024474.382 | 1.65551233 | 0.521460355 | 0.8747 | 1.655512334 | 149.4391383 | 149.4391383 | 59037.7035 | 4.771129456 | TRUE | FALSE | FALSE |
| TMEM145 | 95542.277 | 2.5048E-05 | 0.18175 | 9335.561505 | 10717554.4 | 33877657.76 | 0.18934491 | 0.854499566 | 1 | 16.26735962 | 156.6772478 | 156.6772478 | 504593.8573 | 5.702941959 | TRUE | TRUE | TRUE |
| AKR7A2 | 95192.964 | 2.5622E-05 | 0.38295 | 9292.185759 | 10699601.79 | 117274727.9 | 150.829079 | 0.018660743 | 0.31235 | 15.65984485 | 1489.089288 | 1489.089288 | 631.1313794 | 2.800119774 | TRUE | FALSE | FALSE |
| TMCO4 | 90272.451 | 2.6228E-05 | 0.69505 | 9781148.905 | 12956698910 | 1.4827E+11 | 182.8072 | 0.01497335 | 0.9542 | 180927.4587 | 37253149.09 | 2441430641 | 493.8123369 | 2.693561936 | TRUE | FALSE | FALSE |
| LRP8 | 89525.175 | 2.6836E-05 | 0.10795 | 535.4433445 | 651198.3328 | 10392360.34 | 0.5594875 | 0.728925589 | 0.9269 | 52.58123867 | 8452.041746 | 33792.10272 | 160012.8244 | 5.204154791 | TRUE | TRUE | TRUE |
| GCNT3 | 82821.326 | 2.7495E-05 | 0.1792 | 8141.912048 | 9178477.023 | 96182284.71 | 111103.677 | 1.7514E-05 | 0.17575 | 10854.55594 | 12466479.04 | 136522382.7 | 0.745441812 | -0.127586251 | TRUE | FALSE | FALSE |
| ZNF713 | 77725.135 | 2.82E-05 | 0.03675 | 766.0063859 | 77725.13539 | 810686.7571 | 1094.96012 | 0.002264028 | 0.3509 | 111.806018 | 10985.34588 | 112840.6301 | 70.98444396 | 1.851163185 | TRUE | FALSE | FALSE |
| OR51G1 | 71702.574 | 2.8966E-05 | 0.61775 | 71702.57365 | 86811616.74 | 1185992630 | 1.37210554 | 0.548610917 | 0.97035 | 123.0340542 | 1209.454153 | 12179.64479 | 52257.33134 | 4.718147228 | TRUE | FALSE | FALSE |
| MROH2B | 71037.337 | 2.9735E-05 | 0.41345 | 17762.04055 | 61292560.27 | 608119391.8 | 9.87174764 | 0.200315501 | 0.91185 | 774.0226394 | 635968.99 | 6218275.5 | 7196.024421 | 3.857092629 | TRUE | FALSE | FALSE |
| ALG10 | 67785.909 | 3.054E-05 | 0.01795 | 68.32625836 | 6636.913376 | 67785.90887 | 0.13714087 | 0.867348438 | 1 | 0.137140867 | 0.137140867 | 2.723123081 | 494279.4264 | 5.693972534 | TRUE | TRUE | TRUE |
| NUDT6 | 66945.128 | 3.1352E-05 | 0.14575 | 663.9209903 | 66945.12754 | 696063.931 | 98.7354459 | 0.029854298 | 0.34555 | 98.7354459 | 9646.971144 | 98816.33146 | 678.0252718 | 2.831245881 | TRUE | FALSE | FALSE |
| AAR2 | 62746.008 | 3.2219E-05 | 0.39095 | 62746.00817 | 75071120.02 | 8332606391.6 | 75486.4745 | 2.61012E-05 | 0.7769 | 8370852.285 | 5187102084 | 7.19558E+11 | 0.831221866 | -0.080283041 | TRUE | FALSE | FALSE |
| KIF16B | 61143.277 | 3.3105E-05 | 0.2936 | 5643.996159 | 6111335.916 | 97744994.31 | 356.04223 | 0.006556125 | 0.81405 | 18223.17632 | 18368960.73 | 293867483.5 | 171.7304068 | 2.234847199 | TRUE | FALSE | FALSE |
| MLNR | 60270.505 | 3.4001E-05 | 0.9495 | 72733501.99 | 2.05012E+11 | 1.74539E+13 | 11780077.2 | 1.64534E-07 | 0.19745 | 103856.7337 | 130541580.9 | 2032219142 | 0.005116308 | -2.291043309 | TRUE | FALSE | FALSE |
| UGGT2 | 53864.606 | 3.5009E-05 | 0.3257 | 22929.95683 | 20278091.85 | 518702449.7 | 2.54363483 | 0.444464515 | 0.98865 | 17220.84573 | 16242249.18 | 1734143468 | 21176.23366 | 4.32584872 | TRUE | FALSE | FALSE |
| ZCCHC4 | 52649.805 | 3.6037E-05 | 0.71205 | 543977.6948 | 113876686.8 | 3269987942 | 91837.9587 | 2.31226E-05 | 0.06175 | 900.0271533 | 91837.95868 | 178196.8112 | 0.573290233 | -0.241625457 | TRUE | FALSE | FALSE |
| DHCR7 | 50340.574 | 3.711E-05 | 0.32165 | 4954.564299 | 5565983.222 | 88905632.28 | 0.10938192 | 0.873551599 | 0.9822 | 259.4361527 | 74957.1528 | 968276.5766 | 460227.5376 | 5.662972601 | TRUE | TRUE | TRUE |
| PPP1CB | 49076.566 | 3.8207E-05 | 0.2276 | 4722.609804 | 522139.7249 | 1662916.011 | 0.69918011 | 0.666755041 | 1 | 0.699180107 | 0.699180107 | 0.699180107 | 70191.59324 | 4.8462851 | TRUE | TRUE | TRUE |
| PDE3B | 48902.282 | 3.9301E-05 | 0.18 | 1914.347124 | 511312.6254 | 5465359.6 | 11.5337971 | 0.181991273 | 0.3221 | 1.224847744 | 111.3289997 | 111.3289997 | 4239.911776 | 3.62735682 | TRUE | FALSE | FALSE |
| SLC25A30 | 42856.696 | 4.0559E-05 | 0.90405 | 10483827.04 | 6092525484 | 43857142851 | 2217.2623 | 0.001079636 | 0.30495 | 1171.81965 | 383630.9258 | 1537930.236 | 19.3286541 | 1.286201614 | TRUE | FALSE | FALSE |
| MICAL2 | 42132.197 | 4.1833E-05 | 0.51075 | 42132.19721 | 52494718.76 | 236161784.1 | 98.4257743 | 0.030027917 | 0.35435 | 10.12556146 | 980.1297586 | 9993.026079 | 428.0606124 | 2.631505268 | TRUE | FALSE | FALSE |
| TRIM58 | 41364.097 | 4.3125E-05 | 0.09865 | 4126.973047 | 424256.1391 | 4450624.489 | 0.87210298 | 0.602570798 | 0.8777 | 8.030346587 | 75.84329433 | 75.84329433 | 47430.28985 | 4.676055779 | TRUE | TRUE | TRUE |
| OPN4 | 41255.949 | 4.4412E-05 | 0.84645 | 4388453.29 | 8157163191 | 2.34406E+11 | 281.902332 | 0.008873769 | 0.99315 | 43721419.71 | 2.16934E+11 | 1.06825E+13 | 146.3483769 | 2.16538791 | TRUE | FALSE | FALSE |
| C9 | 40880 | 4.5705E-05 | 0.61935 | 163648.0611 | 152180884 | 1755474655 | 10720716.3 | 1.90955E-07 | 0.343 | 2680571.963 | 4852086657 | 77707649548 | 0.003813178 | -2.418712911 | TRUE | FALSE | FALSE |
| RASAL3 | 40773.37 | 4.6994E-05 | 0.5063 | 419032.6111 | 518244437.1 | 5801386083 | 341603265 | 6.38565E-09 | 0.17115 | 7892169.695 | 42630251882 | 5.3374E+11 | 0.000119359 | -3.923145399 | TRUE | FALSE | FALSE |
| IFNB1 | 40269.276 | 4.8292E-05 | 0.22985 | 3912.017129 | 425137.1057 | 425137.1057 | 445.505885 | 0.005180678 | 0.3467 | 445.5058849 | 46374.50574 | 492255.2356 | 90.38999792 | 1.956120376 | TRUE | FALSE | FALSE |
| RPL29 | 38644.669 | 4.9643E-05 | 0.38865 | 38644.66865 | 48490967.74 | 546719009.8 | 492.322992 | 0.004495076 | 0.50075 | 492.322992 | 4980.115389 | 51606.0666 | 78.49454379 | 1.89483947 | TRUE | FALSE | FALSE |
| HSD17B8 | 38462.869 | 5.0993E-05 | 0.32795 | 9615.897192 | 3960370.033 | 41579281.57 | 19.5132538 | 0.13509685 | 0.65885 | 46.07769546 | 1783.66812 | 17687.4455 | 1971.115095 | 3.294711984 | TRUE | FALSE | FALSE |
| DEFA6 | 38456.146 | 5.2335E-05 | 0.36085 | 3735.224797 | 406369.608 | 4383634.166 | 4164.37996 | 0.000457335 | 0.3681 | 4164.379955 | 454826.9362 | 4917314.944 | 9.23454304 | 0.96541541 | TRUE | FALSE | FALSE |
| DCC | 38037.05 | 5.3685E-05 | 0.7875 | 491234.3733 | 312880222.1 | 5033398236 | 806.20988 | 0.002819695 | 0.7464 | 7432.410947 | 345048.5372 | 4976896.472 | 47.18008395 | 1.673758709 | TRUE | FALSE | FALSE |
| CYP26C1 | 34514.648 | 5.5181E-05 | 0.5065 | 34514.64841 | 3877199.143 | 42484553.16 | 883716780 | 2.06071E-09 | 0.09275 | 486092.787 | 883716780.1 | 10149794824 | 3.90562E-05 | -4.408309648 | TRUE | FALSE | FALSE |
| SURF1 | 32560.993 | 5.6766E-05 | 0.00715 | 23.15318025 | 3430.425659 | 13727.20126 | 0.03849361 | 0.889346998 | 0.93725 | 0.153899418 | 5.474264541 | 5.474264541 | 845880.3563 | 5.92730894 | TRUE | TRUE | TRUE |
| ADPRH | 32526.245 | 5.8344E-05 | 0.9275 | 9227317.668 | 66645350018 | 2.30986E+12 | 1787551358 | 1.09966E-09 | 0.31325 | 22639521 | 2.40655E+11 | 2.87884E+12 | 1.8196E-05 | -4.740024594 | TRUE | FALSE | FALSE |
| A4GNT | 31452.283 | 5.9972E-05 | 0.49645 | 7866.133748 | 10187647.17 | 113756448.5 | 23730.9007 | 7.41343E-05 | 0.8611 | 4227512.535 | 6390704939 | 1.02255E+11 | 1.325372488 | 0.122337951 | TRUE | FALSE | FALSE |
| CRYGD | 31299.653 | 6.1599E-05 | 0.5909 | 31299.65269 | 8094125.889 | 32478663.7 | 20.3124426 | 0.131377977 | 0.8055 | 325.5165721 | 31201.48353 | 125048.9728 | 1540.910336 | 3.187777368 | TRUE | FALSE | FALSE |
| LRRC55 | 29785.186 | 6.3308E-05 | 0.01955 | 287.5189676 | 29785.18632 | 315410.444 | 3323.87399 | 0.000609807 | 0.1157 | 329.8122143 | 34315.81838 | 364042.6351 | 8.960985401 | 0.95235577 | TRUE | FALSE | FALSE |
| DCD | 29744.722 | 6.501E-05 | 0.3658 | 2916.230688 | 310620.1542 | 3319296.453 | 386890633 | 5.58712E-09 | 0.0471 | 299120.9886 | 386890633 | 51529692809 | 7.68815E-05 | -4.114178296 | TRUE | FALSE | FALSE |
| PDE5A | 29594.724 | 6.671E-05 | 0.7861 | 308587.3119 | 505455474.7 | 5771033536 | 624135.044 | 2.79661E-06 | 0.83285 | 191100520.1 | 8.03553E+11 | 1.28529E+13 | 0.047417181 | -1.324064268 | TRUE | FALSE | FALSE |
| CYP24A1 | 28494.79 | 6.8474E-05 | 0.39845 | 16050.31603 | 7377489.003 | 145757795.8 | 569.010577 | 0.003814265 | 0.1422 | 86.95790049 | 1393.224817 | 9142.241834 | 50.07778636 | 1.699645123 | TRUE | FALSE | FALSE |
| LLPH | 26842.591 | 7.0347E-05 | 0.25595 | 3959.461125 | 2989604.375 | 35149390.5 | 3456616656 | 3.2243E-10 | 0.15365 | 27171045.28 | 40249548851 | 4.78455E+11 | 7.76557E-06 | -5.109826779 | TRUE | FALSE | FALSE |
| EML3 | 26501.134 | 7.2237E-05 | 0.4043 | 26501.13447 | 30707140.28 | 336958861.7 | 44.8659021 | 0.060708455 | 0.8348 | 4288.680103 | 4777280.711 | 51787260.42 | 590.6742812 | 2.771348061 | TRUE | FALSE | FALSE |
| PEX6 | 25371.768 | 7.4208E-05 | 0.067 | 1579.718372 | 25371.76838 | 348375.3531 | 70330.6815 | 2.77673E-05 | 0.60695 | 281959.6505 | 73019107.59 | 1176818609 | 0.360749645 | -0.442794088 | TRUE | FALSE | FALSE |
| ENTPD1 | 25123.291 | 7.6189E-05 | 0.0027 | 246.2319568 | 2457.378119 | 25123.29141 | 0.38902143 | 0.796028805 | 1 | 0.389021426 | 0.389021426 | 7.721860741 | 64580.73967 | 4.810103015 | TRUE | TRUE | TRUE |
| PCDHA8 | 24779.672 | 7.8189E-05 | 0.5045 | 24779.67225 | 28394990.03 | 310157812.8 | 21480.8692 | 9.73981E-05 | 0.2334 | 2167.766984 | 871126.5874 | 23953003.43 | 1.153569348 | 0.062043708 | TRUE | FALSE | FALSE |
| OR7G1 | 24201.791 | 8.023E-05 | 0.0012 | 238.7570166 | 2374.994697 | 24201.79051 | 3.32617515 | 0.393290212 | 0.751 | 3.326175148 | 31.66386061 | 31.66386061 | 7276.162389 | 3.861902383 | TRUE | FALSE | FALSE |
| PLS1 | 24024.216 | 8.2276E-05 | 0.2831 | 5998.823863 | 96236.68213 | 243058.2473 | 6496.89395 | 0.000262892 | 0.20465 | 405.642196 | 61787.30292 | 247367.7828 | 3.697800292 | 0.567943452 | TRUE | FALSE | FALSE |
| CCDC90B | 23042.389 | 8.4407E-05 | 0.3176 | 5752.383755 | 370008.7275 | 1483258.105 | 0.8590369 | 0.605678512 | 0.80445 | 13.7543269 | 55.05670581 | 220.4395839 | 26823.51471 | 4.428515683 | TRUE | TRUE | TRUE |
| MPPE1 | 23042.338 | 8.6526E-05 | 0.9507 | 25684650.07 | 1.04324E+11 | 9.10455E+12 | 43.5047226 | 0.063611194 | 0.97065 | 419974.5207 | 519218922.6 | 5816488203 | 529.6514072 | 2.72399013 | TRUE | FALSE | FALSE |
| CCDC63 | 21815.459 | 8.8765E-05 | 0.39455 | 21815.45866 | 24126564.78 | 260673103 | 37.8988762 | 0.072632725 | 0.8911 | 35114.40169 | 3748472.702 | 40040888.57 | 575.6228375 | 2.760138016 | TRUE | FALSE | FALSE |
| PLA2G4C | 21646.078 | 9.1011E-05 | 0.2554 | 2190.450227 | 218898.5203 | 2264121.677 | 4.73518685 | 0.318145556 | 0.50695 | 4.73518685 | 44.15739445 | 44.15739445 | 4571.325018 | 3.6600421 | TRUE | FALSE | FALSE |
| ZNF846 | 21307.769 | 9.3283E-05 | 0.29715 | 21307.76857 | 4172759.169 | 231128679.3 | 556.932444 | 0.003869107 | 0.23185 | 59.28147638 | 5357.261962 | 11053.49125 | 38.25916193 | 1.582735453 | TRUE | FALSE | FALSE |
| CHRNA10 | 20897.468 | 9.5592E-05 | 0.1938 | 1809.424123 | 334992.3534 | 2319687.811 | 30.060158 | 0.091946177 | 0.547 | 68.6514265 | 4399.178805 | 17612.83165 | 695.1882232 | 2.842102406 | TRUE | FALSE | FALSE |
| OR4D11 | 20553.126 | 9.7931E-05 | 1 | 2.81736E+33 | 6.08437E+40 | 3.3896E+44 | 33.8576878 | 0.082437374 | 1 | 5.10083E+33 | 1.16648E+41 | 6.68717E+44 | 607.0445985 | 2.783220599 | TRUE | FALSE | FALSE |
| KIR2DL4 | 19955.846 | 0.00010033 | 0.6127 | 19955.84611 | 25066551.55 | 284920198.7 | 21.5588886 | 0.125844781 | 0.9564 | 241633.2563 | 27871272.97 | 3693588421 | 925.6435487 | 2.966443779 | TRUE | FALSE | FALSE |
| CAPN11 | 19375.269 | 0.0001028 | 0.12825 | 648.1672735 | 77656.93503 | 668754.458 | 2654.37538 | 0.00086051 | 0.5159 | 2654.375376 | 204887.5639 | 4660078.013 | 7.299370455 | 0.863285405 | TRUE | FALSE | FALSE |
| DCAF4L2 | 18570.16 | 0.00010538 | 0.6178 | 193062.9175 | 256058233.6 | 2914377111 | 30.7716199 | 0.090199231 | 0.13085 | 30.77161986 | 301.1225087 | 301.1225087 | 603.4833407 | 2.780665286 | TRUE | FALSE | FALSE |
| GBP5 | 18056.15 | 0.00010802 | 0.88835 | 19268657.83 | 65166092809 | 1.13486E+12 | 4023228150 | 2.64292E-10 | 0.2525 | 365982392 | 5.96265E+11 | 2.88147E+13 | 4.48798E-06 | -5.3479495 | TRUE | FALSE | FALSE |

| | | | | | | | | | | | | | | | | | |
|---|---|---|---|---|---|---|---|---|---|---|---|---|---|---|---|---|---|
| MOCOS | 17797.407 | 0.00011069 | 0.11925 | 193.0606605 | 44258.30645 | 450068.1592 | 0.04413543 | 0.888078303 | 0.5056 | 0.044135431 | 0.044135431 | 0.869170877 | 403245.3541 | 5.605569372 | TRUE | TRUE | TRUE |
| SPATA31E1 | 17626.206 | 0.00011337 | 0.7669 | 167545.386 | 163570549.5 | 1691027565 | 393336.139 | 4.76626E-06 | 0.7468 | 39233291.15 | 45616264008 | 4.93149E+11 | 0.044812069 | -1.348605004 | TRUE | FALSE | FALSE |
| FBXW9 | 17585.734 | 0.00011605 | 0.0421 | 185.2773308 | 17585.73397 | 177408.709 | 1016354.27 | 1.49155E-06 | 0.1936 | 25011.01095 | 10561299.01 | 112192276.5 | 0.01730276 | -1.761884617 | TRUE | FALSE | FALSE |
| TGFBR2 | 17554.575 | 0.00011872 | 0.13805 | 1094.429684 | 50625.63354 | 659930.4365 | 14.9404721 | 0.158879522 | 0.499 | 7.860081238 | 239.0942761 | 956.8290255 | 1174.967911 | 3.070026006 | TRUE | FALSE | FALSE |
| CHURC1 | 17412.811 | 0.00012139 | 0.03005 | 116.3855631 | 4347.896733 | 17412.81121 | 33.6468799 | 0.083004655 | 0.4956 | 8.407677811 | 134.6859469 | 539.2724686 | 517.5163722 | 2.713924094 | TRUE | FALSE | FALSE |
| SHISA5 | 16819.863 | 0.00012416 | 1 | 5.92406E+31 | 1.60016E+40 | 8.91432E+43 | 0.45038476 | 0.774245821 | 1 | 7.26095E+28 | 1.18102E+36 | 5.76913E+39 | 37345.54173 | 4.572238764 | TRUE | TRUE | TRUE |
| MGST2 | 16815.621 | 0.00012691 | 0.22895 | 1658.591318 | 174550.2761 | 698133.4196 | 835.763913 | 0.002703096 | 0.57935 | 835.7639132 | 84679.85844 | 883278.0915 | 20.12006124 | 1.303629298 | TRUE | FALSE | FALSE |
| LIPC | 16228.894 | 0.00012976 | 0.13195 | 738.9814993 | 109629.6613 | 1040864.859 | 0.36064212 | 0.805434325 | 0.9765 | 53.71780629 | 3438.918136 | 13763.8495 | 44999.99674 | 4.653212482 | TRUE | TRUE | TRUE |
| WDR90 | 16073.944 | 0.00013262 | 0.59295 | 16073.94358 | 132911788.2 | 1332397067 | 0.16978808 | 0.859759678 | 0.9901 | 940.16195 | 762827.7433 | 73678237.71 | 94670.62191 | 4.97621523 | TRUE | TRUE | TRUE |
| CARS2 | 15788.281 | 0.00013553 | 0.5015 | 31258.93173 | 18039882.29 | 626983872.5 | 2602.0304 | 0.000904668 | 0.8053 | 104895.5913 | 30794832.93 | 1357203647 | 6.067677401 | 0.783022483 | TRUE | FALSE | FALSE |
| CCDC34 | 15763.249 | 0.00013842 | 0.0175 | 48.73939799 | 4533.803171 | 45315.84885 | 0.84544575 | 0.608636534 | 0.94005 | 7.738619628 | 699.2019572 | 699.2019572 | 18644.89713 | 4.270559992 | TRUE | TRUE | TRUE |
| OR6C70 | 15721.223 | 0.00014131 | 0.25665 | 15721.22308 | 1713136.645 | 18502172 | 22.2144594 | 0.121883487 | 0.65845 | 214.7856368 | 21612.03638 | 224652.8403 | 707.7022586 | 2.849850582 | TRUE | FALSE | FALSE |
| OR51E1 | 15332.28 | 0.00014427 | 0.2312 | 1507.714079 | 159634.7295 | 159634.7295 | 19.293831 | 0.13603206 | 0.77775 | 187.5317711 | 199264.8491 | 378981.9139 | 794.6726613 | 2.900188272 | TRUE | FALSE | FALSE |
| TRMT10B | 15010.419 | 0.00014728 | 0.38985 | 15010.41913 | 16586351.66 | 179154386.2 | 247608.56 | 7.36276E-06 | 0.1738 | 2424.782636 | 2588829.083 | 27670219.49 | 0.060621568 | -1.217372836 | TRUE | FALSE | FALSE |
| FAXDC2 | 14540.091 | 0.00015038 | 0.6102 | 147869.2984 | 65401553.46 | 1972123940 | 29907167.6 | 6.11893E-08 | 0.03495 | 25767.31942 | 2783334.706 | 29907167.63 | 0.000486174 | -3.313208173 | TRUE | FALSE | FALSE |
| ZNF7 | 13573.736 | 0.00015372 | 0.5094 | 136551.7217 | 27742514.52 | 1743972575 | 218575.144 | 9.08271E-06 | 0.6072 | 218575.1443 | 261270332.1 | 2894198887 | 0.062101002 | -1.206901391 | TRUE | FALSE | FALSE |
| FCN1 | 13480.159 | 0.00015706 | 0.24415 | 1352.945688 | 215691.0003 | 2200329.203 | 25.5977274 | 0.107352621 | 0.128 | 2.738699839 | 25.59772742 | 25.59772742 | 526.6154713 | 2.721493614 | TRUE | FALSE | FALSE |
| ZNF268 | 12175.421 | 0.00016079 | 0.4976 | 1244.302219 | 1248645.921 | 13073669.95 | 204349.569 | 9.72264E-06 | 0.27145 | 2042.587986 | 2114785.739 | 22373491.39 | 0.059581337 | -1.224889756 | TRUE | FALSE | FALSE |
| ZNF544 | 11170.593 | 0.00016486 | 0.06315 | 126.3049628 | 108701.4947 | 1081416.543 | 2026366056 | 8.08996E-10 | 0.182 | 18100733.93 | 22107855047 | 2.79235E+12 | 5.51262E-06 | -5.258641691 | TRUE | FALSE | FALSE |
| LTB4R2 | 11149.981 | 0.00016893 | 0.31605 | 4543.587515 | 1152663.57 | 12125791.38 | 127235.335 | 1.58848E-05 | 0.50195 | 391018.6001 | 44594350.19 | 4455868213 | 0.087632737 | -1.057333625 | TRUE | FALSE | FALSE |
| ALDH1L2 | 11129.676 | 0.00017298 | 0.0211 | 32.34393245 | 4742.300975 | 44532.03498 | 0.1230587 | 0.870587576 | 0.95235 | 17.74115947 | 2626.085738 | 10508.74267 | 90442.01036 | 4.956370207 | TRUE | TRUE | TRUE |
| DHX58 | 10975.663 | 0.00017707 | 0.2688 | 2744.324603 | 1134352.078 | 11931620.74 | 9.549013 | 0.205050624 | 0.4148 | 9.549013001 | 208.5806619 | 834.418453 | 1149.402876 | 3.060472279 | TRUE | FALSE | FALSE |
| CRB2 | 10940.956 | 0.00018116 | 0.60445 | 27160.96973 | 96219949.01 | 2346769610 | 79.431085 | 0.036732116 | 0.57945 | 183.0285325 | 66768.44447 | 805605.9545 | 137.7414932 | 2.139064787 | TRUE | FALSE | FALSE |
| NPFFR2 | 10800.296 | 0.00018528 | 1 | 2.338E+21 | 1.38397E+27 | 3.14731E+29 | 0.72643907 | 0.653745039 | 1 | 4.10372E+25 | 2.23397E+33 | 5.99871E+35 | 14867.44949 | 4.172236472 | TRUE | TRUE | TRUE |
| TSPAN32 | 10731.284 | 0.00018941 | 0.36415 | 1067.659933 | 110435.3283 | 1162965.888 | 154.021323 | 0.017983209 | 0.1911 | 154.0213234 | 1522.718758 | 15421.89054 | 69.67401529 | 1.84307084 | TRUE | FALSE | FALSE |
| LTBP4 | 10602.342 | 0.00019358 | 0.68495 | 73903.09816 | 10968294.28 | 176519232.2 | 59625.8256 | 3.41037E-05 | 0.5197 | 103409.9946 | 15398677.6 | 247832135.5 | 0.177814599 | -0.750032586 | TRUE | FALSE | FALSE |
| SRSF11 | 10436.449 | 0.00019779 | 0.000025 | 0.246988766 | 0.246988766 | 4.885458625 | 0.29151035 | 0.827595944 | 1 | 0.291510351 | 0.291510351 | 5.773726913 | 35801.29971 | 4.553898793 | TRUE | TRUE | TRUE |
| ZNF816 | 10253.699 | 0.00020207 | 0.85165 | 1058966.591 | 1309938331 | 14664764477 | 225731.934 | 8.77867E-06 | 0.0061 | 227.3542201 | 22107.54499 | 225731.9337 | 0.045424229 | -1.342712437 | TRUE | FALSE | FALSE |
| ZNF135 | 10072.895 | 0.00020641 | 0.71835 | 100440.5386 | 10680666.91 | 1238252703 | 14402.2234 | 0.00014571 | 0.94885 | 15578292.76 | 38553201939 | 4.56382E+11 | 0.699398636 | -0.155275219 | TRUE | FALSE | FALSE |
| SAMD9L | 9939.981 | 0.0002108 | 0.66375 | 39761.27576 | 63054721.98 | 1143561261 | 4675551.84 | 3.63567E-07 | 0.32645 | 1801216.722 | 2149025398 | 23770252129 | 0.002125948 | -2.672447323 | TRUE | FALSE | FALSE |
| UPK2 | 9706.4936 | 0.00021528 | 0.36515 | 977.5554592 | 98679.20624 | 1026588.157 | 12669.0832 | 0.000167645 | 0.14745 | 1267.917412 | 129609.5226 | 1356847.876 | 0.766155956 | -0.115682818 | TRUE | FALSE | FALSE |
| NPHP1 | 9503.8218 | 0.00021984 | 0.61535 | 18699.58354 | 10996289.89 | 482196040.3 | 2.28687318 | 0.46583607 | 0.81035 | 21.63515266 | 209.8080361 | 2084.347285 | 4155.814951 | 3.618656201 | TRUE | FALSE | FALSE |
| GFRAL | 9171.2744 | 0.00022457 | 0.1084 | 93.88574734 | 9171.274392 | 93934.91912 | 452.807657 | 0.00511608 | 0.6396 | 4438.127499 | 458112.2839 | 4819220.749 | 20.25423875 | 1.306515925 | TRUE | FALSE | FALSE |
| FAM83A | 9164.0082 | 0.00022927 | 0.5004 | 9164.008201 | 976263.2006 | 10425386.09 | 1330.93779 | 0.001825429 | 0.11525 | 135.6560279 | 13376.98341 | 137655.6687 | 6.885376832 | 0.837927714 | TRUE | FALSE | FALSE |
| PEX7 | 9145.7784 | 0.00023397 | 0.06005 | 95.99876887 | 9145.77837 | 36586.5146 | 34.1916188 | 0.081588688 | 0.31545 | 34.19161878 | 821.4368236 | 3284.604675 | 267.4859716 | 2.42730101 | TRUE | FALSE | FALSE |
| NLRX1 | 8908.4411 | 0.00023877 | 0.1833 | 909.4498441 | 229015.6091 | 2400417.149 | 0.68856306 | 0.671519946 | 0.38645 | 0.304682723 | 11.00895451 | 13.4604297 | 12937.72737 | 4.111857995 | TRUE | TRUE | TRUE |
| ERCC6 | 8839.82 | 0.0002436 | 0.6455 | 35411.59175 | 19348801.89 | 203794802.3 | 195469515 | 8.67717E-09 | 0.6982 | 1685741414 | 3.28785E+12 | 1.13958E+14 | 4.52235E-05 | -4.344635617 | TRUE | FALSE | FALSE |
| SLC39A5 | 8837.3708 | 0.0002484 | 0.0964 | 138.0837649 | 21390.54704 | 105655.6337 | 44.3247836 | 0.062025683 | 0.3056 | 11.08086549 | 413.534398 | 1654.600361 | 199.3776406 | 2.299676452 | TRUE | FALSE | FALSE |
| GOT1L1 | 8822.0566 | 0.00025319 | 0.0416 | 91.03810457 | 8822.056645 | 89996.95644 | 0.20469376 | 0.850868806 | 0.7535 | 0.46622693 | 1.864100489 | 1.864100489 | 43098.8057 | 4.634465236 | TRUE | TRUE | TRUE |
| MRI1 | 8663.5903 | 0.00025805 | 0.38 | 5216.200658 | 825420.8105 | 5361965.292 | 11.1451738 | 0.186397203 | 0.642 | 25.68979486 | 411.2387098 | 1645.975461 | 777.3400829 | 2.890611062 | TRUE | FALSE | FALSE |
| TAAR8 | 8643.4711 | 0.0002629 | 0.1119 | 215.5207418 | 8643.471101 | 88790.42938 | 0.34599278 | 0.810291687 | 1 | 0.345992779 | 0.345992779 | 0.345992779 | 24981.6517 | 4.397621149 | TRUE | TRUE | TRUE |
| GLIPR1L2 | 8626.1994 | 0.00026774 | 0.01805 | 89.50409554 | 8626.199373 | 87759.48018 | 12.2918728 | 0.17690464 | 0.8897 | 117.0371144 | 11420.95551 | 116915.197 | 701.7807247 | 2.846201436 | TRUE | FALSE | FALSE |
| PTPN13 | 8383.7517 | 0.00027271 | 0.0424 | 27.00447345 | 8383.751726 | 78149.14786 | 0.22333075 | 0.846552092 | 0.98175 | 70.61757611 | 18188.902 | 292349.9948 | 37539.62106 | 4.574489884 | TRUE | TRUE | TRUE |
| METTL6 | 8350.2491 | 0.00027768 | 0.3756 | 834.5160018 | 85546.75442 | 896833.894 | 12.7900468 | 0.172212718 | 0.657 | 108.8001799 | 4838.71518 | 48809.70443 | 652.8708765 | 2.814827296 | TRUE | FALSE | FALSE |
| WRAP53 | 8158.7708 | 0.00028276 | 0.14825 | 433.4192621 | 81478.75545 | 832679.7864 | 3079.54883 | 0.000736315 | 0.3858 | 3079.548834 | 1275638.774 | 13432012.81 | 2.649339644 | 0.423137638 | TRUE | FALSE | FALSE |
| TNNI3K | 8083.0997 | 0.00028786 | 0.85015 | 211800.2994 | 51612512.3 | 828744754.1 | 8380449.14 | 2.12137E-07 | 0.6116 | 22272494.4 | 22601117470 | 3.63096E+11 | 0.000964519 | -3.015689359 | TRUE | FALSE | FALSE |
| FAM135A | 7764.2985 | 0.00029318 | 1 | 6.36513E+45 | 2.05356E+55 | 1.53675E+59 | 128.516465 | 0.02209944 | 1 | 1.60036E+54 | 4.43155E+64 | 4.35358E+68 | 60.41481544 | 1.781143453 | TRUE | FALSE | FALSE |
| RPS27A | 7654.8075 | 0.00029855 | 0.9145 | 122902.0393 | 31776688.35 | 511720634.8 | 2623.90751 | 0.000875227 | 0.01015 | 41.81316834 | 2623.907514 | 3337.698883 | 2.917331276 | 0.464985748 | TRUE | FALSE | FALSE |
| FGGY | 7473.8906 | 0.00030405 | 0.1123 | 466.3148965 | 29932.50032 | 216936.7285 | 236268.712 | 8.48304E-06 | 0.0352 | 918.3972731 | 58972.53587 | 946827.6145 | 0.03163301 | -1.499859475 | TRUE | FALSE | FALSE |
| ATP1A3 | 7473.592 | 0.00030952 | 0.99995 | 2.25855E+13 | 9.26979E+18 | 1.2972E+20 | 0.12769992 | 0.86948532 | 1 | 5.9558E+15 | 2.04115E+20 | 4.19358E+22 | 58524.64266 | 4.767338771 | TRUE | TRUE | TRUE |
| PHOX2B | 7419.4591 | 0.00031501 | 1 | 2.74087E+25 | 3.12646E+30 | 3.147E+32 | 1201.23033 | 0.00212046 | 1 | 1.40021E+22 | 4.74242E+26 | 4.53936E+28 | 6.176549898 | 0.790745954 | TRUE | FALSE | FALSE |
| DNASE2B | 7380.3813 | 0.0003205 | 0.3304 | 1844.198213 | 180810.1737 | 1157151.339 | 987.735668 | 0.002454169 | 0.6394 | 1690.520736 | 253914.6874 | 1496064.224 | 7.472020653 | 0.873438064 | TRUE | FALSE | FALSE |
| OR52M1 | 7047.6649 | 0.00032626 | 0.3906 | 7047.664866 | 7324360.205 | 77514392.15 | 16.6008742 | 0.149536694 | 0.77175 | 36.09458862 | 14695.82171 | 29716.83768 | 424.5357673 | 2.627914286 | TRUE | FALSE | FALSE |
| DPEP2 | 7009.1648 | 0.00033203 | 0.6185 | 71153.28839 | 85298350.6 | 946658180.6 | 9506.1993 | 0.000206619 | 0.80945 | 97125.63584 | 118698400.9 | 1122166505 | 0.737325674 | -0.132340643 | TRUE | FALSE | FALSE |
| MYOM3 | 6950.5083 | 0.00033783 | 0.41045 | 2999.69344 | 3735375.263 | 49239000.25 | 70.2453544 | 0.040835445 | 0.6135 | 649.055812 | 585206.4637 | 5901706.735 | 98.94616392 | 1.995398962 | TRUE | FALSE | FALSE |
| ZNF568 | 6845.117 | 0.0003437 | 0.0127 | 73.2509371 | 6845.116966 | 23661.63111 | 140.433202 | 0.020052057 | 0.0654 | 1.612785413 | 140.4332021 | 140.4332021 | 48.74286752 | 1.687911075 | TRUE | FALSE | FALSE |
| CD1B | 6510.7683 | 0.00034988 | 0.94215 | 679874.8822 | 4325006817 | 50101266267 | 101.750126 | 0.028647171 | 0.92815 | 100519.5584 | 122197324.7 | 872239821.8 | 63.9878152 | 1.806097282 | TRUE | FALSE | FALSE |
| ZNF519 | 6411.4008 | 0.00035615 | 0.6233 | 6411.40084 | 7099900.393 | 76815519.51 | 912.691195 | 0.002592151 | 0.74305 | 9084.065159 | 965327.6013 | 10295642.31 | 7.024720823 | 0.846629069 | TRUE | FALSE | FALSE |
| ALS2CL | 6395.9893 | 0.00036239 | 0.207 | 674.82342 | 614568.2172 | 6224363.057 | 1.12339471 | 0.575553516 | 0.5068 | 1.123394708 | 22.35254334 | 89.39179483 | 5693.447953 | 3.755375355 | TRUE | FALSE | FALSE |
| NPB | 6370.6804 | 0.00036864 | 0.00075 | 63.26246705 | 6370.68038 | 6370.68038 | 66.1501729 | 0.042991373 | 0.68945 | 66.15017285 | 6674.997965 | 6674.997965 | 96.30633006 | 1.983654834 | TRUE | FALSE | FALSE |
| RNASEL | 6336.5268 | 0.0003749 | 0.98745 | 129978744.2 | 1.3932E+11 | 2.02423E+13 | 147.729951 | 0.019070994 | 0.97655 | 564737.5775 | 661298590.3 | 16420556769 | 42.89263429 | 1.63238272 | TRUE | FALSE | FALSE |
| GALC | 6116.227 | 0.00038138 | 0.42525 | 3508.208631 | 1571023.569 | 25216315.75 | 1.05313603 | 0.58305621 | 0.89575 | 37.13565665 | 2378.312173 | 20111.32697 | 5807.632471 | 3.763999125 | TRUE | FALSE | FALSE |
| AIP | 6109.2151 | 0.00038784 | 0.88805 | 548779.5909 | 218541603.9 | 1605439453 | 35.8779169 | 0.077372414 | 0.8801 | 1348.180104 | 94760.37006 | 834088.8472 | 170.2778658 | 2.231158198 | TRUE | FALSE | FALSE |
| AIPL1 | 6086.1264 | 0.00039429 | 0.8446 | 391336.8095 | 36114365.97 | 581349820.2 | 6999847.21 | 2.63501E-07 | 0.81105 | 200700855.9 | 1.29844E+11 | 2.08965E+12 | 0.000869466 | -3.060747592 | TRUE | FALSE | FALSE |
| CCR5 | 5974.2268 | 0.00040085 | 0.1521 | 594.1144403 | 61507.87945 | 648010.8321 | 91.8645296 | 0.032187757 | 0.5059 | 91.86452956 | 908.5740291 | 9205.592137 | 65.03300977 | 1.813133854 | TRUE | FALSE | FALSE |
| KRTAP9-2 | 5932.9877 | 0.00040744 | 0.1143 | 584.3711929 | 61704.13354 | 657019.8035 | 7.3271057 | 0.238154365 | 0.7531 | 7.327105698 | 71.09524055 | 71.09524055 | 809.7314188 | 2.908340991 | TRUE | FALSE | FALSE |
| KRTAP21-2 | 5921.2456 | 0.00041401 | 0.50095 | 61511.19095 | 7120215.616 | 79254099.83 | 6038.86632 | 0.000285183 | 0.62345 | 62741.71402 | 80912509.34 | 920923571.9 | 0.980522722 | -0.008542338 | TRUE | FALSE | FALSE |
| OR13C9 | 5821.9877 | 0.00042067 | 0.81085 | 58564.71609 | 68314134.02 | 751299152.1 | 81552.8377 | 2.45574E-05 | 0.611 | 81552.8377 | 97270088.03 | 448558828.4 | 0.071389149 | -1.146367797 | TRUE | FALSE | FALSE |
| LTK | 5737.6792 | 0.00042741 | 0.8455 | 525542.5174 | 2172263635 | 61636477170 | 23090.2057 | 7.86699E-05 | 0.8148 | 3702011.552 | 11598293357 | 5.16084E+11 | 0.248489739 | -0.60469154 | TRUE | FALSE | FALSE |
| RERE | 5657.0798 | 0.00043424 | 0.5172 | 59164.34395 | 7093640.188 | 136869960.1 | 93.6856236 | 0.030923648 | 0.51185 | 93.68562363 | 3768.04885 | 38820.7948 | 60.38364861 | 1.780919351 | TRUE | FALSE | FALSE |
| SLC26A1 | 5594.9024 | 0.00044111 | 0.38885 | 3587.64707 | 89929.26674 | 1446912.665 | 290.220105 | 0.008605666 | 0.6727 | 2009.172867 | 298087.156 | 2829517.94 | 19.27813501 | 1.285065017 | TRUE | FALSE | FALSE |
| ZNF280A | 5434.2597 | 0.00044819 | 0.02075 | 57.28356024 | 5434.25973 | 54851.22593 | 8.33489764 | 0.220893558 | 0.6592 | 78.80374744 | 7582.545582 | 77078.24598 | 651.9887784 | 2.814240121 | TRUE | FALSE | FALSE |
| ZNF138 | 5373.896 | 0.00045532 | 0.10685 | 56.5368593 | 5373.895953 | 54294.80511 | 7.48626984 | 0.235176322 | 0.9802 | 683.5751518 | 713889.343 | 7581882.101 | 717.8335894 | 2.856023776 | TRUE | FALSE | FALSE |

| | | | | | | | | | | | | | | | | | |
|---|---|---|---|---|---|---|---|---|---|---|---|---|---|---|---|---|---|
| AARSD1 | 5331.3821 | 0.00046249 | 0.4643 | 5331.382086 | 866809.8958 | 3490086.677 | 2.16426653 | 0.474655688 | 0.936 | 8.658592536 | 138.690212 | 138.690212 | 2463.366693 | 3.391529065 | TRUE | FALSE | FALSE |
| LONP1 | 5046.2678 | 0.00047009 | 0.00225 | 9.329825543 | 181.4954775 | 1260.884676 | 13.5043275 | 0.1661712 | 0.63205 | 54.03715365 | 3467.381147 | 13888.48056 | 373.6778283 | 2.57249733 | TRUE | FALSE | FALSE |
| NIPAL4 | 4967.0486 | 0.00047778 | 0.235 | 539.001625 | 19889.04837 | 319131.9497 | 124.338691 | 0.022860268 | 0.18425 | 13.44591078 | 497.5561177 | 617.1701215 | 39.94773091 | 1.601492116 | TRUE | FALSE | FALSE |
| SIRT3 | 4651.4075 | 0.00048604 | 0.1108 | 48.35809411 | 4651.407501 | 47274.70171 | 0.87249133 | 0.602466354 | 0.7525 | 0.872491333 | 8.120718419 | 8.120718419 | 5331.179036 | 3.726823268 | TRUE | TRUE | TRUE |
| FECH | 4637.6513 | 0.00049428 | 0.51115 | 18574.02064 | 1195031.132 | 23711460.42 | 31.8560444 | 0.08789044 | 0.72975 | 127.476322 | 2042.820132 | 8180.74777 | 145.5815184 | 2.163106245 | TRUE | FALSE | FALSE |
| PCDHGA12 | 4604.9523 | 0.00050255 | 0.7909 | 46585.95623 | 55113063.55 | 608423471.2 | 107.802895 | 0.027471426 | 0.64295 | 107.802895 | 10592.07039 | 108714.458 | 42.71640677 | 1.630594713 | TRUE | FALSE | FALSE |
| ROM1 | 4570.8511 | 0.00051085 | 0.0785 | 71.32637903 | 10668.80801 | 42704.46917 | 14678.8111 | 0.000139048 | 0.3763 | 1457.733307 | 1594100.716 | 17174687.75 | 0.311391099 | -0.506693806 | TRUE | FALSE | FALSE |
| C1orf216 | 4553.1616 | 0.00051916 | 0.002 | 45.7610243 | 4553.161648 | 4553.161648 | 490.531428 | 0.004555359 | 0.31725 | 49.70312912 | 4962.253443 | 4962.253443 | 9.282099759 | 0.967646232 | TRUE | FALSE | FALSE |
| TRAP1 | 4354.7576 | 0.00052785 | 0.49275 | 3342.852718 | 1602209.545 | 25663722.5 | 0.00198441 | 0.899096042 | 1 | 0.154396558 | 2.466445907 | 9.861691224 | 2194486.324 | 6.341332879 | TRUE | TRUE | TRUE |
| CHAC2 | 4351.2438 | 0.00053651 | 0.46435 | 4351.243821 | 279276.8136 | 1970936.343 | 314555.271 | 6.03694E-06 | 0.0877 | 3917.714331 | 314555.2706 | 1260986.504 | 0.013833002 | -1.859083547 | TRUE | FALSE | FALSE |
| COL6A5 | 4234.4264 | 0.0005454 | 0.8423 | 3672661.138 | 40120044778 | 4.693E+12 | 31435.7751 | 6.15311E-05 | 0.0637 | 358.6007564 | 303905.6044 | 2999653.576 | 0.134700874 | -0.870629585 | TRUE | FALSE | FALSE |
| SDR39U1 | 4119.0721 | 0.00055454 | 0.82715 | 408134.6737 | 477920732.3 | 5262797153 | 0.51529476 | 0.749328065 | 0.9994 | 4333.665748 | 4885780.386 | 53175782.52 | 7993.623188 | 3.902743672 | TRUE | TRUE | TRUE |
| SLC22A1 | 4082.3144 | 0.00056372 | 0.27675 | 798.1735438 | 114446.9943 | 1049345.662 | 5738.96562 | 0.000300972 | 0.50665 | 5738.965623 | 989752.848 | 8933463.743 | 0.711332778 | -0.147927179 | TRUE | FALSE | FALSE |
| MYH7B | 4076.8247 | 0.00057288 | 0.33395 | 254.7192795 | 7601799.575 | 275491792.6 | 0.00610184 | 0.897839921 | 0.92055 | 13.94196703 | 188497.8267 | 4422945.271 | 668129.9261 | 5.824860925 | TRUE | TRUE | TRUE |
| PCGF3 | 4041.1253 | 0.00058209 | 0.74765 | 41841.26221 | 4808753.936 | 53347457.1 | 51.0300788 | 0.053791068 | 0.508 | 505.829969 | 5139.49131 | 53492.75338 | 79.1910456 | 1.898676077 | TRUE | FALSE | FALSE |
| HOXD12 | 3920.18 | 0.00059158 | 0.50255 | 3920.179994 | 1026626.06 | 7839581.177 | 42.2316308 | 0.064949935 | 0.13185 | 4.440801803 | 42.23163083 | 42.23163083 | 92.82568344 | 1.967668155 | TRUE | FALSE | FALSE |
| PMP22 | 3835.9342 | 0.00060126 | 0.05695 | 239.2831977 | 15364.31602 | 33152.46716 | 2464.74082 | 0.000965729 | 0.82725 | 13271.8987 | 503229.1876 | 3429215.631 | 1.556323554 | 0.19209989 | TRUE | FALSE | FALSE |
| SUSD2 | 3769.6174 | 0.00061109 | 0.2611 | 458.4940815 | 143413.3944 | 1396665.453 | 0.14278749 | 0.86619397 | 0.6389 | 0.261041053 | 2.284065329 | 5.11846036 | 26400.19352 | 4.42160711 | TRUE | TRUE | TRUE |
| DECR2 | 3724.166 | 0.00062101 | 0.83585 | 138968.9626 | 35859932.65 | 143814665.3 | 10.5638554 | 0.192165623 | 0.49815 | 10.56385543 | 666.7342716 | 2670.694518 | 352.5385217 | 2.547206579 | TRUE | FALSE | FALSE |
| ZNF546 | 3705.4287 | 0.00063094 | 0.31415 | 3705.428702 | 3115257.02 | 30668047.54 | 1.95044287 | 0.493944963 | 0.81505 | 149.3092589 | 118795.7208 | 1150214.16 | 1899.788379 | 3.278705227 | TRUE | FALSE | FALSE |
| WNT10A | 3699.5344 | 0.00064085 | 0.41725 | 3699.534392 | 605682.2147 | 6347357.938 | 3.14970505 | 0.403757009 | 0.7712 | 12.59948086 | 470.7317011 | 1710.378896 | 1174.565344 | 3.069877183 | TRUE | FALSE | FALSE |
| NUDT7 | 3644.4023 | 0.00065089 | 0.032 | 21.32059084 | 911.1605565 | 3644.402279 | 5.67164709 | 0.278818666 | 0.6944 | 13.20008844 | 504.1115892 | 504.1115892 | 642.5650644 | 2.80791711 | TRUE | FALSE | FALSE |
| TAS1R1 | 3593.4976 | 0.00066104 | 0.39975 | 2208.912818 | 1418126.125 | 14577010.03 | 527.957373 | 0.004259401 | 0.7718 | 8436.974157 | 8820669.27 | 141098835 | 6.80641621 | 0.832918503 | TRUE | FALSE | FALSE |
| EVPL | 3573.891 | 0.00067121 | 0.5196 | 34091.83013 | 33551132.64 | 3670508927 | 0.03330659 | 0.890512533 | 0.98195 | 89.32756258 | 73808.24341 | 723678.7683 | 107302.8321 | 5.030611185 | TRUE | TRUE | TRUE |
| NME1 | 3556.8265 | 0.00068139 | 0.7952 | 56917.40709 | 67258856.01 | 743251031.5 | 468.610035 | 0.004738855 | 0.1908 | 468.6100351 | 4744.631608 | 49187.8875 | 7.590162973 | 0.880251101 | TRUE | FALSE | FALSE |
| KLC3 | 3523.2787 | 0.00069163 | 0.0562 | 21.72972816 | 3523.27872 | 14100.92237 | 3.96104411 | 0.357593057 | 0.3554 | 1.799095412 | 142.9372231 | 571.6903536 | 889.4823242 | 2.949137322 | TRUE | FALSE | FALSE |
| FMO2 | 3511.9227 | 0.00070187 | 0.18945 | 348.6386549 | 3511.92265 | 36239.93496 | 365.313391 | 0.006479154 | 0.49955 | 365.3133908 | 402492.4644 | 753713.3116 | 9.613451732 | 0.98287935 | TRUE | FALSE | FALSE |
| PTEN | 3340.4594 | 0.00071267 | 0.000025 | 0.238082865 | 0.238082865 | 4.727692765 | 0.27026392 | 0.833707677 | 0.7512 | 2.527994469 | 10.11010885 | 10.11010885 | 12359.99021 | 4.092018127 | TRUE | TRUE | TRUE |
| SMPD1 | 3309.3121 | 0.00072353 | 0.53915 | 3309.312141 | 116221.1218 | 791472.5183 | 2093.85152 | 0.001149441 | 0.0677 | 130.6049879 | 8386.899336 | 33602.01112 | 1.580490362 | 0.198791852 | TRUE | FALSE | FALSE |
| CCDC14 | 3264.0643 | 0.00073451 | 0.0443 | 35.37075338 | 3264.064282 | 32468.95929 | 6.34107876 | 0.260597935 | 0.50355 | 59.47122217 | 5621.309515 | 11305.65414 | 514.7490525 | 2.711595556 | TRUE | FALSE | FALSE |
| TP53AIP1 | 3135.0816 | 0.00074596 | 0.06535 | 32.27202707 | 3135.081577 | 3135.081577 | 0.42594104 | 0.78253487 | 1 | 3.954741982 | 368.3319549 | 368.3319549 | 7360.365213 | 3.866899364 | TRUE | TRUE | TRUE |
| CUTA | 3036.8352 | 0.00075778 | 0.50775 | 3036.835172 | 304072.7958 | 3148164.117 | 53.4183223 | 0.051812573 | 0.34455 | 53.41832226 | 4983.431445 | 49881.32102 | 56.85006649 | 1.754730977 | TRUE | FALSE | FALSE |
| SHPK | 3022.9707 | 0.00076961 | 0.1148 | 128.797682 | 12090.15725 | 30352.15454 | 0.64649605 | 0.691870424 | 0.5021 | 0.64649605 | 0.64649605 | 1.424673565 | 4675.930634 | 3.66986806 | TRUE | TRUE | TRUE |
| RAB33B | 2997.6171 | 0.0007815 | 0.4032 | 1273.400087 | 38326.76097 | 153543.4129 | 36309.0245 | 5.03711E-05 | 0.5933 | 36309.02453 | 9383237.655 | 37645052.69 | 0.082558458 | -1.083238429 | TRUE | FALSE | FALSE |
| PSD2 | 2982.7679 | 0.0007934 | 0.0701 | 53.82793168 | 3442.703122 | 34451.3724 | 7781.8356 | 0.000223288 | 0.0979 | 485.9328303 | 31152.84026 | 263869.362 | 0.383298751 | -0.416462596 | TRUE | FALSE | FALSE |
| ZNF714 | 2948.1346 | 0.00080541 | 0.77635 | 280028.0699 | 305598369.8 | 35958142394 | 81.7124624 | 0.035955323 | 0.63595 | 81.71246242 | 7381.0696 | 72641.59311 | 36.07937576 | 1.557259015 | TRUE | FALSE | FALSE |
| HOXA4 | 2945.2863 | 0.00081739 | 0.3087 | 735.8283578 | 47223.12741 | 189160.7088 | 5255.58968 | 0.000342714 | 0.44965 | 5255.589683 | 1429248.207 | 8692217.311 | 0.560410238 | -0.251493939 | TRUE | FALSE | FALSE |
| ZNF695 | 2905.5324 | 0.0008295 | 0.23155 | 298.2249867 | 28983.89873 | 98412.80943 | 0.0685007 | 0.882757034 | 1 | 5.591336956 | 52.49239714 | 52.49239714 | 42416.09821 | 4.627530716 | TRUE | TRUE | TRUE |
| CHAF1B | 2887.6022 | 0.00084164 | 0.60995 | 14985.29073 | 964554.6412 | 15511106.1 | 23920722.1 | 9.24871E-08 | 0.0885 | 371030.583 | 23920722.07 | 202085904 | 0.000120716 | -3.918236914 | TRUE | FALSE | FALSE |
| CSH2 | 2884.7406 | 0.00085375 | 0.1878 | 28.98938837 | 2884.74057 | 29845.42375 | 0.43095643 | 0.780694482 | 1 | 0.430956429 | 0.430956429 | 0.430956429 | 6693.810271 | 3.825673399 | TRUE | TRUE | TRUE |
| TUBGCP2 | 2800.7861 | 0.00086621 | 0.1623 | 287.149181 | 27970.57045 | 285849.0198 | 0.60776546 | 0.708747625 | 0.87935 | 5.573349845 | 52.42209554 | 109.2909342 | 4608.333885 | 3.663543937 | TRUE | TRUE | TRUE |
| SERPINA10 | 2793.6603 | 0.00087866 | 0.18315 | 174.2937131 | 11188.76494 | 102228.798 | 8.2498728 | 0.223903434 | 0.6397 | 18.50479351 | 296.3478196 | 1186.378246 | 338.6307151 | 2.529726348 | TRUE | FALSE | FALSE |
| ZNF559 | 2784.667 | 0.00089111 | 0.1191 | 305.8584561 | 25932.87898 | 246909.7347 | 10.0175684 | 0.197938607 | 0.6356 | 10.01756837 | 810.0872518 | 7543.773276 | 277.9783397 | 2.444010957 | TRUE | FALSE | FALSE |
| CNBD2 | 2777.6681 | 0.00090353 | 0.3655 | 693.9198607 | 104432.2377 | 515535.1335 | 67.5627375 | 0.041906944 | 0.5024 | 67.56273749 | 6182.917352 | 30594.2406 | 41.11242696 | 1.613973115 | TRUE | FALSE | FALSE |
| GALR3 | 2762.9456 | 0.00091598 | 0.92365 | 246338.9983 | 38118233.55 | 612848363.7 | 6855915.98 | 2.77205E-07 | 0.2548 | 1000889.242 | 110341652.5 | 1038147300 | 0.000403002 | -3.394693145 | TRUE | FALSE | FALSE |
| SLC3A2 | 2739.6827 | 0.0009285 | 0.7927 | 31528.03151 | 15773946 | 105845971.8 | 13455.7828 | 0.000152863 | 0.0973 | 357.5746114 | 53863.14902 | 215666.5965 | 0.203606344 | -0.691208695 | TRUE | FALSE | FALSE |
| OTOP3 | 2611.3258 | 0.00094167 | 0.1159 | 268.3094705 | 26035.9347 | 66451.28193 | 1.1344633 | 0.574259405 | 0.87505 | 2.463385025 | 101.4191992 | 101.4191992 | 2301.816038 | 3.362070612 | TRUE | FALSE | FALSE |
| DNAH7 | 2607.9425 | 0.00095481 | 0.5027 | 2607.942541 | 186453792.8 | 18335844115 | 3.36917022 | 0.390794667 | 0.63315 | 29.11708228 | 1625685.885 | 71647157.14 | 774.060784 | 2.888775065 | TRUE | FALSE | FALSE |
| ENTPD4 | 2513.814 | 0.00096845 | 0.00365 | 27.25688223 | 258.6165991 | 2513.813987 | 0.05408136 | 0.885879288 | 0.96855 | 40.76132869 | 390.102991 | 3824.726516 | 46482.07859 | 4.667285541 | TRUE | TRUE | TRUE |
| TPMT | 2500.7316 | 0.00098211 | 0.5039 | 3292.367509 | 365297.2318 | 1464492.327 | 4490.11156 | 0.000416771 | 0.3434 | 1062.051952 | 38891.0595 | 273991.7933 | 0.556941978 | -0.254190047 | TRUE | FALSE | FALSE |
| ACSM5 | 2495.6431 | 0.00099575 | 0.32325 | 2495.643078 | 160756.428 | 644733.4454 | 1183.9595 | 0.002175855 | 0.7751 | 17296.66842 | 5227803.192 | 38342326.61 | 2.107878762 | 0.323845628 | TRUE | FALSE | FALSE |
| EXOC6B | 2484.4424 | 0.0010094 | 0.1837 | 154.9407694 | 6240.387787 | 25003.1244 | 3.05738631 | 0.408764599 | 0.91435 | 195.9660551 | 1750.49568 | 7012.204327 | 812.6033589 | 2.909878613 | TRUE | FALSE | FALSE |
| GPR68 | 2458.7098 | 0.00102316 | 0.19495 | 245.4517871 | 2458.7098 | 25230.37866 | 28.3182317 | 0.09783474 | 0.65915 | 277.2195361 | 28594.1387 | 300931.587 | 86.8242703 | 1.938641142 | TRUE | FALSE | FALSE |
| FAM120A | 2457.5195 | 0.00103687 | 0.1928 | 242.7433986 | 2457.519491 | 25478.84163 | 3.34830103 | 0.392157987 | 0.5114 | 3.348301026 | 3.348301026 | 65.43825448 | 733.9601404 | 2.865672475 | TRUE | FALSE | FALSE |
| HGSNAT | 2456.7995 | 0.00105052 | 0.0014 | 9.562745075 | 153.2002056 | 613.4234086 | 66.7728294 | 0.042773017 | 0.0451 | 4.169285199 | 66.77282936 | 330.2311641 | 36.79340118 | 1.565769936 | TRUE | FALSE | FALSE |
| SLC12A1 | 2420.161 | 0.00106436 | 0.44805 | 1156.321416 | 2525363.034 | 40782236.1 | 6.75336683 | 0.249952873 | 0.0803 | 0.422263914 | 15.98180984 | 256.8041747 | 358.3636171 | 2.554323911 | TRUE | FALSE | FALSE |
| IGFN1 | 2365.3218 | 0.00107851 | 1 | 1.06819E+43 | 2.97693E+52 | 5.08789E+56 | 1.6E+12 | 4.99409E-13 | 0.9998 | 1.9929E+26 | 1.01095E+34 | 7.77529E+36 | 1.47835E-09 | -8.830221814 | TRUE | FALSE | FALSE |
| RP1 | 2346.2355 | 0.00109273 | 0.34985 | 251.4980263 | 218416.6064 | 2999551.738 | 0.15211018 | 0.864047223 | 0.1489 | 0.038048837 | 1.322111712 | 2.945986845 | 15424.57885 | 4.188213315 | TRUE | TRUE | TRUE |
| TMEM234 | 2328.0113 | 0.00110702 | 0.185 | 24.08964587 | 2328.011288 | 23736.0714 | 3.26369201 | 0.396888852 | 0.2522 | 3.263692006 | 31.08433417 | 31.08433417 | 713.3060607 | 2.853275914 | TRUE | FALSE | FALSE |
| MS4A12 | 2314.6453 | 0.00112135 | 0.91075 | 248155.6291 | 451051718.8 | 5151284020 | 346.954009 | 0.006942251 | 0.75045 | 3503.249273 | 383120.9768 | 4145169.723 | 6.671331727 | 0.824212536 | TRUE | FALSE | FALSE |
| QRFPR | 2278.5344 | 0.00113588 | 1 | 4.23466E+46 | 7.94179E+56 | 6.85579E+60 | 0.00397019 | 0.89836337 | 1 | 2.95785E+77 | 1.08255E+91 | 3.67441E+96 | 573909.9753 | 5.758843773 | TRUE | TRUE | TRUE |
| CCDC89 | 2248.3609 | 0.00115057 | 0.1132 | 226.2873942 | 22884.97813 | 238488.5252 | 3.28057088 | 0.395768218 | 0.7492 | 3.280570877 | 31.22553705 | 31.22553705 | 685.3566056 | 2.835916603 | TRUE | FALSE | FALSE |
| HDAC4 | 2243.4184 | 0.00116524 | 0.21375 | 395.1466749 | 2243.418447 | 18907.12139 | 211.69551 | 0.012547383 | 0.41275 | 211.6955102 | 8004.206425 | 32051.33712 | 10.59738322 | 1.025198639 | TRUE | FALSE | FALSE |
| MED15 | 2214.5122 | 0.00118007 | 0.0074 | 22.38618439 | 2214.512207 | 2214.512207 | 0.33417413 | 0.814251086 | 1 | 0.33417413 | 0.33417413 | 6.57676142 | 6626.821198 | 3.821305253 | TRUE | TRUE | TRUE |
| CCDC81 | 2167.734 | 0.00119521 | 0.6274 | 2167.734036 | 2245503.27 | 23760880.36 | 3702.78232 | 0.00051234 | 0.22705 | 84.52015426 | 37054.36951 | 73354.62461 | 0.585433831 | -0.232522185 | TRUE | FALSE | FALSE |
| MEPE | 2157.1855 | 0.00121038 | 1 | 2.33744E+19 | 1.17431E+25 | 2.5099E+27 | 0.52304061 | 0.745551544 | 1 | 3.16996E+16 | 9.42316E+20 | 1.80237E+23 | 4124.317435 | 3.615352084 | TRUE | TRUE | TRUE |
| OSGEPL1 | 2149.0466 | 0.00122554 | 0.84985 | 2298229.79 | 2997566508 | 32510262919 | 3335.65258 | 0.000597401 | 0.9394 | 346590.1332 | 4963449781 | 56551925500 | 0.644265723 | -0.190934974 | TRUE | FALSE | FALSE |
| CDK11B | 2120.8041 | 0.00124088 | 0.8103 | 228162.8364 | 294979634.7 | 3383896702 | 2709.64025 | 0.000831501 | 0.83035 | 27902.3005 | 34541045.38 | 389639619.5 | 0.782688459 | -0.10641107 | TRUE | FALSE | FALSE |
| POLR3B | 2098.4339 | 0.00125635 | 0.38735 | 524.1286119 | 134870.6711 | 1206380.166 | 36.4608739 | 0.076252874 | 0.50555 | 36.46087394 | 2340.910871 | 20615.48404 | 57.55303301 | 1.760068216 | TRUE | FALSE | FALSE |
| LONP2 | 2097.844 | 0.00127175 | 0.5053 | 2097.844003 | 517432.1345 | 5386682.268 | 1.36064145 | 0.549920236 | 0.8135 | 12.50092597 | 117.7319814 | 1135.905052 | 1541.805153 | 3.188029493 | TRUE | FALSE | FALSE |
| COX6B2 | 2077.9338 | 0.00128727 | 0.88905 | 221379.615 | 27347912.82 | 305420540.3 | 275774.924 | 6.54355E-06 | 0.36885 | 26294.94214 | 33680023.2 | 61281601.64 | 0.00753489 | -2.122923062 | TRUE | FALSE | FALSE |
| ADAP1 | 2074.2838 | 0.00130276 | 0.82845 | 21182.70965 | 26721029.05 | 298702584.9 | 276055.852 | 6.29113E-06 | 0.26085 | 26296.03074 | 2997885.151 | 32918616.71 | 0.007514001 | -2.124128773 | TRUE | FALSE | FALSE |
| MRGPRF | 2065.1633 | 0.00131826 | 0.99625 | 296186545.3 | 4.70268E+11 | 7.08659E+13 | 30965562.7 | 5.73873E-08 | 0.74715 | 3969389040 | 8.37052E+13 | 1.05995E+15 | 6.66923E-05 | -4.175924585 | TRUE | FALSE | FALSE |

| | | | | | | | | | | | | | | | | | |
|---|---|---|---|---|---|---|---|---|---|---|---|---|---|---|---|---|---|
| UTP14C | 2062.332 | 0.00133372 | 0.8112 | 20369.92957 | 22495827.64 | 242938733 | 387.345245 | 0.006255174 | 0.2365 | 387.3452445 | 37933.21998 | 74909.25353 | 5.324273329 | 0.726260343 | TRUE | FALSE | FALSE |
| KRTAP13-1 | 2056.8751 | 0.00134918 | 0.50365 | 20814.44888 | 2286381.166 | 21195029.3 | 250280.452 | 7.0853E-06 | 0.38005 | 24077.20213 | 23506284.72 | 261600288.7 | 0.008218281 | -2.085219002 | TRUE | FALSE | FALSE |
| RNF145 | 1971.1746 | 0.00136535 | 0.50415 | 20288.64814 | 2308987.308 | 25467703.15 | 285.397911 | 0.00878347 | 0.1381 | 28.89234383 | 1141.454257 | 1141.454257 | 6.906758866 | 0.839274294 | TRUE | FALSE | FALSE |
| GPR35 | 1918.7092 | 0.00138196 | 0.23465 | 1918.709188 | 205609.487 | 205609.487 | 2.64431014 | 0.437641059 | 0.9691 | 52.45493429 | 2479.293892 | 25485.67439 | 725.5991486 | 2.860696764 | TRUE | FALSE | FALSE |
| TRIM45 | 1914.3794 | 0.00139855 | 0.3251 | 478.8383012 | 184412.9392 | 1872770.549 | 0.36094838 | 0.80534067 | 0.88825 | 13.2121172 | 1192.588967 | 11742.16183 | 5303.748587 | 3.724582929 | TRUE | TRUE | TRUE |
| PHYHD1 | 1828.0364 | 0.00141598 | 0.5813 | 3160.395934 | 469835.2963 | 4071780.896 | 142.401058 | 0.019909643 | 0.77075 | 2283.242468 | 295565.807 | 4751256.797 | 12.83723906 | 1.108471629 | TRUE | FALSE | FALSE |
| SFXN3 | 1825.1754 | 0.00143337 | 0.8474 | 358182.9992 | 2374655552 | 26745943218 | 3401.23082 | 0.000560182 | 0.63065 | 3401.230824 | 3659757.79 | 39218267.63 | 0.536622037 | -0.270331497 | TRUE | FALSE | FALSE |
| TRAPPC12 | 1791.7316 | 0.00145107 | 0.01345 | 12.43271129 | 447.46355 | 1791.731619 | 0.07050127 | 0.882317223 | 0.9709 | 4.513166786 | 40.82933273 | 163.4120787 | 25414.1768 | 4.405076047 | TRUE | TRUE | TRUE |
| SLCO1B3 | 1788.4905 | 0.00146873 | 0.00845 | 2.45967195 | 194.2046653 | 447.2975178 | 0.17053068 | 0.859477781 | 0.31345 | 0.170530677 | 13.3017763 | 13.3017763 | 10487.79331 | 4.02068412 | TRUE | TRUE | TRUE |
| APPL1 | 1774.7171 | 0.00148648 | 0.1203 | 178.734404 | 18052.1607 | 188001.8053 | 23.0037742 | 0.117917525 | 0.8153 | 223.8430022 | 2232.658255 | 22812.70779 | 77.14895392 | 1.887330042 | TRUE | FALSE | FALSE |
| TYSND1 | 1737.6375 | 0.00150459 | 0.0934 | 108.3857064 | 4026.816123 | 16125.36657 | 8.99568057 | 0.211799538 | 0.7516 | 144.0862823 | 2310.175929 | 11885.20089 | 193.1635402 | 2.285925156 | TRUE | FALSE | FALSE |
| ZFP2 | 1734.57 | 0.00152268 | 0.23625 | 179.7953497 | 17133.97534 | 34420.62321 | 0.42451188 | 0.782946052 | 0.50275 | 0.424511881 | 0.424511881 | 0.952239809 | 4086.034103 | 3.611301987 | TRUE | TRUE | TRUE |
| CD276 | 1700.3662 | 0.00154111 | 0.11305 | 18.67606579 | 1700.366205 | 16814.07332 | 0.04554583 | 0.887698261 | 1 | 0.403337232 | 0.403337232 | 0.89956477 | 37333.08286 | 4.572093854 | TRUE | TRUE | TRUE |
| POLK | 1584.0209 | 0.00156101 | 0.2549 | 165.5869337 | 15514.99131 | 155511.6676 | 238.368195 | 0.010521236 | 0.4015 | 106.0473406 | 20805.43007 | 83270.7009 | 6.645269351 | 0.822512589 | TRUE | FALSE | FALSE |
| OR10AD1 | 1578.6351 | 0.00158092 | 0.8683 | 165474.8377 | 179266300.9 | 23559597680 | 25.4205553 | 0.108252331 | 0.65725 | 247.5209513 | 25258.61597 | 264408.5327 | 62.10073115 | 1.793096713 | TRUE | FALSE | FALSE |
| PLA2G4B | 1547.8467 | 0.0016012 | 0.27615 | 624.0339939 | 62994.19542 | 656413.5873 | 0.54859278 | 0.733823262 | 1 | 5.195384784 | 5.195384784 | 18.5609724 | 2821.485724 | 3.450477857 | TRUE | TRUE | TRUE |
| GP1BA | 1547.1068 | 0.00162141 | 0.3715 | 160.9024353 | 15231.12512 | 153448.5172 | 0.04520171 | 0.887787421 | 1 | 0.399385731 | 0.399385731 | 0.893599077 | 34226.72957 | 4.534365403 | TRUE | TRUE | TRUE |
| OR4F15 | 1540.932 | 0.00164164 | 0.9839 | 18281893.1 | 21305546292 | 2.50087E+11 | 20524762.2 | 1.20221E-07 | 0.597 | 233299776.4 | 6.02054E+11 | 5.20736E+13 | 7.50767E-05 | -4.124494652 | TRUE | FALSE | FALSE |
| CNGB1 | 1501.901 | 0.00166239 | 1 | 1.30919E+14 | 2.20567E+18 | 1.32509E+20 | 749927.869 | 2.33836E-06 | 0.9958 | 2.93536E+12 | 2.97323E+16 | 1.26071E+18 | 0.002002727 | -2.698378174 | TRUE | FALSE | FALSE |
| CLPB | 1498.5742 | 0.00168311 | 0.0898 | 163.9488393 | 6004.985108 | 24068.77309 | 551.402964 | 0.003924343 | 0.0457 | 34.41625461 | 551.4029643 | 2710.847538 | 2.717747873 | 0.434209165 | TRUE | FALSE | FALSE |
| PIGG | 1478.8368 | 0.00170406 | 0.18605 | 153.5163313 | 146996.19 | 1515260.095 | 4.18399242 | 0.345097716 | 0.51235 | 4.183992418 | 156.1517424 | 1493.251538 | 353.4511264 | 2.54832937 | TRUE | FALSE | FALSE |
| NR1H4 | 1460.2041 | 0.00172524 | 0.76465 | 5847.246507 | 107303.5917 | 430144.5525 | 81823002.1 | 2.02152E-08 | 0.08795 | 1269714.206 | 328207337.3 | 5284677316 | 1.78459E-05 | -4.748461847 | TRUE | FALSE | FALSE |
| NLRP8 | 1415.4293 | 0.0017471 | 0.61565 | 1415.42929 | 1327395.174 | 13575731.43 | 25147.4278 | 6.96896E-05 | 0.7759 | 249134.2971 | 276035316.5 | 32806726114 | 0.056285251 | -1.249605391 | TRUE | FALSE | FALSE |
| HCN3 | 1408.5316 | 0.001769 | 0.77295 | 39753.60936 | 16518934.21 | 217392318.7 | 227199957 | 7.9746E-09 | 0.0305 | 385073.0771 | 227199956.6 | 3552824196 | 6.19952E-06 | -5.207641647 | TRUE | FALSE | FALSE |
| EDN3 | 1398.46 | 0.001791 | 0.75035 | 5592.374889 | 6316421.913 | 69278377.09 | 15597.8039 | 0.000122988 | 0.8678 | 653449.9543 | 917073818.4 | 30691040575 | 0.089657496 | -1.047413395 | TRUE | FALSE | FALSE |
| SLC25A45 | 1377.4643 | 0.00181329 | 0.96495 | 3896365.325 | 2265198452 | 20936645108 | 1584328.61 | 9.76767E-07 | 0.087 | 37043.83915 | 6352966.326 | 102226574.5 | 0.000869431 | -3.060764917 | TRUE | FALSE | FALSE |
| COLEC10 | 1344.3352 | 0.00183612 | 0.00205 | 20.98094428 | 335.870278 | 1344.335224 | 45.3299871 | 0.06044477 | 0.367 | 45.32998708 | 726.4296772 | 2909.110521 | 29.65664255 | 1.472121983 | TRUE | FALSE | FALSE |
| POMC | 1341.5297 | 0.00185892 | 0.67925 | 5370.737489 | 810807.7769 | 7827444.037 | 126.089521 | 0.022554586 | 0.8019 | 1190.487754 | 184671.0181 | 1833109.46 | 10.63950221 | 1.026921309 | TRUE | FALSE | FALSE |
| KDM2B | 1325.2704 | 0.00188195 | 0.98505 | 15863350.92 | 18877504480 | 1.6275E+12 | 30.4056197 | 0.09136112 | 0.75135 | 2980.446553 | 321045.2635 | 3447360.164 | 43.58636285 | 1.63935063 | TRUE | FALSE | FALSE |
| METAP1D | 1315.4679 | 0.00190509 | 0.50565 | 1942.586923 | 313423.9454 | 3236120.402 | 115.218783 | 0.025350791 | 0.8512 | 2748.516028 | 445530.346 | 4621172.872 | 11.41713032 | 1.057556958 | TRUE | FALSE | FALSE |
| OR10Q1 | 1291.9379 | 0.00192862 | 0.50025 | 1291.937918 | 133841.9569 | 133841.9569 | 18.870313 | 0.138525934 | 0.3244 | 18.87031303 | 1801.866601 | 1801.866601 | 68.46404277 | 1.83546254 | TRUE | FALSE | FALSE |
| CFHR3 | 1282.8474 | 0.00195225 | 0.34335 | 130.3084512 | 12937.81178 | 133593.0057 | 1.80183965 | 0.506905089 | 0.9847 | 163.5310155 | 16385.11495 | 169960.2256 | 711.9653771 | 2.852458874 | TRUE | FALSE | FALSE |
| CNGB3 | 1279.3257 | 0.00197586 | 0.307 | 124.1030807 | 578860.033 | 25392551.4 | 18.8129974 | 0.139459795 | 0.50745 | 48.21552519 | 235210.9881 | 2619396.144 | 68.00222659 | 1.832523133 | TRUE | FALSE | FALSE |
| LRRIQ3 | 1271.1239 | 0.00199957 | 0.06675 | 133.9604066 | 12349.74202 | 25182.48692 | 23.8163009 | 0.114567824 | 0.3484 | 23.8163009 | 2144.230894 | 21084.77315 | 53.37201167 | 1.727313572 | TRUE | FALSE | FALSE |
| FUT2 | 1267.1255 | 0.00202326 | 0.15215 | 15.09342601 | 12042.63152 | 193373.5122 | 2.8846E-05 | 0.90050583 | 0.99355 | 0.14450127 | 37.16622219 | 267.1349373 | 43926977.17 | 7.642731318 | TRUE | TRUE | TRUE |
| SEMA3D | 1241.241 | 0.00204744 | 0.50855 | 1241.240996 | 79871.12296 | 320221.8924 | 4283.0978 | 0.000447021 | 0.68175 | 17155.09029 | 1103947.29 | 17749802.22 | 0.289799826 | -0.53790188 | TRUE | FALSE | FALSE |
| ZNF467 | 1239.9647 | 0.00207155 | 0.20235 | 309.5854683 | 19906.36681 | 79789.52346 | 29.3124008 | 0.093992479 | 0.2355 | 29.31240078 | 389.0931768 | 1558.501832 | 42.30171094 | 1.626357933 | TRUE | FALSE | FALSE |
| PABPC3 | 1204.8362 | 0.00209637 | 0.89025 | 125338.3939 | 14297385.13 | 128299316.5 | 195.408361 | 0.013785997 | 0.3226 | 20.13682541 | 1943.706009 | 1943.706009 | 6.165734988 | 0.789984854 | TRUE | FALSE | FALSE |
| DRC1 | 1193.7767 | 0.00212136 | 0.2311 | 125.1341013 | 11660.7694 | 23521.34499 | 23.0990242 | 0.11669628 | 0.19515 | 23.09902424 | 217.7716306 | 2103.312901 | 51.68082758 | 1.71332946 | TRUE | FALSE | FALSE |
| MUTYH | 1188.8403 | 0.00214638 | 0.8607 | 76261.4869 | 19651570.28 | 123170139.8 | 2920297753 | 5.80948E-10 | 0.4713 | 2920297753 | 6.90249E+12 | 1.99841E+14 | 4.07096E-07 | -6.390303607 | TRUE | FALSE | FALSE |
| APOL5 | 1180.8249 | 0.0021715 | 0.0038 | 12.6489159 | 1180.824877 | 1180.824877 | 0.21087973 | 0.849381463 | 0.0098 | 0.210879734 | 0.210879734 | 0.210879734 | 5599.518036 | 3.748150648 | TRUE | TRUE | TRUE |
| KCNRG | 1156.2992 | 0.00219713 | 0.7533 | 2358.690192 | 1243145.468 | 13331176.73 | 1424.20522 | 0.001699818 | 0.6255 | 14317.56792 | 16720242.09 | 185561874.1 | 0.811890859 | -0.090502348 | TRUE | FALSE | FALSE |
| STEAP3 | 1148.4363 | 0.00222286 | 0.32155 | 216.2966325 | 18402.87363 | 73694.93894 | 536738.477 | 3.32615E-06 | 0.06015 | 2699.04273 | 536738.4772 | 2150598.159 | 0.002139657 | -2.669655822 | TRUE | FALSE | FALSE |
| PACRG | 1143.5411 | 0.00224863 | 0.1947 | 47.51984298 | 4573.084213 | 46529.02661 | 0.20128025 | 0.851813035 | 1 | 0.804953262 | 0.804953262 | 0.804953262 | 5681.33803 | 3.75445063 | TRUE | TRUE | TRUE |
| LPA | 1125.1971 | 0.00227478 | 0.4196 | 346.284973 | 475411.2949 | 15869183.62 | 0.00065819 | 0.899660592 | 0.87895 | 0.005275387 | 0.365558495 | 1.461217658 | 1709542.427 | 6.232879884 | TRUE | TRUE | TRUE |
| SLC2A8 | 1105.7452 | 0.00230136 | 0.7674 | 10419.55217 | 3986009.245 | 40384471.91 | 108.063231 | 0.027306954 | 0.9452 | 39603.02837 | 41557966.93 | 442072095.5 | 10.23239088 | 1.009977122 | TRUE | FALSE | FALSE |
| ALDH3A1 | 1075.6224 | 0.00232869 | 0.71025 | 17204.60041 | 11142878.6 | 178230557.1 | 0.33484176 | 0.813995246 | 0.9911 | 613.3550632 | 243214.2887 | 2507920.301 | 3212.330448 | 3.506820214 | TRUE | TRUE | TRUE |
| SLC22A5 | 1054.3062 | 0.00235654 | 0.6698 | 3378.677061 | 148992.5148 | 597261.6343 | 7.05151172 | 0.2437471 | 0.8667 | 112.960232 | 1811.350578 | 1811.350578 | 149.5149162 | 2.174684522 | TRUE | FALSE | FALSE |
| SOX4 | 1044.6246 | 0.00238458 | 0.00415 | 10.30902643 | 1044.62457 | 1044.62457 | 13.3733269 | 0.167508766 | 0.31645 | 13.37332687 | 1356.919781 | 1356.919781 | 78.112543 | 1.892720777 | TRUE | FALSE | FALSE |
| L3MBTL1 | 1044.4481 | 0.00241252 | 0.0212 | 6.963340696 | 261.0802426 | 1044.448135 | 0.13113868 | 0.868625406 | 0.9841 | 10.81064824 | 986.4012749 | 3945.290011 | 7964.455137 | 3.90115607 | TRUE | TRUE | TRUE |
| PRKD3 | 1040.5083 | 0.00244049 | 0.5094 | 1040.508274 | 76907.0975 | 2713865.838 | 53.6930556 | 0.051080094 | 0.595 | 118.9865404 | 17364.16035 | 69543.72809 | 19.37882401 | 1.287327419 | TRUE | FALSE | FALSE |
| MGAM | 1032.0552 | 0.00246861 | 0.3422 | 115.7651882 | 263223.1615 | 5606315.149 | 0.01691489 | 0.894621912 | 0.5016 | 0.031721178 | 9.377976938 | 68.66710621 | 61014.58598 | 4.785433669 | TRUE | TRUE | TRUE |
| UBA7 | 1027.5548 | 0.00249677 | 0.9034 | 244412.7581 | 422567284.2 | 4536053234 | 27.3129636 | 0.101967404 | 0.8315 | 607.228567 | 237272.2331 | 2426738.084 | 37.62150546 | 1.57543617 | TRUE | FALSE | FALSE |
| CYP2J2 | 1018.8209 | 0.00252511 | 0.23005 | 110.5819654 | 9627.870984 | 38532.1465 | 12.8175331 | 0.171541119 | 0.1193 | 3.204760735 | 51.27690353 | 205.1859478 | 79.48650313 | 1.900293391 | TRUE | FALSE | FALSE |
| TMTC1 | 1017.7907 | 0.00255337 | 0.6251 | 4068.546587 | 1725634.406 | 18388429.52 | 82.867285 | 0.034987105 | 0.3519 | 20.72841793 | 812.183544 | 8154.689403 | 12.28217705 | 1.089275354 | TRUE | FALSE | FALSE |
| SAMD4A | 997.25141 | 0.0025822 | 0.8862 | 105082.662 | 11967665.39 | 133233328.5 | 126.22775 | 0.022402554 | 0.98255 | 132915.9521 | 158976299.3 | 1813654239 | 7.900413422 | 0.897649818 | TRUE | FALSE | FALSE |
| ABCB6 | 977.16721 | 0.0026116 | 0.29545 | 120.3896484 | 62716.99257 | 528659.5221 | 2.31362177 | 0.463500552 | 0.79185 | 73.21658533 | 18839.31407 | 153435.7078 | 422.353915 | 2.625676524 | TRUE | FALSE | FALSE |
| SIRPG | 974.99267 | 0.00264096 | 0.6332 | 974.9926656 | 99530.46487 | 1041258.596 | 1.59159291 | 0.527383868 | 0.96955 | 142.6630588 | 1402.156392 | 14117.65334 | 612.5892224 | 2.787169352 | TRUE | FALSE | FALSE |
| ALG14 | 971.9108 | 0.00267033 | 0.03365 | 41.18221751 | 971.9107962 | 3887.887099 | 0.36012565 | 0.805621354 | 1 | 0.360125652 | 0.360125652 | 0.360125652 | 2698.810233 | 3.431172348 | TRUE | TRUE | TRUE |
| ZNF433 | 965.53835 | 0.00269981 | 0.26035 | 104.3500136 | 9142.926544 | 88555.42236 | 2028.92606 | 0.001185533 | 0.3763 | 215.9066878 | 191923.0436 | 1929962.543 | 0.475886415 | -0.322496693 | TRUE | FALSE | FALSE |
| ACSM3 | 963.17951 | 0.00272927 | 0.1677 | 34.37568032 | 5148.582408 | 35238.92901 | 0.01237447 | 0.895965135 | 1 | 0.049486275 | 0.049486275 | 0.245228204 | 77836.01472 | 4.891180592 | TRUE | TRUE | TRUE |
| SERPINB4 | 958.43813 | 0.00275879 | 0.3629 | 239.7179733 | 24276.02712 | 97060.18182 | 3353.76403 | 0.000584984 | 0.61735 | 3353.764029 | 3832078.817 | 41894054.19 | 0.285779835 | -0.543968419 | TRUE | FALSE | FALSE |
| PGK2 | 949.78007 | 0.00278852 | 0.52155 | 949.7800703 | 34733.70989 | 47184.18454 | 0.51987097 | 0.747414537 | 0.6588 | 1.193219379 | 19.07223012 | 76.27896691 | 1826.953468 | 3.261727486 | TRUE | TRUE | TRUE |
| OBSL1 | 949.66369 | 0.00281814 | 0.23355 | 101.5939069 | 88633.11487 | 884133.6371 | 12965.612 | 0.000160199 | 0.4674 | 12965.61195 | 12748768.38 | 50967051.34 | 0.073244803 | -1.135223186 | TRUE | FALSE | FALSE |
| ZNF573 | 935.78084 | 0.00284816 | 0.1109 | 10.45093648 | 935.7808352 | 9176.769997 | 18.5428422 | 0.141017782 | 0.3172 | 2.009035621 | 175.4354173 | 175.4354173 | 50.46587913 | 1.702997843 | TRUE | FALSE | FALSE |
| SLC12A9 | 931.81403 | 0.00287822 | 0.5086 | 931.8140343 | 73016.7223 | 961195.1647 | 406.540425 | 0.006183991 | 0.5036 | 406.5404247 | 104427.539 | 418324.7237 | 2.292057512 | 0.360225511 | TRUE | FALSE | FALSE |
| SPTBN5 | 915.12319 | 0.0029088 | 0.9489 | 18299891.72 | 1.17599E+12 | 1.11485E+14 | 0.0396422 | 0.889030277 | 0.0382 | 0.000703328 | 0.039642203 | 0.307992157 | 23084.56975 | 4.363321784 | TRUE | TRUE | TRUE |
| NOXA1 | 911.51278 | 0.00293941 | 0.50475 | 911.512776 | 93848.86648 | 181214.7447 | 13.9418166 | 0.164504634 | 0.6904 | 13.94181657 | 1323.94938 | 1323.94938 | 65.37977111 | 1.815443396 | TRUE | FALSE | FALSE |
| DDO | 905.69188 | 0.00297014 | 0.6928 | 2874.662269 | 185170.8393 | 742650.3863 | 6.16828674 | 0.264420487 | 0.88645 | 98.84979072 | 1687.968789 | 9528.289676 | 146.8303784 | 2.166815918 | TRUE | FALSE | FALSE |
| MPZL2 | 888.57765 | 0.00300144 | 0.37865 | 362.7645618 | 365378.9891 | 3834088.919 | 14609.599 | 0.000142362 | 0.27325 | 2927.102041 | 1592034.088 | 17181820.66 | 0.060821495 | -1.21594291 | TRUE | FALSE | FALSE |
| PEPD | 884.31873 | 0.0030328 | 0.2911 | 131.986517 | 20638.93593 | 135105.6848 | 1475958.24 | 1.14478E-06 | 0.20815 | 63675.07655 | 23636286.42 | 243146619.8 | 0.000599149 | -3.222465247 | TRUE | FALSE | FALSE |
| HSD17B14 | 869.93923 | 0.00306464 | 0.112 | 88.57254899 | 3479.157478 | 35006.38963 | 0.56279652 | 0.727706682 | 0.98375 | 116.3311756 | 4591.762008 | 46424.61459 | 1545.743787 | 3.189137509 | TRUE | TRUE | TRUE |
| ARSK | 868.28559 | 0.00309644 | 0.4186 | 387.9557596 | 55680.85479 | 399580.0008 | 238910.883 | 8.19986E-06 | 0.2294 | 14887.69321 | 8632013.087 | 138646047.5 | 0.003634349 | -2.439573343 | TRUE | FALSE | FALSE |

| | | | | | | | | | | | | | | | | | |
|---|---|---|---|---|---|---|---|---|---|---|---|---|---|---|---|---|---|
| CTSZ | 850.43722 | 0.00312889 | 0.2341 | 88.63082281 | 8359.729108 | 21041.71535 | 0.44461231 | 0.775855074 | 0.7533 | 0.444612306 | 16.40345782 | 16.40345782 | 1912.761325 | 3.281660782 | TRUE | TRUE | TRUE |
| PRKAG3 | 850.06138 | 0.00316125 | 0.3056 | 212.3457297 | 13632.92011 | 54616.09953 | 88.5631996 | 0.033100224 | 0.51015 | 354.486978 | 7023.262417 | 91216.19549 | 9.598359001 | 0.98219699 | TRUE | FALSE | FALSE |
| BIRC6 | 845.70251 | 0.00319368 | 0.21945 | 211.2870848 | 6937.334303 | 62693.11687 | 0.19600076 | 0.853054247 | 0.98945 | 266.4416581 | 68299.18519 | 1095145.75 | 4314.792038 | 3.634959869 | TRUE | TRUE | TRUE |
| OPRL1 | 835.44386 | 0.00322646 | 0.1524 | 52.12243063 | 835.443864 | 2960.971663 | 6153.48201 | 0.00027762 | 0.3948 | 1536.018825 | 98831.3706 | 396227.1675 | 0.135767662 | -0.867203662 | TRUE | FALSE | FALSE |
| TTC23 | 830.57115 | 0.00325934 | 0.00215 | 9.464251001 | 87.59493482 | 830.5711546 | 0.02600693 | 0.892155964 | 0.01095 | 0.026006928 | 0.026006928 | 0.516310258 | 31936.5345 | 4.504287788 | TRUE | TRUE | TRUE |
| SERPINB10 | 810.50304 | 0.00329304 | 0.68505 | 4726.637459 | 1943647.376 | 20405106.5 | 5.01453369 | 0.305937772 | 0.85405 | 192.5079507 | 49316.43472 | 197184.5042 | 161.6307902 | 2.208524096 | TRUE | FALSE | FALSE |
| GBP7 | 804.03698 | 0.00332693 | 0.2289 | 83.45191151 | 7931.751971 | 27204.00524 | 1442.83749 | 0.001627016 | 0.02155 | 15.59444956 | 1442.83749 | 14382.47005 | 0.55726094 | -0.253941397 | TRUE | FALSE | FALSE |
| MSRA | 803.999 | 0.00336071 | 0.1479 | 82.98514432 | 3215.136377 | 31911.21669 | 57.8019204 | 0.048460379 | 0.2991 | 14.4534623 | 550.7750299 | 2203.196078 | 13.90955524 | 1.143313244 | TRUE | FALSE | FALSE |
| USP6 | 802.50818 | 0.00339444 | 0.51205 | 7341.554251 | 6407334.422 | 63866844.37 | 0.05131227 | 0.886336897 | 0.9897 | 32.03708363 | 24752.9013 | 237560.8032 | 15639.69205 | 4.194228197 | TRUE | TRUE | TRUE |
| CDK15 | 801.9442 | 0.00342809 | 0.36915 | 84.23951333 | 7816.816368 | 77971.90123 | 0.02556214 | 0.892276496 | 0.7492 | 0.223650008 | 2.00840282 | 2.00840282 | 31372.34205 | 4.496546941 | TRUE | TRUE | TRUE |
| EHHADH | 791.92212 | 0.0034621 | 0.26485 | 87.04788936 | 12678.99934 | 118241.5756 | 5.48375881 | 0.287657727 | 0.8477 | 196.7894018 | 28997.74507 | 273512.1142 | 144.4122818 | 2.15960413 | TRUE | FALSE | FALSE |
| BRICD5 | 786.99496 | 0.00349625 | 0.26245 | 196.6701722 | 18518.02574 | 74101.69387 | 1.78544904 | 0.509515608 | 0.88085 | 16.77920453 | 161.7352054 | 161.7352054 | 440.7826496 | 2.644224491 | TRUE | FALSE | FALSE |
| SPOCD1 | 778.97341 | 0.00353068 | 0.37175 | 82.19517724 | 7558.8864 | 75061.43767 | 0.22083831 | 0.847030682 | 0.2673 | 0.22083831 | 1.975133701 | 7.455593084 | 3527.347292 | 3.547448221 | TRUE | TRUE | TRUE |
| SERPINB11 | 755.40383 | 0.00356622 | 0.34865 | 303.7899205 | 30764.98456 | 80236.75145 | 1.23864974 | 0.56265331 | 0.8148 | 4.952936198 | 188.6747747 | 188.6747747 | 609.8607282 | 2.785230668 | TRUE | FALSE | FALSE |
| ZNF71 | 752.05861 | 0.00360182 | 0.74795 | 7515.460649 | 805096.6354 | 8621386.13 | 111930.266 | 1.6958E-05 | 0.0654 | 1078.761242 | 111930.2655 | 214222.0634 | 0.006718992 | -2.172695847 | TRUE | FALSE | FALSE |
| NDUFS7 | 739.43333 | 0.00363799 | 1 | 3.61607E+35 | 1.09608E+44 | 5.59574E+47 | 1.28064096 | 0.55700793 | 1 | 5.5862E+30 | 1.03803E+38 | 4.57949E+41 | 577.3931622 | 2.761471636 | TRUE | FALSE | FALSE |
| PLAT | 731.59151 | 0.00367448 | 0.6905 | 2929.080283 | 188264.7849 | 754509.877 | 5.31056625 | 0.294537029 | 0.5106 | 21.24405845 | 340.2170959 | 1362.003011 | 137.7614885 | 2.139127827 | TRUE | FALSE | FALSE |
| OGG1 | 724.28687 | 0.00371127 | 0.5633 | 1317.766918 | 186889.4207 | 1367051.176 | 2583974.95 | 7.02807E-07 | 0.08135 | 40044.1129 | 10369422.65 | 167113514.8 | 0.000280299 | -3.552377687 | TRUE | FALSE | FALSE |
| CUL9 | 716.79252 | 0.00374837 | 0.0951 | 11.19934497 | 1229.524839 | 11483.37474 | 0.30299846 | 0.824407072 | 0.5327 | 0.302998462 | 4.848188386 | 23.04673266 | 2365.663889 | 3.373953041 | TRUE | TRUE | TRUE |
| PPM1J | 707.53342 | 0.00378589 | 0.06645 | 7.733711899 | 707.5334201 | 707.5334201 | 94.6908165 | 0.030382358 | 0.5012 | 94.69081654 | 922.0635586 | 9198.102499 | 7.47203843 | 0.873439097 | TRUE | FALSE | FALSE |
| STON1 | 705.05922 | 0.00382344 | 0.5027 | 705.0592154 | 77924.35472 | 144760.4988 | 0.10610538 | 0.874352793 | 0.01575 | 0.106105381 | 0.106105381 | 2.099782954 | 6644.895962 | 3.822488186 | TRUE | TRUE | TRUE |
| ACOXL | 691.34527 | 0.00386171 | 0.51485 | 1262.541048 | 88615.63851 | 99915.00007 | 2.3060015 | 0.463861284 | 0.99035 | 745.1057976 | 47683.30227 | 108795.5827 | 299.8026122 | 2.476835413 | TRUE | FALSE | FALSE |
| TPO | 676.45817 | 0.00390081 | 0.52055 | 1200.830813 | 574971.8395 | 8019561.772 | 0.01131637 | 0.896324085 | 0.8787 | 0.045241453 | 0.180914901 | 0.887567583 | 59776.93687 | 4.776533657 | TRUE | TRUE | TRUE |
| NELL2 | 668.03868 | 0.00394034 | 0.0801 | 17.93440681 | 668.0386848 | 2673.674967 | 467.654754 | 0.004801452 | 0.1823 | 29.19651373 | 1872.341848 | 7498.135879 | 1.428486888 | 0.154876258 | TRUE | FALSE | FALSE |
| TTLL6 | 665.18189 | 0.00397992 | 0.6877 | 2661.781922 | 942554.6643 | 9187824.755 | 1.6401E+10 | 9.45507E-11 | 0.0395 | 3793363.671 | 6259428775 | 1.75805E+11 | 4.05586E-08 | -7.391917229 | TRUE | FALSE | FALSE |
| BTBD16 | 661.60666 | 0.00401962 | 0.07 | 7.288398664 | 661.6066594 | 661.6066594 | 1.15835989 | 0.571529344 | 0.50025 | 1.158359885 | 1.158359885 | 2.543334633 | 571.1581244 | 2.756756359 | TRUE | FALSE | FALSE |
| INPP5B | 660.07292 | 0.00405928 | 0.8648 | 107663.0029 | 366025743 | 16619697069 | 22.4746086 | 0.12066117 | 0.9418 | 23298.65633 | 29457520.69 | 332179919 | 29.36971804 | 1.467899777 | TRUE | FALSE | FALSE |
| NTHL1 | 652.612 | 0.00409933 | 0.2259 | 66.0033727 | 6606.750894 | 6606.750894 | 99.6020106 | 0.029335051 | 0.92735 | 10017.84762 | 11996052.4 | 50599590.82 | 6.552197043 | 0.816386949 | TRUE | FALSE | FALSE |
| GPR87 | 651.52075 | 0.00413933 | 0.7452 | 1328.954103 | 693908.7946 | 7418091.322 | 5.11423716 | 0.303045777 | 0.3778 | 2.23805809 | 20.45902848 | 20.45902848 | 127.3935357 | 2.105147391 | TRUE | FALSE | FALSE |
| A2ML1 | 632.96294 | 0.00418052 | 0.305 | 75.18333366 | 21856.81937 | 202852.7721 | 0.23491697 | 0.843467376 | 0.5103 | 0.23491697 | 8.082658311 | 71.2525544 | 2694.411308 | 3.430463892 | TRUE | TRUE | TRUE |
| CFHR5 | 621.51441 | 0.00422243 | 0.8388 | 199148.0245 | 623018661.7 | 6709270986 | 1403064.48 | 1.20382E-06 | 0.2274 | 139296.2304 | 151812607.7 | 17845894602 | 0.000442969 | -3.353626424 | TRUE | FALSE | FALSE |
| POU6F2 | 607.961 | 0.00426526 | 0.8067 | 65296.49058 | 21117415.29 | 967143414.6 | 48.6389013 | 0.056065368 | 0.64735 | 119.6550373 | 12456.64867 | 132242.3538 | 12.49948048 | 1.096891963 | TRUE | FALSE | FALSE |
| CXCL17 | 601.04691 | 0.0043085 | 0.12665 | 6.284613874 | 601.0469057 | 601.0469057 | 0.76990023 | 0.632463529 | 1 | 0.769900234 | 0.769900234 | 0.769900234 | 780.6815469 | 2.892473914 | TRUE | TRUE | TRUE |
| CYP3A43 | 598.05414 | 0.00435185 | 0.06725 | 17.657383 | 1131.084 | 4527.946711 | 0.83757722 | 0.610750774 | 0.69025 | 1.906693001 | 71.17874333 | 71.17874333 | 714.0286592 | 2.853715644 | TRUE | TRUE | TRUE |
| FLII | 595.16777 | 0.00439529 | 0.1325 | 15.99033973 | 1418.72947 | 5675.928967 | 0.20283068 | 0.851485928 | 0.7608 | 0.811101561 | 7.429664003 | 36.02333178 | 2934.308332 | 3.467505747 | TRUE | TRUE | TRUE |
| TPRG1L | 594.90501 | 0.00443861 | 0.64095 | 1470.182563 | 154330.447 | 617262.4937 | 81.0841635 | 0.036342366 | 0.50215 | 196.6459259 | 7820.161532 | 79652.66694 | 7.336882929 | 0.865511589 | TRUE | FALSE | FALSE |
| HADH | 576.18828 | 0.00448337 | 0.5093 | 576.1882763 | 93848.2117 | 375404.8678 | 87.7337113 | 0.03347132 | 0.31305 | 8.99629943 | 877.0060617 | 3506.602771 | 6.56746726 | 0.817397916 | TRUE | FALSE | FALSE |
| C3orf20 | 559.51632 | 0.00452948 | 0.36395 | 59.87165397 | 5353.868818 | 52426.53423 | 1.42849974 | 0.543031496 | 0.6941 | 1.428499735 | 119.7671971 | 119.7671971 | 391.6810793 | 2.592932593 | TRUE | FALSE | FALSE |
| SNCG | 559.09857 | 0.00457548 | 0.37595 | 139.6580351 | 8967.234554 | 14840.43651 | 0.72915788 | 0.652012784 | 0.81415 | 11.66717356 | 46.68748433 | 186.8717995 | 766.7729859 | 2.884666804 | TRUE | TRUE | TRUE |
| SARDH | 555.7152 | 0.00462165 | 0.2065 | 26.19630249 | 5312.972931 | 84938.84188 | 1.41691763 | 0.543835856 | 0.85495 | 22.64139034 | 3327.64186 | 28102.47752 | 392.2000719 | 2.593507669 | TRUE | FALSE | FALSE |
| SIGLEC5 | 555.11404 | 0.00466772 | 0.3503 | 57.27033327 | 5512.179125 | 56041.11035 | 78.9568988 | 0.036929394 | 0.3495 | 78.95689878 | 7695.742669 | 78733.60712 | 7.030595786 | 0.846992129 | TRUE | FALSE | FALSE |
| P2RY11 | 553.15876 | 0.00471383 | 0.13385 | 55.5821578 | 553.1587592 | 553.1587592 | 5.95059385 | 0.271042067 | 0.9398 | 231.0847102 | 9206.351836 | 36843.307 | 92.95858074 | 1.968289484 | TRUE | FALSE | FALSE |
| ZNF813 | 542.54585 | 0.0047608 | 0.19045 | 55.78631936 | 542.5458515 | 5405.458735 | 0.11158738 | 0.872964738 | 0.01595 | 0.111587383 | 0.111587383 | 2.207937958 | 4862.071657 | 3.686821355 | TRUE | TRUE | TRUE |
| ZNF488 | 540.44003 | 0.00480782 | 0.95395 | 55928.15351 | 67786850.69 | 8701848381 | 70707.3322 | 2.69346E-05 | 0.5029 | 748435.1292 | 88899593.03 | 11533619552 | 0.007643338 | -2.116716939 | TRUE | FALSE | FALSE |
| SLC14A2 | 539.50465 | 0.00485478 | 0.0366 | 6.015514146 | 56.28378062 | 539.504651 | 0.061779 | 0.88437567 | 0.8789 | 1.220577874 | 45.08110091 | 45.08110091 | 8732.816885 | 3.941154354 | TRUE | TRUE | TRUE |
| FAM98C | 533.57401 | 0.00490218 | 0.06515 | 13.89606818 | 533.5740092 | 533.5740092 | 1.86184896 | 0.502549807 | 0.8706 | 17.50723867 | 168.8495655 | 168.8495655 | 286.5828656 | 2.457250221 | TRUE | FALSE | FALSE |
| NOC3L | 531.9022 | 0.0049496 | 0.8663 | 50003.9527 | 9902084.384 | 577217252.4 | 11.2431712 | 0.184700863 | 0.98075 | 9597.088004 | 10083801.38 | 107105556 | 47.30891282 | 1.674942968 | TRUE | FALSE | FALSE |
| FAM217A | 529.01805 | 0.00499715 | 0.0648 | 5.905946616 | 529.0180489 | 529.0180489 | 0.11090202 | 0.873157538 | 1 | 0.110902017 | 0.110902017 | 2.202147012 | 4770.139123 | 3.678531046 | TRUE | TRUE | TRUE |
| MAPK12 | 519.19681 | 0.00504556 | 0.34885 | 53.76756609 | 5136.10318 | 52021.06447 | 74.5945631 | 0.038547503 | 0.6571 | 74.59456313 | 7221.848103 | 73637.67939 | 6.960250045 | 0.842624842 | TRUE | FALSE | FALSE |
| ENPP3 | 514.92525 | 0.00509427 | 0.29365 | 55.63732459 | 19526.93237 | 78117.66663 | 6.02945738 | 0.268215695 | 0.88615 | 286.8264077 | 73457.35238 | 704405.2537 | 85.40158966 | 1.931465955 | TRUE | FALSE | FALSE |
| PRAMEF4 | 514.07113 | 0.00514291 | 0.85405 | 5225.265204 | 579994.3731 | 6326752.829 | 0.69022571 | 0.670183459 | 1 | 63.44842555 | 6454.346667 | 6454.346667 | 744.7869944 | 2.872032084 | TRUE | TRUE | TRUE |
| ECT2L | 507.84721 | 0.00519207 | 0.92745 | 456288.1605 | 1991981541 | 21374303965 | 11511.4323 | 0.000183336 | 0.83475 | 1133162.177 | 1300019523 | 14181307650 | 0.04411677 | -1.355396295 | TRUE | FALSE | FALSE |
| TRIM59 | 504.43515 | 0.00524145 | 0.63035 | 4825.798252 | 1214724.948 | 12732001.19 | 907.337398 | 0.002627633 | 0.36495 | 226.8673857 | 21814.4014 | 87244.89927 | 0.555951013 | -0.254963474 | TRUE | FALSE | FALSE |
| CTTNBP2NL | 501.94688 | 0.00529094 | 0.000025 | 0.425845499 | 0.425845499 | 8.421012707 | 0.47091435 | 0.766749557 | 1 | 0.470914348 | 0.470914348 | 9.324855025 | 1065.898457 | 3.027715833 | TRUE | TRUE | TRUE |
| INSL6 | 496.6536 | 0.00534087 | 0.50025 | 496.6536049 | 50036.70802 | 130139.3933 | 1.89912909 | 0.499077326 | 0.87495 | 18.02970395 | 175.5611079 | 175.5611079 | 261.5165063 | 2.417499106 | TRUE | FALSE | FALSE |
| CDRT15 | 493.96438 | 0.00539095 | 0.0662 | 50.04283386 | 4997.801458 | 4997.801458 | 0.67419464 | 0.67763074 | 1 | 0.674194644 | 0.674194644 | 0.674194644 | 732.6732517 | 2.864910337 | TRUE | TRUE | TRUE |
| MOCS1 | 490.6863 | 0.00544124 | 0.7407 | 7868.174631 | 1148001.649 | 5678411.706 | 59.4001034 | 0.047298376 | 0.51175 | 237.7453616 | 8063.277196 | 61163.90109 | 8.260697689 | 0.917016729 | TRUE | FALSE | FALSE |
| SCRT2 | 490.06648 | 0.00549145 | 0.31865 | 49.50812887 | 4972.44506 | 4972.44506 | 465.104865 | 0.004864196 | 0.34355 | 465.1048651 | 48839.6337 | 518603.8567 | 1.053668799 | 0.02270412 | TRUE | FALSE | FALSE |
| CHRNG | 485.15095 | 0.00554207 | 0.15095 | 5.280916614 | 485.1509466 | 4815.199031 | 12.9026061 | 0.170195586 | 0.5055 | 12.90260606 | 1214.694333 | 2463.670098 | 37.60100436 | 1.575199445 | TRUE | FALSE | FALSE |
| DPRX | 474.16182 | 0.00559386 | 0.18765 | 48.69295277 | 474.1618189 | 4730.139177 | 0.87186277 | 0.602675184 | 0.5038 | 0.871862771 | 0.871862771 | 0.871862771 | 543.849141 | 2.735478447 | TRUE | TRUE | TRUE |
| PNKD | 471.62545 | 0.00564579 | 0.65 | 1501.732783 | 281957.7338 | 1941465.015 | 0.05479957 | 0.885695341 | 0.96255 | 7.829369306 | 501.2873066 | 4598.773568 | 8606.371727 | 3.9348201 | TRUE | TRUE | TRUE |
| PABPN1L | 449.35246 | 0.00570044 | 0.34375 | 46.73215449 | 4426.397589 | 44643.61605 | 0.88385724 | 0.60089333 | 0.0046 | 0.883857241 | 0.883857241 | 0.883857241 | 508.3993708 | 2.706205004 | TRUE | TRUE | TRUE |
| NIPSNAP3A | 447.78102 | 0.00575513 | 0.8271 | 4389.054868 | 4751608.402 | 51023031.5 | 0.97512637 | 0.59077177 | 0.74765 | 0.975126375 | 9.005083172 | 9.005083172 | 459.2030605 | 2.662004774 | TRUE | TRUE | TRUE |
| TGM5 | 439.85965 | 0.00581075 | 0.21785 | 27.40170769 | 7067.791602 | 34725.24544 | 4.60117012 | 0.323972068 | 0.45535 | 4.601170115 | 124.1886172 | 1186.321961 | 95.59734636 | 1.980445837 | TRUE | FALSE | FALSE |
| LRRC8E | 437.51711 | 0.00586653 | 0.1245 | 45.12142841 | 4346.060446 | 44201.67027 | 0.09317302 | 0.877511638 | 0.5102 | 0.846209269 | 0.846209269 | 16.62381561 | 4695.748851 | 3.671704861 | TRUE | TRUE | TRUE |
| FSCB | 436.65413 | 0.00592225 | 0.9674 | 41706759.41 | 52508439205 | 6.76781E+12 | 9993.50386 | 0.000196972 | 0.50705 | 9993.503857 | 1000331.15 | 10347521.12 | 0.043693798 | -1.359580208 | TRUE | FALSE | FALSE |
| SCFD1 | 434.89437 | 0.00597806 | 0.8385 | 1742.095594 | 112146.715 | 553802.51 | 96477.2956 | 2.0562E-05 | 0.58585 | 96477.29559 | 24955800.28 | 401970229.5 | 0.004507738 | -2.346041337 | TRUE | FALSE | FALSE |
| SLFNL1 | 432.36896 | 0.00603405 | 0.1923 | 45.14829166 | 432.368959 | 4241.904561 | 0.09301429 | 0.877549599 | 1 | 0.834683388 | 0.834683388 | 1.842792434 | 4648.414425 | 3.66730484 | TRUE | TRUE | TRUE |
| PSPN | 431.62488 | 0.00608998 | 0.1906 | 43.76716219 | 4363.094174 | 45180.84174 | 0.59353892 | 0.714591501 | 1 | 2.374031286 | 22.3586407 | 22.3586407 | 727.2056881 | 2.861657267 | TRUE | TRUE | TRUE |
| PCDHB1 | 431.07051 | 0.00614581 | 0.0671 | 11.6611368 | 1020.877515 | 4083.593036 | 0.06945557 | 0.882572671 | 0.8776 | 0.616166343 | 5.610409603 | 12.13579192 | 6206.427847 | 3.792841221 | TRUE | TRUE | TRUE |
| PARD6G | 430.20764 | 0.0062016 | 0.1957 | 430.207637 | 4356.703917 | 45185.7952 | 47.5879622 | 0.058110533 | 0.8118 | 47.58796221 | 4774.528573 | 49586.97434 | 9.040261803 | 0.956181008 | TRUE | FALSE | FALSE |
| TMEM161A | 428.03675 | 0.00625754 | 0.34705 | 107.0157885 | 4202.322173 | 42265.04059 | 14.9146865 | 0.159205823 | 0.6896 | 14.91468654 | 1418.735566 | 1418.735566 | 28.69901065 | 1.457866925 | TRUE | FALSE | FALSE |
| FBXO24 | 426.82112 | 0.00631348 | 0.2134 | 45.70608764 | 1708.206965 | 16361.60693 | 4.15728064 | 0.346135423 | 0.50685 | 4.157280643 | 66.54227779 | 66.6978133 | 102.6683439 | 2.011436557 | TRUE | FALSE | FALSE |

| | | | | | | | | | | | | | | | | | |
|---|---|---|---|---|---|---|---|---|---|---|---|---|---|---|---|---|---|
| SPATA4 | 423.18741 | 0.00636978 | 0.3674 | 423.1874104 | 43794.36354 | 461298.72 | 49336.8111 | 3.73483E-05 | 0.5025 | 49336.81114 | 5623786.08 | 77471048.44 | 0.008577519 | -2.066638337 | TRUE | FALSE | FALSE |
| GSTO2 | 422.39902 | 0.00642603 | 0.18755 | 44.10890353 | 1036.712352 | 4146.309575 | 137.350073 | 0.020340758 | 0.3118 | 14.27253245 | 1354.862059 | 1354.862059 | 3.075346176 | 0.487894009 | TRUE | FALSE | FALSE |
| RARA | 415.23947 | 0.0064832 | 0.01335 | 11.13395423 | 415.2394736 | 880.7270653 | 0.62324508 | 0.701963025 | 0.5068 | 0.623245082 | 2.491464522 | 2.491464522 | 666.2539115 | 2.823639772 | TRUE | TRUE | TRUE |
| EPN3 | 414.82452 | 0.00654025 | 0.1143 | 4.689856643 | 414.8245246 | 4043.209869 | 9.4157797 | 0.206402392 | 0.13525 | 1.024889884 | 9.415779695 | 9.415779695 | 44.05631165 | 1.644008136 | TRUE | FALSE | FALSE |
| MPHOSPH1( | 411.3125 | 0.00659768 | 1 | 8.65559E+23 | 9.3846E+30 | 2.34551E+33 | 0.64939955 | 0.690024538 | 1 | 6.95683E+25 | 3.69173E+32 | 1.59184E+36 | 633.3735581 | 2.801659929 | TRUE | TRUE | TRUE |
| SBK1 | 409.82455 | 0.00665516 | 0.8305 | 42983.48544 | 55048316.19 | 625099738.4 | 0.51983445 | 0.747454714 | 1 | 49196.5206 | 5698595.124 | 63451589.93 | 788.3751195 | 2.89673291 | TRUE | TRUE | TRUE |
| SGCZ | 403.26341 | 0.00671352 | 0.0229 | 25.18074566 | 403.2634126 | 1614.401493 | 0.64503318 | 0.692541367 | 0.6865 | 0.645033178 | 10.31718394 | 10.31718394 | 625.1824344 | 2.796006767 | TRUE | TRUE | TRUE |
| CHMP4A | 400.3937 | 0.00677216 | 0.63825 | 400.3936983 | 39775.46807 | 410499.2685 | 504291.326 | 3.61798E-06 | 0.28875 | 48725.99077 | 57763645.94 | 3988205485 | 0.000793973 | -3.100194264 | TRUE | FALSE | FALSE |
| SLC34A1 | 397.61625 | 0.00683109 | 0.01755 | 6.20562286 | 397.6162543 | 1591.868594 | 0.20495061 | 0.850813705 | 0.63135 | 0.371331239 | 3.276779554 | 13.10717211 | 1940.058878 | 3.28781491 | TRUE | TRUE | TRUE |
| ACAA1 | 394.56311 | 0.00689035 | 0.2937 | 98.56971306 | 6326.850103 | 25344.60743 | 11.4677985 | 0.183346324 | 0.5037 | 45.88725101 | 735.2605482 | 2944.275509 | 34.40617746 | 1.536636425 | TRUE | FALSE | FALSE |
| TSNAXIP1 | 392.16754 | 0.00694984 | 0.8975 | 38195.09843 | 43276246.79 | 5220804515 | 8.6192E+10 | 2.1564E-11 | 0.2763 | 677508983.1 | 1.47195E+13 | 2.1314E+15 | 4.54994E-09 | -8.341993972 | TRUE | FALSE | FALSE |
| ZNF616 | 385.75927 | 0.00701025 | 0.1215 | 40.26562045 | 3786.093752 | 38046.35379 | 6.73222932 | 0.250926979 | 0.139 | 6.732229323 | 63.56526514 | 63.56526514 | 57.30037641 | 1.758157475 | TRUE | FALSE | FALSE |
| RPL28 | 385.62921 | 0.0070705 | 0.65715 | 385.6292149 | 39868.43212 | 420357.1419 | 0.56870048 | 0.72528177 | 1 | 5.331159322 | 5.331159322 | 5.331159322 | 678.0884234 | 2.83128633 | TRUE | TRUE | TRUE |
| UBR4 | 382.81993 | 0.00713106 | 0.05725 | 4.235678457 | 382.8199347 | 2873.83723 | 0.57302456 | 0.72325044 | 0.3167 | 0.062043237 | 2.042830453 | 18.86735876 | 668.0689862 | 2.824821311 | TRUE | TRUE | TRUE |
| SULT1C2 | 378.94153 | 0.00719214 | 0.5065 | 378.9415261 | 7539.674977 | 73075.36071 | 1696.18609 | 0.001380496 | 0.1353 | 146.2619846 | 6796.260254 | 109191.2945 | 0.223407991 | -0.650901296 | TRUE | FALSE | FALSE |
| TRIM15 | 378.43152 | 0.00725312 | 0.5064 | 378.4315174 | 37257.2412 | 125745.2534 | 60.5338841 | 0.046375066 | 0.19665 | 60.53388412 | 584.9018648 | 5789.679505 | 6.2515651 | 0.795988758 | TRUE | FALSE | FALSE |
| AMZ1 | 373.8683 | 0.00731475 | 0.50815 | 3543.491811 | 340978.683 | 3458367.881 | 632.352892 | 0.003352851 | 0.9077 | 59463.12385 | 64144045.73 | 686680627.3 | 0.591233639 | -0.228240864 | TRUE | FALSE | FALSE |
| PDE1B | 371.85602 | 0.00737657 | 0.71025 | 1487.422904 | 391947.7838 | 1567789.886 | 0.15527834 | 0.863223814 | 1 | 133.1089466 | 1329.658969 | 13606.56746 | 2394.770762 | 3.379263947 | TRUE | TRUE | TRUE |
| CD36 | 371.77866 | 0.00743822 | 0.0761 | 0.176996175 | 794.8593846 | 27691.21917 | 4.0695E-06 | 0.90090062 | 0.58765 | 7.49663E-06 | 0.003460224 | 0.028022121 | 91358256.05 | 7.960747801 | TRUE | TRUE | TRUE |
| KRT20 | 366.50945 | 0.00750068 | 0.69135 | 1467.906996 | 17125.39176 | 68825.45338 | 41.6953839 | 0.066027266 | 0.8314 | 1804.964952 | 29000.57 | 116288.7819 | 8.790168511 | 0.943997201 | TRUE | FALSE | FALSE |
| MESP1 | 364.45534 | 0.00756335 | 0.0952 | 22.77888331 | 889.225956 | 3557.315254 | 1027.04461 | 0.002357557 | 0.187 | 105.0279584 | 2573.92626 | 10294.52388 | 0.354858332 | -0.449944993 | TRUE | FALSE | FALSE |
| ZMAT3 | 363.66459 | 0.00762598 | 0.49815 | 37.07505784 | 3656.389893 | 37659.78476 | 4087.56545 | 0.000467811 | 0.03195 | 405.6547155 | 4087.565446 | 42193.44275 | 0.088968506 | -1.050763701 | TRUE | FALSE | FALSE |
| CLIP4 | 358.77575 | 0.00768938 | 0.4171 | 208.2327442 | 31769.23241 | 127159.3821 | 47222.2715 | 3.85524E-05 | 0.3298 | 4612.029888 | 1978151.787 | 55961422.24 | 0.007597596 | -2.119323792 | TRUE | FALSE | FALSE |
| CLCN7 | 358.73817 | 0.0077526 | 0.3321 | 89.74317604 | 9102.219831 | 57080.6902 | 3760927.44 | 4.32631E-07 | 0.1661 | 54758.47878 | 60098001.33 | 670831801.9 | 9.53856E-05 | -4.020517367 | TRUE | FALSE | FALSE |
| IGSF10 | 357.38138 | 0.0078159 | 0.89485 | 2874639.312 | 5.83379E+11 | 1.05131E+14 | 236.257362 | 0.010624323 | 0.2292 | 2.769819532 | 21733.66901 | 2150419.907 | 1.512678301 | 0.179746577 | TRUE | FALSE | FALSE |
| ADAMTS12 | 356.80696 | 0.00787913 | 0.1825 | 22.27262285 | 1428.653359 | 12860.59899 | 0.09900398 | 0.876217689 | 0.8662 | 3.449498126 | 221.021139 | 884.8641762 | 3603.965697 | 3.556780649 | TRUE | TRUE | TRUE |
| ZSCAN32 | 356.39444 | 0.00794225 | 0.0037 | 3.993158803 | 356.3944408 | 356.3944408 | 0.64382696 | 0.69308802 | 0.87485 | 5.907920041 | 55.60548381 | 55.60548381 | 553.5562568 | 2.743161764 | TRUE | TRUE | TRUE |
| PUSL1 | 353.16133 | 0.00800584 | 0.18995 | 88.29494727 | 3431.659989 | 34160.41302 | 11.5083609 | 0.182669019 | 0.93165 | 109.5605639 | 109397.6495 | 109397.6495 | 30.68737013 | 1.486959671 | TRUE | FALSE | FALSE |
| SFI1 | 348.29964 | 0.00807023 | 0.29335 | 348.2996411 | 30350.00394 | 589548.0186 | 7148.94929 | 0.000248962 | 0.61395 | 7148.94929 | 6750269.778 | 69129943.14 | 0.048720396 | -1.31228919 | TRUE | FALSE | FALSE |
| LIPT1 | 347.11768 | 0.00813468 | 0.89 | 34903.18194 | 3848540.912 | 41780394.46 | 503961.078 | 3.76363E-06 | 0.5024 | 503961.078 | 58652934.55 | 4482380276 | 0.000688779 | -3.161920266 | TRUE | FALSE | FALSE |
| IP6K3 | 346.53319 | 0.00819906 | 0.06655 | 3.926643161 | 346.5331919 | 346.5331919 | 0.07815204 | 0.880826432 | 0.01105 | 0.078152043 | 0.078152043 | 1.549335401 | 4434.089966 | 3.646804501 | TRUE | TRUE | TRUE |
| ZNF451 | 344.67266 | 0.00826364 | 0.10875 | 36.46792739 | 3337.340598 | 33086.12015 | 0.7250311 | 0.654370369 | 0.50065 | 0.725031099 | 0.725031099 | 0.725031099 | 475.3901742 | 2.677050201 | TRUE | TRUE | TRUE |
| SPDYA | 343.91374 | 0.0083282 | 0.12735 | 35.02430744 | 343.9137394 | 343.9137394 | 0.48081836 | 0.762838142 | 1 | 0.480818362 | 0.480818362 | 0.480818362 | 715.2674826 | 2.854468482 | TRUE | TRUE | TRUE |
| GATA5 | 340.41784 | 0.00839331 | 0.1481 | 21.25670384 | 340.4178404 | 1362.803247 | 6.89600568 | 0.247017819 | 0.9664 | 441.8728532 | 7084.447193 | 28377.41695 | 49.36449536 | 1.693414702 | TRUE | FALSE | FALSE |
| ELP5 | 337.76565 | 0.0084588 | 0.0701 | 3.827128187 | 337.7656492 | 337.7656492 | 0.07200705 | 0.882025296 | 1 | 0.641115167 | 5.858705287 | 5.858705287 | 4690.730424 | 3.671240475 | TRUE | TRUE | TRUE |
| GALNT4 | 336.74167 | 0.00852433 | 0.0655 | 21.03170619 | 336.7416731 | 336.7416731 | 2.4871716 | 0.447344586 | 0.75915 | 2.487171603 | 9.950968 | 49.22818402 | 135.3914111 | 2.131591115 | TRUE | FALSE | FALSE |
| PKD2L2 | 336.34813 | 0.00858975 | 0.0188 | 3.858313129 | 142.3091477 | 1344.874426 | 62.9378357 | 0.044331498 | 0.50505 | 62.93783574 | 5871.032917 | 23476.15418 | 5.34413242 | 0.727877211 | TRUE | FALSE | FALSE |
| DERL3 | 335.91981 | 0.00865507 | 0.92855 | 84533.8229 | 38132444.02 | 418686988.1 | 1353.4281 | 0.001774082 | 0.1924 | 136.8724692 | 1353.428096 | 13709.86919 | 0.248199227 | -0.605199575 | TRUE | FALSE | FALSE |
| BHLHE22 | 333.86256 | 0.00872066 | 0.06325 | 8.602119727 | 333.8625608 | 333.8625608 | 21609.5021 | 9.2686E-05 | 0.36305 | 2078.637873 | 230025.6444 | 2505509.102 | 0.015449803 | -1.811077041 | TRUE | FALSE | FALSE |
| C15orf48 | 331.6856 | 0.00878654 | 0.77475 | 3329.515419 | 360468.3775 | 360468.3775 | 4.10927781 | 0.349744576 | 0.98215 | 386.7808068 | 40188.35172 | 424983.5166 | 80.71627565 | 1.906961115 | TRUE | FALSE | FALSE |
| CYB561D1 | 325.43553 | 0.00885366 | 0.00025 | 1.045459138 | 9.577621686 | 9.577621686 | 0.31827155 | 0.819303053 | 0.00125 | 0.318271555 | 0.318271555 | 0.318271555 | 1022.509013 | 3.009667145 | TRUE | TRUE | TRUE |
| RAI1 | 322.89916 | 0.00892117 | 0.00005 | 0.278902177 | 0.564634312 | 5.32391202 | 0.16990295 | 0.859710733 | 0.9421 | 1.601925748 | 61.98536309 | 297.5298417 | 1900.491762 | 3.278865991 | TRUE | TRUE | TRUE |
| CNOT3 | 322.77814 | 0.00898852 | 0.1337 | 32.60269585 | 322.7781391 | 639.9326723 | 0.44568708 | 0.775572764 | 1 | 0.445687081 | 0.445687081 | 8.767622803 | 724.2259269 | 2.859874068 | TRUE | TRUE | TRUE |
| LRRK2 | 321.61054 | 0.00905594 | 0.73155 | 3737.347129 | 2273157.375 | 54807002.05 | 3195.49138 | 0.000672198 | 0.0957 | 36.67290998 | 12779.63142 | 123529.9839 | 0.100645096 | -0.997207381 | TRUE | FALSE | FALSE |
| KLHL30 | 317.42785 | 0.00912417 | 0.4576 | 184.7893531 | 11845.16886 | 47429.2489 | 0.92095386 | 0.596319492 | 0.1466 | 0.920953861 | 3.684197914 | 7.800814609 | 344.6729078 | 2.537407148 | TRUE | TRUE | TRUE |
| MYO1H | 316.89989 | 0.00919233 | 0.77285 | 5080.01418 | 697199.7269 | 6116709.972 | 36209.2663 | 5.18857E-05 | 0.3754 | 9043.088679 | 1290696.818 | 5170633.208 | 0.0087519 | -2.057897635 | TRUE | FALSE | FALSE |
| DPY19L4 | 316.51861 | 0.00926039 | 0.367 | 316.5186094 | 28816.84408 | 284725.247 | 123.192516 | 0.023014747 | 0.80305 | 11427.60795 | 4649857.993 | 48488938.52 | 2.569300642 | 0.409814925 | TRUE | FALSE | FALSE |
| AQP12B | 315.84725 | 0.00932842 | 0.3128 | 32.18427163 | 3177.185919 | 3177.185919 | 3.9237607 | 0.360338298 | 0.8736 | 8.330001491 | 370.7006215 | 370.7006215 | 80.49605285 | 1.905774585 | TRUE | FALSE | FALSE |
| ASPSCR1 | 314.66253 | 0.00939654 | 0.19455 | 33.10145788 | 314.6625349 | 3064.352939 | 406.865351 | 0.006116271 | 0.74745 | 3982.062636 | 409186.3081 | 4291580.997 | 0.773382481 | -0.11160567 | TRUE | FALSE | FALSE |
| CPZ | 313.13087 | 0.00946484 | 0.90005 | 114050.577 | 116926938.4 | 3301162372 | 12.4012539 | 0.176569006 | 0.3495 | 1.376174008 | 114.4895768 | 1082.251193 | 25.24993588 | 1.40226028 | TRUE | FALSE | FALSE |
| TMX4 | 310.35901 | 0.00953363 | 0.12855 | 3.295711373 | 310.3590099 | 310.3590099 | 0.41163696 | 0.787785707 | 1 | 3.854271433 | 37.0388 | 37.0388 | 753.9629354 | 2.877349997 | TRUE | TRUE | TRUE |
| TMED3 | 309.77789 | 0.00960237 | 0.64145 | 3105.644265 | 335376.9801 | 3558186.751 | 3793.12991 | 0.000500887 | 0.11645 | 376.7165317 | 39124.88763 | 413110.8002 | 0.081668148 | -1.087947294 | TRUE | FALSE | FALSE |
| KISS1R | 309.29886 | 0.00967103 | 0.35435 | 309.2988576 | 32081.77807 | 339953.1183 | 4.2511252 | 0.340911851 | 0.2579 | 4.251125204 | 40.92587177 | 40.92587177 | 72.75693911 | 1.86187442 | TRUE | FALSE | FALSE |
| EBPL | 305.31482 | 0.00974051 | 0.71075 | 1221.015218 | 194966.4133 | 1992524.655 | 11.4486308 | 0.18368451 | 0.7416 | 76.80731682 | 2930.67941 | 7163.166341 | 26.66823883 | 1.425994336 | TRUE | FALSE | FALSE |
| COLGALT1 | 301.07213 | 0.0098109 | 0.92 | 65513.3412 | 42122972.52 | 761928788.4 | 0.0661388 | 0.88331565 | 0.99925 | 154.7162468 | 21786.86239 | 137921.1614 | 4552.125415 | 3.658214219 | TRUE | TRUE | TRUE |
| PCSK9 | 300.72692 | 0.00988117 | 0.1559 | 15.94327783 | 1203.780093 | 5741.193002 | 0.52908511 | 0.743451837 | 0.3283 | 0.132264845 | 2.116972178 | 2.607095745 | 568.3904501 | 2.754646772 | TRUE | TRUE | TRUE |
| OR6F1 | 299.30425 | 0.00995163 | 0.7808 | 2992.492045 | 321407.1976 | 609339.6636 | 319735.01 | 5.81505E-06 | 0.2731 | 30513.43523 | 39866024.23 | 446728072.4 | 0.000936101 | -3.028677309 | TRUE | FALSE | FALSE |
| ZNF69 | 298.83095 | 0.01002201 | 0.3128 | 30.73243141 | 2978.449213 | 2978.449213 | 4.04086725 | 0.35457563 | 0.50035 | 4.040867254 | 4.040867254 | 4.040867254 | 73.95218105 | 1.868950987 | TRUE | FALSE | FALSE |
| HAO2 | 297.72607 | 0.01009248 | 0.0647 | 3.336958942 | 297.726066 | 297.726066 | 0.06723895 | 0.883054406 | 1 | 0.06723895 | 0.06723895 | 0.06723895 | 4427.880926 | 3.646195933 | TRUE | TRUE | TRUE |
| RNF186 | 294.61915 | 0.01016359 | 0.347 | 294.6191545 | 6606.094917 | 324964.2168 | 3174.71796 | 0.000684883 | 0.6333 | 3174.717955 | 348017.7648 | 3765206.059 | 0.092801678 | -1.032444169 | TRUE | FALSE | FALSE |
| PSG1 | 293.97178 | 0.01023468 | 0.11465 | 31.08584989 | 2848.040914 | 28251.28214 | 0.55758655 | 0.729809165 | 0.9676 | 47.18856179 | 450.6700338 | 4409.318207 | 527.2217944 | 2.721993355 | TRUE | TRUE | TRUE |
| ANO9 | 291.53657 | 0.01030624 | 0.61725 | 2852.466475 | 1169367.287 | 12242734.16 | 37999.2 | 4.88513E-05 | 0.2054 | 2375.361835 | 953472.301 | 9883448.871 | 0.007672177 | -2.115081411 | TRUE | FALSE | FALSE |
| LSM12 | 283.46781 | 0.01037989 | 0.77145 | 2882.774152 | 320314.0667 | 320314.0667 | 3134.79969 | 0.000697688 | 0.10905 | 307.6937415 | 32716.60157 | 349581.2445 | 0.090426133 | -1.043706042 | TRUE | FALSE | FALSE |
| KLHL35 | 283.18419 | 0.01045341 | 0.49945 | 148.5185815 | 27262.42815 | 276983.5622 | 0.63461863 | 0.697565058 | 0.9646 | 128.5951958 | 12623.41686 | 50470.31487 | 446.2273532 | 2.649556189 | TRUE | TRUE | TRUE |
| IDI2 | 274.82145 | 0.01052923 | 0.64205 | 274.8214518 | 26163.12892 | 264334.4671 | 5.27131559 | 0.295425641 | 0.69285 | 5.271315591 | 467.5056175 | 467.5056175 | 52.13526814 | 1.717131612 | TRUE | FALSE | FALSE |
| DOK7 | 271.69161 | 0.01060582 | 0.235 | 27.91602937 | 2707.376471 | 10820.20393 | 0.28224156 | 0.830547609 | 0.92825 | 6.03549742 | 925.6525623 | 7850.329405 | 962.6208625 | 2.98345527 | TRUE | TRUE | TRUE |
| SMYD4 | 271.44286 | 0.01068226 | 0.1151 | 3.157876458 | 271.442859 | 2608.173181 | 55.7015178 | 0.050352637 | 0.3495 | 6.006076758 | 529.2283087 | 5148.404643 | 4.87316809 | 0.687811392 | TRUE | FALSE | FALSE |
| TM2D2 | 267.82714 | 0.01075964 | 0.0027 | 27.52185905 | 267.8271398 | 267.8271398 | 3.54980122 | 0.380859927 | 0.50115 | 3.549801225 | 3.549801225 | 3.549801225 | 75.44848933 | 1.877650549 | TRUE | FALSE | FALSE |
| RPF1 | 267.66077 | 0.01083686 | 0.34995 | 28.20061313 | 2602.594884 | 25910.68241 | 0.06452606 | 0.883688233 | 1 | 0.574906224 | 0.574906224 | 0.574906224 | 4148.103342 | 3.617849567 | TRUE | TRUE | TRUE |
| ZNF429 | 267.42586 | 0.01091394 | 0.03135 | 3.226405938 | 29.02044823 | 267.4258588 | 0.0095459 | 0.896861685 | 1 | 0.701491063 | 57.06621093 | 57.06621093 | 28014.72506 | 4.447386365 | TRUE | TRUE | TRUE |
| NFX1 | 266.52126 | 0.0109911 | 0.07 | 2.893073835 | 266.5212593 | 266.5212593 | 0.05799717 | 0.885064219 | 1 | 0.057997172 | 0.057997172 | 1.138102957 | 4595.4182 | 3.66232504 | TRUE | TRUE | TRUE |
| DNAJC1 | 263.27841 | 0.0110691 | 0.19435 | 27.625076 | 263.2784088 | 2570.525651 | 371.541472 | 0.006329422 | 0.23455 | 371.5414721 | 36731.1793 | 123581.6174 | 0.708611093 | -0.149592053 | TRUE | FALSE | FALSE |
| ETV2 | 260.55994 | 0.0111478 | 0.06775 | 26.75862848 | 2600.679392 | 2600.679392 | 0.38413599 | 0.797867298 | 1 | 3.573757659 | 3.573757659 | 3.573757659 | 678.3012848 | 2.83142264 | TRUE | TRUE | TRUE |

| | | | | | | | | | | | | | | | | | |
|---|---|---|---|---|---|---|---|---|---|---|---|---|---|---|---|---|---|
| GRHPR | 257.0218 | 0.01122747 | 0.73755 | 1809.695612 | 264378.3346 | 4760497.92 | 1.46356786 | 0.539106322 | 0.7033 | 7.508174321 | 259.8423041 | 1018.794792 | 175.6131779 | 2.244557102 | TRUE | FALSE | FALSE |
| ANKRD55 | 253.64036 | 0.01130811 | 0.06625 | 2.784494917 | 253.6403575 | 253.6403575 | 0.05483245 | 0.885678591 | 1 | 0.495580795 | 0.495580795 | 1.084497199 | 4625.734472 | 3.6651807 | TRUE | TRUE | TRUE |
| SETD6 | 253.06157 | 0.01138873 | 0.4989 | 253.0615698 | 252402.8783 | 2639965.94 | 43.9387767 | 0.06281759 | 0.6568 | 43.93877667 | 4174.606603 | 42168.2124 | 5.759413188 | 0.760378237 | TRUE | FALSE | FALSE |
| LAMB1 | 252.12046 | 0.01146946 | 0.7768 | 9873.43772 | 16184975.84 | 452628947.6 | 12.1743149 | 0.177241533 | 0.79215 | 282.9735615 | 299429.2262 | 3184999.718 | 20.70921126 | 1.316163558 | TRUE | FALSE | FALSE |
| ZC3H12D | 247.14103 | 0.01155177 | 0.50575 | 247.141029 | 2487.260651 | 25643.14006 | 32.4048402 | 0.085304618 | 0.25715 | 3.368791623 | 32.40484018 | 32.40484018 | 7.626670203 | 0.882334967 | TRUE | FALSE | FALSE |
| SHANK3 | 245.42768 | 0.01163449 | 0.2123 | 183.9139987 | 1849.130804 | 19045.58865 | 9.36390088 | 0.207414284 | 0.2665 | 2.601802439 | 25.00570664 | 87.68694808 | 26.20998247 | 1.418466731 | TRUE | FALSE | FALSE |
| AAK1 | 245.27099 | 0.01171704 | 0.50855 | 2474.717599 | 270907.1449 | 2933445.369 | 0.38996198 | 0.795719017 | 1 | 3.673170743 | 3.673170743 | 72.44073227 | 628.9612868 | 2.798623915 | TRUE | TRUE | TRUE |
| NOC2L | 243.27829 | 0.01180011 | 0.2262 | 26.30049358 | 2304.143533 | 4770.367158 | 0.08492335 | 0.879143453 | 0.51245 | 0.08492335 | 0.08492335 | 1.669158885 | 2864.680789 | 3.457076236 | TRUE | TRUE | TRUE |
| OR5F1 | 234.94998 | 0.0118862 | 0.3146 | 24.23253422 | 2335.012226 | 2335.012226 | 0.35427267 | 0.807654658 | 1 | 3.283325481 | 3.283325481 | 3.283325481 | 663.189688 | 2.821637765 | TRUE | TRUE | TRUE |
| HYKK | 234.38081 | 0.01197228 | 0.7488 | 1607.219318 | 144219.1682 | 577577.9251 | 8.05422445 | 0.226235862 | 0.911 | 303.0970714 | 19440.46494 | 77856.99335 | 29.10035727 | 1.463898321 | TRUE | FALSE | FALSE |
| EMP1 | 234.34023 | 0.01205815 | 0.80625 | 938.2074807 | 117960.5641 | 472741.6008 | 1.11820646 | 0.576490116 | 0.9849 | 42.05168019 | 1623.447798 | 6496.868393 | 209.5679452 | 2.321324855 | TRUE | FALSE | FALSE |
| RBKS | 233.68451 | 0.01214405 | 0.19135 | 34.74032436 | 2222.352084 | 11045.13862 | 2040.29086 | 0.00116747 | 0.03255 | 21.75969646 | 510.2508072 | 2040.290863 | 0.1145349 | -0.94106216 | TRUE | FALSE | FALSE |
| MAGEF1 | 227.07688 | 0.01223245 | 0.00515 | 23.42648565 | 227.0768767 | 227.0768767 | 258.570611 | 0.009715622 | 0.31885 | 26.60482091 | 2575.922145 | 2575.922145 | 0.878200641 | -0.05640625 | TRUE | FALSE | FALSE |
| SFTPA2 | 223.97465 | 0.01232196 | 0.31215 | 22.78157994 | 2257.07104 | 2257.07104 | 2.79783426 | 0.427438817 | 0.9379 | 26.87082625 | 2672.583954 | 2672.583954 | 80.0528654 | 1.903376882 | TRUE | FALSE | FALSE |
| CNTN3 | 223.57149 | 0.0124114 | 0.3836 | 55.84827632 | 895.2235763 | 4355.384671 | 4.77693686 | 0.315902416 | 0.84465 | 105.0417521 | 6729.996752 | 26943.21568 | 46.80227115 | 1.670266928 | TRUE | FALSE | FALSE |
| OR2B11 | 223.33204 | 0.0125007 | 0.0049 | 22.82923111 | 223.3320431 | 223.3320431 | 0.32138176 | 0.81833142 | 1 | 2.999599753 | 2.999599753 | 2.999599753 | 694.9120096 | 2.841929817 | TRUE | TRUE | TRUE |
| BRD2 | 222.45484 | 0.01259014 | 0.7809 | 2239.859476 | 244999.2953 | 780541.4166 | 30.5192317 | 0.091070236 | 0.51335 | 30.51923168 | 301.4963353 | 601.6351055 | 7.289005316 | 0.862668267 | TRUE | FALSE | FALSE |
| TMPRSS12 | 221.85613 | 0.01267961 | 0.1286 | 23.37752892 | 887.4562509 | 8633.074373 | 586402.58 | 3.05465E-06 | 0.02945 | 4596.081652 | 423926.423 | 2349251.507 | 0.000378334 | -3.422124446 | TRUE | FALSE | FALSE |
| OR52B2 | 218.89484 | 0.01277016 | 0.18945 | 218.8948395 | 2194.916776 | 22546.31621 | 3.04349349 | 0.409843016 | 0.7475 | 3.043493487 | 29.18009912 | 29.18009912 | 71.9222303 | 1.856863146 | TRUE | FALSE | FALSE |
| CD109 | 217.31052 | 0.01286118 | 0.3474 | 54.25317674 | 10120.0883 | 95157.91041 | 50.183984 | 0.054545614 | 0.87635 | 6385.914345 | 1749249.517 | 26602702.94 | 4.330276368 | 0.636515615 | TRUE | FALSE | FALSE |
| CHEK2 | 216.70044 | 0.01295225 | 0.25745 | 90.99336131 | 8443.670551 | 84225.44114 | 341395.094 | 5.17212E-06 | 0.6018 | 1364799.192 | 1937626330 | 25038326263 | 0.00063475 | -3.197397475 | TRUE | FALSE | FALSE |
| SLC44A3 | 214.63467 | 0.01304402 | 0.1778 | 19.41439945 | 858.8155211 | 3437.228458 | 4.40084336 | 0.335613909 | 0.4757 | 2.479642165 | 39.67235851 | 158.7451029 | 48.7712589 | 1.688163966 | TRUE | FALSE | FALSE |
| CRHR2 | 213.41164 | 0.01313612 | 0.6248 | 2084.450446 | 2232919.83 | 23893065.71 | 17.2761283 | 0.14729996 | 0.41665 | 17.27612831 | 1573.290699 | 6293.352075 | 12.35297818 | 1.091771674 | TRUE | FALSE | FALSE |
| ANGEL2 | 212.32296 | 0.01322849 | 0.03605 | 21.69268664 | 212.322961 | 2128.949819 | 0.04638302 | 0.88745442 | 1 | 0.424426669 | 0.424426669 | 0.919768397 | 4577.600828 | 3.660637919 | TRUE | TRUE | TRUE |
| TNK1 | 210.10962 | 0.01332167 | 0.23465 | 210.1096189 | 20720.30859 | 69800.88965 | 0.45522169 | 0.772741734 | 0.26005 | 0.455221691 | 4.197622776 | 9.002840198 | 461.554498 | 2.664222988 | TRUE | TRUE | TRUE |
| BBS7 | 208.0812 | 0.0134156 | 0.41155 | 208.0812 | 13346.23602 | 119751.0121 | 1026.15681 | 0.002389327 | 0.3441 | 256.5046747 | 28707.17034 | 263494.4749 | 0.202777195 | -0.692980888 | TRUE | FALSE | FALSE |
| TTC33 | 207.01404 | 0.0135098 | 0.8554 | 2056.533909 | 218071.2492 | 2325110.592 | 24218.969 | 7.1894E-05 | 0.50525 | 24218.96902 | 2704891.906 | 31395567.88 | 0.008547599 | -2.068155845 | TRUE | FALSE | FALSE |
| WDR37 | 205.43496 | 0.01360453 | 0.19485 | 205.4349623 | 2072.999149 | 21428.71914 | 0.3231314 | 0.817698252 | 1 | 0.323131403 | 0.323131403 | 6.386941218 | 635.7629142 | 2.803295191 | TRUE | TRUE | TRUE |
| NCOR2 | 205.39466 | 0.01369904 | 0.6526 | 205.3946622 | 38902.06222 | 402034.2003 | 4.08458361 | 0.351533108 | 0.2876 | 0.431568115 | 8.158580726 | 77.2170231 | 50.28533671 | 1.701441362 | TRUE | FALSE | FALSE |
| PAXBP1 | 203.51699 | 0.01379424 | 0.91045 | 20899.44547 | 2305223.613 | 25334939.43 | 29528638.1 | 6.87296E-08 | 0.0544 | 24222.46382 | 29528638.08 | 332261333.7 | 6.89219E-06 | -5.161642756 | TRUE | FALSE | FALSE |
| AOX1 | 203.44865 | 0.01388923 | 0.44885 | 87.03386636 | 19065.9155 | 190975.3448 | 1.5970379 | 0.526525791 | 0.20825 | 1.597037904 | 14.80651036 | 140.6339605 | 127.3912494 | 2.105139597 | TRUE | FALSE | FALSE |
| DNAJC5B | 203.10908 | 0.01398413 | 0.63635 | 2030.221249 | 217948.6438 | 2292651.043 | 12.8755793 | 0.170868733 | 0.0033 | 3.219318852 | 12.87557928 | 12.87557928 | 15.77475243 | 1.197962552 | TRUE | FALSE | FALSE |
| MUSK | 202.5006 | 0.0140791 | 0.94515 | 847.17734 | 52212.9839 | 209356.8378 | 6.17119223 | 0.264103013 | 0.4749 | 6.171192233 | 98.81780933 | 411.0126498 | 32.81385437 | 1.516057246 | TRUE | FALSE | FALSE |
| C20orf141 | 201.20411 | 0.01417448 | 0.85795 | 1985.539956 | 207759.4004 | 2200491.387 | 242149.866 | 7.91796E-06 | 0.25815 | 23315.64076 | 2573465.545 | 22226044.23 | 0.000830907 | -3.080447391 | TRUE | FALSE | FALSE |
| PITX3 | 198.36811 | 0.0142711 | 0.25595 | 49.59913578 | 3929.70629 | 19705.84736 | 0.7407942 | 0.646211698 | 0.9688 | 16.75811923 | 268.0088879 | 1072.195477 | 267.7776265 | 2.427774288 | TRUE | TRUE | TRUE |
| CD302 | 198.03817 | 0.01436763 | 0.1257 | 2.045775583 | 198.0381728 | 198.0381728 | 2.17668193 | 0.47326206 | 0.75275 | 2.176681931 | 21.17479966 | 21.17479966 | 90.98167722 | 1.958953939 | TRUE | FALSE | FALSE |
| CPNE1 | 196.29881 | 0.01446485 | 0.24135 | 196.2988148 | 19279.19477 | 37981.01362 | 317.06685 | 0.007659607 | 0.346 | 32.8786416 | 3132.387482 | 31684.10805 | 0.619108603 | -0.208233161 | TRUE | FALSE | FALSE |
| SLC26A5 | 195.17007 | 0.01456242 | 0.31295 | 48.77736977 | 3127.032086 | 12521.37793 | 967001.47 | 1.65939E-06 | 0.11865 | 15053.25127 | 3874718.432 | 15529639.49 | 0.00020183 | -3.695013927 | TRUE | FALSE | FALSE |
| PMM2 | 194.97166 | 0.01465984 | 0.35225 | 112.972515 | 7233.189609 | 50064.69621 | 41.1652109 | 0.066837528 | 0.6101 | 41.16521092 | 5996.505636 | 24010.4031 | 4.736321113 | 0.675441139 | TRUE | FALSE | FALSE |
| ADAM21 | 194.96216 | 0.01475701 | 0.0682 | 2.231457302 | 194.9621553 | 194.9621553 | 119.240902 | 0.024562068 | 0.00215 | 3.523417106 | 32.84875519 | 32.84875519 | 1.635027511 | 0.213525064 | TRUE | FALSE | FALSE |
| SMPD4 | 194.85102 | 0.01485398 | 0.0695 | 2.235031552 | 194.8510248 | 194.8510248 | 0.04655944 | 0.887429983 | 0.5083 | 0.41263108 | 0.41263108 | 8.106730185 | 4184.995431 | 3.621694988 | TRUE | TRUE | TRUE |
| C3orf52 | 193.11692 | 0.01495165 | 0.18995 | 193.1169225 | 1903.169703 | 19213.95857 | 208916.794 | 9.39998E-06 | 0.25585 | 6881.138056 | 2211752.516 | 23948258.71 | 0.000924372 | -3.034153021 | TRUE | FALSE | FALSE |
| PNPLA3 | 191.67199 | 0.01504987 | 0.25205 | 47.88899919 | 3072.768128 | 28004.87422 | 2.74149187 | 0.431680994 | 0.77225 | 19.11785643 | 701.9399568 | 3472.328314 | 69.91521512 | 1.844571698 | TRUE | FALSE | FALSE |
| ACTRT3 | 190.34299 | 0.01514858 | 0.75085 | 3057.753621 | 791466.6783 | 12752541.83 | 15443.6023 | 0.000129353 | 0.1848 | 961.194888 | 61926.93615 | 996474.6247 | 0.012325039 | -1.909211713 | TRUE | FALSE | FALSE |
| HINT2 | 188.93614 | 0.01524784 | 0.73065 | 1760.644457 | 269708.3789 | 2212393.024 | 663.975292 | 0.003206241 | 0.0782 | 17.68573455 | 663.9752923 | 2657.27155 | 0.284552966 | -0.545836883 | TRUE | FALSE | FALSE |
| SPDYE1 | 186.8947 | 0.01534801 | 0.32245 | 19.35589598 | 1849.777756 | 1849.777756 | 0.28359617 | 0.830104245 | 0.75415 | 2.616313149 | 24.77258273 | 24.77258273 | 659.0170171 | 2.818896629 | TRUE | TRUE | TRUE |
| SLC11A1 | 184.74446 | 0.01544918 | 0.5079 | 739.7154574 | 47554.81387 | 190599.1596 | 0.09884157 | 0.876237043 | 0.96595 | 1.580756536 | 25.306043 | 101.2899885 | 1869.096683 | 3.271631767 | TRUE | TRUE | TRUE |
| NPVF | 183.86449 | 0.01555062 | 0.22955 | 183.8644851 | 19159.73156 | 19159.73156 | 1960.27747 | 0.001241237 | 0.5022 | 20267.81396 | 2320373.029 | 26595746.77 | 0.093795132 | -1.027819699 | TRUE | FALSE | FALSE |
| FMNL2 | 182.27114 | 0.01565276 | 0.99525 | 3634749.077 | 1574633585 | 18399499139 | 81.0540936 | 0.036535516 | 0.98185 | 412473.2689 | 486446948 | 7790482871 | 2.248759273 | 0.351942967 | TRUE | FALSE | FALSE |
| RCAN1 | 182.09427 | 0.01575474 | 0.65585 | 182.0942675 | 18262.58835 | 189526.4445 | 2260.56628 | 0.001046158 | 0.03335 | 23.4374829 | 2260.566283 | 23025.81994 | 0.080552501 | -1.093920972 | TRUE | FALSE | FALSE |
| EPSTI1 | 181.03167 | 0.0158571 | 0.1922 | 19.44561564 | 181.0316663 | 1726.595462 | 0.39982612 | 0.792165553 | 0.26185 | 0.399826118 | 3.58732809 | 3.58732809 | 452.7759892 | 2.655883388 | TRUE | TRUE | TRUE |
| ILK | 181.00965 | 0.01595922 | 0.4007 | 45.22767274 | 2901.495556 | 11621.02741 | 19109.7308 | 0.000102391 | 0.06785 | 1191.234055 | 76564.84181 | 306840.3715 | 0.00947212 | -2.02355283 | TRUE | FALSE | FALSE |
| C22orf39 | 176.9425 | 0.01606361 | 0.1905 | 176.9424974 | 1740.066495 | 17530.04593 | 0.28369424 | 0.829979682 | 0.75315 | 2.604785166 | 24.54639164 | 24.54639164 | 623.7084701 | 2.794981642 | TRUE | TRUE | TRUE |
| FAM200A | 176.68734 | 0.0161679 | 0.11115 | 19.04437799 | 1679.377441 | 16343.65303 | 0.39409261 | 0.794102748 | 0.87515 | 0.394092613 | 32.34001828 | 32.34001828 | 448.3396302 | 2.651607129 | TRUE | TRUE | TRUE |
| GLYATL1 | 176.08373 | 0.01627 | 0.0678 | 18.30625814 | 1736.118142 | 1736.118142 | 2.20159968 | 0.471861829 | 0.9848 | 199.6457118 | 19986.87344 | 207233.3644 | 79.97990433 | 1.90298088 | TRUE | FALSE | FALSE |
| CAPN9 | 175.02921 | 0.01637711 | 0.5962 | 621.9120731 | 196661.107 | 3157868.764 | 35.7414859 | 0.077932744 | 0.8702 | 5189.260114 | 7348445.281 | 177646841.9 | 4.897088222 | 0.689937928 | TRUE | FALSE | FALSE |
| LMOD2 | 174.94421 | 0.01648172 | 0.9293 | 796.6563041 | 12798.09928 | 63192.6716 | 278.084952 | 0.00905521 | 0.2635 | 69.47954499 | 1113.285205 | 4458.038193 | 0.629103485 | -0.201277909 | TRUE | FALSE | FALSE |
| IRAK1BP1 | 174.23615 | 0.01658651 | 0.19165 | 18.12350535 | 1717.018426 | 17333.80018 | 0.2953514 | 0.82663839 | 1 | 0.295351396 | 0.295351396 | 0.295351396 | 589.9283189 | 2.770799245 | TRUE | TRUE | TRUE |
| OR10A3 | 173.85331 | 0.01669128 | 0.66075 | 173.8533133 | 17366.39826 | 179865.898 | 24.8377621 | 0.1109489 | 0.0056 | 2.623892443 | 24.83776213 | 24.83776213 | 6.999556259 | 0.845070509 | TRUE | FALSE | FALSE |
| CYP1B1 | 173.44972 | 0.01679606 | 0.50045 | 173.4497244 | 17778.68205 | 34191.19156 | 28.1837675 | 0.098424135 | 0.772 | 276.7749212 | 28680.06366 | 54955.7162 | 6.154241949 | 0.789174566 | TRUE | FALSE | FALSE |
| PPP3R2 | 172.87956 | 0.01690093 | 0.06625 | 17.65162315 | 1735.543937 | 1735.543937 | 2.46738853 | 0.449440157 | 0.00495 | 0.263849253 | 2.46738853 | 2.46738853 | 70.06580367 | 1.845506108 | TRUE | FALSE | FALSE |
| MYF6 | 170.75236 | 0.01700696 | 0.1736 | 42.67472352 | 1212.629839 | 10954.82177 | 30.0458803 | 0.09223812 | 0.8113 | 120.2595824 | 7722.793324 | 30941.59597 | 5.683053978 | 0.754581781 | TRUE | FALSE | FALSE |
| TNN | 168.76948 | 0.01711406 | 0.42765 | 92.47346925 | 24539.16411 | 228323.7755 | 0.09999641 | 0.875907239 | 0.80045 | 1.972544637 | 171.7468507 | 686.5221516 | 1687.755371 | 3.227309499 | TRUE | TRUE | TRUE |
| ACADL | 167.55085 | 0.01722173 | 0.50175 | 167.5508461 | 10747.04352 | 172421.8236 | 17138166.6 | 1.42801E-07 | 0.02405 | 12067.39973 | 4279658.836 | 41778229.43 | 9.77647E-06 | -5.009817736 | TRUE | FALSE | FALSE |
| OARD1 | 165.65777 | 0.01733045 | 0.9451 | 65657.43938 | 19296595.01 | 177167795.5 | 225.731109 | 0.011354919 | 0.77365 | 2236.412705 | 235864.0042 | 235864.0042 | 0.733872123 | -0.134379609 | TRUE | FALSE | FALSE |
| MS4A2 | 163.30342 | 0.01744058 | 0.7712 | 1604.766396 | 166513.0854 | 321844.6947 | 22.0886703 | 0.123103236 | 0.49725 | 22.08867032 | 2108.397089 | 21364.43291 | 7.393085182 | 0.86882571 | TRUE | FALSE | FALSE |
| EXTL1 | 162.27859 | 0.01755117 | 0.1025 | 0.343059023 | 648.5661094 | 3198.37801 | 0.0272772 | 0.891891447 | 0.51065 | 0.0272772 | 12.05751931 | 142.2339185 | 5949.239233 | 3.774461433 | TRUE | TRUE | TRUE |
| ADHFE1 | 161.53558 | 0.01766203 | 0.6412 | 395.5680314 | 40749.80624 | 162937.4072 | 0.085238 | 0.879087836 | 1 | 0.781627119 | 3.125462507 | 3.125462507 | 1895.11223 | 3.277634934 | TRUE | TRUE | TRUE |
| SRPRB | 159.54042 | 0.0177741 | 0.3107 | 16.57860315 | 1573.74276 | 1573.74276 | 0.2564443 | 0.837618829 | 1 | 2.360861817 | 2.360861817 | 2.360861817 | 622.125033 | 2.793877677 | TRUE | TRUE | TRUE |
| ANGPTL6 | 159.15502 | 0.01788617 | 0.7511 | 1566.333133 | 163009.6982 | 1721859.371 | 0.69676269 | 0.667690906 | 0.9683 | 55.23889078 | 2518.62761 | 25577.86486 | 228.420696 | 2.358735451 | TRUE | TRUE | TRUE |
| DOK2 | 159.07419 | 0.01799803 | 0.81495 | 1490.816172 | 95671.50424 | 1283396.738 | 6.40977968 | 0.257394259 | 0.94755 | 102.6743422 | 12802.7443 | 51297.90391 | 24.81741892 | 1.394756612 | TRUE | FALSE | FALSE |
| FMO3 | 157.41704 | 0.01811086 | 0.3522 | 16.49143954 | 1539.35672 | 15412.5635 | 0.61438611 | 0.705674026 | 0.6957 | 1.37558471 | 22.0119475 | 22.0119475 | 256.2184137 | 2.408610338 | TRUE | TRUE | TRUE |
| SLC35F1 | 154.1699 | 0.01822594 | 0.12825 | 15.9797348 | 154.1699002 | 154.1699002 | 0.50641315 | 0.753217981 | 1 | 0.506413146 | 0.506413146 | 10.05228213 | 304.4350278 | 2.48349462 | TRUE | TRUE | TRUE |

| | | | | | | | | | | | | | | | | | |
|---|---|---|---|---|---|---|---|---|---|---|---|---|---|---|---|---|---|
| OR11H6 | 154.12058 | 0.01834078 | 0.49665 | 154.1205761 | 158132.6269 | 158132.6269 | 2.52334609 | 0.445619419 | 0.50085 | 2.523346086 | 2.523346086 | 2.523346086 | 61.07785887 | 1.785883804 | TRUE | FALSE | FALSE |
| STK10 | 150.62129 | 0.01845816 | 0.07285 | 1.708961855 | 150.6212915 | 309.3326352 | 0.03648652 | 0.889771975 | 1 | 0.32494342 | 0.32494342 | 6.361415945 | 4128.135497 | 3.615753944 | TRUE | TRUE | TRUE |
| MRPS9 | 147.60431 | 0.01857779 | 0.0705 | 9.214876259 | 147.6043085 | 590.9713811 | 3.40182125 | 0.389196874 | 0.50925 | 13.61075627 | 218.0465421 | 461.3105602 | 43.38978972 | 1.637387545 | TRUE | FALSE | FALSE |
| EPS8L2 | 144.57151 | 0.01869977 | 0.67245 | 439.0500077 | 37116.15543 | 182333.7273 | 1284.92953 | 0.00190393 | 0.66125 | 2995.305884 | 230304.7811 | 2866943.583 | 0.112513181 | -0.948796596 | TRUE | FALSE | FALSE |
| OR1B1 | 144.33106 | 0.01882165 | 0.06595 | 15.07073948 | 1416.860785 | 1416.860785 | 2.17658244 | 0.473436671 | 0.50045 | 2.176582441 | 2.176582441 | 2.176582441 | 66.31086172 | 1.821584672 | TRUE | FALSE | FALSE |
| TMPRSS11D | 141.56964 | 0.01894572 | 0.5086 | 141.5696404 | 1410.069466 | 14387.61202 | 2.09463713 | 0.480329473 | 0.75455 | 2.094637133 | 19.96325421 | 19.96325421 | 67.58671378 | 1.829861331 | TRUE | FALSE | FALSE |
| ADRA1A | 141.4614 | 0.01906959 | 0.63665 | 141.4614022 | 13751.53429 | 140393.4021 | 26.3335233 | 0.104356081 | 0.50615 | 26.33352331 | 253.2236387 | 2494.526346 | 5.371913227 | 0.730128989 | TRUE | FALSE | FALSE |
| CELA2B | 141.13265 | 0.01919345 | 0.3255 | 14.78426775 | 1381.015714 | 13843.78099 | 0.02108682 | 0.893534701 | 0.75265 | 0.192739464 | 1.808113678 | 1.808113678 | 6692.930914 | 3.825616342 | TRUE | TRUE | TRUE |
| CCDC74A | 140.16483 | 0.01931789 | 0.0703 | 14.59608099 | 1379.69464 | 1379.69464 | 1.86275893 | 0.502079007 | 0.96895 | 168.7225098 | 1666.954262 | 16871.49957 | 75.24582531 | 1.87648241 | TRUE | FALSE | FALSE |
| CBX2 | 138.84563 | 0.01944327 | 0.0726 | 14.41672479 | 1370.685278 | 1370.685278 | 599.188313 | 0.003654829 | 0.75055 | 5919.374274 | 620715.434 | 6581358.67 | 0.23172286 | -0.63503112 | TRUE | FALSE | FALSE |
| OR10G9 | 137.24594 | 0.01956986 | 0.127 | 14.34153475 | 137.245944 | 137.245944 | 2.01983624 | 0.488544504 | 0.4994 | 0.220690296 | 2.019836242 | 2.019836242 | 67.94904516 | 1.832183358 | TRUE | FALSE | FALSE |
| PDZD9 | 135.94864 | 0.0196974 | 0.98855 | 136954.3339 | 172827579.9 | 26179765379 | 1900354.75 | 8.74016E-07 | 0.3849 | 179805.7997 | 259103940.6 | 2916940074 | 7.15386E-05 | -4.145459819 | TRUE | FALSE | FALSE |
| C4orf17 | 135.09642 | 0.01982546 | 0.00345 | 1.520253769 | 135.0964174 | 135.0964174 | 0.2256408 | 0.845938814 | 1 | 0.225640801 | 0.225640801 | 0.225640801 | 598.7233561 | 2.7772262 | TRUE | TRUE | TRUE |
| AK3 | 134.61047 | 0.01995369 | 0.1876 | 14.07254222 | 330.2506758 | 1320.653422 | 0.5478915 | 0.73418197 | 0.7514 | 0.547891499 | 2.1911194 | 2.1911194 | 245.6881812 | 2.390384265 | TRUE | TRUE | TRUE |
| ZNF763 | 134.00812 | 0.0200822 | 0.3514 | 134.0081181 | 13461.94905 | 139820.3232 | 2.01239954 | 0.489039334 | 0.8771 | 19.08026424 | 185.5498552 | 185.5498552 | 66.59120893 | 1.823416899 | TRUE | FALSE | FALSE |
| ZNF799 | 133.85547 | 0.02021055 | 0.1895 | 1.556221347 | 133.8554654 | 1287.650349 | 0.03619232 | 0.889850352 | 0.01415 | 0.036192325 | 0.036192325 | 0.716619472 | 3698.44893 | 3.568019626 | TRUE | TRUE | TRUE |
| RASA1 | 132.75056 | 0.02033969 | 0.62325 | 531.5091758 | 27332.92041 | 109540.5202 | 2.82181274 | 0.425397379 | 0.66585 | 11.29020742 | 180.8734559 | 724.2258207 | 47.04442681 | 1.672508181 | TRUE | FALSE | FALSE |
| AUTS2 | 131.28932 | 0.02047002 | 0.000025 | 0.11236869 | 0.11236869 | 2.177905732 | 0.13018902 | 0.868916822 | 1 | 0.13018902 | 0.13018902 | 2.532701359 | 1008.451539 | 3.003655033 | TRUE | TRUE | TRUE |
| OR52R1 | 131.213 | 0.0206001 | 0.06635 | 13.57811471 | 1299.728894 | 1299.728894 | 1.91304482 | 0.497963445 | 0.7469 | 1.913044822 | 18.12683307 | 18.12683307 | 68.58856537 | 1.836251719 | TRUE | FALSE | FALSE |
| CENPQ | 130.53412 | 0.02073057 | 0.06595 | 1.383506752 | 130.5341203 | 130.5341203 | 0.02007183 | 0.893821226 | 0.8725 | 0.40009691 | 16.69389042 | 16.69389042 | 6503.350789 | 3.81313718 | TRUE | TRUE | TRUE |
| CLSTN3 | 130.49576 | 0.02086075 | 0.3206 | 13.41151936 | 1301.519066 | 1301.519066 | 15.7583749 | 0.15563537 | 0.66205 | 153.5343665 | 15686.89698 | 164312.113 | 8.28104183 | 0.918084978 | TRUE | FALSE | FALSE |
| OR7E24 | 129.93725 | 0.02099121 | 0.96805 | 25340.80669 | 14940389.17 | 165785248.5 | 22.6677683 | 0.120052307 | 0.25155 | 2.398213117 | 22.66776831 | 22.66776831 | 5.73224711 | 0.758324904 | TRUE | FALSE | FALSE |
| UEVLD | 128.49352 | 0.02112286 | 0.35255 | 13.29128723 | 1272.579878 | 12904.23198 | 2.46682542 | 0.449629816 | 0.59265 | 2.466825419 | 93.87308265 | 375.3874583 | 52.08861703 | 1.716742827 | TRUE | FALSE | FALSE |
| CCDC74B | 127.28659 | 0.0212555 | 0.07245 | 13.30905506 | 1247.845405 | 1247.845405 | 1.95265292 | 0.493133725 | 0.51395 | 1.952652916 | 1.952652916 | 4.22716991 | 65.18648798 | 1.814157583 | TRUE | FALSE | FALSE |
| ZBBX | 127.03557 | 0.02138808 | 0.19605 | 13.68184201 | 127.0355733 | 1208.398403 | 200.079477 | 0.013441529 | 0.11525 | 21.33169599 | 1922.543617 | 18914.72549 | 0.634925555 | -0.197277192 | TRUE | FALSE | FALSE |
| OR6C68 | 126.72263 | 0.02152068 | 0.6384 | 1239.367386 | 127379.9182 | 1337133.295 | 18.2946263 | 0.14226538 | 0.50685 | 175.9486952 | 1734.551097 | 17517.34282 | 6.926768006 | 0.840530642 | TRUE | FALSE | FALSE |
| AGR3 | 125.12023 | 0.02165472 | 0.662 | 125.1202312 | 12229.78275 | 125298.6783 | 0.23200577 | 0.844429385 | 1 | 0.232005772 | 0.232005772 | 0.232005772 | 539.2979232 | 2.731828748 | TRUE | TRUE | TRUE |
| GNA14 | 125.10809 | 0.02178844 | 0.38735 | 31.28046741 | 2002.787735 | 8016.268736 | 15.1051485 | 0.157575233 | 0.6077 | 60.40942944 | 5480.241327 | 21922.38855 | 8.282479678 | 0.918160379 | TRUE | FALSE | FALSE |
| MEIOB | 125.02353 | 0.02192194 | 0.1295 | 13.12815485 | 500.0919343 | 500.0919343 | 0.03916527 | 0.889149243 | 0.50065 | 0.35690151 | 0.35690151 | 0.35690151 | 3192.204372 | 3.504090688 | TRUE | TRUE | TRUE |
| LAYN | 123.51271 | 0.02205681 | 0.00555 | 12.9269183 | 123.5127102 | 123.5127102 | 0.20188523 | 0.851676905 | 1 | 1.845123178 | 1.845123178 | 1.845123178 | 611.7966638 | 2.786607104 | TRUE | TRUE | TRUE |
| OR51M1 | 122.58429 | 0.0221924 | 0.3217 | 12.75876863 | 1207.262609 | 1207.262609 | 149.939116 | 0.018797088 | 0.4987 | 15.54901584 | 1482.055915 | 15006.98272 | 0.817560456 | -0.087480123 | TRUE | FALSE | FALSE |
| HEXA | 122.33839 | 0.02232795 | 0.20555 | 13.60818666 | 489.4272514 | 489.4272514 | 0.11002833 | 0.873339704 | 0.07885 | 0.006881676 | 0.440121221 | 1.760955799 | 1111.880834 | 3.046058244 | TRUE | TRUE | TRUE |
| SNX18 | 121.66582 | 0.02246395 | 0.31735 | 12.69888846 | 1194.850141 | 1194.850141 | 16.5102412 | 0.15049775 | 0.5015 | 16.51024119 | 159.0924245 | 159.0924245 | 7.369112081 | 0.867415162 | TRUE | FALSE | FALSE |
| KRTAP5-1 | 120.25737 | 0.02260127 | 0.6395 | 120.2573696 | 10907.24389 | 107565.6151 | 23.1157677 | 0.116390131 | 0.1906 | 5.705553085 | 215.4850452 | 2057.900228 | 5.202395659 | 0.716203379 | TRUE | FALSE | FALSE |
| ZBED3 | 119.1389 | 0.02273959 | 0.8576 | 1159.368858 | 117997.4329 | 1232746.341 | 0.35234287 | 0.808292532 | 1 | 0.352342872 | 0.352342872 | 0.352342872 | 338.1334193 | 2.529088096 | TRUE | TRUE | TRUE |
| TRIM63 | 118.1637 | 0.02287875 | 0.75675 | 1127.938444 | 442640.2302 | 4542755.166 | 0.14263997 | 0.866272253 | 1 | 0.570464324 | 5.227838211 | 5.227838211 | 828.4053027 | 2.91824287 | TRUE | TRUE | TRUE |
| LRRC19 | 117.92175 | 0.02301787 | 0.6469 | 1156.353349 | 119478.555 | 1257509.959 | 154.694268 | 0.01771322 | 0.7752 | 154.6942682 | 159529.5436 | 159529.5436 | 0.762289074 | -0.117880305 | TRUE | FALSE | FALSE |
| ZNF696 | 117.58931 | 0.02315706 | 0.5055 | 117.5893086 | 4652.647958 | 47153.36464 | 1.87613028 | 0.50066094 | 0.01335 | 1.876130277 | 1.876130277 | 4.018403486 | 62.67651564 | 1.797104845 | TRUE | FALSE | FALSE |
| ASPN | 117.58711 | 0.02329592 | 0.76875 | 327.9033569 | 121897.2592 | 233708.8134 | 17.590812 | 0.14508876 | 0.4997 | 6.823444847 | 170.8903475 | 170.8903475 | 6.684575412 | 0.825073827 | TRUE | FALSE | FALSE |
| ZNF77 | 116.64609 | 0.0234356 | 1 | 1.62677E+32 | 4.97457E+40 | 4.71154E+44 | 89.8096794 | 0.03291525 | 1 | 1.16339E+28 | 4.42207E+36 | 2.24705E+40 | 1.298814235 | 0.11354704 | TRUE | FALSE | FALSE |
| KRTAP24-1 | 116.06079 | 0.02357568 | 0.9451 | 11276.3463 | 12871736.09 | 140674683.9 | 1303609.63 | 1.26719E-06 | 0.69215 | 1303609.628 | 19366007858 | 2.26984E+11 | 8.90303E-05 | -4.050462047 | TRUE | FALSE | FALSE |
| RHNO1 | 114.82249 | 0.02371697 | 0.50445 | 1118.142673 | 113933.626 | 113933.626 | 16.2805724 | 0.151459205 | 0.8169 | 155.8389635 | 1529.053333 | 15369.1893 | 7.052730394 | 0.848357282 | TRUE | FALSE | FALSE |
| HBA1 | 113.80484 | 0.02385922 | 0.1894 | 28.43572132 | 113.8048444 | 455.5811309 | 8.00302669 | 0.228229194 | 0.4983 | 2.000603701 | 32.02255614 | 32.02255614 | 14.22022552 | 1.152906484 | TRUE | FALSE | FALSE |
| EPHA7 | 112.7519 | 0.0240025 | 0.119 | 7.040636557 | 112.7518994 | 451.380225 | 2.22047427 | 0.470102475 | 0.81825 | 12.60333758 | 201.5577311 | 806.3418867 | 50.77829574 | 1.705678121 | TRUE | FALSE | FALSE |
| BRSK2 | 112.22607 | 0.02414612 | 0.0063 | 0.868387489 | 8.133324898 | 17.04248424 | 152.424012 | 0.018389197 | 0.00015 | 1.184613343 | 1.184613343 | 23.30098405 | 0.736275526 | -0.132959635 | TRUE | FALSE | FALSE |
| GJB6 | 110.26993 | 0.024292 | 0.5085 | 1118.924913 | 123581.932 | 1339538.478 | 21.4081568 | 0.127064194 | 0.1883 | 21.40815681 | 213.6398958 | 2184.042608 | 5.150837189 | 0.711877823 | TRUE | FALSE | FALSE |
| SYNPO2L | 109.88057 | 0.02443807 | 0.6272 | 1046.23072 | 1038394.386 | 10832536.69 | 213.829307 | 0.012328921 | 0.19545 | 22.66233287 | 213.8293074 | 2066.924324 | 0.513870496 | -0.289146317 | TRUE | FALSE | FALSE |
| GAS2L2 | 108.25942 | 0.02458601 | 0.1218 | 11.44445216 | 1049.141232 | 10410.06532 | 19.5618642 | 0.134786147 | 0.5111 | 19.56186416 | 186.8728209 | 1828.82761 | 5.534207777 | 0.74305546 | TRUE | FALSE | FALSE |
| ID1 | 107.98975 | 0.02473397 | 0.03565 | 26.97923018 | 107.9897522 | 433.9224086 | 7.46269475 | 0.235838535 | 0.50485 | 29.86372702 | 119.536584 | 480.2754997 | 14.47061093 | 1.160486867 | TRUE | FALSE | FALSE |
| RRH | 107.47156 | 0.02488232 | 0.5026 | 107.4715585 | 4210.823453 | 42257.27675 | 15.6309105 | 0.156598961 | 0.31275 | 6.657796127 | 250.1181685 | 250.1181685 | 6.875578903 | 0.83730927 | TRUE | FALSE | FALSE |
| KRT36 | 105.22912 | 0.02503351 | 0.46775 | 45.34386515 | 9734.128057 | 96945.90571 | 138958.283 | 1.3968E-05 | 0.1143 | 1344.429978 | 863253.0305 | 8875786.376 | 0.000757271 | -3.120748512 | TRUE | FALSE | FALSE |
| EIF4G3 | 104.95505 | 0.02518475 | 0.99015 | 13003352.15 | 19531254662 | 2.84237E+12 | 0.1665827 | 0.860586825 | 0.99995 | 1662385.837 | 2376291409 | 46329505827 | 630.0477329 | 2.799373453 | TRUE | TRUE | TRUE |
| OR10Z1 | 103.86006 | 0.02533723 | 0.31535 | 10.95139743 | 1009.656514 | 1009.656514 | 0.18738483 | 0.855153379 | 1 | 0.187384826 | 0.187384826 | 0.187384826 | 554.2607955 | 2.743714161 | TRUE | TRUE | TRUE |
| GLG1 | 103.79249 | 0.02548946 | 0.01895 | 10.64533841 | 103.7924934 | 207.3620883 | 56.1065334 | 0.049639108 | 0.01715 | 5.879845679 | 56.10653344 | 198.0501647 | 1.84991813 | 0.267152509 | TRUE | FALSE | FALSE |
| RNH1 | 103.33028 | 0.02564202 | 0.0074 | 10.82781935 | 103.3302843 | 103.3302843 | 0.1783719 | 0.857397671 | 1 | 0.178371904 | 0.178371904 | 3.532371316 | 579.2968604 | 2.762901175 | TRUE | TRUE | TRUE |
| SYTL2 | 102.9869 | 0.02579474 | 0.6349 | 269.9459302 | 267332.8688 | 2786767.89 | 605.343141 | 0.003603444 | 0.11645 | 6.657296324 | 605.3431409 | 5982.048238 | 0.170129785 | -0.769219647 | TRUE | FALSE | FALSE |
| ZNF783 | 102.3757 | 0.02594801 | 0.5149 | 102.3756966 | 1001.415254 | 10034.9823 | 1.66772639 | 0.520583137 | 0.5087 | 1.667726393 | 1.667726393 | 6.17625239 | 61.38638628 | 1.788072068 | TRUE | FALSE | FALSE |
| BIRC7 | 102.17552 | 0.02610123 | 0.2592 | 102.175521 | 1711.790716 | 6857.738594 | 12068.3849 | 0.000175321 | 0.6758 | 48370.30857 | 7120786.399 | 50217098.7 | 0.008466379 | -2.072302291 | TRUE | FALSE | FALSE |
| CPB2 | 101.33356 | 0.02625538 | 0.0713 | 10.61967032 | 991.1495166 | 991.1495166 | 0.18079347 | 0.856916054 | 1 | 0.180793468 | 0.180793468 | 0.180793468 | 560.4934771 | 2.748570563 | TRUE | TRUE | TRUE |
| FCRL3 | 100.13601 | 0.02641101 | 0.83235 | 3800.354173 | 933329.6196 | 9703008.633 | 75.2225957 | 0.038137952 | 0.6934 | 705.7643348 | 267094.0172 | 2689906.511 | 1.331195936 | 0.124241983 | TRUE | FALSE | FALSE |
| SUPT3H | 99.425995 | 0.02656741 | 0.94415 | 9685.765565 | 11100052.59 | 121472181.5 | 1.68439692 | 0.518524512 | 0.81195 | 15.78084432 | 1493.693383 | 15072.20809 | 59.02765182 | 1.771055507 | TRUE | FALSE | FALSE |
| CEP164 | 99.33234 | 0.02672358 | 0.72455 | 928.2339983 | 86830.77734 | 868328.4816 | 26.2198383 | 0.104655715 | 0.5186 | 26.21983834 | 2320.623109 | 22607.18836 | 3.788442104 | 0.578460655 | TRUE | FALSE | FALSE |
| NAPG | 99.225221 | 0.02687956 | 0.3439 | 24.78940548 | 398.3629668 | 1595.33471 | 28.3463297 | 0.097539884 | 0.068 | 7.083414641 | 113.4651473 | 113.4651473 | 3.500460975 | 0.54412524 | TRUE | FALSE | FALSE |
| PSAP | 98.953586 | 0.02703561 | 0.00205 | 0.730282644 | 11.68274633 | 11.68274633 | 0.2690167 | 0.834081755 | 0.7546 | 0.269016701 | 1.075678637 | 2.281043677 | 367.8343581 | 2.565652293 | TRUE | TRUE | TRUE |
| PMPCA | 98.58399 | 0.02719188 | 0.3166 | 10.33400961 | 964.0235094 | 964.0235094 | 127.729696 | 0.022250269 | 0.3149 | 13.31798348 | 1255.696931 | 1255.696931 | 0.771817306 | -0.112485488 | TRUE | FALSE | FALSE |
| SLC2A3 | 98.140664 | 0.0273485 | 0.3842 | 98.14066427 | 6299.632818 | 25236.01419 | 8.27295485 | 0.222569126 | 0.85835 | 33.10194153 | 530.3523585 | 2123.648692 | 11.86283088 | 1.074188339 | TRUE | FALSE | FALSE |
| KRT81 | 95.562156 | 0.02750899 | 0.5686 | 187.1452608 | 23289.51035 | 195959.7987 | 10814482.8 | 1.81919E-07 | 0.33275 | 1485793.495 | 2978172648 | 34438073389 | 8.8365E-06 | -5.053719813 | TRUE | FALSE | FALSE |
| CCDC87 | 95.531455 | 0.02766915 | 0.97065 | 87683.90929 | 98615037.08 | 1070485132 | 1879.75956 | 0.001298622 | 0.3771 | 194.8234021 | 18570.02267 | 187731.2319 | 0.050821103 | -1.293955911 | TRUE | FALSE | FALSE |
| CMYA5 | 95.23833 | 0.02782944 | 0.88515 | 497292.9947 | 3556678924 | 3.36044E+11 | 0.11979194 | 0.87126769 | 1 | 24190854545 | 2.79554E+15 | 3.33652E+17 | 795.0311957 | 2.90038417 | TRUE | TRUE | TRUE |
| ZNF133 | 94.036066 | 0.0279914 | 0.12805 | 9.817729744 | 94.03606562 | 94.03606562 | 0.16274423 | 0.861416658 | 1 | 0.162744225 | 0.162744225 | 3.215923775 | 577.815068 | 2.761788863 | TRUE | TRUE | TRUE |
| ARHGAP22 | 93.495434 | 0.02815392 | 0.74695 | 895.7491828 | 88350.92164 | 908570.0575 | 337.35946 | 0.007255805 | 0.7505 | 3333.432381 | 349122.6637 | 3696632.521 | 0.277138914 | -0.55730249 | TRUE | FALSE | FALSE |
| PCCB | 92.694055 | 0.02831746 | 0.8743 | 13067.34766 | 4913476.653 | 78743297.69 | 513863.791 | 3.47118E-06 | 0.1183 | 32018.59626 | 8255178.766 | 33081968.73 | 0.000180386 | -3.743796133 | TRUE | FALSE | FALSE |
| DCDC1 | 91.740286 | 0.02848231 | 0.5068 | 91.74028558 | 71783.31802 | 690570.5817 | 0.00116568 | 0.899342522 | 0.7539 | 0.00116568 | 0.009393526 | 0.022987694 | 78701.0618 | 4.895980592 | TRUE | TRUE | TRUE |

| | | | | | | | | | | | | | | | | |
|---|---|---|---|---|---|---|---|---|---|---|---|---|---|---|---|---|
| NMI | 90.954182 | 0.0286482 | 0.00385 | 1.044114386 | 90.95418207 | 90.95418207 | 0.15809908 | 0.862532253 | 1 | 1.431146791 | 1.431146791 | 1.431146791 | 575.2986121 | 2.759893326 | TRUE | TRUE | TRUE |
| KRT28 | 90.599859 | 0.02881434 | 0.50395 | 90.59985907 | 8691.86895 | 88225.39189 | 0.06385995 | 0.883826319 | 0.75165 | 0.063859951 | 0.255308122 | 0.541485206 | 1418.727353 | 3.151898942 | TRUE | TRUE | TRUE |
| FILIP1L | 90.097259 | 0.02898101 | 0.12475 | 9.714142422 | 856.09722 | 8328.996103 | 0.02501138 | 0.892449321 | 0.8778 | 1.948534606 | 17.87207263 | 38.30974689 | 3602.250628 | 3.556573926 | TRUE | TRUE | TRUE |
| AMY2A | 89.094109 | 0.02914916 | 0.71155 | 356.5476854 | 8931.536569 | 35778.49894 | 48.0907766 | 0.05708821 | 0.608 | 111.8937923 | 4279.702173 | 17132.04875 | 1.852623633 | 0.2677872 | TRUE | FALSE | FALSE |
| TRIM7 | 88.638845 | 0.02931777 | 0.88675 | 5046.796626 | 821111.8397 | 8545093.43 | 127788.226 | 1.53961E-05 | 0.00855 | 136.5911827 | 12751.63304 | 31943.79497 | 0.000693639 | -3.158866753 | TRUE | FALSE | FALSE |
| MEGF10 | 84.159747 | 0.02949476 | 0.50945 | 84.15974655 | 12050.28226 | 48261.48879 | 11884.3113 | 0.000179268 | 0.11215 | 185.0576227 | 11884.3113 | 47615.27728 | 0.007081584 | -2.1498696 | TRUE | FALSE | FALSE |
| CHRNA6 | 83.845503 | 0.02967198 | 0.0361 | 8.569004848 | 83.84550349 | 840.4543513 | 0.08247366 | 0.879687887 | 0.75345 | 0.082473663 | 0.329718553 | 0.409393304 | 1016.633677 | 3.007164492 | TRUE | TRUE | TRUE |
| ATP11A | 83.790341 | 0.02984892 | 0.06325 | 2.28836805 | 196.7966025 | 786.9559926 | 0.03882356 | 0.889252158 | 0.6355 | 0.087357325 | 3.222288113 | 6.813586864 | 2158.23447 | 3.334098625 | TRUE | TRUE | TRUE |
| CCDC110 | 82.728495 | 0.03002764 | 0.5068 | 82.72849479 | 7503.316006 | 73939.78029 | 2.0043239 | 0.489204164 | 0.6658 | 2.004323901 | 171.1010741 | 1638.345858 | 41.27501286 | 1.615687217 | TRUE | FALSE | FALSE |
| TTC27 | 82.479491 | 0.03020649 | 0.19505 | 8.75201887 | 329.8093244 | 3184.194592 | 0.19109382 | 0.854078065 | 0.875 | 1.731911798 | 16.10002348 | 16.19345135 | 431.6177893 | 2.635099336 | TRUE | TRUE | TRUE |
| HTRA4 | 81.256641 | 0.03038752 | 0.1501 | 11.52169217 | 184.4120308 | 738.0545041 | 0.1338886 | 0.868006158 | 0.93645 | 0.535364292 | 8.56616821 | 8.56616821 | 606.897399 | 2.783115276 | TRUE | TRUE | TRUE |
| WFDC8 | 80.134141 | 0.03057057 | 0.77395 | 167.5791455 | 79980.4435 | 79980.4435 | 22230.8861 | 8.79438E-05 | 0.0011 | 113.562065 | 1116.180722 | 11238.72064 | 0.003604631 | -2.443139185 | TRUE | FALSE | FALSE |
| TAS2R10 | 80.093782 | 0.0307533 | 0.65165 | 80.09378206 | 7689.173287 | 78074.21117 | 106.058299 | 0.027803721 | 0.6516 | 106.0582991 | 10313.68321 | 105397.0565 | 0.755186371 | -0.121945856 | TRUE | FALSE | FALSE |
| NFIC | 79.963957 | 0.0309359 | 0.37425 | 79.96395668 | 1253.420762 | 6309.641293 | 91.7830392 | 0.032367988 | 0.37035 | 91.78303924 | 1432.216474 | 5736.788237 | 0.871228032 | -0.059868159 | TRUE | FALSE | FALSE |
| THUMPD3 | 79.113288 | 0.03111997 | 0.2328 | 8.611209753 | 7176.227106 | 14758.31204 | 0.21653749 | 0.848093645 | 0.5063 | 0.216537494 | 0.216537494 | 0.496104355 | 365.3560715 | 2.562716329 | TRUE | TRUE | TRUE |
| PSKH2 | 78.139669 | 0.03130582 | 0.03695 | 8.530968036 | 78.1396694 | 312.7420587 | 2.81276521 | 0.426011415 | 0.13505 | 2.812765206 | 11.25510784 | 11.25510784 | 27.780374 | 1.443738088 | TRUE | FALSE | FALSE |
| FAM222B | 78.123794 | 0.03149128 | 0.0023 | 2.308326022 | 2.308326022 | 8.446579572 | 0.30283537 | 0.824483017 | 0.01665 | 0.302835369 | 0.302835369 | 5.98775822 | 257.9744704 | 2.41157673 | TRUE | TRUE | TRUE |
| SLC28A1 | 77.882255 | 0.03167688 | 0.0055 | 0.937865405 | 77.88225525 | 77.88225525 | 0.02299621 | 0.892957326 | 1 | 0.022996208 | 0.022996208 | 0.454318377 | 3386.743373 | 3.52978229 | TRUE | TRUE | TRUE |
| CHDH | 77.130253 | 0.03186378 | 0.0844 | 3.902579679 | 143.8133515 | 575.4369792 | 1.59308438 | 0.526812207 | 0.26 | 0.398417996 | 1.593084377 | 3.487445559 | 48.41567365 | 1.684985979 | TRUE | FALSE | FALSE |
| UGT1A10 | 76.450473 | 0.03205185 | 0.1861 | 2.072677825 | 180.721055 | 722.7644696 | 0.36749555 | 0.803137013 | 0.874 | 3.337267975 | 31.08501017 | 31.08501017 | 208.0310155 | 2.318128089 | TRUE | TRUE | TRUE |
| GIMAP2 | 76.286485 | 0.03223988 | 0.5001 | 160.4821129 | 74616.85173 | 74616.85173 | 1.15158714 | 0.572125539 | 0.99155 | 991.2799554 | 101313.3799 | 101313.3799 | 66.24464856 | 1.8211508 | TRUE | FALSE | FALSE |
| SLC5A3 | 75.605052 | 0.0324291 | 0.97645 | 108407.0232 | 42985693.87 | 337493781.6 | 0.43841289 | 0.777872512 | 0.99925 | 1555.069807 | 251467.9775 | 1006430.497 | 172.45171 | 2.236667505 | TRUE | TRUE | TRUE |
| RASSF9 | 74.360392 | 0.0326209 | 0.07125 | 7.861213525 | 721.0091 | 721.0091 | 1.19723675 | 0.568042682 | 0.7511 | 1.197236751 | 11.13028432 | 11.13028432 | 62.11001431 | 1.793161629 | TRUE | FALSE | FALSE |
| C18orf54 | 74.350672 | 0.03281229 | 0.1283 | 7.757074683 | 74.35067181 | 74.35067181 | 10.3524549 | 0.194881615 | 0.1275 | 10.35245494 | 99.70521095 | 99.70521095 | 7.181936289 | 0.856241548 | TRUE | FALSE | FALSE |
| SIGIRR | 73.906583 | 0.03300434 | 0.3201 | 7.822177579 | 715.7906518 | 715.7906518 | 10.3933538 | 0.194203834 | 0.68915 | 10.39335378 | 963.3126984 | 963.3126984 | 7.110946571 | 0.851927416 | TRUE | FALSE | FALSE |
| IMPACT | 73.786812 | 0.03319626 | 0.0034 | 1.947011926 | 73.78681231 | 73.78681231 | 0.27214585 | 0.833126246 | 1 | 0.272145846 | 0.272145846 | 0.272145846 | 271.1296657 | 2.433177039 | TRUE | TRUE | TRUE |
| DDX51 | 73.744238 | 0.03338785 | 0.0693 | 2.012242103 | 73.74423843 | 73.74423843 | 0.74424038 | 0.644148838 | 0.50395 | 0.744240376 | 0.744240376 | 1.636421781 | 99.08658654 | 1.996014868 | TRUE | TRUE | TRUE |
| FANCA | 73.380179 | 0.03357992 | 0.348 | 18.33962432 | 9432.444904 | 151272.0713 | 0.11430426 | 0.87254688 | 0.7678 | 0.892914943 | 228.9934002 | 1836.160031 | 641.972419 | 2.80751637 | TRUE | TRUE | TRUE |
| TXLNB | 73.241225 | 0.0337719 | 0.3713 | 73.2412253 | 6596.350541 | 64824.08268 | 0.20993365 | 0.849717566 | 0.50515 | 0.209933653 | 0.209933653 | 0.821495785 | 348.8779631 | 2.542673538 | TRUE | TRUE | TRUE |
| OR52E4 | 72.871146 | 0.03396438 | 0.13295 | 20.23868674 | 195.5952968 | 195.5952968 | 0.27697325 | 0.83184665 | 1 | 24.14655938 | 234.2702907 | 234.2702907 | 263.0981343 | 2.420117768 | TRUE | TRUE | TRUE |
| OR4F5 | 71.109659 | 0.03416093 | 0.0054 | 7.309513377 | 71.10965913 | 71.10965913 | 0.45152902 | 0.773927515 | 1 | 0.451529024 | 0.451529024 | 0.451529024 | 157.4863528 | 2.197242925 | TRUE | TRUE | TRUE |
| BHMT | 70.46573 | 0.03435872 | 0.31855 | 7.43480651 | 684.5930026 | 684.5930026 | 1.13846193 | 0.573786908 | 0.7572 | 1.138461929 | 10.6024815 | 10.6024815 | 61.89555203 | 1.791659441 | TRUE | FALSE | FALSE |
| MRPL2 | 69.945127 | 0.03455745 | 0.27185 | 17.48043983 | 279.9439477 | 1120.709822 | 2.58561337 | 0.44058349 | 0.5004 | 2.585613368 | 10.34618908 | 10.34618908 | 27.05165759 | 1.432193882 | TRUE | FALSE | FALSE |
| BRWD1 | 69.763355 | 0.03475623 | 0.1385 | 7.348175231 | 678.5338823 | 6757.183597 | 0.17031845 | 0.859563697 | 0.29415 | 0.17031845 | 1.556550014 | 30.05569517 | 409.6053913 | 2.612365664 | TRUE | TRUE | TRUE |
| HYAL4 | 67.823117 | 0.03495986 | 0.8332 | 6532.726977 | 2765777.838 | 29440965.18 | 92.6895756 | 0.031464064 | 0.91105 | 9040.21333 | 968918.7235 | 10378172.76 | 0.731723245 | -0.135653148 | TRUE | FALSE | FALSE |
| ORC4 | 67.753534 | 0.03516323 | 0.38535 | 16.92701702 | 1086.326203 | 4351.479838 | 60.8429627 | 0.046145879 | 0.3913 | 15.2069792 | 974.6991086 | 3902.683461 | 1.11358046 | 0.046721602 | TRUE | FALSE | FALSE |
| SORBS3 | 67.73187 | 0.03536621 | 0.07485 | 7.127593971 | 659.763664 | 659.763664 | 1.06177496 | 0.582031238 | 0.8779 | 9.902765121 | 94.73107644 | 94.73107644 | 63.7911724 | 1.804760584 | TRUE | FALSE | FALSE |
| KIAA1328 | 67.194951 | 0.03557025 | 0.6558 | 67.19495051 | 6404.924175 | 64802.54342 | 93.7749058 | 0.030743429 | 0.1919 | 93.77490577 | 911.5426736 | 9077.209353 | 0.716555777 | -0.144749998 | TRUE | FALSE | FALSE |
| HOMEZ | 67.038558 | 0.03577428 | 0.99865 | 777285399.4 | 1.55012E+13 | 1.93603E+14 | 1.7893E+11 | 5.06746E-12 | 0.77885 | 2.61513E+13 | 7.0323E+17 | 1.28667E+20 | 3.74656E-10 | -9.426367601 | TRUE | FALSE | FALSE |
| PRTFDC1 | 66.718874 | 0.03597877 | 0.0341 | 3.020945479 | 66.71887389 | 266.7641744 | 11.3758567 | 0.184360985 | 0.2799 | 2.14095309 | 45.48785534 | 181.9346181 | 5.864953791 | 0.768264595 | TRUE | FALSE | FALSE |
| NAALADL2 | 66.535722 | 0.03618333 | 0.314 | 6.983156457 | 649.8336096 | 649.8336096 | 1.04113509 | 0.583623899 | 0.87825 | 9.733252997 | 93.32975277 | 93.32975277 | 63.90690543 | 1.805547788 | TRUE | FALSE | FALSE |
| AMER3 | 66.410308 | 0.0363878 | 0.1367 | 7.010092694 | 66.41030768 | 137.4734282 | 0.12287057 | 0.870650634 | 1 | 0.122870572 | 0.122870572 | 2.415235562 | 540.4899351 | 2.732787611 | TRUE | TRUE | TRUE |
| CLNK | 65.523569 | 0.03659438 | 0.5056 | 65.52356893 | 1051.49576 | 4213.811325 | 1606.72748 | 0.0015108 | 0.19735 | 241.4096501 | 13983.92811 | 56040.73002 | 0.040780761 | -1.389544676 | TRUE | FALSE | FALSE |
| FAM90A1 | 64.674559 | 0.03680301 | 0.06685 | 0.710196218 | 64.67455854 | 64.67455854 | 8.67272488 | 0.215849025 | 0.19385 | 0.914821878 | 84.27876262 | 839.0074587 | 7.457236272 | 0.872577903 | TRUE | FALSE | FALSE |
| F2RL1 | 64.512988 | 0.03701166 | 0.3176 | 16.13102981 | 627.8068952 | 627.8068952 | 20.0174329 | 0.132613261 | 0.13425 | 20.01743292 | 195.404824 | 195.404824 | 3.222840223 | 0.508238775 | TRUE | FALSE | FALSE |
| C11orf65 | 63.905328 | 0.03722168 | 0.1913 | 14.88876629 | 613.112744 | 6026.096717 | 8.63030432 | 0.216523001 | 0.93895 | 783.8438583 | 78414.26486 | 154263.2062 | 7.404759509 | 0.869510958 | TRUE | FALSE | FALSE |
| WDR31 | 63.389374 | 0.0374328 | 0.31915 | 6.693800881 | 615.3280424 | 615.3280424 | 1.0065619 | 0.58800569 | 0.87645 | 2.209049435 | 89.25201769 | 89.25201769 | 62.97613136 | 1.799175978 | TRUE | FALSE | FALSE |
| TMEM67 | 63.258089 | 0.03764387 | 0.26985 | 7.056890295 | 1369.202884 | 9291.723619 | 6.75775122 | 0.249627663 | 0.2753 | 1.690876912 | 133.1581271 | 911.9915528 | 9.360819469 | 0.97131387 | TRUE | FALSE | FALSE |
| PNP | 63.128754 | 0.03785487 | 0.2205 | 11.59164847 | 427.3990168 | 1010.087228 | 1.09960809 | 0.578698977 | 0.68765 | 2.544852345 | 40.70485834 | 40.70485834 | 57.41023019 | 1.758989288 | TRUE | FALSE | FALSE |
| BRI3BP | 62.909576 | 0.03806609 | 0.12855 | 6.488352211 | 625.5998683 | 625.5998683 | 0.78957678 | 0.62408775 | 1 | 0.78957678 | 0.78957678 | 0.78957678 | 79.67505887 | 1.901322393 | TRUE | TRUE | TRUE |
| ZNHIT3 | 62.873408 | 0.03827696 | 0.06685 | 6.695312251 | 605.2195257 | 605.2195257 | 0.12308991 | 0.870577061 | 1 | 0.123089905 | 0.123089905 | 0.123089905 | 510.792564 | 2.708244566 | TRUE | TRUE | TRUE |
| STON2 | 62.850688 | 0.03848744 | 0.0789 | 6.598754917 | 613.6240959 | 613.6240959 | 0.96018441 | 0.591979228 | 0.93815 | 8.965687019 | 842.9554839 | 842.9554839 | 65.45689301 | 1.815955387 | TRUE | TRUE | TRUE |
| METTL2A | 62.61824 | 0.03869818 | 0.0644 | 6.646546775 | 604.719798 | 604.719798 | 0.1217692 | 0.870860408 | 0.0069 | 0.121769198 | 0.121769198 | 0.121769198 | 514.237109 | 2.711163414 | TRUE | TRUE | TRUE |
| CALML4 | 62.436513 | 0.03890903 | 0.72025 | 249.6648308 | 39260.81861 | 395258.7225 | 834.239639 | 0.00274161 | 0.42645 | 338.1452216 | 33715.95024 | 348879.0666 | 0.074842419 | -1.125852181 | TRUE | FALSE | FALSE |
| MMEL1 | 62.239184 | 0.03912003 | 0.5582 | 93.89626803 | 8738.351434 | 18439.35219 | 137.325178 | 0.02048533 | 0.0681 | 8.559003782 | 137.3251776 | 1158.744608 | 0.45322486 | -0.343686277 | TRUE | FALSE | FALSE |
| NDUFB3 | 62.198218 | 0.03933071 | 0.6427 | 62.19821838 | 5888.867533 | 23564.77472 | 0.48476847 | 0.7611706 | 1 | 31.03948806 | 291.5923592 | 1167.13184 | 128.3050004 | 2.108243582 | TRUE | TRUE | TRUE |
| ZNF143 | 61.449357 | 0.03954328 | 0.00005 | 1.815418152 | 1.815418152 | 6.679563489 | 0.24595812 | 0.840320966 | 1 | 0.245958125 | 0.245958125 | 4.866520045 | 249.8366606 | 2.397656166 | TRUE | TRUE | TRUE |
| CALHM2 | 60.903029 | 0.03975714 | 0.7743 | 585.7420308 | 58219.66125 | 114376.652 | 1.08002364 | 0.580427764 | 0.8761 | 9.985330321 | 94.69053803 | 94.69053803 | 56.39045917 | 1.751205631 | TRUE | FALSE | FALSE |
| BTG3 | 60.423818 | 0.03997208 | 0.25355 | 6.227211541 | 60.42381806 | 60.42381806 | 6.69557125 | 0.252221528 | 0.00555 | 6.695571251 | 6.695571251 | 6.695571251 | 9.024445532 | 0.955420528 | TRUE | FALSE | FALSE |
| UQCR10 | 60.15749 | 0.04018742 | 0.8903 | 595.7019569 | 62869.76782 | 669273.0977 | 0.8180673 | 0.61491123 | 1 | 7.740775053 | 7.740775053 | 7.740775053 | 73.5361134 | 1.866500672 | TRUE | TRUE | TRUE |
| LCE4A | 59.724982 | 0.04040372 | 0.50335 | 59.72498216 | 590.9766383 | 590.9766383 | 0.76051131 | 0.635984315 | 0.0022 | 0.760511311 | 0.760511311 | 0.760511311 | 78.53266785 | 1.895050351 | TRUE | TRUE | TRUE |
| TMC8 | 59.642834 | 0.04061983 | 0.15535 | 8.167348796 | 130.8155343 | 523.7344233 | 115.604681 | 0.025191818 | 0.46495 | 115.6046805 | 7413.940549 | 29690.93534 | 0.515920584 | -0.287417144 | TRUE | FALSE | FALSE |
| CCM2L | 59.595818 | 0.04083563 | 0.1344 | 6.362459927 | 59.59581847 | 59.59581847 | 0.11462124 | 0.872465063 | 1 | 0.114621244 | 0.114621244 | 0.114621244 | 519.9369376 | 2.715950672 | TRUE | TRUE | TRUE |
| VSX2 | 59.316983 | 0.04105189 | 0.03715 | 3.706144578 | 14.82506025 | 59.31698341 | 0.08588367 | 0.879004326 | 0.00695 | 0.085883669 | 0.085883669 | 0.085883669 | 690.6666234 | 2.839268469 | TRUE | TRUE | TRUE |
| ACY1 | 58.95593 | 0.04126888 | 0.01055 | 6.067098248 | 58.95592971 | 120.1328553 | 0.71939264 | 0.656155313 | 1 | 0.719392639 | 0.719392639 | 14.26155891 | 81.95236722 | 1.913561503 | TRUE | TRUE | TRUE |
| DMGDH | 58.757926 | 0.04148608 | 0.6998 | 547.9771926 | 204450.3603 | 3270885.797 | 1264.90637 | 0.001930683 | 0.6069 | 3050.79424 | 3134614.229 | 33047478.56 | 0.046452392 | -1.33299192 | TRUE | FALSE | FALSE |
| SURF4 | 58.40021 | 0.04170402 | 0.59555 | 58.40020997 | 2249.095724 | 9002.190973 | 0.45505512 | 0.772838353 | 0.752 | 4.273539354 | 41.19048742 | 41.19048742 | 128.3365626 | 2.108350403 | TRUE | TRUE | TRUE |
| SI | 58.399959 | 0.04192149 | 0.4142 | 14.59670519 | 15001.15507 | 240784.8773 | 18013.7769 | 0.000107999 | 0.1277 | 140.6427508 | 144304.6391 | 2314741.969 | 0.003241961 | -2.489192237 | TRUE | FALSE | FALSE |
| PLBD1 | 57.863042 | 0.04214033 | 0.11645 | 6.372084939 | 231.4920682 | 926.3593826 | 0.03605454 | 0.889889503 | 0.50945 | 0.144209368 | 0.144209368 | 0.715015978 | 1604.87532 | 3.205441298 | TRUE | TRUE | TRUE |
| PDE6H | 57.850735 | 0.04235875 | 0.25215 | 6.002473613 | 57.85073518 | 57.85073518 | 0.6950577 | 0.668264083 | 1 | 0.695057704 | 6.54164211 | 6.54164211 | 83.23155743 | 1.920288021 | TRUE | TRUE | TRUE |
| ASB2 | 57.209416 | 0.04257891 | 0.0651 | 0.648858739 | 57.20941621 | 57.20941621 | 2.16292445 | 0.474829391 | 0.13775 | 0.24282576 | 2.162924449 | 4.780694156 | 26.45002983 | 1.422426166 | TRUE | FALSE | FALSE |
| BLK | 56.668986 | 0.0428005 | 0.298 | 56.66898599 | 1158.662553 | 14568.66483 | 1704.7536 | 0.001359324 | 0.4082 | 425.7429434 | 27353.62822 | 439340.6893 | 0.033241746 | -1.478316175 | TRUE | FALSE | FALSE |
| RSPH1 | 56.195963 | 0.0430233 | 0.31435 | 56.1959625 | 1344.145371 | 5375.87036 | 0.88118446 | 0.601208839 | 0.7579 | 2.035305139 | 8.140270728 | 8.140270728 | 63.77321096 | 1.804638284 | TRUE | TRUE | TRUE |

| | | | | | | | | | | | | | | | | | |
|---|---|---|---|---|---|---|---|---|---|---|---|---|---|---|---|---|---|
| GAL3ST3 | 56.178361 | 0.04324568 | 0.21 | 14.04386608 | 536.6666123 | 2147.313575 | 0.13584114 | 0.867623213 | 1 | 0.543153775 | 2.172315299 | 2.689214441 | 413.5592715 | 2.616537762 | TRUE | TRUE | TRUE |
| MPP4 | 55.917616 | 0.04346853 | 0.83425 | 2113.019229 | 512571.5951 | 5306642.34 | 0.16986222 | 0.85973521 | 0.74915 | 0.169862224 | 1.517772005 | 3.362272444 | 329.1939464 | 2.51745184 | TRUE | TRUE | TRUE |
| ZNF678 | 55.767391 | 0.04369144 | 0.63155 | 528.8479703 | 518567.9627 | 5388326.383 | 7180.4942 | 0.000242506 | 0.9531 | 7636814.293 | 887105986.8 | 81723990216 | 0.007766512 | -2.109774008 | TRUE | FALSE | FALSE |
| PLD2 | 55.558935 | 0.04391463 | 0.73435 | 505.7256382 | 109670.1685 | 1093873.668 | 3.21410658 | 0.39912033 | 0.8339 | 268.161369 | 243669.0033 | 2468306.188 | 17.28596544 | 1.23769364 | TRUE | FALSE | FALSE |
| QPCT | 55.446367 | 0.04413776 | 0.06885 | 5.878826547 | 536.0479303 | 536.0479303 | 0.105868 | 0.874392602 | 0.5032 | 0.957280166 | 0.957280166 | 2.103862257 | 523.7311462 | 2.719108402 | TRUE | TRUE | TRUE |
| WDR49 | 55.388598 | 0.04436063 | 0.373 | 55.38859752 | 5027.624098 | 49601.0269 | 0.15807671 | 0.862544029 | 0.75135 | 0.15807671 | 1.405535277 | 3.129528815 | 350.3906278 | 2.544552481 | TRUE | TRUE | TRUE |
| GZMK | 54.701186 | 0.04458554 | 0.13165 | 8.088186717 | 129.3427172 | 517.428326 | 1.1600113 | 0.571409934 | 0.75095 | 4.638488625 | 44.02801211 | 44.02801211 | 47.15573569 | 1.673534525 | TRUE | FALSE | FALSE |
| PES1 | 54.651679 | 0.04481015 | 0.94615 | 13926.25555 | 6379026.07 | 70474319.04 | 0.18721873 | 0.855179412 | 1 | 17137.91429 | 21740938.45 | 245831896.6 | 291.9135206 | 2.465254211 | TRUE | TRUE | TRUE |
| STPG2 | 53.861454 | 0.04503724 | 0.3229 | 5.711747489 | 520.6373259 | 520.6373259 | 72.3373191 | 0.039787554 | 0.50145 | 72.33731911 | 703.2466172 | 7003.866242 | 0.744587362 | -0.12808434 | TRUE | FALSE | FALSE |
| SPRR4 | 53.475766 | 0.0452653 | 0.25115 | 5.560057672 | 53.47576585 | 53.47576585 | 57.0187053 | 0.049165666 | 0.00125 | 5.919155748 | 57.01870532 | 57.01870532 | 0.937863558 | -0.027860339 | TRUE | FALSE | FALSE |
| KRTAP19-3 | 53.265287 | 0.04549369 | 0.6868 | 53.26528689 | 5313.530385 | 5313.530385 | 6.09156838 | 0.26663776 | 0.8743 | 58.73585482 | 580.8676629 | 580.8676629 | 8.74410062 | 0.941715146 | TRUE | FALSE | FALSE |
| KIF19 | 52.109528 | 0.04572608 | 0.16035 | 2.544291742 | 208.3383502 | 1955.519907 | 3.54510942 | 0.381094154 | 0.39575 | 3.545109423 | 277.1992256 | 1249.25401 | 14.69898987 | 1.167287491 | TRUE | FALSE | FALSE |
| ACSL6 | 51.839828 | 0.04595905 | 0.7784 | 509.1557739 | 52775.09148 | 173204.8929 | 7240.47503 | 0.000236046 | 0.2362 | 716.1441672 | 74949.20734 | 142840.4576 | 0.007159728 | -2.145103505 | TRUE | FALSE | FALSE |
| CGB2 | 51.698502 | 0.04619209 | 0.6857 | 51.69850197 | 5177.608321 | 5177.608321 | 5361.90431 | 0.000334226 | 0.18475 | 110.4571596 | 5361.904314 | 55653.84361 | 0.009641817 | -2.0158411 | TRUE | FALSE | FALSE |
| TPSD1 | 51.693309 | 0.04642467 | 0.06935 | 5.392825374 | 507.9185976 | 507.9185976 | 0.37902603 | 0.799572727 | 0.75285 | 0.379026025 | 3.477171687 | 3.477171687 | 136.3845885 | 2.134765298 | TRUE | TRUE | TRUE |
| TRNT1 | 51.556902 | 0.0466573 | 0.4726 | 30.1352519 | 1928.829097 | 4620.499937 | 2.19528893 | 0.47256255 | 0.72045 | 5.050757779 | 80.80095304 | 323.3026524 | 23.48524679 | 1.370795128 | TRUE | FALSE | FALSE |
| SEMG1 | 51.025799 | 0.04689158 | 0.99985 | 39515293670 | 7.94115E+14 | 1.26218E+17 | 101.549286 | 0.028818438 | 0.98985 | 1052454.267 | 122906028.1 | 12414673670 | 0.502473241 | -0.298887062 | TRUE | FALSE | FALSE |
| PDCL2 | 50.882148 | 0.04712595 | 0.00315 | 5.28679367 | 50.88214775 | 50.88214775 | 0.64201389 | 0.693451351 | 1 | 0.642013888 | 0.642013888 | 0.642013888 | 79.25396741 | 1.899021012 | TRUE | TRUE | TRUE |
| ZNF397 | 50.419921 | 0.04736171 | 0.066 | 5.393901583 | 483.1174328 | 483.1174328 | 0.10235052 | 0.875399647 | 1 | 0.102350521 | 0.102350521 | 0.102350521 | 492.6200706 | 2.692512102 | TRUE | TRUE | TRUE |
| ZNF780A | 50.283908 | 0.04759755 | 0.18965 | 50.28390776 | 486.2952165 | 4817.92981 | 0.10000614 | 0.875897513 | 1 | 0.10000614 | 0.905334 | 0.905334 | 502.8082044 | 2.701402356 | TRUE | TRUE | TRUE |
| THBS4 | 49.977536 | 0.04783416 | 0.55545 | 88.67584506 | 22752.59128 | 205098.7433 | 29.0257764 | 0.094583421 | 0.6969 | 116.1502349 | 16505.73386 | 66099.17706 | 1.721832861 | 0.235990992 | TRUE | FALSE | FALSE |
| HEPACAM | 49.535216 | 0.04807212 | 0.49915 | 10.81558296 | 490.8462753 | 490.8462753 | 0.62329177 | 0.701906031 | 1 | 5.884222662 | 5.884222662 | 12.38737403 | 79.47356088 | 1.900222672 | TRUE | TRUE | TRUE |
| CD82 | 49.489093 | 0.04830978 | 0.37055 | 49.48909268 | 792.6242019 | 3173.283982 | 117.612055 | 0.024876075 | 0.14835 | 29.38335182 | 470.8805985 | 1885.724218 | 0.420782483 | -0.375942348 | TRUE | FALSE | FALSE |
| CHCHD6 | 48.941086 | 0.04854927 | 0.83275 | 195.9516678 | 12595.79738 | 50481.78196 | 240.11891 | 0.010418804 | 0.49615 | 59.97181212 | 3852.211077 | 15435.28105 | 0.203820205 | -0.690752766 | TRUE | FALSE | FALSE |
| TBX1 | 48.928723 | 0.04878831 | 0.1277 | 5.0299396 | 48.92872315 | 48.92872315 | 0.08818724 | 0.878566559 | 1 | 0.088187244 | 0.088187244 | 1.747288641 | 554.8276692 | 2.744158111 | TRUE | TRUE | TRUE |
| ARL11 | 48.887262 | 0.04902704 | 0.5009 | 48.88726216 | 783.4479487 | 973.1931933 | 58.7749053 | 0.0479917 | 0.6351 | 58.77490533 | 941.9685146 | 3772.430626 | 0.831771006 | -0.079996222 | TRUE | FALSE | FALSE |
| COL16A1 | 48.863255 | 0.04926537 | 0.3126 | 3.049353372 | 3140.18243 | 50444.04627 | 8835.95925 | 0.000211903 | 0.33815 | 2207.219599 | 2435282.496 | 146601036.1 | 0.005530045 | -2.257271314 | TRUE | FALSE | FALSE |
| CNTN2 | 48.310569 | 0.04950559 | 0.27355 | 20.48692161 | 773.2060988 | 7480.70681 | 1.86915315 | 0.501134556 | 0.82595 | 42.90828387 | 2744.79353 | 27298.39642 | 25.84623379 | 1.412397268 | TRUE | FALSE | FALSE |
| CRP | 48.128572 | 0.0497461 | 0.25685 | 5.004728461 | 48.12857199 | 48.12857199 | 0.61213846 | 0.706614319 | 1 | 0.612138462 | 0.612138462 | 0.612138462 | 78.62366921 | 1.895553308 | TRUE | TRUE | TRUE |
| AARD | 47.893893 | 0.04998713 | 0.50575 | 471.9041824 | 4766.042274 | 49309.87423 | 461.139964 | 0.005052394 | 0.6368 | 4652.57037 | 510978.9473 | 5532879.478 | 0.103859774 | -0.983552626 | TRUE | FALSE | FALSE |
| SLC25A23 | 47.690764 | 0.05022856 | 0.07385 | 11.91864522 | 190.8754695 | 764.1425501 | 4.42696026 | 0.334280864 | 0.32415 | 1.106819001 | 17.71100262 | 21.86322071 | 10.77280145 | 1.032328656 | TRUE | FALSE | FALSE |
| ASIC3 | 47.430277 | 0.05047064 | 0.9822 | 468083.7998 | 585389509.7 | 6577348726 | 67164.7645 | 2.86378E-05 | 0.93525 | 879300866.5 | 1.55687E+13 | 1.918E+14 | 0.000706178 | -3.151085838 | TRUE | FALSE | FALSE |
| ACTL7A | 47.411932 | 0.0507123 | 0.88205 | 759.6136652 | 195570.3142 | 783882.8017 | 21467.4097 | 9.974E-05 | 0.8774 | 777286.7985 | 250654321.7 | 1794095472 | 0.002208554 | -2.655891993 | TRUE | FALSE | FALSE |
| CD300LB | 47.363003 | 0.05095369 | 0.2567 | 4.916452783 | 47.3630029 | 47.3630029 | 5.3125295 | 0.293943851 | 0.7542 | 5.312529503 | 51.24925651 | 51.24925651 | 8.915339271 | 0.950137875 | TRUE | FALSE | FALSE |
| BCL2L12 | 47.274642 | 0.05119497 | 0.3161 | 5.08649275 | 450.392996 | 450.392996 | 0.09232958 | 0.877682298 | 1 | 0.823270558 | 7.534395447 | 7.534395447 | 512.0205389 | 2.709287382 | TRUE | TRUE | TRUE |
| PLOD3 | 47.225193 | 0.05143598 | 0.2424 | 6.745584702 | 431.6206047 | 1727.215928 | 0.64512926 | 0.692480502 | 0.6668 | 1.448259676 | 23.16775023 | 92.69692073 | 73.20268386 | 1.864527004 | TRUE | TRUE | TRUE |
| FHL3 | 47.112924 | 0.05167699 | 0.5033 | 188.5436893 | 3021.90728 | 12102.58868 | 4302.64936 | 0.000426872 | 0.1295 | 268.4059406 | 4302.649359 | 4302.649359 | 0.010949747 | -1.960595896 | TRUE | FALSE | FALSE |
| ARHGAP11B | 46.972008 | 0.05191814 | 0.31685 | 46.97200827 | 4682.269086 | 4682.269086 | 52.5718662 | 0.052791507 | 0.1314 | 52.57186619 | 519.7390962 | 519.7390962 | 0.89348185 | -0.048914265 | TRUE | FALSE | FALSE |
| BDH1 | 46.944077 | 0.05215892 | 0.11265 | 2.93283943 | 46.94407695 | 187.8839818 | 4.31586978 | 0.338003924 | 0.3237 | 1.079063477 | 17.26625656 | 17.26625656 | 10.87708371 | 1.036512471 | TRUE | FALSE | FALSE |
| KALRN | 46.466636 | 0.05240135 | 0.52465 | 46.46663559 | 4757.056228 | 49838.07855 | 0.11165484 | 0.872954574 | 0.88595 | 1.046054973 | 10.05175448 | 189.8523742 | 416.1631984 | 2.619263673 | TRUE | TRUE | TRUE |
| ANXA9 | 46.43635 | 0.05264342 | 0.5021 | 46.43634962 | 442.3124223 | 4316.138361 | 0.83576536 | 0.611050926 | 0.75295 | 0.835765365 | 7.653924814 | 7.653924814 | 55.56146685 | 1.744773703 | TRUE | TRUE | TRUE |
| VSTM2A | 46.360413 | 0.05288534 | 0.4988 | 4.802081066 | 459.0555859 | 459.0555859 | 5.58921877 | 0.284025581 | 0.50525 | 5.589218766 | 5.589218766 | 5.589218766 | 8.29461406 | 0.918796184 | TRUE | FALSE | FALSE |
| PDE11A | 46.339436 | 0.05312687 | 0.2992 | 11.58901718 | 2966.976342 | 26434.80081 | 133.893938 | 0.021359828 | 0.36365 | 33.48211014 | 8575.43454 | 77773.19163 | 0.346090617 | -0.460810174 | TRUE | FALSE | FALSE |
| TAAR5 | 46.061189 | 0.05336917 | 0.5027 | 46.06118909 | 737.8519694 | 2954.261809 | 0.46368857 | 0.769571024 | 0.7509 | 0.46368857 | 1.854789893 | 1.854789893 | 99.33647735 | 1.997108755 | TRUE | TRUE | TRUE |
| MSS51 | 45.842966 | 0.05361197 | 0.74915 | 4321.328258 | 447670.168 | 4714341.564 | 650740.587 | 2.67429E-06 | 0.02285 | 6189.114243 | 650740.5873 | 6903697.036 | 7.04474E-05 | -4.152135188 | TRUE | FALSE | FALSE |
| TNFAIP6 | 45.835696 | 0.05385431 | 0.3207 | 11.45922858 | 435.861516 | 435.861516 | 0.19053293 | 0.854183661 | 0.0062 | 0.190532928 | 0.190532928 | 0.190532928 | 240.5657483 | 2.381233793 | TRUE | TRUE | TRUE |
| GPR15 | 45.827044 | 0.05409621 | 0.84995 | 1701.372658 | 117604.4634 | 471262.1467 | 34.1383037 | 0.081871443 | 0.59445 | 34.13830371 | 1302.621465 | 5213.48381 | 1.342393718 | 0.127879911 | TRUE | FALSE | FALSE |
| TAGLN | 45.679664 | 0.05433829 | 0.07035 | 2.85348969 | 45.67966395 | 45.67966395 | 3.53734058 | 0.381562091 | 0.4978 | 0.884394686 | 14.15194595 | 14.15194595 | 12.91356117 | 1.111046024 | TRUE | FALSE | FALSE |
| HLA-DOB | 45.61757 | 0.05458016 | 0.50065 | 45.61756992 | 449.5555304 | 449.5555304 | 47.9519744 | 0.057344076 | 0.31 | 47.95197438 | 4780.857525 | 4780.857525 | 0.951317866 | -0.021674347 | TRUE | FALSE | FALSE |
| CPEB1 | 45.596351 | 0.05482165 | 0.50715 | 45.59635057 | 451.9612145 | 451.9612145 | 5.1254884 | 0.302465573 | 0.87785 | 49.58466744 | 491.9919746 | 491.9919746 | 8.896001129 | 0.949194829 | TRUE | FALSE | FALSE |
| DES | 45.50261 | 0.05506308 | 0.37375 | 11.37286916 | 182.1004544 | 728.944016 | 4.20368777 | 0.343273005 | 0.656 | 4.203687769 | 67.28809859 | 269.3114706 | 10.82445049 | 1.034405859 | TRUE | FALSE | FALSE |
| ZNF517 | 45.357933 | 0.05530469 | 0.0751 | 4.851051848 | 434.7322961 | 434.7322961 | 0.0932045 | 0.877502145 | 0.015 | 0.093204496 | 0.093204496 | 1.841960242 | 486.6496216 | 2.68721639 | TRUE | TRUE | TRUE |
| CES1 | 45.224185 | 0.05554643 | 0.99905 | 14286383.42 | 16527062541 | 2.6509E+11 | 2.40630121 | 0.454516754 | 0.99945 | 339931.7139 | 397394184.7 | 6032528685 | 18.79406629 | 1.274020754 | TRUE | FALSE | FALSE |
| ERMP1 | 44.95089 | 0.05578896 | 0.3208 | 4.788839055 | 432.5100635 | 432.5100635 | 7.17171809 | 0.241778203 | 0.258 | 0.776750096 | 7.171718088 | 15.32510608 | 6.2677994 | 0.797115088 | TRUE | FALSE | FALSE |
| PLEKHN1 | 44.777992 | 0.05603181 | 0.06665 | 0.565079211 | 44.77799192 | 44.77799192 | 0.01470987 | 0.895204531 | 1 | 0.124899265 | 0.124899265 | 0.290635595 | 3044.078924 | 3.483455908 | TRUE | TRUE | TRUE |
| WWOX | 44.648089 | 0.05627478 | 0.75045 | 440.0089772 | 45843.66981 | 484149.1 | 0.01397368 | 0.895447334 | 0.50245 | 0.126343403 | 0.126343403 | 0.277070792 | 3195.156035 | 3.504492072 | TRUE | TRUE | TRUE |
| FKBP7 | 44.142619 | 0.05651965 | 0.8315 | 424.4588965 | 42242.91351 | 436706.6217 | 5466.67201 | 0.000309273 | 0.11055 | 138.0653899 | 55443.23049 | 143951.1246 | 0.008074861 | -2.092864925 | TRUE | FALSE | FALSE |
| CA7 | 43.88374 | 0.05676528 | 0.24915 | 4.540910695 | 43.8837403 | 43.8837403 | 2.21438382 | 0.471335174 | 0.00555 | 2.214383818 | 2.214383818 | 2.214383818 | 19.81758534 | 1.297050737 | TRUE | FALSE | FALSE |
| SF1 | 43.805166 | 0.05701079 | 0.6987 | 43.80516638 | 4422.063028 | 4422.063028 | 47.8674431 | 0.057854647 | 0.68765 | 47.86744309 | 4840.619078 | 4840.619078 | 0.915134871 | -0.038514895 | TRUE | FALSE | FALSE |
| LCE3D | 43.574026 | 0.05725693 | 0.0019 | 4.553808401 | 43.57402561 | 43.57402561 | 0.56519124 | 0.726428556 | 1 | 0.56519124 | 0.56519124 | 0.56519124 | 77.09607395 | 1.887032263 | TRUE | TRUE | TRUE |
| CYP1A2 | 43.494201 | 0.05750296 | 0.0196 | 0.30252511 | 10.87619576 | 43.49420142 | 5.59902427 | 0.283112932 | 0.1912 | 0.617570025 | 13.02472667 | 52.06669179 | 7.768175195 | 0.890319012 | TRUE | FALSE | FALSE |
| C1QTNF3 | 43.396204 | 0.05774898 | 0.06875 | 4.496142401 | 429.3410678 | 429.3410678 | 0.08334049 | 0.879522263 | 0.00785 | 0.083340492 | 0.083340492 | 0.083340492 | 520.7097106 | 2.716595677 | TRUE | TRUE | TRUE |
| IFI44 | 43.196137 | 0.05799548 | 0.7776 | 412.258703 | 40351.22066 | 79985.92531 | 7.17484146 | 0.241449935 | 0.31555 | 7.174841461 | 651.2094098 | 651.2094098 | 6.020500499 | 0.779632597 | TRUE | FALSE | FALSE |
| CREB1 | 43.110335 | 0.05824192 | 0.25145 | 4.449106012 | 43.11033502 | 43.11033502 | 0.54889183 | 0.733643524 | 1 | 0.548891831 | 0.548891831 | 0.548891831 | 78.54067525 | 1.895094631 | TRUE | TRUE | TRUE |
| GPR25 | 43.037918 | 0.05848822 | 0.0112 | 4.460683372 | 43.03791786 | 88.20367853 | 0.55405665 | 0.731558161 | 1 | 0.554056651 | 0.554056651 | 10.97060514 | 77.67782916 | 1.89029708 | TRUE | TRUE | TRUE |
| RGS21 | 42.976802 | 0.05873432 | 0.2563 | 4.478744095 | 42.97680245 | 42.97680245 | 0.55597408 | 0.730502466 | 0.00335 | 0.555974083 | 0.555974083 | 0.555974083 | 77.30001043 | 1.888179552 | TRUE | TRUE | TRUE |
| MCL1 | 42.84629 | 0.05898059 | 0.01045 | 4.452282486 | 42.84628983 | 88.1212736 | 0.55202318 | 0.732605117 | 1 | 0.552023182 | 0.552023182 | 10.9394973 | 77.61683058 | 1.889955905 | TRUE | TRUE | TRUE |
| SPRR3 | 42.605587 | 0.05922755 | 0.6852 | 42.60558682 | 4206.144503 | 4206.144503 | 0.59005816 | 0.716553483 | 1 | 0.590058155 | 0.590058155 | 0.590058155 | 72.2057418 | 1.858571734 | TRUE | TRUE | TRUE |
| ZNF587 | 42.568664 | 0.05947421 | 0.8579 | 411.4970456 | 41318.33316 | 428707.2893 | 488152.514 | 3.91323E-06 | 0.698 | 1592148.886 | 2712005658 | 31246690510 | 8.72036E-05 | -4.059465506 | TRUE | FALSE | FALSE |
| SLC2A5 | 42.326397 | 0.05972158 | 0.2519 | 4.479395522 | 358.89979 | 1639.191307 | 0.39155916 | 0.795096507 | 0.51195 | 0.391559161 | 3.546451756 | 3.546451756 | 108.0970684 | 2.033813916 | TRUE | TRUE | TRUE |
| NEBL | 42.17579 | 0.05996922 | 0.49645 | 4.835047846 | 3443.519772 | 32197.33984 | 60571701.3 | 2.78911E-08 | 0.0692 | 62282.97649 | 242252691.5 | 2503254462 | 6.96295E-07 | -6.157206546 | TRUE | FALSE | FALSE |
| RGL4 | 41.622047 | 0.06021917 | 0.31575 | 4.482860853 | 396.1371378 | 396.1371378 | 0.75977679 | 0.636679929 | 0.5065 | 0.759776789 | 0.759776789 | 1.689267589 | 54.78194081 | 1.738637414 | TRUE | TRUE | TRUE |
| ATAD2 | 41.582549 | 0.06046883 | 0.81595 | 4249.726834 | 1903003.361 | 37075030.58 | 0.00877352 | 0.897090536 | 1 | 267442947.2 | 5.23429E+12 | 2.06241E+14 | 4739.550368 | 3.675737143 | TRUE | TRUE | TRUE |

| | | | | | | | | | | | | | | | | | |
|---|---|---|---|---|---|---|---|---|---|---|---|---|---|---|---|---|---|
| BPIFB4 | 41.472934 | 0.06071856 | 0.5069 | 41.47293439 | 395.6334945 | 3866.481674 | 0.73232645 | 0.650421811 | 0.87505 | 6.709894421 | 63.05865699 | 63.05865699 | 56.6317578 | 1.753060042 | TRUE | TRUE | TRUE |
| KLHDC7B | 40.644411 | 0.06097208 | 0.34865 | 40.64441105 | 3899.273436 | 39578.82366 | 0.70517802 | 0.664134323 | 0.68945 | 6.544356632 | 607.8175367 | 607.8175367 | 57.63709315 | 1.76070207 | TRUE | TRUE | TRUE |
| LPCAT2 | 40.422918 | 0.06122628 | 1 | 2.30976E+47 | 9.19944E+56 | 7.18919E+60 | 351.55209 | 0.006786906 | 1 | 1.11847E+36 | 1.36475E+44 | 7.45671E+47 | 0.114984149 | -0.939362023 | TRUE | FALSE | FALSE |
| TRMT5 | 40.402168 | 0.06148008 | 0.85875 | 385.2492609 | 37640.36734 | 385254.2476 | 57079.9326 | 3.50992E-05 | 0.06695 | 573.8630949 | 57079.93257 | 112264.1845 | 0.000707817 | -3.150078779 | TRUE | FALSE | FALSE |
| MSH3 | 40.265241 | 0.06173411 | 0.7846 | 644.5039661 | 349486.1519 | 5607997.292 | 188.756538 | 0.014255736 | 0.3539 | 188.7565376 | 48602.90156 | 194815.1619 | 0.21331839 | -0.670971703 | TRUE | FALSE | FALSE |
| FANCM | 40.213342 | 0.06198792 | 0.6336 | 40.21334206 | 34529.30909 | 343252.2709 | 108.117451 | 0.027142452 | 0.12195 | 5.17967778 | 432.3633581 | 4093.898127 | 0.37194127 | -0.42952563 | TRUE | FALSE | FALSE |
| LRAT | 40.07844 | 0.06224195 | 0.2573 | 4.184708976 | 40.07843971 | 40.07843971 | 0.46281197 | 0.76990217 | 0.93785 | 41.38233492 | 4094.948124 | 4094.948124 | 86.5976723 | 1.937506219 | TRUE | TRUE | TRUE |
| DEFB134 | 39.80787 | 0.06249693 | 0.00115 | 4.16844339 | 39.8078697 | 39.8078697 | 0.52357395 | 0.745224523 | 1 | 0.523573951 | 0.523573951 | 0.523573951 | 76.03103562 | 1.880990906 | TRUE | TRUE | TRUE |
| WDR1 | 39.680858 | 0.0627521 | 0.50875 | 393.3560336 | 3996.949785 | 41603.56032 | 4.53420518 | 0.327535274 | 0.97145 | 435.2950605 | 4427.531028 | 46131.49184 | 8.751447401 | 0.942079887 | TRUE | FALSE | FALSE |
| NT5C | 39.258917 | 0.06300905 | 0.50105 | 39.25891661 | 1543.695074 | 1543.695074 | 0.53658412 | 0.739784284 | 1 | 0.536584117 | 0.536584117 | 0.536584117 | 73.16451483 | 1.864300497 | TRUE | TRUE | TRUE |
| ARL9 | 39.142973 | 0.06326614 | 0.6961 | 39.14297347 | 3911.502355 | 3911.502355 | 44.7765317 | 0.060972019 | 0.12315 | 4.640262487 | 443.1571977 | 443.1571977 | 0.874185026 | -0.058396637 | TRUE | FALSE | FALSE |
| PIAS2 | 38.880785 | 0.06352417 | 0.69365 | 38.88078537 | 3884.028508 | 3884.028508 | 4.61496886 | 0.322591805 | 0.88305 | 44.52602787 | 440.6143607 | 440.6143607 | 8.424929081 | 0.925566254 | TRUE | FALSE | FALSE |
| UGT1A4 | 38.758889 | 0.06378237 | 0.3241 | 9.692152943 | 375.5575844 | 375.5575844 | 0.55904934 | 0.729112116 | 0.9704 | 48.87628099 | 1182.556773 | 4729.18062 | 69.32999678 | 1.84092118 | TRUE | TRUE | TRUE |
| EIF6 | 38.707337 | 0.06404035 | 0.06945 | 2.417700725 | 38.70733674 | 38.70733674 | 2.9859645 | 0.413060486 | 0.5017 | 11.94648571 | 47.80839919 | 191.3712595 | 12.9630934 | 1.11270865 | TRUE | FALSE | FALSE |
| MTFMT | 38.659979 | 0.06429809 | 0.50935 | 38.659979 | 1491.962386 | 5969.362037 | 0.63501688 | 0.697270957 | 0.87485 | 0.635016885 | 10.1634714 | 10.1634714 | 60.88023787 | 1.784476341 | TRUE | TRUE | TRUE |
| ENPP7 | 38.53833 | 0.06455602 | 0.0626 | 1.084557692 | 38.53832976 | 154.203223 | 0.00874392 | 0.897111294 | 0.5078 | 0.034947803 | 0.034947803 | 0.172830123 | 4407.441052 | 3.644186512 | TRUE | TRUE | TRUE |
| C20orf96 | 38.401215 | 0.0648142 | 0.195 | 38.40121462 | 364.0220501 | 3535.145656 | 0.08246686 | 0.879697076 | 1 | 0.733298597 | 0.733298597 | 0.733298597 | 465.6563468 | 2.668065527 | TRUE | TRUE | TRUE |
| RBMS3 | 38.281686 | 0.06507256 | 0.2544 | 3.976246805 | 38.28168636 | 38.28168636 | 4.40627777 | 0.335347658 | 0.7459 | 4.406277773 | 42.48339328 | 42.48339328 | 8.687987531 | 0.938919189 | TRUE | FALSE | FALSE |
| DRD5 | 38.202688 | 0.06533085 | 0.24205 | 38.20268838 | 3562.13176 | 12353.18049 | 626.582633 | 0.003451911 | 0.63505 | 626.5826328 | 641362.4427 | 6759578.755 | 0.060969913 | -1.214884427 | TRUE | FALSE | FALSE |
| ZNF547 | 38.091806 | 0.06558927 | 0.6382 | 361.6339426 | 35025.72833 | 356932.6027 | 51.348571 | 0.053041556 | 0.9467 | 4806.664009 | 5189063.8 | 55667375.17 | 0.741827961 | -0.129696801 | TRUE | FALSE | FALSE |
| CAGE1 | 38.032277 | 0.06584753 | 0.12815 | 4.102432686 | 38.03227696 | 38.03227696 | 0.68209519 | 0.674021074 | 0.50495 | 0.682095192 | 0.682095192 | 1.52103186 | 55.75801943 | 1.746307339 | TRUE | TRUE | TRUE |
| CRELD1 | 37.858363 | 0.06610628 | 0.06545 | 1.029154211 | 37.85836284 | 37.85836284 | 130.902519 | 0.021949159 | 0.37225 | 130.902519 | 12636.62847 | 128560.7298 | 0.289210346 | -0.538786175 | TRUE | FALSE | FALSE |
| SLC25A34 | 37.699431 | 0.06636542 | 0.0677 | 1.026883927 | 37.69943097 | 37.69943097 | 1.51332244 | 0.534286154 | 0.7831 | 6.052603672 | 132.3541029 | 529.3564074 | 24.9116976 | 1.396403323 | TRUE | FALSE | FALSE |
| ADAM8 | 37.69165 | 0.06662411 | 0.9284 | 3269.349639 | 3136893.967 | 32327139.83 | 1.05698836 | 0.582715064 | 0.81785 | 9.452532696 | 86.65387679 | 813.8252221 | 35.6594749 | 1.552174944 | TRUE | FALSE | FALSE |
| ACOX2 | 37.63659 | 0.06688262 | 0.9104 | 8320.785864 | 3441969.288 | 97288750.78 | 199.839477 | 0.013554945 | 0.3716 | 199.8394771 | 19330.93013 | 196867.2273 | 0.188334112 | -0.725071011 | TRUE | FALSE | FALSE |
| P2RX5 | 37.372584 | 0.06714214 | 0.6596 | 37.37258434 | 3515.323129 | 35331.69131 | 6.5053195 | 0.25545972 | 0.25505 | 0.705953044 | 6.505319498 | 6.505319498 | 5.744926802 | 0.7592845 | TRUE | FALSE | FALSE |
| ULK4 | 37.354819 | 0.06740127 | 0.9515 | 27991.66785 | 86795348.54 | 2654297308 | 490.251688 | 0.004615459 | 0.77485 | 10397.3474 | 9574431.296 | 388501093.3 | 0.076195186 | -1.118072466 | TRUE | FALSE | FALSE |
| PLEKHO1 | 37.329356 | 0.06766004 | 0.26105 | 3.88778953 | 37.32935606 | 37.72620416 | 0.49064623 | 0.758567643 | 1 | 0.49064623 | 0.49064623 | 9.73048061 | 76.08201953 | 1.881282032 | TRUE | TRUE | TRUE |
| OR4F4 | 37.327185 | 0.06791833 | 0.1336 | 37.3271846 | 372.0349078 | 372.0349078 | 0.3146277 | 0.820403607 | 1 | 0.314627695 | 0.314627695 | 0.314627695 | 118.6392207 | 2.074228285 | TRUE | TRUE | TRUE |
| CACNA2D4 | 36.999532 | 0.06817801 | 0.9935 | 2866170.305 | 34327378449 | 3.7747E+11 | 0.01016852 | 0.896708603 | 0.99705 | 54.87817594 | 18694.74864 | 178645.2349 | 3638.636566 | 3.56093868 | TRUE | TRUE | TRUE |
| STK32B | 36.907932 | 0.06843773 | 0.1316 | 3.963210288 | 36.90793245 | 36.90793245 | 0.14280192 | 0.866182779 | 0.75355 | 1.302474269 | 12.1927201 | 12.1927201 | 258.4554446 | 2.412385685 | TRUE | TRUE | TRUE |
| MPZL3 | 36.739679 | 0.06869794 | 0.3323 | 9.185143654 | 347.5475945 | 1390.503844 | 0.375702 | 0.800562183 | 0.75445 | 0.375701997 | 1.502499857 | 1.502499857 | 97.78941521 | 1.990291849 | TRUE | TRUE | TRUE |
| CAMP | 36.708968 | 0.06895782 | 0.81385 | 359.8821984 | 3616.438553 | 37228.53178 | 43.6933663 | 0.063082177 | 0.13055 | 43.69336626 | 430.1045485 | 430.1045485 | 0.840149698 | -0.075643324 | TRUE | FALSE | FALSE |
| CPT1B | 36.578759 | 0.06921797 | 0.11985 | 0.991503674 | 146.1766473 | 1011.769243 | 0.0975494 | 0.876574824 | 0.5066 | 0.0975494 | 1.557512622 | 6.225837139 | 374.9767709 | 2.574004365 | TRUE | TRUE | TRUE |
| CAPNS2 | 36.257022 | 0.06947953 | 0.8977 | 1337.76475 | 47424.93417 | 413849.9317 | 27.8514618 | 0.099606493 | 0.71895 | 111.439107 | 1785.428131 | 7149.199868 | 1.301799618 | 0.11454414 | TRUE | FALSE | FALSE |
| IL17D | 36.112103 | 0.06974144 | 0.69175 | 36.11210285 | 3568.849525 | 3568.849525 | 0.47945822 | 0.763189814 | 0.74865 | 4.490020362 | 43.15514866 | 43.15514866 | 75.31856046 | 1.876902011 | TRUE | TRUE | TRUE |
| CNGA4 | 36.019097 | 0.07000341 | 0.02865 | 0.416139016 | 21.81429848 | 87.14354461 | 0.03987659 | 0.888982625 | 0.8259 | 0.275252633 | 10.15992344 | 21.71763659 | 903.2642951 | 2.955814843 | TRUE | TRUE | TRUE |
| ORAI3 | 35.985685 | 0.07026508 | 0.69095 | 35.98568529 | 3545.151813 | 3545.151813 | 0.49354797 | 0.757539207 | 1 | 4.61944343 | 4.61944343 | 4.61944343 | 72.91223511 | 1.862800412 | TRUE | TRUE | TRUE |
| CCDC102B | 35.836266 | 0.07052715 | 0.50545 | 35.83626593 | 339.6507923 | 3297.914923 | 473398457 | 4.88769E-09 | 0.0034 | 38259.38154 | 43435693.81 | 473398456.7 | 7.57E-08 | -7.120904087 | TRUE | FALSE | FALSE |
| GABRR2 | 35.778093 | 0.07078907 | 0.7252 | 143.2237192 | 573.4837759 | 3057.437797 | 13.2316719 | 0.168513005 | 0.8907 | 212.0069356 | 848.9466394 | 4769.491632 | 2.703973691 | 0.432002462 | TRUE | FALSE | FALSE |
| ZNF233 | 35.505196 | 0.07105214 | 0.5045 | 35.50519625 | 3091.52225 | 6480.037483 | 0.01170257 | 0.89621875 | 1 | 7.694633475 | 70.36909301 | 659.2953281 | 3033.964846 | 3.482010544 | TRUE | TRUE | TRUE |
| SLC25A10 | 35.359659 | 0.0713156 | 0.4666 | 21.00227797 | 1343.884911 | 2806.949615 | 0.09366921 | 0.877407132 | 1 | 0.093669206 | 0.093669206 | 0.093669206 | 377.4950207 | 2.576911227 | TRUE | TRUE | TRUE |
| DLX6 | 35.164421 | 0.07157975 | 0.49835 | 3.627162161 | 349.6540955 | 349.6540955 | 0.48142099 | 0.762520709 | 1 | 0.481420991 | 0.481420991 | 0.481420991 | 73.04297494 | 1.863578453 | TRUE | TRUE | TRUE |
| ZNF224 | 35.050753 | 0.07184409 | 0.1949 | 3.819277891 | 35.05075259 | 329.5500507 | 0.0118976 | 0.896113219 | 0.01685 | 0.011897598 | 0.011897598 | 0.235244817 | 2946.036155 | 3.469238072 | TRUE | TRUE | TRUE |
| KLHDC1 | 34.902772 | 0.07210885 | 0.1284 | 1.594762831 | 139.5659672 | 139.5659672 | 0.25449974 | 0.838024263 | 0.9398 | 5.355761581 | 204.1526905 | 204.1526905 | 137.1426652 | 2.137172585 | TRUE | TRUE | TRUE |
| SLC7A9 | 34.841912 | 0.07237348 | 0.24335 | 4.775657788 | 305.8448455 | 2590.37285 | 0.01908329 | 0.894077175 | 0.9685 | 0.305014674 | 1.219880482 | 4.880027976 | 1825.781465 | 3.261448794 | TRUE | TRUE | TRUE |
| NDC1 | 34.779621 | 0.072638 | 0.50765 | 34.77962113 | 342.2176482 | 342.2176482 | 0.47620272 | 0.764585711 | 1 | 0.476202717 | 0.476202717 | 9.432806424 | 73.03532694 | 1.863532978 | TRUE | TRUE | TRUE |
| TINAG | 34.60695 | 0.07290309 | 0.0671 | 1.203359406 | 73.16984415 | 171.3946548 | 0.50620803 | 0.753332717 | 0.07455 | 0.126565153 | 2.025127711 | 2.025127711 | 68.36507607 | 1.834834301 | TRUE | TRUE | TRUE |
| SPDYE3 | 34.566672 | 0.07316794 | 0.1444 | 34.56667225 | 340.5514655 | 340.5514655 | 0.44405666 | 0.776042882 | 1 | 4.166897801 | 40.13036778 | 82.37298012 | 77.84293166 | 1.891219184 | TRUE | TRUE | TRUE |
| CORO1C | 34.316354 | 0.07343385 | 0.13275 | 3.545330679 | 340.6780557 | 340.6780557 | 1.7784569 | 0.510278207 | 0.75485 | 4.204923769 | 16.8175791 | 16.8175791 | 19.29557807 | 1.285457794 | TRUE | FALSE | FALSE |
| OR10H4 | 34.302898 | 0.07369935 | 0.0037 | 3.537753475 | 34.30289813 | 34.30289813 | 0.05148458 | 0.886312018 | 1 | 0.051484584 | 0.051484584 | 0.051484584 | 666.2751286 | 2.823653602 | TRUE | TRUE | TRUE |
| KRT33B | 34.298679 | 0.07396437 | 0.6039 | 137.2670239 | 8812.168633 | 141460.5547 | 621.717833 | 0.00350182 | 0.40225 | 343.1491358 | 39900.40143 | 286032.1474 | 0.055167598 | -1.258315925 | TRUE | FALSE | FALSE |
| GTPBP2 | 34.014337 | 0.07423069 | 0.00695 | 1.005583776 | 4.023016754 | 19.82266551 | 0.30251145 | 0.824615757 | 1 | 0.302511453 | 0.302511453 | 5.973599157 | 112.4398318 | 2.050920187 | TRUE | TRUE | TRUE |
| PLD1 | 33.854947 | 0.07449753 | 0.50385 | 33.85494737 | 2171.795724 | 19221.08134 | 71.9248675 | 0.040203096 | 0.41335 | 32.9180262 | 2582.195346 | 10337.45551 | 0.47069878 | -0.327256928 | TRUE | FALSE | FALSE |
| C9orf152 | 33.819778 | 0.07476408 | 0.12615 | 33.81977796 | 330.1691783 | 330.1691783 | 38.0283282 | 0.071520986 | 0.4974 | 3.984316399 | 372.2753289 | 372.2753289 | 0.88933118 | -0.050936481 | TRUE | FALSE | FALSE |
| CX3CR1 | 33.643094 | 0.07503127 | 0.25325 | 3.509242118 | 33.6430938 | 33.6430938 | 0.43488821 | 0.77936871 | 1 | 4.073863044 | 4.073863044 | 4.073863044 | 77.36032694 | 1.888518296 | TRUE | TRUE | TRUE |
| SECTM1 | 33.619873 | 0.07529811 | 0.68925 | 33.61987324 | 3291.085677 | 3291.085677 | 335.120457 | 0.007414367 | 0.77425 | 3356.678987 | 361829.6396 | 361829.6396 | 0.100321757 | -0.998604869 | TRUE | FALSE | FALSE |
| SLC25A33 | 33.521049 | 0.07556508 | 0.1938 | 8.377528562 | 33.52104905 | 134.1614568 | 0.16876302 | 0.859942938 | 0.75255 | 1.579066881 | 6.31400228 | 6.31400228 | 198.6279308 | 2.298040318 | TRUE | TRUE | TRUE |
| PCDHGA2 | 33.301317 | 0.07583297 | 0.19565 | 3.725448169 | 133.2749826 | 533.5121539 | 0.40584378 | 0.789999406 | 0.14705 | 0.405843779 | 1.623522687 | 7.939761609 | 82.0545203 | 1.914102511 | TRUE | TRUE | TRUE |
| PPP2R2C | 33.169047 | 0.07610122 | 0.68835 | 33.16904678 | 3330.115344 | 3330.115344 | 3.98600315 | 0.356840775 | 0.7539 | 3.986003148 | 38.57857145 | 78.86479147 | 8.321379976 | 0.920195353 | TRUE | FALSE | FALSE |
| C8A | 33.129203 | 0.07636923 | 0.9772 | 45061.39351 | 46592354.31 | 749680409.8 | 62.4499439 | 0.045008876 | 0.59085 | 62.44994391 | 16128.30027 | 79605.24883 | 0.53049212 | -0.275321063 | TRUE | FALSE | FALSE |
| GLRB | 33.067802 | 0.07663714 | 0.32175 | 33.06780247 | 810.0606534 | 3238.852272 | 0.14158681 | 0.86643972 | 1 | 0.141586814 | 0.141586814 | 2.81091757 | 233.5514267 | 2.368382525 | TRUE | TRUE | TRUE |
| ASTL | 33.054361 | 0.07690464 | 0.5046 | 33.05436067 | 776.4826595 | 3105.397388 | 1.21761904 | 0.56560845 | 0.8769 | 11.43153039 | 110.0796282 | 110.0796282 | 27.14671796 | 1.433717331 | TRUE | FALSE | FALSE |
| PHC1 | 32.992114 | 0.07717205 | 0.2654 | 3.404126482 | 32.99211389 | 114.8327975 | 0.43355163 | 0.77985272 | 0.0254 | 0.433551629 | 0.433551629 | 8.521706776 | 76.09731273 | 1.881369321 | TRUE | TRUE | TRUE |
| OR5AU1 | 32.814092 | 0.07744013 | 0.0057 | 3.429543223 | 32.81409249 | 32.81409249 | 3.51044418 | 0.382496109 | 0.97015 | 329.2548313 | 3310.16781 | 34091.08201 | 9.347561384 | 0.970698326 | TRUE | FALSE | FALSE |
| ZNF446 | 32.78452 | 0.0777079 | 0.0069 | 3.541335935 | 32.78452045 | 32.78452045 | 0.59951895 | 0.712653651 | 0.5104 | 0.59951895 | 0.59951895 | 1.336226789 | 54.68471086 | 1.73786592 | TRUE | TRUE | TRUE |
| HAO1 | 32.703061 | 0.07797572 | 0.1883 | 3.489465468 | 130.8126916 | 130.8126916 | 0.13252018 | 0.868311034 | 1 | 1.216619511 | 1.216619511 | 1.216619511 | 246.777979 | 2.392306403 | TRUE | TRUE | TRUE |
| ABCF3 | 32.669475 | 0.07824326 | 0.22875 | 8.169841211 | 522.786466 | 2565.207378 | 1.28796423 | 0.556371862 | 0.6322 | 2.990760881 | 191.0978348 | 764.358844 | 25.36520371 | 1.404238354 | TRUE | FALSE | FALSE |
| TPT1 | 32.63902 | 0.0785105 | 0.0033 | 3.430203853 | 32.63901979 | 32.63901979 | 35.6623374 | 0.078492132 | 0.5082 | 35.66233735 | 348.6980346 | 348.6980346 | 0.915223797 | -0.038472696 | TRUE | FALSE | FALSE |
| FNDC4 | 32.627512 | 0.07877733 | 0.5022 | 321.0778348 | 3238.676822 | 33465.62333 | 0.46438473 | 0.76933864 | 1 | 4.365087329 | 4.365087329 | 4.365087329 | 70.25965641 | 1.846706022 | TRUE | TRUE | TRUE |
| GRAMD1B | 32.597863 | 0.07904387 | 0.0181 | 3.381742103 | 32.59786252 | 66.46306426 | 0.41928013 | 0.784747957 | 1 | 3.946688154 | 3.946688154 | 77.72802254 | 77.747215 | 1.890684841 | TRUE | TRUE | TRUE |
| SCARF1 | 32.450614 | 0.07931088 | 0.0778 | 3.460993431 | 311.8856173 | 311.8856173 | 442.046137 | 0.005245597 | 0.191 | 45.70162262 | 1767.94999 | 17518.32572 | 0.073410017 | -1.134244678 | TRUE | FALSE | FALSE |
| UCP3 | 32.299644 | 0.0795784 | 0.40545 | 14.13948954 | 1210.605874 | 4843.070116 | 0.05022204 | 0.886543843 | 0.688 | 0.200786443 | 3.211737127 | 3.211737127 | 643.1368059 | 2.808303364 | TRUE | TRUE | TRUE |

| | | | | | | | | | | | | | | | | | |
|---|---|---|---|---|---|---|---|---|---|---|---|---|---|---|---|---|---|
| EGR4 | 32.11684 | 0.07984664 | 0.6921 | 32.11683952 | 3180.546835 | 3180.546835 | 3370.17966 | 0.00057258 | 0.1159 | 334.4587087 | 34788.50233 | 368714.0298 | 0.009529711 | -2.020920252 | TRUE | FALSE | FALSE |
| AURKC | 31.94891 | 0.08011552 | 0.50415 | 31.94890987 | 312.7659254 | 312.7659254 | 355.09202 | 0.00670967 | 0.1292 | 36.18444112 | 355.0920196 | 355.0920196 | 0.089973607 | -1.045884868 | TRUE | FALSE | FALSE |
| FERMT3 | 31.922788 | 0.08038408 | 0.1423 | 7.984096055 | 313.761874 | 313.761874 | 0.52140769 | 0.746689048 | 0.5101 | 0.521407688 | 0.521407688 | 10.260514 | 61.22423767 | 1.786923386 | TRUE | TRUE | TRUE |
| KRTAP10-3 | 31.728399 | 0.08065346 | 0.00445 | 3.336361898 | 31.72839947 | 31.72839947 | 3.42489378 | 0.387362436 | 0.96885 | 317.7830813 | 3177.667539 | 32550.75642 | 9.264053586 | 0.966801058 | TRUE | FALSE | FALSE |
| DEFB126 | 31.529182 | 0.08092369 | 0.00285 | 3.318455855 | 31.52918191 | 31.52918191 | 3.63246637 | 0.375905819 | 0.87685 | 34.58355962 | 337.7087659 | 337.7087659 | 8.679827603 | 0.938511099 | TRUE | FALSE | FALSE |
| ZNF782 | 31.51482 | 0.08119353 | 0.50185 | 31.51482033 | 300.5159231 | 2935.721421 | 49.4891622 | 0.05555442 | 0.0072 | 1.275866458 | 49.48916222 | 49.48916222 | 0.636802462 | -0.195995266 | TRUE | FALSE | FALSE |
| REG3G | 31.46007 | 0.08146325 | 0.5051 | 31.46006964 | 306.4906285 | 306.4906285 | 3.86538408 | 0.363309294 | 0.25305 | 3.865384081 | 36.85793676 | 36.85793676 | 8.138924615 | 0.910567026 | TRUE | FALSE | FALSE |
| ACOT1 | 31.192237 | 0.08173431 | 0.25665 | 3.252932281 | 31.19223653 | 31.19223653 | 0.42249482 | 0.783647248 | 1 | 0.422494823 | 0.422494823 | 8.365699959 | 73.82868335 | 1.868225123 | TRUE | TRUE | TRUE |
| B3GALNT1 | 31.170481 | 0.08200502 | 0.8763 | 722.4073957 | 115414.8182 | 461823.1163 | 0.15201917 | 0.86410485 | 1 | 0.152019172 | 0.152019172 | 0.152019172 | 205.0430923 | 2.311845143 | TRUE | TRUE | TRUE |
| FAM149A | 31.134801 | 0.08227549 | 0.20175 | 31.1348008 | 294.8738231 | 2861.037239 | 4283.2994 | 0.000436972 | 0.0026 | 4.913115706 | 437.5836848 | 4283.299399 | 0.007268883 | -2.138532341 | TRUE | FALSE | FALSE |
| GLP1R | 31.106347 | 0.08254566 | 0.26205 | 3.241564002 | 31.10634708 | 64.16092831 | 0.4181384 | 0.785322621 | 1 | 0.418138396 | 0.418138396 | 8.286591706 | 74.39246763 | 1.871528965 | TRUE | TRUE | TRUE |
| GALM | 31.047404 | 0.08281575 | 0.1244 | 3.319047456 | 31.04740394 | 31.04740394 | 4.22541919 | 0.342225334 | 0.77585 | 16.90211017 | 382.1360794 | 1528.583513 | 7.347768948 | 0.866155491 | TRUE | FALSE | FALSE |
| IFNA21 | 30.969375 | 0.08308588 | 0.49835 | 3.262898246 | 301.4868381 | 301.4868381 | 0.42354919 | 0.783268013 | 1 | 3.93600617 | 3.93600617 | 3.93600617 | 73.11872053 | 1.864028583 | TRUE | TRUE | TRUE |
| WDR5B | 30.932553 | 0.08335578 | 0.6537 | 30.93255267 | 2907.993616 | 29219.68414 | 42.8503698 | 0.064145316 | 0.85615 | 412.8973189 | 41194.04855 | 426047.5952 | 0.721873645 | -0.141538813 | TRUE | FALSE | FALSE |
| KRT12 | 30.90265 | 0.08362539 | 0.265 | 7.721896717 | 123.7017586 | 495.2955514 | 55.9449202 | 0.050114606 | 0.2303 | 13.96241469 | 224.2172925 | 3604.216274 | 0.552376329 | -0.257764941 | TRUE | FALSE | FALSE |
| CETN1 | 30.616519 | 0.08389651 | 0.3235 | 30.61651863 | 3054.210928 | 3054.210928 | 37.2746888 | 0.074024276 | 0.003 | 3.862267643 | 37.2746888 | 37.2746888 | 0.821375567 | -0.08545822 | TRUE | FALSE | FALSE |
| OR2M3 | 30.603807 | 0.08416722 | 0.2585 | 3.211591532 | 30.60380718 | 30.60380718 | 0.41672885 | 0.785865588 | 0.00755 | 0.416728852 | 0.416728852 | 0.416728852 | 73.4381769 | 1.865921887 | TRUE | TRUE | TRUE |
| MTR | 30.563355 | 0.08443773 | 0.9592 | 40764.50448 | 24797180.73 | 378908349.7 | 2381.91628 | 0.000996928 | 0.904 | 515449.3068 | 844914798.3 | 10805953055 | 0.012831414 | -1.891725467 | TRUE | FALSE | FALSE |
| G3BP2 | 30.477985 | 0.08470835 | 0.2563 | 3.139807782 | 30.47798478 | 62.1521758 | 0.39058206 | 0.795460113 | 1 | 3.699411965 | 3.699411965 | 7.741069882 | 78.03221879 | 1.892273956 | TRUE | TRUE | TRUE |
| SSBP4 | 30.455268 | 0.08497863 | 0.50515 | 30.45526806 | 299.7140544 | 299.7140544 | 3.38377491 | 0.389882502 | 0.9411 | 32.49734495 | 3232.012272 | 3232.012272 | 9.000382377 | 0.954260961 | TRUE | FALSE | FALSE |
| OR5H1 | 30.264777 | 0.08524977 | 0.8154 | 295.4926239 | 2957.263993 | 30318.61651 | 349.221372 | 0.006864437 | 0.31685 | 35.6380258 | 3507.677978 | 3507.677978 | 0.086663588 | -1.062163335 | TRUE | FALSE | FALSE |
| IRAK4 | 30.250519 | 0.08552052 | 0.5046 | 30.25051919 | 296.2055658 | 2971.243736 | 0.06025291 | 0.884699588 | 0.75235 | 0.551054026 | 5.172561861 | 5.172561861 | 502.0590638 | 2.700754812 | TRUE | TRUE | TRUE |
| IREB2 | 30.145195 | 0.08579153 | 0.00405 | 0.893782068 | 0.893782068 | 17.57275174 | 0.26982468 | 0.833826885 | 1 | 0.269824683 | 0.269824683 | 5.315875881 | 111.7214131 | 2.04813642 | TRUE | TRUE | TRUE |
| BTN2A1 | 30.115757 | 0.08606226 | 0.34225 | 3.351309918 | 277.2601313 | 2613.660505 | 0.74723744 | 0.64273145 | 0.6959 | 0.747237437 | 60.78576755 | 60.78576755 | 40.30279452 | 1.60533516 | TRUE | TRUE | TRUE |
| KHK | 29.620728 | 0.08633604 | 0.5795 | 29.62072772 | 1104.106309 | 4419.108934 | 4.85365119 | 0.314212081 | 0.7762 | 19.42237631 | 311.2399804 | 659.0949979 | 6.102772234 | 0.785527162 | TRUE | FALSE | FALSE |
| OR1F1 | 29.60311 | 0.08660945 | 0.5008 | 61.84564362 | 2893.180386 | 29664.52976 | 3.77643029 | 0.367718158 | 0.49745 | 3.776430286 | 354.2493465 | 354.2493465 | 7.838913418 | 0.894255868 | TRUE | FALSE | FALSE |
| SERPINB5 | 29.495631 | 0.08688316 | 0.23645 | 29.49563108 | 472.6302358 | 1892.631471 | 0.60237206 | 0.711115329 | 0.75735 | 5.601021293 | 89.62562223 | 89.62562223 | 48.96580245 | 1.689892876 | TRUE | TRUE | TRUE |
| FADS1 | 29.484243 | 0.08715645 | 0.13185 | 6.580114457 | 288.7741912 | 288.7741912 | 26777.2894 | 6.54305E-05 | 0.6201 | 26777.28943 | 34260950.49 | 384214677.7 | 0.001101091 | -2.958176626 | TRUE | FALSE | FALSE |
| HMOX2 | 29.43876 | 0.08742959 | 0.5041 | 29.43875981 | 287.6011769 | 287.6011769 | 0.41569685 | 0.786235461 | 1 | 0.415696853 | 0.415696853 | 0.415696853 | 70.81785578 | 1.850142773 | TRUE | TRUE | TRUE |
| ITGB1 | 29.279151 | 0.08770339 | 0.00495 | 0.157210262 | 1.485236028 | 3.099159301 | 0.26309165 | 0.835794484 | 0.50795 | 0.263091649 | 0.263091649 | 0.962532794 | 111.2887906 | 2.046451423 | TRUE | TRUE | TRUE |
| PRRG2 | 29.259524 | 0.08797685 | 0.5047 | 29.2595245 | 468.3662441 | 1874.594945 | 18.6112165 | 0.140394768 | 0.5041 | 18.61121646 | 298.1097521 | 371.15981 | 1.572144656 | 0.196492504 | TRUE | FALSE | FALSE |
| SDR42E1 | 29.203031 | 0.08825023 | 0.06715 | 1.824387086 | 29.20303058 | 61.85622747 | 0.01262545 | 0.89585912 | 1 | 1.820065459 | 29.11139765 | 116.46993 | 2313.028324 | 3.364180951 | TRUE | TRUE | TRUE |
| EXOSC5 | 29.199469 | 0.08852316 | 0.24895 | 3.042808254 | 29.199469 | 29.199469 | 0.38320819 | 0.798069482 | 1 | 3.592806804 | 3.592806804 | 3.592806804 | 76.19740249 | 1.881940167 | TRUE | TRUE | TRUE |
| NT5C1B | 29.161497 | 0.08879587 | 0.32185 | 7.28716138 | 116.742348 | 143.4396924 | 2.04831513 | 0.485054496 | 0.75465 | 2.048315128 | 8.194946746 | 40.39552084 | 14.23682148 | 1.153413039 | TRUE | FALSE | FALSE |
| PGM2 | 29.077399 | 0.08906871 | 0.1551 | 7.267361338 | 116.3704818 | 465.8419165 | 0.36680194 | 0.803472014 | 0.5088 | 0.366801943 | 1.467346666 | 1.817338835 | 79.27275023 | 1.899123926 | TRUE | TRUE | TRUE |
| SGCA | 29.063378 | 0.08934117 | 0.6221 | 116.2905881 | 7460.886485 | 29882.87975 | 110.091848 | 0.026319437 | 0.03555 | 2.936636028 | 27.52409491 | 110.0918481 | 0.263992101 | -0.578409067 | TRUE | FALSE | FALSE |
| NREP | 28.982001 | 0.08961374 | 0.93735 | 2796.468896 | 297178.6301 | 297178.6301 | 36.5677862 | 0.075973993 | 0.6891 | 36.56778622 | 3571.955921 | 3571.955921 | 0.79255553 | -0.100970299 | TRUE | FALSE | FALSE |
| SLC27A6 | 28.936144 | 0.08988616 | 0.83025 | 273.6191187 | 266006.8339 | 2755935.559 | 5.06854009 | 0.304493564 | 0.9892 | 446199.8078 | 540900406.9 | 2162707227 | 5.708970173 | 0.756557774 | TRUE | FALSE | FALSE |
| ZNF492 | 28.914048 | 0.09015826 | 0.25415 | 3.035627276 | 28.91404768 | 60.20209801 | 0.39673213 | 0.793045902 | 1 | 0.396732127 | 0.396732127 | 7.875886803 | 72.88053 | 1.862611522 | TRUE | TRUE | TRUE |
| CR2 | 28.723179 | 0.09043128 | 0.50725 | 275.3825391 | 27156.54233 | 279028.9139 | 10.9996422 | 0.18809182 | 0.5855 | 25.43162615 | 2340.381942 | 23266.80359 | 2.611283062 | 0.416853952 | TRUE | FALSE | FALSE |
| RPN2 | 28.53276 | 0.09070523 | 0.26135 | 2.985722405 | 28.53276027 | 58.95903931 | 0.39008812 | 0.79558965 | 1 | 0.390088117 | 0.390088117 | 7.714792781 | 73.14439751 | 1.864181067 | TRUE | TRUE | TRUE |
| SLC25A26 | 28.445667 | 0.09097935 | 0.12865 | 3.17815953 | 65.31323226 | 261.313736 | 0.3050953 | 0.823854478 | 0.49855 | 0.076291235 | 0.3050953 | 0.3050953 | 93.23534945 | 1.969580603 | TRUE | TRUE | TRUE |
| UTS2 | 28.340383 | 0.09125377 | 0.5035 | 28.34038258 | 275.0129156 | 275.0129156 | 3.64888586 | 0.374236673 | 0.5035 | 3.648885857 | 3.648885857 | 3.648885857 | 7.76685917 | 0.890245431 | TRUE | FALSE | FALSE |
| DACH1 | 28.331973 | 0.09152777 | 0.2544 | 2.920984657 | 28.33197257 | 57.67616596 | 0.37861377 | 0.799796219 | 1 | 0.378613772 | 0.378613772 | 7.487349584 | 74.830803 | 1.874080406 | TRUE | TRUE | TRUE |
| TFIP11 | 28.292361 | 0.09180158 | 0.0132 | 3.043709255 | 28.29236135 | 59.72827084 | 0.05980058 | 0.884767574 | 1 | 0.239189877 | 0.239189877 | 4.704385481 | 473.1118391 | 2.674963816 | TRUE | TRUE | TRUE |
| OR1Q1 | 28.209534 | 0.09207553 | 0.5073 | 28.20953366 | 274.8815515 | 274.8815515 | 3.62473416 | 0.376618276 | 0.5069 | 3.624734158 | 3.624734158 | 3.624734158 | 7.782511055 | 0.891119746 | TRUE | FALSE | FALSE |
| RAB25 | 27.917509 | 0.09235121 | 0.73355 | 67.52161156 | 10703.3872 | 27176.70392 | 328.253818 | 0.007495084 | 0.8435 | 8282.273948 | 1378688.341 | 14684401.99 | 0.085048543 | -1.070333123 | TRUE | FALSE | FALSE |
| GDF15 | 27.761317 | 0.09262759 | 0.38305 | 12.07904614 | 111.0922069 | 444.6677494 | 524.342447 | 0.00431627 | 0.2879 | 130.9977593 | 2067.940043 | 8281.447357 | 0.052945011 | -1.276174955 | TRUE | FALSE | FALSE |
| ZNF493 | 27.688144 | 0.09290404 | 0.0355 | 3.034408278 | 27.68814443 | 258.8352393 | 53.4903895 | 0.051568596 | 0.1928 | 5.787504965 | 53.49038945 | 506.4785486 | 0.517628395 | -0.285981908 | TRUE | FALSE | FALSE |
| PAFAH2 | 27.588406 | 0.09318077 | 0.83775 | 656.6867408 | 53356.15682 | 499774.2464 | 44461.2995 | 4.34853E-05 | 0.20495 | 5728.195542 | 714158.5987 | 6575651.607 | 0.000620504 | -3.207255541 | TRUE | FALSE | FALSE |
| C1orf74 | 27.534271 | 0.09345744 | 0.7772 | 110.1659424 | 4128.285366 | 5129.785003 | 8.20209034 | 0.22456934 | 0.00645 | 2.050032207 | 8.202090336 | 8.202090336 | 3.356982205 | 0.525949039 | TRUE | FALSE | FALSE |
| ATOH7 | 27.315185 | 0.09373531 | 0.5015 | 27.31518454 | 271.9469143 | 271.9469143 | 1.47704841 | 0.537738144 | 0.752 | 1.477048407 | 5.908011758 | 5.908011758 | 18.4930869 | 1.26700941 | TRUE | FALSE | FALSE |
| GIPC3 | 27.245855 | 0.09401324 | 0.127 | 7.610223627 | 73.98412209 | 73.98412209 | 0.12324468 | 0.870513933 | 0.5041 | 1.14390963 | 1.14390963 | 1.14390963 | 221.0712489 | 2.344532265 | TRUE | TRUE | TRUE |
| ACOT6 | 27.21891 | 0.09429088 | 0.658 | 265.2264148 | 27105.39449 | 283951.3288 | 3.62637863 | 0.376380996 | 0.8773 | 34.64760726 | 339.528775 | 339.528775 | 7.505810356 | 0.875397587 | TRUE | FALSE | FALSE |
| LHCGR | 27.131245 | 0.09456873 | 0.0945 | 1.696765652 | 27.13124499 | 108.5316453 | 5.1623306 | 0.29925627 | 0.04425 | 1.290683076 | 5.162330599 | 20.65287383 | 5.255619429 | 0.720623909 | TRUE | FALSE | FALSE |
| FPGT | 27.113271 | 0.09484623 | 0.32005 | 6.778476791 | 255.2234512 | 255.2234512 | 0.12170949 | 0.870881352 | 1 | 0.121709491 | 0.121709491 | 2.413002674 | 222.7703893 | 2.347857464 | TRUE | TRUE | TRUE |
| RGL3 | 27.035101 | 0.09512386 | 0.12225 | 3.011832559 | 248.6222498 | 2341.10264 | 48.7723008 | 0.05580987 | 0.3685 | 48.77230079 | 4333.436062 | 42297.61225 | 0.554312609 | -0.256245242 | TRUE | FALSE | FALSE |
| NDUFA4L2 | 26.910127 | 0.09540198 | 0.002 | 2.809754744 | 26.91012723 | 26.91012723 | 30.1037515 | 0.091653768 | 0.50465 | 30.10375146 | 296.0504722 | 296.0504722 | 0.893912749 | -0.048704869 | TRUE | FALSE | FALSE |
| GNLY | 26.896959 | 0.09567972 | 0.0677 | 2.819909757 | 262.9752702 | 262.9752702 | 2504.36863 | 0.000935042 | 0.61425 | 25638.53231 | 31521292.53 | 385627464 | 0.010740016 | -1.968995076 | TRUE | FALSE | FALSE |
| CSHL1 | 26.893858 | 0.095957 | 0.0661 | 2.833875605 | 261.6201342 | 261.6201342 | 3.98832901 | 0.356589706 | 0.89075 | 38.00735015 | 3715.246701 | 38065.04514 | 6.743139388 | 0.828862137 | TRUE | FALSE | FALSE |
| APOBEC3H | 26.869077 | 0.09623399 | 0.8903 | 262.0013712 | 26813.35199 | 281088.4944 | 274.477297 | 0.009147253 | 0.74875 | 28100.47174 | 3302583.516 | 36220710.63 | 0.097891801 | -1.009253681 | TRUE | FALSE | FALSE |
| LRIG1 | 26.798489 | 0.09651106 | 0.6376 | 26.7984888 | 2554.227543 | 25821.32101 | 6.10188894 | 0.266321659 | 0.51165 | 6.101888936 | 58.16054164 | 567.9159285 | 4.391834903 | 0.642646006 | TRUE | FALSE | FALSE |
| CCDC73 | 26.684787 | 0.09678854 | 0.8325 | 243.8855624 | 214114.1916 | 2144274.843 | 0.78482062 | 0.626127094 | 0.5057 | 0.784820616 | 6.974139602 | 6.974139602 | 34.00112891 | 1.531493337 | TRUE | TRUE | TRUE |
| GOLGA6B | 26.663091 | 0.09706572 | 0.8178 | 261.3246053 | 2625.330864 | 27018.62638 | 29.706859 | 0.093405383 | 0.89215 | 291.5203352 | 30215.61081 | 316500.2794 | 0.897539899 | -0.046946236 | TRUE | FALSE | FALSE |
| C5AR1 | 26.546559 | 0.09734333 | 0.00875 | 6.63670372 | 26.54655866 | 26.54655866 | 6.77785013 | 0.248976324 | 0.2607 | 6.777850129 | 65.92456807 | 65.92456807 | 3.916663566 | 0.592916268 | TRUE | FALSE | FALSE |
| SDHA | 26.427582 | 0.09762141 | 0.8436 | 619.9521105 | 158574.3337 | 1592728.241 | 0.81746896 | 0.615107143 | 0.9384 | 77.43752949 | 12486.80697 | 49925.14498 | 32.32854479 | 1.509586156 | TRUE | TRUE | TRUE |
| OR2V2 | 26.224735 | 0.09790063 | 0.66245 | 256.0772074 | 26280.56416 | 275888.6871 | 0.36419644 | 0.80435171 | 1 | 32.45521091 | 3200.80114 | 3200.80114 | 72.00711652 | 1.85737542 | TRUE | TRUE | TRUE |
| HLA-DPB1 | 26.078134 | 0.09818056 | 0.2549 | 2.752938543 | 26.07813405 | 26.07813405 | 0.36531821 | 0.803972298 | 1 | 0.365318208 | 0.365318208 | 0.365318208 | 71.38470929 | 1.853605195 | TRUE | TRUE | TRUE |
| GINS1 | 25.77718 | 0.09846244 | 0.1876 | 6.441382111 | 103.1598964 | 413.0324865 | 0.45800139 | 0.771868304 | 1 | 7.329344772 | 29.33169715 | 29.33169715 | 56.2818804 | 1.750368599 | TRUE | TRUE | TRUE |
| PADI2 | 25.772677 | 0.09874388 | 0.8599 | 244.4454949 | 23630.37955 | 240577.7885 | 131.669544 | 0.021653206 | 1 | 5.57976E+17 | 2.75999E+23 | 5.88183E+25 | 0.195737573 | -0.708325801 | TRUE | FALSE | FALSE |
| MFN1 | 25.734279 | 0.09902514 | 0.59775 | 25.73427935 | 1649.752856 | 26458.277 | 0.06598477 | 0.883350402 | 0.98175 | 4.216916201 | 270.1031101 | 1081.24288 | 390.0032909 | 2.591068272 | TRUE | TRUE | TRUE |
| IL36B | 25.646851 | 0.09930664 | 0.12715 | 2.717048604 | 248.301864 | 248.301864 | 289.174311 | 0.008694079 | 0.12905 | 3.141051861 | 289.1743113 | 289.1743113 | 0.088689935 | -1.052125664 | TRUE | FALSE | FALSE |
| PYROXD2 | 25.627359 | 0.09958781 | 0.0716 | 2.71848007 | 247.645164 | 247.645164 | 68.2068977 | 0.041474022 | 0.0675 | 1.839827806 | 68.2068977 | 68.2068977 | 0.375729726 | -0.425124444 | TRUE | FALSE | FALSE |

| | | | | | | | | | | | | | | | | | |
|---|---|---|---|---|---|---|---|---|---|---|---|---|---|---|---|---|---|
| CORO1B | 25.522856 | 0.09986936 | 0.66245 | 61.52014709 | 2430.49443 | 24599.61014 | 312.536495 | 0.007743901 | 0.69965 | 1250.110522 | 326189.0161 | 1304719.059 | 0.081663601 | -1.087971474 | TRUE | FALSE | FALSE |
| PDYN | 25.507307 | 0.10015056 | 0.2681 | 3.509127999 | 102.0640613 | 408.4976542 | 0.2674673 | 0.834471065 | 0.8739 | 1.06975919 | 4.279666135 | 4.279666135 | 95.36607497 | 1.979393909 | FALSE | TRUE | FALSE |
| IGF2 | 25.421528 | 0.10043197 | 0.69 | 25.42152781 | 2555.384987 | 2555.384987 | 30.6264052 | 0.090779651 | 0.12415 | 3.165579591 | 303.9045827 | 303.9045827 | 0.830052617 | -0.080894377 | FALSE | FALSE | FALSE |
| RAP1GAP | 25.414878 | 0.10071296 | 0.15015 | 1.591261875 | 101.6070817 | 406.3202028 | 0.00943075 | 0.896917249 | 0.5161 | 0.087194687 | 0.087194687 | 0.187122793 | 2694.895808 | 3.430541979 | FALSE | TRUE | FALSE |
| ENPP2 | 25.397245 | 0.10099362 | 0.00755 | 0.139618112 | 1.293304412 | 4.71329727 | 0.02675802 | 0.89199739 | 1 | 0.026758016 | 0.026758016 | 0.526606924 | 949.1452832 | 2.977332694 | FALSE | TRUE | FALSE |
| ZNF2 | 25.36764 | 0.10127403 | 0.5073 | 25.36763984 | 248.0366231 | 248.0366231 | 2.89815225 | 0.420024352 | 0.9679 | 184.3945602 | 2713.686063 | 27882.75006 | 8.753039062 | 0.942158867 | FALSE | FALSE | FALSE |
| HAS1 | 25.243092 | 0.101555 | 0.1975 | 25.24309177 | 240.2554053 | 2342.605628 | 0.05322961 | 0.886029402 | 0.8752 | 4.335957966 | 40.66509796 | 40.66509796 | 474.2302269 | 2.675989232 | FALSE | TRUE | FALSE |
| ALDH1B1 | 25.235118 | 0.10183555 | 0.5024 | 25.23511831 | 3965.746392 | 15854.85035 | 0.05483567 | 0.885670214 | 0.9988 | 450.4185028 | 498919.2791 | 5398328.362 | 460.1952847 | 2.662942164 | FALSE | TRUE | FALSE |
| OR2M2 | 25.071387 | 0.10211699 | 0.50675 | 243.3078331 | 2420.307763 | 24663.99466 | 3.07317595 | 0.40811588 | 0.968 | 284.1215149 | 2835.954729 | 28998.13412 | 8.158135979 | 0.91159094 | FALSE | FALSE | FALSE |
| OR10W1 | 25.042249 | 0.1023982 | 0.50485 | 25.04224875 | 242.794711 | 242.794711 | 3.07250637 | 0.408332165 | 0.877 | 29.14624001 | 283.5816808 | 283.5816808 | 8.150430209 | 0.911180533 | FALSE | FALSE | FALSE |
| NFE2L2 | 25.026611 | 0.10267905 | 0.26045 | 2.620100349 | 25.0266105 | 51.87325817 | 0.34775657 | 0.809779078 | 1 | 0.347756571 | 0.347756571 | 6.893309156 | 71.9658881 | 1.857126689 | FALSE | TRUE | FALSE |
| NUDT1 | 24.98492 | 0.10295977 | 0.25425 | 2.635916503 | 24.98492039 | 24.98492039 | 3.15930388 | 0.403098862 | 0.50195 | 3.15930388 | 3.15930388 | 3.15930388 | 7.908362516 | 0.898086569 | FALSE | FALSE | FALSE |
| BDNF | 24.938129 | 0.10324041 | 0.4944 | 2.623878164 | 243.1031023 | 243.1031023 | 30.0364353 | 0.09252956 | 0.2544 | 3.148270849 | 30.03643535 | 30.03643535 | 0.830262591 | -0.08078453 | FALSE | FALSE | FALSE |
| IDH2 | 24.909983 | 0.1035208 | 0.20005 | 6.226063626 | 24.90998279 | 99.6877468 | 7.93165829 | 0.229553911 | 0.6421 | 31.73552288 | 508.433777 | 2035.829551 | 3.140576899 | 0.497009432 | FALSE | FALSE | FALSE |
| IL17RB | 24.872169 | 0.10380103 | 0.82705 | 245.7966793 | 272708.4585 | 2952359.276 | 0.05083305 | 0.886428039 | 0.25465 | 0.050833054 | 0.472902766 | 0.472902766 | 489.2912539 | 2.689567453 | FALSE | TRUE | FALSE |
| ELFN2 | 24.851579 | 0.10408096 | 0.50765 | 24.85157949 | 245.5229662 | 484.7947258 | 0.35194934 | 0.808496636 | 0.02705 | 0.351949339 | 0.351949339 | 6.886056258 | 70.61124077 | 1.848873843 | FALSE | TRUE | FALSE |
| ZNF611 | 24.822265 | 0.10436067 | 0.1294 | 0.661021331 | 24.82226463 | 24.82226463 | 0.05401176 | 0.885895984 | 1 | 0.05401176 | 0.05401176 | 1.068325064 | 459.5714824 | 2.662353072 | FALSE | TRUE | FALSE |
| CUTC | 24.819265 | 0.10463992 | 0.18935 | 4.585235938 | 99.25894932 | 397.0625837 | 3.64078664 | 0.375191579 | 0.6186 | 3.640786638 | 137.8031755 | 137.8031755 | 6.81700619 | 0.833593688 | FALSE | FALSE | FALSE |
| TMEM231 | 24.67435 | 0.10491992 | 0.77215 | 231.6813144 | 22026.44842 | 55680.21015 | 2.14188954 | 0.477249384 | 0.75485 | 2.141889537 | 20.25186883 | 20.25186883 | 11.51989825 | 1.061448643 | FALSE | FALSE | FALSE |
| CLDN14 | 24.611444 | 0.10519997 | 0.19495 | 1.537972427 | 24.61144409 | 98.49044591 | 32.4658982 | 0.0844391 | 0.19125 | 2.02853723 | 32.46589821 | 129.9308787 | 0.758070635 | -0.120290326 | FALSE | FALSE | FALSE |
| KRT1 | 24.602376 | 0.10547963 | 0.12085 | 2.587073093 | 98.36834527 | 393.4070975 | 1.93764019 | 0.495076532 | 0.1401 | 0.484558485 | 1.937640192 | 4.103237937 | 12.6970817 | 1.103703914 | FALSE | FALSE | FALSE |
| OR5M10 | 24.431857 | 0.10576026 | 0.50315 | 24.43185664 | 237.108983 | 237.108983 | 0.35637786 | 0.806920348 | 1 | 0.356377863 | 0.356377863 | 0.356377863 | 68.55604453 | 1.836045752 | FALSE | TRUE | FALSE |
| GBE1 | 24.371494 | 0.10604093 | 0.6857 | 175.4907153 | 30120.8537 | 133659.0618 | 0.02476015 | 0.892516811 | 0.8911 | 0.395727614 | 6.331017338 | 25.33230856 | 984.3032217 | 2.993128907 | FALSE | TRUE | FALSE |
| WNT2B | 24.303813 | 0.10632171 | 0.07335 | 1.518466505 | 24.30381298 | 29.9987645 | 0.12968754 | 0.869035195 | 0.5069 | 0.51854868 | 0.51854868 | 2.565863563 | 187.4028321 | 2.27277615 | FALSE | TRUE | FALSE |
| CDHR3 | 24.302359 | 0.10660203 | 0.3599 | 2.662435892 | 227.2622961 | 2176.054079 | 46.2017678 | 0.059661094 | 0.5039 | 46.20176777 | 4236.694566 | 42010.57751 | 0.526004965 | -0.279010157 | FALSE | FALSE | FALSE |
| PRDM7 | 24.171977 | 0.10688298 | 0.85545 | 229.3243332 | 22180.40643 | 225875.729 | 4.59108672 | 0.324522396 | 0.50715 | 4.591086722 | 43.04609372 | 43.04609372 | 5.264979316 | 0.721396669 | FALSE | FALSE | FALSE |
| LIX1 | 24.100929 | 0.10716408 | 0.00565 | 2.545847207 | 24.10092894 | 24.10092894 | 0.33073069 | 0.815162075 | 1 | 3.063622189 | 3.063622189 | 3.063622189 | 72.87176407 | 1.862559283 | FALSE | TRUE | FALSE |
| TUFT1 | 24.094 | 0.10744476 | 0.69155 | 24.09399994 | 2345.699101 | 2345.699101 | 3.25695147 | 0.397112543 | 0.50735 | 3.25695147 | 3.25695147 | 3.25695147 | 7.397715367 | 0.869097617 | FALSE | FALSE | FALSE |
| GOLGA8B | 24.052204 | 0.10772534 | 0.1372 | 24.05220427 | 239.6777074 | 239.6777074 | 2.61433888 | 0.439409621 | 0.87605 | 2.614338877 | 253.3157281 | 253.3157281 | 9.200109628 | 0.963793002 | FALSE | FALSE | FALSE |
| C7orf57 | 24.051443 | 0.10800544 | 0.2501 | 2.543271524 | 24.05144316 | 24.05144316 | 0.33065802 | 0.815183155 | 1 | 3.060105325 | 3.060105325 | 3.060105325 | 72.73812155 | 1.861762081 | FALSE | TRUE | FALSE |
| SLC23A3 | 23.823748 | 0.10828705 | 0.1293 | 2.487458914 | 23.82374782 | 23.82374782 | 0.03958937 | 0.889046153 | 0.011 | 0.03958937 | 0.03958937 | 0.785939095 | 601.7713257 | 2.77943149 | FALSE | TRUE | FALSE |
| HDAC10 | 23.747937 | 0.10856884 | 0.4298 | 10.38434507 | 890.4489773 | 7535.396793 | 42.3485789 | 0.064681503 | 0.42335 | 18.33560705 | 1603.814 | 13598.82305 | 0.56077295 | -0.251212944 | FALSE | FALSE | FALSE |
| PMS2 | 23.725804 | 0.10885035 | 0.51525 | 23.72580437 | 1522.513284 | 6098.520368 | 0.79677646 | 0.621647385 | 0.7908 | 12.75246464 | 452.7317219 | 1812.344665 | 29.77724058 | 1.47388445 | FALSE | TRUE | FALSE |
| TMEM120B | 23.723659 | 0.10913142 | 0.00815 | 2.493940634 | 23.72365882 | 23.72365882 | 3.011264 | 0.410704246 | 0.4984 | 0.323688794 | 3.011264002 | 3.011264002 | 7.878305857 | 0.896432837 | FALSE | FALSE | FALSE |
| PIGL | 23.687424 | 0.10941232 | 0.3455 | 5.91873509 | 94.82334194 | 379.6829777 | 0.50442058 | 0.753676267 | 0.87875 | 2.017813036 | 8.073792471 | 8.073792471 | 46.95967061 | 1.671725042 | FALSE | TRUE | FALSE |
| PRSS38 | 23.597715 | 0.10969354 | 0.0068 | 1.474023644 | 23.59771544 | 23.59771544 | 0.12605608 | 0.869900727 | 0.49985 | 0.126056079 | 0.50407778 | 0.50407778 | 187.2001381 | 2.272306165 | FALSE | TRUE | FALSE |
| RPS15 | 23.486897 | 0.10997527 | 0.00185 | 6.551845042 | 6.551845042 | 6.551845042 | 0.72228194 | 0.655381605 | 1 | 0.722281943 | 6.847033012 | 6.847033012 | 32.51762989 | 1.512118884 | FALSE | TRUE | FALSE |
| TECTB | 23.437362 | 0.11025695 | 0.1268 | 2.491910469 | 23.43736244 | 23.43736244 | 0.04987692 | 0.886667684 | 0.00865 | 0.049876923 | 0.049876923 | 0.049876923 | 469.9039368 | 2.672009083 | FALSE | TRUE | FALSE |
| PIK3R3 | 23.37271 | 0.11053874 | 0.77845 | 93.55141818 | 3460.899614 | 12464.19731 | 1.73575872 | 0.514058078 | 0.9387 | 6.944197417 | 111.2277204 | 111.2277204 | 13.4654144 | 1.129219723 | FALSE | FALSE | FALSE |
| CEMP1 | 23.315807 | 0.11082056 | 0.8123 | 225.5413809 | 2236.352218 | 22716.10635 | 0.35461754 | 0.807517405 | 1 | 3.282779168 | 3.282779168 | 3.282779168 | 65.74916402 | 1.817890235 | FALSE | TRUE | FALSE |
| PLXNB1 | 23.280423 | 0.11110222 | 0.1518 | 2.435858185 | 93.10407177 | 326.4874691 | 0.04827193 | 0.88705409 | 1 | 0.048271927 | 0.924248365 | 1.620602813 | 482.2766368 | 2.683296224 | FALSE | TRUE | FALSE |
| RPL35 | 23.163513 | 0.11138446 | 0.00115 | 0.68205071 | 0.68205071 | 0.68205071 | 0.71373825 | 0.659213336 | 1 | 0.713738245 | 0.713738245 | 0.713738245 | 32.45379239 | 1.511265454 | FALSE | TRUE | FALSE |
| TM7SF2 | 23.0013 | 0.11166766 | 0.283 | 5.749451832 | 92.04214233 | 368.408321 | 0.78381441 | 0.626587354 | 0.8318 | 3.134677415 | 200.8085948 | 803.8880236 | 29.34533922 | 1.467539134 | FALSE | TRUE | FALSE |
| ERCC2 | 22.899503 | 0.11195131 | 0.36115 | 12.46862525 | 1780.920335 | 12775.40562 | 0.6965882 | 0.66783439 | 0.40485 | 0.315745075 | 25.17670492 | 100.6430913 | 32.87380256 | 1.516849942 | FALSE | TRUE | FALSE |
| LIPK | 22.828965 | 0.11223513 | 0.7096 | 91.3564029 | 2946.020967 | 26363.03016 | 3.99253438 | 0.356338414 | 0.50215 | 3.992534384 | 15.97804133 | 63.9597702 | 5.717913123 | 0.757237552 | FALSE | FALSE | FALSE |
| IL10RA | 22.776878 | 0.11251894 | 0.01145 | 5.691991048 | 22.77687822 | 28.14319757 | 1.64450442 | 0.522043867 | 0.5021 | 1.644504418 | 1.644504418 | 8.143626692 | 13.8502992 | 1.141459155 | FALSE | FALSE | FALSE |
| AKR1C3 | 22.529062 | 0.11280453 | 0.66645 | 22.52906225 | 2031.034075 | 19985.20913 | 0.424469 | 0.783004662 | 0.97065 | 35.40815718 | 335.7338815 | 3261.220329 | 53.07587163 | 1.724897135 | FALSE | TRUE | FALSE |
| RASGEF1C | 22.347783 | 0.11309131 | 0.2538 | 2.358992678 | 22.34778263 | 22.34778263 | 2.86497563 | 0.422723185 | 0.0108 | 2.864975627 | 2.864975627 | 6.129150713 | 7.800339529 | 0.892113507 | FALSE | FALSE | FALSE |
| FZD5 | 22.249021 | 0.11337852 | 0.3834 | 9.434948092 | 89.00600721 | 356.1526983 | 0.78127835 | 0.627687075 | 0.756 | 0.781278355 | 3.124847305 | 15.36082101 | 28.47771329 | 1.454505113 | FALSE | TRUE | FALSE |
| RNF103 | 22.130797 | 0.11366635 | 0.50275 | 22.1307971 | 217.3851936 | 217.3851936 | 0.31073926 | 0.821823842 | 1 | 2.910821444 | 2.910821444 | 10.51252819 | 71.21982991 | 1.852600932 | FALSE | TRUE | FALSE |
| NTSR2 | 22.098173 | 0.113954 | 0.2523 | 2.338474954 | 22.09817283 | 22.09817283 | 2.75269613 | 0.430877608 | 0.7517 | 2.752696126 | 26.10534626 | 26.10534626 | 8.027828653 | 0.904598094 | FALSE | FALSE | FALSE |
| APOL6 | 22.085511 | 0.11424131 | 0.0071 | 2.341595697 | 22.08551077 | 22.08551077 | 0.31767252 | 0.819363436 | 0.00945 | 0.317672517 | 0.317672517 | 0.317672517 | 69.52288792 | 1.842127804 | FALSE | TRUE | FALSE |
| C10orf95 | 21.806297 | 0.11453074 | 0.8167 | 210.2338062 | 2077.604344 | 21033.15069 | 0.33622001 | 0.813416939 | 0.50485 | 3.103251861 | 3.103251861 | 3.103251861 | 64.85722605 | 1.81195837 | FALSE | TRUE | FALSE |
| WNT9B | 21.791544 | 0.11481984 | 0.50445 | 21.79154387 | 818.6725989 | 3276.826421 | 0.11684976 | 0.872013341 | 1 | 4.387826263 | 42.29774795 | 42.29774795 | 186.4919908 | 2.270660185 | FALSE | TRUE | FALSE |
| ZKSCAN7 | 21.746635 | 0.11510888 | 0.91375 | 2012.463389 | 204679.181 | 2135718.413 | 0.45653957 | 0.772224977 | 0.9386 | 4.180023147 | 377.8874646 | 377.8874646 | 47.63362476 | 1.677913631 | FALSE | TRUE | FALSE |
| WEE2 | 21.688907 | 0.11539799 | 0.12715 | 2.331125969 | 21.68890739 | 21.68890739 | 1.44320311 | 0.541550274 | 0.50895 | 1.443203109 | 13.16407523 | 13.16407523 | 15.02831255 | 1.176910219 | FALSE | FALSE | FALSE |
| KRT6C | 21.687461 | 0.11568664 | 0.9324 | 3300.741615 | 846627.1317 | 8259426.667 | 48.2970615 | 0.056832429 | 0.14465 | 5.166815331 | 193.1438712 | 772.5909835 | 0.449043069 | -0.347712002 | FALSE | FALSE | FALSE |
| ALG10B | 21.592245 | 0.11597572 | 0.8595 | 205.7820409 | 20084.69192 | 205462.1848 | 16.9282114 | 0.147934631 | 0.12485 | 1.842765072 | 16.92821143 | 16.92821143 | 1.275518416 | 0.105686733 | FALSE | FALSE | FALSE |
| ZNF697 | 21.568369 | 0.11626454 | 0.81325 | 44.85467787 | 2093.068602 | 21385.11836 | 0.29893434 | 0.825763851 | 1 | 26.42397239 | 2584.949669 | 2584.949669 | 72.15085718 | 1.858241495 | FALSE | TRUE | FALSE |
| CRYBA2 | 21.551505 | 0.11655306 | 0.1916 | 5.387783461 | 203.8515108 | 203.8515108 | 0.77881093 | 0.629142659 | 0.7529 | 0.778810928 | 3.115382884 | 3.115382884 | 27.67232011 | 1.442045573 | FALSE | TRUE | FALSE |
| FAM210B | 21.486336 | 0.11684172 | 0.0029 | 5.371635013 | 21.48633644 | 21.48633644 | 0.61792532 | 0.70461572 | 1 | 5.80458462 | 5.80458462 | 5.80458462 | 34.77173647 | 1.541226379 | FALSE | TRUE | FALSE |
| MRPL22 | 21.473808 | 0.11713002 | 0.2505 | 2.275555956 | 21.4738082 | 21.4738082 | 0.30963149 | 0.82217534 | 0.0069 | 0.309631489 | 0.309631489 | 0.309631489 | 69.35279175 | 1.841063948 | FALSE | TRUE | FALSE |
| TTLL7 | 21.414781 | 0.11741842 | 0.011 | 0.635265846 | 6.037625703 | 21.41478136 | 0.08688996 | 0.878818393 | 0.0212 | 0.086889965 | 0.086889965 | 1.712614417 | 246.4586268 | 2.391744024 | FALSE | TRUE | FALSE |
| OR5I1 | 21.210501 | 0.11770829 | 0.0056 | 2.235257735 | 21.2105007 | 21.2105007 | 0.27665697 | 0.831916313 | 1 | 24.37812902 | 237.7826304 | 237.7826304 | 76.66714722 | 1.884609304 | FALSE | TRUE | FALSE |
| PGAP3 | 21.195138 | 0.11799784 | 0.3192 | 5.296794357 | 84.83359703 | 84.83359703 | 0.38989438 | 0.795744869 | 1 | 1.559423945 | 1.559423945 | 1.559423945 | 54.36122932 | 1.735289269 | FALSE | TRUE | FALSE |
| VOPP1 | 21.191109 | 0.11828695 | 0.81365 | 44.93538994 | 2016.696003 | 20404.92059 | 27.7000436 | 0.100196957 | 0.0033 | 2.928334166 | 27.7000436 | 27.7000436 | 0.765020781 | -0.116326767 | FALSE | FALSE | FALSE |
| EPO | 21.067144 | 0.11857679 | 0.24875 | 2.244411968 | 21.06714414 | 21.06714414 | 0.10763984 | 0.873863187 | 1 | 1.011756359 | 1.011756359 | 1.011756359 | 195.7188265 | 2.291632603 | FALSE | TRUE | FALSE |
| DNA2 | 21.012555 | 0.11886668 | 0.0568 | 0.267803819 | 38.41021471 | 153.5862466 | 4.113E-06 | 0.900894173 | 0.02305 | 4.11299E-06 | 4.11299E-06 | 8.09265E-05 | 5108830.222 | 6.70832147 | FALSE | TRUE | FALSE |
| CNGA1 | 20.982444 | 0.11915639 | 0.88525 | 4399.306387 | 4560741.258 | 72897313.34 | 26.0674066 | 0.105254907 | 0.9485 | 8825.794038 | 5403543.465 | 52871271.55 | 0.804930236 | -0.094241759 | FALSE | FALSE | FALSE |
| AAAS | 20.918444 | 0.11944624 | 0.05 | 0.245381621 | 19.46258825 | 83.59565557 | 2.40515322 | 0.455075343 | 0.1282 | 0.150599907 | 2.405153218 | 9.615338667 | 8.697343776 | 0.939386637 | FALSE | FALSE | FALSE |
| ACSM1 | 20.892294 | 0.11973588 | 0.49805 | 4.909666289 | 198.9192639 | 1940.272871 | 0.04776299 | 0.887193269 | 1 | 0.426383576 | 0.426383576 | 0.945404036 | 437.4159411 | 2.640894606 | FALSE | TRUE | FALSE |
| FNBP4 | 20.768268 | 0.12002626 | 0.1456 | 20.76826813 | 203.9708207 | 408.7852662 | 0.30327789 | 0.824254976 | 1 | 0.303277889 | 0.303277889 | 5.980852322 | 68.47933481 | 1.835559533 | FALSE | TRUE | FALSE |
| TUBB1 | 20.756833 | 0.12031627 | 0.2122 | 2.304998382 | 83.02336342 | 332.1604909 | 24.5998564 | 0.111850936 | 0.22855 | 2.647873131 | 98.36134748 | 393.3913878 | 0.843778642 | -0.073771472 | FALSE | FALSE | FALSE |

| | | | | | | | | | | | | | | | | | |
|---|---|---|---|---|---|---|---|---|---|---|---|---|---|---|---|---|---|
| PCLO | 20.585077 | 0.1206075 | 0.3811 | 2.13526765 | 378.4920786 | 3738.071544 | 4.45795084 | 0.331868147 | 0.2581 | 4.457950844 | 146.346195 | 1421.853198 | 4.617609661 | 0.664417218 | FALSE | FALSE | FALSE |
| OR4B1 | 20.47905 | 0.12089929 | 0.99485 | 209382.8251 | 26400188.4 | 294465356.2 | 0.39136103 | 0.795174535 | 1 | 0.391361034 | 0.391361034 | 0.391361034 | 52.32776884 | 1.718732218 | FALSE | TRUE | FALSE |
| ATP6V0A1 | 20.470456 | 0.1211907 | 0.1038 | 1.280741181 | 20.47045622 | 81.86972478 | 0.99671262 | 0.588672434 | 0.5084 | 0.996712621 | 3.985804621 | 19.62885863 | 20.53797233 | 1.312557564 | FALSE | TRUE | FALSE |
| PODN | 20.448778 | 0.12148186 | 0.011 | 2.218415931 | 20.44877754 | 43.61950561 | 0.09155113 | 0.877824181 | 1 | 0.091551133 | 0.091551133 | 1.803777659 | 223.3590865 | 2.349003625 | FALSE | TRUE | FALSE |
| CCSAP | 20.421139 | 0.12177281 | 0.0026 | 0.602120041 | 0.602120041 | 0.602120041 | 0.63609709 | 0.69668089 | 1 | 0.636097095 | 0.636097095 | 0.636097095 | 32.10380872 | 1.506556559 | FALSE | TRUE | FALSE |
| PPM1N | 20.384165 | 0.12206366 | 0.8131 | 197.6089858 | 1963.622151 | 19988.86621 | 0.32459843 | 0.817307955 | 1 | 0.324598431 | 0.324598431 | 6.444656502 | 62.79809986 | 1.797946503 | FALSE | TRUE | FALSE |
| RNF182 | 20.362442 | 0.12235426 | 0.2552 | 2.126020536 | 20.36244175 | 20.36244175 | 0.28865211 | 0.828506723 | 1 | 0.288652106 | 0.288652106 | 0.288652106 | 70.54319489 | 1.848455125 | FALSE | TRUE | FALSE |
| HAX1 | 20.339087 | 0.12264462 | 0.2553 | 2.159286351 | 20.33908733 | 20.33908733 | 2.5645296 | 0.442530471 | 0.25945 | 2.5645296 | 24.25126017 | 24.25126017 | 7.93092321 | 0.899323745 | FALSE | FALSE | FALSE |
| DNTT | 20.111597 | 0.12293677 | 0.07005 | 1.256646856 | 20.11159666 | 20.11159666 | 0.84676867 | 0.608332577 | 0.31515 | 0.846768667 | 13.55928903 | 13.55928903 | 23.75099297 | 1.375681771 | FALSE | TRUE | FALSE |
| MLLT3 | 20.011846 | 0.12322945 | 0.255 | 5.582944516 | 54.1576071 | 110.5033114 | 0.64908796 | 0.690333825 | 1 | 6.144276654 | 6.144276654 | 12.86340283 | 30.830715 | 1.488983597 | FALSE | TRUE | FALSE |
| HSD3B2 | 19.852111 | 0.12352327 | 0.19175 | 2.124308961 | 79.39810751 | 79.39810751 | 0.0847764 | 0.879180506 | 0.7432 | 0.338993162 | 3.104274559 | 3.104274559 | 234.1702532 | 2.369531725 | FALSE | TRUE | FALSE |
| DTWD2 | 19.844307 | 0.12381669 | 0.3255 | 4.960845386 | 186.9354041 | 186.9354041 | 0.18568321 | 0.855633526 | 1 | 0.185683211 | 0.185683211 | 0.185683211 | 106.8718435 | 2.028863301 | FALSE | TRUE | FALSE |
| NGDN | 19.833722 | 0.12410976 | 0.2534 | 2.099982428 | 19.83372157 | 19.83372157 | 0.28830243 | 0.828651548 | 1 | 0.288302429 | 0.288302429 | 0.288302429 | 68.79484727 | 1.837555911 | FALSE | TRUE | FALSE |
| REM2 | 19.83361 | 0.12440235 | 0.25275 | 8.353194854 | 79.32410243 | 79.32410243 | 2.36642605 | 0.458770147 | 0.9371 | 22.49952452 | 2193.197474 | 2193.197474 | 8.38125054 | 0.923308823 | FALSE | FALSE | FALSE |
| BAHD1 | 19.658889 | 0.12469625 | 0.2637 | 2.042942929 | 19.65888941 | 68.9923323 | 2.53040434 | 0.444849941 | 0.0242 | 2.530404337 | 2.530404337 | 49.6851482 | 7.769070392 | 0.890369056 | FALSE | FALSE | FALSE |
| RFNG | 19.658409 | 0.12498968 | 0.3819 | 8.542921149 | 78.65351814 | 314.7722591 | 11.0296746 | 0.187414585 | 0.50045 | 11.02967459 | 44.14200256 | 176.7054092 | 1.782319925 | 0.250985662 | FALSE | FALSE | FALSE |
| ITGAD | 19.633845 | 0.12528289 | 0.3838 | 6.146564306 | 48488.38955 | 483539.5548 | 28522.3865 | 6.34192E-05 | 0.5081 | 287472.5322 | 334464004.1 | 3665600286 | 0.000688366 | -3.162180494 | FALSE | FALSE | FALSE |
| GLIS3 | 19.607843 | 0.1255759 | 0.50685 | 19.60784347 | 749.5622712 | 7340.145977 | 307.841925 | 0.007913901 | 0.0702 | 3.394027828 | 307.8419254 | 307.8419254 | 0.063694519 | -1.195897935 | FALSE | FALSE | FALSE |
| OTOP1 | 19.570577 | 0.12586881 | 0.77905 | 183.0928951 | 17221.26947 | 34610.4807 | 28.4225189 | 0.097244686 | 0.91245 | 2592.03639 | 259791.1456 | 2690986.909 | 0.688558845 | -0.162058938 | FALSE | FALSE | FALSE |
| OR4D10 | 19.319669 | 0.12616384 | 0.68865 | 19.31966886 | 1846.578754 | 1846.578754 | 224.667379 | 0.011566601 | 0.06775 | 23.17387705 | 2232.631178 | 2232.631178 | 0.085992319 | -1.065540341 | FALSE | FALSE | FALSE |
| RIMS1 | 19.311388 | 0.12645848 | 0.3724 | 19.31138847 | 748.898589 | 7439.627947 | 0.37937692 | 0.799398534 | 0.76075 | 1.517212812 | 24.28443416 | 117.8881256 | 50.90290839 | 1.706742597 | FALSE | TRUE | FALSE |
| OR4M1 | 19.304635 | 0.12675272 | 0.25485 | 2.028203574 | 19.30463454 | 19.30463454 | 0.26139261 | 0.836308064 | 1 | 2.430796397 | 23.20027529 | 23.20027529 | 73.85302413 | 1.868368283 | FALSE | TRUE | FALSE |
| TRPM2 | 19.28195 | 0.12704672 | 0.1506 | 0.463767863 | 170.7326601 | 1771.781697 | 0.54286908 | 0.736971002 | 0.7469 | 10.82959258 | 6385.401106 | 60502.19492 | 35.51860039 | 1.550455844 | FALSE | TRUE | FALSE |
| VSX1 | 19.279425 | 0.12734029 | 0.1268 | 2.803665102 | 44.81981319 | 179.2688405 | 28.7710756 | 0.095175945 | 0.3825 | 7.192793792 | 460.679162 | 460.679162 | 0.670097465 | -0.173862025 | FALSE | FALSE | FALSE |
| CYP1A1 | 19.252067 | 0.12763367 | 1 | 2.28387E+15 | 3.94829E+20 | 7.1596E+22 | 9.45169344 | 0.205727123 | 1 | 7.55476E+13 | 7.77182E+18 | 1.19246E+21 | 2.036890708 | 0.308967727 | FALSE | FALSE | FALSE |
| NUP54 | 19.178097 | 0.12792734 | 0.50575 | 19.17809693 | 186.4725004 | 186.4725004 | 0.28711926 | 0.828976517 | 1 | 0.287119262 | 0.287119262 | 5.698376242 | 66.79488086 | 1.82474318 | FALSE | TRUE | FALSE |
| LRRC14B | 19.105716 | 0.12822131 | 0.89175 | 426.0304553 | 27306.09769 | 267743.6142 | 0.75367976 | 0.639775124 | 0.87485 | 3.015345162 | 27.61145756 | 27.61145756 | 25.34991297 | 1.403976473 | FALSE | TRUE | FALSE |
| RPS6KA5 | 19.057718 | 0.1285153 | 0.01555 | 1.992752025 | 19.05771795 | 39.22843301 | 0.27301096 | 0.832937274 | 1 | 0.273010959 | 0.273010959 | 5.384543752 | 69.80568849 | 1.843890815 | FALSE | TRUE | FALSE |
| CES2 | 18.960432 | 0.12880984 | 1 | 4.16126E+24 | 2.11965E+31 | 1.78649E+34 | 0.01407647 | 0.89539743 | 1 | 2.08639E+13 | 6.82616E+17 | 1.10035E+20 | 1346.959581 | 3.129354564 | FALSE | TRUE | FALSE |
| DDR2 | 18.897267 | 0.12910458 | 0.2403 | 4.723965794 | 75.61356747 | 302.627837 | 0.12393444 | 0.870387507 | 0.87345 | 0.495464541 | 1.981260927 | 9.760290041 | 152.4779317 | 2.183206992 | FALSE | TRUE | FALSE |
| TLX1 | 18.856604 | 0.12939927 | 0.31845 | 4.713122955 | 75.46172096 | 75.46172096 | 12.7446496 | 0.172548398 | 0.3776 | 3.186930605 | 50.97905198 | 50.97905198 | 1.479570227 | 0.170135584 | FALSE | FALSE | FALSE |
| C5 | 18.833628 | 0.12969374 | 0.2358 | 2.127414052 | 170.818471 | 1586.383583 | 0.00874819 | 0.897104375 | 0.87945 | 0.073150342 | 0.627823871 | 2.459137983 | 2152.859631 | 3.333015714 | FALSE | TRUE | FALSE |
| ERAP2 | 18.825523 | 0.12998783 | 0.6309 | 171.1118234 | 147768.1766 | 1471746.38 | 4.54798392 | 0.326168683 | 0.7741 | 41.08939025 | 3604.349488 | 7497.178673 | 4.139311658 | 0.616928127 | FALSE | FALSE | FALSE |
| GYG1 | 18.769573 | 0.13028205 | 0.0215 | 1.172005017 | 18.76957325 | 75.14144711 | 27.6331276 | 0.100786796 | 0.72655 | 110.6291698 | 3936.966168 | 3936.966168 | 0.679241724 | -0.167975644 | FALSE | FALSE | FALSE |
| GAL3ST1 | 18.732238 | 0.13057619 | 0.0026 | 0.552572683 | 0.552572683 | 10.88305213 | 0.59284945 | 0.714851316 | 1 | 0.592849448 | 0.592849448 | 11.69772841 | 31.59695622 | 1.499645248 | FALSE | TRUE | FALSE |
| OXGR1 | 18.730879 | 0.13086988 | 0.5043 | 18.73087916 | 182.1091872 | 182.1091872 | 0.24845456 | 0.839942623 | 1 | 21.85256713 | 2127.126236 | 2127.126236 | 75.38955727 | 1.877311193 | FALSE | TRUE | FALSE |
| HCAR3 | 18.422667 | 0.13116637 | 0.81735 | 178.9464767 | 1781.682133 | 18172.52467 | 2.5762461 | 0.441558747 | 0.74995 | 2.576246105 | 24.54499433 | 24.54499433 | 7.150973301 | 0.854365157 | FALSE | FALSE | FALSE |
| CYP4X1 | 18.38976 | 0.13146275 | 0.00775 | 1.953999595 | 18.38975989 | 18.38975989 | 0.14977664 | 0.864621401 | 0.8762 | 1.349356224 | 12.47698859 | 12.47698859 | 122.7812261 | 2.089131966 | FALSE | TRUE | FALSE |
| AHCYL2 | 18.350181 | 0.13175909 | 0.13335 | 1.959262909 | 18.35018111 | 22.71613977 | 0.16387429 | 0.86112907 | 1 | 0.655259646 | 0.655259646 | 3.247935574 | 111.9771834 | 2.049129539 | FALSE | TRUE | FALSE |
| PDE10A | 18.27838 | 0.13205573 | 0.88905 | 179.2783764 | 18562.94736 | 196409.5165 | 2.62622735 | 0.438428494 | 0.75765 | 2.626227352 | 25.26142689 | 51.8046416 | 6.959938112 | 0.842605378 | FALSE | FALSE | FALSE |
| XKR9 | 18.184264 | 0.13235292 | 0.78355 | 164.4342764 | 14527.97629 | 58155.62751 | 14.3588534 | 0.160856839 | 0.60225 | 57.45201847 | 2145.954564 | 6381.061006 | 1.266414792 | 0.102575974 | FALSE | FALSE | FALSE |
| NAIP | 18.065918 | 0.13265092 | 0.26335 | 1.87124132 | 18.06591784 | 62.92632091 | 0.25651098 | 0.837553778 | 0.03215 | 0.256510984 | 0.256510984 | 5.0195308 | 70.42941234 | 1.847754065 | FALSE | TRUE | FALSE |
| CCRL2 | 18.022428 | 0.13294892 | 0.0059 | 1.910749519 | 18.02242762 | 18.02242762 | 0.26553041 | 0.835060021 | 1 | 0.265530408 | 0.265530408 | 0.265530408 | 67.87330974 | 1.831699027 | FALSE | TRUE | FALSE |
| APPL2 | 17.91123 | 0.13324767 | 0.1969 | 2.039490251 | 40.3301218 | 161.3485729 | 0.22476983 | 0.846202234 | 0.51165 | 0.224769827 | 0.224769827 | 0.512613451 | 79.68698767 | 1.90138741 | FALSE | TRUE | FALSE |
| DHODH | 17.906448 | 0.133546 | 0.0661 | 0.475146433 | 17.90644814 | 17.90644814 | 12.1642896 | 0.177578055 | 0.5822 | 12.16428964 | 460.4556714 | 1842.060446 | 1.472050457 | 0.167922697 | FALSE | FALSE | FALSE |
| PLA2G12A | 17.893241 | 0.13384401 | 0.50725 | 17.89324143 | 71.61680082 | 286.7142581 | 20.3214392 | 0.130760572 | 0.77605 | 81.33625665 | 1175.977593 | 1175.977593 | 0.88051054 | -0.055265441 | FALSE | FALSE | FALSE |
| PRKAA2 | 17.811621 | 0.13414245 | 0.2002 | 7.793738795 | 71.27173853 | 285.2592751 | 0.66373544 | 0.683633917 | 0.4978 | 0.165953304 | 0.663735438 | 3.289089796 | 26.83542214 | 1.428708431 | FALSE | TRUE | FALSE |
| PNPLA2 | 17.807492 | 0.13444046 | 0.00605 | 1.113114054 | 17.80749226 | 17.80749226 | 0.04939214 | 0.886815957 | 1 | 0.049392137 | 0.049392137 | 0.978504864 | 360.5329386 | 2.556944948 | FALSE | TRUE | FALSE |
| C14orf119 | 17.777826 | 0.13473834 | 0.2527 | 1.901798253 | 17.77782628 | 17.77782628 | 0.26582265 | 0.834959356 | 1 | 0.265822648 | 0.265822648 | 0.265822648 | 66.87852388 | 1.825286679 | FALSE | TRUE | FALSE |
| TCF25 | 17.736202 | 0.13503621 | 0.8316 | 70.98086159 | 4556.525766 | 4556.525766 | 99.5230776 | 0.029508064 | 0.37445 | 24.86650601 | 398.420137 | 1595.391189 | 0.178211948 | -0.749063183 | FALSE | FALSE | FALSE |
| HORMAD2 | 17.710749 | 0.13533389 | 0.77555 | 1673.933979 | 174253.9079 | 174253.9079 | 2.35238589 | 0.460234408 | 0.9853 | 213.8114841 | 21454.35106 | 222704.5871 | 7.528845076 | 0.876728361 | FALSE | FALSE | FALSE |
| RAPGEF4 | 17.683515 | 0.13563141 | 0.2039 | 4.419223495 | 21.64746074 | 86.60063643 | 0.0978905 | 0.876507406 | 1 | 0.391436781 | 1.565637668 | 13.20449982 | 180.645869 | 2.256828034 | FALSE | TRUE | FALSE |
| CD48 | 17.587267 | 0.13592953 | 0.25365 | 1.845891501 | 17.58726658 | 17.58726658 | 19.6976515 | 0.13385491 | 0.0027 | 2.061948632 | 19.69765154 | 19.69765154 | 0.892861088 | -0.049216104 | FALSE | FALSE | FALSE |
| UBE4B | 17.586783 | 0.13622719 | 0.03055 | 1.837806788 | 17.58678329 | 341.5237527 | 2.23023269 | 0.469395482 | 0.76305 | 2.230232685 | 21.38780742 | 75.15986604 | 7.885627094 | 0.896836236 | FALSE | FALSE | FALSE |
| OR6M1 | 17.58624 | 0.13652439 | 0.32015 | 17.58624023 | 1725.217311 | 1725.217311 | 21.0804346 | 0.12737153 | 0.2506 | 2.200588941 | 21.08043461 | 21.08043461 | 0.834244671 | -0.078706559 | FALSE | FALSE | FALSE |
| ZNF44 | 17.547285 | 0.13682156 | 0.6635 | 17.54728456 | 1605.252076 | 15911.47758 | 28.4998465 | 0.09665416 | 0.1975 | 28.49984649 | 272.0033693 | 2659.482204 | 0.615697511 | -0.210632602 | FALSE | FALSE | FALSE |
| PYHIN1 | 17.494315 | 0.13711885 | 0.8109 | 168.4782001 | 1663.146077 | 16818.93623 | 2.47418605 | 0.448680165 | 0.75375 | 2.474186055 | 23.39320188 | 23.39320188 | 7.070735409 | 0.849464586 | FALSE | FALSE | FALSE |
| ADCY4 | 17.414186 | 0.13741657 | 0.3758 | 17.41418589 | 1683.690391 | 17142.8206 | 0.45370028 | 0.773224001 | 0.6925 | 1.813427156 | 40.8000424 | 163.0766112 | 38.38257654 | 1.584134125 | FALSE | TRUE | FALSE |
| CR1L | 17.36574 | 0.13771437 | 0.502 | 17.36573995 | 1514.059988 | 3169.238232 | 0.48680096 | 0.760309567 | 0.5006 | 0.486800957 | 4.352659973 | 4.352659973 | 35.67318365 | 1.55234187 | FALSE | TRUE | FALSE |
| NEK11 | 17.293789 | 0.13801251 | 0.0036 | 0.234194337 | 2.045115058 | 2.045115058 | 0.01258085 | 0.895873267 | 1 | 0.108441186 | 0.108441186 | 0.249152888 | 1374.612052 | 3.138180147 | FALSE | TRUE | FALSE |
| SEMA6A | 17.257827 | 0.13831059 | 0.26675 | 1.806475864 | 17.25782708 | 60.99979282 | 0.25056811 | 0.839244494 | 1 | 0.250568106 | 0.250568106 | 4.910418592 | 68.87479547 | 1.838060322 | FALSE | TRUE | FALSE |
| MANSC1 | 17.206275 | 0.13860878 | 0.69115 | 17.20627529 | 1633.912342 | 1633.912342 | 170504.868 | 1.22356E-05 | 0.18105 | 16493.23774 | 19515097.06 | 35823257.41 | 0.000100914 | -3.996049916 | FALSE | FALSE | FALSE |
| ZNF485 | 17.015668 | 0.13890866 | 0.1324 | 17.01566768 | 164.3842578 | 164.3842578 | 0.25294787 | 0.838652721 | 1 | 2.334659546 | 2.334659546 | 2.334659546 | 67.26946398 | 1.827817967 | FALSE | TRUE | FALSE |
| TAF1B | 17.013088 | 0.1392081 | 0.66035 | 17.01308821 | 1571.106157 | 15646.41714 | 0.30689324 | 0.823182899 | 0.939 | 2.799960684 | 251.3437021 | 251.3437021 | 55.43650359 | 1.743795831 | FALSE | TRUE | FALSE |
| NUDT18 | 16.947775 | 0.13950782 | 0.69185 | 16.94777545 | 1608.815133 | 1608.815133 | 2.40727676 | 0.454330246 | 0.4974 | 0.26148359 | 2.407276759 | 2.407276759 | 7.040227256 | 0.847586678 | FALSE | FALSE | FALSE |
| SPZ1 | 16.929733 | 0.13980729 | 0.50025 | 16.92973327 | 162.6118553 | 162.6118553 | 0.25405212 | 0.838169733 | 1 | 2.332815578 | 2.332815578 | 2.332815578 | 66.63882041 | 1.823727301 | FALSE | TRUE | FALSE |
| UCP1 | 16.822797 | 0.1401075 | 0.16235 | 1.845791026 | 16.82279736 | 67.29943257 | 0.04715975 | 0.887299492 | 1 | 0.047159751 | 0.047159751 | 0.047159751 | 356.7193881 | 2.552326714 | FALSE | TRUE | FALSE |
| SLCO1A2 | 16.796027 | 0.14040756 | 0.0694 | 1.807753705 | 159.9659549 | 159.9659549 | 0.33086657 | 0.815140991 | 0.7511 | 0.330866567 | 3.027133934 | 3.062397312 | 50.76374795 | 1.705553679 | FALSE | TRUE | FALSE |
| HTR6 | 16.712233 | 0.14070812 | 0.2589 | 1.746913802 | 16.71223299 | 34.45180587 | 0.24302903 | 0.841134801 | 1 | 0.243029033 | 0.243029033 | 4.800934657 | 68.76640529 | 1.837376323 | FALSE | TRUE | FALSE |
| ADD2 | 16.646 | 0.14100897 | 0.01635 | 1.762512632 | 16.64599995 | 34.70036791 | 2.21491457 | 0.470983661 | 0.51225 | 2.214914569 | 2.214914569 | 8.103956598 | 7.515413995 | 0.875952909 | FALSE | FALSE | FALSE |
| POLG | 16.585986 | 0.14131005 | 1 | 1.83243E+17 | 4.22833E+23 | 1.57445E+26 | 0.00098389 | 0.899462026 | 0.99985 | 6136899.313 | 5.01286E+11 | 5.20525E+13 | 16857.6256 | 4.226796404 | FALSE | TRUE | FALSE |
| DQX1 | 16.573512 | 0.14161081 | 0.1264 | 1.81621622 | 16.57351233 | 16.57351233 | 23.7825704 | 0.114870708 | 0.35335 | 23.78257037 | 2164.645782 | 21401.64296 | 0.696876413 | -0.156844235 | FALSE | FALSE | FALSE |
| SNX27 | 16.559026 | 0.14191128 | 0.19535 | 7.255686904 | 66.25960712 | 265.1987319 | 0.15754392 | 0.862638173 | 1 | 0.157543919 | 0.157543919 | 3.120949403 | 105.1073608 | 2.021633131 | FALSE | TRUE | FALSE |

| | | | | | | | | | | | | | | | | | |
|---|---|---|---|---|---|---|---|---|---|---|---|---|---|---|---|---|---|
| TAF4 | 16.53127 | 0.14221161 | 0.99885 | 1822489.007 | 2565232052 | 30004905594 | 0.32326973 | 0.817616223 | 1 | 3.035418759 | 29.25214982 | 59.81466225 | 51.13769789 | 1.708741173 | FALSE | TRUE | FALSE |
| OR10AG1 | 16.508731 | 0.14251174 | 0.81425 | 35.1009639 | 1558.977125 | 15712.89478 | 0.25644566 | 0.837602572 | 1 | 2.355393666 | 22.20370644 | 22.20370644 | 64.37516064 | 1.808718326 | FALSE | TRUE | FALSE |
| TRUB2 | 16.181191 | 0.14281521 | 0.98135 | 15477.99408 | 1692231.423 | 18306660.32 | 1968040.47 | 8.2907E-07 | 0.1174 | 185567.874 | 18027047.36 | 199398223.6 | 8.22198E-06 | -5.085023527 | FALSE | FALSE | FALSE |
| ZNF30 | 16.140014 | 0.1431187 | 0.20015 | 16.14001412 | 153.0007801 | 1485.868069 | 25.6733824 | 0.106752575 | 0.4998 | 10.2421662 | 245.5080961 | 2405.132384 | 0.628667227 | -0.201579179 | FALSE | FALSE | FALSE |
| NEK1 | 16.08242 | 0.14342241 | 0.6421 | 59.70958587 | 5680.882023 | 55290.63969 | 335.373161 | 0.007335189 | 0.0249 | 3.863807392 | 335.3731612 | 3238.109366 | 0.047953808 | -1.319176897 | FALSE | FALSE | FALSE |
| GSTK1 | 16.08065 | 0.14372567 | 0.50915 | 156.4971436 | 1561.150748 | 15953.66372 | 2.21041755 | 0.471510784 | 0.5014 | 21.07651363 | 206.1233592 | 206.1233592 | 7.274937862 | 0.861829288 | FALSE | FALSE | FALSE |
| MIOS | 16.056719 | 0.14402876 | 0.01205 | 1.755023054 | 16.05671939 | 34.54395044 | 0.0747842 | 0.881428556 | 1 | 0.074784196 | 0.074784196 | 1.474779419 | 214.7073874 | 2.331846987 | FALSE | TRUE | FALSE |
| OGFOD3 | 15.962457 | 0.14433249 | 0.12735 | 3.991090274 | 148.9425664 | 148.9425664 | 0.07724518 | 0.881006746 | 1 | 0.077245179 | 0.077245179 | 0.077245179 | 206.6466373 | 2.315228342 | FALSE | TRUE | FALSE |
| SMCR8 | 15.883871 | 0.1446367 | 0.2658 | 1.661230157 | 15.88387057 | 32.60788607 | 0.23257574 | 0.844175026 | 1 | 0.232575741 | 0.232575741 | 4.575376808 | 68.29547445 | 1.834391926 | FALSE | TRUE | FALSE |
| EDEM3 | 15.852979 | 0.1449408 | 0.23195 | 6.802953667 | 151.5532756 | 606.1665519 | 0.07279211 | 0.881829953 | 1 | 0.665604624 | 0.665604624 | 13.13836982 | 217.7843061 | 2.338026581 | FALSE | TRUE | FALSE |
| ISX | 15.837672 | 0.14524463 | 0.0365 | 3.959073983 | 15.83767203 | 63.3720203 | 5.4387867 | 0.28946211 | 0.34665 | 5.438786702 | 87.0704113 | 348.5118688 | 2.911986238 | 0.464189318 | FALSE | FALSE | FALSE |
| CRTAP | 15.748028 | 0.14554907 | 0.25385 | 1.655011451 | 15.74802779 | 15.74802779 | 0.23258746 | 0.844160037 | 1 | 0.232587459 | 0.232587459 | 4.615978614 | 67.70798314 | 1.830639877 | FALSE | TRUE | FALSE |
| ZNF439 | 15.683503 | 0.14585381 | 0.9483 | 1403.59897 | 1416769.557 | 14862901.27 | 2203199.37 | 7.42215E-07 | 0.197 | 21038.54653 | 23309993.42 | 251999603.5 | 7.11851E-06 | -5.147610738 | FALSE | FALSE | FALSE |
| USP44 | 15.660365 | 0.14615837 | 0.0421 | 1.785433636 | 15.66036469 | 62.680878 | 0.04473967 | 0.887949194 | 0.5163 | 0.178941647 | 0.178941647 | 1.512260111 | 350.0330728 | 2.54410908 | FALSE | TRUE | FALSE |
| ASPG | 15.658054 | 0.1464625 | 0.4984 | 3.577854263 | 151.3151436 | 151.3151436 | 0.22786982 | 0.845409308 | 1 | 2.101891992 | 19.89877217 | 19.89877217 | 68.71490989 | 1.837050981 | FALSE | TRUE | FALSE |
| LMBRD1 | 15.6424 | 0.14676635 | 0.6584 | 15.64239957 | 1475.948665 | 14857.56016 | 0.26417366 | 0.83547812 | 0.9381 | 2.433505902 | 222.6873089 | 222.6873089 | 59.21256397 | 1.772413867 | FALSE | TRUE | FALSE |
| RAB26 | 15.613634 | 0.14707009 | 0.389 | 15.61363436 | 570.0549471 | 570.0549471 | 1.45882096 | 0.53924278 | 0.247 | 0.364742739 | 1.458820956 | 1.458820956 | 10.7029134 | 1.029502011 | FALSE | FALSE | FALSE |
| PON2 | 15.531522 | 0.14737435 | 0.00505 | 1.662089676 | 15.53152242 | 15.53152242 | 133017.676 | 1.49061E-05 | 0.61435 | 133017.6764 | 164771239 | 2462600081 | 0.000116763 | -3.932695329 | FALSE | FALSE | FALSE |
| SCAMP4 | 15.491969 | 0.14767864 | 0.0024 | 0.456583143 | 0.456583143 | 0.456583143 | 0.48515336 | 0.761099092 | 1 | 4.543238136 | 4.543238136 | 4.543238136 | 31.93210651 | 1.504227569 | FALSE | TRUE | FALSE |
| RGS6 | 15.460603 | 0.14798284 | 0.0109 | 1.646168291 | 15.46060288 | 32.59009824 | 0.233491 | 0.843829546 | 0.0123 | 0.233491002 | 0.233491002 | 4.628178295 | 66.21498378 | 1.820956277 | FALSE | TRUE | FALSE |
| CA12 | 15.447877 | 0.14828674 | 0.31095 | 1.659973515 | 147.4522735 | 147.4522735 | 0.6048014 | 0.709990379 | 0.5021 | 5.658135384 | 54.29311615 | 54.29311615 | 25.54206646 | 1.407256031 | FALSE | TRUE | FALSE |
| XIRP2 | 15.447635 | 0.14859018 | 0.14625 | 0.245531045 | 1056.457853 | 9005.06917 | 0.04444396 | 0.888013783 | 0.51855 | 0.044443959 | 22.88496571 | 191.5074691 | 347.5755945 | 2.541049274 | FALSE | TRUE | FALSE |
| EAF2 | 15.338933 | 0.14889448 | 0.0052 | 1.641223188 | 15.33893301 | 15.33893301 | 0.22399138 | 0.84639193 | 1 | 2.051327029 | 8.205056266 | 8.205056266 | 68.48001449 | 1.835563844 | FALSE | TRUE | FALSE |
| ABCA5 | 15.262515 | 0.14919925 | 0.15805 | 0.003891755 | 141.2960579 | 1339.369812 | 1.57270664 | 0.529090938 | 0.18 | 0.080165383 | 6.285040632 | 25.12331633 | 9.704616549 | 0.98697838 | FALSE | FALSE | FALSE |
| ZNF791 | 15.208294 | 0.14950422 | 0.01155 | 1.612524344 | 15.20829425 | 31.9157229 | 0.22866496 | 0.845232034 | 1 | 0.228664956 | 0.228664956 | 4.531479003 | 66.50907286 | 1.822880894 | FALSE | TRUE | FALSE |
| UFSP1 | 15.177566 | 0.14980911 | 0.25045 | 1.629845479 | 15.17756579 | 15.17756579 | 1650.38541 | 0.001445006 | 0.06505 | 17.52766429 | 1650.385413 | 1650.385413 | 0.009196377 | -2.036383252 | FALSE | FALSE | FALSE |
| DIAPH3 | 15.170663 | 0.15011362 | 0.54185 | 32.81224862 | 10572.63672 | 98352.65672 | 0.00327632 | 0.898566007 | 0.50725 | 0.003276318 | 0.494961762 | 4.469921801 | 4630.399657 | 3.665618477 | FALSE | TRUE | FALSE |
| MAPK9 | 15.162701 | 0.15041778 | 0.50655 | 15.16270139 | 148.4900338 | 148.4900338 | 0.22372563 | 0.846450207 | 0.5052 | 2.090111386 | 2.090111386 | 2.090111386 | 67.77364363 | 1.831060835 | FALSE | TRUE | FALSE |
| ZNF774 | 15.146739 | 0.15072167 | 0.64435 | 143.3207227 | 13789.06606 | 140052.07 | 0.3337182 | 0.814442488 | 0.50845 | 3.043324203 | 28.466634 | 28.466634 | 45.38781096 | 1.656939237 | FALSE | TRUE | FALSE |
| ZNF354C | 15.123288 | 0.1510254 | 0.00715 | 1.657442528 | 15.12328765 | 15.12328765 | 5.18677418 | 0.298962665 | 0.6953 | 5.186774181 | 471.0470596 | 471.0470596 | 2.91574052 | 0.464748872 | FALSE | FALSE | FALSE |
| BCL11B | 14.992156 | 0.15133028 | 0.00565 | 0.443491207 | 0.443491207 | 8.642454821 | 0.48842947 | 0.759514492 | 1 | 0.488429469 | 0.488429469 | 9.54758564 | 30.69461898 | 1.487062247 | FALSE | TRUE | FALSE |
| TECRL | 14.977364 | 0.15163488 | 0.2581 | 1.604837786 | 14.97736392 | 14.97736392 | 2.03334693 | 0.486722601 | 0.50925 | 2.033346933 | 2.033346933 | 2.033346933 | 7.365867416 | 0.867223897 | FALSE | FALSE | FALSE |
| PROSER2 | 14.941446 | 0.15193947 | 0.13015 | 14.94144646 | 142.9544799 | 142.9544799 | 0.22887744 | 0.84520245 | 1 | 2.094339351 | 2.094339351 | 2.094339351 | 65.28143014 | 1.81478966 | FALSE | TRUE | FALSE |
| MMP19 | 14.634256 | 0.15224742 | 0.07535 | 1.615581752 | 58.54803069 | 58.54803069 | 9.56056935 | 0.204375623 | 0.13465 | 1.073460721 | 9.560569346 | 9.560569346 | 1.530688786 | 0.1848869 | FALSE | FALSE | FALSE |
| LACTB2 | 14.58768 | 0.1525555 | 0.25215 | 3.647050005 | 58.36322176 | 58.36322176 | 0.13905659 | 0.866950881 | 1 | 0.139056595 | 0.139056595 | 0.139056595 | 104.904624 | 2.020794631 | FALSE | TRUE | FALSE |
| SERPIND1 | 14.583693 | 0.15286316 | 0.14865 | 0.91140349 | 14.58369303 | 58.35909941 | 0.45578897 | 0.772515958 | 0.51045 | 0.455788968 | 1.822802949 | 7.29162148 | 31.99659063 | 1.505103705 | FALSE | TRUE | FALSE |
| OR7C2 | 14.581108 | 0.1531704 | 0.13055 | 14.58110788 | 139.8163306 | 139.8163306 | 1.98373106 | 0.490191833 | 0.2553 | 1.983731061 | 18.65065388 | 18.65065388 | 7.350345095 | 0.866307729 | FALSE | FALSE | FALSE |
| TPRX1 | 14.562495 | 0.15347742 | 0.0059 | 1.555123172 | 14.56249515 | 14.56249515 | 0.21573467 | 0.848221946 | 1 | 1.97912776 | 1.97912776 | 1.97912776 | 67.50187612 | 1.829315844 | FALSE | TRUE | FALSE |
| INCENP | 14.489962 | 0.15378489 | 0.0232 | 1.527936085 | 14.4899622 | 51.6150274 | 0.21248417 | 0.848973348 | 1 | 0.212484166 | 0.212484166 | 4.164901393 | 68.19313864 | 1.83374068 | FALSE | TRUE | FALSE |
| TULP2 | 14.48047 | 0.15409203 | 0.3494 | 3.620604923 | 133.309035 | 533.29753 | 185.746759 | 0.014612946 | 0.3972 | 46.40436113 | 2978.319669 | 11930.50624 | 0.077958128 | -1.1081386 | FALSE | FALSE | FALSE |
| OR52W1 | 14.452928 | 0.15439905 | 0.6861 | 14.45292823 | 1364.631556 | 1364.631556 | 0.23517429 | 0.843315992 | 1 | 0.235174294 | 0.235174294 | 0.235174294 | 61.45624156 | 1.788565997 | FALSE | TRUE | FALSE |
| RGS20 | 14.399889 | 0.15470629 | 0.5008 | 30.57659953 | 1354.934113 | 13631.76876 | 2.15731875 | 0.475349867 | 0.49815 | 0.235049184 | 2.157318751 | 2.157318751 | 6.674900767 | 0.824444814 | FALSE | FALSE | FALSE |
| AGL | 14.395148 | 0.15501314 | 0.0283 | 0.027344999 | 7.609617987 | 58.66505084 | 5.5338E-05 | 0.900317041 | 0.46755 | 2.66138E-05 | 0.13947843 | 0.557539425 | 260132.6609 | 5.415194883 | FALSE | TRUE | FALSE |
| PIP5K1C | 14.289133 | 0.15532087 | 0.813 | 138.1063683 | 1368.232851 | 13886.29337 | 2.1324946 | 0.478108196 | 0.51325 | 2.132494601 | 2.132494601 | 7.808407392 | 6.700665359 | 0.826117929 | FALSE | FALSE | FALSE |
| CYP3A4 | 14.244912 | 0.1556287 | 0.6577 | 14.2449121 | 1329.385387 | 13308.8878 | 2.78481586 | 0.428454322 | 0.25975 | 0.303273749 | 2.784815858 | 2.784815858 | 5.115207908 | 0.70886329 | FALSE | FALSE | FALSE |
| NRP2 | 14.210323 | 0.15593652 | 0.471 | 14.21032348 | 1585.953911 | 14536.7294 | 2.01991167 | 0.488379416 | 0.7958 | 8.077830147 | 517.4066719 | 2535.795645 | 7.035121227 | 0.847271585 | FALSE | FALSE | FALSE |
| ZNF552 | 14.168055 | 0.15624443 | 0.88935 | 136.3646812 | 13597.12854 | 140700.637 | 176.096326 | 0.01534587 | 0.19335 | 176.0963259 | 1746.426933 | 17743.05813 | 0.080456276 | -1.094440073 | FALSE | FALSE | FALSE |
| MAPK15 | 14.125507 | 0.15655242 | 0.99275 | 1120957.576 | 3479089663 | 55652553191 | 0.01242252 | 0.895958073 | 0.99825 | 317.8681387 | 29468.44821 | 293803.6332 | 1137.089073 | 3.055794486 | FALSE | TRUE | FALSE |
| SPATA2 | 14.090418 | 0.15686041 | 0.00015 | 0.416044726 | 0.416044726 | 8.203883907 | 0.42966954 | 0.781232089 | 1 | 4.064076511 | 4.064076511 | 8.486886973 | 32.79361581 | 1.515789304 | FALSE | TRUE | FALSE |
| POLR3G | 14.087669 | 0.15716798 | 0.2497 | 1.502830899 | 14.08766914 | 14.08766914 | 1.86138172 | 0.502706514 | 0.75365 | 1.861381724 | 17.52687304 | 17.52687304 | 7.568393391 | 0.879003698 | FALSE | FALSE | FALSE |
| C1orf174 | 14.001048 | 0.15747621 | 0.2563 | 1.504134015 | 14.00104754 | 14.00104754 | 0.21027347 | 0.849633682 | 1 | 1.918791244 | 1.918791244 | 1.918791244 | 66.58494705 | 1.823376059 | FALSE | TRUE | FALSE |
| PEX1 | 13.972832 | 0.15778434 | 0.2195 | 0.87531917 | 127.8932004 | 1093.11388 | 2.72362305 | 0.43367923 | 0.1771 | 0.170451688 | 10.89136976 | 96.16254966 | 5.130236985 | 0.710137427 | FALSE | FALSE | FALSE |
| EDDM3A | 13.961564 | 0.15809217 | 0.25485 | 1.502495371 | 13.96156384 | 13.96156384 | 1.8574725 | 0.503332298 | 0.75235 | 1.857472498 | 17.35434239 | 17.35434239 | 7.516430986 | 0.876011674 | FALSE | FALSE | FALSE |
| NINL | 13.922171 | 0.15840005 | 0.4021 | 13.9221709 | 10231.0158 | 96568.71555 | 0.53976607 | 0.738450616 | 0.67025 | 0.539766073 | 41.02377631 | 370.6665409 | 25.79297146 | 1.411501378 | FALSE | TRUE | FALSE |
| SBDS | 13.912272 | 0.15870761 | 0.8818 | 1344.97128 | 572567.5275 | 6117289.56 | 92.2736711 | 0.03164519 | 0.46185 | 38.70788153 | 3609.04413 | 9044.96924 | 0.150771848 | -0.821679743 | FALSE | FALSE | FALSE |
| RAX2 | 13.892907 | 0.15901496 | 0.65835 | 55.56980223 | 2126.24374 | 8508.94606 | 0.14478849 | 0.865699857 | 1 | 0.578890538 | 2.315087618 | 2.315087618 | 95.95311639 | 1.982059084 | FALSE | TRUE | FALSE |
| FCN3 | 13.885087 | 0.15932197 | 0.50145 | 13.88508667 | 134.9567785 | 134.9567785 | 0.2160665 | 0.848164948 | 1 | 0.216066501 | 0.216066501 | 0.216066501 | 64.26302383 | 1.807961157 | FALSE | TRUE | FALSE |
| TTC22 | 13.878369 | 0.15962861 | 0.50495 | 13.87836916 | 133.3965384 | 133.3965384 | 1.89638142 | 0.499236122 | 0.7518 | 1.896381418 | 17.86737852 | 17.86737852 | 7.318342727 | 0.864412744 | FALSE | FALSE | FALSE |
| PRKAR2A | 13.866523 | 0.15993495 | 0.50875 | 13.8665234 | 55.49281614 | 222.1336848 | 1.00360825 | 0.588228115 | 0.99675 | 64.27954412 | 1030.277451 | 2091.76681 | 13.81666937 | 1.140403365 | FALSE | FALSE | FALSE |
| SPRED3 | 13.84023 | 0.16024119 | 1 | 26445399516 | 4.71439E+13 | 7.64362E+15 | 17766.8652 | 0.000110823 | 0.99505 | 3004672758 | 6.33722E+13 | 8.02506E+14 | 0.000778991 | -3.108467507 | FALSE | FALSE | FALSE |
| CD70 | 13.790761 | 0.16054761 | 0.00005 | 0.406289809 | 0.406289809 | 0.406289809 | 0.4379079 | 0.778118228 | 1 | 4.124309812 | 4.124309812 | 4.124309812 | 31.49237696 | 1.498205441 | FALSE | TRUE | FALSE |
| ASAP2 | 13.615947 | 0.16085586 | 0.0253 | 1.426535615 | 13.61594697 | 48.13885298 | 0.20361422 | 0.851212275 | 1 | 0.203614218 | 0.203614218 | 3.988849793 | 66.87129765 | 1.825239751 | FALSE | TRUE | FALSE |
| RUNX1T1 | 13.595349 | 0.16116393 | 0.0003 | 0.401713845 | 0.401713845 | 7.924094364 | 0.44732683 | 0.775289747 | 0.0162 | 0.447326832 | 0.447326832 | 8.838406087 | 30.39242862 | 1.482765405 | FALSE | TRUE | FALSE |
| MTDH | 13.591364 | 0.16147159 | 0.00015 | 0.401661751 | 0.401661751 | 7.930555665 | 0.44733196 | 0.77525826 | 1 | 0.447331956 | 0.447331956 | 8.845869088 | 30.38317225 | 1.482633116 | FALSE | TRUE | FALSE |
| ZNF221 | 13.58895 | 0.16177884 | 0.6438 | 127.6906213 | 12115.98993 | 122209.3336 | 0.29693676 | 0.826230105 | 0.9374 | 2.688792668 | 237.7596929 | 237.7596929 | 45.76378518 | 1.660521938 | FALSE | TRUE | FALSE |
| ACOT8 | 13.559268 | 0.16208603 | 0.19805 | 5.82767614 | 129.3033636 | 516.9985285 | 2.42736908 | 0.452646433 | 0.5003 | 2.42736908 | 91.10684935 | 91.10684935 | 5.585993694 | 0.747100441 | FALSE | FALSE | FALSE |
| DPT | 13.515807 | 0.16239335 | 0.3168 | 13.51580731 | 1265.360133 | 1265.360133 | 0.21638853 | 0.848122173 | 1 | 1.979832827 | 1.979832827 | 1.979832827 | 62.46083075 | 1.795607756 | FALSE | TRUE | FALSE |
| PRR23B | 13.447468 | 0.1627011 | 0.00715 | 1.445815831 | 13.4474678 | 13.4474678 | 1.79613949 | 0.508137218 | 0.2565 | 1.796139493 | 16.79527165 | 16.79527165 | 7.486872738 | 0.874300451 | FALSE | FALSE | FALSE |
| DTWD1 | 13.305284 | 0.16301029 | 0.4981 | 1.428136928 | 127.1431744 | 127.1431744 | 0.2134395 | 0.848803854 | 1 | 0.213439503 | 0.213439503 | 0.213439503 | 62.33749613 | 1.794749354 | FALSE | TRUE | FALSE |
| LHFPL5 | 13.285389 | 0.16331929 | 0.0665 | 3.320557177 | 53.16748611 | 53.16748611 | 1.03330708 | 0.584642708 | 0.5046 | 1.033307077 | 1.033307077 | 1.033307077 | 12.85715523 | 1.109144887 | FALSE | FALSE | FALSE |
| ADAM33 | 13.276527 | 0.16362796 | 0.5969 | 13.27652731 | 1044.112334 | 4795.045079 | 0.06791132 | 0.882897146 | 1 | 0.067911319 | 0.067911319 | 1.337818044 | 195.4980035 | 2.291142327 | FALSE | TRUE | FALSE |
| NCCRP1 | 13.178595 | 0.16393747 | 0.2527 | 1.415901472 | 13.17859475 | 13.17859475 | 0.20556343 | 0.850689589 | 1 | 0.205563427 | 0.205563427 | 0.205563427 | 64.10962754 | 1.806923254 | FALSE | TRUE | FALSE |
| OR4A47 | 13.168285 | 0.16424668 | 0.947 | 12532.18661 | 14747595.53 | 133250396.7 | 0.26058303 | 0.836555426 | 1 | 0.260583026 | 0.260583026 | 0.260583026 | 50.53393097 | 1.703583083 | FALSE | TRUE | FALSE |

| | | | | | | | | | | | | | | | | | |
|---|---|---|---|---|---|---|---|---|---|---|---|---|---|---|---|---|---|
| PKP3 | 13.069895 | 0.16455675 | 0.19625 | 1.395921589 | 52.26932722 | 52.26932722 | 0.06032676 | 0.88468258 | 1 | 0.241212984 | 0.241212984 | 2.041160704 | 216.6517144 | 2.33576213 | FALSE | TRUE | FALSE |
| BRSK1 | 13.003691 | 0.16486726 | 0.9696 | 138608.1666 | 16447442.98 | 185354194 | 0.23558778 | 0.843133935 | 1 | 21.34859956 | 211.1640776 | 419.7189917 | 55.19679737 | 1.74191388 | FALSE | TRUE | FALSE |
| FAM83H | 12.847071 | 0.16517944 | 0.945 | 12598.62204 | 16544098.82 | 184523912.4 | 2008058.74 | 7.85381E-07 | 0.0946 | 16613.4908 | 2008058.74 | 21998332.01 | 6.39776E-06 | -5.193972293 | FALSE | FALSE | FALSE |
| APCDD1L | 12.826433 | 0.16549144 | 0.50585 | 12.82643328 | 123.0585168 | 123.0585168 | 1.77124832 | 0.510886689 | 0.7551 | 1.771248325 | 16.66104679 | 35.0245302 | 7.241465299 | 0.859826454 | FALSE | FALSE | FALSE |
| SIAE | 12.795487 | 0.16580341 | 0.6117 | 27.61355506 | 1770.302838 | 7088.799728 | 0.508974 | 0.752064452 | 0.11985 | 0.127276867 | 2.035870764 | 8.145415893 | 25.13976482 | 1.400361211 | FALSE | TRUE | FALSE |
| LRFN4 | 12.790639 | 0.16611499 | 0.6309 | 12.790639 | 490.8483376 | 490.8483376 | 0.41347968 | 0.787055775 | 1 | 1.653861462 | 6.616870679 | 32.66774377 | 30.9341419 | 1.490438073 | FALSE | TRUE | FALSE |
| RPS6KB2 | 12.681015 | 0.16642762 | 0.50825 | 50.70351775 | 4722.835856 | 11814.0013 | 0.12631028 | 0.869805085 | 0.7533 | 0.282618674 | 1.130070812 | 2.497031437 | 100.395748 | 2.00171532 | FALSE | TRUE | FALSE |
| PRSS53 | 12.674415 | 0.16673988 | 0.1991 | 5.367634134 | 122.6379045 | 490.262634 | 0.26326547 | 0.835744612 | 0.50515 | 0.263265469 | 0.263265469 | 0.576118431 | 48.14309566 | 1.682534012 | FALSE | TRUE | FALSE |
| EXO5 | 12.595692 | 0.16705278 | 0.1334 | 12.59569239 | 120.1528686 | 120.1528686 | 0.19776793 | 0.852624794 | 1 | 0.197767935 | 1.804875065 | 1.804875065 | 63.68925481 | 1.804066167 | FALSE | TRUE | FALSE |
| OGDHL | 12.593358 | 0.16736525 | 0.00525 | 0.04045856 | 0.788438474 | 5.460799835 | 0.10458538 | 0.874828878 | 0.8513 | 2.170072235 | 138.5738803 | 1171.638633 | 120.4122243 | 2.080670579 | FALSE | TRUE | FALSE |
| REM1 | 12.551408 | 0.16767785 | 0.0067 | 0.783994927 | 12.55140825 | 12.55140825 | 1.12557886 | 0.57531874 | 0.5056 | 1.125578856 | 4.503286732 | 4.503286732 | 11.15106969 | 1.04731653 | FALSE | FALSE | FALSE |
| AMPD1 | 12.532654 | 0.16799026 | 0.23245 | 1.453860763 | 61.75504868 | 802.5069218 | 0.01752626 | 0.894419136 | 0.88755 | 1.119641404 | 17.9148955 | 88.39221685 | 715.0786414 | 2.854353806 | FALSE | TRUE | FALSE |
| MRPS15 | 12.440636 | 0.16830348 | 0.2576 | 1.330351106 | 12.44063605 | 12.44063605 | 0.17705526 | 0.857813421 | 1 | 15.20232552 | 146.3753637 | 146.3753637 | 70.26414148 | 1.846733744 | FALSE | TRUE | FALSE |
| CDCA5 | 12.438607 | 0.16861629 | 0.5062 | 12.43860717 | 121.7944808 | 121.7944808 | 0.17108612 | 0.859330258 | 1 | 0.171086117 | 0.171086117 | 0.171086117 | 72.70377846 | 1.861556982 | FALSE | TRUE | FALSE |
| CERKL | 12.417575 | 0.16892893 | 0.133 | 1.374237166 | 12.41757544 | 12.41757544 | 20.1837867 | 0.1319948 | 0.00645 | 0.565331095 | 20.18378675 | 20.18378675 | 0.615225259 | -0.210965842 | FALSE | FALSE | FALSE |
| DCAF4 | 12.348394 | 0.16924209 | 0.1254 | 3.085450322 | 49.4323042 | 197.933678 | 155.212089 | 0.017578206 | 0.02205 | 9.692256828 | 155.2120892 | 621.3532744 | 0.079558195 | -1.099315078 | FALSE | FALSE | FALSE |
| KRT74 | 12.220804 | 0.16955657 | 0.06565 | 0.545840542 | 12.22080386 | 48.82368952 | 0.00405722 | 0.898309207 | 0.0157 | 0.004057222 | 0.004057222 | 0.080234458 | 3012.110974 | 3.478870968 | FALSE | TRUE | FALSE |
| GUF1 | 12.124481 | 0.16987194 | 0.85915 | 457.0951193 | 174419.4078 | 1763078.226 | 21567.142 | 9.50438E-05 | 0.608 | 21567.14197 | 24398746.43 | 265366120.7 | 0.000562174 | -3.250129441 | FALSE | FALSE | FALSE |
| USP42 | 12.084926 | 0.17018742 | 0.26965 | 1.28207031 | 12.08492631 | 43.23466693 | 0.18688989 | 0.855270458 | 1 | 0.18688989 | 0.18688989 | 3.653768138 | 64.6633496 | 1.810658198 | FALSE | TRUE | FALSE |
| CRMP1 | 12.060282 | 0.1705028 | 0.073 | 0.754032394 | 12.06028238 | 14.86203671 | 4.38048304 | 0.336145935 | 0.14075 | 1.095342489 | 4.380483041 | 5.409184136 | 2.753185498 | 0.439835473 | FALSE | FALSE | FALSE |
| DNAJB9 | 12.016569 | 0.17081834 | 0.00385 | 1.416405485 | 1.416405485 | 1.416405485 | 0.40458185 | 0.790735698 | 1 | 0.40458185 | 0.40458185 | 0.40458185 | 29.70120719 | 1.472774101 | FALSE | TRUE | FALSE |
| OR6S1 | 11.996914 | 0.17113371 | 0.50065 | 11.99691434 | 114.342084 | 114.342084 | 1.62522112 | 0.523062256 | 0.87735 | 15.1844149 | 145.5107391 | 145.5107391 | 7.381712077 | 0.868157102 | FALSE | FALSE | FALSE |
| ZNF683 | 11.985748 | 0.17144878 | 0.13595 | 5.52392124 | 51.69789515 | 51.69789515 | 0.88223303 | 0.601103717 | 0.50215 | 0.882233027 | 0.882233027 | 1.949107099 | 13.58569438 | 1.133081841 | FALSE | TRUE | FALSE |
| POU2F3 | 11.969499 | 0.17176364 | 0.19495 | 5.054835164 | 116.1354026 | 464.2347538 | 0.95560934 | 0.59219817 | 0.5025 | 0.955609345 | 8.889219632 | 8.889219632 | 12.52551438 | 1.09779557 | FALSE | TRUE | FALSE |
| USP31 | 11.95593 | 0.17207824 | 0.5141 | 11.95593017 | 115.6969591 | 232.7800121 | 0.17925171 | 0.857207926 | 1 | 1.655443891 | 15.69124647 | 55.82593675 | 66.69911397 | 1.824120065 | FALSE | TRUE | FALSE |
| SLC10A1 | 11.950888 | 0.17239246 | 0.9738 | 33241.81617 | 28460329.84 | 715224415.3 | 0.01182887 | 0.896155456 | 0.9998 | 2227.004549 | 1890944.943 | 18727253.62 | 1010.315182 | 3.004456879 | FALSE | TRUE | FALSE |
| DPYS | 11.907246 | 0.17270685 | 0.72815 | 190.3472425 | 65334.92742 | 627258.4201 | 0.11768753 | 0.871807032 | 0.65545 | 0.470441885 | 16.64855436 | 66.58387287 | 101.1767842 | 2.005080872 | FALSE | TRUE | FALSE |
| VPS37A | 11.901993 | 0.17302087 | 0.68575 | 11.90199267 | 1118.500285 | 1118.500285 | 0.40538284 | 0.790354598 | 1 | 0.405382845 | 0.405382845 | 0.405382845 | 29.35988243 | 1.467754312 | FALSE | TRUE | FALSE |
| ZNF410 | 11.892299 | 0.17333459 | 0.00025 | 0.351013318 | 0.351013318 | 6.960794095 | 3.58123707 | 0.379451834 | 0.51245 | 3.581237075 | 3.581237075 | 7.635447167 | 3.320723611 | 0.52123273 | FALSE | FALSE | FALSE |
| OR10A6 | 11.887255 | 0.17364792 | 0.5076 | 11.88725513 | 113.0278716 | 113.0278716 | 0.17772252 | 0.857536522 | 1 | 15.05143823 | 143.9318238 | 143.9318238 | 66.88660011 | 1.825339121 | FALSE | TRUE | FALSE |
| FNDC9 | 11.854753 | 0.17396128 | 0.25535 | 1.280931689 | 11.85475269 | 11.85475269 | 0.1888262 | 0.854709424 | 1 | 0.188826198 | 0.188826198 | 0.188826198 | 62.78129202 | 1.797830249 | FALSE | TRUE | FALSE |
| PCDH20 | 11.850219 | 0.17427425 | 0.2589 | 1.251051086 | 11.85021911 | 42.25816845 | 0.18233281 | 0.856367292 | 0.02045 | 0.182332811 | 0.182332811 | 3.590247712 | 64.99224721 | 1.812861554 | FALSE | TRUE | FALSE |
| RGS14 | 11.844216 | 0.17458686 | 0.5115 | 11.84421569 | 113.2073157 | 113.2073157 | 0.39143498 | 0.795148533 | 1 | 0.391434982 | 0.391434982 | 7.725750642 | 30.25844965 | 1.480846672 | FALSE | TRUE | FALSE |
| CEACAM1 | 11.82625 | 0.17489929 | 0.81525 | 112.9794657 | 1106.356465 | 11098.78101 | 15.0037419 | 0.158553036 | 0.8197 | 144.0786617 | 1418.204375 | 14300.74268 | 0.788220056 | -0.103352519 | FALSE | FALSE | FALSE |
| VSIG2 | 11.808153 | 0.17521153 | 0.3167 | 2.951096062 | 47.25950146 | 47.25950146 | 2.02325208 | 0.488048887 | 0.50575 | 8.091808867 | 76.87461226 | 76.87461226 | 5.83622454 | 0.766131992 | FALSE | FALSE | FALSE |
| DHX38 | 11.807144 | 0.17552334 | 0.0789 | 0.737821806 | 11.80714363 | 57.20666119 | 0.03450097 | 0.890248527 | 1 | 0.034500969 | 0.034500969 | 1.162234824 | 342.2264358 | 2.534313554 | FALSE | TRUE | FALSE |
| ZNF169 | 11.796755 | 0.17583486 | 0.1936 | 1.294145926 | 110.2306356 | 1055.22241 | 0.03096316 | 0.891021797 | 1 | 0.030963156 | 0.030963156 | 0.612678486 | 380.9933069 | 2.580917346 | FALSE | TRUE | FALSE |
| TULP1 | 11.762527 | 0.17614643 | 0.15815 | 1.278903367 | 11.7625269 | 47.04809325 | 8.27201006 | 0.223236881 | 0.04525 | 2.067905519 | 8.272010058 | 33.09785852 | 1.42196719 | 0.152889576 | FALSE | FALSE | FALSE |
| TDRP | 11.710804 | 0.17645829 | 0.49755 | 1.266971931 | 111.0258374 | 111.0258374 | 0.19288396 | 0.853654232 | 0.0048 | 0.192883961 | 0.192883961 | 0.192883961 | 60.71424307 | 1.783290585 | FALSE | TRUE | FALSE |
| GABRR1 | 11.683743 | 0.17677009 | 0.02255 | 0.329990169 | 11.68374333 | 46.7419684 | 0.63419665 | 0.697741229 | 0.0714 | 0.158588183 | 2.536796116 | 2.536796116 | 18.42290307 | 1.265358067 | FALSE | TRUE | FALSE |
| BPIFC | 11.661623 | 0.17708177 | 0.5118 | 111.0507291 | 1083.99735 | 10839.79156 | 0.19687205 | 0.852853242 | 1 | 1.797113678 | 1.797113678 | 3.905718711 | 59.23452743 | 1.772574928 | FALSE | TRUE | FALSE |
| PLEKHB2 | 11.624038 | 0.17739355 | 0.6875 | 11.62403752 | 1093.743857 | 1093.743857 | 13.8625498 | 0.165167202 | 0.89155 | 133.2262743 | 13244.67711 | 136848.9135 | 0.838520885 | -0.076486116 | FALSE | FALSE | FALSE |
| ANKRD26 | 11.6177 | 0.17770498 | 0.187 | 1.412944079 | 158.9843036 | 636.776037 | 1.68971899 | 0.518081088 | 0.50455 | 1.68971899 | 58.03961704 | 283.1830247 | 6.875521671 | 0.837305655 | FALSE | FALSE | FALSE |
| PAXIP1 | 11.616761 | 0.17801598 | 0.89545 | 114.0540665 | 11833.12542 | 125463.6831 | 0.19976416 | 0.852152307 | 1 | 0.199764161 | 0.199764161 | 3.937968797 | 58.15238031 | 1.764567496 | FALSE | TRUE | FALSE |
| CERS4 | 11.594875 | 0.17832686 | 0.3405 | 2.900320844 | 185.4520803 | 1687.883678 | 0.04495086 | 0.887868362 | 0.26105 | 0.011242738 | 0.044950861 | 0.055791067 | 257.9455631 | 2.411528062 | FALSE | TRUE | FALSE |
| LMCD1 | 11.578424 | 0.17863754 | 0.0391 | 2.894535332 | 11.5784244 | 46.32640007 | 0.26099132 | 0.836390602 | 0.96685 | 4.173649693 | 16.69643432 | 66.80976622 | 44.36325462 | 1.6470234 | FALSE | TRUE | FALSE |
| ZNF35 | 11.453544 | 0.17894957 | 0.51025 | 11.45354437 | 438.1373901 | 438.1373901 | 0.18893534 | 0.854657016 | 0.01155 | 0.188935342 | 0.188935342 | 3.749038493 | 60.62150272 | 1.782626698 | FALSE | TRUE | FALSE |
| GPRC5C | 11.394986 | 0.179262 | 0.12025 | 2.848021964 | 45.60292816 | 182.5494208 | 1.02591416 | 0.585881377 | 0.8899 | 4.103914836 | 65.72013692 | 263.0941661 | 11.1071533 | 1.045602766 | FALSE | FALSE | FALSE |
| CUL5 | 11.317172 | 0.17957512 | 0.00765 | 0.335272806 | 0.335272806 | 6.615759298 | 0.38163773 | 0.798573164 | 1 | 0.381637732 | 0.381637732 | 7.542775892 | 29.65422601 | 1.472086593 | FALSE | TRUE | FALSE |
| MYBPH | 11.281785 | 0.17988831 | 0.3175 | 1.228047671 | 106.2424441 | 106.2424441 | 2.01383595 | 0.488709565 | 0.69245 | 2.013835951 | 177.5502402 | 177.5502402 | 5.602136984 | 0.748353724 | FALSE | FALSE | FALSE |
| CRLF1 | 11.270579 | 0.18020123 | 0.12675 | 0.704757135 | 11.27057907 | 11.27057907 | 0.10928793 | 0.873601948 | 1 | 0.109287929 | 0.109287929 | 2.164623624 | 103.1273922 | 2.013374036 | FALSE | TRUE | FALSE |
| CSGALNACT | 11.240507 | 0.18051414 | 0.6929 | 11.24050741 | 1060.016097 | 1060.016097 | 0.18298462 | 0.856239078 | 0.5047 | 1.679639877 | 1.679639877 | 3.620793222 | 61.42870017 | 1.788371326 | FALSE | TRUE | FALSE |
| OR8K3 | 11.225845 | 0.18082683 | 0.6909 | 11.22584493 | 1044.747625 | 1044.747625 | 1217.26898 | 0.002092898 | 0.4982 | 124.2791524 | 125476.9592 | 125476.9592 | 0.009222156 | -2.035167516 | FALSE | FALSE | FALSE |
| PGAP1 | 11.148986 | 0.18114021 | 0.2002 | 2.786052994 | 11.14898576 | 54.85965585 | 3.89641965 | 0.360835358 | 0.6642 | 8.858796793 | 141.8375196 | 567.7564203 | 2.861341118 | 0.456569636 | FALSE | FALSE | FALSE |
| STRC | 11.145372 | 0.1814532 | 0.5168 | 11.14537238 | 855.680871 | 3844.106778 | 0.03420843 | 0.890310748 | 0.5212 | 0.308182561 | 0.308182561 | 5.978919561 | 325.8077997 | 2.512961477 | FALSE | TRUE | FALSE |
| KCNT1 | 11.128537 | 0.18176599 | 0.5486 | 11.12853672 | 716.8076918 | 2874.798205 | 0.01581206 | 0.894910358 | 0.9978 | 4.987355055 | 319.2822664 | 1277.889326 | 703.8005779 | 2.847449619 | FALSE | TRUE | FALSE |
| TTC9C | 11.102935 | 0.18207873 | 0.25345 | 1.171566543 | 11.10293537 | 11.10293537 | 0.16673392 | 0.860514224 | 1 | 1.545972553 | 1.545972553 | 1.545972553 | 66.59074175 | 1.823413852 | FALSE | TRUE | FALSE |
| HAGHL | 11.096462 | 0.18239113 | 0.79295 | 44.37450175 | 4107.770666 | 40992.29721 | 0.05771001 | 0.885106488 | 0.49905 | 0.057710007 | 0.525845532 | 0.525845532 | 192.2796867 | 2.283933406 | FALSE | TRUE | FALSE |
| FCGR1A | 11.051165 | 0.18270375 | 0.00765 | 1.15924956 | 11.05116517 | 11.05116517 | 0.14952969 | 0.86467857 | 1 | 0.149529685 | 0.149529685 | 0.149529685 | 73.90616223 | 1.868680651 | FALSE | TRUE | FALSE |
| DHCR24 | 10.967727 | 0.18301716 | 0.35665 | 2.74203941 | 43.88014207 | 175.6013637 | 0.06662476 | 0.883211281 | 1 | 0.066624758 | 0.066624758 | 1.316613058 | 164.619384 | 2.216480972 | FALSE | TRUE | FALSE |
| NBPF3 | 10.908582 | 0.183331 | 0.50725 | 10.90858212 | 104.0315568 | 104.0315568 | 1.40397901 | 0.545037983 | 0.96765 | 125.3934999 | 1230.207466 | 12364.12741 | 7.769761554 | 0.890407691 | FALSE | FALSE | FALSE |
| SLC12A4 | 10.895446 | 0.18364461 | 0.90955 | 696.9364975 | 219941.9054 | 2870851.034 | 50.7444153 | 0.054041505 | 0.65895 | 451.8002285 | 115817.767 | 1857114.929 | 0.214712218 | -0.668143241 | FALSE | FALSE | FALSE |
| KRT84 | 10.799472 | 0.1839592 | 0.7947 | 43.20553949 | 2770.770085 | 11096.11246 | 85.9364473 | 0.034033688 | 0.5115 | 158.9811534 | 5513.616725 | 5513.616725 | 0.125668119 | -0.900774885 | FALSE | FALSE | FALSE |
| BMS1 | 10.749019 | 0.1842741 | 0.66795 | 103.8488337 | 10433.36893 | 108370.2226 | 0.18507358 | 0.85572402 | 1 | 1.712862872 | 1.712862872 | 33.51328244 | 58.07970703 | 1.764024417 | FALSE | TRUE | FALSE |
| GPR182 | 10.709764 | 0.18458915 | 0.68485 | 42.83056188 | 1023.16353 | 4091.842808 | 0.18108535 | 0.856814273 | 1 | 0.181085352 | 0.181085352 | 3.591733463 | 59.14207518 | 1.771896559 | FALSE | TRUE | FALSE |
| ALDH1A3 | 10.676393 | 0.18490426 | 0.94645 | 4182.423042 | 1805133.755 | 19073858.86 | 1435.45842 | 0.001651297 | 0.5044 | 1435.458416 | 154344.8232 | 154344.8232 | 0.007437619 | -2.128566059 | FALSE | FALSE | FALSE |
| TBC1D7 | 10.670983 | 0.18521901 | 0.2596 | 2.667103862 | 10.67098294 | 10.67098294 | 0.48534888 | 0.761027544 | 1 | 0.485348885 | 0.485348885 | 0.485348885 | 21.98621091 | 1.34215039 | FALSE | TRUE | FALSE |
| LDHB | 10.667552 | 0.18553339 | 0.46935 | 6.343475653 | 101.4154495 | 405.6536198 | 0.42721228 | 0.78203342 | 0.50445 | 0.427212277 | 0.427212277 | 0.427212277 | 24.97014418 | 1.39742105 | FALSE | TRUE | FALSE |
| CPEB2 | 10.666035 | 0.18584735 | 0.97085 | 9056.404042 | 912266.4166 | 9466504.746 | 0.23065762 | 0.844682945 | 0.9978 | 1687.669426 | 166716.4786 | 1714565.06 | 46.2418515 | 1.665035215 | FALSE | TRUE | FALSE |
| PPT2 | 10.640926 | 0.18616125 | 0.0051 | 0.536020653 | 0.536020653 | 2.661327279 | 0.17188883 | 0.859182442 | 1 | 0.171888833 | 0.171888833 | 0.171888833 | 61.9058588 | 1.791731753 | FALSE | TRUE | FALSE |
| PARL | 10.625175 | 0.18647495 | 0.69055 | 10.62517468 | 1004.008548 | 1004.008548 | 1.59713649 | 0.526382491 | 0.5019 | 1.597136491 | 1.597136491 | 1.597136491 | 6.652640362 | 0.822994046 | FALSE | FALSE | FALSE |
| TNFSF10 | 10.583235 | 0.18678885 | 0.6868 | 10.58323464 | 1029.314335 | 1029.314335 | 0.17483684 | 0.858351615 | 0.0055 | 0.174836836 | 0.174836836 | 0.174836836 | 60.53206453 | 1.781985486 | FALSE | TRUE | FALSE |
| SUMF2 | 10.544602 | 0.1871029 | 0.72335 | 54.79902999 | 3509.015404 | 33251.3617 | 12809.5364 | 0.000163909 | 0.3409 | 4971.362195 | 1246654.312 | 4989198.255 | 0.000823184 | -3.084503227 | FALSE | FALSE | FALSE |

| | | | | | | | | | | | | | | | | | |
|---|---|---|---|---|---|---|---|---|---|---|---|---|---|---|---|---|---|
| ZFR | 10.518432 | 0.1874169 | 0.51195 | 11.72974721 | 112.9872794 | 394.8754258 | 0.38617193 | 0.797207391 | 1 | 0.386171931 | 0.386171931 | 7.588846789 | 27.23769081 | 1.435170286 | FALSE | TRUE | FALSE |
| STX10 | 10.506115 | 0.18773066 | 0.50085 | 22.39888008 | 982.8466819 | 9859.703057 | 125.570093 | 0.022706851 | 0.3543 | 125.5700928 | 12441.52541 | 128334.2228 | 0.083667338 | -1.077444048 | FALSE | FALSE | FALSE |
| GALT | 10.503152 | 0.18804403 | 0.61095 | 42.02479432 | 2695.961474 | 10823.16955 | 342.212995 | 0.007176663 | 0.14085 | 85.49459922 | 2898.367964 | 21979.61451 | 0.030691857 | -1.512976832 | FALSE | FALSE | FALSE |
| CATSPERG | 10.49223 | 0.18835714 | 0.64315 | 10.49222977 | 858.404407 | 8040.516605 | 0.32278018 | 0.817821157 | 0.3274 | 0.32278018 | 24.97960109 | 24.97960109 | 32.5058056 | 1.511960934 | FALSE | TRUE | FALSE |
| PKLR | 10.46745 | 0.18867018 | 0.9367 | 1491.597073 | 383182.5766 | 2768690.46 | 2.03582636 | 0.486223319 | 0.78835 | 8.141361738 | 521.4543574 | 2566.860872 | 5.141621964 | 0.711100142 | FALSE | FALSE | FALSE |
| KCNE3 | 10.467428 | 0.1889828 | 0.25355 | 2.617749144 | 10.4674283 | 10.4674283 | 1.33112148 | 0.552131534 | 0.4967 | 0.332880436 | 1.331121482 | 1.331121482 | 7.863616088 | 0.895622303 | FALSE | FALSE | FALSE |
| KRTAP10-8 | 10.447874 | 0.18929528 | 0.89755 | 98.88367432 | 9534.542513 | 97022.76682 | 14.303308 | 0.161518957 | 0.31145 | 14.303308 | 1334.420635 | 1334.420635 | 0.73045156 | -0.136408579 | FALSE | FALSE | FALSE |
| ZAN | 10.436379 | 0.18960751 | 0.98245 | 1047526.131 | 1373649800 | 11494375986 | 168.310753 | 0.016110166 | 0.9831 | 1826865.648 | 1935016644 | 22058682367 | 0.06200661 | -1.207562012 | FALSE | FALSE | FALSE |
| ITIH3 | 10.407015 | 0.18991975 | 0.1375 | 1.125851202 | 10.40701507 | 22.07724475 | 1.90717873 | 0.498441386 | 0.1405 | 0.838120981 | 7.626497901 | 16.47368948 | 5.456759208 | 0.73693479 | FALSE | FALSE | FALSE |
| CLIC3 | 10.399212 | 0.19023168 | 0.6173 | 10.39921156 | 666.4269711 | 826.5995077 | 1.58617412 | 0.527954193 | 0.9364 | 25.39641021 | 407.0312222 | 504.9746194 | 6.556160157 | 0.816649554 | FALSE | FALSE | FALSE |
| TGDS | 10.346791 | 0.19054398 | 0.8303 | 41.4047038 | 2657.249401 | 10644.12523 | 3.94208279 | 0.358594254 | 0.34525 | 3.942082786 | 63.12147722 | 252.6769075 | 2.624701577 | 0.419079932 | FALSE | FALSE | FALSE |
| C8orf74 | 10.323596 | 0.1908562 | 0.9714 | 105479.4233 | 130871689.2 | 1777052199 | 1399.9541 | 0.001748672 | 0.62 | 14359.25133 | 17754471.86 | 256427970.8 | 0.007374239 | -2.132282778 | FALSE | FALSE | FALSE |
| KANSL1L | 10.318107 | 0.19116807 | 0.5113 | 98.81163213 | 969.9699592 | 9754.211889 | 1.51076797 | 0.534425181 | 0.88105 | 14.20025604 | 136.9001119 | 280.1415284 | 6.829709772 | 0.834402249 | FALSE | FALSE | FALSE |
| ARF5 | 10.290594 | 0.19147993 | 0.0013 | 0.516557956 | 0.516557956 | 0.516557956 | 0.55859714 | 0.729437878 | 1 | 0.558597136 | 0.558597136 | 0.558597136 | 18.42221106 | 1.265341754 | FALSE | TRUE | FALSE |
| GM2A | 10.281069 | 0.19179152 | 0.25525 | 2.570679056 | 10.28106918 | 10.28106918 | 0.33499597 | 0.813952535 | 1 | 0.334995974 | 0.334995974 | 0.334995974 | 30.69012761 | 1.486998694 | FALSE | TRUE | FALSE |
| BRD1 | 10.259 | 0.19210301 | 0.03245 | 1.075569263 | 10.25900016 | 198.9132466 | 0.15878535 | 0.862367189 | 1 | 0.158785352 | 0.158785352 | 3.090051366 | 64.60923517 | 1.8102946 | FALSE | TRUE | FALSE |
| SLC35G3 | 10.247561 | 0.19241425 | 0.25125 | 1.109509007 | 10.24756125 | 10.24756125 | 1.46472067 | 0.538696615 | 0.0087 | 1.464720673 | 1.464720673 | 1.464720673 | 6.996256313 | 0.844865712 | FALSE | FALSE | FALSE |
| ULBP3 | 10.201185 | 0.19272577 | 0.253 | 1.106638091 | 10.20118506 | 10.20118506 | 0.16636664 | 0.860635185 | 1 | 0.166366635 | 0.166366635 | 0.166366635 | 61.31749345 | 1.787584393 | FALSE | TRUE | FALSE |
| SH3RF2 | 10.073511 | 0.1930388 | 0.1354 | 1.426106009 | 22.80853865 | 91.25004771 | 0.12178659 | 0.870849934 | 0.7597 | 0.121786592 | 0.487129995 | 2.398771346 | 82.71445347 | 1.917581405 | FALSE | TRUE | FALSE |
| GPR152 | 10.068554 | 0.19335149 | 0.5059 | 40.27275915 | 936.8871423 | 3747.413008 | 1.45366094 | 0.540195745 | 0.87285 | 1.453660937 | 130.3012151 | 130.3012151 | 6.926342824 | 0.840503983 | FALSE | FALSE | FALSE |
| EDN2 | 10.054576 | 0.19366397 | 0.4997 | 5.366066512 | 96.27700022 | 96.27700022 | 0.3243785 | 0.817349116 | 1 | 3.035395717 | 12.13962097 | 12.13962097 | 30.99643226 | 1.491311709 | FALSE | TRUE | FALSE |
| GPD1 | 10.040184 | 0.19397624 | 0.62865 | 40.17735207 | 2578.403032 | 10328.1867 | 230.009824 | 0.011143328 | 0.61705 | 230.0098241 | 14774.79031 | 64847.15363 | 0.04365111 | -1.360004704 | FALSE | FALSE | FALSE |
| MFSD6L | 10.028481 | 0.19428827 | 0.07535 | 1.069539764 | 96.38816331 | 96.38816331 | 0.02449265 | 0.892629118 | 1 | 0.024492651 | 0.024492651 | 0.48418716 | 409.4485938 | 2.612199384 | FALSE | TRUE | FALSE |
| WBP2NL | 9.9896972 | 0.19460047 | 0.0065 | 1.077691861 | 9.989697157 | 9.989697157 | 0.15080208 | 0.864438158 | 0.87765 | 5.478814459 | 51.07720921 | 51.07720921 | 66.24376386 | 1.821145 | FALSE | TRUE | FALSE |
| RPE | 9.9773426 | 0.19491243 | 0.1296 | 4.264875238 | 39.91064296 | 39.91064296 | 0.31787972 | 0.819343312 | 1 | 5.085352332 | 20.34758977 | 20.34758977 | 31.38716274 | 1.496752059 | FALSE | TRUE | FALSE |
| ENTPD8 | 9.9730473 | 0.19522404 | 0.50935 | 9.973047343 | 94.89230003 | 94.89230003 | 111.839212 | 0.02599276 | 0.664 | 111.8392121 | 10963.70864 | 112490.8205 | 0.089173083 | -1.049766218 | FALSE | FALSE | FALSE |
| ZBTB48 | 9.9720265 | 0.19553524 | 0.261 | 1.068908372 | 9.972026494 | 21.03052 | 0.16026422 | 0.861964734 | 1 | 0.160264223 | 0.160264223 | 3.159333373 | 62.2224118 | 1.793946841 | FALSE | TRUE | FALSE |
| ACADSB | 9.9513753 | 0.19584634 | 0.3035 | 2.49043661 | 159.0098068 | 565.9819917 | 271.475315 | 0.009240096 | 0.4144 | 113.7099568 | 9040.521076 | 69557.74404 | 0.036656649 | -1.435847241 | FALSE | FALSE | FALSE |
| SORD | 9.9293555 | 0.19615736 | 0.1284 | 1.115069328 | 9.929355477 | 9.929355477 | 0.09964973 | 0.876014125 | 1 | 0.099649729 | 0.099649729 | 0.099649729 | 99.64257359 | 1.998444936 | FALSE | TRUE | FALSE |
| OR9A4 | 9.9141978 | 0.19646819 | 0.5031 | 9.91419784 | 94.01989325 | 94.01989325 | 0.16155421 | 0.861655457 | 1 | 1.466683256 | 1.466683256 | 1.466683256 | 61.36762311 | 1.787939303 | FALSE | TRUE | FALSE |
| CHST14 | 9.9108642 | 0.19677865 | 0.49755 | 2.478244292 | 95.48933191 | 95.48933191 | 5.14849766 | 0.301593804 | 0.135 | 5.148497663 | 20.60026117 | 20.60026117 | 1.925001212 | 0.284431007 | FALSE | FALSE | FALSE |
| SUCLG1 | 9.8808401 | 0.19708915 | 0.2685 | 2.471628585 | 39.51054693 | 158.0304192 | 420.539698 | 0.005642998 | 0.3892 | 105.0797045 | 6740.754043 | 26997.38829 | 0.023495618 | -1.629013127 | FALSE | FALSE | FALSE |
| SLC9C1 | 9.7679577 | 0.19740097 | 0.044 | 0.490827682 | 9.767957734 | 39.06207582 | 0.55572007 | 0.73064067 | 0.26485 | 0.062330265 | 0.555720065 | 2.104206031 | 17.57711904 | 1.244947694 | FALSE | TRUE | FALSE |
| ACOX1 | 9.7562916 | 0.19771256 | 0.3838 | 4.218055319 | 39.02653325 | 156.1506095 | 5.49434787 | 0.287054463 | 0.8668 | 204.0208289 | 6907.696935 | 13089.5163 | 1.77569601 | 0.249368619 | FALSE | FALSE | FALSE |
| IDE | 9.7224832 | 0.19802424 | 0.66585 | 93.38552852 | 9273.608868 | 95765.8849 | 114.150799 | 0.025669616 | 0.8597 | 1127.027987 | 118042.5366 | 1250850.367 | 0.085172275 | -1.069701752 | FALSE | FALSE | FALSE |
| CARD6 | 9.7082008 | 0.19833573 | 0.3598 | 9.708200813 | 861.2843102 | 8407.348191 | 1336.45024 | 0.00179975 | 0.5035 | 1336.450244 | 132458.613 | 1364401.448 | 0.007264169 | -2.138814044 | FALSE | FALSE | FALSE |
| TIGD6 | 9.6809548 | 0.19864723 | 0.51035 | 9.68095485 | 91.7963234 | 91.7963234 | 0.1534766 | 0.863688875 | 1 | 1.391961617 | 12.95718406 | 12.95718406 | 63.07772511 | 1.799876022 | FALSE | TRUE | FALSE |
| LGALS3BP | 9.6718141 | 0.19895844 | 0.32295 | 9.671814062 | 913.9738486 | 913.9738486 | 1.38053512 | 0.547293941 | 0.8746 | 12.99482571 | 125.4591056 | 125.4591056 | 7.005844261 | 0.845460479 | FALSE | FALSE | FALSE |
| FIGLA | 9.6543898 | 0.19926951 | 0.1323 | 2.413336381 | 9.654389787 | 9.654389787 | 0.76264495 | 0.635198363 | 0.7513 | 0.762644947 | 3.050299614 | 3.050299614 | 12.65908838 | 1.102402432 | FALSE | TRUE | FALSE |
| DPM1 | 9.6492135 | 0.19958025 | 0.25185 | 1.018398765 | 9.649213491 | 9.649213491 | 0.15202138 | 0.864093328 | 1 | 0.152021376 | 0.152021376 | 0.152021376 | 63.47274145 | 1.802587256 | FALSE | TRUE | FALSE |
| DUSP19 | 9.6453429 | 0.19989062 | 0.0034 | 2.411741417 | 9.645342853 | 9.645342853 | 1.25335122 | 0.560911697 | 0.50045 | 1.253351223 | 1.253351223 | 1.253351223 | 7.695642428 | 0.88624488 | FALSE | FALSE | FALSE |
| PARP15 | 9.618083 | 0.20020101 | 0.3177 | 1.075544559 | 88.16888867 | 88.16888867 | 1.82528716 | 0.50520139 | 0.5087 | 1.825287157 | 16.53925271 | 16.53925271 | 5.269353329 | 0.721757321 | FALSE | FALSE | FALSE |
| ZNF548 | 9.6069532 | 0.20051115 | 0.50335 | 9.606953153 | 91.7822221 | 91.7822221 | 1.37664762 | 0.547953356 | 0.7533 | 1.376647623 | 12.90073638 | 12.90073638 | 6.978512868 | 0.843762884 | FALSE | FALSE | FALSE |
| SMTNL1 | 9.5775389 | 0.20082134 | 0.8754 | 223.2874673 | 35744.14769 | 142971.621 | 0.04739907 | 0.887242316 | 1 | 4.049075631 | 382.8233947 | 382.8233947 | 202.0617618 | 2.305484135 | FALSE | TRUE | FALSE |
| PRAMEF1 | 9.5733505 | 0.20113117 | 1 | 1.9321E+11 | 5.5826E+16 | 7.30336E+17 | 0.20636792 | 0.85055146 | 1 | 1858628.561 | 2464403912 | 28371180881 | 46.38972328 | 1.666421782 | FALSE | TRUE | FALSE |
| MYNN | 9.5356332 | 0.20144118 | 0.3834 | 4.112254216 | 38.14975106 | 38.14975106 | 0.31440623 | 0.82056255 | 1 | 1.257562236 | 1.257562236 | 6.228536666 | 30.32902113 | 1.481858394 | FALSE | TRUE | FALSE |
| PNPLA5 | 9.4855261 | 0.20175156 | 0.8948 | 91.81457919 | 9259.067523 | 96353.58451 | 0.1536757 | 0.8636541 | 0.93765 | 13.50697251 | 526.1862977 | 526.1862977 | 61.72430685 | 1.790456222 | FALSE | TRUE | FALSE |
| RGS4 | 9.480181 | 0.2020616 | 0.2477 | 1.017505732 | 9.480181029 | 9.480181029 | 0.15353116 | 0.863677285 | 0.00495 | 0.153531159 | 0.153531159 | 0.153531159 | 61.74760286 | 1.790620102 | FALSE | TRUE | FALSE |
| CACNG7 | 9.4739904 | 0.20237133 | 0.3855 | 4.007680036 | 37.89304524 | 37.89304524 | 0.31164364 | 0.821510213 | 1 | 0.311643638 | 0.311643638 | 0.311643638 | 30.40007632 | 1.482874674 | FALSE | TRUE | FALSE |
| ADI1 | 9.448641 | 0.20268105 | 0.37685 | 9.448640965 | 87.00251642 | 348.1885739 | 418.718446 | 0.005777049 | 0.21925 | 61.94125535 | 1676.192085 | 3967.656649 | 0.022565619 | -1.646552747 | FALSE | FALSE | FALSE |
| BTN3A3 | 9.428932 | 0.20299066 | 0.1324 | 1.049198616 | 9.428931987 | 9.428931987 | 1.79030462 | 0.509056938 | 0.00945 | 0.201767621 | 1.790304622 | 1.790304622 | 5.266663488 | 0.72153557 | FALSE | FALSE | FALSE |
| BCAS2 | 9.4089334 | 0.20330017 | 0.25285 | 2.352296709 | 9.408933376 | 9.408933376 | 0.31259258 | 0.821175749 | 1 | 0.312592578 | 0.312592578 | 0.312592578 | 30.09966978 | 1.478561731 | FALSE | TRUE | FALSE |
| HAVCR1 | 9.3965038 | 0.20360946 | 0.50685 | 9.396503776 | 89.08301494 | 89.08301494 | 1.19835263 | 0.567800454 | 0.9835 | 105.7253702 | 10298.08011 | 105323.2664 | 7.841184266 | 0.89438166 | FALSE | FALSE | FALSE |
| SLC25A29 | 9.3827354 | 0.20391856 | 0.5086 | 15.73378008 | 366.9330764 | 1467.499212 | 0.60582334 | 0.70950541 | 0.7533 | 0.605823338 | 2.423025273 | 3.004582127 | 15.4875766 | 1.189983467 | FALSE | TRUE | FALSE |
| VANGL2 | 9.3480429 | 0.20422779 | 0.7497 | 37.39624348 | 598.9186969 | 2397.73084 | 0.05856322 | 0.884937247 | 0.75145 | 0.234074967 | 0.935822684 | 4.62058326 | 159.6231084 | 2.203095764 | FALSE | TRUE | FALSE |
| GRIK3 | 9.3077094 | 0.20453724 | 0.51125 | 9.307709366 | 223.1213945 | 892.1869188 | 16.7549861 | 0.148574489 | 0.0217 | 0.85633564 | 8.294163253 | 28.92124646 | 0.555518777 | -0.255301257 | FALSE | FALSE | FALSE |
| SMO | 9.2978228 | 0.20484644 | 0.19065 | 1.017273406 | 37.18311367 | 148.7368901 | 0.00781688 | 0.897380463 | 1 | 0.499053385 | 4.581408959 | 18.31979946 | 1189.455148 | 3.07534807 | FALSE | TRUE | FALSE |
| LGALS9C | 9.2970222 | 0.20515525 | 0.0094 | 0.959500507 | 9.297022243 | 9.297022243 | 85.2571934 | 0.03422297 | 0.35335 | 85.2571934 | 9008.217424 | 95951.71693 | 0.109046778 | -0.962387162 | FALSE | FALSE | FALSE |
| FAM227B | 9.2847613 | 0.20546384 | 0.91165 | 815.9151967 | 78807.02176 | 801374.9465 | 1239.03797 | 0.002011534 | 0.72285 | 1239.037973 | 1252135.074 | 13140657.76 | 0.007493524 | -2.125313874 | FALSE | FALSE | FALSE |
| OR5H2 | 9.2509048 | 0.20577255 | 0.69455 | 9.250904824 | 844.5830089 | 844.5830089 | 12.4335308 | 0.1762345 | 0.509 | 12.43353082 | 118.2127668 | 118.2127668 | 0.744028784 | -0.128410262 | FALSE | FALSE | FALSE |
| GPR32 | 9.219561 | 0.20608135 | 0.4988 | 2.167438354 | 87.95523051 | 87.95523051 | 0.15482032 | 0.863328724 | 1 | 0.154820324 | 0.154820324 | 0.154820324 | 59.55006942 | 1.774882272 | FALSE | TRUE | FALSE |
| ERMARD | 9.2075184 | 0.20638992 | 0.80835 | 769.5049903 | 698343.6963 | 7064593.491 | 1.30235425 | 0.554325018 | 0.3153 | 0.325661286 | 11.47149268 | 11.47149268 | 7.069903086 | 0.849413461 | FALSE | FALSE | FALSE |
| UPB1 | 9.2024641 | 0.20669818 | 0.255 | 1.283684355 | 82.10477114 | 328.5150976 | 0.31278255 | 0.821116596 | 0.50355 | 0.312782555 | 1.25075053 | 5.002733276 | 29.42128317 | 1.46866161 | FALSE | TRUE | FALSE |
| RASGRP2 | 9.1955059 | 0.20700613 | 0.25885 | 0.970058494 | 9.195505855 | 19.11157517 | 0.14397526 | 0.865879928 | 1 | 0.575755704 | 0.575755704 | 4.876469552 | 63.86865396 | 1.805287763 | FALSE | TRUE | FALSE |
| ITGA7 | 9.1742211 | 0.20731402 | 0.8069 | 85.40935853 | 31811.22828 | 317905.7587 | 2.31827555 | 0.463139435 | 0.4203 | 1.031553554 | 21.37120458 | 85.46640512 | 3.957347126 | 0.597404147 | FALSE | FALSE | FALSE |
| RLN3 | 9.1596421 | 0.20762173 | 0.4977 | 0.985132769 | 87.35296089 | 87.35296089 | 114.779965 | 0.025509963 | 0.12675 | 11.97114615 | 114.7799647 | 114.7799647 | 0.07980175 | -1.097987583 | FALSE | FALSE | FALSE |
| MYBPC2 | 9.085459 | 0.2079302 | 0.93955 | 7524.547424 | 7604769.669 | 318150615 | 32.4270569 | 0.085016563 | 0.37225 | 3.536247917 | 304.463972 | 2925.451005 | 0.280181424 | -0.552560661 | FALSE | FALSE | FALSE |
| YOD1 | 9.0822779 | 0.20823832 | 0.25395 | 2.27071932 | 9.082277943 | 9.082277943 | 0.3021055 | 0.824823926 | 0.00885 | 0.302105499 | 0.302105499 | 0.302105499 | 30.06326587 | 1.478036158 | FALSE | TRUE | FALSE |
| RMDN2 | 9.0579727 | 0.20854643 | 0.31805 | 1.012785031 | 83.04460059 | 83.04460059 | 0.20177402 | 0.851690529 | 0.75425 | 0.201774023 | 1.784152055 | 1.784152055 | 44.89166907 | 1.652165753 | FALSE | TRUE | FALSE |
| ERI2 | 9.0510034 | 0.20885423 | 0.5308 | 9.051003375 | 709.7067138 | 1630.422026 | 17.9898639 | 0.143517572 | 0.5892 | 31.71572021 | 2678.61008 | 10719.26557 | 0.503116834 | -0.298331151 | FALSE | FALSE | FALSE |
| ACTR1B | 9.047177 | 0.2091617 | 0.50105 | 9.047177014 | 331.6880341 | 1327.348432 | 63.0083382 | 0.044105804 | 0.50205 | 63.00833818 | 1009.518962 | 4042.362721 | 0.14358698 | -0.842884938 | FALSE | FALSE | FALSE |
| CCDC8 | 9.0392123 | 0.20946889 | 0.01485 | 0.977054609 | 9.039212305 | 19.26086809 | 1.27009714 | 0.558779659 | 0.75535 | 1.270097139 | 11.82021111 | 25.08088457 | 7.116945648 | 0.852293649 | FALSE | FALSE | FALSE |
| IL37 | 9.030095 | 0.20977582 | 0.50435 | 9.030094966 | 89.63086442 | 911.3854112 | 0.01402768 | 0.895425952 | 1 | 0.014027681 | 0.129975841 | 0.129975841 | 643.7339852 | 2.808706438 | FALSE | TRUE | FALSE |

| | | | | | | | | | | | | | | | | | |
|---|---|---|---|---|---|---|---|---|---|---|---|---|---|---|---|---|---|
| CLDN19 | 9.0143544 | 0.21008259 | 0.50835 | 9.014354381 | 36.0676876 | 144.3478936 | 0.17413733 | 0.858613033 | 1 | 11.13955932 | 104.6197917 | 418.6228608 | 51.76577733 | 1.71404274 | FALSE | TRUE | FALSE |
| PTPMT1 | 8.9572636 | 0.21038987 | 0.00005 | 0.263809964 | 0.263809964 | 0.263809964 | 0.3085553 | 0.822447773 | 1 | 0.308555299 | 0.308555299 | 0.308555299 | 29.02968652 | 1.462842346 | FALSE | TRUE | FALSE |
| RASGRP1 | 8.9322019 | 0.21069715 | 0.1367 | 8.932201852 | 86.19182895 | 176.0876365 | 0.14811018 | 0.865031982 | 1 | 0.148110177 | 0.148110177 | 2.924842933 | 60.3078197 | 1.780373628 | FALSE | TRUE | FALSE |
| SCN2A | 8.8977887 | 0.21100458 | 0.00065 | 0.014209333 | 0.226712913 | 1.093447749 | 0.39817066 | 0.792726657 | 0.0007 | 0.00249456 | 0.009960824 | 0.19276917 | 22.34667101 | 1.349212835 | FALSE | TRUE | FALSE |
| CIZ1 | 8.8756132 | 0.21131195 | 0.26585 | 0.947576802 | 8.875613208 | 31.97732401 | 0.14376708 | 0.865924876 | 1 | 0.143767085 | 0.143767085 | 2.830982969 | 61.73605889 | 1.790538902 | FALSE | TRUE | FALSE |
| AHDC1 | 8.8509667 | 0.21161932 | 0.0015 | 0.263311903 | 5.039045378 | 8.850966656 | 0.30826095 | 0.822583671 | 1 | 0.308260948 | 0.308260948 | 5.92997928 | 28.71257843 | 1.458072195 | FALSE | TRUE | FALSE |
| EPHB6 | 8.8200029 | 0.21192679 | 0.01675 | 0.929942209 | 8.820002862 | 18.1350552 | 0.02067312 | 0.893681813 | 1 | 0.020673116 | 0.020673116 | 0.404363499 | 426.641199 | 2.630062791 | FALSE | TRUE | FALSE |
| NDUFA8 | 8.8189651 | 0.21223387 | 0.5044 | 8.818965121 | 141.1232332 | 141.1232332 | 0.29417177 | 0.826878755 | 1 | 1.176379495 | 1.176379495 | 1.176379495 | 29.97896502 | 1.476816635 | FALSE | TRUE | FALSE |
| CEP55 | 8.816166 | 0.21254059 | 0.2557 | 0.956413403 | 8.816165984 | 18.97809823 | 11.1899557 | 0.185379966 | 0.50645 | 11.18995565 | 106.3768809 | 106.3768809 | 0.787864247 | -0.103548607 | FALSE | FALSE | FALSE |
| ARPC3 | 8.8101732 | 0.21284701 | 0.00385 | 0.441795309 | 0.441795309 | 0.441795309 | 0.48667608 | 0.760381571 | 1 | 0.486676082 | 0.486676082 | 0.486676082 | 18.10274528 | 1.257744441 | FALSE | TRUE | FALSE |
| OR5H6 | 8.8020265 | 0.21315317 | 0.2526 | 0.954940055 | 8.802026491 | 8.802026491 | 11.5332405 | 0.182330212 | 0.25235 | 1.2433425 | 11.53324051 | 11.53324051 | 0.76318763 | -0.117368678 | FALSE | FALSE | FALSE |
| NDUFAF2 | 8.7894298 | 0.21345912 | 0.4972 | 0.948979667 | 83.49902378 | 83.49902378 | 106.629434 | 0.027637333 | 0.31935 | 11.16918179 | 1043.455699 | 1043.455699 | 0.082429677 | -1.083916404 | FALSE | FALSE | FALSE |
| OR2G6 | 8.77123 | 0.21376497 | 0.6876 | 8.771229981 | 799.3554549 | 799.3554549 | 10.448101 | 0.193525012 | 0.77215 | 98.91076239 | 97119.0652 | 189780.7524 | 0.83950471 | -0.075976863 | FALSE | FALSE | FALSE |
| ELP4 | 8.7711433 | 0.21407043 | 0.13235 | 8.771143251 | 83.29798796 | 83.29798796 | 0.14917474 | 0.864781355 | 1 | 0.149174737 | 0.149174737 | 0.149174737 | 58.79777929 | 1.769360924 | FALSE | TRUE | FALSE |
| SLC17A5 | 8.7577468 | 0.2143757 | 0.50615 | 8.757746844 | 197.1463232 | 788.4822301 | 0.09346189 | 0.87743565 | 0.0127 | 0.093461888 | 0.093461888 | 1.85442384 | 93.7039363 | 1.971757835 | FALSE | TRUE | FALSE |
| MYF5 | 8.7407783 | 0.21468084 | 0.06765 | 0.9165512 | 34.93998752 | 34.93998752 | 0.03854657 | 0.889315401 | 1 | 1.436445931 | 22.9630711 | 91.84660089 | 226.7589327 | 2.355564404 | FALSE | TRUE | FALSE |
| HSP90AB1 | 8.7327432 | 0.21498572 | 0.26235 | 2.183519297 | 8.732743241 | 43.03387291 | 0.28605429 | 0.82931819 | 1 | 1.144063018 | 1.144063018 | 9.682321104 | 30.52827199 | 1.484702222 | FALSE | TRUE | FALSE |
| POMGNT2 | 8.7163385 | 0.21529047 | 0.5078 | 8.716338532 | 139.5917644 | 558.8371928 | 0.21225734 | 0.849057958 | 0.8795 | 0.848761991 | 3.394827165 | 7.189037752 | 41.06495755 | 1.613471378 | FALSE | TRUE | FALSE |
| MDH1 | 8.6633937 | 0.21559567 | 0.5048 | 20.88910982 | 826.0697026 | 826.0697026 | 26.040958 | 0.105554418 | 0.25855 | 2.694895383 | 26.04095801 | 26.04095801 | 0.332683371 | -0.477968907 | FALSE | FALSE | FALSE |
| SPIRE2 | 8.6553665 | 0.21590061 | 0.20315 | 8.655366506 | 81.78186477 | 791.6377925 | 0.09095361 | 0.877928049 | 0.5086 | 0.090953608 | 0.090953608 | 1.789600618 | 95.16243122 | 1.978465529 | FALSE | TRUE | FALSE |
| C2orf42 | 8.6334683 | 0.2162055 | 0.1324 | 0.933568292 | 8.633468277 | 8.633468277 | 0.08606881 | 0.878976468 | 0.014 | 0.086068805 | 0.086068805 | 1.704044439 | 100.3089127 | 2.001339523 | FALSE | TRUE | FALSE |
| ANK1 | 8.6244117 | 0.21651015 | 0.4233 | 8.624411723 | 552.6877608 | 2633.552332 | 0.30194405 | 0.824880611 | 0.23165 | 0.301944046 | 4.828634611 | 23.15617405 | 28.56294683 | 1.455803011 | FALSE | TRUE | FALSE |
| SEMA5B | 8.6021894 | 0.21681476 | 0.65865 | 8.602189358 | 786.7345444 | 7797.197436 | 0.18541201 | 0.855672321 | 0.7021 | 1.684307181 | 149.8922363 | 305.7291006 | 46.39499504 | 1.666471133 | FALSE | TRUE | FALSE |
| KIAA0825 | 8.5713762 | 0.21711947 | 0.3145 | 0.954343734 | 78.91537801 | 78.91537801 | 0.1911944 | 0.854051644 | 0.75355 | 0.191194401 | 1.697008885 | 1.697008885 | 44.83068639 | 1.651575388 | FALSE | TRUE | FALSE |
| INTS4 | 8.564678 | 0.2174239 | 0.35585 | 8.564677967 | 780.4797137 | 7721.267489 | 3.17180567 | 0.401778099 | 0.1339 | 0.358022674 | 3.171805674 | 7.046547444 | 2.700253057 | 0.431404466 | FALSE | FALSE | FALSE |
| BRIX1 | 8.557422 | 0.21772806 | 0.1327 | 8.557421978 | 34.24400314 | 34.24400314 | 2.32989865 | 0.462234767 | 0.751 | 2.329898647 | 9.321287068 | 9.321287068 | 3.672873062 | 0.565005919 | FALSE | FALSE | FALSE |
| KRT6A | 8.5432218 | 0.21803205 | 0.36545 | 2.136202116 | 34.17507473 | 136.7431854 | 0.90395955 | 0.598246111 | 0.86195 | 8.388505892 | 319.4809991 | 1277.874672 | 9.450889452 | 0.975472683 | FALSE | TRUE | FALSE |
| VPS72 | 8.5429741 | 0.21833565 | 0.50895 | 8.542974061 | 136.7750355 | 136.7750355 | 0.28962466 | 0.828270849 | 0.50555 | 1.158450941 | 1.158450941 | 5.743960312 | 29.49670776 | 1.469773545 | FALSE | TRUE | FALSE |
| CERCAM | 8.4842059 | 0.21863981 | 0.4002 | 8.484205911 | 307.1057879 | 1229.166472 | 232.406797 | 0.010935176 | 0.3048 | 232.4067967 | 9727.495014 | 14927.26976 | 0.036505842 | -1.437637625 | FALSE | FALSE | FALSE |
| TJP1 | 8.4462955 | 0.21894419 | 0.5152 | 8.44629552 | 82.41050984 | 282.8061767 | 11.5684345 | 0.181311948 | 0.2651 | 1.211576974 | 11.56843445 | 224.4235522 | 0.730115692 | -0.136608318 | FALSE | FALSE | FALSE |
| IFIT1 | 8.4414897 | 0.21924826 | 0.50035 | 8.441489716 | 81.47519626 | 81.47519626 | 91901.387 | 2.2478E-05 | 0.25605 | 8868.105195 | 974566.1186 | 8986996.093 | 9.18538E-05 | -4.03690297 | FALSE | FALSE | FALSE |
| ZNF17 | 8.4202827 | 0.21955229 | 0.50725 | 8.420282747 | 80.21655553 | 80.21655553 | 1.26689106 | 0.559534435 | 0.50505 | 1.266891064 | 1.266891064 | 4.706605692 | 6.64641419 | 0.822587402 | FALSE | FALSE | FALSE |
| ZNF589 | 8.4047537 | 0.21985617 | 0.2514 | 0.912235802 | 8.404753708 | 8.404753708 | 0.14037273 | 0.866729079 | 1 | 0.140372726 | 0.140372726 | 0.140372726 | 59.87454938 | 1.777242258 | FALSE | TRUE | FALSE |
| EIF3M | 8.3954702 | 0.22015982 | 0.2544 | 2.098388637 | 8.395470202 | 8.395470202 | 0.57889954 | 0.721286235 | 1 | 0.578899541 | 0.578899541 | 0.578899541 | 14.50246478 | 1.161441819 | FALSE | TRUE | FALSE |
| SMAD4 | 8.3777297 | 0.22046337 | 0.25865 | 4.88106202 | 78.10189588 | 78.10189588 | 0.17757089 | 0.857624742 | 1 | 0.177570889 | 0.177570889 | 3.517850945 | 47.17963479 | 1.673754575 | FALSE | TRUE | FALSE |
| ZNF454 | 8.3578482 | 0.22076685 | 0.0105 | 0.897398212 | 8.357848172 | 17.79102378 | 0.13338009 | 0.86814787 | 1 | 1.217708806 | 1.217708806 | 2.647056631 | 62.66188864 | 1.79700348 | FALSE | TRUE | FALSE |
| TG | 8.3342519 | 0.22107032 | 0.2485 | 1.001327241 | 506.1839053 | 4204.131083 | 1.67949403 | 0.519261805 | 0.10075 | 0.026276987 | 3.405374823 | 31.94010427 | 4.962358739 | 0.695688157 | FALSE | FALSE | FALSE |
| EFS | 8.3021517 | 0.22137393 | 0.0117 | 0.894756441 | 8.302151669 | 17.6727988 | 0.13746099 | 0.867282333 | 0.014 | 0.137460987 | 0.137460987 | 2.719122077 | 60.39642116 | 1.781011205 | FALSE | TRUE | FALSE |
| NDUFA12 | 8.2993412 | 0.2216772 | 0.00355 | 2.075014723 | 8.299341221 | 8.299341221 | 1.09424107 | 0.579507218 | 0.7502 | 1.094241068 | 4.377708307 | 4.377708307 | 7.584563825 | 0.87993061 | FALSE | FALSE | FALSE |
| NAGK | 8.2935739 | 0.22198017 | 0.25085 | 0.897452307 | 8.293573913 | 8.293573913 | 0.13814284 | 0.867116804 | 0.00925 | 0.138142839 | 0.138142839 | 0.138142839 | 60.03622028 | 1.778413342 | FALSE | TRUE | FALSE |
| VSIG10 | 8.2749139 | 0.22228306 | 0.00905 | 0.897373736 | 8.274913885 | 8.274913885 | 0.13829914 | 0.86706154 | 1 | 0.138299138 | 0.138299138 | 2.741952846 | 59.83344529 | 1.776944011 | FALSE | TRUE | FALSE |
| PASK | 8.2211071 | 0.22258645 | 0.32015 | 2.056282869 | 649.4364694 | 5977.599402 | 0.0230118 | 0.892949887 | 0.759 | 0.091987171 | 6.897089647 | 27.57726341 | 357.2562024 | 2.552979777 | FALSE | TRUE | FALSE |
| SLC25A38 | 8.1696302 | 0.22289029 | 0.5025 | 8.169630238 | 79.24823232 | 79.24823232 | 1.18136799 | 0.569009483 | 0.2542 | 1.181367987 | 11.22174994 | 11.22174994 | 6.915398356 | 0.839817202 | FALSE | FALSE | FALSE |
| ALOXE3 | 8.1619513 | 0.22319386 | 0.80175 | 73.37752116 | 8379.92778 | 75337.17211 | 4.95790737 | 0.307665656 | 0.6097 | 8.967957593 | 317.4809283 | 1563.558711 | 1.646249247 | 0.216495589 | FALSE | FALSE | FALSE |
| AK4 | 8.1456364 | 0.22349732 | 0.5045 | 8.14563643 | 32.59541275 | 32.59541275 | 0.57030586 | 0.724656405 | 1 | 0.570305861 | 0.570305861 | 0.570305861 | 14.28292604 | 1.154817187 | FALSE | TRUE | FALSE |
| SGCG | 8.1147785 | 0.2238009 | 0.3 | 2.030499743 | 69.08937949 | 129.7029055 | 0.04690469 | 0.887356616 | 0.9391 | 0.187477834 | 2.997387055 | 2.997387055 | 173.0056962 | 2.238060402 | FALSE | TRUE | FALSE |
| CANX | 8.099505 | 0.22410435 | 0.99995 | 12921181958 | 2.71185E+14 | 5.36786E+15 | 0.12555853 | 0.869996235 | 1 | 18349683589 | 3.93519E+14 | 4.99941E+15 | 64.5078061 | 1.809612272 | FALSE | TRUE | FALSE |
| RAPH1 | 8.0956649 | 0.22440747 | 0.69565 | 8.095664948 | 801.1746326 | 1553.872792 | 0.13631473 | 0.867535399 | 0.02985 | 0.13631473 | 0.13631473 | 2.670248099 | 59.38950931 | 1.773709737 | FALSE | TRUE | FALSE |
| GNPDA1 | 8.0821137 | 0.22471043 | 0.71815 | 19.15370603 | 306.4620363 | 1226.313273 | 0.2672814 | 0.834538586 | 1 | 2.476357436 | 94.18150294 | 94.18150294 | 30.23821962 | 1.480556217 | FALSE | TRUE | FALSE |
| SLC16A3 | 8.0529546 | 0.22501349 | 0.2606 | 0.863451636 | 8.052954615 | 17.05747661 | 0.13280196 | 0.868256687 | 0.0123 | 0.132801957 | 0.132801957 | 2.627368134 | 60.6388247 | 1.782750775 | FALSE | TRUE | FALSE |
| ZBED4 | 8.0462733 | 0.22531627 | 0.2697 | 2.011713208 | 8.046273329 | 156.1552782 | 0.26038742 | 0.836637709 | 1 | 2.443607754 | 2.443607754 | 47.59297109 | 30.90115984 | 1.48997478 | FALSE | TRUE | FALSE |
| SIMC1 | 8.0134528 | 0.22561921 | 0.07085 | 0.868817577 | 75.76473232 | 75.76473232 | 0.01703603 | 0.894592989 | 1 | 0.151000242 | 0.151000242 | 0.151000242 | 470.3825088 | 2.672451164 | FALSE | TRUE | FALSE |
| DCPS | 7.9875982 | 0.22592219 | 0.74845 | 31.96383955 | 512.2355676 | 2051.340416 | 0.79235829 | 0.623246679 | 0.691 | 0.792358293 | 12.68110063 | 12.68110063 | 10.08079087 | 1.003494605 | FALSE | TRUE | FALSE |
| SGSH | 7.96984 | 0.22622508 | 0.204 | 0.864243991 | 31.8596446 | 127.3915675 | 7.86151145 | 0.230876591 | 0.6194 | 7.861511449 | 503.2661786 | 2014.242266 | 1.013779602 | 0.005943549 | FALSE | FALSE | FALSE |
| MPHOSPH8 | 7.9353388 | 0.22652816 | 0.26185 | 0.853360204 | 7.935338785 | 16.79597895 | 0.1316568 | 0.868527991 | 1 | 0.131656796 | 0.131656796 | 2.596248169 | 60.27291446 | 1.780122192 | FALSE | TRUE | FALSE |
| UBALD1 | 7.9325264 | 0.2268309 | 0.0064 | 0.836200896 | 7.932526448 | 7.932526448 | 0.77706453 | 0.629414263 | 1 | 0.777064531 | 0.777064531 | 0.777064531 | 10.20832393 | 1.008954443 | FALSE | TRUE | FALSE |
| SIGLEC15 | 7.9297726 | 0.22713331 | 0.8108 | 18.65837032 | 1782.135931 | 7128.224441 | 0.29580851 | 0.826490147 | 0.0075 | 0.295808508 | 0.295808508 | 0.295808508 | 26.80711473 | 1.428250073 | FALSE | TRUE | FALSE |
| ADNP | 7.896571 | 0.22743588 | 0.511 | 21.1336062 | 208.5541977 | 414.465369 | 93.9969971 | 0.030562913 | 0.0046 | 2.786066298 | 2.786066298 | 54.76717974 | 0.084008759 | -1.075675433 | FALSE | FALSE | FALSE |
| TTC39B | 7.8791732 | 0.22773836 | 0.4778 | 7.879173168 | 504.2617283 | 2492.841566 | 0.18663614 | 0.85536139 | 0.5098 | 0.186636142 | 0.186636142 | 0.423101906 | 42.21675979 | 1.625484897 | FALSE | TRUE | FALSE |
| SERPINB7 | 7.8608759 | 0.22804076 | 0.1967 | 3.415551423 | 31.44235779 | 125.7962737 | 18.8603432 | 0.138837527 | 0.38565 | 18.8603432 | 697.3456378 | 1477.134146 | 0.416793895 | -0.380078651 | FALSE | FALSE | FALSE |
| NRL | 7.8400128 | 0.22834313 | 0.49335 | 0.833697222 | 75.61988865 | 75.61988865 | 0.12805915 | 0.869442574 | 1 | 0.128059148 | 1.180285897 | 1.180285897 | 61.22180967 | 1.786906163 | FALSE | TRUE | FALSE |
| SMYD1 | 7.7906522 | 0.22864594 | 0.5075 | 7.790652181 | 71.3456983 | 285.5483557 | 0.16784058 | 0.860198757 | 0.01385 | 0.167840578 | 0.167840578 | 3.32273865 | 46.41697652 | 1.666676848 | FALSE | TRUE | FALSE |
| DVL2 | 7.762931 | 0.22894883 | 0.0005 | 0.22953094 | 0.22953094 | 4.504754342 | 0.27288597 | 0.83300604 | 1 | 0.272885969 | 0.272885969 | 5.367698803 | 28.44752716 | 1.454044521 | FALSE | TRUE | FALSE |
| LRP11 | 7.7342208 | 0.22925182 | 0.008 | 0.806434572 | 7.734220751 | 7.734220751 | 0.12270816 | 0.870734617 | 1 | 0.122708157 | 0.122708157 | 0.122708157 | 63.0293936 | 1.799543129 | FALSE | TRUE | FALSE |
| UPF3A | 7.7286983 | 0.22955452 | 0.50075 | 7.728698338 | 73.80421356 | 73.80421356 | 0.12455105 | 0.870229149 | 0.74935 | 1.13675505 | 10.6483543 | 10.6483543 | 62.05245484 | 1.792758967 | FALSE | TRUE | FALSE |
| GOLPH3L | 7.7098571 | 0.22985715 | 0.505 | 7.709857124 | 30.85499326 | 123.5131108 | 0.63111035 | 0.698793808 | 0.8774 | 2.524545309 | 10.10112097 | 10.10112097 | 12.2163377 | 1.08694103 | FALSE | TRUE | FALSE |
| PPP2R5D | 7.6949664 | 0.23015965 | 0.0055 | 0.389953986 | 0.389953986 | 7.694966382 | 0.43609344 | 0.778791052 | 1 | 0.436093436 | 0.436093436 | 8.619260777 | 17.64522403 | 1.246627177 | FALSE | TRUE | FALSE |
| ARID1B | 7.6848786 | 0.23046195 | 0.99825 | 874298.0188 | 1221395458 | 14250465061 | 921.768859 | 0.002556761 | 0.94475 | 12444568.9 | 18644918130 | 2.22466E+11 | 0.008337099 | -2.078985022 | FALSE | FALSE | FALSE |
| IQCD | 7.6337152 | 0.23076472 | 0.64245 | 74.28311776 | 7558.114248 | 78940.76281 | 0.01412127 | 0.895383164 | 0.87605 | 0.280668033 | 11.53908249 | 11.53908249 | 540.5825744 | 2.732862042 | FALSE | TRUE | FALSE |
| DEFB123 | 7.5765101 | 0.23106807 | 0.00175 | 0.800210986 | 7.576510108 | 7.576510108 | 0.84829599 | 0.607824929 | 1 | 0.848295988 | 0.848295988 | 0.848295988 | 8.931446354 | 0.950921794 | FALSE | TRUE | FALSE |
| ATP6V0E1 | 7.5736824 | 0.23137109 | 0.501 | 7.573682449 | 7.573682449 | 7.573682449 | 0.84776335 | 0.608231153 | 1 | 0.84776335 | 0.84776335 | 0.84776335 | 8.933722421 | 0.951032454 | FALSE | TRUE | FALSE |
| LPAR6 | 7.5710525 | 0.23167378 | 1 | 9.65761E+29 | 2.43166E+38 | 6.94421E+40 | 1.88472868 | 0.500502905 | 1 | 1.32629E+26 | 1.20045E+32 | 5.36587E+35 | 4.01705167 | 0.603907418 | FALSE | FALSE | FALSE |
| SLC25A19 | 7.5708914 | 0.23197609 | 0.00165 | 0.223115747 | 0.223115747 | 0.265167932 | 0.26642831 | 0.834740818 | 1 | 0.266428307 | 0.266428307 | 0.266428307 | 28.41624258 | 1.453566651 | FALSE | TRUE | FALSE |

| | | | | | | | | | | | | | | | | | |
|---|---|---|---|---|---|---|---|---|---|---|---|---|---|---|---|---|---|
| PSG5 | 7.5671477 | 0.23227808 | 0.1339 | 7.567147712 | 71.15122925 | 71.15122925 | 10.0798316 | 0.196917776 | 0.49705 | 1.091961799 | 95.43653009 | 95.43653009 | 0.750721637 | -0.124521067 | FALSE | FALSE | FALSE |
| CYB5D1 | 7.557914 | 0.23257986 | 0.63115 | 7.55791403 | 286.8744428 | 286.8744428 | 0.26786864 | 0.83436968 | 1 | 0.267868642 | 0.267868642 | 0.267868642 | 28.21500106 | 1.450480071 | FALSE | TRUE | FALSE |
| PAAF1 | 7.5466932 | 0.23288144 | 1 | 1.97018E+24 | 1.54575E+30 | 6.37267E+33 | 6.39546134 | 0.258037296 | 1 | 2.14771E+38 | 9.63151E+46 | 5.92776E+50 | 1.180007638 | 0.071884818 | FALSE | FALSE | FALSE |
| ZNF197 | 7.5322038 | 0.2331829 | 0.8171 | 30.14313655 | 285.5712812 | 1143.114234 | 19.3215836 | 0.135720527 | 0.386 | 19.32158357 | 751.1968939 | 751.1968939 | 0.389833667 | -0.409120656 | FALSE | FALSE | FALSE |
| COX17 | 7.5175552 | 0.23348422 | 0.25145 | 7.517555242 | 72.90533638 | 72.90533638 | 7.98323111 | 0.228891552 | 0.50005 | 7.983231111 | 7.983231111 | 7.983231111 | 0.941668247 | -0.026102074 | FALSE | FALSE | FALSE |
| STK11 | 7.5124297 | 0.23378525 | 0.0128 | 0.3796991 | 0.3796991 | 7.51242967 | 0.42091343 | 0.784228016 | 1 | 1.683440073 | 1.683440073 | 8.33862297 | 17.84792118 | 1.251587639 | FALSE | TRUE | FALSE |
| ZNF438 | 7.4852018 | 0.23408637 | 0.69215 | 7.485201803 | 690.4660854 | 690.4660854 | 873.578655 | 0.002664509 | 0.34265 | 89.46268469 | 8738.735996 | 89501.91725 | 0.008568435 | -2.067098501 | FALSE | FALSE | FALSE |
| ATP6V1G1 | 7.4757933 | 0.23438728 | 0.4962 | 0.789828015 | 7.475793302 | 7.475793302 | 0.84046825 | 0.610149056 | 1 | 0.840468247 | 0.840468247 | 0.840468247 | 8.894795642 | 0.949135975 | FALSE | TRUE | FALSE |
| PSEN2 | 7.4685199 | 0.23468793 | 0.3975 | 7.468519863 | 478.0966991 | 1913.485645 | 0.06607486 | 0.883333028 | 0.69125 | 0.153196005 | 2.445907532 | 2.445907532 | 113.0311944 | 2.053198317 | FALSE | TRUE | FALSE |
| RPS10 | 7.4536776 | 0.23498845 | 0.5047 | 7.453677579 | 7.453677579 | 7.453677579 | 0.81631165 | 0.615400646 | 1 | 0.816311647 | 0.816311647 | 0.816311647 | 9.130921515 | 0.96051461 | FALSE | TRUE | FALSE |
| SLC27A3 | 7.4482649 | 0.2352887 | 0.37125 | 7.448264917 | 637.9002683 | 6113.243236 | 1418.76306 | 0.001724111 | 0.26065 | 149.3621642 | 13791.27695 | 137119.7401 | 0.00524983 | -2.279854757 | FALSE | FALSE | FALSE |
| DENND3 | 7.4446967 | 0.23558863 | 0.13875 | 1.989083812 | 18.37007506 | 38.61543339 | 0.16101322 | 0.861798363 | 0.76065 | 0.356906399 | 1.427585302 | 12.04037534 | 46.23655453 | 1.664985464 | FALSE | TRUE | FALSE |
| ANKLE1 | 7.4123796 | 0.23588874 | 0.1339 | 0.801660376 | 7.412379591 | 7.412379591 | 0.15072479 | 0.864449624 | 0.87625 | 1.370686145 | 51.11833432 | 51.11833432 | 49.17823777 | 1.691772962 | FALSE | TRUE | FALSE |
| DLAT | 7.4078304 | 0.23618855 | 0.2637 | 0.786949303 | 7.407830387 | 15.52262865 | 0.12165455 | 0.870891822 | 0.01695 | 0.121654551 | 0.121654551 | 2.403614066 | 60.89234085 | 1.78456267 | FALSE | TRUE | FALSE |
| NUTF2 | 7.399168 | 0.23648813 | 0.50775 | 71.88320746 | 716.2571774 | 716.2571774 | 7.84237353 | 0.231206895 | 0.74685 | 7.842373534 | 76.29029842 | 76.29029842 | 0.943485794 | -0.025264635 | FALSE | FALSE | FALSE |
| IL22RA2 | 7.387735 | 0.23678754 | 0.2518 | 0.796189687 | 7.387735026 | 7.387735026 | 1.06099339 | 0.582259408 | 0.75215 | 1.060993389 | 9.900147826 | 9.900147826 | 6.963035871 | 0.842798632 | FALSE | FALSE | FALSE |
| EGFL8 | 7.3612463 | 0.23708703 | 0.12955 | 0.808972052 | 7.361246251 | 7.361246251 | 0.29554845 | 0.826582829 | 0.75015 | 0.295548455 | 1.182053971 | 1.182053971 | 24.90707068 | 1.396322653 | FALSE | TRUE | FALSE |
| LYPD2 | 7.3357296 | 0.23738657 | 0.5074 | 7.335729616 | 7.335729616 | 7.335729616 | 0.82348222 | 0.613928765 | 1 | 0.823482219 | 0.823482219 | 0.823482219 | 8.908182168 | 0.94978909 | FALSE | TRUE | FALSE |
| PPIL2 | 7.3040386 | 0.23768629 | 0.01305 | 0.789262096 | 7.304038559 | 15.58408396 | 1.08560494 | 0.580197859 | 0.5061 | 1.085604939 | 1.085604939 | 2.364877393 | 6.728081546 | 0.827891247 | FALSE | FALSE | FALSE |
| CSTF3 | 7.297251 | 0.23798574 | 0.2624 | 1.997634498 | 18.95572733 | 39.39845754 | 2.47568167 | 0.447917789 | 0.50925 | 2.475681672 | 2.475681672 | 48.90828357 | 2.947572412 | 0.469464483 | FALSE | FALSE | FALSE |
| C16orf89 | 7.2854772 | 0.23828503 | 0.50705 | 68.37945777 | 657.8747732 | 6484.138259 | 0.8026448 | 0.620034016 | 0.9999 | 678391.5428 | 851727173.7 | 8655872136 | 9.076838469 | 0.957934607 | FALSE | TRUE | FALSE |
| IFNA13 | 7.2771766 | 0.23858409 | 0.7532 | 7.277176566 | 70.62968279 | 70.62968279 | 7.76408605 | 0.232197977 | 0.5058 | 7.764086054 | 7.764086054 | 7.764086054 | 0.937286954 | -0.028127428 | FALSE | FALSE | FALSE |
| AQP1 | 7.2439555 | 0.23888335 | 0.11735 | 1.811185506 | 28.97991428 | 115.9649837 | 1.59197162 | 0.527098234 | 0.5129 | 2.735449135 | 25.4524612 | 101.809848 | 4.550304427 | 0.658040453 | FALSE | FALSE | FALSE |
| SMARCC2 | 7.2044666 | 0.23918291 | 0.01025 | 0.21401126 | 0.21401126 | 7.204466559 | 0.25677699 | 0.837521233 | 1 | 0.256776993 | 0.256776993 | 5.018288161 | 28.05729004 | 1.448045722 | FALSE | TRUE | FALSE |
| ALDH5A1 | 7.1936708 | 0.23948228 | 0.4037 | 7.193670796 | 258.083358 | 1032.975312 | 0.05532486 | 0.885569607 | 0.7521 | 0.055324862 | 0.22118846 | 0.274437395 | 130.0260055 | 2.114030221 | FALSE | TRUE | FALSE |
| CDKL3 | 7.1746272 | 0.23978161 | 0.3176 | 1.793888785 | 65.61902503 | 65.61902503 | 0.03593294 | 0.889897331 | 0.87385 | 2.83944909 | 26.22908135 | 26.22908135 | 199.6671415 | 2.300306601 | FALSE | TRUE | FALSE |
| CLPSL2 | 7.1577162 | 0.24008086 | 0.50075 | 7.157716161 | 7.157716161 | 7.157716161 | 0.8087231 | 0.617924945 | 1 | 0.808723101 | 0.808723101 | 0.808723101 | 8.850638932 | 0.946974624 | FALSE | TRUE | FALSE |
| PCDHAC1 | 7.1079139 | 0.2403806 | 0.7791 | 68.02010187 | 6693.626711 | 68755.78942 | 1.47691378 | 0.537875274 | 0.33065 | 1.47691378 | 134.6955134 | 134.6955134 | 4.812680363 | 0.682387019 | FALSE | FALSE | FALSE |
| QSOX2 | 7.0875389 | 0.24068031 | 0.25715 | 0.762033272 | 7.08753886 | 14.99109421 | 4.03138363 | 0.355079971 | 0.0001 | 0.119157892 | 0.119157892 | 2.348753757 | 1.758090897 | 0.245041325 | FALSE | FALSE | FALSE |
| ACSM2A | 7.085708 | 0.24097969 | 0.50285 | 65.6680304 | 6061.262072 | 12328.57865 | 0.15110023 | 0.86435785 | 0.9362 | 115.3951925 | 10891.09569 | 21937.1079 | 46.89409169 | 1.671118128 | FALSE | TRUE | FALSE |
| MAPK1IP1L | 7.0492753 | 0.24127933 | 0.0053 | 7.049275314 | 7.049275314 | 7.049275314 | 0.79658709 | 0.621930637 | 1 | 0.796587088 | 0.796587088 | 0.796587088 | 8.8493467 | 0.94691121 | FALSE | TRUE | FALSE |
| HMGCS2 | 7.0485363 | 0.24157861 | 0.18375 | 0.794829522 | 28.19388204 | 112.8026412 | 0.09926582 | 0.876169279 | 0.77415 | 1.587399367 | 25.41011184 | 101.7019818 | 71.00667755 | 1.851299192 | FALSE | TRUE | FALSE |
| RAB3GAP2 | 7.0102958 | 0.24187818 | 0.3222 | 1.753174899 | 66.15588113 | 136.9373459 | 0.03574056 | 0.889959914 | 0.5167 | 0.323056513 | 0.323056513 | 6.328228918 | 196.1439747 | 2.292574972 | FALSE | TRUE | FALSE |
| GALNT7 | 7.0009718 | 0.24217755 | 0.01335 | 0.741054929 | 7.000971775 | 14.63622219 | 0.11511099 | 0.872352405 | 1 | 0.115110989 | 0.115110989 | 2.276914526 | 60.81931765 | 1.784041543 | FALSE | TRUE | FALSE |
| TCP10L | 6.9901189 | 0.24247673 | 0.0056 | 0.739245839 | 6.990118922 | 6.990118922 | 0.74229171 | 0.645059672 | 1 | 7.016260557 | 68.06436834 | 68.06436834 | 9.416943208 | 0.973909951 | FALSE | TRUE | FALSE |
| COX6A2 | 6.9867289 | 0.24277561 | 0.2523 | 6.986728919 | 67.63691173 | 67.63691173 | 0.80840596 | 0.618021347 | 1 | 7.64286324 | 7.64286324 | 7.64286324 | 8.642599445 | 0.936644385 | FALSE | TRUE | FALSE |
| DNASE1L3 | 6.98288 | 0.24307419 | 0.68745 | 16.61660865 | 648.6212154 | 648.6212154 | 0.25429919 | 0.838121272 | 1 | 0.254299193 | 0.254299193 | 0.254299193 | 27.45930837 | 1.438689594 | FALSE | TRUE | FALSE |
| C2CD4B | 6.982875 | 0.2433724 | 0.5042 | 6.982875014 | 65.87947931 | 65.87947931 | 8.39516817 | 0.220221601 | 0.3783 | 8.395168169 | 318.0357385 | 318.0357385 | 0.831773095 | -0.079995132 | FALSE | FALSE | FALSE |
| TNS4 | 6.9765898 | 0.24367036 | 0.32225 | 1.74353637 | 27.92311554 | 34.36350048 | 0.57109753 | 0.724221703 | 0.8787 | 2.284668548 | 9.142072374 | 19.31625131 | 12.21610917 | 1.086932905 | FALSE | TRUE | FALSE |
| POMP | 6.9582638 | 0.24396827 | 0.2531 | 6.958263753 | 67.6009898 | 67.6009898 | 0.81322418 | 0.616569997 | 1 | 0.813224178 | 0.813224178 | 0.813224178 | 8.556390648 | 0.932290604 | FALSE | TRUE | FALSE |
| ZCCHC24 | 6.9170288 | 0.24426653 | 0.00585 | 6.917028834 | 6.917028834 | 6.917028834 | 0.75910752 | 0.637199752 | 1 | 0.759107524 | 7.181450114 | 7.181450114 | 9.112054112 | 0.95961629 | FALSE | TRUE | FALSE |
| NACA | 6.9020336 | 0.24456468 | 0.9886 | 256298.316 | 77354552.07 | 3311300294 | 1.22215822 | 0.564995965 | 0.8923 | 109.2020799 | 10772.29308 | 110872.3008 | 5.64741415 | 0.751849638 | FALSE | FALSE | FALSE |
| PARP3 | 6.894918 | 0.24486259 | 0.81445 | 65.21327442 | 632.2516178 | 6279.60694 | 1.0666467 | 0.581917055 | 0.5038 | 9.915316992 | 94.53786089 | 94.53786089 | 6.464106685 | 0.810508516 | FALSE | FALSE | FALSE |
| PLBD2 | 6.8938127 | 0.24516016 | 0.50855 | 6.893812655 | 65.05336917 | 65.05336917 | 0.11813711 | 0.871682947 | 0.5108 | 1.067783951 | 1.067783951 | 2.340516641 | 58.3543354 | 1.766073127 | FALSE | TRUE | FALSE |
| NIT1 | 6.8891016 | 0.24545744 | 0.4088 | 6.889101588 | 258.0842053 | 2478.286812 | 0.33292969 | 0.814591079 | 0.5017 | 0.332929686 | 0.332929686 | 0.332929686 | 20.69236203 | 1.315810068 | FALSE | TRUE | FALSE |
| PHACTR4 | 6.883324 | 0.24575447 | 1 | 1.13992E+11 | 2.5623E+15 | 4.30404E+17 | 539.775295 | 0.004146912 | 1 | 2.58168E+18 | 1.26395E+24 | 2.67988E+26 | 0.012752203 | -1.894414788 | FALSE | FALSE | FALSE |
| IL18RAP | 6.8756343 | 0.24605126 | 0.01165 | 0.74728016 | 6.875634303 | 14.78660962 | 0.11785916 | 0.871755358 | 1 | 0.117859156 | 0.117859156 | 2.335091561 | 58.33771899 | 1.765949444 | FALSE | TRUE | FALSE |
| ATXN7 | 6.8447376 | 0.24634823 | 0.50725 | 6.844737572 | 67.21891346 | 134.439403 | 8.14453332 | 0.224902946 | 0.26275 | 0.848086425 | 8.144533325 | 16.69347546 | 0.840408812 | -0.075509403 | FALSE | FALSE | FALSE |
| MAP4 | 6.8220776 | 0.24664523 | 0.0212 | 0.723153117 | 6.822077632 | 24.41275332 | 0.10934346 | 0.873561672 | 1 | 1.009732272 | 1.009732272 | 19.82959543 | 62.39127296 | 1.795123847 | FALSE | TRUE | FALSE |
| OR5D14 | 6.8180001 | 0.24694194 | 0.6861 | 6.818000062 | 617.7688562 | 617.7688562 | 9.79483843 | 0.201668634 | 0.0048 | 1.059324802 | 9.794838426 | 9.794838426 | 0.696080912 | -0.157340275 | FALSE | FALSE | FALSE |
| TGM6 | 6.8151595 | 0.24723834 | 0.81565 | 593.4395782 | 1486625.679 | 58533566.5 | 3648.46338 | 0.000523924 | 0.14255 | 38.88918712 | 58479.66646 | 983634.6213 | 0.001867953 | -2.728633965 | FALSE | FALSE | FALSE |
| CRCP | 6.7889961 | 0.24753484 | 0.0034 | 1.697383284 | 6.788996107 | 6.788996107 | 0.23548716 | 0.843194668 | 0.003 | 0.235487162 | 0.235487162 | 0.235487162 | 28.82958059 | 1.459838324 | FALSE | TRUE | FALSE |
| ZNF345 | 6.7631953 | 0.24783142 | 0.5075 | 6.763195338 | 64.75214485 | 64.75214485 | 0.11370562 | 0.872628604 | 1 | 1.041097173 | 1.041097173 | 1.041097173 | 59.47987042 | 1.774370014 | FALSE | TRUE | FALSE |
| PGAM1 | 6.7615972 | 0.24812767 | 0.31775 | 3.898807713 | 62.38432101 | 62.38432101 | 0.57381483 | 0.723055323 | 0.2546 | 0.573814826 | 2.295137629 | 2.295137629 | 11.78358748 | 1.07127753 | FALSE | TRUE | FALSE |
| TRAPPC3 | 6.7491417 | 0.24842377 | 0.7559 | 6.749141704 | 65.57756714 | 65.57756714 | 0.79039988 | 0.62371443 | 1 | 0.790399877 | 0.790399877 | 0.790399877 | 8.538895188 | 0.931401683 | FALSE | TRUE | FALSE |
| PTK6 | 6.7465674 | 0.24871957 | 0.62245 | 15.66725348 | 1453.702781 | 5811.727713 | 0.04555976 | 0.887690149 | 0.7502 | 0.045559759 | 0.182159834 | 0.902213724 | 148.0817174 | 2.170501443 | FALSE | TRUE | FALSE |
| C1QTNF4 | 6.7457799 | 0.24901502 | 0.87345 | 64.31414451 | 6303.759883 | 25215.59925 | 1.8955822 | 0.499394809 | 0.99165 | 706.4776856 | 191019.4191 | 763927.0428 | 3.558684992 | 0.551289547 | FALSE | FALSE | FALSE |
| CISD2 | 6.7348357 | 0.2493103 | 0.49935 | 0.70985215 | 6.73483575 | 6.73483575 | 0.56021631 | 0.728692055 | 1 | 0.56021631 | 0.56021631 | 0.56021631 | 12.02184876 | 1.07997126 | FALSE | TRUE | FALSE |
| PIRT | 6.7283566 | 0.24960534 | 0.7497 | 6.728356604 | 65.11603643 | 65.11603643 | 0.79090409 | 0.62352747 | 1 | 0.790904089 | 0.790904089 | 0.790904089 | 8.507171343 | 0.92978518 | FALSE | TRUE | FALSE |
| TRIM21 | 6.7253842 | 0.24990007 | 0.8561 | 26.91795986 | 425.7066922 | 1705.155824 | 4.93937707 | 0.309385392 | 0.7791 | 34.6943708 | 1269.411253 | 2761.945341 | 1.361585504 | 0.134044919 | FALSE | FALSE | FALSE |
| PRAMEF17 | 6.6999466 | 0.25019489 | 0.75295 | 6.699946641 | 65.17045842 | 65.17045842 | 611.653255 | 0.003552439 | 0.19565 | 611.6532547 | 6239.25403 | 65196.96589 | 0.010953831 | -1.960433947 | FALSE | FALSE | FALSE |
| TUB | 6.6810878 | 0.25048969 | 0.0085 | 0.41771514 | 1.670357601 | 6.681087806 | 0.10501942 | 0.874720082 | 0.8101 | 0.419693142 | 15.72806147 | 15.72806147 | 63.61763981 | 1.803577553 | FALSE | TRUE | FALSE |
| CGB8 | 6.6771919 | 0.25078419 | 0.25335 | 6.677191891 | 64.74596147 | 64.74596147 | 0.75993442 | 0.636593135 | 1 | 7.190911039 | 7.190911039 | 7.190911039 | 8.78653698 | 0.943817741 | FALSE | TRUE | FALSE |
| ENKUR | 6.6629072 | 0.25107859 | 0.3787 | 2.894984207 | 26.65406043 | 26.65406043 | 0.23302377 | 0.84408505 | 1 | 0.233023774 | 0.233023774 | 0.233023774 | 28.59325089 | 1.456263535 | FALSE | TRUE | FALSE |
| C1orf52 | 6.618562 | 0.2513734 | 1 | 2.91802E+27 | 4.27426E+34 | 1.99176E+38 | 0.13038665 | 0.86885217 | 1 | 1.97575E+24 | 2.57646E+31 | 1.14193E+35 | 50.76104201 | 1.705530529 | FALSE | TRUE | FALSE |
| LRRC42 | 6.6124195 | 0.25166797 | 0.504 | 6.612419497 | 61.9974981 | 61.9974981 | 0.11490054 | 0.872413878 | 0.50935 | 1.032577457 | 1.032577457 | 2.281777993 | 57.54907475 | 1.760038346 | FALSE | TRUE | FALSE |
| RND1 | 6.5848317 | 0.25196266 | 0.0084 | 1.646112409 | 6.584831727 | 6.584831727 | 0.45730892 | 0.772030568 | 1 | 1.828925996 | 7.31629409 | 7.31629409 | 14.39908877 | 1.158335009 | FALSE | TRUE | FALSE |
| DHRS7B | 6.5669958 | 0.25225731 | 0.47085 | 3.834239851 | 61.30685294 | 245.2374042 | 45.3940379 | 0.060182888 | 0.6416 | 445.3777096 | 46066.67089 | 485103.324 | 0.144666482 | -0.839632081 | FALSE | FALSE | FALSE |
| TBXA2R | 6.5623924 | 0.25255169 | 0.38585 | 2.861465263 | 26.25419546 | 26.25419546 | 3.45763879 | 0.385516838 | 0.5079 | 7.961078536 | 127.4552621 | 127.4552621 | 1.897940421 | 0.278282575 | FALSE | FALSE | FALSE |
| NPNT | 6.5522983 | 0.25284589 | 0.19705 | 1.638400238 | 61.36145993 | 61.36145993 | 0.06944329 | 0.882581462 | 0.50625 | 0.277664248 | 0.277664248 | 1.37559475 | 94.35466683 | 1.974763385 | FALSE | TRUE | FALSE |
| TBCA | 6.5374339 | 0.25314 | 0.751 | 6.537433907 | 63.32376568 | 63.32376568 | 0.77062346 | 0.631929263 | 0.0032 | 0.770623462 | 0.770623462 | 0.770623462 | 8.483305049 | 0.928565084 | FALSE | TRUE | FALSE |
| MT1X | 6.5346752 | 0.25343381 | 0.75255 | 6.534675215 | 63.10988777 | 63.10988777 | 0.77189062 | 0.631572161 | 1 | 0.771890625 | 0.771890625 | 0.771890625 | 8.465804619 | 0.927668241 | FALSE | TRUE | FALSE |
| DAPL1 | 6.5001366 | 0.25372786 | 0.7486 | 6.500136644 | 63.02813691 | 63.02813691 | 0.76602198 | 0.633615582 | 1 | 0.766021978 | 0.766021978 | 0.766021978 | 8.485574603 | 0.928681256 | FALSE | TRUE | FALSE |

| | | | | | | | | | | | | | | | | |
|---|---|---|---|---|---|---|---|---|---|---|---|---|---|---|---|---|
| CAPZA2 | 6.4983822 | 0.25402159 | 0.0064 | 6.498382213 | 6.498382213 | 6.498382213 | 0.72970143 | 0.651854381 | 0.0063 | 0.729701429 | 0.729701429 | 0.729701429 | 8.90553582 | 0.949660055 | FALSE | TRUE | FALSE |
| XPR1 | 6.4935966 | 0.25431506 | 0.2614 | 1.623059511 | 6.493596596 | 31.98666542 | 0.46186485 | 0.770331156 | 1 | 0.461864854 | 0.461864854 | 9.118278659 | 14.05951664 | 1.14797039 | FALSE | TRUE | FALSE |
| XRCC3 | 6.4826028 | 0.25460837 | 0.19435 | 6.482602754 | 25.94451749 | 103.8604658 | 0.13982879 | 0.866840062 | 1 | 0.139828794 | 0.139828794 | 0.139828794 | 46.36100036 | 1.666152798 | FALSE | TRUE | FALSE |
| APEX1 | 6.4497666 | 0.2549019 | 0.3153 | 6.449766598 | 148.2837255 | 592.7774796 | 1.17127783 | 0.570452738 | 0.62585 | 1.171277826 | 43.58348248 | 43.58348248 | 5.506606936 | 0.740884077 | FALSE | FALSE | FALSE |
| NOP2 | 6.4440048 | 0.25519519 | 0.50955 | 6.444004848 | 61.30277981 | 126.6764674 | 0.97083508 | 0.591101589 | 0.7578 | 0.970835082 | 9.063401865 | 19.12399498 | 6.637589606 | 0.822010397 | FALSE | TRUE | FALSE |
| TRMT61B | 6.420769 | 0.25548853 | 0.50825 | 6.420769015 | 60.2489581 | 60.2489581 | 0.97543834 | 0.590661721 | 0.7518 | 0.97543834 | 8.995673413 | 8.995673413 | 6.582444789 | 0.818387225 | FALSE | TRUE | FALSE |
| BET1 | 6.4205908 | 0.25578152 | 0.5046 | 6.420590795 | 6.420590795 | 6.420590795 | 56.9541941 | 0.049401011 | 0.5024 | 563.196638 | 5708.508449 | 59249.63094 | 0.112732537 | -0.947950719 | FALSE | FALSE | FALSE |
| EXT2 | 6.4185212 | 0.2560742 | 0.1443 | 1.605358845 | 102.6799842 | 410.8409194 | 4659.37872 | 0.000397531 | 0.0292 | 72.68513778 | 4659.378722 | 18656.86678 | 0.001377549 | -2.86089303 | FALSE | FALSE | FALSE |
| ATP8B2 | 6.4113441 | 0.25636666 | 0.13305 | 0.401110731 | 14.89431881 | 59.55487277 | 0.0933067 | 0.877483153 | 0.4001 | 0.093306703 | 0.8498636 | 4.149628189 | 68.71257774 | 1.837036241 | FALSE | TRUE | FALSE |
| UTS2R | 6.4077609 | 0.25665883 | 0.69235 | 6.407760872 | 590.3630184 | 590.3630184 | 0.10355871 | 0.875036104 | 1 | 8.724864823 | 813.5746194 | 813.5746194 | 61.87563441 | 1.791519665 | FALSE | TRUE | FALSE |
| TNFRSF12A | 6.4039391 | 0.25695073 | 0.7491 | 6.403939132 | 61.96270619 | 61.96270619 | 6.70843958 | 0.251898233 | 0.7514 | 6.708439577 | 64.96605167 | 64.96605167 | 0.954609348 | -0.020174317 | FALSE | FALSE | FALSE |
| DOCK10 | 6.4025179 | 0.2572423 | 0.51405 | 6.402517874 | 410.1324969 | 1981.34913 | 0.01076757 | 0.896541139 | 1 | 0.010767573 | 0.208291144 | 0.360900764 | 594.6110397 | 2.774232968 | FALSE | TRUE | FALSE |
| SH2B2 | 6.3974041 | 0.25753361 | 0.51025 | 6.397404117 | 6.397404117 | 13.36275998 | 0.71440423 | 0.658834238 | 1 | 0.714404227 | 0.714404227 | 0.714404227 | 8.954879993 | 0.95205977 | FALSE | TRUE | FALSE |
| VIPAS39 | 6.3909484 | 0.2578247 | 0.0028 | 0.188971899 | 0.188971899 | 1.598047802 | 0.06799495 | 0.882870897 | 0.01515 | 0.067994946 | 0.067994946 | 1.347416243 | 93.99152181 | 1.973088681 | FALSE | TRUE | FALSE |
| FOLR3 | 6.3857263 | 0.25811552 | 0.8772 | 61.90484179 | 615.5120463 | 615.5120463 | 0.77890886 | 0.629052041 | 1 | 0.778908861 | 0.778908861 | 0.778908861 | 8.198297212 | 0.913723659 | FALSE | TRUE | FALSE |
| KAZALD1 | 6.3853884 | 0.25840601 | 0.68705 | 6.385388406 | 600.2468038 | 600.2468038 | 9.3009316 | 0.208423899 | 0.2556 | 0.989541319 | 9.300931601 | 9.300931601 | 0.686532132 | -0.163339131 | FALSE | FALSE | FALSE |
| PKP1 | 6.3836372 | 0.25869619 | 0.01665 | 0.325040477 | 1.299930456 | 6.383637197 | 0.11180563 | 0.87294441 | 0.0204 | 0.111805631 | 0.111805631 | 2.20057371 | 57.09584675 | 1.756604518 | FALSE | TRUE | FALSE |
| PLEKHG5 | 6.3785199 | 0.25898611 | 0.3316 | 1.595254329 | 60.14388147 | 124.3235731 | 0.27951912 | 0.831217113 | 0.87945 | 2.586693223 | 24.55234776 | 50.57181298 | 22.81961909 | 1.358308391 | FALSE | TRUE | FALSE |
| ZNF117 | 6.3642187 | 0.25927594 | 0.26005 | 6.364218726 | 61.87256573 | 61.87256573 | 582.793739 | 0.003707603 | 0.5 | 393.7551531 | 5942.14831 | 62063.97897 | 0.010920191 | -1.96176978 | FALSE | FALSE | FALSE |
| HELT | 6.3608606 | 0.25956549 | 0.25355 | 1.590075259 | 6.360860646 | 7.868038369 | 0.45360871 | 0.773256078 | 1 | 0.453608714 | 0.453608714 | 8.988121013 | 14.02279201 | 1.146834492 | FALSE | TRUE | FALSE |
| CLCNKB | 6.3570559 | 0.25985476 | 0.72975 | 58.75293857 | 21550.30738 | 142359.4578 | 0.089737 | 0.878191702 | 0.75085 | 3.346930171 | 313.8081574 | 3146.409004 | 70.8409655 | 1.850284472 | FALSE | TRUE | FALSE |
| SPESP1 | 6.3459958 | 0.26014388 | 1 | 7.36478E+47 | 1.14602E+58 | 9.16074E+61 | 0.97491184 | 0.590881755 | 1 | 4.27145E+53 | 1.0832E+64 | 1.03157E+68 | 6.509302234 | 0.813534437 | FALSE | TRUE | FALSE |
| ATP6V1C2 | 6.3437664 | 0.2604327 | 0.5048 | 6.343766377 | 60.17302997 | 60.17302997 | 0.98936841 | 0.589558845 | 0.01175 | 0.989368414 | 0.989368414 | 2.162707804 | 6.411935419 | 0.80698914 | FALSE | TRUE | FALSE |
| OGN | 6.3429627 | 0.26072119 | 0.37895 | 2.739486189 | 25.37220579 | 25.37220579 | 0.22221813 | 0.846813483 | 1 | 0.222218127 | 0.222218127 | 0.222218127 | 28.54385834 | 1.455512677 | FALSE | TRUE | FALSE |
| PRKCZ | 6.3280036 | 0.2610096 | 0.2634 | 0.671523173 | 6.328003553 | 13.24522183 | 0.94561724 | 0.593289892 | 0.5062 | 0.945617241 | 0.945617241 | 2.025106217 | 6.691929121 | 0.825551332 | FALSE | TRUE | FALSE |
| LCE3E | 6.3263249 | 0.2612977 | 0.75225 | 6.326324904 | 61.09547767 | 61.09547767 | 0.70438265 | 0.664574144 | 1 | 6.635266088 | 64.14936701 | 64.14936701 | 8.981375254 | 0.953342842 | FALSE | TRUE | FALSE |
| DTNBP1 | 6.3220542 | 0.26158554 | 0.66145 | 6.32205416 | 529.7539556 | 5025.794699 | 0.01806023 | 0.894317484 | 1 | 1.344683434 | 12.05758041 | 12.05758041 | 350.053838 | 2.544134844 | FALSE | TRUE | FALSE |
| IGSF3 | 6.298304 | 0.26187345 | 0.34155 | 6.298303974 | 591.031196 | 1168.389822 | 9.18548522 | 0.209098426 | 0.0277 | 0.978019836 | 9.185485215 | 32.95793881 | 0.685680051 | -0.163878485 | FALSE | FALSE | FALSE |
| BATF | 6.2900503 | 0.26216117 | 0.50405 | 6.290050306 | 6.290050306 | 6.290050306 | 62.0725268 | 0.045460157 | 0.2545 | 6.391931527 | 62.07252677 | 62.07252677 | 0.101333885 | -0.994245306 | FALSE | FALSE | FALSE |
| ZNF608 | 6.2751369 | 0.26244881 | 0.01135 | 0.186489843 | 0.186489843 | 6.2751369 | 0.22753477 | 0.845468314 | 1 | 0.22753477 | 0.22753477 | 4.432301814 | 27.57880437 | 1.440575434 | FALSE | TRUE | FALSE |
| GPR151 | 6.2629863 | 0.26273633 | 0.3148 | 0.703069811 | 57.19162434 | 57.19162434 | 0.14225055 | 0.866372777 | 0.87165 | 0.445892781 | 11.30811876 | 11.30811876 | 44.02785367 | 1.643727514 | FALSE | TRUE | FALSE |
| CLEC5A | 6.2429811 | 0.26302386 | 0.0055 | 6.242981144 | 6.242981144 | 6.242981144 | 6.35011489 | 0.259959402 | 0.74515 | 6.350114886 | 61.43393553 | 61.43393553 | 0.98312885 | -0.007389559 | FALSE | FALSE | FALSE |
| IFITM3 | 6.2426187 | 0.26331106 | 0.50175 | 6.242618687 | 6.242618687 | 6.242618687 | 0.69383799 | 0.668621293 | 1 | 6.560085546 | 6.560085546 | 6.560085546 | 8.997228135 | 0.954108733 | FALSE | TRUE | FALSE |
| TRIM34 | 6.2309767 | 0.26359812 | 0.0126 | 6.230976652 | 6.230976652 | 13.04783762 | 0.71199411 | 0.6601948 | 1 | 0.711994115 | 0.711994115 | 14.13438234 | 8.751444042 | 0.94207972 | FALSE | TRUE | FALSE |
| ATAD3C | 6.2160474 | 0.26388511 | 0.5625 | 6.216047381 | 536.759032 | 3649.072173 | 3.67181749 | 0.373278729 | 0.29635 | 0.918689244 | 34.05343408 | 136.1387955 | 1.692907502 | 0.22863323 | FALSE | FALSE | FALSE |
| CDC42EP5 | 6.2044108 | 0.26417197 | 0.4976 | 0.658722868 | 6.204410762 | 6.204410762 | 0.72820376 | 0.652565978 | 1 | 0.728203758 | 0.728203758 | 0.728203758 | 8.520157567 | 0.930447626 | FALSE | TRUE | FALSE |
| MICB | 6.1916314 | 0.26445871 | 0.00295 | 0.182611548 | 0.182611548 | 3.622236105 | 0.20290322 | 0.851444944 | 1 | 17.35036519 | 166.715856 | 166.715856 | 30.51519522 | 1.484516153 | FALSE | TRUE | FALSE |
| ZNF584 | 6.1841024 | 0.26474525 | 0.0087 | 0.673033704 | 6.184102382 | 6.184102382 | 0.88503836 | 0.600577339 | 0.8712 | 0.885038363 | 77.53692043 | 77.53692043 | 6.987383422 | 0.844314575 | FALSE | TRUE | FALSE |
| SPATC1L | 6.1667197 | 0.26503176 | 0.25585 | 0.677745671 | 6.166719674 | 6.166719674 | 8.39640764 | 0.219885905 | 0.25435 | 0.91566018 | 8.396407643 | 8.396407643 | 0.734447389 | -0.134039309 | FALSE | FALSE | FALSE |
| GABRB2 | 6.1591984 | 0.26531807 | 0.00405 | 0.311354287 | 0.311354287 | 6.159198408 | 0.35355545 | 0.807882968 | 1 | 1.41398 | 1.41398 | 7.003192949 | 17.42074254 | 1.241066662 | FALSE | TRUE | FALSE |
| PTPN6 | 6.1591958 | 0.26560404 | 0.32365 | 3.602000784 | 57.62355016 | 70.96091482 | 0.13103393 | 0.868668659 | 1 | 2.095408859 | 8.382510397 | 10.34021757 | 47.00458866 | 1.672140256 | FALSE | TRUE | FALSE |
| KRTAP9-8 | 6.1564048 | 0.26588973 | 0.0041 | 6.156404768 | 6.156404768 | 6.156404768 | 0.70977345 | 0.661618377 | 1 | 0.709773454 | 0.709773454 | 0.709773454 | 8.67376025 | 0.938207414 | FALSE | TRUE | FALSE |
| PIANP | 6.1560949 | 0.26617509 | 0.50055 | 6.15609492 | 6.15609492 | 6.15609492 | 0.70849622 | 0.662213973 | 1 | 0.708496219 | 0.708496219 | 0.708496219 | 8.688959457 | 0.938967771 | FALSE | TRUE | FALSE |
| APBB2 | 6.1496725 | 0.26646024 | 0.2624 | 6.149672548 | 58.26363919 | 120.9726136 | 0.21434843 | 0.84847795 | 1 | 1.986946434 | 1.986946434 | 7.247220252 | 28.69007523 | 1.457731687 | FALSE | TRUE | FALSE |
| BRD4 | 6.1458473 | 0.26674511 | 0.0105 | 0.182520296 | 0.182520296 | 6.145847308 | 0.22089024 | 0.847001756 | 1 | 0.220890242 | 0.220890242 | 4.296622941 | 27.823082 | 1.444405236 | FALSE | TRUE | FALSE |
| CATSPERB | 6.1406197 | 0.26702975 | 0.50405 | 6.140619654 | 56.56280836 | 533.7727527 | 0.15107723 | 0.864369328 | 0.5073 | 0.151077225 | 0.151077225 | 2.969270294 | 40.64556816 | 1.609013199 | FALSE | TRUE | FALSE |
| ADAM32 | 6.1385606 | 0.26731409 | 0.01285 | 0.383331128 | 6.138560638 | 7.576442144 | 342.425078 | 0.00709848 | 0.4133 | 151.3816529 | 14306.48218 | 139438.7784 | 0.017926726 | -1.746499013 | FALSE | FALSE | FALSE |
| ZNF285 | 6.1093252 | 0.26759861 | 0.01635 | 6.109325235 | 6.109325235 | 21.81424097 | 6.42544786 | 0.257072297 | 0.50415 | 6.425447863 | 6.425447863 | 13.42545209 | 0.950801464 | -0.021910158 | FALSE | FALSE | FALSE |
| SULT1B1 | 6.0379478 | 0.26788405 | 0.12375 | 0.675160459 | 6.037947818 | 6.037947818 | 0.25364974 | 0.838379393 | 0.5025 | 0.253649737 | 0.253649737 | 0.253649737 | 23.80427392 | 1.376654939 | FALSE | TRUE | FALSE |
| MIS12 | 6.0362235 | 0.2681692 | 0.0052 | 6.036223511 | 6.036223511 | 6.036223511 | 0.69691441 | 0.667619113 | 1 | 0.696914412 | 0.696914412 | 0.696914412 | 8.661355547 | 0.937585867 | FALSE | TRUE | FALSE |
| CACNB1 | 6.0300066 | 0.26845412 | 0.35365 | 1.507651826 | 24.12364909 | 96.53319222 | 0.14522861 | 0.865598366 | 0.9925 | 9.287577064 | 148.7116683 | 595.2905736 | 41.52079073 | 1.618265616 | FALSE | TRUE | FALSE |
| CD86 | 6.0290222 | 0.26873873 | 0.50635 | 6.029022228 | 6.029022228 | 6.029022228 | 0.68310018 | 0.673537876 | 1 | 0.683100176 | 0.683100176 | 0.683100176 | 8.825970833 | 0.945762488 | FALSE | TRUE | FALSE |
| SRRT | 6.0202518 | 0.26902316 | 0.00625 | 0.178260046 | 0.178260046 | 3.477699968 | 0.21819704 | 0.847635763 | 0.0272 | 0.21819704 | 0.21819704 | 4.269425655 | 27.59089571 | 1.4407658 | FALSE | TRUE | FALSE |
| CAD | 6.0194817 | 0.26930727 | 0.0974 | 0.005808227 | 24.15382364 | 389.1911444 | 4.5307E-05 | 0.900382221 | 0.4461 | 1.13228E-05 | 0.011643462 | 0.09735839 | 132860.1363 | 5.123394694 | FALSE | TRUE | FALSE |
| OR51I2 | 6.0100093 | 0.26959123 | 0.5045 | 56.48981251 | 544.2701096 | 5372.169666 | 0.10846038 | 0.873742732 | 1 | 0.977723482 | 9.046099368 | 9.046099368 | 55.4120237 | 1.743604011 | FALSE | TRUE | FALSE |
| MTNR1A | 6.0081211 | 0.26987489 | 0.25825 | 0.645966921 | 6.008121106 | 6.008121106 | 0.90087273 | 0.598778088 | 0.50335 | 0.900872727 | 3.60263825 | 3.60263825 | 6.669222996 | 0.824075239 | FALSE | TRUE | FALSE |
| CPLX3 | 6.0004427 | 0.27015835 | 0.4986 | 0.635155202 | 6.000442736 | 6.000442736 | 0.69173352 | 0.669546285 | 1 | 0.691733518 | 0.691733518 | 0.691733518 | 8.674500478 | 0.938244475 | FALSE | TRUE | FALSE |
| PDE7B | 5.9981878 | 0.27044153 | 0.2562 | 1.499857203 | 5.998187805 | 5.998187805 | 0.20930716 | 0.849926876 | 1 | 0.209307159 | 0.209307159 | 4.153470068 | 28.65734656 | 1.457235976 | FALSE | TRUE | FALSE |
| KDELR3 | 5.9975385 | 0.27072439 | 0.3178 | 1.499578987 | 55.33747949 | 55.33747949 | 4.86057913 | 0.313647996 | 0.68775 | 4.860579135 | 450.2330742 | 450.2330742 | 1.23391438 | 0.091285026 | FALSE | FALSE | FALSE |
| RAP1B | 5.9793979 | 0.27100724 | 0.50045 | 5.979397935 | 5.979397935 | 5.979397935 | 6.3161651 | 0.261235833 | 0.50895 | 6.316165103 | 6.316165103 | 6.316165103 | 0.946681703 | -0.023796017 | FALSE | FALSE | FALSE |
| KRTAP9-4 | 5.9768716 | 0.27128982 | 0.8748 | 57.66195916 | 570.564747 | 570.564747 | 0.73706674 | 0.64792487 | 1 | 0.737066737 | 0.737066737 | 0.737066737 | 8.10899656 | 0.908967116 | FALSE | TRUE | FALSE |
| SMAD9 | 5.9668394 | 0.27157224 | 0.8919 | 103.3475896 | 15471.48867 | 15471.48867 | 0.39003919 | 0.795667291 | 0.9814 | 24.94777802 | 399.5058369 | 399.5058369 | 15.29805085 | 1.1846361 | FALSE | TRUE | FALSE |
| SCGB3A1 | 5.9529641 | 0.27185458 | 0.8776 | 57.37214275 | 567.1134727 | 567.1134727 | 6.51725372 | 0.254814053 | 0.75355 | 6.517253724 | 62.95747768 | 62.95747768 | 0.913416037 | -0.039331368 | FALSE | FALSE | FALSE |
| GPR89B | 5.9514183 | 0.27213662 | 0.5064 | 5.951418345 | 5.951418345 | 12.48255191 | 6.08731909 | 0.266953537 | 0.7548 | 6.087319093 | 59.04007081 | 59.04007081 | 0.977674778 | -0.009805588 | FALSE | FALSE | FALSE |
| RAB20 | 5.9513306 | 0.27241834 | 0.1322 | 5.951330561 | 23.81308019 | 23.81308019 | 1.71015299 | 0.516299877 | 0.0058 | 0.427600717 | 1.710152986 | 1.710152986 | 3.47999893 | 0.54157911 | FALSE | FALSE | FALSE |
| MMP9 | 5.9418573 | 0.2726999 | 0.99635 | 100269.1648 | 624170615.8 | 6927277033 | 74.1485007 | 0.038752749 | 0.99995 | 859525661.4 | 1.22734E+13 | 1.9119E+15 | 0.080134557 | -1.096180159 | FALSE | FALSE | FALSE |
| WSCD1 | 5.9394374 | 0.27298118 | 0.51125 | 5.939437396 | 95.0520173 | 380.392623 | 3.93764876 | 0.359093691 | 0.3219 | 0.984362983 | 15.75531834 | 19.42815369 | 1.508371556 | 0.178508334 | FALSE | FALSE | FALSE |
| TMEM87B | 5.924359 | 0.2732624 | 0.0111 | 0.641400437 | 5.924359039 | 12.6994824 | 0.10276307 | 0.875242687 | 1 | 0.102763065 | 0.102763065 | 2.037121424 | 57.65066496 | 1.760804321 | FALSE | TRUE | FALSE |
| APOL1 | 5.8897548 | 0.27354391 | 0.5044 | 5.889754846 | 55.01414532 | 55.01414532 | 0.09830982 | 0.876382019 | 1 | 1.951724333 | 75.92227304 | 75.92227304 | 59.9101359 | 1.777500305 | FALSE | TRUE | FALSE |
| CCZ1 | 5.8653314 | 0.27382552 | 0.6257 | 5.865331435 | 93.86517417 | 93.86517417 | 25.1476325 | 0.110347988 | 0.25365 | 6.286988307 | 25.14763253 | 25.14763253 | 0.233235929 | -0.632204548 | FALSE | FALSE | FALSE |
| FSTL3 | 5.8635868 | 0.27410685 | 0.7486 | 5.86358676 | 56.96144857 | 56.96144857 | 0.69862191 | 0.667043579 | 1 | 0.698621914 | 0.698621914 | 0.698621914 | 8.39307592 | 0.923921152 | FALSE | TRUE | FALSE |
| PPP1R1C | 5.8386403 | 0.27438829 | 0.49955 | 0.621849811 | 5.838640259 | 5.838640259 | 0.67829434 | 0.676074778 | 1 | 0.678294338 | 0.678294338 | 0.678294338 | 8.607826911 | 0.934893525 | FALSE | TRUE | FALSE |
| FLT3LG | 5.83057 | 0.27466954 | 0.5025 | 5.830569969 | 5.830569969 | 5.830569969 | 0.67674549 | 0.676888534 | 1 | 0.676745489 | 0.676745489 | 0.676745489 | 8.615602269 | 0.935285642 | FALSE | TRUE | FALSE |

| | | | | | | | | | | | | | | | | | |
|---|---|---|---|---|---|---|---|---|---|---|---|---|---|---|---|---|---|
| FAM174B | 5.8266964 | 0.27495055 | 0.50355 | 5.82669643 | 5.82669643 | 5.82669643 | 0.6557406 | 0.68708473 | 1 | 6.170624895 | 6.170624895 | 6.170624895 | 8.885672822 | 0.948690318 | FALSE | TRUE | FALSE |
| CHTF8 | 5.8216965 | 0.27523132 | 0.75025 | 5.82169653 | 56.2762732 | 56.2762732 | 0.6966656 | 0.667762665 | 1 | 0.696665602 | 0.696665602 | 0.696665602 | 8.356514967 | 0.922025195 | FALSE | TRUE | FALSE |
| GALR2 | 5.7941449 | 0.27551226 | 0.32525 | 1.448071732 | 23.18980715 | 28.56144538 | 0.12523002 | 0.870091611 | 1 | 0.125230018 | 0.125230018 | 2.474114437 | 46.26801964 | 1.665280911 | FALSE | TRUE | FALSE |
| CCDC86 | 5.7859052 | 0.27579302 | 0.26165 | 0.614408524 | 5.785905168 | 12.15957499 | 0.0838591 | 0.87938393 | 1 | 70.25373736 | 696.4379042 | 7072.508568 | 68.99555304 | 1.8388211 | FALSE | TRUE | FALSE |
| TMEM14C | 5.7813751 | 0.27607354 | 0.49955 | 0.615091124 | 5.781375085 | 5.781375085 | 0.67202204 | 0.679037719 | 1 | 0.672022039 | 0.672022039 | 0.672022039 | 8.602954595 | 0.934647631 | FALSE | TRUE | FALSE |
| JSRP1 | 5.7518302 | 0.27635427 | 0.25565 | 0.630459347 | 5.751830244 | 5.751830244 | 0.09890605 | 0.876227367 | 1 | 0.886529277 | 0.886529277 | 1.961939747 | 58.15448621 | 1.764583223 | FALSE | TRUE | FALSE |
| FGFR1OP2 | 5.7442852 | 0.2766348 | 0.25505 | 1.435738458 | 5.744285198 | 5.744285198 | 0.41376509 | 0.786971216 | 1 | 0.413765093 | 0.413765093 | 0.413765093 | 13.88296231 | 1.142482145 | FALSE | TRUE | FALSE |
| EFCAB12 | 5.7342594 | 0.2769152 | 0.50545 | 5.734259444 | 56.36375978 | 567.5521201 | 0.08736211 | 0.878706581 | 0.75315 | 0.087362108 | 0.822845859 | 0.822845859 | 65.63783253 | 1.817154232 | FALSE | TRUE | FALSE |
| MNDA | 5.7139469 | 0.27719563 | 0.8143 | 53.27309157 | 509.1339195 | 4984.82724 | 8.62591057 | 0.216859359 | 0.25695 | 0.939085016 | 8.625910567 | 8.625910567 | 0.662416664 | -0.178868751 | FALSE | FALSE | FALSE |
| OR2L3 | 5.7120751 | 0.27747578 | 0.5007 | 5.712075103 | 53.22217532 | 53.22217532 | 0.10497263 | 0.874729979 | 1 | 0.104972627 | 0.104972627 | 0.104972627 | 54.41490115 | 1.735717845 | FALSE | TRUE | FALSE |
| SH3GLB1 | 5.7060378 | 0.27775571 | 0.50395 | 5.706037783 | 5.706037783 | 5.706037783 | 0.66204671 | 0.68472897 | 1 | 0.662046708 | 0.662046708 | 0.662046708 | 8.618784313 | 0.935446013 | FALSE | TRUE | FALSE |
| TRIP6 | 5.7022409 | 0.2780354 | 0.4126 | 5.702240869 | 214.9783673 | 860.0633748 | 0.06257826 | 0.884153084 | 1 | 0.250188806 | 0.250188806 | 1.238918765 | 91.12175237 | 1.959622063 | FALSE | TRUE | FALSE |
| SCNN1D | 5.6755129 | 0.27831524 | 0.50855 | 5.675512872 | 52.00891358 | 488.2688326 | 0.14187968 | 0.866395096 | 0.5065 | 0.141879679 | 0.141879679 | 0.556762036 | 40.0022957 | 1.602084916 | FALSE | TRUE | FALSE |
| RPS6KA2 | 5.6723423 | 0.27859482 | 0.50975 | 5.672342322 | 54.72000629 | 111.3156441 | 0.39253586 | 0.794653171 | 0.51365 | 0.392535865 | 0.392535865 | 7.720660142 | 14.45050715 | 1.159883089 | FALSE | TRUE | FALSE |
| ECE1 | 5.6718923 | 0.27887408 | 0.588 | 22.67317442 | 2199.390515 | 7460.278304 | 0.12153955 | 0.870923221 | 0.8801 | 0.485898581 | 1.943041939 | 9.571962327 | 46.66705101 | 1.669010358 | FALSE | TRUE | FALSE |
| ADAM17 | 5.6472194 | 0.27915347 | 0.018 | 0.607437177 | 5.647219373 | 11.95131825 | 0.86291647 | 0.604648009 | 0.5117 | 0.862916466 | 0.862916466 | 17.01108274 | 6.544340732 | 0.815865903 | FALSE | TRUE | FALSE |
| HOXD1 | 5.6445691 | 0.27943259 | 0.1312 | 2.412690618 | 22.57039114 | 22.57039114 | 0.09771463 | 0.876555569 | 1 | 0.097714633 | 0.097714633 | 0.097714633 | 57.76585318 | 1.761671192 | FALSE | TRUE | FALSE |
| MRPS7 | 5.6307507 | 0.27971164 | 0.13145 | 5.63075074 | 22.53141592 | 22.53141592 | 0.40704352 | 0.789505504 | 1 | 1.628004634 | 1.628004634 | 1.628004634 | 13.83328926 | 1.140925458 | FALSE | TRUE | FALSE |
| SH3BGRL2 | 5.6292886 | 0.2799904 | 0.5013 | 5.629288638 | 22.52501284 | 22.52501284 | 1.60924688 | 0.524222219 | 0.7518 | 1.609246882 | 6.43776357 | 6.43776357 | 3.498088889 | 0.543830841 | FALSE | FALSE | FALSE |
| UGT2A2 | 5.6176482 | 0.28026904 | 0.5058 | 54.48766377 | 542.0500052 | 542.0500052 | 60.0371932 | 0.046605491 | 0.25635 | 6.180342381 | 60.0371932 | 60.0371932 | 0.093569468 | -1.028865838 | FALSE | FALSE | FALSE |
| UGT3A1 | 5.6127133 | 0.28054746 | 0.31645 | 5.61271331 | 213.7973077 | 213.7973077 | 3.54251276 | 0.381328192 | 0.00325 | 0.885862247 | 3.542512757 | 3.542512757 | 1.584387607 | 0.199861437 | FALSE | FALSE | FALSE |
| TSR1 | 5.6062133 | 0.28082568 | 0.01035 | 0.170129174 | 5.606213314 | 12.42382597 | 0.12954132 | 0.869067444 | 0.75975 | 0.12954132 | 1.137260776 | 2.554297398 | 43.27741371 | 1.636261299 | FALSE | TRUE | FALSE |
| NPDC1 | 5.5991515 | 0.28110372 | 0.5047 | 5.599151538 | 5.599151538 | 5.599151538 | 0.6527621 | 0.688655769 | 1 | 0.652762103 | 0.652762103 | 0.652762103 | 8.577629603 | 0.933367289 | FALSE | TRUE | FALSE |
| SGTA | 5.5921252 | 0.28138156 | 0.7521 | 5.592125153 | 54.12660761 | 54.12660761 | 0.67196932 | 0.679104387 | 1 | 0.671969324 | 0.671969324 | 0.671969324 | 8.321994696 | 0.920227435 | FALSE | TRUE | FALSE |
| GSK3A | 5.5850708 | 0.28165921 | 0.5062 | 5.585070823 | 5.585070823 | 11.7299082 | 0.64850897 | 0.690457357 | 1 | 2.593991589 | 2.593991589 | 12.87673361 | 8.612172109 | 0.9351127 | FALSE | TRUE | FALSE |
| ATP6V1F | 5.5772177 | 0.28193669 | 0.00425 | 0.590219045 | 5.577217671 | 5.577217671 | 0.64836221 | 0.690580785 | 1 | 0.648362205 | 0.648362205 | 0.648362205 | 8.602009226 | 0.934599904 | FALSE | TRUE | FALSE |
| ZNF664 | 5.5594503 | 0.28221417 | 0.752 | 5.559450333 | 53.74151855 | 53.74151855 | 0.66910678 | 0.680695554 | 1 | 0.669106776 | 0.669106776 | 0.669106776 | 8.308764054 | 0.919536426 | FALSE | TRUE | FALSE |
| SOCS2 | 5.5339442 | 0.28249179 | 0.49885 | 0.586400715 | 5.533944155 | 5.533944155 | 2.54941216 | 0.444078774 | 0.4955 | 0.63738866 | 2.549412158 | 2.549412158 | 2.170674576 | 0.33659472 | FALSE | FALSE | FALSE |
| NKAPL | 5.5304139 | 0.28276916 | 0.68955 | 5.530413923 | 498.3181016 | 498.3181016 | 72.1503582 | 0.039995263 | 0.3165 | 7.639950572 | 698.44349 | 698.44349 | 0.076651233 | -1.115480854 | FALSE | FALSE | FALSE |
| IFNA6 | 5.5290925 | 0.28304624 | 0.00195 | 0.565674655 | 5.529092472 | 5.529092472 | 0.0703618 | 0.882378978 | 0.0038 | 0.070361799 | 0.070361799 | 0.070361799 | 78.58088512 | 1.895316916 | FALSE | TRUE | FALSE |
| TFEC | 5.5271163 | 0.28332305 | 0.0062 | 1.382297199 | 5.527116339 | 5.527116339 | 155.254052 | 0.017443273 | 0.12475 | 15.78184596 | 155.2540517 | 155.2540517 | 0.035600464 | -1.448544337 | FALSE | FALSE | FALSE |
| OR4C3 | 5.526633 | 0.28359955 | 0.50895 | 5.526633029 | 5.526633029 | 5.526633029 | 0.59776885 | 0.713127554 | 1 | 5.672149314 | 5.672149314 | 5.672149314 | 9.245435013 | 0.96592735 | FALSE | TRUE | FALSE |
| COX8C | 5.5171046 | 0.28387591 | 0.4982 | 0.588682251 | 5.51710464 | 5.51710464 | 0.64629637 | 0.691931546 | 1 | 0.646296369 | 0.646296369 | 0.646296369 | 8.536493331 | 0.931279505 | FALSE | TRUE | FALSE |
| HES6 | 5.49834 | 0.2841523 | 0.50605 | 21.99282629 | 212.4941411 | 212.4941411 | 6.13644003 | 0.265372188 | 0.00655 | 6.136440029 | 6.136440029 | 6.136440029 | 0.896014628 | -0.0476849 | FALSE | FALSE | FALSE |
| ZFP36 | 5.4955134 | 0.28442842 | 0.498 | 0.582466879 | 5.49551344 | 11.54337099 | 0.64108143 | 0.693693096 | 1 | 0.641081433 | 0.641081433 | 12.7188987 | 8.57225487 | 0.933095075 | FALSE | TRUE | FALSE |
| ABHD12 | 5.4949575 | 0.28470424 | 0.25215 | 0.60310125 | 5.494957521 | 5.494957521 | 0.09807296 | 0.876478492 | 0.0098 | 0.098072965 | 0.098072965 | 0.098072965 | 56.02927906 | 1.748415034 | FALSE | TRUE | FALSE |
| TRIML1 | 5.4905899 | 0.28497984 | 0.01015 | 0.595176986 | 5.490589862 | 11.7914348 | 0.84564807 | 0.608535267 | 0.51045 | 0.845648067 | 0.845648067 | 1.852820349 | 6.492759905 | 0.812429343 | FALSE | TRUE | FALSE |
| ARL14 | 5.4870854 | 0.28525518 | 0.06555 | 1.371599439 | 21.95657767 | 21.95657767 | 0.11465406 | 0.872444594 | 1 | 1.054870894 | 4.218136583 | 4.218136583 | 47.85775214 | 1.679952296 | FALSE | TRUE | FALSE |
| SIRT5 | 5.4840035 | 0.28553027 | 0.06995 | 0.593583058 | 504.7916376 | 504.7916376 | 7.3669209 | 0.236830749 | 0.5038 | 70.1740933 | 685.1876508 | 6853.709558 | 0.744409176 | -0.128188282 | FALSE | FALSE | FALSE |
| TMEM86A | 5.4808468 | 0.28580511 | 0.0074 | 5.480846779 | 5.480846779 | 5.480846779 | 0.64023116 | 0.694175367 | 1 | 0.640231156 | 0.640231156 | 0.640231156 | 8.560731114 | 0.932510856 | FALSE | TRUE | FALSE |
| HILPDA | 5.4774007 | 0.2860797 | 0.7461 | 5.477400704 | 52.6346311 | 52.6346311 | 0.66302917 | 0.684279056 | 1 | 0.663029171 | 0.663029171 | 0.663029171 | 8.261176038 | 0.917041877 | FALSE | TRUE | FALSE |
| NKPD1 | 5.4589394 | 0.28635431 | 0.69985 | 5.45893936 | 498.5502817 | 498.5502817 | 0.10250213 | 0.875340832 | 1 | 0.102502135 | 0.102502135 | 2.019026866 | 53.25683584 | 1.72637536 | FALSE | TRUE | FALSE |
| TMEM150B | 5.448686 | 0.2866288 | 0.2517 | 1.362243217 | 5.448685994 | 5.448685994 | 0.19478755 | 0.853268094 | 0.00605 | 0.194787551 | 0.194787551 | 0.194787551 | 27.97245498 | 1.446730584 | FALSE | TRUE | FALSE |
| TXNDC11 | 5.4137606 | 0.2869036 | 0.0181 | 0.590354821 | 5.413760617 | 11.58815082 | 0.78806814 | 0.624924856 | 0.51685 | 7.270584697 | 68.80035017 | 143.0339098 | 6.869660566 | 0.836935279 | FALSE | TRUE | FALSE |
| GMPS | 5.4127361 | 0.28717812 | 0.1311 | 2.311005679 | 21.64883223 | 21.64883223 | 0.1916626 | 0.853945868 | 1 | 0.191662604 | 0.191662604 | 3.792456514 | 28.24096084 | 1.450879469 | FALSE | TRUE | FALSE |
| CMTM4 | 5.4011607 | 0.28745253 | 0.0043 | 5.401160701 | 5.401160701 | 5.401160701 | 0.63401949 | 0.697917178 | 1 | 0.634019486 | 0.634019486 | 0.634019486 | 8.518919086 | 0.930384493 | FALSE | TRUE | FALSE |
| LRRC10 | 5.396757 | 0.28772671 | 0.503 | 5.396757024 | 5.396757024 | 5.396757024 | 0.63203087 | 0.698385374 | 1 | 0.632030874 | 0.632030874 | 0.632030874 | 8.538755379 | 0.931394572 | FALSE | TRUE | FALSE |
| MYL3 | 5.3955708 | 0.28800061 | 0.68995 | 12.97671068 | 511.7155278 | 511.7155278 | 0.19672367 | 0.852893486 | 1 | 0.786584353 | 3.145882873 | 3.145882873 | 27.42715603 | 1.438180777 | FALSE | TRUE | FALSE |
| LANCL1 | 5.3940119 | 0.28827424 | 0.85885 | 21.58801891 | 346.0508915 | 1827.039217 | 0.11105442 | 0.873137269 | 0.99805 | 28.45258494 | 2499.86011 | 10015.15229 | 48.57088945 | 1.686376057 | FALSE | TRUE | FALSE |
| HIVEP3 | 5.3841915 | 0.28854773 | 0.3586 | 5.384191498 | 516.5604544 | 1740.108152 | 0.09429153 | 0.877264364 | 0.52945 | 0.872180334 | 1.802466832 | 16.67250566 | 57.10153719 | 1.7566478 | FALSE | TRUE | FALSE |
| CYP2F1 | 5.3808854 | 0.28882098 | 0.69565 | 5.380885446 | 508.5353925 | 508.5353925 | 0.09839654 | 0.876353049 | 1 | 0.098396535 | 0.098396535 | 1.94409326 | 54.68572065 | 1.73787394 | FALSE | TRUE | FALSE |
| MATN4 | 5.3703278 | 0.28909411 | 0.7902 | 126.9237339 | 20436.21517 | 210904.7274 | 2481.00558 | 0.000950363 | 0.24355 | 245.2191319 | 39642.83547 | 410649.4649 | 0.002164577 | -2.664626949 | FALSE | FALSE | FALSE |
| ZMYM2 | 5.3561717 | 0.28936719 | 0.00065 | 0.159446253 | 0.159446253 | 5.356171732 | 0.19780742 | 0.852611334 | 1 | 0.197807419 | 0.197807419 | 6.654398842 | 27.07770906 | 1.432611918 | FALSE | TRUE | FALSE |
| CLEC2L | 5.3495179 | 0.28964008 | 0.7518 | 5.349517895 | 51.57486249 | 51.57486249 | 0.62840999 | 0.699723044 | 1 | 0.628409988 | 5.916270241 | 5.916270241 | 8.512783056 | 0.930071566 | FALSE | TRUE | FALSE |
| LSM14B | 5.3426927 | 0.28991279 | 0.50445 | 5.342692713 | 5.342692713 | 11.18247188 | 0.62396091 | 0.701620711 | 1 | 0.623960909 | 0.623960909 | 12.36733209 | 8.562543963 | 0.932602814 | FALSE | TRUE | FALSE |
| RPH3AL | 5.3424743 | 0.2901852 | 0.0077 | 0.577143509 | 5.342474253 | 5.342474253 | 0.09072774 | 0.877984636 | 0.5019 | 0.823820181 | 0.823820181 | 0.823820181 | 58.88468048 | 1.770002323 | FALSE | TRUE | FALSE |
| SPINK13 | 5.3291164 | 0.29045755 | 0.49975 | 0.568746913 | 5.329116435 | 5.329116435 | 0.62693715 | 0.700416004 | 1 | 0.626937153 | 0.626937153 | 0.626937153 | 8.500240276 | 0.929431202 | FALSE | TRUE | FALSE |
| NANOS3 | 5.3225475 | 0.29072972 | 0.50275 | 5.322547507 | 5.322547507 | 5.322547507 | 0.62507916 | 0.701048332 | 1 | 0.625079163 | 0.625079163 | 0.625079163 | 8.514997492 | 0.930184524 | FALSE | TRUE | FALSE |
| SIGLEC7 | 5.3141447 | 0.29100173 | 0.5055 | 5.314144705 | 49.63024073 | 49.63024073 | 0.09820412 | 0.876391674 | 0.0117 | 0.098204119 | 0.098204119 | 1.948085742 | 54.11325658 | 1.733303671 | FALSE | TRUE | FALSE |
| NDUFB6 | 5.2979624 | 0.29127373 | 0.5036 | 5.297962413 | 5.297962413 | 5.297962413 | 0.62389806 | 0.70167782 | 1 | 0.623898064 | 0.623898064 | 0.623898064 | 8.491711583 | 0.928995235 | FALSE | TRUE | FALSE |
| RAP2A | 5.2977983 | 0.29154543 | 0.2576 | 5.297798314 | 21.1886823 | 21.1886823 | 2.47171064 | 0.449250343 | 0.50195 | 2.471710638 | 2.471710638 | 2.471710638 | 2.143373189 | 0.331097794 | FALSE | FALSE | FALSE |
| CACNB4 | 5.2952129 | 0.29181687 | 0.3272 | 1.32368058 | 21.18810728 | 26.20651017 | 6.94416387 | 0.24571107 | 0.35285 | 6.944163866 | 111.219563 | 445.2710792 | 0.762541468 | -0.117736534 | FALSE | FALSE | FALSE |
| ZBTB34 | 5.2802437 | 0.29208828 | 0.0154 | 5.280243716 | 5.280243716 | 11.05116826 | 0.61828313 | 0.704447894 | 1 | 0.618283132 | 0.618283132 | 12.23641585 | 8.540171072 | 0.93146657 | FALSE | TRUE | FALSE |
| ARHGEF25 | 5.2686859 | 0.2923596 | 0.51165 | 5.268685868 | 49.55804577 | 49.55804577 | 3.33762338 | 0.392611373 | 0.26395 | 0.834466671 | 3.337623378 | 7.32536121 | 1.578574114 | 0.198264977 | FALSE | FALSE | FALSE |
| KRTAP5-8 | 5.2659145 | 0.29263067 | 0.0047 | 0.561939415 | 5.265914482 | 5.265914482 | 0.56470692 | 0.726761153 | 1 | 50.87032244 | 501.6828069 | 501.6828069 | 9.32503975 | 0.969650692 | FALSE | TRUE | FALSE |
| ALKBH8 | 5.2589451 | 0.29290156 | 0.50795 | 5.258945138 | 49.50324966 | 49.50324966 | 0.19456526 | 0.853321466 | 1 | 0.194565258 | 0.194565258 | 0.194565258 | 27.02920964 | 1.431833347 | FALSE | TRUE | FALSE |
| GYPB | 5.2579543 | 0.29317217 | 0.49895 | 0.561795638 | 5.257954303 | 5.257954303 | 0.60095325 | 0.711913193 | 1 | 0.600953248 | 5.634083905 | 5.634083905 | 8.749356658 | 0.94197612 | FALSE | TRUE | FALSE |
| CACNA2D1 | 5.2558041 | 0.29344252 | 0.26315 | 1.314109036 | 5.255804101 | 25.7957627 | 0.1864624 | 0.85538735 | 0.0241 | 0.186462396 | 0.186462396 | 3.668385733 | 28.18693851 | 1.450047908 | FALSE | TRUE | FALSE |
| RRP8 | 5.2545569 | 0.2937126 | 0.5095 | 49.2667046 | 473.5014005 | 4662.092455 | 667.681953 | 0.003158725 | 0.19195 | 68.97011717 | 667.6819527 | 6621.626935 | 0.00786985 | -2.10403354 | FALSE | FALSE | FALSE |
| OCM2 | 5.2530036 | 0.2939824 | 0.003 | 5.253003578 | 5.253003578 | 5.253003578 | 0.59491605 | 0.714383303 | 1 | 5.634847538 | 5.634847538 | 5.634847538 | 8.829823286 | 0.945952012 | FALSE | TRUE | FALSE |
| TSGA13 | 5.2514448 | 0.29425194 | 0.69065 | 5.251444814 | 464.3753338 | 464.3753338 | 60430.7489 | 3.31492E-05 | 0.00085 | 63.90020674 | 6011.929154 | 60430.7489 | 8.69002E-05 | -4.06097917 | FALSE | FALSE | FALSE |
| NUP214 | 5.247839 | 0.29452124 | 0.01585 | 0.063218624 | 5.247838955 | 10.95316891 | 1.37172609 | 0.548742173 | 0.14335 | 0.150537376 | 1.371726094 | 5.053897137 | 3.825719273 | 0.582713099 | FALSE | FALSE | FALSE |
| CYP27A1 | 5.2432066 | 0.29479033 | 0.2389 | 1.310446062 | 20.98375698 | 83.99974138 | 0.0390226 | 0.889173008 | 0.6937 | 0.156030914 | 2.496462468 | 2.496462468 | 134.3633204 | 2.128280728 | FALSE | TRUE | FALSE |

| | | | | | | | | | | | | | | | | | |
|---|---|---|---|---|---|---|---|---|---|---|---|---|---|---|---|---|---|
| CCL25 | 5.232294 | 0.29505931 | 0.49925 | 0.558475418 | 5.23229404 | 5.23229404 | 0.61690791 | 0.704950771 | 1 | 0.616907906 | 0.616907906 | 0.616907906 | 8.481483196 | 0.928471806 | FALSE | TRUE | FALSE |
| KCNJ1 | 5.2300394 | 0.29532805 | 0.46445 | 3.135750727 | 83.6116293 | 248.8069885 | 0.06425959 | 0.88374005 | 0.8791 | 0.600776348 | 5.764676731 | 5.764676731 | 81.38924418 | 1.910567015 | FALSE | TRUE | FALSE |
| RPN1 | 5.192166 | 0.29559716 | 0.3774 | 2.217922717 | 20.7660523 | 20.7660523 | 0.18075173 | 0.856928767 | 1 | 0.722767327 | 2.890833806 | 6.124230176 | 28.72540162 | 1.458266109 | FALSE | TRUE | FALSE |
| TRIT1 | 5.1857179 | 0.29586609 | 0.818 | 48.99414423 | 474.4898461 | 4707.590209 | 0.74819065 | 0.642228304 | 0.9843 | 263.3093256 | 25580.45298 | 86878.48465 | 6.931011293 | 0.840796606 | FALSE | TRUE | FALSE |
| MCF2L2 | 5.175885 | 0.2961349 | 0.01615 | 0.574363614 | 5.175884958 | 11.23023119 | 0.01448262 | 0.895297489 | 1 | 0.014482622 | 0.014482622 | 0.283934453 | 357.3859017 | 2.553137416 | FALSE | TRUE | FALSE |
| LDHAL6A | 5.1672952 | 0.29640357 | 0.0632 | 0.560625764 | 20.65583807 | 20.65583807 | 0.05442169 | 0.885795743 | 0.75295 | 0.054421694 | 0.217616392 | 0.217616392 | 94.94918001 | 1.977491218 | FALSE | TRUE | FALSE |
| LYPLA2 | 5.1367029 | 0.29667249 | 0.0059 | 5.136702907 | 5.136702907 | 5.136702907 | 49.8404339 | 0.055049708 | 0.1296 | 49.84043393 | 492.2055636 | 492.2055636 | 0.103062965 | -0.986897367 | FALSE | FALSE | FALSE |
| FAM131A | 5.1286227 | 0.29694126 | 0.7596 | 5.128622676 | 49.42867214 | 49.42867214 | 0.60515888 | 0.709882755 | 1 | 5.695537091 | 5.695537091 | 5.695537091 | 8.474836676 | 0.928131337 | FALSE | TRUE | FALSE |
| NDUFS8 | 5.1042433 | 0.29721017 | 0.99995 | 334452.6043 | 145019551.4 | 718197517.9 | 7.44590602 | 0.2364994 | 0.99675 | 7651.269042 | 1215776.345 | 4873901.128 | 0.685510038 | -0.163986181 | FALSE | FALSE | FALSE |
| KISS1 | 5.0945304 | 0.29747896 | 0.5039 | 5.094530428 | 5.094530428 | 5.094530428 | 0.58166829 | 0.720043809 | 1 | 5.482381773 | 5.482381773 | 5.482381773 | 8.758480631 | 0.942428774 | FALSE | TRUE | FALSE |
| HRC | 5.0826732 | 0.29774767 | 0.9899 | 4466.992093 | 4855410.548 | 52207248.62 | 0.10626134 | 0.874303 | 0.87745 | 8.835001763 | 83.71205875 | 83.71205875 | 47.8318197 | 1.679716903 | FALSE | TRUE | FALSE |
| C6 | 5.0820358 | 0.2980161 | 0.23175 | 0.574050357 | 46.06857436 | 171.4553979 | 0.09665843 | 0.87677667 | 0.50465 | 0.09665843 | 1.836317032 | 7.339620452 | 52.57726424 | 1.720797985 | FALSE | TRUE | FALSE |
| PTCD1 | 5.0669376 | 0.2982845 | 0.5092 | 5.066937586 | 5.066937586 | 99.64924414 | 2.22751023 | 0.46974916 | 0.9354 | 21.11653608 | 338.1021275 | 416.1231836 | 2.274709012 | 0.356925848 | FALSE | FALSE | FALSE |
| NUMBL | 5.0626717 | 0.29855269 | 0.5069 | 5.062671675 | 5.062671675 | 18.12229165 | 5.45572083 | 0.288560988 | 0.5014 | 5.455720832 | 5.455720832 | 19.52925208 | 0.927956512 | -0.032472376 | FALSE | FALSE | FALSE |
| KHNYN | 5.0550798 | 0.29882072 | 0.26985 | 0.551040484 | 5.055079784 | 10.84816002 | 0.09024138 | 0.878107069 | 1 | 0.090241377 | 0.090241377 | 1.779903555 | 56.01731663 | 1.748322301 | FALSE | TRUE | FALSE |
| DENND1C | 5.0487835 | 0.29908857 | 0.6929 | 5.048783522 | 455.0600034 | 455.0600034 | 62.4118241 | 0.045234202 | 0.65555 | 62.41182414 | 5972.358796 | 60543.77003 | 0.080894664 | -1.092080126 | FALSE | FALSE | FALSE |
| SLC19A1 | 5.046853 | 0.29935617 | 0.7377 | 20.18122327 | 1840.157013 | 9037.083455 | 0.26024627 | 0.836654156 | 0.5028 | 0.260246269 | 0.260246269 | 0.974297961 | 19.39260477 | 1.287636146 | FALSE | TRUE | FALSE |
| STK33 | 5.0246803 | 0.29962387 | 0.77495 | 461.6751752 | 46722.84876 | 91070.08692 | 0.10346907 | 0.875075503 | 1 | 0.103469069 | 0.103469069 | 0.103469069 | 48.56214835 | 1.686297891 | FALSE | TRUE | FALSE |
| C7orf33 | 5.024378 | 0.29989128 | 0.504 | 5.024377965 | 5.024377965 | 5.024377965 | 5.4132961 | 0.289762357 | 0.5084 | 5.413296097 | 5.413296097 | 5.413296097 | 0.928155023 | -0.032379481 | FALSE | FALSE | FALSE |
| TTC23L | 5.0217445 | 0.30015845 | 0.25465 | 0.551532608 | 5.021744454 | 5.021744454 | 0.09067601 | 0.878022332 | 1 | 0.090676015 | 0.090676015 | 0.090676015 | 55.38117739 | 1.743362184 | FALSE | TRUE | FALSE |
| DNAJC28 | 5.0214704 | 0.30042533 | 0.008 | 0.547208813 | 5.021470422 | 5.021470422 | 0.08702218 | 0.878771829 | 0.50175 | 0.784068226 | 0.784068226 | 0.784068226 | 57.70333996 | 1.761200952 | FALSE | TRUE | FALSE |
| VMP1 | 5.0210028 | 0.30069194 | 0.50175 | 5.021002802 | 5.021002802 | 5.021002802 | 0.52196741 | 0.746284063 | 1 | 47.46347399 | 4776.235627 | 4776.235627 | 9.619379804 | 0.983147072 | FALSE | TRUE | FALSE |
| KHDRBS3 | 5.0135179 | 0.30095838 | 0.7521 | 5.013517882 | 48.38421177 | 48.38421177 | 0.61165326 | 0.706999646 | 1 | 0.611653261 | 0.611653261 | 0.611653261 | 8.196666643 | 0.913637273 | FALSE | TRUE | FALSE |
| VLDLR | 5.0105945 | 0.30122459 | 0.41175 | 2.225979106 | 142.401058 | 1281.332973 | 0.10474681 | 0.874799223 | 0.63205 | 0.418617485 | 26.76071073 | 107.0552212 | 47.83529406 | 1.679748448 | FALSE | TRUE | FALSE |
| GTF2H2C | 5.0078389 | 0.30149056 | 0.753 | 5.007838883 | 48.67436728 | 48.67436728 | 0.58587296 | 0.718234056 | 1 | 0.585872964 | 0.585872964 | 0.585872964 | 8.547653148 | 0.931846891 | FALSE | TRUE | FALSE |
| TUBA1A | 5.0044643 | 0.30175631 | 0.9384 | 48.5060104 | 4913.530689 | 4913.530689 | 0.58971617 | 0.716809483 | 0.8754 | 54.15286806 | 539.1988402 | 539.1988402 | 8.486225235 | 0.928714554 | FALSE | TRUE | FALSE |
| GPR137B | 5.0035784 | 0.30202178 | 0.69185 | 5.003578438 | 458.950609 | 458.950609 | 62.5280102 | 0.044783141 | 0.6642 | 62.52801019 | 6077.253891 | 62087.45769 | 0.080021392 | -1.096793896 | FALSE | FALSE | FALSE |
| TCP11L2 | 5.0023565 | 0.30228698 | 0.2563 | 0.548401973 | 5.002356479 | 10.86839277 | 0.08749289 | 0.878687928 | 1 | 0.784109464 | 0.784109464 | 1.735854706 | 57.17443603 | 1.757201889 | FALSE | TRUE | FALSE |
| MMP25 | 5.0021348 | 0.3025519 | 0.81675 | 47.39301499 | 460.2773564 | 4579.443793 | 6.79402069 | 0.248650278 | 0.8923 | 64.70703652 | 6317.833418 | 64692.72246 | 0.736255452 | -0.132971476 | FALSE | FALSE | FALSE |
| AGAP1 | 5.0021023 | 0.30281654 | 1 | 1.21494E+11 | 2.51342E+15 | 1.01916E+17 | 24.8535569 | 0.110648651 | 0.9848 | 265008.5151 | 347402569.1 | 4000161663 | 0.201263036 | -0.696235981 | FALSE | FALSE | FALSE |
| TEK | 4.9954325 | 0.30308101 | 0.69955 | 4.99543252 | 496.1051314 | 962.3075846 | 0.75796552 | 0.638234551 | 0.8834 | 7.293976274 | 71.99125751 | 143.1171776 | 6.590580183 | 0.818923648 | FALSE | TRUE | FALSE |
| FPR1 | 4.992615 | 0.30334524 | 0.5112 | 4.992614981 | 19.97782257 | 19.97782257 | 0.35482853 | 0.807448705 | 1 | 3.321110498 | 13.283003 | 13.283003 | 14.07050048 | 1.148309545 | FALSE | TRUE | FALSE |
| MRPL36 | 4.9902066 | 0.30360923 | 0.7516 | 4.990206645 | 48.04130533 | 48.04130533 | 0.59119157 | 0.715834025 | 0.5008 | 5.55861571 | 5.55861571 | 5.55861571 | 8.440929991 | 0.926390298 | FALSE | TRUE | FALSE |
| KIF3B | 4.9868014 | 0.30387299 | 0.5081 | 4.986801404 | 24.53940366 | 98.16124761 | 0.11024165 | 0.87326892 | 1 | 0.440793145 | 0.440793145 | 8.693156058 | 45.23518435 | 1.655476365 | FALSE | TRUE | FALSE |
| FAM209B | 4.9848172 | 0.30413651 | 0.4993 | 0.532589605 | 4.984817211 | 4.984817211 | 5.37572917 | 0.290960782 | 0.49615 | 0.573374932 | 5.37572917 | 5.37572917 | 0.927282058 | -0.032788143 | FALSE | FALSE | FALSE |
| RRAGD | 4.9587503 | 0.3044002 | 0.5059 | 4.958750278 | 4.958750278 | 4.958750278 | 0.58438181 | 0.71894076 | 1 | 0.584381809 | 0.584381809 | 0.584381809 | 8.485463098 | 0.928675549 | FALSE | TRUE | FALSE |
| MYOC | 4.9462974 | 0.30466384 | 0.9109 | 463.369856 | 47706.33857 | 500834.6341 | 0.11546531 | 0.872280617 | 0.88675 | 10.00268868 | 956.7025133 | 9695.96823 | 42.83795303 | 1.63182871 | FALSE | TRUE | FALSE |
| RFPL3 | 4.9397859 | 0.30492731 | 0.00725 | 0.542267813 | 4.939785918 | 4.939785918 | 0.0893091 | 0.878285616 | 1 | 0.089309098 | 0.089309098 | 0.089309098 | 55.31111622 | 1.742812423 | FALSE | TRUE | FALSE |
| MMP21 | 4.9372274 | 0.30519053 | 0.69045 | 11.51871744 | 440.9820623 | 440.9820623 | 13.4666434 | 0.166839555 | 0.6577 | 129.1929583 | 12798.45794 | 132005.5563 | 0.36662643 | -0.43577623 | FALSE | FALSE | FALSE |
| TIMM8B | 4.931725 | 0.30545358 | 0.7527 | 4.931724998 | 47.43109897 | 47.43109897 | 0.60421361 | 0.710312735 | 1 | 0.604213609 | 0.604213609 | 0.604213609 | 8.162220978 | 0.911808348 | FALSE | TRUE | FALSE |
| TMPRSS11B | 4.9023589 | 0.30571686 | 0.318 | 1.225782255 | 45.21500698 | 45.21500698 | 67.8044007 | 0.041690375 | 0.0038 | 1.826600545 | 67.80440068 | 67.80440068 | 0.072301486 | -1.140852776 | FALSE | FALSE | FALSE |
| TLCD1 | 4.898835 | 0.30597993 | 0.2537 | 0.527832417 | 4.898835021 | 4.898835021 | 0.32114904 | 0.818392456 | 0.74995 | 0.730283034 | 27.25516707 | 27.25516707 | 15.25408602 | 1.183386191 | FALSE | TRUE | FALSE |
| SLC25A44 | 4.8969176 | 0.30624274 | 0.3846 | 3.943385272 | 36.89458423 | 36.89458423 | 0.30682862 | 0.82320216 | 0.0102 | 0.306828624 | 0.306828624 | 6.100207571 | 15.95978093 | 1.203026926 | FALSE | TRUE | FALSE |
| GEMIN6 | 4.8877264 | 0.30650544 | 0.5026 | 4.887726361 | 4.887726361 | 4.887726361 | 0.5818522 | 0.719943909 | 1 | 0.581852202 | 0.581852202 | 0.581852202 | 8.400288494 | 0.924294201 | FALSE | TRUE | FALSE |
| TGM2 | 4.8811666 | 0.30676798 | 0.6388 | 11.49959954 | 1095.296829 | 8700.203191 | 0.93999342 | 0.594267876 | 0.8724 | 18.80725216 | 2797.218348 | 13755.85471 | 5.192766741 | 0.715398815 | FALSE | TRUE | FALSE |
| RBM14 | 4.8790669 | 0.30703027 | 0.9814 | 5059.302264 | 583761.0211 | 6487351.051 | 801931.156 | 2.13035E-06 | 0.06 | 6943.268853 | 8916256.141 | 101339305.9 | 6.08415E-06 | -5.215800314 | FALSE | FALSE | FALSE |
| OLFM3 | 4.8754966 | 0.30729234 | 0.0748 | 1.218956699 | 19.50553886 | 19.50553886 | 1.60175928 | 0.52566465 | 0.81495 | 6.407031904 | 59.92419751 | 239.7564902 | 3.043838561 | 0.483421615 | FALSE | FALSE | FALSE |
| DDI1 | 4.8745086 | 0.30755415 | 0.2553 | 0.532121115 | 4.874508587 | 10.53372196 | 0.08756996 | 0.878669269 | 1 | 0.087569955 | 0.087569955 | 1.73564041 | 55.66416685 | 1.745575713 | FALSE | TRUE | FALSE |
| ANKRD36 | 4.8705666 | 0.30781575 | 1 | 1.30998E+42 | 7.64896E+51 | 4.25169E+55 | 109.204591 | 0.02664865 | 1 | 2.1696E+45 | 1.03371E+54 | 6.93448E+57 | 0.044600383 | -1.350661408 | FALSE | FALSE | FALSE |
| TVP23C | 4.8695673 | 0.30807708 | 0.74705 | 4.869567336 | 46.80816995 | 46.80816995 | 5.40476926 | 0.290361943 | 0.49635 | 0.57560992 | 5.404769265 | 5.404769265 | 0.900975989 | -0.045286783 | FALSE | FALSE | FALSE |
| OSM | 4.8628136 | 0.30833826 | 0.5063 | 4.862813587 | 4.862813587 | 4.862813587 | 0.5786275 | 0.721385127 | 1 | 0.578627497 | 0.578627497 | 0.578627497 | 8.404048568 | 0.924488554 | FALSE | TRUE | FALSE |
| OR10H5 | 4.8507403 | 0.30859937 | 0.0103 | 4.850740273 | 4.850740273 | 10.25256407 | 0.57620278 | 0.72212455 | 1 | 0.576202775 | 0.576202775 | 0.576202775 | 8.418460445 | 0.925232676 | FALSE | TRUE | FALSE |
| C8orf76 | 4.8481855 | 0.30886025 | 0.502 | 4.848185514 | 4.848185514 | 4.848185514 | 0.57649894 | 0.721976992 | 1 | 0.576498939 | 0.576498939 | 0.576498939 | 8.409704136 | 0.924780717 | FALSE | TRUE | FALSE |
| UMPS | 4.8477612 | 0.30912086 | 0.25365 | 0.51107528 | 4.847761203 | 10.11842982 | 0.08021044 | 0.880255144 | 1 | 0.744305353 | 0.744305353 | 1.589957975 | 60.43803223 | 1.781310316 | FALSE | TRUE | FALSE |
| OR2A2 | 4.8329807 | 0.30938146 | 0.00865 | 4.832980729 | 4.832980729 | 4.832980729 | 0.5739572 | 0.722908797 | 1 | 0.573957196 | 0.573957196 | 0.573957196 | 8.420454987 | 0.925335559 | FALSE | TRUE | FALSE |
| U2AF2 | 4.8202194 | 0.309642 | 0.5093 | 4.820219375 | 4.820219375 | 17.32094676 | 0.5714555 | 0.723931082 | 1 | 0.571455496 | 0.571455496 | 11.27953704 | 8.434986465 | 0.92608439 | FALSE | TRUE | FALSE |
| MPND | 4.8117727 | 0.30990242 | 0.13335 | 4.811772746 | 45.4625888 | 45.4625888 | 6.56016821 | 0.253195446 | 0.69445 | 6.56016821 | 606.9523763 | 606.9523763 | 0.733483135 | -0.134609867 | FALSE | FALSE | FALSE |
| FAM89A | 4.8092158 | 0.31016261 | 0.8734 | 46.15617874 | 454.3470866 | 454.3470866 | 493.776339 | 0.004434811 | 0.0018 | 5.20772647 | 493.776339 | 493.776339 | 0.009739664 | -2.011456011 | FALSE | FALSE | FALSE |
| OR6C65 | 4.8048451 | 0.31042259 | 0.8142 | 10.5771747 | 423.5293604 | 4124.380254 | 0.09345583 | 0.877445153 | 1 | 0.09345583 | 0.8330687 | 0.8330687 | 51.41300557 | 1.711072993 | FALSE | TRUE | FALSE |
| RIMKLB | 4.7968844 | 0.31068244 | 0.51005 | 4.796884427 | 4.796884427 | 10.11045144 | 8.79764193 | 0.213823815 | 0.9641 | 3959.072239 | 434944.4976 | 4711833.583 | 0.545246609 | -0.263407027 | FALSE | FALSE | FALSE |
| SEC14L3 | 4.7711116 | 0.31094248 | 0.2838 | 1.193424436 | 19.07887571 | 76.31229586 | 3.58810601 | 0.378981244 | 0.13425 | 0.401077088 | 3.588106013 | 3.588106013 | 1.32970195 | 0.123754306 | FALSE | FALSE | FALSE |
| PCDHA9 | 4.7677123 | 0.3112023 | 0.32005 | 0.520158066 | 44.79641811 | 44.79641811 | 0.11444701 | 0.872495756 | 0.51025 | 0.114447009 | 0.114447009 | 2.236266355 | 41.65868901 | 1.619705599 | FALSE | TRUE | FALSE |
| HES7 | 4.7669784 | 0.31146186 | 0.50245 | 4.766978395 | 4.766978395 | 4.766978395 | 0.55893193 | 0.729158705 | 1 | 2.235433256 | 2.235433256 | 2.235433256 | 8.528727962 | 0.930884262 | FALSE | TRUE | FALSE |
| STMN3 | 4.7638805 | 0.31172119 | 0.49735 | 0.505908774 | 4.763880473 | 4.763880473 | 0.56594999 | 0.726142792 | 1 | 0.565949993 | 0.565949993 | 0.565949993 | 8.417493657 | 0.925182798 | FALSE | TRUE | FALSE |
| DDAH1 | 4.760262 | 0.31198032 | 0.0043 | 0.506631702 | 4.760261974 | 4.760261974 | 0.56009846 | 0.728738795 | 1 | 2.24026503 | 2.24026503 | 2.24026503 | 8.498973569 | 0.929366479 | FALSE | TRUE | FALSE |
| TMEM45A | 4.7369737 | 0.31223958 | 0.5014 | 4.736973749 | 4.736973749 | 4.736973749 | 5.14074263 | 0.302175104 | 0.49995 | 0.547444289 | 5.140742633 | 5.140742633 | 0.92145709 | -0.035524884 | FALSE | FALSE | FALSE |
| SOAT2 | 4.7287004 | 0.31249872 | 0.47825 | 2.641617684 | 168.9796845 | 1429.27634 | 2.46373807 | 0.450575732 | 0.0082 | 0.278927831 | 2.463738073 | 2.463738073 | 1.919319433 | 0.283147261 | FALSE | FALSE | FALSE |
| SERPINF1 | 4.719679 | 0.31275775 | 0.503 | 4.719678969 | 75.52820241 | 302.2527259 | 1.83186148 | 0.504890491 | 0.3797 | 0.802221243 | 7.32589158 | 7.32589158 | 2.576438793 | 0.41101983 | FALSE | FALSE | FALSE |
| FAM83E | 4.7045061 | 0.31301677 | 0.25915 | 0.517489581 | 4.704506071 | 10.21892932 | 6.5388415 | 0.254490765 | 0.13525 | 2.854478289 | 26.15477198 | 26.15477198 | 0.719470884 | -0.142986777 | FALSE | FALSE | FALSE |
| ARSJ | 4.6865285 | 0.31327585 | 0.6036 | 18.7412101 | 1625.375716 | 6501.425402 | 38.322253 | 0.071242045 | 0.7729 | 613.4448215 | 39361.81704 | 157661.2705 | 0.122292615 | -0.912599767 | FALSE | FALSE | FALSE |
| NUDT14 | 4.6582454 | 0.31353515 | 0.5054 | 4.658245411 | 18.63273141 | 18.63273141 | 2.20441044 | 0.471686355 | 0.7518 | 2.204410437 | 8.819708654 | 8.819708654 | 2.113147956 | 0.324929906 | FALSE | FALSE | FALSE |
| ANKRD52 | 4.6516656 | 0.3137943 | 0.2738 | 1.162757682 | 4.65166556 | 39.2363161 | 21.0748032 | 0.127678382 | 0.3299 | 5.266634101 | 84.35336543 | 177.8790947 | 0.220721661 | -0.656155045 | FALSE | FALSE | FALSE |
| RNASE2 | 4.6494075 | 0.31405321 | 0.68585 | 44.61271964 | 4429.111095 | 4429.111095 | 463.117536 | 0.004927005 | 0.18545 | 47.01390747 | 4676.121915 | 48367.28433 | 0.010039368 | -1.998293612 | FALSE | FALSE | FALSE |

| | | | | | | | | | | | | | | | | | |
|---|---|---|---|---|---|---|---|---|---|---|---|---|---|---|---|---|---|
| FBXO22 | 4.6493431 | 0.31431186 | 0.50485 | 4.649343128 | 4.649343128 | 4.649343128 | 0.5540189 | 0.731649544 | 1 | 0.554018897 | 0.554018897 | 0.554018897 | 8.392029856 | 0.92386702 | FALSE | TRUE | FALSE |
| ESR2 | 4.6469184 | 0.31457027 | 0.4389 | 2.044381425 | 1198.163396 | 8489.597929 | 1250.84598 | 0.001984485 | 0.03825 | 3.81130432 | 1250.845978 | 11761.54524 | 0.00371502 | -2.430038792 | FALSE | FALSE | FALSE |
| TSGA10 | 4.6442688 | 0.31482846 | 0.2624 | 0.503875395 | 4.644268822 | 9.947000497 | 0.08071993 | 0.880118361 | 0.50985 | 0.730150094 | 0.730150094 | 2.726289058 | 57.53558929 | 1.759936566 | FALSE | TRUE | FALSE |
| HHIPL1 | 4.641459 | 0.31508642 | 0.0121 | 0.504436064 | 4.641458985 | 9.97235915 | 0.07848298 | 0.880727047 | 1 | 0.708209761 | 6.559162154 | 14.02025537 | 59.13968665 | 1.771879019 | FALSE | TRUE | FALSE |
| ZNF529 | 4.6390468 | 0.31534416 | 0.68935 | 4.639046788 | 420.5299095 | 420.5299095 | 6.3448986 | 0.26027884 | 0.81105 | 6.344898599 | 582.1553694 | 5783.67573 | 0.73114593 | -0.135995933 | FALSE | FALSE | FALSE |
| NUTM2A | 4.6372766 | 0.31560166 | 0.87245 | 45.02427752 | 448.3609899 | 884.3858607 | 0.51693573 | 0.748614059 | 0.93965 | 47.44003662 | 4817.8824 | 4817.8824 | 8.970702408 | 0.95282645 | FALSE | TRUE | FALSE |
| AP1S1 | 4.6284382 | 0.31585904 | 0.50465 | 4.62843818 | 4.62843818 | 4.62843818 | 0.53632464 | 0.739869841 | 1 | 5.036391163 | 5.036391163 | 5.036391163 | 8.6299189 | 0.936006714 | FALSE | TRUE | FALSE |
| ZBTB7B | 4.6269376 | 0.31611618 | 0.50255 | 4.626937591 | 4.626937591 | 16.60793996 | 0.55077572 | 0.733057619 | 1 | 0.550775719 | 0.550775719 | 10.88124455 | 8.400765384 | 0.924318856 | FALSE | TRUE | FALSE |
| MYL2 | 4.6235891 | 0.31637311 | 0.49935 | 1.155685093 | 18.50237441 | 74.06007957 | 0.41032267 | 0.788315818 | 0.00395 | 0.410322669 | 0.410322669 | 0.410322669 | 11.26817866 | 1.051853724 | FALSE | TRUE | FALSE |
| RUSC2 | 4.6116851 | 0.31662998 | 0.0134 | 0.137275752 | 0.137275752 | 4.611685087 | 0.17332611 | 0.858761997 | 1 | 0.173326111 | 0.173326111 | 5.831131471 | 26.60698421 | 1.424995652 | FALSE | TRUE | FALSE |
| NUDT5 | 4.6066896 | 0.31688667 | 0.75075 | 4.60668958 | 44.32288047 | 44.32288047 | 0.56881434 | 0.725233772 | 1 | 0.568814342 | 0.568814342 | 0.568814342 | 8.098757784 | 0.90841841 | FALSE | TRUE | FALSE |
| RNF144A | 4.6047358 | 0.31714313 | 0.5042 | 4.604735832 | 4.604735832 | 4.604735832 | 0.53096424 | 0.742367382 | 1 | 5.017126615 | 5.017126615 | 5.017126615 | 8.672402963 | 0.938139449 | FALSE | TRUE | FALSE |
| CLCNKA | 4.6027602 | 0.31739935 | 0.85015 | 176.6001305 | 179974.9665 | 1894098.263 | 0.16827994 | 0.8600405 | 0.91015 | 3.704957082 | 1433.317907 | 14606.64434 | 27.35180575 | 1.436986004 | FALSE | TRUE | FALSE |
| PAK6 | 4.6024808 | 0.31765531 | 0.7605 | 4.602480831 | 44.66602276 | 90.50724284 | 0.56499653 | 0.726571204 | 1 | 0.564996531 | 0.564996531 | 11.13300234 | 8.146033782 | 0.910946207 | FALSE | TRUE | FALSE |
| TPM1 | 4.6014815 | 0.31791102 | 0.00565 | 0.487845605 | 4.601481485 | 4.601481485 | 0.07970092 | 0.880437062 | 0.006 | 0.079700921 | 0.079700921 | 0.079700921 | 57.73435784 | 1.76143434 | FALSE | TRUE | FALSE |
| ASTE1 | 4.5939803 | 0.31816659 | 0.49995 | 1.912871048 | 43.02449091 | 43.02449091 | 0.72703807 | 0.653274446 | 0.75575 | 0.727038068 | 6.693376332 | 14.38832245 | 6.31876177 | 0.800631982 | FALSE | TRUE | FALSE |
| SPINK8 | 4.5710999 | 0.3184223 | 0.50475 | 4.571099925 | 4.571099925 | 4.571099925 | 0.53237673 | 0.741695141 | 1 | 0.532376734 | 4.978362939 | 4.978362939 | 8.586212797 | 0.933801648 | FALSE | TRUE | FALSE |
| HOXC13 | 4.569908 | 0.31867777 | 0.6293 | 4.569907972 | 176.3802054 | 176.3802054 | 0.1702929 | 0.859588224 | 1 | 0.170292897 | 0.170292897 | 0.170292897 | 26.83557604 | 1.428710922 | FALSE | TRUE | FALSE |
| CRIM1 | 4.5665789 | 0.31893303 | 0.25615 | 1.141948214 | 4.56657893 | 22.28718628 | 0.16347849 | 0.861225059 | 1 | 0.163478487 | 0.163478487 | 3.199866591 | 27.93382189 | 1.44613036 | FALSE | TRUE | FALSE |
| ANGPT1 | 4.5610708 | 0.31918813 | 0.8732 | 103.1894486 | 10335.00508 | 41342.95841 | 0.38363148 | 0.797943163 | 1 | 0.383631479 | 0.383631479 | 7.611762006 | 11.88919844 | 1.075152576 | FALSE | TRUE | FALSE |
| MLIP | 4.555461 | 0.31944305 | 0.6858 | 4.555460983 | 401.83419 | 401.83419 | 0.08649629 | 0.878902127 | 1 | 0.086496288 | 0.770605476 | 1.718186226 | 52.66654898 | 1.721534862 | FALSE | TRUE | FALSE |
| PRR19 | 4.5505685 | 0.3196978 | 0.5061 | 4.550568471 | 4.550568471 | 9.633748312 | 0.5456499 | 0.735429298 | 1 | 0.545649905 | 0.545649905 | 10.83310695 | 8.339721919 | 0.92115157 | FALSE | TRUE | FALSE |
| CNTN1 | 4.5483352 | 0.31995232 | 0.14325 | 1.136991665 | 4.548335178 | 22.31698637 | 0.10047478 | 0.875800182 | 0.0231 | 0.100474776 | 0.100474776 | 1.976544751 | 45.26842804 | 1.655795414 | FALSE | TRUE | FALSE |
| MAP1LC3C | 4.5468858 | 0.3202066 | 0.0037 | 0.486729509 | 4.546885844 | 4.546885844 | 0.54634433 | 0.73507419 | 1 | 0.546344329 | 0.546344329 | 0.546344329 | 8.322381333 | 0.920247611 | FALSE | TRUE | FALSE |
| FBLN5 | 4.5465778 | 0.32046062 | 0.6381 | 18.18674676 | 291.2200154 | 1165.781001 | 8.10928985 | 0.225902488 | 0.77995 | 129.6207814 | 13088.35161 | 64851.52064 | 0.560662878 | -0.251298198 | FALSE | FALSE | FALSE |
| OR8K5 | 4.5402651 | 0.32071449 | 0.7453 | 4.540265098 | 43.65977397 | 43.65977397 | 0.56186217 | 0.728223767 | 1 | 0.561862174 | 0.561862174 | 0.561862174 | 8.080745262 | 0.907451416 | FALSE | TRUE | FALSE |
| PDCD5 | 4.5356254 | 0.32096818 | 0.50095 | 4.535625387 | 4.535625387 | 4.535625387 | 0.54434027 | 0.736312636 | 1 | 0.544340274 | 0.544340274 | 0.544340274 | 8.332334762 | 0.92076671 | FALSE | TRUE | FALSE |
| NUDT22 | 4.533034 | 0.32122165 | 0.50485 | 4.533034029 | 4.533034029 | 9.598129293 | 0.54350017 | 0.736751936 | 0.00885 | 0.543500165 | 0.543500165 | 0.543500165 | 8.340446458 | 0.921189299 | FALSE | TRUE | FALSE |
| NAT14 | 4.5328672 | 0.32147486 | 0.49995 | 0.485912434 | 4.532867234 | 4.532867234 | 0.55081925 | 0.73301244 | 1 | 0.550819247 | 0.550819247 | 0.550819247 | 8.229318893 | 0.915363892 | FALSE | TRUE | FALSE |
| WHAMM | 4.5305396 | 0.32172784 | 0.1318 | 1.906301807 | 42.20452738 | 42.20452738 | 0.08562877 | 0.879032174 | 1 | 0.085628769 | 0.085628769 | 1.692789216 | 52.90908239 | 1.72353023 | FALSE | TRUE | FALSE |
| GTF2H1 | 4.523361 | 0.3219807 | 0.508 | 4.523360986 | 4.523360986 | 16.29489324 | 0.54157329 | 0.737364382 | 1 | 0.541573287 | 0.541573287 | 10.73965353 | 8.352260158 | 0.921804013 | FALSE | TRUE | FALSE |
| STX5 | 4.5220627 | 0.32223331 | 0.50555 | 4.522062723 | 4.522062723 | 4.522062723 | 4.48015624 | 0.331059706 | 0.9376 | 43.09266562 | 4298.869596 | 4298.869596 | 1.0093538 | 0.004043423 | FALSE | FALSE | FALSE |
| NRSN1 | 4.516953 | 0.32248575 | 0.7473 | 4.516953031 | 43.3467741 | 43.3467741 | 4.77170429 | 0.316464382 | 0.87295 | 45.83324549 | 451.5324063 | 451.5324063 | 0.946612103 | -0.023827947 | FALSE | FALSE | FALSE |
| MRPL54 | 4.5162834 | 0.32273794 | 0.5062 | 4.516283373 | 4.516283373 | 4.516283373 | 2.07870454 | 0.48253366 | 0.5036 | 4.872902892 | 19.49093687 | 19.49093687 | 2.172643247 | 0.33698842 | FALSE | FALSE | FALSE |
| AGPAT3 | 4.5082112 | 0.32299001 | 0.00245 | 0.227717355 | 0.227717355 | 4.508211174 | 0.26831213 | 0.834251241 | 1 | 1.072961134 | 1.072961134 | 5.318355005 | 16.80211448 | 1.225363939 | FALSE | TRUE | FALSE |
| ZCRB1 | 4.5042752 | 0.32324189 | 0.51055 | 4.504275245 | 41.84864077 | 41.84864077 | 2.94647137 | 0.414764181 | 0.81915 | 11.78461835 | 450.2952737 | 450.2952737 | 1.528701513 | 0.184322696 | FALSE | FALSE | FALSE |
| CSTA | 4.4982784 | 0.32349361 | 0.50045 | 4.498278418 | 4.498278418 | 4.498278418 | 0.54119573 | 0.737538828 | 1 | 0.541195726 | 0.541195726 | 0.541195726 | 8.311740468 | 0.919691974 | FALSE | TRUE | FALSE |
| TMCO5A | 4.4977676 | 0.32374509 | 0.25365 | 0.49004482 | 4.497767624 | 4.497767624 | 0.65794843 | 0.686515079 | 0.0064 | 0.657948432 | 0.657948432 | 0.657948432 | 6.836048849 | 0.834805157 | FALSE | TRUE | FALSE |
| ATP6V0D2 | 4.4952634 | 0.32399635 | 0.2608 | 0.493219104 | 4.495263385 | 4.495263385 | 0.07971684 | 0.880418894 | 1 | 0.713892418 | 0.713892418 | 0.713892418 | 56.39038638 | 1.75120507 | FALSE | TRUE | FALSE |
| OR8B3 | 4.4941159 | 0.32424737 | 0.87615 | 43.17817058 | 425.4867944 | 425.4867944 | 0.50710732 | 0.753103134 | 1 | 45.68013401 | 4553.782714 | 4553.782714 | 8.862258053 | 0.947544392 | FALSE | TRUE | FALSE |
| AKR1C2 | 4.4809322 | 0.32449836 | 0.6921 | 37.9719661 | 973.2432005 | 3892.637733 | 0.17978549 | 0.857081163 | 1 | 0.179785487 | 0.179785487 | 0.179785487 | 24.92377052 | 1.396613744 | FALSE | TRUE | FALSE |
| DEFB1 | 4.4638905 | 0.3247494 | 0.7457 | 4.463890513 | 43.02449699 | 43.02449699 | 4.72337586 | 0.319817465 | 0.8758 | 45.54431461 | 450.4178275 | 450.4178275 | 0.945063582 | -0.024538972 | FALSE | FALSE | FALSE |
| EEF1E1 | 4.4596553 | 0.32500025 | 0.5027 | 4.459655338 | 4.459655338 | 4.459655338 | 4.87320981 | 0.31280019 | 0.49585 | 0.519794751 | 4.873209815 | 4.873209815 | 0.915137149 | -0.038513814 | FALSE | FALSE | FALSE |
| CYP46A1 | 4.4569439 | 0.32525089 | 0.50725 | 4.456943882 | 4.456943882 | 9.382570373 | 0.53430612 | 0.740637262 | 1 | 0.534306117 | 0.534306117 | 10.58867135 | 8.341555034 | 0.921247019 | FALSE | TRUE | FALSE |
| NUFIP1 | 4.4544685 | 0.32550132 | 0.5075 | 4.454468537 | 41.51679968 | 41.51679968 | 0.73144129 | 0.650741242 | 0.0129 | 0.731441286 | 0.731441286 | 1.622007079 | 6.089987838 | 0.784616425 | FALSE | TRUE | FALSE |
| NFYA | 4.4474291 | 0.32575162 | 0.5074 | 4.447429063 | 4.447429063 | 4.447429063 | 0.53414169 | 0.740722234 | 1 | 0.534141686 | 0.534141686 | 0.534141686 | 8.326309635 | 0.920452557 | FALSE | TRUE | FALSE |
| GPHN | 4.4467686 | 0.32600167 | 0.0006 | 0.1315724 | 0.1315724 | 2.590034539 | 1.52762549 | 0.53345086 | 0.5124 | 1.527625488 | 1.527625488 | 5.48363179 | 2.910902337 | 0.464027635 | FALSE | FALSE | FALSE |
| SLC29A2 | 4.4271003 | 0.32625181 | 0.64335 | 10.09901821 | 378.0407587 | 1512.466323 | 1.7162493 | 0.516150926 | 0.71985 | 3.948665045 | 63.16107254 | 252.70388 | 2.579520548 | 0.411538992 | FALSE | FALSE | FALSE |
| CDX1 | 4.4164724 | 0.32650188 | 0.687 | 4.416472376 | 70.699221 | 70.699221 | 0.33648301 | 0.813352453 | 1 | 0.33648301 | 0.33648301 | 0.33648301 | 13.12539488 | 1.118112378 | FALSE | TRUE | FALSE |
| CD180 | 4.4078562 | 0.32675185 | 0.6904 | 17.62621011 | 671.0973987 | 2684.937479 | 551.357837 | 0.003979377 | 0.11645 | 56.34168224 | 5527.388017 | 56733.90988 | 0.007994547 | -2.097206137 | FALSE | FALSE | FALSE |
| TTC36 | 4.3989369 | 0.32700172 | 0.49865 | 0.470520824 | 4.398936885 | 4.398936885 | 0.53030061 | 0.742869118 | 1 | 0.530300606 | 0.530300606 | 0.530300606 | 8.29517605 | 0.918825608 | FALSE | TRUE | FALSE |
| ZC3H11A | 4.3968556 | 0.32725138 | 0.51845 | 4.396855612 | 4.396855612 | 86.62241359 | 0.5263359 | 0.744403193 | 0.01835 | 0.526335901 | 0.526335901 | 10.387998 | 8.353706454 | 0.92187921 | FALSE | TRUE | FALSE |
| CSTL1 | 4.3925003 | 0.32750085 | 0.3123 | 4.392500264 | 429.0981529 | 429.0981529 | 0.51599436 | 0.748892239 | 0.93915 | 4.924805746 | 483.7452277 | 483.7452277 | 8.512690413 | 0.930066839 | FALSE | TRUE | FALSE |
| HFM1 | 4.3786523 | 0.32775031 | 0.048 | 0.496990916 | 4.378652322 | 39.52326032 | 0.01731696 | 0.894506129 | 0.51175 | 0.017316964 | 0.017316964 | 0.339668344 | 252.8533465 | 2.402868706 | FALSE | TRUE | FALSE |
| VAPB | 4.3766114 | 0.32799956 | 0.7497 | 4.376611436 | 42.29409514 | 42.29409514 | 2.14349411 | 0.476905073 | 0.504 | 2.143494112 | 2.143494112 | 2.143494112 | 2.041811737 | 0.310015696 | FALSE | FALSE | FALSE |
| DNAJC8 | 4.3741058 | 0.32824859 | 0.50175 | 4.374105809 | 4.374105809 | 4.374105809 | 0.52628871 | 0.744485572 | 1 | 0.526288709 | 0.526288709 | 0.526288709 | 8.31122867 | 0.919665231 | FALSE | TRUE | FALSE |
| GRPEL1 | 4.3733403 | 0.32849738 | 0.005 | 4.373340284 | 4.373340284 | 4.373340284 | 0.52516237 | 0.744896642 | 0.00535 | 0.525162375 | 0.525162375 | 0.525162375 | 8.327596368 | 0.920519667 | FALSE | TRUE | FALSE |
| IGLON5 | 4.3644566 | 0.32874607 | 0.50735 | 4.364456571 | 4.364456571 | 4.364456571 | 0.52441573 | 0.745101667 | 0.00825 | 0.524415727 | 0.524415727 | 0.524415727 | 8.322512747 | 0.920254469 | FALSE | TRUE | FALSE |
| UBLCP1 | 4.3386316 | 0.32899497 | 0.50015 | 4.33863162 | 4.33863162 | 4.33863162 | 0.52204783 | 0.746202898 | 0.0055 | 0.522047833 | 0.522047833 | 0.522047833 | 8.310793273 | 0.91964248 | FALSE | TRUE | FALSE |
| GMPR2 | 4.3367286 | 0.32924365 | 0.19485 | 1.084988191 | 17.3383606 | 69.3365703 | 3.1362448 | 0.404414036 | 0.47395 | 1.822916113 | 29.13572224 | 116.5247433 | 1.382777444 | 0.140752287 | FALSE | FALSE | FALSE |
| SYNGR1 | 4.3255797 | 0.32949227 | 0.50405 | 4.325579713 | 4.325579713 | 4.325579713 | 0.52058042 | 0.747132922 | 1 | 0.520580417 | 0.520580417 | 0.520580417 | 8.309147962 | 0.919556493 | FALSE | TRUE | FALSE |
| SHE | 4.3220786 | 0.3297407 | 0.50065 | 4.322078579 | 4.322078579 | 9.102025993 | 0.52012372 | 0.747334143 | 1 | 0.520123724 | 0.520123724 | 10.30858939 | 8.309712437 | 0.919585995 | FALSE | TRUE | FALSE |
| ZNF396 | 4.3149496 | 0.32998901 | 0.0035 | 0.127335521 | 0.127335521 | 0.127335521 | 0.16140983 | 0.861715035 | 1 | 0.161409827 | 0.161409827 | 0.161409827 | 26.73288009 | 1.42704575 | FALSE | TRUE | FALSE |
| RNASE7 | 4.3063433 | 0.33023721 | 0.75165 | 4.306343251 | 41.28725763 | 41.28725763 | 0.53736962 | 0.739570143 | 1 | 0.537369621 | 0.537369621 | 0.537369621 | 8.013745254 | 0.903835533 | FALSE | TRUE | FALSE |
| B3GNTL1 | 4.3047376 | 0.33048519 | 0.25705 | 0.471085765 | 4.304737608 | 4.304737608 | 0.07876024 | 0.880699916 | 0.0119 | 0.07876024 | 0.07876024 | 1.56211421 | 54.65622746 | 1.737639652 | FALSE | TRUE | FALSE |
| OR2M5 | 4.2956695 | 0.33073308 | 0.50745 | 4.295669494 | 4.295669494 | 4.295669494 | 0.50360707 | 0.75405687 | 1 | 4.713514893 | 4.713514893 | 4.713514893 | 8.529803743 | 0.930939039 | FALSE | TRUE | FALSE |
| BAD | 4.2855348 | 0.3309809 | 0.968 | 402.177435 | 4044.438496 | 41665.11714 | 0.57110496 | 0.724124903 | 1 | 0.571104964 | 5.35389682 | 5.35389682 | 7.50393545 | 0.87528909 | FALSE | TRUE | FALSE |
| RPP14 | 4.2785412 | 0.33122859 | 0.50315 | 4.278541201 | 4.278541201 | 4.278541201 | 0.5174313 | 0.748375113 | 1 | 0.517431301 | 0.517431301 | 0.517431301 | 8.268810172 | 0.917443022 | FALSE | TRUE | FALSE |
| OR51A4 | 4.2711455 | 0.33147615 | 0.5056 | 4.27114553 | 4.27114553 | 4.27114553 | 0.51666684 | 0.748693602 | 1 | 0.516666836 | 0.516666836 | 0.516666836 | 8.266730583 | 0.917333784 | FALSE | TRUE | FALSE |
| MAP2K7 | 4.2683276 | 0.33172352 | 0.507 | 4.268327648 | 17.07173753 | 17.07173753 | 0.51851028 | 0.747895791 | 1 | 0.518510282 | 0.518510282 | 10.2663002 | 8.231905507 | 0.915500377 | FALSE | TRUE | FALSE |
| NFKBIB | 4.2631471 | 0.33197073 | 0.5052 | 4.263147105 | 4.263147105 | 8.986467007 | 0.51454903 | 0.749841034 | 1 | 0.514549032 | 0.514549032 | 10.1915433 | 8.285210622 | 0.918303553 | FALSE | TRUE | FALSE |
| OR1L6 | 4.26278 | 0.33221769 | 0.4944 | 0.453993743 | 4.26278002 | 4.26278002 | 4.53618015 | 0.326989441 | 0.7521 | 4.536180154 | 43.73283382 | 43.73283382 | 0.939728996 | -0.026997373 | FALSE | FALSE | FALSE |
| NAALADL1 | 4.2597663 | 0.33246445 | 0.5031 | 4.259766328 | 39.37824599 | 372.9344133 | 0.1027055 | 0.875281963 | 0.8738 | 0.914912822 | 8.359739148 | 18.02113213 | 41.47554334 | 1.617792084 | FALSE | TRUE | FALSE |

| | | | | | | | | | | | | | | | | | |
|---|---|---|---|---|---|---|---|---|---|---|---|---|---|---|---|---|---|
| GTF3C3 | 4.2460881 | 0.33271121 | 0.25655 | 0.462399465 | 4.246088144 | 15.60554011 | 0.077286 | 0.88097973 | 0.02115 | 0.077286004 | 0.077286004 | 1.522684932 | 54.93993672 | 1.739888155 | FALSE | TRUE | FALSE |
| CSTB | 4.245408 | 0.33295773 | 0.752 | 4.24540803 | 40.57963881 | 40.57963881 | 0.53171851 | 0.74203172 | 1 | 0.53171851 | 0.53171851 | 0.53171851 | 7.984314921 | 0.902237658 | FALSE | TRUE | FALSE |
| PTGER1 | 4.2399603 | 0.33320409 | 0.50315 | 4.2399603 | 4.2399603 | 4.2399603 | 0.49700226 | 0.75620338 | 1 | 4.660996964 | 4.660996964 | 4.660996964 | 8.531068476 | 0.931003428 | FALSE | TRUE | FALSE |
| CSDE1 | 4.2228969 | 0.33345051 | 0.5142 | 4.222896946 | 4.222896946 | 83.1252521 | 0.50836174 | 0.752411721 | 0.0198 | 0.508361743 | 0.508361743 | 10.02580995 | 8.306873998 | 0.919437623 | FALSE | TRUE | FALSE |
| CST11 | 4.2224922 | 0.33369669 | 0.5001 | 4.222492199 | 4.222492199 | 4.222492199 | 0.49715869 | 0.756054009 | 1 | 0.497158688 | 4.640277897 | 4.640277897 | 8.493248336 | 0.929073823 | FALSE | TRUE | FALSE |
| ZBED2 | 4.2064528 | 0.3339429 | 0.50165 | 4.206452818 | 4.206452818 | 4.206452818 | 0.51094844 | 0.75121122 | 1 | 0.510948436 | 0.510948436 | 0.510948436 | 8.232636649 | 0.915538948 | FALSE | TRUE | FALSE |
| RECK | 4.202836 | 0.33418893 | 0.2605 | 0.458127902 | 4.202835982 | 15.45990936 | 0.67046229 | 0.679902101 | 0.50855 | 0.67046229 | 0.67046229 | 13.20263486 | 6.268564309 | 0.797168086 | FALSE | TRUE | FALSE |
| KBTBD13 | 4.2026048 | 0.33443471 | 0.59805 | 7.318086653 | 158.3594516 | 633.4480015 | 10.8138322 | 0.189450086 | 0.3782 | 4.793210147 | 43.26769959 | 173.1635534 | 0.38863233 | -0.410461073 | FALSE | FALSE | FALSE |
| CCL11 | 4.2009733 | 0.33468028 | 0.49885 | 0.451539917 | 4.200973341 | 4.200973341 | 0.5109369 | 0.751250136 | 1 | 0.510936901 | 0.510936901 | 0.510936901 | 8.222098136 | 0.914982656 | FALSE | TRUE | FALSE |
| TRIP4 | 4.1918198 | 0.33492575 | 0.0749 | 1.047762193 | 16.77455452 | 20.75382537 | 0.37257828 | 0.801541805 | 0.5063 | 0.372578282 | 0.372578282 | 1.847612938 | 11.25084316 | 1.051185071 | FALSE | TRUE | FALSE |
| DNPH1 | 4.1868858 | 0.33517107 | 0.50065 | 4.1868858 | 4.1868858 | 4.1868858 | 0.50961492 | 0.75171615 | 1 | 0.509614919 | 0.509614919 | 0.509614919 | 8.215783423 | 0.914648982 | FALSE | TRUE | FALSE |
| PCMTD1 | 4.1768841 | 0.33541632 | 0.75535 | 4.176884115 | 40.10333693 | 40.10333693 | 4.60016561 | 0.324247301 | 0.259 | 4.600165609 | 44.24527328 | 44.24527328 | 0.907985596 | -0.041921041 | FALSE | FALSE | FALSE |
| TSHB | 4.1763684 | 0.33566133 | 0.87715 | 40.333739 | 399.5193234 | 399.5193234 | 4.70016017 | 0.320650533 | 0.5037 | 18.79832424 | 181.9116331 | 181.9116331 | 0.888558731 | -0.051313862 | FALSE | FALSE | FALSE |
| TXNRD2 | 4.1674653 | 0.33590625 | 0.942 | 157.0630901 | 15034.44685 | 60154.87265 | 0.62451156 | 0.70139204 | 0.87475 | 13.635897 | 529.5187135 | 529.5187135 | 6.673159563 | 0.824331509 | FALSE | TRUE | FALSE |
| SRFBP1 | 4.1646816 | 0.33615097 | 0.50255 | 38.8531294 | 371.5543545 | 3640.092091 | 0.68890414 | 0.671239553 | 0.877 | 6.325913249 | 59.58061274 | 59.58061274 | 6.045371678 | 0.781423007 | FALSE | TRUE | FALSE |
| OR1J2 | 4.1603864 | 0.33639553 | 0.4978 | 4.160386372 | 392.2308369 | 392.2308369 | 433.76127 | 0.005311639 | 0.00315 | 44.02610723 | 433.7612701 | 433.7612701 | 0.00959142 | -2.018117107 | FALSE | FALSE | FALSE |
| MS4A3 | 4.1515442 | 0.33663999 | 0.0051 | 4.151544215 | 4.151544215 | 4.151544215 | 4.57256735 | 0.325620416 | 0.0051 | 4.572567349 | 4.572567349 | 4.572567349 | 0.907924126 | -0.041950443 | FALSE | FALSE | FALSE |
| FAM177B | 4.1467733 | 0.3368843 | 0.49345 | 0.444498181 | 4.146773258 | 4.146773258 | 4.42453032 | 0.33481451 | 0.75435 | 4.424530316 | 42.41361692 | 42.41361692 | 0.93722338 | -0.028156886 | FALSE | FALSE | FALSE |
| RPS15A | 4.1464504 | 0.33712836 | 1 | 51594350.71 | 79564176807 | 1.17832E+13 | 0.55342345 | 0.731877717 | 1 | 554095.8969 | 762456057.4 | 8859116913 | 7.492364867 | 0.874618919 | FALSE | TRUE | FALSE |
| SP100 | 4.1365832 | 0.33737235 | 0.13 | 0.4544459 | 4.136583196 | 4.184700025 | 0.86152632 | 0.604957732 | 0.26085 | 0.095989407 | 0.861526323 | 1.893599892 | 4.801458857 | 0.681373212 | FALSE | TRUE | FALSE |
| MMRN2 | 4.1318471 | 0.33761618 | 0.5154 | 4.131847108 | 38.95250238 | 80.84525491 | 0.65968726 | 0.68587963 | 0.7585 | 0.659687256 | 6.109138199 | 12.94139695 | 6.26334232 | 0.796806149 | FALSE | TRUE | FALSE |
| RGS9BP | 4.1264019 | 0.33785987 | 0.1324 | 4.126401933 | 39.66315764 | 39.66315764 | 0.06370219 | 0.88384356 | 1 | 0.063702195 | 0.063702195 | 0.063702195 | 64.77644854 | 1.811417134 | FALSE | TRUE | FALSE |
| DNAJC25 | 4.1103029 | 0.33810359 | 0.50195 | 4.110302883 | 4.110302883 | 4.110302883 | 0.49938797 | 0.755304331 | 1 | 0.499387972 | 0.499387972 | 0.499387972 | 8.230680583 | 0.915435748 | FALSE | TRUE | FALSE |
| TRAF4 | 4.0988472 | 0.33834728 | 0.7565 | 4.098847161 | 39.50971029 | 81.00361905 | 4.52654074 | 0.328080062 | 0.757 | 4.526540738 | 43.6896194 | 89.58371982 | 0.905514254 | -0.043104709 | FALSE | FALSE | FALSE |
| ZNF878 | 4.0954603 | 0.33859078 | 0.6935 | 4.095460262 | 365.3924843 | 365.3924843 | 0.65840344 | 0.686261225 | 0.875 | 6.040699282 | 56.84600557 | 56.84600557 | 6.220289915 | 0.793810627 | FALSE | TRUE | FALSE |
| SMCP | 4.0891791 | 0.33883415 | 0.7551 | 4.089179139 | 39.00804403 | 39.00804403 | 0.51517341 | 0.749525626 | 1 | 0.515173407 | 0.515173407 | 0.515173407 | 7.937481022 | 0.8996827 | FALSE | TRUE | FALSE |
| ZNF765 | 4.0753702 | 0.33907752 | 0.50725 | 4.075370247 | 38.00074108 | 38.00074108 | 5.6476734 | 0.280665237 | 0.3222 | 5.647673403 | 510.4258812 | 510.4258812 | 0.721601615 | -0.141702504 | FALSE | FALSE | FALSE |
| CSRNP2 | 4.072853 | 0.3393207 | 0.25995 | 0.42422711 | 4.072852978 | 8.380252169 | 0.05371039 | 0.885946047 | 1 | 0.053710391 | 0.053710391 | 1.062573108 | 75.82988834 | 1.879840416 | FALSE | TRUE | FALSE |
| CD19 | 4.0499108 | 0.33956404 | 0.25335 | 0.427129018 | 4.049910794 | 8.440858431 | 0.05741652 | 0.88519094 | 1 | 0.05741652 | 0.05741652 | 1.136282114 | 70.53563628 | 1.848408588 | FALSE | TRUE | FALSE |
| HOXB13 | 4.0406165 | 0.3398073 | 0.3191 | 1.010180782 | 16.16607834 | 16.16607834 | 0.09044337 | 0.878097658 | 1 | 0.361609591 | 0.361609591 | 0.361609591 | 44.67565288 | 1.650070908 | FALSE | TRUE | FALSE |
| ATG14 | 4.0405372 | 0.34005032 | 0.01565 | 4.04053723 | 4.04053723 | 14.57942191 | 0.49094153 | 0.758457888 | 1 | 0.49094153 | 0.49094153 | 9.718601966 | 8.230180141 | 0.915409341 | FALSE | TRUE | FALSE |
| ZNF444 | 4.0387281 | 0.34029313 | 0.4978 | 0.428902502 | 4.038728107 | 4.038728107 | 0.46615551 | 0.768504644 | 0.005 | 0.466155505 | 0.466155505 | 0.466155505 | 8.663907351 | 0.937713799 | FALSE | TRUE | FALSE |
| CYP11B2 | 4.0244394 | 0.34053595 | 0.2599 | 1.726061059 | 16.09167541 | 16.09167541 | 0.07290461 | 0.881812164 | 0.014 | 0.072904614 | 0.072904614 | 1.442049073 | 55.20143613 | 1.741950377 | FALSE | TRUE | FALSE |
| CLDN9 | 4.0238429 | 0.34077854 | 0.0008 | 0.118525771 | 0.118525771 | 0.118525771 | 0.04488006 | 0.887884537 | 1 | 0.044880056 | 0.179397129 | 0.179397129 | 89.65770596 | 1.952587623 | FALSE | TRUE | FALSE |
| TPD52L3 | 4.0237365 | 0.34102089 | 0.4999 | 0.432916633 | 4.023736475 | 4.023736475 | 0.49233145 | 0.757870756 | 1 | 0.492331447 | 0.492331447 | 0.492331447 | 8.172820361 | 0.912371953 | FALSE | TRUE | FALSE |
| ZCCHC10 | 4.0218389 | 0.34126303 | 0.50445 | 38.67171595 | 381.3851144 | 381.3851144 | 0.52100045 | 0.747012029 | 1 | 0.52100045 | 0.52100045 | 0.52100045 | 7.719453742 | 0.887586569 | FALSE | TRUE | FALSE |
| OBP2B | 4.0162761 | 0.34150503 | 0.7494 | 4.016276104 | 38.34322695 | 38.34322695 | 4.58513481 | 0.325071876 | 0.5049 | 4.585134811 | 4.585134811 | 4.585134811 | 0.875934137 | -0.057528548 | FALSE | FALSE | FALSE |
| VPS26B | 4.0150081 | 0.34174681 | 0.25675 | 4.015008089 | 38.87616128 | 38.87616128 | 0.50172751 | 0.754474124 | 1 | 0.501727512 | 0.501727512 | 0.501727512 | 8.002367799 | 0.903218508 | FALSE | TRUE | FALSE |
| NAA20 | 4.0095988 | 0.34198845 | 0.50615 | 4.009598842 | 4.009598842 | 4.009598842 | 0.48871915 | 0.759296679 | 1 | 0.488719153 | 0.488719153 | 0.488719153 | 8.204300601 | 0.914041564 | FALSE | TRUE | FALSE |
| C6orf136 | 4.0066666 | 0.34222989 | 0.01 | 4.006666635 | 4.006666635 | 4.051755994 | 4.29360033 | 0.338534286 | 0.2546 | 4.293600326 | 41.19073475 | 41.19073475 | 0.933171775 | -0.030038406 | FALSE | FALSE | FALSE |
| TAOK3 | 4.0058033 | 0.34247112 | 0.00775 | 0.118769426 | 0.118769426 | 2.330617459 | 0.15268838 | 0.863920273 | 0.0234 | 0.152688377 | 0.152688377 | 3.003010696 | 26.23515547 | 1.418883642 | FALSE | TRUE | FALSE |
| PGRMC2 | 4.003731 | 0.34271213 | 0.9991 | 39322.78769 | 49837794.22 | 563800006.1 | 44.4265531 | 0.061762101 | 0.98055 | 45946.3143 | 5288909.377 | 58706461.75 | 0.090120227 | -1.045177725 | FALSE | FALSE | FALSE |
| MACROD1 | 4.0032202 | 0.34295292 | 0.5027 | 4.003220173 | 4.003220173 | 4.003220173 | 0.4731733 | 0.765793021 | 1 | 4.430648173 | 4.430648173 | 4.430648173 | 8.460367822 | 0.927389245 | FALSE | TRUE | FALSE |
| TMEM38B | 4.0014156 | 0.3431935 | 0.504 | 4.00141561 | 4.00141561 | 4.00141561 | 0.48869199 | 0.75933301 | 1 | 0.48869199 | 0.48869199 | 0.48869199 | 8.188011445 | 0.913178441 | FALSE | TRUE | FALSE |
| NAGA | 3.9981067 | 0.3434339 | 0.7297 | 15.99471429 | 256.1804359 | 1267.802371 | 121.223777 | 0.023787195 | 0.38565 | 30.29020909 | 1943.05039 | 7782.062606 | 0.032981208 | -1.481733435 | FALSE | FALSE | FALSE |
| PELI2 | 3.9947396 | 0.34367412 | 0.5071 | 3.994739578 | 3.994739578 | 8.403753276 | 4.42579606 | 0.334547824 | 0.50685 | 4.42579606 | 4.42579606 | 9.311789343 | 0.902603627 | -0.044502925 | FALSE | FALSE | FALSE |
| ZPBP | 3.9928312 | 0.34391413 | 0.0093 | 0.427253825 | 3.992831201 | 3.992831201 | 0.47213659 | 0.766340624 | 1 | 4.420589601 | 4.420589601 | 4.420589601 | 8.456940797 | 0.92721329 | FALSE | TRUE | FALSE |
| PTPN23 | 3.9914795 | 0.34415393 | 0.9568 | 34319.95498 | 11762052.41 | 423570617.2 | 0.01204806 | 0.896049801 | 0.99635 | 82.0236593 | 7843.284848 | 79417.29884 | 331.296321 | 2.520216613 | FALSE | TRUE | FALSE |
| RHO | 3.9909118 | 0.34439351 | 0.85145 | 93.48536573 | 15017.9972 | 74394.39942 | 0.13675253 | 0.867469469 | 0.99245 | 478.8343243 | 49645.86215 | 130163.8221 | 29.18345782 | 1.465136748 | FALSE | TRUE | FALSE |
| NOXO1 | 3.9885917 | 0.34463288 | 0.0022 | 0.117591561 | 0.117591561 | 2.328929633 | 1.33113926 | 0.552001952 | 0.5087 | 1.331139261 | 1.331139261 | 2.898380885 | 2.996374476 | 0.476596089 | FALSE | FALSE | FALSE |
| B3GNT6 | 3.980796 | 0.34487216 | 0.50845 | 3.980795979 | 3.980795979 | 8.386294893 | 0.61684125 | 0.705006537 | 1 | 0.616841246 | 0.616841246 | 12.20364447 | 6.45351783 | 0.809796514 | FALSE | TRUE | FALSE |
| AP3S1 | 3.9791162 | 0.34511123 | 0.5071 | 3.979116201 | 3.979116201 | 3.979116201 | 0.48709983 | 0.760165423 | 1 | 0.487099831 | 0.487099831 | 0.487099831 | 8.168995247 | 0.912168643 | FALSE | TRUE | FALSE |
| PADI3 | 3.9742756 | 0.34535015 | 0.7799 | 15.9088218 | 567.1787239 | 2271.471432 | 786.252051 | 0.002860352 | 0.0151 | 21.89330727 | 786.2520509 | 3148.842426 | 0.005054709 | -2.296303806 | FALSE | FALSE | FALSE |
| CLPP | 3.9730698 | 0.34558885 | 0.50175 | 3.973069829 | 3.973069829 | 3.973069829 | 0.48121451 | 0.762626619 | 0.00735 | 0.481214507 | 0.481214507 | 0.481214507 | 8.256338437 | 0.916787487 | FALSE | TRUE | FALSE |
| DEFB118 | 3.9602017 | 0.34582755 | 0.5048 | 3.96020174 | 3.96020174 | 3.96020174 | 0.46990798 | 0.767021376 | 1 | 4.386139655 | 4.386139655 | 4.386139655 | 8.427611132 | 0.925704488 | FALSE | TRUE | FALSE |
| TIMM10 | 3.9573958 | 0.34606606 | 0.505 | 3.957395834 | 3.957395834 | 3.957395834 | 4.24591597 | 0.34117493 | 0.7489 | 4.245915974 | 40.71371139 | 40.71371139 | 0.93204761 | -0.030561903 | FALSE | FALSE | FALSE |
| KIAA0408 | 3.9571547 | 0.34630434 | 0.5021 | 3.957154657 | 36.73913465 | 36.73913465 | 45368.7146 | 4.22315E-05 | 0.01865 | 48.15217427 | 4519.080699 | 45368.71461 | 8.72221E-05 | -4.059373451 | FALSE | FALSE | FALSE |
| OR2D3 | 3.9504165 | 0.34654251 | 0.6885 | 9.739003836 | 901.5145111 | 901.5145111 | 0.16841757 | 0.859991735 | 0.0066 | 0.168417568 | 0.168417568 | 0.168417568 | 23.45608339 | 1.370255497 | FALSE | TRUE | FALSE |
| SUPT4H1 | 3.9482407 | 0.34678047 | 0.50235 | 3.948240706 | 3.948240706 | 3.948240706 | 0.48103965 | 0.762661901 | 0.0038 | 0.481039655 | 0.481039655 | 0.481039655 | 8.20772397 | 0.914222742 | FALSE | TRUE | FALSE |
| MISP | 3.9297938 | 0.34701853 | 0.89295 | 37.06598992 | 3549.548396 | 35996.47493 | 0.69248906 | 0.668977711 | 0.7569 | 0.69248906 | 6.432663947 | 13.6157841 | 5.674882147 | 0.753956847 | FALSE | TRUE | FALSE |
| MCEE | 3.9252233 | 0.34725644 | 0.754 | 3.925223322 | 37.78788993 | 37.78788993 | 0.47315119 | 0.765827325 | 1 | 4.448683864 | 17.79294205 | 17.79294205 | 8.295917599 | 0.91886443 | FALSE | TRUE | FALSE |
| FSCN3 | 3.8886642 | 0.34749477 | 0.50065 | 3.88866425 | 36.05899839 | 36.05899839 | 484.136444 | 0.004676169 | 0.0654 | 5.410009927 | 484.136444 | 484.136444 | 0.008032166 | -2.095167329 | FALSE | FALSE | FALSE |
| RAET1E | 3.8859996 | 0.34773291 | 0.5059 | 3.885999629 | 3.885999629 | 3.885999629 | 4.17722452 | 0.345876078 | 0.7482 | 4.177224517 | 39.93247829 | 39.93247829 | 0.930282683 | -0.031385063 | FALSE | FALSE | FALSE |
| FABP3 | 3.8848091 | 0.34797083 | 0.8776 | 37.0582929 | 362.5818334 | 362.5818334 | 0.50794449 | 0.752642651 | 1 | 0.507944491 | 0.507944491 | 0.507944491 | 7.648097667 | 0.883553425 | FALSE | TRUE | FALSE |
| KRTAP21-1 | 3.8753408 | 0.34820869 | 0.5027 | 3.875340835 | 3.875340835 | 3.875340835 | 0.47706817 | 0.764203573 | 1 | 0.477068174 | 0.477068174 | 0.477068174 | 8.123243274 | 0.90972946 | FALSE | TRUE | FALSE |
| RAB31 | 3.8615484 | 0.34844656 | 0.50345 | 3.861548425 | 3.861548425 | 3.861548425 | 0.47390296 | 0.765518202 | 1 | 0.473902958 | 0.473902958 | 0.473902958 | 8.14839485 | 0.911072066 | FALSE | TRUE | FALSE |
| ZMYM5 | 3.8494504 | 0.34868442 | 0.5061 | 3.84945042 | 35.66248469 | 35.66248469 | 0.07043635 | 0.882334874 | 1 | 0.625711556 | 5.705016001 | 12.39551332 | 54.65147712 | 1.737601905 | FALSE | TRUE | FALSE |
| STARD7 | 3.8408481 | 0.34892219 | 0.75685 | 15.36187096 | 148.4127675 | 148.4127675 | 4.18566672 | 0.344837753 | 0.8751 | 40.44393597 | 400.8123635 | 400.8123635 | 0.9176192 | -0.037337508 | FALSE | FALSE | FALSE |
| EML1 | 3.8358827 | 0.34915982 | 0.26715 | 0.959336679 | 3.835882749 | 18.84039835 | 1.27224863 | 0.558148574 | 0.5141 | 1.272248634 | 1.272248634 | 25.03997565 | 3.015041752 | 0.479293331 | FALSE | FALSE | FALSE |
| C10orf71 | 3.8278486 | 0.34939736 | 0.50795 | 3.827848552 | 35.69233849 | 75.52711747 | 0.62052188 | 0.703380277 | 0.7546 | 0.620521876 | 5.690428981 | 12.26331078 | 6.168756811 | 0.790197649 | FALSE | TRUE | FALSE |
| MKX | 3.8238215 | 0.34963474 | 0.5077 | 3.823821458 | 3.823821458 | 8.0633894 | 0.46703309 | 0.768102069 | 1 | 0.46703309 | 0.46703309 | 9.263139171 | 8.187474377 | 0.913149954 | FALSE | TRUE | FALSE |
| CES3 | 3.8219584 | 0.34987192 | 0.69115 | 3.821958422 | 353.1070126 | 353.1070126 | 5.90092563 | 0.272604334 | 0.01135 | 0.632676596 | 5.900925633 | 12.51170701 | 0.647687949 | -0.188634183 | FALSE | FALSE | FALSE |
| MED10 | 3.8192754 | 0.35010892 | 0.50875 | 3.819275387 | 3.819275387 | 3.819275387 | 0.46817355 | 0.767731764 | 1 | 0.468173549 | 0.468173549 | 0.468173549 | 8.157819668 | 0.911574101 | FALSE | TRUE | FALSE |

| | | | | | | | | | | | | | | | | | |
|---|---|---|---|---|---|---|---|---|---|---|---|---|---|---|---|---|---|
| UNC119 | 3.817477 | 0.35034571 | 0.8532 | 15.2709562 | 244.5520178 | 979.0083865 | 88.1807832 | 0.033285533 | 0.03655 | 5.50815905 | 22.0361628 | 88.18078318 | 0.043291484 | -1.363597523 | FALSE | FALSE | FALSE |
| ZNF175 | 3.8159754 | 0.3505823 | 0.51145 | 35.84041333 | 345.0564163 | 3403.285459 | 5.87187358 | 0.273539786 | 0.5016 | 5.871873576 | 55.63724253 | 55.63724253 | 0.649873556 | -0.187171135 | FALSE | FALSE | FALSE |
| ID3 | 3.8086127 | 0.35081879 | 0.5004 | 3.808612685 | 15.23648299 | 15.23648299 | 1.12864866 | 0.575201265 | 0.87535 | 4.51426371 | 18.06024397 | 18.06024397 | 3.37448916 | 0.528208037 | FALSE | FALSE | FALSE |
| FXYD3 | 3.8049095 | 0.35105511 | 0.0029 | 3.804909527 | 3.804909527 | 3.804909527 | 4.10101067 | 0.350767712 | 0.24975 | 4.101010672 | 39.27424247 | 39.27424247 | 0.927798007 | -0.032546565 | FALSE | FALSE | FALSE |
| OR1E1 | 3.8035106 | 0.35129123 | 0.25725 | 3.8035106 | 36.35189358 | 36.35189358 | 0.48279221 | 0.761954192 | 1 | 0.482792206 | 0.482792206 | 0.482792206 | 7.878152441 | 0.89642438 | FALSE | TRUE | FALSE |
| KCNQ3 | 3.7799729 | 0.35152753 | 0.9012 | 141.3967627 | 21720.64121 | 249140.9755 | 135404.838 | 1.44341E-05 | 0.43195 | 33808.94599 | 8711464.078 | 140045760 | 2.79161E-05 | -4.554145497 | FALSE | FALSE | FALSE |
| CLEC4C | 3.7696852 | 0.35176379 | 0.69045 | 3.769685178 | 360.5099569 | 360.5099569 | 41.7098855 | 0.06575819 | 0.4925 | 4.305035304 | 414.2258301 | 4214.177298 | 0.090378699 | -1.043933916 | FALSE | FALSE | FALSE |
| CPQ | 3.7612915 | 0.35199997 | 0.00795 | 0.41425622 | 3.761291505 | 3.761291505 | 0.61133883 | 0.707109539 | 0.50105 | 0.611338828 | 0.611338828 | 1.359920756 | 6.152548033 | 0.789055013 | FALSE | TRUE | FALSE |
| HPGDS | 3.7545657 | 0.35223604 | 0.00535 | 3.754565652 | 3.754565652 | 3.754565652 | 0.46324695 | 0.769703602 | 1 | 0.463246946 | 0.463246946 | 0.463246946 | 8.104890237 | 0.908747138 | FALSE | TRUE | FALSE |
| GPM6A | 3.7524944 | 0.35247192 | 0.37825 | 0.938737853 | 15.00390608 | 15.00390608 | 0.13785664 | 0.867216167 | 1 | 0.13785664 | 0.13785664 | 0.13785664 | 27.22026588 | 1.434892363 | FALSE | TRUE | FALSE |
| FCHO2 | 3.732639 | 0.35270792 | 0.01645 | 3.732638987 | 3.732638987 | 13.46799796 | 0.45829868 | 0.771770834 | 1 | 0.458298682 | 0.458298682 | 9.074209229 | 8.144555353 | 0.910867379 | FALSE | TRUE | FALSE |
| STX3 | 3.7307984 | 0.35294372 | 0.5019 | 3.730798421 | 3.730798421 | 3.730798421 | 0.45855183 | 0.771543059 | 1 | 0.458551826 | 0.458551826 | 0.458551826 | 8.136045276 | 0.910413357 | FALSE | TRUE | FALSE |
| LYZ | 3.7294808 | 0.35317932 | 0.256 | 3.729480849 | 35.78721748 | 35.78721748 | 4.29815377 | 0.338269208 | 0.505 | 4.298153771 | 4.298153771 | 4.298153771 | 0.867693677 | -0.061633567 | FALSE | FALSE | FALSE |
| RTP2 | 3.724269 | 0.35341478 | 0.50755 | 3.724268953 | 3.724268953 | 3.724268953 | 0.45958791 | 0.77108606 | 0.00715 | 0.459587912 | 0.459587912 | 0.459587912 | 8.103496311 | 0.908672439 | FALSE | TRUE | FALSE |
| DEFB128 | 3.7135813 | 0.35365021 | 0.5003 | 3.713581282 | 3.713581282 | 3.713581282 | 0.45837408 | 0.771705806 | 0.0015 | 0.458374078 | 0.458374078 | 0.458374078 | 8.101638954 | 0.908572885 | FALSE | TRUE | FALSE |
| AGPAT1 | 3.7054647 | 0.35388555 | 0.5055 | 3.705464737 | 14.81905538 | 14.81905538 | 0.4571921 | 0.772095408 | 1 | 0.457192099 | 0.457192099 | 0.457192099 | 8.104831083 | 0.908743968 | FALSE | TRUE | FALSE |
| FGFBP2 | 3.7005754 | 0.35412074 | 0.50655 | 3.700575379 | 3.700575379 | 3.700575379 | 0.45745407 | 0.77196569 | 1 | 0.457454067 | 0.457454067 | 0.457454067 | 8.089501546 | 0.907921762 | FALSE | TRUE | FALSE |
| ASCL4 | 3.699644 | 0.35435573 | 0.49535 | 0.924980045 | 14.80116984 | 14.80116984 | 4.44966964 | 0.333210704 | 0.25055 | 1.112453733 | 4.44966964 | 4.44966964 | 0.831442399 | -0.080167832 | FALSE | FALSE | FALSE |
| ZSWIM7 | 3.699504 | 0.35459049 | 0.49305 | 0.398169054 | 3.699504016 | 3.699504016 | 3.99935171 | 0.356086934 | 0.74755 | 3.999351706 | 38.20024218 | 38.20024218 | 0.925025926 | -0.033846095 | FALSE | FALSE | FALSE |
| PTGDR2 | 3.6970936 | 0.35482507 | 0.257 | 3.697093602 | 35.51221203 | 35.51221203 | 0.46945292 | 0.767224811 | 1 | 0.46945292 | 0.46945292 | 0.46945292 | 7.875323477 | 0.896268401 | FALSE | TRUE | FALSE |
| BOP1 | 3.695683 | 0.35505944 | 0.02405 | 3.695683008 | 3.695683008 | 72.63993871 | 0.43984381 | 0.777472006 | 1 | 4.131314993 | 4.131314993 | 14.89211217 | 8.402262243 | 0.924396232 | FALSE | TRUE | FALSE |
| RHBG | 3.6727987 | 0.35529399 | 0.0084 | 0.407648289 | 3.67279866 | 3.67279866 | 0.58311132 | 0.719543554 | 0.7512 | 0.58311132 | 5.300305599 | 5.300305599 | 6.298623498 | 0.799245649 | FALSE | TRUE | FALSE |
| PTCD2 | 3.6692581 | 0.35552838 | 0.2538 | 0.917322499 | 3.669258081 | 3.669258081 | 1.21076532 | 0.566341204 | 0.5049 | 1.210765317 | 1.210765317 | 1.210765317 | 3.030527905 | 0.481518287 | FALSE | FALSE | FALSE |
| TAP1 | 3.6623916 | 0.35576267 | 1 | 7.84764E+11 | 3.33226E+15 | 5.40674E+16 | 1.34776798 | 0.550832789 | 1 | 1.33507E+14 | 5.71382E+17 | 3.75051E+19 | 2.717375466 | 0.43414965 | FALSE | FALSE | FALSE |
| CYP2A7 | 3.6583437 | 0.3559968 | 0.77945 | 328.3612119 | 32463.97785 | 63734.45319 | 0.07945318 | 0.880518752 | 1 | 0.079453185 | 0.079453185 | 1.571731336 | 46.04401549 | 1.663173191 | FALSE | TRUE | FALSE |
| NKTR | 3.6574808 | 0.35623072 | 0.26745 | 0.394047514 | 3.657480801 | 13.21648267 | 0.06670922 | 0.883176454 | 0.03635 | 0.066709219 | 0.066709219 | 2.241089315 | 54.82721664 | 1.738996199 | FALSE | TRUE | FALSE |
| PTHLH | 3.6546558 | 0.35646447 | 0.50365 | 3.654655841 | 3.654655841 | 3.654655841 | 368.625417 | 0.006404049 | 0.1209 | 3.835268783 | 368.6254173 | 368.6254173 | 0.009914281 | -2.003738792 | FALSE | FALSE | FALSE |
| POLE3 | 3.65428 | 0.35669799 | 0.49605 | 0.389975387 | 3.654280003 | 3.654280003 | 0.43034825 | 0.781023352 | 1 | 0.430348253 | 0.430348253 | 0.430348253 | 8.491448451 | 0.928981777 | FALSE | TRUE | FALSE |
| TRIM46 | 3.6440653 | 0.35693148 | 0.75955 | 3.644065349 | 35.17636509 | 71.45727537 | 0.46161852 | 0.770462813 | 1 | 0.461618523 | 0.461618523 | 9.073278313 | 7.894105559 | 0.897302929 | FALSE | TRUE | FALSE |
| SORT1 | 3.6352657 | 0.35716489 | 0.02105 | 3.635265677 | 3.635265677 | 71.69544228 | 4.07137891 | 0.352550503 | 0.5112 | 4.07137891 | 4.07137891 | 14.66498231 | 0.892883163 | -0.049205367 | FALSE | FALSE | FALSE |
| TFB1M | 3.6350427 | 0.35739808 | 0.25725 | 0.403887613 | 3.635042668 | 3.635042668 | 0.59591098 | 0.71412264 | 0.50555 | 0.595910985 | 0.595910985 | 0.595910985 | 6.099975936 | 0.785328122 | FALSE | TRUE | FALSE |
| MMD2 | 3.6313738 | 0.35763112 | 0.26215 | 0.907628432 | 3.631373797 | 3.631373797 | 2.46099879 | 0.450764508 | 0.50255 | 2.460998795 | 2.460998795 | 2.460998795 | 1.475569108 | 0.168959554 | FALSE | FALSE | FALSE |
| DEFB108B | 3.6195689 | 0.35786413 | 0.5052 | 34.46160189 | 336.5269219 | 336.5269219 | 37.411237 | 0.073745917 | 0.6915 | 37.411237 | 3670.090458 | 3670.090458 | 0.096750847 | -1.014345223 | FALSE | FALSE | FALSE |
| TPSB2 | 3.6153848 | 0.358097 | 0.0114 | 3.615384782 | 3.615384782 | 7.665220091 | 4.0504707 | 0.354070638 | 0.5083 | 4.050470698 | 4.050470698 | 4.050470698 | 0.892583863 | -0.049350969 | FALSE | FALSE | FALSE |
| MMD | 3.6145919 | 0.35832966 | 0.75405 | 3.614591946 | 34.56820288 | 34.56820288 | 0.46135982 | 0.770561452 | 1 | 0.461359822 | 0.461359822 | 0.461359822 | 7.834648296 | 0.894019505 | FALSE | TRUE | FALSE |
| OR8J1 | 3.6145506 | 0.3585621 | 0.502 | 3.614550635 | 3.614550635 | 3.614550635 | 0.43296516 | 0.780124 | 1 | 4.048341612 | 4.048341612 | 4.048341612 | 8.348363764 | 0.921601364 | FALSE | TRUE | FALSE |
| ADAM20 | 3.6060319 | 0.35879446 | 0.50605 | 3.606031944 | 33.81034029 | 33.81034029 | 0.53532935 | 0.740040787 | 0.9694 | 46.39011661 | 448.3132645 | 4438.381973 | 6.736099868 | 0.828408517 | FALSE | TRUE | FALSE |
| POLDIP3 | 3.5954115 | 0.35902679 | 0.5081 | 3.595411534 | 3.595411534 | 7.623889809 | 4.03051916 | 0.355331799 | 0.0115 | 4.030519159 | 4.030519159 | 8.539877547 | 0.892046754 | -0.049612383 | FALSE | FALSE | FALSE |
| CELF6 | 3.5951036 | 0.3592589 | 0.5074 | 3.595103577 | 3.595103577 | 7.645880788 | 0.43085368 | 0.780784277 | 1 | 4.029593958 | 4.029593958 | 4.029593958 | 8.344140329 | 0.921381599 | FALSE | TRUE | FALSE |
| ERV3-1 | 3.5904169 | 0.35949087 | 0.6406 | 34.38745003 | 3389.59345 | 34846.47397 | 5.34006748 | 0.292156944 | 0.6584 | 5.340067479 | 511.3667826 | 5185.743295 | 0.672354222 | -0.172401864 | FALSE | FALSE | FALSE |
| CYP4A11 | 3.5721663 | 0.35972295 | 0.6319 | 14.29536626 | 917.5648157 | 3675.648889 | 27.6852445 | 0.100491941 | 0.781 | 347.9052212 | 28425.63754 | 266996.1758 | 0.129027804 | -0.889316695 | FALSE | FALSE | FALSE |
| MBOAT7 | 3.5646136 | 0.35995494 | 0.253 | 0.89152404 | 3.56461361 | 7.59350966 | 0.1320102 | 0.868473814 | 1 | 0.132010196 | 0.132010196 | 2.615404044 | 27.00256273 | 1.431404984 | FALSE | TRUE | FALSE |
| HM13 | 3.5617886 | 0.36018675 | 0.49965 | 0.384254392 | 3.561788587 | 3.561788587 | 35.6719024 | 0.078212697 | 0.13275 | 35.67190236 | 348.4249931 | 348.4249931 | 0.099848574 | -1.000658133 | FALSE | FALSE | FALSE |
| LMBR1L | 3.551444 | 0.36041853 | 0.5079 | 3.551443997 | 32.77013217 | 32.77013217 | 0.58388492 | 0.719041417 | 0.7542 | 0.583884917 | 5.304644626 | 5.304644626 | 6.082438321 | 0.784077714 | FALSE | TRUE | FALSE |
| OR2T27 | 3.5506588 | 0.3606501 | 0.6592 | 33.77413918 | 3289.357232 | 33640.12996 | 42.8593223 | 0.063878524 | 0.659 | 42.8593223 | 4221.540645 | 43417.06982 | 0.082844492 | -1.081736363 | FALSE | FALSE | FALSE |
| TIMP4 | 3.5470175 | 0.36088151 | 0.1302 | 0.887014405 | 3.547017541 | 3.547017541 | 0.08083668 | 0.880100101 | 1 | 0.08083668 | 0.08083668 | 0.08083668 | 43.87881283 | 1.642254869 | FALSE | TRUE | FALSE |
| LRFN1 | 3.5462757 | 0.36111272 | 0.512 | 3.546275714 | 3.546275714 | 69.37379911 | 3.72897752 | 0.369662371 | 0.51325 | 36.08629094 | 358.1732496 | 707.7962988 | 0.951004852 | -0.021817267 | FALSE | FALSE | FALSE |
| PSMC1 | 3.5457787 | 0.36134371 | 0.2556 | 0.886524886 | 3.54577868 | 4.397491958 | 0.27106254 | 0.833502926 | 1 | 0.271062542 | 0.271062542 | 5.381795051 | 13.08103528 | 1.116642117 | FALSE | TRUE | FALSE |
| ISOC2 | 3.5434587 | 0.36157452 | 0.50575 | 3.543458685 | 3.543458685 | 3.543458685 | 0.43830807 | 0.777964723 | 1 | 0.438308066 | 0.438308066 | 0.438308066 | 8.084402183 | 0.907647911 | FALSE | TRUE | FALSE |
| KANK3 | 3.5386116 | 0.3618052 | 0.2625 | 0.884742787 | 3.53861156 | 7.739626013 | 0.1334179 | 0.868115193 | 1 | 0.133417897 | 0.133417897 | 2.644573273 | 26.52276526 | 1.423618802 | FALSE | TRUE | FALSE |
| LNP1 | 3.5381431 | 0.36203566 | 0.50605 | 3.538143101 | 3.538143101 | 3.538143101 | 0.44034973 | 0.777379378 | 1 | 0.440349732 | 0.440349732 | 0.440349732 | 8.034847853 | 0.904977657 | FALSE | TRUE | FALSE |
| COL4A3 | 3.5381259 | 0.36226591 | 0.76825 | 107.5530987 | 69003.44873 | 1104595.81 | 2980.62218 | 0.00076239 | 0.17695 | 99.28122256 | 25433.59281 | 407687.202 | 0.001187043 | -2.925533646 | FALSE | FALSE | FALSE |
| NLRP11 | 3.5367421 | 0.36249596 | 0.26425 | 0.389455983 | 3.536742129 | 13.14386009 | 0.06682714 | 0.883141607 | 1 | 0.066827137 | 0.066827137 | 1.312536301 | 52.92374133 | 1.723650538 | FALSE | TRUE | FALSE |
| CLEC16A | 3.5325421 | 0.36272586 | 0.008 | 0.104674641 | 0.104674641 | 2.039094751 | 0.13642676 | 0.867513429 | 1 | 0.136426762 | 0.136426762 | 2.665958792 | 25.89332226 | 1.413187776 | FALSE | TRUE | FALSE |
| LTV1 | 3.525241 | 0.36295568 | 0.66515 | 33.17363848 | 3162.20568 | 31994.69989 | 5.8394462 | 0.273851571 | 0.25335 | 0.629924989 | 5.839446205 | 5.839446205 | 0.603694411 | -0.219182845 | FALSE | FALSE | FALSE |
| MAEA | 3.5227614 | 0.36318531 | 0.26135 | 3.522761447 | 33.82419645 | 69.59448584 | 0.44983287 | 0.774499807 | 0.01335 | 0.449832868 | 0.449832868 | 8.899809341 | 7.831267335 | 0.89383205 | FALSE | TRUE | FALSE |
| IFNL2 | 3.5142815 | 0.36341489 | 0.75475 | 3.514281522 | 33.44872363 | 33.44872363 | 31837.3318 | 5.98251E-05 | 0.0617 | 311.4701572 | 333551.9557 | 333551.9557 | 0.000110382 | -3.957100115 | FALSE | FALSE | FALSE |
| NFKBIE | 3.51313 | 0.36364426 | 0.93875 | 33.51704623 | 3289.665924 | 3289.665924 | 3.70206604 | 0.370872463 | 0.9847 | 346.0858475 | 35716.13847 | 375983.5427 | 0.948964703 | -0.022749941 | FALSE | FALSE | FALSE |
| CGB7 | 3.5093624 | 0.36387348 | 0.93855 | 33.58522715 | 3316.863187 | 3316.863187 | 0.47833625 | 0.763610407 | 1 | 0.478336252 | 0.478336252 | 0.478336252 | 7.336601393 | 0.865494924 | FALSE | TRUE | FALSE |
| RIN3 | 3.5065018 | 0.36410254 | 0.99855 | 3107070.706 | 15867865650 | 8.76168E+11 | 0.14801447 | 0.865077455 | 1 | 571317860.4 | 4.88235E+12 | 2.35983E+14 | 23.69026346 | 1.37456989 | FALSE | TRUE | FALSE |
| ZNF442 | 3.502458 | 0.36433144 | 0.69095 | 3.502457961 | 312.345578 | 312.345578 | 4.94198976 | 0.308813328 | 0.8151 | 46.41672956 | 446.8831036 | 4407.593807 | 0.708714128 | -0.14952891 | FALSE | FALSE | FALSE |
| TRAPPC1 | 3.5007869 | 0.36456016 | 0.75325 | 3.500786911 | 33.55842339 | 33.55842339 | 0.42059242 | 0.784314851 | 1 | 3.935996251 | 37.80374145 | 37.80374145 | 8.323466434 | 0.920304233 | FALSE | TRUE | FALSE |
| ST7 | 3.4975713 | 0.36478872 | 0.5019 | 3.497571283 | 3.497571283 | 12.63395714 | 35.4499663 | 0.079051481 | 0.50535 | 35.44996635 | 349.4125579 | 349.4125579 | 0.098662189 | -1.005849253 | FALSE | FALSE | FALSE |
| TMEM207 | 3.4964119 | 0.36501709 | 0.5074 | 3.496411911 | 3.496411911 | 3.496411911 | 0.43539565 | 0.779034665 | 1 | 0.435395651 | 0.435395651 | 0.435395651 | 8.03042451 | 0.904738504 | FALSE | TRUE | FALSE |
| ANKRD42 | 3.4944852 | 0.36524527 | 0.81435 | 32.82393697 | 316.0441389 | 3117.424452 | 0.07147118 | 0.882122749 | 0.00805 | 0.071471184 | 0.071471184 | 0.071471184 | 48.89362397 | 1.689252228 | FALSE | TRUE | FALSE |
| NDUFV3 | 3.4910427 | 0.36547329 | 0.50725 | 3.491042707 | 32.79418329 | 32.79418329 | 0.54205279 | 0.73723339 | 0.50695 | 0.542052792 | 0.542052792 | 1.194364674 | 6.440410891 | 0.808913576 | FALSE | TRUE | FALSE |
| LCE5A | 3.4738498 | 0.36570141 | 0.7543 | 3.473849811 | 33.00080102 | 33.00080102 | 0.44750233 | 0.775100677 | 1 | 0.447502332 | 0.447502332 | 0.447502332 | 7.762752422 | 0.890015735 | FALSE | TRUE | FALSE |
| ZSCAN21 | 3.4720806 | 0.36592934 | 0.25215 | 0.385030674 | 3.472080647 | 3.472080647 | 0.53815213 | 0.739226754 | 0.87705 | 4.891270576 | 45.59938845 | 45.59938845 | 6.451857144 | 0.809684743 | FALSE | TRUE | FALSE |
| SAMD13 | 3.4693871 | 0.3661571 | 0.0029 | 3.469387087 | 3.469387087 | 3.469387087 | 3.6929384 | 0.371837258 | 0.4977 | 0.393153168 | 3.692938405 | 3.692938405 | 0.939465192 | -0.027119306 | FALSE | FALSE | FALSE |
| ANKRD22 | 3.4659036 | 0.36638471 | 0.25435 | 3.465903609 | 33.18520357 | 33.18520357 | 0.43919944 | 0.77762622 | 0.4992 | 0.439199438 | 1.756670145 | 1.756670145 | 7.891411754 | 0.897154704 | FALSE | TRUE | FALSE |
| OCIAD1 | 3.4636337 | 0.36661214 | 0.50905 | 3.463633711 | 3.463633711 | 3.463633711 | 0.43185401 | 0.780334534 | 1 | 0.431854007 | 0.431854007 | 0.431854007 | 8.020381083 | 0.904195004 | FALSE | TRUE | FALSE |
| C1orf53 | 3.4562207 | 0.36683949 | 0.50085 | 3.456220742 | 3.456220742 | 3.456220742 | 3.64760044 | 0.374475718 | 0.87245 | 3.64760044 | 338.9668958 | 338.9668958 | 0.947532713 | -0.023405787 | FALSE | FALSE | FALSE |
| GLIPR1L1 | 3.4541183 | 0.36706666 | 0.00605 | 3.454118315 | 3.454118315 | 3.454118315 | 0.41537446 | 0.78640572 | 1 | 3.890193294 | 3.890193294 | 3.890193294 | 8.315673399 | 0.919897424 | FALSE | TRUE | FALSE |

| | | | | | | | | | | | | | | | | | |
|---|---|---|---|---|---|---|---|---|---|---|---|---|---|---|---|---|---|
| DGKG | 3.4468829 | 0.36729374 | 1 | 1.61833E+17 | 1.90961E+23 | 1.7078E+25 | 0.08423816 | 0.879282296 | 1 | 791646804.1 | 1.04887E+13 | 5.1664E+14 | 40.91830613 | 1.611917647 | FALSE | TRUE | FALSE |
| OC90 | 3.4432471 | 0.36752068 | 0.264 | 0.375958009 | 3.443247142 | 3.443247142 | 0.56285622 | 0.727659569 | 0.50145 | 0.562856224 | 0.562856224 | 0.562856224 | 6.117454152 | 0.786570723 | FALSE | TRUE | FALSE |
| SNAI3 | 3.4408835 | 0.36774744 | 0.68985 | 3.440883502 | 55.12680713 | 55.12680713 | 2.4060385 | 0.454889295 | 0.5092 | 2.406038499 | 2.406038499 | 5.175661037 | 1.43010326 | 0.155367397 | FALSE | FALSE | FALSE |
| MOGAT3 | 3.4407245 | 0.36797399 | 0.25165 | 0.377006601 | 3.440724494 | 3.440724494 | 0.064757 | 0.883653665 | 1 | 0.064757 | 0.064757 | 0.064757 | 53.13285778 | 1.725363175 | FALSE | TRUE | FALSE |
| ZNF676 | 3.4344772 | 0.36820044 | 0.9119 | 3003.398929 | 309348.5509 | 3248327.056 | 5.68708747 | 0.277891399 | 0.81345 | 53.66561086 | 519.0956811 | 5143.816388 | 0.603907933 | -0.219029266 | FALSE | FALSE | FALSE |
| HSPB3 | 3.4336761 | 0.36842668 | 0.3812 | 1.504999657 | 13.7341889 | 13.7341889 | 1.09853962 | 0.578814638 | 0.8928 | 17.57171972 | 281.3500153 | 1126.227203 | 3.125673391 | 0.494943596 | FALSE | FALSE | FALSE |
| TMEM53 | 3.4323332 | 0.36865274 | 0.7494 | 3.432333247 | 33.03488047 | 33.03488047 | 0.43879925 | 0.777749335 | 1 | 0.438799254 | 0.438799254 | 0.438799254 | 7.822103648 | 0.893323566 | FALSE | TRUE | FALSE |
| NDUFB1 | 3.4282483 | 0.36887866 | 0.5119 | 3.428248293 | 3.428248293 | 3.428248293 | 0.42592938 | 0.782564292 | 0.0034 | 0.425929384 | 0.425929384 | 0.425929384 | 8.048865419 | 0.905734666 | FALSE | TRUE | FALSE |
| OR5B3 | 3.4160933 | 0.36910458 | 0.51115 | 3.416093329 | 3.416093329 | 3.416093329 | 0.42594166 | 0.782505439 | 1 | 0.425941661 | 0.425941661 | 0.425941661 | 8.020096747 | 0.904179607 | FALSE | TRUE | FALSE |
| VPS37B | 3.4098276 | 0.3693304 | 0.0097 | 0.8524049 | 3.409827606 | 3.409827606 | 0.26139302 | 0.836291547 | 1 | 0.261393024 | 0.261393024 | 0.261393024 | 13.04483019 | 1.11543843 | FALSE | TRUE | FALSE |
| UBXN6 | 3.403554 | 0.36955612 | 1 | 6.85513E+21 | 4.05886E+27 | 9.23132E+29 | 0.10487819 | 0.874759664 | 1 | 9.6555E+11 | 2.3781E+16 | 3.12224E+17 | 32.4524476 | 1.511247457 | FALSE | TRUE | FALSE |
| ACER2 | 3.4031279 | 0.36978163 | 0.5047 | 3.403127925 | 3.403127925 | 3.403127925 | 0.42368034 | 0.783209548 | 1 | 0.423680343 | 0.423680343 | 0.423680343 | 8.032300722 | 0.90483996 | FALSE | TRUE | FALSE |
| CERS6 | 3.3844592 | 0.37000727 | 0.25575 | 0.84656854 | 3.384459236 | 3.384459236 | 1.96005252 | 0.492319717 | 0.2617 | 0.49012422 | 1.960052524 | 1.960052524 | 1.726718643 | 0.237221578 | FALSE | FALSE | FALSE |
| LRRC47 | 3.3840273 | 0.3702327 | 0.0148 | 3.384027342 | 3.384027342 | 7.159619228 | 0.42106884 | 0.784141107 | 1 | 0.421068838 | 0.421068838 | 8.336808751 | 8.036755606 | 0.905080762 | FALSE | TRUE | FALSE |
| IL1F10 | 3.3817978 | 0.37045795 | 0.7535 | 3.38179782 | 32.12555944 | 32.12555944 | 0.43695924 | 0.778455167 | 1 | 0.436959242 | 0.436959242 | 0.436959242 | 7.739389612 | 0.88870671 | FALSE | TRUE | FALSE |
| RFPL4B | 3.3806612 | 0.37068302 | 0.2631 | 3.380661157 | 32.21155836 | 32.21155836 | 0.43604142 | 0.778852007 | 1 | 0.436041415 | 0.436041415 | 0.436041415 | 7.753073532 | 0.889473903 | FALSE | TRUE | FALSE |
| SP140L | 3.3698158 | 0.37090807 | 0.26095 | 0.362749776 | 3.369815844 | 7.179398601 | 0.5476577 | 0.734405631 | 0.5053 | 0.547657697 | 0.547657697 | 1.191434696 | 6.153142492 | 0.789096972 | FALSE | TRUE | FALSE |
| TAPBP | 3.3668014 | 0.37113296 | 0.2534 | 0.892011019 | 8.217412768 | 17.63929393 | 10.8536936 | 0.188770608 | 0.2608 | 1.1706627 | 10.85369356 | 23.18092786 | 0.310198682 | -0.508360052 | FALSE | FALSE | FALSE |
| TRH | 3.3665101 | 0.37135764 | 0.5009 | 3.366510114 | 3.366510114 | 3.366510114 | 0.41995585 | 0.784546067 | 0.00805 | 0.419955852 | 0.419955852 | 0.419955852 | 8.016342901 | 0.903976286 | FALSE | TRUE | FALSE |
| DBT | 3.3625446 | 0.37158218 | 0.5033 | 3.362544579 | 53.81398204 | 53.81398204 | 0.12820482 | 0.869410499 | 1 | 0.512709266 | 0.512709266 | 2.544064186 | 26.22791143 | 1.418763708 | FALSE | TRUE | FALSE |
| CELF4 | 3.3586685 | 0.37180659 | 0.5135 | 3.358668515 | 3.358668515 | 12.08240285 | 0.41606248 | 0.786093361 | 0.01485 | 0.416062483 | 0.416062483 | 8.221302731 | 8.072509899 | 0.907008586 | FALSE | TRUE | FALSE |
| RAB3IL1 | 3.3509752 | 0.37203091 | 0.8173 | 13.40314541 | 514.2892682 | 514.2892682 | 136.262748 | 0.020630677 | 0.599 | 136.262748 | 5363.075123 | 54079.65216 | 0.024592012 | -1.609205933 | FALSE | FALSE | FALSE |
| VTN | 3.3500459 | 0.37225505 | 0.81455 | 30.98982015 | 293.8614394 | 2854.716001 | 0.57110266 | 0.724173312 | 0.8734 | 5.204703456 | 48.65165456 | 48.65165456 | 5.865925933 | 0.768336575 | FALSE | TRUE | FALSE |
| FBXO48 | 3.3499702 | 0.37247897 | 0.5 | 1.855951161 | 3.349970179 | 3.349970179 | 0.42017585 | 0.784430523 | 1 | 0.420175854 | 0.420175854 | 0.420175854 | 7.972781278 | 0.90160985 | FALSE | TRUE | FALSE |
| FIBP | 3.3466813 | 0.37270275 | 0.50345 | 3.346681332 | 3.346681332 | 7.141028943 | 0.41875826 | 0.784978217 | 1 | 0.418758257 | 0.418758257 | 0.418758257 | 7.991917235 | 0.902650978 | FALSE | TRUE | FALSE |
| CYP2B6 | 3.3398734 | 0.37292644 | 0.81535 | 31.74502863 | 309.2904596 | 3087.051911 | 5.41175858 | 0.290062266 | 0.25625 | 0.579115053 | 5.411758584 | 5.411758584 | 0.617151212 | -0.209608414 | FALSE | FALSE | FALSE |
| PDXK | 3.3274544 | 0.37315013 | 0.69625 | 32.14959858 | 3236.154086 | 3236.154086 | 4.15302996 | 0.346653345 | 0.0083 | 4.153029961 | 4.153029961 | 4.153029961 | 0.80121126 | -0.096252956 | FALSE | FALSE | FALSE |
| C10orf67 | 3.3163622 | 0.37337382 | 0.75475 | 3.316362168 | 31.49805303 | 31.49805303 | 3.86981658 | 0.362816044 | 0.004 | 3.869816577 | 3.869816577 | 3.869816577 | 0.856981746 | -0.067028428 | FALSE | FALSE | FALSE |
| WAC | 3.3160102 | 0.37359731 | 0.01715 | 3.316010183 | 3.316010183 | 11.97157347 | 0.41296404 | 0.787280922 | 0.015 | 0.412964035 | 0.412964035 | 8.173797175 | 8.029779595 | 0.904703625 | FALSE | TRUE | FALSE |
| RBPJL | 3.3153059 | 0.37382059 | 0.32775 | 3.315305903 | 305.0736051 | 305.0736051 | 2.14205942 | 0.477077297 | 0.5078 | 4.98289105 | 190.1110624 | 190.1110624 | 1.547718923 | 0.189692093 | FALSE | FALSE | FALSE |
| JOSD2 | 3.3115906 | 0.37404374 | 0.50405 | 3.311590566 | 3.311590566 | 3.311590566 | 3.74495513 | 0.368448175 | 0.49985 | 0.402381274 | 3.744955127 | 3.744955127 | 0.88428044 | -0.053409982 | FALSE | FALSE | FALSE |
| GABRA6 | 3.304433 | 0.3742668 | 0.76215 | 13.21537065 | 846.5981583 | 4193.723104 | 2.76968498 | 0.429264335 | 0.8883 | 44.34104709 | 710.5836615 | 2845.656173 | 1.193071782 | 0.076666574 | FALSE | FALSE | FALSE |
| ELK4 | 3.2984075 | 0.37448977 | 0.0097 | 3.298407488 | 3.298407488 | 3.298407488 | 0.41278828 | 0.787365226 | 1 | 0.41278828 | 0.41278828 | 0.41278828 | 7.990555083 | 0.90257695 | FALSE | TRUE | FALSE |
| PDCD1LG2 | 3.29107 | 0.37471265 | 0.75255 | 3.29106999 | 31.29028255 | 31.29028255 | 0.42639897 | 0.782387632 | 0.00565 | 0.426398972 | 0.426398972 | 0.426398972 | 7.718287812 | 0.887520969 | FALSE | TRUE | FALSE |
| TMPRSS5 | 3.2879171 | 0.37493539 | 0.6595 | 30.57168355 | 1165.381702 | 11379.29343 | 0.60265212 | 0.710848315 | 0.51125 | 0.602652122 | 0.602652122 | 0.602652122 | 5.455746371 | 0.736854172 | FALSE | TRUE | FALSE |
| GLOD4 | 3.2869565 | 0.37515793 | 0.25655 | 0.821961446 | 3.28695648 | 3.28695648 | 1.9240757 | 0.496684565 | 0.1278 | 1.924075704 | 7.698256785 | 7.698256785 | 1.708330121 | 0.232571798 | FALSE | FALSE | FALSE |
| SLC7A10 | 3.2863911 | 0.37538028 | 0.9121 | 52.58866917 | 3372.597519 | 16623.9716 | 409.40284 | 0.005912528 | 0.2691 | 288.5573127 | 6563.043696 | 6563.043696 | 0.00802728 | -2.095431607 | FALSE | FALSE | FALSE |
| KRTAP19-6 | 3.285241 | 0.37560244 | 0.74825 | 3.285241 | 31.12151046 | 31.12151046 | 0.42677082 | 0.782210677 | 1 | 0.426770823 | 0.426770823 | 0.426770823 | 7.697904411 | 0.886372514 | FALSE | TRUE | FALSE |
| CMIP | 3.2798896 | 0.37582449 | 0.51195 | 3.279889602 | 3.279889602 | 64.44229303 | 3.71978522 | 0.370146871 | 0.02035 | 3.719785219 | 3.719785219 | 73.24023182 | 0.881741662 | -0.054658638 | FALSE | FALSE | FALSE |
| FN1 | 3.2744469 | 0.37604643 | 0.9699 | 473.2057804 | 30503.5111 | 263283.8123 | 4897.85038 | 0.000360898 | 0.1527 | 484.6442773 | 42728.26137 | 203888.7153 | 0.000668548 | -3.174867559 | FALSE | FALSE | FALSE |
| LIMS2 | 3.2667353 | 0.3762683 | 0.1541 | 0.204466668 | 3.266735266 | 13.0623834 | 0.38176129 | 0.798497777 | 0.5035 | 0.381761295 | 6.100971591 | 6.100971591 | 8.557010128 | 0.932322046 | FALSE | TRUE | FALSE |
| ESYT2 | 3.263874 | 0.37649002 | 0.5117 | 13.06002354 | 209.2620717 | 837.9661057 | 4.84228894 | 0.314493862 | 0.01725 | 1.210379388 | 4.842288939 | 10.2316852 | 0.674035367 | -0.171317315 | FALSE | FALSE | FALSE |
| ACTN4 | 3.2613767 | 0.37671157 | 0.5103 | 3.261376663 | 15.9187931 | 63.66520434 | 0.30352823 | 0.824216909 | 0.5139 | 0.303528226 | 0.303528226 | 5.94226249 | 10.74488758 | 1.031201876 | FALSE | TRUE | FALSE |
| SRD5A3 | 3.2605267 | 0.37693294 | 0.75545 | 3.260526724 | 31.15052759 | 31.15052759 | 3.46653104 | 0.385285421 | 0.68715 | 33.13588779 | 3264.69994 | 3264.69994 | 0.940573352 | -0.02660733 | FALSE | FALSE | FALSE |
| ELMOD3 | 3.2381238 | 0.37715449 | 0.13375 | 3.238123787 | 29.74799994 | 29.74799994 | 0.0629945 | 0.884015706 | 1 | 0.555893118 | 0.555893118 | 0.555893118 | 51.40328044 | 1.710990836 | FALSE | TRUE | FALSE |
| RP9 | 3.2372261 | 0.37737586 | 0.49965 | 0.345349176 | 3.237226118 | 3.237226118 | 0.40392454 | 0.791034108 | 1 | 0.403924537 | 0.403924537 | 0.403924537 | 8.014432946 | 0.9038728 | FALSE | TRUE | FALSE |
| GMFB | 3.2336938 | 0.37759709 | 0.00355 | 0.80843099 | 3.23369376 | 3.23369376 | 0.83783942 | 0.610650622 | 1 | 0.837839424 | 0.837839424 | 0.837839424 | 3.859562663 | 0.586538096 | FALSE | TRUE | FALSE |
| LCMT2 | 3.231563 | 0.37781815 | 0.3219 | 0.364910786 | 29.33650511 | 29.33650511 | 0.08476082 | 0.879189766 | 0.02225 | 0.084760821 | 0.084760821 | 1.666460579 | 38.12566943 | 1.581217478 | FALSE | TRUE | FALSE |
| PGPEP1 | 3.2313597 | 0.378039 | 0.50545 | 3.231359736 | 3.231359736 | 3.231359736 | 0.40170647 | 0.791655248 | 1 | 1.606774771 | 1.606774771 | 1.606774771 | 8.044081796 | 0.905476478 | FALSE | TRUE | FALSE |
| RFESD | 3.2288576 | 0.3782597 | 0.50955 | 3.228857586 | 3.228857586 | 3.228857586 | 1.58632283 | 0.527811749 | 0.75535 | 1.58632283 | 6.346485476 | 6.346485476 | 2.035435363 | 0.308657316 | FALSE | FALSE | FALSE |
| SGIP1 | 3.2201974 | 0.37848035 | 0.0215 | 3.220197395 | 3.220197395 | 63.37873293 | 0.40220948 | 0.791520535 | 1 | 0.402209482 | 0.402209482 | 7.931306922 | 8.006269213 | 0.903430189 | FALSE | TRUE | FALSE |
| CHMP1B | 3.2165426 | 0.37870085 | 0.504 | 3.216542576 | 3.216542576 | 3.216542576 | 0.39891216 | 0.79240641 | 1 | 0.398912159 | 0.398912159 | 0.398912159 | 8.063285377 | 0.906512031 | FALSE | TRUE | FALSE |
| STYXL1 | 3.2106284 | 0.37892126 | 0.69295 | 7.373148982 | 118.0390406 | 118.0390406 | 9.02481161 | 0.211462443 | 0.13235 | 3.914942603 | 36.10572929 | 36.10572929 | 0.355755723 | -0.448848104 | FALSE | FALSE | FALSE |
| ZFAND4 | 3.2089457 | 0.3791415 | 0.5112 | 3.208945684 | 29.90356046 | 29.90356046 | 0.54784146 | 0.734226735 | 0.5089 | 0.547841465 | 0.547841465 | 2.07416916 | 5.857434844 | 0.767707466 | FALSE | TRUE | FALSE |
| ASB16 | 3.2061654 | 0.37936158 | 0.5037 | 3.206165438 | 120.1905481 | 120.1905481 | 0.51321661 | 0.750351812 | 0.8754 | 4.724566124 | 178.4081553 | 178.4081553 | 6.247197285 | 0.795685221 | FALSE | TRUE | FALSE |
| PPP1CA | 3.2009971 | 0.37958155 | 0.50335 | 3.200997106 | 3.200997106 | 6.769891269 | 0.4000605 | 0.792085143 | 0.01035 | 0.400060501 | 0.400060501 | 7.938004885 | 8.001282561 | 0.903159608 | FALSE | TRUE | FALSE |
| TSPAN11 | 3.2004113 | 0.37980133 | 0.13235 | 0.800132528 | 12.80436985 | 12.80436985 | 0.24191235 | 0.84146115 | 1 | 3.869604997 | 15.48223813 | 15.48223813 | 13.22963173 | 1.121547755 | FALSE | TRUE | FALSE |
| TMEM241 | 3.199671 | 0.38002092 | 0.0069 | 3.199671036 | 3.199671036 | 3.199671036 | 0.40282692 | 0.791331637 | 1 | 0.402826918 | 0.402826918 | 0.402826918 | 7.943041771 | 0.899986846 | FALSE | TRUE | FALSE |
| TPPP2 | 3.1931658 | 0.38024042 | 0.9383 | 30.42778961 | 2979.29032 | 2979.29032 | 3074.41692 | 0.000749156 | 0.50215 | 3074.416923 | 331469.5859 | 331469.5859 | 0.001038625 | -2.983541295 | FALSE | FALSE | FALSE |
| CST2 | 3.1911625 | 0.38045975 | 0.4997 | 0.344822184 | 3.191162501 | 3.191162501 | 3.62311211 | 0.376855344 | 0.508 | 3.623112112 | 3.623112112 | 3.623112112 | 0.880779397 | -0.055132853 | FALSE | FALSE | FALSE |
| CLEC4D | 3.1904687 | 0.3806789 | 0.5035 | 3.190468697 | 3.190468697 | 3.190468697 | 0.40228594 | 0.791493571 | 1 | 0.40228594 | 0.40228594 | 0.40228594 | 7.930848127 | 0.899319633 | FALSE | TRUE | FALSE |
| UBXN4 | 3.1826894 | 0.38089798 | 0.7526 | 3.182689446 | 30.39215662 | 30.39215662 | 0.41289558 | 0.787309031 | 1 | 0.412895585 | 0.412895585 | 8.189471543 | 7.708218647 | 0.886954025 | FALSE | TRUE | FALSE |
| ANKRD24 | 3.1802561 | 0.3811169 | 0.51075 | 3.180256121 | 29.57108008 | 62.55296749 | 0.52784081 | 0.743907744 | 0.7567 | 0.527840811 | 4.828439697 | 10.40286048 | 6.025028864 | 0.779959132 | FALSE | TRUE | FALSE |
| OR13J1 | 3.1784291 | 0.38133566 | 0.50795 | 3.178429079 | 3.178429079 | 3.178429079 | 2695.08685 | 0.000845915 | 0.63685 | 2695.086854 | 290600.3424 | 3099368.625 | 0.001179342 | -2.92836024 | FALSE | FALSE | FALSE |
| TULP3 | 3.1703833 | 0.38155435 | 0.6168 | 5.515069485 | 202.7488181 | 811.1927788 | 0.68876082 | 0.6713097 | 0.318 | 0.172292297 | 2.75409875 | 2.75409875 | 4.603025014 | 0.663043335 | FALSE | TRUE | FALSE |
| CLEC10A | 3.1702052 | 0.38177285 | 0.4975 | 0.350154445 | 29.44018568 | 29.44018568 | 0.5133155 | 0.750234138 | 0.51085 | 0.513315501 | 0.513315501 | 0.513315501 | 6.175938929 | 0.790702993 | FALSE | TRUE | FALSE |
| NDST2 | 3.1679941 | 0.38199119 | 0.0041 | 0.09383145 | 0.09383145 | 1.837611878 | 0.03677747 | 0.8897249 | 0.02555 | 0.036777469 | 0.036777469 | 0.722060182 | 86.13953614 | 1.935202529 | FALSE | TRUE | FALSE |
| KIF12 | 3.1577676 | 0.38220951 | 0.51005 | 3.157767558 | 29.69929678 | 29.69929678 | 0.53755847 | 0.739527271 | 0.49985 | 0.059651005 | 0.537558465 | 1.181159128 | 5.874277425 | 0.768954453 | FALSE | TRUE | FALSE |
| ZNHIT1 | 3.1354952 | 0.38242802 | 0.50445 | 3.135495223 | 12.54180891 | 12.54180891 | 0.38866248 | 0.796183337 | 1 | 0.388662481 | 3.607612849 | 3.607612849 | 8.067398771 | 0.906733525 | FALSE | TRUE | FALSE |
| DEFB113 | 3.1323227 | 0.38264639 | 0.00255 | 0.331900332 | 3.132322697 | 3.132322697 | 0.3542806 | 0.807631797 | 1 | 0.354280596 | 0.354280596 | 0.354280596 | 8.841361137 | 0.94651913 | FALSE | TRUE | FALSE |
| LILRA1 | 3.1307905 | 0.38286458 | 0.50955 | 3.130790464 | 28.79379286 | 28.79379286 | 41.3293106 | 0.066567213 | 0.31295 | 4.455804653 | 392.9532664 | 392.9532664 | 0.075752303 | -1.120604159 | FALSE | FALSE | FALSE |
| DUS4L | 3.1305877 | 0.38308258 | 0.75115 | 3.130587745 | 29.88202263 | 29.88202263 | 0.40697408 | 0.789560502 | 1 | 0.406974078 | 0.406974078 | 0.406974078 | 7.692351713 | 0.886059133 | FALSE | TRUE | FALSE |
| SYNC | 3.1297102 | 0.3833004 | 0.25335 | 0.336658402 | 3.12971023 | 3.12971023 | 0.49702724 | 0.756091371 | 0.25495 | 0.497027238 | 4.645560357 | 4.645560357 | 6.296858587 | 0.79912394 | FALSE | TRUE | FALSE |

| | | | | | | | | | | | | | | | | | |
|---|---|---|---|---|---|---|---|---|---|---|---|---|---|---|---|---|---|
| HIGD1B | 3.1294376 | 0.38351802 | 0.49985 | 0.338781863 | 3.129437633 | 3.129437633 | 0.39625229 | 0.793311176 | 1 | 0.396252285 | 0.396252285 | 0.396252285 | 7.89758886 | 0.897494521 | FALSE | TRUE | FALSE |
| MIEF1 | 3.1282131 | 0.38373546 | 0.7544 | 3.128213079 | 29.85324975 | 61.81511322 | 0.40682807 | 0.789642942 | 0.0147 | 0.406828066 | 0.406828066 | 8.050715537 | 7.689275493 | 0.885885421 | FALSE | TRUE | FALSE |
| DROSHA | 3.127218 | 0.38395273 | 0.154 | 3.12721795 | 29.7488459 | 104.963337 | 0.48227987 | 0.762202395 | 0.6987 | 4.494363483 | 420.8928911 | 838.4038339 | 6.48423897 | 0.811859012 | FALSE | TRUE | FALSE |
| DDX59 | 3.1039941 | 0.3841702 | 0.80975 | 29.49491623 | 287.2887824 | 2866.66486 | 0.52642408 | 0.744361981 | 0.87555 | 4.913843385 | 47.04555184 | 47.04555184 | 5.896375657 | 0.770585145 | FALSE | TRUE | FALSE |
| RNPEPL1 | 3.101387 | 0.38438753 | 0.01635 | 3.101387013 | 3.101387013 | 11.23363039 | 0.37791363 | 0.799969697 | 1 | 3.53845481 | 3.53845481 | 7.476707476 | 8.20660272 | 0.91416341 | FALSE | TRUE | FALSE |
| ITGB8 | 3.0989895 | 0.38460469 | 0.20005 | 0.775061458 | 3.098989463 | 12.39402988 | 0.32334716 | 0.817595704 | 0.89495 | 1.292712681 | 20.67728106 | 82.72786862 | 9.58409376 | 0.981551054 | FALSE | TRUE | FALSE |
| PCBD1 | 3.0967964 | 0.3848217 | 0.9051 | 49.54112782 | 1177.495127 | 4710.207227 | 0.42270412 | 0.783559854 | 0.0022 | 0.422704122 | 0.422704122 | 0.422704122 | 7.326156214 | 0.864876175 | FALSE | TRUE | FALSE |
| SLC22A25 | 3.0916295 | 0.38503861 | 0.3162 | 3.091629456 | 275.8639453 | 275.8639453 | 40.2707075 | 0.068195706 | 0.6597 | 40.27070751 | 3819.275839 | 38544.57741 | 0.076771173 | -1.114801823 | FALSE | FALSE | FALSE |
| INTS3 | 3.0883093 | 0.38525537 | 1 | 3.1574E+21 | 1.88383E+27 | 1.71962E+30 | 0.02568893 | 0.892231326 | 1 | 3.4938E+36 | 1.87135E+45 | 1.24881E+49 | 120.2194699 | 2.079974809 | FALSE | TRUE | FALSE |
| SLC17A1 | 3.0852169 | 0.38547199 | 0.13075 | 3.085216911 | 28.42047014 | 28.42047014 | 0.06015852 | 0.88472509 | 1 | 0.532161878 | 0.532161878 | 1.194150353 | 51.284789 | 1.709988573 | FALSE | TRUE | FALSE |
| TMEM30B | 3.0849995 | 0.38568842 | 0.013 | 3.084999515 | 3.084999515 | 6.503831633 | 0.38679416 | 0.796850196 | 1 | 0.38679416 | 0.38679416 | 7.664602961 | 7.97581721 | 0.901775192 | FALSE | TRUE | FALSE |
| KLF13 | 3.0829684 | 0.38590468 | 0.5084 | 3.082968389 | 3.082968389 | 3.082968389 | 0.60907137 | 0.708094726 | 1 | 0.609071367 | 0.609071367 | 0.609071367 | 5.061752294 | 0.704300888 | FALSE | TRUE | FALSE |
| FAS | 3.0788798 | 0.38612082 | 0.2476 | 3.078879771 | 29.71740202 | 29.71740202 | 0.3978293 | 0.792806559 | 0.0062 | 0.397829295 | 0.397829295 | 0.397829295 | 7.739198208 | 0.888695969 | FALSE | TRUE | FALSE |
| SLC35G5 | 3.0780479 | 0.38633678 | 0.00005 | 0.090748264 | 0.090748264 | 0.090748264 | 0.01218011 | 0.896028646 | 1 | 0.012180107 | 0.012180107 | 0.012180107 | 252.7110774 | 2.402624279 | FALSE | TRUE | FALSE |
| TMEM208 | 3.073282 | 0.38655263 | 0.50205 | 3.073282032 | 3.073282032 | 3.073282032 | 0.38981055 | 0.795796553 | 1 | 0.389810555 | 0.389810555 | 0.389810555 | 7.88404007 | 0.896748823 | FALSE | TRUE | FALSE |
| OR6N2 | 3.0729876 | 0.38676829 | 0.00845 | 3.072987599 | 3.072987599 | 3.072987599 | 0.37642294 | 0.800389732 | 1 | 3.506911466 | 3.506911466 | 3.506911466 | 8.163656621 | 0.911884729 | FALSE | TRUE | FALSE |
| SLC5A8 | 3.070337 | 0.38698379 | 0.66255 | 5.664469431 | 437.467241 | 1750.701539 | 0.01329305 | 0.895717463 | 0.93835 | 0.05313848 | 0.849774922 | 1.051482002 | 230.9731607 | 2.363561517 | FALSE | TRUE | FALSE |
| ADA | 3.0702006 | 0.38719911 | 0.50425 | 4.08412744 | 138.6449013 | 323.9388907 | 1.30493315 | 0.554196471 | 0.67825 | 2.984765528 | 112.045954 | 448.1661184 | 2.352764632 | 0.371578483 | FALSE | FALSE | FALSE |
| NUTM2D | 3.0689542 | 0.38741425 | 0.5114 | 3.068954192 | 3.068954192 | 6.520354373 | 0.38772872 | 0.796466015 | 1 | 0.38772872 | 0.38772872 | 7.671623763 | 7.915209878 | 0.898462435 | FALSE | TRUE | FALSE |
| OR52H1 | 3.0654371 | 0.38762925 | 0.5077 | 3.065437072 | 3.065437072 | 3.065437072 | 0.37587235 | 0.800463678 | 1 | 3.498865826 | 3.498865826 | 3.498865826 | 8.155526894 | 0.911452024 | FALSE | TRUE | FALSE |
| FGF5 | 3.0583739 | 0.38784419 | 0.2584 | 0.764803401 | 3.05837392 | 3.05837392 | 0.11688063 | 0.871992738 | 0.00635 | 0.116880626 | 0.116880626 | 0.116880626 | 26.16664567 | 1.417748053 | FALSE | TRUE | FALSE |
| TNFAIP8 | 3.0555485 | 0.38805898 | 0.2505 | 0.316407272 | 3.055548479 | 3.055548479 | 0.037723 | 0.889520493 | 0.00455 | 0.037722997 | 0.037722997 | 0.037722997 | 80.99962081 | 1.908482986 | FALSE | TRUE | FALSE |
| DET1 | 3.0544024 | 0.3882736 | 0.5027 | 3.054402392 | 3.054402392 | 10.973793 | 0.38240234 | 0.798296459 | 1 | 0.382402336 | 0.382402336 | 7.556622042 | 7.98740516 | 0.902405715 | FALSE | TRUE | FALSE |
| TMEM106A | 3.0530532 | 0.38848805 | 0.5006 | 3.053053193 | 3.053053193 | 3.053053193 | 0.38756628 | 0.796491675 | 0.0057 | 0.387566279 | 0.387566279 | 0.387566279 | 7.877499553 | 0.896388387 | FALSE | TRUE | FALSE |
| CEP85L | 3.0518688 | 0.38870232 | 0.5033 | 3.051868819 | 28.14891334 | 28.14891334 | 4.36199551 | 0.33694328 | 0.6854 | 4.361995509 | 386.657204 | 386.657204 | 0.699649693 | -0.155119352 | FALSE | FALSE | FALSE |
| CBFB | 3.0471787 | 0.38891649 | 0.00665 | 3.047178715 | 3.047178715 | 3.047178715 | 0.46542254 | 0.768838998 | 1 | 0.465422538 | 0.465422538 | 0.465422538 | 6.547123238 | 0.816050516 | FALSE | TRUE | FALSE |
| ONECUT1 | 3.0454578 | 0.38913048 | 0.01625 | 3.045457795 | 3.045457795 | 6.386155204 | 0.3818585 | 0.798447486 | 1 | 0.3818585 | 0.3818585 | 7.544994546 | 7.975356834 | 0.901750123 | FALSE | TRUE | FALSE |
| LDHA | 3.0411249 | 0.38934437 | 0.1281 | 0.190100427 | 3.041124886 | 3.041124886 | 0.62248516 | 0.702588401 | 0.00725 | 0.622485161 | 0.622485161 | 0.622485161 | 4.885457641 | 0.688905252 | FALSE | TRUE | FALSE |
| ITIH4 | 3.0408997 | 0.38955806 | 0.35665 | 0.759919731 | 12.17152383 | 48.72998325 | 296.467033 | 0.008341447 | 0.32985 | 92.4016059 | 5925.799479 | 29048.02098 | 0.010257126 | -1.98897431 | FALSE | FALSE | FALSE |
| DMRT3 | 3.0297966 | 0.38977176 | 0.50895 | 3.029796627 | 28.02793972 | 28.02793972 | 0.47558784 | 0.764862814 | 0.0133 | 0.475587845 | 0.475587845 | 2.353637916 | 6.37063512 | 0.804182731 | FALSE | TRUE | FALSE |
| HOXA9 | 3.022556 | 0.38998539 | 0.07235 | 0.755856015 | 12.08977314 | 12.08977314 | 0.27743379 | 0.831707155 | 0.75505 | 0.277433786 | 1.109250426 | 1.109250426 | 10.89469336 | 1.037215011 | FALSE | TRUE | FALSE |
| CSNK2B | 3.0186898 | 0.3901989 | 0.0069 | 3.018689754 | 3.018689754 | 3.018689754 | 0.78922229 | 0.624274123 | 1 | 0.789222294 | 0.789222294 | 0.789222294 | 3.824891638 | 0.582619136 | FALSE | TRUE | FALSE |
| GAS7 | 3.0160643 | 0.39041226 | 0.0157 | 3.016064268 | 3.016064268 | 6.389924951 | 0.38120432 | 0.798798973 | 1 | 0.381204323 | 0.381204323 | 7.538680987 | 7.911936158 | 0.898282774 | FALSE | TRUE | FALSE |
| TLR10 | 3.0124158 | 0.39062549 | 0.5116 | 3.012415848 | 28.31703547 | 28.31703547 | 0.5002073 | 0.755078493 | 0.75195 | 0.500207298 | 4.620306576 | 9.889912705 | 6.02233486 | 0.7797649 | FALSE | TRUE | FALSE |
| DDX25 | 3.0093818 | 0.39083858 | 0.25145 | 0.332325971 | 3.009381815 | 3.009381815 | 0.48788577 | 0.759768078 | 0.7509 | 0.487885768 | 4.454814039 | 4.454814039 | 6.168209882 | 0.790159143 | FALSE | TRUE | FALSE |
| ENTPD7 | 3.0089702 | 0.39105149 | 0.8111 | 27.7757766 | 262.8273555 | 2547.841834 | 4.73100933 | 0.318703953 | 0.5106 | 4.731009326 | 44.14021478 | 44.14021478 | 0.636010202 | -0.196535918 | FALSE | FALSE | FALSE |
| ARHGEF18 | 3.0025987 | 0.39126432 | 0.50915 | 3.002598707 | 28.79457507 | 58.67280144 | 0.47748612 | 0.76399458 | 0.88075 | 4.485669003 | 43.22182508 | 87.89552224 | 6.288347583 | 0.798536539 | FALSE | TRUE | FALSE |
| MSRB3 | 3.0001693 | 0.391477 | 0.0042 | 0.320601056 | 3.000169342 | 3.000169342 | 304.793707 | 0.008168997 | 0.00235 | 30.9437849 | 304.793707 | 304.793707 | 0.009843278 | -2.006860227 | FALSE | FALSE | FALSE |
| ALPL | 3.0001138 | 0.39168949 | 0.15485 | 0.187699772 | 3.000113769 | 14.80424382 | 0.11337501 | 0.872669431 | 0.5021 | 0.113375014 | 0.453189969 | 1.811972921 | 26.46186013 | 1.42262037 | FALSE | TRUE | FALSE |
| SLC25A2 | 2.9956398 | 0.39190187 | 0.49885 | 0.320508619 | 28.71790565 | 28.71790565 | 1.9012008 | 0.498918453 | 0.72205 | 4.469973441 | 71.46546938 | 285.8608748 | 1.575656697 | 0.1974616 | FALSE | FALSE | FALSE |
| SIRPD | 2.9955704 | 0.39211406 | 0.4955 | 0.324302074 | 2.995570426 | 2.995570426 | 0.38131301 | 0.79869868 | 1 | 0.381313011 | 0.381313011 | 0.381313011 | 7.855935511 | 0.89519791 | FALSE | TRUE | FALSE |
| XCR1 | 2.9953124 | 0.39232607 | 0.75355 | 2.995312401 | 28.7441717 | 28.7441717 | 29.8756307 | 0.093113084 | 0.5061 | 293.7904761 | 2961.343027 | 30578.18138 | 0.100259386 | -0.998874958 | FALSE | FALSE | FALSE |
| RABL2A | 2.9899921 | 0.39253797 | 0.49935 | 0.318183795 | 2.989992129 | 2.989992129 | 32.0139766 | 0.086741204 | 0.2526 | 3.318289941 | 32.01397659 | 32.01397659 | 0.093396461 | -1.029669578 | FALSE | FALSE | FALSE |
| DNAJB14 | 2.9897436 | 0.39274969 | 0.74885 | 2.989743629 | 28.7237633 | 28.7237633 | 0.37282543 | 0.801395482 | 1 | 1.491134764 | 13.99481148 | 13.99481148 | 8.019151604 | 0.904128424 | FALSE | TRUE | FALSE |
| LSR | 2.9849475 | 0.39296131 | 0.1441 | 2.984947497 | 28.00312915 | 58.61240111 | 33.4296766 | 0.083289452 | 0.9468 | 3174.028558 | 3500123.741 | 37815185.75 | 0.089290349 | -1.049195479 | FALSE | FALSE | FALSE |
| SUPT7L | 2.98185 | 0.39317279 | 0.50615 | 2.981850012 | 2.981850012 | 6.383941408 | 0.37908305 | 0.79952299 | 1 | 0.379083053 | 0.379083053 | 7.524770585 | 7.865954414 | 0.895751425 | FALSE | TRUE | FALSE |
| CENPN | 2.9793324 | 0.39338412 | 0.00895 | 2.979332383 | 2.979332383 | 2.979332383 | 0.37890323 | 0.799647284 | 1 | 0.378903234 | 0.378903234 | 0.378903234 | 7.863042894 | 0.895590645 | FALSE | TRUE | FALSE |
| SYT14 | 2.9782994 | 0.39359528 | 0.25595 | 2.978299427 | 28.64850384 | 28.64850384 | 0.38738238 | 0.796619869 | 1 | 0.387382384 | 0.387382384 | 7.689766297 | 7.68826758 | 0.88582849 | FALSE | TRUE | FALSE |
| ELOVL2 | 2.9772225 | 0.39380628 | 0.00735 | 0.14974464 | 0.14974464 | 0.14974464 | 0.18665093 | 0.855348407 | 1 | 0.186650932 | 0.186650932 | 0.186650932 | 15.95075057 | 1.202781124 | FALSE | TRUE | FALSE |
| ZNF266 | 2.9682734 | 0.39401724 | 0.5031 | 26.62900037 | 2303.324888 | 4848.249755 | 52.8130071 | 0.052546206 | 0.3502 | 5.796930557 | 492.9338821 | 4710.804691 | 0.056203453 | -1.250237002 | FALSE | FALSE | FALSE |
| TRIB3 | 2.9680718 | 0.39422802 | 0.3184 | 2.968071802 | 263.878251 | 263.878251 | 0.52750986 | 0.744031791 | 0.50965 | 0.527509855 | 0.527509855 | 1.169120059 | 5.626571282 | 0.750243825 | FALSE | TRUE | FALSE |
| MGAT4C | 2.9647674 | 0.39443866 | 0.25525 | 2.964767415 | 28.34968284 | 28.34968284 | 0.38752227 | 0.796517327 | 1 | 0.387522273 | 0.387522273 | 7.692062444 | 7.650572938 | 0.88369396 | FALSE | TRUE | FALSE |
| REG4 | 2.9604129 | 0.3946492 | 0.00495 | 2.96041289 | 2.96041289 | 2.96041289 | 0.37760442 | 0.800043952 | 1 | 0.377604418 | 0.377604418 | 0.377604418 | 7.839984783 | 0.89431522 | FALSE | TRUE | FALSE |
| OR5W2 | 2.9565672 | 0.39485961 | 0.25555 | 2.956567177 | 28.20399524 | 28.20399524 | 0.38712271 | 0.796696704 | 1 | 0.387122714 | 0.387122714 | 0.387122714 | 7.637286758 | 0.882939097 | FALSE | TRUE | FALSE |
| CLDN6 | 2.9544098 | 0.39506987 | 0.3218 | 2.954409765 | 47.2438754 | 47.2438754 | 0.44362911 | 0.776230361 | 0.74655 | 0.443629106 | 1.773937934 | 1.773937934 | 6.659639159 | 0.823450698 | FALSE | TRUE | FALSE |
| FSIP1 | 2.9526463 | 0.39527998 | 0.5039 | 27.0997164 | 254.962373 | 2457.462893 | 37.928452 | 0.072355344 | 0.7743 | 359.652119 | 34749.67358 | 69293.8646 | 0.077847792 | -1.1087537 | FALSE | FALSE | FALSE |
| CMTM5 | 2.9516298 | 0.39548992 | 0.0046 | 2.951629791 | 2.951629791 | 2.951629791 | 3.38003511 | 0.39011079 | 0.49855 | 0.365106761 | 3.380035106 | 3.380035106 | 0.873254182 | -0.058859326 | FALSE | FALSE | FALSE |
| DYRK1B | 2.9496719 | 0.3956997 | 0.75605 | 2.949671948 | 28.25246918 | 58.14339221 | 0.38534853 | 0.797537899 | 1 | 0.38534853 | 0.38534853 | 7.609092252 | 7.654556119 | 0.883920011 | FALSE | TRUE | FALSE |
| JAKMIP2 | 2.9494931 | 0.3959093 | 0.7602 | 2.949493093 | 28.35591123 | 58.05674768 | 0.37244393 | 0.80161488 | 1 | 3.49757463 | 3.49757463 | 68.9760721 | 7.919294341 | 0.898686485 | FALSE | TRUE | FALSE |
| EIF3C | 2.946031 | 0.39611878 | 0.51805 | 2.946030965 | 2.946030965 | 57.60118305 | 3.21650907 | 0.398675066 | 0.50795 | 3.216509073 | 3.216509073 | 63.05926428 | 0.915909422 | -0.038147473 | FALSE | FALSE | FALSE |
| MARK2 | 2.9382926 | 0.3963282 | 0.5107 | 2.938292572 | 2.938292572 | 57.65967239 | 0.36924229 | 0.80241513 | 1 | 0.369242292 | 0.369242292 | 7.262261084 | 7.957627383 | 0.900783599 | FALSE | TRUE | FALSE |
| TP53TG5 | 2.9296608 | 0.39653759 | 0.50155 | 2.929660812 | 2.929660812 | 6.289037409 | 0.3621422 | 0.804941596 | 1 | 3.360382528 | 3.360382528 | 3.360382528 | 8.08980774 | 0.9079382 | FALSE | TRUE | FALSE |
| IGSF6 | 2.9289444 | 0.3967468 | 0.5073 | 2.92894442 | 2.92894442 | 2.92894442 | 3.36051278 | 0.391476985 | 0.50405 | 3.360512779 | 3.360512779 | 3.360512779 | 0.871576635 | -0.059694421 | FALSE | FALSE | FALSE |
| ZPLD1 | 2.9246998 | 0.39695591 | 0.87765 | 27.64783877 | 2664.253264 | 10655.42283 | 0.11995639 | 0.871236443 | 0.5052 | 1.11175828 | 1.11175828 | 2.381926194 | 24.38135826 | 1.387057896 | FALSE | TRUE | FALSE |
| ACCS | 2.9217164 | 0.39716488 | 0.0727 | 0.730494614 | 11.68873728 | 11.68873728 | 0.26274425 | 0.835894136 | 0.69025 | 1.050693166 | 16.81461223 | 16.81461223 | 11.1200012 | 1.046104834 | FALSE | TRUE | FALSE |
| ST6GALNAC( | 2.9167637 | 0.39737375 | 0.50165 | 2.916763667 | 2.916763667 | 6.217817483 | 0.37099142 | 0.802100638 | 1 | 0.370991421 | 0.370991421 | 7.355734666 | 7.862078489 | 0.895537375 | FALSE | TRUE | FALSE |
| MGAT5 | 2.9139219 | 0.39758248 | 0.5126 | 2.913921887 | 2.913921887 | 10.47089259 | 0.36735593 | 0.803208898 | 1 | 0.367355925 | 0.367355925 | 7.256315817 | 7.932148865 | 0.899390856 | FALSE | TRUE | FALSE |
| ADIPOR2 | 2.9132176 | 0.39779105 | 0.50015 | 2.913217614 | 2.913217614 | 2.913217614 | 0.368408 | 0.802728616 | 1 | 0.368408001 | 0.368408001 | 0.368408001 | 7.907585084 | 0.898043874 | FALSE | TRUE | FALSE |
| CAPN1 | 2.9059912 | 0.39799955 | 0.51005 | 11.62376047 | 186.1129671 | 1174.558346 | 2102.95628 | 0.001131856 | 0.41635 | 2102.956278 | 21797.12938 | 135518.8397 | 0.00138186 | -2.859535955 | FALSE | FALSE | FALSE |
| ZFPL1 | 2.9055702 | 0.39820788 | 0.2578 | 0.726475789 | 2.905570206 | 3.601359178 | 0.22482117 | 0.846158393 | 0.50415 | 0.898998872 | 0.898998872 | 0.898998872 | 12.92391729 | 1.11139417 | FALSE | TRUE | FALSE |
| NDUFA9 | 2.9048447 | 0.39841603 | 0.6431 | 6.861591494 | 265.6699888 | 1062.317426 | 4.72467652 | 0.319539545 | 0.59785 | 4.724676522 | 179.226729 | 716.9098136 | 0.614824039 | -0.21124916 | FALSE | FALSE | FALSE |
| TPRG1 | 2.9040744 | 0.39862402 | 0.8784 | 27.52727271 | 1070.494205 | 1070.494205 | 3.22830887 | 0.398229086 | 0.8149 | 122.6865248 | 2985.450237 | 11941.75543 | 0.899565231 | -0.045967339 | FALSE | FALSE | FALSE |
| RARB | 2.8867389 | 0.39883212 | 0.2581 | 0.722115527 | 2.886738925 | 2.886738925 | 0.10883433 | 0.873682431 | 0.0126 | 0.108834325 | 0.108834325 | 2.15958928 | 26.52415883 | 1.42364162 | FALSE | TRUE | FALSE |

| | | | | | | | | | | | | | | | | | |
|---|---|---|---|---|---|---|---|---|---|---|---|---|---|---|---|---|---|
| METTL24 | 2.8797366 | 0.39904016 | 0.49945 | 2.879736555 | 265.3271225 | 265.3271225 | 306.761479 | 0.00808382 | 0.0033 | 31.45450037 | 306.7614785 | 306.7614785 | 0.009387543 | -2.027448063 | FALSE | FALSE | FALSE |
| AP5S1 | 2.8783504 | 0.39924804 | 0.5086 | 2.878350376 | 2.878350376 | 2.878350376 | 0.33579451 | 0.813588681 | 1 | 3.103360042 | 286.3771591 | 286.3771591 | 8.571761212 | 0.933070064 | FALSE | TRUE | FALSE |
| CLEC4E | 2.8761963 | 0.39945578 | 0.49845 | 0.311122145 | 2.876196288 | 2.876196288 | 0.36766721 | 0.802993075 | 1 | 0.367667213 | 0.367667213 | 0.367667213 | 7.822825069 | 0.893363619 | FALSE | TRUE | FALSE |
| SKA1 | 2.8731549 | 0.39966339 | 0.74565 | 2.873154898 | 27.25193143 | 27.25193143 | 0.35621816 | 0.807012451 | 1 | 3.302396488 | 31.421927 | 31.421927 | 8.065717081 | 0.906642984 | FALSE | TRUE | FALSE |
| SNTB1 | 2.8698432 | 0.39987087 | 0.13975 | 1.253650227 | 11.47882363 | 14.17427128 | 0.11011466 | 0.873309377 | 0.5091 | 0.440344563 | 0.440344563 | 3.730285456 | 26.06231823 | 1.416013043 | FALSE | TRUE | FALSE |
| SEC31A | 2.8639353 | 0.40007827 | 0.02005 | 0.317181311 | 2.86393533 | 14.0226582 | 0.29130083 | 0.827687461 | 0.51445 | 0.291300825 | 0.291300825 | 5.718271065 | 9.831538676 | 0.992621492 | FALSE | TRUE | FALSE |
| MTFR1L | 2.8620244 | 0.40028552 | 0.51065 | 2.862024368 | 2.862024368 | 2.862024368 | 0.3650454 | 0.804185899 | 1 | 0.365045396 | 0.365045396 | 0.365045396 | 7.840187538 | 0.894326451 | FALSE | TRUE | FALSE |
| PURB | 2.861767 | 0.40049259 | 0.5032 | 2.861767015 | 2.861767015 | 6.001497827 | 0.32827057 | 0.815876109 | 1 | 0.328270574 | 0.328270574 | 0.328270574 | 8.717708009 | 0.940402319 | FALSE | TRUE | FALSE |
| CAPZA1 | 2.8472197 | 0.40069973 | 0.5065 | 2.847219708 | 2.847219708 | 2.847219708 | 0.74962961 | 0.641470762 | 1 | 0.749629607 | 0.749629607 | 0.749629607 | 3.798168699 | 0.57957425 | FALSE | TRUE | FALSE |
| ASL | 2.8462442 | 0.40090671 | 0.70955 | 3.518393776 | 729.0654634 | 2918.508648 | 102.821387 | 0.028475796 | 0.50675 | 102.8213871 | 14965.6656 | 59920.74989 | 0.027681441 | -1.557811302 | FALSE | FALSE | FALSE |
| GALNT5 | 2.8460791 | 0.40111351 | 0.7215 | 11.38023678 | 981.0735528 | 9440.44409 | 0.07351116 | 0.881678597 | 0.81605 | 2.600522958 | 54.89316863 | 219.5040873 | 38.7162863 | 1.587893693 | FALSE | TRUE | FALSE |
| GNS | 2.8454324 | 0.40132014 | 0.06765 | 0.17816745 | 2.845432358 | 2.845432358 | 0.00937371 | 0.896924191 | 0.87795 | 0.149674188 | 0.598311652 | 0.743016589 | 303.5546041 | 2.482236824 | FALSE | TRUE | FALSE |
| DEFB135 | 2.8451958 | 0.40152659 | 0.75085 | 2.845195805 | 26.94879543 | 26.94879543 | 0.37603396 | 0.800439035 | 1 | 0.376033963 | 0.376033963 | 0.376033963 | 7.566326677 | 0.878885088 | FALSE | TRUE | FALSE |
| EPHA6 | 2.8427699 | 0.4017329 | 0.136 | 0.299742572 | 2.842769941 | 5.864006717 | 0.00762237 | 0.897401115 | 1 | 0.007622373 | 0.007622373 | 0.149512182 | 372.9507694 | 2.571651507 | FALSE | TRUE | FALSE |
| NR2E1 | 2.839999 | 0.40193908 | 0.01305 | 2.839999043 | 2.839999043 | 5.960792135 | 0.34710531 | 0.809890751 | 1 | 3.276790104 | 3.276790104 | 6.880282925 | 8.181952257 | 0.912856941 | FALSE | TRUE | FALSE |
| PTGR2 | 2.8370318 | 0.40214513 | 0.44655 | 0.709414829 | 26.34927418 | 26.34927418 | 3.91084943 | 0.360586822 | 0.5087 | 3.910849427 | 62.59563267 | 62.59563267 | 0.725425982 | -0.139406894 | FALSE | FALSE | FALSE |
| MRPL16 | 2.8364368 | 0.40235101 | 0.50345 | 2.836436776 | 2.836436776 | 2.836436776 | 3.26690437 | 0.39644111 | 0.5035 | 3.266904371 | 3.266904371 | 3.266904371 | 0.868233794 | -0.061363314 | FALSE | FALSE | FALSE |
| CHCHD4 | 2.8355588 | 0.40255672 | 0.50515 | 2.835558769 | 2.835558769 | 2.835558769 | 0.35117377 | 0.80876808 | 1 | 3.26569319 | 3.26569319 | 3.26569319 | 8.074517636 | 0.907116587 | FALSE | TRUE | FALSE |
| OR10A4 | 2.8354332 | 0.40276225 | 0.7497 | 2.835433174 | 26.88157159 | 26.88157159 | 0.37469506 | 0.800955193 | 1 | 0.37469506 | 0.37469506 | 0.37469506 | 7.56730867 | 0.878941449 | FALSE | TRUE | FALSE |
| ZNF34 | 2.8248036 | 0.40296779 | 0.51575 | 2.82480362 | 2.82480362 | 6.000485375 | 0.35994882 | 0.805668055 | 1 | 0.359948821 | 0.359948821 | 7.131612005 | 7.847792406 | 0.894747506 | FALSE | TRUE | FALSE |
| TRIM10 | 2.8242978 | 0.40317316 | 0.0115 | 0.307835248 | 2.824297809 | 6.089541594 | 0.05341373 | 0.885979402 | 1 | 0.213595575 | 0.213595575 | 1.058007443 | 52.87587959 | 1.723257605 | FALSE | TRUE | FALSE |
| ZDHHC24 | 2.8162873 | 0.40337849 | 0.5039 | 26.90431805 | 263.6158013 | 263.6158013 | 0.37082646 | 0.802149081 | 1 | 3.466064871 | 3.466064871 | 3.466064871 | 7.594623405 | 0.880506243 | FALSE | TRUE | FALSE |
| NRBP2 | 2.8144696 | 0.40358367 | 0.0093 | 0.301761711 | 2.814469632 | 2.814469632 | 0.35853183 | 0.806157043 | 0.00795 | 0.35853183 | 0.35853183 | 0.35853183 | 7.849985413 | 0.89486885 | FALSE | TRUE | FALSE |
| VPS28 | 2.8118288 | 0.40378871 | 0.5021 | 2.811828835 | 2.811828835 | 2.811828835 | 0.34764937 | 0.809801422 | 1 | 0.347649368 | 3.243365433 | 3.243365433 | 8.088117216 | 0.907847437 | FALSE | TRUE | FALSE |
| ACADVL | 2.8087125 | 0.40399363 | 0.40035 | 1.528296665 | 97.68375542 | 481.4398817 | 0.15484763 | 0.863293772 | 0.1587 | 0.038747042 | 0.618983398 | 3.057453817 | 18.13855692 | 1.258602732 | FALSE | TRUE | FALSE |
| OVGP1 | 2.7936403 | 0.40419864 | 0.256 | 0.310726122 | 2.793640297 | 6.134459674 | 0.4728057 | 0.765964437 | 0.50805 | 0.472805704 | 0.472805704 | 1.056712663 | 5.908643395 | 0.77148778 | FALSE | TRUE | FALSE |
| TAAR2 | 2.7927656 | 0.40440348 | 0.6908 | 25.81496355 | 594.7603946 | 2378.630985 | 9.06678604 | 0.210450451 | 0.3142 | 3.895915884 | 346.1765326 | 346.1765326 | 0.308021564 | -0.511418878 | FALSE | FALSE | FALSE |
| GFRA1 | 2.7906072 | 0.40460817 | 0.5044 | 2.790607237 | 2.790607237 | 5.91523708 | 0.3555385 | 0.807265265 | 0.01195 | 0.355538495 | 0.355538495 | 7.04451535 | 7.84895946 | 0.894812086 | FALSE | TRUE | FALSE |
| C3orf62 | 2.7871724 | 0.40481275 | 0.7545 | 2.787172396 | 26.36814415 | 26.36814415 | 3.21382703 | 0.399342651 | 0.7511 | 3.213827032 | 30.50917381 | 30.50917381 | 0.867244058 | -0.061858667 | FALSE | FALSE | FALSE |
| FBXO41 | 2.7842014 | 0.40501721 | 0.5092 | 2.78420137 | 2.78420137 | 9.999313598 | 0.31446755 | 0.820542699 | 1 | 2.9608426 | 28.61141617 | 58.44604638 | 8.853700176 | 0.947124811 | FALSE | TRUE | FALSE |
| RPRD1A | 2.7746867 | 0.40522165 | 0.5053 | 2.774686664 | 2.774686664 | 2.774686664 | 0.32693777 | 0.816397565 | 1 | 0.326937765 | 0.326937765 | 0.326937765 | 8.486895547 | 0.928748857 | FALSE | TRUE | FALSE |
| ARF6 | 2.7738014 | 0.40542593 | 0.5021 | 2.773801416 | 2.773801416 | 2.773801416 | 0.73243256 | 0.650181824 | 1 | 0.732432564 | 0.732432564 | 0.732432564 | 3.787108261 | 0.57830772 | FALSE | TRUE | FALSE |
| SLC4A1AP | 2.7716834 | 0.40563006 | 0.50515 | 2.771683383 | 26.1764721 | 54.51864447 | 0.05446822 | 0.88577902 | 0.0202 | 0.054468217 | 0.054468217 | 1.073511574 | 50.88625128 | 1.706600458 | FALSE | TRUE | FALSE |
| ITSN1 | 2.7695798 | 0.40583406 | 1 | 1.59423E+21 | 1.23793E+27 | 3.3113E+30 | 66373.8066 | 2.95137E-05 | 1 | 2.00274E+30 | 4.42799E+37 | 2.0511E+41 | 4.1727E-05 | -4.379582845 | FALSE | FALSE | FALSE |
| PANK3 | 2.7663862 | 0.40603793 | 0.25575 | 0.691991144 | 2.766386202 | 2.766386202 | 0.9534828 | 0.592416941 | 0.50965 | 0.953482796 | 0.953482796 | 0.953482796 | 2.901348838 | 0.462599949 | FALSE | TRUE | FALSE |
| IGFBPL1 | 2.7652275 | 0.40624164 | 0.7533 | 11.05946735 | 105.6736978 | 105.6736978 | 1.36982003 | 0.549135443 | 0.87265 | 3.195964927 | 122.4189127 | 122.4189127 | 2.018679388 | 0.305067349 | FALSE | FALSE | FALSE |
| EPYC | 2.7638088 | 0.4064452 | 0.878 | 26.37839913 | 1032.794041 | 1032.794041 | 0.38053018 | 0.799149157 | 0.0076 | 0.380530179 | 0.380530179 | 0.380530179 | 7.263047648 | 0.861118893 | FALSE | TRUE | FALSE |
| ASAH2 | 2.7631276 | 0.4066486 | 0.51085 | 2.763127557 | 2.763127557 | 54.49740124 | 0.32917354 | 0.815582764 | 1 | 3.096476573 | 29.89479388 | 61.17383434 | 8.39413639 | 0.923976021 | FALSE | TRUE | FALSE |
| BPIFA3 | 2.7607462 | 0.40685186 | 0.00535 | 0.299591669 | 2.760746164 | 2.760746164 | 0.3554136 | 0.807334095 | 1 | 0.355413596 | 0.355413596 | 0.355413596 | 7.767699931 | 0.89029244 | FALSE | TRUE | FALSE |
| DNAL4 | 2.7596552 | 0.40705496 | 0.0036 | 0.689946146 | 2.759655219 | 2.759655219 | 0.72899184 | 0.652250095 | 1 | 0.728991842 | 0.728991842 | 0.728991842 | 3.785577643 | 0.578132158 | FALSE | TRUE | FALSE |
| MOGAT2 | 2.7538368 | 0.40725799 | 0.74745 | 2.753836751 | 26.23009763 | 26.23009763 | 0.36404068 | 0.80444633 | 1 | 0.364040675 | 0.364040675 | 0.364040675 | 7.56464027 | 0.87878828 | FALSE | TRUE | FALSE |
| DEPDC1B | 2.7520412 | 0.40746087 | 0.2564 | 0.68814995 | 2.752041225 | 6.013677941 | 0.1068322 | 0.87411344 | 0.0128 | 0.106832204 | 0.106832204 | 2.118008294 | 25.76040847 | 1.410952745 | FALSE | TRUE | FALSE |
| DBH | 2.7519961 | 0.40766358 | 0.8902 | 44.07831812 | 706.7013522 | 3472.227733 | 1.24603341 | 0.561908594 | 0.5096 | 1.246033414 | 4.985352348 | 19.95126823 | 2.208605359 | 0.344118122 | FALSE | FALSE | FALSE |
| CACNG6 | 2.7489106 | 0.40786616 | 0.00685 | 2.748910613 | 2.748910613 | 2.748910613 | 0.35378669 | 0.80779171 | 1 | 0.353786692 | 0.353786692 | 0.353786692 | 7.769966129 | 0.890419126 | FALSE | TRUE | FALSE |
| SLC4A4 | 2.7453729 | 0.40806863 | 0.075 | 0.28776488 | 10.97153241 | 13.46876662 | 0.0143591 | 0.895326061 | 0.7588 | 0.133928816 | 0.535170611 | 2.631476089 | 191.1939754 | 2.281474203 | FALSE | TRUE | FALSE |
| NUDT8 | 2.7451345 | 0.40827093 | 0.5044 | 2.745134481 | 2.745134481 | 2.745134481 | 0.35282587 | 0.808087976 | 1 | 0.35282587 | 0.35282587 | 0.35282587 | 7.780422903 | 0.891003204 | FALSE | TRUE | FALSE |
| HLA-A | 2.7447884 | 0.40847306 | 0.87845 | 26.0716293 | 254.0002847 | 254.0002847 | 0.35260609 | 0.808247113 | 1 | 3.276014493 | 31.23859101 | 31.5749996 | 7.784290963 | 0.891219061 | FALSE | TRUE | FALSE |
| SMIM8 | 2.7422489 | 0.40867506 | 0.7501 | 2.742248908 | 10.97164533 | 10.97164533 | 25.5513353 | 0.107652548 | 0.49465 | 6.387857669 | 246.8127168 | 246.8127168 | 0.107323116 | -0.969306728 | FALSE | FALSE | FALSE |
| HSPA14 | 2.7419048 | 0.40887689 | 0.5077 | 2.741904812 | 2.741904812 | 5.859414605 | 2.79185715 | 0.42784544 | 0.9707 | 258.9699954 | 2589.18739 | 26518.71617 | 0.982107846 | -0.00784082 | FALSE | FALSE | FALSE |
| CSH1 | 2.7412803 | 0.40907855 | 0.93985 | 26.49632819 | 2669.232115 | 2669.232115 | 0.35475875 | 0.80749451 | 1 | 3.350491204 | 129.8752285 | 129.8752285 | 7.727167547 | 0.888020329 | FALSE | TRUE | FALSE |
| SWI5 | 2.740523 | 0.40928005 | 0.00825 | 2.740523036 | 2.740523036 | 2.740523036 | 0.35262037 | 0.808224397 | 1 | 0.352620374 | 0.352620374 | 0.352620374 | 7.771879442 | 0.890526055 | FALSE | TRUE | FALSE |
| IQCK | 2.7357674 | 0.40948146 | 0.8773 | 25.89819173 | 251.4593814 | 251.4593814 | 0.37490517 | 0.80085709 | 1 | 0.374905174 | 0.374905174 | 0.374905174 | 7.297225064 | 0.863157741 | FALSE | TRUE | FALSE |
| ZNF585B | 2.73488 | 0.40968271 | 0.66185 | 25.55823671 | 2402.76968 | 24143.26437 | 3564.71377 | 0.000535751 | 0.23135 | 360.8045763 | 36059.40942 | 70452.17361 | 0.000767209 | -3.115086387 | FALSE | FALSE | FALSE |
| SIRT6 | 2.7339895 | 0.40988381 | 0.01145 | 2.733989455 | 2.733989455 | 5.825119975 | 3.16433515 | 0.402439232 | 0.5038 | 3.164335152 | 3.164335152 | 6.733088515 | 0.864001227 | -0.063485641 | FALSE | FALSE | FALSE |
| MAP3K3 | 2.7336839 | 0.41008473 | 0.0232 | 2.733683927 | 2.733683927 | 53.79215498 | 0.34838074 | 0.809600131 | 1 | 0.348380736 | 0.348380736 | 6.868578704 | 7.846828603 | 0.894694166 | FALSE | TRUE | FALSE |
| DNAJC24 | 2.7328892 | 0.41028549 | 0.5069 | 2.732889158 | 2.732889158 | 2.732889158 | 0.33657058 | 0.813330947 | 0.00385 | 0.33657058 | 0.33657058 | 0.33657058 | 8.119809999 | 0.909545867 | FALSE | TRUE | FALSE |
| FBXW4 | 2.7324545 | 0.41048609 | 0.5 | 1.955299347 | 10.92928052 | 10.92928052 | 3.19720509 | 0.40045166 | 0.50595 | 3.19720509 | 3.19720509 | 3.19720509 | 0.854638468 | -0.068217563 | FALSE | FALSE | FALSE |
| C14orf180 | 2.7318063 | 0.41068653 | 0.75655 | 2.731806273 | 25.76532159 | 25.76532159 | 0.36382964 | 0.804469971 | 1 | 0.363829638 | 0.363829638 | 0.363829638 | 7.508476461 | 0.875551824 | FALSE | TRUE | FALSE |
| TTC32 | 2.7315897 | 0.41088679 | 0.00465 | 2.731589745 | 2.731589745 | 2.731589745 | 0.35072956 | 0.808926054 | 1 | 0.35072956 | 0.35072956 | 0.35072956 | 7.788307743 | 0.891443104 | FALSE | TRUE | FALSE |
| NMT2 | 2.7313363 | 0.41108689 | 0.25635 | 0.682976698 | 2.731336336 | 5.964719522 | 0.10611912 | 0.874342837 | 0.01305 | 0.106119118 | 0.106119118 | 2.103063797 | 25.73840034 | 1.410581552 | FALSE | TRUE | FALSE |
| ELOVL7 | 2.7295338 | 0.41128685 | 0.75555 | 2.729533758 | 26.07880569 | 26.07880569 | 3.12455122 | 0.404851413 | 0.75195 | 12.49687491 | 119.6763236 | 119.6763236 | 0.873576255 | -0.058699179 | FALSE | FALSE | FALSE |
| MIA | 2.7287333 | 0.41148665 | 0.0045 | 2.728733257 | 2.728733257 | 2.728733257 | 3.1601374 | 0.402879168 | 0.00415 | 3.160137403 | 3.160137403 | 3.160137403 | 0.863485636 | -0.063744882 | FALSE | FALSE | FALSE |
| MOXD1 | 2.7272509 | 0.4116863 | 0.13465 | 0.305780046 | 2.727250872 | 2.727250872 | 0.27501092 | 0.832298447 | 0.5026 | 0.275010921 | 1.099590055 | 1.099590055 | 9.916882057 | 0.996375148 | FALSE | TRUE | FALSE |
| EXOC3L1 | 2.7263934 | 0.41188579 | 0.5119 | 2.726393385 | 25.14448028 | 53.51442102 | 36.7121369 | 0.075695219 | 0.31835 | 3.948210887 | 349.9201308 | 349.9201308 | 0.074264089 | -1.129221145 | FALSE | FALSE | FALSE |
| OR2C1 | 2.7165511 | 0.41208528 | 0.5029 | 2.716551138 | 2.716551138 | 2.716551138 | 3.14360752 | 0.40397615 | 0.50805 | 3.143607517 | 3.143607517 | 3.143607517 | 0.86415086 | -0.063410434 | FALSE | FALSE | FALSE |
| SGMS1 | 2.7126928 | 0.41228467 | 0.99635 | 2579.185388 | 282257.6393 | 3053792.994 | 2.94872959 | 0.414551656 | 0.99925 | 28964.62781 | 36836049.99 | 416725081.5 | 0.919953053 | -0.036234335 | FALSE | FALSE | FALSE |
| ACO1 | 2.7116081 | 0.41248391 | 0.82195 | 52.21736184 | 4103.610895 | 53677.94369 | 1.44002798 | 0.541955105 | 0.83705 | 5.76220863 | 369.7376984 | 1480.967168 | 1.883024615 | 0.274855997 | FALSE | FALSE | FALSE |
| RNF5 | 2.7114149 | 0.41268297 | 0.50505 | 2.711414929 | 2.711414929 | 2.711414929 | 0.3466352 | 0.810002295 | 0.00605 | 0.346635197 | 0.346635197 | 0.346635197 | 7.822099292 | 0.893323324 | FALSE | TRUE | FALSE |
| SERINC3 | 2.7087425 | 0.41288192 | 0.01385 | 2.708742451 | 2.708742451 | 5.797406937 | 0.34839162 | 0.809577739 | 1 | 0.348391618 | 0.348391618 | 6.911474612 | 7.774993163 | 0.890700016 | FALSE | TRUE | FALSE |
| UBE2L6 | 2.7010472 | 0.41308082 | 0.74935 | 5.807500286 | 102.9179561 | 102.9179561 | 0.36184953 | 0.805012143 | 1 | 0.361849527 | 0.361849527 | 0.361849527 | 7.464559154 | 0.873004164 | FALSE | TRUE | FALSE |
| MRPL17 | 2.6979715 | 0.41327961 | 0.50395 | 2.697971538 | 2.697971538 | 2.697971538 | 0.34596477 | 0.810336126 | 1 | 0.345964771 | 0.345964771 | 0.345964771 | 7.798399624 | 0.892005487 | FALSE | TRUE | FALSE |
| MOSPD3 | 2.6947216 | 0.41347828 | 0.5049 | 2.694721599 | 10.77805227 | 10.77805227 | 0.35008381 | 0.809083769 | 1 | 0.350083811 | 0.350083811 | 0.350083811 | 7.697361358 | 0.886341875 | FALSE | TRUE | FALSE |
| NAA11 | 2.6921397 | 0.41367683 | 0.5077 | 2.692139748 | 2.692139748 | 2.692139748 | 0.34211136 | 0.811484189 | 1 | 0.342111363 | 0.342111363 | 0.342111363 | 7.869191252 | 0.8959301 | FALSE | TRUE | FALSE |

| | | | | | | | | | | | | | | | | | |
|---|---|---|---|---|---|---|---|---|---|---|---|---|---|---|---|---|---|
| CNBP | 2.6908752 | 0.41387523 | 0.0061 | 2.690875174 | 2.690875174 | 2.690875174 | 0.71304236 | 0.659515976 | 1 | 0.713042365 | 0.713042365 | 0.713042365 | 3.773794246 | 0.576778218 | FALSE | TRUE | FALSE |
| OR2J2 | 2.6884081 | 0.4140735 | 0.87835 | 25.48863231 | 247.8593793 | 247.8593793 | 3.21507677 | 0.398897797 | 0.25675 | 3.215076774 | 30.60689655 | 30.60689655 | 0.836187837 | -0.077696154 | FALSE | FALSE | FALSE |
| VWA1 | 2.6848135 | 0.41427166 | 0.69535 | 2.684813495 | 254.3895731 | 254.3895731 | 0.47474313 | 0.765242713 | 0.0111 | 0.474743131 | 0.474743131 | 1.024405686 | 5.655297187 | 0.752455432 | FALSE | TRUE | FALSE |
| TMEM182 | 2.6839067 | 0.41446967 | 0.50165 | 2.683906651 | 2.683906651 | 2.683906651 | 0.33435665 | 0.814187193 | 0.5032 | 3.111814114 | 3.111814114 | 3.111814114 | 8.02707717 | 0.904557438 | FALSE | TRUE | FALSE |
| ZNF124 | 2.6829083 | 0.41466752 | 0.5002 | 2.682908288 | 24.61705352 | 24.61705352 | 37.1228882 | 0.07458064 | 0.1268 | 4.007222654 | 37.12288824 | 37.12288824 | 0.072271001 | -1.141035931 | FALSE | FALSE | FALSE |
| GPR85 | 2.6804537 | 0.41486525 | 0.50045 | 2.680453671 | 2.680453671 | 2.680453671 | 2.81116282 | 0.426419945 | 0.5026 | 2.811162823 | 2.811162823 | 2.811162823 | 0.953503529 | -0.020677695 | FALSE | FALSE | FALSE |
| GLT6D1 | 2.6790005 | 0.41506284 | 0.7486 | 2.679000456 | 25.34051854 | 25.34051854 | 3.00660715 | 0.411134104 | 0.877 | 11.04218556 | 277.325631 | 277.325631 | 0.891037746 | -0.050103898 | FALSE | FALSE | FALSE |
| FCRL6 | 2.6753548 | 0.41526032 | 0.68685 | 2.675354841 | 229.2490005 | 229.2490005 | 4.11777503 | 0.348717925 | 0.50245 | 4.117775032 | 38.05845065 | 38.05845065 | 0.649708841 | -0.187281224 | FALSE | FALSE | FALSE |
| PLA2G7 | 2.6752431 | 0.41545763 | 0.497 | 2.675243095 | 235.6781072 | 235.6781072 | 0.10442313 | 0.874878272 | 1 | 0.949170397 | 8.855013225 | 8.855013225 | 25.61925885 | 1.408566562 | FALSE | TRUE | FALSE |
| NAALAD2 | 2.6731015 | 0.41565481 | 0.6591 | 2.673101526 | 226.1991395 | 2156.4974 | 0.62892465 | 0.699491288 | 0.5046 | 0.628924652 | 5.664371654 | 5.664371654 | 4.250273096 | 0.628416836 | FALSE | TRUE | FALSE |
| SLC10A7 | 2.672763 | 0.41585183 | 0.2549 | 2.672763005 | 25.48253322 | 25.48253322 | 3.10090431 | 0.405506944 | 0.2593 | 3.100904307 | 29.64026453 | 29.64026453 | 0.861930179 | -0.064527913 | FALSE | FALSE | FALSE |
| MRPS23 | 2.6705457 | 0.41604872 | 0.50205 | 2.670545656 | 2.670545656 | 2.670545656 | 0.34295659 | 0.811220498 | 0.0051 | 0.342956595 | 0.342956595 | 0.342956595 | 7.786832787 | 0.891360849 | FALSE | TRUE | FALSE |
| CLEC4G | 2.6697486 | 0.41624545 | 0.50805 | 2.669748613 | 2.669748613 | 5.73730919 | 0.34464497 | 0.810801453 | 0.00805 | 0.344644967 | 0.344644967 | 0.344644967 | 7.746373421 | 0.889098429 | FALSE | TRUE | FALSE |
| GNAO1 | 2.6673978 | 0.41644206 | 0.261 | 1.271314924 | 12.31864749 | 12.31864749 | 0.16035661 | 0.861940989 | 1 | 14.6372402 | 1478.583368 | 1478.583368 | 16.63416131 | 1.221000909 | FALSE | TRUE | FALSE |
| SEMA6B | 2.6653477 | 0.41663853 | 0.51775 | 2.665347712 | 2.665347712 | 52.27379743 | 0.33122567 | 0.815014381 | 0.5081 | 3.095437191 | 3.095437191 | 60.85056719 | 8.046923855 | 0.905629892 | FALSE | TRUE | FALSE |
| TCEA3 | 2.6616755 | 0.4168349 | 0.5017 | 2.661675493 | 24.69653741 | 24.69653741 | 0.94521991 | 0.593398776 | 0.5092 | 0.94521991 | 0.94521991 | 0.94521991 | 2.815932529 | 0.449622245 | FALSE | TRUE | FALSE |
| KRAS | 2.6616092 | 0.4170311 | 0.49845 | 0.287361835 | 2.661609161 | 2.661609161 | 3.0879704 | 0.406596905 | 0.50645 | 3.087970401 | 3.087970401 | 3.087970401 | 0.861928327 | -0.064528846 | FALSE | FALSE | FALSE |
| TOB1 | 2.6601041 | 0.41722716 | 0.75055 | 2.660104111 | 10.64264226 | 10.64264226 | 0.70598269 | 0.663177082 | 1 | 2.82383961 | 2.82383961 | 2.82383961 | 3.767945246 | 0.576104583 | FALSE | TRUE | FALSE |
| RFX1 | 2.6583276 | 0.41742309 | 0.02925 | 0.664585257 | 2.658327563 | 22.44030333 | 0.21018735 | 0.849661654 | 1 | 0.210187351 | 0.210187351 | 4.119388099 | 12.64741932 | 1.102001918 | FALSE | TRUE | FALSE |
| THTPA | 2.6567126 | 0.41761888 | 0.00805 | 0.664074875 | 2.656712648 | 2.656712648 | 0.20685214 | 0.850454629 | 1 | 0.827342699 | 0.827342699 | 0.827342699 | 12.84353467 | 1.108684562 | FALSE | TRUE | FALSE |
| KRTAP10-4 | 2.6562312 | 0.4178145 | 0.9373 | 25.17673029 | 2439.031899 | 2439.031899 | 0.32250302 | 0.817943889 | 1 | 276.3634617 | 2760.348416 | 28243.76075 | 8.236298605 | 0.915732083 | FALSE | TRUE | FALSE |
| LCOR | 2.6520028 | 0.41801004 | 0.03215 | 2.652002777 | 2.652002777 | 51.84042169 | 28.739926 | 0.09547195 | 0.0209 | 2.985496833 | 28.73992599 | 100.7005615 | 0.092275908 | -1.034911671 | FALSE | FALSE | FALSE |
| NRIP3 | 2.6511667 | 0.41820542 | 0.5026 | 2.651166655 | 2.651166655 | 2.651166655 | 3.07390864 | 0.407682724 | 0.5002 | 3.073908641 | 3.073908641 | 3.073908641 | 0.862474122 | -0.064253927 | FALSE | FALSE | FALSE |
| KDELR2 | 2.6420278 | 0.4184008 | 0.96665 | 25.2722884 | 2493.438369 | 25687.12686 | 0.37243319 | 0.801663565 | 1 | 3.491325411 | 3.491325411 | 3.491325411 | 7.09396432 | 0.850889 | FALSE | TRUE | FALSE |
| SOS1 | 2.639924 | 0.41859604 | 0.1524 | 2.639924007 | 25.92218966 | 51.94083766 | 6.54214932 | 0.254167467 | 0.703 | 15.71064652 | 1005.15768 | 4022.208411 | 0.40352549 | -0.394129027 | FALSE | FALSE | FALSE |
| DNAJB1 | 2.6389559 | 0.41879114 | 0.2541 | 2.638955916 | 25.34335164 | 25.34335164 | 0.31735971 | 0.819544341 | 1 | 28.56597942 | 282.0649328 | 282.0649328 | 8.315346455 | 0.919880349 | FALSE | TRUE | FALSE |
| SERPINB13 | 2.6388861 | 0.41898607 | 0.07205 | 0.659783757 | 10.55718636 | 10.55718636 | 0.2334167 | 0.843859651 | 0.874 | 2.148657487 | 20.29997301 | 20.29997301 | 11.30547273 | 1.053288727 | FALSE | TRUE | FALSE |
| DIO1 | 2.6388389 | 0.41918083 | 0.5057 | 2.638838932 | 2.638838932 | 2.638838932 | 2.80256694 | 0.427031677 | 0.50455 | 2.802566943 | 2.802566943 | 2.802566943 | 0.941579268 | -0.026143112 | FALSE | FALSE | FALSE |
| METAP1 | 2.6387342 | 0.41937544 | 0.5108 | 2.638734222 | 2.638734222 | 2.638734222 | 0.33689052 | 0.813137169 | 0.007 | 0.336890515 | 0.336890515 | 0.336890515 | 7.832616537 | 0.893906865 | FALSE | TRUE | FALSE |
| GPS1 | 2.630694 | 0.41957001 | 0.50755 | 2.630694037 | 2.630694037 | 5.548861739 | 0.33730616 | 0.81292136 | 1 | 0.337306157 | 0.337306157 | 6.662643952 | 7.79912842 | 0.892046071 | FALSE | TRUE | FALSE |
| PPP1R16A | 2.62863 | 0.41976446 | 0.5107 | 2.628629961 | 2.628629961 | 9.505739326 | 2.86743526 | 0.42251659 | 0.51425 | 27.54604745 | 271.4101896 | 271.4101896 | 0.916718155 | -0.037764168 | FALSE | FALSE | FALSE |
| GABRG1 | 2.6278543 | 0.41995876 | 0.13595 | 1.133761746 | 10.50873418 | 10.50873418 | 1.57993303 | 0.528665195 | 0.50335 | 1.57993303 | 6.320051201 | 6.320051201 | 1.663269427 | 0.220962605 | FALSE | FALSE | FALSE |
| APBA2 | 2.6271847 | 0.4201529 | 0.51185 | 2.627184673 | 2.627184673 | 51.54989856 | 0.33723724 | 0.812964563 | 1 | 0.337237245 | 0.337237245 | 6.632247875 | 7.790315905 | 0.891555069 | FALSE | TRUE | FALSE |
| OXSR1 | 2.6263265 | 0.42034689 | 0.0139 | 2.626326497 | 2.626326497 | 5.556588564 | 0.33618203 | 0.813438424 | 1 | 0.336182031 | 0.336182031 | 6.669981379 | 7.812215577 | 0.892774219 | FALSE | TRUE | FALSE |
| SCNN1A | 2.6257835 | 0.42054073 | 0.3138 | 0.291013047 | 24.28673919 | 24.28673919 | 9.0575495 | 0.210787927 | 0.19295 | 1.017129835 | 20.68702343 | 82.7317391 | 0.289899989 | -0.537751801 | FALSE | FALSE | FALSE |
| ACAT2 | 2.6235938 | 0.42073444 | 0.31535 | 0.655905134 | 10.4968909 | 10.4968909 | 0.96409988 | 0.591540806 | 0.1294 | 0.964099878 | 3.856606489 | 3.856606489 | 2.721288432 | 0.434774576 | FALSE | TRUE | FALSE |
| PTPN1 | 2.6183579 | 0.42092808 | 0.50805 | 2.618357937 | 2.618357937 | 5.539752396 | 0.33523251 | 0.813909805 | 0.0097 | 0.335232506 | 0.335232506 | 0.335232506 | 7.81057293 | 0.892682892 | FALSE | TRUE | FALSE |
| CLDN15 | 2.6172955 | 0.42112157 | 0.25075 | 0.654465393 | 2.617295485 | 2.617295485 | 0.20742461 | 0.850316089 | 1 | 0.207424606 | 0.207424606 | 0.207424606 | 12.61805695 | 1.100992483 | FALSE | TRUE | FALSE |
| TMEM161B | 2.6127191 | 0.42131498 | 0.509 | 2.612719081 | 2.612719081 | 5.607772103 | 3.00375383 | 0.411348796 | 0.26245 | 3.003753825 | 12.01486348 | 12.01486348 | 0.869817979 | -0.06057162 | FALSE | FALSE | FALSE |
| OR5D18 | 2.6096347 | 0.42150828 | 0.5031 | 2.609634717 | 2.609634717 | 2.609634717 | 0.3378924 | 0.812683369 | 1 | 0.337892402 | 0.337892402 | 0.337892402 | 7.723271381 | 0.887801295 | FALSE | TRUE | FALSE |
| FCRL1 | 2.6084318 | 0.42170144 | 0.99905 | 24411.3286 | 31297012.28 | 349615652.5 | 0.41961735 | 0.784719138 | 1 | 3.914457032 | 37.47811206 | 37.47811206 | 6.216215471 | 0.79352606 | FALSE | TRUE | FALSE |
| PHLDA3 | 2.60581 | 0.42189447 | 0.50085 | 2.605809971 | 2.605809971 | 2.605809971 | 0.69306869 | 0.668835234 | 1 | 0.693068687 | 0.693068687 | 0.693068687 | 3.759814894 | 0.575166464 | FALSE | TRUE | FALSE |
| POLR2H | 2.604614 | 0.42208737 | 0.4998 | 0.651196916 | 2.604613985 | 2.604613985 | 2.74040223 | 0.431881479 | 0.00325 | 0.685140854 | 2.740402231 | 2.740402231 | 0.95044952 | -0.022070944 | FALSE | FALSE | FALSE |
| AGPAT2 | 2.6029178 | 0.42228013 | 0.13285 | 1.102226386 | 10.40426008 | 10.40426008 | 0.42675167 | 0.78224019 | 0.503 | 0.738270979 | 6.821108291 | 6.821108291 | 6.099373342 | 0.785285217 | FALSE | TRUE | FALSE |
| BCCIP | 2.600665 | 0.42247277 | 0.00865 | 2.600665045 | 2.600665045 | 2.600665045 | 0.33675698 | 0.813244875 | 1 | 0.336756981 | 0.336756981 | 0.336756981 | 7.72267597 | 0.887767813 | FALSE | TRUE | FALSE |
| MAD2L1BP | 2.5992344 | 0.42266527 | 0.50045 | 2.599234433 | 10.39684835 | 10.39684835 | 0.33997069 | 0.812118394 | 1 | 0.339970694 | 0.339970694 | 0.339970694 | 7.645466145 | 0.88340397 | FALSE | TRUE | FALSE |
| TIRAP | 2.5879598 | 0.4228578 | 0.50565 | 2.587959753 | 2.587959753 | 2.587959753 | 2.825021 | 0.425192459 | 0.87375 | 26.72233812 | 259.2591852 | 259.2591852 | 0.916085137 | -0.038064163 | FALSE | FALSE | FALSE |
| MPST | 2.5844908 | 0.42305023 | 0.7524 | 2.58449081 | 24.59434803 | 24.59434803 | 11.9035213 | 0.178931615 | 0.50375 | 11.90352129 | 113.590085 | 113.590085 | 0.217119854 | -0.663300461 | FALSE | FALSE | FALSE |
| GRB14 | 2.5814276 | 0.42324255 | 0.5935 | 10.3240724 | 236.0892665 | 944.2072475 | 0.10756062 | 0.873883243 | 1 | 0.107560622 | 0.107560622 | 2.132977068 | 23.99974614 | 1.380206648 | FALSE | TRUE | FALSE |
| THOC1 | 2.5809899 | 0.42343471 | 0.5053 | 2.580989926 | 2.580989926 | 5.460093595 | 0.33136998 | 0.814929876 | 1 | 0.331369979 | 0.331369979 | 6.566616144 | 7.788846576 | 0.891473149 | FALSE | TRUE | FALSE |
| ENPP4 | 2.5761033 | 0.4236268 | 0.01095 | 0.644035242 | 2.576103324 | 3.196665391 | 0.20421614 | 0.851047607 | 1 | 0.20421614 | 0.20421614 | 0.20421614 | 12.61514928 | 1.100873215 | FALSE | TRUE | FALSE |
| ARHGAP21 | 2.5757161 | 0.42381874 | 0.27505 | 0.27720994 | 2.575716126 | 49.60507534 | 0.17646194 | 0.857976542 | 0.9411 | 3.745531346 | 143.4121966 | 289.7791179 | 14.59644008 | 1.164246949 | FALSE | TRUE | FALSE |
| CBX7 | 2.5738674 | 0.42401054 | 0.0083 | 2.573867423 | 2.573867423 | 2.573867423 | 0.68574519 | 0.6726365 | 1 | 0.685745191 | 0.685745191 | 0.685745191 | 3.753387494 | 0.574423403 | FALSE | TRUE | FALSE |
| KCNK15 | 2.572169 | 0.42420221 | 0.8187 | 23.89779447 | 556.2791653 | 2224.590123 | 0.42329836 | 0.783355652 | 0.7484 | 0.965972735 | 3.863084399 | 3.863084399 | 6.076491701 | 0.783652909 | FALSE | TRUE | FALSE |
| OTUD4 | 2.5687997 | 0.42439378 | 0.50585 | 2.568799713 | 23.77417819 | 50.50978984 | 1.86852591 | 0.501449782 | 0.2667 | 1.868525912 | 7.473426646 | 36.81502186 | 1.374773396 | 0.138231119 | FALSE | FALSE | FALSE |
| OR11H4 | 2.5681593 | 0.4245852 | 0.75125 | 2.568159312 | 24.29595368 | 24.29595368 | 0.34389229 | 0.810933981 | 1 | 0.343892292 | 0.343892292 | 0.343892292 | 7.467917629 | 0.873199519 | FALSE | TRUE | FALSE |
| C10orf53 | 2.565194 | 0.42477651 | 0.69305 | 24.19827918 | 2322.042148 | 2322.042148 | 0.36598244 | 0.803758211 | 0.0034 | 0.365982438 | 0.365982438 | 0.365982438 | 7.009063178 | 0.845659975 | FALSE | TRUE | FALSE |
| AURKB | 2.5649972 | 0.42496766 | 0.5018 | 24.44907485 | 239.0249388 | 239.0249388 | 0.34178235 | 0.811528068 | 0.5128 | 3.188191071 | 3.188191071 | 3.188191071 | 7.504767849 | 0.875337262 | FALSE | TRUE | FALSE |
| RASD1 | 2.5649787 | 0.42515865 | 0.0085 | 0.51775912 | 2.070762415 | 2.564978729 | 2.63563323 | 0.43783808 | 0.2575 | 0.658965811 | 2.635633228 | 2.635633228 | 0.973192591 | -0.011801206 | FALSE | FALSE | FALSE |
| DALRD3 | 2.5646836 | 0.42534949 | 0.0076 | 0.283904713 | 2.564683591 | 2.564683591 | 0.04922798 | 0.88685708 | 1 | 0.04922798 | 0.437377098 | 0.437377098 | 52.09808707 | 1.716821777 | FALSE | TRUE | FALSE |
| KCNIP2 | 2.5569187 | 0.4255403 | 0.50325 | 2.556918674 | 2.556918674 | 2.556918674 | 0.33064674 | 0.81520423 | 1 | 0.330646738 | 0.330646738 | 0.330646738 | 7.733083012 | 0.888352672 | FALSE | TRUE | FALSE |
| HEBP2 | 2.554631 | 0.42573099 | 0.7546 | 2.55463099 | 24.09411433 | 24.09411433 | 0.34288896 | 0.811242501 | 0.0037 | 0.342888961 | 0.342888961 | 0.342888961 | 7.450315643 | 0.872174673 | FALSE | TRUE | FALSE |
| SPINT4 | 2.551829 | 0.42592157 | 0.50285 | 2.551828982 | 2.551828982 | 2.551828982 | 0.33257037 | 0.814654694 | 1 | 0.332570372 | 0.332570372 | 0.332570372 | 7.673049667 | 0.884968009 | FALSE | TRUE | FALSE |
| RAB11FIP5 | 2.5505218 | 0.42611201 | 0.8793 | 24.36417562 | 238.7144568 | 479.3781894 | 3.07365351 | 0.407899402 | 0.75215 | 3.073653511 | 29.45681991 | 60.5938828 | 0.829801331 | -0.081025873 | FALSE | FALSE | FALSE |
| COG8 | 2.5474634 | 0.42630235 | 0.019 | 0.636707193 | 2.547463367 | 12.52918077 | 0.90797603 | 0.597391965 | 0.5064 | 0.907976033 | 8.18119009 | 10.08324676 | 2.805650452 | 0.448033562 | FALSE | TRUE | FALSE |
| PYGL | 2.5433922 | 0.42649259 | 0.9991 | 218497055.5 | 3.07509E+12 | 1.98821E+14 | 1969.10884 | 0.001204136 | 0.53585 | 4437.669792 | 8173121.953 | 296014107.8 | 0.001291646 | -2.88885639 | FALSE | FALSE | FALSE |
| WSCD2 | 2.5428381 | 0.42668269 | 0.50515 | 2.542838118 | 2.542838118 | 50.07930293 | 0.32791472 | 0.816001536 | 0.01955 | 0.327914724 | 0.327914724 | 6.469919422 | 7.754571343 | 0.889557796 | FALSE | TRUE | FALSE |
| RHOG | 2.5417579 | 0.42687265 | 0.87885 | 21.57852288 | 233.3735139 | 233.3735139 | 0.72250164 | 0.654993417 | 1 | 0.722501637 | 0.722501637 | 0.722501637 | 3.51799603 | 0.546295345 | FALSE | TRUE | FALSE |
| PELI3 | 2.5402248 | 0.42706247 | 0.50685 | 2.540224758 | 2.540224758 | 5.36943468 | 11.1009966 | 0.186736236 | 0.0001 | 0.327537172 | 0.327537172 | 6.467742652 | 0.228828532 | -0.640489826 | FALSE | FALSE | FALSE |
| TMEM176B | 2.5352293 | 0.42725222 | 0.50565 | 2.535229283 | 2.535229283 | 2.535229283 | 0.33015294 | 0.81535162 | 0.0075 | 0.330152939 | 0.330152939 | 0.330152939 | 7.678954151 | 0.885302075 | FALSE | TRUE | FALSE |
| CDC42EP4 | 2.5346841 | 0.42744182 | 0.49975 | 1.077374849 | 10.13639633 | 21.31752717 | 0.32839539 | 0.815834261 | 1 | 0.328395392 | 0.328395392 | 6.506153191 | 7.718391268 | 0.88752679 | FALSE | TRUE | FALSE |
| LRRC32 | 2.5322678 | 0.42763131 | 0.51015 | 2.532267781 | 10.13154617 | 49.84785738 | 0.78000391 | 0.628507347 | 0.75385 | 1.82156643 | 7.284677427 | 15.38353196 | 3.246480882 | 0.51141285 | FALSE | TRUE | FALSE |
| ASB1 | 2.5321181 | 0.42782064 | 0.75535 | 2.532118097 | 24.17003914 | 24.17003914 | 0.33756542 | 0.812813261 | 1 | 0.33756542 | 0.33756542 | 0.33756542 | 7.501118154 | 0.875126006 | FALSE | TRUE | FALSE |

| | | | | | | | | | | | | | | | | | |
|---|---|---|---|---|---|---|---|---|---|---|---|---|---|---|---|---|---|
| AP1AR | 2.5281794 | 0.42800988 | 0.74825 | 2.52817938 | 23.87705053 | 23.87705053 | 0.33949362 | 0.812227309 | 1 | 0.339493621 | 0.339493621 | 0.339493621 | 7.446912773 | 0.871976267 | FALSE | TRUE | FALSE |
| FOXL1 | 2.5259284 | 0.428199 | 0.26015 | 0.631568086 | 2.52592836 | 3.130262752 | 0.20092583 | 0.85188101 | 0.0099 | 0.200925834 | 0.200925834 | 0.200925834 | 12.57144646 | 1.09938525 | FALSE | TRUE | FALSE |
| DNAJC12 | 2.5247636 | 0.42838798 | 0.75035 | 2.524763621 | 23.88063116 | 23.88063116 | 2.85347996 | 0.423548106 | 0.50745 | 27.0591177 | 263.1833726 | 263.1833726 | 0.884801594 | -0.053154104 | FALSE | FALSE | FALSE |
| PTGDR | 2.5243704 | 0.42857682 | 0.5071 | 2.524370368 | 2.524370368 | 5.392724662 | 11.6587789 | 0.17995172 | 0.0086 | 2.914820095 | 11.65877885 | 11.65877885 | 0.216520992 | -0.664499991 | FALSE | FALSE | FALSE |
| MAS1 | 2.5209453 | 0.42876555 | 0.5054 | 2.520945255 | 2.520945255 | 2.520945255 | 0.31640522 | 0.819845014 | 1 | 2.944948404 | 2.944948404 | 2.944948404 | 7.967457862 | 0.901319775 | FALSE | TRUE | FALSE |
| OCEL1 | 2.5208025 | 0.42895413 | 0.5014 | 2.520802451 | 2.520802451 | 2.520802451 | 0.31832457 | 0.819262778 | 0.50735 | 2.941040568 | 2.941040568 | 2.941040568 | 7.918969077 | 0.898668647 | FALSE | TRUE | FALSE |
| HEMK1 | 2.5206105 | 0.42914256 | 0.75045 | 2.520610463 | 23.88105263 | 23.88105263 | 3.00357244 | 0.411563287 | 0.5068 | 3.00357244 | 12.01400118 | 12.01400118 | 0.839204152 | -0.076132376 | FALSE | FALSE | FALSE |
| RSBN1L | 2.5198866 | 0.42933084 | 0.0224 | 2.519886576 | 2.519886576 | 49.55554323 | 0.31516395 | 0.820204511 | 1 | 2.946250078 | 2.946250078 | 58.05687716 | 7.995478587 | 0.902844465 | FALSE | TRUE | FALSE |
| C6orf58 | 2.5176311 | 0.42951901 | 0.499 | 0.273513342 | 2.517631144 | 2.517631144 | 0.327906 | 0.816022423 | 1 | 0.327905997 | 0.327905997 | 0.327905997 | 7.677905152 | 0.885242743 | FALSE | TRUE | FALSE |
| VRK1 | 2.5176037 | 0.42970702 | 0.8819 | 23.87050267 | 232.136391 | 232.136391 | 0.34803884 | 0.809644901 | 0.00895 | 0.348038838 | 0.348038838 | 0.348038838 | 7.233685969 | 0.859359651 | FALSE | TRUE | FALSE |
| ILF2 | 2.5164086 | 0.42989489 | 0.5149 | 2.516408614 | 2.516408614 | 2.516408614 | 0.67202828 | 0.678971021 | 1 | 0.672028276 | 0.672028276 | 0.672028276 | 3.744498117 | 0.573393617 | FALSE | TRUE | FALSE |
| NCBP2 | 2.514631 | 0.43008264 | 0.50735 | 2.514631045 | 2.514631045 | 2.514631045 | 0.32588693 | 0.816833233 | 1 | 0.325886931 | 0.325886931 | 0.325886931 | 7.71626845 | 0.887407328 | FALSE | TRUE | FALSE |
| TMX1 | 2.5094602 | 0.43027032 | 0.50235 | 2.509460228 | 2.509460228 | 2.509460228 | 1.24762434 | 0.561659814 | 0.75195 | 2.899829646 | 11.59833462 | 11.59833462 | 2.011390895 | 0.30349648 | FALSE | FALSE | FALSE |
| OTX1 | 2.507179 | 0.43045788 | 0.93755 | 94.44720324 | 3652.466926 | 31015.04463 | 0.75658544 | 0.638663859 | 0.9964 | 275.456041 | 27749.71722 | 111013.556 | 3.313808123 | 0.520327358 | FALSE | TRUE | FALSE |
| OGFRL1 | 2.5056727 | 0.43064532 | 0.5037 | 2.505672666 | 2.505672666 | 5.392770579 | 0.32617233 | 0.816667516 | 1 | 0.32617233 | 0.32617233 | 0.32617233 | 7.682051598 | 0.88547722 | FALSE | TRUE | FALSE |
| ADORA2A | 2.5050727 | 0.43083261 | 0.51035 | 2.505072718 | 2.505072718 | 5.302084461 | 1.27718619 | 0.557388761 | 0.0125 | 1.277186192 | 1.277186192 | 6.326562475 | 1.961399782 | 0.292566123 | FALSE | FALSE | FALSE |
| OR5K1 | 2.5026065 | 0.43101979 | 0.5058 | 2.502606457 | 2.502606457 | 2.502606457 | 0.3251645 | 0.817060587 | 1 | 0.325164499 | 0.325164499 | 0.325164499 | 7.696431996 | 0.886289436 | FALSE | TRUE | FALSE |
| GCG | 2.5005563 | 0.43120684 | 0.5093 | 2.500556259 | 2.500556259 | 2.500556259 | 0.32548376 | 0.816998638 | 1 | 0.325483763 | 0.325483763 | 0.325483763 | 7.682583711 | 0.885507301 | FALSE | TRUE | FALSE |
| RPL23A | 2.4984654 | 0.43139378 | 0.5054 | 2.49846538 | 2.49846538 | 2.49846538 | 0.65965241 | 0.685943294 | 0.0044 | 0.659652409 | 0.659652409 | 0.659652409 | 3.787548332 | 0.578358183 | FALSE | TRUE | FALSE |
| ABHD17B | 2.4980912 | 0.43158057 | 0.00845 | 2.498091171 | 2.498091171 | 2.498091171 | 0.66784495 | 0.681550401 | 1 | 0.667844948 | 0.667844948 | 0.667844948 | 3.74052567 | 0.57293264 | FALSE | TRUE | FALSE |
| AP2A2 | 2.4978035 | 0.43176721 | 0.5134 | 2.497803516 | 2.497803516 | 48.89835035 | 2.92381096 | 0.41709075 | 0.02465 | 2.923810961 | 2.923810961 | 57.38407692 | 0.854297199 | -0.068391017 | FALSE | FALSE | FALSE |
| CISH | 2.4962923 | 0.43195372 | 0.2574 | 0.624113621 | 2.496292258 | 2.496292258 | 0.19644231 | 0.852987303 | 0.5014 | 0.785553865 | 0.785553865 | 0.785553865 | 12.70750844 | 1.104060407 | FALSE | TRUE | FALSE |
| SRSF7 | 2.4948957 | 0.43214011 | 0.50475 | 2.494895734 | 2.494895734 | 12.39424564 | 0.66711981 | 0.682040711 | 1 | 0.667119812 | 0.667119812 | 0.667119812 | 3.739801591 | 0.572848562 | FALSE | TRUE | FALSE |
| WRNIP1 | 2.4943902 | 0.43232634 | 1 | 2.70851E+15 | 1.15352E+19 | 3.28716E+20 | 0.86494093 | 0.604441274 | 1 | 6.09933E+16 | 6.0237E+20 | 1.71727E+22 | 2.883884978 | 0.459977935 | FALSE | TRUE | FALSE |
| KLRC2 | 2.488112 | 0.43251253 | 0.50755 | 2.488112007 | 2.488112007 | 2.488112007 | 0.324882 | 0.81720497 | 1 | 0.324882002 | 0.324882002 | 0.324882002 | 7.658509826 | 0.884144274 | FALSE | TRUE | FALSE |
| CPNE9 | 2.4878037 | 0.43269858 | 0.50755 | 2.487803652 | 2.487803652 | 9.096897595 | 1.27842938 | 0.557261881 | 0.5143 | 1.278429384 | 1.278429384 | 6.314622244 | 1.945984412 | 0.289139357 | FALSE | FALSE | FALSE |
| MAP2K3 | 2.4850449 | 0.43288451 | 0.9926 | 376.4066656 | 14628.98294 | 36393.37891 | 21.8855445 | 0.124320829 | 0.9805 | 21336.89827 | 2369010.24 | 25845408.95 | 0.113547317 | -0.944823122 | FALSE | FALSE | FALSE |
| MRAP | 2.4846215 | 0.4330703 | 0.50255 | 2.484621491 | 2.484621491 | 2.484621491 | 0.32248459 | 0.817964327 | 1 | 0.322484586 | 0.322484586 | 0.322484586 | 7.704620917 | 0.886751275 | FALSE | TRUE | FALSE |
| ZNF570 | 2.4837048 | 0.43325595 | 0.2529 | 1.095211503 | 9.932599442 | 9.932599442 | 0.04958004 | 0.886750103 | 0.01135 | 0.049580039 | 0.049580039 | 0.984255989 | 50.09485289 | 1.699793106 | FALSE | TRUE | FALSE |
| GNMT | 2.4788199 | 0.43344154 | 0.49985 | 0.619730752 | 2.478819884 | 2.478819884 | 0.66722064 | 0.682008663 | 1 | 0.667220635 | 0.667220635 | 0.667220635 | 3.715142718 | 0.569975502 | FALSE | TRUE | FALSE |
| ANKRD37 | 2.478156 | 0.43362698 | 0.5102 | 2.478155997 | 2.478155997 | 2.478155997 | 0.32349911 | 0.817513578 | 1 | 0.323499105 | 0.323499105 | 0.323499105 | 7.66047249 | 0.884255557 | FALSE | TRUE | FALSE |
| BCL7C | 2.47181 | 0.43381237 | 0.9796 | 2241.902682 | 2526999.238 | 29404015.87 | 28.2832021 | 0.098129313 | 0.96325 | 2742.523555 | 292879.101 | 3136710.771 | 0.087394985 | -1.05851349 | FALSE | FALSE | FALSE |
| HBA2 | 2.4714823 | 0.43399762 | 0.74895 | 2.471482273 | 9.887560488 | 9.887560488 | 0.66161994 | 0.685113509 | 1 | 0.66161994 | 2.646291306 | 2.646291306 | 3.735501494 | 0.572348914 | FALSE | TRUE | FALSE |
| LONRF1 | 2.469591 | 0.43418275 | 0.5049 | 2.469590997 | 2.469590997 | 5.252838445 | 0.31968119 | 0.818838791 | 1 | 0.319681195 | 0.319681195 | 6.343160814 | 7.725168194 | 0.887907944 | FALSE | TRUE | FALSE |
| FAM193A | 2.4693388 | 0.43436773 | 0.5175 | 2.469338832 | 2.469338832 | 48.14117078 | 0.30884817 | 0.822350574 | 1 | 2.894827619 | 2.894827619 | 56.60785185 | 7.99531642 | 0.902835656 | FALSE | TRUE | FALSE |
| CYTH2 | 2.466748 | 0.43455261 | 0.5055 | 2.46674795 | 2.46674795 | 5.220419094 | 0.31849734 | 0.819182171 | 0.01235 | 0.318497344 | 0.318497344 | 6.311116038 | 7.74495611 | 0.889018961 | FALSE | TRUE | FALSE |
| PROP1 | 2.4661806 | 0.43473734 | 0.81825 | 39.4737589 | 1484.560172 | 1484.560172 | 69.9212788 | 0.041046996 | 0.2896 | 17.48685536 | 279.6504793 | 1118.742938 | 0.035270817 | -1.452584485 | FALSE | FALSE | FALSE |
| TAS2R50 | 2.4661703 | 0.43492192 | 0.50765 | 2.466170278 | 2.466170278 | 2.466170278 | 0.30303235 | 0.824369074 | 1 | 2.793072742 | 26.42214475 | 26.42214475 | 8.138307021 | 0.91053407 | FALSE | TRUE | FALSE |
| TGOLN2 | 2.4554066 | 0.43510653 | 0.50625 | 2.455406591 | 2.455406591 | 5.256857017 | 0.31986486 | 0.818818551 | 1 | 0.319864855 | 0.319864855 | 6.3384342 | 7.676387547 | 0.885156892 | FALSE | TRUE | FALSE |
| USP40 | 2.455091 | 0.435291 | 0.0812 | 0.281673082 | 21.93635882 | 21.93635882 | 4.6056401 | 0.32369654 | 0.2024 | 4.605640097 | 41.7396841 | 387.5384704 | 0.533061833 | -0.273222412 | FALSE | FALSE | FALSE |
| UBE2V2 | 2.4545384 | 0.43547532 | 0.0034 | 0.61367326 | 2.454538398 | 2.454538398 | 5.99729257 | 0.269787635 | 0.50205 | 5.997292566 | 5.997292566 | 5.997292566 | 0.409274414 | -0.387985405 | FALSE | FALSE | FALSE |
| ALDH8A1 | 2.4523601 | 0.43565953 | 0.34975 | 0.613034226 | 9.812783732 | 39.27431987 | 1.08791734 | 0.580082878 | 0.13575 | 0.271998375 | 1.087917338 | 1.347423619 | 2.254178673 | 0.352988337 | FALSE | FALSE | FALSE |
| ANP32B | 2.4509995 | 0.43584361 | 0.5028 | 2.450999474 | 2.450999474 | 2.450999474 | 0.6567369 | 0.686831811 | 1 | 0.656736896 | 0.656736896 | 0.656736896 | 3.73208737 | 0.571951802 | FALSE | TRUE | FALSE |
| HIGD1A | 2.4505029 | 0.43602755 | 0.4975 | 0.612628314 | 2.450502854 | 2.450502854 | 0.0254802 | 0.89230659 | 0.0036 | 0.025480202 | 0.025480202 | 0.025480202 | 96.17282028 | 1.983052352 | FALSE | TRUE | FALSE |
| DDX5 | 2.4490768 | 0.43621136 | 0.5106 | 2.44907677 | 2.44907677 | 8.91890918 | 0.31797701 | 0.819323185 | 1 | 0.317977007 | 0.317977007 | 6.293968724 | 7.702056178 | 0.886606682 | FALSE | TRUE | FALSE |
| PLD4 | 2.4481704 | 0.43639504 | 0.68945 | 2.448170408 | 212.5515055 | 212.5515055 | 3.58865664 | 0.378745684 | 0.81395 | 33.30573344 | 316.8521846 | 3088.065806 | 0.682196892 | -0.166090263 | FALSE | FALSE | FALSE |
| CD63 | 2.4472788 | 0.43657859 | 0.8733 | 23.42986667 | 230.0697742 | 230.0697742 | 0.33723788 | 0.812942964 | 1 | 0.337237875 | 0.337237875 | 0.337237875 | 7.256832437 | 0.860747095 | FALSE | TRUE | FALSE |
| PTPN3 | 2.4454159 | 0.43676201 | 0.00175 | 0.018129601 | 0.072471415 | 0.611750367 | 0.67266761 | 0.67850329 | 0.14225 | 0.291536336 | 2.688549519 | 5.739809269 | 3.635400173 | 0.560552224 | FALSE | TRUE | FALSE |
| MFF | 2.443317 | 0.43694532 | 0.5089 | 2.443317004 | 2.443317004 | 5.264123026 | 0.31923061 | 0.81900054 | 1 | 0.31923061 | 0.31923061 | 0.31923061 | 7.653767927 | 0.88387529 | FALSE | TRUE | FALSE |
| SERPINA6 | 2.4395245 | 0.43712855 | 0.5029 | 2.439524476 | 23.17058587 | 23.17058587 | 0.38637042 | 0.797054471 | 0.50415 | 0.386370421 | 0.386370421 | 0.386370421 | 6.313952468 | 0.800301308 | FALSE | TRUE | FALSE |
| LYPD5 | 2.4363125 | 0.43731168 | 0.50235 | 2.436312491 | 2.436312491 | 2.436312491 | 0.31721616 | 0.819624621 | 1 | 0.317216156 | 0.317216156 | 0.317216156 | 7.680291333 | 0.885377694 | FALSE | TRUE | FALSE |
| OR8A1 | 2.4334515 | 0.43749471 | 0.49995 | 0.26022112 | 2.43345147 | 2.43345147 | 0.3146598 | 0.820363824 | 1 | 0.314659796 | 0.314659796 | 0.314659796 | 7.733595139 | 0.888381433 | FALSE | TRUE | FALSE |
| GDE1 | 2.4326589 | 0.4376776 | 0.7501 | 2.432658856 | 22.9836712 | 22.9836712 | 2.94197832 | 0.41561254 | 0.50605 | 2.94197832 | 2.94197832 | 2.94197832 | 0.82687858 | -0.082558258 | FALSE | FALSE | FALSE |
| ABHD15 | 2.4320424 | 0.43786035 | 0.13355 | 0.604919381 | 22.51768223 | 22.51768223 | 0.43045223 | 0.780963635 | 0.01495 | 0.430452229 | 0.430452229 | 0.957935197 | 5.649970416 | 0.752046174 | FALSE | TRUE | FALSE |
| PODXL | 2.4308436 | 0.43804298 | 0.50045 | 2.430843641 | 2.430843641 | 5.209157487 | 0.31739705 | 0.819504173 | 1 | 0.317397047 | 0.317397047 | 6.286288177 | 7.658683852 | 0.884154142 | FALSE | TRUE | FALSE |
| CRHR1 | 2.4293574 | 0.43822548 | 0.50125 | 2.429357399 | 2.429357399 | 2.429357399 | 0.64426264 | 0.69284532 | 1 | 2.576978665 | 2.576978665 | 2.576978665 | 3.77075631 | 0.576428466 | FALSE | TRUE | FALSE |
| AP1G2 | 2.4290228 | 0.43840784 | 0.8605 | 21.62483921 | 1842.0483 | 17605.42351 | 4.92785184 | 0.309956522 | 0.66265 | 19.7105733 | 1655.875018 | 3932.455883 | 0.49291717 | -0.307226054 | FALSE | FALSE | FALSE |
| GSC2 | 2.4288667 | 0.43859006 | 0.50455 | 2.428866687 | 2.428866687 | 2.428866687 | 104.843259 | 0.028138085 | 0.0024 | 2.710455456 | 104.8432585 | 104.8432585 | 0.023166646 | -1.635136832 | FALSE | FALSE | FALSE |
| TECPR1 | 2.4287109 | 0.43877213 | 0.9525 | 879.8668091 | 225437.2972 | 2225724.106 | 2.14418916 | 0.476732733 | 0.74695 | 8.576129019 | 387.9225161 | 3013.928932 | 1.132694305 | 0.054112717 | FALSE | FALSE | FALSE |
| VSTM2L | 2.4255766 | 0.4389541 | 0.99955 | 251251.9403 | 30003455.64 | 338952285.7 | 0.24478254 | 0.840650595 | 1 | 300101944.1 | 4.8478E+11 | 7.38975E+13 | 9.90910806 | 0.996034564 | FALSE | TRUE | FALSE |
| TFPI | 2.4250156 | 0.43913593 | 0.50265 | 2.425015572 | 2.425015572 | 2.425015572 | 0.31621897 | 0.819865024 | 1 | 0.316218969 | 0.316218969 | 0.316218969 | 7.668785895 | 0.884726613 | FALSE | TRUE | FALSE |
| AGTR1 | 2.4249674 | 0.43931762 | 0.3189 | 2.42496744 | 38.83719502 | 38.83719502 | 1.76022635 | 0.511947995 | 0.50115 | 1.760226354 | 1.760226354 | 1.760226354 | 1.37764523 | 0.139137393 | FALSE | FALSE | FALSE |
| EPDR1 | 2.4199503 | 0.43949925 | 0.4999 | 0.605008345 | 2.419950295 | 2.419950295 | 0.64925032 | 0.690148333 | 1 | 0.649250324 | 0.649250324 | 0.649250324 | 3.727299326 | 0.571394271 | FALSE | TRUE | FALSE |
| COMMD9 | 2.4193449 | 0.43968074 | 0.49985 | 0.263011845 | 2.419344874 | 2.419344874 | 0.31675715 | 0.81978496 | 1 | 0.316757147 | 0.316757147 | 0.316757147 | 7.637854111 | 0.882971359 | FALSE | TRUE | FALSE |
| SULT1A1 | 2.418182 | 0.4398621 | 0.63175 | 2.418182036 | 89.29713532 | 89.29713532 | 0.09979439 | 0.875955845 | 1 | 0.099794389 | 0.099794389 | 0.099794389 | 24.23164323 | 1.384382866 | FALSE | TRUE | FALSE |
| SDC4 | 2.4173144 | 0.44004332 | 0.2516 | 2.417314395 | 9.671540008 | 9.671540008 | 0.63411669 | 0.697858553 | 1 | 10.14807661 | 40.61195314 | 40.61195314 | 3.812097076 | 0.581163952 | FALSE | TRUE | FALSE |
| SPAG11A | 2.416812 | 0.44022441 | 0.49945 | 0.256188092 | 2.416812022 | 2.416812022 | 0.27575558 | 0.832142324 | 1 | 0.275755582 | 0.275755582 | 0.275755582 | 8.764326738 | 0.94271856 | FALSE | TRUE | FALSE |
| OR5K2 | 2.4129168 | 0.44040542 | 0.5067 | 2.412916839 | 2.412916839 | 2.412916839 | 0.30661983 | 0.82327916 | 1 | 0.306619825 | 2.828873257 | 2.828873257 | 7.869409086 | 0.895942122 | FALSE | TRUE | FALSE |
| RABL2B | 2.4097046 | 0.44058634 | 0.75265 | 2.409704551 | 22.9562334 | 22.9562334 | 2.92183218 | 0.417511735 | 0.0059 | 2.921832184 | 2.921832184 | 2.921832184 | 0.824723803 | -0.083691471 | FALSE | FALSE | FALSE |
| XYLT1 | 2.408489 | 0.44076713 | 0.02815 | 2.40848895 | 2.40848895 | 47.13930354 | 0.31248373 | 0.821195459 | 1 | 0.312483729 | 0.312483729 | 6.13173777 | 7.707565943 | 0.886917249 | FALSE | TRUE | FALSE |
| RWDD3 | 2.4081333 | 0.44094778 | 0.0057 | 2.40813326 | 2.40813326 | 2.40813326 | 0.31527556 | 0.820184577 | 0.0068 | 0.315275564 | 0.315275564 | 0.315275564 | 7.638185567 | 0.882990205 | FALSE | TRUE | FALSE |
| ERN1 | 2.4072762 | 0.44112829 | 0.8762 | 23.13123317 | 227.9677815 | 452.7028928 | 0.33171943 | 0.814781798 | 1 | 0.331719434 | 0.331719434 | 6.508874394 | 7.256964685 | 0.86075501 | FALSE | TRUE | FALSE |
| C6orf89 | 2.4069939 | 0.44130867 | 0.00975 | 2.406993926 | 2.406993926 | 2.406993926 | 24.2594063 | 0.113354425 | 0.68665 | 24.25940627 | 2335.018224 | 2335.018224 | 0.099218996 | -1.003405173 | FALSE | FALSE | FALSE |

| | | | | | | | | | | | | | | | | | |
|---|---|---|---|---|---|---|---|---|---|---|---|---|---|---|---|---|---|
| PNKP | 2.4055375 | 0.44148893 | 0.12995 | 0.275108575 | 2.405537468 | 2.405537468 | 0.0136876 | 0.895561233 | 0.87745 | 1.042718805 | 9.45744544 | 9.45744544 | 175.7457666 | 2.244884872 | FALSE | TRUE | FALSE |
| ACER1 | 2.4044348 | 0.44166905 | 0.5059 | 2.404434803 | 2.404434803 | 2.404434803 | 0.29513065 | 0.826675411 | 1 | 2.734397791 | 26.00157897 | 26.00157897 | 8.147018319 | 0.910998693 | FALSE | TRUE | FALSE |
| CLDN1 | 2.4028073 | 0.44184906 | 0.0066 | 0.120726938 | 0.120726938 | 0.120726938 | 0.15332735 | 0.863712049 | 0.49935 | 0.153327352 | 0.613015726 | 0.613015726 | 15.67109383 | 1.195099311 | FALSE | TRUE | FALSE |
| UBFD1 | 2.3982733 | 0.44202901 | 0.00855 | 2.398273282 | 2.398273282 | 2.398273282 | 0.64424325 | 0.692906032 | 1 | 0.644243254 | 0.644243254 | 0.644243254 | 3.722620712 | 0.570848789 | FALSE | TRUE | FALSE |
| GTDC1 | 2.3947093 | 0.44220886 | 0.3241 | 2.394709289 | 215.7917938 | 215.7917938 | 31.3015842 | 0.089040783 | 0.7781 | 302.2322418 | 30276.47159 | 59392.06659 | 0.076504412 | -1.11631352 | FALSE | FALSE | FALSE |
| OR2AG2 | 2.3919577 | 0.44238862 | 0.75405 | 2.391957723 | 22.63346047 | 22.63346047 | 2.72090234 | 0.433878262 | 0.87525 | 2.72090234 | 251.1314047 | 251.1314047 | 0.879104585 | -0.055959455 | FALSE | FALSE | FALSE |
| STEAP2 | 2.3915808 | 0.44256823 | 0.68755 | 5.557547369 | 211.8365978 | 211.8365978 | 0.86055915 | 0.605060873 | 0.7496 | 0.86055915 | 8.047773029 | 8.047773029 | 2.779100968 | 0.443904326 | FALSE | TRUE | FALSE |
| PPP1R42 | 2.3913012 | 0.44274771 | 0.50485 | 2.391301207 | 2.391301207 | 2.391301207 | 0.31383662 | 0.820701386 | 1 | 0.313836616 | 0.313836616 | 0.313836616 | 7.619573654 | 0.881930671 | FALSE | TRUE | FALSE |
| MYL9 | 2.3896885 | 0.44292707 | 0.1267 | 2.389688507 | 9.560528003 | 9.560528003 | 11.9554713 | 0.178254545 | 0.1286 | 2.988202489 | 11.95547127 | 11.95547127 | 0.199882418 | -0.699225405 | FALSE | FALSE | FALSE |
| LYZL1 | 2.3873809 | 0.44310633 | 0.5016 | 2.387380895 | 2.387380895 | 2.387380895 | 0.31081264 | 0.821765123 | 1 | 0.310812636 | 0.310812636 | 0.310812636 | 7.681093435 | 0.885423048 | FALSE | TRUE | FALSE |
| DUSP15 | 2.3835677 | 0.4432855 | 0.499 | 1.367725618 | 21.88049921 | 21.88049921 | 1.75344877 | 0.512703424 | 0.00465 | 1.753448766 | 1.753448766 | 1.753448766 | 1.359359776 | 0.133334415 | FALSE | FALSE | FALSE |
| THAP5 | 2.3818053 | 0.44346456 | 0.6857 | 2.38180532 | 222.3963994 | 222.3963994 | 0.04693481 | 0.8873403 | 1 | 0.046934805 | 0.428919342 | 0.428919342 | 50.74710122 | 1.70541124 | FALSE | TRUE | FALSE |
| FOXB2 | 2.3753453 | 0.44364358 | 0.81815 | 22.3153943 | 859.8224476 | 3439.002265 | 0.02858612 | 0.891580124 | 0.98425 | 1.826830761 | 29.23723248 | 117.0086386 | 83.0943485 | 1.919571487 | FALSE | TRUE | FALSE |
| SSR3 | 2.3726373 | 0.44382251 | 0.50575 | 2.372637267 | 2.372637267 | 2.372637267 | 0.6379843 | 0.695552904 | 1 | 0.637984296 | 0.637984296 | 0.637984296 | 3.718958729 | 0.570421359 | FALSE | TRUE | FALSE |
| TTR | 2.3715 | 0.44400131 | 0.25305 | 0.59300333 | 2.371500046 | 2.371500046 | 0.1879473 | 0.855049159 | 1 | 0.751482757 | 0.751482757 | 0.751482757 | 12.61789913 | 1.100987051 | FALSE | TRUE | FALSE |
| ABTB2 | 2.3712146 | 0.44417997 | 0.78785 | 9.483277596 | 220.1880843 | 880.6055634 | 0.09931252 | 0.876140217 | 1 | 0.099312519 | 0.099312519 | 1.948476932 | 23.87629049 | 1.377966854 | FALSE | TRUE | FALSE |
| GGT7 | 2.3704967 | 0.4443585 | 0.51105 | 2.370496701 | 2.370496701 | 46.62244514 | 0.30965013 | 0.822155849 | 1 | 0.309650128 | 0.309650128 | 6.102313437 | 7.655403581 | 0.883968091 | FALSE | TRUE | FALSE |
| SCARA5 | 2.3694581 | 0.4445369 | 0.3254 | 2.369458087 | 230.7706706 | 230.7706706 | 0.4169057 | 0.785808568 | 0.5042 | 0.4169057 | 1.666468754 | 3.535675608 | 5.683438931 | 0.754611198 | FALSE | TRUE | FALSE |
| KRBA1 | 2.3640694 | 0.44471525 | 1 | 1.60033E+17 | 5.59377E+21 | 1.15818E+24 | 0.05004175 | 0.886618179 | 1 | 1.20244E+12 | 2.99879E+16 | 5.29472E+18 | 47.24194276 | 1.674327749 | FALSE | TRUE | FALSE |
| RAB23 | 2.3638905 | 0.44489346 | 0.93525 | 22.58661477 | 2223.514206 | 2223.514206 | 0.31752072 | 0.819443885 | 1 | 2.968246857 | 28.47837862 | 28.47837862 | 7.444838615 | 0.871855288 | FALSE | TRUE | FALSE |
| SLC22A9 | 2.3598473 | 0.4450716 | 0.25825 | 0.262972644 | 2.359847347 | 5.206499239 | 3.26608705 | 0.396665069 | 0.50935 | 30.26452754 | 287.4690633 | 2797.312128 | 0.72253045 | -0.141143845 | FALSE | FALSE | FALSE |
| TNF | 2.3572813 | 0.44524964 | 0.8759 | 9.43069043 | 37.73845386 | 37.73845386 | 2.53848349 | 0.44465724 | 0.75275 | 2.538483494 | 10.15575823 | 10.15575823 | 0.928617929 | -0.032162935 | FALSE | FALSE | FALSE |
| BLMH | 2.356878 | 0.44542754 | 0.75385 | 2.356877951 | 22.48095619 | 22.48095619 | 2.77598682 | 0.428859589 | 0.2585 | 2.775986818 | 26.545113 | 26.545113 | 0.849023466 | -0.071080306 | FALSE | FALSE | FALSE |
| CHRNA2 | 2.3567162 | 0.4456053 | 1 | 8.37579E+18 | 1.38159E+25 | 2.95314E+27 | 0.0541289 | 0.885870939 | 1 | 1.20734E+22 | 2.8334E+28 | 2.67761E+31 | 43.5389601 | 1.638878052 | FALSE | TRUE | FALSE |
| SLC4A10 | 2.356258 | 0.44578292 | 0.00375 | 0.069880857 | 0.069880857 | 1.371218724 | 0.02841291 | 0.891633392 | 1 | 0.028412907 | 0.028412907 | 0.558792863 | 82.92914271 | 1.918707176 | FALSE | TRUE | FALSE |
| MRPL19 | 2.3550363 | 0.44596042 | 0.2551 | 0.588734014 | 2.355036322 | 2.355036322 | 0.18838952 | 0.854840284 | 1 | 0.18838952 | 0.18838952 | 0.18838952 | 12.50088818 | 1.09694087 | FALSE | TRUE | FALSE |
| SLC44A5 | 2.3511244 | 0.44613785 | 0.2536 | 0.258304558 | 2.351124356 | 5.104746683 | 3.28718004 | 0.395318934 | 0.2575 | 0.35836455 | 3.287180041 | 7.092301614 | 0.715240518 | -0.145547891 | FALSE | FALSE | FALSE |
| TNKS | 2.3498758 | 0.44631516 | 0.03025 | 0.587650536 | 2.34987583 | 19.7929112 | 0.0889765 | 0.878332525 | 1 | 0.824474001 | 0.824474001 | 16.08167377 | 26.41007205 | 1.421769586 | FALSE | TRUE | FALSE |
| E2F8 | 2.3449118 | 0.4464924 | 0.50945 | 2.344911779 | 2.344911779 | 46.12570955 | 2.7644372 | 0.429870451 | 0.5113 | 2.764437204 | 2.764437204 | 10.01451678 | 0.848242013 | -0.071480221 | FALSE | FALSE | FALSE |
| UBE2D4 | 2.3434028 | 0.44666954 | 0.50625 | 2.343402799 | 9.372428316 | 9.372428316 | 0.30063971 | 0.825351531 | 1 | 0.300639712 | 2.790770319 | 2.790770319 | 7.794721412 | 0.891800598 | FALSE | TRUE | FALSE |
| HEXIM2 | 2.3410789 | 0.44684656 | 0.75035 | 2.34107891 | 22.09659211 | 22.09659211 | 0.3174342 | 0.819484082 | 1 | 0.317434196 | 0.317434196 | 0.317434196 | 7.375005392 | 0.867762342 | FALSE | TRUE | FALSE |
| KBTBD11 | 2.3338069 | 0.44702357 | 0.94045 | 22.35038983 | 2210.483011 | 2210.483011 | 0.35612865 | 0.807035462 | 1 | 3.347894296 | 3.347894296 | 3.347894296 | 6.553269253 | 0.816458012 | FALSE | TRUE | FALSE |
| RAB3B | 2.3328153 | 0.44720045 | 0.5014 | 2.332815267 | 2.332815267 | 2.332815267 | 0.30127926 | 0.825106972 | 1 | 0.301279263 | 0.301279263 | 0.301279263 | 7.743032973 | 0.888911109 | FALSE | TRUE | FALSE |
| ABCD4 | 2.3326931 | 0.4473772 | 0.91845 | 149.3780724 | 9587.337226 | 153875.0843 | 426.480436 | 0.005575884 | 0.2361 | 58.48705304 | 3747.879875 | 31712.29404 | 0.005469637 | -2.262041514 | FALSE | FALSE | FALSE |
| TAOK1 | 2.3314257 | 0.44755382 | 0.02995 | 0.475469212 | 0.475469212 | 9.32316796 | 0.15465024 | 0.863410208 | 1 | 0.154650236 | 0.154650236 | 3.039587636 | 15.07547442 | 1.178270988 | FALSE | TRUE | FALSE |
| TMUB1 | 2.3289707 | 0.44773035 | 0.8776 | 21.90759614 | 211.5002607 | 211.5002607 | 0.67000868 | 0.680365462 | 1 | 0.670008679 | 0.670008679 | 0.670008679 | 3.476030628 | 0.541083594 | FALSE | TRUE | FALSE |
| NOL7 | 2.3280458 | 0.44790675 | 0.50735 | 2.328045801 | 2.328045801 | 2.328045801 | 0.30691294 | 0.823163633 | 1 | 0.306912944 | 0.306912944 | 0.306912944 | 7.585362061 | 0.879976315 | FALSE | TRUE | FALSE |
| NME7 | 2.3279251 | 0.44808301 | 0.891 | 51.07605855 | 1986.27084 | 19787.25176 | 0.10442805 | 0.874868397 | 1 | 0.104428053 | 0.104428053 | 0.104428053 | 22.29214341 | 1.348151828 | FALSE | TRUE | FALSE |
| TLK1 | 2.3275361 | 0.44825914 | 0.51195 | 2.327536087 | 2.327536087 | 45.90080477 | 0.30239137 | 0.824691515 | 0.01755 | 0.30239137 | 0.30239137 | 5.973410556 | 7.69709825 | 0.88632703 | FALSE | TRUE | FALSE |
| ARCN1 | 2.3268824 | 0.44843513 | 0.015 | 2.326882366 | 2.326882366 | 19.68009776 | 0.62757015 | 0.70024308 | 1 | 0.627570145 | 0.627570145 | 12.44164433 | 3.707764598 | 0.569112153 | FALSE | TRUE | FALSE |
| STARD4 | 2.3262098 | 0.448611 | 0.50295 | 2.326209831 | 9.307651696 | 9.307651696 | 0.18624083 | 0.855465177 | 1 | 0.744838988 | 0.744838988 | 0.744838988 | 12.49033189 | 1.096573979 | FALSE | TRUE | FALSE |
| EFCAB2 | 2.3231562 | 0.44878678 | 0.5013 | 2.323156196 | 2.323156196 | 2.323156196 | 2.73646212 | 0.432482022 | 0.5004 | 2.736462125 | 2.736462125 | 2.736462125 | 0.848963403 | -0.071111031 | FALSE | FALSE | FALSE |
| VCAM1 | 2.3226205 | 0.44896243 | 0.51105 | 2.322620492 | 2.322620492 | 4.908654125 | 0.30226645 | 0.824767204 | 1 | 0.30226645 | 0.30226645 | 5.977565511 | 7.684016849 | 0.885588309 | FALSE | TRUE | FALSE |
| R3HDM1 | 2.3219273 | 0.44913795 | 0.8825 | 22.18793835 | 217.4650462 | 435.1707078 | 0.32247786 | 0.817984761 | 0.02335 | 0.322477861 | 0.322477861 | 6.339479279 | 7.200268818 | 0.857348711 | FALSE | TRUE | FALSE |
| IFT20 | 2.3217611 | 0.44931334 | 0.002 | 2.321761073 | 2.321761073 | 2.321761073 | 0.6257181 | 0.700646263 | 1 | 0.625718098 | 0.625718098 | 0.625718098 | 3.71055445 | 0.569438809 | FALSE | TRUE | FALSE |
| SNAP29 | 2.3188949 | 0.44948863 | 0.50665 | 2.318894935 | 2.318894935 | 2.318894935 | 2.73750104 | 0.432081834 | 0.0075 | 2.737501036 | 2.737501036 | 2.737501036 | 0.847084587 | -0.07207322 | FALSE | FALSE | FALSE |
| SFMBT2 | 2.3187458 | 0.44966379 | 0.02555 | 2.318745796 | 2.318745796 | 45.4643869 | 0.30186799 | 0.824937258 | 0.02515 | 0.301867993 | 0.301867993 | 5.932816907 | 7.681323799 | 0.885436073 | FALSE | TRUE | FALSE |
| SNRNP40 | 2.314572 | 0.44983888 | 0.5094 | 2.314572001 | 9.257081291 | 9.257081291 | 0.30598556 | 0.82354812 | 1 | 0.305985558 | 0.305985558 | 0.305985558 | 7.564317782 | 0.878769766 | FALSE | TRUE | FALSE |
| HES2 | 2.3136183 | 0.45001384 | 0.5033 | 2.313618339 | 2.313618339 | 2.313618339 | 0.62401901 | 0.701563577 | 1 | 0.624019006 | 0.624019006 | 0.624019006 | 3.707608772 | 0.569093901 | FALSE | TRUE | FALSE |
| LDHAL6B | 2.3128724 | 0.45018868 | 0.2573 | 0.24892768 | 2.312872387 | 2.312872387 | 3.33735229 | 0.39283776 | 0.69205 | 3.337352287 | 312.9896064 | 312.9896064 | 0.693026144 | -0.159250382 | FALSE | FALSE | FALSE |
| ZNF174 | 2.3128546 | 0.45036338 | 0.75575 | 2.312854553 | 21.87144373 | 21.87144373 | 0.31367933 | 0.820780625 | 1 | 0.313679327 | 0.313679327 | 6.22769808 | 7.373308844 | 0.867662425 | FALSE | TRUE | FALSE |
| RGL1 | 2.3101747 | 0.45053799 | 0.50915 | 2.310174669 | 2.310174669 | 45.4850109 | 0.29334413 | 0.827081639 | 0.50025 | 2.726768791 | 2.726768791 | 9.918902921 | 7.875305468 | 0.896267408 | FALSE | TRUE | FALSE |
| KIAA1191 | 2.3071845 | 0.4507125 | 0.75005 | 9.228618345 | 147.7648194 | 147.7648194 | 0.11981154 | 0.871257276 | 0.00845 | 0.119811541 | 0.119811541 | 0.119811541 | 19.2567803 | 1.284583676 | FALSE | TRUE | FALSE |
| NDUFS6 | 2.3053096 | 0.45088691 | 0.50535 | 2.305309645 | 2.305309645 | 2.305309645 | 0.3004438 | 0.825407868 | 1 | 0.300443795 | 0.300443795 | 0.300443795 | 7.67301466 | 0.884966028 | FALSE | TRUE | FALSE |
| IL10RB | 2.3027577 | 0.45106123 | 0.7535 | 2.302757747 | 21.73507187 | 21.73507187 | 0.31286463 | 0.821077138 | 1 | 0.312864633 | 0.312864633 | 0.312864633 | 7.360236672 | 0.866891779 | FALSE | TRUE | FALSE |
| LIN37 | 2.2996069 | 0.45123546 | 0.5026 | 2.299606858 | 2.299606858 | 2.299606858 | 0.29363635 | 0.826971033 | 0.50545 | 2.712173155 | 2.712173155 | 2.712173155 | 7.831478733 | 0.893843773 | FALSE | TRUE | FALSE |
| ANKRD11 | 2.2987136 | 0.45140956 | 0.0034 | 0.068979834 | 1.287719948 | 2.298713633 | 0.85896657 | 0.605781253 | 0.5387 | 0.858966575 | 1.721422403 | 28.69306863 | 2.676138631 | 0.427508607 | FALSE | TRUE | FALSE |
| SPRY2 | 2.297992 | 0.45158354 | 0.5014 | 2.29799205 | 2.29799205 | 11.40885514 | 0.62042612 | 0.703436672 | 1 | 0.620426122 | 0.620426122 | 0.620426122 | 3.703893125 | 0.568658447 | FALSE | TRUE | FALSE |
| SCCPDH | 2.2974936 | 0.4517574 | 0.50775 | 2.297493599 | 2.297493599 | 4.930365688 | 0.29231514 | 0.827375909 | 1 | 2.712268365 | 2.712268365 | 2.712268365 | 7.859646279 | 0.895403001 | FALSE | TRUE | FALSE |
| CD38 | 2.2970569 | 0.45193112 | 0.2545 | 2.297056943 | 21.65934961 | 21.65934961 | 0.30290157 | 0.824464037 | 1 | 2.795929382 | 2.795929382 | 2.795929382 | 7.583509556 | 0.879870238 | FALSE | TRUE | FALSE |
| CCNL2 | 2.2952058 | 0.45210473 | 0.0208 | 2.295205828 | 2.295205828 | 45.19546641 | 0.29847903 | 0.825801244 | 1 | 0.298479034 | 0.298479034 | 5.888345957 | 7.689671868 | 0.885907808 | FALSE | TRUE | FALSE |
| PPP4R1 | 2.2946368 | 0.45227822 | 0.7548 | 2.294636781 | 21.8633432 | 77.40355237 | 0.30996071 | 0.821999768 | 0.02355 | 0.309960711 | 0.309960711 | 6.102168256 | 7.402992384 | 0.869407303 | FALSE | TRUE | FALSE |
| MYCT1 | 2.2936837 | 0.45245159 | 0.50495 | 2.293683722 | 2.293683722 | 2.293683722 | 24.7337302 | 0.111249549 | 0.00495 | 2.620040842 | 24.73373021 | 24.73373021 | 0.092735051 | -1.032756087 | FALSE | FALSE | FALSE |
| PPP1R7 | 2.2921462 | 0.45262484 | 0.50385 | 2.292146171 | 2.292146171 | 11.36135641 | 0.6121459 | 0.706559183 | 1 | 2.448518898 | 2.448518898 | 2.448518898 | 3.744444221 | 0.573387365 | FALSE | TRUE | FALSE |
| COX7A2 | 2.2916997 | 0.45279797 | 0.49875 | 0.243322803 | 2.291699713 | 2.291699713 | 0.25374458 | 0.838298822 | 1 | 0.253744579 | 2.392229602 | 2.392229602 | 9.031521858 | 0.955760937 | FALSE | TRUE | FALSE |
| ST8SIA2 | 2.2802677 | 0.45297115 | 0.7583 | 5.383846543 | 21.53458798 | 21.53458798 | 0.61625151 | 0.705340662 | 1 | 5.823780191 | 5.823780191 | 5.823780191 | 3.700222561 | 0.568227847 | FALSE | TRUE | FALSE |
| OR51G2 | 2.2796057 | 0.4531442 | 0.75245 | 2.279605668 | 21.58861261 | 21.58861261 | 210.875169 | 0.012657095 | 0.49825 | 210.8751685 | 222855.7938 | 222855.7938 | 0.010810214 | -1.966165714 | FALSE | FALSE | FALSE |
| APOB | 2.2775021 | 0.45331716 | 0.05405 | 0.030126702 | 2.277502147 | 19.17214493 | 0.00565652 | 0.897955898 | 0.53115 | 0.011797054 | 0.403494433 | 1.886464196 | 402.6331666 | 2.604909547 | FALSE | TRUE | FALSE |
| TLR5 | 2.2736502 | 0.45349004 | 0.6905 | 2.273650236 | 199.7461988 | 199.7461988 | 3.6998677 | 0.37135525 | 0.01405 | 0.405702908 | 3.699867697 | 8.012972042 | 0.614522038 | -0.211462538 | FALSE | FALSE | FALSE |
| ZFP42 | 2.2732087 | 0.45366279 | 0.0106 | 2.273208651 | 2.273208651 | 4.897673831 | 24.6768196 | 0.111550165 | 0.0058 | 2.601740399 | 24.6768196 | 24.6768196 | 0.092119191 | -1.035649886 | FALSE | FALSE | FALSE |
| C3AR1 | 2.2729643 | 0.45383541 | 0.8158 | 21.09411526 | 489.9229671 | 1959.185071 | 7.12592604 | 0.242434953 | 0.8162 | 68.14989447 | 2671.717282 | 26831.00725 | 0.31897108 | -0.496248692 | FALSE | FALSE | FALSE |
| CLDND2 | 2.2719087 | 0.45400791 | 0.5027 | 2.271908741 | 2.271908741 | 2.271908741 | 0.28729131 | 0.828922441 | 1 | 2.688764125 | 2.688764125 | 2.688764125 | 7.908031466 | 0.898068389 | FALSE | TRUE | FALSE |
| DPY19L2 | 2.2656195 | 0.45418038 | 0.14345 | 2.265619503 | 9.066589089 | 44.61955214 | 0.18390247 | 0.855969117 | 1 | 0.183902466 | 0.183902466 | 3.628476681 | 12.31967981 | 1.090599421 | FALSE | TRUE | FALSE |

| | | | | | | | | | | | | | | | | | |
|---|---|---|---|---|---|---|---|---|---|---|---|---|---|---|---|---|---|
| MMP27 | 2.2646971 | 0.45435273 | 0.69115 | 9.056925637 | 199.3138565 | 797.0914813 | 0.0468101 | 0.887397385 | 1 | 0.416615032 | 1.666157843 | 1.666157843 | 48.38052367 | 1.684670565 | FALSE | TRUE | FALSE |
| KRTAP10-12 | 2.2638385 | 0.45452496 | 0.5111 | 2.26383847 | 2.26383847 | 2.26383847 | 0.29936967 | 0.825651591 | 1 | 0.299369669 | 0.299369669 | 0.299369669 | 7.562016811 | 0.878637639 | FALSE | TRUE | FALSE |
| POLI | 2.2634972 | 0.45469706 | 0.6971 | 2.263497233 | 201.1201772 | 201.1201772 | 0.04789044 | 0.887152367 | 0.01355 | 0.047890438 | 0.047890438 | 0.948083682 | 47.26407409 | 1.674531154 | FALSE | TRUE | FALSE |
| TYRP1 | 2.2504011 | 0.45486924 | 0.24245 | 0.140987213 | 19.7785065 | 143.7783124 | 0.00789094 | 0.897346026 | 0.91495 | 0.267191952 | 32.23910047 | 148.4901319 | 285.1879196 | 2.455131125 | FALSE | TRUE | FALSE |
| COQ3 | 2.2446534 | 0.45504138 | 0.2543 | 2.244653412 | 21.20825021 | 21.20825021 | 2127.69194 | 0.001114278 | 0.1121 | 214.1943316 | 21651.44505 | 225577.6685 | 0.001054971 | -2.976759455 | FALSE | FALSE | FALSE |
| GRK6 | 2.2436766 | 0.4552134 | 0.51045 | 2.243676601 | 2.243676601 | 44.14074909 | 0.2933011 | 0.82710006 | 1 | 0.293301099 | 0.293301099 | 5.781573274 | 7.649738127 | 0.883646568 | FALSE | TRUE | FALSE |
| MDGA1 | 2.2415474 | 0.45538532 | 0.51305 | 2.24154742 | 2.24154742 | 43.81421833 | 2.57330147 | 0.442142167 | 0.7571 | 2.573301475 | 24.6809451 | 50.43624886 | 0.871078435 | -0.059942738 | FALSE | FALSE | FALSE |
| R3HDML | 2.2406206 | 0.45555712 | 0.87535 | 2.240620642 | 204.4062432 | 204.4062432 | 1907.31224 | 0.001260271 | 0.8304 | 19325.75125 | 18637958.9 | 206547875.2 | 0.001174753 | -2.930053462 | FALSE | FALSE | FALSE |
| MFSD9 | 2.2404118 | 0.45572879 | 0.50965 | 21.09454082 | 203.5921447 | 2012.992674 | 0.39692458 | 0.792992761 | 0.87445 | 3.674856087 | 34.89692704 | 34.89692704 | 5.644426867 | 0.75161985 | FALSE | TRUE | FALSE |
| OR52N2 | 2.240021 | 0.45590034 | 0.8118 | 44.51598722 | 2058.726527 | 21022.52197 | 0.33418157 | 0.814229793 | 1 | 0.334181567 | 0.334181567 | 0.334181567 | 6.703005996 | 0.826269608 | FALSE | TRUE | FALSE |
| TAS2R13 | 2.2365082 | 0.4560718 | 0.4975 | 0.243658963 | 2.23650823 | 2.23650823 | 0.29603322 | 0.826397364 | 1 | 0.296033225 | 0.296033225 | 0.296033225 | 7.55492304 | 0.878230045 | FALSE | TRUE | FALSE |
| HSD17B13 | 2.2357883 | 0.45624315 | 0.6003 | 3.839688955 | 85.3250969 | 341.1782469 | 0.04141579 | 0.888624063 | 0.8767 | 3.532727338 | 33.90325162 | 33.90325162 | 53.98396483 | 1.732264778 | FALSE | TRUE | FALSE |
| ZMAT5 | 2.2335455 | 0.4564144 | 0.49765 | 0.243123329 | 2.233545537 | 2.233545537 | 0.29217478 | 0.827467657 | 1 | 1.168653341 | 1.168653341 | 1.168653341 | 7.644552807 | 0.883352085 | FALSE | TRUE | FALSE |
| PRELID2 | 2.2321836 | 0.45658554 | 0.50115 | 2.23218362 | 2.23218362 | 2.23218362 | 2.55613816 | 0.443305784 | 0.74935 | 2.556138162 | 24.08433255 | 24.08433255 | 0.873264072 | -0.058854407 | FALSE | FALSE | FALSE |
| CYFIP2 | 2.2291612 | 0.45675659 | 0.5201 | 2.229161243 | 2.229161243 | 43.30698052 | 0.29084466 | 0.827833685 | 1 | 0.290844662 | 0.290844662 | 5.669878461 | 7.664439243 | 0.884480386 | FALSE | TRUE | FALSE |
| CALHM1 | 2.2281793 | 0.45692752 | 0.94025 | 21.0573192 | 2027.981498 | 2027.981498 | 2.81155597 | 0.42621577 | 0.75685 | 2.811555968 | 26.71309323 | 26.71309323 | 0.792507543 | -0.100996596 | FALSE | FALSE | FALSE |
| P2RY1 | 2.2277789 | 0.45709833 | 0.011 | 2.227778916 | 2.227778916 | 11.04089862 | 0.58493958 | 0.718789642 | 1 | 5.524017643 | 5.524017643 | 11.6077029 | 3.808562475 | 0.580761084 | FALSE | TRUE | FALSE |
| SYT9 | 2.2227915 | 0.45726909 | 0.50595 | 2.222791478 | 2.222791478 | 4.703058022 | 0.29033185 | 0.827943191 | 1 | 0.290331853 | 0.290331853 | 5.757634079 | 7.656037234 | 0.884004037 | FALSE | TRUE | FALSE |
| GDPGP1 | 2.2205926 | 0.45743975 | 0.7513 | 2.220592597 | 20.96504655 | 20.96504655 | 0.30292923 | 0.824445053 | 1 | 0.302929232 | 0.302929232 | 6.010977127 | 7.330400514 | 0.865127704 | FALSE | TRUE | FALSE |
| GRM8 | 2.2197091 | 0.4576103 | 0.06615 | 0.060561022 | 2.219709138 | 8.870735528 | 0.00268573 | 0.898808187 | 0.9401 | 0.010725194 | 0.171166022 | 0.362109051 | 826.4837812 | 2.917234335 | FALSE | TRUE | FALSE |
| GIMAP7 | 2.217602 | 0.45778074 | 0.99155 | 2037.632617 | 217500.3454 | 217500.3454 | 2612.60216 | 0.000889948 | 0.1816 | 26.68899401 | 2612.602156 | 26809.23778 | 0.00084881 | -3.071189673 | FALSE | FALSE | FALSE |
| SLC6A15 | 2.2157733 | 0.45795108 | 0.0151 | 0.261896772 | 1.047332604 | 5.172160986 | 0.36525996 | 0.804043554 | 0.50195 | 0.365259959 | 0.365259959 | 3.09199412 | 6.066291383 | 0.782923267 | FALSE | TRUE | FALSE |
| MFSD10 | 2.2137731 | 0.45812133 | 0.4763 | 1.265828593 | 20.23625453 | 80.9413724 | 0.24125575 | 0.84169343 | 0.5091 | 0.24125575 | 0.24125575 | 0.532177214 | 9.176043071 | 0.962655442 | FALSE | TRUE | FALSE |
| NOX4 | 2.2131963 | 0.45829145 | 0.1523 | 0.138458094 | 2.213196285 | 10.96493129 | 1.38125947 | 0.547161831 | 0.0389 | 0.086420453 | 0.345455144 | 1.381259473 | 1.602303063 | 0.204744663 | FALSE | FALSE | FALSE |
| FERMT1 | 2.2124742 | 0.45846144 | 0.74925 | 8.851872061 | 141.7993714 | 567.7491674 | 61.0350134 | 0.045917249 | 0.3322 | 15.25020045 | 244.3379707 | 301.4700784 | 0.036249261 | -1.440700839 | FALSE | FALSE | FALSE |
| RNF185 | 2.2085666 | 0.45863138 | 0.5115 | 2.208566638 | 8.831165967 | 8.831165967 | 1.13535784 | 0.573905139 | 0.75095 | 1.13535784 | 4.541143496 | 4.541143496 | 1.945260393 | 0.288977744 | FALSE | FALSE | FALSE |
| TPTE | 2.2060914 | 0.45880122 | 1 | 19312091.37 | 2.91139E+11 | 4.96978E+12 | 303.636713 | 0.008254242 | 0.9929 | 32808326.25 | 44704327124 | 7.79143E+11 | 0.007265562 | -2.138730772 | FALSE | FALSE | FALSE |
| ANKRD16 | 2.2034969 | 0.45897097 | 0.5065 | 2.203496922 | 8.816846482 | 8.816846482 | 0.17958607 | 0.857144572 | 1 | 0.179586065 | 0.179586065 | 0.179586065 | 12.2698658 | 1.088839813 | FALSE | TRUE | FALSE |
| SLC41A1 | 2.2024259 | 0.4591406 | 0.5085 | 2.202425871 | 2.202425871 | 18.64932927 | 0.59099979 | 0.715937044 | 0.50785 | 2.363978282 | 2.363978282 | 11.69721402 | 3.726610242 | 0.571313973 | FALSE | TRUE | FALSE |
| ATG3 | 2.2018459 | 0.45931012 | 0.00865 | 2.20184592 | 2.20184592 | 2.20184592 | 0.29041903 | 0.827906704 | 1 | 0.290419028 | 0.290419028 | 0.290419028 | 7.581617288 | 0.879761858 | FALSE | TRUE | FALSE |
| PHB2 | 2.1993645 | 0.45947954 | 0.75365 | 2.199364501 | 8.79939346 | 10.91533678 | 0.6035344 | 0.710634329 | 1 | 0.603534404 | 0.603534404 | 0.603534404 | 3.644141058 | 0.561595179 | FALSE | TRUE | FALSE |
| ANGPTL4 | 2.1955898 | 0.45964889 | 0.255 | 0.548996351 | 2.195589787 | 2.195589787 | 0.77170946 | 0.631661501 | 0.50435 | 0.771709457 | 0.771709457 | 1.697349398 | 2.845098978 | 0.45409738 | FALSE | TRUE | FALSE |
| LTBR | 2.1954148 | 0.45981812 | 0.01255 | 2.195414837 | 2.195414837 | 4.645196196 | 0.28707397 | 0.828994534 | 1 | 0.287073965 | 0.287073965 | 0.287073965 | 7.647558135 | 0.883522787 | FALSE | TRUE | FALSE |
| FABP12 | 2.1953734 | 0.45998722 | 0.7481 | 2.195373446 | 20.76986875 | 20.76986875 | 0.29945646 | 0.825614139 | 1 | 0.299456456 | 0.299456456 | 0.299456456 | 7.331194252 | 0.865174727 | FALSE | TRUE | FALSE |
| DPP10 | 2.194143 | 0.4601562 | 0.5057 | 2.194143008 | 2.194143008 | 7.915245778 | 0.28687384 | 0.829084563 | 1 | 0.28687384 | 0.28687384 | 5.671212783 | 7.648459703 | 0.883573983 | FALSE | TRUE | FALSE |
| C1QL2 | 2.1870002 | 0.46032517 | 0.5013 | 2.187000157 | 2.187000157 | 2.187000157 | 0.28772951 | 0.828796131 | 1 | 0.287729513 | 0.287729513 | 0.287729513 | 7.600889228 | 0.880864403 | FALSE | TRUE | FALSE |
| PDCD7 | 2.1856323 | 0.46049403 | 0.5052 | 2.185632306 | 2.185632306 | 2.185632306 | 0.28976114 | 0.828216328 | 0.0086 | 0.289761144 | 0.289761144 | 0.289761144 | 7.542875755 | 0.877536954 | FALSE | TRUE | FALSE |
| PIGM | 2.1836942 | 0.4606628 | 0.51225 | 2.183694215 | 2.183694215 | 4.656529959 | 0.28760712 | 0.828814187 | 1 | 0.287607117 | 0.287607117 | 5.700262775 | 7.592629279 | 0.880392195 | FALSE | TRUE | FALSE |
| FHDC1 | 2.1815923 | 0.46083146 | 0.03345 | 0.111908249 | 0.111908249 | 2.181592304 | 0.14454786 | 0.865767437 | 0.0296 | 0.144547855 | 0.144547855 | 2.826467691 | 15.09252629 | 1.178761941 | FALSE | TRUE | FALSE |
| COX11 | 2.1809963 | 0.46100001 | 0.50485 | 2.180996301 | 2.180996301 | 2.180996301 | 0.2887471 | 0.828452348 | 1 | 0.288747102 | 0.288747102 | 0.288747102 | 7.553309743 | 0.878137294 | FALSE | TRUE | FALSE |
| DND1 | 2.178503 | 0.46116847 | 0.50525 | 2.178503035 | 2.178503035 | 2.178503035 | 0.25516296 | 0.837878533 | 1 | 0.25516296 | 0.25516296 | 0.25516296 | 8.537693066 | 0.931340538 | FALSE | TRUE | FALSE |
| FBXO4 | 2.1762173 | 0.46133683 | 0.50515 | 2.176217289 | 2.176217289 | 2.176217289 | 22.1294888 | 0.12249376 | 0.6866 | 22.12948876 | 2118.224702 | 2118.224702 | 0.098340152 | -1.007269125 | FALSE | FALSE | FALSE |
| RIC8A | 2.1760027 | 0.46150507 | 0.51075 | 2.176002688 | 8.703206089 | 31.74362774 | 0.28987674 | 0.828198147 | 1 | 0.289876737 | 0.289876737 | 5.726875791 | 7.506648216 | 0.875446064 | FALSE | TRUE | FALSE |
| SEMA7A | 2.1757144 | 0.46167318 | 0.51195 | 2.17571438 | 2.17571438 | 7.961459664 | 0.28750642 | 0.828868331 | 1 | 0.28750642 | 0.28750642 | 5.671619978 | 7.567533213 | 0.878954336 | FALSE | TRUE | FALSE |
| REPIN1 | 2.1740886 | 0.46184119 | 0.7542 | 2.174088616 | 20.73788819 | 42.71025725 | 0.29530268 | 0.826656902 | 1 | 0.295302676 | 0.295302676 | 5.813653376 | 7.3622381 | 0.867009859 | FALSE | TRUE | FALSE |
| MYLK2 | 2.1736535 | 0.46200908 | 0.50645 | 2.17365347 | 2.17365347 | 4.633773407 | 0.28357306 | 0.830122024 | 1 | 1.134207846 | 1.134207846 | 5.611016735 | 7.665232704 | 0.884525344 | FALSE | TRUE | FALSE |
| UAP1 | 2.1688175 | 0.46217692 | 0.5094 | 2.168817455 | 2.168817455 | 4.652881131 | 0.28200597 | 0.830565295 | 1 | 0.282005974 | 0.282005974 | 5.594469462 | 7.690679115 | 0.885964691 | FALSE | TRUE | FALSE |
| INHBA | 2.1650364 | 0.46234469 | 0.50305 | 2.165036357 | 2.165036357 | 10.71609322 | 0.57800397 | 0.72143455 | 1 | 2.311849083 | 2.311849083 | 11.45663148 | 3.745711929 | 0.573534374 | FALSE | TRUE | FALSE |
| LEFTY1 | 2.163988 | 0.46251235 | 0.25935 | 2.163988012 | 8.654614773 | 8.654614773 | 1.10460407 | 0.577887662 | 0.50615 | 2.574827503 | 10.29790644 | 10.29790644 | 1.959062132 | 0.29204821 | FALSE | FALSE | FALSE |
| HUS1B | 2.1591719 | 0.46267995 | 0.51005 | 2.159171914 | 2.159171914 | 4.663353014 | 2.48440658 | 0.447535758 | 0.7491 | 2.484406585 | 23.46180025 | 23.46180025 | 0.869089596 | -0.060935449 | FALSE | FALSE | FALSE |
| ARHGEF26 | 2.154988 | 0.4628475 | 0.69475 | 8.617819415 | 193.7603489 | 774.8496386 | 0.04334064 | 0.88823128 | 1 | 0.38910619 | 3.585442434 | 7.646887172 | 49.72210567 | 1.696549512 | FALSE | TRUE | FALSE |
| GDA | 2.1533342 | 0.46301494 | 0.50965 | 2.153334238 | 8.613421049 | 8.613421049 | 0.17076767 | 0.85941635 | 0.87335 | 2.7306036 | 10.92315209 | 10.92315209 | 12.60973046 | 1.100705803 | FALSE | TRUE | FALSE |
| AGAP2 | 2.1508755 | 0.4631823 | 0.7609 | 2.150875536 | 20.70877459 | 72.45449318 | 0.27291702 | 0.832988854 | 1 | 2.563962127 | 24.72167933 | 86.46567282 | 7.88106047 | 0.89658466 | FALSE | TRUE | FALSE |
| OR4E2 | 2.1504285 | 0.46334953 | 0.0082 | 2.150428531 | 2.150428531 | 2.150428531 | 0.2762347 | 0.832020698 | 1 | 2.557580846 | 2.557580846 | 2.557580846 | 7.784787812 | 0.89124678 | FALSE | TRUE | FALSE |
| MAS1L | 2.1474433 | 0.46351669 | 1 | 4.44306E+23 | 4.7554E+30 | 1.18277E+33 | 4.4702242 | 0.331598799 | 1 | 6.66289E+19 | 3.51097E+25 | 7.64878E+27 | 0.480388268 | -0.318407607 | FALSE | FALSE | FALSE |
| RACGAP1 | 2.1447226 | 0.46368376 | 0.6894 | 5.035846761 | 193.9039028 | 193.9039028 | 0.0906384 | 0.878031753 | 1 | 0.090638399 | 0.090638399 | 1.792941882 | 23.66240584 | 1.374058899 | FALSE | TRUE | FALSE |
| ADD1 | 2.1445051 | 0.46385071 | 0.51355 | 2.14450506 | 2.14450506 | 42.26484135 | 2.5529481 | 0.443692577 | 0.0177 | 2.552948097 | 2.552948097 | 9.31023241 | 0.840011226 | -0.07571491 | FALSE | FALSE | FALSE |
| HDAC1 | 2.1424452 | 0.46401756 | 0.2549 | 0.53567758 | 2.142445218 | 4.536760539 | 0.16465716 | 0.86097282 | 0.8731 | 2.633436522 | 24.64562508 | 24.64562508 | 13.01155173 | 1.114329093 | FALSE | TRUE | FALSE |
| OR1M1 | 2.1376007 | 0.46418436 | 0.5089 | 2.137600687 | 2.137600687 | 2.137600687 | 0.28420182 | 0.829801404 | 1 | 0.284201824 | 0.284201824 | 0.284201824 | 7.521417903 | 0.876299719 | FALSE | TRUE | FALSE |
| CYP20A1 | 2.1373027 | 0.46435104 | 0.2636 | 0.534249174 | 2.137302679 | 4.524900041 | 0.67515464 | 0.677361261 | 0.75415 | 0.675154638 | 2.70103925 | 2.70103925 | 3.165649111 | 0.500462775 | FALSE | TRUE | FALSE |
| CACNG1 | 2.1348957 | 0.46451763 | 0.5097 | 2.134895662 | 8.538701648 | 8.538701648 | 0.28611365 | 0.829300241 | 0.00725 | 0.286113654 | 0.286113654 | 0.286113654 | 7.461704938 | 0.872838072 | FALSE | TRUE | FALSE |
| STK31 | 2.1328319 | 0.46468413 | 0.9395 | 195.571232 | 19752.39698 | 205544.396 | 31.8317636 | 0.088176905 | 0.7768 | 310.5513837 | 31760.25648 | 61204.62805 | 0.067003258 | -1.17390408 | FALSE | FALSE | FALSE |
| ZBTB8OS | 2.1261858 | 0.46485061 | 0.75465 | 4.999814456 | 19.99810835 | 19.99810835 | 0.59766958 | 0.713337653 | 1 | 0.597669582 | 0.597669582 | 0.597669582 | 3.557460214 | 0.551140052 | FALSE | TRUE | FALSE |
| PROK1 | 2.1254188 | 0.46501697 | 0.24895 | 2.12541878 | 20.31460316 | 20.31460316 | 0.28899516 | 0.828379793 | 1 | 0.288995161 | 0.288995161 | 0.288995161 | 7.354513396 | 0.866553943 | FALSE | TRUE | FALSE |
| CLDN20 | 2.1236036 | 0.46518323 | 0.1292 | 0.530838694 | 2.123603632 | 2.123603632 | 0.19105325 | 0.854104477 | 0.9421 | 1.716682273 | 27.46896859 | 27.46896859 | 11.11524489 | 1.045919036 | FALSE | TRUE | FALSE |
| AADAT | 2.1229094 | 0.46534938 | 0.503 | 2.12290941 | 2.12290941 | 4.523648476 | 0.27980339 | 0.831129315 | 1 | 0.279803387 | 0.279803387 | 0.279803387 | 7.587146931 | 0.880078495 | FALSE | TRUE | FALSE |
| METTL21A | 2.122906 | 0.46551541 | 0.5052 | 2.122905979 | 2.122905979 | 2.122905979 | 20.3138196 | 0.131069538 | 0.6616 | 196.1944046 | 19698.32855 | 204537.2279 | 0.104505505 | -0.980860831 | FALSE | FALSE | FALSE |
| ELL3 | 2.122007 | 0.46568132 | 0.0082 | 0.235345755 | 2.122006986 | 2.122006986 | 0.03970361 | 0.889006456 | 1 | 0.039703607 | 0.039703607 | 0.039703607 | 53.44620135 | 1.727916843 | FALSE | TRUE | FALSE |
| FRZB | 2.1184218 | 0.46584716 | 0.2569 | 0.529651303 | 2.118421836 | 2.626630776 | 0.61231476 | 0.706504023 | 0.9705 | 22.75174996 | 530.3460947 | 2121.284924 | 3.459694232 | 0.539037718 | FALSE | TRUE | FALSE |
| OR10T2 | 2.1178815 | 0.46601288 | 0.2555 | 2.117881499 | 19.91748397 | 19.91748397 | 0.29130987 | 0.827669166 | 1 | 0.291309867 | 0.291309867 | 0.291309867 | 7.270201731 | 0.861546462 | FALSE | TRUE | FALSE |
| LCP2 | 2.1170192 | 0.46617849 | 0.5085 | 2.117019173 | 2.117019173 | 4.46666863 | 0.27757588 | 0.83163732 | 0.0124 | 0.277575883 | 0.277575883 | 5.503649055 | 7.626812349 | 0.882343061 | FALSE | TRUE | FALSE |
| AVP | 2.1168215 | 0.46634398 | 0.7534 | 2.116821481 | 8.468502881 | 8.468502881 | 0.58355981 | 0.719192262 | 0.0031 | 0.583559812 | 0.583559812 | 0.583559812 | 3.627428479 | 0.559598858 | FALSE | TRUE | FALSE |
| NAT8 | 2.1150585 | 0.46650937 | 0.0072 | 2.11505852 | 2.11505852 | 2.11505852 | 0.28193656 | 0.830600657 | 0.00545 | 0.281936556 | 0.281936556 | 0.281936556 | 7.501895291 | 0.875170998 | FALSE | TRUE | FALSE |

| | | | | | | | | | | | | | | | | | |
|---|---|---|---|---|---|---|---|---|---|---|---|---|---|---|---|---|---|
| EEF2K | 2.1141522 | 0.46667465 | 0.9674 | 181.8346452 | 17227.81033 | 173641.149 | 34.1071162 | 0.082153834 | 0.3242 | 3.674693888 | 324.5034948 | 324.5034948 | 0.061985663 | -1.207708752 | FALSE | FALSE | FALSE |
| TMCO2 | 2.1131126 | 0.46683982 | 0.74705 | 2.11311258 | 19.81102783 | 19.81102783 | 0.29122474 | 0.827705753 | 1 | 0.291224738 | 0.291224738 | 0.291224738 | 7.25595152 | 0.860694372 | FALSE | TRUE | FALSE |
| ALDOC | 2.1028856 | 0.46700503 | 0.5076 | 2.102885555 | 8.411326502 | 33.65284807 | 0.19904588 | 0.852355157 | 0.69935 | 0.795841014 | 12.73204413 | 12.73204413 | 10.56482829 | 1.023862443 | FALSE | TRUE | FALSE |
| ERLEC1 | 2.102444 | 0.46717012 | 0.50405 | 2.102443977 | 2.102443977 | 4.533342924 | 0.28011336 | 0.831041424 | 1 | 0.280113361 | 0.280113361 | 5.557265708 | 7.505689737 | 0.875390608 | FALSE | TRUE | FALSE |
| RASGRF2 | 2.0936097 | 0.46733523 | 0.2714 | 0.523527978 | 2.09360968 | 40.7401961 | 0.08335493 | 0.87951305 | 1 | 0.083354933 | 0.083354933 | 1.627314939 | 25.11680612 | 1.399964413 | FALSE | TRUE | FALSE |
| TBC1D15 | 2.0895616 | 0.46750028 | 0.5055 | 2.089561551 | 2.089561551 | 7.661313564 | 0.26955607 | 0.833928927 | 1 | 2.493771772 | 2.493771772 | 9.118465861 | 7.75186222 | 0.889406045 | FALSE | TRUE | FALSE |
| SLC10A6 | 2.0892589 | 0.46766521 | 0.50055 | 2.089258851 | 2.089258851 | 2.089258851 | 9.85507341 | 0.200991979 | 0.2582 | 2.463863277 | 9.855073414 | 9.855073414 | 0.211998304 | -0.673667613 | FALSE | FALSE | FALSE |
| RPUSD4 | 2.0877869 | 0.46783003 | 0.7518 | 2.087786907 | 19.76131019 | 19.76131019 | 0.28645135 | 0.82921044 | 1 | 0.286451354 | 0.286451354 | 5.680967635 | 7.288451875 | 0.86263529 | FALSE | TRUE | FALSE |
| PHF14 | 2.0841965 | 0.46799479 | 0.75335 | 2.084196451 | 19.93466563 | 41.08799272 | 0.2840423 | 0.829854927 | 1 | 0.284042304 | 0.284042304 | 5.609264349 | 7.337626909 | 0.865555626 | FALSE | TRUE | FALSE |
| STX8 | 2.0836533 | 0.46815944 | 0.50195 | 2.083653285 | 2.083653285 | 2.083653285 | 0.26157428 | 0.83620891 | 1 | 2.407664491 | 22.74521043 | 22.74521043 | 7.965818613 | 0.901230413 | FALSE | TRUE | FALSE |
| RAPGEF5 | 2.0835474 | 0.46832396 | 0.88305 | 19.97522987 | 196.4187572 | 394.8583328 | 2.65749817 | 0.436852069 | 0.51645 | 2.657498175 | 2.657498175 | 9.58762296 | 0.784025913 | -0.105669583 | FALSE | FALSE | FALSE |
| SALL2 | 2.0783689 | 0.46848844 | 0.93655 | 180.0828563 | 17188.18965 | 58696.83564 | 0.4117733 | 0.787729742 | 0.75605 | 0.411773299 | 3.741732855 | 8.087729617 | 5.047361942 | 0.703064449 | FALSE | TRUE | FALSE |
| KCNIP4 | 2.0734949 | 0.46865287 | 0.50035 | 2.073494908 | 2.073494908 | 2.073494908 | 0.56655572 | 0.725999675 | 1 | 0.566555716 | 0.566555716 | 0.566555716 | 3.659825238 | 0.563460348 | FALSE | TRUE | FALSE |
| PRPF31 | 2.0733947 | 0.46881718 | 0.50245 | 2.073394677 | 8.293301783 | 10.24987637 | 0.65338242 | 0.688217595 | 0.93565 | 2.612943968 | 41.81970607 | 41.81970607 | 3.173324868 | 0.501514535 | FALSE | TRUE | FALSE |
| TOR1AIP2 | 2.0712008 | 0.4689814 | 0.50555 | 2.071200787 | 2.071200787 | 4.48297878 | 2.47181263 | 0.449060441 | 0.5119 | 2.47181263 | 2.47181263 | 5.333460646 | 0.837927909 | -0.076793344 | FALSE | FALSE | FALSE |
| PPP2CB | 2.0704205 | 0.46914551 | 0.5042 | 2.070420502 | 2.070420502 | 2.070420502 | 0.56589716 | 0.726190464 | 0.00805 | 0.56589716 | 0.56589716 | 0.56589716 | 3.658651516 | 0.563321045 | FALSE | TRUE | FALSE |
| PGA5 | 2.0659336 | 0.46930957 | 0.50595 | 2.065933632 | 2.065933632 | 4.396612325 | 0.273298 | 0.832851235 | 1 | 0.273298003 | 0.273298003 | 5.421440408 | 7.55927086 | 0.878479907 | FALSE | TRUE | FALSE |
| MYOZ3 | 2.0659106 | 0.4694735 | 0.74765 | 2.065910587 | 19.55953928 | 19.55953928 | 0.27508853 | 0.832281114 | 1 | 0.275088526 | 2.54843459 | 2.54843459 | 7.509984562 | 0.875639044 | FALSE | TRUE | FALSE |
| MCM9 | 2.0626291 | 0.46963737 | 0.8919 | 18.8076996 | 1683.399292 | 16505.14129 | 3.478254 | 0.384358678 | 0.49435 | 0.386278595 | 32.12518362 | 32.12518362 | 0.593007045 | -0.226940147 | FALSE | FALSE | FALSE |
| PLA2G1B | 2.0583501 | 0.46980117 | 0.0043 | 2.058350145 | 2.058350145 | 2.058350145 | 0.27428184 | 0.832471586 | 1 | 0.274281835 | 0.274281835 | 0.274281835 | 7.504507707 | 0.875322208 | FALSE | TRUE | FALSE |
| DHDDS | 2.0566924 | 0.46996489 | 0.50645 | 2.056692391 | 8.224926154 | 8.224926154 | 2.49108069 | 0.447153297 | 0.50125 | 2.491080692 | 2.491080692 | 2.491080692 | 0.825622549 | -0.083218454 | FALSE | FALSE | FALSE |
| ZBTB41 | 2.0564345 | 0.47012848 | 0.93825 | 19.56373306 | 1909.290676 | 1909.290676 | 2.70877668 | 0.434474731 | 0.5079 | 2.708776685 | 2.708776685 | 10.25387412 | 0.759174629 | -0.119658314 | FALSE | FALSE | FALSE |
| ARL16 | 2.053297 | 0.470292 | 0.50445 | 2.053297003 | 2.053297003 | 2.053297003 | 0.27275468 | 0.833057576 | 1 | 0.272754683 | 0.272754683 | 0.272754683 | 7.527999087 | 0.876679558 | FALSE | TRUE | FALSE |
| IL33 | 2.0499318 | 0.47045545 | 0.7555 | 2.049931795 | 19.21644417 | 19.21644417 | 0.2836185 | 0.830050885 | 1 | 0.283618496 | 0.283618496 | 0.283618496 | 7.227778945 | 0.859004862 | FALSE | TRUE | FALSE |
| C2orf80 | 2.0442432 | 0.47061886 | 0.49915 | 0.223888494 | 2.044243183 | 2.044243183 | 2.36567663 | 0.458953576 | 0.7444 | 2.365676633 | 22.24964428 | 22.24964428 | 0.86412621 | -0.063422822 | FALSE | FALSE | FALSE |
| PRAMEF2 | 2.042996 | 0.47078218 | 0.50605 | 19.22412987 | 185.5395533 | 185.5395533 | 23.8374398 | 0.114264704 | 0.25305 | 2.520789589 | 23.8374398 | 23.8374398 | 0.085705346 | -1.066992088 | FALSE | FALSE | FALSE |
| TMCC3 | 2.0406007 | 0.47094541 | 0.50675 | 2.040600734 | 2.040600734 | 4.370214597 | 0.26058887 | 0.836522488 | 1 | 2.415821244 | 9.662586711 | 9.662586711 | 7.830728672 | 0.893802176 | FALSE | TRUE | FALSE |
| PHTF2 | 2.0394194 | 0.47110853 | 0.5069 | 2.039419411 | 2.039419411 | 7.443970426 | 0.27134255 | 0.833451662 | 1 | 0.271342553 | 0.271342553 | 5.367485786 | 7.516032371 | 0.875988642 | FALSE | TRUE | FALSE |
| KCNV1 | 2.0368981 | 0.47127158 | 0.2639 | 0.509372877 | 2.036898109 | 10.07602538 | 0.16453012 | 0.860984851 | 1 | 0.657808773 | 0.657808773 | 3.258891896 | 12.38009268 | 1.092723896 | FALSE | TRUE | FALSE |
| MEOX1 | 2.0339766 | 0.47143455 | 0.503 | 2.03397661 | 19.07223321 | 19.07223321 | 0.16416425 | 0.861093042 | 1 | 1.541647258 | 6.164045395 | 6.164045395 | 12.38988752 | 1.093067364 | FALSE | TRUE | FALSE |
| THEMIS | 2.0318149 | 0.47159743 | 0.5052 | 2.031814889 | 19.0694926 | 19.0694926 | 0.31945019 | 0.818980337 | 0.5056 | 0.319450195 | 0.319450195 | 0.702856752 | 6.360349506 | 0.803480981 | FALSE | TRUE | FALSE |
| ISL2 | 2.028458 | 0.47176024 | 0.32305 | 2.028458036 | 32.46526763 | 40.09989641 | 160.428763 | 0.01650335 | 0.2346 | 40.08424824 | 642.2427051 | 2571.72514 | 0.01264398 | -1.898116208 | FALSE | FALSE | FALSE |
| APH1B | 2.0258012 | 0.47192297 | 0.50855 | 2.025801205 | 2.025801205 | 2.025801205 | 2.42422797 | 0.453208835 | 0.5071 | 2.424227968 | 2.424227968 | 2.424227968 | 0.83564798 | -0.077976632 | FALSE | FALSE | FALSE |
| PRRT2 | 2.0246249 | 0.4720856 | 0.75225 | 2.024624869 | 19.18793523 | 19.18793523 | 0.27856769 | 0.831374925 | 0.00645 | 0.278567688 | 0.278567688 | 0.278567688 | 7.267981743 | 0.861413828 | FALSE | TRUE | FALSE |
| ZC3HAV1 | 2.0237167 | 0.47224812 | 0.7617 | 2.02371668 | 19.25618473 | 68.35231729 | 0.26922478 | 0.834013867 | 0.5153 | 2.505226737 | 2.505226737 | 49.2372553 | 7.516829304 | 0.876034688 | FALSE | TRUE | FALSE |
| RABEP1 | 2.0217244 | 0.47241055 | 0.5127 | 2.021724406 | 2.021724406 | 39.81874236 | 2.42358337 | 0.453582997 | 0.51495 | 2.423583369 | 2.423583369 | 47.8210224 | 0.8341881 | -0.07873601 | FALSE | FALSE | FALSE |
| OR5D16 | 2.0207088 | 0.47257288 | 0.5033 | 2.020708788 | 2.020708788 | 2.020708788 | 0.27102856 | 0.833554158 | 1 | 0.271028563 | 0.271028563 | 0.271028563 | 7.455704175 | 0.872488668 | FALSE | TRUE | FALSE |
| SESN1 | 2.0198781 | 0.47273509 | 0.504 | 2.019878068 | 2.019878068 | 4.348202876 | 0.27034367 | 0.833673587 | 1 | 0.270343666 | 0.270343666 | 5.356994426 | 7.471519849 | 0.873408955 | FALSE | TRUE | FALSE |
| DKK4 | 2.0195714 | 0.47289719 | 0.2562 | 0.504858347 | 2.019571386 | 2.019571386 | 0.16243828 | 0.861488377 | 1 | 0.649651449 | 0.649651449 | 0.649651449 | 12.43285344 | 1.094570814 | FALSE | TRUE | FALSE |
| FAM136A | 2.0187136 | 0.47305918 | 0.87305 | 2.018713613 | 182.7914276 | 182.7914276 | 0.28485479 | 0.829640636 | 1 | 1.139366721 | 1.139366721 | 1.139366721 | 7.086816498 | 0.850451188 | FALSE | TRUE | FALSE |
| C22orf42 | 2.0136051 | 0.47322114 | 0.7503 | 2.013605126 | 18.871674 | 18.871674 | 22.6785434 | 0.119748018 | 0.00505 | 2.40895484 | 22.67854337 | 22.67854337 | 0.088788997 | -1.051640849 | FALSE | FALSE | FALSE |
| METTL18 | 2.0134445 | 0.47338297 | 0.5054 | 2.013444503 | 2.013444503 | 2.013444503 | 0.26962133 | 0.833894927 | 1 | 0.269621331 | 0.269621331 | 0.269621331 | 7.467675116 | 0.873185416 | FALSE | TRUE | FALSE |
| OR14I1 | 2.0116202 | 0.47354471 | 0.25835 | 2.011620214 | 18.87973142 | 18.87973142 | 0.27868027 | 0.831339877 | 1 | 0.278680267 | 0.278680267 | 0.278680267 | 7.218380543 | 0.858439774 | FALSE | TRUE | FALSE |
| CLEC4A | 2.0113544 | 0.47370633 | 0.49635 | 0.219813505 | 2.01135443 | 2.01135443 | 0.27013663 | 0.833741754 | 1 | 0.270136634 | 0.270136634 | 0.270136634 | 7.445692936 | 0.871905122 | FALSE | TRUE | FALSE |
| FSD1 | 2.0099652 | 0.47386786 | 0.50525 | 2.009965204 | 2.009965204 | 7.296374846 | 0.2662636 | 0.834808123 | 1 | 0.266263597 | 0.266263597 | 5.260692776 | 7.548779581 | 0.877876744 | FALSE | TRUE | FALSE |
| NIT2 | 2.0098172 | 0.47402927 | 0.63745 | 8.041054769 | 128.810529 | 515.742816 | 12.0450528 | 0.177915875 | 0.365 | 12.04505276 | 192.9750097 | 772.6982081 | 0.166858313 | -0.777652152 | FALSE | FALSE | FALSE |
| OR9A2 | 2.0087678 | 0.47419057 | 0.50545 | 2.008767753 | 2.008767753 | 2.008767753 | 0.2616592 | 0.836175832 | 1 | 0.261659197 | 2.405053633 | 2.405053633 | 7.677038597 | 0.885193724 | FALSE | TRUE | FALSE |
| KLRC3 | 2.0061387 | 0.47435179 | 0.754 | 2.006138651 | 18.82474934 | 18.82474934 | 0.27807923 | 0.831514969 | 0.0058 | 0.278079235 | 0.278079235 | 0.278079235 | 7.214269895 | 0.858192386 | FALSE | TRUE | FALSE |
| ZKSCAN8 | 2.0059958 | 0.4745129 | 0.01555 | 2.005995767 | 2.005995767 | 4.297795884 | 0.26782357 | 0.834386586 | 1 | 0.267823573 | 0.267823573 | 5.307163075 | 7.489989569 | 0.874481213 | FALSE | TRUE | FALSE |
| MAPK10 | 2.0027134 | 0.47467394 | 0.9848 | 188.4681846 | 19579.28811 | 207623.9809 | 235.191793 | 0.010727579 | 0.19585 | 235.1917926 | 2375.395362 | 24576.54481 | 0.008515235 | -2.069803358 | FALSE | FALSE | FALSE |
| ENC1 | 1.9980601 | 0.47483493 | 0.5094 | 1.998060102 | 1.998060102 | 7.214305054 | 0.26403551 | 0.835528152 | 0.01365 | 0.264035507 | 0.264035507 | 5.224581745 | 7.5673917 | 0.878946214 | FALSE | TRUE | FALSE |
| SLC7A6 | 1.9955601 | 0.47499584 | 0.50205 | 1.995560131 | 46.07551255 | 184.1623733 | 0.09658349 | 0.876805456 | 0.7488 | 0.224463407 | 0.89705884 | 0.89705884 | 20.66150331 | 1.315161917 | FALSE | TRUE | FALSE |
| VWA3A | 1.9948137 | 0.47515665 | 0.07865 | 0.124598972 | 1.994813688 | 2.443708008 | 0.02850194 | 0.891602959 | 0.94165 | 0.114029166 | 1.833139622 | 2.241623691 | 69.98868808 | 1.845027853 | FALSE | TRUE | FALSE |
| MYOZ1 | 1.9885381 | 0.47531743 | 0.13405 | 1.988538132 | 7.956243835 | 7.956243835 | 39.2914123 | 0.06902032 | 0.3432 | 9.817652703 | 157.2881815 | 629.8005058 | 0.050609994 | -1.295763716 | FALSE | FALSE | FALSE |
| STOML2 | 1.9863039 | 0.47547813 | 0.3238 | 0.496726662 | 7.94478981 | 7.94478981 | 0.17642037 | 0.858001605 | 0.9363 | 3.911775923 | 152.9575416 | 152.9575416 | 11.25892605 | 1.051496967 | FALSE | TRUE | FALSE |
| TRPT1 | 1.9862198 | 0.47563871 | 0.0076 | 1.986219811 | 1.986219811 | 1.986219811 | 0.26591352 | 0.834925773 | 0.00615 | 0.265913524 | 0.265913524 | 0.265913524 | 7.469420061 | 0.873286884 | FALSE | TRUE | FALSE |
| FRMPD2 | 1.9833769 | 0.47579922 | 0.2093 | 0.870002506 | 18.52836924 | 74.0668543 | 0.00596243 | 0.897887705 | 1 | 0.052238028 | 0.469742509 | 1.761845527 | 332.6455827 | 2.521981761 | FALSE | TRUE | FALSE |
| RNF6 | 1.9810931 | 0.47595965 | 0.51305 | 1.981093149 | 1.981093149 | 7.241325507 | 2.37988035 | 0.457666866 | 0.50845 | 2.379880355 | 2.379880355 | 8.678439872 | 0.832433927 | -0.079650228 | FALSE | FALSE | FALSE |
| MALT1 | 1.9804262 | 0.47611996 | 0.2597 | 1.980426191 | 18.72751268 | 39.11468125 | 0.27360005 | 0.832713384 | 1 | 0.273600055 | 0.273600055 | 5.411882557 | 7.23839837 | 0.859642481 | FALSE | TRUE | FALSE |
| SLC39A8 | 1.9792569 | 0.47628018 | 0.50575 | 1.979256902 | 1.979256902 | 1.979256902 | 0.54373577 | 0.736576404 | 1 | 0.543735773 | 0.543735773 | 0.543735773 | 3.640107934 | 0.561114261 | FALSE | TRUE | FALSE |
| TEX35 | 1.9771382 | 0.47644031 | 0.50645 | 1.977138158 | 1.977138158 | 1.977138158 | 0.26662563 | 0.834723983 | 1 | 0.266625633 | 0.266625633 | 0.266625633 | 7.415409157 | 0.870135119 | FALSE | TRUE | FALSE |
| UGT1A5 | 1.9750913 | 0.47660035 | 0.509 | 1.975091334 | 1.975091334 | 4.20463158 | 2.22837788 | 0.469572387 | 0.8803 | 21.26036227 | 208.0445031 | 208.0445031 | 0.88633591 | -0.052401655 | FALSE | FALSE | FALSE |
| ADCK2 | 1.9748801 | 0.47676028 | 0.8142 | 18.21092434 | 172.1391165 | 1666.959279 | 263.33482 | 0.009524205 | 0.12035 | 27.6315223 | 2571.007031 | 25700.61009 | 0.007499502 | -2.124967549 | FALSE | FALSE | FALSE |
| NUSAP1 | 1.9734708 | 0.47692011 | 0.5029 | 1.973470809 | 1.973470809 | 4.271897792 | 0.26540724 | 0.835177305 | 1 | 0.265407244 | 0.265407244 | 5.270489216 | 7.435632805 | 0.871317935 | FALSE | TRUE | FALSE |
| SFTPC | 1.969047 | 0.47707989 | 0.4997 | 0.211270723 | 1.969046955 | 1.969046955 | 0.26172615 | 0.836159287 | 1 | 0.261726153 | 0.261726153 | 0.261726153 | 7.523309888 | 0.876408951 | FALSE | TRUE | FALSE |
| MBL2 | 1.965657 | 0.47723961 | 0.49895 | 0.211946114 | 1.965656988 | 1.965656988 | 206.515971 | 0.012988775 | 0.12745 | 21.13326008 | 206.5159707 | 206.5159707 | 0.009518184 | -2.021445908 | FALSE | FALSE | FALSE |
| NCALD | 1.9619453 | 0.47739927 | 0.50775 | 1.96194527 | 1.96194527 | 1.96194527 | 0.53983448 | 0.738320778 | 1 | 0.539834482 | 0.539834482 | 0.539834482 | 3.634345965 | 0.560426267 | FALSE | TRUE | FALSE |
| ERRFI1 | 1.9605699 | 0.47755883 | 0.5042 | 1.960569934 | 1.960569934 | 4.209327876 | 0.26253945 | 0.835960505 | 1 | 0.262539452 | 0.262539452 | 5.209825995 | 7.4677155 | 0.873187764 | FALSE | TRUE | FALSE |
| WDR12 | 1.9597489 | 0.47771829 | 0.5039 | 1.95974889 | 1.95974889 | 4.19963764 | 0.25378057 | 0.838282697 | 1 | 2.358241035 | 2.358241035 | 5.043543058 | 7.722217908 | 0.887742053 | FALSE | TRUE | FALSE |
| KCNA6 | 1.9586629 | 0.47787765 | 0.96655 | 768.8424293 | 83166.33455 | 889247.8695 | 0.04308799 | 0.888311642 | 1 | 0.40421276 | 1.615598537 | 3.398435281 | 45.45728394 | 1.657603483 | FALSE | TRUE | FALSE |
| FAM117B | 1.9573924 | 0.4780369 | 0.50715 | 1.957392389 | 1.957392389 | 4.207907244 | 0.26207734 | 0.83612619 | 1 | 0.262077342 | 0.262077342 | 0.262077342 | 7.468758556 | 0.87324842 | FALSE | TRUE | FALSE |
| CD1C | 1.9543498 | 0.47819609 | 0.684 | 1.954349802 | 187.6841192 | 187.6841192 | 0.02980387 | 0.891313079 | 0.00905 | 0.02980387 | 0.02980387 | 0.02980387 | 65.57369307 | 1.816729644 | FALSE | TRUE | FALSE |
| RHOQ | 1.9520007 | 0.4783552 | 0.87965 | 7.809096179 | 31.24856784 | 31.24856784 | 2.17322399 | 0.473785565 | 0.49555 | 0.543354633 | 2.17322399 | 2.17322399 | 0.898205 | -0.046624532 | FALSE | FALSE | FALSE |

| | | | | | | | | | | | | | | | | | |
|---|---|---|---|---|---|---|---|---|---|---|---|---|---|---|---|---|---|
| CPXM1 | 1.9513528 | 0.4785142 | 0.85065 | 31.21608803 | 2671.435531 | 10688.79467 | 7.22352917 | 0.240133859 | 0.40875 | 7.223529173 | 259.59523 | 1039.054907 | 0.270138423 | -0.56841364 | FALSE | FALSE | FALSE |
| PA2G4 | 1.9504752 | 0.4786731 | 0.0112 | 1.950475231 | 1.950475231 | 9.677602492 | 0.53706337 | 0.739741483 | 1 | 0.537063366 | 0.537063366 | 0.537063366 | 3.631741342 | 0.56011491 | FALSE | TRUE | FALSE |
| CD9 | 1.9499816 | 0.47883189 | 0.50075 | 1.949981571 | 1.949981571 | 1.949981571 | 0.25770361 | 0.837293064 | 1 | 0.257703609 | 0.257703609 | 0.257703609 | 7.566760825 | 0.878910007 | FALSE | TRUE | FALSE |
| PLG | 1.947085 | 0.4789906 | 0.9113 | 31.1656223 | 499.3447703 | 1999.518694 | 4.07085275 | 0.352804274 | 0.4817 | 4.070852752 | 65.07497585 | 260.2802055 | 0.47829905 | -0.320300482 | FALSE | FALSE | FALSE |
| ZNF613 | 1.9425606 | 0.47914927 | 0.2598 | 0.216495241 | 1.942560591 | 4.282861952 | 0.04006311 | 0.888942888 | 0.01565 | 0.040063111 | 0.040063111 | 0.793592683 | 48.48751288 | 1.685629908 | FALSE | TRUE | FALSE |
| OR4A16 | 1.9425189 | 0.47930783 | 0.874 | 18.24945175 | 175.8513236 | 175.8513236 | 0.25401837 | 0.838185881 | 1 | 22.0295905 | 213.1727761 | 213.1727761 | 7.647159059 | 0.883500124 | FALSE | TRUE | FALSE |
| TNNI1 | 1.9420558 | 0.47946627 | 0.9854 | 31.09305627 | 498.1342893 | 1994.926946 | 0.55283245 | 0.732105483 | 1 | 2.211263934 | 8.84700333 | 8.84700333 | 3.512919307 | 0.545668174 | FALSE | TRUE | FALSE |
| EXD3 | 1.9413259 | 0.47962461 | 0.50945 | 4.358578555 | 160.4200821 | 1512.616306 | 0.10348996 | 0.875065656 | 0.9388 | 0.917507226 | 77.77609024 | 77.77609024 | 18.75859058 | 1.273200205 | FALSE | TRUE | FALSE |
| CNR2 | 1.9410125 | 0.47978284 | 0.75485 | 1.941012506 | 18.30639705 | 18.30639705 | 0.25315952 | 0.838588492 | 0.75665 | 2.335176485 | 22.10738769 | 22.10738769 | 7.667152006 | 0.884634073 | FALSE | TRUE | FALSE |
| ZSCAN2 | 1.9405273 | 0.47994095 | 0.51055 | 1.940527268 | 1.940527268 | 7.099112058 | 1.0277987 | 0.585544265 | 0.50385 | 1.027798696 | 1.027798696 | 5.081158079 | 1.888042158 | 0.276011687 | FALSE | FALSE | FALSE |
| TBCEL | 1.9404936 | 0.48009895 | 0.506 | 1.940493644 | 1.940493644 | 4.074368276 | 0.22376441 | 0.846435642 | 1 | 0.223764407 | 0.223764407 | 4.444116753 | 8.672038927 | 0.938121219 | FALSE | TRUE | FALSE |
| STK16 | 1.9402998 | 0.48025684 | 0.00235 | 0.09769765 | 0.09769765 | 0.09769765 | 0.12437889 | 0.87026085 | 1 | 1.143891598 | 1.143891598 | 1.143891598 | 15.59991286 | 1.193122172 | FALSE | TRUE | FALSE |
| AGPAT5 | 1.9361003 | 0.48041468 | 0.37865 | 0.838672059 | 7.741992732 | 7.741992732 | 0.07828927 | 0.880781277 | 0.00995 | 0.078289266 | 0.078289266 | 0.078289266 | 24.73008572 | 1.393225622 | FALSE | TRUE | FALSE |
| KCNJ14 | 1.9287877 | 0.48057252 | 0.199 | 0.281993154 | 4.501325522 | 17.99095286 | 6.82644111 | 0.24799761 | 0.02295 | 0.426892194 | 6.826441106 | 27.3083279 | 0.282546591 | -0.548909928 | FALSE | FALSE | FALSE |
| RHBDL2 | 1.9279009 | 0.48073025 | 0.50785 | 1.927900935 | 1.927900935 | 1.927900935 | 0.26056994 | 0.836571889 | 1 | 0.260569937 | 0.260569937 | 0.260569937 | 7.398784962 | 0.869160405 | FALSE | TRUE | FALSE |
| SLC33A1 | 1.927116 | 0.48088788 | 0.50695 | 1.927116047 | 1.927116047 | 6.998460931 | 0.24121179 | 0.841724349 | 1 | 2.252299542 | 21.58447463 | 21.58447463 | 7.989311149 | 0.902509335 | FALSE | TRUE | FALSE |
| SLC22A12 | 1.9263155 | 0.48104541 | 0.25935 | 0.841102071 | 7.7022373 | 7.7022373 | 0.03822115 | 0.889386472 | 0.7537 | 0.152831094 | 0.611263195 | 3.01867715 | 50.39920114 | 1.702423653 | FALSE | TRUE | FALSE |
| LSMEM2 | 1.9257267 | 0.48120284 | 0.5062 | 1.925726691 | 1.925726691 | 1.925726691 | 0.2606454 | 0.836506014 | 0.00505 | 0.260645396 | 0.260645396 | 0.260645396 | 7.388301208 | 0.868544593 | FALSE | TRUE | FALSE |
| KIR3DL3 | 1.9220087 | 0.4813602 | 1 | 7.39162E+15 | 2.37548E+20 | 4.73463E+22 | 245.492123 | 0.010112046 | 1 | 8.34031E+13 | 2.38606E+18 | 8.46347E+19 | 0.007829207 | -2.106282205 | FALSE | FALSE | FALSE |
| OSTC | 1.9212842 | 0.48151746 | 0.5023 | 1.921284162 | 1.921284162 | 1.921284162 | 0.52953074 | 0.74328563 | 0.00395 | 0.529530738 | 0.529530738 | 0.529530738 | 3.628276931 | 0.559700427 | FALSE | TRUE | FALSE |
| APOF | 1.9207509 | 0.48167461 | 0.4971 | 1.920750941 | 182.3399788 | 1842.634267 | 0.02842967 | 0.891625785 | 1 | 2.468671107 | 23.90359395 | 23.90359395 | 67.56148407 | 1.829699181 | FALSE | TRUE | FALSE |
| PHC2 | 1.9205266 | 0.48183166 | 0.02745 | 1.920526569 | 1.920526569 | 37.6654591 | 0.25686039 | 0.837456105 | 1 | 0.256860392 | 0.256860392 | 5.049325843 | 7.476927671 | 0.873723179 | FALSE | TRUE | FALSE |
| SYTL1 | 1.9169299 | 0.48198864 | 0.8145 | 17.69103574 | 167.3618031 | 1622.023006 | 26.9399325 | 0.102561587 | 0.348 | 26.93993255 | 2510.310316 | 25112.12632 | 0.071155706 | -1.147790267 | FALSE | FALSE | FALSE |
| SP7 | 1.912721 | 0.48214557 | 0.5058 | 1.912720998 | 1.912720998 | 9.476632619 | 2.08696551 | 0.481857546 | 0.5135 | 2.086965511 | 2.086965511 | 10.35124255 | 0.916508197 | -0.037863646 | FALSE | FALSE | FALSE |
| INHBC | 1.9125158 | 0.4823024 | 0.13415 | 1.912515815 | 7.652513056 | 7.652513056 | 0.15660395 | 0.862826091 | 1 | 0.626321208 | 0.626321208 | 0.626321208 | 12.21243633 | 1.086802313 | FALSE | TRUE | FALSE |
| OR9K2 | 1.9086252 | 0.48245917 | 0.87135 | 17.96838652 | 173.5033735 | 173.5033735 | 0.27392563 | 0.832609844 | 1 | 0.273925632 | 0.273925632 | 0.273925632 | 6.967676617 | 0.843087986 | FALSE | TRUE | FALSE |
| ANKS1A | 1.9069283 | 0.48261585 | 0.81555 | 7.629976286 | 70.84100879 | 345.5873143 | 215.638773 | 0.012001481 | 0.07195 | 5.647206934 | 215.6387732 | 244.5098357 | 0.008843161 | -2.053392488 | FALSE | FALSE | FALSE |
| COQ10A | 1.906126 | 0.48277243 | 0.50515 | 1.906125951 | 1.906125951 | 1.906125951 | 0.25682587 | 0.837488676 | 0.0077 | 0.256825871 | 0.256825871 | 0.256825871 | 7.421861153 | 0.870512825 | FALSE | TRUE | FALSE |
| LY6G6F | 1.9059232 | 0.48292889 | 0.4988 | 0.208635247 | 1.905923155 | 1.905923155 | 2.29535436 | 0.465657071 | 0.49565 | 0.250148619 | 2.295354356 | 2.295354356 | 0.830339398 | -0.080744355 | FALSE | FALSE | FALSE |
| SDC3 | 1.9032519 | 0.48308529 | 0.0131 | 1.903251891 | 1.903251891 | 9.422831497 | 0.52550785 | 0.744732382 | 1 | 0.525507846 | 0.525507846 | 10.4185132 | 3.621738297 | 0.558917065 | FALSE | TRUE | FALSE |
| RANBP2 | 1.9023455 | 0.48324159 | 0.24485 | 0.830990314 | 30.45039069 | 144.2715681 | 0.20796983 | 0.850218965 | 0.53925 | 0.207969834 | 0.431436228 | 6.918943002 | 9.147218307 | 0.961289044 | FALSE | TRUE | FALSE |
| NOXRED1 | 1.8962329 | 0.48339786 | 0.50755 | 1.896232892 | 1.896232892 | 1.896232892 | 0.25610972 | 0.837683829 | 1 | 0.256109722 | 0.256109722 | 0.256109722 | 7.403986385 | 0.869465611 | FALSE | TRUE | FALSE |
| FBXO17 | 1.89469 | 0.48355405 | 0.4975 | 0.207134774 | 1.894690043 | 1.894690043 | 2.28396392 | 0.466193695 | 0.50315 | 2.283963918 | 2.283963918 | 2.283963918 | 0.829562161 | -0.081151066 | FALSE | FALSE | FALSE |
| FNDC8 | 1.8919102 | 0.48371017 | 0.49535 | 0.206895209 | 1.89191018 | 1.89191018 | 0.25606601 | 0.83770007 | 1 | 0.256066005 | 0.256066005 | 0.256066005 | 7.388369182 | 0.868548588 | FALSE | TRUE | FALSE |
| UFL1 | 1.8918723 | 0.48386617 | 0.0071 | 0.056006367 | 0.056006367 | 1.103910463 | 0.67806474 | 0.676210708 | 0.5124 | 0.678064745 | 0.678064745 | 2.55970661 | 2.79010572 | 0.445620659 | FALSE | TRUE | FALSE |
| CERS2 | 1.8895969 | 0.4840221 | 0.5115 | 1.889596944 | 1.889596944 | 3.993342477 | 0.99363199 | 0.58911595 | 0.50665 | 0.993631986 | 0.993631986 | 4.929657354 | 1.901707041 | 0.279143614 | FALSE | TRUE | FALSE |
| CPSF6 | 1.8895321 | 0.48417791 | 0.0197 | 1.889532125 | 1.889532125 | 37.31379035 | 0.52218298 | 0.746081044 | 1 | 0.522182975 | 0.522182975 | 10.3275354 | 3.61852495 | 0.558531571 | FALSE | TRUE | FALSE |
| CHTOP | 1.888907 | 0.48433362 | 0.50345 | 1.888907012 | 1.888907012 | 3.975508787 | 0.24995379 | 0.839483169 | 1 | 0.249953789 | 0.249953789 | 0.249953789 | 7.557024903 | 0.878350853 | FALSE | TRUE | FALSE |
| SIX2 | 1.885893 | 0.48448926 | 0.89495 | 42.48000717 | 1675.387488 | 16935.89164 | 0.15846346 | 0.862449767 | 1 | 14.34636544 | 567.1206435 | 567.1206435 | 11.90112261 | 1.07558793 | FALSE | TRUE | FALSE |
| DOLK | 1.8845963 | 0.48464481 | 0.6758 | 4.346078229 | 164.5182925 | 658.1039901 | 0.04804282 | 0.887127812 | 1 | 0.767930212 | 3.071363206 | 3.799697378 | 39.22742484 | 1.593589799 | FALSE | TRUE | FALSE |
| APP | 1.8791569 | 0.48480033 | 0.0116 | 0.197202737 | 1.879156914 | 3.876064274 | 0.04105157 | 0.888671993 | 0.8733 | 0.164059329 | 0.655813896 | 3.231315612 | 45.77551757 | 1.660633263 | FALSE | TRUE | FALSE |
| GJB2 | 1.8775778 | 0.48495577 | 0.9662 | 481.7926157 | 66026.59325 | 264558.585 | 23.4952877 | 0.116083586 | 0.13325 | 2.652803841 | 213.4446094 | 853.7020474 | 0.079912953 | -1.097382823 | FALSE | FALSE | FALSE |
| COA1 | 1.8763608 | 0.48511111 | 0.0031 | 0.200930245 | 1.876360835 | 1.876360835 | 2.27335127 | 0.466729534 | 0.24925 | 0.242253586 | 2.273351267 | 2.273351267 | 0.825372155 | -0.083350187 | FALSE | FALSE | FALSE |
| OR10A2 | 1.8763276 | 0.48526633 | 0.9684 | 63.68204214 | 1668.369595 | 16849.99185 | 22.7487608 | 0.119443283 | 0.4989 | 2.410950552 | 220.1582746 | 220.1582746 | 0.082480429 | -1.083649089 | FALSE | FALSE | FALSE |
| PGLYRP1 | 1.8759997 | 0.48542145 | 0.00685 | 1.875999684 | 1.875999684 | 1.875999684 | 0.24547931 | 0.840540869 | 0.5075 | 2.266313692 | 2.266313692 | 2.266313692 | 7.642190679 | 0.883217869 | FALSE | TRUE | FALSE |
| JTB | 1.8753103 | 0.48557647 | 0.00535 | 1.875310302 | 1.875310302 | 1.875310302 | 0.21359138 | 0.848704816 | 0.004 | 0.213591378 | 0.213591378 | 0.213591378 | 8.779897032 | 0.943489423 | FALSE | TRUE | FALSE |
| PROSER1 | 1.8749 | 0.48573138 | 0.51365 | 1.874900023 | 1.874900023 | 36.86878907 | 0.25289529 | 0.838684815 | 1 | 0.252895293 | 0.252895293 | 4.983080812 | 7.413740299 | 0.870037369 | FALSE | TRUE | FALSE |
| CASD1 | 1.8747612 | 0.48588618 | 0.7507 | 1.87476125 | 17.8012958 | 37.01790101 | 0.26000972 | 0.836703479 | 1 | 0.26000972 | 0.26000972 | 5.141858889 | 7.210350644 | 0.857956385 | FALSE | TRUE | FALSE |
| FUT5 | 1.874243 | 0.48604087 | 0.5116 | 1.874243034 | 1.874243034 | 4.047893107 | 0.25371746 | 0.838347174 | 1 | 0.253717459 | 0.253717459 | 5.025504011 | 7.387126778 | 0.868475552 | FALSE | TRUE | FALSE |
| METTL2B | 1.8707495 | 0.48619551 | 0.94045 | 17.63419298 | 1689.637806 | 1689.637806 | 22.8299405 | 0.118528608 | 0.25095 | 2.408340573 | 22.82994054 | 22.82994054 | 0.081942809 | -1.08648915 | FALSE | FALSE | FALSE |
| MFSD3 | 1.8694092 | 0.48635005 | 0.5045 | 1.869409205 | 17.19449286 | 17.19449286 | 0.31175119 | 0.821431631 | 0.0064 | 0.311751188 | 0.311751188 | 0.311751188 | 5.996478212 | 0.77789626 | FALSE | TRUE | FALSE |
| KRT86 | 1.869385 | 0.48650449 | 0.13865 | 0.467457262 | 1.869385035 | 2.303166047 | 2.80443641 | 0.426827905 | 0.07895 | 0.701217287 | 11.21881758 | 13.83794965 | 0.66658136 | -0.176146835 | FALSE | FALSE | FALSE |
| ZNF334 | 1.8684352 | 0.48665882 | 0.6959 | 1.868435187 | 160.9986944 | 160.9986944 | 2.91316911 | 0.418351773 | 0.696 | 2.913169107 | 255.9244658 | 255.9244658 | 0.641375464 | -0.192887658 | FALSE | FALSE | FALSE |
| IFITM5 | 1.8675133 | 0.48681306 | 0.50385 | 1.867513334 | 1.867513334 | 1.867513334 | 20.7649508 | 0.128908169 | 0.25175 | 2.187338507 | 20.76495084 | 20.76495084 | 0.089935842 | -1.046067196 | FALSE | FALSE | FALSE |
| INPP4A | 1.867429 | 0.48696719 | 0.75455 | 1.867429006 | 17.75733018 | 63.03217546 | 0.25887208 | 0.836966026 | 1 | 0.258872077 | 0.258872077 | 5.077976227 | 7.213713529 | 0.858158892 | FALSE | TRUE | FALSE |
| MPPED1 | 1.8636841 | 0.48712127 | 0.7477 | 1.863684114 | 7.456064185 | 9.238249096 | 4.75870416 | 0.316745174 | 0.51005 | 4.758704164 | 4.758704164 | 4.758704164 | 0.391636893 | -0.407116403 | FALSE | FALSE | FALSE |
| PGLS | 1.8636832 | 0.48727523 | 0.50405 | 1.863683195 | 1.863683195 | 1.863683195 | 0.51568582 | 0.749011264 | 1 | 0.515685817 | 0.515685817 | 0.515685817 | 3.613989629 | 0.557986902 | FALSE | TRUE | FALSE |
| C1orf105 | 1.8636644 | 0.48742908 | 0.50445 | 1.863664438 | 1.863664438 | 1.863664438 | 0.24590144 | 0.840352419 | 0.5002 | 2.248331991 | 2.248331991 | 2.248331991 | 7.5789081 | 0.879606641 | FALSE | TRUE | FALSE |
| MRPL4 | 1.8620854 | 0.48758284 | 0.00825 | 0.206500985 | 1.862085368 | 1.862085368 | 0.03721797 | 0.889638502 | 1 | 0.330139056 | 0.330139056 | 0.330139056 | 50.03188796 | 1.699246891 | FALSE | TRUE | FALSE |
| HNRNPDL | 1.8606609 | 0.48773652 | 0.75125 | 1.860660901 | 17.84381086 | 17.84381086 | 0.25647782 | 0.837570046 | 1 | 0.256477817 | 0.256477817 | 5.093783946 | 7.254666018 | 0.860617424 | FALSE | TRUE | FALSE |
| TRIM35 | 1.8540202 | 0.48789019 | 0.51175 | 1.854020152 | 1.854020152 | 6.753515622 | 0.24871305 | 0.839800317 | 1 | 0.248713055 | 0.248713055 | 4.902604228 | 7.454454506 | 0.872415869 | FALSE | TRUE | FALSE |
| RMND1 | 1.8500222 | 0.48804381 | 0.2568 | 0.462457628 | 1.850022171 | 2.294093288 | 105.48715 | 0.027970596 | 0.596 | 175.7760687 | 16611.0989 | 66468.54094 | 0.017537891 | -1.756022625 | FALSE | FALSE | FALSE |
| ZFP57 | 1.8494118 | 0.48819733 | 0.51275 | 1.849411755 | 1.849411755 | 3.996950848 | 0.24308477 | 0.841103651 | 1 | 2.236854875 | 2.236854875 | 4.820152466 | 7.608093938 | 0.881275866 | FALSE | TRUE | FALSE |
| PADI4 | 1.849379 | 0.48835074 | 0.5108 | 1.849379 | 17.05107418 | 17.05107418 | 0.03940728 | 0.889085823 | 1 | 0.039407279 | 0.039407279 | 0.777656063 | 46.92988377 | 1.671449479 | FALSE | TRUE | FALSE |
| PIWIL4 | 1.8481263 | 0.48850405 | 0.69765 | 4.204931572 | 157.172179 | 157.172179 | 1.43282877 | 0.542493844 | 0.51055 | 1.432828768 | 1.432828768 | 5.291782896 | 1.289844483 | 0.11053735 | FALSE | FALSE | FALSE |
| RRAS | 1.8468153 | 0.48865728 | 0.8079 | 12.62729773 | 118.1749962 | 472.908036 | 3.82284028 | 0.365764799 | 0.81205 | 15.29482809 | 143.3859706 | 573.8172684 | 0.483100291 | -0.315962701 | FALSE | FALSE | FALSE |
| LACTB | 1.8447182 | 0.48881043 | 0.96965 | 167.8240953 | 1661.729323 | 16855.68837 | 0.2827011 | 0.830335087 | 1 | 0.2827011 | 0.2827011 | 5.612857527 | 6.525330804 | 0.814602533 | FALSE | TRUE | FALSE |
| EFEMP1 | 1.8433411 | 0.48896349 | 0.261 | 0.46097446 | 1.843341055 | 9.12312994 | 0.15236084 | 0.863978005 | 1 | 0.152360843 | 0.152360843 | 3.019159481 | 12.09852231 | 1.08273233 | FALSE | TRUE | FALSE |
| TDRKH | 1.8422235 | 0.48911646 | 0.7539 | 1.842223507 | 17.34679091 | 36.45374196 | 0.24206623 | 0.84139909 | 0.7476 | 2.229665391 | 21.07841712 | 21.07841712 | 7.610411124 | 0.881408119 | FALSE | TRUE | FALSE |
| CYB561 | 1.8409003 | 0.48926933 | 0.50195 | 1.840900326 | 1.840900326 | 1.840900326 | 0.51005636 | 0.751638607 | 1 | 0.510056362 | 0.510056362 | 0.510056362 | 3.609209615 | 0.557412106 | FALSE | TRUE | FALSE |
| OR4K14 | 1.8379405 | 0.48942215 | 0.87855 | 17.24837425 | 166.0261735 | 166.0261735 | 17.8295929 | 0.144458891 | 0.966 | 1692.951606 | 177043.1574 | 1874613.699 | 0.103083706 | -0.986809978 | FALSE | FALSE | FALSE |
| LMAN2 | 1.8370299 | 0.48957487 | 0.25975 | 0.459298307 | 1.837029863 | 9.079162071 | 0.15153662 | 0.864219962 | 1 | 0.151536616 | 0.151536616 | 2.999684642 | 12.12267976 | 1.083598633 | FALSE | TRUE | FALSE |
| ECHDC3 | 1.8365982 | 0.48972748 | 0.2521 | 0.459296612 | 1.836598178 | 1.836598178 | 9.8969755 | 0.199637448 | 0.2833 | 5.749696057 | 91.96479684 | 367.9364329 | 0.185571661 | -0.731488346 | FALSE | FALSE | FALSE |

| | | | | | | | | | | | | | | | | | |
|---|---|---|---|---|---|---|---|---|---|---|---|---|---|---|---|---|---|
| SAYSD1 | 1.8350639 | 0.48988001 | 0.752 | 1.835063922 | 17.17062576 | 17.17062576 | 194.58101 | 0.013901648 | 0.00345 | 20.19817119 | 194.5810099 | 194.5810099 | 0.009430848 | -2.025449256 | FALSE | FALSE | FALSE |
| SNRNP25 | 1.8348028 | 0.49003243 | 0.497 | 0.201322504 | 1.834802807 | 1.834802807 | 0.2497756 | 0.839546695 | 0.00475 | 0.249775598 | 0.249775598 | 0.249775598 | 7.345804876 | 0.866039388 | FALSE | TRUE | FALSE |
| MTRF1 | 1.8325086 | 0.49018478 | 0.25595 | 0.478692143 | 4.343756564 | 4.343756564 | 0.6734757 | 0.677966935 | 0.759 | 0.673475702 | 6.160726319 | 6.160726319 | 2.720972061 | 0.434724083 | FALSE | TRUE | FALSE |
| ARID4A | 1.83236 | 0.49033702 | 0.9976 | 10935.53865 | 2954982.061 | 31226857.23 | 0.28795755 | 0.828705796 | 1 | 18.44178593 | 172.6527213 | 847.4300873 | 6.363299092 | 0.803682337 | FALSE | TRUE | FALSE |
| CASP14 | 1.8323595 | 0.49048915 | 0.50235 | 1.832359467 | 1.832359467 | 1.832359467 | 2.21648253 | 0.470631588 | 0.50835 | 2.21648253 | 2.21648253 | 2.21648253 | 0.826697004 | -0.082653636 | FALSE | FALSE | FALSE |
| NT5M | 1.8302683 | 0.4906412 | 0.25635 | 1.830268298 | 17.17506303 | 17.17506303 | 8.70808898 | 0.214499333 | 0.00125 | 0.256451312 | 0.256451312 | 0.256451312 | 0.210180248 | -0.677408101 | FALSE | FALSE | FALSE |
| ZFAND5 | 1.828351 | 0.49079318 | 0.7547 | 4.269090019 | 17.07593685 | 17.07593685 | 0.52274624 | 0.745673946 | 1 | 0.522746235 | 0.522746235 | 0.522746235 | 3.49758817 | 0.543768671 | FALSE | TRUE | FALSE |
| RHBDD3 | 1.8262689 | 0.49094507 | 0.5074 | 1.826268905 | 1.826268905 | 1.826268905 | 0.23503921 | 0.843391721 | 0.00875 | 0.235039206 | 0.235039206 | 0.235039206 | 7.770060739 | 0.890424414 | FALSE | TRUE | FALSE |
| TAAR6 | 1.8241656 | 0.49109689 | 0.5036 | 1.82416555 | 7.298834231 | 7.298834231 | 2.37628387 | 0.457851047 | 0.25995 | 0.594026235 | 2.376283874 | 2.376283874 | 0.767654728 | -0.114834071 | FALSE | FALSE | FALSE |
| OR2Y1 | 1.8199345 | 0.49124867 | 0.2617 | 1.819934462 | 17.11412814 | 17.11412814 | 0.25487745 | 0.837943334 | 1 | 0.254877453 | 0.254877453 | 0.254877453 | 7.140429425 | 0.853724331 | FALSE | TRUE | FALSE |
| APIP | 1.8184618 | 0.49140037 | 0.5025 | 1.818461807 | 1.818461807 | 1.818461807 | 0.93808151 | 0.594484582 | 0.8729 | 3.753093664 | 34.45605725 | 34.45605725 | 1.938490193 | 0.287463608 | FALSE | TRUE | FALSE |
| SLC36A1 | 1.8174178 | 0.49155197 | 0.01 | 0.196703025 | 1.817417759 | 1.83817416 | 0.0298096 | 0.891305431 | 1 | 0.0298096 | 0.0298096 | 0.590717898 | 60.96753231 | 1.785098617 | FALSE | TRUE | FALSE |
| MFRP | 1.8167057 | 0.49170347 | 0.1344 | 1.81670569 | 16.96697107 | 16.96697107 | 0.31170737 | 0.821451283 | 0.88005 | 2.860093286 | 26.91727686 | 26.91727686 | 5.828241095 | 0.765537509 | FALSE | TRUE | FALSE |
| TAF1A | 1.8118014 | 0.49185494 | 0.511 | 1.811801356 | 1.811801356 | 3.932006222 | 0.24647218 | 0.840226532 | 1 | 0.246472181 | 0.246472181 | 0.246472181 | 7.350936526 | 0.866342673 | FALSE | TRUE | FALSE |
| MTX3 | 1.8096312 | 0.49200634 | 0.49915 | 0.198393246 | 1.809631248 | 1.809631248 | 2.19202686 | 0.472737468 | 0.49595 | 0.239207922 | 2.192026864 | 2.192026864 | 0.825551583 | -0.083255785 | FALSE | FALSE | FALSE |
| DTX2 | 1.809515 | 0.49215762 | 0.5129 | 1.809514979 | 1.809514979 | 35.6021874 | 19.6563303 | 0.134475445 | 0.5109 | 19.65633033 | 192.1496203 | 192.1496203 | 0.09205762 | -1.035940259 | FALSE | FALSE | FALSE |
| KCTD20 | 1.8078328 | 0.49230883 | 0.50655 | 1.807832756 | 16.80726516 | 16.80726516 | 0.07606975 | 0.881231444 | 1 | 0.076069754 | 0.076069754 | 0.076069754 | 23.76546093 | 1.375946242 | FALSE | TRUE | FALSE |
| SSNA1 | 1.8073906 | 0.49245994 | 0.2495 | 1.807390615 | 17.10025524 | 17.10025524 | 20.8265988 | 0.128600226 | 0.2475 | 2.193712588 | 20.82659883 | 20.82659883 | 0.086782803 | -1.061566329 | FALSE | FALSE | FALSE |
| FAM50B | 1.8058746 | 0.49261096 | 0.5123 | 1.805874645 | 1.805874645 | 3.823597311 | 0.24236889 | 0.841321446 | 1 | 0.242368889 | 0.242368889 | 4.795042877 | 7.450934211 | 0.872210729 | FALSE | TRUE | FALSE |
| AQP3 | 1.8051757 | 0.49276188 | 0.00945 | 1.805175683 | 1.805175683 | 1.805175683 | 0.24322733 | 0.841056902 | 1 | 0.243227334 | 0.243227334 | 0.243227334 | 7.421763234 | 0.870507096 | FALSE | TRUE | FALSE |
| SLC16A12 | 1.8048905 | 0.4929127 | 0.50475 | 1.804890473 | 1.804890473 | 3.85858183 | 0.24336876 | 0.840994527 | 0.0106 | 0.243368764 | 0.243368764 | 4.828963228 | 7.416278257 | 0.870186016 | FALSE | TRUE | FALSE |
| TRIM17 | 1.8044224 | 0.49306342 | 0.50365 | 1.804422402 | 16.56172926 | 16.56172926 | 0.33223363 | 0.814739447 | 0.0114 | 0.332233626 | 0.332233626 | 0.746344941 | 5.431185356 | 0.734894625 | FALSE | TRUE | FALSE |
| LRP2BP | 1.8034925 | 0.49321404 | 0.2516 | 1.80349252 | 17.0013822 | 17.0013822 | 0.25244664 | 0.838797046 | 1 | 0.252446642 | 0.252446642 | 0.252446642 | 7.144054738 | 0.853944747 | FALSE | TRUE | FALSE |
| DTD1 | 1.8024407 | 0.49336458 | 0.8773 | 7.210388206 | 28.85126334 | 28.85126334 | 7.83642883 | 0.231536851 | 0.32015 | 7.836428827 | 125.5025533 | 125.5025533 | 0.230007925 | -0.6382572 | FALSE | FALSE | FALSE |
| ARGFX | 1.8024105 | 0.49351501 | 0.50345 | 1.802410533 | 1.802410533 | 1.802410533 | 0.2366542 | 0.842966695 | 1 | 0.236654196 | 2.18854249 | 2.18854249 | 7.616220463 | 0.881739507 | FALSE | TRUE | FALSE |
| TEKT3 | 1.8013579 | 0.49366534 | 0.01145 | 0.203687257 | 1.801357938 | 4.026260505 | 0.03712304 | 0.889654219 | 0.50735 | 0.323627222 | 0.323627222 | 0.734835615 | 48.52398507 | 1.68595646 | FALSE | TRUE | FALSE |
| GLYATL2 | 1.7984955 | 0.49381561 | 0.50735 | 1.798495481 | 1.798495481 | 1.798495481 | 2.1794317 | 0.473087307 | 0.5059 | 2.179431695 | 2.179431695 | 2.179431695 | 0.825213052 | -0.083433912 | FALSE | FALSE | FALSE |
| RBM15 | 1.7938241 | 0.49396585 | 0.5221 | 1.793824146 | 1.793824146 | 34.89253765 | 0.24176725 | 0.841554145 | 1 | 0.241767248 | 0.241767248 | 4.718278294 | 7.419632553 | 0.870382398 | FALSE | TRUE | FALSE |
| WDR73 | 1.7929048 | 0.494116 | 0.811 | 1.792904777 | 164.144548 | 1629.009354 | 202.457174 | 0.013102005 | 0.23045 | 202.4571737 | 20604.56256 | 40116.32469 | 0.008855724 | -2.052775945 | FALSE | FALSE | FALSE |
| GDAP2 | 1.7925478 | 0.49426604 | 0.26095 | 1.792547785 | 16.88709446 | 35.5383977 | 186.79834 | 0.01449358 | 0.07075 | 19.29117297 | 1854.058798 | 1854.058798 | 0.009596166 | -2.01790227 | FALSE | FALSE | FALSE |
| SCAMP3 | 1.7920782 | 0.49441599 | 0.87425 | 16.76229057 | 160.9154109 | 160.9154109 | 0.04274344 | 0.888359808 | 0.0088 | 0.042743442 | 0.042743442 | 0.042743442 | 41.92638845 | 1.622487454 | FALSE | TRUE | FALSE |
| KCNK4 | 1.7916569 | 0.49456583 | 0.507 | 1.791656872 | 1.791656872 | 3.859635645 | 0.243195 | 0.841088071 | 0.01035 | 0.243194996 | 0.243194996 | 4.822556117 | 7.367161739 | 0.867300204 | FALSE | TRUE | FALSE |
| HOXA10 | 1.7914925 | 0.49471557 | 0.50545 | 1.791492524 | 7.163422261 | 7.163422261 | 0.24190831 | 0.841476657 | 1 | 0.241908306 | 0.241908306 | 4.804293976 | 7.405667679 | 0.86956422 | FALSE | TRUE | FALSE |
| PDE4C | 1.7889169 | 0.49486524 | 0.50855 | 1.788916932 | 28.62651238 | 114.5566398 | 0.10475028 | 0.874789336 | 0.514 | 0.418915195 | 1.675735846 | 8.262703002 | 17.077921 | 1.232435 | FALSE | TRUE | FALSE |
| NUDCD3 | 1.7861259 | 0.49501485 | 0.875 | 16.72768338 | 160.7854872 | 160.7854872 | 2.02652533 | 0.487883462 | 0.50455 | 4.756968061 | 19.0275389 | 19.0275389 | 0.881373595 | -0.054839965 | FALSE | FALSE | FALSE |
| ANKK1 | 1.7814156 | 0.49516443 | 0.9532 | 265.7709252 | 40685.15725 | 162851.4804 | 27.0649892 | 0.102264243 | 0.577 | 46.0895754 | 4168.310663 | 20575.91077 | 0.065819928 | -1.181642599 | FALSE | FALSE | FALSE |
| TAS2R39 | 1.7808444 | 0.49531391 | 0.7527 | 1.780844408 | 16.67928425 | 16.67928425 | 0.25066638 | 0.839164774 | 1 | 0.25066638 | 0.25066638 | 0.25066638 | 7.104440616 | 0.851529888 | FALSE | TRUE | FALSE |
| KCNK6 | 1.780053 | 0.49546329 | 0.98405 | 161.3900434 | 16154.41488 | 167484.1039 | 0.28280865 | 0.830299613 | 0.00785 | 0.282808649 | 0.282808649 | 0.282808649 | 6.294195909 | 0.798940256 | FALSE | TRUE | FALSE |
| IGFBP1 | 1.7797098 | 0.49561258 | 0.9694 | 164.0242367 | 1634.659171 | 16688.7554 | 21.9789309 | 0.123712258 | 0.13065 | 21.97893092 | 214.7636454 | 214.7636454 | 0.080973446 | -1.091657378 | FALSE | FALSE | FALSE |
| TRADD | 1.7777634 | 0.49576178 | 0.50015 | 1.777763387 | 1.777763387 | 1.777763387 | 0.49484331 | 0.756984487 | 1 | 0.494843309 | 0.494843309 | 0.494843309 | 3.592578404 | 0.555406255 | FALSE | TRUE | FALSE |
| KDM4C | 1.774333 | 0.49591094 | 0.782 | 16.78814959 | 1617.783209 | 16457.55552 | 0.01148418 | 0.896253883 | 1 | 0.104321994 | 0.416964611 | 2.049439751 | 154.5024248 | 2.1889353 | FALSE | TRUE | FALSE |
| DHX9 | 1.7730876 | 0.49606001 | 0.52215 | 1.773087623 | 1.773087623 | 34.62015647 | 2.16131615 | 0.475002979 | 0.02985 | 2.161316147 | 2.161316147 | 42.32112578 | 0.820374023 | -0.0859881 | FALSE | FALSE | FALSE |
| NGEF | 1.7718542 | 0.49620899 | 0.50935 | 1.771854245 | 1.771854245 | 34.79835119 | 0.23102423 | 0.844593541 | 1 | 2.159899875 | 2.159899875 | 42.51143779 | 7.669560294 | 0.884770466 | FALSE | TRUE | FALSE |
| SOD3 | 1.7693363 | 0.4963579 | 0.4978 | 0.188217318 | 1.769336273 | 1.769336273 | 0.23726543 | 0.842799109 | 1 | 0.23726543 | 0.23726543 | 0.23726543 | 7.457202153 | 0.872575916 | FALSE | TRUE | FALSE |
| MTO1 | 1.7679658 | 0.49650674 | 0.63045 | 1.767965808 | 113.333938 | 453.8079841 | 2.649309 | 0.437246849 | 0.19395 | 0.50818317 | 10.58860846 | 42.33053196 | 0.667330919 | -0.175658753 | FALSE | FALSE | FALSE |
| MED31 | 1.7676747 | 0.49665547 | 0.74905 | 4.131769639 | 16.52617397 | 16.52617397 | 0.5074716 | 0.752911488 | 1 | 0.507471603 | 0.507471603 | 0.507471603 | 3.483297854 | 0.541990612 | FALSE | TRUE | FALSE |
| TRUB1 | 1.7673849 | 0.49680409 | 0.51025 | 1.767384863 | 1.767384863 | 1.767384863 | 0.24075022 | 0.8418016 | 1 | 0.240750219 | 0.240750219 | 0.240750219 | 7.341155793 | 0.865764441 | FALSE | TRUE | FALSE |
| DCN | 1.7667321 | 0.49695263 | 0.01 | 1.766732067 | 1.766732067 | 1.786737465 | 0.94099074 | 0.593942382 | 0.0084 | 0.940990738 | 0.940990738 | 0.940990738 | 1.877523333 | 0.273585343 | FALSE | TRUE | FALSE |
| ABCC5 | 1.7647509 | 0.49710109 | 0.46945 | 0.441571741 | 159.353946 | 637.0205579 | 2.33349125 | 0.461872065 | 0.2002 | 0.479213524 | 17.62427627 | 70.48444082 | 0.756270639 | -0.12132276 | FALSE | FALSE | FALSE |
| ST6GALNAC | 1.7632814 | 0.49724946 | 0.5048 | 1.763281444 | 1.763281444 | 1.763281444 | 0.49079583 | 0.758531069 | 1 | 0.490795828 | 0.490795828 | 0.490795828 | 3.592698521 | 0.555420775 | FALSE | TRUE | FALSE |
| DEPDC4 | 1.7626962 | 0.49739774 | 0.0083 | 1.762696216 | 1.762696216 | 1.762696216 | 18.8478139 | 0.13914873 | 0.5054 | 18.84781391 | 181.2322825 | 181.2322825 | 0.093522582 | -1.029083513 | FALSE | FALSE | FALSE |
| CLRN3 | 1.7623363 | 0.49754592 | 0.5047 | 1.762336314 | 7.048160928 | 7.048160928 | 0.24174128 | 0.841585118 | 0.0062 | 0.241741279 | 0.241741279 | 0.241741279 | 7.290175366 | 0.862737975 | FALSE | TRUE | FALSE |
| CELF3 | 1.7607883 | 0.49769402 | 0.5042 | 1.76078827 | 1.76078827 | 8.724563155 | 0.49064185 | 0.758604204 | 1 | 0.49064185 | 0.49064185 | 9.73413327 | 3.588744559 | 0.554942547 | FALSE | TRUE | FALSE |
| CUZD1 | 1.7603834 | 0.49784202 | 0.2594 | 0.440264997 | 1.760383356 | 3.83176065 | 0.28437569 | 0.829747847 | 0.5074 | 0.284375687 | 0.284375687 | 1.409082263 | 6.1903441 | 0.791714791 | FALSE | TRUE | FALSE |
| PPRC1 | 1.7601257 | 0.49798992 | 0.048 | 1.760125731 | 1.760125731 | 34.07983541 | 0.23679729 | 0.842905795 | 1 | 0.236797287 | 0.236797287 | 7.960490971 | 7.433048551 | 0.87116697 | FALSE | TRUE | FALSE |
| CYGB | 1.7555329 | 0.49813779 | 0.81755 | 7.018946695 | 272.3443053 | 272.3443053 | 3.58503549 | 0.379216623 | 0.81825 | 20.58603209 | 329.1928532 | 1316.897849 | 0.48968354 | -0.310084494 | FALSE | FALSE | FALSE |
| CLDN23 | 1.7516573 | 0.49828561 | 0.26095 | 0.438134338 | 1.751657265 | 1.751657265 | 0.07143752 | 0.882131601 | 0.0079 | 0.071437523 | 0.071437523 | 0.071437523 | 24.5201288 | 1.389522747 | FALSE | TRUE | FALSE |
| RBFA | 1.7501699 | 0.49843335 | 0.5033 | 1.750169884 | 1.750169884 | 1.750169884 | 2.12834321 | 0.478622128 | 0.50605 | 2.128343214 | 2.128343214 | 2.128343214 | 0.822315627 | -0.084961457 | FALSE | FALSE | FALSE |
| TBCE | 1.7496143 | 0.498581 | 0.26015 | 0.196537432 | 1.749614321 | 3.890287604 | 0.31447454 | 0.820502982 | 0.01245 | 0.314474542 | 0.314474542 | 0.710723012 | 5.563611956 | 0.745356832 | FALSE | TRUE | FALSE |
| YIPF7 | 1.749494 | 0.49872854 | 0.7478 | 1.749494009 | 16.44817307 | 16.44817307 | 0.2387377 | 0.842416915 | 1 | 2.197952835 | 2.197952835 | 2.197952835 | 7.328101062 | 0.86499145 | FALSE | TRUE | FALSE |
| KCNG2 | 1.7489972 | 0.49887599 | 0.26055 | 0.83087093 | 13.27853548 | 13.27853548 | 0.14725783 | 0.86524772 | 0.0185 | 0.147257825 | 0.147257825 | 1.249555374 | 11.87710894 | 1.07471074 | FALSE | TRUE | FALSE |
| REXO4 | 1.7489175 | 0.49902333 | 0.69095 | 1.748917524 | 152.7961223 | 152.7961223 | 0.03745071 | 0.889591326 | 0.5083 | 0.332221668 | 0.332221668 | 0.742226588 | 46.69917594 | 1.669309217 | FALSE | TRUE | FALSE |
| PPHLN1 | 1.7488975 | 0.49917057 | 0.01305 | 1.748897475 | 1.748897475 | 3.787605001 | 2.06273284 | 0.484216589 | 0.7531 | 2.06273284 | 19.43666594 | 19.43666594 | 0.847854575 | -0.071678632 | FALSE | FALSE | FALSE |
| TMEM59 | 1.748495 | 0.49931771 | 0.5054 | 1.748494993 | 1.748494993 | 1.748494993 | 0.23881196 | 0.842370931 | 1 | 0.238811962 | 0.238811962 | 0.238811962 | 7.321639075 | 0.864608316 | FALSE | TRUE | FALSE |
| STK19 | 1.7434715 | 0.49946483 | 0.5105 | 1.743471533 | 1.743471533 | 3.771697127 | 0.23838389 | 0.842478189 | 1 | 0.238383895 | 0.238383895 | 4.720743738 | 7.313713609 | 0.86413795 | FALSE | TRUE | FALSE |
| ALLC | 1.7414798 | 0.49961187 | 0.25625 | 0.191272193 | 1.741479845 | 1.741479845 | 22.7948982 | 0.118833851 | 0.6601 | 22.79489821 | 2158.072159 | 21760.39829 | 0.07639779 | -1.116919205 | FALSE | FALSE | FALSE |
| OR2T4 | 1.7409919 | 0.49975882 | 0.25765 | 1.74099187 | 16.29154288 | 16.29154288 | 0.24583638 | 0.840399574 | 1 | 0.245836379 | 0.245836379 | 0.245836379 | 7.081913092 | 0.850150593 | FALSE | TRUE | FALSE |
| AIRE | 1.7389016 | 0.4999057 | 0.79345 | 16.4899734 | 1302.623429 | 6379.925724 | 290.48186 | 0.008517306 | 0.3638 | 29.75728542 | 7442.211992 | 29742.32319 | 0.005986266 | -2.222844015 | FALSE | FALSE | FALSE |
| OR10H3 | 1.7386003 | 0.50005247 | 0.96835 | 155.8634836 | 1532.021834 | 15426.547 | 2.39932302 | 0.455818203 | 0.5045 | 2.399323025 | 2.399323025 | 2.399323025 | 0.724621204 | -0.139888962 | FALSE | FALSE | FALSE |
| USP38 | 1.7361571 | 0.50019919 | 0.78815 | 6.945251111 | 151.4736076 | 605.9487507 | 2.67135449 | 0.436061355 | 0.26965 | 0.667930596 | 2.671354487 | 13.13680236 | 0.649916413 | -0.187142495 | FALSE | FALSE | FALSE |
| OR6C6 | 1.7356744 | 0.5003458 | 0.9961 | 1547.374713 | 161885.1228 | 1714473.306 | 2.39918403 | 0.45600356 | 0.93705 | 22.69613164 | 2190.169169 | 2190.169169 | 0.723443649 | -0.140595291 | FALSE | FALSE | FALSE |
| HSPBAP1 | 1.7355246 | 0.50049232 | 0.5072 | 1.735524638 | 1.735524638 | 3.75349212 | 0.23701769 | 0.842860089 | 0.0121 | 0.237017685 | 0.237017685 | 4.702576703 | 7.322342362 | 0.864650031 | FALSE | TRUE | FALSE |
| ITGB3BP | 1.7344448 | 0.50063875 | 0.2591 | 1.734444766 | 16.16116256 | 16.16116256 | 17.3722129 | 0.146352131 | 0.89115 | 166.293311 | 16401.22413 | 168792.5817 | 0.099840174 | -1.00069467 | FALSE | FALSE | FALSE |

| | | | | | | | | | | | | | | | | | |
|---|---|---|---|---|---|---|---|---|---|---|---|---|---|---|---|---|---|
| ZNF257 | 1.7339546 | 0.50078508 | 1 | 1.86972E+15 | 5.34804E+19 | 1.00344E+22 | 2.63172103 | 0.438034996 | 1 | 2.46282E+15 | 1.00309E+21 | 1.98302E+23 | 0.658867191 | -0.181202118 | FALSE | FALSE | FALSE |
| FAM118A | 1.7312322 | 0.50093135 | 0.5083 | 1.731232248 | 1.731232248 | 3.758120187 | 0.23702119 | 0.842844848 | 1 | 0.237021192 | 0.237021192 | 4.703666887 | 7.304124288 | 0.863568155 | FALSE | TRUE | FALSE |
| STAP1 | 1.7304692 | 0.50107753 | 0.5034 | 1.730469234 | 1.730469234 | 1.730469234 | 0.23583466 | 0.84310355 | 1 | 0.235834662 | 0.235834662 | 0.235834662 | 7.337637406 | 0.865556247 | FALSE | TRUE | FALSE |
| HDGFL1 | 1.7302153 | 0.50122362 | 0.00445 | 1.730215263 | 1.730215263 | 1.730215263 | 4.3648247 | 0.336411993 | 0.49745 | 0.467658674 | 4.364824704 | 4.364824704 | 0.396399714 | -0.401866668 | FALSE | FALSE | FALSE |
| TDRD1 | 1.7296965 | 0.5013696 | 0.5107 | 1.729696468 | 1.729696468 | 33.9124191 | 0.23509449 | 0.843361439 | 0.0245 | 0.235094494 | 0.235094494 | 4.620196762 | 7.357452066 | 0.866727441 | FALSE | TRUE | FALSE |
| CMC1 | 1.7296233 | 0.50151549 | 0.4992 | 0.432419171 | 1.729623275 | 1.729623275 | 0.48226768 | 0.762237808 | 1 | 0.482267684 | 0.482267684 | 0.482267684 | 3.586438266 | 0.55466336 | FALSE | TRUE | FALSE |
| OSBP | 1.7269677 | 0.50166132 | 0.2655 | 1.726967678 | 16.57008831 | 34.03857766 | 0.24008323 | 0.842079074 | 0.01915 | 0.240083228 | 0.240083228 | 4.740303965 | 7.193204178 | 0.856922388 | FALSE | TRUE | FALSE |
| STXBP5 | 1.7245901 | 0.50180708 | 0.5244 | 1.7245901 | 16.10365898 | 78.84257894 | 0.14638938 | 0.865417588 | 1 | 0.585401787 | 0.585401787 | 11.49254486 | 11.78084196 | 1.07117633 | FALSE | TRUE | FALSE |
| OR5T2 | 1.7244843 | 0.50195273 | 0.8771 | 16.13723486 | 154.8858834 | 154.8858834 | 0.23645565 | 0.843027547 | 1 | 2.167883898 | 20.399374 | 20.399374 | 7.293056025 | 0.86290955 | FALSE | TRUE | FALSE |
| CTRC | 1.7242393 | 0.5020983 | 0.31365 | 0.431156481 | 6.89713378 | 6.89713378 | 1.5209792 | 0.53400794 | 0.2536 | 0.380483533 | 1.520979197 | 1.520979197 | 1.133637638 | 0.054474257 | FALSE | FALSE | FALSE |
| GMPPA | 1.7242239 | 0.50224375 | 0.0129 | 0.431101656 | 1.724223888 | 2.134260271 | 0.15973225 | 0.86211888 | 0.9374 | 1.460394222 | 23.35885463 | 23.35885463 | 10.79446325 | 1.033201052 | FALSE | TRUE | FALSE |
| SNAPC2 | 1.7223444 | 0.50238914 | 0.0101 | 1.722344391 | 1.722344391 | 3.748628145 | 19.0949902 | 0.137277776 | 0.00735 | 2.033589911 | 19.09499025 | 19.09499025 | 0.090198757 | -1.044799446 | FALSE | FALSE | FALSE |
| ARHGEF2 | 1.7221643 | 0.50253443 | 0.7583 | 1.722164254 | 16.54967164 | 58.01095616 | 1.97578805 | 0.491339591 | 0.69605 | 18.98403247 | 1889.80264 | 3659.433956 | 0.87163411 | -0.059665783 | FALSE | FALSE | FALSE |
| WDR41 | 1.7200368 | 0.50267964 | 0.50285 | 3.076804118 | 63.10433579 | 252.4349602 | 11.1734631 | 0.185719395 | 0.193 | 1.236292823 | 44.69792113 | 44.69792113 | 0.153939451 | -0.812650068 | FALSE | FALSE | FALSE |
| HLA-G | 1.7193345 | 0.50282477 | 0.75095 | 1.719334512 | 16.12143051 | 16.12143051 | 0.24289223 | 0.841197064 | 0.0118 | 0.242892228 | 0.242892228 | 4.814375353 | 7.078590052 | 0.849946761 | FALSE | TRUE | FALSE |
| LCE1F | 1.7182178 | 0.50296982 | 0.50245 | 1.718217781 | 1.718217781 | 1.718217781 | 2.09156865 | 0.480839934 | 0.50305 | 2.091568648 | 2.091568648 | 2.091568648 | 0.821497197 | -0.085393914 | FALSE | FALSE | FALSE |
| STUB1 | 1.7143998 | 0.50311482 | 0.756 | 1.714399816 | 6.859193736 | 8.502611877 | 0.48431066 | 0.761456172 | 1 | 0.484310656 | 0.484310656 | 9.617010007 | 3.539876303 | 0.548988086 | FALSE | TRUE | FALSE |
| FGF12 | 1.7110396 | 0.50325977 | 0.50715 | 1.711039585 | 1.711039585 | 1.711039585 | 1.89212477 | 0.499870234 | 0.51015 | 1.892124766 | 1.892124766 | 1.892124766 | 0.904295327 | -0.043689713 | FALSE | FALSE | FALSE |
| SDF4 | 1.7098043 | 0.50340464 | 0.01185 | 1.709804262 | 1.709804262 | 3.676882453 | 0.23319523 | 0.843994971 | 1 | 0.233195233 | 0.233195233 | 4.621765189 | 7.33207209 | 0.865226726 | FALSE | TRUE | FALSE |
| KLHL18 | 1.7090797 | 0.50354941 | 0.51095 | 1.709079723 | 15.87239622 | 63.48626123 | 0.04327885 | 0.888255399 | 1 | 0.172994246 | 0.691665574 | 3.414505626 | 39.48995169 | 1.596486602 | FALSE | TRUE | FALSE |
| OR5P2 | 1.7089324 | 0.50369409 | 0.5063 | 1.708932418 | 1.708932418 | 1.708932418 | 0.23338757 | 0.843889743 | 1 | 0.233387568 | 0.233387568 | 0.233387568 | 7.322294121 | 0.86464717 | FALSE | TRUE | FALSE |
| CLDN16 | 1.7020043 | 0.50383878 | 0.0089 | 0.425603829 | 1.702004299 | 1.702004299 | 0.56084284 | 0.728598524 | 0.5038 | 0.560842842 | 0.560842842 | 0.560842842 | 3.0347259 | 0.482119471 | FALSE | TRUE | FALSE |
| SNCB | 1.7012974 | 0.50398338 | 0.50355 | 1.701297375 | 1.701297375 | 1.701297375 | 0.47583963 | 0.764724349 | 1 | 0.475839631 | 0.475839631 | 0.475839631 | 3.57535872 | 0.553319622 | FALSE | TRUE | FALSE |
| IRAK2 | 1.7009371 | 0.50412788 | 0.50755 | 1.700937052 | 15.83442911 | 15.83442911 | 0.0361408 | 0.889866016 | 1 | 0.036140796 | 0.036140796 | 0.713737849 | 47.06418301 | 1.672690524 | FALSE | TRUE | FALSE |
| OR2A5 | 1.7008609 | 0.50427228 | 0.5001 | 1.700860871 | 1.700860871 | 1.700860871 | 0.22625899 | 0.845792086 | 1 | 2.076175823 | 2.076175823 | 2.076175823 | 7.517318492 | 0.876062951 | FALSE | TRUE | FALSE |
| GCH1 | 1.700354 | 0.50441659 | 0.6857 | 1.700354038 | 27.21293959 | 27.21293959 | 0.56025436 | 0.728645298 | 0.87475 | 2.240615144 | 8.963091364 | 8.963091364 | 3.034967976 | 0.482154113 | FALSE | TRUE | FALSE |
| MAB21L3 | 1.6999862 | 0.5045608 | 0.01035 | 1.699986203 | 1.699986203 | 3.701795632 | 0.23357901 | 0.843769303 | 1 | 0.233579009 | 0.233579009 | 0.233579009 | 7.277992183 | 0.862011585 | FALSE | TRUE | FALSE |
| MCUR1 | 1.697429 | 0.50470495 | 0.2504 | 1.697428956 | 15.84503656 | 15.84503656 | 0.22653504 | 0.84571862 | 1 | 2.070548928 | 19.42366593 | 19.42366593 | 7.493008312 | 0.874656214 | FALSE | TRUE | FALSE |
| ABHD11 | 1.6964478 | 0.50484902 | 0.01235 | 0.42416457 | 1.6943347 | 2.117106711 | 0.14126671 | 0.866528881 | 1 | 0.141266711 | 0.141266711 | 0.141266711 | 12.00882949 | 1.079500678 | FALSE | TRUE | FALSE |
| DUSP8 | 1.6958492 | 0.50499299 | 0.5033 | 1.695849183 | 1.695849183 | 8.402641435 | 0.47449039 | 0.765311647 | 0.0102 | 0.474490388 | 0.474490388 | 9.413751016 | 3.574043277 | 0.553159807 | FALSE | TRUE | FALSE |
| FICD | 1.6955016 | 0.50513687 | 0.82365 | 11.85740608 | 434.2970785 | 1738.522366 | 0.28974307 | 0.828234506 | 0.51205 | 0.289743075 | 0.289743075 | 1.433643932 | 5.85174162 | 0.767285142 | FALSE | TRUE | FALSE |
| RBL1 | 1.6943347 | 0.50528067 | 0.1249 | 0.106069945 | 1.6943347 | 8.304531538 | 0.00224673 | 0.898982423 | 0.749 | 0.002246732 | 0.00897595 | 0.075790419 | 754.1328362 | 2.877447851 | FALSE | TRUE | FALSE |
| FGF21 | 1.692534 | 0.5054244 | 0.49835 | 0.182142964 | 1.692533987 | 1.692533987 | 0.22955617 | 0.845009889 | 0.00635 | 0.229556173 | 0.229556173 | 0.229556173 | 7.373071123 | 0.867648423 | FALSE | TRUE | FALSE |
| DHDH | 1.6916354 | 0.50556804 | 0.0124 | 1.691635366 | 1.691635366 | 3.647531927 | 2.00423741 | 0.489368865 | 0.754 | 2.004237414 | 18.95387512 | 18.95387512 | 0.844029432 | -0.073642409 | FALSE | FALSE | FALSE |
| DCAF10 | 1.6914511 | 0.50571158 | 0.0154 | 1.691451078 | 1.691451078 | 14.30748825 | 0.47335417 | 0.765758705 | 1 | 0.473354175 | 0.473354175 | 9.376916358 | 3.573330855 | 0.553073229 | FALSE | TRUE | FALSE |
| B4GALT4 | 1.6882751 | 0.50585507 | 0.50175 | 1.68827511 | 1.68827511 | 3.633051667 | 0.23010779 | 0.844891159 | 1 | 0.230107789 | 0.230107789 | 0.230107789 | 7.33688815 | 0.865511898 | FALSE | TRUE | FALSE |
| WNK4 | 1.6859242 | 0.5059985 | 0.27215 | 0.193291284 | 15.07434073 | 60.27900893 | 0.05728217 | 0.885224689 | 0.39055 | 0.027173271 | 0.229073894 | 1.113008656 | 29.43192064 | 1.468818604 | FALSE | TRUE | FALSE |
| HNRNPU | 1.6823514 | 0.50614188 | 0.514 | 1.682351361 | 1.682351361 | 33.00749523 | 0.22711848 | 0.845512542 | 1 | 0.227118478 | 0.227118478 | 4.466231411 | 7.407373365 | 0.869664236 | FALSE | TRUE | FALSE |
| KCNA2 | 1.6808015 | 0.50628519 | 0.754 | 1.680801501 | 16.22155372 | 33.28864948 | 2.13122649 | 0.478279627 | 0.50665 | 2.131226487 | 2.131226487 | 4.484576715 | 0.788654567 | -0.103113178 | FALSE | FALSE | FALSE |
| NDUFA5 | 1.6806266 | 0.5064284 | 0.5055 | 1.680626616 | 1.680626616 | 1.680626616 | 0.47063509 | 0.766783571 | 1 | 0.470635086 | 0.470635086 | 0.470635086 | 3.570976038 | 0.552786936 | FALSE | TRUE | FALSE |
| C1orf56 | 1.6797241 | 0.50657153 | 0.96645 | 150.8226419 | 1483.640117 | 14951.10986 | 2.32768179 | 0.462415928 | 0.00945 | 0.253241182 | 2.327681792 | 2.327681792 | 0.721629622 | -0.141685648 | FALSE | FALSE | FALSE |
| ZDHHC16 | 1.6792971 | 0.50671456 | 0.75365 | 1.679297072 | 15.87083858 | 15.87083858 | 0.23665014 | 0.842981913 | 1 | 0.236650144 | 0.236650144 | 4.696458755 | 7.09611684 | 0.851020758 | FALSE | TRUE | FALSE |
| SOCS4 | 1.6792054 | 0.5068575 | 0.5057 | 1.679205395 | 1.679205395 | 3.611909416 | 0.22665582 | 0.845659798 | 0.5046 | 0.906517017 | 0.906517017 | 0.906517017 | 7.408613548 | 0.869736941 | FALSE | TRUE | FALSE |
| BSG | 1.6781486 | 0.50700035 | 0.26185 | 1.678148607 | 15.78685944 | 33.19800326 | 0.23727563 | 0.842768601 | 1 | 0.237275628 | 0.237275628 | 4.700049604 | 7.072570504 | 0.849577285 | FALSE | TRUE | FALSE |
| FAM161A | 1.6741543 | 0.50714317 | 0.2584 | 0.188827749 | 1.674154343 | 3.730434864 | 2.55646673 | 0.442918415 | 0.5073 | 2.556466725 | 23.47566491 | 23.47566491 | 0.654870383 | -0.183844651 | FALSE | FALSE | FALSE |
| TATDN3 | 1.6730405 | 0.5072859 | 0.8147 | 6.694453762 | 26.79367902 | 107.264999 | 2.14948029 | 0.476387711 | 0.65965 | 8.600993363 | 137.817508 | 443.1344004 | 0.778346508 | -0.108827019 | FALSE | FALSE | FALSE |
| BOLA1 | 1.6726516 | 0.50742854 | 0.874 | 6.691435333 | 26.77574755 | 26.77574755 | 7.50019782 | 0.23484493 | 0.00495 | 1.874782362 | 7.500197816 | 7.500197816 | 0.223014336 | -0.651667219 | FALSE | FALSE | FALSE |
| USB1 | 1.6726246 | 0.50757108 | 0.25265 | 1.672624616 | 15.86960735 | 15.86960735 | 2.11629928 | 0.479476776 | 0.4994 | 0.228012053 | 2.116299279 | 2.116299279 | 0.790353535 | -0.1021786 | FALSE | FALSE | FALSE |
| CLIC1 | 1.668413 | 0.50771359 | 0.50695 | 1.668412983 | 1.668412983 | 1.668412983 | 1.8497816 | 0.503800745 | 0.5045 | 1.8497816 | 1.8497816 | 1.8497816 | 0.901951335 | -0.044816894 | FALSE | FALSE | FALSE |
| HOXC4 | 1.6683105 | 0.507856 | 0.50495 | 1.668310501 | 1.668310501 | 3.526778316 | 0.22248485 | 0.846740962 | 1 | 0.88965599 | 0.88965599 | 0.88965599 | 7.498535396 | 0.874976446 | FALSE | TRUE | FALSE |
| GPR78 | 1.6674044 | 0.50799833 | 0.50845 | 1.667404371 | 1.667404371 | 3.571221915 | 2.04360583 | 0.485388967 | 0.50705 | 2.043605832 | 2.043605832 | 4.368690014 | 0.815912905 | -0.088356198 | FALSE | FALSE | FALSE |
| PCDHGB1 | 1.6669153 | 0.50814056 | 0.3306 | 1.666915292 | 157.0694822 | 157.0694822 | 0.03509736 | 0.890084887 | 0.02865 | 0.035097359 | 0.035097359 | 0.686919606 | 47.49403705 | 1.676639087 | FALSE | TRUE | FALSE |
| BAMBI | 1.6635812 | 0.50828275 | 0.50505 | 6.653618915 | 62.61986323 | 62.61986323 | 8.41095005 | 0.219549755 | 0.25465 | 2.102934292 | 8.410950046 | 8.410950046 | 0.19778755 | -0.703801049 | FALSE | FALSE | FALSE |
| SEMG2 | 1.6619039 | 0.50842487 | 0.891 | 15.02129637 | 1321.118378 | 12840.13882 | 1821.36454 | 0.001318415 | 0.50885 | 17825.55808 | 1828781.057 | 19151157.87 | 0.00091245 | -3.039790968 | FALSE | FALSE | FALSE |
| SIN3B | 1.6618909 | 0.50856689 | 0.75735 | 1.661890929 | 15.93307035 | 308.5255661 | 18.9696105 | 0.137902146 | 0.15155 | 18.96961046 | 186.7244614 | 368.7337875 | 0.087608068 | -1.057455895 | FALSE | FALSE | FALSE |
| MTHFD2L | 1.6610999 | 0.50870882 | 0.0077 | 1.661099901 | 1.661099901 | 1.661099901 | 0.21926644 | 0.847362614 | 0.51095 | 2.038387054 | 2.038387054 | 2.038387054 | 7.575714229 | 0.879423584 | FALSE | TRUE | FALSE |
| MRPS18B | 1.6606727 | 0.50885066 | 0.5068 | 1.660672654 | 1.660672654 | 1.660672654 | 0.22183907 | 0.846827981 | 1 | 0.221839073 | 2.032150922 | 2.032150922 | 7.485933969 | 0.874245992 | FALSE | TRUE | FALSE |
| PPIE | 1.6581743 | 0.50899244 | 0.5031 | 15.77726903 | 153.971192 | 153.971192 | 8.30994122 | 0.221564602 | 0.5048 | 8.309941223 | 79.36010363 | 79.36010363 | 0.199541044 | -0.69996776 | FALSE | FALSE | FALSE |
| RBM11 | 1.6577561 | 0.50913413 | 0.87575 | 1.65775607 | 150.2233811 | 150.2233811 | 0.23216781 | 0.844414445 | 0.75255 | 2.138925424 | 8.555006728 | 8.555006728 | 7.140335448 | 0.853718615 | FALSE | TRUE | FALSE |
| COPE | 1.6554771 | 0.50927576 | 0.74565 | 1.655477101 | 15.5350973 | 15.5350973 | 4.36455022 | 0.33667776 | 0.4999 | 0.464446928 | 4.364550216 | 4.364550216 | 0.379300734 | -0.421016317 | FALSE | FALSE | FALSE |
| HEY1 | 1.6542719 | 0.50941731 | 0.5016 | 1.654271871 | 1.654271871 | 8.204947051 | 0.46434189 | 0.769371867 | 1 | 0.46434189 | 0.46434189 | 0.46434189 | 3.562616054 | 0.551769021 | FALSE | TRUE | FALSE |
| ATL1 | 1.6537844 | 0.50955876 | 0.75475 | 6.613449782 | 63.32727071 | 63.32727071 | 0.92538391 | 0.595134057 | 0.50625 | 0.925383915 | 0.925383915 | 4.588372622 | 1.787133303 | 0.252156948 | FALSE | TRUE | FALSE |
| RSPO1 | 1.6513119 | 0.50970016 | 0.50645 | 1.651311854 | 15.42200705 | 15.42200705 | 0.14089687 | 0.8666068 | 1 | 0.563344306 | 0.563344306 | 0.563344306 | 11.72000372 | 1.068927749 | FALSE | TRUE | FALSE |
| MAP2K2 | 1.6488242 | 0.5098415 | 0.25885 | 0.41230997 | 1.648824162 | 2.044129714 | 0.54547572 | 0.735562203 | 0.01095 | 0.545475718 | 0.545475718 | 2.70707223 | 3.022726968 | 0.480398921 | FALSE | TRUE | FALSE |
| PRKG2 | 1.6487846 | 0.50998274 | 0.50895 | 1.648784586 | 6.593676456 | 23.86764999 | 0.22614227 | 0.845821453 | 0.01825 | 0.22614227 | 0.22614227 | 4.47041867 | 7.290917291 | 0.862782172 | FALSE | TRUE | FALSE |
| BARX1 | 1.6485411 | 0.51012389 | 0.5044 | 1.648541056 | 1.648541056 | 1.648541056 | 0.46304932 | 0.769736722 | 1 | 0.463049318 | 0.463049318 | 0.463049318 | 3.560184607 | 0.551472518 | FALSE | TRUE | FALSE |
| OR51A7 | 1.6446206 | 0.510265 | 0.2561 | 1.644620646 | 15.45147722 | 15.45147722 | 0.23322009 | 0.843964921 | 1 | 0.233220086 | 0.233220086 | 0.233220086 | 7.051796765 | 0.848299787 | FALSE | TRUE | FALSE |
| PHPT1 | 1.6443625 | 0.51040602 | 0.00485 | 0.178218201 | 1.644362522 | 1.644362522 | 0.22491675 | 0.84612915 | 0.00415 | 0.224916751 | 0.224916751 | 0.224916751 | 7.310982919 | 0.863975769 | FALSE | TRUE | FALSE |
| RNASEH2C | 1.6433872 | 0.51054696 | 0.5011 | 1.64338724 | 1.64338724 | 1.64338724 | 0.22301119 | 0.846624787 | 0.00635 | 0.223011187 | 0.223011187 | 0.223011187 | 7.369079842 | 0.867413262 | FALSE | TRUE | FALSE |
| GORASP2 | 1.6417537 | 0.51068783 | 0.50435 | 1.641753674 | 1.641753674 | 8.143379683 | 0.4609001 | 0.770725656 | 1 | 0.460900104 | 0.460900104 | 9.153174378 | 3.562059673 | 0.551701191 | FALSE | TRUE | FALSE |
| ROPN1L | 1.6410493 | 0.5108286 | 0.9699 | 146.470814 | 1436.532363 | 14433.239 | 158.726235 | 0.016904484 | 0.7403 | 1559.002479 | 1703553.117 | 18367336.05 | 0.010338866 | -1.985527096 | FALSE | FALSE | FALSE |
| ZNRF4 | 1.6407559 | 0.51096929 | 0.5127 | 15.48835709 | 149.9602143 | 149.9602143 | 0.23283211 | 0.84413005 | 1 | 0.232832109 | 0.232832109 | 4.600122188 | 7.04694845 | 0.848001095 | FALSE | TRUE | FALSE |
| MIPEP | 1.6407357 | 0.51110988 | 0.6969 | 1.640735746 | 139.8132 | 139.8132 | 2.59219345 | 0.440192476 | 0.6884 | 2.592193453 | 225.474893 | 225.474893 | 0.632952662 | -0.19862877 | FALSE | FALSE | FALSE |

| | | | | | | | | | | | | | | | | | |
|---|---|---|---|---|---|---|---|---|---|---|---|---|---|---|---|---|---|
| LSG1 | 1.6399492 | 0.51125039 | 0.55215 | 3.627076835 | 232.3322239 | 887.72548 | 0.70682332 | 0.662955341 | 0.73555 | 2.827466851 | 45.27895575 | 181.2627541 | 2.320168439 | 0.365519515 | FALSE | TRUE | FALSE |
| UBE2U | 1.6363057 | 0.51139085 | 0.93445 | 15.2754141 | 1435.5555 | 1435.5555 | 2.13468195 | 0.477936647 | 0.75245 | 2.134681954 | 20.06201787 | 20.06201787 | 0.766533706 | -0.115468744 | FALSE | FALSE | FALSE |
| SYT13 | 1.6349128 | 0.51153124 | 0.50035 | 1.634912781 | 1.634912781 | 1.634912781 | 0.45936588 | 0.771216826 | 1 | 0.459365877 | 0.459365877 | 0.459365877 | 3.559064488 | 0.551335857 | FALSE | TRUE | FALSE |
| CD6 | 1.6346126 | 0.51167154 | 0.75355 | 1.63461257 | 15.37169255 | 32.29430814 | 2.07051186 | 0.483544772 | 0.5072 | 2.070511864 | 2.070511864 | 4.448748891 | 0.789472689 | -0.102662889 | FALSE | FALSE | FALSE |
| ADRM1 | 1.6337044 | 0.51181176 | 0.50005 | 1.633704377 | 1.633704377 | 8.079101274 | 4.19577641 | 0.343795474 | 0.50665 | 4.195776409 | 4.195776409 | 8.796297502 | 0.389368788 | -0.409638864 | FALSE | FALSE | FALSE |
| TAS2R8 | 1.6333566 | 0.51195189 | 0.87585 | 15.22156838 | 145.4964208 | 145.4964208 | 19.3558953 | 0.135408766 | 0.00495 | 2.064547265 | 19.35589533 | 19.35589533 | 0.084385483 | -1.073732262 | FALSE | FALSE | FALSE |
| CYP2S1 | 1.6321881 | 0.51209193 | 0.01555 | 1.632188051 | 1.632188051 | 5.931543682 | 0.21494356 | 0.848378488 | 1 | 0.214943557 | 2.008877284 | 4.25596765 | 7.593565843 | 0.880445763 | FALSE | TRUE | FALSE |
| UNG | 1.6272579 | 0.51223196 | 0.6898 | 3.79156955 | 144.8559033 | 144.8559033 | 87.0423523 | 0.033658118 | 0.09965 | 9.29516429 | 209.0332794 | 836.0162999 | 0.018695013 | -1.728274223 | FALSE | FALSE | FALSE |
| IFI30 | 1.6272013 | 0.51237189 | 0.9353 | 15.23630251 | 1440.530556 | 1440.530556 | 2.16961392 | 0.474307969 | 0.2588 | 2.169613916 | 8.677946429 | 8.677946429 | 0.749995808 | -0.124941164 | FALSE | FALSE | FALSE |
| ZMYND10 | 1.625174 | 0.51251176 | 0.5124 | 1.625173957 | 6.50006623 | 14.01180079 | 0.22575456 | 0.84590949 | 1 | 0.225754561 | 0.225754561 | 4.478019481 | 7.198853255 | 0.857263321 | FALSE | TRUE | FALSE |
| IAH1 | 1.6238498 | 0.51265155 | 0.50425 | 1.623849816 | 1.623849816 | 1.623849816 | 18.2065952 | 0.14257811 | 0.00435 | 1.931964606 | 18.20659518 | 18.20659518 | 0.089190197 | -1.049682875 | FALSE | FALSE | FALSE |
| MOB1A | 1.6237095 | 0.51279125 | 0.25625 | 1.623709469 | 6.495541807 | 6.495541807 | 0.46173104 | 0.770397005 | 0.0037 | 0.461731042 | 0.461731042 | 0.461731042 | 3.516569868 | 0.546119251 | FALSE | TRUE | FALSE |
| TSHR | 1.6213371 | 0.51293089 | 0.2325 | 0.706372291 | 15.25825776 | 60.99249472 | 7.3277805 | 0.237824081 | 0.13335 | 0.773342297 | 29.28759844 | 29.28759844 | 0.221258963 | -0.655099127 | FALSE | FALSE | FALSE |
| ZBP1 | 1.6184318 | 0.51307048 | 0.00885 | 0.179298878 | 1.618431762 | 1.618431762 | 0.02932938 | 0.891427672 | 1 | 0.260398879 | 0.260398879 | 0.582051023 | 55.18124231 | 1.741791474 | FALSE | TRUE | FALSE |
| SLC35F4 | 1.6174988 | 0.51320999 | 0.7502 | 1.617498806 | 6.471410667 | 8.024399078 | 0.46011526 | 0.770955138 | 1 | 0.460115257 | 0.460115257 | 9.138827754 | 3.515420931 | 0.545977334 | FALSE | TRUE | FALSE |
| HMGCR | 1.6154828 | 0.51334943 | 0.93965 | 25.85884182 | 414.3331784 | 1659.138663 | 0.5665791 | 0.725951936 | 0.50845 | 0.566579098 | 0.566579098 | 11.16127691 | 2.851292663 | 0.455041797 | FALSE | TRUE | FALSE |
| CDH13 | 1.6135289 | 0.51348881 | 0.5106 | 1.613528927 | 1.613528927 | 5.873639448 | 0.22009605 | 0.847117402 | 1 | 0.220096052 | 0.220096052 | 4.341336361 | 7.331021662 | 0.865164503 | FALSE | TRUE | FALSE |
| CHRNB3 | 1.6133914 | 0.51362809 | 0.5154 | 2.014218411 | 103.3991545 | 103.3991545 | 0.19438142 | 0.85336147 | 0.49985 | 0.048616032 | 0.777389241 | 0.777389241 | 8.300131757 | 0.919084986 | FALSE | TRUE | FALSE |
| CD3G | 1.6107697 | 0.51376732 | 0.5018 | 1.610769719 | 1.610769719 | 1.610769719 | 0.45290597 | 0.773512344 | 1 | 0.452905968 | 0.452905968 | 0.452905968 | 3.556521292 | 0.551025413 | FALSE | TRUE | FALSE |
| GNAL | 1.6099268 | 0.51390647 | 0.7507 | 1.609926806 | 6.440155056 | 13.64139559 | 0.45868186 | 0.771477891 | 1 | 0.458681858 | 0.458681858 | 9.101049521 | 3.509898587 | 0.545294568 | FALSE | TRUE | FALSE |
| GAR1 | 1.6075032 | 0.51404556 | 0.509 | 1.607503242 | 1.607503242 | 1.607503242 | 0.45279221 | 0.773544336 | 0.00685 | 0.452792215 | 0.452792215 | 0.452792215 | 3.550200712 | 0.550252907 | FALSE | TRUE | FALSE |
| ZNF594 | 1.6074837 | 0.51418455 | 0.132 | 0.184844295 | 1.607483667 | 1.607483667 | 0.34861222 | 0.809532939 | 0.8163 | 3.073296455 | 27.77358909 | 257.1382994 | 4.6110939 | 0.663803966 | FALSE | TRUE | FALSE |
| GRTP1 | 1.6070426 | 0.51432346 | 0.00845 | 0.182641255 | 1.607042627 | 1.607042627 | 2.45811819 | 0.451141666 | 0.5056 | 2.458118187 | 22.42008682 | 22.42008682 | 0.653769471 | -0.184575364 | FALSE | FALSE | FALSE |
| SYT6 | 1.6043439 | 0.51446231 | 0.50665 | 1.604343886 | 1.604343886 | 3.401573613 | 1.97893069 | 0.490848363 | 0.5068 | 1.978930688 | 1.978930688 | 4.192585209 | 0.81071252 | -0.09113312 | FALSE | FALSE | FALSE |
| CUEDC1 | 1.6041644 | 0.51460107 | 0.50225 | 1.604164369 | 1.604164369 | 3.477344365 | 1.97173402 | 0.491503158 | 0.01235 | 1.971734024 | 1.971734024 | 4.260212668 | 0.813580508 | -0.089599465 | FALSE | FALSE | FALSE |
| RPS5 | 1.6031149 | 0.51473975 | 0.50595 | 1.603114899 | 1.603114899 | 1.603114899 | 0.44874489 | 0.774784827 | 1 | 0.448744887 | 0.448744887 | 0.448744887 | 3.572441597 | 0.552965138 | FALSE | TRUE | FALSE |
| KIF4B | 1.6020811 | 0.51487836 | 1 | 1.83442E+12 | 2.82522E+17 | 1.42366E+19 | 0.0052676 | 0.89806484 | 1 | 2.76689E+15 | 6.60617E+20 | 1.14017E+23 | 304.1386873 | 2.483071667 | FALSE | TRUE | FALSE |
| GJB3 | 1.600781 | 0.51501689 | 0.35135 | 0.400444033 | 6.400744895 | 25.59985816 | 0.74617701 | 0.643316534 | 0.3133 | 0.186675305 | 2.983357981 | 2.983357981 | 2.14531005 | 0.331490067 | FALSE | TRUE | FALSE |
| DEAF1 | 1.5993165 | 0.51515535 | 0.51225 | 1.599316475 | 1.599316475 | 5.873919119 | 0.18828141 | 0.854931743 | 1 | 159.3494003 | 1585.022694 | 16150.92503 | 8.494287856 | 0.929126974 | FALSE | TRUE | FALSE |
| TAF13 | 1.598381 | 0.51529373 | 0.75185 | 1.598380978 | 6.394781658 | 6.394781658 | 1.72495917 | 0.515554775 | 0.87655 | 4.024375449 | 64.40005232 | 64.40005232 | 0.926619601 | -0.033098517 | FALSE | FALSE | FALSE |
| RNF167 | 1.5965667 | 0.51543204 | 0.50895 | 1.596566683 | 1.596566683 | 1.596566683 | 0.20808723 | 0.850191194 | 1 | 1.901677963 | 17.83709626 | 17.83709626 | 7.672583785 | 0.88494164 | FALSE | TRUE | FALSE |
| NHLRC2 | 1.5936999 | 0.5155703 | 0.25425 | 0.398611726 | 1.593699936 | 5.901864504 | 0.25679593 | 0.837504956 | 0.7534 | 0.256795928 | 1.027012109 | 5.08821588 | 6.206095046 | 0.792818422 | FALSE | TRUE | FALSE |
| SLC16A8 | 1.5936377 | 0.51570847 | 0.50555 | 1.593637717 | 1.593637717 | 3.428863842 | 0.2191328 | 0.847391411 | 0.0105 | 0.2191328 | 0.2191328 | 4.346024617 | 7.272474581 | 0.861682212 | FALSE | TRUE | FALSE |
| FSHB | 1.5929564 | 0.51584655 | 0.5008 | 1.592956405 | 1.592956405 | 1.592956405 | 0.43524432 | 0.779156262 | 1 | 4.122256809 | 4.122256809 | 4.122256809 | 3.659913115 | 0.563470776 | FALSE | TRUE | FALSE |
| SMG5 | 1.5926285 | 0.51598455 | 0.51245 | 1.592628503 | 1.592628503 | 31.15113551 | 0.21884806 | 0.847434588 | 0.02695 | 0.218848056 | 0.218848056 | 4.291930132 | 7.277325328 | 0.86197179 | FALSE | TRUE | FALSE |
| TFB2M | 1.5921426 | 0.51612246 | 0.504 | 1.592142648 | 1.592142648 | 3.464142176 | 0.22045942 | 0.847088504 | 0.01035 | 0.220459418 | 0.220459418 | 4.374702266 | 7.22193075 | 0.85865332 | FALSE | TRUE | FALSE |
| ARHGAP28 | 1.5903931 | 0.5162603 | 0.7499 | 1.590393136 | 14.92069349 | 31.4772348 | 1.95741423 | 0.492645639 | 0.2542 | 1.957414228 | 18.44452051 | 38.78979888 | 0.812496973 | -0.090178248 | FALSE | FALSE | FALSE |
| THNSL2 | 1.5898492 | 0.51639806 | 0.50225 | 1.589849239 | 14.76092935 | 14.76092935 | 2.61855604 | 0.439213617 | 0.25765 | 0.286639439 | 2.618556044 | 2.618556044 | 0.607147303 | -0.21670593 | FALSE | FALSE | FALSE |
| ATL3 | 1.5896171 | 0.51653573 | 0.0109 | 0.172974005 | 1.58961708 | 3.427161595 | 0.12317311 | 0.870524458 | 0.63115 | 0.4925858 | 4.443966693 | 4.443966693 | 12.90555317 | 1.110776624 | FALSE | TRUE | FALSE |
| LRRC26 | 1.5877988 | 0.51667333 | 0.49685 | 0.173132942 | 1.587798801 | 1.587798801 | 0.19347908 | 0.853494682 | 1 | 1.779367949 | 16.79556626 | 16.79556626 | 8.20656573 | 0.914161452 | FALSE | TRUE | FALSE |
| GLT8D1 | 1.5865455 | 0.51681086 | 0.01085 | 1.586545487 | 1.586545487 | 3.452242041 | 1.95253483 | 0.493296206 | 0.50325 | 1.952534826 | 1.952534826 | 1.952534826 | 0.812556819 | -0.090146261 | FALSE | FALSE | FALSE |
| AMH | 1.5859882 | 0.51694831 | 0.72115 | 13.99709743 | 224.213342 | 897.7096158 | 65147.464 | 3.0402E-05 | 0.184 | 4056.916169 | 579723.5049 | 9318706.983 | 2.43446E-05 | -4.613597551 | FALSE | FALSE | FALSE |
| ZNF330 | 1.5853396 | 0.51708567 | 0.50565 | 1.585339563 | 1.585339563 | 1.585339563 | 0.10567642 | 0.874501947 | 0.50455 | 0.105676425 | 0.105676425 | 0.105676425 | 15.00182813 | 1.176144186 | FALSE | TRUE | FALSE |
| SH2D2A | 1.5851105 | 0.51722294 | 0.5056 | 1.585110533 | 1.585110533 | 1.585110533 | 0.21786244 | 0.84772182 | 0.0086 | 0.217862438 | 0.217862438 | 0.217862438 | 7.275740371 | 0.861877193 | FALSE | TRUE | FALSE |
| P2RY2 | 1.5845529 | 0.51736012 | 0.7531 | 1.584552865 | 15.00272123 | 31.29428015 | 0.22462455 | 0.846246051 | 0.0124 | 0.224624549 | 0.224624549 | 4.442927454 | 7.054228355 | 0.848449514 | FALSE | TRUE | FALSE |
| ADH7 | 1.5839195 | 0.51749723 | 0.8762 | 14.84480459 | 142.7013402 | 142.7013402 | 0.233004 | 0.844100053 | 1 | 0.233003996 | 0.233003996 | 0.233003996 | 6.79782138 | 0.832369749 | FALSE | TRUE | FALSE |
| RNF220 | 1.5828845 | 0.51763426 | 0.26615 | 0.395806954 | 1.582884497 | 7.808536134 | 0.12900789 | 0.869185564 | 1 | 1.20937215 | 1.20937215 | 2.549249307 | 12.26967191 | 1.08883295 | FALSE | TRUE | FALSE |
| TP53RK | 1.5828284 | 0.51777119 | 0.5009 | 1.582828372 | 1.582828372 | 1.582828372 | 1.88666464 | 0.500186786 | 0.74675 | 1.886664645 | 17.67423008 | 17.67423008 | 0.838955866 | -0.076260885 | FALSE | FALSE | FALSE |
| ADAD2 | 1.5812407 | 0.51790806 | 0.16385 | 0.178936246 | 6.529064711 | 26.09571311 | 0.04819359 | 0.887070481 | 0.84805 | 0.433411314 | 15.97757478 | 63.869705 | 32.81018504 | 1.51600868 | FALSE | TRUE | FALSE |
| CABP5 | 1.5778155 | 0.51804488 | 0.0053 | 1.577815453 | 1.577815453 | 1.577815453 | 1.74089985 | 0.513607202 | 0.75355 | 1.74089985 | 6.964588869 | 6.964588869 | 0.906321781 | -0.042717583 | FALSE | FALSE | FALSE |
| PRKAR2B | 1.5748275 | 0.51818166 | 0.50875 | 1.574827536 | 1.574827536 | 1.574827536 | 1.75851524 | 0.512250453 | 0.00805 | 1.758515243 | 1.758515243 | 1.758515243 | 0.895543864 | -0.047913137 | FALSE | FALSE | FALSE |
| NEDD9 | 1.5722932 | 0.51831839 | 0.5128 | 1.57229318 | 1.57229318 | 30.88304101 | 0.21676347 | 0.847993714 | 1 | 0.216763466 | 0.216763466 | 4.266851333 | 7.253497132 | 0.860547444 | FALSE | TRUE | FALSE |
| PLCZ1 | 1.5703543 | 0.51845505 | 0.14185 | 1.570354311 | 14.67169777 | 14.67169777 | 0.28262151 | 0.830388269 | 0.75365 | 0.282621509 | 2.596345626 | 2.596345626 | 5.5563864 | 0.74479244 | FALSE | TRUE | FALSE |
| METTL23 | 1.5692885 | 0.51859164 | 0.75535 | 1.569288451 | 14.58103652 | 14.58103652 | 1.92993092 | 0.49572095 | 0.74925 | 1.929930922 | 18.03001455 | 18.03001455 | 0.813131928 | -0.089838986 | FALSE | FALSE | FALSE |
| DYNC1I2 | 1.5676081 | 0.51872816 | 0.25275 | 0.392043714 | 1.567608094 | 5.852181336 | 0.06509233 | 0.883575823 | 0.0141 | 0.065092326 | 0.065092326 | 1.288827453 | 24.08284017 | 1.381707703 | FALSE | TRUE | FALSE |
| RSAD2 | 1.5671147 | 0.51886459 | 0.38195 | 0.69439756 | 6.266346223 | 25.06319905 | 0.01886588 | 0.894128231 | 0.8744 | 0.301488517 | 2.736548861 | 2.736548861 | 83.06607874 | 1.919423709 | FALSE | TRUE | FALSE |
| CLTB | 1.5666839 | 0.51900094 | 0.50735 | 1.56668394 | 1.56668394 | 1.56668394 | 0.44264501 | 0.776448679 | 1 | 0.442645008 | 0.442645008 | 0.442645008 | 3.53936882 | 0.548925821 | FALSE | TRUE | FALSE |
| BCL2L11 | 1.5652241 | 0.51913722 | 0.00505 | 0.172787098 | 1.565224136 | 1.565224136 | 1432.70595 | 0.001675527 | 0.1128 | 146.4245119 | 14360.94663 | 147382.3586 | 0.001092495 | -2.961580527 | FALSE | FALSE | FALSE |
| KREMEN2 | 1.5624286 | 0.51927345 | 0.252 | 1.562428553 | 14.77151499 | 14.77151499 | 1.99168216 | 0.489533649 | 0.5077 | 1.991682162 | 1.991682162 | 1.991682162 | 0.784476852 | -0.105419867 | FALSE | FALSE | FALSE |
| GIN1 | 1.5624197 | 0.51940959 | 0.5085 | 1.562419734 | 1.562419734 | 3.396558529 | 0.2167385 | 0.848022279 | 1 | 0.2167385 | 0.2167385 | 4.301780317 | 7.20877801 | 0.857861652 | FALSE | TRUE | FALSE |
| RHOBTB2 | 1.5618114 | 0.51954565 | 0.99405 | 67380.38339 | 7708941.814 | 67434597.42 | 0.12477971 | 0.870165703 | 1 | 105.4668125 | 10680.28548 | 111374.4673 | 12.51654932 | 1.097484615 | FALSE | TRUE | FALSE |
| AAMDC | 1.5617573 | 0.51968162 | 0.00335 | 0.168558909 | 1.561757322 | 1.561757322 | 7.31067797 | 0.238814079 | 0.37825 | 7.310677967 | 278.6243568 | 278.6243568 | 0.21362688 | -0.670344103 | FALSE | FALSE | FALSE |
| MPZL1 | 1.5596037 | 0.51981752 | 0.75635 | 1.559603654 | 14.94834514 | 14.94834514 | 0.21956751 | 0.847232896 | 1 | 0.219567506 | 0.219567506 | 0.219567506 | 7.103071307 | 0.851446175 | FALSE | TRUE | FALSE |
| BTBD7 | 1.5566348 | 0.51995338 | 0.72765 | 8.468646029 | 165.6570716 | 625.7856117 | 0.13914119 | 0.866928731 | 1 | 0.556485043 | 0.556485043 | 10.91429617 | 11.18744746 | 1.048731009 | FALSE | TRUE | FALSE |
| WIPF1 | 1.5556154 | 0.52008917 | 0.51095 | 1.555615374 | 1.555615374 | 5.621897146 | 0.21176398 | 0.849156559 | 1 | 0.211763984 | 0.211763984 | 4.190771411 | 7.345986522 | 0.866050127 | FALSE | TRUE | FALSE |
| RPUSD1 | 1.5546457 | 0.52022488 | 0.7466 | 1.554645745 | 14.55848149 | 14.55848149 | 0.22242585 | 0.846769979 | 0.0101 | 0.222425853 | 0.222425853 | 4.412372751 | 6.989501098 | 0.844446177 | FALSE | TRUE | FALSE |
| SLC22A15 | 1.5544996 | 0.5203605 | 0.26415 | 0.388737575 | 1.554499646 | 3.424293494 | 0.06492475 | 0.883627728 | 1 | 0.064924746 | 0.064924746 | 1.28641174 | 23.94309953 | 1.379180371 | FALSE | TRUE | FALSE |
| PREP | 1.5534072 | 0.52049604 | 0.01935 | 1.553407233 | 1.553407233 | 30.63918388 | 0.21399983 | 0.848591466 | 0.0165 | 0.213999826 | 0.213999826 | 4.227915669 | 7.258918211 | 0.860871903 | FALSE | TRUE | FALSE |
| C8orf34 | 1.552368 | 0.52063151 | 0.503 | 1.552368023 | 1.552368023 | 1.552368023 | 1.91445191 | 0.497803942 | 0.50245 | 1.914451911 | 1.914451911 | 1.914451911 | 0.810868121 | -0.091049774 | FALSE | FALSE | FALSE |
| FGF1 | 1.5503754 | 0.52076692 | 0.50695 | 1.550375448 | 1.550375448 | 1.550375448 | 0.4383981 | 0.777903258 | 1 | 0.438398099 | 0.438398099 | 0.438398099 | 3.536455683 | 0.54856822 | FALSE | TRUE | FALSE |
| AKAP7 | 1.5495944 | 0.52090225 | 0.50385 | 1.549594414 | 1.549594414 | 1.549594414 | 156.446517 | 0.017307981 | 0.0696 | 16.28898438 | 1540.199017 | 1540.199017 | 0.009904947 | -2.004147856 | FALSE | FALSE | FALSE |
| PBX3 | 1.5480732 | 0.52103752 | 0.50495 | 1.548073212 | 1.548073212 | 3.296108288 | 0.21198877 | 0.849086143 | 1 | 0.211988773 | 0.211988773 | 4.205423362 | 7.30261885 | 0.863478634 | FALSE | TRUE | FALSE |
| GORAB | 1.5444608 | 0.52117274 | 0.7559 | 1.544460814 | 14.4576491 | 30.63430645 | 0.21453212 | 0.848435337 | 0.50755 | 1.967492083 | 1.967492083 | 4.259457563 | 7.199205399 | 0.857284565 | FALSE | TRUE | FALSE |

| | | | | | | | | | | | | | | | | | |
|---|---|---|---|---|---|---|---|---|---|---|---|---|---|---|---|---|---|
| ALDH7A1 | 1.5443719 | 0.52130788 | 0.231 | 0.386152114 | 6.178083814 | 24.72089207 | 0.00323331 | 0.898606445 | 1 | 0.206463345 | 1.828614036 | 7.312583736 | 477.6438354 | 2.679104177 | FALSE | TRUE | FALSE |
| UBA5 | 1.5416752 | 0.52144297 | 0.4991 | 0.385457607 | 1.541675225 | 1.541675225 | 0.43619441 | 0.778760561 | 1 | 0.436194408 | 0.436194408 | 0.436194408 | 3.534376406 | 0.548312799 | FALSE | TRUE | FALSE |
| TBX18 | 1.5411864 | 0.52157797 | 0.25945 | 0.385428473 | 1.541186378 | 7.608080141 | 0.13012488 | 0.86893836 | 1 | 0.130124885 | 0.130124885 | 2.57260063 | 11.84390198 | 1.073494804 | FALSE | TRUE | FALSE |
| FAM83F | 1.5379153 | 0.52171293 | 0.5062 | 1.537915309 | 1.537915309 | 3.345798989 | 0.21400473 | 0.848577285 | 0.01145 | 0.214004735 | 0.214004735 | 4.243089482 | 7.186361134 | 0.856509038 | FALSE | TRUE | FALSE |
| RBM48 | 1.53773 | 0.52184781 | 0.7562 | 6.150217611 | 57.9233952 | 57.9233952 | 0.87790707 | 0.601733646 | 0.50925 | 0.877907073 | 0.877907073 | 4.360650462 | 1.751586302 | 0.24343154 | FALSE | TRUE | FALSE |
| OR2A14 | 1.5367521 | 0.52198261 | 0.50565 | 1.536752149 | 1.536752149 | 1.536752149 | 0.21388722 | 0.848605644 | 1 | 0.213887219 | 0.213887219 | 0.213887219 | 7.184871342 | 0.856418996 | FALSE | TRUE | FALSE |
| BMP8A | 1.5349949 | 0.52211735 | 0.7527 | 1.534994857 | 14.74170708 | 30.39592621 | 1.9660062 | 0.491666693 | 0.50485 | 1.966006195 | 1.966006195 | 4.154407429 | 0.780768067 | -0.107477957 | FALSE | FALSE | FALSE |
| XRRA1 | 1.5318545 | 0.52225204 | 0.8953 | 14.06578826 | 1276.651075 | 12604.59759 | 0.30537987 | 0.82379712 | 0.74965 | 0.305379867 | 2.769406064 | 6.023753103 | 5.016226275 | 0.700377119 | FALSE | TRUE | FALSE |
| PCDHAC2 | 1.5305824 | 0.52238666 | 0.5232 | 1.530582373 | 1.530582373 | 29.91716824 | 0.20886114 | 0.850010439 | 1 | 0.208861143 | 0.208861143 | 4.093613116 | 7.328229419 | 0.864999057 | FALSE | TRUE | FALSE |
| CCDC121 | 1.5284549 | 0.52252123 | 0.50745 | 1.528454905 | 1.528454905 | 1.528454905 | 0.20068088 | 0.851948924 | 1 | 1.828954137 | 17.10788122 | 17.10788122 | 7.616345472 | 0.881746635 | FALSE | TRUE | FALSE |
| ACAP3 | 1.5258976 | 0.52265574 | 0.75505 | 1.52589756 | 14.48658065 | 30.05211414 | 0.21689068 | 0.84793655 | 1 | 0.216890685 | 0.216890685 | 4.279447939 | 7.035330092 | 0.847284479 | FALSE | TRUE | FALSE |
| KRT6B | 1.5228334 | 0.52279021 | 0.54195 | 2.12066789 | 135.2895172 | 707.5243851 | 0.09359708 | 0.877416639 | 0.505 | 0.093597079 | 0.855474087 | 0.855474087 | 16.27009482 | 1.211390084 | FALSE | TRUE | FALSE |
| SPACA7 | 1.522744 | 0.5229246 | 0.74705 | 1.522744021 | 14.14619927 | 14.14619927 | 0.21944 | 0.84730499 | 1 | 0.219440001 | 0.219440001 | 0.219440001 | 6.939227175 | 0.841311106 | FALSE | TRUE | FALSE |
| RIPPLY3 | 1.5226044 | 0.52305889 | 0.25085 | 1.522604422 | 14.14677974 | 14.14677974 | 1.93899773 | 0.494915194 | 0.4977 | 0.212838056 | 1.938997729 | 1.938997729 | 0.785253329 | -0.104990214 | FALSE | FALSE | FALSE |
| LFNG | 1.5221156 | 0.5231931 | 0.96975 | 24.36085008 | 97.49393902 | 390.2754875 | 0.4317603 | 0.780424636 | 1 | 6.907125353 | 110.6078885 | 547.8908534 | 3.525371765 | 0.547204922 | FALSE | TRUE | FALSE |
| OR1L3 | 1.5213442 | 0.52332724 | 0.7497 | 1.521344176 | 14.20640362 | 14.20640362 | 0.21201442 | 0.849072052 | 0.51205 | 1.940167129 | 1.940167129 | 1.940167129 | 7.175663829 | 0.855862084 | FALSE | TRUE | FALSE |
| SF3A1 | 1.5205691 | 0.5234613 | 0.5128 | 1.520569063 | 1.520569063 | 12.84715646 | 0.41777527 | 0.785437167 | 1 | 3.933244835 | 3.933244835 | 77.52307068 | 3.639681832 | 0.561063421 | FALSE | TRUE | FALSE |
| SOHLH2 | 1.5199804 | 0.52359527 | 0.50605 | 1.519980351 | 1.519980351 | 7.540960187 | 0.43027511 | 0.78114268 | 0.00955 | 0.430275107 | 0.430275107 | 0.430275107 | 3.532577938 | 0.548091752 | FALSE | TRUE | FALSE |
| ZCCHC17 | 1.5193689 | 0.52372917 | 0.0067 | 1.519368933 | 1.519368933 | 1.519368933 | 0.21180415 | 0.849128401 | 1 | 0.211804146 | 0.211804146 | 0.211804146 | 7.173461713 | 0.855728785 | FALSE | TRUE | FALSE |
| FTL | 1.5141845 | 0.52386305 | 0.00575 | 1.514184503 | 1.514184503 | 1.514184503 | 0.82869078 | 0.612941177 | 0.4981 | 0.207208013 | 0.828690781 | 0.828690781 | 1.827200854 | 0.26178629 | FALSE | TRUE | FALSE |
| TMEM192 | 1.5140367 | 0.52399685 | 0.5051 | 1.514036675 | 1.514036675 | 1.514036675 | 0.21132103 | 0.849240974 | 1 | 0.211321031 | 0.211321031 | 0.211321031 | 7.164628459 | 0.855193674 | FALSE | TRUE | FALSE |
| GOLPH3 | 1.512541 | 0.52413058 | 0.7502 | 1.512541041 | 6.050531412 | 6.050531412 | 0.43402692 | 0.779701704 | 0.00735 | 0.434026919 | 0.434026919 | 0.434026919 | 3.484901456 | 0.542190502 | FALSE | TRUE | FALSE |
| ZCWPW2 | 1.5121083 | 0.52426423 | 0.7568 | 1.512108302 | 14.12779036 | 14.12779036 | 0.20447291 | 0.850923867 | 0.7487 | 1.870390721 | 17.55998807 | 17.55998807 | 7.395152208 | 0.868947117 | FALSE | TRUE | FALSE |
| SHF | 1.5092026 | 0.52439784 | 0.7544 | 1.509202552 | 14.14063415 | 14.14063415 | 1.86846518 | 0.501607216 | 0.7503 | 1.868465185 | 17.58609658 | 17.58609658 | 0.807723133 | -0.092737479 | FALSE | FALSE | FALSE |
| USP14 | 1.5082904 | 0.52453137 | 0.5066 | 1.508290376 | 1.508290376 | 7.47616881 | 0.42782666 | 0.781826242 | 1 | 0.427826659 | 0.427826659 | 8.490828796 | 3.525470761 | 0.547217117 | FALSE | TRUE | FALSE |
| CRIP1 | 1.5077084 | 0.52466482 | 0.5019 | 1.507708371 | 1.507708371 | 1.507708371 | 0.20800856 | 0.850205081 | 1 | 0.208008556 | 0.208008556 | 0.208008556 | 7.248299797 | 0.860236148 | FALSE | TRUE | FALSE |
| SGPP2 | 1.5070911 | 0.52479819 | 0.25395 | 1.507091066 | 6.027186633 | 6.027186633 | 0.21064799 | 0.849423559 | 1 | 0.210647992 | 0.210647992 | 0.210647992 | 7.154547519 | 0.854582173 | FALSE | TRUE | FALSE |
| AGPS | 1.5066842 | 0.52493147 | 0.25995 | 0.376802641 | 1.506684161 | 7.462639537 | 0.12612484 | 0.86984761 | 0.5093 | 0.504234602 | 0.504234602 | 2.500850611 | 11.94597486 | 1.077221597 | FALSE | TRUE | FALSE |
| TPD52L1 | 1.5053141 | 0.52506469 | 0.00475 | 0.376330246 | 1.505314137 | 1.505314137 | 0.42163093 | 0.78396707 | 1 | 1.686516941 | 1.686516941 | 1.686516941 | 3.570217554 | 0.552694681 | FALSE | TRUE | FALSE |
| SIM2 | 1.5019717 | 0.52519788 | 0.0174 | 1.501971659 | 1.501971659 | 5.455054075 | 0.20621236 | 0.850606739 | 1 | 0.206212363 | 0.206212363 | 4.076677226 | 7.283615961 | 0.862347039 | FALSE | TRUE | FALSE |
| ESD | 1.5017159 | 0.52533097 | 0.5082 | 1.501715865 | 1.501715865 | 1.501715865 | 1.86254299 | 0.502392986 | 0.50235 | 1.862542987 | 1.862542987 | 1.862542987 | 0.806271788 | -0.093518536 | FALSE | FALSE | FALSE |
| ITGA4 | 1.4929113 | 0.52546411 | 0.3869 | 0.668236892 | 5.972257309 | 29.27073711 | 0.06354914 | 0.88388664 | 1 | 0.063549141 | 0.063549141 | 1.248927459 | 23.49223374 | 1.370924313 | FALSE | TRUE | FALSE |
| FPGS | 1.4891988 | 0.52559722 | 0.50505 | 1.489198799 | 5.956383485 | 12.81742715 | 0.20933433 | 0.849899002 | 0.0157 | 0.209334329 | 0.209334329 | 4.138185061 | 7.113973163 | 0.852112222 | FALSE | TRUE | FALSE |
| FRA10AC1 | 1.4890583 | 0.52573024 | 0.5038 | 1.48905833 | 5.956075753 | 5.956075753 | 0.20433736 | 0.850992638 | 1 | 1.865441759 | 1.865441759 | 1.865441759 | 7.287254406 | 0.862563931 | FALSE | TRUE | FALSE |
| TXNL4B | 1.4888966 | 0.52586318 | 0.751 | 1.488896623 | 5.956916322 | 5.956916322 | 3.80470615 | 0.366254143 | 0.50655 | 3.804706152 | 15.21852258 | 15.21852258 | 0.391330254 | -0.407456576 | FALSE | FALSE | FALSE |
| ALDH9A1 | 1.4878406 | 0.52599605 | 0.8013 | 10.49213172 | 380.8869338 | 1524.498922 | 0.29926264 | 0.825707738 | 0.8175 | 1.196738607 | 4.786902842 | 19.15218834 | 4.971688623 | 0.696503921 | FALSE | TRUE | FALSE |
| NKX2-4 | 1.4865708 | 0.52612884 | 0.7499 | 1.48657082 | 5.946516172 | 5.946516172 | 0.41404842 | 0.786914804 | 0.50765 | 3.909559864 | 3.909559864 | 3.909559864 | 3.590330891 | 0.555134476 | FALSE | TRUE | FALSE |
| EPHX4 | 1.484862 | 0.52626157 | 0.00985 | 1.484862007 | 1.484862007 | 1.484862007 | 0.42175647 | 0.783850884 | 0.0104 | 0.421756474 | 0.421756474 | 8.382039478 | 3.52066204 | 0.546624338 | FALSE | TRUE | FALSE |
| CBR4 | 1.4840897 | 0.52639423 | 0.50135 | 1.484089672 | 1.484089672 | 1.484089672 | 0.77998162 | 0.628598247 | 0.7504 | 1.826806611 | 7.304786909 | 7.304786909 | 1.902723908 | 0.279375775 | FALSE | TRUE | FALSE |
| HSF2 | 1.4830722 | 0.52652682 | 0.50825 | 1.483072156 | 1.483072156 | 7.348642016 | 0.41700098 | 0.785780047 | 1 | 1.667835634 | 1.667835634 | 8.273091776 | 3.556519567 | 0.551025202 | FALSE | TRUE | FALSE |
| IFIT2 | 1.4823293 | 0.52665932 | 0.8128 | 5.928795352 | 222.959586 | 222.959586 | 1.92095345 | 0.497164863 | 0.5154 | 1.920953449 | 7.683245951 | 7.683245951 | 0.771663316 | -0.112572145 | FALSE | FALSE | FALSE |
| DERL1 | 1.478246 | 0.52679181 | 0.87595 | 5.914226644 | 23.66779123 | 23.66779123 | 0.41176114 | 0.787757728 | 1 | 3.840650604 | 15.3621775 | 15.3621775 | 3.590057069 | 0.555101352 | FALSE | TRUE | FALSE |
| TTC39C | 1.4753889 | 0.52692424 | 0.5103 | 1.475388862 | 1.475388862 | 5.439978356 | 0.20518525 | 0.850772355 | 1 | 0.205185251 | 0.205185251 | 4.071084955 | 7.190521031 | 0.856760361 | FALSE | TRUE | FALSE |
| ANKRD54 | 1.4724604 | 0.52705664 | 0.75215 | 3.451133393 | 13.80284125 | 13.80284125 | 0.43222935 | 0.780244355 | 0.00595 | 0.432229347 | 0.432229347 | 0.432229347 | 3.406664509 | 0.532329366 | FALSE | TRUE | FALSE |
| TYMS | 1.4715951 | 0.52718895 | 0.5018 | 1.471595052 | 1.471595052 | 1.471595052 | 0.41890551 | 0.784891931 | 1 | 0.418905505 | 0.418905505 | 0.418905505 | 3.512952285 | 0.545672251 | FALSE | TRUE | FALSE |
| SLC25A32 | 1.4692394 | 0.52732122 | 0.50455 | 1.469239436 | 5.877800022 | 5.877800022 | 0.12293193 | 0.870640128 | 0.7458 | 0.491574885 | 1.966179879 | 1.966179879 | 11.95165069 | 1.077427892 | FALSE | TRUE | FALSE |
| ZNF407 | 1.4685552 | 0.52745341 | 0.52765 | 1.468555249 | 2.989648237 | 28.07454047 | 0.20085964 | 0.851908183 | 1 | 0.200859636 | 3.860330874 | 6.734452429 | 7.311350732 | 0.863997618 | FALSE | TRUE | FALSE |
| SMCO1 | 1.4676084 | 0.52758553 | 0.94035 | 13.6238461 | 1266.062866 | 1266.062866 | 1.93628648 | 0.495237767 | 0.74755 | 1.93628648 | 18.10671165 | 18.10671165 | 0.757950058 | -0.12035941 | FALSE | FALSE | FALSE |
| CEACAM3 | 1.466962 | 0.52771757 | 0.49945 | 0.161774025 | 1.466961967 | 1.466961967 | 0.20575716 | 0.850675787 | 1 | 0.205757156 | 0.205757156 | 0.205757156 | 7.129579346 | 0.853063907 | FALSE | TRUE | FALSE |
| TNK2 | 1.4667868 | 0.52784952 | 0.3658 | 1.466786822 | 130.9573787 | 1282.384944 | 0.00508253 | 0.898105639 | 1 | 0.005082528 | 0.005082528 | 0.099050802 | 288.5939692 | 2.460287251 | FALSE | TRUE | FALSE |
| ARID3C | 1.4622368 | 0.52798146 | 0.50095 | 1.462236816 | 1.462236816 | 3.179766571 | 15.989004 | 0.153707916 | 0.13305 | 15.98900397 | 153.7100934 | 153.7100934 | 0.091452652 | -1.038803696 | FALSE | FALSE | FALSE |
| WFDC1 | 1.4600181 | 0.52811334 | 0.50875 | 1.460018079 | 1.460018079 | 1.460018079 | 0.17884282 | 0.857321831 | 1 | 15.30548966 | 588.4654101 | 588.4654101 | 8.163694019 | 0.911886719 | FALSE | TRUE | FALSE |
| OR1S2 | 1.4580462 | 0.52824517 | 0.75275 | 1.458046214 | 13.65846022 | 13.65846022 | 1.87013593 | 0.500976769 | 0.5045 | 1.870135935 | 1.870135935 | 1.870135935 | 0.779647183 | -0.108101886 | FALSE | FALSE | FALSE |
| MKRN1 | 1.4574083 | 0.52837691 | 0.5061 | 1.457408332 | 1.457408332 | 7.223200316 | 0.41533525 | 0.786434068 | 1 | 0.415335248 | 0.415335248 | 8.241512507 | 3.508992651 | 0.545182459 | FALSE | TRUE | FALSE |
| GPR20 | 1.4551333 | 0.52850861 | 0.75165 | 3.397998028 | 13.59135019 | 13.74328668 | 1.67219924 | 0.520290084 | 0.75035 | 1.672199242 | 6.690165449 | 8.280968074 | 0.870191333 | -0.060385247 | FALSE | FALSE | FALSE |
| RPUSD2 | 1.4542295 | 0.52864023 | 1 | 1.92044E+30 | 3.50599E+37 | 1.80485E+41 | 27.4555627 | 0.10137668 | 1 | 2.52168E+26 | 3.43314E+33 | 9.21915E+35 | 0.052966663 | -1.275997385 | FALSE | FALSE | FALSE |
| PDCD2L | 1.4541957 | 0.52877177 | 0.5023 | 3.279723244 | 52.50101281 | 52.50101281 | 10.1545681 | 0.196577392 | 0.24655 | 1.09115293 | 10.15456812 | 10.15456812 | 0.143206059 | -0.844038607 | FALSE | FALSE | FALSE |
| KANK4 | 1.4540949 | 0.52890322 | 0.00685 | 0.043073727 | 0.043073727 | 0.844467022 | 0.06211928 | 0.884281596 | 0.02515 | 0.06211928 | 0.06211928 | 1.220753548 | 23.40810958 | 1.369366342 | FALSE | TRUE | FALSE |
| KLF17 | 1.4523639 | 0.52903461 | 0.8762 | 13.57036566 | 130.0532122 | 130.0532122 | 0.20976188 | 0.84977344 | 0.5087 | 1.922282091 | 1.922282091 | 4.16336472 | 6.923869499 | 0.840348874 | FALSE | TRUE | FALSE |
| ZNF555 | 1.4507586 | 0.52916594 | 0.6924 | 1.450758564 | 123.2629043 | 123.2629043 | 0.03142578 | 0.890883324 | 0.75825 | 0.27508085 | 2.471409018 | 5.446244991 | 46.16459684 | 1.664309047 | FALSE | TRUE | FALSE |
| LRRIQ1 | 1.4495603 | 0.5292972 | 0.36255 | 0.17215329 | 12.50516623 | 110.4659714 | 0.45030871 | 0.774277602 | 0.51275 | 0.450308711 | 3.875206574 | 34.1663773 | 3.219036789 | 0.50772594 | FALSE | TRUE | FALSE |
| SFXN5 | 1.4484018 | 0.52942839 | 0.5051 | 1.448401782 | 1.448401782 | 3.140064559 | 0.19622219 | 0.853014087 | 1 | 1.802567178 | 1.802567178 | 3.895051871 | 7.38143742 | 0.868140942 | FALSE | TRUE | FALSE |
| TMEM143 | 1.4476449 | 0.52955951 | 0.0171 | 1.447644945 | 1.447644945 | 3.135846219 | 15.3550211 | 0.156923122 | 0.69655 | 15.35502107 | 1454.13227 | 1454.13227 | 0.094278278 | -1.025588358 | FALSE | FALSE | FALSE |
| PIP4K2C | 1.4466851 | 0.52969055 | 0.986 | 131.3196832 | 13159.97953 | 136518.8437 | 0.22824539 | 0.84535026 | 1 | 0.228245392 | 2.110032079 | 4.526480316 | 6.338288354 | 0.801971993 | FALSE | TRUE | FALSE |
| SAG | 1.4466618 | 0.52982151 | 0.75785 | 5.786491087 | 54.04675459 | 54.04675459 | 1.81918172 | 0.505667091 | 0.75205 | 1.81918172 | 17.07443684 | 17.07443684 | 0.795226656 | -0.099509071 | FALSE | FALSE | FALSE |
| VPS8 | 1.4457767 | 0.5299524 | 0.8902 | 13.46307221 | 1256.695834 | 12582.54263 | 0.27985763 | 0.83109417 | 0.87755 | 2.567815436 | 24.1661819 | 50.37214746 | 5.166114936 | 0.713164064 | FALSE | TRUE | FALSE |
| ZNF621 | 1.4447703 | 0.53008321 | 0.8777 | 13.50009664 | 129.3863275 | 129.3863275 | 0.21528189 | 0.848293138 | 1 | 0.215281889 | 0.215281889 | 0.215281889 | 6.711062726 | 0.826791298 | FALSE | TRUE | FALSE |
| ACTR3 | 1.4430804 | 0.53021397 | 0.50855 | 1.443080386 | 1.443080386 | 1.443080386 | 0.4118379 | 0.787673747 | 1 | 0.411837902 | 0.411837902 | 0.411837902 | 3.504000913 | 0.544564211 | FALSE | TRUE | FALSE |
| TSNARE1 | 1.440965 | 0.53034467 | 0.0125 | 0.15461058 | 1.44096497 | 3.047546816 | 0.19537529 | 0.853161244 | 0.7548 | 0.195375291 | 1.830225417 | 3.857759456 | 7.375369544 | 0.867783786 | FALSE | TRUE | FALSE |
| SLC1A4 | 1.4355389 | 0.53047537 | 0.5054 | 1.43553894 | 1.43553894 | 5.210408607 | 0.19785972 | 0.85257094 | 0.01255 | 0.197859721 | 0.197859721 | 3.918125739 | 7.255336938 | 0.860657586 | FALSE | TRUE | FALSE |
| GDF6 | 1.4350916 | 0.53060598 | 0.5059 | 1.435091646 | 1.435091646 | 7.099254457 | 1.62021263 | 0.523787782 | 0.5043 | 1.620212626 | 1.620212626 | 8.025270373 | 0.885742786 | -0.052692376 | FALSE | FALSE | FALSE |
| SERPINB3 | 1.4344825 | 0.53073652 | 0.50505 | 1.434482502 | 13.37305378 | 13.37305378 | 0.25336797 | 0.838492053 | 0.5005 | 0.564174366 | 21.80922824 | 21.80922824 | 5.661656799 | 0.75294354 | FALSE | TRUE | FALSE |
| GRM2 | 1.4314081 | 0.53086703 | 0.23725 | 0.358117992 | 5.722808941 | 22.88566072 | 0.19942121 | 0.852219985 | 0.5121 | 0.199421211 | 0.976959628 | 3.903815975 | 7.177812616 | 0.855992116 | FALSE | TRUE | FALSE |

| | | | | | | | | | | | | | | | | | |
|---|---|---|---|---|---|---|---|---|---|---|---|---|---|---|---|---|---|
| HDHD3 | 1.4311352 | 0.53099745 | 0.93825 | 13.43010411 | 1275.38211 | 1275.38211 | 1610.1172 | 0.001488868 | 0.0629 | 16.80862252 | 1610.117203 | 1610.117203 | 0.000888839 | -3.051176831 | FALSE | FALSE | FALSE |
| GDF1 | 1.4295772 | 0.5311278 | 0.5033 | 3.242111926 | 120.625078 | 120.625078 | 4.60724202 | 0.323144731 | 0.6912 | 4.60724202 | 414.3265495 | 414.3265495 | 0.310289152 | -0.508233408 | FALSE | FALSE | FALSE |
| GTF2H4 | 1.4294679 | 0.53125808 | 0.5055 | 1.429467924 | 1.429467924 | 5.27358183 | 7.05230912 | 0.243419229 | 0.01035 | 1.763261965 | 7.052309123 | 15.15791397 | 0.202695018 | -0.693156926 | FALSE | FALSE | FALSE |
| SRC | 1.4287343 | 0.53138828 | 0.50635 | 1.428734349 | 1.428734349 | 12.10115528 | 0.40824045 | 0.789174889 | 1 | 0.40824045 | 0.40824045 | 8.068287174 | 3.499737347 | 0.544035452 | FALSE | TRUE | FALSE |
| PIGK | 1.4250721 | 0.53151845 | 0.5111 | 1.425072083 | 1.425072083 | 3.098163722 | 0.19972925 | 0.852165847 | 1 | 0.199729251 | 0.199729251 | 0.199729251 | 7.13501941 | 0.853395159 | FALSE | TRUE | FALSE |
| FKBP14 | 1.4219113 | 0.53164858 | 0.96955 | 13.42399814 | 1290.199181 | 13118.74376 | 17.9513455 | 0.14383117 | 0.49865 | 1.891110802 | 174.7763494 | 174.7763494 | 0.079209175 | -1.101224511 | FALSE | FALSE | FALSE |
| DDX52 | 1.4213623 | 0.53177864 | 0.2566 | 0.159227639 | 1.42136233 | 3.150124376 | 0.0306332 | 0.891113932 | 1 | 0.030633204 | 0.030633204 | 0.606828731 | 46.39940162 | 1.66651238 | FALSE | TRUE | FALSE |
| POLE2 | 1.4200618 | 0.53190863 | 0.75545 | 1.420061776 | 13.25675646 | 28.13888377 | 0.81385731 | 0.616278369 | 0.5087 | 0.813857306 | 0.813857306 | 4.036257155 | 1.744853509 | 0.241758971 | FALSE | TRUE | FALSE |
| C1QTNF9 | 1.4174957 | 0.53203857 | 0.25665 | 1.417495727 | 13.51517552 | 13.51517552 | 0.20257465 | 0.851513239 | 1 | 0.202574651 | 0.202574651 | 0.202574651 | 6.997399339 | 0.84493666 | FALSE | TRUE | FALSE |
| OSGIN2 | 1.4165425 | 0.53216844 | 0.2602 | 1.416542515 | 13.29486611 | 28.06571603 | 0.19246685 | 0.853747149 | 1 | 1.767655047 | 16.66222316 | 35.06167131 | 7.359929689 | 0.866873665 | FALSE | TRUE | FALSE |
| XAF1 | 1.4162841 | 0.53229823 | 0.4983 | 0.156161489 | 1.41628414 | 1.41628414 | 0.19945006 | 0.852206454 | 1 | 0.199450062 | 0.199450062 | 0.199450062 | 7.100946114 | 0.851316217 | FALSE | TRUE | FALSE |
| AP5M1 | 1.4160023 | 0.53242795 | 0.50605 | 1.416002279 | 1.416002279 | 3.091683714 | 0.19924987 | 0.852314631 | 1 | 0.199249872 | 0.199249872 | 3.955364956 | 7.106665932 | 0.851665901 | FALSE | TRUE | FALSE |
| SLC4A3 | 1.4134077 | 0.53255761 | 0.374 | 0.818592765 | 31.05119186 | 124.1313668 | 0.34753385 | 0.809823761 | 0.1552 | 0.08692366 | 0.34753385 | 1.689349244 | 4.066964227 | 0.609270352 | FALSE | TRUE | FALSE |
| TACR3 | 1.4121426 | 0.53268721 | 0.875 | 13.3947471 | 130.3160064 | 130.3160064 | 1.76598436 | 0.511645155 | 0.5007 | 3.774058503 | 163.9950337 | 163.9950337 | 0.799634826 | -0.0971083 | FALSE | FALSE | FALSE |
| FEM1C | 1.4111439 | 0.53281674 | 0.75245 | 1.411143863 | 5.645250653 | 27.90448199 | 0.40827637 | 0.789119685 | 1 | 0.408276373 | 0.408276373 | 8.084365975 | 3.456344666 | 0.538617044 | FALSE | TRUE | FALSE |
| DNM1 | 1.40948 | 0.53294621 | 0.75995 | 3.330007315 | 13.31888881 | 65.26345239 | 15.0838286 | 0.158226868 | 0.26365 | 3.771241961 | 15.08382856 | 74.08968628 | 0.09344312 | -1.02945267 | FALSE | FALSE | FALSE |
| GALNT12 | 1.4062314 | 0.53307565 | 0.5069 | 1.406231445 | 13.25437604 | 13.25437604 | 0.23898283 | 0.84232492 | 0.5004 | 0.238982833 | 0.238982833 | 1.184762415 | 5.884236233 | 0.7696901 | FALSE | TRUE | FALSE |
| GRXCR1 | 1.4035627 | 0.53320504 | 0.753 | 1.40356274 | 13.12724475 | 13.12724475 | 0.20340857 | 0.851253388 | 0.009 | 0.203408566 | 0.203408566 | 0.203408566 | 6.900214522 | 0.838862593 | FALSE | TRUE | FALSE |
| SEMA3E | 1.4029037 | 0.53333435 | 0.0217 | 1.402903703 | 1.402903703 | 27.62363923 | 0.19551387 | 0.853121137 | 0.01985 | 0.195513869 | 0.195513869 | 3.856907735 | 7.175468981 | 0.855850291 | FALSE | TRUE | FALSE |
| CHCHD5 | 1.4025956 | 0.53346359 | 0.74975 | 3.245564039 | 12.9820234 | 12.9820234 | 0.37654711 | 0.800365073 | 1 | 33.18585536 | 323.7204209 | 323.7204209 | 3.724887391 | 0.571113148 | FALSE | TRUE | FALSE |
| PSMA8 | 1.4025529 | 0.53359274 | 0.876 | 13.19628954 | 127.3488565 | 127.3488565 | 0.20882442 | 0.850024358 | 1 | 0.208824418 | 0.208824418 | 0.208824418 | 6.716421977 | 0.827137974 | FALSE | TRUE | FALSE |
| DMXL1 | 1.3993355 | 0.53372186 | 0.8942 | 13.36941933 | 1313.604571 | 13493.39681 | 10.2703995 | 0.19589814 | 0.1513 | 1.093016597 | 10.27039951 | 85.59575678 | 0.136249373 | -0.865665486 | FALSE | FALSE | FALSE |
| RAB3D | 1.3976586 | 0.53385092 | 0.512 | 1.397658608 | 5.591286025 | 5.591286025 | 0.12030939 | 0.871163482 | 0.00745 | 0.120309391 | 0.120309391 | 0.120309391 | 11.61720292 | 1.065101575 | FALSE | TRUE | FALSE |
| OBSCN | 1.3971453 | 0.5339799 | 0.40705 | 0.179603646 | 12244.3939 | 3746485.289 | 0.00019351 | 0.9000358 | 0.6647 | 0.011033542 | 366.8802961 | 26510.33313 | 7220.111655 | 3.858543914 | FALSE | TRUE | FALSE |
| CRY2 | 1.3966968 | 0.53410881 | 0.51145 | 1.396696797 | 1.396696797 | 27.49255077 | 0.18175451 | 0.856610282 | 1 | 1.695833665 | 16.23937474 | 33.44424551 | 7.684523278 | 0.88561693 | FALSE | TRUE | FALSE |
| CDC23 | 1.3960922 | 0.53423764 | 0.75095 | 3.263333194 | 13.05283989 | 27.57856297 | 1.58045329 | 0.528523163 | 0.76075 | 3.701027511 | 14.80361697 | 14.80361697 | 0.883349218 | -0.053867571 | FALSE | FALSE | FALSE |
| PIGP | 1.3959703 | 0.53436639 | 0.49985 | 0.154654371 | 1.39597032 | 1.39597032 | 0.19746745 | 0.852705503 | 0.00315 | 0.197467455 | 0.197467455 | 0.197467455 | 7.069369083 | 0.849380656 | FALSE | TRUE | FALSE |
| HTT | 1.3953041 | 0.53449506 | 0.99025 | 9045.026633 | 4787091.914 | 59488189.17 | 108103.188 | 1.80832E-05 | 0.70505 | 12390946.14 | 9237350965 | 1.07732E+11 | 1.29072E-05 | -4.889169634 | FALSE | FALSE | FALSE |
| FSIP2 | 1.3938229 | 0.53462368 | 0.79105 | 22.4607661 | 14595.39233 | 284977.4402 | 0.02263637 | 0.8930391 | 0.86615 | 0.18060294 | 25.38929795 | 241.9679038 | 61.57449297 | 1.789400844 | FALSE | TRUE | FALSE |
| CCDC170 | 1.3931869 | 0.53475222 | 0.51125 | 1.393186907 | 12.57492951 | 12.57492951 | 0.26637569 | 0.834774477 | 0.5075 | 0.266375685 | 0.266375685 | 1.037394807 | 5.230157944 | 0.718514804 | FALSE | TRUE | FALSE |
| PTOV1 | 1.3929602 | 0.53488068 | 0.5064 | 1.392960176 | 1.392960176 | 6.898516295 | 0.02064773 | 0.893696503 | 0.0118 | 0.020647731 | 0.020647731 | 0.409491183 | 67.46311001 | 1.829066358 | FALSE | TRUE | FALSE |
| SEC61A1 | 1.3902636 | 0.53500909 | 0.50145 | 1.390263609 | 1.390263609 | 11.77312963 | 0.39836725 | 0.792646692 | 1 | 0.398367253 | 0.398367253 | 7.895163685 | 3.489904352 | 0.542813524 | FALSE | TRUE | FALSE |
| KCNJ8 | 1.3900771 | 0.53513743 | 0.5041 | 1.390077114 | 1.390077114 | 2.931564122 | 0.19145751 | 0.853998774 | 0.01115 | 0.191457514 | 0.191457514 | 3.797145662 | 7.260499128 | 0.860966478 | FALSE | TRUE | FALSE |
| DDX43 | 1.3889985 | 0.5352657 | 0.2586 | 1.38899847 | 13.10983051 | 27.41364748 | 1.79525551 | 0.508597517 | 0.5084 | 1.795255508 | 1.795255508 | 6.574845749 | 0.773705171 | -0.111424501 | FALSE | FALSE | FALSE |
| NCKAP1 | 1.3888958 | 0.53539389 | 0.5158 | 1.388895776 | 1.388895776 | 27.22107627 | 0.39801628 | 0.792753298 | 1 | 0.39801628 | 0.39801628 | 7.819584937 | 3.489545142 | 0.542768821 | FALSE | TRUE | FALSE |
| SLC9A2 | 1.3877436 | 0.53552201 | 0.02375 | 1.387743568 | 1.387743568 | 27.2620127 | 1.73651645 | 0.513907878 | 0.51065 | 1.736516453 | 1.736516453 | 34.18653938 | 0.799153711 | -0.097369679 | FALSE | FALSE | FALSE |
| DCUN1D2 | 1.3858465 | 0.53565008 | 0.50035 | 1.385846524 | 1.385846524 | 1.385846524 | 1.73318772 | 0.514807538 | 0.5028 | 1.733187721 | 1.733187721 | 1.733187721 | 0.799594012 | -0.097130467 | FALSE | FALSE | FALSE |
| HRG | 1.3854686 | 0.53577807 | 0.5113 | 1.385468613 | 1.385468613 | 3.019595972 | 145.901587 | 0.019347394 | 0.007 | 15.20652111 | 145.9015867 | 145.9015867 | 0.009495912 | -2.022463323 | FALSE | FALSE | FALSE |
| APCS | 1.3850566 | 0.53590598 | 0.4979 | 0.346278954 | 1.385056563 | 1.385056563 | 0.39636221 | 0.793284681 | 1 | 0.396362209 | 0.396362209 | 0.396362209 | 3.494421345 | 0.543375269 | FALSE | TRUE | FALSE |
| SSTR4 | 1.3818621 | 0.53603386 | 0.5064 | 1.381862105 | 1.381862105 | 2.968229329 | 0.19352026 | 0.853481371 | 0.0137 | 0.193520262 | 0.193520262 | 3.825823364 | 7.140658496 | 0.853738263 | FALSE | TRUE | FALSE |
| DCBLD1 | 1.3800869 | 0.53616168 | 1 | 4.2333E+46 | 4.96845E+56 | 4.65107E+60 | 983.740055 | 0.002487091 | 1 | 8.81954E+47 | 4.41303E+57 | 1.47889E+62 | 0.001402898 | -2.852973908 | FALSE | FALSE | FALSE |
| ZNF623 | 1.3778501 | 0.53628945 | 0.5055 | 1.377850075 | 1.377850075 | 2.988390765 | 0.18798702 | 0.855036122 | 0.4989 | 0.187987025 | 1.724545059 | 3.727903655 | 7.329495627 | 0.86507409 | FALSE | TRUE | FALSE |
| PGAM2 | 1.3772844 | 0.53641715 | 0.7824 | 5.506924115 | 125.9196953 | 503.4763992 | 123.0217 | 0.023169019 | 0.09715 | 8.381232201 | 134.1100917 | 536.662174 | 0.011195459 | -1.950958087 | FALSE | FALSE | FALSE |
| RBM23 | 1.3758082 | 0.53654479 | 0.50805 | 1.375808249 | 12.85632029 | 12.85632029 | 2.10245632 | 0.479988579 | 0.5062 | 19.76231373 | 190.4123214 | 1879.491167 | 0.654381372 | -0.184169072 | FALSE | FALSE | FALSE |
| SLC25A42 | 1.3752511 | 0.53667236 | 0.7461 | 1.375251143 | 5.50171177 | 6.815927583 | 0.39890452 | 0.792433137 | 0.00955 | 0.398904516 | 0.398904516 | 0.398904516 | 3.447569747 | 0.537513061 | FALSE | TRUE | FALSE |
| SPA17 | 1.3745034 | 0.53679985 | 0.75165 | 1.374503366 | 5.498999241 | 5.498999241 | 0.39824144 | 0.792700009 | 1 | 0.398241439 | 0.398241439 | 0.398241439 | 3.451432303 | 0.537999359 | FALSE | TRUE | FALSE |
| FETUB | 1.373703 | 0.53692727 | 0.75155 | 1.373703026 | 12.79739801 | 12.79739801 | 130.980082 | 0.021801345 | 0.5018 | 1283.832496 | 132530.1501 | 132530.1501 | 0.010487877 | -1.979312403 | FALSE | FALSE | FALSE |
| C12orf42 | 1.3714729 | 0.53705465 | 0.5074 | 12.74248566 | 121.432998 | 121.432998 | 16.5350124 | 0.149857382 | 0.25225 | 1.768412095 | 16.53501235 | 16.53501235 | 0.082943566 | -1.081217299 | FALSE | FALSE | FALSE |
| TMED8 | 1.3692623 | 0.53718197 | 0.50355 | 1.369262326 | 1.369262326 | 1.369262326 | 0.39239025 | 0.794731552 | 1 | 0.392390253 | 0.392390253 | 0.392390253 | 3.489542148 | 0.542768448 | FALSE | TRUE | FALSE |
| PPEF2 | 1.3676578 | 0.53730924 | 0.8159 | 12.49635224 | 468.1097201 | 4491.735917 | 182.852254 | 0.014853022 | 0.23195 | 19.3605415 | 17526.81152 | 35134.8108 | 0.007479579 | -2.126122869 | FALSE | FALSE | FALSE |
| SPOCK1 | 1.3659236 | 0.53743644 | 0.01195 | 1.365923625 | 1.365923625 | 6.769878896 | 0.39198359 | 0.794888129 | 1 | 0.391983589 | 0.391983589 | 7.778695492 | 3.484644922 | 0.542158531 | FALSE | TRUE | FALSE |
| ADAP2 | 1.3658602 | 0.53756357 | 0.5075 | 1.365860236 | 1.365860236 | 2.984892228 | 0.19301857 | 0.853627664 | 1 | 0.193018572 | 0.193018572 | 3.834608741 | 7.076315092 | 0.849807163 | FALSE | TRUE | FALSE |
| NOP58 | 1.3654275 | 0.53769063 | 0.5098 | 1.365427535 | 1.365427535 | 2.917148448 | 0.19026409 | 0.854249584 | 1 | 0.190264088 | 0.190264088 | 3.774020257 | 7.176485856 | 0.855911833 | FALSE | TRUE | FALSE |
| TSPAN15 | 1.3653982 | 0.5378176 | 0.65695 | 5.461829087 | 87.46235459 | 350.1268258 | 112.984535 | 0.02583055 | 0.5025 | 112.984535 | 1811.227029 | 1811.227029 | 0.012084824 | -1.91775968 | FALSE | FALSE | FALSE |
| KRTAP10-11 | 1.3645722 | 0.5379445 | 0.50675 | 1.364572165 | 1.364572165 | 1.364572165 | 1.70611953 | 0.516448783 | 0.50745 | 1.706119529 | 1.706119529 | 1.706119529 | 0.799810413 | -0.097012946 | FALSE | FALSE | FALSE |
| KIAA1614 | 1.3645696 | 0.53807132 | 0.0286 | 0.153351503 | 1.364569583 | 26.51656513 | 19.6921064 | 0.13416525 | 0.1388 | 2.137507562 | 19.69210635 | 41.67860307 | 0.069295258 | -1.159296486 | FALSE | FALSE | FALSE |
| TSFM | 1.3641589 | 0.53819807 | 0.5037 | 1.364158866 | 1.364158866 | 1.364158866 | 15.7899771 | 0.155314355 | 0.2524 | 1.659392079 | 15.78997708 | 15.78997708 | 0.086393974 | -1.06351655 | FALSE | FALSE | FALSE |
| CLCA1 | 1.363696 | 0.53832474 | 0.6925 | 5.454004085 | 117.0886597 | 468.2876863 | 0.02955234 | 0.891343658 | 0.8769 | 1.039473596 | 9.382928004 | 20.48700105 | 46.14511207 | 1.664125705 | FALSE | TRUE | FALSE |
| CENPV | 1.3604567 | 0.53845138 | 0.99555 | 1179.519711 | 121133.3838 | 1271061.979 | 155666.522 | 1.26469E-05 | 0.03695 | 1499.691452 | 155666.5222 | 1642136.337 | 8.73956E-06 | -5.058510513 | FALSE | FALSE | FALSE |
| KCNK17 | 1.3603 | 0.53857794 | 0.8739 | 12.69691451 | 121.5557595 | 121.5557595 | 1.75771653 | 0.51240153 | 0.2492 | 1.757716532 | 16.50236276 | 16.50236276 | 0.773901827 | -0.111314128 | FALSE | FALSE | FALSE |
| POLA2 | 1.3562124 | 0.53870447 | 0.5038 | 1.356212351 | 1.356212351 | 4.992810002 | 0.19043045 | 0.854236404 | 1 | 0.19043045 | 0.19043045 | 3.76926707 | 7.121825047 | 0.852591301 | FALSE | TRUE | FALSE |
| PLEKHA1 | 1.3556998 | 0.53883094 | 0.5114 | 1.355699763 | 1.355699763 | 1.355699763 | 143.40702 | 0.019628627 | 0.00335 | 1.595288808 | 143.4070201 | 143.4070201 | 0.00945351 | -2.024406891 | FALSE | FALSE | FALSE |
| ADK | 1.3552948 | 0.53895733 | 0.87465 | 13.05299554 | 128.9399255 | 128.9399255 | 0.19332688 | 0.853534601 | 1 | 1.820059227 | 1.820059227 | 1.820059227 | 7.010379616 | 0.845741536 | FALSE | TRUE | FALSE |
| LRR1 | 1.3547856 | 0.53908364 | 0.87525 | 2.950750726 | 121.5437484 | 121.5437484 | 1.75244449 | 0.513004949 | 0.75555 | 1.752444487 | 16.48188212 | 16.48188212 | 0.773083339 | -0.111773686 | FALSE | FALSE | FALSE |
| KLHDC9 | 1.3546267 | 0.53920988 | 0.5036 | 1.354626669 | 1.354626669 | 2.94746797 | 0.19108163 | 0.854091272 | 1 | 0.191081635 | 0.191081635 | 0.191081635 | 7.089256231 | 0.850600674 | FALSE | TRUE | FALSE |
| PCSK1 | 1.3542063 | 0.53933604 | 0.62505 | 5.417089384 | 198.6615849 | 795.0821724 | 0.56827894 | 0.725425664 | 0.5074 | 0.568278943 | 2.272871848 | 4.804765934 | 2.382995654 | 0.37712325 | FALSE | TRUE | FALSE |
| ZNF445 | 1.3524763 | 0.53946215 | 0.515 | 1.35247632 | 1.35247632 | 26.52119822 | 0.38880776 | 0.796106102 | 1 | 0.38880776 | 0.38880776 | 7.642205827 | 3.478521929 | 0.541394746 | FALSE | TRUE | FALSE |
| WDR75 | 1.3493006 | 0.53958822 | 0.50845 | 1.349300594 | 1.349300594 | 26.62751745 | 0.18397706 | 0.855956236 | 1 | 1.69467237 | 1.69467237 | 6.220507471 | 7.334069953 | 0.865345048 | FALSE | TRUE | FALSE |
| MTX1 | 1.3490447 | 0.53971422 | 0.50465 | 1.349044658 | 1.349044658 | 2.934404495 | 1.6916019 | 0.517933121 | 0.50525 | 1.691601898 | 1.691601898 | 3.666120471 | 0.797495356 | -0.098271837 | FALSE | FALSE | FALSE |
| METTL8 | 1.3487766 | 0.53984014 | 0.93785 | 12.57370641 | 1178.368864 | 1178.368864 | 0.20911893 | 0.849968668 | 0.0068 | 0.209118933 | 0.209118933 | 0.209118933 | 6.449806407 | 0.809546679 | FALSE | TRUE | FALSE |
| UQCC1 | 1.3467266 | 0.53996601 | 0.75455 | 1.34672663 | 12.59191632 | 12.59191632 | 0.19025671 | 0.854262762 | 1 | 1.743041739 | 1.743041739 | 1.743041739 | 7.078470955 | 0.849939454 | FALSE | TRUE | FALSE |
| WDR26 | 1.3461722 | 0.54009181 | 0.50105 | 1.346172202 | 1.346172202 | 11.38674188 | 1.53126435 | 0.532752652 | 0.50605 | 1.531264345 | 1.531264345 | 7.586516288 | 0.879124631 | -0.055949552 | FALSE | FALSE | FALSE |
| ZHX3 | 1.3460183 | 0.54021753 | 0.5223 | 1.346018303 | 1.346018303 | 26.41531844 | 0.18677656 | 0.855322433 | 1 | 0.186776561 | 0.186776561 | 3.673730276 | 7.206569696 | 0.857728591 | FALSE | TRUE | FALSE |

| | | | | | | | | | | | | | | | | | |
|---|---|---|---|---|---|---|---|---|---|---|---|---|---|---|---|---|---|
| CALCOCO1 | 1.3445227 | 0.54034319 | 0.93835 | 12.58887047 | 1190.118718 | 1190.118718 | 0.208138 | 0.850163413 | 1 | 0.208138001 | 0.208138001 | 4.111648531 | 6.459765785 | 0.810216772 | FALSE | TRUE | FALSE |
| ZKSCAN3 | 1.3442745 | 0.54046877 | 0.5078 | 1.344274452 | 1.344274452 | 2.922497648 | 0.18997946 | 0.854354938 | 0.0144 | 0.189979459 | 0.189979459 | 3.766728503 | 7.075893669 | 0.849781298 | FALSE | TRUE | FALSE |
| NDUFB2 | 1.3419616 | 0.54059431 | 0.50315 | 3.121430739 | 12.48454274 | 12.48454274 | 0.38560128 | 0.797410913 | 1 | 0.385601283 | 3.595936029 | 3.595936029 | 3.480179209 | 0.541601608 | FALSE | TRUE | FALSE |
| NKD1 | 1.3416925 | 0.54071977 | 0.5082 | 1.341692465 | 1.341692465 | 11.3570423 | 0.38624839 | 0.797181922 | 1 | 0.386248393 | 0.386248393 | 7.645136246 | 3.473651906 | 0.540786296 | FALSE | TRUE | FALSE |
| SLC6A7 | 1.3407784 | 0.54084517 | 0.1986 | 0.335281798 | 1.340778383 | 6.606383437 | 0.01016213 | 0.89671557 | 1 | 0.040612929 | 0.040612929 | 0.344081114 | 131.9387148 | 2.120372249 | FALSE | TRUE | FALSE |
| UCHL5 | 1.3383504 | 0.54097052 | 0.5067 | 1.338350397 | 1.338350397 | 1.338350397 | 0.18947583 | 0.85443386 | 1 | 0.189475835 | 0.189475835 | 0.189475835 | 7.063435812 | 0.849016003 | FALSE | TRUE | FALSE |
| RAB2A | 1.3382602 | 0.54109579 | 1 | 3.13643E+51 | 4.4739E+60 | 7.01421E+64 | 0.25037977 | 0.839324132 | 1 | 2.40131E+49 | 1.14446E+58 | 1.24532E+62 | 5.344921216 | 0.727941308 | FALSE | TRUE | FALSE |
| GGA2 | 1.3382141 | 0.54122098 | 0.50245 | 1.338214075 | 1.338214075 | 2.906819328 | 1.67954367 | 0.519114543 | 0.5109 | 1.679543673 | 1.679543673 | 3.63556415 | 0.796772419 | -0.098665708 | FALSE | FALSE | FALSE |
| MSTO1 | 1.3378754 | 0.5413461 | 0.9387 | 29.85592508 | 1172.353254 | 1172.353254 | 0.38473656 | 0.797715402 | 1 | 34.48791237 | 339.2845499 | 339.2845499 | 3.477380441 | 0.541252207 | FALSE | TRUE | FALSE |
| KDM4B | 1.3375084 | 0.54147114 | 0.75645 | 1.337508431 | 12.8622823 | 45.13793289 | 0.19131287 | 0.854025214 | 0.03275 | 0.191312869 | 0.191312869 | 3.728703086 | 6.991209923 | 0.844552343 | FALSE | TRUE | FALSE |
| TSPAN19 | 1.3336147 | 0.54159616 | 0.7446 | 1.333614675 | 12.43879476 | 12.43879476 | 15.6863187 | 0.155956788 | 0.2548 | 1.673367501 | 15.68631874 | 15.68631874 | 0.085017696 | -1.070490669 | FALSE | FALSE | FALSE |
| ADAMTS13 | 1.3328268 | 0.54172112 | 0.07805 | 0.036908968 | 1.332826757 | 2.849417197 | 0.08185007 | 0.879807242 | 0.7271 | 0.186645869 | 6.345767353 | 14.46688106 | 16.28375833 | 1.211754648 | FALSE | TRUE | FALSE |
| JAKMIP3 | 1.331997 | 0.541846 | 0.758 | 1.331996977 | 12.50220115 | 26.08985603 | 1.67442833 | 0.519849929 | 0.75505 | 1.67442833 | 15.78439035 | 32.87863373 | 0.795493574 | -0.099363324 | FALSE | FALSE | FALSE |
| NENF | 1.3317094 | 0.54197082 | 0.75105 | 1.33170944 | 12.59942273 | 12.59942273 | 0.18687148 | 0.855283455 | 0.49645 | 0.186871479 | 1.730104705 | 1.730104705 | 7.12633863 | 0.852866455 | FALSE | TRUE | FALSE |
| PRELP | 1.3316256 | 0.54209555 | 0.75655 | 1.331625644 | 12.54406196 | 26.40219385 | 1.62284871 | 0.523207549 | 0.87775 | 15.33372975 | 148.6020902 | 148.6020902 | 0.820548237 | -0.085895883 | FALSE | FALSE | FALSE |
| NT5DC3 | 1.3302356 | 0.54222022 | 0.7507 | 1.330235623 | 12.42325961 | 26.33904178 | 1.61924646 | 0.523932604 | 0.8722 | 15.18298405 | 146.0196775 | 146.0196775 | 0.821515228 | -0.085384382 | FALSE | FALSE | FALSE |
| SLC25A52 | 1.330057 | 0.54234482 | 0.5016 | 1.330057015 | 1.330057015 | 1.330057015 | 0.1818101 | 0.856584743 | 0.5062 | 1.672545551 | 1.672545551 | 1.672545551 | 7.315638733 | 0.864252251 | FALSE | TRUE | FALSE |
| ZDHHC6 | 1.3284891 | 0.54246936 | 0.5063 | 1.328489083 | 1.328489083 | 2.88158646 | 0.18737403 | 0.855166397 | 1 | 0.187374025 | 0.187374025 | 0.187374025 | 7.090038668 | 0.850648604 | FALSE | TRUE | FALSE |
| GGCT | 1.3281532 | 0.54259383 | 0.50315 | 1.328153179 | 1.328153179 | 1.328153179 | 0.18243553 | 0.856341666 | 1 | 1.667903773 | 1.667903773 | 1.667903773 | 7.280123319 | 0.862138736 | FALSE | TRUE | FALSE |
| PMFBP1 | 1.3249707 | 0.54271826 | 0.9945 | 9180.600421 | 8857245.828 | 91362743.99 | 262.259405 | 0.009619335 | 0.50935 | 262.2594048 | 23847.45614 | 235295.8905 | 0.005052138 | -2.296524794 | FALSE | FALSE | FALSE |
| CD72 | 1.3224757 | 0.54284265 | 0.503 | 1.322475659 | 1.322475659 | 1.322475659 | 3.42561079 | 0.387132251 | 0.50645 | 3.42561079 | 3.42561079 | 3.42561079 | 0.386055434 | -0.413350331 | FALSE | FALSE | FALSE |
| HOMER3 | 1.3218303 | 0.54296697 | 0.96935 | 21.15763857 | 198.9435485 | 796.2297214 | 0.38444049 | 0.797791378 | 1 | 24.61081354 | 98.50130103 | 394.3360164 | 3.438322153 | 0.536346565 | FALSE | TRUE | FALSE |
| GRIN2D | 1.3217054 | 0.54309122 | 0.8802 | 12.66207737 | 124.4166112 | 247.15288 | 1.77908461 | 0.510125877 | 0.51535 | 1.779084606 | 1.779084606 | 34.82680581 | 0.742913171 | -0.129061942 | FALSE | FALSE | FALSE |
| SHPRH | 1.3211536 | 0.54321539 | 0.3337 | 0.756563954 | 48.35708911 | 193.4410535 | 0.01849374 | 0.894208373 | 0.99115 | 2.652759924 | 118.9278441 | 923.0551555 | 71.43790215 | 1.853928693 | FALSE | TRUE | FALSE |
| NDUFAF3 | 1.3185124 | 0.54333953 | 0.49965 | 0.141940985 | 1.31851243 | 1.31851243 | 1.66338655 | 0.521022044 | 0.5034 | 1.663386553 | 1.663386553 | 1.663386553 | 0.792667482 | -0.100908958 | FALSE | FALSE | FALSE |
| DSC3 | 1.3175919 | 0.5434636 | 0.60005 | 1.317591858 | 111.2976652 | 445.058545 | 84.6989046 | 0.034413021 | 0.4548 | 84.69890464 | 12728.44169 | 122442.3264 | 0.015556185 | -1.808096891 | FALSE | FALSE | FALSE |
| GLUL | 1.3165771 | 0.5435876 | 0.5055 | 1.316577068 | 1.316577068 | 2.765579597 | 0.17429008 | 0.858513558 | 1 | 1.645495397 | 6.579152692 | 6.579152692 | 7.553941432 | 0.878173613 | FALSE | TRUE | FALSE |
| STK17A | 1.3155167 | 0.54371155 | 0.5066 | 1.315516654 | 1.315516654 | 2.875174037 | 0.18099737 | 0.856852458 | 1 | 1.654029644 | 1.654029644 | 1.654029644 | 7.268153387 | 0.861424084 | FALSE | TRUE | FALSE |
| PDHA2 | 1.3141642 | 0.54383543 | 0.9102 | 48.18645041 | 7251.050517 | 69918.39143 | 1.3546445 | 0.550442101 | 0.34565 | 0.764444968 | 12.22029619 | 48.87786406 | 0.970117396 | -0.013175707 | FALSE | FALSE | FALSE |
| ARRDC4 | 1.3118449 | 0.54395927 | 0.5017 | 1.31184489 | 1.31184489 | 1.31184489 | 1.528 | 0.533171864 | 0.507 | 1.527999998 | 1.527999998 | 1.527999998 | 0.858537233 | -0.066240866 | FALSE | FALSE | FALSE |
| MCPH1 | 1.3111222 | 0.54408304 | 0.32595 | 0.595008006 | 11.84077874 | 47.33969518 | 0.03783172 | 0.889473226 | 0.7483 | 0.037831722 | 0.330028365 | 0.330028365 | 34.65668855 | 1.539787064 | FALSE | TRUE | FALSE |
| ZNF263 | 1.3108373 | 0.54420674 | 0.5092 | 1.310837328 | 5.245375911 | 11.07863214 | 0.11327219 | 0.872700036 | 1 | 0.113272188 | 0.113272188 | 2.236172319 | 11.57245529 | 1.063425511 | FALSE | TRUE | FALSE |
| DDR1 | 1.3099832 | 0.54433037 | 0.24555 | 0.327572599 | 5.240013069 | 20.96626134 | 0.73034435 | 0.651457713 | 0.2034 | 0.182646602 | 0.894072452 | 3.574211781 | 1.793651519 | 0.25373807 | FALSE | TRUE | FALSE |
| RASAL1 | 1.3098613 | 0.54445392 | 0.12785 | 0.327456066 | 5.240901989 | 20.97467452 | 0.5975212 | 0.713390126 | 0.7818 | 2.390346721 | 38.28260472 | 153.261976 | 2.192158652 | 0.340871982 | FALSE | TRUE | FALSE |
| TBC1D10C | 1.3086756 | 0.54457742 | 0.75365 | 1.308675647 | 12.20718468 | 12.34454718 | 0.18581995 | 0.855568823 | 0.50935 | 1.698812386 | 1.698812386 | 3.687624562 | 7.04270798 | 0.847739681 | FALSE | TRUE | FALSE |
| CACNG2 | 1.3085947 | 0.54470084 | 0.74845 | 1.308594652 | 5.234344026 | 6.486015336 | 0.3701868 | 0.802221704 | 1 | 3.505039227 | 3.505039227 | 3.543441923 | 3.534957618 | 0.548384211 | FALSE | TRUE | FALSE |
| CD320 | 1.3084848 | 0.54482418 | 0.7542 | 1.308484824 | 12.49041903 | 12.49041903 | 0.7458723 | 0.643483326 | 0.4959 | 0.186525439 | 0.745872297 | 0.745872297 | 1.754301413 | 0.244104213 | FALSE | TRUE | FALSE |
| CHN1 | 1.3056925 | 0.54494748 | 0.75415 | 1.305692492 | 12.43298344 | 12.43298344 | 0.18863679 | 0.854722521 | 1 | 0.188636786 | 0.188636786 | 0.188636786 | 6.921727834 | 0.840214519 | FALSE | TRUE | FALSE |
| LIPE | 1.3049475 | 0.54507072 | 0.50995 | 1.304947499 | 11.87549952 | 25.4610143 | 0.0294837 | 0.891366584 | 1 | 0.029483696 | 0.029483696 | 0.576954871 | 44.25997054 | 1.646011121 | FALSE | TRUE | FALSE |
| MRGPRX3 | 1.3033967 | 0.5451939 | 0.01085 | 1.303396701 | 1.303396701 | 2.831066434 | 0.18452482 | 0.855827314 | 1 | 0.184524817 | 0.184524817 | 0.184524817 | 7.063530654 | 0.849021834 | FALSE | TRUE | FALSE |
| CAPS | 1.2993757 | 0.54531706 | 0.4984 | 0.142099351 | 1.299375659 | 1.299375659 | 0.18389355 | 0.855981995 | 0.0061 | 0.18389355 | 0.18389355 | 0.18389355 | 7.06591211 | 0.849168231 | FALSE | TRUE | FALSE |
| RIPK1 | 1.2981372 | 0.54544017 | 0.51065 | 1.298137234 | 5.192693435 | 10.98502112 | 0.43666115 | 0.778577432 | 0.8762 | 1.74635392 | 6.985998786 | 8.632777573 | 2.972870914 | 0.473176052 | FALSE | TRUE | FALSE |
| CALB2 | 1.2978267 | 0.5455632 | 0.50705 | 1.297826703 | 5.189264777 | 5.189264777 | 0.18102708 | 0.856827003 | 1 | 0.723857079 | 0.723857079 | 0.723857079 | 7.16924048 | 0.855473148 | FALSE | TRUE | FALSE |
| FAM98B | 1.2939015 | 0.54568621 | 0.74925 | 3.006945832 | 12.02716571 | 12.02716571 | 0.38089198 | 0.799074225 | 1 | 1.523498905 | 1.523498905 | 1.523498905 | 3.397029997 | 0.531099382 | FALSE | TRUE | FALSE |
| IMMP2L | 1.2935668 | 0.54580914 | 0.87485 | 5.174997105 | 48.18866302 | 48.18866302 | 0.37757662 | 0.800068691 | 1 | 3.523993228 | 3.523993228 | 3.523993228 | 3.42597165 | 0.534783765 | FALSE | TRUE | FALSE |
| STIP1 | 1.2934478 | 0.54593201 | 0.50495 | 12.30375411 | 120.0417934 | 120.0417934 | 0.19313237 | 0.8535745 | 1 | 0.193132369 | 0.193132369 | 3.823522087 | 6.697209029 | 0.825893854 | FALSE | TRUE | FALSE |
| GJD3 | 1.2926144 | 0.5460548 | 0.5004 | 4.728899545 | 120.2608169 | 120.2608169 | 16.1620756 | 0.152422913 | 0.25045 | 1.69080179 | 16.16207561 | 16.16207561 | 0.079978244 | -1.097028137 | FALSE | FALSE | FALSE |
| SAMSN1 | 1.2923268 | 0.54617752 | 0.75265 | 1.292326798 | 12.14719827 | 12.14719827 | 0.18283523 | 0.856264739 | 0.5115 | 1.682931856 | 1.682931856 | 1.682931856 | 7.068259336 | 0.849312476 | FALSE | TRUE | FALSE |
| SCARB1 | 1.2896384 | 0.54630021 | 0.25415 | 0.322612884 | 1.289638353 | 2.768521381 | 0.84777229 | 0.608129683 | 0.2565 | 0.212008739 | 0.847772285 | 4.194330326 | 1.521208437 | 0.182188725 | FALSE | TRUE | FALSE |
| TBC1D24 | 1.2893674 | 0.54642282 | 0.7567 | 1.289367362 | 12.04823705 | 25.38973155 | 0.18884708 | 0.854683225 | 0.01685 | 0.188847082 | 0.188847082 | 3.725606619 | 6.827573657 | 0.834266394 | FALSE | TRUE | FALSE |
| SUN5 | 1.2876911 | 0.54654538 | 0.50395 | 1.287691131 | 1.287691131 | 2.801653197 | 0.17728594 | 0.857700274 | 0.50145 | 1.624114777 | 1.624114777 | 1.624114777 | 7.263357476 | 0.861137419 | FALSE | TRUE | FALSE |
| FKBP4 | 1.2870105 | 0.54666787 | 0.5061 | 1.287010544 | 1.287010544 | 2.733385577 | 0.17988566 | 0.857043091 | 1 | 0.179885658 | 0.179885658 | 3.56661681 | 7.154603436 | 0.854585567 | FALSE | TRUE | FALSE |
| SUSD5 | 1.2869706 | 0.54679029 | 0.5141 | 1.286970633 | 1.286970633 | 2.793619899 | 0.18288146 | 0.85625191 | 1 | 0.182881458 | 0.182881458 | 3.619394387 | 7.037184875 | 0.847398961 | FALSE | TRUE | FALSE |
| STRBP | 1.2853995 | 0.54691265 | 0.50225 | 1.285399518 | 1.285399518 | 10.87139437 | 0.37185189 | 0.80178517 | 1 | 0.371851889 | 0.371851889 | 7.35725356 | 3.45675135 | 0.538668141 | FALSE | TRUE | FALSE |
| ODF4 | 1.2846655 | 0.54703495 | 0.75115 | 1.284665483 | 11.88977615 | 11.88977615 | 0.18931083 | 0.854538961 | 1 | 0.189310834 | 0.189310834 | 0.189310834 | 6.786011413 | 0.831614586 | FALSE | TRUE | FALSE |
| CRBN | 1.284295 | 0.54715717 | 0.01075 | 1.28429502 | 1.28429502 | 2.794024561 | 1.62038496 | 0.52364287 | 0.50475 | 1.620384962 | 1.620384962 | 1.620384962 | 0.792586361 | -0.100953406 | FALSE | FALSE | FALSE |
| IL18BP | 1.2831695 | 0.54727933 | 0.74805 | 1.283169475 | 11.87401064 | 11.87401064 | 0.1891459 | 0.854591454 | 0.00445 | 0.189145896 | 0.189145896 | 0.189145896 | 6.784019665 | 0.831487098 | FALSE | TRUE | FALSE |
| DLGAP4 | 1.2823062 | 0.54740143 | 0.5154 | 1.282306241 | 1.282306241 | 24.9436986 | 0.17304408 | 0.858799193 | 0.51625 | 1.624873028 | 1.624873028 | 31.71173442 | 7.410286728 | 0.869835013 | FALSE | TRUE | FALSE |
| UBQLNL | 1.281806 | 0.54752346 | 0.01265 | 1.281805987 | 1.281805987 | 2.790527026 | 0.18211799 | 0.856431318 | 1 | 0.182117989 | 0.182117989 | 3.616567829 | 7.038327151 | 0.84746945 | FALSE | TRUE | FALSE |
| NSF | 1.2817837 | 0.54764542 | 0.50945 | 1.281783691 | 1.281783691 | 25.29122229 | 0.7072349 | 0.662807343 | 0.5107 | 0.707234903 | 0.707234903 | 5.980720503 | 1.81238749 | 0.258251056 | FALSE | TRUE | FALSE |
| IRF3 | 1.2807681 | 0.54776731 | 0.1304 | 0.138583494 | 1.280768087 | 1.280768087 | 0.00793731 | 0.897311568 | 1 | 0.007937311 | 0.007937311 | 0.007937311 | 161.3604475 | 2.20779709 | FALSE | TRUE | FALSE |
| ZNF333 | 1.2795279 | 0.54788914 | 0.81615 | 11.68023159 | 109.298216 | 1047.794544 | 0.26121676 | 0.836324578 | 0.5034 | 0.261216759 | 0.261216759 | 0.587544149 | 4.898337776 | 0.69004873 | FALSE | TRUE | FALSE |
| ZNF687 | 1.2792975 | 0.54801091 | 0.5101 | 1.279297493 | 1.279297493 | 24.92393258 | 0.18029059 | 0.857005 | 1 | 0.180290587 | 0.180290587 | 3.523467294 | 7.095753098 | 0.850998495 | FALSE | TRUE | FALSE |
| HOXC10 | 1.2792049 | 0.54813259 | 0.50865 | 2.990670162 | 47.83282994 | 47.83282994 | 0.11156028 | 0.872974899 | 1 | 1.044872252 | 1.044872252 | 1.044872252 | 11.46649073 | 1.059430524 | FALSE | TRUE | FALSE |
| GKN1 | 1.2785077 | 0.54825421 | 0.50065 | 1.27850765 | 1.27850765 | 1.27850765 | 0.18267438 | 0.85632885 | 1 | 0.182674384 | 0.182674384 | 0.182674384 | 6.998833792 | 0.84502568 | FALSE | TRUE | FALSE |
| RNFT1 | 1.2779976 | 0.54837576 | 0.9358 | 11.90254071 | 1113.349442 | 1113.349442 | 1.77040321 | 0.51103858 | 0.5045 | 1.77040321 | 1.77040321 | 3.845702986 | 0.721868117 | -0.14154214 | FALSE | FALSE | FALSE |
| NT5C3B | 1.2763984 | 0.54849726 | 0.25375 | 0.140194751 | 1.276398421 | 1.276398421 | 1.7990884 | 0.507675948 | 0.71835 | 4.221550222 | 67.49429281 | 269.9774002 | 0.709469539 | -0.149066246 | FALSE | FALSE | FALSE |
| STT3B | 1.2756868 | 0.5486187 | 0.5044 | 1.275686818 | 5.100818344 | 25.16343504 | 0.17750645 | 0.857637336 | 1 | 0.709789019 | 0.709789019 | 6.001572502 | 7.186706997 | 0.856529939 | FALSE | TRUE | FALSE |
| SURF2 | 1.2754386 | 0.54874006 | 0.5037 | 1.275438627 | 1.275438627 | 1.275438627 | 0.17690099 | 0.857876203 | 0.50205 | 1.606870694 | 1.606870694 | 1.606870694 | 7.209900916 | 0.857929296 | FALSE | TRUE | FALSE |
| BTN3A1 | 1.2743629 | 0.54886136 | 0.75505 | 1.274362947 | 11.83116301 | 11.83116301 | 0.18762645 | 0.855101286 | 1 | 0.187626449 | 0.187626449 | 0.187626449 | 6.792021863 | 0.831999075 | FALSE | TRUE | FALSE |
| AP5B1 | 1.2735332 | 0.54898259 | 0.5136 | 1.273533212 | 1.273533212 | 25.06516146 | 0.1812663 | 0.856737843 | 1 | 0.181266302 | 0.181266302 | 3.574446463 | 7.02575823 | 0.846693201 | FALSE | TRUE | FALSE |
| TNFRSF6B | 1.2704507 | 0.5491038 | 0.50385 | 1.270450694 | 1.270450694 | 2.750367438 | 0.17482229 | 0.858364085 | 1 | 1.605633059 | 1.605633059 | 3.465192482 | 7.267097774 | 0.861361003 | FALSE | TRUE | FALSE |
| C19orf12 | 1.2698966 | 0.54922494 | 0.74875 | 1.269896566 | 5.079773261 | 5.079773261 | 52.9468577 | 0.052301368 | 0.0031 | 5.63632938 | 52.9468577 | 52.9468577 | 0.023984361 | -1.620071842 | FALSE | FALSE | FALSE |

| | | | | | | | | | | | | | | | | | |
|---|---|---|---|---|---|---|---|---|---|---|---|---|---|---|---|---|---|
| WNT8B | 1.2695458 | 0.549346 | 0.01435 | 1.269545824 | 1.269545824 | 2.692780398 | 0.67937618 | 0.675393278 | 0.9388 | 2.717289909 | 43.50236461 | 43.50236461 | 1.868693447 | 0.271538063 | FALSE | TRUE | FALSE |
| TBC1D14 | 1.2664464 | 0.54946704 | 0.5153 | 1.266446419 | 1.266446419 | 24.93284789 | 0.36665882 | 0.803519774 | 1 | 0.366658816 | 0.366658816 | 7.232091292 | 3.454018737 | 0.538324689 | FALSE | TRUE | FALSE |
| B3GNT8 | 1.2653592 | 0.54958802 | 0.5055 | 1.265359246 | 1.265359246 | 2.752996564 | 0.18030729 | 0.856992299 | 1 | 0.180307294 | 0.180307294 | 3.572251839 | 7.017792879 | 0.846200546 | FALSE | TRUE | FALSE |
| WNT9A | 1.2626272 | 0.54970896 | 0.5059 | 1.262627195 | 1.262627195 | 2.644872835 | 0.17541992 | 0.85823929 | 1 | 0.175419921 | 0.175419921 | 3.474414353 | 7.197741212 | 0.857196228 | FALSE | TRUE | FALSE |
| BCAT2 | 1.2609487 | 0.54982985 | 0.5112 | 1.260948731 | 5.04345651 | 5.04345651 | 0.71860172 | 0.657002105 | 0.01045 | 0.718601725 | 0.718601725 | 3.562926107 | 1.754725444 | 0.244209174 | FALSE | TRUE | FALSE |
| EFCAB5 | 1.2601908 | 0.54995067 | 0.94635 | 101.0497383 | 86530.63211 | 859412.6267 | 2.8678191 | 0.422102921 | 0.51445 | 25.8441936 | 2255.607019 | 21820.8332 | 0.439424802 | -0.357115434 | FALSE | FALSE | FALSE |
| LYPD3 | 1.2601601 | 0.55007142 | 0.50915 | 1.26016015 | 1.26016015 | 2.745043439 | 0.17972975 | 0.857106533 | 0.01155 | 0.179729746 | 0.179729746 | 3.562300793 | 7.011416754 | 0.845805782 | FALSE | TRUE | FALSE |
| LRRC27 | 1.2579762 | 0.55019213 | 0.5116 | 11.33324888 | 104.6648316 | 990.2685353 | 19.1862529 | 0.136654914 | 0.31375 | 2.103405085 | 179.3962564 | 179.3962564 | 0.065566538 | -1.183317747 | FALSE | FALSE | FALSE |
| DPF2 | 1.2554586 | 0.5503128 | 0.87475 | 12.17201847 | 121.0376206 | 121.0376206 | 0.73411971 | 0.649379289 | 0.5083 | 0.734119709 | 0.734119709 | 3.641402216 | 1.710155157 | 0.233035514 | FALSE | TRUE | FALSE |
| BMPR1A | 1.2506419 | 0.55043346 | 0.69605 | 1.250641936 | 20.00850378 | 24.72815168 | 6.54408742 | 0.253843805 | 0.5074 | 26.1802365 | 419.3222654 | 1678.795336 | 0.191110212 | -0.718716105 | FALSE | FALSE | FALSE |
| PGLYRP2 | 1.2488797 | 0.55055407 | 0.51325 | 1.248879722 | 1.248879722 | 24.60450198 | 0.17293726 | 0.858836371 | 1 | 1.580016441 | 1.580016441 | 5.856204506 | 7.22157673 | 0.85863203 | FALSE | TRUE | FALSE |
| STRADB | 1.2479819 | 0.55067462 | 0.5027 | 1.247981888 | 1.247981888 | 1.247981888 | 0.36175048 | 0.805082636 | 1 | 0.361750482 | 0.361750482 | 0.361750482 | 3.44984167 | 0.537799164 | FALSE | TRUE | FALSE |
| FAM53A | 1.2460118 | 0.55079513 | 0.75785 | 1.246011819 | 11.60900211 | 24.65034125 | 0.18359505 | 0.856059215 | 1 | 0.183595051 | 0.183595051 | 3.636865323 | 6.786739702 | 0.831661193 | FALSE | TRUE | FALSE |
| EPX | 1.2445566 | 0.55091558 | 0.7533 | 25.82273664 | 8306.85355 | 37802.11277 | 15.8944752 | 0.154350524 | 0.6642 | 63.53318518 | 23686.806 | 94727.90391 | 0.07830121 | -1.106231524 | FALSE | FALSE | FALSE |
| KCTD14 | 1.2444611 | 0.55103595 | 0.75555 | 4.976555112 | 47.08379229 | 47.08379229 | 0.18360976 | 0.856046351 | 1 | 0.183609756 | 0.183609756 | 0.183609756 | 6.777750627 | 0.831085586 | FALSE | TRUE | FALSE |
| PRSS35 | 1.2439542 | 0.55115626 | 0.74785 | 1.243954215 | 11.6706253 | 11.6706253 | 1.57618612 | 0.528807185 | 0.7522 | 1.576186119 | 14.85240468 | 14.85240468 | 0.789217847 | -0.102803103 | FALSE | FALSE | FALSE |
| RAG2 | 1.2439077 | 0.5512765 | 0.9008 | 79.50841365 | 7340.885403 | 29365.70038 | 0.2684375 | 0.834217371 | 0.81825 | 1.073185187 | 4.291554856 | 17.16576438 | 4.633882146 | 0.665944984 | FALSE | TRUE | FALSE |
| OR4K1 | 1.2428738 | 0.55139667 | 0.75095 | 1.242873756 | 11.53750065 | 11.53750065 | 0.18357046 | 0.856084937 | 0.0077 | 0.18357046 | 0.18357046 | 0.18357046 | 6.770554231 | 0.830624221 | FALSE | TRUE | FALSE |
| RNF181 | 1.2427602 | 0.55151678 | 0.8776 | 11.69417171 | 112.8657585 | 112.8657585 | 1.67843413 | 0.519408977 | 0.5047 | 1.678434127 | 1.678434127 | 1.678434127 | 0.740428331 | -0.130516972 | FALSE | FALSE | FALSE |
| TMED6 | 1.2398343 | 0.55163685 | 0.75335 | 1.239834336 | 11.47256434 | 11.47256434 | 0.18347251 | 0.856123503 | 1 | 0.183472514 | 0.183472514 | 0.183472514 | 6.757602609 | 0.829792649 | FALSE | TRUE | FALSE |
| RNF135 | 1.2392478 | 0.55175685 | 0.8756 | 1.239247776 | 109.4579051 | 109.4579051 | 0.18311399 | 0.85621341 | 1 | 1.667854271 | 1.667854271 | 1.667854271 | 6.767630185 | 0.830436619 | FALSE | TRUE | FALSE |
| ZSCAN1 | 1.2375437 | 0.5518768 | 0.98315 | 109.7211676 | 10740.10799 | 110115.7599 | 16.1193309 | 0.152744061 | 0.13545 | 16.11933094 | 154.9546029 | 154.9546029 | 0.076773889 | -1.114786461 | FALSE | FALSE | FALSE |
| COL23A1 | 1.2375005 | 0.55199668 | 0.0096 | 0.077389508 | 1.237500504 | 1.237500504 | 0.03726498 | 0.889622781 | 0.4999 | 0.009323709 | 0.037264984 | 0.184981727 | 33.2081323 | 1.521244451 | FALSE | TRUE | FALSE |
| KRT26 | 1.2366304 | 0.5521165 | 0.66 | 1.2366304 | 112.5587074 | 1112.886122 | 0.03685026 | 0.889693496 | 0.51095 | 0.036850257 | 0.036850257 | 0.082591854 | 33.55825755 | 1.525799403 | FALSE | TRUE | FALSE |
| SYNDIG1L | 1.2353049 | 0.55223627 | 0.69515 | 1.23530487 | 19.76101421 | 19.76101421 | 25.886326 | 0.10645205 | 0.22445 | 6.470926541 | 414.575585 | 515.3793866 | 0.047720363 | -1.321296263 | FALSE | FALSE | FALSE |
| ABI3 | 1.2343353 | 0.55235597 | 0.50165 | 1.23433533 | 1.23433533 | 1.23433533 | 0.1750947 | 0.85827675 | 1 | 0.175094698 | 0.175094698 | 0.175094698 | 7.049530025 | 0.848160165 | FALSE | TRUE | FALSE |
| NNMT | 1.233984 | 0.55247561 | 0.50115 | 1.233983966 | 1.233983966 | 1.233983966 | 0.17199724 | 0.859108425 | 1 | 1.560177645 | 1.560177645 | 1.560177645 | 7.174440697 | 0.85578805 | FALSE | TRUE | FALSE |
| TBC1D28 | 1.2333313 | 0.55259517 | 0.74935 | 1.233331327 | 11.40046871 | 11.40046871 | 0.18275224 | 0.85629039 | 0.006 | 0.182752243 | 0.182752243 | 0.182752243 | 6.748652196 | 0.829217047 | FALSE | TRUE | FALSE |
| NPRL2 | 1.232552 | 0.55271468 | 0.87565 | 4.930714202 | 19.72981153 | 19.72981153 | 1.44845578 | 0.54073851 | 0.01145 | 1.448455781 | 1.448455781 | 7.18842381 | 0.85094211 | -0.070099984 | FALSE | FALSE | FALSE |
| RDH16 | 1.2324696 | 0.55283412 | 0.9814 | 1215.770736 | 521891.4803 | 5595784.053 | 4.1212028 | 0.348203126 | 0.7742 | 38.63224059 | 3660.671858 | 12602.26605 | 0.29905581 | -0.524247756 | FALSE | FALSE | FALSE |
| SOX15 | 1.2320666 | 0.55295348 | 0.7479 | 1.23206659 | 4.928384693 | 4.928384693 | 0.36218244 | 0.80491807 | 1 | 0.362182436 | 0.362182436 | 0.362182436 | 3.401784483 | 0.531706796 | FALSE | TRUE | FALSE |
| SHOX2 | 1.2318117 | 0.55307278 | 0.1342 | 0.077063064 | 1.231811684 | 1.528430509 | 0.1254886 | 0.870006839 | 0.50215 | 0.1254886 | 0.1254886 | 0.623311028 | 9.816124208 | 0.991940045 | FALSE | TRUE | FALSE |
| GLIS1 | 1.230819 | 0.55319202 | 0.99125 | 1156.360365 | 125269.483 | 234025.5361 | 0.19646436 | 0.852973908 | 1 | 1.840995548 | 17.70549603 | 36.29178537 | 6.26484616 | 0.796910411 | FALSE | TRUE | FALSE |
| ZNF598 | 1.2256392 | 0.55331125 | 0.9684 | 110.8590135 | 1094.512834 | 11070.11601 | 6.22833103 | 0.26251097 | 0.26445 | 1.696888497 | 16.04619665 | 33.27692722 | 0.19678453 | -0.706009045 | FALSE | FALSE | FALSE |
| HS3ST2 | 1.2250062 | 0.55343043 | 0.8793 | 11.54249125 | 111.6208904 | 111.6208904 | 0.35637164 | 0.806943382 | 1 | 3.361044504 | 32.53338545 | 32.53338545 | 3.437440154 | 0.536235146 | FALSE | TRUE | FALSE |
| JMJD7 | 1.2249254 | 0.55354953 | 0.5025 | 1.224925367 | 1.224925367 | 1.224925367 | 12.8527489 | 0.171204931 | 0.0077 | 3.21377739 | 12.85274886 | 12.85274886 | 0.095304544 | -1.020886393 | FALSE | FALSE | FALSE |
| PEX2 | 1.2245877 | 0.55366856 | 0.00725 | 0.306224133 | 1.224587727 | 1.224587727 | 0.41910009 | 0.78480557 | 0.5023 | 0.419100085 | 0.419100085 | 0.419100085 | 2.921945784 | 0.465672153 | FALSE | TRUE | FALSE |
| PPA1 | 1.2215906 | 0.55378756 | 0.5037 | 1.221590553 | 1.221590553 | 1.221590553 | 0.35507696 | 0.807379954 | 0.0051 | 0.35507696 | 0.35507696 | 0.35507696 | 3.440354322 | 0.536603173 | FALSE | TRUE | FALSE |
| CHST13 | 1.2210176 | 0.55390649 | 0.5118 | 1.221017644 | 1.221017644 | 6.04726962 | 0.35549278 | 0.807331511 | 1 | 0.355492784 | 0.355492784 | 0.355492784 | 3.434718505 | 0.53589115 | FALSE | TRUE | FALSE |
| ZNF418 | 1.2201051 | 0.55402537 | 0.7819 | 10.7261247 | 890.9441426 | 3259.468319 | 0.30832022 | 0.822544865 | 0.8111 | 2.692924154 | 24.11096057 | 221.1641278 | 3.957265964 | 0.59739524 | FALSE | TRUE | FALSE |
| ACTN2 | 1.2186136 | 0.55414419 | 0.51035 | 1.218613605 | 46.94990477 | 95.26025343 | 0.02533622 | 0.892381755 | 1 | 0.101275091 | 0.404922621 | 3.423402419 | 48.09769001 | 1.682124219 | FALSE | TRUE | FALSE |
| GABRA2 | 1.2185293 | 0.55426295 | 0.49955 | 0.304675442 | 1.218529256 | 6.046208212 | 0.35448328 | 0.807563178 | 1 | 0.354483278 | 0.354483278 | 0.354483278 | 3.437480215 | 0.536240207 | FALSE | TRUE | FALSE |
| HOXC11 | 1.2176709 | 0.55438164 | 0.93945 | 11.62527794 | 1142.595069 | 1142.595069 | 0.18844326 | 0.854814131 | 1 | 0.18844326 | 0.18844326 | 0.18844326 | 6.46173761 | 0.810349319 | FALSE | TRUE | FALSE |
| PPCS | 1.2165677 | 0.55450027 | 0.50605 | 1.216567742 | 1.216567742 | 2.626079537 | 0.17297435 | 0.858811588 | 0.01015 | 0.17297435 | 0.17297435 | 3.434068771 | 7.033226248 | 0.847154588 | FALSE | TRUE | FALSE |
| PRKAR1B | 1.2135323 | 0.55461888 | 0.1354 | 0.980365809 | 3.92064152 | 4.850686827 | 5.21878235 | 0.297493272 | 0.7767 | 20.87740645 | 334.3618209 | 1338.595625 | 0.232531701 | -0.633517831 | FALSE | FALSE | FALSE |
| OVCH2 | 1.2130275 | 0.55473741 | 0.9945 | 10065.10562 | 1055298.649 | 11171941.37 | 2.40197133 | 0.455447034 | 0.8182 | 22.42323516 | 214.5741523 | 2103.520513 | 0.505013312 | -0.296697174 | FALSE | FALSE | FALSE |
| FKBP6 | 1.2117666 | 0.5548559 | 0.7509 | 1.211766585 | 11.40004398 | 11.40004398 | 1.54092275 | 0.53163147 | 0.75265 | 1.540922745 | 14.55571747 | 14.55571747 | 0.786390225 | -0.104361893 | FALSE | FALSE | FALSE |
| PAPSS2 | 1.2116685 | 0.55497431 | 0.63015 | 1.211668527 | 45.38071996 | 45.38071996 | 0.05263643 | 0.88609601 | 1 | 0.210447254 | 0.84160565 | 4.159372337 | 23.01957934 | 1.362097383 | FALSE | TRUE | FALSE |
| IL6R | 1.2110031 | 0.55509266 | 0.50735 | 1.211003069 | 1.211003069 | 2.648198087 | 0.17399983 | 0.858637881 | 1 | 0.173999828 | 0.173999828 | 3.447246076 | 6.95979465 | 0.842596426 | FALSE | TRUE | FALSE |
| CBFA2T3 | 1.2074121 | 0.55521099 | 0.51065 | 1.207412052 | 1.207412052 | 10.2194194 | 0.35150178 | 0.808609833 | 1 | 0.351501782 | 0.351501782 | 6.932827037 | 3.435009757 | 0.535927975 | FALSE | TRUE | FALSE |
| ECE2 | 1.2071603 | 0.55532925 | 0.2092 | 0.075507399 | 4.828533489 | 19.31852168 | 0.00103718 | 0.899402308 | 0.9258 | 0.077048332 | 2.756923363 | 11.01659741 | 1163.888926 | 3.065911536 | FALSE | TRUE | FALSE |
| ZNF670 | 1.2061641 | 0.55544745 | 0.0094 | 1.206164062 | 1.206164062 | 1.206164062 | 0.16739979 | 0.860320252 | 1 | 1.532925802 | 1.532925802 | 1.532925802 | 7.205290032 | 0.857651467 | FALSE | TRUE | FALSE |
| KRTCAP3 | 1.2054932 | 0.55556558 | 0.8772 | 11.14563316 | 105.6969065 | 105.6969065 | 1.52775878 | 0.53331141 | 0.87485 | 14.21024767 | 135.5693201 | 135.5693201 | 0.789059876 | -0.10289004 | FALSE | FALSE | FALSE |
| LRFN2 | 1.2046594 | 0.55568366 | 0.51365 | 1.204659377 | 1.204659377 | 23.59742279 | 0.35102653 | 0.808835817 | 0.0227 | 0.351026527 | 0.351026527 | 6.893110526 | 3.431818636 | 0.535524328 | FALSE | TRUE | FALSE |
| TIMM21 | 1.2015128 | 0.55580171 | 0.8757 | 11.14646106 | 106.0624901 | 106.0624901 | 0.17290947 | 0.858873529 | 1 | 1.573102133 | 14.6890087 | 14.6890087 | 6.948796955 | 0.841909622 | FALSE | TRUE | FALSE |
| DHRS4 | 1.2009765 | 0.55591969 | 0.9645 | 106.6257677 | 10476.65004 | 41904.58601 | 0.40407378 | 0.790925697 | 0.99235 | 143.9038147 | 14182.34866 | 36514.29907 | 2.972171405 | 0.473073851 | FALSE | TRUE | FALSE |
| GPR137 | 1.200212 | 0.55603762 | 0.49985 | 0.12990558 | 1.200212011 | 2.576508667 | 0.15431332 | 0.863491599 | 1 | 0.154313325 | 0.154313325 | 0.154313325 | 7.777760027 | 0.890854539 | FALSE | TRUE | FALSE |
| RUFY2 | 1.2001381 | 0.55615547 | 0.5074 | 1.200138116 | 1.200138116 | 2.590619064 | 0.16607224 | 0.860719736 | 1 | 1.528026068 | 1.528026068 | 5.616986212 | 7.226602966 | 0.858934195 | FALSE | TRUE | FALSE |
| NACA2 | 1.1989088 | 0.55627326 | 0.50035 | 1.198908827 | 1.198908827 | 1.198908827 | 1.27499004 | 0.55751559 | 0.50225 | 1.274990041 | 1.274990041 | 1.274990041 | 0.940327994 | -0.026720635 | FALSE | FALSE | FALSE |
| RAB2B | 1.1988956 | 0.55639099 | 0.49935 | 0.299785293 | 1.198895583 | 1.198895583 | 0.34946281 | 0.809263692 | 1 | 0.349462809 | 0.349462809 | 0.349462809 | 3.430681468 | 0.535380397 | FALSE | TRUE | FALSE |
| STARD13 | 1.1987686 | 0.55650864 | 0.81335 | 11.02762498 | 251.6512814 | 1006.380336 | 4.25386358 | 0.340385071 | 0.02165 | 0.491162553 | 1.9642632 | 9.621619578 | 0.281807026 | -0.550048184 | FALSE | FALSE | FALSE |
| UNC13A | 1.1984698 | 0.55662623 | 0.157 | 0.503191036 | 4.790132897 | 23.09024521 | 3.21009199 | 0.399786773 | 0.43675 | 3.210091991 | 107.9758727 | 362.6062077 | 0.373344367 | -0.427890397 | FALSE | FALSE | FALSE |
| CPNE5 | 1.1970251 | 0.55674377 | 0.7568 | 1.197025097 | 4.788989745 | 23.58665083 | 0.35281934 | 0.808110727 | 1 | 0.352819341 | 0.352819341 | 6.964457729 | 3.392742289 | 0.530550872 | FALSE | TRUE | FALSE |
| C1QA | 1.1960642 | 0.55686124 | 0.4995 | 0.299047704 | 1.196064243 | 1.196064243 | 12.4555631 | 0.175899957 | 0.2569 | 3.114181451 | 12.45556311 | 12.45556311 | 0.096026509 | -1.01760886 | FALSE | FALSE | FALSE |
| NUDT19 | 1.1960256 | 0.55697865 | 0.5086 | 1.196025646 | 1.196025646 | 2.621529437 | 1017.73582 | 0.002421266 | 0.7448 | 10159.97211 | 1086140.211 | 11618830.79 | 0.001175183 | -2.92989457 | FALSE | FALSE | FALSE |
| NRG1 | 1.1956779 | 0.55709599 | 0.50645 | 1.195677919 | 1.195677919 | 2.569017594 | 0.1648028 | 0.860924678 | 1 | 1.524490009 | 1.524490009 | 3.268069273 | 7.255203936 | 0.860649625 | FALSE | TRUE | FALSE |
| ENDOV | 1.1956001 | 0.55721326 | 0.5056 | 1.195600098 | 4.782165806 | 4.782165806 | 1.53583605 | 0.532332795 | 0.50105 | 1.535836051 | 1.535836051 | 1.535836051 | 0.778468573 | -0.108758915 | FALSE | FALSE | FALSE |
| IKBKE | 1.1932062 | 0.55733049 | 0.50255 | 1.193206178 | 1.193206178 | 23.47605176 | 0.17056637 | 0.859453215 | 1 | 0.170566373 | 0.170566373 | 3.36242276 | 6.995553426 | 0.844822078 | FALSE | TRUE | FALSE |
| NFU1 | 1.1925413 | 0.55744766 | 0.2529 | 0.298137311 | 1.192541343 | 1.192541343 | 0.39534097 | 0.793681355 | 0.7488 | 0.898154182 | 3.591816958 | 3.591816958 | 3.016488098 | 0.479501616 | FALSE | TRUE | FALSE |
| GRK1 | 1.1914478 | 0.55756477 | 0.2657 | 1.191447782 | 11.10803401 | 11.10803401 | 1.33743457 | 0.551612711 | 0.98515 | 119.6042278 | 11808.47191 | 121588.7344 | 0.890845661 | -0.050197531 | FALSE | FALSE | FALSE |
| MRPL38 | 1.1894653 | 0.55768184 | 0.50435 | 1.189465314 | 1.189465314 | 2.591586386 | 0.66795607 | 0.681419208 | 0.75965 | 0.667956072 | 2.672306184 | 5.660841053 | 1.7807538 | 0.25060388 | FALSE | TRUE | FALSE |
| BSDC1 | 1.1878051 | 0.55779886 | 0.5066 | 1.187805087 | 1.187805087 | 5.886941403 | 0.34607845 | 0.810224985 | 0.00985 | 0.346078451 | 0.346078451 | 0.346078451 | 3.432184471 | 0.535570622 | FALSE | TRUE | FALSE |
| RNF215 | 1.1859076 | 0.55791584 | 0.5043 | 1.185907563 | 1.185907563 | 1.200131081 | 0.16592712 | 0.860780069 | 1 | 1.507057534 | 1.507057534 | 1.507057534 | 7.147159337 | 0.854133464 | FALSE | TRUE | FALSE |

| | | | | | | | | | | | | | | | | | |
|---|---|---|---|---|---|---|---|---|---|---|---|---|---|---|---|---|---|
| KAT8 | 1.1845824 | 0.55803277 | 0.49935 | 0.296190443 | 1.184582424 | 5.871423825 | 1.36758937 | 0.549266405 | 0.50505 | 1.367589368 | 1.367589368 | 6.78559248 | 0.866182827 | -0.062390431 | FALSE | FALSE | FALSE |
| ALB | 1.1838174 | 0.55814963 | 0.75455 | 1.183817418 | 11.29889483 | 23.41604493 | 0.1708101 | 0.859391763 | 1 | 0.683044792 | 0.683044792 | 3.383112107 | 6.930605696 | 0.840771191 | FALSE | TRUE | FALSE |
| VAX2 | 1.1815679 | 0.55826645 | 0.7778 | 4.725907178 | 172.8478416 | 691.6834421 | 0.49804928 | 0.755867035 | 0.6886 | 0.498049283 | 7.967239747 | 7.967239747 | 2.372391627 | 0.375186383 | FALSE | TRUE | FALSE |
| NRN1L | 1.1784473 | 0.55838325 | 0.255 | 0.123430126 | 1.178447319 | 1.178447319 | 0.01468245 | 0.895218843 | 0.75745 | 0.136912126 | 1.31030851 | 1.31030851 | 80.26230921 | 1.904511651 | FALSE | TRUE | FALSE |
| TFPI2 | 1.1771615 | 0.5585 | 0.7496 | 1.177161513 | 11.125159 | 11.125159 | 0.17109523 | 0.859317951 | 1 | 0.6842157 | 0.6842157 | 0.6842157 | 6.880154003 | 0.837598159 | FALSE | TRUE | FALSE |
| CLK1 | 1.1768504 | 0.55861668 | 0.5047 | 1.176850395 | 1.176850395 | 2.559072675 | 0.16391874 | 0.861117063 | 0.5113 | 1.499915746 | 1.499915746 | 3.249809552 | 7.179474289 | 0.856092645 | FALSE | TRUE | FALSE |
| ZC3H3 | 1.1756136 | 0.5587333 | 0.7573 | 1.175613554 | 11.10052316 | 39.728137 | 1.45458638 | 0.539787849 | 0.8767 | 13.77263182 | 133.7521964 | 269.1644919 | 0.808211577 | -0.092474933 | FALSE | FALSE | FALSE |
| DNAJC30 | 1.1752914 | 0.55884986 | 0.818 | 18.81429198 | 75.30465307 | 301.3072759 | 5.19212622 | 0.298669184 | 0.9809 | 83.15796623 | 5452.70367 | 21846.48556 | 0.22636033 | -0.645199682 | FALSE | FALSE | FALSE |
| PKN1 | 1.1724427 | 0.55896639 | 0.03015 | 1.172442693 | 1.172442693 | 22.88688837 | 0.1674122 | 0.860308112 | 1 | 0.167412201 | 0.167412201 | 3.277438238 | 7.003328817 | 0.845304518 | FALSE | TRUE | FALSE |
| MECR | 1.1717004 | 0.55908286 | 0.50695 | 1.171700402 | 1.171700402 | 1.171700402 | 0.1678425 | 0.860186596 | 0.0076 | 0.167842497 | 0.167842497 | 0.167842497 | 6.980951913 | 0.843914646 | FALSE | TRUE | FALSE |
| DBX2 | 1.1716738 | 0.55919926 | 0.506 | 1.171673847 | 1.171673847 | 1.171673847 | 0.16649574 | 0.860611009 | 1 | 0.166495744 | 0.166495744 | 0.166495744 | 7.037260036 | 0.847403599 | FALSE | TRUE | FALSE |
| PCDHB15 | 1.1697512 | 0.55931561 | 0.26345 | 0.128729833 | 1.169751243 | 2.522548299 | 0.21829432 | 0.847592699 | 0.51245 | 0.218294321 | 0.218294321 | 4.288040168 | 5.358596756 | 0.729051077 | FALSE | TRUE | FALSE |
| SDF2 | 1.1689876 | 0.5594319 | 0.6903 | 1.168987601 | 18.71391197 | 18.71391197 | 0.41025502 | 0.78837146 | 0.50595 | 0.410255023 | 0.410255023 | 0.410255023 | 2.849416912 | 0.454755998 | FALSE | TRUE | FALSE |
| ZBTB46 | 1.1675009 | 0.55954815 | 0.51235 | 1.167500914 | 1.167500914 | 22.91015631 | 0.34127434 | 0.811725261 | 1 | 0.341274339 | 0.341274339 | 6.712101099 | 3.421004106 | 0.534153595 | FALSE | TRUE | FALSE |
| GATA2 | 1.1648169 | 0.55966436 | 0.38535 | 0.49203104 | 4.655835796 | 5.754809174 | 0.18988363 | 0.854368098 | 0.881 | 0.759060546 | 3.035105955 | 3.754604813 | 6.134372454 | 0.787770141 | FALSE | TRUE | FALSE |
| EPHB4 | 1.1626685 | 0.55978053 | 0.39815 | 0.503913424 | 4.649255809 | 22.64084544 | 0.04790557 | 0.887144183 | 0.7591 | 0.443468365 | 1.773011869 | 8.66315312 | 24.2700095 | 1.385069946 | FALSE | TRUE | FALSE |
| MYO1G | 1.1621652 | 0.55989664 | 0.93585 | 24654.30795 | 118219980.3 | 3424113705 | 0.02306322 | 0.892920119 | 0.8459 | 15.97965622 | 74412.8961 | 2097377.967 | 50.39042188 | 1.702347994 | FALSE | TRUE | FALSE |
| ACTR10 | 1.1613935 | 0.56001269 | 0.12885 | 0.290464422 | 1.161393451 | 1.441459332 | 16.5126994 | 0.150177817 | 0.1597 | 1.786409774 | 16.51269937 | 66.04766077 | 0.070333349 | -1.152838702 | FALSE | FALSE | FALSE |
| NEK10 | 1.1603794 | 0.56012869 | 0.50885 | 1.160379385 | 10.57436531 | 10.57436531 | 0.02654096 | 0.892035182 | 1 | 0.026540961 | 0.026540961 | 0.525575223 | 43.72032297 | 1.640683361 | FALSE | TRUE | FALSE |
| NUGGC | 1.1571575 | 0.56024465 | 0.3221 | 1.157157523 | 104.090223 | 104.090223 | 1.80632773 | 0.506750594 | 0.66705 | 17.01623939 | 1625.459971 | 16463.3103 | 0.640613275 | -0.193404066 | FALSE | FALSE | FALSE |
| FRS3 | 1.1556957 | 0.56036057 | 0.87565 | 4.623023082 | 18.49767537 | 22.84167041 | 0.33853453 | 0.812466478 | 1 | 1.353906647 | 5.416054542 | 11.47031579 | 3.413819317 | 0.533240532 | FALSE | TRUE | FALSE |
| APRT | 1.1551174 | 0.56047643 | 0.25565 | 0.288812257 | 1.155117434 | 1.155117434 | 0.09817311 | 0.876420627 | 1 | 0.392566392 | 1.570154057 | 1.570154057 | 11.76612917 | 1.070633612 | FALSE | TRUE | FALSE |
| IRGQ | 1.1539741 | 0.56059223 | 0.51545 | 1.153974123 | 1.153974123 | 22.6329089 | 0.16623937 | 0.860695589 | 0.0208 | 0.166239368 | 0.166239368 | 3.268052113 | 6.941641666 | 0.841462191 | FALSE | TRUE | FALSE |
| UGT2B17 | 1.1531128 | 0.56070798 | 0.01265 | 1.153112847 | 1.153112847 | 2.506582994 | 109.126982 | 0.026812808 | 0.66085 | 109.1269819 | 10808.72187 | 111474.5258 | 0.010566707 | -1.976060333 | FALSE | FALSE | FALSE |
| GUCA1A | 1.1528384 | 0.56082366 | 0.8774 | 4.611796599 | 43.02913624 | 43.02913624 | 1.37676034 | 0.547821637 | 0.75345 | 1.376760339 | 5.507670262 | 5.507670262 | 0.837355918 | -0.077089906 | FALSE | FALSE | FALSE |
| JPH1 | 1.1527683 | 0.56093927 | 0.5134 | 1.152768303 | 1.152768303 | 22.61622129 | 0.33397971 | 0.814314932 | 0.51045 | 1.335757513 | 1.335757513 | 26.26640524 | 3.451611825 | 0.538021948 | FALSE | TRUE | FALSE |
| HDDC2 | 1.1461667 | 0.5610549 | 0.50605 | 1.146166706 | 1.146166706 | 1.146166706 | 0.33524338 | 0.813888433 | 1 | 0.335243375 | 0.335243375 | 0.335243375 | 3.418909339 | 0.533887585 | FALSE | TRUE | FALSE |
| DPAGT1 | 1.1450798 | 0.56117048 | 0.50535 | 1.145079844 | 10.42408926 | 41.69586051 | 0.03096253 | 0.891029481 | 0.0091 | 0.030962534 | 0.030962534 | 0.030962534 | 36.98275581 | 1.56799927 | FALSE | TRUE | FALSE |
| APPBP2 | 1.1450622 | 0.56128599 | 0.50225 | 1.145062152 | 1.145062152 | 9.68722535 | 0.33580926 | 0.813567231 | 1 | 0.335809261 | 0.335809261 | 6.653333489 | 3.409858769 | 0.532736392 | FALSE | TRUE | FALSE |
| BTN1A1 | 1.1446137 | 0.56140143 | 0.7506 | 1.144613688 | 10.7152292 | 22.62244701 | 1.46412948 | 0.538833276 | 0.7535 | 1.464129485 | 13.76980097 | 28.97847855 | 0.781770806 | -0.106920551 | FALSE | FALSE | FALSE |
| NXN | 1.139261 | 0.56151689 | 0.938 | 10.69985916 | 412.5842997 | 412.5842997 | 0.35992268 | 0.805714733 | 1 | 0.359922677 | 0.359922677 | 7.124661585 | 3.165293663 | 0.500414008 | FALSE | TRUE | FALSE |
| RNF216 | 1.1385378 | 0.56163228 | 0.87915 | 10.67456114 | 102.6513407 | 209.5660577 | 0.16381843 | 0.86115308 | 1 | 1.504490265 | 14.18109782 | 50.7847896 | 6.949998435 | 0.841984707 | FALSE | TRUE | FALSE |
| MAMSTR | 1.1379806 | 0.56174761 | 0.8752 | 10.56992813 | 100.6989618 | 100.6989618 | 0.20910116 | 0.849982594 | 1 | 0.209101164 | 0.836400637 | 0.836400637 | 5.44224906 | 0.735778413 | FALSE | TRUE | FALSE |
| PDE4A | 1.1363696 | 0.56186289 | 0.2695 | 0.284158495 | 1.136369572 | 9.595910858 | 0.0993818 | 0.876111141 | 1 | 0.099381801 | 0.099381801 | 1.948798718 | 11.43438291 | 1.058212731 | FALSE | TRUE | FALSE |
| TSPAN12 | 1.1351274 | 0.56197812 | 0.7615 | 1.135127379 | 4.540636927 | 4.540636927 | 0.33684164 | 0.813180267 | 1 | 0.336841643 | 0.336841643 | 0.336841643 | 3.369914031 | 0.527618822 | FALSE | TRUE | FALSE |
| TGIF1 | 1.1348831 | 0.56209329 | 0.5067 | 1.134883113 | 1.134883113 | 2.44451605 | 0.1627179 | 0.861428617 | 1 | 0.162717901 | 0.162717901 | 3.223857546 | 6.974543718 | 0.843515801 | FALSE | TRUE | FALSE |
| BTBD8 | 1.1343764 | 0.56220839 | 0.87905 | 10.56867823 | 100.9947305 | 100.9947305 | 0.17420571 | 0.858538441 | 1 | 0.174205713 | 0.174205713 | 0.174205713 | 6.511706085 | 0.81369479 | FALSE | TRUE | FALSE |
| ARHGAP9 | 1.1328157 | 0.56232345 | 0.82025 | 10.48834495 | 99.54272515 | 967.8509266 | 0.23323425 | 0.843949891 | 0.0214 | 0.233234246 | 0.233234246 | 4.590619496 | 4.856987031 | 0.686366944 | FALSE | TRUE | FALSE |
| C11orf54 | 1.130656 | 0.56243846 | 0.7563 | 1.130656037 | 10.57711487 | 10.57711487 | 13.1771636 | 0.168848166 | 0.1313 | 13.17716357 | 126.9179455 | 126.9179455 | 0.085804204 | -1.066491431 | FALSE | FALSE | FALSE |
| OR52N4 | 1.129142 | 0.56255343 | 0.93625 | 10.45109713 | 965.5356696 | 965.5356696 | 14.3192938 | 0.161188055 | 0.0055 | 1.535287405 | 14.31929379 | 14.31929379 | 0.078854585 | -1.103173052 | FALSE | FALSE | FALSE |
| AQP6 | 1.1277603 | 0.56266835 | 0.3799 | 0.495680167 | 4.509509744 | 4.509509744 | 0.18558864 | 0.855659392 | 0.97005 | 2.968490357 | 11.87655666 | 47.52848014 | 6.076666735 | 0.783665419 | FALSE | TRUE | FALSE |
| OR5A1 | 1.1271799 | 0.56278321 | 0.51205 | 1.127179948 | 1.127179948 | 1.127179948 | 0.15883112 | 0.862343577 | 1 | 1.440523647 | 1.440523647 | 1.440523647 | 7.096719923 | 0.851057666 | FALSE | TRUE | FALSE |
| SLC5A11 | 1.125279 | 0.56289803 | 0.9977 | 53683.17225 | 32848845.64 | 511466106.9 | 22.2668234 | 0.121272037 | 0.97275 | 44999.85045 | 29019251.58 | 465007072.7 | 0.050536128 | -1.29639804 | FALSE | FALSE | FALSE |
| CHI3L1 | 1.1249189 | 0.56301278 | 0.50845 | 2.615332332 | 10.46037242 | 10.46037242 | 1.34689065 | 0.550962918 | 0.0107 | 1.346890648 | 1.346890648 | 6.684633079 | 0.83519694 | -0.078211106 | FALSE | FALSE | FALSE |
| CABP4 | 1.1245026 | 0.56312747 | 0.3817 | 1.124502617 | 10.22353778 | 10.22353778 | 0.10101197 | 0.875683197 | 0.0102 | 0.101011967 | 0.101011967 | 2.005679477 | 11.1323702 | 1.04658764 | FALSE | TRUE | FALSE |
| CDH10 | 1.1238033 | 0.56324209 | 0.505 | 1.123803287 | 1.123803287 | 22.12491375 | 0.15756506 | 0.862626411 | 1 | 0.630064208 | 0.630064208 | 12.42773722 | 7.13231257 | 0.853230368 | FALSE | TRUE | FALSE |
| TREML2 | 1.1227885 | 0.56335667 | 0.5055 | 1.122788537 | 1.122788537 | 1.122788537 | 1.39217453 | 0.54583656 | 0.75 | 1.392174533 | 12.96953378 | 12.96953378 | 0.806499839 | -0.093395715 | FALSE | FALSE | FALSE |
| FAM91A1 | 1.1204028 | 0.56347121 | 0.99945 | 101957.3127 | 46325778.75 | 474038059.7 | 0.14499564 | 0.86564349 | 1 | 1285101.37 | 2080694562 | 23915385238 | 7.727148244 | 0.888019244 | FALSE | TRUE | FALSE |
| ZFYVE27 | 1.1188082 | 0.5635857 | 0.5072 | 1.118808245 | 1.118808245 | 2.436822455 | 0.16178634 | 0.861619685 | 1 | 0.161786342 | 0.161786342 | 3.2077538 | 6.915344221 | 0.839813803 | FALSE | TRUE | FALSE |
| SLC26A2 | 1.1186695 | 0.56370013 | 0.5417 | 1.411023548 | 641.1533405 | 5850.687753 | 1.1484E-05 | 0.900706852 | 0.9981 | 0.047159858 | 3.03226731 | 12.15390147 | 97407.70352 | 4.988593305 | FALSE | TRUE | FALSE |
| PPA2 | 1.1156371 | 0.56381453 | 0.5065 | 1.115637073 | 1.115637073 | 1.115637073 | 0.16094017 | 0.861822153 | 1 | 0.160940168 | 0.160940168 | 0.160940168 | 6.931998924 | 0.840858487 | FALSE | TRUE | FALSE |
| PIK3C2G | 1.1147888 | 0.56392887 | 0.9564 | 1630.155039 | 1711517.196 | 22145394.19 | 47.9175955 | 0.057599529 | 0.101 | 5.286126467 | 444.5183801 | 4217.909145 | 0.023264707 | -1.633302404 | FALSE | FALSE | FALSE |
| CDK12 | 1.113954 | 0.56404316 | 0.88025 | 10.72614374 | 105.930468 | 208.1349656 | 0.15766475 | 0.862614648 | 1 | 1.482670369 | 14.30999918 | 49.97598691 | 7.065333016 | 0.849132637 | FALSE | TRUE | FALSE |
| NKX2-8 | 1.1131574 | 0.56415739 | 0.0073 | 1.113157362 | 1.113157362 | 1.113157362 | 0.32751397 | 0.816147645 | 1 | 0.327513975 | 0.327513975 | 0.327513975 | 3.398808748 | 0.531326727 | FALSE | TRUE | FALSE |
| SLC8B1 | 1.1102441 | 0.56427159 | 0.5056 | 1.110244106 | 10.20576975 | 10.20576975 | 0.02523406 | 0.892396776 | 1 | 0.025234064 | 0.025234064 | 0.498846997 | 43.9978327 | 1.643431284 | FALSE | TRUE | FALSE |
| CUL1 | 1.1098236 | 0.56438573 | 0.99245 | 985.1180674 | 103063.1459 | 272986.7481 | 0.33690535 | 0.813115611 | 1 | 301.3316741 | 3052.90983 | 31669.71844 | 3.294170194 | 0.517746033 | FALSE | TRUE | FALSE |
| APOBEC3D | 1.1076758 | 0.56449983 | 0.0126 | 1.107675761 | 1.107675761 | 2.410039287 | 12.9340833 | 0.169521006 | 0.0084 | 1.378063126 | 12.93408333 | 12.93408333 | 0.085640067 | -1.067323003 | FALSE | FALSE | FALSE |
| NKX2-5 | 1.1063224 | 0.56461388 | 0.50525 | 1.106322442 | 1.106322442 | 1.106322442 | 1.4248298 | 0.543433942 | 0.5047 | 1.424829803 | 1.424829803 | 1.424829803 | 0.776459363 | -0.109881269 | FALSE | FALSE | FALSE |
| HEG1 | 1.1062112 | 0.56472787 | 0.26245 | 0.117033622 | 1.106211221 | 3.940835034 | 0.20286564 | 0.851458608 | 0.51085 | 0.20286564 | 0.20286564 | 3.975191408 | 5.452925484 | 0.736629563 | FALSE | TRUE | FALSE |
| AKR1C1 | 1.1056096 | 0.56484179 | 0.50425 | 1.105609566 | 1.105609566 | 1.105609566 | 0.15558828 | 0.863130435 | 1 | 1.418122976 | 1.418122976 | 1.418122976 | 7.105995216 | 0.851624911 | FALSE | TRUE | FALSE |
| NKD2 | 1.1054371 | 0.56495565 | 0.5036 | 1.10543709 | 1.10543709 | 2.391509446 | 5.24144118 | 0.296017732 | 0.0068 | 1.420838143 | 1.420838143 | 3.064505718 | 0.210903271 | -0.675916684 | FALSE | FALSE | FALSE |
| RSPO3 | 1.1050632 | 0.56506945 | 0.7537 | 2.568526065 | 10.27298007 | 10.27298007 | 0.32493496 | 0.817163742 | 1 | 0.324934958 | 3.0281907 | 3.0281907 | 3.400875208 | 0.531590696 | FALSE | TRUE | FALSE |
| ZNF572 | 1.1049982 | 0.56518318 | 0.7541 | 1.104998225 | 10.29511839 | 10.29511839 | 0.16513597 | 0.860888549 | 1 | 0.165135966 | 0.165135966 | 3.276234999 | 6.691444961 | 0.82551991 | FALSE | TRUE | FALSE |
| ERCC3 | 1.1019151 | 0.56529688 | 0.92915 | 86.42962727 | 29947.78426 | 119835.2537 | 0.08163623 | 0.879889745 | 0.26145 | 0.020415017 | 0.081636226 | 0.402147199 | 13.49786927 | 1.130265218 | FALSE | TRUE | FALSE |
| FUT10 | 1.1003062 | 0.56541054 | 0.5001 | 1.10030622 | 1.10030622 | 2.39665594 | 0.61124501 | 0.707164454 | 0.75795 | 1.397550339 | 5.589668824 | 5.589668824 | 1.800106669 | 0.255298241 | FALSE | TRUE | FALSE |
| LPAR2 | 1.0974323 | 0.56552417 | 0.50905 | 1.097432259 | 1.097432259 | 5.429293682 | 0.31300958 | 0.820998166 | 0.5037 | 2.910091572 | 2.910091572 | 6.201690693 | 3.506066086 | 0.544820098 | FALSE | TRUE | FALSE |
| IZUMO2 | 1.0967985 | 0.56563774 | 0.0069 | 1.096798474 | 1.096798474 | 1.096798474 | 0.16020971 | 0.861988471 | 1 | 0.160209708 | 0.160209708 | 0.160209708 | 6.846017569 | 0.835438009 | FALSE | TRUE | FALSE |
| CA13 | 1.0958656 | 0.56575126 | 0.8135 | 4.383686228 | 17.54002244 | 70.19873848 | 0.88720856 | 0.600155236 | 0.51075 | 0.88720856 | 0.88720856 | 0.88720856 | 1.235183724 | 0.09173156 | FALSE | TRUE | FALSE |
| PRDM4 | 1.0957727 | 0.56586471 | 0.93645 | 10.23466043 | 962.8276148 | 962.8276148 | 0.16876681 | 0.859930734 | 0.5107 | 1.547456695 | 1.547456695 | 5.712230837 | 6.492820881 | 0.812433422 | FALSE | TRUE | FALSE |
| PARP16 | 1.0942931 | 0.56597812 | 0.5015 | 1.094293146 | 1.094293146 | 2.397686476 | 1.36742257 | 0.54939728 | 0.503 | 1.36742257 | 1.36742257 | 1.36742257 | 0.800259678 | -0.096769065 | FALSE | FALSE | FALSE |
| PLEKHF1 | 1.0940731 | 0.56609146 | 0.75005 | 1.094073115 | 4.376760048 | 4.376760048 | 2.67723336 | 0.435665261 | 0.9362 | 25.57560586 | 2516.770806 | 2516.770806 | 0.408658106 | -0.388639882 | FALSE | FALSE | FALSE |
| SLC25A31 | 1.0932898 | 0.56620474 | 0.51265 | 1.093289824 | 10.22482824 | 10.22482824 | 0.19018833 | 0.85430228 | 0.7465 | 0.190188334 | 0.760533512 | 0.760533512 | 5.748458901 | 0.759551431 | FALSE | TRUE | FALSE |
| RAD18 | 1.0929914 | 0.56631796 | 0.5115 | 1.092991448 | 1.092991448 | 2.38381156 | 0.15384937 | 0.863619309 | 0.5004 | 1.404473211 | 1.404473211 | 3.051073749 | 7.10429576 | 0.851521033 | FALSE | TRUE | FALSE |

| | | | | | | | | | | | | | | | | | |
|---|---|---|---|---|---|---|---|---|---|---|---|---|---|---|---|---|---|
| ARHGAP5 | 1.0927939 | 0.56643111 | 0.03875 | 1.092793949 | 1.092793949 | 21.29845134 | 1.41122807 | 0.544237208 | 0.5165 | 1.411228068 | 1.411228068 | 27.58924273 | 0.774356728 | -0.111058924 | FALSE | FALSE | FALSE |
| ZSWIM1 | 1.0926379 | 0.5665442 | 0.50705 | 1.092637903 | 1.092637903 | 2.399166789 | 0.15920899 | 0.862272695 | 1 | 0.159208992 | 0.159208992 | 3.158015378 | 6.862915772 | 0.836508669 | FALSE | TRUE | FALSE |
| UCKL1 | 1.0921903 | 0.56665723 | 0.25915 | 0.27314211 | 1.092190304 | 4.133011055 | 0.18843366 | 0.854827209 | 0.5131 | 0.188433661 | 0.188433661 | 1.597570956 | 5.796152872 | 0.763139831 | FALSE | TRUE | FALSE |
| HRAS | 1.0918152 | 0.5667702 | 0.0063 | 1.091815204 | 1.091815204 | 1.091815204 | 0.32161772 | 0.818249973 | 1 | 0.321617723 | 0.321617723 | 0.321617723 | 3.394760694 | 0.530809165 | FALSE | TRUE | FALSE |
| ODC1 | 1.0905351 | 0.56688312 | 0.01255 | 1.09053508 | 1.09053508 | 5.398540564 | 0.32131307 | 0.81835177 | 1 | 0.321313067 | 0.321313067 | 6.369355966 | 3.393995423 | 0.530711252 | FALSE | TRUE | FALSE |
| TIMM22 | 1.0880187 | 0.566996 | 0.503 | 1.088018713 | 1.088018713 | 1.088018713 | 0.32025903 | 0.818676743 | 1 | 0.320259028 | 0.320259028 | 0.320259028 | 3.397308478 | 0.531134983 | FALSE | TRUE | FALSE |
| IKBIP | 1.0878561 | 0.56710882 | 0.506 | 1.087856076 | 1.087856076 | 1.087856076 | 0.14952708 | 0.864689998 | 1 | 1.352967844 | 12.56476137 | 12.56476137 | 7.275311617 | 0.8618516 | FALSE | TRUE | FALSE |
| MRPL13 | 1.0875284 | 0.56722158 | 0.5016 | 1.087528354 | 1.087528354 | 1.087528354 | 2.87449519 | 0.42106564 | 0.5058 | 2.874495187 | 2.874495187 | 2.874495187 | 0.378337163 | -0.422120997 | FALSE | FALSE | FALSE |
| PAX1 | 1.0869513 | 0.56733428 | 0.5059 | 1.086951311 | 10.87050619 | 111.3693179 | 0.00324993 | 0.898592969 | 0.0138 | 0.003249926 | 0.003249926 | 0.064307509 | 334.4541475 | 2.524336586 | FALSE | TRUE | FALSE |
| CLOCK | 1.0865491 | 0.56744691 | 0.75535 | 1.086549058 | 4.346933072 | 21.41657891 | 0.32403865 | 0.817472488 | 1 | 0.324038652 | 0.324038652 | 6.398131186 | 3.35314646 | 0.525452523 | FALSE | TRUE | FALSE |
| ACVR1C | 1.0863431 | 0.56755949 | 0.89295 | 4.34504048 | 69.56213747 | 278.4358529 | 5.72334686 | 0.276651205 | 0.85565 | 53.47047365 | 856.1481059 | 3427.117751 | 0.18980907 | -0.721683039 | FALSE | FALSE | FALSE |
| DNAH6 | 1.0833749 | 0.56767203 | 0.22335 | 0.00833935 | 86.87243742 | 804.979461 | 0.08857431 | 0.878482405 | 0.14835 | 0.001435753 | 2.52431537 | 61.14939477 | 12.2312534 | 1.087470964 | FALSE | TRUE | FALSE |
| FUBP3 | 1.0821298 | 0.56778453 | 1 | 4.63036E+15 | 1.95341E+21 | 3.92505E+23 | 0.08873806 | 0.878398142 | 1 | 1.90608E+25 | 2.71745E+32 | 7.43131E+34 | 12.19465249 | 1.086169429 | FALSE | TRUE | FALSE |
| ASB13 | 1.0814609 | 0.56789697 | 0.5078 | 1.081460878 | 4.324646526 | 4.324646526 | 0.62302606 | 0.702133868 | 0.5046 | 0.623026058 | 0.623026058 | 0.623026058 | 1.735819657 | 0.239504602 | FALSE | TRUE | FALSE |
| LGI4 | 1.0813119 | 0.56800935 | 0.75725 | 4.323531529 | 41.33540356 | 41.33540356 | 13.3880828 | 0.167174321 | 0.13535 | 5.586856876 | 53.53358641 | 53.53358641 | 0.080766751 | -1.092767389 | FALSE | FALSE | FALSE |
| GAD2 | 1.080779 | 0.56812166 | 0.01745 | 1.080779032 | 1.080779032 | 9.145891671 | 0.31904426 | 0.819061124 | 1 | 0.319044262 | 0.319044262 | 6.308652756 | 3.387552017 | 0.529885973 | FALSE | TRUE | FALSE |
| AKR1B10 | 1.0806053 | 0.56823391 | 0.7538 | 1.080605262 | 9.987815491 | 9.987815491 | 0.1626875 | 0.861452526 | 1 | 0.162687496 | 0.162687496 | 0.162687496 | 6.642214595 | 0.822312903 | FALSE | TRUE | FALSE |
| HGS | 1.0798225 | 0.56834611 | 0.0273 | 1.079822527 | 1.079822527 | 21.15260772 | 0.15209113 | 0.864081805 | 1 | 1.389997094 | 1.389997094 | 27.29600549 | 7.099838999 | 0.8512485 | FALSE | TRUE | FALSE |
| WDR64 | 1.0794627 | 0.56845825 | 0.77585 | 9.482752472 | 786.5109205 | 1686.750441 | 18.5228044 | 0.141328976 | 0.64005 | 18.5228044 | 1585.983452 | 15196.93102 | 0.058277498 | -1.234499105 | FALSE | FALSE | FALSE |
| LPXN | 1.078509 | 0.56857033 | 0.69295 | 1.078509037 | 17.26324347 | 17.26324347 | 0.09568882 | 0.876968346 | 0.50365 | 0.382618033 | 0.382618033 | 0.382618033 | 11.271004 | 1.051962604 | FALSE | TRUE | FALSE |
| DLG2 | 1.0770193 | 0.56868236 | 0.9651 | 223.7542088 | 22404.78621 | 232327.4574 | 0.11234048 | 0.872842686 | 1 | 0.449254845 | 0.449254845 | 8.843557373 | 9.587098415 | 0.981687186 | FALSE | TRUE | FALSE |
| PAPSS1 | 1.0766922 | 0.56879434 | 0.26275 | 0.269169316 | 1.076692159 | 5.313833982 | 0.08871354 | 0.878435606 | 1 | 0.808134271 | 7.555685925 | 15.97866937 | 12.13672798 | 1.084101618 | FALSE | TRUE | FALSE |
| ADAMTS8 | 1.0759885 | 0.56890626 | 0.67995 | 5.366704059 | 421.3506465 | 1889.411023 | 1.30564343 | 0.553939202 | 0.6039 | 2.337679694 | 83.52253486 | 765.1506805 | 0.824105959 | -0.084016946 | FALSE | FALSE | FALSE |
| OLFML2B | 1.0752045 | 0.56901812 | 0.33925 | 0.26888216 | 9.532917727 | 38.12970375 | 2.23158256 | 0.469041309 | 0.33755 | 0.558011126 | 19.99578102 | 79.98655266 | 0.481812549 | -0.317121893 | FALSE | FALSE | FALSE |
| REXO1 | 1.0751614 | 0.56912991 | 0.5227 | 1.075161381 | 1.075161381 | 20.89883064 | 0.15559711 | 0.863107071 | 1 | 0.155597108 | 0.155597108 | 3.034722029 | 6.909905935 | 0.839472135 | FALSE | TRUE | FALSE |
| CAV3 | 1.0748535 | 0.56924165 | 0.9685 | 17.20227146 | 68.84411677 | 275.5854923 | 1232.00254 | 0.002065637 | 0.0914 | 307.6860193 | 2113.692079 | 8467.777447 | 0.000872444 | -3.059262333 | FALSE | FALSE | FALSE |
| SDR9C7 | 1.0741396 | 0.56935333 | 0.6912 | 1.074139587 | 17.19053734 | 17.19053734 | 0.09545523 | 0.877016181 | 1 | 0.381656952 | 0.381656952 | 0.381656952 | 11.25280969 | 1.051260974 | FALSE | TRUE | FALSE |
| COIL | 1.0733516 | 0.56946495 | 0.5119 | 1.07335162 | 1.07335162 | 4.003824209 | 0.15282583 | 0.863874055 | 1 | 0.152825829 | 0.152825829 | 3.026701842 | 7.023365277 | 0.846545256 | FALSE | TRUE | FALSE |
| GNA15 | 1.0700117 | 0.56957655 | 0.63115 | 1.836479147 | 17.11079773 | 17.11079773 | 1.41576242 | 0.543969703 | 0.50445 | 1.41576242 | 1.41576242 | 3.005367178 | 0.755784784 | -0.121601856 | FALSE | FALSE | FALSE |
| IVD | 1.070001 | 0.56968809 | 0.588 | 1.07000096 | 39.292765 | 157.1237852 | 0.06702719 | 0.883106741 | 0.50485 | 0.267936276 | 1.071323279 | 4.284676596 | 15.96368566 | 1.203133168 | FALSE | TRUE | FALSE |
| OSBPL6 | 1.068933 | 0.56979958 | 0.7572 | 1.068933016 | 10.06832542 | 36.05173427 | 0.15753406 | 0.862661689 | 1 | 0.630071044 | 0.630071044 | 12.39900715 | 6.785409095 | 0.831576037 | FALSE | TRUE | FALSE |
| PFN4 | 1.0687621 | 0.569911 | 0.75025 | 2.493578136 | 9.972288346 | 9.972288346 | 1.27349832 | 0.557895609 | 0.7508 | 1.273498325 | 5.094335791 | 5.094335791 | 0.839233252 | -0.076117317 | FALSE | FALSE | FALSE |
| OR4C13 | 1.0673137 | 0.57002238 | 0.9996 | 91501.46526 | 110517354.9 | 1183164614 | 0.18740573 | 0.85514036 | 1 | 152.7546014 | 1500.52953 | 15099.97295 | 5.695203205 | 0.755509224 | FALSE | TRUE | FALSE |
| APOBEC3F | 1.06661 | 0.5701337 | 0.50475 | 1.066609967 | 1.066609967 | 2.319454192 | 0.15482636 | 0.863317075 | 0.0104 | 0.15482636 | 0.15482636 | 3.073117051 | 6.889072166 | 0.838160734 | FALSE | TRUE | FALSE |
| ADH4 | 1.0653897 | 0.57024497 | 0.8781 | 1.065389706 | 93.72611597 | 93.72611597 | 0.16541462 | 0.860864455 | 1 | 0.165414622 | 0.165414622 | 0.165414622 | 6.440722662 | 0.808934599 | FALSE | TRUE | FALSE |
| CDH9 | 1.0647859 | 0.57035619 | 0.63405 | 1.815719962 | 17.02700742 | 35.98872515 | 0.15329191 | 0.863723633 | 1 | 0.612950437 | 0.612950437 | 5.184799025 | 6.946132653 | 0.841743073 | FALSE | TRUE | FALSE |
| SMG8 | 1.0645076 | 0.57046734 | 0.51295 | 1.064507609 | 1.064507609 | 20.88775936 | 0.15429194 | 0.863514836 | 1 | 0.154291943 | 0.154291943 | 3.034405678 | 6.899307835 | 0.838805523 | FALSE | TRUE | FALSE |
| SLC30A5 | 1.063623 | 0.57057845 | 0.63185 | 1.85192146 | 17.01554291 | 21.00118793 | 2.45840539 | 0.450953166 | 0.26255 | 0.614554113 | 2.45840539 | 12.15296259 | 0.432647532 | -0.363865769 | FALSE | FALSE | FALSE |
| PRAMEF12 | 1.0628976 | 0.57068949 | 0.88145 | 9.824349743 | 93.13948808 | 93.13948808 | 0.14647022 | 0.865394961 | 1 | 12.31841612 | 1146.641651 | 1146.641651 | 7.256748767 | 0.860742088 | FALSE | TRUE | FALSE |
| MFSD5 | 1.0612006 | 0.57080049 | 0.2553 | 0.265461 | 1.061200579 | 1.073637458 | 0.04321366 | 0.888263437 | 0.753 | 0.395675091 | 3.718359133 | 3.718359133 | 24.55706302 | 1.390176425 | FALSE | TRUE | FALSE |
| PRRX2 | 1.0611025 | 0.57091143 | 0.75335 | 1.06110251 | 4.244343851 | 4.244343851 | 0.307623 | 0.822951432 | 1 | 0.307623 | 2.892345139 | 2.892345139 | 3.44936013 | 0.537738539 | FALSE | TRUE | FALSE |
| PRRC1 | 1.0610024 | 0.57102231 | 0.8777 | 9.795843048 | 92.76526244 | 92.76526244 | 12.6804042 | 0.173219382 | 0.5007 | 12.68040416 | 120.8221674 | 120.8221674 | 0.083672604 | -1.077416717 | FALSE | FALSE | FALSE |
| EBNA1BP2 | 1.0597942 | 0.57113314 | 0.50965 | 1.059794152 | 1.059794152 | 2.324656177 | 0.15003539 | 0.864552739 | 0.5033 | 1.365790967 | 1.365790967 | 1.365790967 | 7.063627644 | 0.849027798 | FALSE | TRUE | FALSE |
| DLL3 | 1.0594506 | 0.5712439 | 0.67735 | 4.236798316 | 157.3561605 | 629.5900506 | 0.0491573 | 0.886889958 | 1 | 0.049157296 | 0.049157296 | 0.971893749 | 21.55225602 | 1.333492737 | FALSE | TRUE | FALSE |
| LRRN4CL | 1.0587432 | 0.57135461 | 0.25525 | 1.058743191 | 10.06973825 | 10.06973825 | 11.057825 | 0.187075443 | 0.4972 | 1.160420065 | 108.0758543 | 108.0758543 | 0.095746062 | -1.018879081 | FALSE | FALSE | FALSE |
| CEP44 | 1.0582393 | 0.57146526 | 0.50365 | 1.05823931 | 1.05823931 | 1.05823931 | 0.1547014 | 0.863386936 | 1 | 0.154701401 | 0.154701401 | 0.154701401 | 6.84052828 | 0.835089643 | FALSE | TRUE | FALSE |
| OR52I1 | 1.0581782 | 0.57157585 | 0.50435 | 9.852757531 | 94.09609893 | 94.09609893 | 1.40785552 | 0.544771344 | 0.74795 | 1.407855524 | 13.18890707 | 13.18890707 | 0.751624135 | -0.123999283 | FALSE | FALSE | FALSE |
| KRTAP4-3 | 1.0577065 | 0.57168638 | 1 | 1.78661E+12 | 4.33561E+16 | 7.55126E+18 | 0.1506057 | 0.864461088 | 1 | 2.3819E+12 | 5.91607E+16 | 1.04243E+19 | 7.02301754 | 0.846523753 | FALSE | TRUE | FALSE |
| CRYGN | 1.0552872 | 0.57179688 | 0.93735 | 9.891600619 | 937.1806222 | 937.1806222 | 1.40658104 | 0.544904689 | 0.51075 | 13.25533326 | 128.1223325 | 128.1223325 | 0.75024982 | -0.1247941 | FALSE | FALSE | FALSE |
| P4HTM | 1.0522308 | 0.57190736 | 0.5172 | 1.052230839 | 9.632669728 | 11.8758177 | 0.36898213 | 0.802535824 | 0.88 | 1.475849507 | 5.904557401 | 12.4965122 | 2.851712192 | 0.455105692 | FALSE | TRUE | FALSE |
| ACSM4 | 1.0515869 | 0.57201778 | 0.5026 | 1.051586906 | 9.527450041 | 9.527450041 | 1.64343833 | 0.522189568 | 0.81655 | 15.0282947 | 140.8708621 | 1352.786755 | 0.639870027 | -0.193908243 | FALSE | FALSE | FALSE |
| BIN3 | 1.0514904 | 0.57212813 | 0.50385 | 1.051490352 | 1.051490352 | 1.051490352 | 1.22864598 | 0.563766061 | 0.5076 | 1.228645985 | 1.228645985 | 1.228645985 | 0.855812305 | -0.067621474 | FALSE | FALSE | FALSE |
| CACTIN | 1.0510602 | 0.57223843 | 0.5179 | 1.051060234 | 1.051060234 | 20.53198605 | 2.82591371 | 0.42498741 | 0.5139 | 2.825913713 | 2.825913713 | 55.3577302 | 0.371936421 | -0.429531292 | FALSE | FALSE | FALSE |
| SLFN13 | 1.0510107 | 0.57234866 | 0.50095 | 1.051010732 | 9.254841328 | 83.49323105 | 1.80940927 | 0.506441451 | 0.92805 | 1371.724545 | 1280354.297 | 13073724.21 | 0.580858488 | -0.23592966 | FALSE | FALSE | FALSE |
| AMBP | 1.0498318 | 0.57245885 | 0.008 | 0.262542963 | 1.049831755 | 1.049831755 | 0.04570331 | 0.887625212 | 1 | 0.182761011 | 0.182761011 | 0.182761011 | 22.97058523 | 1.36117206 | FALSE | TRUE | FALSE |
| OR52A5 | 1.0498097 | 0.57256897 | 0.96825 | 93.87034407 | 921.4809366 | 9266.761753 | 100.415752 | 0.02916172 | 0.88665 | 9904.848093 | 11564388.93 | 128247422.2 | 0.010454631 | -1.980691283 | FALSE | FALSE | FALSE |
| CPSF2 | 1.0489422 | 0.57267904 | 0.5107 | 1.04894222 | 4.195073652 | 9.023070734 | 1.37260277 | 0.548216637 | 0.51295 | 1.372602768 | 1.372602768 | 2.945322361 | 0.764199406 | -0.116793304 | FALSE | FALSE | FALSE |
| CDC6 | 1.0480326 | 0.57278905 | 0.5159 | 1.048032641 | 1.048032641 | 2.290427005 | 0.15303593 | 0.863781526 | 1 | 0.153035931 | 0.153035931 | 3.034928382 | 6.848278283 | 0.8355814 | FALSE | TRUE | FALSE |
| CYP2A13 | 1.0477524 | 0.57289901 | 0.13575 | 1.047752396 | 9.654797825 | 9.654797825 | 0.02390068 | 0.8927636 | 0.0145 | 0.023900681 | 0.023900681 | 0.472493135 | 43.83776348 | 1.641848389 | FALSE | TRUE | FALSE |
| RIMS3 | 1.0476114 | 0.5730089 | 0.75465 | 1.047611383 | 4.190671885 | 5.196277979 | 0.31374924 | 0.820741015 | 0.00995 | 0.313749237 | 0.313749237 | 0.313749237 | 3.339008544 | 0.52361753 | FALSE | TRUE | FALSE |
| ADH6 | 1.0468346 | 0.57311874 | 0.7479 | 1.046834591 | 9.666925544 | 9.666925544 | 0.15826809 | 0.862496914 | 0.00795 | 0.15826809 | 0.15826809 | 0.15826809 | 6.614312407 | 0.820484704 | FALSE | TRUE | FALSE |
| LYZL2 | 1.0465994 | 0.57322852 | 0.00565 | 1.046599416 | 1.046599416 | 1.046599416 | 0.15216361 | 0.864024158 | 1 | 0.152163615 | 0.152163615 | 0.152163615 | 6.878118782 | 0.837469672 | FALSE | TRUE | FALSE |
| GDPD4 | 1.0456873 | 0.57333825 | 0.6891 | 1.045687302 | 86.06933881 | 86.06933881 | 1.67783501 | 0.519556049 | 0.50425 | 15.20523144 | 141.2520545 | 1344.299342 | 0.623236074 | -0.205347417 | FALSE | FALSE | FALSE |
| FOXI2 | 1.0454426 | 0.57344791 | 0.2504 | 0.26138866 | 1.045442648 | 1.045442648 | 0.35922753 | 0.805877933 | 0.75035 | 0.359227531 | 1.436816687 | 1.436816687 | 2.910252017 | 0.463930599 | FALSE | TRUE | FALSE |
| CLVS1 | 1.0451192 | 0.57355752 | 0.5064 | 1.045119193 | 1.045119193 | 5.189766546 | 0.30988265 | 0.822077845 | 1 | 0.309882647 | 0.309882647 | 0.309882647 | 3.372628973 | 0.527968567 | FALSE | TRUE | FALSE |
| ACTR3B | 1.0445765 | 0.57366707 | 0.50435 | 1.04457645 | 1.04457645 | 5.174882115 | 0.30980706 | 0.822136354 | 1 | 0.309807062 | 0.309807062 | 6.144260789 | 3.371699928 | 0.527848917 | FALSE | TRUE | FALSE |
| TDRD10 | 1.044436 | 0.57377655 | 0.9678 | 91.27997313 | 885.8864653 | 8807.827349 | 1.47759465 | 0.537600919 | 0.74955 | 1.477594653 | 13.76017275 | 13.76017275 | 0.706848819 | -0.150673463 | FALSE | FALSE | FALSE |
| ARMC5 | 1.0428195 | 0.573886 | 0.88155 | 9.75517535 | 93.59958781 | 190.9400106 | 0.16138039 | 0.861726945 | 0.02225 | 0.161380393 | 0.161380393 | 3.166850324 | 6.461872565 | 0.810358389 | FALSE | TRUE | FALSE |
| NLRP5 | 1.0383964 | 0.57399544 | 0.69715 | 1.038396356 | 91.01162726 | 187.1862139 | 0.20970091 | 0.849787401 | 0.0302 | 0.209700905 | 0.209700905 | 4.100544625 | 4.951797202 | 0.69476285 | FALSE | TRUE | FALSE |
| VASN | 1.0383625 | 0.57410482 | 0.51385 | 1.038362542 | 1.038362542 | 20.39490392 | 0.14848655 | 0.86495233 | 0.0206 | 0.148486548 | 0.148486548 | 2.922781165 | 6.992973805 | 0.844661901 | FALSE | TRUE | FALSE |
| EMC9 | 1.0379058 | 0.57421414 | 1 | 39955282.3 | 2.10187E+11 | 9.99896E+12 | 0.17382444 | 0.858675136 | 1 | 144095.0321 | 245256664.6 | 2758817472 | 5.971000387 | 0.776047099 | FALSE | TRUE | FALSE |
| ADAMTS4 | 1.0361809 | 0.57432342 | 0.326 | 0.25906766 | 4.1454001 | 5.079133233 | 1.6970895 | 0.51734049 | 0.1431 | 0.42427822 | 1.697089502 | 8.337920603 | 0.610563485 | -0.214269173 | FALSE | FALSE | FALSE |
| CENPT | 1.035707 | 0.57443265 | 0.50745 | 1.035707041 | 1.035707041 | 3.868929789 | 0.15146497 | 0.864242963 | 1 | 0.151464969 | 0.151464969 | 2.995130846 | 6.837931231 | 0.834924729 | FALSE | TRUE | FALSE |

| | | | | | | | | | | | | | | | | | |
|---|---|---|---|---|---|---|---|---|---|---|---|---|---|---|---|---|---|
| KLHL32 | 1.0351049 | 0.57454182 | 0.5085 | 1.035104919 | 1.035104919 | 3.792006032 | 0.14946055 | 0.86472427 | 1 | 0.149460549 | 0.149460549 | 2.956928928 | 6.925606304 | 0.8404578 | FALSE | TRUE | FALSE |
| OR52K2 | 1.0350625 | 0.57465092 | 0.50065 | 1.035062519 | 1.035062519 | 1.035062519 | 0.15154244 | 0.864208459 | 1 | 0.151542442 | 0.151542442 | 0.151542442 | 6.830182397 | 0.834432301 | FALSE | TRUE | FALSE |
| PFDN5 | 1.0347297 | 0.57475997 | 0.4997 | 0.258707044 | 1.034729724 | 1.034729724 | 0.30631932 | 0.823413745 | 0.004 | 0.306319324 | 0.306319324 | 0.306319324 | 3.377944658 | 0.52865253 | FALSE | TRUE | FALSE |
| TMPRSS15 | 1.0343292 | 0.57486896 | 0.2872 | 0.120541975 | 9.092990682 | 81.85324419 | 1.52800647 | 0.533032222 | 0.37035 | 0.382256206 | 29.95916204 | 274.8808709 | 0.676914118 | -0.169466428 | FALSE | FALSE | FALSE |
| ROCK1 | 1.0341428 | 0.57497789 | 1 | 5.45834E+11 | 2.28613E+15 | 6.26747E+16 | 0.19259633 | 0.853707341 | 1 | 2018720.87 | 901189580.6 | 8419622251 | 5.369483778 | 0.729932535 | FALSE | TRUE | FALSE |
| SUCNR1 | 1.0333073 | 0.57508676 | 0.50565 | 1.033307334 | 1.033307334 | 1.033307334 | 0.15135619 | 0.864300429 | 1 | 0.151356191 | 0.151356191 | 0.151356191 | 6.826990883 | 0.834229323 | FALSE | TRUE | FALSE |
| CES5A | 1.0315494 | 0.5751956 | 0.2284 | 0.118299356 | 4.124394949 | 16.49449478 | 2710.8822 | 0.000817108 | 0.03525 | 8.075514163 | 677.8123863 | 6435.659489 | 0.000380522 | -3.41962061 | FALSE | FALSE | FALSE |
| FANK1 | 1.0301768 | 0.57530439 | 0.5072 | 1.030176834 | 1.030176834 | 1.030176834 | 1.33243154 | 0.551742536 | 0.5023 | 1.33243154 | 1.33243154 | 1.33243154 | 0.773155545 | -0.111733125 | FALSE | FALSE | FALSE |
| LCLAT1 | 1.0291765 | 0.57541313 | 0.01045 | 0.257330516 | 1.029176472 | 1.277203223 | 0.09075353 | 0.877975208 | 1 | 0.090753529 | 0.090753529 | 1.802710127 | 11.34034654 | 1.054626326 | FALSE | TRUE | FALSE |
| MYO1E | 1.0291359 | 0.57552181 | 0.71395 | 4.114068875 | 263.2197095 | 1053.296637 | 0.70865182 | 0.662065285 | 0.6265 | 0.708651816 | 45.30709949 | 181.2567386 | 1.452244714 | 0.162039804 | FALSE | TRUE | FALSE |
| WDR24 | 1.0289616 | 0.57563043 | 0.51485 | 1.02896162 | 1.02896162 | 20.0948853 | 11.5404442 | 0.18165182 | 0.69235 | 11.54044423 | 1123.635137 | 1123.635137 | 0.089161353 | -1.049823351 | FALSE | FALSE | FALSE |
| JAK3 | 1.0283372 | 0.57573899 | 0.15465 | 0.257124358 | 1.028337191 | 4.994933537 | 0.10567315 | 0.874511878 | 0.75805 | 0.105673148 | 0.422555357 | 3.566488985 | 9.731300793 | 0.988170897 | FALSE | TRUE | FALSE |
| TM9SF4 | 1.0263289 | 0.57584752 | 0.75335 | 1.026328944 | 4.105738259 | 20.22117788 | 0.3081929 | 0.822661227 | 1 | 0.308192898 | 0.308192898 | 6.08302382 | 3.33015118 | 0.52246395 | FALSE | TRUE | FALSE |
| ZNF554 | 1.0258803 | 0.57595599 | 0.75575 | 1.025880324 | 9.527255134 | 20.30585434 | 1.32709383 | 0.552649133 | 0.75395 | 1.32709383 | 12.39268153 | 26.30056238 | 0.773027725 | -0.11180493 | FALSE | FALSE | FALSE |
| SCP2D1 | 1.025139 | 0.57606441 | 0.49825 | 0.109052353 | 1.025138991 | 1.025138991 | 0.11871133 | 0.871537895 | 1 | 0.118711327 | 0.118711327 | 0.118711327 | 8.635561691 | 0.936290591 | FALSE | TRUE | FALSE |
| MRPL18 | 1.0251057 | 0.57617277 | 0.50635 | 1.025105702 | 1.025105702 | 1.025105702 | 0.30365391 | 0.824197871 | 1 | 0.303653912 | 0.303653912 | 0.303653912 | 3.375901518 | 0.528389769 | FALSE | TRUE | FALSE |
| MOV10 | 1.0215394 | 0.57628111 | 0.09245 | 0.255483165 | 4.085606067 | 8.619582852 | 0.42131193 | 0.784054125 | 0.1522 | 0.421311935 | 1.684781225 | 8.248981782 | 2.424662933 | 0.384651373 | FALSE | TRUE | FALSE |
| SMURF1 | 1.0210244 | 0.5763894 | 0.02135 | 1.021024364 | 1.021024364 | 20.07368742 | 1.32909057 | 0.552390481 | 0.0204 | 1.329090567 | 1.329090567 | 4.796328482 | 0.768212783 | -0.11451847 | FALSE | FALSE | FALSE |
| CKLF | 1.0203003 | 0.57649763 | 0.5047 | 1.020300284 | 1.020300284 | 1.020300284 | 0.11784379 | 0.871765696 | 1 | 0.117843785 | 0.117843785 | 0.117843785 | 8.658074587 | 0.937421323 | FALSE | TRUE | FALSE |
| IFIT3 | 1.0186994 | 0.57660582 | 0.5093 | 1.018699396 | 1.018699396 | 2.208687285 | 0.14226364 | 0.866361615 | 1 | 1.307250687 | 5.228026179 | 5.228026179 | 7.160644832 | 0.854952133 | FALSE | TRUE | FALSE |
| FOXRED2 | 1.0186625 | 0.57671395 | 0.50845 | 1.018662481 | 1.018662481 | 19.98858428 | 0.14848763 | 0.864940944 | 0.02045 | 0.148487634 | 0.148487634 | 2.920305603 | 6.86025127 | 0.836340023 | FALSE | TRUE | FALSE |
| GLIPR1 | 1.0185227 | 0.57682202 | 0.75245 | 1.018522742 | 9.430813005 | 9.430813005 | 1.31757849 | 0.553165685 | 0.25725 | 1.317578493 | 12.27130712 | 12.27130712 | 0.773026236 | -0.111805766 | FALSE | FALSE | FALSE |
| OR11H1 | 1.0183417 | 0.57693003 | 0.99245 | 994.3627368 | 110519.5696 | 110519.5696 | 1.14828833 | 0.572601402 | 0.96865 | 22.85409248 | 1130.395766 | 11809.75591 | 0.886834505 | -0.052157418 | FALSE | FALSE | FALSE |
| TMEM171 | 1.0178055 | 0.57703798 | 0.50465 | 1.017805547 | 1.017805547 | 1.017805547 | 0.14130628 | 0.866517742 | 1 | 1.274922208 | 11.80608829 | 11.80608829 | 7.202833099 | 0.857503352 | FALSE | TRUE | FALSE |
| SLC43A2 | 1.0177754 | 0.57714588 | 0.518 | 1.017775373 | 9.391887741 | 9.391887741 | 7.0127843 | 0.24505668 | 0.8274 | 100.1820502 | 4364.248032 | 17462.75755 | 0.145131424 | -0.838238543 | FALSE | FALSE | FALSE |
| CLIP3 | 1.0148687 | 0.57725374 | 0.9977 | 260.2018554 | 2712.10376 | 10865.09467 | 1.24892322 | 0.561286099 | 0.96815 | 19.98973138 | 80.00282813 | 320.2670078 | 0.81259492 | -0.090125897 | FALSE | FALSE | FALSE |
| TRDMT1 | 1.0130123 | 0.57736158 | 0.2544 | 0.253275586 | 1.01301228 | 1.01301228 | 0.08937041 | 0.878266844 | 1 | 0.089370406 | 0.089370406 | 0.089370406 | 11.33498574 | 1.054420978 | FALSE | TRUE | FALSE |
| ZP2 | 1.0127331 | 0.57746935 | 0.6942 | 1.012733058 | 90.60997144 | 90.60997144 | 0.02276648 | 0.89299451 | 0.511 | 0.204121867 | 0.204121867 | 0.449863743 | 44.48351229 | 1.648199071 | FALSE | TRUE | FALSE |
| AGK | 1.0123716 | 0.57757707 | 0.50805 | 1.012371554 | 1.012371554 | 2.224635581 | 0.14874331 | 0.864906776 | 1 | 0.148743313 | 0.148743313 | 2.955524344 | 6.806165165 | 0.832902484 | FALSE | TRUE | FALSE |
| ENDOG | 1.0107077 | 0.57768474 | 0.5018 | 1.010707703 | 1.010707703 | 1.010707703 | 1.17377443 | 0.569972668 | 0.7508 | 1.173774432 | 4.695655943 | 4.695655943 | 0.861074901 | -0.064959069 | FALSE | FALSE | FALSE |
| IL17B | 1.0096282 | 0.57779237 | 0.8757 | 4.039058832 | 37.38845888 | 37.38845888 | 0.31292093 | 0.821037661 | 1 | 0.312920934 | 0.312920934 | 0.312920934 | 3.226464299 | 0.508726864 | FALSE | TRUE | FALSE |
| CALR | 1.0094779 | 0.57789994 | 0.50515 | 1.009477941 | 1.009477941 | 4.999649178 | 4.70045763 | 0.320373121 | 0.2584 | 1.175103215 | 4.700457633 | 5.826587431 | 0.214761629 | -0.668043309 | FALSE | FALSE | FALSE |
| PDCD2 | 1.0075322 | 0.57800747 | 0.008 | 1.007532203 | 1.007532203 | 1.007532203 | 1.22765963 | 0.564381866 | 0.8776 | 11.47627856 | 110.0361474 | 110.0361474 | 0.820693439 | -0.085819039 | FALSE | FALSE | FALSE |
| LRRFIP2 | 1.0058831 | 0.57811497 | 0.25735 | 0.251456223 | 4.024770555 | 4.965134038 | 6.73592299 | 0.250277982 | 0.46535 | 3.712462745 | 237.8279138 | 952.094506 | 0.149331139 | -0.825849622 | FALSE | FALSE | FALSE |
| PITPNM2 | 1.0055626 | 0.5782224 | 0.69595 | 2.358585524 | 90.80843604 | 192.61921 | 0.32089342 | 0.818494091 | 0.9841 | 71.28816149 | 2881.244656 | 29835.02271 | 3.133634183 | 0.496048296 | FALSE | TRUE | FALSE |
| SIGLEC8 | 1.0016772 | 0.57832983 | 0.5075 | 1.001677236 | 1.001677236 | 2.199399708 | 0.14750593 | 0.865213699 | 1 | 0.147505935 | 0.147505935 | 2.924812521 | 6.790758876 | 0.83191831 | FALSE | TRUE | FALSE |
| DYNC2LI1 | 1.0012244 | 0.5784372 | 0.49915 | 0.111206024 | 1.001224361 | 1.001224361 | 0.14759281 | 0.865202355 | 1 | 0.147592812 | 0.147592812 | 0.147592812 | 6.783693225 | 0.8314662 | FALSE | TRUE | FALSE |
| SLC13A5 | 0.9994528 | 0.57854454 | 0.51 | 0.999452818 | 0.999452818 | 3.756560754 | 1.29562314 | 0.555606427 | 0.5074 | 1.295623141 | 1.295623141 | 2.825042273 | 0.771407045 | -0.112716399 | FALSE | FALSE | FALSE |
| NMUR1 | 0.9990031 | 0.57865181 | 0.0163 | 0.249761145 | 0.999003077 | 4.937442025 | 1.26568361 | 0.559910127 | 0.9642 | 25.05568413 | 739.5311193 | 2961.005356 | 0.789299211 | -0.102758332 | FALSE | FALSE | FALSE |
| KLK8 | 0.9981398 | 0.57875904 | 0.00845 | 0.9981398 | 0.9981398 | 0.9981398 | 0.1440664 | 0.865857444 | 1 | 0.144066401 | 0.144066401 | 0.144066401 | 6.928331601 | 0.840628666 | FALSE | TRUE | FALSE |
| RPAIN | 0.9980109 | 0.57886621 | 0.00585 | 0.998010935 | 0.998010935 | 0.998010935 | 0.29663218 | 0.826267301 | 1 | 0.296632184 | 0.296632184 | 0.296632184 | 3.364472871 | 0.526917031 | FALSE | TRUE | FALSE |
| KIRREL2 | 0.9979486 | 0.57897331 | 0.51615 | 2.185996737 | 34.99970834 | 140.0988646 | 1.56333078 | 0.529940365 | 0.7084 | 7.827374685 | 501.3251882 | 1870.82918 | 0.638347683 | -0.194942714 | FALSE | FALSE | FALSE |
| HIF1A | 0.9977401 | 0.57908036 | 0.7499 | 0.997740106 | 3.991554926 | 19.65366937 | 1.18861479 | 0.568768042 | 0.5072 | 1.188614788 | 1.188614788 | 10.04411112 | 0.839414179 | -0.076023699 | FALSE | FALSE | FALSE |
| WDR55 | 0.997646 | 0.57918735 | 0.50965 | 0.997646037 | 0.997646037 | 2.197936139 | 0.13449891 | 0.867896951 | 0.87755 | 11.27059921 | 107.2074242 | 107.2074242 | 7.417502586 | 0.870257706 | FALSE | TRUE | FALSE |
| OR6B1 | 0.9955769 | 0.5792943 | 0.0084 | 0.995576877 | 0.995576877 | 0.995576877 | 0.14655349 | 0.865383645 | 1 | 0.146553488 | 0.146553488 | 0.146553488 | 6.793266342 | 0.832078642 | FALSE | TRUE | FALSE |
| MIPOL1 | 0.9909019 | 0.57940126 | 0.8175 | 86.74576857 | 842.5823205 | 8384.249839 | 0.15539265 | 0.863177141 | 0.7518 | 1.410987576 | 13.14963846 | 13.14963846 | 6.376761829 | 0.804600196 | FALSE | TRUE | FALSE |
| CD14 | 0.9900342 | 0.57950817 | 0.505 | 0.990034187 | 0.990034187 | 2.130501424 | 0.14431848 | 0.865823704 | 1 | 0.144318478 | 0.144318478 | 2.855443501 | 6.86006535 | 0.836328253 | FALSE | TRUE | FALSE |
| TPRA1 | 0.9893136 | 0.57961502 | 0.01115 | 0.247353158 | 0.989313558 | 4.903010543 | 0.27383914 | 0.832644371 | 1 | 10.16438057 | 96.81342432 | 96.81342432 | 3.612754395 | 0.557838438 | FALSE | TRUE | FALSE |
| ATG7 | 0.9874379 | 0.57972184 | 0.02045 | 0.987437885 | 0.987437885 | 19.48101982 | 0.1362198 | 0.867557362 | 1 | 1.245406574 | 11.68631364 | 24.61053316 | 7.248857297 | 0.86026955 | FALSE | TRUE | FALSE |
| SLC27A5 | 0.9871756 | 0.5798286 | 0.13975 | 0.987175636 | 9.171468568 | 19.41983819 | 0.19412241 | 0.853388129 | 0.5031 | 0.194122406 | 0.194122406 | 0.737109693 | 5.085325573 | 0.706318763 | FALSE | TRUE | FALSE |
| PPOX | 0.9863062 | 0.57993531 | 0.8524 | 37.320981 | 3605.01972 | 17838.10077 | 0.04894543 | 0.886947452 | 1 | 0.048945434 | 0.048945434 | 0.969530458 | 20.15113783 | 1.304299573 | FALSE | TRUE | FALSE |
| CSRP3 | 0.986046 | 0.58004197 | 0.2581 | 0.986046017 | 3.942570319 | 3.942570319 | 0.14450138 | 0.865778694 | 1 | 0.14450138 | 0.14450138 | 0.14450138 | 6.823782681 | 0.834025187 | FALSE | TRUE | FALSE |
| HFE | 0.9842461 | 0.58014858 | 0.50915 | 0.984246127 | 0.984246127 | 0.984246127 | 1.28135406 | 0.556880849 | 0.50775 | 1.281354062 | 1.281354062 | 1.281354062 | 0.768129712 | -0.114565435 | FALSE | FALSE | FALSE |
| ZNF586 | 0.9834498 | 0.58025515 | 0.75145 | 0.983449765 | 9.098982205 | 9.098982205 | 0.14517365 | 0.865632212 | 0.5017 | 1.31760689 | 1.31760689 | 1.31760689 | 6.774299297 | 0.83086438 | FALSE | TRUE | FALSE |
| BRAP | 0.9825707 | 0.58036167 | 0.0188 | 0.982570681 | 0.982570681 | 8.308980238 | 0.27531777 | 0.832246437 | 1 | 2.557370067 | 24.38049969 | 50.55548838 | 3.56886038 | 0.552529558 | FALSE | TRUE | FALSE |
| OLIG1 | 0.9820378 | 0.58046813 | 0.5039 | 0.982037766 | 0.982037766 | 0.982037766 | 0.28126392 | 0.830883002 | 1 | 0.281263921 | 0.281263921 | 0.281263921 | 3.491517016 | 0.543014163 | FALSE | TRUE | FALSE |
| QSOX1 | 0.9795193 | 0.58057457 | 0.51065 | 0.979519297 | 0.979519297 | 3.650858475 | 1.27484609 | 0.557642337 | 0.50935 | 1.274846094 | 1.274846094 | 4.729955795 | 0.768343176 | -0.114444761 | FALSE | FALSE | FALSE |
| PRSS1 | 0.9791854 | 0.58068095 | 1 | 1.65193E+17 | 6.74317E+22 | 3.91828E+24 | 3.68837031 | 0.372318417 | 1 | 7.88891E+11 | 6.13152E+15 | 1.05097E+18 | 0.265479156 | -0.575969572 | FALSE | FALSE | FALSE |
| ZNF83 | 0.9791539 | 0.58078727 | 0.68835 | 0.979153894 | 82.69409283 | 82.69409283 | 1195.41877 | 0.002148062 | 0.3696 | 124.4903301 | 11753.16903 | 118251.1473 | 0.000819089 | -3.086669115 | FALSE | FALSE | FALSE |
| CHRNA9 | 0.9769164 | 0.58089355 | 0.72765 | 3.90758974 | 136.8479137 | 547.6543853 | 35.1151745 | 0.079332817 | 0.2765 | 8.77546041 | 310.2001781 | 1241.581867 | 0.027820349 | -1.555637432 | FALSE | FALSE | FALSE |
| MAP3K12 | 0.9768999 | 0.58099978 | 0.509 | 0.976899882 | 0.976899882 | 19.17627675 | 0.29191426 | 0.827522659 | 1 | 0.291914263 | 0.291914263 | 5.742971132 | 3.34653015 | 0.524594742 | FALSE | TRUE | FALSE |
| EFNA3 | 0.9751101 | 0.58110597 | 0.5022 | 0.975110086 | 0.975110086 | 0.975110086 | 0.29004692 | 0.828125382 | 1 | 0.290046919 | 0.290046919 | 0.290046919 | 3.361904652 | 0.526585392 | FALSE | TRUE | FALSE |
| TSEN54 | 0.9731422 | 0.58121213 | 0.5063 | 0.973142232 | 0.973142232 | 2.100457797 | 0.14232976 | 0.866328118 | 1 | 0.142329762 | 0.142329762 | 2.819062124 | 6.837236424 | 0.834880598 | FALSE | TRUE | FALSE |
| SLC41A3 | 0.9702404 | 0.58131827 | 0.6341 | 0.97024036 | 34.55262953 | 34.55262953 | 0.17580198 | 0.858128799 | 0.50455 | 0.703351731 | 2.814685605 | 3.48659462 | 5.518938716 | 0.741855572 | FALSE | TRUE | FALSE |
| PAPOLG | 0.9684784 | 0.58142437 | 0.51 | 0.968478423 | 0.968478423 | 8.185039944 | 1.13186364 | 0.57461308 | 0.75965 | 1.131863639 | 4.52815071 | 22.36781517 | 0.855649382 | -0.067704159 | FALSE | FALSE | FALSE |
| MED17 | 0.9649028 | 0.58153045 | 0.51515 | 0.964902756 | 0.964902756 | 3.615509482 | 10.6353325 | 0.191486193 | 0.6889 | 10.63533247 | 989.8935094 | 989.8935094 | 0.090726149 | -1.042267524 | FALSE | FALSE | FALSE |
| ZP3 | 0.9647243 | 0.58163648 | 0.8121 | 3.857703242 | 145.7797958 | 145.7797958 | 2.27130117 | 0.46708613 | 0.6883 | 2.271301173 | 36.36586266 | 36.36586266 | 0.424745203 | -0.371871517 | FALSE | FALSE | FALSE |
| AXDND1 | 0.9644954 | 0.58174246 | 0.51525 | 0.964495415 | 8.713110835 | 18.96955368 | 1.66662596 | 0.520875814 | 0.26155 | 0.186953136 | 1.666625956 | 3.684317983 | 0.578711385 | -0.237537974 | FALSE | FALSE | FALSE |
| GABRB3 | 0.9630077 | 0.58184839 | 0.5057 | 0.963007681 | 0.963007681 | 4.765382862 | 0.2878597 | 0.828778071 | 1 | 0.287859704 | 0.287859704 | 5.704323273 | 3.345406347 | 0.524448877 | FALSE | TRUE | FALSE |
| IRX4 | 0.9624779 | 0.58195426 | 0.01165 | 0.104129857 | 0.962477876 | 2.06399401 | 2.11826585 | 0.479306061 | 0.5045 | 8.474533565 | 33.9124877 | 135.7412905 | 0.454370671 | -0.34258971 | FALSE | FALSE | FALSE |
| ZBTB45 | 0.9603503 | 0.58206011 | 0.51415 | 0.960350296 | 0.960350296 | 18.87551769 | 1.21453292 | 0.565975111 | 0.7542 | 1.21453292 | 11.37322169 | 23.92103409 | 0.79071574 | -0.101979616 | FALSE | FALSE | FALSE |
| TTC29 | 0.9596159 | 0.58216591 | 0.7539 | 0.959615937 | 8.971334146 | 8.971334146 | 0.14555789 | 0.865508031 | 1 | 0.145557893 | 0.145557893 | 0.145557893 | 6.592675369 | 0.819061691 | FALSE | TRUE | FALSE |
| SSH3 | 0.9595018 | 0.58227164 | 0.38535 | 0.425718918 | 15.34354063 | 15.34354063 | 0.08397673 | 0.879328513 | 0.7572 | 0.186189585 | 0.744497379 | 2.840974428 | 11.42580546 | 1.057886825 | FALSE | TRUE | FALSE |

| | | | | | | | | | | | | | | | | | |
|---|---|---|---|---|---|---|---|---|---|---|---|---|---|---|---|---|---|
| C1orf127 | 0.9587978 | 0.58237733 | 0.51485 | 0.958797794 | 0.958797794 | 3.590856075 | 1.10351122 | 0.578003761 | 0.97035 | 97.51830985 | 951.6775429 | 9514.306379 | 0.86886094 | -0.061049726 | FALSE | FALSE | FALSE |
| ATRN | 0.958378 | 0.58248296 | 0.63735 | 1.646891604 | 15.33044952 | 74.78632434 | 0.14046619 | 0.866684637 | 0.5092 | 0.561746734 | 0.561746734 | 10.99367226 | 6.822837877 | 0.833965052 | FALSE | TRUE | FALSE |
| GET4 | 0.9574644 | 0.58258854 | 0.75305 | 0.957464425 | 3.829799338 | 4.75252634 | 4.53497124 | 0.327262505 | 0.25695 | 1.133724636 | 4.534971236 | 4.534971236 | 0.211129107 | -0.67545189 | FALSE | FALSE | FALSE |
| SLC36A4 | 0.9572222 | 0.58269407 | 0.7492 | 0.957222152 | 35.27239485 | 35.27239485 | 6313.90713 | 0.000270172 | 0.11415 | 394.3382882 | 25274.10044 | 245932.205 | 0.000151605 | -3.819285449 | FALSE | FALSE | FALSE |
| MSTN | 0.9565951 | 0.58279955 | 0.50435 | 0.956595123 | 0.956595123 | 0.956595123 | 1.1325025 | 0.574495249 | 0.49435 | 0.283181519 | 1.132502496 | 1.132502496 | 0.844673744 | -0.073311006 | FALSE | FALSE | FALSE |
| MCAT | 0.9560967 | 0.58290497 | 0.0123 | 0.956096741 | 0.956096741 | 4.736599326 | 2.57707965 | 0.441363965 | 0.50705 | 2.577079655 | 2.577079655 | 5.498786175 | 0.371000073 | -0.430626005 | FALSE | FALSE | FALSE |
| ZNF486 | 0.9558357 | 0.58301033 | 0.69155 | 0.95583567 | 80.72348988 | 80.72348988 | 1.60677731 | 0.52508875 | 0.69905 | 1.606777313 | 138.6469973 | 138.6469973 | 0.594877499 | -0.225572458 | FALSE | FALSE | FALSE |
| SCAPER | 0.9551576 | 0.58311564 | 0.75805 | 0.955157558 | 8.92176351 | 32.15090305 | 1.24785722 | 0.561535319 | 0.7588 | 1.247857224 | 11.71262115 | 42.05374429 | 0.765438176 | -0.116089881 | FALSE | FALSE | FALSE |
| PUS1 | 0.9542382 | 0.58322091 | 0.5041 | 0.954238198 | 0.954238198 | 2.061906445 | 0.13521445 | 0.867743787 | 0.5026 | 1.249311999 | 1.249311999 | 2.674700405 | 7.057220799 | 0.848633705 | FALSE | TRUE | FALSE |
| HS6ST3 | 0.9537818 | 0.58332612 | 0.51005 | 0.953781777 | 0.953781777 | 2.022448381 | 0.13810198 | 0.867149942 | 0.01215 | 0.138101977 | 0.138101977 | 2.735083417 | 6.90635863 | 0.839249126 | FALSE | TRUE | FALSE |
| UBR5 | 0.9535165 | 0.58343128 | 0.5383 | 0.953516506 | 7.89975427 | 72.37436328 | 0.08590718 | 0.878995042 | 1 | 0.085907177 | 1.639732894 | 2.855846368 | 11.09938119 | 1.045298767 | FALSE | TRUE | FALSE |
| SAMD3 | 0.9533442 | 0.58353638 | 0.50245 | 0.953344216 | 8.485338157 | 8.485338157 | 0.02110908 | 0.893519969 | 0.8736 | 1.588846298 | 14.32443777 | 14.32443777 | 45.16274783 | 1.654780358 | FALSE | TRUE | FALSE |
| ZDHHC1 | 0.9532004 | 0.58364142 | 0.50225 | 0.953200429 | 0.953200429 | 2.0774592 | 0.1406635 | 0.866673522 | 1 | 0.140663498 | 0.140663498 | 2.785565523 | 6.776459023 | 0.831002816 | FALSE | TRUE | FALSE |
| AIM2 | 0.9528837 | 0.5837464 | 0.93585 | 8.756688134 | 797.4950127 | 797.4950127 | 11.8419171 | 0.179270462 | 0.1302 | 11.84191713 | 112.4644169 | 112.4644169 | 0.08046701 | -1.094382136 | FALSE | FALSE | FALSE |
| FANCC | 0.9516143 | 0.58385135 | 0.1398 | 0.102740853 | 0.951614316 | 0.951614316 | 0.14278215 | 0.866205159 | 0.31855 | 0.142782154 | 12.75145548 | 12.75145548 | 6.664798727 | 0.823787039 | FALSE | TRUE | FALSE |
| DUSP18 | 0.9512542 | 0.58395623 | 0.75245 | 0.951254239 | 3.80496704 | 3.80496704 | 1.13956659 | 0.573431923 | 0.49805 | 0.28495671 | 1.139566585 | 1.139566585 | 0.834750905 | -0.078443101 | FALSE | FALSE | FALSE |
| SSH1 | 0.9510818 | 0.58406106 | 0.51955 | 0.951081796 | 8.744704852 | 18.58788504 | 12.677579 | 0.173554475 | 0.5075 | 12.67757898 | 291.8414475 | 1167.053395 | 0.075020775 | -1.124818456 | FALSE | FALSE | FALSE |
| NME5 | 0.9508575 | 0.58416584 | 0.50025 | 0.95085746 | 0.95085746 | 0.95085746 | 0.28424833 | 0.829783555 | 1 | 0.284248333 | 0.284248333 | 0.284248333 | 3.345164591 | 0.524417491 | FALSE | TRUE | FALSE |
| FAM187B | 0.9501447 | 0.58427056 | 0.5033 | 8.748705729 | 82.62605974 | 82.62605974 | 1.19968286 | 0.567436581 | 0.93825 | 11.10820444 | 1027.000542 | 1027.000542 | 0.791996533 | -0.101276719 | FALSE | FALSE | FALSE |
| DPH7 | 0.9492748 | 0.58437524 | 0.5017 | 0.949274819 | 0.949274819 | 2.09064421 | 0.14083766 | 0.866617925 | 1 | 0.140837658 | 0.140837658 | 0.140837658 | 6.740205942 | 0.828673166 | FALSE | TRUE | FALSE |
| DDHD1 | 0.9492696 | 0.58447985 | 0.7595 | 0.949269567 | 8.882537331 | 32.02881414 | 0.14407225 | 0.865846199 | 1 | 0.144072255 | 0.144072255 | 2.828890393 | 6.588843681 | 0.818809204 | FALSE | TRUE | FALSE |
| DFFB | 0.949239 | 0.58458441 | 0.50675 | 0.94923903 | 0.94923903 | 2.089429685 | 1.23779136 | 0.562901034 | 0.50105 | 1.237791359 | 1.237791359 | 2.711046 | 0.766881287 | -0.11527186 | FALSE | FALSE | FALSE |
| PRKCG | 0.9488657 | 0.58468891 | 0.5093 | 0.94886568 | 0.94886568 | 18.62337745 | 1.1243124 | 0.575436158 | 0.5145 | 1.124312399 | 1.124312399 | 22.11642596 | 0.843951984 | -0.073682261 | FALSE | FALSE | FALSE |
| SLCO4A1 | 0.9485817 | 0.58479336 | 0.8947 | 8.796989396 | 814.4361202 | 8121.119014 | 50.0454794 | 0.054797481 | 0.80195 | 528.2899565 | 207945.8612 | 2136094.3 | 0.018954394 | -1.72229009 | FALSE | FALSE | FALSE |
| SNX8 | 0.9448936 | 0.5848978 | 0.87645 | 3.78036939 | 15.12879583 | 18.67456203 | 0.2891528 | 0.828343493 | 1 | 0.289152796 | 0.289152796 | 5.722844331 | 3.267800246 | 0.514255501 | FALSE | TRUE | FALSE |
| DGKQ | 0.9437233 | 0.58500219 | 0.9999 | 8611848.811 | 6440156581 | 2.21923E+11 | 0.04274765 | 0.888351783 | 1 | 1398350.906 | 1268301544 | 14547853938 | 22.07661312 | 1.343932447 | FALSE | TRUE | FALSE |
| KCNJ15 | 0.942483 | 0.58510655 | 0.25935 | 0.235733034 | 0.942483003 | 1.170769912 | 5.15407956 | 0.300719665 | 0.0678 | 1.288710976 | 20.61841207 | 20.61841207 | 0.182861555 | -0.737877591 | FALSE | FALSE | FALSE |
| DRD4 | 0.9417101 | 0.58521085 | 0.9386 | 18.68905045 | 6998.860421 | 6998.860421 | 1.7856822 | 0.509362819 | 0.50475 | 1.785682204 | 16.23697133 | 16.23697133 | 0.527367136 | -0.277886938 | FALSE | FALSE | FALSE |
| PEX26 | 0.940639 | 0.58531511 | 0.2588 | 0.235280782 | 0.940638988 | 1.168709428 | 0.08419098 | 0.879300786 | 1 | 0.084190981 | 0.084190981 | 0.084190981 | 11.1726811 | 1.048157403 | FALSE | TRUE | FALSE |
| ATP2B2 | 0.9400556 | 0.58541932 | 0.9734 | 1908.979429 | 2422060.19 | 27399561.41 | 0.0036719 | 0.898444515 | 0.99965 | 132.2608689 | 39564.52397 | 491062.5911 | 256.01343 | 2.408262748 | FALSE | TRUE | FALSE |
| SIGLEC10 | 0.9386737 | 0.58552348 | 0.50665 | 0.938673703 | 0.938673703 | 18.48070803 | 1.23060967 | 0.563519204 | 0.50955 | 1.230609674 | 1.230609674 | 4.520951401 | 0.762771269 | -0.117605674 | FALSE | FALSE | FALSE |
| C22orf23 | 0.9386348 | 0.58562759 | 0.9704 | 81.426913 | 787.3384105 | 7799.099225 | 0.15301505 | 0.863816239 | 1 | 1.384986629 | 1.384986629 | 1.384986629 | 6.134264292 | 0.787762483 | FALSE | TRUE | FALSE |
| ZNF667 | 0.9376473 | 0.58573166 | 0.75425 | 0.937647301 | 8.748427506 | 18.5512342 | 0.14276669 | 0.866227531 | 0.01375 | 0.142766686 | 0.142766686 | 2.828275981 | 6.567689733 | 0.817412628 | FALSE | TRUE | FALSE |
| PMAIP1 | 0.9357216 | 0.58583569 | 1 | 0.935721611 | 0.935721611 | 0.935721611 | 0.9430261 | 0.593725097 | 1 | 0.943026102 | 0.943026102 | 0.943026102 | 0.9922542 | -0.003377054 | FALSE | TRUE | FALSE |
| DPY19L3 | 0.9357089 | 0.58593966 | 0.51055 | 0.935708913 | 0.935708913 | 3.489198914 | 10.4029232 | 0.193864599 | 0.68995 | 10.40292323 | 976.8114436 | 976.8114436 | 0.089946729 | -1.046014627 | FALSE | FALSE | FALSE |
| RCC2 | 0.9343323 | 0.58604359 | 0.51045 | 0.934332334 | 0.934332334 | 7.908484658 | 1.11001177 | 0.577306292 | 0.5087 | 1.110011768 | 1.110011768 | 5.495176543 | 0.841731918 | -0.074826205 | FALSE | FALSE | FALSE |
| PM20D1 | 0.9324022 | 0.58614749 | 0.5014 | 0.932402193 | 8.427311863 | 8.427311863 | 0.02211253 | 0.893187488 | 0.01195 | 0.022112533 | 0.022112533 | 0.438248707 | 42.16623092 | 1.624964783 | FALSE | TRUE | FALSE |
| DNAJB2 | 0.9312853 | 0.58625134 | 0.5069 | 0.931285307 | 0.931285307 | 4.617780403 | 0.27948483 | 0.831269746 | 1 | 0.279484832 | 0.279484832 | 0.279484832 | 3.332149732 | 0.522724508 | FALSE | TRUE | FALSE |
| TIMD4 | 0.9310879 | 0.58635514 | 0.87565 | 8.639408667 | 82.22304133 | 82.22304133 | 1.29643082 | 0.555350767 | 0.5106 | 1.29643082 | 1.29643082 | 1.29643082 | 0.718193254 | -0.143758679 | FALSE | FALSE | FALSE |
| ABAT | 0.9309888 | 0.58645888 | 0.26555 | 0.232816847 | 0.930988837 | 4.600168998 | 0.08235135 | 0.879724634 | 0.50945 | 0.329247091 | 0.329247091 | 1.629521002 | 11.30508311 | 1.05327376 | FALSE | TRUE | FALSE |
| TRIOBP | 0.9299798 | 0.58656258 | 0.8377 | 8.138431681 | 6273.449291 | 60163.63353 | 0.30375677 | 0.824159781 | 0.9207 | 23.64935362 | 2023.972353 | 19374.27764 | 3.061593753 | 0.485947563 | FALSE | TRUE | FALSE |
| PLIN2 | 0.9299665 | 0.58666622 | 0.5022 | 0.929966543 | 0.929966543 | 2.056133419 | 0.13861348 | 0.866995163 | 1 | 0.138613485 | 0.138613485 | 2.751654173 | 6.709062579 | 0.826661843 | FALSE | TRUE | FALSE |
| MAP6D1 | 0.9292806 | 0.58676981 | 0.5027 | 0.929280587 | 0.929280587 | 0.929280587 | 0.98840523 | 0.589669174 | 0.4994 | 0.105228348 | 0.988405229 | 0.988405229 | 0.940181779 | -0.02678817 | FALSE | TRUE | FALSE |
| FIBCD1 | 0.9261297 | 0.58687338 | 0.54335 | 1.200433469 | 19.1866954 | 76.73509555 | 7.8904438 | 0.230215613 | 0.1338 | 1.973731407 | 75.03757204 | 75.03757204 | 0.117373586 | -0.930429628 | FALSE | FALSE | FALSE |
| ZDHHC17 | 0.9257841 | 0.5869769 | 0.5067 | 0.925784069 | 0.925784069 | 7.829421859 | 0.27784607 | 0.831584904 | 1 | 0.277846071 | 0.277846071 | 5.500037034 | 3.332003458 | 0.522705444 | FALSE | TRUE | FALSE |
| ZNF518B | 0.9249264 | 0.58708037 | 0.757 | 0.924926443 | 8.596032431 | 31.21601146 | 0.14138213 | 0.866484315 | 1 | 0.141382129 | 0.141382129 | 2.77978429 | 6.542032236 | 0.81571268 | FALSE | TRUE | FALSE |
| KLHL42 | 0.9231604 | 0.58718381 | 0.5108 | 0.923160437 | 0.923160437 | 4.561461367 | 1.09727643 | 0.579161204 | 0.50805 | 1.097276427 | 1.097276427 | 5.429472684 | 0.841319848 | -0.075038865 | FALSE | FALSE | FALSE |
| FIGNL1 | 0.9221183 | 0.5872872 | 0.46375 | 0.527753083 | 33.69939244 | 134.7625835 | 0.2648916 | 0.835327843 | 0.8926 | 2.493010251 | 237.6924209 | 2405.171004 | 3.481115622 | 0.541718448 | FALSE | TRUE | FALSE |
| SMR3B | 0.9175376 | 0.5873906 | 1 | 0.917537641 | 0.917537641 | 0.917537641 | 0.92685587 | 0.594809515 | 1 | 0.926855871 | 0.926855871 | 0.926855871 | 0.989946408 | -0.004388316 | FALSE | TRUE | FALSE |
| MFNG | 0.9174599 | 0.58749394 | 0.5107 | 0.917459921 | 3.670570313 | 14.68887294 | 1.50043936 | 0.535811122 | 0.19945 | 1.500439358 | 6.003342001 | 24.02570842 | 0.611460847 | -0.213631347 | FALSE | FALSE | FALSE |
| LRRC52 | 0.917221 | 0.58759723 | 0.50745 | 0.917220975 | 0.917220975 | 0.917220975 | 1.20093389 | 0.567072041 | 0.50225 | 1.200933893 | 1.200933893 | 1.200933893 | 0.763756424 | -0.117045124 | FALSE | FALSE | FALSE |
| TMEM167B | 0.9154207 | 0.58770048 | 1 | 0.915420671 | 0.915420671 | 0.915420671 | 0.92500063 | 0.595242119 | 1 | 0.925000627 | 0.925000627 | 0.925000627 | 0.989643298 | -0.004521312 | FALSE | TRUE | FALSE |
| GFM1 | 0.9145512 | 0.58780368 | 0.2279 | 0.234257538 | 4.73642537 | 18.93371435 | 1.01181071 | 0.586890048 | 0.83775 | 9.285551585 | 841.8379192 | 8306.843603 | 0.903875829 | -0.043891227 | FALSE | FALSE | FALSE |
| SPRR2B | 0.9141408 | 0.58790683 | 1 | 0.914140766 | 0.914140766 | 0.914140766 | 8.47722806 | 0.218877668 | 0.50285 | 8.477228057 | 8.477228057 | 8.477228057 | 0.107834868 | -0.96724079 | FALSE | FALSE | FALSE |
| CERS3 | 0.9137156 | 0.58800993 | 0.0116 | 0.913715643 | 0.913715646 | 1.935349583 | 0.13247648 | 0.86833276 | 0.0089 | 0.132476484 | 0.132476484 | 0.132476484 | 6.89719123 | 0.838672267 | FALSE | TRUE | FALSE |
| IQSEC1 | 0.9136479 | 0.58811297 | 0.5109 | 0.913647907 | 0.913647907 | 17.75449484 | 0.1337734 | 0.868038886 | 1 | 0.133773397 | 0.133773397 | 2.608448123 | 6.829817642 | 0.834409108 | FALSE | TRUE | FALSE |
| CNDP2 | 0.9123368 | 0.58821598 | 0.0153 | 0.228077577 | 0.912336802 | 4.509594911 | 0.08130379 | 0.879990437 | 1 | 0.081303788 | 0.081303788 | 1.609757446 | 11.22133206 | 1.050044414 | FALSE | TRUE | FALSE |
| IGIP | 0.911671 | 0.58831893 | 1 | 0.911671024 | 0.911671024 | 0.911671024 | 0.92132448 | 0.59621202 | 1 | 0.921324481 | 0.921324481 | 0.921324481 | 0.989522197 | -0.004574459 | FALSE | TRUE | FALSE |
| RPS27 | 0.9105533 | 0.58842184 | 1 | 0.910553341 | 0.910553341 | 0.910553341 | 0.92068055 | 0.596426903 | 1 | 0.920680553 | 0.920680553 | 0.920680553 | 0.989000297 | -0.004803578 | FALSE | TRUE | FALSE |
| ZBTB10 | 0.9094888 | 0.58852471 | 1 | 1.31253E+20 | 7.76406E+25 | 1.77152E+28 | 1.68051408 | 0.518967173 | 1 | 3.88621E+13 | 1.06369E+18 | 1.96406E+20 | 0.541196763 | -0.266644809 | FALSE | FALSE | FALSE |
| ZIC4 | 0.9078422 | 0.58862754 | 0.50615 | 0.907842247 | 3.630840296 | 7.899542464 | 0.1346432 | 0.867864158 | 1 | 0.538503624 | 0.538503624 | 2.664934073 | 6.742577893 | 0.828825972 | FALSE | TRUE | FALSE |
| OSBP2 | 0.9077531 | 0.58873031 | 0.50675 | 0.907753077 | 8.371779519 | 17.72597841 | 0.15043984 | 0.864495469 | 0.93915 | 3.151627957 | 119.8579356 | 119.8579356 | 6.033993814 | 0.780604861 | FALSE | TRUE | FALSE |
| MTNR1B | 0.9076161 | 0.58883303 | 0.9385 | 8.459229678 | 135.3998545 | 135.3998545 | 0.28193795 | 0.830582978 | 1 | 1.127574724 | 4.510718172 | 5.590653464 | 3.219205268 | 0.50774867 | FALSE | TRUE | FALSE |
| EMC10 | 0.907156 | 0.5889357 | 0.7493 | 0.907156043 | 8.347705329 | 8.347705329 | 10.3236291 | 0.195220478 | 0.69075 | 10.32362909 | 951.7428496 | 951.7428496 | 0.087871817 | -1.056150395 | FALSE | FALSE | FALSE |
| GABRB1 | 0.9070992 | 0.58903831 | 0.5095 | 0.907099249 | 0.907099249 | 7.67901933 | 0.27348533 | 0.832765106 | 1 | 0.273485331 | 0.273485331 | 5.41687379 | 3.316811342 | 0.52072077 | FALSE | TRUE | FALSE |
| MPL | 0.9060974 | 0.58914087 | 0.01345 | 0.226511034 | 0.906097375 | 1.120282988 | 0.08670555 | 0.878855621 | 0.9388 | 1.754137341 | 65.55199076 | 65.55199076 | 10.45028077 | 1.019127959 | FALSE | TRUE | FALSE |
| CDC45 | 0.9053823 | 0.58924339 | 0.2624 | 0.905382288 | 3.621435126 | 7.93550007 | 10.8196091 | 0.189110558 | 0.254 | 1.164220203 | 10.81960906 | 23.01742813 | 0.08367976 | -1.077379575 | FALSE | FALSE | FALSE |
| UBE3B | 0.9051881 | 0.58934586 | 0.6894 | 0.90518808 | 79.55188078 | 163.5016973 | 0.18568353 | 0.855620591 | 0.51505 | 0.185683527 | 0.185683527 | 3.632996599 | 4.874897055 | 0.687965449 | FALSE | TRUE | FALSE |
| ANGPTL1 | 0.9051721 | 0.58944826 | 0.00845 | 0.18256773 | 0.18256773 | 0.905172088 | 0.55813115 | 0.729530799 | 0.7536 | 2.23183873 | 20.76922086 | 20.76922086 | 1.62179103 | 0.209994894 | FALSE | TRUE | FALSE |
| SMAD7 | 0.904658 | 0.58955062 | 0.3866 | 0.90465799 | 8.541991485 | 8.541991485 | 0.76029692 | 0.636245512 | 0.2602 | 0.760296918 | 3.039049327 | 3.039049327 | 1.189874598 | 0.075501193 | FALSE | TRUE | FALSE |
| RNF217 | 0.904427 | 0.58965292 | 0.87685 | 8.33149673 | 78.72116203 | 78.72116203 | 1.26079788 | 0.560286535 | 0.5003 | 1.260797879 | 1.260797879 | 1.260797879 | 0.71734499 | -0.144271931 | FALSE | FALSE | FALSE |
| OXSM | 0.9031077 | 0.58975518 | 0.0107 | 0.22582716 | 0.903107744 | 1.120913284 | 0.08003511 | 0.880327975 | 1 | 0.32000826 | 0.32000826 | 1.589775674 | 11.28389441 | 1.052459013 | FALSE | TRUE | FALSE |
| ALG2 | 0.9029893 | 0.58985739 | 0.6315 | 0.902989307 | 33.77101363 | 33.77101363 | 0.64981888 | 0.689776625 | 0.2642 | 0.162509153 | 0.649818878 | 0.649818878 | 1.38960153 | 0.142890284 | FALSE | TRUE | FALSE |

| | | | | | | | | | | | | | | | | | |
|---|---|---|---|---|---|---|---|---|---|---|---|---|---|---|---|---|---|
| MESP2 | 0.9014244 | 0.58995956 | 0.8758 | 8.322802101 | 78.86843064 | 78.86843064 | 0.28541956 | 0.829461649 | 0.5001 | 1.141585433 | 1.141585433 | 1.141585433 | 3.158243293 | 0.499445582 | FALSE | TRUE | FALSE |
| PTMS | 0.9014077 | 0.59006167 | 1 | 0.901407735 | 0.901407735 | 0.901407735 | 0.88422752 | 0.600682729 | 1 | 8.381741651 | 8.381741651 | 8.381741651 | 1.019429629 | 0.008357252 | FALSE | TRUE | FALSE |
| BLCAP | 0.9012349 | 0.59016373 | 1 | 0.901234868 | 0.901234868 | 0.901234868 | 0.91233575 | 0.596641706 | 1 | 0.912335754 | 0.912335754 | 0.912335754 | 0.987832456 | -0.005316709 | FALSE | TRUE | FALSE |
| BARX2 | 0.9006371 | 0.59026574 | 0.69 | 0.900637054 | 14.40536902 | 14.40536902 | 1.29516896 | 0.555734137 | 0.2591 | 0.323935203 | 1.295168959 | 1.295168959 | 0.695381902 | -0.157776617 | FALSE | FALSE | FALSE |
| ZNF706 | 0.9001891 | 0.59036769 | 1 | 0.900189146 | 0.900189146 | 0.900189146 | 0.91132311 | 0.596963562 | 1 | 0.911323105 | 0.911323105 | 0.911323105 | 0.987782644 | -0.005338609 | FALSE | TRUE | FALSE |
| HOPX | 0.8998417 | 0.5904696 | 0.93575 | 8.337418823 | 317.1568399 | 317.1568399 | 0.28834727 | 0.828633458 | 1 | 1.153197792 | 1.153197792 | 1.153197792 | 3.120687486 | 0.494250279 | FALSE | TRUE | FALSE |
| NCKAP1L | 0.8992742 | 0.59057145 | 0.8808 | 8.368829024 | 79.88261266 | 163.7301398 | 1.21945329 | 0.565241178 | 0.7576 | 1.219453289 | 11.41862911 | 23.92033961 | 0.737440466 | -0.132273035 | FALSE | FALSE | FALSE |
| POLR3D | 0.8974454 | 0.59067328 | 0.51045 | 0.897445428 | 0.897445428 | 1.978438204 | 0.13406231 | 0.867951575 | 0.01 | 0.134062312 | 0.134062312 | 0.159348017 | 6.694241001 | 0.825701343 | FALSE | TRUE | FALSE |
| ASB15 | 0.8974332 | 0.59077504 | 0.9804 | 731.4409153 | 71890.59566 | 737437.0924 | 13113.8527 | 0.000156518 | 0.14475 | 136.725867 | 13113.85266 | 132986.3583 | 6.8434E-05 | -4.164728171 | FALSE | FALSE | FALSE |
| MRPS14 | 0.8970948 | 0.59087676 | 0.50145 | 0.897094801 | 0.897094801 | 0.897094801 | 0.26711076 | 0.834639762 | 1 | 1.068320561 | 1.068320561 | 1.068320561 | 3.358512418 | 0.526146958 | FALSE | TRUE | FALSE |
| DOK6 | 0.8967884 | 0.59097842 | 0.50375 | 0.896788417 | 0.896788417 | 4.452563911 | 1.05827561 | 0.582601238 | 0.7528 | 1.058275611 | 4.233111066 | 4.233111066 | 0.847405353 | -0.071908797 | FALSE | FALSE | FALSE |
| CCL18 | 0.8963714 | 0.59108003 | 0.49655 | 0.096437974 | 0.896371445 | 0.896371445 | 0.10933799 | 0.873571743 | 1 | 0.109337994 | 0.109337994 | 0.109337994 | 8.198169873 | 0.913716913 | FALSE | TRUE | FALSE |
| SH3RF1 | 0.8959839 | 0.59118159 | 0.5052 | 0.895983908 | 0.895983908 | 17.5859974 | 0.13211088 | 0.868430443 | 1 | 0.132110879 | 0.132110879 | 2.598831184 | 6.782060001 | 0.831361628 | FALSE | TRUE | FALSE |
| NRG3 | 0.8937871 | 0.59128312 | 0.81425 | 79.06338593 | 771.9661992 | 7721.594216 | 1.33182715 | 0.551872283 | 0.51345 | 1.331827154 | 1.331827154 | 4.933585742 | 0.671098394 | -0.1732138 | FALSE | FALSE | FALSE |
| PRSS50 | 0.8924082 | 0.59138462 | 0.49895 | 0.392384941 | 3.568980901 | 3.568980901 | 0.1325216 | 0.868300168 | 0.5073 | 0.530000988 | 0.530000988 | 2.628658397 | 6.734058694 | 0.828276897 | FALSE | TRUE | FALSE |
| SMTNL2 | 0.8919445 | 0.59148606 | 0.50415 | 0.891944451 | 3.567558849 | 4.425938389 | 0.31687106 | 0.819744902 | 0.7539 | 0.316871059 | 1.267178293 | 1.574127566 | 2.814849845 | 0.449455233 | FALSE | TRUE | FALSE |
| CABYR | 0.8914344 | 0.59158745 | 0.6932 | 0.891434438 | 72.85701096 | 72.85701096 | 0.18576396 | 0.855581769 | 0.50045 | 0.185763959 | 0.185763959 | 0.185763959 | 4.79874806 | 0.68112795 | FALSE | TRUE | FALSE |
| GNRH2 | 0.8912305 | 0.59168879 | 0.30465 | 0.891230468 | 2.7606E+306 | 1.2442E+308 | 1.6E+288 | 0 | 0.21545 | 5.331E+276 | 2.6529E+301 | 2.5625E+306 | 5.5701E-289 | -288.2541348 | FALSE | FALSE | FALSE |
| TCN1 | 0.8909368 | 0.59179008 | 0.74955 | 0.890936779 | 8.20976812 | 8.20976812 | 0.13340324 | 0.868126087 | 1 | 1.206424726 | 1.206424726 | 2.649609037 | 6.678524218 | 0.824680505 | FALSE | TRUE | FALSE |
| CCNDBP1 | 0.8909214 | 0.59189131 | 0.5048 | 0.890921407 | 0.890921407 | 0.890921407 | 0.1334032 | 0.868136979 | 1 | 0.1334032 | 0.1334032 | 0.1334032 | 6.678411045 | 0.824673146 | FALSE | TRUE | FALSE |
| RPL39L | 0.8906465 | 0.59199249 | 1 | 0.890646451 | 0.890646451 | 0.890646451 | 0.90291 | 0.598352622 | 1 | 0.902910001 | 0.902910001 | 0.902910001 | 0.986417749 | -0.005939122 | FALSE | TRUE | FALSE |
| GRK7 | 0.8898249 | 0.59209362 | 0.5039 | 0.889824904 | 0.889824904 | 1.933338955 | 1.17175575 | 0.570332828 | 0.51395 | 1.171755752 | 1.171755752 | 2.536998177 | 0.759394527 | -0.119532537 | FALSE | FALSE | FALSE |
| TMEM132D | 0.8896171 | 0.5921947 | 1 | 2.60372E+14 | 7.44967E+18 | 1.40358E+21 | 0.05688134 | 0.885300558 | 1 | 4.64954E+28 | 7.69452E+35 | 3.79532E+39 | 15.63987792 | 1.194233359 | FALSE | TRUE | FALSE |
| SNURF | 0.8893408 | 0.59229573 | 1 | 0.889340781 | 0.889340781 | 0.889340781 | 0.90179694 | 0.598671801 | 1 | 0.901796944 | 0.901796944 | 0.901796944 | 0.986187398 | -0.006040552 | FALSE | TRUE | FALSE |
| ERI1 | 0.8888677 | 0.59239671 | 0.74775 | 2.041254244 | 8.1646284 | 8.1646284 | 0.27608359 | 0.832038084 | 1 | 0.27608359 | 0.27608359 | 0.27608359 | 3.219559986 | 0.507796521 | FALSE | TRUE | FALSE |
| TM7SF3 | 0.8886248 | 0.59249763 | 0.50835 | 0.888624757 | 0.888624757 | 3.348437745 | 0.13307326 | 0.868202298 | 1 | 0.133073263 | 0.133073263 | 2.638940538 | 6.677710716 | 0.824627601 | FALSE | TRUE | FALSE |
| SH3PXD2B | 0.8883403 | 0.5925985 | 0.5169 | 0.888340274 | 0.888340274 | 17.37604399 | 0.13019748 | 0.868906051 | 1 | 0.520740016 | 0.520740016 | 10.21272474 | 6.823021781 | 0.833976758 | FALSE | TRUE | FALSE |
| SYNGR2 | 0.887971 | 0.59269932 | 1 | 7.23487E+13 | 6.30411E+18 | 1.17858E+21 | 0.10277285 | 0.875232864 | 1 | 2.43886E+18 | 1.28216E+24 | 1.87943E+25 | 8.64013177 | 0.936520366 | FALSE | TRUE | FALSE |
| HTN1 | 0.887903 | 0.59280009 | 1 | 0.887902962 | 0.887902962 | 0.887902962 | 0.90045148 | 0.598884317 | 1 | 0.900451483 | 0.900451483 | 0.900451483 | 0.98606419 | -0.006094813 | FALSE | TRUE | FALSE |
| ZNF764 | 0.8878103 | 0.5929008 | 0.8768 | 8.222278703 | 78.10541634 | 78.10541634 | 1.24258178 | 0.562405322 | 0.5068 | 1.242581783 | 1.242581783 | 2.70050913 | 0.714488391 | -0.146004823 | FALSE | FALSE | FALSE |
| LPAR5 | 0.8869699 | 0.59300147 | 0.74985 | 2.068731518 | 8.273271359 | 8.273271359 | 2.47549794 | 0.44810862 | 0.5071 | 2.475497945 | 9.900245051 | 9.900245051 | 0.358299581 | -0.445753699 | FALSE | FALSE | FALSE |
| GCDH | 0.8868957 | 0.59310208 | 0.18325 | 0.000306418 | 7.976064963 | 31.89323534 | 1.6336E-05 | 0.900629134 | 0.63435 | 6.5265E-05 | 0.519295274 | 4.699177442 | 54289.72491 | 4.734717641 | FALSE | TRUE | FALSE |
| TRIM16 | 0.8854668 | 0.59320265 | 0.93715 | 8.272749956 | 778.7067739 | 778.7067739 | 0.13155254 | 0.868560476 | 1 | 11.31709629 | 109.0711138 | 109.0711138 | 6.730898895 | 0.828073067 | FALSE | TRUE | FALSE |
| NEDD8 | 0.8852153 | 0.59330317 | 1 | 0.885215327 | 0.885215327 | 0.885215327 | 0.89694356 | 0.599308694 | 1 | 0.896943561 | 0.896943561 | 0.896943561 | 0.986924222 | -0.005716192 | FALSE | TRUE | FALSE |
| PLGRKT | 0.8850736 | 0.59340364 | 0.5004 | 0.885073646 | 0.885073646 | 0.885073646 | 1.02281346 | 0.58621796 | 0.75265 | 1.022813462 | 9.426491367 | 9.426491367 | 0.865332418 | -0.062817026 | FALSE | FALSE | FALSE |
| NABP1 | 0.8840132 | 0.59350407 | 0.0064 | 0.884013191 | 0.884013191 | 0.884013191 | 1.01931103 | 0.586330094 | 0.93665 | 4.077862109 | 65.31451902 | 65.31451902 | 0.867265402 | -0.061847979 | FALSE | FALSE | FALSE |
| CKAP4 | 0.8827968 | 0.59360446 | 0.5083 | 0.882796799 | 0.882796799 | 4.368921461 | 1.05532523 | 0.58294254 | 0.50455 | 1.055325226 | 1.055325226 | 5.229172152 | 0.836516343 | -0.07752557 | FALSE | FALSE | FALSE |
| SBK2 | 0.8822916 | 0.59370479 | 0.5117 | 1.964571375 | 31.44706533 | 31.44706533 | 0.71416779 | 0.658985984 | 0.49995 | 0.079596346 | 0.714167791 | 0.714167791 | 1.235412152 | 0.091811869 | FALSE | TRUE | FALSE |
| CCDC85B | 0.8822385 | 0.59380508 | 1 | 0.882238525 | 0.882238525 | 0.882238525 | 0.89541931 | 0.599414679 | 1 | 0.89541931 | 0.89541931 | 0.89541931 | 0.985279762 | -0.006440438 | FALSE | TRUE | FALSE |
| MIEF2 | 0.8811064 | 0.59390532 | 0.13395 | 0.881106402 | 8.614224244 | 8.614224244 | 1.01145381 | 0.587001882 | 0.13775 | 1.01145381 | 9.91411169 | 9.91411169 | 0.87112866 | -0.059917698 | FALSE | FALSE | FALSE |
| MLLT11 | 0.8810224 | 0.59400551 | 1 | 0.881022381 | 0.881022381 | 0.881022381 | 0.89432512 | 0.599626482 | 1 | 0.894325121 | 0.894325121 | 0.894325121 | 0.985125387 | -0.006508489 | FALSE | TRUE | FALSE |
| IZUMO1 | 0.8801078 | 0.59410565 | 0.2596 | 0.880107754 | 8.132590352 | 8.132590352 | 0.1357759 | 0.867645149 | 1 | 0.1357759 | 0.1357759 | 2.695769258 | 6.482061674 | 0.811713159 | FALSE | TRUE | FALSE |
| DENND4B | 0.8800508 | 0.59420574 | 0.88025 | 8.293195426 | 80.157843 | 278.799939 | 1.23840229 | 0.562777208 | 0.5226 | 1.238402292 | 1.238402292 | 23.98973889 | 0.710634051 | -0.148353986 | FALSE | FALSE | FALSE |
| PRSS48 | 0.8793006 | 0.59430579 | 0.5085 | 0.87930057 | 0.87930057 | 0.87930057 | 1.15641982 | 0.571767973 | 0.5006 | 1.156419816 | 1.156419816 | 1.156419816 | 0.760364496 | -0.11897817 | FALSE | FALSE | FALSE |
| TAS2R3 | 0.8791468 | 0.59440578 | 0.99155 | 729.9811657 | 72842.40577 | 72842.40577 | 88553554.9 | 1.86564E-08 | 0.30085 | 774034.6133 | 6766660123 | 77226628212 | 9.92785E-09 | -8.003144578 | FALSE | FALSE | FALSE |
| ASIC1 | 0.8780786 | 0.59450573 | 0.92455 | 75.63182354 | 2917.769029 | 11675.11033 | 1.35720267 | 0.550181304 | 0.3884 | 1.357202667 | 12.5070367 | 12.53646443 | 0.646976796 | -0.189111295 | FALSE | FALSE | FALSE |
| OCA2 | 0.8770259 | 0.59460564 | 0.7184 | 3.50744598 | 224.6887826 | 899.4839614 | 2.43190376 | 0.452270819 | 0.3827 | 0.608114144 | 38.92222554 | 155.7701161 | 0.360633455 | -0.442933988 | FALSE | FALSE | FALSE |
| XKR8 | 0.875375 | 0.59470552 | 0.5041 | 0.875374968 | 3.500754667 | 3.500754667 | 0.13166296 | 0.868517159 | 1 | 0.131662965 | 0.131662965 | 0.131662965 | 6.648604407 | 0.822730493 | FALSE | TRUE | FALSE |
| KBTBD3 | 0.8749466 | 0.59480534 | 0.5075 | 0.874946616 | 0.874946616 | 3.236270571 | 0.12584684 | 0.869943191 | 0.50605 | 1.156065988 | 1.156065988 | 2.495172931 | 6.95247163 | 0.842139225 | FALSE | TRUE | FALSE |
| ZNF776 | 0.8720078 | 0.59490515 | 0.7575 | 0.872007768 | 8.049036748 | 17.2950369 | 0.13077569 | 0.868765872 | 1 | 1.184458039 | 1.184458039 | 2.596345943 | 6.66796517 | 0.823993323 | FALSE | TRUE | FALSE |
| POLR3E | 0.8661893 | 0.59500498 | 0.02385 | 0.8661893 | 0.8661893 | 16.99703611 | 0.26208111 | 0.836109637 | 1 | 0.262081109 | 0.262081109 | 5.154438547 | 3.305042868 | 0.519177097 | FALSE | TRUE | FALSE |
| NFATC3 | 0.8657941 | 0.59510477 | 0.5099 | 0.865794054 | 0.865794054 | 17.01402414 | 0.12894402 | 0.869196293 | 1 | 0.128944015 | 0.128944015 | 2.539242494 | 6.714495848 | 0.82701341 | FALSE | TRUE | FALSE |
| SMYD5 | 0.8655738 | 0.5952045 | 0.50695 | 0.865573784 | 0.865573784 | 1.880208919 | 0.1287733 | 0.869271343 | 0.0114 | 0.1287733 | 0.1287733 | 2.554141949 | 6.72168674 | 0.827478269 | FALSE | TRUE | FALSE |
| NDUFC2 | 0.8654764 | 0.59530417 | 1 | 8.211571617 | 8.211571617 | 8.211571617 | 0.64620079 | 0.691992642 | 1 | 0.646200786 | 0.646200786 | 0.646200786 | 1.339330409 | 0.126887729 | FALSE | TRUE | FALSE |
| PTCHD4 | 0.8653468 | 0.5954038 | 0.8501 | 8.17936169 | 1575.956608 | 8022.072758 | 0.04688567 | 0.887381081 | 0.94065 | 1.032446045 | 40.20580295 | 40.20580295 | 18.45653107 | 1.266150078 | FALSE | TRUE | FALSE |
| TAF5 | 0.8652033 | 0.59550337 | 0.7552 | 0.865203269 | 3.460899696 | 17.05475974 | 0.26527369 | 0.83526097 | 1 | 0.26527369 | 0.26527369 | 5.238102982 | 3.261549489 | 0.513423973 | FALSE | TRUE | FALSE |
| TNFRSF13C | 0.8650382 | 0.59560289 | 1 | 0.865038183 | 0.865038183 | 0.865038183 | 0.88007693 | 0.601313913 | 1 | 0.880076931 | 0.880076931 | 0.880076931 | 0.982912008 | -0.007485359 | FALSE | TRUE | FALSE |
| SAA1 | 0.8643107 | 0.59570236 | 1 | 0.86431066 | 0.86431066 | 0.86431066 | 0.87940569 | 0.60152389 | 1 | 0.879405694 | 0.879405694 | 0.879405694 | 0.98283496 | -0.007519404 | FALSE | TRUE | FALSE |
| GGTLC1 | 0.8626913 | 0.5958018 | 0.7491 | 0.862691322 | 7.905872329 | 7.905872329 | 10.4745344 | 0.192506073 | 0.00505 | 1.135035043 | 10.47453439 | 10.47453439 | 0.082360828 | -1.084279298 | FALSE | FALSE | FALSE |
| SLC2A13 | 0.8626623 | 0.59590118 | 0.5073 | 0.862662318 | 0.862662318 | 7.301904382 | 0.26165327 | 0.836192373 | 1 | 0.261653267 | 0.261653267 | 5.184546201 | 3.296967497 | 0.518114666 | FALSE | TRUE | FALSE |
| NTPCR | 0.861883 | 0.59600052 | 0.62175 | 1.976272387 | 31.62435946 | 31.62435946 | 0.27159056 | 0.833366152 | 0.0049 | 0.271590561 | 0.271590561 | 0.271590561 | 3.17346463 | 0.501533662 | FALSE | TRUE | FALSE |
| TRAF3IP1 | 0.8609069 | 0.59609982 | 0.5091 | 0.860906887 | 0.860906887 | 3.239771732 | 0.1293427 | 0.869110419 | 0.0156 | 0.129342703 | 0.129342703 | 2.558266983 | 6.656014338 | 0.823214249 | FALSE | TRUE | FALSE |
| XPO7 | 0.8607161 | 0.59619907 | 0.5165 | 0.860716093 | 0.860716093 | 16.84551021 | 0.23028552 | 0.844801995 | 1 | 20.69773855 | 2058.724571 | 2058.724571 | 3.737604067 | 0.572593294 | FALSE | TRUE | FALSE |
| SRP14 | 0.8603233 | 0.59629827 | 1 | 0.860323346 | 0.860323346 | 0.860323346 | 8.04384582 | 0.227233546 | 0.00405 | 8.043845816 | 8.043845816 | 8.043845816 | 0.106954231 | -0.970802029 | FALSE | FALSE | FALSE |
| TCF12 | 0.8600534 | 0.59639741 | 0.96995 | 77.62738155 | 765.6069879 | 7735.326511 | 0.1445667 | 0.865756179 | 0.01645 | 0.144566697 | 0.144566697 | 2.85790155 | 5.949180483 | 0.774457145 | FALSE | TRUE | FALSE |
| ERGIC3 | 0.8600387 | 0.5964965 | 0.75175 | 1.980083394 | 7.919838008 | 7.919838008 | 0.26532665 | 0.835244244 | 1 | 1.061248027 | 1.061248027 | 5.265505235 | 3.241433388 | 0.510737101 | FALSE | TRUE | FALSE |
| ZNF3 | 0.8587824 | 0.59659556 | 0.8732 | 7.897743531 | 74.49764012 | 74.49764012 | 0.11817371 | 0.871672597 | 1 | 93.19758361 | 905.4960961 | 9012.674088 | 7.267118871 | 0.861362264 | FALSE | TRUE | FALSE |
| SDE2 | 0.8586389 | 0.59669456 | 0.5028 | 0.858638935 | 0.858638935 | 1.873800889 | 1.135132 | 0.574023298 | 0.505 | 1.135132001 | 1.135132001 | 2.467265286 | 0.756422103 | -0.121235789 | FALSE | FALSE | FALSE |
| TMEM170B | 0.8570018 | 0.59679353 | 1 | 0.857001827 | 0.857001827 | 0.857001827 | 0.8725878 | 0.602361853 | 1 | 0.872587804 | 0.872587804 | 0.872587804 | 0.982138214 | -0.007827391 | FALSE | TRUE | FALSE |
| DYNLL1 | 0.8569409 | 0.59689244 | 1 | 0.856940944 | 0.856940944 | 0.856940944 | 0.87276488 | 0.602257293 | 1 | 0.87276488 | 0.87276488 | 0.87276488 | 0.981869188 | -0.007946368 | FALSE | TRUE | FALSE |
| CCL19 | 0.8556658 | 0.59699132 | 1 | 0.855665796 | 0.855665796 | 0.855665796 | 0.8714117 | 0.602883782 | 1 | 0.871411704 | 0.871411704 | 0.871411704 | 0.981930575 | -0.007919217 | FALSE | TRUE | FALSE |
| MYCBP | 0.855469 | 0.59709015 | 1 | 8.107722609 | 8.107722609 | 8.107722609 | 0.8991209 | 0.599202668 | 1 | 0.899120901 | 0.899120901 | 0.899120901 | 0.951450415 | -0.02161384 | FALSE | TRUE | FALSE |
| S100A9 | 0.8547614 | 0.59718893 | 1 | 0.854761379 | 0.854761379 | 0.854761379 | 0.87080752 | 0.602987998 | 1 | 0.870807523 | 0.870807523 | 0.870807523 | 0.981573261 | -0.008077281 | FALSE | TRUE | FALSE |

| | | | | | | | | | | | | | | | | | |
|---|---|---|---|---|---|---|---|---|---|---|---|---|---|---|---|---|---|
| SERP2 | 0.854185 | 0.59728766 | 1 | 0.854184982 | 0.854184982 | 0.854184982 | 0.87020655 | 0.603092161 | 1 | 0.870206548 | 0.870206548 | 0.870206548 | 0.981588778 | -0.008070415 | FALSE | TRUE | FALSE |
| SPRR2G | 0.8537942 | 0.59738635 | 1 | 0.853794246 | 0.853794246 | 0.853794246 | 0.86986055 | 0.603300313 | 1 | 0.869860551 | 0.869860551 | 0.869860551 | 0.981530023 | -0.008096412 | FALSE | TRUE | FALSE |
| CHMP4C | 0.8522412 | 0.597485 | 0.872 | 2.130652132 | 13.64005254 | 13.64005254 | 0.26139708 | 0.836275026 | 1 | 1.045392057 | 1.045392057 | 1.045392057 | 3.260331991 | 0.513261825 | FALSE | TRUE | FALSE |
| TAGLN2 | 0.851858 | 0.5975836 | 0.50015 | 0.851857964 | 0.851857964 | 0.851857964 | 0.25864596 | 0.837047908 | 1 | 0.25864596 | 0.25864596 | 0.25864596 | 3.293528981 | 0.517661489 | FALSE | TRUE | FALSE |
| KCNE1 | 0.8516345 | 0.59768215 | 1 | 0.851634471 | 8.080575575 | 8.080575575 | 0.57025037 | 0.724704614 | 1 | 0.570250374 | 0.570250374 | 0.570250374 | 1.493439566 | 0.174187653 | FALSE | TRUE | FALSE |
| FAM72B | 0.85125 | 0.59778066 | 1 | 0.851249965 | 0.851249965 | 0.851249965 | 0.86766034 | 0.603508256 | 1 | 0.867660344 | 0.867660344 | 0.867660344 | 0.981086632 | -0.008292642 | FALSE | TRUE | FALSE |
| SLC48A1 | 0.8510518 | 0.59787911 | 1 | 0.851051784 | 0.851051784 | 0.851051784 | 0.85831455 | 0.606294084 | 1 | 0.858314553 | 0.858314553 | 0.858314553 | 0.991538337 | -0.00369049 | FALSE | TRUE | FALSE |
| OTOF | 0.850435 | 0.59797751 | 0.9949 | 881542.2784 | 4495446416 | 1.2834E+11 | 21.6819107 | 0.125234791 | 0.27875 | 2.527723022 | 1384.460328 | 22145.07089 | 0.039223251 | -1.406456408 | FALSE | FALSE | FALSE |
| PKIB | 0.8502525 | 0.59807587 | 1 | 0.850252493 | 0.850252493 | 0.850252493 | 0.86668304 | 0.603819769 | 1 | 0.866683035 | 0.866683035 | 0.866683035 | 0.98104204 | -0.008312382 | FALSE | TRUE | FALSE |
| ZNF28 | 0.8502349 | 0.59817417 | 0.50175 | 0.850234861 | 7.623579424 | 7.623579424 | 11.9227028 | 0.178593195 | 0.34665 | 11.92270284 | 1051.791842 | 10237.89244 | 0.071312258 | -1.146835812 | FALSE | FALSE | FALSE |
| BASP1 | 0.8498983 | 0.59827242 | 1 | 0.849898287 | 0.849898287 | 0.849898287 | 0.86647438 | 0.603923496 | 1 | 0.866474377 | 0.866474377 | 0.866474377 | 0.980869498 | -0.008388771 | FALSE | TRUE | FALSE |
| PRDM1 | 0.8495004 | 0.59837062 | 0.8776 | 3.398073361 | 31.97584589 | 67.58945014 | 0.26934347 | 0.833996886 | 1 | 0.269343474 | 0.269343474 | 5.300853103 | 3.153966963 | 0.49885714 | FALSE | TRUE | FALSE |
| TOMM22 | 0.848838 | 0.59846878 | 1 | 0.848837962 | 0.848837962 | 0.848837962 | 0.86549479 | 0.604234329 | 1 | 0.865494789 | 0.865494789 | 0.865494789 | 0.980754561 | -0.008439664 | FALSE | TRUE | FALSE |
| MORC3 | 0.8485916 | 0.59856689 | 0.5101 | 0.848591577 | 0.848591577 | 16.67225895 | 0.25770951 | 0.837276743 | 1 | 0.257709509 | 0.257709509 | 5.073973057 | 3.292822136 | 0.517568273 | FALSE | TRUE | FALSE |
| IL15 | 0.8483923 | 0.59866494 | 1 | 0.848392273 | 0.848392273 | 0.848392273 | 0.86510462 | 0.604337831 | 1 | 0.865104619 | 0.865104619 | 0.865104619 | 0.980681705 | -0.008471926 | FALSE | TRUE | FALSE |
| SPRR1B | 0.8474919 | 0.59876296 | 1 | 0.847491927 | 0.847491927 | 0.847491927 | 0.83730439 | 0.610850872 | 0.4965 | 0.837304392 | 7.923745725 | 7.923745725 | 1.012167063 | 0.005252201 | FALSE | TRUE | FALSE |
| AGBL2 | 0.8460092 | 0.59886094 | 0.89505 | 30.26694151 | 2606.8025 | 21991.75197 | 1.49420907 | 0.536638708 | 0.8119 | 13.54177429 | 125.8049217 | 1197.344354 | 0.566191991 | -0.247036278 | FALSE | FALSE | FALSE |
| DUS2 | 0.8455912 | 0.59895887 | 0.50625 | 0.845591222 | 0.845591222 | 3.127836416 | 0.12588716 | 0.869921962 | 0.01285 | 0.125887158 | 0.125887158 | 2.494591815 | 6.717057065 | 0.827179038 | FALSE | TRUE | FALSE |
| COX8A | 0.845566 | 0.59905675 | 1 | 0.845565958 | 0.845565958 | 0.845565958 | 0.86248222 | 0.604751305 | 1 | 0.86248222 | 0.86248222 | 0.86248222 | 0.980386538 | -0.008602661 | FALSE | TRUE | FALSE |
| KLHL11 | 0.8445039 | 0.59915459 | 0.51155 | 0.844503886 | 0.844503886 | 7.137732903 | 0.2565726 | 0.837537507 | 0.0215 | 0.256572601 | 0.256572601 | 5.056278378 | 3.291481169 | 0.517391375 | FALSE | TRUE | FALSE |
| IQCB1 | 0.8444318 | 0.59925238 | 0.6917 | 1.921249523 | 71.69142522 | 71.69142522 | 0.33137167 | 0.814908737 | 0.87625 | 3.038309733 | 28.57370388 | 28.57370388 | 2.548292107 | 0.406249209 | FALSE | TRUE | FALSE |
| RPS12 | 0.8432954 | 0.59935012 | 1 | 0.843295394 | 0.843295394 | 0.843295394 | 0.86052182 | 0.605163955 | 1 | 0.860521823 | 0.860521823 | 0.860521823 | 0.979981416 | -0.00878216 | FALSE | TRUE | FALSE |
| SETDB1 | 0.8430977 | 0.59944782 | 0.51715 | 8.249533211 | 82.73974651 | 850.1075906 | 1.6217839 | 0.523352752 | 0.26895 | 0.169117289 | 1.621783904 | 5.696891391 | 0.519858233 | -0.284115074 | FALSE | FALSE | FALSE |
| ENTPD3 | 0.8430161 | 0.59954547 | 0.2593 | 0.097099244 | 0.843016061 | 0.843016061 | 0.16695749 | 0.860417304 | 0.01145 | 0.166957494 | 0.166957494 | 0.386096685 | 5.049285542 | 0.703229931 | FALSE | TRUE | FALSE |
| NXT1 | 0.8429777 | 0.59964306 | 1 | 0.842977735 | 0.842977735 | 0.842977735 | 0.8602212 | 0.605369942 | 1 | 0.860221202 | 0.860221202 | 0.860221202 | 0.979954612 | -0.008794039 | FALSE | TRUE | FALSE |
| INS | 0.8420805 | 0.59974061 | 1 | 0.842080482 | 0.842080482 | 0.842080482 | 0.85941819 | 0.605575711 | 1 | 0.859418193 | 0.859418193 | 0.859418193 | 0.979826223 | -0.008850942 | FALSE | TRUE | FALSE |
| MZT1 | 0.8418936 | 0.59983811 | 1 | 0.841893566 | 0.841893566 | 0.841893566 | 0.83899846 | 0.610249478 | 1 | 3.35589988 | 13.42657965 | 13.42657965 | 1.003450674 | 0.001496029 | FALSE | TRUE | FALSE |
| FUT4 | 0.8418742 | 0.59993556 | 0.5102 | 0.841874247 | 0.841874247 | 3.155291211 | 1.08031313 | 0.580312846 | 0.7493 | 1.08031313 | 10.06810503 | 21.33667198 | 0.779287248 | -0.10830243 | FALSE | FALSE | FALSE |
| TFF3 | 0.8415988 | 0.60003296 | 1 | 0.841598844 | 0.841598844 | 0.841598844 | 0.85884361 | 0.60598656 | 1 | 0.858843608 | 0.858843608 | 0.858843608 | 0.979920949 | -0.008808958 | FALSE | TRUE | FALSE |
| GOLGA5 | 0.8412144 | 0.60013031 | 0.0206 | 0.841214438 | 0.841214438 | 16.60475901 | 0.25529446 | 0.8378299 | 1 | 0.255294457 | 0.255294457 | 5.047177964 | 3.295075217 | 0.517865333 | FALSE | TRUE | FALSE |
| S100A10 | 0.841204 | 0.60022761 | 1 | 0.841203992 | 0.841203992 | 0.841203992 | 0.85852081 | 0.606191633 | 1 | 0.85852081 | 0.85852081 | 0.85852081 | 0.979829472 | -0.008849502 | FALSE | TRUE | FALSE |
| PDILT | 0.8404961 | 0.60032487 | 0.5135 | 0.840496127 | 0.840496127 | 1.852359014 | 0.126805 | 0.869666701 | 1 | 0.126805003 | 0.126805003 | 2.509371317 | 6.628256812 | 0.821399327 | FALSE | TRUE | FALSE |
| YPEL2 | 0.84041 | 0.60042207 | 1 | 0.840410009 | 0.840410009 | 0.840410009 | 0.85312201 | 0.607009928 | 1 | 0.853122007 | 0.853122007 | 0.853122007 | 0.985099437 | -0.006519929 | FALSE | TRUE | FALSE |
| CD300LF | 0.8401352 | 0.60051922 | 0.50535 | 0.840135153 | 0.840135153 | 0.840135153 | 0.12695078 | 0.869624063 | 0.007 | 0.126950778 | 0.126950778 | 0.126950778 | 6.617802309 | 0.820713789 | FALSE | TRUE | FALSE |
| KCNJ2 | 0.8393823 | 0.60061633 | 0.5071 | 0.839382325 | 0.839382325 | 4.157675173 | 0.25548871 | 0.837765009 | 1 | 0.255488711 | 0.255488711 | 5.066037722 | 3.285398876 | 0.516588104 | FALSE | TRUE | FALSE |
| TMEM127 | 0.8392914 | 0.60071339 | 0.497 | 0.209895117 | 0.839291405 | 0.839291405 | 0.2553049 | 0.837813682 | 1 | 0.255304898 | 0.255304898 | 0.255304898 | 3.287408155 | 0.516853628 | FALSE | TRUE | FALSE |
| RTKN2 | 0.8386783 | 0.6008104 | 0.9326 | 30.02401748 | 2593.045387 | 21905.27313 | 2.90064173 | 0.419815862 | 0.88995 | 26.85742242 | 2478.565414 | 24675.24182 | 0.289135425 | -0.538898695 | FALSE | FALSE | FALSE |
| FBXO28 | 0.8383068 | 0.60090737 | 0.50675 | 0.838306787 | 0.838306787 | 0.838306787 | 0.0955762 | 0.876977916 | 1 | 0.095576196 | 0.095576196 | 0.095576196 | 8.771083436 | 0.943053242 | FALSE | TRUE | FALSE |
| QPCTL | 0.8380766 | 0.60100429 | 0.5039 | 0.838076579 | 0.838076579 | 1.840974163 | 0.12608651 | 0.869879485 | 1 | 0.126086512 | 0.126086512 | 0.126086512 | 6.646837695 | 0.822615074 | FALSE | TRUE | FALSE |
| SLCO3A1 | 0.8375091 | 0.60110116 | 0.94145 | 18.6483616 | 731.3405889 | 731.3405889 | 0.27962171 | 0.831182005 | 1 | 0.279621707 | 0.279621707 | 5.506241117 | 2.995150515 | 0.476418652 | FALSE | TRUE | FALSE |
| FBXO16 | 0.8374058 | 0.60119798 | 0.75135 | 0.837405779 | 7.700399712 | 7.700399712 | 1.14151868 | 0.573313453 | 0.5009 | 1.14151868 | 1.14151868 | 1.14151868 | 0.733589204 | -0.134547069 | FALSE | FALSE | FALSE |
| FAM98A | 0.8360549 | 0.60129476 | 0.75435 | 1.94650127 | 7.784677018 | 16.55019124 | 0.26223009 | 0.83604339 | 1 | 0.262230085 | 0.262230085 | 5.197378802 | 3.188249411 | 0.503552288 | FALSE | TRUE | FALSE |
| PFDN4 | 0.8359022 | 0.6013915 | 1 | 0.835902227 | 0.835902227 | 0.835902227 | 0.85374113 | 0.60670344 | 1 | 0.853741135 | 0.853741135 | 0.853741135 | 0.979105015 | -0.009170725 | FALSE | TRUE | FALSE |
| SNRPD3 | 0.8351811 | 0.60148819 | 1 | 0.835181125 | 0.835181125 | 0.835181125 | 0.8532535 | 0.606907823 | 1 | 0.853253505 | 0.853253505 | 0.853253505 | 0.978819449 | -0.00929741 | FALSE | TRUE | FALSE |
| SPCS3 | 0.834988 | 0.60158483 | 1 | 0.83498797 | 0.83498797 | 0.83498797 | 0.85306868 | 0.607111975 | 1 | 0.853068677 | 0.853068677 | 0.853068677 | 0.978805098 | -0.009303777 | FALSE | TRUE | FALSE |
| ENY2 | 0.8329028 | 0.60168145 | 1 | 0.832902795 | 0.832902795 | 0.832902795 | 0.85112745 | 0.607315926 | 1 | 0.851127455 | 0.851127455 | 0.851127455 | 0.978587625 | -0.009400281 | FALSE | TRUE | FALSE |
| MRPS5 | 0.8324433 | 0.60177802 | 0.50745 | 0.832443267 | 0.832443267 | 4.124808457 | 0.2529027 | 0.83866877 | 0.01305 | 0.252902696 | 0.252902696 | 5.017596327 | 3.29155553 | 0.517401186 | FALSE | TRUE | FALSE |
| MOB4 | 0.8318972 | 0.60187454 | 1 | 0.831897187 | 0.831897187 | 0.831897187 | 0.84667558 | 0.608433944 | 1 | 0.846675582 | 0.846675582 | 0.846675582 | 0.982545387 | -0.007647379 | FALSE | TRUE | FALSE |
| TOMM7 | 0.8318755 | 0.60197102 | 1 | 0.831875506 | 0.831875506 | 0.831875506 | 0.85007846 | 0.607519687 | 1 | 0.850078465 | 0.850078465 | 0.850078465 | 0.97858673 | -0.009400678 | FALSE | TRUE | FALSE |
| COX16 | 0.8315716 | 0.60206744 | 1 | 0.831571615 | 0.831571615 | 0.831571615 | 0.84998026 | 0.607621481 | 1 | 0.849980262 | 0.849980262 | 0.849980262 | 0.978342266 | -0.009509184 | FALSE | TRUE | FALSE |
| NPAP1 | 0.8312853 | 0.60216382 | 0.99895 | 7153.763613 | 8205517.151 | 89822324.17 | 0.12125753 | 0.870975523 | 1 | 9700.891536 | 11314598.65 | 124986679.4 | 6.855535439 | 0.83604138 | FALSE | TRUE | FALSE |
| GCNT2 | 0.8307132 | 0.60226015 | 0.6603 | 0.830713248 | 62.7571334 | 565.3453115 | 0.2267913 | 0.84560093 | 0.50605 | 0.226791303 | 1.940907744 | 1.940907744 | 3.662897287 | 0.563824741 | FALSE | TRUE | FALSE |
| TEX12 | 0.8294963 | 0.60235645 | 1 | 0.829496334 | 0.829496334 | 0.829496334 | 0.8480203 | 0.608028155 | 1 | 0.8480203 | 0.8480203 | 0.8480203 | 0.978156224 | -0.009591777 | FALSE | TRUE | FALSE |
| OR56B1 | 0.8284364 | 0.6024527 | 0.9371 | 7.600127761 | 689.81769 | 689.81769 | 1.12986381 | 0.574848551 | 0.87575 | 10.44593233 | 99.05657962 | 99.05657962 | 0.733217943 | -0.134766915 | FALSE | FALSE | FALSE |
| UBE2G1 | 0.8270764 | 0.60254893 | 1 | 0.827076362 | 0.827076362 | 0.827076362 | 0.84274818 | 0.609746906 | 1 | 0.842748177 | 0.842748177 | 0.842748177 | 0.981403916 | -0.008152213 | FALSE | TRUE | FALSE |
| C16orf54 | 0.8268426 | 0.6026451 | 1 | 7.835499023 | 7.835499023 | 7.835499023 | 0.83351508 | 0.611450437 | 1 | 0.833515081 | 0.833515081 | 0.833515081 | 0.991994826 | -0.003490593 | FALSE | TRUE | FALSE |
| TBC1D16 | 0.8263681 | 0.60274123 | 0.5157 | 0.826368116 | 0.826368116 | 16.15896566 | 1.07680212 | 0.580542658 | 0.5091 | 1.07680212 | 1.07680212 | 21.11254184 | 0.767428018 | -0.114962349 | FALSE | FALSE | FALSE |
| ORC3 | 0.8259831 | 0.60283731 | 0.8745 | 7.611052617 | 71.93449044 | 71.93449044 | 4.61278733 | 0.322868373 | 0.25525 | 1.153205631 | 4.612787327 | 17.23837496 | 0.17906377 | -0.746992276 | FALSE | FALSE | FALSE |
| YPEL5 | 0.8255662 | 0.60293335 | 1 | 0.825566196 | 0.825566196 | 0.825566196 | 0.84458999 | 0.608737753 | 1 | 0.844589992 | 0.844589992 | 0.844589992 | 0.977475703 | -0.009894029 | FALSE | TRUE | FALSE |
| C9orf85 | 0.8255363 | 0.60302933 | 0.49825 | 0.09281959 | 0.825536252 | 0.825536252 | 0.12536442 | 0.870049238 | 1 | 0.125364416 | 0.125364416 | 0.125364416 | 6.585092335 | 0.818561869 | FALSE | TRUE | FALSE |
| NUP37 | 0.8255352 | 0.60312526 | 0.75175 | 0.825535204 | 3.30276805 | 3.30276805 | 0.25359033 | 0.838411597 | 1 | 0.253590332 | 0.253590332 | 0.253590332 | 3.255389109 | 0.512602906 | FALSE | TRUE | FALSE |
| RPRML | 0.8254175 | 0.60322114 | 1 | 0.825417539 | 0.825417539 | 0.825417539 | 0.84435227 | 0.608838918 | 1 | 0.844352274 | 0.844352274 | 0.844352274 | 0.97757484 | -0.009849985 | FALSE | TRUE | FALSE |
| OR5K3 | 0.8252667 | 0.60331697 | 1 | 2.2214E+256 | 1.393E+282 | 3.7192E+293 | 2.439E+264 | 0 | 0.00005 | 8.5107E+211 | 3.2376E+235 | 4.648E+246 | 3.3843E-265 | -264.4705357 | FALSE | FALSE | FALSE |
| MUSTN1 | 0.8250456 | 0.60341276 | 1 | 0.825045559 | 0.825045559 | 0.825045559 | 0.84401217 | 0.608940028 | 1 | 0.844012168 | 0.844012168 | 0.844012168 | 0.977528039 | -0.009870777 | FALSE | TRUE | FALSE |
| SNRPD2 | 0.824674 | 0.60350849 | 1 | 0.824673961 | 0.824673961 | 0.824673961 | 0.84375161 | 0.609041084 | 1 | 0.843751608 | 0.843751608 | 0.843751608 | 0.977389499 | -0.009932331 | FALSE | TRUE | FALSE |
| UBE2E3 | 0.8244393 | 0.60360418 | 1 | 0.824439295 | 0.824439295 | 0.824439295 | 0.84348834 | 0.609142084 | 1 | 0.843488336 | 0.843488336 | 0.843488336 | 0.977416355 | -0.009920398 | FALSE | TRUE | FALSE |
| NRN1 | 0.8239654 | 0.60369982 | 1 | 0.823965351 | 0.823965351 | 0.823965351 | 7.73575457 | 0.232858152 | 0.0052 | 7.735754575 | 7.735754575 | 7.735754575 | 0.106513895 | -0.972593734 | FALSE | FALSE | FALSE |
| C5orf24 | 0.8239418 | 0.60379541 | 1 | 0.823941757 | 0.823941757 | 0.823941757 | 0.8431185 | 0.60934392 | 1 | 0.843118497 | 0.843118497 | 0.843118497 | 0.977254989 | -0.009992104 | FALSE | TRUE | FALSE |
| SPRR2D | 0.8239226 | 0.60389095 | 1 | 0.823922599 | 0.823922599 | 0.823922599 | 7.23409647 | 0.239474462 | 0.8729 | 7.234096468 | 696.1080752 | 696.1080752 | 0.113894334 | -0.943497881 | FALSE | FALSE | FALSE |
| PCP4L1 | 0.8238541 | 0.60398644 | 1 | 0.823854141 | 0.823854141 | 0.823854141 | 0.84285971 | 0.609545528 | 1 | 0.842859706 | 0.842859706 | 0.842859706 | 0.977451092 | -0.009904964 | FALSE | TRUE | FALSE |
| TMEM100 | 0.823841 | 0.60408188 | 1 | 0.823840957 | 0.823840957 | 0.823840957 | 0.84295001 | 0.609444752 | 1 | 0.842950011 | 0.842950011 | 0.842950011 | 0.977330739 | -0.009958442 | FALSE | TRUE | FALSE |
| MT1E | 0.8237527 | 0.60417727 | 1 | 7.7914493 | 7.7914493 | 7.7914493 | 0.86987912 | 0.603196268 | 1 | 0.869879119 | 0.869879119 | 0.869879119 | 0.94697371 | -0.023662078 | FALSE | TRUE | FALSE |
| RPLP1 | 0.8235796 | 0.6042726 | 1 | 0.823579571 | 0.823579571 | 0.823579571 | 0.8427272 | 0.609847508 | 1 | 0.842727195 | 0.842727195 | 0.842727195 | 0.977278977 | -0.009981443 | FALSE | TRUE | FALSE |

| | | | | | | | | | | | | | | | | | |
|---|---|---|---|---|---|---|---|---|---|---|---|---|---|---|---|---|---|
| B4GALNT1 | 0.8227022 | 0.6043679 | 0.50895 | 0.82270217 | 0.82270217 | 1.774985463 | 0.11569473 | 0.872239563 | 0.7634 | 1.060878187 | 9.984168048 | 20.94983271 | 7.110973696 | 0.851929072 | FALSE | TRUE | FALSE |
| STOML3 | 0.8226271 | 0.60446315 | 0.19035 | 0.089628721 | 3.287288314 | 3.287288314 | 0.07892392 | 0.880672775 | 0.89195 | 0.710594863 | 26.25060391 | 104.9665159 | 10.42303984 | 1.017994398 | FALSE | TRUE | FALSE |
| PIH1D2 | 0.821905 | 0.60455836 | 0.50895 | 0.821904998 | 0.821904998 | 0.821904998 | 0.12465005 | 0.870186858 | 1 | 0.124650047 | 0.124650047 | 0.124650047 | 6.593699863 | 0.819129175 | FALSE | TRUE | FALSE |
| STX19 | 0.8218599 | 0.60465351 | 0.50335 | 0.82185992 | 0.82185992 | 0.82185992 | 0.1247514 | 0.870176281 | 0.0068 | 0.124751404 | 0.124751404 | 0.124751404 | 6.587981308 | 0.818752358 | FALSE | TRUE | FALSE |
| ARHGAP11A | 0.8216839 | 0.60474862 | 0.51045 | 0.821683888 | 7.550095608 | 16.14744192 | 0.072227 | 0.881980952 | 0.8782 | 0.63692816 | 5.764862909 | 12.5429353 | 11.3764087 | 1.056005186 | FALSE | TRUE | FALSE |
| CUL3 | 0.8213871 | 0.60484368 | 0.0202 | 0.821387087 | 0.821387087 | 6.940626289 | 0.25053718 | 0.839260428 | 1 | 0.250537182 | 0.250537182 | 4.943080502 | 3.278503739 | 0.515675683 | FALSE | TRUE | FALSE |
| PDZRN4 | 0.8210121 | 0.60493869 | 0.26095 | 0.08833803 | 0.821012076 | 2.978920353 | 0.14816861 | 0.865009233 | 0.87895 | 1.383344798 | 13.24696169 | 27.20735786 | 5.541066018 | 0.743593325 | FALSE | TRUE | FALSE |
| GRWD1 | 0.8203657 | 0.60503366 | 0.5091 | 0.82036569 | 3.280458574 | 7.040896198 | 0.46844873 | 0.767630556 | 0.8758 | 1.873750484 | 17.29296157 | 17.33373219 | 1.751239018 | 0.243345425 | FALSE | TRUE | FALSE |
| SLC36A2 | 0.8203309 | 0.60512858 | 0.5077 | 0.820330948 | 0.820330948 | 1.814063885 | 9.73655792 | 0.202007256 | 0.0078 | 1.05369912 | 9.736557918 | 9.736557918 | 0.084252665 | -1.074416355 | FALSE | FALSE | FALSE |
| ZNF432 | 0.8197579 | 0.60522346 | 0.69375 | 0.819757944 | 68.56806347 | 68.56806347 | 119.83395 | 0.024406587 | 0.0052 | 1.400741287 | 119.8339502 | 119.8339502 | 0.006840782 | -2.164894242 | FALSE | FALSE | FALSE |
| C1QB | 0.8194168 | 0.60531829 | 0.94235 | 3.277467159 | 52.47239648 | 52.47239648 | 1.01050376 | 0.587113661 | 0.751 | 1.01050376 | 4.041946558 | 4.041946558 | 0.810899301 | -0.091033074 | FALSE | FALSE | FALSE |
| S100A6 | 0.8193272 | 0.60541307 | 1 | 0.819327159 | 0.819327159 | 0.819327159 | 0.83878848 | 0.610349845 | 1 | 0.838788477 | 0.838788477 | 0.838788477 | 0.9767983 | -0.010195105 | FALSE | TRUE | FALSE |
| LCORL | 0.8184467 | 0.60550781 | 0.93585 | 7.81282199 | 767.6891229 | 767.6891229 | 0.08928055 | 0.878295 | 1 | 777.9772424 | 84138.23011 | 84138.23011 | 9.167133387 | 0.962233551 | FALSE | TRUE | FALSE |
| GTF2F1 | 0.8184122 | 0.6056025 | 0.7565 | 0.818412206 | 7.56923061 | 16.12522237 | 0.12351638 | 0.870471816 | 0.5001 | 1.120675079 | 1.120675079 | 4.185895841 | 6.625940483 | 0.821247531 | FALSE | TRUE | FALSE |
| MMP17 | 0.8183793 | 0.60569714 | 0.8789 | 7.606722556 | 72.51970092 | 72.51970092 | 1.15773795 | 0.571648687 | 0.50705 | 1.157737945 | 1.157737945 | 2.50270687 | 0.706877846 | -0.150655629 | FALSE | FALSE | FALSE |
| CCDC178 | 0.818109 | 0.60579174 | 0.1998 | 0.818108976 | 7.198964017 | 64.90072198 | 1.56308761 | 0.530081651 | 0.6563 | 1.563087614 | 127.9708836 | 1200.035479 | 0.523392911 | -0.281172164 | FALSE | FALSE | FALSE |
| GREM1 | 0.8167709 | 0.6058863 | 1 | 0.816770893 | 0.816770893 | 0.816770893 | 0.83664285 | 0.610950922 | 1 | 0.836642849 | 0.836642849 | 0.836642849 | 0.976247981 | -0.010439851 | FALSE | TRUE | FALSE |
| SREBF2 | 0.8166573 | 0.60598081 | 0.03405 | 0.816657314 | 0.816657314 | 15.9295881 | 0.48132161 | 0.762591327 | 0.5149 | 0.481321615 | 0.481321615 | 9.416536309 | 1.696697778 | 0.229604491 | FALSE | TRUE | FALSE |
| UBE2K | 0.8160602 | 0.60607528 | 1 | 0.816060188 | 0.816060188 | 0.816060188 | 0.83347274 | 0.611550186 | 1 | 0.833472741 | 0.833472741 | 0.833472741 | 0.979108432 | -0.009169209 | FALSE | TRUE | FALSE |
| ZNF98 | 0.815779 | 0.6061697 | 0.99625 | 683.46618 | 68609.0434 | 711777.0234 | 1.22531812 | 0.5646277 | 0.9361 | 11.37936758 | 1058.341947 | 1058.341947 | 0.66576912 | -0.176676352 | FALSE | FALSE | FALSE |
| C11orf87 | 0.815042 | 0.60626408 | 1 | 0.815041989 | 0.815041989 | 0.815041989 | 0.83512497 | 0.611150882 | 1 | 0.835124966 | 0.835124966 | 0.835124966 | 0.975952129 | -0.010571484 | FALSE | TRUE | FALSE |
| NHLRC1 | 0.8147201 | 0.60635841 | 0.0147 | 0.814720051 | 0.814720051 | 1.739941353 | 0.11709105 | 0.871910264 | 1 | 1.08786638 | 1.08786638 | 2.319612339 | 6.958004756 | 0.842484721 | FALSE | TRUE | FALSE |
| ASF1A | 0.8146556 | 0.60645269 | 1 | 0.814655609 | 0.814655609 | 0.814655609 | 0.83470274 | 0.611250786 | 1 | 0.834702738 | 0.834702738 | 0.834702738 | 0.975982912 | -0.010557786 | FALSE | TRUE | FALSE |
| E2F2 | 0.8136625 | 0.60654694 | 0.01465 | 0.813662472 | 0.813662472 | 4.028002154 | 0.2482522 | 0.839958419 | 1 | 0.248252196 | 0.248252196 | 4.920702665 | 3.277564045 | 0.515551187 | FALSE | TRUE | FALSE |
| MAGOH | 0.8126212 | 0.60664115 | 1 | 0.812621165 | 0.812621165 | 0.812621165 | 0.8329183 | 0.611649885 | 1 | 0.832918302 | 0.832918302 | 0.832918302 | 0.975631299 | -0.010714276 | FALSE | TRUE | FALSE |
| HLA-DRA | 0.81227 | 0.60673531 | 1 | 0.812270006 | 0.812270006 | 0.812270006 | 7.65423706 | 0.233189169 | 0.5084 | 7.65423706 | 7.65423706 | 7.65423706 | 0.106120309 | -0.974201492 | FALSE | FALSE | FALSE |
| S100A7 | 0.8119726 | 0.60682943 | 1 | 0.811972611 | 0.811972611 | 0.811972611 | 0.83216067 | 0.611849121 | 0.00155 | 0.832160672 | 0.832160672 | 0.832160672 | 0.975740188 | -0.010665807 | FALSE | TRUE | FALSE |
| NUDT4 | 0.8118787 | 0.6069235 | 1 | 0.8118787 | 0.8118787 | 0.8118787 | 0.83214437 | 0.611948659 | 1 | 0.832144372 | 0.832144372 | 0.832144372 | 0.975646448 | -0.010707532 | FALSE | TRUE | FALSE |
| ZNF556 | 0.8118092 | 0.60701752 | 0.5008 | 0.811809223 | 0.811809223 | 1.787066281 | 9.70137975 | 0.202345976 | 0.25985 | 1.045277418 | 9.701379752 | 9.701379752 | 0.08367977 | -1.077379524 | FALSE | FALSE | FALSE |
| TAF10 | 0.8117515 | 0.60711149 | 1 | 0.811751473 | 0.811751473 | 0.811751473 | 3.18983859 | 0.400894369 | 0.501 | 7.559745487 | 30.23858848 | 30.23858848 | 0.254480423 | -0.594345622 | FALSE | FALSE | FALSE |
| CFC1 | 0.8116539 | 0.60720541 | 0.81335 | 78.28514188 | 798.1029153 | 8335.013142 | 8.49492684 | 0.218204998 | 0.69005 | 8.494926839 | 864.0213771 | 864.0213771 | 0.095545719 | -1.019788765 | FALSE | FALSE | FALSE |
| CRADD | 0.8113236 | 0.60729928 | 0.5058 | 0.811323593 | 0.811323593 | 0.811323593 | 0.23929914 | 0.842217456 | 1 | 0.956992537 | 15.31683537 | 15.31683537 | 3.390415882 | 0.530252974 | FALSE | TRUE | FALSE |
| BTF3 | 0.8100382 | 0.60739312 | 1 | 0.810038203 | 0.810038203 | 0.810038203 | 0.83056636 | 0.612445547 | 1 | 0.830566363 | 0.830566363 | 0.830566363 | 0.975284142 | -0.010868837 | FALSE | TRUE | FALSE |
| UBC | 0.8097209 | 0.60748692 | 0.01795 | 0.809720876 | 0.809720876 | 2.992478508 | 0.11387988 | 0.872597968 | 0.7504 | 1.046551356 | 9.871149186 | 20.66125794 | 7.110306898 | 0.851888346 | FALSE | TRUE | FALSE |
| SLC6A9 | 0.8094788 | 0.60758067 | 0.26245 | 0.347869706 | 3.235212987 | 6.836319657 | 0.15743007 | 0.862696948 | 0.50765 | 0.157430072 | 0.157430072 | 0.584526191 | 5.141830656 | 0.711117769 | FALSE | TRUE | FALSE |
| SLC16A10 | 0.8088316 | 0.60767437 | 0.7593 | 0.808831616 | 7.482522068 | 7.482522068 | 0.49819408 | 0.755829606 | 0.50895 | 0.498194084 | 0.498194084 | 2.470490545 | 1.62352714 | 0.210459553 | FALSE | TRUE | FALSE |
| SMOC2 | 0.8088093 | 0.60776803 | 0.7553 | 0.80880932 | 3.235684369 | 6.854115065 | 0.24936646 | 0.839594308 | 1 | 0.249366458 | 0.249366458 | 4.942125201 | 3.243456748 | 0.511008111 | FALSE | TRUE | FALSE |
| SMCO4 | 0.8084351 | 0.60786164 | 1 | 0.808435056 | 0.808435056 | 0.808435056 | 0.82880022 | 0.612842162 | 1 | 0.828800218 | 0.828800218 | 0.828800218 | 0.975428142 | -0.010804719 | FALSE | TRUE | FALSE |
| MRGBP | 0.8081476 | 0.60795521 | 1 | 0.808147561 | 0.808147561 | 0.808147561 | 0.82889952 | 0.612743091 | 1 | 0.828899518 | 0.828899518 | 0.828899518 | 0.974964448 | -0.011011221 | FALSE | TRUE | FALSE |
| TRMT2A | 0.8070227 | 0.60804874 | 0.51445 | 0.807022657 | 12.91331231 | 12.91331231 | 0.03641976 | 0.889811176 | 1 | 0.3257346 | 0.3257346 | 6.42877099 | 22.15892505 | 1.345548689 | FALSE | TRUE | FALSE |
| AP3S2 | 0.8067261 | 0.60814222 | 1 | 7.643500628 | 7.643500628 | 7.643500628 | 0.85246879 | 0.607213971 | 1 | 0.852468792 | 0.852468792 | 0.852468792 | 0.946340963 | -0.023952361 | FALSE | TRUE | FALSE |
| NDUFAB1 | 0.8062745 | 0.60823566 | 0.00455 | 0.806274503 | 0.806274503 | 0.806274503 | 0.82712302 | 0.613040157 | 1 | 0.827123018 | 0.827123018 | 0.827123018 | 0.974793937 | -0.011087181 | FALSE | TRUE | FALSE |
| ZNF655 | 0.8060579 | 0.60832906 | 0.50675 | 0.806057915 | 0.806057915 | 1.785029276 | 1.03769736 | 0.584077305 | 0.7518 | 1.037697356 | 9.594705881 | 9.594705881 | 0.776775531 | -0.109704463 | FALSE | FALSE | FALSE |
| FAM168A | 0.8057415 | 0.60842241 | 1 | 0.805741518 | 0.805741518 | 0.805741518 | 0.82674305 | 0.613139086 | 1 | 0.826743053 | 0.826743053 | 0.826743053 | 0.974597264 | -0.011174812 | FALSE | TRUE | FALSE |
| CNIH2 | 0.8055138 | 0.60851571 | 1 | 0.805513771 | 0.805513771 | 0.805513771 | 0.82636591 | 0.613237964 | 1 | 0.826365911 | 0.826365911 | 0.826365911 | 0.974766457 | -0.011099424 | FALSE | TRUE | FALSE |
| ENKD1 | 0.8053401 | 0.60860896 | 0.5008 | 0.805340107 | 0.805340107 | 1.748305827 | 0.98140961 | 0.590331282 | 0.502 | 0.981409614 | 0.981409614 | 2.123237475 | 0.820595291 | -0.08587098 | FALSE | TRUE | FALSE |
| NME2 | 0.8048698 | 0.60870217 | 1 | 0.804869803 | 0.804869803 | 0.804869803 | 0.82521605 | 0.6133368 | 1 | 0.82521605 | 0.82521605 | 0.82521605 | 0.975344338 | -0.010842033 | FALSE | TRUE | FALSE |
| PPP1R14A | 0.8043558 | 0.60879534 | 1 | 0.804355788 | 0.804355788 | 0.804355788 | 7.31689168 | 0.238484389 | 0.7513 | 7.31689168 | 70.92744387 | 70.92744387 | 0.109931351 | -0.958878435 | FALSE | FALSE | FALSE |
| ZNF154 | 0.8037628 | 0.60888846 | 0.99985 | 71158.10322 | 87613828.18 | 1333086169 | 0.15943985 | 0.862213575 | 0.7504 | 1.463705968 | 13.79128742 | 13.79128742 | 5.041166083 | 0.702531006 | FALSE | TRUE | FALSE |
| HMGA1 | 0.803448 | 0.60898154 | 1 | 0.803448045 | 0.803448045 | 0.803448045 | 0.81263972 | 0.616667108 | 1 | 0.812639716 | 0.812639716 | 0.812639716 | 0.988689118 | -0.004940246 | FALSE | TRUE | FALSE |
| MSRB1 | 0.8034163 | 0.60907458 | 1 | 3.213617595 | 12.85749279 | 12.85749279 | 0.84313194 | 0.609243031 | 1 | 0.843131944 | 0.843131944 | 0.843131944 | 0.952895112 | -0.020954901 | FALSE | TRUE | FALSE |
| VAMP2 | 0.8034048 | 0.60916756 | 1 | 0.803404827 | 0.803404827 | 0.803404827 | 0.81478685 | 0.615791267 | 1 | 0.814786848 | 0.814786848 | 0.814786848 | 0.986030678 | -0.006109573 | FALSE | TRUE | FALSE |
| DTNA | 0.8033998 | 0.60926049 | 0.26475 | 0.200892926 | 0.803399819 | 3.961291344 | 2.5812062 | 0.441169025 | 0.25825 | 0.645563272 | 2.581206201 | 5.530526618 | 0.311249763 | -0.50689097 | FALSE | FALSE | FALSE |
| FDCSP | 0.8029813 | 0.60935338 | 1 | 0.802981312 | 0.802981312 | 0.802981312 | 0.82399577 | 0.613435597 | 0.00165 | 0.82399577 | 0.82399577 | 0.82399577 | 0.974496887 | -0.011219544 | FALSE | TRUE | FALSE |
| SLC22A8 | 0.8025487 | 0.60944622 | 0.2609 | 0.802548715 | 3.209096571 | 6.881915269 | 0.12100325 | 0.87105912 | 1 | 0.121003246 | 0.121003246 | 2.394107786 | 6.632456062 | 0.821674382 | FALSE | TRUE | FALSE |
| C9orf40 | 0.8023272 | 0.60953902 | 1 | 0.802327179 | 0.802327179 | 0.802327179 | 7.52401547 | 0.23451334 | 0.0048 | 7.524015471 | 7.524015471 | 7.524015471 | 0.106635504 | -0.972098176 | FALSE | FALSE | FALSE |
| PSMB3 | 0.8022244 | 0.60963177 | 1 | 0.802224431 | 0.802224431 | 0.802224431 | 0.82354359 | 0.613633031 | 1 | 0.823543588 | 0.823543588 | 0.823543588 | 0.974112899 | -0.011390706 | FALSE | TRUE | FALSE |
| UBE2R2 | 0.8021598 | 0.60972448 | 1 | 0.802159772 | 0.802159772 | 0.802159772 | 0.82349257 | 0.613830243 | 1 | 0.823492571 | 0.823492571 | 0.823492571 | 0.974094728 | -0.011398807 | FALSE | TRUE | FALSE |
| IFNAR2 | 0.8021229 | 0.60981713 | 0.50515 | 0.802122932 | 0.802122932 | 1.773930402 | 0.12175425 | 0.870870881 | 1 | 0.121754254 | 0.121754254 | 2.415224062 | 6.588048498 | 0.818756788 | FALSE | TRUE | FALSE |
| ECD | 0.8016831 | 0.60990974 | 0.7613 | 0.801683131 | 7.395786995 | 15.85638809 | 9.5867312 | 0.204037488 | 0.50665 | 9.586731195 | 91.20054788 | 91.20054788 | 0.083624242 | -1.077667805 | FALSE | FALSE | FALSE |
| GCHFR | 0.8007312 | 0.61000232 | 1 | 0.800731209 | 3.202834635 | 3.202834635 | 0.82207435 | 0.614322344 | 1 | 3.288215718 | 3.288215718 | 3.288215718 | 0.974037453 | -0.011424343 | FALSE | TRUE | FALSE |
| PPP3R1 | 0.8007069 | 0.61009484 | 1 | 0.800706941 | 0.800706941 | 0.800706941 | 0.82210159 | 0.614224028 | 1 | 0.822101589 | 0.822101589 | 0.822101589 | 0.973975664 | -0.011451894 | FALSE | TRUE | FALSE |
| NMUR2 | 0.7998542 | 0.61018733 | 0.50405 | 0.799854162 | 3.19849017 | 3.19849017 | 0.12119007 | 0.871006884 | 0.00995 | 0.121190073 | 0.121190073 | 0.121190073 | 6.599997348 | 0.819543761 | FALSE | TRUE | FALSE |
| MT1G | 0.799522 | 0.61027977 | 1 | 0.799521957 | 0.799521957 | 0.799521957 | 0.82082216 | 0.61451883 | 0.00125 | 0.820822156 | 0.820822156 | 0.820822156 | 0.974050165 | -0.011418676 | FALSE | TRUE | FALSE |
| RPL32 | 0.7992594 | 0.61037217 | 1 | 0.7992594 | 0.7992594 | 0.7992594 | 0.82079833 | 0.614616996 | 1 | 0.820798335 | 0.820798335 | 0.820798335 | 0.973758555 | -0.011548714 | FALSE | TRUE | FALSE |
| OR7A5 | 0.7991041 | 0.61046452 | 0.00675 | 0.799104107 | 0.799104107 | 0.799104107 | 0.1033876 | 0.875095194 | 1 | 0.103387603 | 0.103387603 | 0.103387603 | 7.72920625 | 0.888134896 | FALSE | TRUE | FALSE |
| FUCA1 | 0.7988956 | 0.61055682 | 0.81805 | 3.19590791 | 12.7881283 | 51.18328928 | 1.134138 | 0.574377353 | 0.50655 | 4.537226959 | 18.15614374 | 72.67167997 | 0.704407763 | -0.152175867 | FALSE | FALSE | FALSE |
| C1orf21 | 0.7987551 | 0.61064908 | 1 | 0.798755063 | 0.798755063 | 0.798755063 | 0.80684643 | 0.618694735 | 1 | 0.806846427 | 0.806846427 | 0.806846427 | 0.989971618 | -0.004377256 | FALSE | TRUE | FALSE |
| KIAA1549 | 0.7985188 | 0.61074129 | 0.76255 | 0.798518778 | 7.555384664 | 144.6347949 | 0.12272176 | 0.870713632 | 0.0507 | 0.122721755 | 2.361449442 | 4.125619723 | 6.506741831 | 0.813363576 | FALSE | TRUE | FALSE |
| SARNP | 0.7977621 | 0.61083346 | 1 | 0.797762077 | 0.797762077 | 0.797762077 | 0.81949755 | 0.614813195 | 1 | 0.819497548 | 0.819497548 | 0.819497548 | 0.973477075 | -0.011674271 | FALSE | TRUE | FALSE |
| TMEM11 | 0.7977593 | 0.61092558 | 1 | 0.7977593 | 0.7977593 | 0.7977593 | 0.8195044 | 0.614715123 | 1 | 0.819504404 | 0.819504404 | 0.819504404 | 0.973465543 | -0.011679416 | FALSE | TRUE | FALSE |
| MTAP | 0.7975752 | 0.61101766 | 0.4996 | 0.086101421 | 0.797575222 | 0.797575222 | 0.11853105 | 0.87157937 | 1 | 0.118531053 | 0.118531053 | 0.118531053 | 6.7288293 | 0.827939511 | FALSE | TRUE | FALSE |
| C16orf74 | 0.7975492 | 0.61110969 | 1 | 0.797549209 | 0.797549209 | 0.797549209 | 0.81061161 | 0.617248737 | 1 | 0.810611607 | 0.810611607 | 0.810611607 | 0.98388575 | -0.007055329 | FALSE | TRUE | FALSE |

| | | | | | | | | | | | | | | | | |
|---|---|---|---|---|---|---|---|---|---|---|---|---|---|---|---|---|
| POLR2J2 | 0.7974399 | 0.61120167 | 1 | 0.797439919 | 0.797439919 | 0.797439919 | 0.80731635 | 0.618406451 | 1 | 0.807316354 | 0.807316354 | 0.807316354 | 0.987766339 | -0.005345778 | FALSE | TRUE | FALSE |
| RHOBTB3 | 0.7959035 | 0.61129362 | 0.75325 | 0.795903476 | 7.381080325 | 15.72055703 | 0.12402488 | 0.870355865 | 1 | 0.124024881 | 0.124024881 | 2.453376845 | 6.417288746 | 0.807351581 | FALSE | TRUE | FALSE |
| RARG | 0.7958073 | 0.61138553 | 0.50735 | 0.795807338 | 3.181816197 | 6.779417816 | 0.11944624 | 0.871361344 | 1 | 0.119446243 | 0.119446243 | 2.36605225 | 6.66247277 | 0.823635447 | FALSE | TRUE | FALSE |
| CBX1 | 0.794912 | 0.6114774 | 1 | 0.794912025 | 0.794912025 | 0.794912025 | 0.81460976 | 0.615888793 | 1 | 0.814609759 | 0.814609759 | 0.814609759 | 0.975819423 | -0.010630542 | FALSE | TRUE | FALSE |
| UPP2 | 0.7948633 | 0.61156922 | 0.5051 | 0.794863285 | 3.178522696 | 3.178522696 | 0.12070106 | 0.871090445 | 0.00835 | 0.120701061 | 0.120701061 | 0.120701061 | 6.585387745 | 0.818581351 | FALSE | TRUE | FALSE |
| RPL26 | 0.7946601 | 0.61166099 | 1 | 0.794660107 | 0.794660107 | 0.794660107 | 0.81667359 | 0.61520503 | 1 | 0.816673587 | 0.816673587 | 0.816673587 | 0.973044946 | -0.011867099 | FALSE | TRUE | FALSE |
| YWHAZ | 0.7944154 | 0.61175272 | 1 | 0.794415417 | 0.794415417 | 0.794415417 | 0.81635676 | 0.615302865 | 1 | 0.816356762 | 0.816356762 | 0.816356762 | 0.973122847 | -0.011832331 | FALSE | TRUE | FALSE |
| KLF16 | 0.7943762 | 0.6118444 | 1 | 0.794376244 | 0.794376244 | 0.794376244 | 0.8161835 | 0.615498373 | 1 | 0.816183499 | 0.816183499 | 0.816183499 | 0.973281431 | -0.011761562 | FALSE | TRUE | FALSE |
| CERK | 0.7933698 | 0.61193605 | 0.75065 | 0.793369839 | 7.342958965 | 15.68397029 | 0.12012257 | 0.871205184 | 0.51245 | 1.090587772 | 1.090587772 | 2.377981841 | 6.604669241 | 0.819851073 | FALSE | TRUE | FALSE |
| IFIT5 | 0.7933531 | 0.61202765 | 0.5092 | 0.793353135 | 0.793353135 | 1.716171177 | 0.11885163 | 0.871506772 | 1 | 0.11885163 | 0.11885163 | 2.359364339 | 6.675155692 | 0.8244614 | FALSE | TRUE | FALSE |
| RPS23 | 0.7929042 | 0.6121192 | 1 | 0.792904197 | 7.512718732 | 7.512718732 | 0.81503304 | 0.615693687 | 1 | 7.72473877 | 7.72473877 | 7.72473877 | 0.972849145 | -0.011954499 | FALSE | TRUE | FALSE |
| NUDT21 | 0.7922857 | 0.61221072 | 1 | 0.792285665 | 0.792285665 | 0.792285665 | 0.81455084 | 0.615986264 | 1 | 0.814550839 | 0.814550839 | 0.814550839 | 0.972665703 | -0.012036398 | FALSE | TRUE | FALSE |
| ZIK1 | 0.7920675 | 0.61230219 | 0.7454 | 0.792067521 | 7.313644238 | 7.313644238 | 9.80151128 | 0.2013305 | 0.00855 | 1.054998536 | 9.801511284 | 9.801511284 | 0.080810754 | -1.092530839 | FALSE | FALSE | FALSE |
| GCM2 | 0.7918887 | 0.61239361 | 0.8781 | 3.167754464 | 12.67498317 | 15.66783845 | 240.319556 | 0.010214573 | 0.0369 | 14.99889138 | 60.03021481 | 240.3195562 | 0.003295149 | -2.482124948 | FALSE | FALSE | FALSE |
| NUS1 | 0.7917723 | 0.61248499 | 1 | 0.791772336 | 0.791772336 | 0.791772336 | 0.80728028 | 0.618502597 | 1 | 0.807280279 | 0.807280279 | 0.807280279 | 0.98078989 | -0.008424019 | FALSE | TRUE | FALSE |
| RPS21 | 0.7917611 | 0.61257632 | 1 | 0.79176111 | 0.79176111 | 0.79176111 | 0.81388045 | 0.616181054 | 1 | 0.813880454 | 0.813880454 | 0.813880454 | 0.972822368 | -0.011966452 | FALSE | TRUE | FALSE |
| ZNF14 | 0.7910004 | 0.61266761 | 0.6943 | 0.791000387 | 67.2570573 | 67.2570573 | 1.36207705 | 0.549658904 | 0.68895 | 1.36207705 | 118.2459748 | 118.2459748 | 0.580731014 | -0.236024979 | FALSE | FALSE | FALSE |
| EPHA4 | 0.7894728 | 0.61275888 | 0.51635 | 0.789472838 | 0.789472838 | 15.46123064 | 0.11514103 | 0.872331901 | 1 | 0.460381165 | 0.460381165 | 9.038792814 | 6.85657282 | 0.836107093 | FALSE | TRUE | FALSE |
| C6orf62 | 0.7893818 | 0.61285009 | 1 | 0.789381822 | 0.789381822 | 0.789381822 | 0.81188681 | 0.616764175 | 1 | 0.811886811 | 0.811886811 | 0.811886811 | 0.972280632 | -0.012208365 | FALSE | TRUE | FALSE |
| PLA2R1 | 0.7893342 | 0.61294126 | 0.51565 | 0.789334237 | 59.60957702 | 536.4623418 | 0.03241099 | 0.890705928 | 0.81735 | 0.271180195 | 2.325961741 | 20.43937542 | 24.35390567 | 1.386568619 | FALSE | TRUE | FALSE |
| FXYD7 | 0.7886337 | 0.61303239 | 1 | 0.78863371 | 0.78863371 | 0.78863371 | 0.81101628 | 0.617055065 | 1 | 0.811016276 | 0.811016276 | 0.811016276 | 0.972401828 | -0.012154233 | FALSE | TRUE | FALSE |
| PAM16 | 0.788057 | 0.61312348 | 1 | 0.788057048 | 7.475445227 | 7.475445227 | 7.61619303 | 0.23385072 | 0.00405 | 7.616193028 | 7.616193028 | 7.616193028 | 0.103471255 | -0.985180285 | FALSE | FALSE | FALSE |
| MREG | 0.7878455 | 0.61321453 | 0.7518 | 0.787845464 | 7.429269925 | 7.429269925 | 0.94757427 | 0.593180947 | 0.5025 | 0.947574269 | 0.947574269 | 0.947574269 | 0.831434 | -0.08017222 | FALSE | TRUE | FALSE |
| TAC1 | 0.7877278 | 0.61330553 | 1 | 0.787727803 | 0.787727803 | 0.787727803 | 0.81023258 | 0.617538845 | 1 | 0.810232581 | 0.810232581 | 0.810232581 | 0.972224299 | -0.012233529 | FALSE | TRUE | FALSE |
| SCGB2A1 | 0.7875296 | 0.61339648 | 1 | 0.787529553 | 0.787529553 | 0.787529553 | 0.809978 | 0.617635444 | 1 | 0.809977997 | 0.809977997 | 0.809977997 | 0.972285118 | -0.012206362 | FALSE | TRUE | FALSE |
| AGBL1 | 0.7875087 | 0.61348738 | 0.3201 | 0.092301159 | 6.887860796 | 6.887860796 | 0.00299739 | 0.898667046 | 1 | 0.024751594 | 0.209792175 | 0.835183215 | 262.7311573 | 2.419511579 | FALSE | TRUE | FALSE |
| CTXN3 | 0.7874168 | 0.61357824 | 1 | 0.787416823 | 0.787416823 | 0.787416823 | 0.80987936 | 0.61773199 | 0.0026 | 0.80987936 | 0.80987936 | 0.80987936 | 0.972264341 | -0.012215642 | FALSE | TRUE | FALSE |
| SZRD1 | 0.787264 | 0.61366906 | 1 | 0.787264041 | 0.787264041 | 0.787264041 | 0.80482128 | 0.619556839 | 1 | 0.804821281 | 0.804821281 | 0.804821281 | 0.978184921 | -0.009579036 | FALSE | TRUE | FALSE |
| NTNG2 | 0.7869643 | 0.61375983 | 0.75345 | 0.786964279 | 3.147782215 | 15.48991368 | 0.24394429 | 0.840822747 | 0.01805 | 0.243944289 | 0.243944289 | 4.81074266 | 3.226000016 | 0.508664365 | FALSE | TRUE | FALSE |
| GRIK2 | 0.7868166 | 0.61385055 | 0.6979 | 7.551279249 | 297.1880111 | 297.1880111 | 0.12525468 | 0.870070428 | 1 | 0.125254681 | 0.125254681 | 2.469261936 | 6.281733836 | 0.798079531 | FALSE | TRUE | FALSE |
| SCGN | 0.7866512 | 0.61394123 | 0.00775 | 0.786651223 | 0.786651223 | 0.786651223 | 0.24031047 | 0.841971285 | 1 | 0.240310469 | 0.240310469 | 0.240310469 | 3.273478782 | 0.51500953 | FALSE | TRUE | FALSE |
| FRG2B | 0.7858828 | 0.61403187 | 1 | 0.785882766 | 0.785882766 | 0.785882766 | 0.80452564 | 0.619747855 | 1 | 0.804525636 | 0.804525636 | 0.804525636 | 0.976827501 | -0.010182122 | FALSE | TRUE | FALSE |
| RIBC2 | 0.7854088 | 0.61412247 | 0.0118 | 0.785408799 | 0.785408799 | 1.65606006 | 0.84073428 | 0.60994808 | 0.00865 | 0.840734279 | 0.840734279 | 0.840734279 | 0.934193858 | -0.029562993 | FALSE | TRUE | FALSE |
| OTUB1 | 0.7853993 | 0.61421302 | 1 | 0.785399305 | 0.785399305 | 0.785399305 | 0.8069816 | 0.618598692 | 1 | 0.806981596 | 0.806981596 | 0.806981596 | 0.973255534 | -0.011773118 | FALSE | TRUE | FALSE |
| CCL3 | 0.7852446 | 0.61430353 | 1 | 0.785244621 | 0.785244621 | 0.785244621 | 0.80779867 | 0.618213998 | 1 | 0.807798672 | 0.807798672 | 0.807798672 | 0.972079614 | -0.012298165 | FALSE | TRUE | FALSE |
| MTHFS | 0.7852381 | 0.61439399 | 1 | 0.785238092 | 0.785238092 | 0.785238092 | 7.1899169 | 0.241121279 | 0.504 | 7.189916904 | 7.189916904 | 7.189916904 | 0.109213792 | -0.961722512 | FALSE | FALSE | FALSE |
| CORO2A | 0.7846226 | 0.61448441 | 0.26105 | 0.196235864 | 0.784622643 | 1.751492735 | 0.27743203 | 0.831724604 | 0.95375 | 10.19328655 | 928.1232863 | 3712.06859 | 2.828161664 | 0.451504231 | FALSE | TRUE | FALSE |
| PSMA2 | 0.7843787 | 0.61457479 | 1 | 0.784378737 | 0.784378737 | 0.784378737 | 0.8073959 | 0.618310251 | 1 | 0.807395904 | 0.807395904 | 0.807395904 | 0.971492093 | -0.01256073 | FALSE | TRUE | FALSE |
| ZNF480 | 0.7839597 | 0.61466512 | 0.7548 | 0.783959728 | 7.270293264 | 15.54373009 | 9.45363917 | 0.205389104 | 0.135 | 9.453639173 | 90.3538413 | 90.3538413 | 0.082926767 | -1.081305268 | FALSE | FALSE | FALSE |
| TMEM258 | 0.7838446 | 0.61475541 | 1 | 0.783844633 | 0.783844633 | 0.783844633 | 0.80668497 | 0.618790727 | 1 | 0.806684965 | 0.806684965 | 0.806684965 | 0.971686181 | -0.012473974 | FALSE | TRUE | FALSE |
| TUSC2 | 0.7835458 | 0.61484565 | 1 | 0.783545837 | 0.783545837 | 0.783545837 | 0.79260418 | 0.62305925 | 1 | 0.792604175 | 0.792604175 | 0.792604175 | 0.988571423 | -0.004991948 | FALSE | TRUE | FALSE |
| COPS8 | 0.7833129 | 0.61493585 | 1 | 0.783312946 | 0.783312946 | 0.783312946 | 0.80637043 | 0.618982555 | 0.00375 | 0.806370428 | 0.806370428 | 0.806370428 | 0.971405844 | -0.012599288 | FALSE | TRUE | FALSE |
| ZNF550 | 0.7832232 | 0.61502601 | 0.9832 | 68.09176779 | 6535.927907 | 66359.17735 | 0.11914074 | 0.871444484 | 1 | 94.980395 | 926.1401854 | 9251.313723 | 6.573932645 | 0.81782525 | FALSE | TRUE | FALSE |
| CSF2 | 0.7828495 | 0.61511612 | 1 | 0.782849496 | 0.782849496 | 0.782849496 | 0.80582441 | 0.619078396 | 1 | 0.80582441 | 0.80582441 | 0.80582441 | 0.971488933 | -0.012562142 | FALSE | TRUE | FALSE |
| C9orf72 | 0.7827088 | 0.61520619 | 0.0122 | 0.782708797 | 0.782708797 | 1.690340098 | 0.10401007 | 0.874917764 | 0.0103 | 0.104010069 | 0.104010069 | 2.064931421 | 7.525317533 | 0.87652483 | FALSE | TRUE | FALSE |
| GNG4 | 0.7826136 | 0.61529621 | 1 | 0.782613642 | 0.782613642 | 0.782613642 | 0.80556672 | 0.619174187 | 1 | 0.805566722 | 0.805566722 | 0.805566722 | 0.971506917 | -0.012554103 | FALSE | TRUE | FALSE |
| ACTR1A | 0.7823013 | 0.61538619 | 0.50655 | 0.782301272 | 0.782301272 | 3.877474768 | 0.24008679 | 0.842063684 | 1 | 0.240086793 | 0.240086793 | 4.763351738 | 3.258410274 | 0.513005766 | FALSE | TRUE | FALSE |
| LCE3A | 0.7822418 | 0.61547612 | 1 | 0.782241777 | 0.782241777 | 0.782241777 | 0.80519593 | 0.619365616 | 1 | 0.80519593 | 0.80519593 | 0.80519593 | 0.971492463 | -0.012560564 | FALSE | TRUE | FALSE |
| SNAPC5 | 0.7822311 | 0.61556601 | 0.0027 | 0.19556784 | 0.782231123 | 0.782231123 | 3.69558682 | 0.371596354 | 0.50425 | 3.695586816 | 14.78895762 | 14.78895762 | 0.211666283 | -0.674348317 | FALSE | FALSE | FALSE |
| TJAP1 | 0.78207 | 0.61565585 | 0.0188 | 0.782069958 | 0.782069958 | 2.942013201 | 0.11873091 | 0.871527522 | 0.01345 | 0.118730906 | 0.118730906 | 2.347203908 | 6.586911389 | 0.818681821 | FALSE | TRUE | FALSE |
| PRIMA1 | 0.781828 | 0.61574565 | 1 | 7.393418173 | 71.75670349 | 71.75670349 | 0.80488863 | 0.619461254 | 1 | 7.615512446 | 73.95115657 | 73.95115657 | 0.971349333 | -0.012624554 | FALSE | TRUE | FALSE |
| PTRH1 | 0.7816795 | 0.6158354 | 0.7519 | 0.781679533 | 7.363737474 | 7.363737474 | 0.12084359 | 0.871069563 | 0.0052 | 0.120843589 | 0.120843589 | 0.120843589 | 6.468522992 | 0.810805126 | FALSE | TRUE | FALSE |
| NDFIP1 | 0.7815655 | 0.61592511 | 1 | 0.78156551 | 0.78156551 | 0.78156551 | 0.80463135 | 0.619652373 | 1 | 0.80463135 | 0.80463135 | 0.80463135 | 0.971333655 | -0.012631564 | FALSE | TRUE | FALSE |
| LAPTM5 | 0.7806076 | 0.61601478 | 1 | 0.78060755 | 0.78060755 | 0.78060755 | 0.79936299 | 0.62098476 | 1 | 0.799362988 | 0.799362988 | 0.799362988 | 0.976537021 | -0.010311288 | FALSE | TRUE | FALSE |
| WSB1 | 0.7805878 | 0.6161044 | 0.01335 | 0.780587792 | 0.780587792 | 3.870749932 | 0.2393234 | 0.842202092 | 1 | 0.239323399 | 0.239323399 | 4.750715064 | 3.261644264 | 0.513436592 | FALSE | TRUE | FALSE |
| DCUN1D5 | 0.7805536 | 0.61619398 | 1 | 0.780553619 | 0.780553619 | 0.780553619 | 0.80389542 | 0.61993867 | 1 | 0.803895423 | 0.803895423 | 0.803895423 | 0.970964128 | -0.012796815 | FALSE | TRUE | FALSE |
| TAS2R31 | 0.7801669 | 0.61628352 | 1 | 7.39356575 | 71.91217796 | 71.91217796 | 0.82920269 | 0.612544783 | 1 | 7.867692224 | 7.867692224 | 7.867692224 | 0.940863969 | -0.026473163 | FALSE | TRUE | FALSE |
| ROMO1 | 0.7792263 | 0.61637303 | 1 | 0.779226293 | 0.779226293 | 0.779226293 | 0.80247239 | 0.620129312 | 1 | 0.80247239 | 0.80247239 | 0.80247239 | 0.971031904 | -0.012766501 | FALSE | TRUE | FALSE |
| UBL3 | 0.7785096 | 0.61646249 | 1 | 0.778509627 | 0.778509627 | 0.778509627 | 0.80196677 | 0.620319751 | 1 | 0.801966772 | 0.801966772 | 0.801966772 | 0.970750478 | -0.012892387 | FALSE | TRUE | FALSE |
| DYNLRB1 | 0.7780749 | 0.61655192 | 1 | 0.778074861 | 0.778074861 | 0.778074861 | 0.80128965 | 0.620414902 | 1 | 0.801289653 | 0.801289653 | 0.801289653 | 0.971028214 | -0.012768151 | FALSE | TRUE | FALSE |
| GADD45B | 0.777676 | 0.6166413 | 1 | 0.777676022 | 0.777676022 | 0.777676022 | 0.80125152 | 0.62051 | 1 | 0.801251518 | 0.801251518 | 0.801251518 | 0.97057666 | -0.012970157 | FALSE | TRUE | FALSE |
| LCE2B | 0.7775272 | 0.61673064 | 1 | 0.777527159 | 0.777527159 | 0.777527159 | 0.80092168 | 0.620605049 | 1 | 0.800921676 | 0.800921676 | 0.800921676 | 0.970790505 | -0.01287448 | FALSE | TRUE | FALSE |
| KRTAP19-2 | 0.7773922 | 0.61681994 | 1 | 0.777392193 | 0.777392193 | 0.777392193 | 0.80070721 | 0.620700049 | 1 | 0.800707213 | 0.800707213 | 0.800707213 | 0.970881965 | -0.012833566 | FALSE | TRUE | FALSE |
| GCFC2 | 0.777318 | 0.61690918 | 0.50935 | 0.777318013 | 6.903132623 | 6.903132623 | 0.01871618 | 0.894150098 | 0.50835 | 0.160220927 | 0.160220927 | 0.632919933 | 41.53187165 | 1.618381503 | FALSE | TRUE | FALSE |
| STAU2 | 0.7772764 | 0.61699839 | 0.01125 | 0.777276352 | 0.777276352 | 1.670746382 | 0.45441268 | 0.772967053 | 0.25725 | 0.45441268 | 1.81747192 | 2.25574309 | 1.71050762 | 0.233125013 | FALSE | TRUE | FALSE |
| CAMK2N2 | 0.7770972 | 0.61708755 | 1 | 0.777097227 | 0.777097227 | 0.777097227 | 0.78622636 | 0.625665657 | 1 | 0.786226363 | 0.786226363 | 0.786226363 | 0.988388667 | -0.005072243 | FALSE | TRUE | FALSE |
| GNGT2 | 0.7769949 | 0.61717666 | 1 | 0.776994866 | 0.776994866 | 0.776994866 | 0.80040805 | 0.620794998 | 1 | 0.800408049 | 0.800408049 | 0.800408049 | 0.970748442 | -0.012893298 | FALSE | TRUE | FALSE |
| KLK10 | 0.7763973 | 0.61726574 | 0.01015 | 0.194136769 | 0.776397283 | 0.963591712 | 0.06915734 | 0.882599041 | 1 | 0.276521292 | 1.105929553 | 1.105929553 | 11.22653549 | 1.050245753 | FALSE | TRUE | FALSE |
| FOXI1 | 0.7758617 | 0.61735477 | 0.81315 | 3.102309097 | 12.407787 | 49.63774833 | 0.2872953 | 0.828904408 | 0.7562 | 0.287295305 | 1.148602971 | 1.148602971 | 2.700572135 | 0.431455782 | FALSE | TRUE | FALSE |
| PRSS27 | 0.7756816 | 0.61744377 | 0.50375 | 0.775681559 | 0.775681559 | 0.775681559 | 8.31466045 | 0.221229274 | 0.31915 | 8.314660451 | 133.0760539 | 133.0760539 | 0.093290828 | -1.030161052 | FALSE | FALSE | FALSE |
| SNAI2 | 0.7754497 | 0.61753271 | 1 | 0.775449651 | 0.775449651 | 0.775449651 | 0.79929244 | 0.621079565 | 1 | 0.799292437 | 0.799292437 | 0.799292437 | 0.970170135 | -0.013152099 | FALSE | TRUE | FALSE |
| C16orf46 | 0.7753472 | 0.61762162 | 0.5037 | 0.775347185 | 0.775347185 | 0.775347185 | 1.00133745 | 0.588450352 | 0.7563 | 1.001337446 | 9.214646158 | 9.214646158 | 0.774311585 | -0.111084243 | FALSE | FALSE | FALSE |
| CYP2R1 | 0.7753007 | 0.61771048 | 0.72565 | 1.775744042 | 28.39374734 | 113.581265 | 0.14547318 | 0.86554192 | 0.7499 | 0.145473177 | 0.581812106 | 0.721501153 | 5.329509802 | 0.726687265 | FALSE | TRUE | FALSE |

| | | | | | | | | | | | | | | | | | |
|---|---|---|---|---|---|---|---|---|---|---|---|---|---|---|---|---|---|
| CDC42SE2 | 0.7752218 | 0.61779929 | 1 | 0.775221837 | 0.775221837 | 0.775221837 | 0.7849201 | 0.626034899 | 1 | 0.784920103 | 0.784920103 | 0.784920103 | 0.987644264 | -0.005399454 | FALSE | TRUE | FALSE |
| KRTAP19-1 | 0.7751364 | 0.61788806 | 1 | 0.775136354 | 0.775136354 | 0.775136354 | 0.79874928 | 0.621269029 | 0.0022 | 0.798749283 | 0.798749283 | 0.798749283 | 0.970437621 | -0.013032375 | FALSE | TRUE | FALSE |
| CEND1 | 0.7746532 | 0.61797679 | 1 | 0.774653175 | 0.774653175 | 0.774653175 | 0.79839023 | 0.621363688 | 1 | 0.798390226 | 0.798390226 | 0.798390226 | 0.970268861 | -0.013107906 | FALSE | TRUE | FALSE |
| POLE4 | 0.7746133 | 0.61806547 | 1 | 0.774613258 | 0.774613258 | 0.774613258 | 0.79815533 | 0.621458297 | 1 | 0.798155329 | 0.798155329 | 0.798155329 | 0.970504399 | -0.013002492 | FALSE | TRUE | FALSE |
| RAB10 | 0.7745054 | 0.61815411 | 1 | 0.774505383 | 0.774505383 | 0.774505383 | 0.79494486 | 0.622495829 | 1 | 0.794944858 | 0.794944858 | 0.794944858 | 0.974288186 | -0.011312564 | FALSE | TRUE | FALSE |
| CXCL10 | 0.7739132 | 0.61824271 | 1 | 0.773913186 | 0.773913186 | 0.773913186 | 0.79743296 | 0.621552863 | 1 | 0.797432956 | 0.797432956 | 0.797432956 | 0.970505646 | -0.013001934 | FALSE | TRUE | FALSE |
| CALM1 | 0.7738987 | 0.61833127 | 1 | 3.095521481 | 3.095521481 | 3.095521481 | 0.79991051 | 0.620889902 | 1 | 0.799910512 | 0.799910512 | 0.799910512 | 0.96748154 | -0.014357313 | FALSE | TRUE | FALSE |
| EVA1B | 0.7737942 | 0.61841978 | 1 | 7.341198111 | 7.341198111 | 7.341198111 | 0.66037452 | 0.685497058 | 1 | 0.660374523 | 0.660374523 | 0.660374523 | 1.171750567 | 0.068835172 | FALSE | TRUE | FALSE |
| PRPF6 | 0.7735425 | 0.61850825 | 0.51545 | 0.773542489 | 0.773542489 | 15.09328793 | 0.1128452 | 0.872781583 | 0.5163 | 1.037264986 | 1.037264986 | 20.29853357 | 6.854899562 | 0.836001096 | FALSE | TRUE | FALSE |
| RALA | 0.7730561 | 0.61859668 | 1 | 0.773056084 | 0.773056084 | 0.773056084 | 0.79502087 | 0.622401756 | 1 | 0.795020868 | 0.795020868 | 0.795020868 | 0.972372066 | -0.012167526 | FALSE | TRUE | FALSE |
| PABPN1 | 0.7729409 | 0.61868506 | 1 | 0.772940901 | 0.772940901 | 0.772940901 | 0.79677581 | 0.621741854 | 1 | 0.796775811 | 0.796775811 | 0.796775811 | 0.970085802 | -0.013189852 | FALSE | TRUE | FALSE |
| CCL15 | 0.7728558 | 0.6187734 | 0.0027 | 0.772855783 | 0.772855783 | 0.772855783 | 0.79672302 | 0.621836271 | 1 | 0.796723019 | 0.796723019 | 0.796723019 | 0.970043245 | -0.013208904 | FALSE | TRUE | FALSE |
| OR6C76 | 0.7720221 | 0.6188617 | 1 | 0.772022111 | 0.772022111 | 2.4755E+301 | 6.113E+295 | 0 | 0.2492 | 1.773E+284 | 5.5099E+305 | 2.5473E+307 | 1.263E-296 | -295.898598 | FALSE | FALSE | FALSE |
| RRM2 | 0.7715798 | 0.61894997 | 0.5037 | 0.771579812 | 0.771579812 | 0.771579812 | 1.03773922 | 0.583964044 | 0.0029 | 0.110938219 | 1.037739219 | 1.037739219 | 0.74351995 | -0.128707374 | FALSE | FALSE | FALSE |
| FAM124B | 0.7711841 | 0.61903819 | 0.7488 | 0.771184104 | 7.045263681 | 7.045263681 | 0.12026201 | 0.87117391 | 1 | 0.481040531 | 0.481040531 | 0.481040531 | 6.412532882 | 0.807029605 | FALSE | TRUE | FALSE |
| LYRM7 | 0.7711288 | 0.61912637 | 1 | 0.771128759 | 0.771128759 | 0.771128759 | 0.79507063 | 0.622307632 | 1 | 0.795070635 | 0.795070635 | 0.795070635 | 0.969887108 | -0.013278813 | FALSE | TRUE | FALSE |
| CNIH1 | 0.771022 | 0.61921451 | 1 | 0.771022003 | 0.771022003 | 0.771022003 | 0.77026775 | 0.6323746 | 1 | 7.28042105 | 7.28042105 | 7.28042105 | 1.000979206 | 0.000425056 | FALSE | TRUE | FALSE |
| IQCJ | 0.7708485 | 0.6193026 | 0.00405 | 0.770848495 | 0.770848495 | 0.770848495 | 0.7949118 | 0.622683818 | 1 | 0.794911795 | 0.794911795 | 0.794911795 | 0.969728339 | -0.013349912 | FALSE | TRUE | FALSE |
| PPIA | 0.7707883 | 0.61939065 | 1 | 0.770788347 | 0.770788347 | 0.770788347 | 0.79493054 | 0.62258985 | 0.00405 | 0.794930542 | 0.794930542 | 0.794930542 | 0.969629805 | -0.013394044 | FALSE | TRUE | FALSE |
| COCH | 0.7707411 | 0.61947865 | 0.13795 | 0.192709661 | 0.770741086 | 0.954215379 | 0.32581868 | 0.816874613 | 0.01125 | 0.081479021 | 0.325818681 | 0.404006882 | 2.365552168 | 0.37393253 | FALSE | TRUE | FALSE |
| ERC2 | 0.770631 | 0.61956661 | 0.51095 | 1.788471559 | 7.153417536 | 60.25901301 | 0.24396552 | 0.840775832 | 1 | 0.243965522 | 0.243965522 | 4.800866555 | 3.158770063 | 0.499518013 | FALSE | TRUE | FALSE |
| UBE2H | 0.7695935 | 0.61965453 | 1 | 0.769593513 | 0.769593513 | 0.769593513 | 0.79389245 | 0.622777748 | 1 | 0.793892452 | 0.793892452 | 0.793892452 | 0.969392657 | -0.013500274 | FALSE | TRUE | FALSE |
| PIWIL2 | 0.7695826 | 0.61974241 | 0.5113 | 0.769582556 | 0.769582556 | 15.09327979 | 0.11681116 | 0.872033937 | 0.0249 | 0.116811162 | 0.116811162 | 2.296283044 | 6.588262131 | 0.81877087 | FALSE | TRUE | FALSE |
| WDR83OS | 0.7695256 | 0.61983025 | 1 | 0.769525639 | 0.769525639 | 0.769525639 | 0.79371214 | 0.622871628 | 1 | 0.793712139 | 0.793712139 | 0.793712139 | 0.969527366 | -0.013439928 | FALSE | TRUE | FALSE |
| THRA | 0.7693477 | 0.61991804 | 0.69795 | 1.780905236 | 28.48294104 | 28.48294104 | 0.07179167 | 0.88206961 | 1 | 0.287021491 | 1.147792394 | 5.673488538 | 10.71639196 | 1.03004859 | FALSE | TRUE | FALSE |
| ELOF1 | 0.7692282 | 0.62000579 | 1 | 0.7692282 | 0.7692282 | 0.7692282 | 0.79342876 | 0.622965459 | 0.0025 | 0.793428761 | 0.793428761 | 0.793428761 | 0.96949876 | -0.013452742 | FALSE | TRUE | FALSE |
| ABHD17C | 0.7688025 | 0.6200935 | 1 | 0.768802505 | 0.768802505 | 0.768802505 | 7.26186597 | 0.239144235 | 0.50765 | 7.261865966 | 7.261865966 | 7.261865966 | 0.105868451 | -0.975233439 | FALSE | FALSE | FALSE |
| PARVA | 0.7687171 | 0.62018117 | 0.7523 | 7.15913539 | 68.4296546 | 68.4296546 | 0.21024666 | 0.849647669 | 1 | 299.1399899 | 29444.34686 | 75740.47107 | 3.656262772 | 0.5630374 | FALSE | TRUE | FALSE |
| GADD45G | 0.7684082 | 0.6202688 | 1 | 0.768408205 | 0.768408205 | 0.768408205 | 0.78734907 | 0.62511035 | 1 | 0.787349072 | 0.787349072 | 0.787349072 | 0.975943494 | -0.010575327 | FALSE | TRUE | FALSE |
| RPL21 | 0.7680347 | 0.62035638 | 1 | 0.76803466 | 0.76803466 | 0.76803466 | 0.79250606 | 0.623152989 | 1 | 0.79250606 | 0.79250606 | 0.79250606 | 0.969121498 | -0.013621772 | FALSE | TRUE | FALSE |
| EMC6 | 0.7675372 | 0.62044393 | 1 | 0.767537221 | 0.767537221 | 0.767537221 | 0.79192047 | 0.623340322 | 1 | 0.791920468 | 0.791920468 | 0.791920468 | 0.96920998 | -0.013582123 | FALSE | TRUE | FALSE |
| SLC35F2 | 0.7668001 | 0.62053144 | 0.00925 | 0.766800129 | 0.766800129 | 0.766800129 | 8.00534634 | 0.227897677 | 0.3808 | 8.005346338 | 304.2167868 | 304.2167868 | 0.095786003 | -1.018697949 | FALSE | FALSE | FALSE |
| FITM1 | 0.7667762 | 0.6206189 | 0.75085 | 0.766776182 | 7.039663633 | 7.039663633 | 0.12068417 | 0.871100883 | 0.00925 | 0.120684167 | 0.120684167 | 0.120684167 | 6.353577298 | 0.803018318 | FALSE | TRUE | FALSE |
| SNRPF | 0.7664744 | 0.62070632 | 1 | 0.7664744 | 0.7664744 | 0.7664744 | 0.77429607 | 0.63076606 | 1 | 0.774296067 | 0.774296067 | 0.774296067 | 0.989898351 | -0.004409399 | FALSE | TRUE | FALSE |
| RPL36 | 0.7664695 | 0.6207937 | 1 | 0.766469509 | 0.766469509 | 0.766469509 | 0.7779224 | 0.62932377 | 1 | 0.777922405 | 0.777922405 | 0.777922405 | 0.985277585 | -0.006441397 | FALSE | TRUE | FALSE |
| TCAP | 0.7663512 | 0.62088104 | 0.5007 | 0.766351151 | 0.766351151 | 0.766351151 | 0.2355836 | 0.843149123 | 0.0054 | 0.2355836 | 0.2355836 | 0.2355836 | 3.252990234 | 0.51228276 | FALSE | TRUE | FALSE |
| RBPMS2 | 0.7658918 | 0.62096834 | 1 | 0.76589178 | 0.76589178 | 0.76589178 | 0.79039847 | 0.623807834 | 1 | 0.790398474 | 0.790398474 | 0.790398474 | 0.968994507 | -0.013678685 | FALSE | TRUE | FALSE |
| PTMA | 0.7657592 | 0.62105559 | 1 | 0.765759199 | 0.765759199 | 0.765759199 | 0.7902711 | 0.623901188 | 1 | 0.790271102 | 0.790271102 | 0.790271102 | 0.968982919 | -0.013683879 | FALSE | TRUE | FALSE |
| MIF | 0.7655394 | 0.6211428 | 1 | 0.765539407 | 0.765539407 | 0.765539407 | 0.79015004 | 0.623994491 | 1 | 0.790150043 | 0.790150043 | 0.790150043 | 0.968853212 | -0.013742017 | FALSE | TRUE | FALSE |
| LY6H | 0.7647493 | 0.62122998 | 1 | 0.764749325 | 0.764749325 | 0.764749325 | 0.76467861 | 0.634232809 | 1 | 0.764678605 | 7.228526044 | 7.228526044 | 1.000092483 | 4.01631E-05 | FALSE | TRUE | FALSE |
| BBC3 | 0.7644351 | 0.62131712 | 1 | 0.764435105 | 0.764435105 | 0.764435105 | 0.78896614 | 0.624460295 | 1 | 0.788966141 | 0.788966141 | 0.788966141 | 0.968907366 | -0.013717743 | FALSE | TRUE | FALSE |
| SCAMP1 | 0.7644192 | 0.62140421 | 1 | 0.764419196 | 0.764419196 | 0.764419196 | 0.78928557 | 0.624180962 | 0.00745 | 0.789285567 | 0.789285567 | 0.789285567 | 0.96849509 | -0.013902577 | FALSE | TRUE | FALSE |
| CMTM6 | 0.7643599 | 0.62149126 | 1 | 0.764359879 | 0.764359879 | 0.764359879 | 0.78904159 | 0.624367234 | 1 | 0.789041586 | 0.789041586 | 0.789041586 | 0.968719384 | -0.01380201 | FALSE | TRUE | FALSE |
| PSMA6 | 0.7642526 | 0.62157826 | 1 | 0.764252589 | 0.764252589 | 0.764252589 | 0.78883105 | 0.624646266 | 1 | 0.788831054 | 0.788831054 | 0.788831054 | 0.968841915 | -0.013747081 | FALSE | TRUE | FALSE |
| SPATA32 | 0.764116 | 0.62166522 | 0.7514 | 0.764116041 | 6.986919707 | 6.986919707 | 1.05113984 | 0.583283368 | 0.50435 | 1.051139839 | 1.051139839 | 2.324710594 | 0.726940425 | -0.13850118 | FALSE | FALSE | FALSE |
| MT1F | 0.763759 | 0.62175214 | 0.00195 | 0.763759043 | 0.763759043 | 0.763759043 | 0.78831236 | 0.62483204 | 1 | 0.788312365 | 0.788312365 | 0.788312365 | 0.968853309 | -0.013741973 | FALSE | TRUE | FALSE |
| SH2D6 | 0.7637329 | 0.62183902 | 1 | 1.85211E+12 | 4.5009E+16 | 5.88859E+17 | 0.16742763 | 0.860283825 | 0.99975 | 144925.6047 | 148948374.6 | 1686981446 | 4.5615701 | 0.659114353 | FALSE | TRUE | FALSE |
| EPPIN | 0.7637097 | 0.62192586 | 0.00375 | 0.763709691 | 0.763709691 | 0.763709691 | 6.97391835 | 0.245383813 | 0.74985 | 6.973918352 | 67.57927842 | 67.57927842 | 0.109509411 | -0.960548557 | FALSE | FALSE | FALSE |
| RAB14 | 0.763586 | 0.62201265 | 1 | 0.763586019 | 0.763586019 | 0.763586019 | 15.6712134 | 0.156277876 | 0.00125 | 0.786296648 | 0.786296648 | 0.786296648 | 0.048725392 | -1.312244655 | FALSE | FALSE | FALSE |
| PRAMEF6 | 0.7635678 | 0.62209939 | 1 | 0.763567849 | 0.763567849 | 15.13736047 | 0.78864176 | 0.624739176 | 1 | 0.788641761 | 0.788641761 | 15.65093765 | 0.968206209 | -0.014032137 | FALSE | TRUE | FALSE |
| PPP1R11 | 0.7627164 | 0.62218611 | 1 | 0.76271643 | 0.76271643 | 0.76271643 | 0.78729689 | 0.625203022 | 1 | 0.78729689 | 0.78729689 | 0.78729689 | 0.968778664 | -0.013775434 | FALSE | TRUE | FALSE |
| EIF4E3 | 0.7622516 | 0.62227278 | 1 | 0.762251552 | 0.762251552 | 0.762251552 | 0.78721704 | 0.625295644 | 1 | 0.78721704 | 0.78721704 | 0.78721704 | 0.968286398 | -0.013996169 | FALSE | TRUE | FALSE |
| TNFSF13B | 0.7621898 | 0.62235941 | 1 | 7.228246297 | 7.228246297 | 7.228246297 | 0.79603981 | 0.622024958 | 1 | 0.796039811 | 0.796039811 | 0.796039811 | 0.95747699 | -0.018871654 | FALSE | TRUE | FALSE |
| NKAIN3 | 0.7620304 | 0.622446 | 1 | 0.762030445 | 0.762030445 | 0.762030445 | 7.20617966 | 0.240463265 | 0.4973 | 0.762444527 | 7.206179659 | 7.206179659 | 0.105746801 | -0.975732763 | FALSE | FALSE | FALSE |
| GSR | 0.7620051 | 0.62253254 | 0.38 | 0.331760184 | 3.046340404 | 3.046340404 | 0.03312652 | 0.890582226 | 1 | 0.305360475 | 1.220583319 | 1.220583319 | 23.00286997 | 1.361782025 | FALSE | TRUE | FALSE |
| SERPINA4 | 0.7618194 | 0.62261905 | 0.2606 | 0.422726161 | 6.760585811 | 6.760585811 | 134.561399 | 0.021067792 | 0.1906 | 13.99257328 | 134.5613989 | 1325.659508 | 0.005661501 | -2.247068437 | FALSE | FALSE | FALSE |
| SPINK1 | 0.7617008 | 0.62270551 | 0.00225 | 0.761700797 | 0.761700797 | 0.761700797 | 0.78663441 | 0.625480748 | 1 | 0.786634411 | 0.786634411 | 0.786634411 | 0.968303429 | -0.01398853 | FALSE | TRUE | FALSE |
| HMGA2 | 0.7616893 | 0.62279192 | 1 | 0.761689346 | 0.761689346 | 0.761689346 | 0.78643028 | 0.625573227 | 1 | 0.786430277 | 0.786430277 | 0.786430277 | 0.968540211 | -0.013882344 | FALSE | TRUE | FALSE |
| PAIP2 | 0.7616465 | 0.62287829 | 1 | 7.195502706 | 69.76736994 | 69.76736994 | 0.81172752 | 0.616861188 | 1 | 7.680570266 | 7.680570266 | 7.680570266 | 0.938303108 | -0.027656845 | FALSE | TRUE | FALSE |
| AP1M2 | 0.7614771 | 0.62296462 | 0.5026 | 0.761477132 | 3.044912995 | 3.044912995 | 0.45821803 | 0.771803333 | 0.5087 | 0.45821803 | 0.45821803 | 2.271259872 | 1.661822718 | 0.220584692 | FALSE | TRUE | FALSE |
| ZNF771 | 0.7612835 | 0.62305091 | 1 | 0.761283514 | 0.761283514 | 0.761283514 | 3.04094301 | 0.410058242 | 0.5021 | 3.040943009 | 3.040943009 | 3.040943009 | 0.250344552 | -0.601461856 | FALSE | FALSE | FALSE |
| NRARP | 0.7612699 | 0.62313716 | 1 | 0.761269909 | 0.761269909 | 0.761269909 | 0.77628178 | 0.62977577 | 1 | 0.776281777 | 0.776281777 | 0.776281777 | 0.980661831 | -0.008480728 | FALSE | TRUE | FALSE |
| ASTN2 | 0.7611504 | 0.62322336 | 0.5162 | 0.761150362 | 0.761150362 | 14.80763771 | 0.1106397 | 0.873187932 | 0.51475 | 1.023930859 | 1.023930859 | 19.98536138 | 6.879540886 | 0.837559456 | FALSE | TRUE | FALSE |
| FLYWCH2 | 0.7610166 | 0.62330952 | 1 | 0.761016598 | 0.761016598 | 0.761016598 | 0.78592934 | 0.625758041 | 0.00335 | 0.785929337 | 0.785929337 | 0.785929337 | 0.968301553 | -0.013989372 | FALSE | TRUE | FALSE |
| KLF2 | 0.760857 | 0.62339564 | 1 | 0.760856959 | 0.760856959 | 0.760856959 | 0.7858869 | 0.625942655 | 1 | 0.785886898 | 0.785886898 | 0.785886898 | 0.968150711 | -0.014057031 | FALSE | TRUE | FALSE |
| THAP1 | 0.7604973 | 0.62348172 | 0.50605 | 0.760497256 | 0.760497256 | 0.760497256 | 0.22683758 | 0.845571479 | 1 | 2.138219845 | 2.138219845 | 2.138219845 | 3.352607027 | 0.525382651 | FALSE | TRUE | FALSE |
| KLRF1 | 0.7603506 | 0.62356776 | 0.5008 | 0.760350555 | 0.760350555 | 0.760350555 | 0.11626468 | 0.872126544 | 1 | 0.116264677 | 0.116264677 | 0.116264677 | 6.539824274 | 0.815566079 | FALSE | TRUE | FALSE |
| RAN | 0.760118 | 0.62365375 | 1 | 0.760117996 | 0.760117996 | 0.760117996 | 0.78335037 | 0.626862925 | 1 | 0.783350368 | 0.783350368 | 0.783350368 | 0.970342298 | -0.013075037 | FALSE | TRUE | FALSE |
| MRFAP1 | 0.7600582 | 0.62373971 | 1 | 7.188979024 | 7.188979024 | 7.188979024 | 0.81040116 | 0.617442195 | 1 | 0.810401161 | 0.810401161 | 0.810401161 | 0.937878975 | -0.0278532 | FALSE | TRUE | FALSE |
| S100A5 | 0.7592298 | 0.62382563 | 1 | 0.75922984 | 0.75922984 | 0.75922984 | 0.78421143 | 0.626311345 | 1 | 0.784211434 | 0.784211434 | 0.784211434 | 0.968144313 | -0.014059901 | FALSE | TRUE | FALSE |
| PDGFC | 0.759194 | 0.6239115 | 0.0097 | 0.759193959 | 0.759193959 | 0.759193959 | 0.78174003 | 0.627504261 | 1 | 0.781740033 | 0.781740033 | 0.781740033 | 0.971159115 | -0.01270961 | FALSE | TRUE | FALSE |
| IPP | 0.7589086 | 0.62399734 | 0.86045 | 3.035737878 | 48.61152405 | 194.5984484 | 20.6260026 | 0.129524598 | 0.23865 | 5.154583276 | 82.55531314 | 330.5090952 | 0.036793781 | -1.434225576 | FALSE | FALSE | FALSE |
| TOMM40 | 0.7587319 | 0.62408313 | 1 | 0.758731918 | 0.758731918 | 0.758731918 | 0.78410538 | 0.626403397 | 1 | 0.784105383 | 0.784105383 | 0.784105383 | 0.967640237 | -0.014286081 | FALSE | TRUE | FALSE |

| | | | | | | | | | | | | | | | | | |
|---|---|---|---|---|---|---|---|---|---|---|---|---|---|---|---|---|---|
| STAT1 | 0.7586709 | 0.62416888 | 0.5035 | 0.758670908 | 0.758670908 | 6.413174738 | 0.23355275 | 0.843799431 | 0.0185 | 0.233552749 | 0.233552749 | 4.610864079 | 3.248392116 | 0.511668448 | FALSE | TRUE | FALSE |
| CARD18 | 0.758616 | 0.62425459 | 1 | 0.75861602 | 0.75861602 | 0.75861602 | 0.78365212 | 0.626679259 | 1 | 0.783652122 | 0.783652122 | 0.783652122 | 0.968052019 | -0.014101305 | FALSE | TRUE | FALSE |
| NQO2 | 0.7585447 | 0.62434026 | 0.7524 | 1.725082525 | 6.900297896 | 6.900297896 | 0.90425294 | 0.598032929 | 0.0062 | 0.098915557 | 0.904252944 | 0.904252944 | 0.838863403 | -0.076308752 | FALSE | TRUE | FALSE |
| SUMO3 | 0.7584296 | 0.62442588 | 1 | 0.758429609 | 0.758429609 | 0.758429609 | 0.7591391 | 0.637113227 | 1 | 7.169700424 | 7.169700424 | 7.169700424 | 0.999065403 | -0.00040608 | FALSE | TRUE | FALSE |
| ELP3 | 0.7584283 | 0.62451147 | 0.9488 | 26.59253992 | 1706.154993 | 8425.357473 | 0.08701387 | 0.878781144 | 0.63115 | 0.348020928 | 3.079984959 | 3.079984959 | 8.716177033 | 0.940326043 | FALSE | TRUE | FALSE |
| ARPP19 | 0.7582172 | 0.62459701 | 1 | 0.758217192 | 0.758217192 | 0.758217192 | 0.78350555 | 0.626771117 | 1 | 0.783505553 | 0.783505553 | 0.783505553 | 0.967724082 | -0.014248451 | FALSE | TRUE | FALSE |
| ATPAF2 | 0.7575588 | 0.62468251 | 0.8775 | 3.030335598 | 28.12542117 | 28.12542117 | 0.24300209 | 0.841150371 | 1 | 0.243002093 | 0.243002093 | 0.243002093 | 3.117498935 | 0.493806314 | FALSE | TRUE | FALSE |
| RANBP1 | 0.7574077 | 0.62476797 | 1 | 0.757407725 | 0.757407725 | 0.757407725 | 0.78282026 | 0.627046399 | 1 | 0.782820262 | 0.782820262 | 0.782820262 | 0.967537201 | -0.014332328 | FALSE | TRUE | FALSE |
| ACBD7 | 0.7574002 | 0.62485339 | 1 | 0.757400154 | 0.757400154 | 0.757400154 | 0.78254401 | 0.627138067 | 1 | 0.782544013 | 0.782544013 | 0.782544013 | 0.96786908 | -0.014183384 | FALSE | TRUE | FALSE |
| UBE2D2 | 0.7573673 | 0.62493877 | 1 | 0.757367281 | 0.757367281 | 0.757367281 | 3.09557038 | 0.405943433 | 0.49665 | 0.773904951 | 3.095570381 | 3.095570381 | 0.244661626 | -0.611434143 | FALSE | FALSE | FALSE |
| MAN2B2 | 0.757316 | 0.62502411 | 0.42435 | 0.347260651 | 27.04494052 | 247.5738482 | 0.0040719 | 0.898302433 | 0.81745 | 0.016276853 | 0.167603161 | 0.669803278 | 185.9860104 | 2.269480278 | FALSE | TRUE | FALSE |
| S100A4 | 0.7572426 | 0.6251094 | 1 | 0.757242635 | 0.757242635 | 0.757242635 | 0.78240935 | 0.627229687 | 1 | 0.78240935 | 0.78240935 | 0.78240935 | 0.967834338 | -0.014198974 | FALSE | TRUE | FALSE |
| HBEGF | 0.7567383 | 0.62519465 | 1 | 0.756738298 | 0.756738298 | 0.756738298 | 0.78216724 | 0.627321259 | 1 | 0.78216724 | 0.78216724 | 0.78216724 | 0.967489124 | -0.014353908 | FALSE | TRUE | FALSE |
| EMP3 | 0.7566089 | 0.62527987 | 0.75345 | 3.025081602 | 28.64168393 | 28.64168393 | 0.45372659 | 0.773191916 | 0.50335 | 1.051282292 | 4.203469563 | 4.203469563 | 1.667543667 | 0.222077215 | FALSE | TRUE | FALSE |
| HBG2 | 0.7561997 | 0.62536504 | 1 | 0.756199688 | 0.756199688 | 0.756199688 | 0.77292977 | 0.631214345 | 1 | 3.091562188 | 3.091562188 | 3.091562188 | 0.978354974 | -0.009503543 | FALSE | TRUE | FALSE |
| DENR | 0.7555808 | 0.62545018 | 1 | 0.755580788 | 0.755580788 | 0.755580788 | 0.78101242 | 0.627869696 | 1 | 0.781012424 | 0.781012424 | 0.781012424 | 0.967437604 | -0.014377036 | FALSE | TRUE | FALSE |
| BTG1 | 0.7554784 | 0.62553528 | 1 | 0.755478358 | 0.755478358 | 0.755478358 | 0.78096882 | 0.627960933 | 1 | 0.780968819 | 0.780968819 | 0.780968819 | 0.967360463 | -0.014411667 | FALSE | TRUE | FALSE |
| ZNF474 | 0.7554368 | 0.62562034 | 0.50655 | 0.755436764 | 0.755436764 | 1.671700271 | 3.87784392 | 0.362569093 | 0.5071 | 3.877843919 | 35.93648488 | 35.93648488 | 0.19480845 | -0.710392209 | FALSE | FALSE | FALSE |
| FXYD1 | 0.7551842 | 0.62570535 | 1 | 0.755184176 | 0.755184176 | 0.755184176 | 0.7806373 | 0.628143263 | 1 | 0.780637299 | 0.780637299 | 0.780637299 | 0.967394431 | -0.014396417 | FALSE | TRUE | FALSE |
| SRP9 | 0.7551559 | 0.62579033 | 1 | 0.755155881 | 0.755155881 | 0.755155881 | 0.78039175 | 0.6283254 | 1 | 0.78039175 | 0.78039175 | 0.78039175 | 0.967662563 | -0.014276061 | FALSE | TRUE | FALSE |
| C11orf58 | 0.7550609 | 0.62587526 | 1 | 0.75506087 | 0.75506087 | 0.75506087 | 0.78057504 | 0.628234355 | 1 | 0.780575042 | 0.780575042 | 0.780575042 | 0.967313621 | -0.014432697 | FALSE | TRUE | FALSE |
| NXPH2 | 0.7548748 | 0.62596015 | 0.75725 | 7.145991754 | 69.42768646 | 69.42768646 | 0.83158352 | 0.61214757 | 1 | 0.831583523 | 0.831583523 | 0.831583523 | 0.907755822 | -0.042030957 | FALSE | TRUE | FALSE |
| LSM5 | 0.7545371 | 0.626045 | 1 | 0.754537104 | 0.754537104 | 0.754537104 | 0.77990652 | 0.628689099 | 1 | 0.779906523 | 0.779906523 | 0.779906523 | 0.967471204 | -0.014361952 | FALSE | TRUE | FALSE |
| S100A7A | 0.754217 | 0.62612981 | 1 | 0.754216977 | 0.754216977 | 0.754216977 | 0.77965964 | 0.628779905 | 1 | 0.779659642 | 0.779659642 | 0.779659642 | 0.96736696 | -0.01440875 | FALSE | TRUE | FALSE |
| CNRIP1 | 0.753743 | 0.62621459 | 1 | 0.75374303 | 0.75374303 | 0.75374303 | 0.779291 | 0.628961374 | 1 | 0.779290998 | 0.779290998 | 0.779290998 | 0.967216395 | -0.01447635 | FALSE | TRUE | FALSE |
| MCAM | 0.7533338 | 0.62629933 | 0.507 | 0.753333809 | 0.753333809 | 2.942277567 | 0.11463081 | 0.872454829 | 1 | 0.114630811 | 0.114630811 | 2.261803703 | 6.571826575 | 0.817686094 | FALSE | TRUE | FALSE |
| DPY30 | 0.7532489 | 0.62638402 | 1 | 0.753248948 | 0.753248948 | 0.753248948 | 0.76721324 | 0.633261883 | 1 | 0.767213239 | 0.767213239 | 0.767213239 | 0.981798683 | -0.007977555 | FALSE | TRUE | FALSE |
| SPCS2 | 0.7531243 | 0.62646868 | 1 | 0.753124266 | 0.753124266 | 0.753124266 | 0.76592902 | 0.633703893 | 1 | 0.765929017 | 0.765929017 | 0.765929017 | 0.983282066 | -0.007321882 | FALSE | TRUE | FALSE |
| GABARAP | 0.7514721 | 0.62655331 | 0.00495 | 0.751472078 | 0.751472078 | 0.751472078 | 7.12480204 | 0.242762771 | 0.5016 | 7.124802039 | 7.124802039 | 7.124802039 | 0.105472696 | -0.976859953 | FALSE | FALSE | FALSE |
| DDX27 | 0.7511276 | 0.62663791 | 0.7531 | 0.751127622 | 7.056739684 | 25.35579583 | 0.11691857 | 0.871982434 | 1 | 0.11691857 | 0.11691857 | 2.3002616 | 6.424365431 | 0.807830237 | FALSE | TRUE | FALSE |
| ANKAR | 0.7508864 | 0.62672246 | 0.517 | 6.539312885 | 533.0549594 | 1963.435347 | 0.02451771 | 0.892606675 | 0.9383 | 0.20717113 | 15.95444833 | 15.95444833 | 30.62628829 | 1.486094366 | FALSE | TRUE | FALSE |
| RPL14 | 0.7506764 | 0.62680698 | 1 | 0.750676408 | 0.750676408 | 0.750676408 | 0.77670143 | 0.629504711 | 0.0061 | 0.776701431 | 0.776701431 | 0.776701431 | 0.966492887 | -0.014801338 | FALSE | TRUE | FALSE |
| ZMYND19 | 0.7505458 | 0.62689145 | 1 | 0.750545782 | 0.750545782 | 0.750545782 | 0.77657643 | 0.629595112 | 1 | 0.776576434 | 0.776576434 | 0.776576434 | 0.966480245 | -0.014807019 | FALSE | TRUE | FALSE |
| GREM2 | 0.7505332 | 0.62697588 | 1 | 0.750533249 | 0.750533249 | 0.750533249 | 0.77649797 | 0.629685464 | 1 | 0.776497974 | 0.776497974 | 0.776497974 | 0.966561761 | -0.014770391 | FALSE | TRUE | FALSE |
| DMRTA1 | 0.7504056 | 0.62706027 | 0.7489 | 0.750405646 | 6.921384834 | 6.921384834 | 0.2380123 | 0.842570012 | 1 | 0.2380123 | 0.2380123 | 4.723821885 | 3.152801958 | 0.498696692 | FALSE | TRUE | FALSE |
| TSPAN18 | 0.75026 | 0.62714462 | 0.49665 | 0.187634167 | 0.750260027 | 0.750260027 | 8.23695769 | 0.224236307 | 0.38295 | 3.505146815 | 32.94523055 | 32.94523055 | 0.091084604 | -1.040555027 | FALSE | FALSE | FALSE |
| G0S2 | 0.7501774 | 0.62722893 | 1 | 0.750177403 | 0.750177403 | 0.750177403 | 0.77601449 | 0.62986603 | 1 | 0.776014486 | 0.776014486 | 0.776014486 | 0.966705412 | -0.01470585 | FALSE | TRUE | FALSE |
| NLGN2 | 0.7501619 | 0.6273132 | 1 | 3.07863E+19 | 1.96884E+25 | 4.34514E+27 | 0.08034195 | 0.88020958 | 1 | 4.35844E+16 | 1.45429E+21 | 2.94709E+23 | 9.337113829 | 0.970212653 | FALSE | TRUE | FALSE |
| SMIM19 | 0.7496331 | 0.62739743 | 1 | 0.749633061 | 0.749633061 | 0.749633061 | 0.77547728 | 0.629956248 | 1 | 0.77547728 | 0.77547728 | 0.77547728 | 0.966673145 | -0.014720346 | FALSE | TRUE | FALSE |
| IER5L | 0.7493959 | 0.62748162 | 1 | 0.749395894 | 0.749395894 | 14.87353888 | 7.3312494 | 0.237493358 | 0.0094 | 7.331249402 | 7.331249402 | 7.331249402 | 0.102219397 | -0.990466685 | FALSE | FALSE | FALSE |
| DDA1 | 0.7493227 | 0.62756577 | 1 | 0.749322706 | 0.749322706 | 0.749322706 | 0.77532783 | 0.630046419 | 1 | 0.775327831 | 0.775327831 | 0.775327831 | 0.966459188 | -0.014816481 | FALSE | TRUE | FALSE |
| ZFAND6 | 0.7491808 | 0.62764987 | 1 | 0.749180837 | 0.749180837 | 0.749180837 | 0.77526482 | 0.630136542 | 1 | 0.775264822 | 0.775264822 | 0.775264822 | 0.966354742 | -0.014863418 | FALSE | TRUE | FALSE |
| HMGB1 | 0.7491556 | 0.62773394 | 1 | 0.7491556 | 0.7491556 | 0.7491556 | 0.7751867 | 0.630406617 | 1 | 0.775186702 | 0.775186702 | 0.775186702 | 0.96641957 | -0.014834284 | FALSE | TRUE | FALSE |
| PEA15 | 0.7491418 | 0.62781796 | 1 | 0.749141828 | 0.749141828 | 0.749141828 | 0.775203 | 0.63031664 | 0.00285 | 0.775203003 | 0.775203003 | 0.775203003 | 0.966381483 | -0.0148514 | FALSE | TRUE | FALSE |
| MED9 | 0.7490864 | 0.62790195 | 1 | 0.749086351 | 0.749086351 | 0.749086351 | 0.77507218 | 0.630496546 | 1 | 0.775072183 | 0.775072183 | 0.775072183 | 0.966473016 | -0.014810267 | FALSE | TRUE | FALSE |
| PCNP | 0.7488361 | 0.62798589 | 1 | 0.748836137 | 0.748836137 | 0.748836137 | 0.77487587 | 0.630586428 | 1 | 0.774875875 | 0.774875875 | 0.774875875 | 0.966394956 | -0.014845345 | FALSE | TRUE | FALSE |
| KRTAP12-3 | 0.7485213 | 0.6280698 | 1 | 0.748521261 | 0.748521261 | 0.748521261 | 0.77447669 | 0.630676267 | 1 | 0.774476688 | 0.774476688 | 0.774476688 | 0.966486497 | -0.014804209 | FALSE | TRUE | FALSE |
| YIPF5 | 0.748017 | 0.62815367 | 1 | 0.748017043 | 0.748017043 | 0.748017043 | 0.77427211 | 0.630855804 | 1 | 0.774272114 | 0.774272114 | 0.774272114 | 0.966090641 | -0.014982125 | FALSE | TRUE | FALSE |
| C4orf46 | 0.7479882 | 0.62823749 | 1 | 0.747988243 | 0.747988243 | 0.747988243 | 0.77397485 | 0.630945503 | 1 | 0.773974847 | 0.773974847 | 0.773974847 | 0.966424485 | -0.014832075 | FALSE | TRUE | FALSE |
| LCE3C | 0.7472059 | 0.62832129 | 1 | 0.74720594 | 0.74720594 | 0.74720594 | 0.77294255 | 0.631124779 | 1 | 0.772942551 | 0.772942551 | 0.772942551 | 0.966703074 | -0.0147069 | FALSE | TRUE | FALSE |
| CDKN2C | 0.7469053 | 0.62840505 | 1 | 0.746905339 | 0.746905339 | 0.746905339 | 0.77323165 | 0.631035163 | 1 | 0.773231649 | 0.773231649 | 0.773231649 | 0.965952881 | -0.015044058 | FALSE | TRUE | FALSE |
| HOGA1 | 0.7468865 | 0.62848876 | 0.039 | 0.18680384 | 0.746886463 | 2.986976852 | 0.00592488 | 0.897921811 | 1 | 0.094624268 | 0.378291815 | 0.378291815 | 126.0594036 | 2.100575248 | FALSE | TRUE | FALSE |
| CNPY3 | 0.7466248 | 0.62857244 | 1 | 0.746624814 | 0.746624814 | 0.746624814 | 0.7488039 | 0.641976172 | 1 | 7.083026777 | 7.083026777 | 7.083026777 | 0.997089918 | -0.001265675 | FALSE | TRUE | FALSE |
| CCDC24 | 0.7466062 | 0.62865607 | 0.25585 | 0.081351442 | 0.74660616 | 0.74660616 | 0.01254026 | 0.895880339 | 0.0069 | 0.012540255 | 0.012540255 | 0.012540255 | 59.53675924 | 1.774785191 | FALSE | TRUE | FALSE |
| PGP | 0.7462943 | 0.62873967 | 1 | 0.746294349 | 0.746294349 | 0.746294349 | 0.77265396 | 0.631303867 | 1 | 0.772653962 | 0.772653962 | 0.772653962 | 0.965884323 | -0.015074883 | FALSE | TRUE | FALSE |
| RAB11A | 0.7454895 | 0.62882323 | 1 | 0.745489469 | 0.745489469 | 0.745489469 | 0.77201297 | 0.631482775 | 1 | 0.772012967 | 0.772012967 | 0.772012967 | 0.965643715 | -0.015183082 | FALSE | TRUE | FALSE |
| KCNMB4 | 0.7450696 | 0.62890676 | 1 | 0.745069626 | 0.745069626 | 0.745069626 | 0.760346 | 0.636071427 | 1 | 0.760346002 | 0.760346002 | 0.760346002 | 0.979908653 | -0.008814407 | FALSE | TRUE | FALSE |
| TAF12 | 0.7448022 | 0.62899025 | 1 | 0.744802169 | 0.744802169 | 0.744802169 | 0.77120756 | 0.6317508 | 1 | 0.771207556 | 0.771207556 | 0.771207556 | 0.965760986 | -0.015130343 | FALSE | TRUE | FALSE |
| NUCKS1 | 0.7443727 | 0.6290737 | 1 | 0.744372728 | 0.744372728 | 0.744372728 | 0.77090626 | 0.631840053 | 1 | 0.770906263 | 0.770906263 | 0.770906263 | 0.965581373 | -0.015211121 | FALSE | TRUE | FALSE |
| MT4 | 0.7443414 | 0.62915711 | 1 | 0.744341404 | 0.744341404 | 0.744341404 | 0.77057227 | 0.632018424 | 1 | 0.770572268 | 0.770572268 | 0.770572268 | 0.965959242 | -0.015041198 | FALSE | TRUE | FALSE |
| VAMP4 | 0.7441158 | 0.62924049 | 1 | 0.744115783 | 0.744115783 | 0.744115783 | 0.77037047 | 0.632196607 | 1 | 0.770370469 | 0.770370469 | 0.770370469 | 0.965919402 | -0.01505911 | FALSE | TRUE | FALSE |
| SAA4 | 0.7436798 | 0.62932382 | 1 | 0.743679817 | 0.743679817 | 0.743679817 | 0.75259857 | 0.640370515 | 1 | 0.752598573 | 0.752598573 | 0.752598573 | 0.988149385 | -0.005177395 | FALSE | TRUE | FALSE |
| IL17A | 0.7436753 | 0.62940711 | 1 | 0.743675283 | 0.743675283 | 0.743675283 | 0.77030754 | 0.632285628 | 1 | 0.770307538 | 0.770307538 | 0.770307538 | 0.965426465 | -0.0152808 | FALSE | TRUE | FALSE |
| ZNF622 | 0.7431827 | 0.62949037 | 0.51185 | 0.743182696 | 0.743182696 | 3.671744779 | 0.22887838 | 0.845187655 | 0.0142 | 0.228878382 | 0.228878382 | 4.529164261 | 3.247063738 | 0.511490814 | FALSE | TRUE | FALSE |
| OXA1L | 0.7430483 | 0.62957359 | 0.50565 | 0.743048303 | 0.743048303 | 2.806176499 | 0.1104086 | 0.873218314 | 1 | 0.996401363 | 0.996401363 | 3.740324426 | 6.729985965 | 0.828014159 | FALSE | TRUE | FALSE |
| LCE2A | 0.7429272 | 0.62965677 | 1 | 0.742927201 | 0.742927201 | 0.742927201 | 0.7454691 | 0.643816407 | 1 | 0.745469105 | 7.03502566 | 7.03502566 | 0.996590195 | -0.00148339 | FALSE | TRUE | FALSE |
| KRTDAP | 0.7424015 | 0.62973991 | 1 | 0.742401457 | 0.742401457 | 0.742401457 | 0.76893353 | 0.632818794 | 1 | 0.76893353 | 0.76893353 | 0.76893353 | 0.965494972 | -0.015249984 | FALSE | TRUE | FALSE |
| DEFB114 | 0.742366 | 0.62982302 | 1 | 0.742365996 | 0.742365996 | 0.742365996 | 0.76881186 | 0.632907497 | 1 | 0.768811858 | 0.768811858 | 0.768811858 | 0.965601646 | -0.015202002 | FALSE | TRUE | FALSE |
| ZNF619 | 0.7423591 | 0.62990608 | 0.49825 | 0.082627256 | 0.742359134 | 1.637577081 | 0.11354501 | 0.872659226 | 1 | 0.113545011 | 0.113545011 | 2.252782891 | 6.538016317 | 0.815446 | FALSE | TRUE | FALSE |
| CTNNBIP1 | 0.7422136 | 0.6299891 | 1 | 0.742213573 | 0.742213573 | 0.742213573 | 0.76870755 | 0.632996154 | 0.0038 | 0.768707549 | 0.768707549 | 0.768707549 | 0.965534388 | -0.015232254 | FALSE | TRUE | FALSE |
| OR2T8 | 0.7412725 | 0.63007209 | 1 | 1.58563E+11 | 2.9978E+14 | 5.04517E+16 | 415.297062 | 0.005844375 | 1 | 1.12717E+21 | 7.46118E+26 | 2.80743E+30 | 0.001784921 | -2.748380994 | FALSE | FALSE | FALSE |
| YWHAG | 0.740997 | 0.63015504 | 1 | 0.740996993 | 0.740996993 | 0.740996993 | 0.76794844 | 0.633084773 | 1 | 0.767948437 | 0.767948437 | 0.767948437 | 0.964904617 | -0.015515615 | FALSE | TRUE | FALSE |
| AP2S1 | 0.7406871 | 0.63023796 | 1 | 0.740687144 | 0.740687144 | 0.740687144 | 0.76750914 | 0.63317335 | 1 | 0.767509136 | 0.767509136 | 0.767509136 | 0.965053195 | -0.015448747 | FALSE | TRUE | FALSE |

| | | | | | | | | | | | | | | | | | |
|---|---|---|---|---|---|---|---|---|---|---|---|---|---|---|---|---|---|
| UBE2L3 | 0.7403842 | 0.63032083 | 1 | 0.740384231 | 0.740384231 | 0.740384231 | 0.75294974 | 0.640115611 | 1 | 0.752949743 | 0.752949743 | 0.752949743 | 0.98331162 | -0.007308829 | FALSE | TRUE | FALSE |
| CDC20 | 0.7402599 | 0.63040367 | 0.50705 | 0.740259867 | 0.740259867 | 2.789212751 | 0.1132853 | 0.872689836 | 1 | 0.113285298 | 0.113285298 | 2.242077303 | 6.534474303 | 0.815210654 | FALSE | TRUE | FALSE |
| SPRR2E | 0.7399825 | 0.63048647 | 1 | 0.739982479 | 0.739982479 | 0.739982479 | 0.76659931 | 0.63343882 | 1 | 0.766599307 | 0.766599307 | 0.766599307 | 0.965279348 | -0.015346985 | FALSE | TRUE | FALSE |
| SNUPN | 0.7398622 | 0.63056923 | 0.9375 | 27.28992495 | 625.8386536 | 2503.060705 | 1.03589038 | 0.584529734 | 0.8788 | 9.622013057 | 91.67090081 | 91.67090081 | 0.714228308 | -0.146162941 | FALSE | FALSE | FALSE |
| KRTAP20-2 | 0.7393857 | 0.63065195 | 0.50275 | 6.972304239 | 6.972304239 | 6.972304239 | 0.79064404 | 0.623620974 | 1 | 0.790644042 | 0.790644042 | 0.790644042 | 0.935168848 | -0.029109969 | FALSE | TRUE | FALSE |
| IL1RN | 0.7392162 | 0.63073464 | 1 | 0.739216154 | 0.739216154 | 0.739216154 | 0.76583052 | 0.633792158 | 1 | 0.765830523 | 0.765830523 | 0.765830523 | 0.965247704 | -0.015361223 | FALSE | TRUE | FALSE |
| SPACA1 | 0.7387378 | 0.63081729 | 1 | 0.738737793 | 0.738737793 | 0.738737793 | 0.76576465 | 0.633880376 | 1 | 0.765764654 | 0.765764654 | 0.765764654 | 0.964706048 | -0.015604999 | FALSE | TRUE | FALSE |
| RPL22 | 0.7385463 | 0.6308999 | 1 | 0.738546256 | 0.738546256 | 0.738546256 | 0.7654284 | 0.634056677 | 1 | 0.765428405 | 0.765428405 | 0.765428405 | 0.964879603 | -0.015526874 | FALSE | TRUE | FALSE |
| UBA52 | 0.7385411 | 0.63098247 | 1 | 0.73854106 | 0.73854106 | 0.73854106 | 0.7655996 | 0.633968549 | 1 | 0.765599596 | 0.765599596 | 0.765599596 | 0.964657066 | -0.01562705 | FALSE | TRUE | FALSE |
| PATE2 | 0.7382469 | 0.631065 | 0.5071 | 0.73824687 | 0.73824687 | 0.73824687 | 0.22450592 | 0.846304433 | 1 | 0.897950545 | 0.897950545 | 0.897950545 | 3.288318022 | 0.516973813 | FALSE | TRUE | FALSE |
| CBX3 | 0.7379371 | 0.6311475 | 1 | 0.737937107 | 0.737937107 | 0.737937107 | 0.74116831 | 0.645883377 | 1 | 7.012246437 | 7.012246437 | 7.012246437 | 0.995640388 | -0.001897495 | FALSE | TRUE | FALSE |
| SNRPN | 0.7375535 | 0.63122996 | 1 | 0.737553452 | 0.737553452 | 0.737553452 | 0.76472393 | 0.634144766 | 1 | 0.764723927 | 0.764723927 | 0.764723927 | 0.964470218 | -0.015711179 | FALSE | TRUE | FALSE |
| TMIGD1 | 0.7374331 | 0.63131238 | 0.50635 | 1.694202883 | 6.775942012 | 6.775942012 | 2.07119441 | 0.483376554 | 0.50235 | 2.071194415 | 8.283839979 | 8.283839979 | 0.356042445 | -0.448498225 | FALSE | FALSE | FALSE |
| ZNF428 | 0.7373816 | 0.63139476 | 1 | 0.737381609 | 0.737381609 | 0.737381609 | 0.75050657 | 0.641132981 | 1 | 0.750506572 | 0.750506572 | 0.750506572 | 0.982511862 | -0.007662198 | FALSE | TRUE | FALSE |
| PRSS23 | 0.7373685 | 0.63147709 | 0.50035 | 0.73736845 | 0.73736845 | 1.60118585 | 81.897256 | 0.035375175 | 0.06875 | 8.498399427 | 808.9830071 | 808.9830071 | 0.009003579 | -2.045584799 | FALSE | FALSE | FALSE |
| HEMGN | 0.7372563 | 0.63155939 | 0.75245 | 0.737256337 | 6.739742953 | 6.739742953 | 0.11686426 | 0.87200304 | 0.00925 | 0.116864256 | 0.116864256 | 0.116864256 | 6.308655526 | 0.799936814 | FALSE | TRUE | FALSE |
| PSMB2 | 0.7369347 | 0.63164165 | 1 | 0.736934684 | 0.736934684 | 0.736934684 | 0.76401908 | 0.634320813 | 1 | 0.764019084 | 0.764019084 | 0.764019084 | 0.9645501 | -0.01567521 | FALSE | TRUE | FALSE |
| TBPL1 | 0.7368263 | 0.63172388 | 1 | 0.736826268 | 0.736826268 | 0.736826268 | 0.76392033 | 0.63440877 | 1 | 0.763920328 | 0.763920328 | 0.763920328 | 0.964532873 | -0.015682966 | FALSE | TRUE | FALSE |
| ZNF140 | 0.7367729 | 0.63180606 | 0.50365 | 0.73677293 | 0.73677293 | 1.628881571 | 0.10953285 | 0.873491127 | 1 | 0.989118894 | 0.989118894 | 2.174846979 | 6.726501988 | 0.827789275 | FALSE | TRUE | FALSE |
| C1orf115 | 0.7367673 | 0.6318882 | 1 | 0.736767288 | 0.736767288 | 0.736767288 | 0.76373408 | 0.634496683 | 1 | 0.763734081 | 0.763734081 | 0.763734081 | 0.96469086 | -0.015611836 | FALSE | TRUE | FALSE |
| ISY1 | 0.7367258 | 0.6319703 | 1 | 0.736725848 | 0.736725848 | 0.736725848 | 0.76165086 | 0.635722727 | 1 | 0.76165086 | 0.76165086 | 0.76165086 | 0.967275017 | -0.014450029 | FALSE | TRUE | FALSE |
| UBE2F | 0.7366526 | 0.63205236 | 1 | 0.73665256 | 0.73665256 | 0.73665256 | 0.76005797 | 0.636506297 | 1 | 0.760057972 | 0.760057972 | 0.760057972 | 0.969205754 | -0.013584016 | FALSE | TRUE | FALSE |
| S100A12 | 0.7364566 | 0.63213438 | 1 | 0.73645662 | 0.73645662 | 0.73645662 | 0.7633862 | 0.634584553 | 1 | 0.7633862 | 0.7633862 | 0.7633862 | 0.964723518 | -0.015597134 | FALSE | TRUE | FALSE |
| EMC8 | 0.7361586 | 0.63221637 | 1 | 0.736158632 | 0.736158632 | 0.736158632 | 0.7633759 | 0.634672376 | 1 | 0.763375902 | 0.763375902 | 0.763375902 | 0.964346176 | -0.015767037 | FALSE | TRUE | FALSE |
| PSMD14 | 0.7359739 | 0.63229832 | 1 | 0.735973919 | 0.735973919 | 0.735973919 | 0.76327871 | 0.634760154 | 1 | 0.763278706 | 0.763278706 | 0.763278706 | 0.964226976 | -0.015820722 | FALSE | TRUE | FALSE |
| MED30 | 0.7358427 | 0.63238022 | 1 | 0.735842705 | 0.735842705 | 0.735842705 | 0.76305077 | 0.634847887 | 1 | 0.763050771 | 0.763050771 | 0.763050771 | 0.964343047 | -0.015768447 | FALSE | TRUE | FALSE |
| SEC61B | 0.7356755 | 0.63246209 | 1 | 0.735675537 | 0.735675537 | 0.735675537 | 0.76278446 | 0.635023218 | 1 | 0.762784456 | 0.762784456 | 0.762784456 | 0.964460578 | -0.015715519 | FALSE | TRUE | FALSE |
| GABARAPL1 | 0.7350601 | 0.63254393 | 1 | 0.735060125 | 0.735060125 | 0.735060125 | 0.76231852 | 0.63537333 | 0.0025 | 0.762318518 | 0.762318518 | 0.762318518 | 0.964242777 | -0.015813606 | FALSE | TRUE | FALSE |
| UBL5 | 0.7347975 | 0.63262573 | 1 | 0.734797505 | 0.734797505 | 0.734797505 | 0.76192818 | 0.63554812 | 1 | 0.761928176 | 0.761928176 | 0.761928176 | 0.964392089 | -0.015746361 | FALSE | TRUE | FALSE |
| IFITM1 | 0.7346323 | 0.63270749 | 1 | 0.734632301 | 0.734632301 | 0.734632301 | 0.76195505 | 0.635460748 | 1 | 0.761955051 | 0.761955051 | 0.761955051 | 0.964141257 | -0.015859333 | FALSE | TRUE | FALSE |
| VSNL1 | 0.7344937 | 0.63278921 | 1 | 0.734493652 | 0.734493652 | 0.734493652 | 0.76190849 | 0.635635445 | 1 | 0.761908488 | 0.761908488 | 0.761908488 | 0.964018203 | -0.015914765 | FALSE | TRUE | FALSE |
| FBXO45 | 0.7344919 | 0.63287089 | 1 | 0.734491926 | 0.734491926 | 0.734491926 | 0.75854461 | 0.637717952 | 1 | 0.758544606 | 0.758544606 | 0.758544606 | 0.968291015 | -0.013994098 | FALSE | TRUE | FALSE |
| GIPC2 | 0.7339315 | 0.63295254 | 0.50725 | 0.733931453 | 2.935183094 | 2.935183094 | 0.44805005 | 0.774974447 | 0.5085 | 0.448050049 | 0.448050049 | 0.448050049 | 1.638056854 | 0.214328971 | FALSE | TRUE | FALSE |
| RPL27 | 0.7338011 | 0.63303415 | 1 | 0.733801139 | 0.733801139 | 0.733801139 | 0.76126755 | 0.635897159 | 0.0035 | 0.76126755 | 0.76126755 | 0.76126755 | 0.96392016 | -0.015958937 | FALSE | TRUE | FALSE |
| LRRC39 | 0.7335979 | 0.63311572 | 0.87625 | 6.771862569 | 64.11746595 | 64.11746595 | 0.11926249 | 0.871402927 | 1 | 0.119262491 | 0.119262491 | 0.119262491 | 6.151120217 | 0.788954215 | FALSE | TRUE | FALSE |
| CFD | 0.733177 | 0.63319725 | 1 | 4.03747E+25 | 3.7959E+32 | 3.78016E+35 | 0.00893421 | 0.897035142 | 1 | 5.23772E+48 | 7.04309E+58 | 7.13387E+62 | 82.06399499 | 1.914152655 | FALSE | TRUE | FALSE |
| TMEM89 | 0.7327377 | 0.63327876 | 1 | 0.73273773 | 0.73273773 | 0.73273773 | 0.76015953 | 0.636419414 | 1 | 0.76015953 | 0.76015953 | 0.76015953 | 0.963926256 | -0.01595619 | FALSE | TRUE | FALSE |
| LIN28B | 0.7326945 | 0.63336022 | 1 | 0.732694481 | 0.732694481 | 0.732694481 | 0.76023357 | 0.636332486 | 1 | 0.760233571 | 0.760233571 | 0.760233571 | 0.963775488 | -0.016024123 | FALSE | TRUE | FALSE |
| ZFAND3 | 0.7326454 | 0.63344164 | 1 | 0.732645441 | 0.732645441 | 0.732645441 | 0.75413869 | 0.639604631 | 1 | 0.754138689 | 0.754138689 | 0.754138689 | 0.971499608 | -0.01255737 | FALSE | TRUE | FALSE |
| PSMA3 | 0.7319474 | 0.63352303 | 1 | 0.731947354 | 0.731947354 | 0.731947354 | 0.75953378 | 0.636853383 | 1 | 0.759533775 | 0.759533775 | 0.759533775 | 0.963679796 | -0.016067246 | FALSE | TRUE | FALSE |
| CTLA4 | 0.7319323 | 0.63360438 | 1 | 6.921734004 | 6.921734004 | 6.921734004 | 0.75946942 | 0.636940041 | 1 | 0.759469416 | 7.185916906 | 7.185916906 | 0.963741697 | -0.01603935 | FALSE | TRUE | FALSE |
| PHF13 | 0.7318278 | 0.63368569 | 1 | 0.731827785 | 0.731827785 | 0.731827785 | 0.75500912 | 0.639177658 | 0.0103 | 0.755009124 | 0.755009124 | 14.99221494 | 0.969296612 | -0.013543305 | FALSE | TRUE | FALSE |
| CCL24 | 0.7317954 | 0.63376696 | 1 | 0.731795442 | 0.731795442 | 0.731795442 | 0.75904969 | 0.637372665 | 1 | 0.759049694 | 0.759049694 | 0.759049694 | 0.964094245 | -0.01588051 | FALSE | TRUE | FALSE |
| TMEM160 | 0.7315493 | 0.63384819 | 1 | 6.896154398 | 6.896154398 | 6.896154398 | 0.78324459 | 0.626954685 | 1 | 0.783244589 | 0.783244589 | 0.783244589 | 0.933998492 | -0.029653825 | FALSE | TRUE | FALSE |
| CFL1 | 0.7314009 | 0.63392939 | 1 | 0.731400863 | 0.731400863 | 0.731400863 | 0.75900354 | 0.637459053 | 1 | 0.759003539 | 0.759003539 | 0.759003539 | 0.963633007 | -0.016088333 | FALSE | TRUE | FALSE |
| VWC2 | 0.7312086 | 0.63401054 | 1 | 0.731208647 | 0.731208647 | 0.731208647 | 0.7588901 | 0.637545397 | 1 | 0.758890099 | 0.758890099 | 0.758890099 | 0.963523767 | -0.016137568 | FALSE | TRUE | FALSE |
| RPL34 | 0.7311535 | 0.63409166 | 1 | 0.731153492 | 0.731153492 | 0.731153492 | 0.75872642 | 0.637631696 | 1 | 0.758726425 | 0.758726425 | 0.758726425 | 0.963658927 | -0.016076651 | FALSE | TRUE | FALSE |
| AP1B1 | 0.731059 | 0.63417274 | 0.7587 | 0.731059024 | 2.92452168 | 24.70155065 | 0.22844997 | 0.84527639 | 1 | 0.228449969 | 0.228449969 | 4.479214239 | 3.200083714 | 0.50516134 | FALSE | TRUE | FALSE |
| ARL4C | 0.7307768 | 0.63425378 | 1 | 0.730776841 | 0.730776841 | 0.730776841 | 0.75852397 | 0.637804163 | 1 | 0.758523966 | 0.758523966 | 0.758523966 | 0.96341958 | -0.016184531 | FALSE | TRUE | FALSE |
| ISCA1 | 0.7306125 | 0.63433479 | 1 | 0.730612479 | 0.730612479 | 0.730612479 | 0.75820549 | 0.638062529 | 1 | 0.758205486 | 0.758205486 | 0.758205486 | 0.963607481 | -0.016099837 | FALSE | TRUE | FALSE |
| APH1A | 0.7305336 | 0.63441575 | 1 | 0.730533589 | 0.730533589 | 0.730533589 | 0.75825051 | 0.637976452 | 0.00615 | 0.75825051 | 0.75825051 | 0.75825051 | 0.963446221 | -0.016172522 | FALSE | TRUE | FALSE |
| UBE2Z | 0.7305197 | 0.63449668 | 1 | 0.730519651 | 0.730519651 | 0.730519651 | 0.75834801 | 0.63789033 | 1 | 0.758348007 | 0.758348007 | 0.758348007 | 0.963303977 | -0.016236647 | FALSE | TRUE | FALSE |
| PSMB7 | 0.7301519 | 0.63457757 | 1 | 0.730151927 | 0.730151927 | 0.730151927 | 0.75799003 | 0.638148563 | 1 | 0.757990028 | 0.757990028 | 0.757990028 | 0.96327379 | -0.016250256 | FALSE | TRUE | FALSE |
| CYS1 | 0.7297355 | 0.63465843 | 1 | 0.729735466 | 0.729735466 | 0.729735466 | 0.75725223 | 0.638492258 | 1 | 0.757252234 | 0.757252234 | 0.757252234 | 0.963662348 | -0.016075109 | FALSE | TRUE | FALSE |
| IFNA1 | 0.7296674 | 0.63473924 | 1 | 0.72966745 | 0.72966745 | 0.72966745 | 0.74000018 | 0.646457513 | 1 | 0.740000177 | 0.740000177 | 0.740000177 | 0.986036859 | -0.006106851 | FALSE | TRUE | FALSE |
| PTPN5 | 0.7290193 | 0.63482003 | 0.87675 | 2.916379131 | 11.66964024 | 24.67935678 | 0.23022545 | 0.844816863 | 1 | 0.230225455 | 0.230225455 | 4.552219476 | 3.166545189 | 0.50058569 | FALSE | TRUE | FALSE |
| HSPE1 | 0.7285026 | 0.63490078 | 1 | 0.728502616 | 0.728502616 | 0.728502616 | 0.7405659 | 0.646293676 | 1 | 0.740565903 | 0.740565903 | 0.740565903 | 0.983710718 | -0.007132597 | FALSE | TRUE | FALSE |
| CXCL14 | 0.7282988 | 0.63498149 | 1 | 0.728298827 | 0.728298827 | 0.728298827 | 0.75581856 | 0.638835296 | 1 | 0.755818559 | 0.755818559 | 0.755818559 | 0.9635895 | -0.016107941 | FALSE | TRUE | FALSE |
| S100A2 | 0.7282457 | 0.63506217 | 0.495 | 0.728245702 | 6.86645102 | 6.86645102 | 66.8238189 | 0.042556096 | 0.00135 | 6.905782942 | 66.82381891 | 66.82381891 | 0.010897996 | -1.962653362 | FALSE | FALSE | FALSE |
| LAX1 | 0.7281696 | 0.6351428 | 0.50645 | 0.728169607 | 0.728169607 | 1.624602423 | 0.11225662 | 0.872852865 | 1 | 0.112256618 | 0.112256618 | 0.133438072 | 6.486651942 | 0.812020595 | FALSE | TRUE | FALSE |
| ELOVL1 | 0.7279749 | 0.6352234 | 1 | 0.727974897 | 0.727974897 | 0.727974897 | 0.7559812 | 0.638749599 | 1 | 0.755981197 | 0.755981197 | 0.755981197 | 0.962953708 | -0.01639459 | FALSE | TRUE | FALSE |
| FIGN | 0.7279595 | 0.63530396 | 0.32025 | 0.182134189 | 2.910257518 | 3.587592524 | 0.02027893 | 0.893755229 | 1 | 0.02027893 | 0.02027893 | 0.400171844 | 35.89733009 | 1.555062149 | FALSE | TRUE | FALSE |
| TEX2 | 0.7278432 | 0.63538448 | 0.5155 | 0.727843183 | 0.727843183 | 14.23897293 | 0.10768965 | 0.873843125 | 1 | 0.981080446 | 0.981080446 | 19.24360442 | 6.758710743 | 0.82986386 | FALSE | TRUE | FALSE |
| GUCA2B | 0.7271479 | 0.63546497 | 1 | 0.727147876 | 0.727147876 | 0.727147876 | 0.75500303 | 0.639263139 | 1 | 0.755003035 | 0.755003035 | 0.755003035 | 0.963105898 | -0.016325958 | FALSE | TRUE | FALSE |
| RPS3A | 0.7271049 | 0.63554542 | 0.0087 | 0.727104883 | 0.727104883 | 0.727104883 | 0.74857494 | 0.642144301 | 1 | 0.748574941 | 0.748574941 | 0.748574941 | 0.971318758 | -0.012638224 | FALSE | TRUE | FALSE |
| WBP1 | 0.7270362 | 0.63562584 | 0.8773 | 2.776625514 | 61.96545557 | 61.96545557 | 1.03629912 | 0.584303656 | 0.5101 | 1.036299118 | 1.036299118 | 1.036299118 | 0.701569818 | -0.153929103 | FALSE | FALSE | FALSE |
| VDAC1 | 0.7268961 | 0.63570621 | 1 | 0.726896106 | 0.726896106 | 0.726896106 | 0.75499134 | 0.639348574 | 0.00705 | 0.754991341 | 0.754991341 | 0.754991341 | 0.962787342 | -0.016469628 | FALSE | TRUE | FALSE |
| ZNF740 | 0.7263405 | 0.63578655 | 1 | 0.726340535 | 0.726340535 | 0.726340535 | 0.7543235 | 0.63951932 | 1 | 0.754323496 | 0.754323496 | 0.754323496 | 0.962903236 | -0.016417354 | FALSE | TRUE | FALSE |
| RAB43 | 0.7257213 | 0.63586686 | 1 | 2.902734453 | 46.47375716 | 46.47375716 | 0.76240585 | 0.635285869 | 1 | 3.049494918 | 12.20051534 | 12.20051534 | 0.95188323 | -0.021416324 | FALSE | TRUE | FALSE |
| ZNF577 | 0.7253737 | 0.63594714 | 0.939 | 6.703557218 | 617.410412 | 617.410412 | 86.5846583 | 0.033845432 | 0.3192 | 9.079370244 | 846.384433 | 846.384433 | 0.008377623 | -2.076879163 | FALSE | FALSE | FALSE |
| CXCL1 | 0.7252505 | 0.63602737 | 1 | 0.72525049 | 0.72525049 | 0.72525049 | 0.75312385 | 0.639945455 | 1 | 0.753123851 | 0.753123851 | 0.753123851 | 0.962989672 | -0.016378371 | FALSE | TRUE | FALSE |
| PGF | 0.7247903 | 0.63610758 | 1 | 0.724790258 | 0.724790258 | 0.724790258 | 0.75297532 | 0.640030556 | 1 | 0.752975323 | 0.752975323 | 0.752975323 | 0.962568407 | -0.016568397 | FALSE | TRUE | FALSE |
| B3GNT2 | 0.7246523 | 0.63618774 | 0.0104 | 0.724652337 | 0.724652337 | 14.37806901 | 0.7432307 | 0.644480625 | 1 | 7.049879294 | 7.049879294 | 7.126988077 | 0.97500324 | -0.010993941 | FALSE | TRUE | FALSE |

| | | | | | | | | | | | | | | | | |
|---|---|---|---|---|---|---|---|---|---|---|---|---|---|---|---|---|
| KNCN | 0.7241049 | 0.63626787 | 1 | 0.724104895 | 0.724104895 | 0.724104895 | 0.72878583 | 0.652329121 | 1 | 6.890682989 | 6.890682989 | 6.890682989 | 0.993577078 | -0.002798436 | FALSE | TRUE | FALSE |
| FCGR2B | 0.7239579 | 0.63634797 | 1 | 0.723957919 | 0.723957919 | 0.723957919 | 0.75220832 | 0.64054024 | 1 | 0.752208316 | 0.752208316 | 0.752208316 | 0.962443386 | -0.016624808 | FALSE | TRUE | FALSE |
| RPS19 | 0.7236188 | 0.63642803 | 1 | 0.723618755 | 0.723618755 | 0.723618755 | 0.75190239 | 0.64062504 | 1 | 0.751902394 | 0.751902394 | 0.751902394 | 0.962383895 | -0.016651653 | FALSE | TRUE | FALSE |
| SMAGP | 0.7228397 | 0.63650806 | 1 | 0.722839732 | 0.722839732 | 0.722839732 | 0.75104155 | 0.640794521 | 1 | 0.751041552 | 0.751041552 | 0.751041552 | 0.962449721 | -0.016621949 | FALSE | TRUE | FALSE |
| CCL13 | 0.7227699 | 0.63658805 | 1 | 0.722769875 | 0.722769875 | 0.722769875 | 0.75070749 | 0.640963838 | 1 | 0.750707489 | 0.750707489 | 0.750707489 | 0.962784954 | -0.016470705 | FALSE | TRUE | FALSE |
| TMEM65 | 0.722532 | 0.636668 | 1 | 0.722531963 | 0.722531963 | 0.722531963 | 0.75083486 | 0.640879201 | 1 | 0.750834855 | 0.750834855 | 0.750834855 | 0.962304771 | -0.016687361 | FALSE | TRUE | FALSE |
| CHMP3 | 0.7224118 | 0.63674792 | 1 | 0.722411814 | 0.722411814 | 0.722411814 | 0.74974068 | 0.641386378 | 1 | 0.749740685 | 0.749740685 | 0.749740685 | 0.963548903 | -0.016126239 | FALSE | TRUE | FALSE |
| ELAVL1 | 0.7222525 | 0.6368278 | 1 | 0.722252461 | 0.722252461 | 14.31182553 | 0.74663362 | 0.643066031 | 1 | 7.082506 | 7.082506 | 14.81136347 | 0.967345217 | -0.014418511 | FALSE | TRUE | FALSE |
| S100P | 0.7222441 | 0.63690764 | 1 | 0.722244062 | 0.722244062 | 0.722244062 | 0.75039844 | 0.641217488 | 1 | 0.750398439 | 0.750398439 | 0.750398439 | 0.962480763 | -0.016607942 | FALSE | TRUE | FALSE |
| CPLX2 | 0.7220978 | 0.63698745 | 1 | 0.722097826 | 0.722097826 | 0.722097826 | 0.73676521 | 0.648087109 | 1 | 0.736765209 | 0.736765209 | 0.736765209 | 0.980092189 | -0.008733072 | FALSE | TRUE | FALSE |
| CPLX1 | 0.722056 | 0.63706721 | 1 | 0.722056005 | 0.722056005 | 0.722056005 | 0.74702297 | 0.642898825 | 1 | 0.747022966 | 0.747022966 | 0.747022966 | 0.966578053 | -0.01476307 | FALSE | TRUE | FALSE |
| ASB3 | 0.7217047 | 0.63714694 | 1 | 0.721704656 | 0.721704656 | 14.31568387 | 0.75026853 | 0.641301952 | 1 | 0.75026853 | 0.75026853 | 0.75026853 | 0.961928466 | -0.016857223 | FALSE | TRUE | FALSE |
| PCBP2 | 0.7213022 | 0.63722664 | 1 | 0.72130218 | 0.72130218 | 14.31002137 | 0.74938439 | 0.641555104 | 1 | 0.749384391 | 0.749384391 | 0.749384391 | 0.962526293 | -0.016587398 | FALSE | TRUE | FALSE |
| MAF1 | 0.7212488 | 0.6373063 | 1 | 0.72124881 | 0.72124881 | 0.72124881 | 0.72628946 | 0.653979814 | 1 | 0.726289459 | 6.875511285 | 6.875511285 | 0.993059725 | -0.003024631 | FALSE | TRUE | FALSE |
| KCNK12 | 0.7211221 | 0.63738592 | 1 | 0.721122142 | 0.721122142 | 0.721122142 | 0.74771961 | 0.642396189 | 1 | 0.747719613 | 0.747719613 | 0.747719613 | 0.96442855 | -0.015729942 | FALSE | TRUE | FALSE |
| DESI2 | 0.720716 | 0.63746551 | 1 | 2.882752442 | 2.882752442 | 2.882752442 | 0.75770714 | 0.638406398 | 1 | 0.757707135 | 0.757707135 | 0.757707135 | 0.95118019 | -0.021737203 | FALSE | TRUE | FALSE |
| CD58 | 0.7205273 | 0.63754506 | 1 | 0.720527291 | 0.720527291 | 0.720527291 | 0.74898018 | 0.641892044 | 1 | 0.748980176 | 0.748980176 | 0.748980176 | 0.962011163 | -0.016819888 | FALSE | TRUE | FALSE |
| CRYAA | 0.7199081 | 0.63762458 | 1 | 0.719908116 | 0.719908116 | 0.719908116 | 0.76282157 | 0.634935575 | 1 | 7.230677475 | 70.342356 | 70.342356 | 0.94374378 | -0.025145898 | FALSE | TRUE | FALSE |
| RPAP2 | 0.7196693 | 0.63770406 | 0.8812 | 6.621460459 | 62.48762614 | 62.48762614 | 0.11747885 | 0.871868989 | 1 | 0.117478849 | 0.117478849 | 2.330267174 | 6.125947637 | 0.78717328 | FALSE | TRUE | FALSE |
| UBE2E1 | 0.7193916 | 0.63778351 | 1 | 6.80942838 | 6.80942838 | 6.80942838 | 0.77252355 | 0.631393342 | 1 | 0.772523547 | 0.772523547 | 0.772523547 | 0.931222885 | -0.03094636 | FALSE | TRUE | FALSE |
| MAL2 | 0.718956 | 0.63786292 | 0.00465 | 0.718956009 | 0.718956009 | 0.718956009 | 0.7474653 | 0.642563904 | 1 | 0.747465298 | 0.747465298 | 0.747465298 | 0.961858712 | -0.016888717 | FALSE | TRUE | FALSE |
| RHBDF2 | 0.7188851 | 0.6379423 | 0.51405 | 0.718885106 | 0.718885106 | 14.04991764 | 0.10880771 | 0.873702537 | 0.0245 | 0.108807707 | 0.108807707 | 2.132382679 | 6.606931845 | 0.819999827 | FALSE | TRUE | FALSE |
| GABRD | 0.718867 | 0.63802164 | 0.75715 | 0.718866955 | 2.875193541 | 14.18744774 | 0.22531415 | 0.846055999 | 1 | 0.225314146 | 0.225314146 | 4.45383283 | 3.190509645 | 0.503860062 | FALSE | TRUE | FALSE |
| ORAI2 | 0.7187609 | 0.63810094 | 1 | 0.718760944 | 0.718760944 | 0.718760944 | 2.95761257 | 0.414126196 | 0.5036 | 2.957612573 | 2.957612573 | 2.957612573 | 0.243020655 | -0.614356813 | FALSE | FALSE | FALSE |
| CDC42 | 0.7185819 | 0.6381802 | 1 | 0.718581877 | 0.718581877 | 0.718581877 | 0.74732627 | 0.642647699 | 1 | 0.747326272 | 0.747326272 | 0.747326272 | 0.961537021 | -0.01703399 | FALSE | TRUE | FALSE |
| CYCS | 0.7183327 | 0.63825943 | 1 | 0.718332682 | 0.718332682 | 0.718332682 | 0.74701451 | 0.642982447 | 1 | 0.747014512 | 0.747014512 | 0.747014512 | 0.961604722 | -0.017003413 | FALSE | TRUE | FALSE |
| NPBWR1 | 0.7182731 | 0.63833862 | 0.8748 | 6.701519101 | 64.13142716 | 64.13142716 | 0.11635228 | 0.87211626 | 0.0141 | 0.116352284 | 0.116352284 | 2.301587692 | 6.173261619 | 0.790514682 | FALSE | TRUE | FALSE |
| TESC | 0.7177352 | 0.63841778 | 1 | 0.717735187 | 0.717735187 | 0.717735187 | 0.74646954 | 0.643149574 | 1 | 0.74646954 | 0.74646954 | 0.74646954 | 0.961506329 | -0.017047853 | FALSE | TRUE | FALSE |
| UBE2D1 | 0.7172505 | 0.6384969 | 1 | 0.717250453 | 0.717250453 | 0.717250453 | 0.74602936 | 0.643399951 | 1 | 0.746029361 | 0.746029361 | 0.746029361 | 0.961423894 | -0.017085089 | FALSE | TRUE | FALSE |
| LALBA | 0.7168951 | 0.63857599 | 1 | 0.716895145 | 0.716895145 | 0.716895145 | 0.7455585 | 0.643649952 | 1 | 0.745558502 | 0.745558502 | 0.745558502 | 0.961554517 | -0.017026088 | FALSE | TRUE | FALSE |
| SLC22A2 | 0.7167659 | 0.63865505 | 0.8556 | 11.45460507 | 1034.089268 | 5107.375369 | 2.27601103 | 0.466372435 | 0.74575 | 5.254654574 | 336.1535418 | 1661.835298 | 0.314921995 | -0.501797006 | FALSE | FALSE | FALSE |
| GRB2 | 0.7167087 | 0.63873406 | 1 | 0.716708704 | 0.716708704 | 0.716708704 | 0.74563943 | 0.64356666 | 1 | 0.745639428 | 0.745639428 | 0.745639428 | 0.961200115 | -0.017186186 | FALSE | TRUE | FALSE |
| NUPR1 | 0.7165763 | 0.63881304 | 1 | 6.756686789 | 6.756686789 | 6.756686789 | 0.7691476 | 0.632730044 | 1 | 0.769147595 | 0.769147595 | 0.769147595 | 0.931649976 | -0.030747223 | FALSE | TRUE | FALSE |
| RIPPLY2 | 0.7163916 | 0.63889199 | 0.00395 | 0.716391633 | 0.716391633 | 0.716391633 | 0.74503028 | 0.643982699 | 1 | 0.745030277 | 0.745030277 | 0.745030277 | 0.961560429 | -0.017023417 | FALSE | TRUE | FALSE |
| IGFBP2 | 0.7160903 | 0.63897089 | 1 | 0.716090316 | 0.716090316 | 0.716090316 | 0.74509884 | 0.643899575 | 1 | 0.745098839 | 0.745098839 | 0.745098839 | 0.961067551 | -0.017246086 | FALSE | TRUE | FALSE |
| INPP4B | 0.7154328 | 0.63904977 | 0.51195 | 0.715432791 | 0.715432791 | 14.07729679 | 0.22128979 | 0.846943871 | 1 | 0.221289788 | 0.221289788 | 4.362709492 | 3.233013136 | 0.509607469 | FALSE | TRUE | FALSE |
| APBA3 | 0.71508 | 0.63912862 | 0.87935 | 6.578420104 | 248.2777159 | 248.2777159 | 1.03626543 | 0.584416727 | 0.49875 | 0.114655499 | 1.036265426 | 2.273114671 | 0.690054872 | -0.161116373 | FALSE | FALSE | FALSE |
| CSNK1A1 | 0.715026 | 0.63920742 | 1 | 0.715025974 | 0.715025974 | 0.715025974 | 0.7441859 | 0.644231843 | 1 | 0.744185905 | 0.744185905 | 0.744185905 | 0.960816335 | -0.017359622 | FALSE | TRUE | FALSE |
| RPS8 | 0.7146646 | 0.6392862 | 1 | 0.714664583 | 0.714664583 | 0.714664583 | 0.74168835 | 0.645389654 | 1 | 0.741688348 | 0.741688348 | 0.741688348 | 0.963564527 | -0.016119197 | FALSE | TRUE | FALSE |
| PFAS | 0.7146452 | 0.63936493 | 0.1405 | 0.080844665 | 0.714645208 | 2.721120431 | 0.00260339 | 0.898841737 | 1 | 0.010408651 | 0.010408651 | 0.202185986 | 274.5053872 | 2.438550872 | FALSE | TRUE | FALSE |
| SMIM7 | 0.7145917 | 0.63944363 | 0.0027 | 0.714591662 | 0.714591662 | 0.714591662 | 0.74313914 | 0.644563471 | 1 | 0.743139137 | 0.743139137 | 0.743139137 | 0.961585289 | -0.017012189 | FALSE | TRUE | FALSE |
| IL23A | 0.7141325 | 0.63952229 | 1 | 0.714132459 | 0.714132459 | 0.714132459 | 0.7430558 | 0.644729035 | 1 | 0.743055796 | 0.743055796 | 0.743055796 | 0.961075148 | -0.017242653 | FALSE | TRUE | FALSE |
| COTL1 | 0.714025 | 0.63960092 | 1 | 0.714024985 | 0.714024985 | 0.714024985 | 0.74293352 | 0.644811753 | 1 | 0.742933521 | 0.742933521 | 0.742933521 | 0.961088664 | -0.017236545 | FALSE | TRUE | FALSE |
| TAX1BP3 | 0.7136572 | 0.63967951 | 1 | 0.713657209 | 0.713657209 | 0.713657209 | 0.7424755 | 0.644894435 | 1 | 0.742475497 | 0.742475497 | 0.742475497 | 0.961186209 | -0.017192469 | FALSE | TRUE | FALSE |
| KRTAP6-1 | 0.7134549 | 0.63975807 | 0.5039 | 6.720849234 | 6.720849234 | 6.720849234 | 0.7660397 | 0.633527225 | 1 | 0.766039703 | 0.766039703 | 0.766039703 | 0.931354963 | -0.030884767 | FALSE | TRUE | FALSE |
| ASCL3 | 0.7131133 | 0.63983659 | 0.8919 | 67.37367136 | 2717.523542 | 28089.46294 | 37.2044383 | 0.074302492 | 0.74435 | 148.7702816 | 14857.00314 | 59423.90429 | 0.019167425 | -1.717436231 | FALSE | FALSE | FALSE |
| ZNF146 | 0.7131094 | 0.63991508 | 1 | 65.44730295 | 6647.086624 | 6647.086624 | 0.74200544 | 0.645224744 | 1 | 68.17143806 | 6931.065782 | 6931.065782 | 0.96105679 | -0.017250949 | FALSE | TRUE | FALSE |
| CST6 | 0.7129309 | 0.63999353 | 1 | 0.71293089 | 0.71293089 | 0.71293089 | 0.74178747 | 0.64530722 | 1 | 0.74178747 | 0.74178747 | 0.74178747 | 0.961098588 | -0.017232061 | FALSE | TRUE | FALSE |
| FGF6 | 0.7129077 | 0.64007194 | 0.8759 | 2.851452918 | 11.40795104 | 11.40795104 | 0.22092054 | 0.846987288 | 0.749 | 0.883442007 | 3.533690882 | 3.533690882 | 3.226986873 | 0.508797199 | FALSE | TRUE | FALSE |
| GJA10 | 0.7127385 | 0.64015032 | 0.3687 | 0.178227561 | 2.85097874 | 11.40686207 | 0.1081949 | 0.873792944 | 0.32325 | 0.027064408 | 0.432636941 | 0.432636941 | 6.587541819 | 0.818723385 | FALSE | TRUE | FALSE |
| MRPL34 | 0.7125065 | 0.64022866 | 1 | 0.712506496 | 0.712506496 | 0.712506496 | 0.74147236 | 0.645718968 | 1 | 0.741472356 | 0.741472356 | 0.741472356 | 0.960934672 | -0.017306136 | FALSE | TRUE | FALSE |
| HPCA | 0.7122903 | 0.64030696 | 1 | 0.712290288 | 0.712290288 | 0.712290288 | 0.74159168 | 0.645554394 | 1 | 0.741591683 | 0.741591683 | 0.741591683 | 0.960488506 | -0.017507828 | FALSE | TRUE | FALSE |
| LMO4 | 0.712215 | 0.64038523 | 1 | 0.712215002 | 0.712215002 | 0.712215002 | 0.7415146 | 0.645636702 | 1 | 0.741514596 | 0.741514596 | 0.741514596 | 0.960486829 | -0.017508586 | FALSE | TRUE | FALSE |
| CMTM8 | 0.7121656 | 0.64046346 | 1 | 0.712165603 | 0.712165603 | 0.712165603 | 0.74124729 | 0.645801193 | 1 | 0.741247288 | 0.741247288 | 0.741247288 | 0.960766555 | -0.017382123 | FALSE | TRUE | FALSE |
| RPL30 | 0.7121325 | 0.64054166 | 1 | 0.712132469 | 0.712132469 | 0.712132469 | 0.72325777 | 0.654624274 | 1 | 0.723257771 | 0.723257771 | 0.723257771 | 0.984617791 | -0.006732321 | FALSE | TRUE | FALSE |
| TMEM229B | 0.7118698 | 0.64061982 | 1 | 0.711869784 | 0.711869784 | 0.711869784 | 0.74109326 | 0.645965519 | 1 | 0.74109326 | 0.74109326 | 0.74109326 | 0.960567073 | -0.017472305 | FALSE | TRUE | FALSE |
| UBE2B | 0.7117643 | 0.64069794 | 1 | 0.711764303 | 0.711764303 | 0.711764303 | 0.74096004 | 0.64604762 | 1 | 0.740960044 | 0.740960044 | 0.740960044 | 0.960597416 | -0.017458586 | FALSE | TRUE | FALSE |
| NDUFA2 | 0.711442 | 0.64077603 | 1 | 0.711441998 | 0.711441998 | 0.711441998 | 0.74055503 | 0.646375613 | 1 | 0.740555035 | 0.740555035 | 0.740555035 | 0.960687544 | -0.01741784 | FALSE | TRUE | FALSE |
| HECTD3 | 0.7110819 | 0.64085408 | 0.5135 | 0.711081869 | 0.711081869 | 13.926488 | 0.96239139 | 0.591760126 | 0.0226 | 0.962391391 | 0.962391391 | 18.89548164 | 0.738869732 | -0.131432124 | FALSE | TRUE | FALSE |
| SNRPA1 | 0.7107633 | 0.6409321 | 1 | 0.710763331 | 0.710763331 | 0.710763331 | 0.73850476 | 0.647192809 | 1 | 0.738504758 | 0.738504758 | 0.738504758 | 0.962435683 | -0.016628283 | FALSE | TRUE | FALSE |
| FKBP1A | 0.710748 | 0.64101009 | 1 | 0.710748039 | 0.710748039 | 0.710748039 | 6.76903546 | 0.249302103 | 0.0019 | 6.769035456 | 6.769035456 | 6.769035456 | 0.104999899 | -0.978811119 | FALSE | FALSE | FALSE |
| MDFI | 0.7107009 | 0.64108803 | 1 | 0.710700875 | 0.710700875 | 0.710700875 | 0.73998829 | 0.646539372 | 1 | 0.739988291 | 0.739988291 | 0.739988291 | 0.960421786 | -0.017537997 | FALSE | TRUE | FALSE |
| LGALS7B | 0.7106616 | 0.64116594 | 1 | 0.710661636 | 0.710661636 | 0.710661636 | 0.72248909 | 0.655071132 | 1 | 0.72248909 | 0.72248909 | 0.72248909 | 0.983629574 | -0.007168422 | FALSE | TRUE | FALSE |
| CCDC126 | 0.710024 | 0.64124382 | 1 | 0.710024003 | 0.710024003 | 0.710024003 | 0.7392695 | 0.646866414 | 1 | 0.739269501 | 0.739269501 | 0.739269501 | 0.960440004 | -0.017529759 | FALSE | TRUE | FALSE |
| ANO10 | 0.7099844 | 0.64132167 | 0.50685 | 0.709984398 | 6.578120417 | 6.578120417 | 4.87212124 | 0.313083039 | 0.4032 | 1.218660457 | 45.52830555 | 182.0645597 | 0.145723877 | -0.836469282 | FALSE | FALSE | FALSE |
| MAPK1 | 0.7099321 | 0.64139947 | 1 | 0.709932061 | 0.709932061 | 0.709932061 | 0.73889354 | 0.647029692 | 1 | 0.738893541 | 0.738893541 | 0.738893541 | 0.960804259 | -0.017365081 | FALSE | TRUE | FALSE |
| CNIH3 | 0.7098835 | 0.64147724 | 0.0036 | 0.709883549 | 0.709883549 | 0.709883549 | 0.73898452 | 0.646948074 | 1 | 0.738984523 | 0.738984523 | 0.738984523 | 0.960620321 | -0.017448231 | FALSE | TRUE | FALSE |
| PSMA7 | 0.7098561 | 0.64155497 | 1 | 0.709856092 | 0.709856092 | 0.709856092 | 0.73939896 | 0.646702976 | 1 | 0.739398963 | 0.739398963 | 0.739398963 | 0.96004475 | -0.017708523 | FALSE | TRUE | FALSE |
| ABHD13 | 0.7097822 | 0.64163267 | 1 | 0.709782248 | 0.709782248 | 0.709782248 | 0.73932904 | 0.646784716 | 1 | 0.73932904 | 0.73932904 | 0.73932904 | 0.960035666 | -0.017712632 | FALSE | TRUE | FALSE |
| CIB3 | 0.7097047 | 0.64171032 | 0.7524 | 0.709704726 | 6.607377378 | 6.607377378 | 0.10521636 | 0.874640844 | 0.7534 | 0.959424509 | 8.979139919 | 8.979139919 | 6.745193766 | 0.82899443 | FALSE | TRUE | FALSE |
| LSM7 | 0.709673 | 0.64178795 | 1 | 0.709672981 | 0.709672981 | 0.709672981 | 2.92254498 | 0.417301334 | 0.5004 | 2.92254498 | 2.92254498 | 2.92254498 | 0.242827052 | -0.614702933 | FALSE | FALSE | FALSE |
| GHRH | 0.7095436 | 0.64186553 | 1 | 0.7095436 | 0.7095436 | 0.7095436 | 0.73876142 | 0.64711127 | 1 | 0.738761421 | 0.738761421 | 0.738761421 | 0.960450261 | -0.017525121 | FALSE | TRUE | FALSE |
| TMEM60 | 0.7095145 | 0.64194308 | 0.50155 | 0.709514516 | 0.709514516 | 0.709514516 | 0.06242432 | 0.884195951 | 1 | 0.062424321 | 0.062424321 | 0.062424321 | 11.36599495 | 1.055607459 | FALSE | TRUE | FALSE |

| | | | | | | | | | | | | | | | | | |
|---|---|---|---|---|---|---|---|---|---|---|---|---|---|---|---|---|---|
| PCBP1 | 0.7093503 | 0.64202059 | 1 | 0.709350332 | 0.709350332 | 14.05422028 | 0.73578169 | 0.648572869 | 1 | 0.735781694 | 0.735781694 | 0.735781694 | 0.964077168 | -0.015888202 | FALSE | TRUE | FALSE |
| NPIPA1 | 0.7088557 | 0.64209807 | 1 | 65.24269759 | 650.3646937 | 650.3646937 | 0.71545707 | 0.65853031 | 0.93875 | 65.77774089 | 6692.308143 | 6692.308143 | 0.990773264 | -0.004025721 | FALSE | TRUE | FALSE |
| TXNDC9 | 0.7084556 | 0.64217552 | 1 | 0.708455633 | 0.708455633 | 0.708455633 | 0.73790645 | 0.647437194 | 1 | 0.737906454 | 0.737906454 | 0.737906454 | 0.960088679 | -0.017688651 | FALSE | TRUE | FALSE |
| EMC7 | 0.7082593 | 0.64225294 | 1 | 0.708259291 | 0.708259291 | 0.708259291 | 0.73785035 | 0.647518573 | 1 | 0.737850354 | 0.737850354 | 0.737850354 | 0.959895578 | -0.017776009 | FALSE | TRUE | FALSE |
| TBC1D32 | 0.7080734 | 0.64233032 | 0.0234 | 0.081428113 | 0.708073362 | 2.741313497 | 0.14310126 | 0.866070767 | 0.02635 | 0.143101259 | 0.143101259 | 2.810427182 | 4.948058239 | 0.694434803 | FALSE | TRUE | FALSE |
| DKK2 | 0.7078383 | 0.64240766 | 1 | 0.70783832 | 0.70783832 | 0.70783832 | 0.73757494 | 0.647599913 | 1 | 0.737574936 | 0.737574936 | 0.737574936 | 0.959683262 | -0.01787208 | FALSE | TRUE | FALSE |
| EBAG9 | 0.7078305 | 0.64248497 | 1 | 0.707830549 | 0.707830549 | 0.707830549 | 0.73732718 | 0.647762473 | 1 | 0.73732718 | 0.73732718 | 0.73732718 | 0.959995194 | -0.017730941 | FALSE | TRUE | FALSE |
| MEMO1 | 0.7076989 | 0.64256224 | 1 | 0.707698884 | 0.707698884 | 0.707698884 | 0.73740965 | 0.647681214 | 1 | 0.737409649 | 0.737409649 | 0.737409649 | 0.959709281 | -0.017860305 | FALSE | TRUE | FALSE |
| PRR18 | 0.7076625 | 0.64263947 | 0.0058 | 0.707662484 | 0.707662484 | 0.707662484 | 0.73551087 | 0.648815211 | 1 | 0.735510873 | 0.735510873 | 0.735510873 | 0.962137352 | -0.016762925 | FALSE | TRUE | FALSE |
| C9orf153 | 0.7075338 | 0.64271667 | 1 | 0.707533777 | 0.707533777 | 0.707533777 | 0.73683811 | 0.64800601 | 1 | 0.73683811 | 0.73683811 | 0.73683811 | 0.960229619 | -0.017624902 | FALSE | TRUE | FALSE |
| HMGN3 | 0.7072221 | 0.64279384 | 1 | 0.707222135 | 0.707222135 | 0.707222135 | 0.7365428 | 0.648249186 | 1 | 0.736542805 | 0.736542805 | 0.736542805 | 0.960191492 | -0.017642146 | FALSE | TRUE | FALSE |
| HEPN1 | 0.707181 | 0.64287097 | 1 | 0.707180965 | 0.707180965 | 0.707180965 | 0.73651057 | 0.648330163 | 1 | 0.736510569 | 0.736510569 | 0.736510569 | 0.96017762 | -0.017648421 | FALSE | TRUE | FALSE |
| PSMB1 | 0.7068652 | 0.64294807 | 1 | 0.706865216 | 0.706865216 | 0.706865216 | 0.72568819 | 0.654136143 | 1 | 0.72568819 | 0.72568819 | 0.72568819 | 0.974061898 | -0.011413444 | FALSE | TRUE | FALSE |
| ZNRF2 | 0.706509 | 0.64302513 | 1 | 0.706509037 | 0.706509037 | 0.706509037 | 0.71317631 | 0.659364727 | 1 | 6.74100276 | 6.74100276 | 6.74100276 | 0.990651292 | -0.00407919 | FALSE | TRUE | FALSE |
| UBE2G2 | 0.7062547 | 0.64310216 | 1 | 0.706254709 | 0.706254709 | 0.706254709 | 0.73589889 | 0.648411106 | 1 | 0.735898887 | 0.735898887 | 0.735898887 | 0.959717051 | -0.017856789 | FALSE | TRUE | FALSE |
| PPP1R17 | 0.7061898 | 0.64317915 | 1 | 6.658153693 | 6.658153693 | 6.658153693 | 0.75926821 | 0.637026657 | 1 | 0.75926821 | 0.75926821 | 0.75926821 | 0.930092712 | -0.031473759 | FALSE | TRUE | FALSE |
| PDGFA | 0.7060719 | 0.64325611 | 1 | 0.706071907 | 0.706071907 | 0.706071907 | 0.73581412 | 0.648492008 | 1 | 0.735814118 | 0.735814118 | 0.735814118 | 0.959579179 | -0.017919184 | FALSE | TRUE | FALSE |
| ARPC5L | 0.7058516 | 0.64333303 | 1 | 0.705851605 | 0.705851605 | 0.705851605 | 0.73557027 | 0.648734472 | 1 | 0.735570274 | 0.735570274 | 0.735570274 | 0.959597784 | -0.017910764 | FALSE | TRUE | FALSE |
| WFDC11 | 0.7057461 | 0.64340992 | 1 | 0.705746137 | 0.705746137 | 0.705746137 | 0.73519118 | 0.648976573 | 1 | 0.735191184 | 0.735191184 | 0.735191184 | 0.959949129 | -0.017751781 | FALSE | TRUE | FALSE |
| ABLIM1 | 0.7056903 | 0.64348677 | 0.2645 | 0.176476545 | 0.705690319 | 3.467998392 | 0.25498217 | 0.83791094 | 0.7611 | 0.254982171 | 1.019683606 | 5.021255461 | 2.767606516 | 0.442104344 | FALSE | TRUE | FALSE |
| IL18 | 0.7056001 | 0.64356359 | 1 | 0.705600124 | 0.705600124 | 0.705600124 | 0.73514621 | 0.649057193 | 1 | 0.73514621 | 0.73514621 | 0.73514621 | 0.959809239 | -0.017815074 | FALSE | TRUE | FALSE |
| UBE2J2 | 0.7055817 | 0.64364037 | 1 | 0.705581705 | 0.705581705 | 0.705581705 | 0.73532015 | 0.648895912 | 1 | 0.735320145 | 0.735320145 | 0.735320145 | 0.959557153 | -0.017929153 | FALSE | TRUE | FALSE |
| KRTAP3-2 | 0.7054004 | 0.64371711 | 1 | 0.705400391 | 0.705400391 | 0.705400391 | 0.73489566 | 0.649137774 | 1 | 0.734895655 | 0.734895655 | 0.734895655 | 0.959864691 | -0.017789984 | FALSE | TRUE | FALSE |
| DCUN1D1 | 0.7052114 | 0.64379382 | 1 | 0.705211389 | 2.820744811 | 2.820744811 | 0.74339632 | 0.644397738 | 1 | 0.743396317 | 0.743396317 | 0.743396317 | 0.948634493 | -0.022901088 | FALSE | TRUE | FALSE |
| ZDHHC12 | 0.7051891 | 0.6438705 | 0.50465 | 0.705189139 | 0.705189139 | 0.705189139 | 0.10905635 | 0.873632139 | 0.00835 | 0.109056354 | 0.109056354 | 0.109056354 | 6.466281972 | 0.810654639 | FALSE | TRUE | FALSE |
| BOK | 0.7048747 | 0.64394714 | 1 | 0.704874655 | 0.704874655 | 0.704874655 | 0.73469779 | 0.649218317 | 1 | 0.734697785 | 0.734697785 | 0.734697785 | 0.959407621 | -0.017996836 | FALSE | TRUE | FALSE |
| RBP3 | 0.7046126 | 0.64402374 | 0.76335 | 0.70461255 | 6.561926003 | 127.1676124 | 0.11109352 | 0.873116994 | 1 | 0.111093523 | 0.111093523 | 2.161003506 | 6.34251694 | 0.802261636 | FALSE | TRUE | FALSE |
| NXPH4 | 0.7045535 | 0.64410032 | 1 | 0.704553497 | 0.704553497 | 13.97541706 | 0.73449183 | 0.649298821 | 1 | 0.734491826 | 0.734491826 | 0.734491826 | 0.959239398 | -0.018072992 | FALSE | TRUE | FALSE |
| CD79B | 0.7040874 | 0.64417686 | 1 | 0.704087443 | 0.704087443 | 0.704087443 | 0.73406373 | 0.649540101 | 1 | 0.734063726 | 0.734063726 | 0.734063726 | 0.959163922 | -0.018107165 | FALSE | TRUE | FALSE |
| RPS11 | 0.7036339 | 0.64425336 | 1 | 0.703633941 | 0.703633941 | 0.703633941 | 0.72784271 | 0.65288124 | 0.00555 | 0.727842709 | 0.727842709 | 0.727842709 | 0.966739011 | -0.014690756 | FALSE | TRUE | FALSE |
| CBX5 | 0.703076 | 0.64432984 | 1 | 2.812220364 | 2.812220364 | 2.812220364 | 2.88365973 | 0.420441214 | 0.00455 | 0.720933801 | 2.88365973 | 2.88365973 | 0.24381377 | -0.61294177 | FALSE | FALSE | FALSE |
| CCNYL1 | 0.7030522 | 0.64440628 | 1 | 0.703052211 | 0.703052211 | 0.703052211 | 0.73309068 | 0.649781039 | 1 | 0.733090678 | 0.733090678 | 0.733090678 | 0.959024897 | -0.018170118 | FALSE | TRUE | FALSE |
| APOA2 | 0.7029576 | 0.64448269 | 1 | 0.702957572 | 0.702957572 | 0.702957572 | 0.73264705 | 0.649941472 | 1 | 0.732647049 | 0.732647049 | 0.732647049 | 0.959476425 | -0.017965692 | FALSE | TRUE | FALSE |
| LELP1 | 0.7026604 | 0.64455907 | 1 | 0.702660439 | 0.702660439 | 0.702660439 | 0.73237625 | 0.65026186 | 1 | 0.732376245 | 0.732376245 | 0.732376245 | 0.959425492 | -0.017988747 | FALSE | TRUE | FALSE |
| ADCYAP1 | 0.7023341 | 0.64463541 | 1 | 0.702334053 | 0.702334053 | 0.702334053 | 0.73234797 | 0.650341856 | 1 | 0.732347965 | 0.732347965 | 0.732347965 | 0.95901687 | -0.018173753 | FALSE | TRUE | FALSE |
| PPP1R14C | 0.7022571 | 0.64471171 | 1 | 0.702257077 | 0.702257077 | 0.702257077 | 0.73219961 | 0.650501727 | 1 | 0.732199609 | 0.732199609 | 0.732199609 | 0.959106054 | -0.018133368 | FALSE | TRUE | FALSE |
| RPL19 | 0.7021139 | 0.64478798 | 1 | 0.702113888 | 0.702113888 | 0.702113888 | 0.73106939 | 0.650980416 | 0.0053 | 0.731069391 | 0.731069391 | 0.731069391 | 0.96039295 | -0.017551037 | FALSE | TRUE | FALSE |
| SMG7 | 0.7020552 | 0.64486422 | 0.9386 | 6.584567961 | 624.602306 | 624.602306 | 8.97758475 | 0.212136464 | 0.2614 | 0.951293688 | 8.977584752 | 32.06603536 | 0.078200899 | -1.106788256 | FALSE | FALSE | FALSE |
| RPL18A | 0.7019628 | 0.64494042 | 1 | 0.7019628 | 0.7019628 | 0.7019628 | 0.73214161 | 0.650581603 | 1 | 0.732141609 | 0.732141609 | 0.732141609 | 0.958780092 | -0.018280992 | FALSE | TRUE | FALSE |
| RAB7A | 0.7015034 | 0.64501659 | 1 | 0.701503402 | 0.701503402 | 0.701503402 | 0.72640042 | 0.653823336 | 1 | 0.726400421 | 0.726400421 | 0.726400421 | 0.965725489 | -0.015146306 | FALSE | TRUE | FALSE |
| SNAP25 | 0.7013261 | 0.64509272 | 1 | 0.701326128 | 0.701326128 | 0.701326128 | 0.73022836 | 0.651616496 | 1 | 0.730228362 | 0.730228362 | 0.730228362 | 0.96042028 | -0.017538678 | FALSE | TRUE | FALSE |
| CAAP1 | 0.7008658 | 0.64516883 | 1 | 0.700865787 | 0.700865787 | 0.700865787 | 0.73091485 | 0.651060063 | 1 | 0.730914846 | 0.730914846 | 0.730914846 | 0.958888428 | -0.018231923 | FALSE | TRUE | FALSE |
| CMTM7 | 0.7006219 | 0.6452449 | 1 | 0.700621865 | 0.700621865 | 0.700621865 | 0.73058116 | 0.651139673 | 1 | 0.730581155 | 0.730581155 | 0.730581155 | 0.958992522 | -0.018184779 | FALSE | TRUE | FALSE |
| CLPS | 0.7006069 | 0.64532093 | 1 | 0.700606873 | 0.700606873 | 0.700606873 | 0.73048906 | 0.651298772 | 1 | 0.730489062 | 0.730489062 | 0.730489062 | 0.959092901 | -0.018139324 | FALSE | TRUE | FALSE |
| ADO | 0.7003405 | 0.64539693 | 1 | 0.700340524 | 0.700340524 | 0.700340524 | 0.73057648 | 0.651219242 | 1 | 0.730576479 | 0.730576479 | 0.730576479 | 0.958613566 | -0.018356429 | FALSE | TRUE | FALSE |
| KDELR1 | 0.7002777 | 0.6454729 | 1 | 0.700277709 | 0.700277709 | 0.700277709 | 0.73043038 | 0.651378262 | 1 | 0.730430378 | 0.730430378 | 0.730430378 | 0.958719311 | -0.018308525 | FALSE | TRUE | FALSE |
| FAM168B | 0.7001731 | 0.64554883 | 0.00585 | 0.700173056 | 0.700173056 | 0.700173056 | 0.73023567 | 0.651537124 | 1 | 0.730235669 | 0.730235669 | 0.730235669 | 0.95883163 | -0.018257648 | FALSE | TRUE | FALSE |
| C1orf226 | 0.6998288 | 0.64562473 | 1 | 0.699828845 | 0.699828845 | 0.699828845 | 0.72998549 | 0.651695829 | 1 | 0.729985492 | 0.729985492 | 0.729985492 | 0.958688703 | -0.01832239 | FALSE | TRUE | FALSE |
| RMDN1 | 0.6996622 | 0.64570059 | 0.50155 | 0.699662188 | 0.699662188 | 0.699662188 | 1.86882656 | 0.501292227 | 0.75085 | 1.868826564 | 17.59042137 | 17.59042137 | 0.374385832 | -0.426680595 | FALSE | FALSE | FALSE |
| SCGB1D2 | 0.6993022 | 0.64577643 | 1 | 0.699302226 | 0.699302226 | 0.699302226 | 0.72928377 | 0.651933602 | 1 | 0.729283771 | 0.729283771 | 0.729283771 | 0.958889054 | -0.018231639 | FALSE | TRUE | FALSE |
| FAM163A | 0.6990183 | 0.64585223 | 1 | 0.699018297 | 0.699018297 | 0.699018297 | 0.72911869 | 0.652091927 | 1 | 0.72911869 | 0.72911869 | 0.72911869 | 0.958716744 | -0.018309688 | FALSE | TRUE | FALSE |
| SPINK9 | 0.6984061 | 0.645928 | 1 | 0.69840613 | 0.69840613 | 0.69840613 | 0.70578348 | 0.663324742 | 1 | 6.648357489 | 6.648357489 | 6.648357489 | 0.989547283 | -0.004563449 | FALSE | TRUE | FALSE |
| SLC35D2 | 0.6982594 | 0.64600373 | 0.62795 | 1.24677145 | 11.17211627 | 11.17211627 | 0.43573016 | 0.778973813 | 0.49955 | 0.108944292 | 0.435730165 | 0.435730165 | 1.602504116 | 0.204799154 | FALSE | TRUE | FALSE |
| ZNRF1 | 0.6982288 | 0.64607943 | 1 | 0.698228825 | 0.698228825 | 0.698228825 | 0.70097083 | 0.666103897 | 1 | 0.700970828 | 6.611756342 | 6.611756342 | 0.996088278 | -0.001702171 | FALSE | TRUE | FALSE |
| SLC15A3 | 0.6979608 | 0.6461551 | 0.7552 | 0.697960844 | 6.436763835 | 13.81357773 | 8.48147687 | 0.218545134 | 0.4966 | 0.914716374 | 80.66228133 | 80.66228133 | 0.082292371 | -1.084640423 | FALSE | FALSE | FALSE |
| LYRM9 | 0.6978974 | 0.64623074 | 1 | 0.697897406 | 0.697897406 | 0.697897406 | 0.72801855 | 0.652723687 | 1 | 0.72801855 | 0.72801855 | 0.72801855 | 0.958625857 | -0.018350861 | FALSE | TRUE | FALSE |
| HCST | 0.6978589 | 0.64630633 | 1 | 0.697858897 | 0.697858897 | 0.697858897 | 0.72796573 | 0.652802483 | 1 | 0.727965726 | 0.727965726 | 0.727965726 | 0.958642519 | -0.018343313 | FALSE | TRUE | FALSE |
| CHIC2 | 0.697827 | 0.6463819 | 1 | 0.697827022 | 0.697827022 | 0.697827022 | 0.72808067 | 0.652644852 | 1 | 0.728080667 | 0.728080667 | 0.728080667 | 0.958447399 | -0.018431717 | FALSE | TRUE | FALSE |
| RPL9 | 0.6978214 | 0.64645742 | 1 | 0.697821426 | 0.697821426 | 0.697821426 | 0.72830166 | 0.652408113 | 1 | 0.728301662 | 0.728301662 | 0.728301662 | 0.958148886 | -0.018567001 | FALSE | TRUE | FALSE |
| RPS13 | 0.6976873 | 0.64653291 | 1 | 0.69768732 | 0.69768732 | 0.69768732 | 0.72675725 | 0.653588327 | 1 | 0.726757249 | 0.726757249 | 0.726757249 | 0.960000496 | -0.017728542 | FALSE | TRUE | FALSE |
| AKIRIN2 | 0.697405 | 0.64660837 | 1 | 0.69740505 | 0.69740505 | 0.69740505 | 0.72780063 | 0.652959958 | 1 | 0.727800633 | 0.727800633 | 0.727800633 | 0.958236388 | -0.018527342 | FALSE | TRUE | FALSE |
| FXYD6 | 0.6973709 | 0.6466838 | 1 | 0.697370901 | 0.697370901 | 0.697370901 | 0.70357338 | 0.665012724 | 1 | 0.70357338 | 0.70357338 | 0.70357338 | 0.991184317 | -0.003845578 | FALSE | TRUE | FALSE |
| STMN2 | 0.6973549 | 0.64675918 | 1 | 0.697354873 | 0.697354873 | 0.697354873 | 0.72778716 | 0.653038636 | 1 | 0.727787157 | 0.727787157 | 0.727787157 | 0.958185187 | -0.018550547 | FALSE | TRUE | FALSE |
| SMDT1 | 0.6973436 | 0.64683454 | 1 | 0.697343583 | 0.697343583 | 0.697343583 | 0.72755253 | 0.653117277 | 1 | 0.727552526 | 0.727552526 | 0.727552526 | 0.958478677 | -0.018417544 | FALSE | TRUE | FALSE |
| CISD1 | 0.6973072 | 0.64690986 | 1 | 0.697307176 | 0.697307176 | 0.697307176 | 0.72749236 | 0.653195879 | 1 | 0.727492362 | 0.727492362 | 0.727492362 | 0.958507899 | -0.018404304 | FALSE | TRUE | FALSE |
| RNF7 | 0.6966952 | 0.64698514 | 1 | 0.696695194 | 0.696695194 | 0.696695194 | 0.72687805 | 0.653431464 | 1 | 0.72687805 | 0.72687805 | 0.72687805 | 0.958476039 | -0.01841874 | FALSE | TRUE | FALSE |
| CAV1 | 0.6965963 | 0.6470604 | 0.75095 | 0.696596306 | 2.785708062 | 2.785708062 | 0.81345223 | 0.616472839 | 0.87395 | 7.683141845 | 74.47816208 | 74.47816208 | 0.856345685 | -0.067350886 | FALSE | TRUE | FALSE |
| RFWD3 | 0.6963359 | 0.64713562 | 0.50775 | 0.696335901 | 0.696335901 | 13.72896665 | 0.10743751 | 0.873963403 | 1 | 0.107437508 | 0.107437508 | 2.121858996 | 6.481311005 | 0.811662862 | FALSE | TRUE | FALSE |
| TEF | 0.6962646 | 0.64721081 | 1 | 0.696264601 | 0.696264601 | 0.696264601 | 0.72683462 | 0.653509915 | 1 | 0.72683462 | 0.72683462 | 0.72683462 | 0.957940888 | -0.018661289 | FALSE | TRUE | FALSE |
| SEPHS2 | 0.696093 | 0.64728596 | 1 | 0.696092978 | 0.696092978 | 13.79954689 | 0.72132953 | 0.655536631 | 1 | 0.721329532 | 0.721329532 | 0.721329532 | 0.965013835 | -0.015466461 | FALSE | TRUE | FALSE |
| GABARAPL2 | 0.6958633 | 0.64736108 | 1 | 0.695863277 | 0.695863277 | 0.695863277 | 0.72616335 | 0.654057995 | 1 | 0.726163347 | 0.726163347 | 0.726163347 | 0.958273754 | -0.018510406 | FALSE | TRUE | FALSE |
| KBTBD4 | 0.6952383 | 0.64743617 | 0.50585 | 0.695238301 | 0.695238301 | 1.51829658 | 0.93155567 | 0.594701219 | 0.50545 | 0.931555667 | 3.725586852 | 3.725586852 | 0.746319652 | -0.127075122 | FALSE | TRUE | FALSE |
| ADAMTS1 | 0.6950828 | 0.64751123 | 0.271 | 0.173809827 | 0.695082839 | 5.865849057 | 1.00562446 | 0.588116922 | 0.2734 | 0.251448627 | 1.005624458 | 8.493598199 | 0.691195241 | -0.160399261 | FALSE | FALSE | FALSE |

| | | | | | | | | | | | | | | | | | |
|---|---|---|---|---|---|---|---|---|---|---|---|---|---|---|---|---|---|
| OR51T1 | 0.6950513 | 0.64758625 | 0.5019 | 0.69505126 | 0.69505126 | 0.69505126 | 0.94024282 | 0.594050943 | 0.50445 | 0.940242821 | 0.940242821 | 0.940242821 | 0.739225277 | -0.131223191 | FALSE | TRUE | FALSE |
| TFF2 | 0.6945366 | 0.64766125 | 1 | 0.694536628 | 0.694536628 | 0.694536628 | 0.72504424 | 0.654292331 | 1 | 0.725044237 | 0.725044237 | 0.725044237 | 0.957923108 | -0.01866935 | FALSE | TRUE | FALSE |
| DEFB124 | 0.6944523 | 0.64773621 | 1 | 0.694452349 | 0.694452349 | 0.694452349 | 0.72487013 | 0.654448369 | 1 | 0.72487013 | 0.72487013 | 0.72487013 | 0.958036923 | -0.018617753 | FALSE | TRUE | FALSE |
| DCST2 | 0.6941771 | 0.64781113 | 0.2718 | 0.694177112 | 2.7766945 | 13.63050427 | 0.6273699 | 0.700300745 | 0.5127 | 0.627369898 | 0.627369898 | 12.34574599 | 1.106487759 | 0.043946614 | FALSE | TRUE | FALSE |
| MAPK6 | 0.6940511 | 0.64788603 | 0.01645 | 0.694051148 | 0.694051148 | 5.8701809 | 1621.89935 | 0.001466894 | 0.0665 | 16.70897937 | 1621.899353 | 1621.899353 | 0.000427925 | -3.368632424 | FALSE | FALSE | FALSE |
| ADRA2B | 0.6939274 | 0.64796089 | 0.8789 | 6.503875371 | 62.52339622 | 62.52339622 | 3.58954643 | 0.378509894 | 0.2533 | 0.972249515 | 9.161580076 | 19.2215348 | 0.193318976 | -0.713725514 | FALSE | FALSE | FALSE |
| RPL35A | 0.693887 | 0.64803571 | 1 | 0.693886973 | 0.693886973 | 0.693886973 | 0.71879075 | 0.656694708 | 1 | 0.718790745 | 0.718790745 | 0.718790745 | 0.965353237 | -0.015313743 | FALSE | TRUE | FALSE |
| UBE2S | 0.693681 | 0.6481105 | 1 | 2.774599088 | 11.10067136 | 11.10067136 | 0.74092153 | 0.64612968 | 1 | 0.740921532 | 0.740921532 | 0.740921532 | 0.93624083 | -0.028612423 | FALSE | TRUE | FALSE |
| BIRC5 | 0.6935944 | 0.64818526 | 1 | 0.693594362 | 6.551438556 | 6.551438556 | 0.7478728 | 0.642312268 | 1 | 0.747872799 | 0.747872799 | 0.747872799 | 0.927422903 | -0.032722183 | FALSE | TRUE | FALSE |
| ZSCAN22 | 0.6935099 | 0.64825998 | 0.5024 | 0.693509874 | 0.693509874 | 1.530508242 | 0.1071121 | 0.874053466 | 1 | 0.107112097 | 0.107112097 | 2.120749079 | 6.474617669 | 0.811214128 | FALSE | TRUE | FALSE |
| NRIP2 | 0.6934099 | 0.64833467 | 0.9391 | 6.32163026 | 566.6360928 | 566.6360928 | 8.58766601 | 0.217531641 | 0.68905 | 8.58766601 | 781.4317531 | 781.4317531 | 0.080744858 | -1.092885124 | FALSE | FALSE | FALSE |
| RBM22 | 0.6931098 | 0.64840933 | 1 | 0.69310979 | 0.69310979 | 13.74396115 | 0.71916075 | 0.656309615 | 1 | 0.719160753 | 0.719160753 | 14.27516491 | 0.963775884 | -0.016023945 | FALSE | TRUE | FALSE |
| ARHGAP18 | 0.6930329 | 0.64848395 | 0.5064 | 0.693032919 | 0.693032919 | 2.591359663 | 0.1032915 | 0.875124718 | 0.5039 | 0.939066576 | 0.939066576 | 3.494076741 | 6.709486601 | 0.82668929 | FALSE | TRUE | FALSE |
| PHF11 | 0.6929756 | 0.64855854 | 1 | 2.771883216 | 26.20186655 | 26.20186655 | 6.73573479 | 0.250602663 | 0.4988 | 0.71217357 | 6.735734789 | 6.735734789 | 0.102880473 | -0.987667049 | FALSE | FALSE | FALSE |
| FGF7 | 0.6929082 | 0.64863309 | 1 | 2.77151685 | 2.77151685 | 2.77151685 | 0.73203325 | 0.650661439 | 1 | 0.73203325 | 0.73203325 | 0.73203325 | 0.946552929 | -0.023855096 | FALSE | TRUE | FALSE |
| SND1 | 0.6924067 | 0.64870761 | 0.02945 | 0.692406678 | 0.692406678 | 13.54813021 | 0.2150203 | 0.84836427 | 0.0269 | 0.2150203 | 0.2150203 | 4.21822195 | 3.220192136 | 0.507881785 | FALSE | TRUE | FALSE |
| CYP4V2 | 0.6924006 | 0.6487821 | 0.47425 | 0.389503396 | 6.224649007 | 24.89312913 | 0.03694617 | 0.889677788 | 0.9384 | 0.326558962 | 11.84555167 | 11.84555167 | 18.74079647 | 1.272788044 | FALSE | TRUE | FALSE |
| MAP2K4 | 0.6921905 | 0.64885656 | 1 | 0.692190492 | 0.692190492 | 13.7500209 | 0.72319846 | 0.65476004 | 1 | 0.723198462 | 0.723198462 | 0.723198462 | 0.957123843 | -0.019031865 | FALSE | TRUE | FALSE |
| EIF3F | 0.6921788 | 0.64893098 | 1 | 0.692178788 | 0.692178788 | 13.71896121 | 0.72319884 | 0.654682177 | 1 | 0.723198844 | 0.723198844 | 14.34929641 | 0.957107155 | -0.019039437 | FALSE | TRUE | FALSE |
| ZMAT2 | 0.6915238 | 0.64900537 | 1 | 0.691523811 | 0.691523811 | 0.691523811 | 0.72247831 | 0.655148808 | 1 | 0.722478309 | 0.722478309 | 0.722478309 | 0.957155118 | -0.019017674 | FALSE | TRUE | FALSE |
| OR1D5 | 0.6914343 | 0.64907973 | 1 | 6.532267488 | 6.532267488 | 6.532267488 | 0.74550534 | 0.643733201 | 1 | 0.745505338 | 0.745505338 | 0.745505338 | 0.927470647 | -0.032699826 | FALSE | TRUE | FALSE |
| RAD23B | 0.6913984 | 0.64915406 | 1 | 0.69139842 | 0.69139842 | 0.69139842 | 0.72246634 | 0.655226445 | 1 | 0.722466341 | 0.722466341 | 0.722466341 | 0.956997414 | -0.019089236 | FALSE | TRUE | FALSE |
| NKX2-1 | 0.6912928 | 0.64922835 | 0.50565 | 0.691292776 | 0.691292776 | 3.424375613 | 0.21288732 | 0.848902768 | 1 | 0.851224018 | 0.851224018 | 4.221318809 | 3.247223821 | 0.511512224 | FALSE | TRUE | FALSE |
| DLK1 | 0.691042 | 0.64930261 | 0.50885 | 0.691041991 | 0.691041991 | 3.419122779 | 0.21519061 | 0.848321599 | 0.0114 | 0.215190612 | 0.215190612 | 4.262731925 | 3.211301762 | 0.506681118 | FALSE | TRUE | FALSE |
| TPTE2 | 0.6909934 | 0.64937684 | 0.9648 | 62.39156357 | 6205.201411 | 64077.21649 | 1073.4217 | 0.002294591 | 0.2579 | 1073.4217 | 113255.9801 | 1203621.023 | 0.00064373 | -3.191296455 | FALSE | FALSE | FALSE |
| COX19 | 0.6909436 | 0.64945103 | 1 | 0.690943598 | 0.690943598 | 0.690943598 | 2.85253401 | 0.423753977 | 0.00185 | 0.713148308 | 2.852534012 | 2.852534012 | 0.242220985 | -0.615788234 | FALSE | FALSE | FALSE |
| YWHAE | 0.6904828 | 0.64952519 | 1 | 0.690482786 | 0.690482786 | 0.690482786 | 0.72154866 | 0.655459136 | 1 | 0.721548664 | 0.721548664 | 0.721548664 | 0.956945553 | -0.019112771 | FALSE | TRUE | FALSE |
| ADAMTS10 | 0.6904143 | 0.64959932 | 0.27535 | 0.690414266 | 6.563311181 | 127.4192947 | 0.10767791 | 0.873853156 | 1 | 0.107677908 | 0.107677908 | 2.097831755 | 6.411846973 | 0.806983149 | FALSE | TRUE | FALSE |
| IL32 | 0.690264 | 0.64967341 | 0.8749 | 6.274293688 | 58.49765425 | 58.49765425 | 0.95908055 | 0.592088707 | 0.7565 | 0.959080547 | 8.794301711 | 8.794301711 | 0.719714277 | -0.142839882 | FALSE | TRUE | FALSE |
| PIM3 | 0.6899451 | 0.64974747 | 1 | 0.68994506 | 0.68994506 | 13.69371621 | 0.72086071 | 0.655614093 | 1 | 0.720860711 | 0.720860711 | 0.720860711 | 0.957112865 | -0.019036846 | FALSE | TRUE | FALSE |
| PCNA | 0.689739 | 0.6498215 | 1 | 0.689739011 | 0.689739011 | 0.689739011 | 0.71744535 | 0.657538639 | 1 | 0.717445351 | 0.717445351 | 0.717445351 | 0.961381951 | -0.017104036 | FALSE | TRUE | FALSE |
| GNGT1 | 0.68943 | 0.6498955 | 1 | 0.689429967 | 0.689429967 | 0.689429967 | 0.69775119 | 0.667187659 | 1 | 0.697751192 | 6.572312914 | 6.572312914 | 0.988074223 | -0.00521043 | FALSE | TRUE | FALSE |
| NUP205 | 0.6894007 | 0.64996947 | 0.5212 | 0.689400672 | 2.757554194 | 53.111474 | 0.10624475 | 0.874312961 | 1 | 0.106244749 | 0.106244749 | 3.560148466 | 6.488797579 | 0.812164226 | FALSE | TRUE | FALSE |
| KLF5 | 0.6893428 | 0.6500434 | 1 | 0.689342777 | 0.689342777 | 13.68131154 | 0.72059851 | 0.655691519 | 1 | 0.720598512 | 0.720598512 | 0.720598512 | 0.956625313 | -0.019258131 | FALSE | TRUE | FALSE |
| GPR88 | 0.6890161 | 0.6501173 | 1 | 0.68901611 | 0.68901611 | 13.66022812 | 0.72016812 | 0.65584626 | 1 | 0.720168117 | 0.720168117 | 14.29280375 | 0.956743425 | -0.019204514 | FALSE | TRUE | FALSE |
| TNFSF4 | 0.6885497 | 0.65019117 | 1 | 0.688549683 | 0.688549683 | 0.688549683 | 0.71971735 | 0.65592358 | 1 | 0.719717346 | 0.719717346 | 0.719717346 | 0.956694579 | -0.019226687 | FALSE | TRUE | FALSE |
| SPRR2F | 0.6883504 | 0.65026501 | 1 | 0.688350378 | 0.688350378 | 0.688350378 | 0.71913926 | 0.656540782 | 1 | 0.719139264 | 0.719139264 | 0.719139264 | 0.957186476 | -0.019003446 | FALSE | TRUE | FALSE |
| HAUS3 | 0.6881967 | 0.65033881 | 0.8748 | 6.308960497 | 59.32309536 | 59.32309536 | 73.7667743 | 0.038958526 | 0.34865 | 73.76677428 | 7081.385491 | 71900.22239 | 0.009329359 | -2.030148193 | FALSE | FALSE | FALSE |
| RAP2B | 0.6881004 | 0.65041259 | 1 | 0.688100395 | 0.688100395 | 0.688100395 | 0.71935486 | 0.656232092 | 1 | 0.719354859 | 0.719354859 | 0.719354859 | 0.956552092 | -0.019291374 | FALSE | TRUE | FALSE |
| GRK5 | 0.6879784 | 0.65048632 | 0.2612 | 0.687978382 | 2.752198134 | 13.56815218 | 0.85546747 | 0.606601164 | 0.5108 | 0.855467474 | 0.855467474 | 4.225425351 | 0.804213372 | -0.09462871 | FALSE | TRUE | FALSE |
| COPS5 | 0.6877891 | 0.65056003 | 1 | 0.687789139 | 0.687789139 | 0.687789139 | 0.71872965 | 0.656771614 | 1 | 0.718729649 | 0.718729649 | 0.718729649 | 0.956951116 | -0.019110247 | FALSE | TRUE | FALSE |
| IER2 | 0.6877575 | 0.6506337 | 1 | 6.501453939 | 6.501453939 | 6.501453939 | 6.79999495 | 0.248324105 | 0.50025 | 6.799994949 | 6.799994949 | 6.799994949 | 0.101140878 | -0.99507328 | FALSE | FALSE | FALSE |
| EID2 | 0.6876472 | 0.65070734 | 1 | 0.687647157 | 0.687647157 | 0.687647157 | 0.71890141 | 0.656617764 | 0.00785 | 0.718901405 | 0.718901405 | 0.718901405 | 0.956524986 | -0.019303681 | FALSE | TRUE | FALSE |
| UQCRFS1 | 0.6874846 | 0.65078095 | 1 | 0.687484577 | 0.687484577 | 0.687484577 | 0.7186837 | 0.656848483 | 1 | 0.718683695 | 0.718683695 | 0.718683695 | 0.956588527 | -0.019274832 | FALSE | TRUE | FALSE |
| NRAS | 0.6874653 | 0.65085452 | 1 | 0.687465275 | 0.687465275 | 0.687465275 | 0.71864368 | 0.656925313 | 1 | 0.718643685 | 0.718643685 | 0.718643685 | 0.956614926 | -0.019262847 | FALSE | TRUE | FALSE |
| ANKS4B | 0.6873231 | 0.65092806 | 0.5094 | 0.687323119 | 0.687323119 | 3.412098323 | 0.21352288 | 0.848775571 | 0.008 | 0.213522883 | 0.213522883 | 0.213522883 | 3.218967022 | 0.507716527 | FALSE | TRUE | FALSE |
| P2RX6 | 0.6870115 | 0.65100157 | 0.00885 | 0.073697179 | 0.68701149 | 0.68701149 | 0.01062481 | 0.896597016 | 1 | 0.010624809 | 0.010624809 | 0.210965342 | 64.66106907 | 1.810642881 | FALSE | TRUE | FALSE |
| SDC2 | 0.6868045 | 0.65107504 | 1 | 0.686804535 | 0.686804535 | 0.686804535 | 0.71793392 | 0.657232265 | 1 | 0.717933921 | 0.717933921 | 0.717933921 | 0.956640319 | -0.019251319 | FALSE | TRUE | FALSE |
| POU2AF1 | 0.6866114 | 0.65114849 | 1 | 0.686611425 | 0.686611425 | 0.686611425 | 0.71784912 | 0.657308913 | 1 | 0.717849119 | 0.717849119 | 0.717849119 | 0.956484317 | -0.019322146 | FALSE | TRUE | FALSE |
| GRIA4 | 0.6865439 | 0.6512219 | 0.2651 | 0.171684526 | 0.686543908 | 5.791461899 | 0.0636364 | 0.883869411 | 1 | 0.063636396 | 0.063636396 | 1.254867669 | 10.78854166 | 1.032962743 | FALSE | TRUE | FALSE |
| TMEFF1 | 0.6863393 | 0.65129528 | 1 | 0.686339334 | 0.686339334 | 0.686339334 | 0.71304649 | 0.659440371 | 1 | 0.71304649 | 0.71304649 | 0.71304649 | 0.962545 | -0.016578958 | FALSE | TRUE | FALSE |
| DERL2 | 0.6862415 | 0.65136862 | 1 | 0.686241528 | 0.686241528 | 0.686241528 | 0.71758962 | 0.657385526 | 1 | 0.717589622 | 0.717589622 | 0.717589622 | 0.956314734 | -0.019399153 | FALSE | TRUE | FALSE |
| NINJ1 | 0.6862096 | 0.65144194 | 1 | 0.686209643 | 0.686209643 | 0.686209643 | 0.71727117 | 0.657691604 | 1 | 0.71727117 | 0.71727117 | 0.71727117 | 0.956694862 | -0.019226558 | FALSE | TRUE | FALSE |
| PODXL2 | 0.6859831 | 0.65151522 | 1 | 0.685983115 | 0.685983115 | 13.584251 | 0.71752924 | 0.657462101 | 1 | 0.717529243 | 0.717529243 | 14.22581557 | 0.956035063 | -0.01952618 | FALSE | TRUE | FALSE |
| RPL7 | 0.6858212 | 0.65158846 | 0.00715 | 0.685821236 | 0.685821236 | 0.685821236 | 0.71729698 | 0.657615141 | 1 | 0.717296976 | 0.717296976 | 0.717296976 | 0.956118956 | -0.019488071 | FALSE | TRUE | FALSE |
| SNRPB | 0.6858124 | 0.65166168 | 1 | 0.685812409 | 0.685812409 | 0.685812409 | 0.71714106 | 0.657844419 | 1 | 0.717141059 | 0.717141059 | 0.717141059 | 0.956314522 | -0.019399249 | FALSE | TRUE | FALSE |
| FNTB | 0.6856763 | 0.65173486 | 1 | 0.685676287 | 0.685676287 | 13.58338654 | 0.71724315 | 0.65776803 | 1 | 0.717243148 | 0.717243148 | 14.22495107 | 0.95598862 | -0.019547277 | FALSE | TRUE | FALSE |
| C1orf141 | 0.6855582 | 0.65180801 | 0.74875 | 0.685558157 | 6.243674471 | 6.243674471 | 0.10982521 | 0.873410412 | 0.0084 | 0.10982521 | 0.10982521 | 0.10982521 | 6.242265827 | 0.795342259 | FALSE | TRUE | FALSE |
| PARS2 | 0.6854278 | 0.65188112 | 0.94485 | 43.89249807 | 2817.052727 | 11284.43919 | 65.0252247 | 0.043212813 | 0.3708 | 16.24795298 | 260.2996131 | 1042.254091 | 0.010540952 | -1.977120166 | FALSE | FALSE | FALSE |
| SSU72 | 0.6854055 | 0.6519542 | 1 | 0.68540545 | 0.68540545 | 0.68540545 | 0.71671053 | 0.657920774 | 1 | 0.716710528 | 0.716710528 | 0.716710528 | 0.956321169 | -0.019396231 | FALSE | TRUE | FALSE |
| POLR2D | 0.685363 | 0.65202725 | 0.0037 | 0.68536303 | 0.68536303 | 0.68536303 | 0.71658776 | 0.658149616 | 1 | 0.716587764 | 0.716587764 | 0.716587764 | 0.956425806 | -0.019348714 | FALSE | TRUE | FALSE |
| UCN2 | 0.6852418 | 0.65210027 | 1 | 0.685241753 | 0.685241753 | 0.685241753 | 0.71634197 | 0.658225824 | 1 | 0.716341972 | 0.716341972 | 0.716341972 | 0.956584675 | -0.019276581 | FALSE | TRUE | FALSE |
| ATP6V0C | 0.6849632 | 0.65217325 | 1 | 0.684963231 | 0.684963231 | 0.684963231 | 0.69932872 | 0.666682824 | 1 | 0.699328723 | 0.699328723 | 0.699328723 | 0.979458171 | -0.009014106 | FALSE | TRUE | FALSE |
| EXOSC6 | 0.6845861 | 0.65224621 | 1 | 0.684586069 | 0.684586069 | 0.684586069 | 0.71581819 | 0.658302001 | 1 | 0.71581819 | 0.71581819 | 0.71581819 | 0.95636864 | -0.019374673 | FALSE | TRUE | FALSE |
| UBE2N | 0.6845499 | 0.65231913 | 1 | 0.684549941 | 0.684549941 | 0.684549941 | 0.784341 | 0.626219244 | 1 | 0.784340999 | 7.441908318 | 7.441908318 | 0.872770826 | -0.05909978 | FALSE | TRUE | FALSE |
| KLHL9 | 0.6839451 | 0.65239202 | 0.5117 | 0.683945059 | 0.683945059 | 5.785785187 | 0.21301578 | 0.848874519 | 1 | 0.213015776 | 0.213015776 | 4.217681631 | 3.210771862 | 0.506609448 | FALSE | TRUE | FALSE |
| SIGLEC9 | 0.6836219 | 0.65246488 | 0.752 | 0.68362191 | 6.286918015 | 6.286918015 | 0.10574976 | 0.874482081 | 1 | 0.954650514 | 0.954650514 | 2.096166803 | 6.46452448 | 0.810536584 | FALSE | TRUE | FALSE |
| GOLT1B | 0.6828818 | 0.65253772 | 0.4979 | 0.682881753 | 2.731417761 | 2.731417761 | 0.72243862 | 0.655304044 | 0.00295 | 0.722438618 | 0.722438618 | 0.722438618 | 0.945245362 | -0.024455445 | FALSE | TRUE | FALSE |
| SSR1 | 0.6827985 | 0.65261053 | 1 | 0.682798529 | 0.682798529 | 0.682798529 | 0.71436595 | 0.658910129 | 1 | 0.714365952 | 0.714365952 | 0.714365952 | 0.955810572 | -0.01962817 | FALSE | TRUE | FALSE |
| TOMM20 | 0.6827567 | 0.6526833 | 1 | 0.682756668 | 0.682756668 | 0.682756668 | 0.69198648 | 0.669475327 | 1 | 0.691986475 | 6.52748663 | 6.52748663 | 0.986661867 | -0.005831656 | FALSE | TRUE | FALSE |
| COA4 | 0.6826736 | 0.65275604 | 1 | 0.68267358 | 0.68267358 | 0.68267358 | 0.71409392 | 0.659061803 | 1 | 0.714093922 | 0.714093922 | 0.714093922 | 0.955999706 | -0.019542241 | FALSE | TRUE | FALSE |
| MED19 | 0.6822383 | 0.65282875 | 1 | 0.682238253 | 0.682238253 | 0.682238253 | 0.71374533 | 0.659137588 | 1 | 0.71374533 | 0.71374533 | 0.71374533 | 0.955856696 | -0.019607213 | FALSE | TRUE | FALSE |
| PCGF1 | 0.6818661 | 0.65290143 | 1 | 0.681866117 | 0.681866117 | 0.681866117 | 0.70455586 | 0.664500926 | 1 | 0.704555858 | 0.704555858 | 0.704555858 | 0.967795681 | -0.01421632 | FALSE | TRUE | FALSE |

| | | | | | | | | | | | | | | | | | |
|---|---|---|---|---|---|---|---|---|---|---|---|---|---|---|---|---|---|
| NXPH1 | 0.6817832 | 0.65297408 | 1 | 0.681783236 | 0.681783236 | 0.681783236 | 0.71356341 | 0.659289048 | 1 | 0.713563407 | 0.713563407 | 0.713563407 | 0.955462724 | -0.019786252 | FALSE | TRUE | FALSE |
| PKDREJ | 0.6816045 | 0.65304669 | 0.39085 | 0.681604525 | 54.20664221 | 500.6086599 | 0.24758729 | 0.840037359 | 0.09435 | 0.028885029 | 2.175489641 | 4.771887988 | 2.752986788 | 0.439804127 | FALSE | TRUE | FALSE |
| DAPK2 | 0.6813403 | 0.65311928 | 0.5101 | 0.681340258 | 0.681340258 | 3.37522186 | 0.21179861 | 0.849142481 | 1 | 0.21179861 | 0.21179861 | 4.20105457 | 3.216925068 | 0.507440945 | FALSE | TRUE | FALSE |
| MANBAL | 0.6813181 | 0.65319183 | 1 | 0.68131812 | 0.68131812 | 0.68131812 | 0.7126726 | 0.659818039 | 1 | 0.712672596 | 0.712672596 | 0.712672596 | 0.956004375 | -0.01954012 | FALSE | TRUE | FALSE |
| RPL17 | 0.681114 | 0.65326435 | 1 | 0.681113969 | 0.681113969 | 0.681113969 | 0.71059338 | 0.661020544 | 1 | 0.71059338 | 0.71059338 | 0.71059338 | 0.958514375 | -0.01840137 | FALSE | TRUE | FALSE |
| CGGBP1 | 0.681066 | 0.65333684 | 1 | 0.681066036 | 0.681066036 | 0.681066036 | 0.71249815 | 0.659968849 | 1 | 0.712498147 | 0.712498147 | 0.712498147 | 0.955884641 | -0.019594517 | FALSE | TRUE | FALSE |
| GRAP | 0.6810439 | 0.65340929 | 1 | 0.681043939 | 0.681043939 | 0.681043939 | 0.71280113 | 0.659667081 | 1 | 0.712801126 | 0.712801126 | 0.712801126 | 0.955447339 | -0.019793245 | FALSE | TRUE | FALSE |
| YKT6 | 0.6810351 | 0.65348171 | 1 | 0.681035131 | 0.681035131 | 0.681035131 | 0.71268084 | 0.659742578 | 1 | 0.712680837 | 0.712680837 | 0.712680837 | 0.955596244 | -0.019725566 | FALSE | TRUE | FALSE |
| TIMM9 | 0.6808242 | 0.6535541 | 1 | 0.680824249 | 0.680824249 | 0.680824249 | 0.71226473 | 0.660044202 | 1 | 0.712264727 | 0.712264727 | 0.712264727 | 0.955858437 | -0.019606422 | FALSE | TRUE | FALSE |
| GPSM3 | 0.680693 | 0.65362646 | 1 | 6.41686403 | 6.41686403 | 6.41686403 | 0.73561193 | 0.648653691 | 1 | 0.735611929 | 0.735611929 | 0.735611929 | 0.925342539 | -0.033697473 | FALSE | TRUE | FALSE |
| CCL26 | 0.6806839 | 0.65369878 | 1 | 0.680683937 | 0.680683937 | 0.680683937 | 0.7121889 | 0.660119519 | 1 | 0.7121889 | 0.7121889 | 0.7121889 | 0.955763193 | -0.019649698 | FALSE | TRUE | FALSE |
| KDSR | 0.6805783 | 0.65377107 | 0.0102 | 0.680578256 | 0.680578256 | 1.442026586 | 0.90006843 | 0.598990488 | 0.75145 | 0.900068428 | 8.665335228 | 8.665335228 | 0.756140572 | -0.121397458 | FALSE | TRUE | FALSE |
| FABP5 | 0.6805621 | 0.65384333 | 1 | 0.680562116 | 0.680562116 | 0.680562116 | 0.71187163 | 0.660345253 | 0.0032 | 0.71187163 | 0.71187163 | 0.71187163 | 0.956018033 | -0.019533916 | FALSE | TRUE | FALSE |
| GOLGA7 | 0.6802423 | 0.65391556 | 1 | 0.6802423 | 0.6802423 | 0.6802423 | 0.71192141 | 0.660270045 | 1 | 0.711921405 | 0.711921405 | 0.711921405 | 0.955501963 | -0.019768416 | FALSE | TRUE | FALSE |
| ASB8 | 0.6800921 | 0.65398776 | 0.0078 | 0.680092132 | 0.680092132 | 0.680092132 | 8.04837891 | 0.226901377 | 0.13275 | 0.867003257 | 76.63192484 | 76.63192484 | 0.084500511 | -1.073140664 | FALSE | FALSE | FALSE |
| ITIH2 | 0.6799406 | 0.65405992 | 0.83515 | 12.57517061 | 806.0328743 | 3227.373771 | 48.4117178 | 0.05632136 | 0.0473 | 3.023031815 | 48.41171777 | 237.8364155 | 0.014044959 | -1.85247953 | FALSE | FALSE | FALSE |
| NEFM | 0.6798353 | 0.65413205 | 0.908 | 63.46173134 | 17221.06976 | 183448.9334 | 0.01313859 | 0.895731644 | 0.51815 | 0.013138591 | 0.013138591 | 0.257849889 | 51.74339355 | 1.713854908 | FALSE | TRUE | FALSE |
| GNRH1 | 0.6797842 | 0.65420415 | 1 | 0.67978423 | 0.67978423 | 0.67978423 | 0.711426 | 0.660495568 | 1 | 0.711425996 | 0.711425996 | 0.711425996 | 0.955523461 | -0.019758645 | FALSE | TRUE | FALSE |
| TMEM204 | 0.6797562 | 0.65427622 | 1 | 0.679756235 | 0.679756235 | 0.679756235 | 0.71153926 | 0.660420428 | 1 | 0.71153926 | 0.71153926 | 0.71153926 | 0.955332014 | -0.019845668 | FALSE | TRUE | FALSE |
| DRAP1 | 0.6795772 | 0.65434826 | 1 | 0.679577176 | 0.679577176 | 0.679577176 | 0.71125276 | 0.660570672 | 1 | 0.71125276 | 0.71125276 | 0.71125276 | 0.955465081 | -0.01978518 | FALSE | TRUE | FALSE |
| FGFR1 | 0.6795501 | 0.65442026 | 0.7427 | 2.716251126 | 102.4629047 | 991.5530974 | 9.55934569 | 0.204713283 | 0.11815 | 0.59770606 | 9.559345693 | 38.24380674 | 0.071087515 | -1.148206669 | FALSE | FALSE | FALSE |
| C2orf66 | 0.6794652 | 0.65449223 | 1 | 0.679465166 | 0.679465166 | 0.679465166 | 0.71096777 | 0.660795769 | 1 | 0.710967767 | 0.710967767 | 0.710967767 | 0.955690536 | -0.019682715 | FALSE | TRUE | FALSE |
| RAC2 | 0.6792195 | 0.65456417 | 1 | 0.679219519 | 0.679219519 | 0.679219519 | 0.71116188 | 0.66064574 | 1 | 0.711161883 | 0.711161883 | 0.711161883 | 0.955084258 | -0.019958313 | FALSE | TRUE | FALSE |
| LAMTOR3 | 0.6792137 | 0.65463607 | 1 | 0.679213735 | 0.679213735 | 0.679213735 | 0.71103738 | 0.660720772 | 1 | 0.711037382 | 0.711037382 | 0.711037382 | 0.955243356 | -0.019885974 | FALSE | TRUE | FALSE |
| ORM1 | 0.6790089 | 0.65470795 | 0.00585 | 0.679008851 | 0.679008851 | 0.679008851 | 0.66729313 | 0.68181244 | 1 | 6.289125695 | 60.83423376 | 60.83423376 | 1.01755709 | 0.007558785 | FALSE | TRUE | FALSE |
| JAZF1 | 0.6789956 | 0.65477979 | 1 | 0.678995552 | 0.678995552 | 0.678995552 | 0.70893244 | 0.661916454 | 1 | 0.708932439 | 0.708932439 | 0.708932439 | 0.957771877 | -0.018737919 | FALSE | TRUE | FALSE |
| VAMP8 | 0.6788204 | 0.6548516 | 1 | 0.678820357 | 0.678820357 | 0.678820357 | 0.71049233 | 0.661095397 | 1 | 0.710492334 | 0.710492334 | 0.710492334 | 0.955422494 | -0.019804538 | FALSE | TRUE | FALSE |
| KCTD6 | 0.67871 | 0.65492338 | 1 | 0.67871002 | 0.67871002 | 0.67871002 | 0.71065444 | 0.660945654 | 1 | 0.710654438 | 0.710654438 | 0.710654438 | 0.955049294 | -0.019974212 | FALSE | TRUE | FALSE |
| COX5B | 0.6784147 | 0.65499513 | 0.0025 | 0.678414682 | 0.678414682 | 0.678414682 | 0.71024221 | 0.661244999 | 1 | 0.710242212 | 0.710242212 | 0.710242212 | 0.95518778 | -0.019911242 | FALSE | TRUE | FALSE |
| IGFBP3 | 0.6783856 | 0.65506684 | 1 | 0.678385586 | 0.678385586 | 0.678385586 | 0.7102891 | 0.661170216 | 1 | 0.710289097 | 0.710289097 | 0.710289097 | 0.955083767 | -0.019958536 | FALSE | TRUE | FALSE |
| PSMA1 | 0.6780923 | 0.65513853 | 1 | 0.67809227 | 0.67809227 | 0.67809227 | 0.71012144 | 0.661394456 | 1 | 0.710121438 | 0.710121438 | 0.710121438 | 0.95489621 | -0.02004383 | FALSE | TRUE | FALSE |
| KXD1 | 0.6780243 | 0.65521018 | 1 | 0.678024277 | 0.678024277 | 0.678024277 | 0.70985019 | 0.661543773 | 1 | 0.709850185 | 0.709850185 | 0.709850185 | 0.955165317 | -0.019921456 | FALSE | TRUE | FALSE |
| EMG1 | 0.6779975 | 0.65528181 | 1 | 0.67799746 | 0.67799746 | 0.67799746 | 0.70990397 | 0.661469133 | 1 | 0.709903971 | 0.709903971 | 0.709903971 | 0.955055172 | -0.019971539 | FALSE | TRUE | FALSE |
| SGK3 | 0.6776702 | 0.6553534 | 1 | 6.42191103 | 6.42191103 | 13.4442794 | 0.7325231 | 0.65002163 | 1 | 0.732523105 | 0.732523105 | 0.732523105 | 0.925117912 | -0.03380291 | FALSE | TRUE | FALSE |
| EGR3 | 0.6773544 | 0.65542496 | 1 | 0.677354351 | 0.677354351 | 13.37439298 | 0.70948122 | 0.661692947 | 1 | 0.709481224 | 0.709481224 | 14.03022779 | 0.954717796 | -0.020124982 | FALSE | TRUE | FALSE |
| PPTC7 | 0.6771497 | 0.65549649 | 1 | 0.677149749 | 0.677149749 | 0.677149749 | 0.68709205 | 0.67214898 | 1 | 6.493919354 | 6.493919354 | 6.493919354 | 0.985529882 | -0.006330203 | FALSE | TRUE | FALSE |
| NSG2 | 0.6771157 | 0.65556799 | 0.00505 | 0.677115667 | 0.677115667 | 0.677115667 | 0.70914634 | 0.661767486 | 1 | 0.709146344 | 0.709146344 | 0.709146344 | 0.954832063 | -0.020073006 | FALSE | TRUE | FALSE |
| SLC30A3 | 0.6770971 | 0.65563946 | 0.50375 | 0.677097077 | 0.677097077 | 3.351628833 | 0.83469408 | 0.611350632 | 0.503 | 0.834694085 | 0.834694085 | 4.136705165 | 0.811191895 | -0.090876397 | FALSE | TRUE | FALSE |
| PSMB5 | 0.6770242 | 0.65571089 | 1 | 0.677024246 | 0.677024246 | 0.677024246 | 0.70914573 | 0.661841987 | 0.00515 | 0.709145728 | 0.709145728 | 0.709145728 | 0.954703976 | -0.020131269 | FALSE | TRUE | FALSE |
| PRAMEF19 | 0.6767038 | 0.6557823 | 1 | 6.414131466 | 62.39634226 | 126.9959298 | 0.73120567 | 0.650900731 | 1 | 6.938901501 | 6.938901501 | 14.49489818 | 0.925463002 | -0.033640939 | FALSE | TRUE | FALSE |
| EMC1 | 0.6764119 | 0.65585367 | 0.2663 | 0.077315483 | 0.676411866 | 2.606374789 | 0.01634201 | 0.894773525 | 0.02775 | 0.016342013 | 0.016342013 | 0.320449978 | 41.3909752 | 1.616905659 | FALSE | TRUE | FALSE |
| KRTAP19-5 | 0.6759872 | 0.65592502 | 1 | 0.675987201 | 0.675987201 | 0.675987201 | 0.70776056 | 0.662510937 | 1 | 0.707760564 | 0.707760564 | 0.707760564 | 0.955107186 | -0.019947887 | FALSE | TRUE | FALSE |
| GRM1 | 0.6759283 | 0.65599634 | 0.26785 | 0.184796078 | 2.951723253 | 6.239920955 | 0.00998851 | 0.896757355 | 1 | 0.009988506 | 0.009988506 | 0.196227537 | 67.67061186 | 1.830400103 | FALSE | TRUE | FALSE |
| SIDT1 | 0.6759202 | 0.65606762 | 0.8816 | 6.2098734 | 58.51807289 | 122.0261565 | 8.47363278 | 0.219213388 | 0.507 | 8.473632776 | 80.38644101 | 80.38644101 | 0.079767466 | -1.098174204 | FALSE | FALSE | FALSE |
| JUN | 0.6756668 | 0.65613887 | 1 | 0.675666777 | 0.675666777 | 13.37214882 | 0.70797453 | 0.662362525 | 1 | 0.707974532 | 0.707974532 | 14.02916323 | 0.954365937 | -0.02028507 | FALSE | TRUE | FALSE |
| MSC | 0.6755993 | 0.65621009 | 0.51005 | 0.675599346 | 0.675599346 | 0.675599346 | 0.21101712 | 0.849353386 | 1 | 0.211017118 | 0.211017118 | 0.211017118 | 3.201632895 | 0.505371533 | FALSE | TRUE | FALSE |
| BNIP3L | 0.6755568 | 0.65628128 | 1 | 0.675556796 | 0.675556796 | 0.675556796 | 0.70762704 | 0.66258509 | 1 | 0.707627042 | 0.707627042 | 0.707627042 | 0.954679169 | -0.020142553 | FALSE | TRUE | FALSE |
| SAR1A | 0.675491 | 0.65635244 | 1 | 0.675491047 | 2.701766913 | 2.701766913 | 0.71570184 | 0.658378141 | 1 | 0.715701845 | 0.715701845 | 0.715701845 | 0.943816272 | -0.025112539 | FALSE | TRUE | FALSE |
| KRTAP6-3 | 0.6753914 | 0.65642356 | 1 | 6.355512414 | 61.38055884 | 61.38055884 | 0.75317486 | 0.639860312 | 1 | 0.753174859 | 0.753174859 | 0.753174859 | 0.896725916 | -0.047340279 | FALSE | TRUE | FALSE |
| RPL22L1 | 0.6752687 | 0.65649465 | 1 | 0.675268691 | 0.675268691 | 0.675268691 | 0.70725362 | 0.662733294 | 1 | 0.707253615 | 0.707253615 | 0.707253615 | 0.954775877 | -0.020098562 | FALSE | TRUE | FALSE |
| ARF1 | 0.6745891 | 0.65656572 | 1 | 0.6745891 | 0.6745891 | 0.6745891 | 0.70678362 | 0.663029286 | 1 | 0.706783619 | 0.706783619 | 0.706783619 | 0.954449256 | -0.020247156 | FALSE | TRUE | FALSE |
| GNB1 | 0.6745087 | 0.65663676 | 1 | 0.674508727 | 0.674508727 | 13.36638061 | 0.70689738 | 0.662881359 | 1 | 0.706897375 | 0.706897375 | 14.02367057 | 0.954181966 | -0.020368796 | FALSE | TRUE | FALSE |
| SRSF3 | 0.6741834 | 0.65670777 | 1 | 0.674183438 | 0.674183438 | 0.674183438 | 0.70467637 | 0.664427675 | 1 | 0.704676369 | 0.704676369 | 0.704676369 | 0.956727751 | -0.019211628 | FALSE | TRUE | FALSE |
| MDM4 | 0.6740631 | 0.65677874 | 1 | 0.674063133 | 0.674063133 | 13.36816024 | 0.70576505 | 0.663472259 | 1 | 0.705765053 | 0.705765053 | 14.0110019 | 0.955081483 | -0.019959575 | FALSE | TRUE | FALSE |
| S100A14 | 0.6737147 | 0.65684969 | 1 | 0.673714717 | 0.673714717 | 0.673714717 | 0.70566355 | 0.663693265 | 1 | 0.705663554 | 0.705663554 | 0.705663554 | 0.954725114 | -0.020121653 | FALSE | TRUE | FALSE |
| LCE1E | 0.6736249 | 0.65692061 | 1 | 0.673624884 | 0.673624884 | 0.673624884 | 0.70558268 | 0.663766863 | 1 | 0.70558268 | 0.70558268 | 0.70558268 | 0.954707228 | -0.020129789 | FALSE | TRUE | FALSE |
| CNOT7 | 0.6735854 | 0.65699149 | 1 | 0.673585374 | 0.673585374 | 0.673585374 | 2.79220448 | 0.427642218 | 0.5081 | 2.792204477 | 2.792204477 | 2.792204477 | 0.241237839 | -0.617554571 | FALSE | FALSE | FALSE |
| CRB3 | 0.6734983 | 0.65706234 | 1 | 0.673498295 | 0.673498295 | 0.673498295 | 0.70578007 | 0.663398518 | 1 | 0.70578007 | 0.70578007 | 0.70578007 | 0.954260857 | -0.02033289 | FALSE | TRUE | FALSE |
| YWHAH | 0.6734358 | 0.65713316 | 1 | 0.673435802 | 0.673435802 | 0.673435802 | 0.70573132 | 0.663545963 | 0.00605 | 0.70573132 | 0.70573132 | 0.70573132 | 0.954238225 | -0.02034319 | FALSE | TRUE | FALSE |
| LIN28A | 0.6732255 | 0.65720395 | 1 | 0.673225537 | 0.673225537 | 0.673225537 | 0.70549582 | 0.663913952 | 1 | 0.705495815 | 0.705495815 | 0.705495815 | 0.954258725 | -0.020333861 | FALSE | TRUE | FALSE |
| PPP2CA | 0.6730834 | 0.65727471 | 1 | 0.673083398 | 0.673083398 | 0.673083398 | 0.70547677 | 0.663987443 | 1 | 0.705476769 | 0.705476769 | 0.705476769 | 0.954083009 | -0.020413838 | FALSE | TRUE | FALSE |
| UBE2E2 | 0.6730051 | 0.65734544 | 1 | 6.370183975 | 61.88255733 | 61.88255733 | 0.75156879 | 0.6407098 | 1 | 0.751568786 | 0.751568786 | 0.751568786 | 0.895467037 | -0.047950396 | FALSE | TRUE | FALSE |
| RWDD1 | 0.6727346 | 0.65741614 | 1 | 0.672734638 | 0.672734638 | 0.672734638 | 0.70475901 | 0.664281069 | 1 | 0.70475901 | 0.70475901 | 0.70475901 | 0.954559827 | -0.020196847 | FALSE | TRUE | FALSE |
| ETV3 | 0.6726809 | 0.65748681 | 1 | 0.6726809 | 0.6726809 | 0.6726809 | 0.70496492 | 0.664207713 | 1 | 0.70496492 | 0.70496492 | 0.70496492 | 0.954204784 | -0.02035841 | FALSE | TRUE | FALSE |
| AJAP1 | 0.6720786 | 0.65755745 | 1 | 0.672078576 | 0.672078576 | 13.28084921 | 0.70468706 | 0.66435439 | 1 | 0.704687056 | 0.704687056 | 13.94525423 | 0.953726297 | -0.020576243 | FALSE | TRUE | FALSE |
| TMEM141 | 0.6720782 | 0.65762806 | 1 | 0.672078227 | 0.672078227 | 0.672078227 | 0.70414519 | 0.664720474 | 1 | 0.704145189 | 0.704145189 | 0.704145189 | 0.95445973 | -0.020242391 | FALSE | TRUE | FALSE |
| PBOV1 | 0.671802 | 0.65769864 | 1 | 0.671802048 | 0.671802048 | 0.671802048 | 0.70389852 | 0.664939711 | 1 | 0.703898522 | 0.703898522 | 0.703898522 | 0.954401846 | -0.02026873 | FALSE | TRUE | FALSE |
| ZBTB43 | 0.6717319 | 0.65776918 | 1 | 0.671731885 | 0.671731885 | 13.29469091 | 0.70436282 | 0.664647326 | 1 | 0.704362822 | 0.704362822 | 13.95798022 | 0.953673114 | -0.020600461 | FALSE | TRUE | FALSE |
| ZBTB39 | 0.6715202 | 0.6578397 | 0.8757 | 2.686354153 | 24.8776741 | 52.9147261 | 0.21845898 | 0.847563976 | 1 | 0.218458984 | 0.218458984 | 4.310948649 | 3.073896083 | 0.487689182 | FALSE | TRUE | FALSE |
| ANAPC16 | 0.6706718 | 0.6579102 | 1 | 0.67067177 | 0.67067177 | 0.67067177 | 0.70285405 | 0.665377287 | 1 | 0.702854048 | 0.702854048 | 0.702854048 | 0.954212005 | -0.020355124 | FALSE | TRUE | FALSE |
| GTSF1 | 0.6706343 | 0.65798066 | 0.00455 | 0.670634258 | 0.670634258 | 0.670634258 | 0.70300063 | 0.665231565 | 1 | 0.703000631 | 0.703000631 | 0.703000631 | 0.953959682 | -0.02046998 | FALSE | TRUE | FALSE |
| GPX3 | 0.6706279 | 0.65805109 | 1 | 0.670627939 | 0.670627939 | 0.670627939 | 0.69720674 | 0.66747543 | 1 | 0.697206738 | 0.697206738 | 0.697206738 | 0.961878168 | -0.016879933 | FALSE | TRUE | FALSE |
| RPRM | 0.6706256 | 0.65812149 | 1 | 0.670625605 | 0.670625605 | 0.670625605 | 0.70290665 | 0.665304444 | 1 | 0.702906652 | 0.702906652 | 0.702906652 | 0.954074917 | -0.020417522 | FALSE | TRUE | FALSE |

| | | | | | | | | | | | | | | | | | |
|---|---|---|---|---|---|---|---|---|---|---|---|---|---|---|---|---|---|
| CXCL3 | 0.6704735 | 0.65819186 | 1 | 0.670473476 | 0.670473476 | 0.670473476 | 0.7027148 | 0.665522871 | 1 | 0.702714804 | 0.702714804 | 0.702714804 | 0.9541189 | -0.020397501 | FALSE | TRUE | FALSE |
| MAPK3 | 0.6703911 | 0.6582622 | 0.99935 | 648181.8336 | 260517395.3 | 8409909626 | 0.10090151 | 0.875712462 | 1 | 918757.9683 | 94832819.58 | 12360414801 | 6.644014374 | 0.822430563 | FALSE | TRUE | FALSE |
| CSF1 | 0.6702891 | 0.65833251 | 1 | 0.670289056 | 0.670289056 | 13.26024669 | 0.70302914 | 0.665158653 | 1 | 0.703029138 | 0.703029138 | 13.92610357 | 0.953429978 | -0.020711197 | FALSE | TRUE | FALSE |
| DIRAS1 | 0.670271 | 0.65840278 | 1 | 0.670270981 | 0.670270981 | 0.670270981 | 0.68031256 | 0.67511982 | 1 | 0.680312564 | 6.449230648 | 6.449230648 | 0.98523975 | -0.006458075 | FALSE | TRUE | FALSE |
| PFN1 | 0.6698085 | 0.65847303 | 1 | 0.669808533 | 0.669808533 | 0.669808533 | 0.70243089 | 0.665595613 | 1 | 0.702430893 | 0.702430893 | 0.702430893 | 0.953557908 | -0.020652928 | FALSE | TRUE | FALSE |
| NCMAP | 0.6696722 | 0.65854324 | 0.004 | 0.669672164 | 0.669672164 | 0.669672164 | 0.70193251 | 0.665740998 | 1 | 0.701932512 | 0.701932512 | 0.701932512 | 0.954040669 | -0.020433112 | FALSE | TRUE | FALSE |
| ASRGL1 | 0.6696221 | 0.65861343 | 0.8142 | 0.669622094 | 10.71379727 | 42.87093766 | 0.95110385 | 0.592744697 | 0.93655 | 15.21948014 | 243.7842327 | 516.5370392 | 0.704047293 | -0.152398167 | FALSE | TRUE | FALSE |
| GRM7 | 0.6688996 | 0.65868359 | 0.1338 | 0.071858448 | 0.668899571 | 3.27321364 | 0.00050918 | 0.899766186 | 0.5248 | 0.000509176 | 0.000509176 | 0.009982481 | 1313.689378 | 3.118492688 | FALSE | TRUE | FALSE |
| CSNK1E | 0.6687952 | 0.65875372 | 1 | 0.668795212 | 0.668795212 | 13.21695471 | 0.68065421 | 0.674777319 | 1 | 6.453800689 | 6.453800689 | 13.47054817 | 0.982577064 | -0.007633378 | FALSE | TRUE | FALSE |
| PLOD2 | 0.6687392 | 0.65882382 | 0.13035 | 0.072970916 | 0.668739245 | 0.82655281 | 0.14044935 | 0.86669575 | 0.1402 | 0.140449349 | 0.561471535 | 1.188198919 | 4.761426457 | 0.677737081 | FALSE | TRUE | FALSE |
| IFT81 | 0.6684003 | 0.65889389 | 0.5097 | 0.668400339 | 0.668400339 | 13.20963659 | 0.10218108 | 0.875448619 | 0.51055 | 0.408682378 | 0.408682378 | 3.454217998 | 6.541331951 | 0.815666189 | FALSE | TRUE | FALSE |
| MYPOP | 0.6683625 | 0.65896393 | 1 | 6.318021144 | 6.318021144 | 6.318021144 | 0.72458484 | 0.654526333 | 1 | 0.724584842 | 0.724584842 | 0.724584842 | 0.922407549 | -0.035077151 | FALSE | TRUE | FALSE |
| DBP | 0.6683533 | 0.65903394 | 1 | 0.668353314 | 0.668353314 | 0.668353314 | 0.67925141 | 0.675529822 | 1 | 6.437634027 | 6.437634027 | 6.437634027 | 0.983955726 | -0.007024443 | FALSE | TRUE | FALSE |
| VGLL2 | 0.6683532 | 0.65910392 | 0.00595 | 0.668353246 | 0.668353246 | 0.668353246 | 0.70098537 | 0.666031385 | 1 | 0.700985371 | 0.700985371 | 0.700985371 | 0.953448209 | -0.020702893 | FALSE | TRUE | FALSE |
| RBM24 | 0.6683411 | 0.65917386 | 1 | 2.673272498 | 10.69539548 | 10.69539548 | 0.70100489 | 0.665958838 | 1 | 2.803934553 | 11.21820185 | 11.21820185 | 0.953404325 | -0.020722882 | FALSE | TRUE | FALSE |
| CDC26 | 0.6680198 | 0.65924378 | 1 | 0.668019751 | 0.668019751 | 0.668019751 | 0.6787316 | 0.675802547 | 1 | 0.678731603 | 0.678731603 | 0.678731603 | 0.984217838 | -0.006908768 | FALSE | TRUE | FALSE |
| GPR27 | 0.6677377 | 0.65931367 | 0.9928 | 171.1767127 | 2744.792253 | 5860.402446 | 0.73288252 | 0.649861275 | 1 | 11.72864005 | 46.93720277 | 46.93720277 | 0.911111531 | -0.040428457 | FALSE | TRUE | FALSE |
| FAM114A2 | 0.6675144 | 0.65938352 | 0.75335 | 0.667514392 | 6.113154048 | 6.113154048 | 0.10380055 | 0.874967098 | 1 | 0.933565669 | 0.933565669 | 0.933565669 | 6.43074018 | 0.808260963 | FALSE | TRUE | FALSE |
| KRTAP1-3 | 0.6670935 | 0.65945335 | 1 | 0.667093458 | 0.667093458 | 0.667093458 | 0.69964393 | 0.666538289 | 1 | 0.699643933 | 0.699643933 | 0.699643933 | 0.953475657 | -0.02069039 | FALSE | TRUE | FALSE |
| CCND2 | 0.6670893 | 0.65952315 | 1 | 0.667089259 | 0.667089259 | 0.667089259 | 0.69990109 | 0.66639362 | 1 | 0.699901091 | 0.699901091 | 0.699901091 | 0.953119331 | -0.020852722 | FALSE | TRUE | FALSE |
| DSTN | 0.6669371 | 0.65959292 | 1 | 0.666937098 | 0.666937098 | 0.666937098 | 0.69946807 | 0.666610573 | 1 | 0.699468067 | 0.699468067 | 0.699468067 | 0.953491844 | -0.020683017 | FALSE | TRUE | FALSE |
| CCND3 | 0.6668706 | 0.65966266 | 1 | 0.666870617 | 0.666870617 | 0.666870617 | 0.69965477 | 0.666465972 | 1 | 0.699654769 | 0.699654769 | 0.699654769 | 0.953142388 | -0.020842216 | FALSE | TRUE | FALSE |
| PRR15 | 0.6664391 | 0.65973237 | 1 | 6.288966582 | 6.288966582 | 6.288966582 | 0.72266599 | 0.654837871 | 1 | 0.722665988 | 0.722665988 | 0.722665988 | 0.922195172 | -0.035177156 | FALSE | TRUE | FALSE |
| CCL20 | 0.6663572 | 0.65980205 | 1 | 0.666357235 | 0.666357235 | 0.666357235 | 0.69872188 | 0.666971495 | 1 | 0.698721882 | 0.698721882 | 0.698721882 | 0.953680216 | -0.020597226 | FALSE | TRUE | FALSE |
| IGFBP5 | 0.6663451 | 0.6598717 | 1 | 0.666345134 | 0.666345134 | 0.666345134 | 0.69428222 | 0.668478503 | 1 | 0.694282219 | 0.694282219 | 0.694282219 | 0.959761198 | -0.017836812 | FALSE | TRUE | FALSE |
| GADD45A | 0.6663031 | 0.65994132 | 1 | 0.666303148 | 0.666303148 | 0.666303148 | 0.69903207 | 0.666827225 | 1 | 0.699032067 | 0.699032067 | 0.699032067 | 0.95317966 | -0.020825234 | FALSE | TRUE | FALSE |
| PBX2 | 0.6662077 | 0.66001091 | 0.5034 | 0.666207685 | 0.666207685 | 5.643622173 | 0.20814891 | 0.850149518 | 1 | 0.208148908 | 0.208148908 | 4.124102232 | 3.200630212 | 0.5052355 | FALSE | TRUE | FALSE |
| DNHD1 | 0.6659129 | 0.66008046 | 0.9382 | 1278.766158 | 3233868.528 | 128976777.2 | 2527.30157 | 0.000919812 | 0.3484 | 2527.30157 | 1649801.365 | 137035337.7 | 0.000263488 | -3.579239632 | FALSE | FALSE | FALSE |
| TAF7 | 0.665899 | 0.66014999 | 1 | 0.665898951 | 0.665898951 | 0.665898951 | 0.69879996 | 0.666899377 | 1 | 0.698799959 | 0.698799959 | 0.698799959 | 0.952917845 | -0.02094454 | FALSE | TRUE | FALSE |
| CCL17 | 0.6649428 | 0.6602195 | 1 | 0.66494278 | 0.66494278 | 0.66494278 | 0.69740435 | 0.667259653 | 1 | 0.697404355 | 0.697404355 | 0.697404355 | 0.953453725 | -0.02070038 | FALSE | TRUE | FALSE |
| INTS9 | 0.664559 | 0.66028898 | 0.5078 | 0.664559022 | 0.664559022 | 13.10252833 | 0.90414701 | 0.59813955 | 0.5071 | 0.904147008 | 0.904147008 | 3.378882411 | 0.735012134 | -0.133705491 | FALSE | TRUE | FALSE |
| NANOS1 | 0.664531 | 0.66035842 | 1 | 0.664531002 | 0.664531002 | 0.664531002 | 0.66131445 | 0.685241465 | 1 | 6.242046129 | 6.242046129 | 6.242046129 | 1.004863875 | 0.002107233 | FALSE | TRUE | FALSE |
| YIPF4 | 0.6641957 | 0.66042784 | 1 | 0.664195678 | 0.664195678 | 0.664195678 | 0.68931591 | 0.670958643 | 1 | 2.757210983 | 2.757210983 | 2.757210983 | 0.963557736 | -0.016122258 | FALSE | TRUE | FALSE |
| OR51A2 | 0.6640497 | 0.66049723 | 1 | 0.664049684 | 0.664049684 | 0.664049684 | 0.68413237 | 0.673122488 | 1 | 0.684132369 | 0.684132369 | 0.684132369 | 0.970645031 | -0.012939564 | FALSE | TRUE | FALSE |
| SMU1 | 0.6640343 | 0.66056659 | 1 | 0.664034332 | 0.664034332 | 13.1565883 | 0.69716066 | 0.667547287 | 1 | 0.697160658 | 0.697160658 | 13.82844868 | 0.952483943 | -0.021142337 | FALSE | TRUE | FALSE |
| KLF6 | 0.6635825 | 0.66063593 | 1 | 6.274685467 | 6.274685467 | 6.274685467 | 6.37974843 | 0.258999797 | 0.25525 | 6.37974843 | 61.88978586 | 61.88978586 | 0.104013891 | -0.982908655 | FALSE | FALSE | FALSE |
| SIX3 | 0.6634695 | 0.66070523 | 0.99285 | 42.46911631 | 680.4305822 | 2724.59388 | 0.21519122 | 0.84830737 | 1 | 3.441328239 | 13.76701192 | 13.76701192 | 3.083162693 | 0.488996442 | FALSE | TRUE | FALSE |
| TIAL1 | 0.6634444 | 0.6607745 | 1 | 0.663444365 | 0.663444365 | 13.17194542 | 0.69656923 | 0.667906081 | 1 | 0.696569228 | 0.696569228 | 13.84175239 | 0.952445698 | -0.021159775 | FALSE | TRUE | FALSE |
| TRIM54 | 0.6632864 | 0.66084374 | 0.75655 | 0.663286376 | 6.094287907 | 6.094287907 | 0.10616544 | 0.874322922 | 1 | 0.106165441 | 0.106165441 | 0.106165441 | 6.247667503 | 0.795717909 | FALSE | TRUE | FALSE |
| PHF23 | 0.6630407 | 0.66091295 | 1 | 0.663040749 | 0.663040749 | 13.14609744 | 0.69627956 | 0.667977741 | 1 | 0.696279563 | 0.696279563 | 13.81948617 | 0.952262258 | -0.021243428 | FALSE | TRUE | FALSE |
| IL12A | 0.662459 | 0.66098214 | 1 | 0.662458995 | 0.662458995 | 0.662458995 | 0.6954765 | 0.668049376 | 1 | 0.695476499 | 0.695476499 | 0.695476499 | 0.952525349 | -0.021123458 | FALSE | TRUE | FALSE |
| ANKRD46 | 0.6622052 | 0.6610513 | 1 | 0.662205232 | 0.662205232 | 0.662205232 | 0.69524805 | 0.668120978 | 1 | 0.695248049 | 0.695248049 | 0.695248049 | 0.952473341 | -0.021147171 | FALSE | TRUE | FALSE |
| CDK6 | 0.6618866 | 0.66112043 | 1 | 0.661886564 | 0.661886564 | 13.12905038 | 0.69518631 | 0.668192547 | 1 | 0.695186309 | 0.695186309 | 13.80319267 | 0.952099539 | -0.021317645 | FALSE | TRUE | FALSE |
| LURAP1 | 0.6618203 | 0.66118953 | 1 | 0.661820348 | 0.661820348 | 0.661820348 | 0.694911 | 0.668335587 | 1 | 0.694911005 | 0.694911005 | 0.694911005 | 0.952381446 | -0.021189074 | FALSE | TRUE | FALSE |
| ARL8A | 0.6611511 | 0.66125861 | 1 | 0.661151121 | 0.661151121 | 0.661151121 | 6.36979575 | 0.259639648 | 0.4969 | 0.672821636 | 6.369795751 | 6.369795751 | 0.103794713 | -0.983824768 | FALSE | FALSE | FALSE |
| FAM180A | 0.6610984 | 0.66132766 | 1 | 0.661098439 | 0.661098439 | 0.661098439 | 0.6725714 | 0.678637077 | 1 | 6.335248187 | 6.335248187 | 6.335248187 | 0.982941642 | -0.007472266 | FALSE | TRUE | FALSE |
| NCS1 | 0.6604949 | 0.66139668 | 1 | 0.66049489 | 2.641846139 | 2.641846139 | 0.70157277 | 0.665813644 | 0.0062 | 0.701572773 | 0.701572773 | 0.701572773 | 0.941448864 | -0.026203264 | FALSE | TRUE | FALSE |
| CARTPT | 0.6604665 | 0.66146567 | 1 | 0.660466511 | 0.660466511 | 0.660466511 | 0.69359701 | 0.66876395 | 1 | 0.693597011 | 0.693597011 | 0.693597011 | 0.952233797 | -0.021256411 | FALSE | TRUE | FALSE |
| RNF126 | 0.6604445 | 0.66153463 | 1 | 6.243019737 | 60.56704435 | 60.56704435 | 0.69372989 | 0.668692638 | 1 | 6.561423897 | 63.69253959 | 63.69253959 | 0.952019616 | -0.021354103 | FALSE | TRUE | FALSE |
| RPL24 | 0.6603718 | 0.66160356 | 1 | 0.660371784 | 0.660371784 | 0.660371784 | 0.68594888 | 0.67242775 | 0.50185 | 2.74366191 | 2.74366191 | 2.74366191 | 0.962712829 | -0.016503241 | FALSE | TRUE | FALSE |
| RAB11B | 0.6601105 | 0.66167246 | 1 | 0.660110535 | 0.660110535 | 0.660110535 | 0.69003466 | 0.670395267 | 1 | 0.690034658 | 0.690034658 | 0.690034658 | 0.956633884 | -0.01925424 | FALSE | TRUE | FALSE |
| MRPS21 | 0.6595849 | 0.66174134 | 1 | 6.20585746 | 59.92640051 | 59.92640051 | 0.71478815 | 0.658682345 | 1 | 6.738870565 | 6.738870565 | 6.738870565 | 0.92276977 | -0.034906641 | FALSE | TRUE | FALSE |
| NPTN | 0.6593147 | 0.66181019 | 1 | 0.659314708 | 0.659314708 | 13.05652788 | 0.69240551 | 0.669191174 | 1 | 0.692405508 | 0.692405508 | 13.72806223 | 0.952208929 | -0.02126775 | FALSE | TRUE | FALSE |
| KLHL23 | 0.6592646 | 0.661879 | 1 | 0.659264634 | 0.659264634 | 13.05810105 | 0.67265809 | 0.678570199 | 1 | 6.378418646 | 6.378418646 | 13.3388455 | 0.980088763 | -0.00873459 | FALSE | TRUE | FALSE |
| NOP10 | 0.6591672 | 0.66194779 | 1 | 0.659167208 | 0.659167208 | 0.659167208 | 7.02220902 | 0.244075083 | 0.0028 | 0.740129603 | 7.022209021 | 7.022209021 | 0.093868924 | -1.027478159 | FALSE | FALSE | FALSE |
| RTN4RL1 | 0.6590221 | 0.66201655 | 1 | 0.659022112 | 0.659022112 | 13.01947429 | 0.69259562 | 0.668906489 | 1 | 0.692595618 | 0.692595618 | 13.70284992 | 0.951525096 | -0.021579753 | FALSE | TRUE | FALSE |
| NDN | 0.6590207 | 0.66208528 | 1 | 0.659020658 | 0.659020658 | 13.05712011 | 0.6924571 | 0.669120054 | 1 | 0.692457104 | 0.692457104 | 13.73502986 | 0.951713333 | -0.021493847 | FALSE | TRUE | FALSE |
| SCD | 0.6589359 | 0.66215397 | 1 | 0.658935942 | 0.658935942 | 0.658935942 | 0.69246034 | 0.6690489 | 1 | 0.692460341 | 0.692460341 | 0.692460341 | 0.951586544 | -0.021551708 | FALSE | TRUE | FALSE |
| TOR4A | 0.6588091 | 0.66222264 | 1 | 0.658809132 | 0.658809132 | 13.04689544 | 0.69226613 | 0.669262262 | 1 | 0.692266129 | 0.692266129 | 13.72565056 | 0.951670325 | -0.021513473 | FALSE | TRUE | FALSE |
| POP7 | 0.6588031 | 0.66229128 | 1 | 0.658803127 | 0.658803127 | 0.658803127 | 6.31578435 | 0.26155417 | 0.0048 | 0.670513842 | 6.315784352 | 6.315784352 | 0.10431058 | -0.981671641 | FALSE | FALSE | FALSE |
| FTO | 0.6587367 | 0.66235988 | 0.7478 | 2.633902858 | 24.86655139 | 24.86655139 | 33.0486487 | 0.08386098 | 0.32305 | 8.265050869 | 320.9088292 | 320.9088292 | 0.019932334 | -1.700441839 | FALSE | FALSE | FALSE |
| DNAJC5 | 0.6585659 | 0.66242846 | 1 | 0.658565881 | 0.658565881 | 0.658565881 | 0.69208141 | 0.669404338 | 1 | 0.692081409 | 0.692081409 | 0.692081409 | 0.951572853 | -0.021557956 | FALSE | TRUE | FALSE |
| PSG9 | 0.6578911 | 0.66249701 | 0.9924 | 203.3281859 | 53248.06412 | 104929.471 | 1.00932363 | 0.587337033 | 0.8745 | 9.309061022 | 88.06373954 | 88.06373954 | 0.651813811 | -0.185876442 | FALSE | FALSE | FALSE |
| PARN | 0.6577591 | 0.66256554 | 1 | 0.657759137 | 0.657759137 | 13.00956816 | 0.6915351 | 0.669688101 | 1 | 0.691535102 | 0.691535102 | 5.8529E+305 | 0.951157988 | -0.02174734 | FALSE | TRUE | FALSE |
| PCDHGB3 | 0.6577446 | 0.66263403 | 0.88175 | 6.064860303 | 57.3591838 | 118.4964401 | 0.10518846 | 0.874660662 | 1 | 0.9526496 | 0.9526496 | 18.66483535 | 6.253010586 | 0.796089164 | FALSE | TRUE | FALSE |
| CXXC5 | 0.657642 | 0.6627025 | 1 | 0.657642029 | 0.657642029 | 13.03284219 | 0.69113666 | 0.669758964 | 1 | 0.691136661 | 0.691136661 | 0.691136661 | 0.95153689 | -0.02157437 | FALSE | TRUE | FALSE |
| DEDD | 0.657471 | 0.66277093 | 1 | 0.657470962 | 0.657470962 | 0.657470962 | 0.69101839 | 0.669829794 | 1 | 0.691018388 | 0.691018388 | 13.7197995 | 0.951452194 | -0.021613028 | FALSE | TRUE | FALSE |
| USP25 | 0.6573281 | 0.66283934 | 0.51775 | 0.657328132 | 0.657328132 | 12.91572269 | 0.08849835 | 0.878501115 | 1 | 30.28188872 | 716.9787049 | 2867.452496 | 7.427574847 | 0.870847037 | FALSE | TRUE | FALSE |
| TNFRSF14 | 0.6572557 | 0.66290771 | 1 | 0.657255687 | 0.657255687 | 0.657255687 | 0.69083241 | 0.669971355 | 1 | 0.690832412 | 0.690832412 | 0.690832412 | 0.951396714 | -0.021638353 | FALSE | TRUE | FALSE |
| LTA | 0.6572197 | 0.66297606 | 1 | 0.657219665 | 0.657219665 | 0.657219665 | 0.690713 | 0.670042087 | 1 | 0.690713003 | 0.690713003 | 0.690713003 | 0.951509039 | -0.021587082 | FALSE | TRUE | FALSE |
| CTDSPL2 | 0.6571546 | 0.66304437 | 1 | 0.657154595 | 0.657154595 | 13.0294845 | 0.69086523 | 0.669900591 | 1 | 0.690865228 | 0.690865228 | 13.71210958 | 0.951205196 | -0.021725786 | FALSE | TRUE | FALSE |
| DAZAP2 | 0.6570482 | 0.66311266 | 1 | 0.657048217 | 0.657048217 | 0.657048217 | 0.69022137 | 0.670254094 | 1 | 0.690221367 | 0.690221367 | 0.690221367 | 0.95193839 | -0.021391159 | FALSE | TRUE | FALSE |
| RGS10 | 0.6568967 | 0.66318092 | 1 | 0.656896654 | 0.656896654 | 0.656896654 | 0.69018623 | 0.670324696 | 1 | 0.690186233 | 0.690186233 | 0.690186233 | 0.951767252 | -0.021469242 | FALSE | TRUE | FALSE |

| | | | | | | | | | | | | | | | | | |
|---|---|---|---|---|---|---|---|---|---|---|---|---|---|---|---|---|---|
| ARF3 | 0.6567506 | 0.66324914 | 1 | 0.656750564 | 0.656750564 | 0.656750564 | 0.69032061 | 0.670112789 | 1 | 0.69032061 | 0.69032061 | 0.69032061 | 0.951370355 | -0.021650385 | FALSE | TRUE | FALSE |
| ZNF580 | 0.6566707 | 0.66331734 | 0.00295 | 0.656670665 | 0.656670665 | 0.656670665 | 0.68325337 | 0.673468723 | 1 | 0.683253374 | 0.683253374 | 0.683253374 | 0.961093921 | -0.01723417 | FALSE | TRUE | FALSE |
| BATF3 | 0.6565927 | 0.66338551 | 1 | 0.656592683 | 0.656592683 | 0.656592683 | 0.68995237 | 0.670677212 | 1 | 0.689952373 | 0.689952373 | 0.689952373 | 0.951649286 | -0.021523074 | FALSE | TRUE | FALSE |
| YWHAQ | 0.6563762 | 0.66345365 | 1 | 0.656376162 | 0.656376162 | 0.656376162 | 0.69000093 | 0.670536306 | 1 | 0.690000929 | 0.690000929 | 0.690000929 | 0.95126852 | -0.021696875 | FALSE | TRUE | FALSE |
| WDR5 | 0.6563129 | 0.66352176 | 1 | 0.656312884 | 0.656312884 | 13.02961617 | 0.6899593 | 0.670606776 | 0.0103 | 0.689959301 | 0.689959301 | 13.70971714 | 0.951234201 | -0.021712543 | FALSE | TRUE | FALSE |
| AMMECR1L | 0.6562901 | 0.66358984 | 1 | 0.656290077 | 0.656290077 | 13.03355236 | 0.66836449 | 0.681156467 | 1 | 6.325528281 | 6.325528281 | 6.325528281 | 0.981934383 | -0.007917532 | FALSE | TRUE | FALSE |
| JUND | 0.6562814 | 0.66365789 | 1 | 0.656281381 | 0.656281381 | 0.656281381 | 0.63659356 | 0.69608834 | 1 | 6.006095499 | 58.157603 | 58.157603 | 1.030926831 | 0.013227843 | FALSE | TRUE | FALSE |
| CDKN1C | 0.6561884 | 0.66372591 | 1 | 0.656188361 | 0.656188361 | 0.656188361 | 0.6765072 | 0.676956152 | 1 | 0.676507203 | 0.676507203 | 0.676507203 | 0.969965077 | -0.013243902 | FALSE | TRUE | FALSE |
| DTX3 | 0.6558981 | 0.6637939 | 1 | 0.655898055 | 0.655898055 | 13.00325934 | 0.68960312 | 0.670747619 | 1 | 0.689603115 | 0.689603115 | 13.68584366 | 0.951123973 | -0.021762872 | FALSE | TRUE | FALSE |
| CMTM3 | 0.6558963 | 0.66386186 | 1 | 0.655896287 | 0.655896287 | 0.655896287 | 0.68920705 | 0.671099162 | 1 | 0.689207046 | 0.689207046 | 0.689207046 | 0.951667995 | -0.021514536 | FALSE | TRUE | FALSE |
| EIF2S2 | 0.6558235 | 0.66392979 | 1 | 0.655823463 | 0.655823463 | 0.655823463 | 0.68945275 | 0.670817993 | 1 | 0.689452753 | 0.689452753 | 0.689452753 | 0.951223212 | -0.02171756 | FALSE | TRUE | FALSE |
| SOCS1 | 0.6557761 | 0.66399769 | 1 | 0.65577613 | 0.65577613 | 0.65577613 | 0.68942289 | 0.670888334 | 1 | 0.689422894 | 0.689422894 | 0.689422894 | 0.951195754 | -0.021730097 | FALSE | TRUE | FALSE |
| SDSL | 0.6556525 | 0.66406557 | 0.3519 | 0.65565251 | 10.48956216 | 41.97221325 | 5.93958806 | 0.271667513 | 0.13095 | 0.635918882 | 5.939588059 | 5.939588059 | 0.110386866 | -0.957082597 | FALSE | FALSE | FALSE |
| SPINK2 | 0.6554666 | 0.66413341 | 1 | 0.655466613 | 0.655466613 | 0.655466613 | 0.68874949 | 0.671379814 | 1 | 0.688749488 | 0.688749488 | 0.688749488 | 0.951676371 | -0.021510714 | FALSE | TRUE | FALSE |
| TEKT4 | 0.6551629 | 0.66420123 | 1 | 0.655162874 | 0.655162874 | 12.91919211 | 0.68864132 | 0.671449896 | 1 | 0.68864132 | 0.68864132 | 13.60231211 | 0.951384784 | -0.021643799 | FALSE | TRUE | FALSE |
| SPOP | 0.6547298 | 0.66426902 | 1 | 0.654729804 | 0.654729804 | 12.9918937 | 0.68823981 | 0.671589966 | 1 | 0.688239809 | 0.688239809 | 0.688239809 | 0.951310568 | -0.021677679 | FALSE | TRUE | FALSE |
| ATRNL1 | 0.6545799 | 0.66433678 | 0.88055 | 6.165961879 | 59.57295527 | 120.4696491 | 0.10661773 | 0.874193322 | 1 | 0.10661773 | 0.10661773 | 3.589451084 | 6.139503353 | 0.788133241 | FALSE | TRUE | FALSE |
| CALY | 0.6545515 | 0.66440452 | 0.00545 | 0.654551497 | 0.654551497 | 0.654551497 | 0.68807515 | 0.671729908 | 1 | 0.688075148 | 0.688075148 | 0.688075148 | 0.951279085 | -0.021692052 | FALSE | TRUE | FALSE |
| NRGN | 0.6545097 | 0.66447222 | 1 | 0.654509654 | 0.654509654 | 0.654509654 | 0.66590639 | 0.682725926 | 1 | 0.665906388 | 0.665906388 | 0.665906388 | 0.98288538 | -0.007497125 | FALSE | TRUE | FALSE |
| HLF | 0.6543417 | 0.6645399 | 1 | 0.654341664 | 0.654341664 | 0.654341664 | 0.68813967 | 0.671659953 | 1 | 0.688139665 | 0.688139665 | 0.688139665 | 0.950884968 | -0.021872018 | FALSE | TRUE | FALSE |
| UBXN7 | 0.654314 | 0.66460754 | 1 | 0.654314007 | 0.654314007 | 12.9556303 | 6.31653026 | 0.260917088 | 0.0126 | 6.316530265 | 6.316530265 | 13.21883106 | 0.103587568 | -0.984692364 | FALSE | FALSE | FALSE |
| YPEL1 | 0.6540367 | 0.66467516 | 1 | 0.654036714 | 0.654036714 | 0.654036714 | 0.68758328 | 0.671939586 | 1 | 0.687583277 | 0.687583277 | 0.687583277 | 0.95121091 | -0.021723177 | FALSE | TRUE | FALSE |
| LRRC59 | 0.6540176 | 0.66474275 | 1 | 0.654017604 | 0.654017604 | 0.654017604 | 0.68778584 | 0.671869725 | 1 | 0.687785844 | 0.687785844 | 0.687785844 | 0.950902973 | -0.021863795 | FALSE | TRUE | FALSE |
| RAB5A | 0.6534958 | 0.66481031 | 1 | 0.65349578 | 0.65349578 | 0.65349578 | 0.68734643 | 0.672009416 | 1 | 0.687346432 | 0.687346432 | 0.687346432 | 0.950751688 | -0.021932895 | FALSE | TRUE | FALSE |
| KERA | 0.6532254 | 0.66487784 | 0.50865 | 0.653225414 | 0.653225414 | 0.653225414 | 0.20422418 | 0.851033869 | 1 | 0.204224184 | 0.204224184 | 0.204224184 | 3.198570321 | 0.504955903 | FALSE | TRUE | FALSE |
| THAP11 | 0.6531007 | 0.66494535 | 1 | 0.653100747 | 0.653100747 | 12.95184977 | 0.6797344 | 0.675256607 | 1 | 0.679734404 | 0.679734404 | 0.679734404 | 0.960817553 | -0.017359071 | FALSE | TRUE | FALSE |
| SF3B5 | 0.6527031 | 0.66501283 | 1 | 0.652703072 | 0.652703072 | 0.652703072 | 0.65947836 | 0.686006931 | 1 | 0.659478362 | 0.659478362 | 0.659478362 | 0.989726289 | -0.004484894 | FALSE | TRUE | FALSE |
| CLEC18C | 0.6524972 | 0.66508028 | 1 | 0.652497233 | 0.652497233 | 12.90495773 | 0.70642516 | 0.6631032 | 1 | 6.701920756 | 6.701920756 | 13.99020776 | 0.923660809 | -0.034487483 | FALSE | TRUE | FALSE |
| RAC1 | 0.6524275 | 0.66514771 | 1 | 0.6524275 | 0.6524275 | 0.6524275 | 0.68635163 | 0.672288423 | 1 | 0.686351633 | 0.686351633 | 0.686351633 | 0.950573247 | -0.022014413 | FALSE | TRUE | FALSE |
| CST7 | 0.6521991 | 0.6652151 | 0.7466 | 1.477609645 | 5.910262546 | 5.910262546 | 0.20320851 | 0.851349227 | 0.50665 | 1.851364946 | 1.851364946 | 1.851364946 | 3.209507 | 0.506438327 | FALSE | TRUE | FALSE |
| LYSMD2 | 0.652133 | 0.66528247 | 1 | 59.37580011 | 588.6914826 | 588.6914826 | 0.68576096 | 0.67256695 | 1 | 6.465408349 | 620.8815788 | 620.8815788 | 0.950962513 | -0.021836603 | FALSE | TRUE | FALSE |
| PURA | 0.6516408 | 0.66534981 | 1 | 59.79938819 | 6072.804383 | 6072.804383 | 0.64306817 | 0.693209221 | 0.98455 | 585.1140476 | 62230.39504 | 665026.2803 | 1.013330865 | 0.005751271 | FALSE | TRUE | FALSE |
| ME1 | 0.6515165 | 0.66541713 | 0.5087 | 0.651516455 | 5.869570182 | 5.869570182 | 0.54783591 | 0.734271483 | 0.50885 | 0.547835911 | 0.547835911 | 1.197392931 | 1.189254742 | 0.075274892 | FALSE | TRUE | FALSE |
| EIF4E2 | 0.6515062 | 0.66548441 | 1 | 0.651506202 | 0.651506202 | 0.651506202 | 0.68543772 | 0.672706021 | 1 | 0.685437718 | 0.685437718 | 0.685437718 | 0.950496574 | -0.022049444 | FALSE | TRUE | FALSE |
| SEC11C | 0.6514315 | 0.66555166 | 1 | 0.651431533 | 0.651431533 | 0.651431533 | 0.68530334 | 0.672844966 | 1 | 0.685303345 | 0.685303345 | 0.685303345 | 0.950573987 | -0.022014074 | FALSE | TRUE | FALSE |
| RBM17 | 0.6512993 | 0.66561889 | 1 | 0.651299258 | 0.651299258 | 12.9162785 | 0.68538613 | 0.67277551 | 1 | 0.685386131 | 0.685386131 | 13.60604186 | 0.950266177 | -0.022154729 | FALSE | TRUE | FALSE |
| UBE2M | 0.6512841 | 0.66568609 | 1 | 0.651284066 | 0.651284066 | 0.651284066 | 0.68504278 | 0.672983785 | 1 | 0.685042776 | 0.685042776 | 0.685042776 | 0.950720288 | -0.021947238 | FALSE | TRUE | FALSE |
| CGB1 | 0.6510531 | 0.66575325 | 1 | 0.651053132 | 0.651053132 | 0.651053132 | 6.26246818 | 0.261873123 | 0.5035 | 6.262468176 | 6.262468176 | 6.262468176 | 0.103961108 | -0.983129099 | FALSE | FALSE | FALSE |
| CTDNEP1 | 0.650805 | 0.6658204 | 1 | 0.650805026 | 0.650805026 | 0.650805026 | 0.66662469 | 0.682335121 | 1 | 0.666624686 | 0.666624686 | 0.666624686 | 0.976269016 | -0.010430494 | FALSE | TRUE | FALSE |
| ZNF24 | 0.6505697 | 0.66588751 | 1 | 0.650569683 | 0.650569683 | 12.92461077 | 0.68464558 | 0.67305315 | 1 | 0.68464558 | 0.68464558 | 0.68464558 | 0.950228412 | -0.022171988 | FALSE | TRUE | FALSE |
| POU4F2 | 0.6505151 | 0.66595459 | 0.8773 | 2.601705013 | 10.40799931 | 12.84088114 | 0.20861532 | 0.8500661 | 1 | 0.20861532 | 0.20861532 | 4.124427145 | 3.118251848 | 0.493911188 | FALSE | TRUE | FALSE |
| TMEM179 | 0.6500672 | 0.66602165 | 1 | 6.122657523 | 6.122657523 | 6.122657523 | 0.70584376 | 0.66325093 | 1 | 0.705843763 | 0.705843763 | 0.705843763 | 0.920978946 | -0.035750298 | FALSE | TRUE | FALSE |
| FNDC5 | 0.6500364 | 0.66608868 | 1 | 0.650036396 | 0.650036396 | 0.650036396 | 6.23542599 | 0.262192185 | 0.004 | 6.235425995 | 6.235425995 | 6.235425995 | 0.104248915 | -0.981928456 | FALSE | FALSE | FALSE |
| PECR | 0.649881 | 0.66615569 | 0.00915 | 0.649881025 | 0.649881025 | 0.649881025 | 0.20304573 | 0.851376587 | 0.00855 | 0.203045734 | 0.203045734 | 0.203045734 | 3.200663285 | 0.505239988 | FALSE | TRUE | FALSE |
| PPY | 0.6497836 | 0.66622266 | 1 | 0.649783638 | 0.649783638 | 0.649783638 | 0.68350525 | 0.673330322 | 1 | 0.683505252 | 0.683505252 | 0.683505252 | 0.950663709 | -0.021973085 | FALSE | TRUE | FALSE |
| TXNL1 | 0.6496595 | 0.6662896 | 1 | 6.133741027 | 59.43590132 | 59.43590132 | 0.68368644 | 0.673261075 | 1 | 6.459600069 | 62.63794027 | 62.63794027 | 0.950230186 | -0.022171177 | FALSE | TRUE | FALSE |
| ODF1 | 0.6494983 | 0.66635652 | 0.87405 | 2.597752676 | 24.25667568 | 24.25667568 | 0.21242986 | 0.848987456 | 0.0085 | 0.212429856 | 0.212429856 | 0.212429856 | 3.057472052 | 0.485362496 | FALSE | TRUE | FALSE |
| ARID4B | 0.6494147 | 0.66642341 | 0.5141 | 0.649414655 | 0.649414655 | 12.71483006 | 0.20330662 | 0.851335543 | 1 | 0.203306616 | 0.203306616 | 3.990616255 | 3.194262276 | 0.504370573 | FALSE | TRUE | FALSE |
| PMP2 | 0.6492298 | 0.66649027 | 0.0033 | 0.649229823 | 0.649229823 | 0.649229823 | 0.68307127 | 0.673676085 | 0.00205 | 0.683071268 | 0.683071268 | 0.683071268 | 0.950456933 | -0.022067557 | FALSE | TRUE | FALSE |
| GSKIP | 0.6491195 | 0.6665571 | 1 | 0.649119488 | 0.649119488 | 0.649119488 | 0.68307826 | 0.673606997 | 1 | 0.683078261 | 0.683078261 | 0.683078261 | 0.950285678 | -0.022145816 | FALSE | TRUE | FALSE |
| ANP32A | 0.648636 | 0.66662391 | 1 | 0.648635994 | 0.648635994 | 0.648635994 | 0.68285353 | 0.673745143 | 1 | 0.682853527 | 0.682853527 | 0.682853527 | 0.949890376 | -0.022326512 | FALSE | TRUE | FALSE |
| SPI1 | 0.6485403 | 0.66669068 | 1 | 0.64854029 | 1.4156E+303 | 2.8222E+306 | 1.289E+279 | 0 | 0.3276 | 3.7681E+272 | 1.6678E+297 | 3.333E+305 | 5.0321E-280 | -279.298255 | FALSE | FALSE | FALSE |
| GCM1 | 0.6483584 | 0.66675744 | 1 | 0.648358431 | 0.648358431 | 12.85226536 | 0.68270227 | 0.67381417 | 1 | 0.682702266 | 0.682702266 | 0.682702266 | 0.949694271 | -0.022416182 | FALSE | TRUE | FALSE |
| SUN3 | 0.6482502 | 0.66682416 | 0.0093 | 0.648250229 | 0.648250229 | 0.648250229 | 0.09816847 | 0.876430275 | 1 | 0.882211059 | 0.882211059 | 0.882211059 | 6.60344609 | 0.819770637 | FALSE | TRUE | FALSE |
| RELL1 | 0.6481724 | 0.66689085 | 1 | 0.648172352 | 0.648172352 | 0.648172352 | 0.68219111 | 0.67388317 | 1 | 0.682191109 | 0.682191109 | 0.682191109 | 0.950133098 | -0.022215553 | FALSE | TRUE | FALSE |
| DDX6 | 0.6481423 | 0.66695751 | 1 | 0.648142342 | 0.648142342 | 12.84890995 | 0.66033534 | 0.685560888 | 1 | 6.253412833 | 6.253412833 | 13.10446489 | 0.981535146 | -0.008094145 | FALSE | TRUE | FALSE |
| MRPL33 | 0.6481271 | 0.66702415 | 0.00185 | 0.6481271 | 0.6481271 | 0.6481271 | 0.68196515 | 0.674089979 | 1 | 0.68196515 | 0.68196515 | 0.68196515 | 0.950381555 | -0.022102001 | FALSE | TRUE | FALSE |
| FRAT1 | 0.6478803 | 0.66709076 | 1 | 0.647880317 | 0.647880317 | 0.647880317 | 0.68180633 | 0.674158853 | 1 | 0.681806334 | 0.681806334 | 0.681806334 | 0.950240977 | -0.022166246 | FALSE | TRUE | FALSE |
| UBE2Q1 | 0.6478508 | 0.66715734 | 1 | 0.647850821 | 0.647850821 | 12.86179256 | 0.68211136 | 0.673952138 | 1 | 0.682111361 | 0.682111361 | 0.682111361 | 0.949772806 | -0.022380269 | FALSE | TRUE | FALSE |
| PINX1 | 0.6478215 | 0.66722388 | 0.2594 | 0.162030647 | 0.647821505 | 0.647821505 | 8.63181114 | 0.216186241 | 0.0031 | 0.254409661 | 2.330514169 | 2.330514169 | 0.075050473 | -1.124646569 | FALSE | FALSE | FALSE |
| EIF4EBP2 | 0.6476123 | 0.66729041 | 1 | 2.590342729 | 2.590342729 | 2.590342729 | 0.68929886 | 0.671028919 | 1 | 0.689298859 | 0.689298859 | 0.689298859 | 0.939523295 | -0.027092447 | FALSE | TRUE | FALSE |
| TMEM150C | 0.6475832 | 0.6673569 | 1 | 0.647583211 | 0.647583211 | 0.647583211 | 0.68151847 | 0.674434032 | 1 | 0.681518466 | 0.681518466 | 0.681518466 | 0.950206405 | -0.022182047 | FALSE | TRUE | FALSE |
| MRPL47 | 0.6475656 | 0.66742336 | 0.7513 | 0.647565558 | 5.920375964 | 5.920375964 | 0.90835565 | 0.597177874 | 0.50495 | 0.908355654 | 0.908355654 | 0.908355654 | 0.712898692 | -0.146972182 | FALSE | TRUE | FALSE |
| FIZ1 | 0.6474166 | 0.6674898 | 1 | 0.647416609 | 0.647416609 | 12.81108054 | 0.68178776 | 0.674227695 | 1 | 0.681787757 | 0.681787757 | 13.50842019 | 0.949586733 | -0.022465362 | FALSE | TRUE | FALSE |
| SRSF2 | 0.647336 | 0.66755621 | 1 | 0.647336023 | 0.647336023 | 12.80986546 | 0.68167118 | 0.674296505 | 1 | 0.681671177 | 0.681671177 | 13.5064629 | 0.949630915 | -0.022445155 | FALSE | TRUE | FALSE |
| GABPB1 | 0.6472374 | 0.66762258 | 1 | 0.647237374 | 0.647237374 | 12.85710527 | 0.68155162 | 0.674365285 | 1 | 0.681551622 | 0.681551622 | 0.681551622 | 0.949652752 | -0.022435169 | FALSE | TRUE | FALSE |
| SAMD11 | 0.647237 | 0.66768893 | 0.68705 | 0.647237025 | 52.8292806 | 52.8292806 | 1.16539052 | 0.571051416 | 0.51305 | 1.165390517 | 10.53941783 | 10.53941783 | 0.555382093 | -0.255408127 | FALSE | FALSE | FALSE |
| TBL1XR1 | 0.6469989 | 0.66775526 | 1 | 0.646998914 | 0.646998914 | 12.84095225 | 13.5356478 | 0.165836507 | 0.0085 | 0.681374505 | 0.681374505 | 0.681374505 | 0.047799627 | -1.320575495 | FALSE | FALSE | FALSE |
| DAZL | 0.6467427 | 0.66782155 | 1 | 0.646742737 | 0.646742737 | 0.646742737 | 0.68325408 | 0.673399539 | 1 | 0.683254081 | 0.683254081 | 0.683254081 | 0.946562568 | -0.023850674 | FALSE | TRUE | FALSE |
| HNRNPH1 | 0.6466985 | 0.66788782 | 1 | 0.646698525 | 0.646698525 | 12.81494813 | 0.68111024 | 0.674571438 | 1 | 0.68111024 | 0.68111024 | 13.51178781 | 0.949477026 | -0.022515539 | FALSE | TRUE | FALSE |
| CENPA | 0.6464836 | 0.66795406 | 1 | 0.646483559 | 0.646483559 | 0.646483559 | 0.68057819 | 0.674845882 | 1 | 0.680578188 | 0.680578188 | 0.680578188 | 0.949903435 | -0.022320542 | FALSE | TRUE | FALSE |
| RPL6 | 0.6463972 | 0.66802027 | 1 | 0.64639725 | 0.64639725 | 0.64639725 | 0.68080406 | 0.674640096 | 1 | 0.680804059 | 0.680804059 | 0.680804059 | 0.94946151 | -0.022522636 | FALSE | TRUE | FALSE |
| STARD3NL | 0.6461788 | 0.66808646 | 1 | 0.646178816 | 0.646178816 | 0.646178816 | 0.68047415 | 0.674982915 | 1 | 0.680474146 | 0.680474146 | 0.680474146 | 0.949600833 | -0.022458913 | FALSE | TRUE | FALSE |

| | | | | | | | | | | | | | | | | | |
|---|---|---|---|---|---|---|---|---|---|---|---|---|---|---|---|---|---|
| COX5A | 0.646098 | 0.66815261 | 1 | 0.646098008 | 0.646098008 | 0.646098008 | 0.6804596 | 0.675051383 | 1 | 0.680459604 | 0.680459604 | 0.680459604 | 0.949502371 | -0.022503946 | FALSE | TRUE | FALSE |
| WDR82 | 0.6460064 | 0.66821874 | 1 | 6.100599478 | 6.100599478 | 12.82808306 | 0.70237021 | 0.66566832 | 1 | 0.702370212 | 0.702370212 | 0.702370212 | 0.919751927 | -0.036329294 | FALSE | TRUE | FALSE |
| RPL7A | 0.645726 | 0.66828484 | 1 | 0.645725961 | 0.645725961 | 0.645725961 | 0.68020143 | 0.675188227 | 1 | 0.680201428 | 0.680201428 | 0.680201428 | 0.949315797 | -0.022589292 | FALSE | TRUE | FALSE |
| NOL12 | 0.6455837 | 0.66835092 | 0.5037 | 0.645583652 | 0.645583652 | 0.645583652 | 0.20254226 | 0.851526891 | 1 | 0.202542256 | 0.202542256 | 0.202542256 | 3.1874023 | 0.503436882 | FALSE | TRUE | FALSE |
| KLF9 | 0.6454776 | 0.66841696 | 1 | 0.645477625 | 0.645477625 | 12.81192855 | 6.561772 | 0.252870844 | 0.50095 | 6.561772003 | 6.561772003 | 6.561772003 | 0.098369408 | -1.007139944 | FALSE | FALSE | FALSE |
| CYP3A7 | 0.6451357 | 0.66848298 | 1 | 6.108998743 | 59.37060203 | 59.37060203 | 0.70155982 | 0.665886255 | 1 | 6.651874292 | 6.651874292 | 13.91847863 | 0.919573334 | -0.036413631 | FALSE | TRUE | FALSE |
| BRF2 | 0.6450763 | 0.66854898 | 0.7556 | 0.645076316 | 5.893729095 | 12.7677847 | 0.10275433 | 0.875252508 | 0.50555 | 0.411013617 | 0.411013617 | 2.036318957 | 6.277850647 | 0.797810979 | FALSE | TRUE | FALSE |
| PISD | 0.6450578 | 0.66861494 | 0.5098 | 0.645057795 | 2.580707434 | 3.191035528 | 0.06052074 | 0.884623017 | 1 | 0.060520736 | 0.060520736 | 1.199020749 | 10.65845927 | 1.02769443 | FALSE | TRUE | FALSE |
| ZNF385A | 0.6449796 | 0.66868087 | 1 | 0.644979563 | 0.644979563 | 0.644979563 | 0.67938663 | 0.675324958 | 0.0088 | 0.679386626 | 0.679386626 | 0.679386626 | 0.949355696 | -0.02257104 | FALSE | TRUE | FALSE |
| MAP1LC3A | 0.6448996 | 0.66874678 | 1 | 6.073669528 | 6.073669528 | 6.073669528 | 0.70082463 | 0.666248819 | 1 | 0.70082463 | 0.70082463 | 0.70082463 | 0.920201143 | -0.036117231 | FALSE | TRUE | FALSE |
| RPL13 | 0.6448748 | 0.66881266 | 1 | 0.644874771 | 0.644874771 | 0.644874771 | 0.67932217 | 0.675461566 | 1 | 0.679322169 | 0.679322169 | 0.679322169 | 0.949291515 | -0.022600401 | FALSE | TRUE | FALSE |
| CCDC117 | 0.6447022 | 0.66887851 | 1 | 0.644702226 | 0.644702226 | 0.644702226 | 0.67891059 | 0.675598051 | 1 | 0.678910587 | 0.678910587 | 0.678910587 | 0.949612861 | -0.022453412 | FALSE | TRUE | FALSE |
| ARF4 | 0.6446845 | 0.66894433 | 1 | 0.6446845 | 0.6446845 | 0.6446845 | 0.6752372 | 0.677293817 | 1 | 0.675237197 | 0.675237197 | 0.675237197 | 0.954752646 | -0.020109129 | FALSE | TRUE | FALSE |
| PSMG3 | 0.6445958 | 0.66901012 | 1 | 0.644595782 | 0.644595782 | 0.644595782 | 0.67872257 | 0.67587065 | 1 | 0.678722566 | 0.678722566 | 0.678722566 | 0.949719096 | -0.022404829 | FALSE | TRUE | FALSE |
| MAPRE1 | 0.6443129 | 0.66907589 | 1 | 0.644312879 | 0.644312879 | 0.644312879 | 0.67880954 | 0.675734413 | 1 | 0.678809542 | 0.678809542 | 0.678809542 | 0.949180644 | -0.022651127 | FALSE | TRUE | FALSE |
| CSNK2A2 | 0.6442459 | 0.66914163 | 1 | 0.644245937 | 0.644245937 | 12.79316712 | 0.67723109 | 0.676685502 | 1 | 0.677231093 | 0.677231093 | 0.677231093 | 0.951294091 | -0.021685201 | FALSE | TRUE | FALSE |
| JMJD4 | 0.6442017 | 0.66920734 | 0.5045 | 0.644201661 | 0.644201661 | 2.43097651 | 0.10023942 | 0.875878058 | 1 | 0.100239423 | 0.100239423 | 1.981456004 | 6.426629802 | 0.807983284 | FALSE | TRUE | FALSE |
| RPL27A | 0.6441636 | 0.66927303 | 1 | 0.644163594 | 0.644163594 | 0.644163594 | 0.6690091 | 0.680761486 | 1 | 0.669009105 | 0.669009105 | 0.669009105 | 0.962862224 | -0.016435852 | FALSE | TRUE | FALSE |
| DAD1 | 0.6440484 | 0.66933868 | 1 | 0.644048377 | 0.644048377 | 0.644048377 | 0.67832314 | 0.676006767 | 1 | 0.678323138 | 0.678323138 | 0.678323138 | 0.949471338 | -0.022518141 | FALSE | TRUE | FALSE |
| DLEU7 | 0.6435015 | 0.66940431 | 1 | 0.643501506 | 0.643501506 | 0.643501506 | 0.65612525 | 0.686958324 | 1 | 6.16757334 | 6.16757334 | 6.16757334 | 0.980760168 | -0.008437181 | FALSE | TRUE | FALSE |
| CCND1 | 0.6433401 | 0.66946992 | 1 | 0.643340128 | 0.643340128 | 12.75689027 | 0.6778166 | 0.676278629 | 1 | 0.677816602 | 0.677816602 | 0.677816602 | 0.949135984 | -0.022671561 | FALSE | TRUE | FALSE |
| USP46 | 0.6431423 | 0.66953549 | 1 | 0.643142307 | 0.643142307 | 0.643142307 | 0.67769396 | 0.676482204 | 1 | 0.677693959 | 0.677693959 | 0.677693959 | 0.949015847 | -0.022726535 | FALSE | TRUE | FALSE |
| ELAVL2 | 0.6431406 | 0.66960104 | 1 | 0.643140607 | 0.643140607 | 0.643140607 | 0.67779328 | 0.676346519 | 1 | 0.677793276 | 0.677793276 | 0.677793276 | 0.948874281 | -0.022791325 | FALSE | TRUE | FALSE |
| KCTD2 | 0.6430776 | 0.66966656 | 1 | 0.643077613 | 0.643077613 | 0.643077613 | 0.67770214 | 0.676414377 | 1 | 0.677702142 | 0.677702142 | 0.677702142 | 0.948908929 | -0.022775467 | FALSE | TRUE | FALSE |
| TSC22D4 | 0.6430412 | 0.66973206 | 1 | 0.6430412 | 0.6430412 | 12.73198515 | 0.67769055 | 0.676549999 | 1 | 0.677690547 | 0.677690547 | 13.43419919 | 0.948871432 | -0.022792629 | FALSE | TRUE | FALSE |
| RFK | 0.6430279 | 0.66979752 | 1 | 0.643027873 | 0.643027873 | 0.643027873 | 0.67742915 | 0.676617766 | 1 | 0.677429149 | 0.677429149 | 0.677429149 | 0.949217897 | -0.022634082 | FALSE | TRUE | FALSE |
| PRR7 | 0.6427824 | 0.66986296 | 1 | 0.642782371 | 0.642782371 | 0.642782371 | 0.60814764 | 0.708475965 | 1 | 5.736867741 | 5.736867741 | 5.736867741 | 1.056951185 | 0.02405493 | FALSE | TRUE | FALSE |
| YTHDF2 | 0.6423612 | 0.66992837 | 0.0149 | 0.642361216 | 0.642361216 | 12.70144948 | 0.67714042 | 0.676753208 | 1 | 0.677140415 | 0.677140415 | 13.40741856 | 0.948638128 | -0.022899424 | FALSE | TRUE | FALSE |
| AZIN1 | 0.6421417 | 0.66999376 | 1 | 0.642141694 | 0.642141694 | 12.74756423 | 0.67683392 | 0.676820886 | 1 | 0.676833921 | 0.676833921 | 13.44824747 | 0.948743367 | -0.022851247 | FALSE | TRUE | FALSE |
| TADA1 | 0.6419327 | 0.67005912 | 0.50115 | 0.641932736 | 0.641932736 | 0.641932736 | 0.71969463 | 0.656000862 | 0.5015 | 0.719694632 | 0.719694632 | 0.719694632 | 0.891951541 | -0.04965874 | FALSE | TRUE | FALSE |
| KRI1 | 0.6418378 | 0.67012445 | 0.2622 | 0.074154553 | 0.641837782 | 1.457501567 | 0.01581334 | 0.894903165 | 1 | 0.015813342 | 0.015813342 | 0.311455557 | 40.58837137 | 1.608401625 | FALSE | TRUE | FALSE |
| COX6B1 | 0.6415259 | 0.67018976 | 1 | 0.641525877 | 0.641525877 | 0.641525877 | 0.6761482 | 0.677023743 | 1 | 0.676148202 | 0.676148202 | 0.676148202 | 0.948794769 | -0.022827718 | FALSE | TRUE | FALSE |
| PPP4R2 | 0.6412636 | 0.67025504 | 1 | 0.641263596 | 0.641263596 | 0.641263596 | 6.0137956 | 0.269159726 | 0.5026 | 6.013795605 | 6.013795605 | 6.013795605 | 0.10663209 | -0.972112077 | FALSE | FALSE | FALSE |
| DYNC1LI2 | 0.641178 | 0.67032029 | 1 | 0.641178023 | 0.641178023 | 12.69430861 | 0.67600143 | 0.677091303 | 1 | 0.676001433 | 0.676001433 | 13.39998388 | 0.948486188 | -0.022968989 | FALSE | TRUE | FALSE |
| BLOC1S3 | 0.6410861 | 0.67038552 | 1 | 6.031526226 | 6.031526226 | 6.031526226 | 0.70014308 | 0.666321236 | 1 | 0.700143077 | 0.700143077 | 0.700143077 | 0.915650161 | -0.038270424 | FALSE | TRUE | FALSE |
| FAM229B | 0.6410583 | 0.67045072 | 1 | 0.641058313 | 0.641058313 | 0.641058313 | 0.67529175 | 0.677226342 | 1 | 0.675291748 | 0.675291748 | 0.675291748 | 0.949305711 | -0.022593906 | FALSE | TRUE | FALSE |
| ARL4A | 0.640913 | 0.67051589 | 1 | 0.640913042 | 0.640913042 | 0.640913042 | 0.67564967 | 0.677158837 | 1 | 0.67564967 | 0.67564967 | 0.67564967 | 0.948587812 | -0.02292246 | FALSE | TRUE | FALSE |
| KCTD17 | 0.64076 | 0.67058103 | 1 | 0.640759982 | 0.640759982 | 0.640759982 | 6.17263918 | 0.263785184 | 0.494 | 0.654324575 | 6.172639181 | 6.172639181 | 0.103806486 | -0.983775511 | FALSE | FALSE | FALSE |
| URAD | 0.6407544 | 0.67064615 | 1 | 0.640754434 | 0.640754434 | 0.640754434 | 0.67352149 | 0.677899755 | 1 | 0.673521486 | 0.673521486 | 0.673521486 | 0.951349655 | -0.021659835 | FALSE | TRUE | FALSE |
| RAP1A | 0.6405028 | 0.67071124 | 1 | 0.640502849 | 0.640502849 | 0.640502849 | 0.67504568 | 0.677428675 | 1 | 0.675045683 | 0.675045683 | 0.675045683 | 0.948828894 | -0.022812099 | FALSE | TRUE | FALSE |
| GLRX5 | 0.6404437 | 0.6707763 | 1 | 0.64044368 | 0.64044368 | 0.64044368 | 0.66538243 | 0.682790965 | 1 | 0.665382431 | 0.665382431 | 0.665382431 | 0.962519673 | -0.016590385 | FALSE | TRUE | FALSE |
| YPEL4 | 0.6401778 | 0.67084133 | 1 | 0.640177817 | 0.640177817 | 0.640177817 | 0.67478584 | 0.677563411 | 1 | 0.674785842 | 0.674785842 | 0.674785842 | 0.94871258 | -0.022865341 | FALSE | TRUE | FALSE |
| HCRTR1 | 0.6397252 | 0.67090635 | 0.5087 | 0.639725228 | 0.639725228 | 1.406680208 | 0.09949876 | 0.876062651 | 0.0119 | 0.099498758 | 0.099498758 | 1.97087106 | 6.429479545 | 0.808175819 | FALSE | TRUE | FALSE |
| EPHX3 | 0.6396785 | 0.67097133 | 0.5042 | 0.639678543 | 0.639678543 | 0.639678543 | 0.09985793 | 0.875946127 | 1 | 0.099857926 | 0.099857926 | 0.099857926 | 6.405886509 | 0.80657924 | FALSE | TRUE | FALSE |
| NKAIN2 | 0.6395293 | 0.67103629 | 1 | 0.639529332 | 0.639529332 | 0.639529332 | 0.67410101 | 0.677765305 | 1 | 0.674101011 | 0.674101011 | 0.674101011 | 0.948714394 | -0.02286451 | FALSE | TRUE | FALSE |
| MPEG1 | 0.6393005 | 0.67110122 | 0.947 | 115.0127276 | 41728.5868 | 411560.0301 | 0.59644008 | 0.713756879 | 0.99675 | 4613.772789 | 4858910.354 | 51648652.91 | 1.071860372 | 0.030138215 | FALSE | TRUE | FALSE |
| FGF10 | 0.6393 | 0.67116612 | 1 | 0.639300049 | 0.639300049 | 0.639300049 | 0.6741905 | 0.677698038 | 1 | 0.674190503 | 0.674190503 | 0.674190503 | 0.948248375 | -0.023077893 | FALSE | TRUE | FALSE |
| PIP | 0.6390744 | 0.671231 | 0.5034 | 0.639074445 | 0.639074445 | 0.639074445 | 0.08566084 | 0.879022893 | 1 | 0.08566084 | 0.08566084 | 0.08566084 | 7.460520443 | 0.872769125 | FALSE | TRUE | FALSE |
| SLC20A1 | 0.6389914 | 0.67129585 | 0.632 | 1.097715661 | 10.21173496 | 40.83801095 | 0.11847104 | 0.871600098 | 0.0159 | 0.118471035 | 0.118471035 | 1.003415001 | 5.393650568 | 0.731882807 | FALSE | TRUE | FALSE |
| CRKL | 0.638938 | 0.67136067 | 1 | 0.638938016 | 0.638938016 | 0.638938016 | 6.18282355 | 0.263466971 | 0.5022 | 6.18282355 | 6.18282355 | 6.18282355 | 0.103340814 | -0.985728123 | FALSE | FALSE | FALSE |
| CENPW | 0.6389088 | 0.67142546 | 1 | 0.638908774 | 0.638908774 | 0.638908774 | 0.67337101 | 0.678101204 | 1 | 0.673371011 | 0.673371011 | 0.673371011 | 0.948821325 | -0.022815563 | FALSE | TRUE | FALSE |
| SEPHS1 | 0.6388889 | 0.67149023 | 1 | 0.638888882 | 0.638888882 | 12.66224437 | 0.67385252 | 0.677832544 | 1 | 0.673852519 | 0.673852519 | 13.36971819 | 0.948113814 | -0.023139526 | FALSE | TRUE | FALSE |
| CCNC | 0.6388142 | 0.67155497 | 1 | 6.03790615 | 6.03790615 | 6.03790615 | 0.66428921 | 0.683245457 | 1 | 6.282373168 | 6.282373168 | 6.282373168 | 0.961650729 | -0.016982635 | FALSE | TRUE | FALSE |
| TCTA | 0.6387249 | 0.67161968 | 1 | 0.638724925 | 0.638724925 | 0.638724925 | 0.67321406 | 0.678168293 | 1 | 0.673214065 | 0.673214065 | 0.673214065 | 0.948769431 | -0.022839316 | FALSE | TRUE | FALSE |
| KLHDC3 | 0.6384551 | 0.67168436 | 1 | 0.638455067 | 0.638455067 | 12.64470779 | 0.67341825 | 0.678034085 | 1 | 0.673418251 | 0.673418251 | 13.35278775 | 0.948081028 | -0.023154544 | FALSE | TRUE | FALSE |
| DMRT1 | 0.6384283 | 0.67174902 | 1 | 0.638428306 | 0.638428306 | 12.6774345 | 0.66489559 | 0.682920962 | 1 | 0.664895595 | 0.664895595 | 0.664895595 | 0.960193315 | -0.017641322 | FALSE | TRUE | FALSE |
| LAMTOR4 | 0.6383975 | 0.67181365 | 1 | 0.638397489 | 0.638397489 | 0.638397489 | 0.67289996 | 0.678302383 | 1 | 0.67289996 | 0.67289996 | 0.67289996 | 0.948725706 | -0.022859332 | FALSE | TRUE | FALSE |
| IL5 | 0.6383303 | 0.67187825 | 1 | 5.999610385 | 5.999610385 | 5.999610385 | 0.69438458 | 0.668407061 | 1 | 0.694384584 | 0.694384584 | 0.694384584 | 0.919274929 | -0.036554584 | FALSE | TRUE | FALSE |
| NPPC | 0.6382755 | 0.67194282 | 1 | 0.63827549 | 0.63827549 | 0.63827549 | 0.67289631 | 0.678369382 | 1 | 0.672896309 | 0.672896309 | 0.672896309 | 0.948549548 | -0.022939979 | FALSE | TRUE | FALSE |
| AADACL3 | 0.6378595 | 0.67200737 | 1 | 0.637859505 | 0.637859505 | 12.66551751 | 0.6646765 | 0.683050846 | 1 | 0.664676501 | 0.664676501 | 0.664676501 | 0.959654064 | -0.017885293 | FALSE | TRUE | FALSE |
| S100A16 | 0.6378262 | 0.6720719 | 1 | 6.0019541 | 6.0019541 | 6.0019541 | 0.69391089 | 0.668549915 | 1 | 0.693910889 | 0.693910889 | 0.693910889 | 0.919176016 | -0.036601316 | FALSE | TRUE | FALSE |
| MEX3A | 0.6377717 | 0.67213639 | 1 | 0.637771711 | 0.637771711 | 12.59488034 | 0.67060014 | 0.679769442 | 1 | 0.67060014 | 0.67060014 | 13.26324225 | 0.951046193 | -0.021798389 | FALSE | TRUE | FALSE |
| RPS3 | 0.637471 | 0.67220086 | 1 | 0.637470979 | 0.637470979 | 0.637470979 | 0.67244605 | 0.678703925 | 1 | 0.67244605 | 0.67244605 | 0.67244605 | 0.947988288 | -0.023197028 | FALSE | TRUE | FALSE |
| ZNF22 | 0.6373949 | 0.6722653 | 1 | 0.637394874 | 0.637394874 | 0.637394874 | 0.67224411 | 0.678770745 | 0.0057 | 0.672244106 | 0.672244106 | 0.672244106 | 0.948159855 | -0.023118437 | FALSE | TRUE | FALSE |
| SRSF9 | 0.6373775 | 0.67232972 | 1 | 6.031798138 | 6.031798138 | 6.031798138 | 0.68516744 | 0.672914391 | 1 | 0.685167437 | 0.685167437 | 0.685167437 | 0.930250789 | -0.031399953 | FALSE | TRUE | FALSE |
| FAM110A | 0.6372526 | 0.67239411 | 1 | 0.63725256 | 0.63725256 | 0.63725256 | 0.67222863 | 0.678837534 | 1 | 0.672228635 | 0.672228635 | 0.672228635 | 0.947969972 | -0.023205419 | FALSE | TRUE | FALSE |
| VEZF1 | 0.6370089 | 0.67245847 | 1 | 0.637008873 | 0.637008873 | 12.61812082 | 0.67213842 | 0.678904292 | 1 | 0.672138415 | 0.672138415 | 13.32932309 | 0.947734662 | -0.023313236 | FALSE | TRUE | FALSE |
| FAM89B | 0.6366749 | 0.67252281 | 0.0022 | 0.636674856 | 0.636674856 | 0.636674856 | 0.6712247 | 0.679503767 | 1 | 0.671224696 | 0.671224696 | 0.671224696 | 0.948527162 | -0.022950228 | FALSE | TRUE | FALSE |
| PLEKHF2 | 0.6365521 | 0.67258712 | 1 | 0.636552114 | 0.636552114 | 0.636552114 | 0.67164496 | 0.679370757 | 1 | 0.671644962 | 0.671644962 | 0.671644962 | 0.947750894 | -0.023305797 | FALSE | TRUE | FALSE |
| BTC | 0.6365325 | 0.6726514 | 1 | 0.636532545 | 0.636532545 | 0.636532545 | 0.67135197 | 0.679437276 | 1 | 0.671351969 | 0.671351969 | 0.671351969 | 0.948135367 | -0.023129653 | FALSE | TRUE | FALSE |
| FXYD5 | 0.6364244 | 0.67271566 | 1 | 0.636424409 | 0.636424409 | 0.636424409 | 2.62600422 | 0.438624986 | 0.74995 | 2.626004221 | 10.50607541 | 10.50607541 | 0.242354679 | -0.615548592 | FALSE | FALSE | FALSE |
| BBS5 | 0.6364072 | 0.67277989 | 0.6295 | 0.636407175 | 23.32323481 | 23.32323481 | 0.02998873 | 0.891236561 | 1 | 0.029988733 | 0.269893095 | 0.269893095 | 21.22154241 | 1.326776946 | FALSE | TRUE | FALSE |
| SCGB2A2 | 0.6362456 | 0.67284409 | 0.0023 | 0.636245646 | 0.636245646 | 0.636245646 | 0.67082638 | 0.679570231 | 1 | 0.670826385 | 0.670826385 | 0.670826385 | 0.948450539 | -0.022985312 | FALSE | TRUE | FALSE |

| | | | | | | | | | | | | | | | | | |
|---|---|---|---|---|---|---|---|---|---|---|---|---|---|---|---|---|---|
| S100B | 0.6360974 | 0.67290827 | 1 | 0.636097397 | 0.636097397 | 0.636097397 | 0.67069225 | 0.679703068 | 1 | 0.670692247 | 0.670692247 | 0.670692247 | 0.948419189 | -0.022999668 | FALSE | TRUE | FALSE |
| KLRD1 | 0.6359936 | 0.67297242 | 1 | 0.635993621 | 0.635993621 | 0.635993621 | 22.7795459 | 0.119138692 | 0.0012 | 0.670796983 | 0.670796983 | 0.670796983 | 0.027919504 | -1.554092302 | FALSE | FALSE | FALSE |
| ITPKA | 0.6356365 | 0.67303654 | 1 | 0.635636521 | 0.635636521 | 12.58389471 | 0.67079852 | 0.679636664 | 1 | 0.670798515 | 0.670798515 | 13.29619703 | 0.947581884 | -0.023383251 | FALSE | TRUE | FALSE |
| AKT1S1 | 0.6353967 | 0.67310064 | 1 | 0.635396736 | 0.635396736 | 0.635396736 | 0.67024581 | 0.680167058 | 1 | 0.670245814 | 0.670245814 | 0.670245814 | 0.948005527 | -0.023189131 | FALSE | TRUE | FALSE |
| KLF12 | 0.635283 | 0.67316472 | 1 | 0.635283032 | 0.635283032 | 12.58318099 | 0.67047671 | 0.679835787 | 1 | 0.67047671 | 0.67047671 | 13.29566797 | 0.947509469 | -0.023416441 | FALSE | TRUE | FALSE |
| MAPRE2 | 0.63521 | 0.67322876 | 1 | 0.635209977 | 0.635209977 | 0.635209977 | 5.93688433 | 0.271979778 | 0.75055 | 5.936884335 | 57.50357641 | 57.50357641 | 0.106993827 | -0.970641277 | FALSE | FALSE | FALSE |
| CBX6 | 0.635111 | 0.67329278 | 1 | 0.63511104 | 0.63511104 | 12.53370378 | 0.67034787 | 0.679968386 | 1 | 0.670347873 | 0.670347873 | 13.25021073 | 0.947435005 | -0.023450573 | FALSE | TRUE | FALSE |
| MXD4 | 0.6350762 | 0.67335677 | 1 | 0.635076201 | 0.635076201 | 0.635076201 | 0.67028253 | 0.680100865 | 1 | 0.670282532 | 0.670282532 | 0.670282532 | 0.947475386 | -0.023432063 | FALSE | TRUE | FALSE |
| ARID3A | 0.634959 | 0.67342074 | 1 | 0.634958986 | 0.634958986 | 12.56292653 | 0.62833217 | 0.699780922 | 1 | 5.946709022 | 57.76257505 | 117.8099659 | 1.010546683 | 0.004556381 | FALSE | TRUE | FALSE |
| GID8 | 0.6348719 | 0.67348468 | 1 | 0.634871929 | 0.634871929 | 0.634871929 | 0.67007905 | 0.680299358 | 1 | 0.670079053 | 0.670079053 | 0.670079053 | 0.947458252 | -0.023439917 | FALSE | TRUE | FALSE |
| LBX1 | 0.6346607 | 0.67354859 | 0.5079 | 0.634660708 | 0.634660708 | 3.146523214 | 0.19970771 | 0.852179385 | 1 | 0.199707709 | 0.199707709 | 3.962670022 | 3.177947962 | 0.502146781 | FALSE | TRUE | FALSE |
| TNFAIP8L2 | 0.6345864 | 0.67361248 | 1 | 0.634586439 | 0.634586439 | 0.634586439 | 0.66946638 | 0.680629591 | 1 | 0.669466379 | 0.669466379 | 0.669466379 | 0.947898892 | -0.023237984 | FALSE | TRUE | FALSE |
| CNFN | 0.6345694 | 0.67367633 | 1 | 0.634569444 | 0.634569444 | 0.634569444 | 0.66952411 | 0.680563603 | 1 | 0.669524109 | 0.669524109 | 0.669524109 | 0.947791775 | -0.023287065 | FALSE | TRUE | FALSE |
| ZNF362 | 0.6345579 | 0.67374017 | 1 | 0.634557888 | 0.634557888 | 12.5776996 | 0.66974922 | 0.680497584 | 1 | 0.669749225 | 0.669749225 | 13.28950821 | 0.947455951 | -0.023440972 | FALSE | TRUE | FALSE |
| ZBTB18 | 0.6344954 | 0.67380397 | 1 | 0.634495423 | 0.634495423 | 12.53833313 | 0.66985787 | 0.680431538 | 1 | 0.669857865 | 0.669857865 | 13.25615777 | 0.947209035 | -0.023554168 | FALSE | TRUE | FALSE |
| TRIM51 | 0.6337375 | 0.67386775 | 0.8757 | 1.417860167 | 54.21890057 | 54.21890057 | 0.10233099 | 0.875409445 | 0.5019 | 0.918898569 | 0.918898569 | 0.918898569 | 6.193016819 | 0.791902259 | FALSE | TRUE | FALSE |
| TINAGL1 | 0.6336188 | 0.67393151 | 0.5076 | 0.633618766 | 0.633618766 | 2.315802986 | 0.38260236 | 0.798246065 | 0.5077 | 0.38260236 | 0.38260236 | 1.892163046 | 1.656076473 | 0.219080388 | FALSE | TRUE | FALSE |
| WASF1 | 0.6335643 | 0.67399524 | 1 | 0.633564308 | 0.633564308 | 12.53491284 | 0.66898515 | 0.680827389 | 1 | 0.668985155 | 0.668985155 | 13.25284201 | 0.947052866 | -0.023625777 | FALSE | TRUE | FALSE |
| KCNK13 | 0.6334927 | 0.67405895 | 1 | 0.633492655 | 0.633492655 | 12.55481414 | 6.12988667 | 0.265688768 | 0.50615 | 6.129886675 | 6.129886675 | 12.85648516 | 0.103344921 | -0.985710862 | FALSE | FALSE | FALSE |
| YTHDF3 | 0.6334841 | 0.67412262 | 1 | 5.993150928 | 58.19124066 | 118.7069432 | 5.91029377 | 0.272292164 | 0.9355 | 57.30345423 | 5808.261642 | 5808.261642 | 0.107183184 | -0.969873344 | FALSE | FALSE | FALSE |
| PITPNB | 0.6333453 | 0.67418627 | 0.0073 | 0.633345262 | 0.633345262 | 0.633345262 | 0.66852818 | 0.681090709 | 1 | 0.668528177 | 0.668528177 | 0.668528177 | 0.947372577 | -0.02347919 | FALSE | TRUE | FALSE |
| RAB27B | 0.6330395 | 0.6742499 | 0.7452 | 0.633039549 | 2.531764643 | 2.531764643 | 0.79593314 | 0.622119227 | 0.4952 | 0.199053715 | 0.795933137 | 0.795933137 | 0.795342623 | -0.099445743 | FALSE | TRUE | FALSE |
| HPGD | 0.6329408 | 0.6743135 | 0.5011 | 0.632940778 | 0.632940778 | 0.632940778 | 0.82350658 | 0.613731665 | 0.8105 | 3.292764031 | 124.2279497 | 124.2279497 | 0.768592252 | -0.114303998 | FALSE | TRUE | FALSE |
| NPY | 0.6329212 | 0.67437707 | 1 | 0.632921158 | 0.632921158 | 0.632921158 | 0.66787256 | 0.68148482 | 1 | 0.667872559 | 0.667872559 | 0.667872559 | 0.94766756 | -0.023343986 | FALSE | TRUE | FALSE |
| CBLN2 | 0.6327108 | 0.67444062 | 1 | 0.632710818 | 0.632710818 | 0.632710818 | 0.66802183 | 0.681353567 | 0.00665 | 0.668021833 | 0.668021833 | 0.668021833 | 0.947140927 | -0.023585397 | FALSE | TRUE | FALSE |
| EFCAB13 | 0.6325365 | 0.67450414 | 0.2571 | 0.073341372 | 0.632536471 | 2.476586621 | 0.12620482 | 0.869826351 | 0.7617 | 0.126204819 | 1.103254521 | 2.48984556 | 5.011983519 | 0.700009634 | FALSE | TRUE | FALSE |
| POLR3K | 0.6324672 | 0.67456763 | 1 | 0.632467165 | 0.632467165 | 0.632467165 | 0.66761367 | 0.681681479 | 1 | 0.667613671 | 0.667613671 | 0.667613671 | 0.947355024 | -0.023487238 | FALSE | TRUE | FALSE |
| LEPROTL1 | 0.63229 | 0.6746311 | 1 | 0.63229001 | 5.963628554 | 5.963628554 | 0.6900148 | 0.670465803 | 1 | 0.690014797 | 0.690014797 | 0.690014797 | 0.916342683 | -0.037942084 | FALSE | TRUE | FALSE |
| GTF2A1L | 0.6322845 | 0.67469454 | 1 | 0.632284459 | 0.632284459 | 12.54723065 | 0.62850306 | 0.699607214 | 1 | 5.945252823 | 5.945252823 | 5.945252823 | 1.006016523 | 0.002605114 | FALSE | TRUE | FALSE |
| SHMT2 | 0.6321072 | 0.67475796 | 0.50975 | 0.632107173 | 0.632107173 | 1.363704729 | 0.09732248 | 0.876632557 | 1 | 0.097322483 | 0.097322483 | 1.926436247 | 6.494975842 | 0.81257754 | FALSE | TRUE | FALSE |
| FAM110B | 0.6319108 | 0.67482135 | 1 | 0.631910822 | 0.631910822 | 12.50821261 | 0.66723974 | 0.681877877 | 1 | 0.66723974 | 0.66723974 | 13.22386574 | 0.947052138 | -0.023626111 | FALSE | TRUE | FALSE |
| MYL1 | 0.6318129 | 0.67488471 | 0.9385 | 2.526958249 | 40.45226301 | 40.45226301 | 190.291275 | 0.014137335 | 0.50215 | 190.2912747 | 3050.954616 | 13043.64966 | 0.003320241 | -2.4788304 | FALSE | FALSE | FALSE |
| HLA-DMA | 0.6316237 | 0.67494805 | 1 | 0.631623716 | 0.631623716 | 0.631623716 | 0.66678315 | 0.682204624 | 1 | 0.66678315 | 0.66678315 | 0.66678315 | 0.947270062 | -0.023526188 | FALSE | TRUE | FALSE |
| SULT1C4 | 0.6314668 | 0.67501137 | 0.50425 | 0.631466846 | 0.631466846 | 0.631466846 | 0.09806405 | 0.876488132 | 1 | 0.098064048 | 0.098064048 | 0.098064048 | 6.439330816 | 0.808840737 | FALSE | TRUE | FALSE |
| RER1 | 0.6314215 | 0.67507465 | 1 | 0.631421484 | 0.631421484 | 0.631421484 | 0.66676518 | 0.682269887 | 1 | 0.666765176 | 0.666765176 | 0.666765176 | 0.946992294 | -0.023653555 | FALSE | TRUE | FALSE |
| FBXL17 | 0.6311183 | 0.67513791 | 1 | 0.631118329 | 0.631118329 | 0.631118329 | 0.65894424 | 0.686070545 | 1 | 0.658944244 | 0.658944244 | 0.658944244 | 0.957771974 | -0.018737875 | FALSE | TRUE | FALSE |
| BRK1 | 0.6310085 | 0.67520115 | 1 | 0.631008483 | 0.631008483 | 0.631008483 | 0.63623586 | 0.696444175 | 1 | 0.636235859 | 0.636235859 | 0.636235859 | 0.991783903 | -0.003582945 | FALSE | TRUE | FALSE |
| RIMS4 | 0.6309472 | 0.67526436 | 1 | 0.63094718 | 0.63094718 | 0.63094718 | 0.66635515 | 0.682530651 | 1 | 0.666355152 | 0.666355152 | 0.666355152 | 0.946863211 | -0.023712757 | FALSE | TRUE | FALSE |
| ZBTB2 | 0.6309087 | 0.67532754 | 1 | 0.630908714 | 0.630908714 | 12.47607914 | 0.66396507 | 0.683375058 | 1 | 0.663965068 | 0.663965068 | 13.14754933 | 0.950213716 | -0.022178705 | FALSE | TRUE | FALSE |
| ALYREF | 0.6306363 | 0.6753907 | 1 | 0.630636282 | 0.630636282 | 0.630636282 | 0.66226428 | 0.684536316 | 1 | 0.662264284 | 0.662264284 | 0.662264284 | 0.952242627 | -0.021252381 | FALSE | TRUE | FALSE |
| PTGES | 0.6305467 | 0.67545383 | 1 | 0.630546738 | 0.630546738 | 0.630546738 | 0.64141294 | 0.693572274 | 1 | 0.641412939 | 0.641412939 | 0.641412939 | 0.983058963 | -0.007420433 | FALSE | TRUE | FALSE |
| GUCA2A | 0.6305254 | 0.67551693 | 1 | 5.926930146 | 5.926930146 | 5.926930146 | 541.016951 | 0.004035355 | 0.31435 | 54.75917114 | 5478.901208 | 5478.901208 | 0.001165445 | -2.933508268 | FALSE | FALSE | FALSE |
| ATP1B1 | 0.6303765 | 0.67558001 | 1 | 0.630376513 | 0.630376513 | 0.630376513 | 0.65992447 | 0.68568847 | 1 | 0.659924474 | 0.659924474 | 0.659924474 | 0.955225239 | -0.019894211 | FALSE | TRUE | FALSE |
| LRRC31 | 0.6303252 | 0.67564307 | 0.75235 | 0.630325168 | 5.766515033 | 12.49530414 | 0.10174487 | 0.87553668 | 1 | 0.101744872 | 0.101744872 | 2.019087812 | 6.195154189 | 0.79205212 | FALSE | TRUE | FALSE |
| PRR13 | 0.6299741 | 0.67570609 | 1 | 5.915606241 | 57.01143249 | 57.01143249 | 0.7081775 | 0.662288266 | 1 | 0.708177499 | 0.708177499 | 0.708177499 | 0.889570968 | -0.050819399 | FALSE | TRUE | FALSE |
| SPR | 0.6298211 | 0.6757691 | 0.62685 | 1.100341934 | 10.07114956 | 10.07114956 | 0.82892251 | 0.612643965 | 0.8757 | 7.812895363 | 124.9435525 | 499.8360607 | 0.759806966 | -0.119296729 | FALSE | TRUE | FALSE |
| ATAD5 | 0.6296943 | 0.67583208 | 0.04075 | 0.629694341 | 0.629694341 | 12.23321129 | 0.09658868 | 0.876795862 | 1 | 0.096588683 | 0.096588683 | 3.245878887 | 6.519338709 | 0.814203545 | FALSE | TRUE | FALSE |
| HAPLN3 | 0.6296422 | 0.67589503 | 0.5094 | 0.62964217 | 0.62964217 | 1.384255772 | 0.09811842 | 0.87645921 | 1 | 0.098118416 | 0.098118416 | 1.943185553 | 6.417166087 | 0.80734328 | FALSE | TRUE | FALSE |
| COPS3 | 0.6294737 | 0.67595795 | 1 | 0.629473691 | 0.629473691 | 12.49256699 | 0.66509413 | 0.682855977 | 1 | 0.66509413 | 0.66509413 | 13.21171032 | 0.94644301 | -0.023905532 | FALSE | TRUE | FALSE |
| FXYD2 | 0.629148 | 0.67602085 | 1 | 0.62914804 | 0.62914804 | 0.62914804 | 0.65986829 | 0.685752218 | 1 | 0.659868292 | 0.659868292 | 0.659868292 | 0.953444873 | -0.020704412 | FALSE | TRUE | FALSE |
| CCT8L2 | 0.6289875 | 0.67608373 | 0.8759 | 5.820414364 | 55.24373011 | 55.24373011 | 0.10399981 | 0.874927633 | 0.013 | 0.103999812 | 0.103999812 | 2.059063253 | 6.047967679 | 0.781609462 | FALSE | TRUE | FALSE |
| BCL10 | 0.6289267 | 0.67614658 | 0.00695 | 0.628926709 | 0.628926709 | 0.628926709 | 6.06059552 | 0.267584853 | 0.50415 | 6.060595522 | 6.060595522 | 6.060595522 | 0.103773088 | -0.983915262 | FALSE | FALSE | FALSE |
| TMEM92 | 0.6285761 | 0.67620941 | 0.00455 | 0.628576066 | 0.628576066 | 0.628576066 | 0.66378567 | 0.683569246 | 1 | 0.663785671 | 0.663785671 | 0.663785671 | 0.946956364 | -0.023670033 | FALSE | TRUE | FALSE |
| PSORS1C2 | 0.6285703 | 0.67627221 | 1 | 0.628570342 | 0.628570342 | 0.628570342 | 0.66380476 | 0.683439817 | 0.0046 | 0.663804757 | 0.663804757 | 0.663804757 | 0.946920515 | -0.023686474 | FALSE | TRUE | FALSE |
| CDRT4 | 0.6283923 | 0.67633498 | 1 | 0.628392311 | 0.628392311 | 0.628392311 | 0.66361949 | 0.68369856 | 1 | 0.663619495 | 0.663619495 | 0.663619495 | 0.946916593 | -0.023688273 | FALSE | TRUE | FALSE |
| LSM4 | 0.6283551 | 0.67639773 | 1 | 0.628355107 | 0.628355107 | 0.628355107 | 0.66378617 | 0.683504546 | 1 | 0.663786171 | 0.663786171 | 0.663786171 | 0.946622775 | -0.023823051 | FALSE | TRUE | FALSE |
| LCE1D | 0.6282575 | 0.67646045 | 1 | 0.628257533 | 0.628257533 | 0.628257533 | 0.66336996 | 0.683827761 | 1 | 0.663369956 | 0.663369956 | 0.663369956 | 0.947069622 | -0.023618093 | FALSE | TRUE | FALSE |
| EIF2S1 | 0.6277415 | 0.67652315 | 1 | 0.627741512 | 0.627741512 | 0.627741512 | 0.66331702 | 0.683892318 | 1 | 0.663317015 | 0.663317015 | 0.663317015 | 0.946367269 | -0.023940289 | FALSE | TRUE | FALSE |
| RLBP1 | 0.6276251 | 0.67658583 | 0.50635 | 0.627625113 | 0.627625113 | 0.627625113 | 0.09404865 | 0.877340544 | 1 | 0.376029093 | 1.503830579 | 1.503830579 | 6.673409414 | 0.82434777 | FALSE | TRUE | FALSE |
| FKBP1B | 0.6276212 | 0.67664847 | 1 | 0.627621187 | 0.627621187 | 0.627621187 | 0.66305531 | 0.684214672 | 1 | 0.663055307 | 0.663055307 | 0.663055307 | 0.946559329 | -0.02385216 | FALSE | TRUE | FALSE |
| LCE2C | 0.6275855 | 0.67671109 | 1 | 0.627585529 | 0.627585529 | 0.627585529 | 0.66278981 | 0.684343413 | 1 | 0.66278981 | 0.66278981 | 0.66278981 | 0.946884699 | -0.023702901 | FALSE | TRUE | FALSE |
| RAB30 | 0.6275526 | 0.67677369 | 1 | 0.627552571 | 0.627552571 | 0.627552571 | 0.66320066 | 0.684021346 | 1 | 0.663200663 | 0.663200663 | 0.663200663 | 0.946248407 | -0.023994839 | FALSE | TRUE | FALSE |
| GP9 | 0.6275268 | 0.67683626 | 1 | 0.627526775 | 0.627526775 | 0.627526775 | 0.66315756 | 0.684085817 | 1 | 0.663157561 | 0.663157561 | 0.663157561 | 0.946271011 | -0.023984464 | FALSE | TRUE | FALSE |
| SAA2 | 0.6274749 | 0.6768988 | 1 | 5.890721817 | 56.75797709 | 56.75797709 | 0.68379461 | 0.673191797 | 1 | 6.434057774 | 6.434057774 | 6.434057774 | 0.917636558 | -0.037329293 | FALSE | TRUE | FALSE |
| CNOT2 | 0.6274602 | 0.67696131 | 1 | 0.627460226 | 0.627460226 | 12.42565942 | 0.66328122 | 0.683956846 | 1 | 0.663281218 | 0.663281218 | 13.15058079 | 0.945994262 | -0.024111498 | FALSE | TRUE | FALSE |
| GSTZ1 | 0.6271194 | 0.67702381 | 0.75035 | 5.680838695 | 52.81723936 | 52.81723936 | 0.20856937 | 0.850080009 | 1 | 0.20856937 | 0.20856937 | 0.20856937 | 3.00676666 | 0.478099726 | FALSE | TRUE | FALSE |
| FBXO8 | 0.6271023 | 0.67708627 | 1 | 0.627102264 | 0.627102264 | 0.627102264 | 0.66268319 | 0.684472041 | 1 | 0.662683194 | 0.662683194 | 0.662683194 | 0.946307784 | -0.023967588 | FALSE | TRUE | FALSE |
| TOB2 | 0.6266365 | 0.67714871 | 1 | 0.62663648 | 0.62663648 | 12.43563319 | 0.7372718 | 0.647843691 | 1 | 6.989237375 | 68.00076749 | 68.00076749 | 0.849939576 | -0.070611948 | FALSE | TRUE | FALSE |
| METRNL | 0.6265258 | 0.67721113 | 1 | 0.626525763 | 0.626525763 | 12.42265163 | 0.66216693 | 0.684600563 | 1 | 0.662166934 | 0.662166934 | 13.14293161 | 0.946174946 | -0.024028556 | FALSE | TRUE | FALSE |
| DIMT1 | 0.6264453 | 0.67727352 | 0.50215 | 0.626445294 | 0.626445294 | 0.626445294 | 0.0983597 | 0.876362707 | 1 | 0.098359695 | 0.098359695 | 0.098359695 | 6.368922681 | 0.804065977 | FALSE | TRUE | FALSE |
| MS4A15 | 0.6264024 | 0.67733589 | 0.0053 | 0.626402418 | 0.626402418 | 0.626402418 | 0.66183732 | 0.684921367 | 1 | 0.661837323 | 0.661837323 | 0.661837323 | 0.946459796 | -0.023897829 | FALSE | TRUE | FALSE |
| EFHD2 | 0.6263026 | 0.67739823 | 1 | 0.626302583 | 0.626302583 | 0.626302583 | 0.66204335 | 0.684793131 | 1 | 0.662043349 | 0.662043349 | 0.662043349 | 0.946014463 | -0.024102224 | FALSE | TRUE | FALSE |
| AGFG1 | 0.626283 | 0.67746054 | 1 | 0.626283032 | 0.626283032 | 12.39622164 | 0.66213414 | 0.684664781 | 1 | 0.662134141 | 0.662134141 | 13.12212612 | 0.945855217 | -0.024175336 | FALSE | TRUE | FALSE |

| | | | | | | | | | | | | | | | | | |
|---|---|---|---|---|---|---|---|---|---|---|---|---|---|---|---|---|---|
| RNF166 | 0.6261672 | 0.67752283 | 1 | 0.626167209 | 0.626167209 | 0.626167209 | 6.03569926 | 0.267900416 | 0.5074 | 6.035699257 | 6.035699257 | 6.035699257 | 0.103743938 | -0.984037271 | FALSE | FALSE | FALSE |
| JOSD1 | 0.6261524 | 0.67758509 | 1 | 0.626152371 | 0.626152371 | 0.626152371 | 0.66176882 | 0.684985443 | 1 | 0.661768818 | 0.661768818 | 0.661768818 | 0.946179925 | -0.02402627 | FALSE | TRUE | FALSE |
| GNPAT | 0.6261191 | 0.67764733 | 0.2574 | 0.626119115 | 2.503269032 | 5.33266608 | 0.36586425 | 0.803829626 | 0.96955 | 5.853063188 | 23.41949925 | 93.73040129 | 1.711342703 | 0.233336987 | FALSE | TRUE | FALSE |
| GTF2A1 | 0.6260345 | 0.67770954 | 1 | 0.626034467 | 0.626034467 | 0.626034467 | 0.66173889 | 0.68504949 | 1 | 0.661738892 | 0.661738892 | 0.661738892 | 0.946044542 | -0.024088415 | FALSE | TRUE | FALSE |
| PHOSPHO1 | 0.6260224 | 0.67777172 | 1 | 0.626022354 | 0.626022354 | 0.626022354 | 0.66140619 | 0.685177501 | 1 | 0.661406186 | 0.661406186 | 0.661406186 | 0.946502115 | -0.023878411 | FALSE | TRUE | FALSE |
| PPP1R8 | 0.6258224 | 0.67783388 | 1 | 5.923254564 | 57.53768559 | 57.53768559 | 0.70528085 | 0.664060901 | 1 | 0.705280849 | 0.705280849 | 0.705280849 | 0.887337815 | -0.05191101 | FALSE | TRUE | FALSE |
| WNT8A | 0.6257199 | 0.67789601 | 0.691 | 0.625719918 | 10.01258976 | 10.01258976 | 0.05986747 | 0.88475908 | 1 | 0.059867472 | 0.059867472 | 0.059867472 | 10.45175108 | 1.019189058 | FALSE | TRUE | FALSE |
| ZBTB44 | 0.6256523 | 0.67795812 | 1 | 0.625652286 | 0.625652286 | 12.40121575 | 0.640935 | 0.693874136 | 0.51095 | 6.065775356 | 6.065775356 | 12.71779396 | 0.976155596 | -0.010480952 | FALSE | TRUE | FALSE |
| ZNF415 | 0.6254871 | 0.6780202 | 0.9351 | 5.774729025 | 530.8120566 | 530.8120566 | 0.91158954 | 0.596749054 | 0.75015 | 0.911589544 | 8.483041893 | 8.483041893 | 0.686149971 | -0.163580951 | FALSE | TRUE | FALSE |
| UCN3 | 0.6253622 | 0.67808225 | 1 | 0.625362217 | 0.625362217 | 0.625362217 | 0.66080939 | 0.685369316 | 1 | 0.660809389 | 0.660809389 | 0.660809389 | 0.946357948 | -0.023944566 | FALSE | TRUE | FALSE |
| CYP4Z1 | 0.6251817 | 0.67814429 | 0.75495 | 0.6251817 | 5.765205978 | 12.38517626 | 0.88307023 | 0.600998549 | 0.0118 | 0.883070225 | 0.883070225 | 1.935366479 | 0.70796374 | -0.149988985 | FALSE | TRUE | FALSE |
| COPS2 | 0.6250735 | 0.67820629 | 1 | 0.625073467 | 0.625073467 | 0.625073467 | 0.6610303 | 0.685305404 | 1 | 0.661030295 | 0.661030295 | 0.661030295 | 0.945604871 | -0.024290299 | FALSE | TRUE | FALSE |
| C12orf56 | 0.625067 | 0.67826827 | 0.87895 | 5.689444628 | 53.11738922 | 53.11738922 | 7.58014199 | 0.234181738 | 0.6881 | 7.580141988 | 686.5955255 | 686.5955255 | 0.082461115 | -1.083750798 | FALSE | FALSE | FALSE |
| COL6A1 | 0.6250448 | 0.67833023 | 0.7687 | 9.988532449 | 1462.867412 | 5853.013013 | 2.34987413 | 0.460416871 | 0.11045 | 0.481220816 | 7.690644312 | 30.7563292 | 0.265990743 | -0.575133478 | FALSE | FALSE | FALSE |
| OVOL1 | 0.624892 | 0.67839216 | 1 | 0.624891985 | 0.624891985 | 0.624891985 | 0.66068556 | 0.6854332 | 1 | 0.660685556 | 0.660685556 | 0.660685556 | 0.945823591 | -0.024189858 | FALSE | TRUE | FALSE |
| COPRS | 0.6248099 | 0.67845406 | 1 | 0.624809911 | 0.624809911 | 0.624809911 | 6.01623916 | 0.268845353 | 0.50565 | 6.016239157 | 6.016239157 | 6.016239157 | 0.103853902 | -0.983577182 | FALSE | FALSE | FALSE |
| DEFB127 | 0.6247425 | 0.67851594 | 1 | 0.624742519 | 0.624742519 | 0.624742519 | 0.66000016 | 0.685624693 | 0.0026 | 0.660000163 | 0.660000163 | 0.660000163 | 0.946579342 | -0.023842978 | FALSE | TRUE | FALSE |
| RBBP4 | 0.6247349 | 0.67857779 | 1 | 2.498883419 | 2.498883419 | 12.39669297 | 0.66813015 | 0.681222196 | 1 | 0.668130155 | 0.668130155 | 0.668130155 | 0.935049645 | -0.02916533 | FALSE | TRUE | FALSE |
| DIS3L2 | 0.6246102 | 0.67863962 | 0.51425 | 0.624610172 | 0.624610172 | 12.24603422 | 0.09711333 | 0.87668063 | 1 | 0.097113328 | 0.097113328 | 1.908511083 | 6.431765683 | 0.808330214 | FALSE | TRUE | FALSE |
| CYTL1 | 0.62434 | 0.67870142 | 1 | 0.624339966 | 0.624339966 | 0.624339966 | 6.01124014 | 0.269473746 | 0.50895 | 6.01124014 | 6.01124014 | 6.01124014 | 0.10386209 | -0.983542941 | FALSE | FALSE | FALSE |
| KRT33A | 0.6240402 | 0.6787632 | 0.0422 | 0.156081126 | 0.624040213 | 2.495647505 | 0.17487557 | 0.858326669 | 0.25725 | 0.174875572 | 1.613289923 | 1.613289923 | 3.568481336 | 0.55248343 | FALSE | TRUE | FALSE |
| SOCS7 | 0.6238248 | 0.67882495 | 1 | 0.623824842 | 0.623824842 | 12.37158126 | 0.6598544 | 0.685815937 | 1 | 0.659854398 | 0.659854398 | 13.09934718 | 0.945397719 | -0.02438545 | FALSE | TRUE | FALSE |
| SEMA3C | 0.6236405 | 0.67888668 | 0.7571 | 0.623640504 | 2.494977502 | 12.28880006 | 0.19215472 | 0.853853196 | 1 | 1.772065167 | 1.772065167 | 6.498160152 | 3.245512321 | 0.511283262 | FALSE | TRUE | FALSE |
| TRIML2 | 0.6235087 | 0.67894839 | 0.50835 | 0.623508657 | 0.623508657 | 0.623508657 | 0.09778318 | 0.876517042 | 1 | 0.097783176 | 0.097783176 | 0.097783176 | 6.376441072 | 0.80457835 | FALSE | TRUE | FALSE |
| S100A11 | 0.6232698 | 0.67901007 | 1 | 0.623269766 | 0.623269766 | 0.623269766 | 0.65865243 | 0.686197692 | 1 | 0.658652426 | 0.658652426 | 0.658652426 | 0.946280225 | -0.023980236 | FALSE | TRUE | FALSE |
| NPR3 | 0.6229461 | 0.67907173 | 0.5083 | 0.622946129 | 0.622946129 | 2.274188625 | 0.09525084 | 0.87708309 | 1 | 0.095250836 | 0.095250836 | 1.883254695 | 6.54005945 | 0.815581696 | FALSE | TRUE | FALSE |
| ADIRF | 0.6228499 | 0.67913337 | 1 | 0.622849899 | 0.622849899 | 0.622849899 | 0.65828162 | 0.686451656 | 1 | 0.658281621 | 0.658281621 | 0.658281621 | 0.946175436 | -0.024028331 | FALSE | TRUE | FALSE |
| RAB6A | 0.622491 | 0.67919498 | 1 | 0.622490972 | 0.622490972 | 0.622490972 | 0.65839016 | 0.68632473 | 1 | 0.658390156 | 0.658390156 | 0.658390156 | 0.945474299 | -0.024350272 | FALSE | TRUE | FALSE |
| DPM2 | 0.6224702 | 0.67925656 | 1 | 0.622470212 | 0.622470212 | 0.622470212 | 0.65095893 | 0.689217269 | 1 | 2.603655487 | 2.603655487 | 2.603655487 | 0.956235767 | -0.019435016 | FALSE | TRUE | FALSE |
| DKK1 | 0.6224115 | 0.67931812 | 1 | 0.622411509 | 0.622411509 | 0.622411509 | 0.65832513 | 0.686388207 | 1 | 0.658325126 | 0.658325126 | 0.658325126 | 0.94544699 | -0.024362817 | FALSE | TRUE | FALSE |
| HSP90AA1 | 0.6221683 | 0.67937966 | 0.87615 | 5.814811488 | 93.01752491 | 372.1706857 | 0.40186977 | 0.791628319 | 0.7493 | 0.401869768 | 1.607561367 | 7.920724083 | 1.548183776 | 0.189822512 | FALSE | TRUE | FALSE |
| CARD10 | 0.6219592 | 0.67944117 | 0.03475 | 0.621959178 | 0.621959178 | 12.12511539 | 0.09580112 | 0.876949202 | 1 | 0.095801119 | 0.095801119 | 1.873328728 | 6.492191137 | 0.812391297 | FALSE | TRUE | FALSE |
| UBTD2 | 0.6217988 | 0.67950266 | 1 | 0.621798752 | 0.621798752 | 0.621798752 | 0.65776003 | 0.686578476 | 1 | 0.657760031 | 0.657760031 | 0.657760031 | 0.945327662 | -0.024417634 | FALSE | TRUE | FALSE |
| TMEM42 | 0.6215094 | 0.67956412 | 1 | 2.485939992 | 9.945854813 | 9.945854813 | 0.66476329 | 0.682985918 | 1 | 0.664763292 | 2.658970803 | 2.658970803 | 0.934933447 | -0.029219303 | FALSE | TRUE | FALSE |
| CDIPT | 0.6214665 | 0.67962556 | 1 | 2.48576234 | 2.48576234 | 2.48576234 | 0.64427058 | 0.692784581 | 1 | 6.084290537 | 6.084290537 | 6.084290537 | 0.964604769 | -0.015650595 | FALSE | TRUE | FALSE |
| MT1M | 0.6212749 | 0.67968698 | 1 | 0.621274915 | 0.621274915 | 0.621274915 | 0.65682349 | 0.686705195 | 1 | 0.656823489 | 0.656823489 | 0.656823489 | 0.945878041 | -0.024164857 | FALSE | TRUE | FALSE |
| TMEM248 | 0.6207717 | 0.67974837 | 1 | 0.620771729 | 0.620771729 | 0.620771729 | 0.65674386 | 0.686768517 | 1 | 0.656743855 | 0.656743855 | 0.656743855 | 0.945226551 | -0.024464088 | FALSE | TRUE | FALSE |
| LUC7L2 | 0.6204051 | 0.67980974 | 1 | 0.620405131 | 0.620405131 | 12.29909586 | 0.65627833 | 0.686895081 | 1 | 0.656278328 | 0.656278328 | 13.02400344 | 0.94533844 | -0.024412682 | FALSE | TRUE | FALSE |
| RNF122 | 0.620188 | 0.67987109 | 1 | 0.62018797 | 0.62018797 | 0.62018797 | 0.65608303 | 0.687021539 | 1 | 0.656083031 | 0.656083031 | 0.656083031 | 0.945288844 | -0.024435467 | FALSE | TRUE | FALSE |
| RFXANK | 0.6199964 | 0.67993242 | 0.7504 | 0.619996383 | 2.480362777 | 2.480362777 | 0.19685371 | 0.852866659 | 1 | 0.196853708 | 0.196853708 | 0.196853708 | 3.149528598 | 0.498245556 | FALSE | TRUE | FALSE |
| MARCKS | 0.6195705 | 0.67999372 | 1 | 0.619570492 | 0.619570492 | 0.619570492 | 5.457075 | 0.288260092 | 0.50615 | 52.57262889 | 519.4703743 | 519.4703743 | 0.113535272 | -0.944869197 | FALSE | FALSE | FALSE |
| GTF2H3 | 0.6194652 | 0.68005499 | 0.93665 | 5.695214372 | 214.9748914 | 214.9748914 | 1.81777263 | 0.505822144 | 0.25425 | 1.817772627 | 17.21658354 | 17.21658354 | 0.340782556 | -0.467522644 | FALSE | FALSE | FALSE |
| TMEM106B | 0.619407 | 0.68011625 | 1 | 0.619406975 | 0.619406975 | 0.619406975 | 0.65521069 | 0.687211036 | 0.00595 | 0.655210686 | 0.655210686 | 0.655210686 | 0.945355422 | -0.02440488 | FALSE | TRUE | FALSE |
| ZSWIM8 | 0.6193455 | 0.68017747 | 0.52255 | 0.619345452 | 1.299852944 | 11.91072127 | 0.09554074 | 0.876987484 | 1 | 0.095540737 | 0.095540737 | 3.210611467 | 6.482527444 | 0.811744364 | FALSE | TRUE | FALSE |
| RNF208 | 0.6189729 | 0.68023868 | 1 | 0.618972918 | 0.618972918 | 12.28537403 | 0.65527339 | 0.687147897 | 1 | 0.655273387 | 0.655273387 | 0.655273387 | 0.944602558 | -0.024750883 | FALSE | TRUE | FALSE |
| BRMS1L | 0.6188768 | 0.68029986 | 1 | 0.618876794 | 0.618876794 | 0.618876794 | 0.65498884 | 0.687274149 | 1 | 0.654988835 | 0.654988835 | 0.654988835 | 0.944866172 | -0.024629699 | FALSE | TRUE | FALSE |
| ANAPC13 | 0.6187227 | 0.68036101 | 1 | 0.618722694 | 0.618722694 | 0.618722694 | 0.65456673 | 0.687400294 | 1 | 0.654566735 | 0.654566735 | 0.654566735 | 0.945240052 | -0.024457885 | FALSE | TRUE | FALSE |
| ARHGAP15 | 0.6186843 | 0.68042214 | 0.509 | 0.618684337 | 0.618684337 | 1.364312869 | 0.38223394 | 0.798372001 | 0.011 | 0.382233935 | 0.382233935 | 1.896740608 | 1.618601282 | 0.20913988 | FALSE | TRUE | FALSE |
| EIF4A3 | 0.6185509 | 0.68048325 | 1 | 0.618550876 | 0.618550876 | 12.24151233 | 0.65497117 | 0.687337234 | 1 | 0.654971167 | 0.654971167 | 12.97861642 | 0.944394054 | -0.024846756 | FALSE | TRUE | FALSE |
| CLEC18A | 0.6184907 | 0.68054433 | 0.7518 | 0.618490732 | 5.976014551 | 12.23164068 | 6.46199449 | 0.256105235 | 0.00935 | 0.668093971 | 6.461994489 | 6.461994489 | 0.095712049 | -1.019033387 | FALSE | FALSE | FALSE |
| RPS14 | 0.6184561 | 0.68060538 | 1 | 0.618456101 | 0.618456101 | 0.618456101 | 0.64910571 | 0.690272021 | 1 | 0.649105711 | 0.649105711 | 0.649105711 | 0.95278179 | -0.021006552 | FALSE | TRUE | FALSE |
| CAPNS1 | 0.6184399 | 0.68066641 | 0.7493 | 0.618439901 | 2.473810164 | 2.473810164 | 0.19679879 | 0.852880073 | 1 | 0.196798787 | 0.196798787 | 0.196798787 | 3.142498538 | 0.497275084 | FALSE | TRUE | FALSE |
| FAM76B | 0.6177882 | 0.68072742 | 1 | 0.617788176 | 0.617788176 | 0.617788176 | 0.65399618 | 0.687778084 | 1 | 0.653996184 | 0.653996184 | 0.653996184 | 0.94463575 | -0.024735622 | FALSE | TRUE | FALSE |
| KRTCAP2 | 0.6177531 | 0.6807884 | 1 | 0.617753073 | 0.617753073 | 0.617753073 | 55.5866451 | 0.05059057 | 0.2505 | 5.766028325 | 55.58664507 | 55.58664507 | 0.011113336 | -1.954155549 | FALSE | FALSE | FALSE |
| TSSK3 | 0.6176469 | 0.68084936 | 0.50175 | 0.617646919 | 0.617646919 | 0.617646919 | 0.84888792 | 0.60772323 | 0.49565 | 0.093087223 | 0.848887915 | 0.848887915 | 0.727595372 | -0.138110072 | FALSE | TRUE | FALSE |
| EED | 0.6176342 | 0.68091029 | 1 | 0.617634241 | 0.617634241 | 12.26161964 | 0.65404037 | 0.687715187 | 1 | 0.654040375 | 0.654040375 | 12.99588521 | 0.944336565 | -0.024873194 | FALSE | TRUE | FALSE |
| SRSF1 | 0.6173483 | 0.68097121 | 1 | 0.617348316 | 0.617348316 | 0.617348316 | 0.65353189 | 0.688092157 | 1 | 0.65353189 | 0.65353189 | 0.65353189 | 0.944633806 | -0.024736516 | FALSE | TRUE | FALSE |
| API5 | 0.6171837 | 0.68103209 | 1 | 0.617183663 | 0.617183663 | 12.23510893 | 0.65360119 | 0.687966609 | 1 | 0.653601195 | 0.653601195 | 12.97075966 | 0.944281724 | -0.024898416 | FALSE | TRUE | FALSE |
| KRTAP5-11 | 0.6171759 | 0.68109295 | 1 | 0.617175895 | 0.617175895 | 0.617175895 | 0.65310349 | 0.688468141 | 1 | 0.653103491 | 0.653103491 | 0.653103491 | 0.944989429 | -0.02457305 | FALSE | TRUE | FALSE |
| KCNK9 | 0.6171115 | 0.68115379 | 1 | 0.617111479 | 0.617111479 | 12.19653942 | 0.65350676 | 0.688154889 | 1 | 0.653506764 | 0.653506764 | 12.93419619 | 0.944307714 | -0.024886462 | FALSE | TRUE | FALSE |
| ETV5 | 0.6170441 | 0.6812146 | 1 | 0.617044058 | 0.617044058 | 12.20771072 | 0.65357458 | 0.688029397 | 1 | 0.653574576 | 0.653574576 | 12.94722288 | 0.94410658 | -0.024978976 | FALSE | TRUE | FALSE |
| ARRB1 | 0.6168639 | 0.68127539 | 1 | 0.61686389 | 0.61686389 | 12.21290511 | 0.65324702 | 0.688405545 | 1 | 0.653247016 | 0.653247016 | 12.94891553 | 0.944304184 | -0.024888086 | FALSE | TRUE | FALSE |
| ADAM15 | 0.6167954 | 0.68133615 | 0.9988 | 45778.34304 | 50411399.36 | 543399671.4 | 0.13808431 | 0.867160983 | 0.99875 | 1000.088734 | 1008238.413 | 10572613.56 | 4.466803165 | 0.649996815 | FALSE | TRUE | FALSE |
| DDX39B | 0.6167373 | 0.68139689 | 1 | 0.616737288 | 0.616737288 | 12.2106903 | 0.65328812 | 0.688280273 | 1 | 0.653288116 | 0.653288116 | 12.95000276 | 0.944050983 | -0.025004551 | FALSE | TRUE | FALSE |
| TMEM215 | 0.6167152 | 0.6814576 | 1 | 0.616715197 | 0.616715197 | 0.616715197 | 0.65283723 | 0.688530712 | 1 | 0.652837231 | 0.652837231 | 0.652837231 | 0.944669157 | -0.024720264 | FALSE | TRUE | FALSE |
| SCNM1 | 0.616553 | 0.68151828 | 1 | 0.616552962 | 0.616552962 | 0.616552962 | 5.95365965 | 0.270728873 | 0.5001 | 5.953659652 | 5.953659652 | 5.953659652 | 0.103558651 | -0.984813616 | FALSE | FALSE | FALSE |
| SYPL1 | 0.6160008 | 0.68157895 | 1 | 5.80048797 | 5.80048797 | 5.80048797 | 0.67311694 | 0.678235352 | 1 | 0.673116937 | 0.673116937 | 0.673116937 | 0.915146849 | -0.038509211 | FALSE | TRUE | FALSE |
| PYDC2 | 0.6157436 | 0.6816396 | 1 | 0.615743634 | 0.615743634 | 0.615743634 | 0.65168203 | 0.688843179 | 1 | 0.651682025 | 0.651682025 | 0.651682025 | 0.944852874 | -0.024635812 | FALSE | TRUE | FALSE |
| CDKN2AIPNI | 0.6157364 | 0.68170022 | 1 | 0.61573643 | 0.61573643 | 0.61573643 | 0.65174769 | 0.688780736 | 1 | 0.651747691 | 0.651747691 | 0.651747691 | 0.944746623 | -0.024684652 | FALSE | TRUE | FALSE |
| MPC2 | 0.6155871 | 0.68176081 | 1 | 0.615587138 | 0.615587138 | 0.615587138 | 0.65189716 | 0.688718266 | 1 | 0.651897159 | 0.651897159 | 0.651897159 | 0.944300999 | -0.024889551 | FALSE | TRUE | FALSE |
| SMCO3 | 0.6154812 | 0.68182138 | 1 | 0.615481167 | 0.615481167 | 0.615481167 | 0.65160705 | 0.688905594 | 1 | 0.651607048 | 0.651607048 | 0.651607048 | 0.944558794 | -0.024771004 | FALSE | TRUE | FALSE |
| GEMIN7 | 0.6153505 | 0.68188193 | 1 | 0.615350485 | 0.615350485 | 0.615350485 | 0.651476 | 0.689092676 | 1 | 0.651475997 | 0.651475997 | 0.651475997 | 0.944548207 | -0.024775872 | FALSE | TRUE | FALSE |
| DEFB136 | 0.6149879 | 0.68194245 | 1 | 0.614987874 | 0.614987874 | 0.614987874 | 0.65097342 | 0.689154986 | 1 | 0.65097342 | 0.65097342 | 0.65097342 | 0.944720407 | -0.024696704 | FALSE | TRUE | FALSE |

| | | | | | | | | | | | | | | | | | |
|---|---|---|---|---|---|---|---|---|---|---|---|---|---|---|---|---|---|
| NAA30 | 0.6149792 | 0.68200295 | 1 | 5.818466264 | 56.49882676 | 56.49882676 | 6.37766439 | 0.259319855 | 0.51025 | 6.377664392 | 6.377664392 | 13.33837103 | 0.09642703 | -1.015801208 | FALSE | FALSE | FALSE |
| YBX1 | 0.6147865 | 0.68206343 | 1 | 0.614786541 | 0.614786541 | 12.20161458 | 0.59239423 | 0.71516243 | 1 | 54.08883226 | 537.0270167 | 537.0270167 | 1.037799671 | 0.016113529 | FALSE | TRUE | FALSE |
| EPHA1 | 0.6145719 | 0.68212388 | 0.8313 | 9.827381629 | 793.6748978 | 7398.442065 | 0.01246708 | 0.895922754 | 0.7569 | 0.01246708 | 0.109581993 | 0.244476632 | 49.29557301 | 1.692807919 | FALSE | TRUE | FALSE |
| MEIG1 | 0.6143529 | 0.68218431 | 1 | 0.614352867 | 0.614352867 | 0.614352867 | 0.65044192 | 0.689466136 | 1 | 0.650441922 | 0.650441922 | 0.650441922 | 0.944516099 | -0.024790635 | FALSE | TRUE | FALSE |
| RHBDD2 | 0.6143448 | 0.68224471 | 1 | 0.61434477 | 0.61434477 | 0.61434477 | 0.65066271 | 0.689403959 | 1 | 0.650662712 | 0.650662712 | 0.650662712 | 0.944183151 | -0.024943754 | FALSE | TRUE | FALSE |
| PRKACA | 0.6142695 | 0.68230509 | 1 | 5.812297886 | 5.812297886 | 5.812297886 | 0.67184279 | 0.679237631 | 0.0097 | 0.671842788 | 0.671842788 | 0.671842788 | 0.914305465 | -0.038908684 | FALSE | TRUE | FALSE |
| RASSF6 | 0.6141877 | 0.68236544 | 0.9679 | 52.21626183 | 499.8292901 | 4901.52274 | 0.94947357 | 0.59285385 | 0.5037 | 0.949473573 | 0.949473573 | 0.949473573 | 0.646871834 | -0.189181758 | FALSE | TRUE | FALSE |
| ARHGDIG | 0.6141715 | 0.68242577 | 1 | 0.614171521 | 0.614171521 | 0.614171521 | 0.65069085 | 0.689341756 | 1 | 0.650690846 | 0.650690846 | 0.650690846 | 0.943876073 | -0.025085023 | FALSE | TRUE | FALSE |
| ZP1 | 0.6139419 | 0.68248608 | 0.6922 | 0.61394186 | 51.66559581 | 51.66559581 | 0.01570072 | 0.894946313 | 0.01455 | 0.015700716 | 0.015700716 | 0.310317451 | 39.10279404 | 1.59220779 | FALSE | TRUE | FALSE |
| HCRT | 0.6139349 | 0.68254636 | 1 | 0.613934916 | 0.613934916 | 0.613934916 | 5.69502713 | 0.277581738 | 0.7475 | 5.695027135 | 55.15838943 | 55.15838943 | 0.10780193 | -0.967373464 | FALSE | FALSE | FALSE |
| CACNA1D | 0.6139022 | 0.68260662 | 0.02305 | 0.009642798 | 0.322284794 | 2.45253172 | 1.2591E-05 | 0.900693908 | 1 | 0.000749888 | 0.011945708 | 0.057346483 | 48756.56134 | 4.688033068 | FALSE | TRUE | FALSE |
| CRK | 0.613735 | 0.68266685 | 1 | 0.613735045 | 0.613735045 | 12.17752111 | 0.65041448 | 0.689528285 | 1 | 0.650414483 | 0.650414483 | 0.650414483 | 0.943606056 | -0.025209281 | FALSE | TRUE | FALSE |
| RPS20 | 0.613728 | 0.68272705 | 1 | 0.613727996 | 0.613727996 | 0.613727996 | 0.65014127 | 0.689590409 | 1 | 0.650141273 | 0.650141273 | 0.650141273 | 0.943991746 | -0.025031803 | FALSE | TRUE | FALSE |
| RAB1A | 0.613329 | 0.68278724 | 1 | 0.613329032 | 0.613329032 | 0.613329032 | 0.64994379 | 0.689652508 | 1 | 0.64994379 | 0.64994379 | 0.64994379 | 0.943664732 | -0.025182276 | FALSE | TRUE | FALSE |
| KLF3 | 0.6132634 | 0.6828474 | 1 | 0.61326341 | 0.61326341 | 12.16695411 | 0.62961067 | 0.699317227 | 1 | 5.939565918 | 5.939565918 | 12.50331981 | 0.974035921 | -0.011425027 | FALSE | TRUE | FALSE |
| MTCH1 | 0.6131828 | 0.68290754 | 1 | 0.613182834 | 0.613182834 | 0.613182834 | 0.64972701 | 0.689838643 | 1 | 0.649727007 | 0.649727007 | 0.649727007 | 0.943754574 | -0.025140931 | FALSE | TRUE | FALSE |
| ABHD4 | 0.6131729 | 0.68296765 | 0.13425 | 0.613172943 | 2.452721455 | 2.452721455 | 0.90810594 | 0.597284951 | 0.2591 | 0.227066578 | 0.908105936 | 1.127611043 | 0.67522182 | -0.170553532 | FALSE | TRUE | FALSE |
| PPAN | 0.6130236 | 0.68302774 | 1 | 0.613023611 | 0.613023611 | 12.11485679 | 0.59808983 | 0.712969758 | 0.75055 | 5.649051427 | 54.76065386 | 54.76065386 | 1.024969119 | 0.010710781 | FALSE | TRUE | FALSE |
| PIN1 | 0.6127127 | 0.6830878 | 1 | 0.612712729 | 0.612712729 | 0.612712729 | 0.64923722 | 0.69021019 | 1 | 0.64923722 | 0.64923722 | 0.64923722 | 0.943742456 | -0.025146507 | FALSE | TRUE | FALSE |
| PGM3 | 0.6123932 | 0.68314785 | 0.5057 | 0.612393185 | 0.612393185 | 1.341695157 | 0.09543528 | 0.877035305 | 0.01275 | 0.09543528 | 0.09543528 | 1.893212609 | 6.416842775 | 0.807321398 | FALSE | TRUE | FALSE |
| PTN | 0.612028 | 0.68320787 | 1 | 0.612027958 | 0.612027958 | 0.612027958 | 0.64836165 | 0.690642459 | 1 | 0.648361651 | 0.648361651 | 0.648361651 | 0.943960762 | -0.025046058 | FALSE | TRUE | FALSE |
| FHIT | 0.6117511 | 0.68326787 | 0.8771 | 2.446448354 | 9.78601493 | 9.78601493 | 0.19321203 | 0.853547903 | 0.7526 | 0.772525698 | 3.089585875 | 3.089585875 | 3.166216472 | 0.500540604 | FALSE | TRUE | FALSE |
| RGS17 | 0.6117226 | 0.68332784 | 1 | 0.61172262 | 0.61172262 | 0.61172262 | 0.64819553 | 0.690704107 | 1 | 0.648195529 | 0.648195529 | 0.648195529 | 0.943731624 | -0.025151492 | FALSE | TRUE | FALSE |
| YPEL3 | 0.6112658 | 0.6833878 | 1 | 0.611265817 | 0.611265817 | 0.611265817 | 0.64791369 | 0.690888891 | 1 | 0.647913694 | 0.647913694 | 0.647913694 | 0.9434371 | -0.025287049 | FALSE | TRUE | FALSE |
| LRTM2 | 0.6111578 | 0.68344773 | 1 | 0.611157762 | 0.611157762 | 12.10614828 | 0.64805351 | 0.690827322 | 1 | 0.648053514 | 0.648053514 | 12.85178032 | 0.943066812 | -0.025457539 | FALSE | TRUE | FALSE |
| CSDC2 | 0.6109926 | 0.68350763 | 1 | 0.610992649 | 0.610992649 | 0.610992649 | 0.6475897 | 0.690950435 | 1 | 0.647589698 | 0.647589698 | 0.647589698 | 0.943487289 | -0.025263946 | FALSE | TRUE | FALSE |
| STMN4 | 0.6109011 | 0.68356752 | 1 | 0.610901128 | 0.610901128 | 0.610901128 | 0.6475432 | 0.691011953 | 1 | 0.6475432 | 0.6475432 | 0.6475432 | 0.943413702 | -0.02529782 | FALSE | TRUE | FALSE |
| CD69 | 0.6107955 | 0.68362738 | 1 | 0.610795492 | 0.610795492 | 0.610795492 | 0.6472753 | 0.691319144 | 1 | 0.647275299 | 0.647275299 | 0.647275299 | 0.943640972 | -0.025193211 | FALSE | TRUE | FALSE |
| IRAK3 | 0.6107439 | 0.68368721 | 0.69035 | 2.442147152 | 51.14494644 | 204.5104026 | 0.12633805 | 0.86979445 | 0.5013 | 1.126766132 | 10.30765823 | 10.30765823 | 4.834204073 | 0.68432498 | FALSE | TRUE | FALSE |
| CREBL2 | 0.6106629 | 0.68374702 | 0.00445 | 0.610662948 | 0.610662948 | 0.610662948 | 0.64727797 | 0.69125776 | 1 | 0.647277967 | 0.647277967 | 0.647277967 | 0.943432311 | -0.025289254 | FALSE | TRUE | FALSE |
| PDE6G | 0.610588 | 0.68380681 | 1 | 0.61058799 | 0.61058799 | 0.61058799 | 0.64698584 | 0.691380503 | 1 | 0.646985843 | 0.646985843 | 0.646985843 | 0.943742428 | -0.02514652 | FALSE | TRUE | FALSE |
| ZRANB2 | 0.610547 | 0.68386657 | 1 | 0.610546998 | 0.610546998 | 12.12621796 | 0.58858309 | 0.717421869 | 1 | 53.65143175 | 532.2414917 | 532.2414917 | 1.037316578 | 0.015911319 | FALSE | TRUE | FALSE |
| MPPED2 | 0.6104641 | 0.6839263 | 1 | 0.610464094 | 0.610464094 | 0.610464094 | 0.64727799 | 0.691196349 | 1 | 0.647277991 | 0.647277991 | 0.647277991 | 0.94312506 | -0.025430715 | FALSE | TRUE | FALSE |
| ATF7 | 0.6104185 | 0.68398602 | 1 | 0.610418547 | 0.610418547 | 12.08740149 | 0.64730232 | 0.69113491 | 1 | 0.647302323 | 0.647302323 | 12.83304506 | 0.943019244 | -0.025479445 | FALSE | TRUE | FALSE |
| ZNF511 | 0.6103074 | 0.6840457 | 0.7565 | 1.413117581 | 5.650903039 | 5.650903039 | 0.75907181 | 0.637286232 | 0.9398 | 3.036269719 | 48.61618668 | 48.61618668 | 0.804017966 | -0.094734247 | FALSE | TRUE | FALSE |
| POLR2I | 0.6102579 | 0.68410537 | 1 | 0.610257937 | 0.610257937 | 0.610257937 | 0.6469661 | 0.691441836 | 0.00385 | 0.646966098 | 0.646966098 | 0.646966098 | 0.943261074 | -0.025368087 | FALSE | TRUE | FALSE |
| SAP30 | 0.6102505 | 0.68416501 | 1 | 0.610250471 | 0.610250471 | 0.610250471 | 0.6468011 | 0.691564421 | 0.0045 | 0.646801095 | 0.646801095 | 0.646801095 | 0.943490164 | -0.025262623 | FALSE | TRUE | FALSE |
| BTG2 | 0.6102183 | 0.68422462 | 1 | 0.610218277 | 0.610218277 | 0.610218277 | 0.6466774 | 0.691625675 | 1 | 0.646677399 | 0.646677399 | 0.646677399 | 0.94362085 | -0.025202471 | FALSE | TRUE | FALSE |
| RBM7 | 0.6102092 | 0.68428421 | 1 | 0.610209155 | 0.610209155 | 0.610209155 | 0.64695761 | 0.691503141 | 1 | 0.646957608 | 0.646957608 | 0.646957608 | 0.943198051 | -0.025397105 | FALSE | TRUE | FALSE |
| PRDX6 | 0.6101552 | 0.68434377 | 1 | 5.757863004 | 5.757863004 | 5.757863004 | 6.10871867 | 0.266005352 | 0.5061 | 6.108718666 | 6.108718666 | 6.108718666 | 0.099882686 | -1.000509789 | FALSE | FALSE | FALSE |
| CDK2AP1 | 0.6100426 | 0.68440332 | 1 | 5.739966296 | 5.739966296 | 5.739966296 | 0.66723426 | 0.681943285 | 1 | 0.667234259 | 0.667234259 | 0.667234259 | 0.914285534 | -0.038918152 | FALSE | TRUE | FALSE |
| USP12 | 0.6099049 | 0.68446283 | 1 | 0.609904876 | 0.609904876 | 0.609904876 | 0.64665035 | 0.691686902 | 1 | 0.646650346 | 0.646650346 | 0.646650346 | 0.943175674 | -0.025407409 | FALSE | TRUE | FALSE |
| TMEM115 | 0.609672 | 0.68452233 | 1 | 2.438525642 | 2.438525642 | 12.09030543 | 0.65391115 | 0.687903793 | 1 | 0.653911153 | 0.653911153 | 0.653911153 | 0.932346804 | -0.030422513 | FALSE | TRUE | FALSE |
| FGF9 | 0.6096273 | 0.6845818 | 1 | 0.609627307 | 0.609627307 | 0.609627307 | 0.64655128 | 0.691748103 | 1 | 0.646551279 | 0.646551279 | 0.646551279 | 0.942890885 | -0.025538563 | FALSE | TRUE | FALSE |
| KLHL26 | 0.609546 | 0.68464125 | 0.86765 | 21.77336682 | 1396.010591 | 5590.190241 | 0.06831351 | 0.882818371 | 0.99675 | 4.371170748 | 70.01659781 | 343.1885853 | 8.922774175 | 0.950499902 | FALSE | TRUE | FALSE |
| LY6K | 0.6094376 | 0.68470067 | 1 | 0.609437586 | 0.609437586 | 0.609437586 | 0.64584821 | 0.692053715 | 1 | 0.645848205 | 0.645848205 | 0.645848205 | 0.943623565 | -0.025201222 | FALSE | TRUE | FALSE |
| C1QL3 | 0.6093656 | 0.68476007 | 1 | 0.609365639 | 0.609365639 | 0.609365639 | 0.63907416 | 0.694716013 | 1 | 0.63907416 | 2.556111246 | 2.556111246 | 0.953513186 | -0.020673297 | FALSE | TRUE | FALSE |
| C19orf25 | 0.6091355 | 0.68481945 | 1 | 0.609135465 | 0.609135465 | 0.609135465 | 0.62273987 | 0.702418127 | 1 | 0.622739865 | 5.846950439 | 5.846950439 | 0.978153959 | -0.009592783 | FALSE | TRUE | FALSE |
| TAF11 | 0.6091132 | 0.6848788 | 1 | 0.609113211 | 0.609113211 | 0.609113211 | 0.64582138 | 0.692114761 | 1 | 0.645821382 | 0.645821382 | 0.645821382 | 0.943160491 | -0.0254144 | FALSE | TRUE | FALSE |
| TGIF2 | 0.6086357 | 0.68493813 | 1 | 0.608635744 | 0.608635744 | 0.608635744 | 0.64543925 | 0.692236779 | 0.0056 | 0.645439253 | 0.645439253 | 0.645439253 | 0.942979128 | -0.02549792 | FALSE | TRUE | FALSE |
| ATXN7L3 | 0.6085515 | 0.68499744 | 1 | 0.608551536 | 0.608551536 | 12.06967088 | 0.6358566 | 0.696799098 | 1 | 0.635856605 | 0.635856605 | 12.62385526 | 0.957057819 | -0.019061824 | FALSE | TRUE | FALSE |
| SEC62 | 0.6084256 | 0.68505672 | 1 | 0.608425571 | 0.608425571 | 0.608425571 | 0.64543803 | 0.692297748 | 1 | 0.645438033 | 0.645438033 | 0.645438033 | 0.942655283 | -0.025647094 | FALSE | TRUE | FALSE |
| MLF2 | 0.6080729 | 0.68511599 | 1 | 0.608072871 | 0.608072871 | 12.07620042 | 0.64501614 | 0.692602206 | 1 | 0.645016138 | 0.645016138 | 0.645016138 | 0.94272505 | -0.025614953 | FALSE | TRUE | FALSE |
| LYZL4 | 0.6075755 | 0.68517523 | 1 | 0.607575454 | 0.607575454 | 0.607575454 | 0.64452587 | 0.692663022 | 1 | 0.644525872 | 0.644525872 | 0.644525872 | 0.942670388 | -0.025640135 | FALSE | TRUE | FALSE |
| TMEM9B | 0.6075527 | 0.68523445 | 1 | 0.607552672 | 0.607552672 | 0.607552672 | 0.64430686 | 0.692723815 | 1 | 0.644306858 | 0.644306858 | 0.644306858 | 0.942955463 | -0.025508819 | FALSE | TRUE | FALSE |
| ZC3H10 | 0.6071794 | 0.68529365 | 1 | 2.428672747 | 2.428672747 | 12.02112748 | 0.65158836 | 0.688967982 | 1 | 0.651588364 | 0.651588364 | 12.91599862 | 0.931845079 | -0.030656284 | FALSE | TRUE | FALSE |
| NUDT3 | 0.6067964 | 0.68535283 | 1 | 0.606796368 | 0.606796368 | 0.606796368 | 0.61985395 | 0.703718301 | 1 | 0.619853953 | 0.619853953 | 0.619853953 | 0.978934417 | -0.009246403 | FALSE | TRUE | FALSE |
| SYNJ2BP | 0.6067843 | 0.68541198 | 1 | 0.606784263 | 0.606784263 | 0.606784263 | 0.61549297 | 0.705451861 | 1 | 0.615492969 | 0.615492969 | 0.615492969 | 0.985850844 | -0.006188787 | FALSE | TRUE | FALSE |
| CHRM1 | 0.6067827 | 0.68547111 | 1 | 0.606782702 | 0.606782702 | 11.99168888 | 0.64395911 | 0.692966721 | 1 | 0.643959111 | 0.643959111 | 12.74456107 | 0.942268992 | -0.0258251 | FALSE | TRUE | FALSE |
| SGCB | 0.6062901 | 0.68553022 | 0.25075 | 0.065451173 | 0.606290086 | 0.606290086 | 0.02744887 | 0.891853567 | 1 | 0.109697631 | 0.109697631 | 0.109697631 | 22.08798263 | 1.344156052 | FALSE | TRUE | FALSE |
| LDLRAD4 | 0.6059019 | 0.68558931 | 1 | 0.605901921 | 0.605901921 | 0.605901921 | 0.64294606 | 0.693269787 | 1 | 0.642946059 | 0.642946059 | 0.642946059 | 0.942383754 | -0.025772209 | FALSE | TRUE | FALSE |
| ELAVL4 | 0.6056825 | 0.68564837 | 1 | 0.605682502 | 0.605682502 | 0.605682502 | 5.87287437 | 0.273228239 | 0.50215 | 5.872874366 | 5.872874366 | 5.872874366 | 0.103132208 | -0.986605683 | FALSE | FALSE | FALSE |
| BLOC1S2 | 0.6054982 | 0.68570742 | 1 | 0.605498247 | 0.605498247 | 0.605498247 | 0.62236084 | 0.702645112 | 0.5016 | 5.850609544 | 5.850609544 | 5.850609544 | 0.972905441 | -0.011929368 | FALSE | TRUE | FALSE |
| CLDND1 | 0.6051668 | 0.68576644 | 1 | 5.703243463 | 5.703243463 | 5.703243463 | 0.66274695 | 0.684407741 | 1 | 0.662746946 | 0.662746946 | 0.662746946 | 0.913118898 | -0.039472669 | FALSE | TRUE | FALSE |
| SMARCE1 | 0.604985 | 0.68582544 | 1 | 0.604984994 | 0.604984994 | 11.99222374 | 0.64220436 | 0.693330333 | 1 | 0.642204359 | 0.642204359 | 12.74359883 | 0.942044359 | -0.025928647 | FALSE | TRUE | FALSE |
| HCFC1R1 | 0.6047633 | 0.68588441 | 1 | 0.604763274 | 0.604763274 | 0.604763274 | 0.6414265 | 0.693511826 | 1 | 0.641426503 | 0.641426503 | 0.641426503 | 0.942841107 | -0.025561491 | FALSE | TRUE | FALSE |
| CHMP4B | 0.6046463 | 0.68594337 | 1 | 0.604646331 | 0.604646331 | 0.604646331 | 0.63486069 | 0.697447491 | 1 | 0.634860691 | 0.634860691 | 0.634860691 | 0.95240789 | -0.021177016 | FALSE | TRUE | FALSE |
| STX1A | 0.6044448 | 0.6860023 | 1 | 0.604444754 | 0.604444754 | 11.98987019 | 0.63123211 | 0.698735532 | 0.00865 | 0.631232106 | 0.631232106 | 0.631232106 | 0.957563387 | -0.018832468 | FALSE | TRUE | FALSE |
| RTN4R | 0.6039923 | 0.68606121 | 1 | 0.603992279 | 0.603992279 | 11.91268225 | 0.63825162 | 0.69549329 | 1 | 0.638251621 | 0.638251621 | 12.60935145 | 0.946323141 | -0.02396054 | FALSE | TRUE | FALSE |
| S1PR1 | 0.6036834 | 0.6861201 | 1 | 0.603683372 | 0.603683372 | 11.95182006 | 0.64095943 | 0.693813815 | 1 | 0.640959434 | 0.640959434 | 12.70522256 | 0.941843336 | -0.02602133 | FALSE | TRUE | FALSE |
| RNF41 | 0.603466 | 0.68617897 | 1 | 0.603466033 | 0.603466033 | 0.603466033 | 0.64056624 | 0.693994704 | 1 | 0.64056624 | 0.64056624 | 0.64056624 | 0.942082169 | -0.025911216 | FALSE | TRUE | FALSE |
| MAPK8IP2 | 0.6034068 | 0.68623781 | 1 | 0.603406798 | 0.603406798 | 11.90628581 | 0.63848202 | 0.695254589 | 1 | 0.63848202 | 0.63848202 | 12.61871467 | 0.945064668 | -0.024538473 | FALSE | TRUE | FALSE |
| DNAJB5 | 0.6031216 | 0.68629664 | 1 | 0.603121602 | 0.603121602 | 11.95773967 | 0.64045739 | 0.69405495 | 1 | 0.640457392 | 0.640457392 | 12.71122932 | 0.941704491 | -0.026085359 | FALSE | TRUE | FALSE |

| | | | | | | | | | | | | | | | | | |
|---|---|---|---|---|---|---|---|---|---|---|---|---|---|---|---|---|---|
| SPATA12 | 0.6031053 | 0.68635543 | 0.00445 | 0.603105263 | 0.603105263 | 0.603105263 | 0.63997511 | 0.694235538 | 1 | 0.639975107 | 0.639975107 | 0.639975107 | 0.942388628 | -0.025769963 | FALSE | TRUE | FALSE |
| TUBAL3 | 0.6029469 | 0.68641421 | 0.68995 | 1.435806321 | 55.99920704 | 55.99920704 | 0.02847447 | 0.891618177 | 1 | 0.265218 | 0.265218 | 0.265218 | 21.17499755 | 1.325823369 | FALSE | TRUE | FALSE |
| WWTR1 | 0.6025771 | 0.68647297 | 1 | 0.602577077 | 0.602577077 | 11.93693756 | 0.63995203 | 0.694295684 | 1 | 0.639952034 | 0.639952034 | 12.69182894 | 0.941597252 | -0.026134817 | FALSE | TRUE | FALSE |
| CMTR2 | 0.6023494 | 0.6865317 | 0.75485 | 0.602349426 | 5.506059582 | 11.89590494 | 0.08679891 | 0.87883701 | 0.93775 | 7.144740721 | 650.9398543 | 650.9398543 | 6.939596924 | 0.841334246 | FALSE | TRUE | FALSE |
| NWD1 | 0.6022989 | 0.68659041 | 0.3597 | 0.070440999 | 5.276269659 | 47.32862929 | 0.02435977 | 0.892666505 | 0.8148 | 0.207343458 | 1.809181953 | 16.17299448 | 24.7251454 | 1.393138854 | FALSE | TRUE | FALSE |
| COPS7B | 0.6022852 | 0.68664909 | 1 | 0.602285153 | 0.602285153 | 0.602285153 | 0.63944946 | 0.694415905 | 1 | 0.639449461 | 0.639449461 | 0.639449461 | 0.941880773 | -0.026004069 | FALSE | TRUE | FALSE |
| CLCF1 | 0.6022218 | 0.68670775 | 1 | 0.602221781 | 0.602221781 | 0.602221781 | 0.63952402 | 0.694355807 | 1 | 0.639524019 | 0.639524019 | 0.639524019 | 0.941671873 | -0.026100402 | FALSE | TRUE | FALSE |
| SPATA13 | 0.602218 | 0.68676639 | 0.5092 | 0.602218016 | 9.628847508 | 38.51654522 | 0.03359634 | 0.89044276 | 0.5045 | 0.134305063 | 0.537033376 | 2.648499881 | 17.92510525 | 1.253461714 | FALSE | TRUE | FALSE |
| PLAG1 | 0.6020742 | 0.686825 | 0.87685 | 22.76657713 | 220.8888147 | 220.8888147 | 0.68901568 | 0.671169373 | 1 | 0.689015676 | 0.689015676 | 13.66668059 | 0.873817875 | -0.058579076 | FALSE | TRUE | FALSE |
| CD164L2 | 0.6020022 | 0.68688359 | 1 | 0.602002193 | 0.602002193 | 0.602002193 | 0.63899001 | 0.694775957 | 1 | 0.638990013 | 0.638990013 | 0.638990013 | 0.942115182 | -0.025895997 | FALSE | TRUE | FALSE |
| EID1 | 0.6018763 | 0.68694216 | 1 | 0.601876323 | 0.601876323 | 0.601876323 | 0.63890752 | 0.694895768 | 1 | 0.638907517 | 0.638907517 | 0.638907517 | 0.942039821 | -0.025930739 | FALSE | TRUE | FALSE |
| DCAF7 | 0.6018294 | 0.6870007 | 1 | 0.60182942 | 0.60182942 | 0.60182942 | 0.63915635 | 0.694596047 | 1 | 0.63915635 | 0.63915635 | 0.63915635 | 0.941599688 | -0.026133694 | FALSE | TRUE | FALSE |
| TFCP2L1 | 0.6018077 | 0.68705922 | 1 | 0.601807668 | 0.601807668 | 11.89446477 | 0.63919072 | 0.694536025 | 1 | 0.639190716 | 0.639190716 | 12.65121735 | 0.941515032 | -0.026172742 | FALSE | TRUE | FALSE |
| CDK1 | 0.601737 | 0.68711772 | 1 | 0.601737 | 0.601737 | 0.601737 | 0.63867992 | 0.695075294 | 1 | 0.638679916 | 0.638679916 | 0.638679916 | 0.942157386 | -0.025876543 | FALSE | TRUE | FALSE |
| CNTFR | 0.6017027 | 0.68717619 | 1 | 0.601702659 | 0.601702659 | 11.93240857 | 0.63912343 | 0.694656043 | 1 | 0.639123429 | 0.639123429 | 12.68737756 | 0.941449855 | -0.026202807 | FALSE | TRUE | FALSE |
| ATP6V1G2 | 0.6016852 | 0.68723464 | 1 | 0.601685166 | 0.601685166 | 0.601685166 | 0.63869017 | 0.695015478 | 1 | 0.638690168 | 0.638690168 | 0.638690168 | 0.942061107 | -0.025920926 | FALSE | TRUE | FALSE |
| CNPY4 | 0.6016648 | 0.68729306 | 0.502 | 0.601664789 | 0.601664789 | 0.601664789 | 7.3346441 | 0.237162248 | 0.00495 | 0.799899236 | 7.3346441 | 7.3346441 | 0.082030536 | -1.086024449 | FALSE | FALSE | FALSE |
| S100A1 | 0.6016412 | 0.68735146 | 1 | 0.601641217 | 0.601641217 | 0.601641217 | 0.63856913 | 0.69519485 | 1 | 0.638569132 | 0.638569132 | 0.638569132 | 0.942170842 | -0.02587034 | FALSE | TRUE | FALSE |
| PLAGL1 | 0.60157 | 0.68740984 | 1 | 0.60157005 | 0.60157005 | 11.92565921 | 0.63893938 | 0.694835876 | 1 | 0.638939379 | 0.638939379 | 12.67986618 | 0.941513498 | -0.026173449 | FALSE | TRUE | FALSE |
| TWF2 | 0.6014222 | 0.68746819 | 1 | 0.60142222 | 0.60142222 | 11.91563493 | 0.63885922 | 0.694955635 | 1 | 0.638859224 | 0.638859224 | 0.638859224 | 0.941400228 | -0.026225701 | FALSE | TRUE | FALSE |
| ACYP1 | 0.601351 | 0.68752652 | 1 | 0.601351001 | 0.601351001 | 0.601351001 | 0.63842794 | 0.695433653 | 1 | 0.638427944 | 0.638427944 | 0.638427944 | 0.941924624 | -0.02598385 | FALSE | TRUE | FALSE |
| KRTAP12-1 | 0.6011235 | 0.68758483 | 1 | 0.601123485 | 5.639345358 | 5.639345358 | 5.80364527 | 0.274785911 | 0.2505 | 5.803645274 | 55.90915496 | 55.90915496 | 0.103576883 | -0.984737164 | FALSE | FALSE | FALSE |
| AP3B2 | 0.6010734 | 0.68764312 | 0.5135 | 0.601073365 | 0.601073365 | 11.73872982 | 0.83136413 | 0.612246947 | 0.02755 | 0.831364127 | 0.831364127 | 16.28222822 | 0.722996513 | -0.140863797 | FALSE | TRUE | FALSE |
| PYGO1 | 0.6009076 | 0.68770138 | 1 | 0.600907555 | 0.600907555 | 11.92302422 | 0.63843035 | 0.695373991 | 0.0095 | 0.638430348 | 0.638430348 | 0.638430348 | 0.941226488 | -0.026305859 | FALSE | TRUE | FALSE |
| RHOB | 0.6008836 | 0.68775962 | 1 | 2.403399822 | 9.615464026 | 9.615464026 | 0.65278896 | 0.688593254 | 1 | 0.652788958 | 0.652788958 | 0.652788958 | 0.920486794 | -0.035982438 | FALSE | TRUE | FALSE |
| CDK19 | 0.6008829 | 0.68781783 | 1 | 0.600882864 | 0.600882864 | 11.89404852 | 21.6291846 | 0.125539697 | 0.0031 | 0.638374614 | 0.638374614 | 12.6518056 | 0.027781115 | -1.556250329 | FALSE | FALSE | FALSE |
| ELMOD1 | 0.6007294 | 0.68787603 | 1 | 0.600729352 | 0.600729352 | 0.600729352 | 2.52385753 | 0.445427268 | 0.5035 | 2.523857525 | 2.523857525 | 2.523857525 | 0.238020311 | -0.623385983 | FALSE | FALSE | FALSE |
| NLK | 0.6006957 | 0.68793419 | 1 | 0.600695693 | 0.600695693 | 11.90305579 | 0.63798245 | 0.695612492 | 0.01105 | 0.637982453 | 0.637982453 | 12.65594049 | 0.941555196 | -0.026154216 | FALSE | TRUE | FALSE |
| RMND5A | 0.6004892 | 0.68799234 | 1 | 0.600489208 | 0.600489208 | 11.92127732 | 0.61807153 | 0.7045598 | 1 | 5.845569151 | 5.845569151 | 5.845569151 | 0.971552932 | -0.012533533 | FALSE | TRUE | FALSE |
| CDK7 | 0.6002213 | 0.68805047 | 0.99965 | 123828.7627 | 56196572.49 | 603035927.5 | 0.21777561 | 0.847779138 | 1 | 639.0757458 | 73184.32715 | 190097.5974 | 2.75614553 | 0.440302145 | FALSE | TRUE | FALSE |
| ZNF654 | 0.6001238 | 0.68810857 | 1 | 0.600123812 | 0.600123812 | 11.88874417 | 0.63764636 | 0.695672058 | 1 | 0.637646356 | 0.637646356 | 12.64648144 | 0.941154618 | -0.026339022 | FALSE | TRUE | FALSE |
| ANKRD9 | 0.6000093 | 0.68816665 | 1 | 2.399810683 | 2.399810683 | 2.399810683 | 2.02766082 | 0.487717962 | 0.9677 | 76.37500992 | 1828.322866 | 7314.477436 | 0.295912065 | -0.528837327 | FALSE | FALSE | FALSE |
| NAIF1 | 0.5999781 | 0.6882247 | 1 | 0.59997812 | 0.59997812 | 11.88832868 | 0.63734154 | 0.695731601 | 1 | 0.637341538 | 0.637341538 | 0.637341538 | 0.941376145 | -0.026236811 | FALSE | TRUE | FALSE |
| FAM219A | 0.5998969 | 0.68828274 | 1 | 0.599896874 | 0.599896874 | 0.599896874 | 0.63723937 | 0.69585061 | 1 | 0.637239375 | 0.637239375 | 0.637239375 | 0.941399571 | -0.026226004 | FALSE | TRUE | FALSE |
| AMACR | 0.5998423 | 0.68834075 | 0.75905 | 0.59984228 | 2.399147015 | 2.972146709 | 0.19190237 | 0.853892926 | 0.0099 | 0.191902375 | 0.191902375 | 0.191902375 | 3.125767883 | 0.494956724 | FALSE | TRUE | FALSE |
| KHDRBS1 | 0.5998312 | 0.68839873 | 1 | 0.599831168 | 0.599831168 | 0.599831168 | 0.63729965 | 0.695791118 | 1 | 0.637299649 | 0.637299649 | 0.637299649 | 0.941207436 | -0.02631465 | FALSE | TRUE | FALSE |
| KRTAP19-7 | 0.5996013 | 0.6884567 | 1 | 0.599601302 | 0.599601302 | 0.599601302 | 0.63652747 | 0.696207052 | 1 | 0.636527475 | 0.636527475 | 0.636527475 | 0.941988093 | -0.025954587 | FALSE | TRUE | FALSE |
| CASZ1 | 0.5995289 | 0.68851464 | 0.7664 | 0.599528917 | 2.855747277 | 45.70210483 | 2.96491767 | 0.413700161 | 0.1719 | 2.964917671 | 11.86541034 | 99.78605346 | 0.20220761 | -0.694202504 | FALSE | FALSE | FALSE |
| TMEM244 | 0.5994623 | 0.68857255 | 1 | 5.622331764 | 5.622331764 | 5.622331764 | 0.63646583 | 0.69626637 | 1 | 0.63646583 | 5.978183361 | 5.978183361 | 0.941860867 | -0.026013247 | FALSE | TRUE | FALSE |
| TTC9 | 0.5994023 | 0.68863045 | 0.0037 | 0.599402283 | 0.599402283 | 0.599402283 | 0.63660616 | 0.696028945 | 1 | 0.636606158 | 0.636606158 | 0.636606158 | 0.94155904 | -0.026152443 | FALSE | TRUE | FALSE |
| ST3GAL2 | 0.5993575 | 0.68868832 | 1 | 0.599357545 | 0.599357545 | 11.86214278 | 0.63690498 | 0.695910079 | 1 | 0.636904979 | 0.636904979 | 12.62107348 | 0.94104704 | -0.026388667 | FALSE | TRUE | FALSE |
| SRSF5 | 0.5991461 | 0.68874617 | 1 | 0.599146073 | 0.599146073 | 11.89669157 | 0.63667612 | 0.695969525 | 1 | 0.636676115 | 0.636676115 | 0.636676115 | 0.941053164 | -0.026385841 | FALSE | TRUE | FALSE |
| MSMP | 0.5989569 | 0.68880399 | 1 | 0.598956935 | 0.598956935 | 0.598956935 | 0.63618136 | 0.69662175 | 1 | 0.636181358 | 0.636181358 | 0.636181358 | 0.941487718 | -0.026185341 | FALSE | TRUE | FALSE |
| CAB39 | 0.5989301 | 0.6888618 | 0.00875 | 0.598930052 | 0.598930052 | 0.598930052 | 0.63632055 | 0.696325664 | 1 | 0.636320545 | 0.636320545 | 0.636320545 | 0.941239531 | -0.026299841 | FALSE | TRUE | FALSE |
| OR1J4 | 0.5988597 | 0.68891958 | 1 | 0.598859698 | 0.598859698 | 0.598859698 | 0.6362075 | 0.696562584 | 1 | 0.636207503 | 0.636207503 | 0.636207503 | 0.941296189 | -0.0262737 | FALSE | TRUE | FALSE |
| COPS6 | 0.5987656 | 0.68897734 | 0.0114 | 0.598765593 | 0.598765593 | 11.88733192 | 0.61649781 | 0.7051737 | 1 | 5.819886041 | 5.819886041 | 12.25018373 | 0.971237175 | -0.012674703 | FALSE | TRUE | FALSE |
| FLI1 | 0.5987011 | 0.68903507 | 1 | 0.598701138 | 0.598701138 | 11.855722 | 0.63628955 | 0.696384932 | 1 | 0.636289553 | 0.636289553 | 12.61503706 | 0.940925614 | -0.026444709 | FALSE | TRUE | FALSE |
| CHRAC1 | 0.5986712 | 0.68909278 | 0.0033 | 0.598671178 | 0.598671178 | 0.598671178 | 0.61935703 | 0.703943202 | 1 | 0.619357031 | 0.619357031 | 0.619357031 | 0.966601085 | -0.014752722 | FALSE | TRUE | FALSE |
| SERTAD1 | 0.5986503 | 0.68915047 | 1 | 0.59865027 | 0.59865027 | 0.59865027 | 0.63604228 | 0.696740006 | 1 | 0.636042281 | 0.636042281 | 0.636042281 | 0.941211438 | -0.026312804 | FALSE | TRUE | FALSE |
| TUBA1B | 0.5985367 | 0.68920813 | 1 | 0.598536733 | 0.598536733 | 11.85272565 | 0.63622275 | 0.696503392 | 1 | 0.636222754 | 0.636222754 | 12.61395577 | 0.940765996 | -0.026518388 | FALSE | TRUE | FALSE |
| IZUMO4 | 0.5982492 | 0.68926578 | 0.94125 | 5.422245722 | 480.3709425 | 480.3709425 | 0.08674544 | 0.878846316 | 1 | 65.80703995 | 6201.605967 | 62388.98783 | 6.896607299 | 0.838635497 | FALSE | TRUE | FALSE |
| TUBB4B | 0.5982236 | 0.6893234 | 1 | 0.598223627 | 0.598223627 | 11.81037562 | 0.62611619 | 0.70058873 | 1 | 2.504372666 | 2.504372666 | 12.38019929 | 0.955451453 | -0.019791375 | FALSE | TRUE | FALSE |
| PYROXD1 | 0.5981269 | 0.689381 | 0.8751 | 5.49264505 | 51.73565382 | 51.73565382 | 0.09991571 | 0.875926686 | 1 | 0.099915708 | 0.099915708 | 1.981990622 | 5.98631458 | 0.777159535 | FALSE | TRUE | FALSE |
| SPACA4 | 0.5978906 | 0.68943857 | 1 | 0.597890588 | 0.597890588 | 0.597890588 | 0.63507254 | 0.697153141 | 1 | 0.635072537 | 0.635072537 | 0.635072537 | 0.941452437 | -0.026201616 | FALSE | TRUE | FALSE |
| PYGO2 | 0.597739 | 0.68949613 | 1 | 0.597738955 | 0.597738955 | 11.8630675 | 0.63533959 | 0.697035225 | 1 | 0.63533959 | 0.63533959 | 0.63533959 | 0.940818051 | -0.026494359 | FALSE | TRUE | FALSE |
| TAAR1 | 0.5977127 | 0.68955366 | 0.50225 | 2.391128033 | 9.568011286 | 9.568011286 | 0.75408952 | 0.639689897 | 0.7546 | 0.754089518 | 3.016766488 | 3.016766488 | 0.792628369 | -0.100930388 | FALSE | TRUE | FALSE |
| C9orf43 | 0.5977055 | 0.68961117 | 0.01265 | 0.597705487 | 0.597705487 | 1.337346145 | 0.09178328 | 0.877786373 | 0.50735 | 0.819632843 | 0.819632843 | 1.821505072 | 6.51213935 | 0.813723685 | FALSE | TRUE | FALSE |
| ZNF148 | 0.5976111 | 0.68966865 | 1 | 0.597611129 | 0.597611129 | 11.78693984 | 0.63540515 | 0.696976231 | 1 | 0.63540515 | 0.63540515 | 12.55322382 | 0.940519806 | -0.026632054 | FALSE | TRUE | FALSE |
| PEBP1 | 0.597568 | 0.68972612 | 1 | 0.597567977 | 0.597567977 | 0.597567977 | 0.63504357 | 0.697212062 | 1 | 0.63504357 | 0.63504357 | 0.63504357 | 0.940987367 | -0.026416207 | FALSE | TRUE | FALSE |
| RBBP5 | 0.5974447 | 0.68978355 | 1 | 0.597444743 | 0.597444743 | 11.83946069 | 0.63519683 | 0.697094195 | 1 | 0.635196826 | 0.635196826 | 12.60145325 | 0.940566322 | -0.026610575 | FALSE | TRUE | FALSE |
| GFRA2 | 0.597308 | 0.68984097 | 1 | 0.597307979 | 0.597307979 | 11.83945035 | 0.63500917 | 0.697329827 | 1 | 0.635009171 | 0.635009171 | 12.60026137 | 0.940628902 | -0.026581681 | FALSE | TRUE | FALSE |
| MT3 | 0.597261 | 0.68989837 | 0.0016 | 0.597260983 | 0.597260983 | 0.597260983 | 0.6343322 | 0.697623807 | 1 | 0.634332199 | 0.634332199 | 0.634332199 | 0.941558672 | -0.026152612 | FALSE | TRUE | FALSE |
| CXCL11 | 0.5971062 | 0.68995574 | 1 | 0.597106212 | 0.597106212 | 0.597106212 | 0.63412981 | 0.697799904 | 1 | 0.634129812 | 0.634129812 | 0.634129812 | 0.941615109 | -0.026126581 | FALSE | TRUE | FALSE |
| DEFB115 | 0.5970428 | 0.69001309 | 1 | 5.595205875 | 5.595205875 | 5.595205875 | 0.65437492 | 0.687526334 | 1 | 0.654374917 | 0.654374917 | 0.654374917 | 0.912386404 | -0.039821195 | FALSE | TRUE | FALSE |
| SPDEF | 0.5966719 | 0.69007042 | 0.0112 | 0.596671887 | 0.596671887 | 11.82852797 | 0.63432836 | 0.69768253 | 1 | 0.634328355 | 0.634328355 | 12.58833244 | 0.940635685 | -0.026578549 | FALSE | TRUE | FALSE |
| FAM131B | 0.5963198 | 0.69012773 | 1 | 0.59631982 | 0.59631982 | 11.83904737 | 5.78759555 | 0.27509713 | 0.009 | 5.787595554 | 5.787595554 | 5.787595554 | 0.103034121 | -0.98701893 | FALSE | FALSE | FALSE |
| VWA8 | 0.5963169 | 0.69018501 | 0.3803 | 0.074025209 | 4.918984678 | 95.07502389 | 0.0043461 | 0.898241428 | 0.5151 | 0.034340152 | 0.666422183 | 2.308454117 | 137.2073599 | 2.137377408 | FALSE | TRUE | FALSE |
| RPH3A | 0.5961234 | 0.69024228 | 0.51605 | 0.596123363 | 0.596123363 | 11.72832253 | 0.17339109 | 0.858749594 | 1 | 6.478536655 | 248.4981238 | 248.4981238 | 3.438027599 | 0.536309359 | FALSE | TRUE | FALSE |
| CARM1 | 0.5960877 | 0.69029952 | 1 | 0.596087731 | 0.596087731 | 11.76380698 | 0.63062986 | 0.69908482 | 1 | 0.630629855 | 0.630629855 | 12.46533851 | 0.945225993 | -0.024464344 | FALSE | TRUE | FALSE |
| GMEB2 | 0.595947 | 0.69035674 | 1 | 0.595947011 | 0.595947011 | 11.74003301 | 0.63352152 | 0.698034359 | 0.0159 | 0.633521519 | 0.633521519 | 12.50279069 | 0.940689454 | -0.026553725 | FALSE | TRUE | FALSE |
| LGALS8 | 0.5954751 | 0.69041394 | 0.75575 | 0.595475058 | 5.407222309 | 5.407222309 | 632.082123 | 0.003402238 | 0.06685 | 7.01993533 | 632.0821234 | 632.0821234 | 0.000942085 | -3.025909933 | FALSE | FALSE | FALSE |
| ZBTB14 | 0.5946288 | 0.69047113 | 1 | 0.594628774 | 0.594628774 | 11.78265598 | 0.63252026 | 0.698151457 | 1 | 0.632520258 | 0.632520258 | 12.54741468 | 0.940094433 | -0.026828519 | FALSE | TRUE | FALSE |
| CLHC1 | 0.594464 | 0.69052829 | 0.74865 | 0.594463989 | 5.41593152 | 5.41593152 | 0.791623 | 0.623433917 | 0.87065 | 7.261059474 | 68.3123538 | 68.3123538 | 0.750943299 | -0.124392854 | FALSE | TRUE | FALSE |

| | | | | | | | | | | | | | | | | | |
|---|---|---|---|---|---|---|---|---|---|---|---|---|---|---|---|---|---|
| PRMT1 | 0.5944391 | 0.69058543 | 1 | 0.594439058 | 0.594439058 | 11.76472489 | 0.63238777 | 0.698209972 | 1 | 0.632387771 | 0.632387771 | 12.5314899 | 0.939991386 | -0.026876126 | FALSE | TRUE | FALSE |
| KRTAP6-2 | 0.5943199 | 0.69064255 | 1 | 0.594319861 | 0.594319861 | 0.594319861 | 0.63159166 | 0.698502189 | 1 | 0.631591658 | 0.631591658 | 0.631591658 | 0.94098751 | -0.026416141 | FALSE | TRUE | FALSE |
| TEX264 | 0.5943126 | 0.69069965 | 1 | 0.594312592 | 0.594312592 | 0.594312592 | 0.63196533 | 0.698443792 | 1 | 0.631965334 | 0.631965334 | 0.631965334 | 0.940419608 | -0.026678324 | FALSE | TRUE | FALSE |
| PRH2 | 0.5943105 | 0.69075672 | 1 | 5.579256884 | 5.579256884 | 5.579256884 | 53.6205685 | 0.051324356 | 0.12755 | 5.567740841 | 529.6439521 | 529.6439521 | 0.011083629 | -1.955318014 | FALSE | FALSE | FALSE |
| BRF1 | 0.5941201 | 0.69081377 | 0.51315 | 0.594120053 | 0.594120053 | 11.6864642 | 0.09252963 | 0.877634935 | 1 | 0.092529627 | 0.092529627 | 1.823689865 | 6.420862972 | 0.807593402 | FALSE | TRUE | FALSE |
| CELF2 | 0.5940862 | 0.6908708 | 1 | 0.594086172 | 0.594086172 | 11.74726787 | 0.63207802 | 0.698326932 | 1 | 0.632078021 | 0.632078021 | 12.51552222 | 0.939893734 | -0.026921246 | FALSE | TRUE | FALSE |
| CCL5 | 0.5936587 | 0.69092781 | 0.00315 | 0.593658737 | 0.593658737 | 0.593658737 | 0.63089145 | 0.698968488 | 1 | 0.630891454 | 0.630891454 | 0.630891454 | 0.940983956 | -0.026417781 | FALSE | TRUE | FALSE |
| TRIM39 | 0.5934466 | 0.69098479 | 1 | 0.593446557 | 0.593446557 | 11.73503978 | 0.63147829 | 0.698560562 | 1 | 0.63147829 | 0.63147829 | 12.50404095 | 0.939773491 | -0.02697681 | FALSE | TRUE | FALSE |
| CBX8 | 0.5933608 | 0.69104176 | 1 | 0.593360845 | 0.593360845 | 11.73504524 | 0.6313844 | 0.698677233 | 1 | 0.631384396 | 0.631384396 | 12.50377797 | 0.939777493 | -0.02697496 | FALSE | TRUE | FALSE |
| CTAGE15 | 0.5932152 | 0.6910987 | 0.0188 | 0.593215179 | 0.593215179 | 11.70829256 | 57.2252774 | 0.048930017 | 0.2591 | 5.882863454 | 57.22527742 | 116.2938487 | 0.010366314 | -1.984375651 | FALSE | FALSE | FALSE |
| NMB | 0.5931339 | 0.69115562 | 1 | 0.593133949 | 0.593133949 | 0.593133949 | 5.75605764 | 0.275719152 | 0.0032 | 5.756057641 | 5.756057641 | 5.756057641 | 0.103045172 | -0.986972352 | FALSE | FALSE | FALSE |
| MVB12B | 0.5931058 | 0.69121252 | 1 | 0.593105766 | 0.593105766 | 0.593105766 | 0.63085521 | 0.699026665 | 1 | 0.630855209 | 0.630855209 | 0.630855209 | 0.94016148 | -0.026797547 | FALSE | TRUE | FALSE |
| RPL23 | 0.5930073 | 0.6912694 | 1 | 0.593007281 | 0.593007281 | 0.593007281 | 0.61119094 | 0.707219346 | 1 | 0.611190935 | 5.792700359 | 5.792700359 | 0.970248815 | -0.013116879 | FALSE | TRUE | FALSE |
| ARHGEF35 | 0.5929438 | 0.69132625 | 1 | 5.593549161 | 5.593549161 | 11.7612004 | 5.9554627 | 0.270415338 | 0.0108 | 5.955462699 | 5.955462699 | 12.52446433 | 0.099563005 | -1.001902006 | FALSE | FALSE | FALSE |
| ZMPSTE24 | 0.59246 | 0.69138309 | 0.5085 | 0.592459958 | 5.404991832 | 5.404991832 | 0.11037099 | 0.873238561 | 0.75285 | 0.247426227 | 0.989264209 | 0.989264209 | 5.367895782 | 0.729804075 | FALSE | TRUE | FALSE |
| CEBPD | 0.59238 | 0.6914399 | 1 | 0.592380048 | 0.592380048 | 0.592380048 | 0.60665237 | 0.709235245 | 1 | 0.606652369 | 0.606652369 | 0.606652369 | 0.976473641 | -0.010339475 | FALSE | TRUE | FALSE |
| TMOD2 | 0.5922564 | 0.69149669 | 0.5007 | 0.59225643 | 0.59225643 | 0.59225643 | 0.18746979 | 0.855127337 | 0.00705 | 0.187469792 | 0.187469792 | 0.187469792 | 3.159209942 | 0.499578487 | FALSE | TRUE | FALSE |
| FKBP15 | 0.5920886 | 0.69155346 | 0.395 | 0.271451639 | 2.368614684 | 11.58369927 | 0.10699904 | 0.874103448 | 0.9374 | 0.942558087 | 15.0884001 | 31.7801965 | 5.533588161 | 0.743006834 | FALSE | TRUE | FALSE |
| STC1 | 0.5920604 | 0.69161021 | 1 | 0.592060434 | 0.592060434 | 0.592060434 | 0.62977799 | 0.699142958 | 1 | 0.629777994 | 0.629777994 | 0.629777994 | 0.940109752 | -0.026821442 | FALSE | TRUE | FALSE |
| MRPS6 | 0.5920296 | 0.69166693 | 1 | 0.592029581 | 0.592029581 | 0.592029581 | 0.6296954 | 0.699201073 | 1 | 0.629695397 | 0.629695397 | 0.629695397 | 0.940184071 | -0.026787111 | FALSE | TRUE | FALSE |
| ARPC4 | 0.5918972 | 0.69172363 | 1 | 0.591897172 | 0.591897172 | 0.591897172 | 0.62212805 | 0.702758468 | 1 | 2.488457329 | 2.488457329 | 2.488457329 | 0.951407307 | -0.021633518 | FALSE | TRUE | FALSE |
| TMEM203 | 0.5917466 | 0.69178031 | 1 | 0.591746607 | 0.591746607 | 0.591746607 | 0.62931538 | 0.69937527 | 1 | 0.629315383 | 0.629315383 | 0.629315383 | 0.94030215 | -0.026732571 | FALSE | TRUE | FALSE |
| ACER3 | 0.5917366 | 0.69183697 | 1 | 2.36689945 | 2.36689945 | 2.36689945 | 0.6365894 | 0.696147709 | 1 | 0.636589402 | 0.636589402 | 0.636589402 | 0.929542092 | -0.03173094 | FALSE | TRUE | FALSE |
| KCNE4 | 0.5916743 | 0.6918936 | 1 | 0.591674294 | 0.591674294 | 0.591674294 | 0.62966746 | 0.699259162 | 1 | 0.629667457 | 0.629667457 | 0.629667457 | 0.939661542 | -0.027028547 | FALSE | TRUE | FALSE |
| EMC3 | 0.5910768 | 0.69195022 | 1 | 0.591076846 | 0.591076846 | 0.591076846 | 0.62900015 | 0.699433291 | 1 | 0.629000154 | 0.629000154 | 0.629000154 | 0.939708588 | -0.027006804 | FALSE | TRUE | FALSE |
| IL4 | 0.5909182 | 0.69200682 | 1 | 0.590918216 | 0.590918216 | 0.590918216 | 0.62842635 | 0.699665141 | 1 | 0.628426354 | 0.628426354 | 0.628426354 | 0.940314187 | -0.026727011 | FALSE | TRUE | FALSE |
| EIF1AD | 0.5907299 | 0.69206339 | 1 | 0.590729892 | 0.590729892 | 0.590729892 | 5.73067691 | 0.276340692 | 0.49675 | 0.609001426 | 5.730676913 | 5.730676913 | 0.103082044 | -0.986816977 | FALSE | FALSE | FALSE |
| PRAC1 | 0.5907244 | 0.69211994 | 1 | 0.590724407 | 0.590724407 | 0.590724407 | 0.62812652 | 0.700012192 | 1 | 0.62812652 | 0.62812652 | 0.62812652 | 0.940454492 | -0.026662215 | FALSE | TRUE | FALSE |
| JUNB | 0.5906623 | 0.69217647 | 1 | 2.362626231 | 2.362626231 | 11.71480846 | 0.61632701 | 0.705229377 | 1 | 0.616327015 | 0.616327015 | 0.616327015 | 0.958358604 | -0.018471954 | FALSE | TRUE | FALSE |
| FAM78B | 0.5906065 | 0.69223298 | 1 | 0.590606462 | 0.590606462 | 0.590606462 | 0.62865637 | 0.699549263 | 1 | 0.628656374 | 0.628656374 | 0.628656374 | 0.93947423 | -0.027115128 | FALSE | TRUE | FALSE |
| TIMM17A | 0.5905205 | 0.69228946 | 1 | 0.59052046 | 0.59052046 | 0.59052046 | 0.62813564 | 0.699954411 | 1 | 0.628135635 | 0.628135635 | 0.628135635 | 0.940116157 | -0.026818483 | FALSE | TRUE | FALSE |
| NKX6-3 | 0.5903425 | 0.69234593 | 1 | 2.361283916 | 9.447152752 | 9.447152752 | 0.65073182 | 0.689279527 | 1 | 0.650731817 | 0.650731817 | 0.650731817 | 0.907197823 | -0.042298001 | FALSE | TRUE | FALSE |
| C16orf92 | 0.590323 | 0.69240237 | 1 | 0.590323036 | 0.590323036 | 0.590323036 | 0.62792608 | 0.70006995 | 1 | 0.62792608 | 0.62792608 | 0.62792608 | 0.940115492 | -0.026818791 | FALSE | TRUE | FALSE |
| RAB6B | 0.5901942 | 0.69245879 | 1 | 0.590194247 | 0.590194247 | 0.590194247 | 0.62665244 | 0.700473601 | 1 | 0.626652442 | 0.626652442 | 0.626652442 | 0.941820709 | -0.026031765 | FALSE | TRUE | FALSE |
| C2orf49 | 0.5901808 | 0.69251519 | 1 | 5.546434711 | 53.49688716 | 53.49688716 | 0.66864596 | 0.681024923 | 1 | 0.668645957 | 0.668645957 | 0.668645957 | 0.882650716 | -0.054211122 | FALSE | TRUE | FALSE |
| PEX14 | 0.590165 | 0.69257156 | 1 | 0.590164977 | 0.590164977 | 11.69249232 | 0.62827946 | 0.699838776 | 1 | 0.628279456 | 0.628279456 | 12.46168113 | 0.93933515 | -0.027179426 | FALSE | TRUE | FALSE |
| TRA2B | 0.5900145 | 0.69262791 | 1 | 0.590014513 | 0.590014513 | 11.69809692 | 0.62484909 | 0.701220292 | 1 | 0.624849085 | 2.49937925 | 2.49937925 | 0.944251224 | -0.024912444 | FALSE | TRUE | FALSE |
| CD79A | 0.5897764 | 0.69268424 | 1 | 0.589776437 | 0.589776437 | 0.589776437 | 0.62775154 | 0.700185393 | 1 | 0.627751544 | 0.627751544 | 0.627751544 | 0.939506151 | -0.027100372 | FALSE | TRUE | FALSE |
| ZNF121 | 0.5897261 | 0.69274055 | 1 | 0.589726115 | 0.589726115 | 11.71664976 | 0.62785589 | 0.700127683 | 1 | 0.62785589 | 0.62785589 | 0.62785589 | 0.939269861 | -0.027209613 | FALSE | TRUE | FALSE |
| USP20 | 0.5896534 | 0.69279684 | 0.5244 | 0.589653377 | 0.589653377 | 11.50637333 | 0.09224621 | 0.877739085 | 1 | 0.092246208 | 0.092246208 | 1.805346291 | 6.392169308 | 0.80564827 | FALSE | TRUE | FALSE |
| PDIK1L | 0.5895894 | 0.69285311 | 1 | 0.589589445 | 0.589589445 | 0.589589445 | 0.62568549 | 0.700703772 | 1 | 0.625685489 | 0.625685489 | 0.625685489 | 0.942309603 | -0.025806383 | FALSE | TRUE | FALSE |
| SLC6A3 | 0.5894272 | 0.69290935 | 0.5128 | 0.589427154 | 0.589427154 | 11.60280432 | 0.18493624 | 0.855749856 | 1 | 0.739498349 | 0.739498349 | 6.264780688 | 3.187191195 | 0.503408117 | FALSE | TRUE | FALSE |
| CMPK1 | 0.5893643 | 0.69296557 | 1 | 0.589364298 | 0.589364298 | 0.589364298 | 0.62734087 | 0.700358384 | 1 | 0.62734087 | 0.62734087 | 0.62734087 | 0.939464215 | -0.027119758 | FALSE | TRUE | FALSE |
| MDM2 | 0.589117 | 0.69302177 | 1 | 5.565812201 | 5.565812201 | 11.6880838 | 0.64741618 | 0.691073445 | 0.0095 | 0.64741618 | 0.64741618 | 0.64741618 | 0.909951024 | -0.040981982 | FALSE | TRUE | FALSE |
| PPIL3 | 0.5889915 | 0.69307795 | 1 | 0.588991473 | 0.588991473 | 0.588991473 | 0.60758356 | 0.708910356 | 1 | 5.753814146 | 5.753814146 | 5.753814146 | 0.969399958 | -0.013497004 | FALSE | TRUE | FALSE |
| CCL16 | 0.5887903 | 0.69313411 | 1 | 0.588790289 | 0.588790289 | 0.588790289 | 0.5883267 | 0.717574522 | 1 | 5.516542591 | 53.08862889 | 53.08862889 | 1.000787973 | 0.000342078 | FALSE | TRUE | FALSE |
| FCRL5 | 0.5883342 | 0.69319025 | 0.757 | 0.588334218 | 5.391159651 | 11.55116378 | 0.09284584 | 0.877578057 | 1 | 0.835475147 | 0.835475147 | 16.43962569 | 6.336677982 | 0.801861638 | FALSE | TRUE | FALSE |
| KRTAP9-9 | 0.5882819 | 0.69324637 | 1 | 0.588281903 | 0.588281903 | 0.588281903 | 0.56987621 | 0.724800981 | 1 | 51.25336038 | 505.0539234 | 505.0539234 | 1.032297703 | 0.013804961 | FALSE | TRUE | FALSE |
| PACSIN1 | 0.5882805 | 0.69330247 | 1 | 0.588280518 | 0.588280518 | 11.63699223 | 0.62209298 | 0.70281511 | 1 | 0.622092976 | 0.622092976 | 12.32203239 | 0.94564726 | -0.024270831 | FALSE | TRUE | FALSE |
| BST2 | 0.5874831 | 0.69335855 | 1 | 0.587483081 | 0.587483081 | 0.587483081 | 0.62534598 | 0.700876158 | 1 | 0.625345978 | 0.625345978 | 0.625345978 | 0.939452883 | -0.027124997 | FALSE | TRUE | FALSE |
| LGI3 | 0.5873199 | 0.69341461 | 0.5058 | 0.587319943 | 9.387304482 | 9.387304482 | 0.98201844 | 0.590110764 | 0.0118 | 0.245697205 | 0.982018436 | 2.097752127 | 0.598074254 | -0.223244893 | FALSE | TRUE | FALSE |
| TANK | 0.5873196 | 0.69347065 | 1 | 0.587319586 | 0.587319586 | 0.698049716 | 0.62545784 | 0.70081872 | 1 | 0.625457842 | 0.625457842 | 0.625457842 | 0.939023458 | -0.027323558 | FALSE | TRUE | FALSE |
| CYP4B1 | 0.5872939 | 0.69352666 | 0.9633 | 51.64532141 | 3303.654587 | 32161.49985 | 61.160566 | 0.045688754 | 0.54545 | 85.04494985 | 5441.447432 | 53282.31151 | 0.009602493 | -2.017616021 | FALSE | FALSE | FALSE |
| SPINK14 | 0.5871765 | 0.69358266 | 0.0021 | 0.587176495 | 0.587176495 | 0.587176495 | 0.62462103 | 0.701277565 | 1 | 0.624621034 | 0.624621034 | 0.624621034 | 0.94005239 | -0.026847942 | FALSE | TRUE | FALSE |
| KCTD12 | 0.5871732 | 0.69363863 | 1 | 0.587173219 | 0.587173219 | 11.63937655 | 0.62549723 | 0.700761258 | 1 | 0.625497233 | 0.625497233 | 0.625497233 | 0.938730323 | -0.027459153 | FALSE | TRUE | FALSE |
| LSM11 | 0.5868798 | 0.69369458 | 1 | 5.541891396 | 5.541891396 | 11.65954786 | 0.64519215 | 0.692358693 | 1 | 0.645192146 | 0.645192146 | 0.645192146 | 0.909620179 | -0.041139914 | FALSE | TRUE | FALSE |
| UBAP1 | 0.5868648 | 0.69375051 | 1 | 0.586864807 | 0.586864807 | 11.6310753 | 0.62508298 | 0.700990965 | 1 | 0.625082982 | 0.625082982 | 12.40200602 | 0.938859037 | -0.027399609 | FALSE | TRUE | FALSE |
| CDV3 | 0.5868521 | 0.69380641 | 1 | 0.586852143 | 0.586852143 | 0.586852143 | 0.62488304 | 0.701162996 | 1 | 0.624883041 | 0.624883041 | 0.624883041 | 0.939139174 | -0.027270044 | FALSE | TRUE | FALSE |
| MORF4L1 | 0.5862227 | 0.6938623 | 1 | 0.586222686 | 0.586222686 | 11.64875436 | 0.62445755 | 0.701449241 | 1 | 0.62445755 | 0.62445755 | 0.62445755 | 0.938771076 | -0.0274403 | FALSE | TRUE | FALSE |
| GTF2A2 | 0.5861914 | 0.69391817 | 1 | 5.504013513 | 53.04027082 | 53.04027082 | 0.66454002 | 0.683180614 | 1 | 0.664540018 | 0.664540018 | 0.664540018 | 0.882100974 | -0.054481698 | FALSE | TRUE | FALSE |
| UGCG | 0.5859655 | 0.69397402 | 1 | 0.585965543 | 0.585965543 | 0.585965543 | 0.62421881 | 0.70150642 | 1 | 0.624218813 | 0.624218813 | 0.624218813 | 0.938718172 | -0.027464775 | FALSE | TRUE | FALSE |
| CAMSAP3 | 0.5859221 | 0.69402984 | 0.51655 | 0.585922069 | 0.585922069 | 11.40792401 | 0.07532657 | 0.881356942 | 1 | 0.075326573 | 0.075326573 | 1.471295144 | 7.778424642 | 0.890891649 | FALSE | TRUE | FALSE |
| MOBP | 0.5855439 | 0.69408565 | 0.0023 | 0.585543917 | 0.585543917 | 0.585543917 | 0.62373022 | 0.701734907 | 1 | 0.623730219 | 0.623730219 | 0.623730219 | 0.938777534 | -0.027437312 | FALSE | TRUE | FALSE |
| CMKLR1 | 0.5855224 | 0.69414143 | 0.5044 | 0.585522362 | 0.585522362 | 0.585522362 | 1.66174574 | 0.521168195 | 0.50205 | 1.661745739 | 1.661745739 | 1.661745739 | 0.352353761 | -0.453021088 | FALSE | FALSE | FALSE |
| EIF5A | 0.5854679 | 0.69419719 | 1 | 0.585467899 | 0.585467899 | 0.585467899 | 0.62304517 | 0.702019997 | 1 | 0.623045165 | 0.623045165 | 0.623045165 | 0.939687732 | -0.027016443 | FALSE | TRUE | FALSE |
| CALML5 | 0.5853982 | 0.69425293 | 1 | 0.585398161 | 0.585398161 | 0.585398161 | 0.61661104 | 0.705118002 | 1 | 2.466326217 | 2.466326217 | 2.466326217 | 0.949379949 | -0.022559945 | FALSE | TRUE | FALSE |
| NEGR1 | 0.5853112 | 0.69430865 | 1 | 5.532028905 | 53.66180947 | 53.66180947 | 0.61670802 | 0.705062281 | 1 | 12.25600922 | 226.4186553 | 226.4186553 | 0.949089733 | -0.022692724 | FALSE | TRUE | FALSE |
| BCL7A | 0.5852755 | 0.69436435 | 1 | 0.585275538 | 0.585275538 | 0.585275538 | 0.62337463 | 0.701849013 | 1 | 0.623374627 | 0.623374627 | 0.623374627 | 0.938882515 | -0.027388749 | FALSE | TRUE | FALSE |
| CALM3 | 0.5848877 | 0.69442003 | 1 | 0.584887653 | 0.584887653 | 0.584887653 | 0.62299882 | 0.702190767 | 1 | 0.622998819 | 0.622998819 | 0.622998819 | 0.938826263 | -0.02741477 | FALSE | TRUE | FALSE |
| PLAC8 | 0.5848736 | 0.69447568 | 0.0041 | 0.584873595 | 0.584873595 | 0.584873595 | 0.62292562 | 0.702304494 | 1 | 0.622925623 | 0.622925623 | 0.622925623 | 0.938914009 | -0.027374181 | FALSE | TRUE | FALSE |
| LY6E | 0.5848695 | 0.69453131 | 1 | 0.584869468 | 0.584869468 | 0.584869468 | 0.62302689 | 0.702076945 | 1 | 0.623026886 | 0.623026886 | 0.623026886 | 0.938754782 | -0.027447838 | FALSE | TRUE | FALSE |
| RPSA | 0.5848169 | 0.69458692 | 1 | 0.58481688 | 0.58481688 | 0.58481688 | 0.61212281 | 0.706669433 | 1 | 0.61212281 | 0.61212281 | 0.61212281 | 0.955391419 | -0.019818664 | FALSE | TRUE | FALSE |
| PGA3 | 0.584599 | 0.69464252 | 1 | 0.584599045 | 0.584599045 | 11.58399662 | 0.60478968 | 0.710044158 | 1 | 0.604789684 | 0.604789684 | 11.99739251 | 0.966615437 | -0.014746274 | FALSE | TRUE | FALSE |

| | | | | | | | | | | | | | | | | | |
|---|---|---|---|---|---|---|---|---|---|---|---|---|---|---|---|---|---|
| RNF4 | 0.5845751 | 0.69469808 | 1 | 0.584575138 | 0.584575138 | 0.584575138 | 0.62298954 | 0.702247642 | 1 | 0.622989541 | 0.622989541 | 0.622989541 | 0.938338607 | -0.027640415 | FALSE | TRUE | FALSE |
| RAB21 | 0.5844774 | 0.69475363 | 1 | 0.58447742 | 0.58447742 | 0.58447742 | 0.62281528 | 0.702361322 | 1 | 0.622815282 | 0.622815282 | 0.622815282 | 0.93844425 | -0.027591522 | FALSE | TRUE | FALSE |
| CST9L | 0.5842356 | 0.69480916 | 1 | 0.584235596 | 0.584235596 | 0.584235596 | 0.62205468 | 0.702871728 | 1 | 0.622054677 | 0.622054677 | 0.622054677 | 0.939202963 | -0.027240546 | FALSE | TRUE | FALSE |
| RRAS2 | 0.5841954 | 0.69486466 | 1 | 0.584195398 | 0.584195398 | 0.584195398 | 0.62267877 | 0.702474908 | 1 | 0.622678769 | 0.622678769 | 0.622678769 | 0.938197073 | -0.027705926 | FALSE | TRUE | FALSE |
| AKIRIN1 | 0.5841409 | 0.69492015 | 1 | 0.584140939 | 0.584140939 | 0.584140939 | 0.62256128 | 0.702531666 | 1 | 0.622561285 | 0.622561285 | 0.622561285 | 0.938286646 | -0.027664465 | FALSE | TRUE | FALSE |
| ACYP2 | 0.5840708 | 0.69497561 | 1 | 0.584070832 | 0.584070832 | 0.584070832 | 0.62215189 | 0.702701802 | 1 | 0.622151888 | 0.622151888 | 0.622151888 | 0.938791383 | -0.027430905 | FALSE | TRUE | FALSE |
| TRIM5 | 0.5838284 | 0.69503105 | 0.8757 | 5.404889611 | 205.2395732 | 205.2395732 | 0.095531 | 0.876997051 | 0.50615 | 0.868278115 | 0.868278115 | 0.868278115 | 6.111402502 | 0.786140888 | FALSE | TRUE | FALSE |
| NR1H3 | 0.5837922 | 0.69508647 | 0.703 | 1.885849814 | 165.9539521 | 345.2150594 | 0.22932002 | 0.845069188 | 0.1336 | 0.22932002 | 0.916755623 | 1.135873535 | 2.545753242 | 0.405816306 | FALSE | TRUE | FALSE |
| TTC9B | 0.5837558 | 0.69514187 | 0.5056 | 5.502939493 | 5.502939493 | 5.502939493 | 0.64204544 | 0.693390855 | 1 | 0.642045439 | 0.642045439 | 0.642045439 | 0.909212669 | -0.041334522 | FALSE | TRUE | FALSE |
| NDUFA3 | 0.5836702 | 0.69519724 | 1 | 0.583670171 | 0.583670171 | 0.583670171 | 5.64846826 | 0.280358287 | 0.00275 | 0.602400795 | 5.648468265 | 5.648468265 | 0.103332469 | -0.985763194 | FALSE | FALSE | FALSE |
| CAPZB | 0.5835977 | 0.6952526 | 0.007 | 0.583597678 | 0.583597678 | 0.583597678 | 0.62200271 | 0.702928323 | 1 | 0.622002706 | 0.622002706 | 0.622002706 | 0.93825585 | -0.027678719 | FALSE | TRUE | FALSE |
| PRR3 | 0.5835525 | 0.69530793 | 1 | 0.583552497 | 0.583552497 | 0.583552497 | 0.62174353 | 0.702984896 | 1 | 0.621743529 | 0.621743529 | 0.621743529 | 0.9385743 | -0.027531342 | FALSE | TRUE | FALSE |
| NME4 | 0.5831971 | 0.69536325 | 0.5027 | 0.583197123 | 0.583197123 | 0.583197123 | 0.80642666 | 0.618886668 | 0.0059 | 0.806426655 | 0.806426655 | 0.806426655 | 0.723186814 | -0.140749501 | FALSE | TRUE | FALSE |
| DMRTB1 | 0.5831808 | 0.69541854 | 1 | 0.583180754 | 0.583180754 | 0.583180754 | 0.62169996 | 0.703097971 | 1 | 0.621699955 | 0.621699955 | 0.621699955 | 0.938042137 | -0.027777653 | FALSE | TRUE | FALSE |
| MOB2 | 0.5831152 | 0.69547381 | 1 | 0.583115153 | 0.583115153 | 0.583115153 | 0.60220845 | 0.71127528 | 0.49885 | 0.60220845 | 5.686839303 | 5.686839303 | 0.968294538 | -0.013992518 | FALSE | TRUE | FALSE |
| BOLA3 | 0.5829631 | 0.69552906 | 1 | 0.58296307 | 0.58296307 | 0.58296307 | 5.68615307 | 0.278200689 | 0.4934 | 0.602031952 | 5.686153072 | 5.686153072 | 0.102523281 | -0.989177503 | FALSE | FALSE | FALSE |
| NTRK2 | 0.5829567 | 0.69558429 | 1 | 2581432.475 | 3703567158 | 59232221493 | 22.1131977 | 0.122798648 | 1 | 6.95624E+11 | 1.58936E+16 | 8.22681E+17 | 0.02636239 | -1.579015227 | FALSE | FALSE | FALSE |
| ZG16B | 0.5828262 | 0.69563949 | 1 | 0.582826188 | 0.582826188 | 0.582826188 | 0.62098375 | 0.703267422 | 1 | 0.620983752 | 0.620983752 | 0.620983752 | 0.93855304 | -0.027541179 | FALSE | TRUE | FALSE |
| RANGAP1 | 0.5822551 | 0.69569468 | 0.5056 | 0.582255083 | 0.582255083 | 2.877421525 | 0.17709489 | 0.857788293 | 1 | 1.647362889 | 6.587921711 | 6.66220176 | 3.287814217 | 0.516907269 | FALSE | TRUE | FALSE |
| RRAD | 0.5822403 | 0.69574985 | 0.50225 | 0.58224028 | 0.58224028 | 2.88808975 | 1.68174211 | 0.51881971 | 0.5063 | 1.681742109 | 1.681742109 | 1.681742109 | 0.346212583 | -0.460657152 | FALSE | FALSE | FALSE |
| LTF | 0.5822083 | 0.695805 | 0.25595 | 0.066681922 | 0.582208337 | 1.312063163 | 0.11675483 | 0.872044232 | 0.7525 | 0.116754827 | 1.031920951 | 2.301793835 | 4.986588997 | 0.697803574 | FALSE | TRUE | FALSE |
| TSSK2 | 0.5820365 | 0.69586012 | 0.81515 | 2.328432126 | 9.317200863 | 37.29201182 | 0.21709368 | 0.847907952 | 0.49695 | 0.217093684 | 1.96212013 | 1.96212013 | 2.681038347 | 0.428303026 | FALSE | TRUE | FALSE |
| RASSF3 | 0.5817709 | 0.69591523 | 1 | 0.58177088 | 0.58177088 | 0.58177088 | 0.61988589 | 0.703662021 | 1 | 0.61988589 | 0.61988589 | 0.61988589 | 0.938512861 | -0.027559771 | FALSE | TRUE | FALSE |
| KLF15 | 0.5817327 | 0.69597031 | 1 | 0.581732664 | 0.581732664 | 11.51891945 | 0.62028637 | 0.703549391 | 1 | 0.620286368 | 0.620286368 | 12.29701981 | 0.937845313 | -0.027868787 | FALSE | TRUE | FALSE |
| SLC2A11 | 0.5816277 | 0.69602538 | 0.5035 | 0.581627725 | 5.238770193 | 5.238770193 | 0.1113454 | 0.873076426 | 0.7508 | 0.111345404 | 0.445357083 | 2.203554788 | 5.223634778 | 0.717972805 | FALSE | TRUE | FALSE |
| MLLT1 | 0.5816011 | 0.69608042 | 1 | 0.581601111 | 0.581601111 | 11.46310913 | 0.62036159 | 0.703493043 | 1 | 0.620361593 | 0.620361593 | 12.24843106 | 0.937519533 | -0.028019675 | FALSE | TRUE | FALSE |
| PIAS1 | 0.5814663 | 0.69613544 | 1 | 0.581466342 | 0.581466342 | 11.48865149 | 0.62022817 | 0.703605716 | 1 | 0.620228165 | 0.620228165 | 12.27234612 | 0.93750393 | -0.028026903 | FALSE | TRUE | FALSE |
| PLAC8L1 | 0.5812246 | 0.69619044 | 1 | 0.581224633 | 0.581224633 | 0.581224633 | 0.61951522 | 0.703830798 | 1 | 0.619515219 | 0.619515219 | 0.619515219 | 0.938192663 | -0.027707967 | FALSE | TRUE | FALSE |
| PTER | 0.5810792 | 0.69624542 | 0.51285 | 0.581079189 | 2.323766467 | 2.323766467 | 0.36103244 | 0.805293808 | 0.75305 | 0.361032439 | 1.444183743 | 1.444183743 | 1.609493019 | 0.206689097 | FALSE | TRUE | FALSE |
| NUMA1 | 0.5810513 | 0.69630038 | 0.5284 | 0.581051278 | 1.207766898 | 19.40430014 | 0.09022054 | 0.878125884 | 1 | 0.09022054 | 1.729240274 | 3.019382672 | 6.440343585 | 0.808909037 | FALSE | TRUE | FALSE |
| RSRC2 | 0.5808854 | 0.69635532 | 0.01355 | 0.580885376 | 0.580885376 | 11.51945381 | 0.61957152 | 0.703774561 | 1 | 0.619571517 | 0.619571517 | 12.29913011 | 0.937559845 | -0.028001001 | FALSE | TRUE | FALSE |
| CNP | 0.5808147 | 0.69641024 | 1 | 0.580814706 | 0.580814706 | 11.48326273 | 5.67190875 | 0.278509869 | 0.0122 | 5.671908751 | 5.671908751 | 11.88444106 | 0.102401984 | -0.989691631 | FALSE | FALSE | FALSE |
| SLC39A9 | 0.5807835 | 0.69646513 | 1 | 0.580783494 | 0.580783494 | 0.580783494 | 0.61942838 | 0.703887011 | 1 | 0.619428376 | 0.619428376 | 0.619428376 | 0.937612026 | -0.027976831 | FALSE | TRUE | FALSE |
| CPLX4 | 0.5806928 | 0.69652001 | 1 | 0.580692779 | 0.580692779 | 0.580692779 | 0.61894812 | 0.70416773 | 1 | 0.618948125 | 0.618948125 | 0.618948125 | 0.938192969 | -0.027707826 | FALSE | TRUE | FALSE |
| ACAP1 | 0.5806627 | 0.69657486 | 0.5089 | 0.58066269 | 0.58066269 | 11.40657034 | 0.08823501 | 0.87852917 | 1 | 0.803942909 | 0.803942909 | 15.82654032 | 6.580865205 | 0.818282995 | FALSE | TRUE | FALSE |
| UCK2 | 0.5806215 | 0.69662969 | 1 | 0.580621475 | 0.580621475 | 0.580621475 | 0.6192728 | 0.704111631 | 1 | 0.619272805 | 0.619272805 | 0.619272805 | 0.93758594 | -0.027988914 | FALSE | TRUE | FALSE |
| RPS7 | 0.580604 | 0.6966845 | 1 | 0.580603983 | 0.580603983 | 0.580603983 | 0.61932959 | 0.703999368 | 1 | 0.619329586 | 0.619329586 | 0.619329586 | 0.937471737 | -0.028041816 | FALSE | TRUE | FALSE |
| C3orf18 | 0.5801627 | 0.69673929 | 1 | 0.580162669 | 0.580162669 | 0.580162669 | 0.61840321 | 0.704391906 | 1 | 0.618403207 | 0.618403207 | 0.618403207 | 0.938162453 | -0.027721952 | FALSE | TRUE | FALSE |
| TTC19 | 0.5801567 | 0.69679406 | 0.5098 | 0.580156736 | 2.321169096 | 2.871314102 | 0.21758294 | 0.847822099 | 0.5059 | 0.217582936 | 0.217582936 | 1.078408149 | 2.666370568 | 0.425920507 | FALSE | TRUE | FALSE |
| ZNF524 | 0.5800896 | 0.69684881 | 1 | 0.580089648 | 0.580089648 | 0.580089648 | 0.61865153 | 0.704279864 | 1 | 0.618651531 | 0.618651531 | 0.618651531 | 0.937667845 | -0.027950977 | FALSE | TRUE | FALSE |
| MAPKAPK2 | 0.5800511 | 0.69690354 | 1 | 0.580051117 | 0.580051117 | 11.50930021 | 0.61867457 | 0.704223809 | 1 | 0.618674572 | 0.618674572 | 12.28733371 | 0.937570645 | -0.027995999 | FALSE | TRUE | FALSE |
| RAMP2 | 0.580013 | 0.69695824 | 1 | 0.580012957 | 0.580012957 | 0.580012957 | 0.61817784 | 0.704503858 | 1 | 0.618177844 | 0.618177844 | 0.618177844 | 0.938262285 | -0.02767574 | FALSE | TRUE | FALSE |
| TSKU | 0.5798749 | 0.69701293 | 0.0114 | 0.579874859 | 0.579874859 | 11.48820104 | 0.61848579 | 0.704335896 | 1 | 0.618485795 | 0.618485795 | 12.2670317 | 0.937571831 | -0.02799545 | FALSE | TRUE | FALSE |
| CHCHD10 | 0.5797814 | 0.69706759 | 1 | 0.579781408 | 0.579781408 | 0.579781408 | 0.59883284 | 0.712917118 | 0.50635 | 5.624772533 | 5.624772533 | 5.624772533 | 0.96818573 | -0.014041323 | FALSE | TRUE | FALSE |
| ADM5 | 0.5796577 | 0.69712224 | 1 | 0.579657711 | 0.579657711 | 0.579657711 | 0.61728563 | 0.704894983 | 1 | 0.617285625 | 0.617285625 | 0.617285625 | 0.939042944 | -0.027314546 | FALSE | TRUE | FALSE |
| RHAG | 0.5796073 | 0.69717686 | 1 | 2.318385907 | 9.27568555 | 11.50614229 | 2.47296379 | 0.448870376 | 0.2588 | 2.472963791 | 9.894161699 | 9.894161699 | 0.234377604 | -0.630083889 | FALSE | FALSE | FALSE |
| BSPRY | 0.5794952 | 0.69723146 | 0.7522 | 0.57949522 | 2.318204708 | 2.874695913 | 2.80853897 | 0.426623981 | 0.8124 | 11.23812706 | 44.97963523 | 180.0720875 | 0.206333338 | -0.685430597 | FALSE | FALSE | FALSE |
| RPP25L | 0.5792956 | 0.69728604 | 1 | 0.57929562 | 0.57929562 | 0.57929562 | 0.61787491 | 0.704671617 | 1 | 0.61787491 | 0.61787491 | 0.61787491 | 0.937561326 | -0.028000316 | FALSE | TRUE | FALSE |
| RPL31 | 0.579179 | 0.6973406 | 1 | 0.579179008 | 0.579179008 | 0.579179008 | 0.61757007 | 0.704783345 | 1 | 0.617570074 | 0.617570074 | 0.617570074 | 0.937835287 | -0.027873431 | FALSE | TRUE | FALSE |
| MT1H | 0.5790812 | 0.69739514 | 1 | 0.57908117 | 0.57908117 | 0.57908117 | 5.60414331 | 0.282197384 | 0.5013 | 5.60414331 | 5.60414331 | 5.60414331 | 0.1033309 | -0.98576979 | FALSE | FALSE | FALSE |
| OPN5 | 0.5789869 | 0.69744966 | 1 | 0.578986911 | 0.578986911 | 0.578986911 | 0.61767203 | 0.704727492 | 1 | 0.617672027 | 0.617672027 | 0.617672027 | 0.937369487 | -0.028089187 | FALSE | TRUE | FALSE |
| NPTX1 | 0.5786752 | 0.69750416 | 1 | 0.578675152 | 0.578675152 | 11.45346416 | 0.61753604 | 0.704839174 | 1 | 0.617536043 | 0.617536043 | 12.23787068 | 0.937071056 | -0.028227476 | FALSE | TRUE | FALSE |
| NUCB1 | 0.5783398 | 0.69755865 | 0.87615 | 2.313622304 | 21.25694315 | 21.25694315 | 0.18539755 | 0.855685249 | 1 | 1.709883872 | 1.709883872 | 3.695006729 | 3.119457397 | 0.494079059 | FALSE | TRUE | FALSE |
| NHLH2 | 0.5782323 | 0.69761311 | 1 | 9.251925025 | 37.02199298 | 37.02199298 | 0.63860572 | 0.695135085 | 1 | 0.638605716 | 0.638605716 | 0.638605716 | 0.905460586 | -0.043130449 | FALSE | TRUE | FALSE |
| DEFB129 | 0.5780142 | 0.69766755 | 1 | 0.578014179 | 0.578014179 | 0.578014179 | 0.61628294 | 0.705285031 | 1 | 0.616282942 | 0.616282942 | 0.616282942 | 0.937903907 | -0.027841655 | FALSE | TRUE | FALSE |
| CAMK1D | 0.5773741 | 0.69772198 | 1 | 0.577374098 | 0.577374098 | 11.44390588 | 0.6162022 | 0.705396269 | 1 | 0.616202201 | 0.616202201 | 0.732306964 | 0.936988049 | -0.028265948 | FALSE | TRUE | FALSE |
| TMED2 | 0.5773315 | 0.69777638 | 1 | 0.577331495 | 0.577331495 | 0.577331495 | 0.59242332 | 0.715058807 | 1 | 0.592423322 | 0.592423322 | 0.592423322 | 0.974525266 | -0.011206897 | FALSE | TRUE | FALSE |
| PRLH | 0.5771004 | 0.69783077 | 1 | 0.57710042 | 5.441395573 | 5.441395573 | 0.6356098 | 0.696858167 | 1 | 0.635609799 | 0.635609799 | 0.635609799 | 0.907947646 | -0.041939193 | FALSE | TRUE | FALSE |
| PIAS4 | 0.5769604 | 0.69788513 | 1 | 0.576960393 | 0.576960393 | 11.38202336 | 0.61250348 | 0.706338414 | 1 | 0.612503476 | 0.612503476 | 12.10300938 | 0.941970806 | -0.025962557 | FALSE | TRUE | FALSE |
| CYSTM1 | 0.5767111 | 0.69793948 | 1 | 0.576711115 | 0.576711115 | 0.576711115 | 0.61497745 | 0.705507434 | 1 | 0.614977452 | 0.614977452 | 0.614977452 | 0.937776033 | -0.027900871 | FALSE | TRUE | FALSE |
| LY86 | 0.5764449 | 0.69799381 | 1 | 0.576444851 | 0.576444851 | 0.576444851 | 0.61482603 | 0.705562986 | 1 | 0.614826032 | 0.614826032 | 0.614826032 | 0.937573918 | -0.027994483 | FALSE | TRUE | FALSE |
| MAK16 | 0.5761075 | 0.69804812 | 1 | 0.576107457 | 0.576107457 | 0.576107457 | 5.62886492 | 0.28097217 | 0.50745 | 5.628864919 | 5.628864919 | 5.628864919 | 0.10234878 | -0.98991733 | FALSE | FALSE | FALSE |
| DRAM1 | 0.5756902 | 0.69810241 | 1 | 0.575690239 | 0.575690239 | 0.575690239 | 5.590489 | 0.283721681 | 0.5017 | 5.590488997 | 5.590488997 | 5.590488997 | 0.102976723 | -0.987260931 | FALSE | FALSE | FALSE |
| CX3CL1 | 0.5754691 | 0.69815668 | 1 | 0.575469097 | 0.575469097 | 11.39220237 | 0.61427711 | 0.705784976 | 1 | 0.614277107 | 0.614277107 | 12.17535344 | 0.936823283 | -0.028342324 | FALSE | TRUE | FALSE |
| MBNL2 | 0.575467 | 0.69821093 | 1 | 0.575467018 | 0.575467018 | 11.40975133 | 0.61432338 | 0.705729512 | 1 | 0.614323382 | 0.614323382 | 12.19282985 | 0.936749333 | -0.028376608 | FALSE | TRUE | FALSE |
| RC3H2 | 0.5753768 | 0.69826516 | 0.51015 | 0.575376771 | 0.575376771 | 11.26275127 | 0.08860898 | 0.87846369 | 1 | 0.088608981 | 0.088608981 | 1.73892657 | 6.493436298 | 0.812474584 | FALSE | TRUE | FALSE |
| DNAL1 | 0.575039 | 0.69831937 | 1 | 0.57503898 | 0.57503898 | 0.57503898 | 0.6136524 | 0.706006609 | 1 | 0.613652402 | 0.613652402 | 0.613652402 | 0.937076068 | -0.028225154 | FALSE | TRUE | FALSE |
| ITK | 0.5749694 | 0.69837356 | 0.017 | 0.116379936 | 0.116379936 | 0.984663963 | 0.04466749 | 0.887957272 | 1 | 0.044667494 | 0.044667494 | 0.883432508 | 12.87221055 | 1.109653135 | FALSE | TRUE | FALSE |
| ARC | 0.574786 | 0.69842773 | 0.01465 | 0.574785971 | 0.574785971 | 11.3517742 | 0.57621635 | 0.722075383 | 1 | 5.435320938 | 52.61973156 | 107.5069706 | 0.997517644 | -0.001079414 | FALSE | TRUE | FALSE |
| LIMD2 | 0.5747503 | 0.69848188 | 1 | 0.574750313 | 0.574750313 | 0.574750313 | 5.62095285 | 0.281278876 | 0.0035 | 5.620952854 | 5.620952854 | 5.620952854 | 0.102251403 | -0.990330726 | FALSE | FALSE | FALSE |
| IL26 | 0.5747074 | 0.69853601 | 1 | 0.574707376 | 0.574707376 | 0.574707376 | 2.42559177 | 0.452834049 | 0.00375 | 2.425591774 | 2.425591774 | 2.425591774 | 0.236934913 | -0.62537094 | FALSE | FALSE | FALSE |
| GMEB1 | 0.574312 | 0.69859012 | 1 | 0.574311989 | 0.574311989 | 11.36862724 | 0.61343594 | 0.706117296 | 1 | 0.613435937 | 0.613435937 | 12.15805102 | 0.936221625 | -0.028621332 | FALSE | TRUE | FALSE |
| UCHL1 | 0.5742892 | 0.69864421 | 1 | 5.428980686 | 5.428980686 | 5.428980686 | 0.63299762 | 0.698092918 | 1 | 0.632997618 | 0.632997618 | 0.632997618 | 0.907253378 | -0.042271406 | FALSE | TRUE | FALSE |

| | | | | | | | | | | | | | | | | | |
|---|---|---|---|---|---|---|---|---|---|---|---|---|---|---|---|---|---|
| SLC38A4 | 0.5742228 | 0.69869828 | 0.01345 | 0.574222838 | 0.574222838 | 11.37162976 | 0.61314207 | 0.706172608 | 1 | 0.613142068 | 0.613142068 | 12.15671631 | 0.936524938 | -0.028480653 | FALSE | TRUE | FALSE |
| AREG | 0.574176 | 0.69875233 | 1 | 0.574175958 | 0.574175958 | 0.574175958 | 0.5871171 | 0.717930033 | 1 | 0.587117103 | 0.587117103 | 0.587117103 | 0.977958153 | -0.009679728 | FALSE | TRUE | FALSE |
| TXNL4A | 0.5741671 | 0.69880636 | 1 | 0.574167087 | 0.574167087 | 0.574167087 | 0.61307489 | 0.706227898 | 0.00395 | 0.61307489 | 0.61307489 | 0.61307489 | 0.936536623 | -0.028475235 | FALSE | TRUE | FALSE |
| MAL | 0.5738598 | 0.69886036 | 1 | 0.57385983 | 0.57385983 | 0.57385983 | 0.61286533 | 0.706283166 | 1 | 0.612865325 | 0.612865325 | 0.612865325 | 0.93635552 | -0.028559225 | FALSE | TRUE | FALSE |
| ARL1 | 0.5737609 | 0.69891435 | 1 | 5.407200758 | 52.29963088 | 52.29963088 | 21.4621621 | 0.12645489 | 0.00265 | 5.969457245 | 5.969457245 | 5.969457245 | 0.026733602 | -1.572942525 | FALSE | FALSE | FALSE |
| CCDC69 | 0.5735911 | 0.69896832 | 0.8767 | 0.573591062 | 48.30549862 | 48.30549862 | 0.09134815 | 0.877852525 | 1 | 0.813812468 | 7.44137662 | 7.44137662 | 6.279175693 | 0.797902635 | FALSE | TRUE | FALSE |
| GYS2 | 0.5735014 | 0.69902227 | 0.2082 | 0.066120073 | 2.292761669 | 9.168364001 | 43307.2232 | 4.4768E-05 | 0.2315 | 2703.112573 | 694528.435 | 6405666.706 | 1.32426E-05 | -4.878025853 | FALSE | FALSE | FALSE |
| PSMD4 | 0.5734048 | 0.69907619 | 1 | 0.573404763 | 0.573404763 | 11.37326461 | 0.61246986 | 0.706448843 | 1 | 0.612469858 | 0.612469858 | 12.16018683 | 0.936217114 | -0.028623424 | FALSE | TRUE | FALSE |
| TFDP1 | 0.5733739 | 0.6991301 | 1 | 0.573373926 | 0.573373926 | 11.34772657 | 0.61250003 | 0.70639364 | 1 | 0.612500031 | 0.612500031 | 12.13730493 | 0.936120648 | -0.028668176 | FALSE | TRUE | FALSE |
| MRFAP1L1 | 0.573355 | 0.69918398 | 1 | 0.573354955 | 0.573354955 | 0.573354955 | 0.61201503 | 0.706724524 | 1 | 0.612015029 | 0.612015029 | 0.612015029 | 0.936831496 | -0.028338517 | FALSE | TRUE | FALSE |
| CXCR5 | 0.5733335 | 0.69923785 | 1 | 2.293265985 | 2.293265985 | 11.35229471 | 0.61932429 | 0.704055511 | 1 | 0.619324285 | 0.619324285 | 12.27764891 | 0.925740395 | -0.033510785 | FALSE | TRUE | FALSE |
| KCTD5 | 0.5727382 | 0.69929169 | 1 | 0.572738217 | 0.572738217 | 0.572738217 | 0.61178155 | 0.706889665 | 1 | 0.611781554 | 0.611781554 | 0.611781554 | 0.936180918 | -0.028640215 | FALSE | TRUE | FALSE |
| RABAC1 | 0.5727063 | 0.69934552 | 1 | 0.572706293 | 0.572706293 | 0.572706293 | 0.61157712 | 0.707054603 | 1 | 0.611577115 | 0.611577115 | 0.611577115 | 0.936441667 | -0.028519271 | FALSE | TRUE | FALSE |
| NUDT2 | 0.5726149 | 0.69939933 | 1 | 0.57261491 | 0.57261491 | 0.57261491 | 5.58159052 | 0.284329252 | 0.50045 | 5.581590515 | 5.581590515 | 5.581590515 | 0.102589917 | -0.98889532 | FALSE | FALSE | FALSE |
| SSTR2 | 0.5725775 | 0.69945311 | 1 | 2.290189029 | 2.290189029 | 11.36673972 | 2.29707658 | 0.465478072 | 0.8748 | 21.69133662 | 210.2231511 | 210.2231511 | 0.249263551 | -0.603341222 | FALSE | FALSE | FALSE |
| VGF | 0.5724662 | 0.69950688 | 1 | 0.57246622 | 0.57246622 | 11.24417046 | 0.61178525 | 0.70683464 | 1 | 0.611785253 | 0.611785253 | 12.04238528 | 0.935730662 | -0.028849139 | FALSE | TRUE | FALSE |
| IRF8 | 0.5724462 | 0.69956062 | 0.69455 | 0.57244625 | 9.153843416 | 11.31075164 | 1.98760595 | 0.48986291 | 0.13845 | 0.84184193 | 7.946061208 | 7.946061208 | 0.288007917 | -0.540595574 | FALSE | FALSE | FALSE |
| MPV17 | 0.5722787 | 0.69961434 | 0.75265 | 1.310907302 | 5.242627921 | 5.242627921 | 409.017547 | 0.005980511 | 0.0334 | 11.10954609 | 102.1961072 | 409.0175466 | 0.001399154 | -2.854134352 | FALSE | FALSE | FALSE |
| SH3BGR | 0.5721435 | 0.69966805 | 1 | 0.572143527 | 0.572143527 | 0.572143527 | 0.61099104 | 0.707274217 | 1 | 0.610991041 | 0.610991041 | 0.610991041 | 0.936418848 | -0.028529853 | FALSE | TRUE | FALSE |
| NAA50 | 0.571821 | 0.69972174 | 1 | 0.571821019 | 0.571821019 | 0.571821019 | 0.59165953 | 0.715576123 | 0.00315 | 0.591659527 | 0.591659527 | 0.591659527 | 0.966469721 | -0.014811747 | FALSE | TRUE | FALSE |
| VDAC3 | 0.5717193 | 0.6997754 | 1 | 0.571719336 | 0.571719336 | 0.571719336 | 0.6021451 | 0.711328554 | 1 | 0.602145096 | 0.602145096 | 0.602145096 | 0.949471049 | -0.022518273 | FALSE | TRUE | FALSE |
| ZNF358 | 0.5716428 | 0.69982905 | 1 | 0.57164276 | 0.57164276 | 11.29002182 | 0.5918912 | 0.715317716 | 1 | 5.599844232 | 5.599844232 | 11.70750406 | 0.965790274 | -0.015117173 | FALSE | TRUE | FALSE |
| C11orf68 | 0.5715459 | 0.69988267 | 1 | 0.571545879 | 0.571545879 | 0.571545879 | 0.61046156 | 0.707329071 | 1 | 0.610461564 | 0.610461564 | 0.610461564 | 0.936252031 | -0.028607227 | FALSE | TRUE | FALSE |
| FAM3D | 0.5714581 | 0.69993628 | 1 | 0.571458113 | 0.571458113 | 0.571458113 | 0.61022706 | 0.707438712 | 1 | 0.610227058 | 0.610227058 | 0.610227058 | 0.936468001 | -0.028507058 | FALSE | TRUE | FALSE |
| TNFAIP8L1 | 0.5714234 | 0.69998986 | 1 | 0.571423445 | 0.571423445 | 0.571423445 | 0.610123 | 0.7074935 | 1 | 0.610122998 | 0.610122998 | 0.610122998 | 0.936570899 | -0.028459341 | FALSE | TRUE | FALSE |
| GAS6 | 0.5713271 | 0.70004342 | 0.3242 | 0.142923193 | 2.284417124 | 2.810882988 | 0.01616127 | 0.89481677 | 1 | 0.064580825 | 0.064580825 | 0.547173769 | 35.35162502 | 1.548409382 | FALSE | TRUE | FALSE |
| KRTAP19-8 | 0.5713073 | 0.70009697 | 1 | 0.571307347 | 0.571307347 | 0.571307347 | 0.60991715 | 0.707603011 | 1 | 0.609917147 | 0.609917147 | 0.609917147 | 0.936696648 | -0.028401034 | FALSE | TRUE | FALSE |
| MAPK13 | 0.571249 | 0.70015049 | 0.51125 | 5.245640008 | 49.40736539 | 49.40736539 | 62.7960559 | 0.044557132 | 0.35065 | 62.79605589 | 6046.13328 | 61479.86984 | 0.009096893 | -2.041106924 | FALSE | FALSE | FALSE |
| FST | 0.5712116 | 0.70020399 | 0.0087 | 0.57121164 | 0.57121164 | 0.57121164 | 0.61034349 | 0.707383902 | 1 | 0.61034349 | 0.61034349 | 0.61034349 | 0.935885529 | -0.028777268 | FALSE | TRUE | FALSE |
| PIK3CG | 0.570867 | 0.70025747 | 0.8854 | 5.307033233 | 494.6069157 | 1977.77715 | 0.1144066 | 0.872505984 | 0.51135 | 0.457505427 | 1.829995289 | 8.947667418 | 4.98980837 | 0.698083867 | FALSE | TRUE | FALSE |
| NUBP2 | 0.5707398 | 0.70031094 | 0.7503 | 0.570739811 | 2.283032694 | 2.283032694 | 0.18331481 | 0.856174892 | 1 | 0.183314807 | 0.183314807 | 0.183314807 | 3.113440858 | 0.49324062 | FALSE | TRUE | FALSE |
| METTL15 | 0.5707176 | 0.70036438 | 0.4966 | 0.253514437 | 2.28231008 | 2.28231008 | 0.09094715 | 0.877937483 | 1 | 0.090947146 | 0.090947146 | 0.090947146 | 6.275266518 | 0.797632176 | FALSE | TRUE | FALSE |
| HS3ST5 | 0.5706947 | 0.7004178 | 1 | 0.570694657 | 0.570694657 | 0.570694657 | 0.6099939 | 0.707548267 | 1 | 0.609993902 | 0.609993902 | 0.609993902 | 0.935574363 | -0.028921687 | FALSE | TRUE | FALSE |
| GCSAML | 0.5706116 | 0.7004712 | 1 | 0.570611649 | 0.570611649 | 0.570611649 | 0.60932846 | 0.707931019 | 1 | 0.609328456 | 0.609328456 | 0.609328456 | 0.936459874 | -0.028510827 | FALSE | TRUE | FALSE |
| PLAGL2 | 0.5703438 | 0.70052459 | 1 | 0.570343835 | 0.570343835 | 11.274081 | 0.60967124 | 0.707712435 | 1 | 0.609671235 | 0.609671235 | 12.06835212 | 0.935494086 | -0.028958954 | FALSE | TRUE | FALSE |
| BCL7B | 0.5702764 | 0.70057795 | 1 | 0.570276351 | 0.570276351 | 0.570276351 | 0.6090591 | 0.708149251 | 1 | 0.609059099 | 0.609059099 | 0.609059099 | 0.936323505 | -0.028574074 | FALSE | TRUE | FALSE |
| FAM53B | 0.5700829 | 0.70063129 | 1 | 0.570082926 | 0.570082926 | 11.27512231 | 0.60949072 | 0.70782177 | 1 | 0.60949072 | 0.60949072 | 12.0706258 | 0.935343079 | -0.029029063 | FALSE | TRUE | FALSE |
| FXYD4 | 0.5698803 | 0.70068462 | 1 | 0.569880342 | 0.569880342 | 0.569880342 | 0.60856788 | 0.708203757 | 0.0016 | 0.608567881 | 0.608567881 | 0.608567881 | 0.936428556 | -0.028525351 | FALSE | TRUE | FALSE |
| NTF3 | 0.5698796 | 0.70073792 | 0.01085 | 0.569879611 | 0.569879611 | 11.31067749 | 0.60923078 | 0.70798561 | 1 | 0.609230779 | 0.609230779 | 0.609230779 | 0.935408437 | -0.028998717 | FALSE | TRUE | FALSE |
| NACC1 | 0.5695348 | 0.70079121 | 1 | 0.5695348 | 0.5695348 | 11.24499152 | 0.58990916 | 0.716604723 | 1 | 5.579979256 | 5.579979256 | 11.66519386 | 0.965461863 | -0.015264877 | FALSE | TRUE | FALSE |
| POLD4 | 0.569528 | 0.70084447 | 1 | 0.569528028 | 0.569528028 | 0.569528028 | 0.60830603 | 0.708367148 | 1 | 0.608306035 | 0.608306035 | 0.608306035 | 0.936252471 | -0.028607023 | FALSE | TRUE | FALSE |
| CRH | 0.5694606 | 0.70089772 | 1 | 0.569460592 | 0.569460592 | 0.569460592 | 0.5761689 | 0.722173698 | 1 | 0.5761689 | 0.5761689 | 0.5761689 | 0.988357046 | -0.005086137 | FALSE | TRUE | FALSE |
| MEAF6 | 0.5693935 | 0.70095094 | 0.0069 | 0.569393493 | 0.569393493 | 0.569393493 | 0.62511737 | 0.700933574 | 1 | 2.500455082 | 2.500455082 | 2.500455082 | 0.91085853 | -0.04054907 | FALSE | TRUE | FALSE |
| PROKR1 | 0.5691127 | 0.70100415 | 0.01325 | 0.569112729 | 0.569112729 | 2.818050225 | 0.17853312 | 0.857372398 | 1 | 0.714063595 | 0.714063595 | 3.539909642 | 3.187715181 | 0.503479511 | FALSE | TRUE | FALSE |
| TASP1 | 0.5689676 | 0.70105733 | 0.5044 | 0.568967641 | 0.568967641 | 2.823914822 | 0.18124717 | 0.856750587 | 1 | 0.181247168 | 0.181247168 | 0.181247168 | 3.13918087 | 0.496816339 | FALSE | TRUE | FALSE |
| DDIT3 | 0.568806 | 0.7011105 | 0.00455 | 0.568806006 | 0.568806006 | 0.568806006 | 0.60773289 | 0.70880189 | 1 | 0.607732891 | 0.607732891 | 0.607732891 | 0.935947379 | -0.028748568 | FALSE | TRUE | FALSE |
| TSPYL5 | 0.5687177 | 0.70116365 | 1 | 0.56871765 | 0.56871765 | 11.25906825 | 0.60793737 | 0.708584695 | 1 | 0.607937369 | 0.607937369 | 12.05018621 | 0.935487238 | -0.028962133 | FALSE | TRUE | FALSE |
| PDPK1 | 0.5686158 | 0.70121677 | 1 | 0.568615848 | 0.568615848 | 11.22337058 | 0.60815726 | 0.708421568 | 1 | 0.608157257 | 0.608157257 | 12.02275966 | 0.934981606 | -0.029196933 | FALSE | TRUE | FALSE |
| ZACN | 0.5684569 | 0.70126988 | 0.25055 | 0.065428863 | 0.568456896 | 0.568456896 | 0.05592395 | 0.885460433 | 0.51245 | 0.055923947 | 0.055923947 | 0.277280841 | 10.16482069 | 1.007099722 | FALSE | TRUE | FALSE |
| CDYL | 0.5684097 | 0.70132297 | 1 | 0.568409687 | 0.568409687 | 11.22825579 | 0.60791831 | 0.708639027 | 1 | 0.607918313 | 0.607918313 | 12.02648794 | 0.935009977 | -0.029183755 | FALSE | TRUE | FALSE |
| GPR183 | 0.5683362 | 0.70137604 | 1 | 0.56833617 | 0.56833617 | 0.56833617 | 0.60769793 | 0.708856134 | 1 | 0.607697927 | 0.607697927 | 0.607697927 | 0.935228087 | -0.029082459 | FALSE | TRUE | FALSE |
| ELAVL3 | 0.5682187 | 0.70142908 | 1 | 0.568218684 | 0.568218684 | 11.23973962 | 0.60755971 | 0.708964555 | 1 | 0.607559708 | 0.607559708 | 12.03378712 | 0.935247477 | -0.029073455 | FALSE | TRUE | FALSE |
| WFDC2 | 0.5681102 | 0.70148211 | 1 | 2.272138615 | 2.272138615 | 2.272138615 | 2.42968438 | 0.452458686 | 0.00255 | 2.429684379 | 2.429684379 | 2.429684379 | 0.233820577 | -0.631117271 | FALSE | FALSE | FALSE |
| RNASE1 | 0.5677027 | 0.70153512 | 1 | 0.567702689 | 0.567702689 | 0.567702689 | 0.60684918 | 0.709181149 | 1 | 0.60684918 | 0.60684918 | 0.60684918 | 0.935492224 | -0.028959818 | FALSE | TRUE | FALSE |
| ST3GAL1 | 0.567545 | 0.70158811 | 1 | 0.567545011 | 0.567545011 | 11.2563371 | 2.32617173 | 0.462596962 | 0.7574 | 5.500990598 | 22.00267349 | 22.00267349 | 0.243982421 | -0.612641464 | FALSE | FALSE | FALSE |
| TSPAN10 | 0.5674447 | 0.70164108 | 1 | 0.567444703 | 5.337235652 | 5.337235652 | 0.62636484 | 0.700531177 | 1 | 0.626364835 | 0.626364835 | 0.626364835 | 0.905933205 | -0.042903822 | FALSE | TRUE | FALSE |
| FAM3C | 0.5673371 | 0.70169403 | 1 | 0.567337083 | 0.567337083 | 0.567337083 | 0.6063957 | 0.709343372 | 1 | 0.606395703 | 0.606395703 | 0.606395703 | 0.935588891 | -0.028914943 | FALSE | TRUE | FALSE |
| PCMT1 | 0.5672097 | 0.70174697 | 0.0064 | 0.567209677 | 0.567209677 | 0.567209677 | 0.59950139 | 0.712706385 | 1 | 2.397979196 | 2.397979196 | 2.397979196 | 0.946135716 | -0.024046563 | FALSE | TRUE | FALSE |
| HPCAL1 | 0.5671703 | 0.70179988 | 1 | 5.367837673 | 52.13965106 | 52.13965106 | 0.60660796 | 0.709289319 | 1 | 5.743571794 | 55.81339273 | 55.81339273 | 0.934986503 | -0.029194658 | FALSE | TRUE | FALSE |
| CINP | 0.5669484 | 0.70185277 | 1 | 0.566948409 | 0.566948409 | 0.566948409 | 0.60596622 | 0.709451418 | 1 | 0.605966215 | 0.605966215 | 0.605966215 | 0.935610591 | -0.028904871 | FALSE | TRUE | FALSE |
| CACUL1 | 0.566786 | 0.70190565 | 1 | 0.566786015 | 0.566786015 | 11.24548071 | 0.60627364 | 0.709397405 | 1 | 0.606273638 | 0.606273638 | 0.606273638 | 0.934868316 | -0.029249559 | FALSE | TRUE | FALSE |
| TMEM150A | 0.5665944 | 0.7019585 | 1 | 0.566594361 | 0.566594361 | 0.566594361 | 0.60573559 | 0.709613329 | 1 | 0.605735592 | 0.605735592 | 0.605735592 | 0.935382316 | -0.029010845 | FALSE | TRUE | FALSE |
| EIF5A2 | 0.5665494 | 0.70201133 | 1 | 0.566549448 | 0.566549448 | 0.566549448 | 0.60575059 | 0.709559381 | 1 | 0.605750587 | 0.605750587 | 0.605750587 | 0.935285017 | -0.029056023 | FALSE | TRUE | FALSE |
| CLEC2D | 0.5663254 | 0.70206415 | 1 | 0.566325449 | 0.566325449 | 0.566325449 | 0.60543102 | 0.709667258 | 1 | 0.605431024 | 0.605431024 | 0.605431024 | 0.935408704 | -0.028998593 | FALSE | TRUE | FALSE |
| CCNY | 0.5660844 | 0.70211695 | 1 | 0.566084412 | 0.566084412 | 11.2284115 | 0.60542452 | 0.709721164 | 1 | 0.605424516 | 0.605424516 | 0.605424516 | 0.935020628 | -0.029178808 | FALSE | TRUE | FALSE |
| MC5R | 0.5660595 | 0.70216973 | 0.00985 | 0.566059461 | 0.566059461 | 0.566059461 | 0.08880609 | 0.878379401 | 1 | 0.088806092 | 0.088806092 | 0.088806092 | 6.374106162 | 0.804419292 | FALSE | TRUE | FALSE |
| ACKR3 | 0.5658188 | 0.70222248 | 1 | 0.565818814 | 0.565818814 | 11.21033177 | 0.60538156 | 0.709775049 | 1 | 0.605381564 | 0.605381564 | 12.00769099 | 0.934648242 | -0.029351807 | FALSE | TRUE | FALSE |
| TMEM39A | 0.5653313 | 0.70227523 | 1 | 0.565331286 | 0.565331286 | 11.202537 | 0.6048828 | 0.709936578 | 1 | 0.604882802 | 0.604882802 | 11.99955721 | 0.934612927 | -0.029368217 | FALSE | TRUE | FALSE |
| SLC17A4 | 0.5650962 | 0.70232795 | 0.9987 | 4688.560938 | 5232962.843 | 56764733.46 | 765.076593 | 0.002984357 | 0.11425 | 78.35679088 | 7652.734948 | 78372.84528 | 0.000738614 | -3.131582556 | FALSE | FALSE | FALSE |
| TGFA | 0.5650333 | 0.70238066 | 1 | 0.565033348 | 0.565033348 | 0.565033348 | 0.60410048 | 0.710420017 | 1 | 0.604100476 | 0.604100476 | 0.604100476 | 0.935330082 | -0.029035098 | FALSE | TRUE | FALSE |
| EGR1 | 0.5648898 | 0.70243334 | 0.0168 | 0.564889796 | 0.564889796 | 11.13548615 | 0.60452653 | 0.710151652 | 0.01675 | 0.604526531 | 0.604526531 | 11.93743673 | 0.934433423 | -0.029451636 | FALSE | TRUE | FALSE |
| CEBPB | 0.5648799 | 0.70248601 | 1 | 5.308416857 | 51.19916369 | 51.19916369 | 0.57270558 | 0.72349393 | 1 | 51.87735759 | 513.0359844 | 513.0359844 | 0.986335654 | -0.005975268 | FALSE | TRUE | FALSE |
| MRPL41 | 0.5647923 | 0.70253865 | 1 | 0.564792271 | 0.564792271 | 0.564792271 | 0.60429833 | 0.710259062 | 1 | 0.604298327 | 0.604298327 | 0.604298327 | 0.934624913 | -0.029362647 | FALSE | TRUE | FALSE |

| | | | | | | | | | | | | | | | | | |
|---|---|---|---|---|---|---|---|---|---|---|---|---|---|---|---|---|---|
| OPALIN | 0.5646391 | 0.70259128 | 1 | 0.564639096 | 0.564639096 | 0.564639096 | 0.60371691 | 0.710580782 | 1 | 0.603716914 | 0.603716914 | 0.603716914 | 0.935271288 | -0.029062398 | FALSE | TRUE | FALSE |
| KHSRP | 0.5644042 | 0.70264388 | 1 | 0.564404208 | 0.564404208 | 11.13781634 | 0.6038267 | 0.710527215 | 1 | 0.6038267 | 0.6038267 | 11.93485855 | 0.934712242 | -0.029322069 | FALSE | TRUE | FALSE |
| FAM86B1 | 0.5642718 | 0.70269647 | 1 | 0.564271791 | 0.564271791 | 0.564271791 | 0.57962722 | 0.720989113 | 1 | 0.579627218 | 0.579627218 | 0.579627218 | 0.973508099 | -0.011660431 | FALSE | TRUE | FALSE |
| GOLGA6A | 0.5640391 | 0.70274904 | 1 | 5.348136541 | 5.348136541 | 105.381967 | 0.5850726 | 0.718638353 | 1 | 5.543709326 | 53.91047914 | 109.4299175 | 0.964049757 | -0.01590055 | FALSE | TRUE | FALSE |
| LCE1A | 0.5639539 | 0.70280159 | 1 | 0.563953927 | 0.563953927 | 0.563953927 | 0.6030134 | 0.710794848 | 1 | 0.6030134 | 0.6030134 | 0.6030134 | 0.935226194 | -0.029083338 | FALSE | TRUE | FALSE |
| HNRNPK | 0.5636942 | 0.70285412 | 1 | 0.563694179 | 0.563694179 | 11.1545281 | 0.60332009 | 0.710687857 | 1 | 0.603320093 | 0.603320093 | 11.95386087 | 0.934320249 | -0.029504239 | FALSE | TRUE | FALSE |
| BLID | 0.5635722 | 0.70290664 | 1 | 0.563572195 | 0.563572195 | 0.563572195 | 0.60264874 | 0.71090176 | 1 | 0.602648737 | 0.602648737 | 0.602648737 | 0.935158676 | -0.029114692 | FALSE | TRUE | FALSE |
| NFIL3 | 0.5635595 | 0.70295913 | 1 | 0.563559455 | 0.563559455 | 11.17229421 | 0.54385595 | 0.736488543 | 1 | 49.5767499 | 491.8308249 | 491.8308249 | 1.036229277 | 0.015455858 | FALSE | TRUE | FALSE |
| CNN2 | 0.5635513 | 0.7030116 | 1 | 8059011596 | 1.67314E+14 | 2.71326E+16 | 1.82355716 | 0.505356702 | 1 | 246620361.9 | 3.95564E+11 | 6.00116E+13 | 0.309039543 | -0.509985947 | FALSE | FALSE | FALSE |
| BET1L | 0.563209 | 0.70306405 | 1 | 0.563209009 | 0.563209009 | 0.563209009 | 0.60253252 | 0.710955185 | 1 | 0.602532524 | 0.602532524 | 0.602532524 | 0.934736278 | -0.029310901 | FALSE | TRUE | FALSE |
| FAHD1 | 0.5631349 | 0.70311649 | 0.68815 | 0.563134884 | 9.011401615 | 9.011401615 | 1.90970369 | 0.498282155 | 0.25395 | 0.47765471 | 1.909703692 | 1.909703692 | 0.294880764 | -0.530353556 | FALSE | FALSE | FALSE |
| ATG5 | 0.5629393 | 0.7031689 | 1 | 0.562939325 | 0.562939325 | 0.562939325 | 0.60247945 | 0.711008588 | 1 | 0.602479454 | 0.602479454 | 0.602479454 | 0.934370992 | -0.029480653 | FALSE | TRUE | FALSE |
| CACYBP | 0.5629316 | 0.7032213 | 1 | 0.562931639 | 0.562931639 | 0.562931639 | 0.60230587 | 0.711168668 | 1 | 0.602305875 | 0.602305875 | 0.602305875 | 0.934627509 | -0.029361441 | FALSE | TRUE | FALSE |
| PCGF5 | 0.5629071 | 0.70327367 | 1 | 0.562907068 | 0.562907068 | 0.562907068 | 0.58935332 | 0.716962845 | 1 | 0.589353324 | 0.589353324 | 0.589353324 | 0.955126654 | -0.019939035 | FALSE | TRUE | FALSE |
| PCBP3 | 0.5628 | 0.70332603 | 1 | 0.562800026 | 0.562800026 | 0.562800026 | 0.60241958 | 0.711061969 | 1 | 0.602419579 | 0.602419579 | 0.602419579 | 0.934232628 | -0.029544969 | FALSE | TRUE | FALSE |
| SLC25A3 | 0.562788 | 0.70337837 | 0.9926 | 517.8098735 | 55321.28222 | 221167.4377 | 0.10637846 | 0.874263138 | 0.00915 | 0.106378456 | 0.106378456 | 0.106378456 | 5.290432266 | 0.723491158 | FALSE | TRUE | FALSE |
| C4orf36 | 0.5627556 | 0.70343068 | 1 | 0.56275559 | 0.56275559 | 0.56275559 | 0.60187492 | 0.711435041 | 1 | 0.601874917 | 0.601874917 | 0.601874917 | 0.935004224 | -0.029186427 | FALSE | TRUE | FALSE |
| SNAP23 | 0.562662 | 0.70348298 | 1 | 0.562661958 | 0.562661958 | 0.562661958 | 0.60223102 | 0.711221985 | 1 | 0.602231021 | 0.602231021 | 0.602231021 | 0.934295873 | -0.02951557 | FALSE | TRUE | FALSE |
| SAP30L | 0.5625069 | 0.70353525 | 1 | 0.562506933 | 0.562506933 | 0.562506933 | 0.60195396 | 0.711381809 | 1 | 0.601953956 | 0.601953956 | 0.601953956 | 0.934468371 | -0.029435394 | FALSE | TRUE | FALSE |
| PSMD12 | 0.562318 | 0.70358751 | 1 | 0.562318031 | 0.562318031 | 11.14313954 | 19.9210916 | 0.132923247 | 0.00415 | 0.588150516 | 0.588150516 | 11.66791364 | 0.02822727 | -1.549331123 | FALSE | FALSE | FALSE |
| SYS1 | 0.5622961 | 0.70363975 | 1 | 0.562296136 | 0.562296136 | 0.562296136 | 0.60170503 | 0.711541445 | 1 | 0.601705027 | 0.601705027 | 0.601705027 | 0.934504635 | -0.02941854 | FALSE | TRUE | FALSE |
| VMAC | 0.5622697 | 0.70369197 | 1 | 0.562269716 | 0.562269716 | 0.562269716 | 0.6014176 | 0.711700889 | 0.00195 | 0.601417595 | 0.601417595 | 0.601417595 | 0.934907326 | -0.029231437 | FALSE | TRUE | FALSE |
| RPA2 | 0.5622434 | 0.70374417 | 1 | 0.562243363 | 0.562243363 | 0.562243363 | 0.6016698 | 0.711594614 | 1 | 0.601669802 | 0.601669802 | 0.601669802 | 0.934471634 | -0.029433877 | FALSE | TRUE | FALSE |
| NPTX2 | 0.5620738 | 0.70379635 | 1 | 0.562073839 | 0.562073839 | 11.14579245 | 0.60172223 | 0.711488254 | 1 | 0.601722229 | 0.601722229 | 11.94422534 | 0.934108483 | -0.029602684 | FALSE | TRUE | FALSE |
| FBXL3 | 0.561921 | 0.70384851 | 1 | 0.561921021 | 0.561921021 | 11.14896006 | 0.54731527 | 0.734628886 | 0.8728 | 49.69611773 | 492.0470927 | 492.0470927 | 1.026686182 | 0.011437717 | FALSE | TRUE | FALSE |
| SYT1 | 0.5615408 | 0.70390065 | 1 | 0.561540806 | 0.561540806 | 11.1414888 | 0.60125431 | 0.711753997 | 1 | 0.601254315 | 0.601254315 | 0.601254315 | 0.9339489 | -0.029676885 | FALSE | TRUE | FALSE |
| TSG101 | 0.5613258 | 0.70395278 | 1 | 2.245246774 | 2.245246774 | 2.245246774 | 0.60780109 | 0.708693337 | 1 | 0.607801093 | 0.607801093 | 0.607801093 | 0.923535352 | -0.034546476 | FALSE | TRUE | FALSE |
| SNRNP70 | 0.5612913 | 0.70400488 | 1 | 0.561291271 | 0.561291271 | 11.12663481 | 0.60114246 | 0.711860148 | 1 | 0.601142459 | 0.601142459 | 11.92928012 | 0.93370758 | -0.029789116 | FALSE | TRUE | FALSE |
| RAMP3 | 0.5611758 | 0.70405697 | 1 | 0.561175814 | 0.561175814 | 0.561175814 | 0.60053013 | 0.712231022 | 1 | 0.600530127 | 0.600530127 | 0.600530127 | 0.934467378 | -0.029435855 | FALSE | TRUE | FALSE |
| CDC34 | 0.5611457 | 0.70410903 | 1 | 0.561145654 | 0.561145654 | 0.561145654 | 0.60065193 | 0.712072202 | 1 | 0.600651929 | 0.600651929 | 0.600651929 | 0.934227673 | -0.029547272 | FALSE | TRUE | FALSE |
| CCDC124 | 0.5610555 | 0.70416108 | 1 | 0.561055505 | 0.561055505 | 0.561055505 | 0.60054111 | 0.712178103 | 1 | 0.600541109 | 0.600541109 | 0.600541109 | 0.934249955 | -0.029536914 | FALSE | TRUE | FALSE |
| RGS1 | 0.5610119 | 0.70421311 | 1 | 50.64163523 | 499.9424224 | 499.9424224 | 0.63935952 | 0.694475977 | 0.5047 | 6.015975892 | 6.015975892 | 6.015975892 | 0.877459274 | -0.056773032 | FALSE | TRUE | FALSE |
| STC2 | 0.5609959 | 0.70426511 | 1 | 0.560995915 | 0.560995915 | 11.12941135 | 0.60064025 | 0.712125163 | 0.00905 | 0.600640247 | 0.600640247 | 0.600640247 | 0.933996543 | -0.029654731 | FALSE | TRUE | FALSE |
| PPP2R5E | 0.5607636 | 0.7043171 | 1 | 0.560763633 | 0.560763633 | 11.11772428 | 0.60071443 | 0.71201922 | 1 | 0.60071443 | 0.60071443 | 11.92224922 | 0.933494528 | -0.029888224 | FALSE | TRUE | FALSE |
| ATP6V1E1 | 0.5607498 | 0.70436907 | 1 | 2.242875282 | 2.242875282 | 2.242875282 | 0.60717376 | 0.709072893 | 1 | 0.607173757 | 0.607173757 | 0.607173757 | 0.923540946 | -0.034543845 | FALSE | TRUE | FALSE |
| RPL11 | 0.560569 | 0.70442102 | 1 | 0.560568973 | 0.560568973 | 0.560568973 | 0.60044435 | 0.712283919 | 1 | 0.600444349 | 0.600444349 | 0.600444349 | 0.933590222 | -0.029843706 | FALSE | TRUE | FALSE |
| SEC14L4 | 0.560417 | 0.70447295 | 0.2613 | 0.140130149 | 0.560417027 | 1.187550862 | 0.04985807 | 0.886692421 | 0.75515 | 0.45189084 | 4.203685144 | 4.203685144 | 11.2402472 | 1.050775862 | FALSE | TRUE | FALSE |
| PF4V1 | 0.5602515 | 0.70452487 | 1 | 0.560251473 | 0.560251473 | 0.560251473 | 0.59957619 | 0.712600896 | 1 | 0.599576187 | 0.599576187 | 0.599576187 | 0.934412482 | -0.029461369 | FALSE | TRUE | FALSE |
| BCL2L15 | 0.5602397 | 0.70457676 | 1 | 0.560239664 | 0.560239664 | 0.560239664 | 0.59957968 | 0.71254812 | 1 | 0.599579683 | 0.599579683 | 0.599579683 | 0.934387338 | -0.029473055 | FALSE | TRUE | FALSE |
| CTDSPL | 0.5602184 | 0.70462863 | 1 | 0.560218351 | 0.560218351 | 0.560218351 | 0.59982221 | 0.712336801 | 1 | 0.599822213 | 0.599822213 | 0.599822213 | 0.933974 | -0.029665213 | FALSE | TRUE | FALSE |
| DPYSL4 | 0.5601679 | 0.70468049 | 0.1346 | 0.06168689 | 0.560167905 | 1.212865741 | 0.11725054 | 0.871889629 | 0.6976 | 0.267759306 | 4.278496538 | 4.278496538 | 4.777529404 | 0.679203369 | FALSE | TRUE | FALSE |
| EEF1G | 0.56015 | 0.70473232 | 0.01305 | 0.560149997 | 0.560149997 | 11.0968192 | 0.58135211 | 0.720143635 | 1 | 5.486176108 | 5.486176108 | 11.52997185 | 0.963529648 | -0.016134917 | FALSE | TRUE | FALSE |
| CTDSP2 | 0.5600978 | 0.70478413 | 1 | 0.560097785 | 0.560097785 | 0.560097785 | 0.59971286 | 0.712389663 | 1 | 0.599712858 | 0.599712858 | 0.599712858 | 0.933943264 | -0.029679506 | FALSE | TRUE | FALSE |
| CAMKV | 0.5598794 | 0.70483593 | 1 | 2.239440333 | 8.959691433 | 44.28226347 | 0.61348685 | 0.706061964 | 1 | 0.613486854 | 0.613486854 | 12.14747717 | 0.912618419 | -0.03971077 | FALSE | TRUE | FALSE |
| ANKMY2 | 0.559831 | 0.7048877 | 0.5033 | 0.559831012 | 0.559831012 | 2.7783178 | 53.7167314 | 0.050835567 | 0.0042 | 1.565219297 | 25.03881141 | 25.03881141 | 0.010421911 | -1.982052626 | FALSE | FALSE | FALSE |
| TTYH3 | 0.5598102 | 0.70493946 | 1 | 0.559810163 | 0.559810163 | 11.06906822 | 0.59961882 | 0.712442503 | 1 | 0.599618821 | 0.599618821 | 11.87239776 | 0.93361006 | -0.029834477 | FALSE | TRUE | FALSE |
| MRPL30 | 0.5597204 | 0.7049912 | 1 | 0.559720391 | 0.559720391 | 0.559720391 | 0.59920303 | 0.71281179 | 1 | 0.599203027 | 0.599203027 | 0.599203027 | 0.934108083 | -0.02960287 | FALSE | TRUE | FALSE |
| MSL2 | 0.5596343 | 0.70504292 | 1 | 0.559634322 | 0.559634322 | 11.06555632 | 0.5995831 | 0.712495323 | 1 | 0.5995831 | 0.5995831 | 11.87165741 | 0.933372407 | -0.029945042 | FALSE | TRUE | FALSE |
| FXR1 | 0.5594397 | 0.70509461 | 1 | 0.559439708 | 0.559439708 | 11.06420169 | 5.48529611 | 0.287356266 | 0.50735 | 5.485296105 | 5.485296105 | 11.49907384 | 0.101988971 | -0.991446788 | FALSE | FALSE | FALSE |
| C1GALT1 | 0.5591064 | 0.7051463 | 1 | 0.55910636 | 0.55910636 | 0.55910636 | 0.59893582 | 0.712864465 | 1 | 0.598935821 | 0.598935821 | 0.598935821 | 0.933499618 | -0.029885855 | FALSE | TRUE | FALSE |
| TNNT2 | 0.5588433 | 0.70519796 | 0.75075 | 0.558843264 | 5.336161583 | 5.336161583 | 0.08636422 | 0.87892072 | 1 | 0.806147938 | 0.806147938 | 0.806147938 | 6.47077287 | 0.810956156 | FALSE | TRUE | FALSE |
| PARP11 | 0.5588148 | 0.70524961 | 0.50415 | 0.558814752 | 0.558814752 | 0.558814752 | 0.07450411 | 0.881509031 | 1 | 0.677288475 | 6.319237113 | 6.319237113 | 7.500455648 | 0.875087647 | FALSE | TRUE | FALSE |
| DHTKD1 | 0.5586874 | 0.70530123 | 0.84385 | 16.63001326 | 2288.835172 | 20174.23342 | 0.21015067 | 0.849675636 | 0.23445 | 0.09881947 | 3.360487029 | 13.44313731 | 2.658508485 | 0.424638051 | FALSE | TRUE | FALSE |
| KIAA1755 | 0.5585957 | 0.70535284 | 0.32475 | 0.066599205 | 4.802988706 | 4.802988706 | 0.00235141 | 0.898922174 | 0.51615 | 0.019135797 | 0.019135797 | 0.374793064 | 237.5578407 | 2.375769369 | FALSE | TRUE | FALSE |
| IRF2BP2 | 0.5585499 | 0.70540443 | 1 | 0.558549902 | 0.558549902 | 11.0560384 | 4.08606509 | 0.351278229 | 0.99915 | 40357.10652 | 51472235.51 | 583068689.2 | 0.136696281 | -0.864243301 | FALSE | FALSE | FALSE |
| BLOC1S4 | 0.5584072 | 0.705456 | 1 | 0.558407207 | 0.558407207 | 0.558407207 | 0.59793107 | 0.713022377 | 1 | 0.597931065 | 0.597931065 | 0.597931065 | 0.933898972 | -0.029700103 | FALSE | TRUE | FALSE |
| CDK2 | 0.558356 | 0.70550755 | 1 | 5.271704966 | 5.271704966 | 5.271704966 | 5.65246928 | 0.279743312 | 0.0079 | 5.652469276 | 5.652469276 | 5.652469276 | 0.098780904 | -1.005327002 | FALSE | FALSE | FALSE |
| KRTAP13-2 | 0.5582035 | 0.70555908 | 0.5008 | 5.230787316 | 5.230787316 | 5.230787316 | 0.59767601 | 0.71328516 | 1 | 0.597676012 | 5.609488813 | 5.609488813 | 0.933956705 | -0.029673256 | FALSE | TRUE | FALSE |
| ARL14EP | 0.5579501 | 0.70561059 | 1 | 0.557950146 | 0.557950146 | 0.557950146 | 0.59772309 | 0.713232646 | 1 | 0.597723095 | 0.597723095 | 0.597723095 | 0.93345924 | -0.029904641 | FALSE | TRUE | FALSE |
| OR2B2 | 0.5578582 | 0.70566209 | 0.81715 | 51.10875016 | 507.8349877 | 5169.264599 | 0.67010976 | 0.680233223 | 0.9386 | 6.361466755 | 618.1946051 | 618.1946051 | 0.832487804 | -0.07962212 | FALSE | TRUE | FALSE |
| ESM1 | 0.5577631 | 0.70571356 | 1 | 0.557763142 | 0.557763142 | 0.557763142 | 0.5974367 | 0.713442579 | 1 | 0.5974367 | 0.5974367 | 0.5974367 | 0.933593705 | -0.029842086 | FALSE | TRUE | FALSE |
| RPL10A | 0.5577344 | 0.70576502 | 1 | 0.557734364 | 0.557734364 | 0.557734364 | 0.57890586 | 0.721236762 | 1 | 5.44210074 | 5.44210074 | 5.44210074 | 0.963428432 | -0.016180541 | FALSE | TRUE | FALSE |
| DYRK2 | 0.5575874 | 0.70581645 | 1 | 0.557587416 | 0.557587416 | 10.99226364 | 0.59773256 | 0.713180111 | 1 | 0.597732561 | 0.597732561 | 11.80396584 | 0.932837614 | -0.030193951 | FALSE | TRUE | FALSE |
| ATCAY | 0.5572674 | 0.70586788 | 1 | 0.557267431 | 0.557267431 | 11.04274936 | 0.59720841 | 0.713495012 | 1 | 0.597208405 | 0.597208405 | 0.597208405 | 0.933120543 | -0.030062249 | FALSE | TRUE | FALSE |
| CDC37L1 | 0.5572164 | 0.70591928 | 1 | 0.557216415 | 0.557216415 | 0.557216415 | 0.59689521 | 0.713599817 | 1 | 0.596895211 | 0.596895211 | 0.596895211 | 0.933524686 | -0.029874193 | FALSE | TRUE | FALSE |
| PDX1 | 0.5570299 | 0.70597066 | 0.8773 | 2.227671387 | 8.911120043 | 8.911120043 | 0.71915415 | 0.65638671 | 0.50385 | 0.719154148 | 0.719154148 | 0.719154148 | 0.774562564 | -0.110943497 | FALSE | TRUE | FALSE |
| SMIM21 | 0.556792 | 0.70602202 | 1 | 0.556792008 | 0.556792008 | 0.556792008 | 0.59625071 | 0.713809193 | 1 | 0.596250707 | 0.596250707 | 0.596250707 | 0.933821967 | -0.029735914 | FALSE | TRUE | FALSE |
| MRPL42 | 0.5567652 | 0.70607337 | 1 | 0.556765172 | 0.556765172 | 0.556765172 | 0.59660416 | 0.713704546 | 1 | 0.596604155 | 0.596604155 | 0.596604155 | 0.933223758 | -0.030014213 | FALSE | TRUE | FALSE |
| STK17B | 0.5567551 | 0.7061247 | 1 | 0.556755138 | 0.556755138 | 0.556755138 | 0.59662161 | 0.713652192 | 1 | 0.596621607 | 0.596621607 | 0.596621607 | 0.933179643 | -0.030034744 | FALSE | TRUE | FALSE |
| IL11 | 0.556637 | 0.706176 | 1 | 0.55663702 | 0.55663702 | 0.55663702 | 0.59616967 | 0.713913759 | 1 | 0.596169667 | 0.596169667 | 0.596169667 | 0.933688933 | -0.029797789 | FALSE | TRUE | FALSE |
| ANAPC15 | 0.5564643 | 0.70622729 | 1 | 0.556464301 | 0.556464301 | 0.556464301 | 0.59605959 | 0.714018241 | 1 | 0.596059592 | 0.596059592 | 0.596059592 | 0.933571589 | -0.029852374 | FALSE | TRUE | FALSE |
| MAB21L2 | 0.5562756 | 0.70627856 | 1 | 0.556275602 | 0.556275602 | 10.99591269 | 0.59615036 | 0.71396601 | 1 | 0.59615036 | 0.59615036 | 11.80063347 | 0.933112918 | -0.030065798 | FALSE | TRUE | FALSE |
| STX16 | 0.556184 | 0.70632982 | 1 | 0.556183999 | 0.556183999 | 11.03006661 | 0.59618275 | 0.713861487 | 1 | 0.596182747 | 0.596182747 | 0.596182747 | 0.932908577 | -0.030160914 | FALSE | TRUE | FALSE |

| | | | | | | | | | | | | | | | | | |
|---|---|---|---|---|---|---|---|---|---|---|---|---|---|---|---|---|---|
| DMRTC2 | 0.5560425 | 0.70638105 | 1 | 0.556042491 | 0.556042491 | 0.556042491 | 0.59599921 | 0.714070451 | 1 | 0.595999211 | 0.595999211 | 0.595999211 | 0.932958435 | -0.030137704 | FALSE | TRUE | FALSE |
| IFNG | 0.5559031 | 0.70643226 | 0.0057 | 0.555903064 | 0.555903064 | 0.555903064 | 0.59541966 | 0.714226965 | 1 | 0.595419657 | 0.595419657 | 0.595419657 | 0.933632369 | -0.0298241 | FALSE | TRUE | FALSE |
| TCIRG1 | 0.5558235 | 0.70648346 | 0.64955 | 2.726113166 | 2311.590359 | 37247.79957 | 0.31439995 | 0.820582397 | 0.4553 | 0.314399947 | 9.604065282 | 81.02625825 | 1.767886781 | 0.247454449 | FALSE | TRUE | FALSE |
| RNASE8 | 0.5555554 | 0.70653464 | 1 | 0.555555429 | 5.211689399 | 5.211689399 | 0.6142311 | 0.705840417 | 1 | 0.614231104 | 0.614231104 | 0.614231104 | 0.904472967 | -0.043604409 | FALSE | TRUE | FALSE |
| NPY4R | 0.5553218 | 0.7065858 | 1 | 52738.64385 | 6116913.995 | 68154170.35 | 0.65427126 | 0.687652263 | 1 | 595.4629317 | 63342.03735 | 676964.5617 | 0.848763795 | -0.071213154 | FALSE | TRUE | FALSE |
| AAMP | 0.5552897 | 0.70663694 | 1 | 0.555289655 | 0.555289655 | 11.00449135 | 2.35491545 | 0.459686266 | 0.50705 | 2.354915452 | 2.354915452 | 11.68036088 | 0.235800251 | -0.627455737 | FALSE | FALSE | FALSE |
| LAMP1 | 0.5551484 | 0.70668807 | 1 | 0.555148399 | 0.555148399 | 10.98987996 | 0.59530924 | 0.714279097 | 1 | 0.595309243 | 0.595309243 | 11.79935486 | 0.932537846 | -0.030333534 | FALSE | TRUE | FALSE |
| ARGLU1 | 0.5547923 | 0.70673917 | 1 | 5.235665285 | 50.71042779 | 51.24015225 | 0.63374299 | 0.69797578 | 1 | 0.633742991 | 0.633742991 | 12.56498811 | 0.875421637 | -0.057782724 | FALSE | TRUE | FALSE |
| PRRG4 | 0.5546454 | 0.70679026 | 0.0053 | 0.554645442 | 0.554645442 | 0.554645442 | 0.17719345 | 0.857775725 | 1 | 0.17719345 | 0.17719345 | 0.17719345 | 3.130168985 | 0.495567784 | FALSE | TRUE | FALSE |
| ARL5B | 0.5544274 | 0.70684134 | 1 | 0.554427425 | 0.554427425 | 0.554427425 | 0.59441092 | 0.71443538 | 1 | 0.594410921 | 0.594410921 | 0.594410921 | 0.932734251 | -0.030242075 | FALSE | TRUE | FALSE |
| CELF5 | 0.554253 | 0.70689239 | 1 | 0.554252992 | 0.554252992 | 10.95815962 | 0.59328553 | 0.714695485 | 1 | 0.593285534 | 0.593285534 | 11.74602914 | 0.934209517 | -0.029555713 | FALSE | TRUE | FALSE |
| KCNJ12 | 0.5540233 | 0.70694342 | 1 | 0.554023336 | 0.554023336 | 10.92163581 | 0.59056297 | 0.716194244 | 1 | 0.590562967 | 0.590562967 | 11.66203725 | 0.93812746 | -0.027738151 | FALSE | TRUE | FALSE |
| NACC2 | 0.5540028 | 0.70699444 | 1 | 0.554002754 | 0.554002754 | 10.93276424 | 0.57573045 | 0.722222831 | 0.50415 | 5.452673276 | 5.452673276 | 11.37971621 | 0.962260651 | -0.016707273 | FALSE | TRUE | FALSE |
| PSCA | 0.5539116 | 0.70704544 | 0.00285 | 0.553911626 | 0.553911626 | 0.553911626 | 0.59363018 | 0.714539479 | 1 | 0.593630176 | 0.593630176 | 0.593630176 | 0.933092098 | -0.030075488 | FALSE | TRUE | FALSE |
| CYB5RL | 0.5537748 | 0.70709642 | 0.93765 | 5.247273263 | 508.026569 | 508.026569 | 120.050461 | 0.024251487 | 0.20705 | 12.60571509 | 480.2084163 | 1921.339928 | 0.00461285 | -2.33603068 | FALSE | FALSE | FALSE |
| GIMAP1 | 0.5534414 | 0.70714738 | 1 | 0.553441398 | 0.553441398 | 10.96829328 | 0.5884162 | 0.717523658 | 1 | 0.588416202 | 0.588416202 | 0.588416202 | 0.940561114 | -0.02661298 | FALSE | TRUE | FALSE |
| FBXO44 | 0.5533607 | 0.70719833 | 0.4971 | 0.138350253 | 0.553360719 | 2.746638492 | 0.17593991 | 0.858089263 | 0.0088 | 0.175939911 | 0.175939911 | 0.175939911 | 3.145168806 | 0.49764396 | FALSE | TRUE | FALSE |
| BTLA | 0.5533308 | 0.70724925 | 1 | 0.553330751 | 0.553330751 | 0.553330751 | 0.59306565 | 0.714747448 | 0.0056 | 0.593065647 | 0.593065647 | 0.593065647 | 0.933000846 | -0.030117962 | FALSE | TRUE | FALSE |
| IL19 | 0.5529848 | 0.70730016 | 1 | 0.552984797 | 0.552984797 | 0.552984797 | 0.59261999 | 0.714955101 | 1 | 0.592619986 | 0.592619986 | 0.592619986 | 0.933118711 | -0.030063102 | FALSE | TRUE | FALSE |
| PRR15L | 0.5529749 | 0.70735105 | 1 | 0.552974893 | 0.552974893 | 0.552974893 | 0.55684541 | 0.730040685 | 1 | 5.255474622 | 50.90645304 | 50.90645304 | 0.993049208 | -0.003029231 | FALSE | TRUE | FALSE |
| B4GALT5 | 0.5526226 | 0.70740192 | 1 | 0.552622608 | 0.552622608 | 10.96308114 | 0.59286577 | 0.714799392 | 1 | 0.592865768 | 0.592865768 | 0.592865768 | 0.932120958 | -0.030527727 | FALSE | TRUE | FALSE |
| SGPP1 | 0.5525059 | 0.70745277 | 0.50385 | 2.209951707 | 2.209951707 | 10.96076916 | 0.59939642 | 0.712759097 | 1 | 0.599396423 | 0.599396423 | 11.9025591 | 0.921770474 | -0.035377207 | FALSE | TRUE | FALSE |
| NPPA | 0.5521977 | 0.70750361 | 0.0041 | 0.552197713 | 0.552197713 | 0.552197713 | 0.59221269 | 0.715214212 | 1 | 0.59221269 | 0.59221269 | 0.59221269 | 0.932431409 | -0.030383106 | FALSE | TRUE | FALSE |
| MAPK14 | 0.551975 | 0.70755443 | 1 | 0.551975025 | 0.551975025 | 0.551975025 | 0.59213875 | 0.715265973 | 1 | 0.592138746 | 0.592138746 | 0.592138746 | 0.932171772 | -0.030504052 | FALSE | TRUE | FALSE |
| LIN54 | 0.5519188 | 0.70760523 | 1 | 0.551918826 | 0.551918826 | 10.89559276 | 0.59240204 | 0.715110629 | 1 | 0.592402041 | 0.592402041 | 11.71298697 | 0.931662601 | -0.030741338 | FALSE | TRUE | FALSE |
| SPTSSB | 0.5518329 | 0.70765602 | 1 | 0.551832867 | 0.551832867 | 0.551832867 | 0.59167282 | 0.715524483 | 1 | 0.591672823 | 0.591672823 | 0.591672823 | 0.932665563 | -0.030274058 | FALSE | TRUE | FALSE |
| ZC3H13 | 0.5517345 | 0.70770678 | 0.52735 | 0.551734548 | 1.161407922 | 18.35206023 | 0.74621589 | 0.643233075 | 0.76265 | 0.746215885 | 7.049568903 | 136.7359856 | 0.739376578 | -0.131134311 | FALSE | TRUE | FALSE |
| ANO5 | 0.5516103 | 0.70775753 | 0.80255 | 53.22977759 | 127472.4766 | 13437411.21 | 0.4626642 | 0.770001317 | 0.9852 | 864.7488245 | 470459.043 | 5498327.755 | 1.192247641 | 0.076366472 | FALSE | TRUE | FALSE |
| GLTP | 0.5514642 | 0.70780825 | 1 | 0.551464196 | 0.551464196 | 0.551464196 | 0.591686 | 0.715472822 | 1 | 0.591686 | 0.591686 | 0.591686 | 0.932021707 | -0.030573973 | FALSE | TRUE | FALSE |
| RNF2 | 0.5513646 | 0.70785896 | 1 | 0.551364555 | 0.551364555 | 0.551364555 | 0.5917733 | 0.71542114 | 1 | 0.591773298 | 0.591773298 | 0.591773298 | 0.931715841 | -0.030716521 | FALSE | TRUE | FALSE |
| IFNA10 | 0.5512612 | 0.70790965 | 1 | 5.165376333 | 5.165376333 | 5.165376333 | 0.60975061 | 0.707657734 | 1 | 0.60975061 | 0.60975061 | 0.60975061 | 0.904076442 | -0.043794847 | FALSE | TRUE | FALSE |
| PSMA4 | 0.5512305 | 0.70796032 | 1 | 0.551230487 | 0.551230487 | 0.551230487 | 0.59120462 | 0.715782484 | 1 | 0.591204622 | 0.591204622 | 0.591204622 | 0.932385279 | -0.030404592 | FALSE | TRUE | FALSE |
| S100A13 | 0.5511843 | 0.70801098 | 1 | 0.551184338 | 0.551184338 | 0.551184338 | 0.59088965 | 0.716039984 | 1 | 0.590889648 | 0.590889648 | 0.590889648 | 0.932804188 | -0.030209513 | FALSE | TRUE | FALSE |
| TUBB | 0.5510849 | 0.70806161 | 1 | 0.551084871 | 0.551084871 | 10.89845603 | 0.59156103 | 0.715627743 | 1 | 0.591561031 | 0.591561031 | 11.71466511 | 0.931577372 | -0.030781069 | FALSE | TRUE | FALSE |
| TFF1 | 0.5510688 | 0.70811222 | 1 | 0.551068811 | 0.551068811 | 0.551068811 | 0.59084461 | 0.716091423 | 1 | 0.590844614 | 0.590844614 | 0.590844614 | 0.932679757 | -0.030267449 | FALSE | TRUE | FALSE |
| NDUFB7 | 0.5510557 | 0.70816282 | 1 | 0.551055663 | 5.168867817 | 5.168867817 | 0.59097153 | 0.715988524 | 1 | 0.590971533 | 5.551379378 | 5.551379378 | 0.932457204 | -0.030371092 | FALSE | TRUE | FALSE |
| RPRD1B | 0.550954 | 0.7082134 | 0.0095 | 0.550953983 | 0.550953983 | 0.550953983 | 0.59114831 | 0.715885544 | 1 | 0.591148311 | 0.591148311 | 0.591148311 | 0.932006356 | -0.030581126 | FALSE | TRUE | FALSE |
| TMEM108 | 0.5508855 | 0.70826395 | 1 | 0.550885522 | 0.550885522 | 10.88003447 | 0.59135037 | 0.715730924 | 1 | 0.591350366 | 0.591350366 | 11.69677898 | 0.931572134 | -0.030783511 | FALSE | TRUE | FALSE |
| EFHB | 0.5507033 | 0.7083145 | 0.8142 | 4.87651574 | 44.26612296 | 411.6657869 | 73.396338 | 0.03916481 | 0.641 | 73.39633795 | 6642.072114 | 65428.69649 | 0.007503145 | -2.124756681 | FALSE | FALSE | FALSE |
| DEFA5 | 0.5505453 | 0.70836502 | 1 | 0.550545279 | 0.550545279 | 0.550545279 | 0.59046828 | 0.716245624 | 1 | 0.590468279 | 0.590468279 | 0.590468279 | 0.932387562 | -0.030403529 | FALSE | TRUE | FALSE |
| TMEM200A | 0.55035 | 0.70841552 | 1 | 0.550349965 | 0.550349965 | 10.89619269 | 0.5906173 | 0.716142843 | 1 | 0.590617299 | 0.590617299 | 11.70759217 | 0.931821615 | -0.03066722 | FALSE | TRUE | FALSE |
| CD300E | 0.5502815 | 0.70846601 | 1 | 5.16539902 | 5.16539902 | 5.16539902 | 0.60915049 | 0.708040179 | 1 | 0.609150486 | 0.609150486 | 0.609150486 | 0.90335893 | -0.044139658 | FALSE | TRUE | FALSE |
| CDR2L | 0.5501587 | 0.70851648 | 1 | 0.550158718 | 0.550158718 | 0.550158718 | 0.59034196 | 0.716399643 | 1 | 0.590341959 | 0.590341959 | 0.590341959 | 0.931932263 | -0.030615653 | FALSE | TRUE | FALSE |
| SP5 | 0.5501101 | 0.70856693 | 1 | 0.550110068 | 0.550110068 | 10.89426126 | 0.59044749 | 0.716296984 | 0.011 | 0.590447494 | 0.590447494 | 11.706894 | 0.931683297 | -0.030731691 | FALSE | TRUE | FALSE |
| TOX | 0.5500697 | 0.70861736 | 1 | 0.550069673 | 0.550069673 | 10.8876026 | 0.57201855 | 0.723688396 | 1 | 5.419651091 | 5.419651091 | 11.33612694 | 0.961629085 | -0.01699241 | FALSE | TRUE | FALSE |
| MAFF | 0.5499216 | 0.70866777 | 1 | 0.549921616 | 0.549921616 | 0.549921616 | 2.3340872 | 0.461690546 | 0.5042 | 2.334087201 | 2.334087201 | 2.334087201 | 0.235604572 | -0.627816286 | FALSE | FALSE | FALSE |
| TIMM13 | 0.5495786 | 0.70871817 | 1 | 0.549578644 | 5.154796827 | 5.154796827 | 0.60838364 | 0.708312706 | 1 | 0.60838364 | 0.60838364 | 0.60838364 | 0.903342248 | -0.044147678 | FALSE | TRUE | FALSE |
| RDH10 | 0.5494024 | 0.70876854 | 0.00765 | 0.549402448 | 0.549402448 | 0.549402448 | 0.58988829 | 0.716655943 | 1 | 0.58988829 | 0.58988829 | 0.58988829 | 0.931366934 | -0.030879185 | FALSE | TRUE | FALSE |
| KCTD16 | 0.549325 | 0.7088189 | 1 | 5.203642734 | 5.203642734 | 10.88968272 | 0.58987215 | 0.716707143 | 1 | 5.589585444 | 5.589585444 | 11.7058738 | 0.931261057 | -0.030928558 | FALSE | TRUE | FALSE |
| PCGF2 | 0.5492516 | 0.70886925 | 0.0098 | 0.54925165 | 0.54925165 | 0.54925165 | 0.58957396 | 0.716860623 | 1 | 0.589573959 | 0.589573959 | 0.589573959 | 0.931607716 | -0.030766923 | FALSE | TRUE | FALSE |
| MAPT | 0.5489214 | 0.70891957 | 0.5181 | 0.548921381 | 0.548921381 | 10.7708689 | 25.9966403 | 0.105853756 | 0.4144 | 11.22878579 | 246.966315 | 988.005635 | 0.021115089 | -1.675407078 | FALSE | FALSE | FALSE |
| TMEM125 | 0.548843 | 0.70896988 | 1 | 0.548842983 | 0.548842983 | 0.548842983 | 5.35621446 | 0.291559369 | 0.00605 | 5.356214459 | 5.356214459 | 5.356214459 | 0.102468448 | -0.989409842 | FALSE | FALSE | FALSE |
| PPP2R2A | 0.5488062 | 0.70902017 | 1 | 0.548806224 | 0.548806224 | 10.89199673 | 0.58930262 | 0.717013926 | 1 | 0.58930262 | 0.58930262 | 11.70649691 | 0.931280816 | -0.030919343 | FALSE | TRUE | FALSE |
| OSR1 | 0.5487962 | 0.70907044 | 1 | 0.548796159 | 0.548796159 | 0.548796159 | 0.56783798 | 0.725521509 | 1 | 0.567837976 | 0.567837976 | 0.567837976 | 0.966466108 | -0.014813371 | FALSE | TRUE | FALSE |
| CDK5 | 0.5487199 | 0.70912069 | 1 | 0.548719893 | 0.548719893 | 0.548719893 | 0.58905949 | 0.717064989 | 1 | 0.58905949 | 0.58905949 | 0.58905949 | 0.931518636 | -0.030808452 | FALSE | TRUE | FALSE |
| ANP32D | 0.5486395 | 0.70917092 | 1 | 5.136461263 | 5.136461263 | 5.136461263 | 0.60729502 | 0.709018735 | 0.00305 | 0.607295017 | 0.607295017 | 0.607295017 | 0.903415141 | -0.044112635 | FALSE | TRUE | FALSE |
| FOSB | 0.5486265 | 0.70922113 | 1 | 0.548626452 | 0.548626452 | 10.87097574 | 0.58900047 | 0.717167055 | 1 | 0.589000474 | 0.589000474 | 11.68397494 | 0.931453328 | -0.030838901 | FALSE | TRUE | FALSE |
| SNN | 0.5485682 | 0.70927133 | 0.00365 | 0.548568199 | 0.548568199 | 0.548568199 | 0.5887108 | 0.717320002 | 1 | 0.588710803 | 0.588710803 | 0.588710803 | 0.931812694 | -0.030671378 | FALSE | TRUE | FALSE |
| ERP44 | 0.5485587 | 0.7093215 | 1 | 0.548558745 | 0.548558745 | 10.89307442 | 0.58901074 | 0.717116032 | 1 | 0.589010738 | 0.589010738 | 11.70636711 | 0.931322147 | -0.030900069 | FALSE | TRUE | FALSE |
| ETF1 | 0.5483787 | 0.70937166 | 1 | 0.548378666 | 0.548378666 | 10.87474144 | 0.5889813 | 0.717269038 | 1 | 0.588981303 | 0.588981303 | 11.69181054 | 0.931062944 | -0.031020958 | FALSE | TRUE | FALSE |
| FKBP2 | 0.5483751 | 0.7094218 | 1 | 0.548375065 | 0.548375065 | 0.548375065 | 0.58868849 | 0.717370945 | 1 | 0.588688491 | 0.588688491 | 0.588688491 | 0.931519936 | -0.030807846 | FALSE | TRUE | FALSE |
| RAB6C | 0.5482348 | 0.70947192 | 1 | 0.548234828 | 0.548234828 | 0.548234828 | 46.9198838 | 0.058626149 | 0.6911 | 46.91988381 | 4693.317694 | 4693.317694 | 0.011684488 | -1.932390306 | FALSE | FALSE | FALSE |
| FABP1 | 0.5481976 | 0.70952202 | 1 | 0.5481976 | 5.131514778 | 5.131514778 | 0.58804866 | 0.717777785 | 1 | 5.513888239 | 5.513888239 | 5.513888239 | 0.932231699 | -0.030476134 | FALSE | TRUE | FALSE |
| TRAT1 | 0.548182 | 0.7095721 | 1 | 0.548182037 | 0.548182037 | 0.548182037 | 0.58825381 | 0.717625367 | 1 | 0.588253805 | 0.588253805 | 0.588253805 | 0.931880138 | -0.030639945 | FALSE | TRUE | FALSE |
| REG3A | 0.548145 | 0.70962217 | 1 | 0.54814496 | 0.54814496 | 0.54814496 | 0.58806181 | 0.717726999 | 1 | 0.588061814 | 0.588061814 | 0.588061814 | 0.932121328 | -0.030527555 | FALSE | TRUE | FALSE |
| SNAI1 | 0.547994 | 0.70967221 | 0.00945 | 0.547994009 | 0.547994009 | 0.547994009 | 0.57988013 | 0.720840305 | 1 | 0.579880127 | 0.579880127 | 0.579880127 | 0.94501257 | -0.024562415 | FALSE | TRUE | FALSE |
| CCR7 | 0.5479484 | 0.70972224 | 1 | 2.191697 | 2.191697 | 10.85215504 | 2.35374872 | 0.459869127 | 0.50395 | 2.353748718 | 2.353748718 | 11.66815272 | 0.232798163 | -0.633020451 | FALSE | FALSE | FALSE |
| IER5 | 0.5478635 | 0.70977225 | 1 | 0.547863491 | 0.547863491 | 0.651012388 | 0.58811645 | 0.717676193 | 1 | 0.588116447 | 0.588116447 | 0.588116447 | 0.931556147 | -0.030790964 | FALSE | TRUE | FALSE |
| LST1 | 0.5478492 | 0.70982224 | 1 | 0.54784916 | 0.54784916 | 0.54784916 | 0.58771924 | 0.717879299 | 1 | 0.587719239 | 0.587719239 | 0.587719239 | 0.93216135 | -0.030508908 | FALSE | TRUE | FALSE |
| NCAPD3 | 0.5478198 | 0.70987221 | 0.6962 | 0.54781979 | 45.91574215 | 96.17184849 | 0.01382945 | 0.895497194 | 1 | 0.120419726 | 0.120419726 | 2.346158019 | 39.61254463 | 1.597832742 | FALSE | TRUE | FALSE |
| FBXO5 | 0.5477927 | 0.70992217 | 1 | 5.154399495 | 5.154399495 | 5.211663008 | 0.60689227 | 0.709127032 | 1 | 0.606892271 | 0.606892271 | 0.606892271 | 0.902619368 | -0.044495352 | FALSE | TRUE | FALSE |
| CREBRF | 0.547554 | 0.7099721 | 1 | 0.547553984 | 0.547553984 | 10.83403125 | 0.58796185 | 0.717828551 | 1 | 0.58796185 | 0.58796185 | 11.64850906 | 0.931274681 | -0.030922204 | FALSE | TRUE | FALSE |
| CLTA | 0.5472397 | 0.71002202 | 1 | 0.547239694 | 0.547239694 | 0.547239694 | 2.32495698 | 0.462777865 | 0.4984 | 0.581299218 | 2.324956985 | 2.324956985 | 0.235376266 | -0.628237331 | FALSE | FALSE | FALSE |

| | | | | | | | | | | | | | | | | | |
|---|---|---|---|---|---|---|---|---|---|---|---|---|---|---|---|---|---|
| RAB24 | 0.5472066 | 0.71007193 | 1 | 2.188687116 | 2.188687116 | 2.188687116 | 0.5942617 | 0.714487437 | 1 | 0.594261702 | 0.594261702 | 0.594261702 | 0.920817498 | -0.035826437 | FALSE | TRUE | FALSE |
| FBLIM1 | 0.5471737 | 0.71012181 | 0.5095 | 0.547173666 | 0.547173666 | 1.17973056 | 2.8391679 | 0.424165616 | 0.692 | 6.712982298 | 260.2565299 | 260.2565299 | 0.19272325 | -0.715065889 | FALSE | FALSE | FALSE |
| PRRC2C | 0.5471693 | 0.71017167 | 0.76195 | 0.547169266 | 10.4579071 | 98.65158976 | 0.78884353 | 0.624553306 | 0.52635 | 0.788843529 | 1.641518237 | 26.24067118 | 0.693634727 | -0.158869172 | FALSE | TRUE | FALSE |
| CLDN12 | 0.5470188 | 0.71022152 | 1 | 0.547018792 | 0.547018792 | 0.547018792 | 0.5692605 | 0.724897281 | 0.49895 | 0.569260503 | 5.376093795 | 5.376093795 | 0.960928765 | -0.017308806 | FALSE | TRUE | FALSE |
| KIN | 0.5467937 | 0.71027135 | 0.50725 | 0.546793745 | 0.546793745 | 1.203766129 | 0.08394313 | 0.879346991 | 0.498 | 0.083943128 | 0.761470803 | 0.761470803 | 6.513859541 | 0.81383839 | FALSE | TRUE | FALSE |
| TPST2 | 0.5466148 | 0.71032116 | 1 | 5.157782882 | 5.157782882 | 10.8037209 | 0.54509457 | 0.735783392 | 1 | 49.70595491 | 1972.727262 | 1972.727262 | 1.002788937 | 0.001209534 | FALSE | TRUE | FALSE |
| CEACAM8 | 0.5464504 | 0.71037096 | 0.75375 | 0.546450372 | 4.947687462 | 4.947687462 | 0.73342898 | 0.649620453 | 0.8762 | 6.690214111 | 62.59526181 | 62.59526181 | 0.745062419 | -0.127807342 | FALSE | TRUE | FALSE |
| DEDD2 | 0.5461253 | 0.71042074 | 1 | 0.5461253 | 0.5461253 | 10.79820541 | 0.58660539 | 0.718031447 | 1 | 0.586605386 | 0.586605386 | 11.61447382 | 0.930992645 | -0.03105375 | FALSE | TRUE | FALSE |
| MXRA7 | 0.5458061 | 0.7104705 | 1 | 0.545806085 | 0.545806085 | 0.545806085 | 0.56768507 | 0.725617281 | 1 | 0.567685067 | 5.317940202 | 5.317940202 | 0.961459295 | -0.017069098 | FALSE | TRUE | FALSE |
| HADHA | 0.5457574 | 0.71052024 | 0.7526 | 2.182794845 | 34.94336877 | 139.8631516 | 1.13153445 | 0.574730841 | 0.3794 | 1.131534446 | 18.1100357 | 89.39687157 | 0.482316236 | -0.316668119 | FALSE | FALSE | FALSE |
| CD274 | 0.5455424 | 0.71056997 | 1 | 0.54554245 | 0.54554245 | 0.54554245 | 0.58599527 | 0.718183432 | 0.00565 | 0.585995268 | 0.585995268 | 0.585995268 | 0.93096733 | -0.031065559 | FALSE | TRUE | FALSE |
| NRF1 | 0.5455208 | 0.71061968 | 1 | 0.545520785 | 0.545520785 | 10.78162911 | 0.58628583 | 0.718082128 | 0.01405 | 0.58628583 | 0.58628583 | 11.60376249 | 0.930468992 | -0.031298095 | FALSE | TRUE | FALSE |
| SS18 | 0.5455105 | 0.71066937 | 1 | 5.132333117 | 5.132333117 | 10.83433729 | 0.60467369 | 0.710097915 | 1 | 0.604673686 | 0.604673686 | 0.604673686 | 0.902156785 | -0.04471798 | FALSE | TRUE | FALSE |
| RCOR1 | 0.5454166 | 0.71071904 | 1 | 0.545416588 | 0.545416588 | 10.8222541 | 0.58616326 | 0.718132789 | 1 | 0.586163259 | 0.586163259 | 11.64179402 | 0.930485798 | -0.031290251 | FALSE | TRUE | FALSE |
| PGD | 0.5453578 | 0.7107687 | 0.99905 | 12140.33861 | 5391307.215 | 59308853.76 | 0.62115298 | 0.703210961 | 0.9999 | 580732.6671 | 763076391.4 | 8762058281 | 0.877976687 | -0.056517016 | FALSE | TRUE | FALSE |
| TRIM27 | 0.5450981 | 0.71081833 | 1 | 0.545098144 | 0.545098144 | 10.76446962 | 0.58583651 | 0.718335243 | 1 | 0.585836506 | 0.585836506 | 11.58651752 | 0.930461208 | -0.031301728 | FALSE | TRUE | FALSE |
| EDF1 | 0.5448172 | 0.71086796 | 1 | 0.544817159 | 0.544817159 | 0.544817159 | 0.58532595 | 0.718436356 | 1 | 0.585325949 | 0.585325949 | 0.585325949 | 0.930792765 | -0.031147001 | FALSE | TRUE | FALSE |
| TMEM9 | 0.5446328 | 0.71091756 | 1 | 0.544632845 | 0.544632845 | 0.544632845 | 0.58506882 | 0.718688802 | 1 | 0.585068815 | 0.585068815 | 0.585068815 | 0.930886813 | -0.031103122 | FALSE | TRUE | FALSE |
| TYROBP | 0.5446121 | 0.71096715 | 1 | 0.54461206 | 0.54461206 | 0.54461206 | 0.58515776 | 0.718537394 | 1 | 0.585157756 | 0.585157756 | 0.585157756 | 0.930709804 | -0.031185711 | FALSE | TRUE | FALSE |
| NAP1L4 | 0.544423 | 0.71101671 | 1 | 0.544422962 | 0.544422962 | 10.8102828 | 0.58495188 | 0.718739232 | 1 | 0.584951878 | 0.584951878 | 0.584951878 | 0.930714103 | -0.031183705 | FALSE | TRUE | FALSE |
| GPR139 | 0.544399 | 0.71106627 | 1 | 0.544399044 | 0.544399044 | 0.544399044 | 0.58486079 | 0.718840032 | 0.00885 | 0.584860794 | 0.584860794 | 0.584860794 | 0.930818153 | -0.031135156 | FALSE | TRUE | FALSE |
| NUFIP2 | 0.544351 | 0.7111158 | 1 | 0.544350993 | 0.544350993 | 10.75888739 | 0.58509724 | 0.718587884 | 1 | 0.585097241 | 0.585097241 | 11.58059641 | 0.93035987 | -0.031349031 | FALSE | TRUE | FALSE |
| CHRM3 | 0.5443329 | 0.71116531 | 1 | 0.544332896 | 0.544332896 | 10.74595122 | 0.58519132 | 0.718486885 | 1 | 0.585191324 | 0.585191324 | 11.5705252 | 0.930179368 | -0.031433298 | FALSE | TRUE | FALSE |
| OMP | 0.5443097 | 0.7112148 | 1 | 0.544309687 | 0.544309687 | 0.544309687 | 0.58456122 | 0.718890405 | 1 | 0.584561216 | 0.584561216 | 0.584561216 | 0.931142321 | -0.030983934 | FALSE | TRUE | FALSE |
| IGFBP4 | 0.5439534 | 0.71126428 | 1 | 5.12671548 | 5.12671548 | 5.12671548 | 0.6032958 | 0.710741362 | 1 | 0.603295803 | 0.603295803 | 0.603295803 | 0.901636358 | -0.044968584 | FALSE | TRUE | FALSE |
| RWDD4 | 0.5433637 | 0.71131375 | 0.00515 | 0.543363736 | 0.543363736 | 0.543363736 | 0.5745164 | 0.722615248 | 1 | 0.574516398 | 0.574516398 | 0.574516398 | 0.945775853 | -0.024211778 | FALSE | TRUE | FALSE |
| CCNA2 | 0.5430993 | 0.7113632 | 1 | 0.543099344 | 0.543099344 | 10.7680093 | 0.58368358 | 0.719091719 | 1 | 0.583683585 | 0.583683585 | 0.583683585 | 0.930468765 | -0.031298201 | FALSE | TRUE | FALSE |
| ZNF385D | 0.5430859 | 0.71141263 | 1 | 0.543085913 | 0.543085913 | 0.645357064 | 0.57158999 | 0.723882581 | 1 | 0.571589987 | 0.571589987 | 11.34696566 | 0.950131958 | -0.022216074 | FALSE | TRUE | FALSE |
| UBE2V1 | 0.5430545 | 0.71146204 | 1 | 0.543054462 | 0.543054462 | 0.543054462 | 0.56924991 | 0.724945406 | 1 | 0.569249914 | 0.569249914 | 0.569249914 | 0.95398251 | -0.020459587 | FALSE | TRUE | FALSE |
| VPS25 | 0.5429945 | 0.71151144 | 1 | 0.542994455 | 0.542994455 | 0.542994455 | 0.58345768 | 0.719292728 | 1 | 0.583457681 | 0.583457681 | 0.583457681 | 0.930649252 | -0.031213967 | FALSE | TRUE | FALSE |
| SMIM15 | 0.5427128 | 0.71156082 | 1 | 0.542712773 | 0.542712773 | 0.542712773 | 0.58273658 | 0.719643752 | 1 | 0.582736584 | 0.582736584 | 0.582736584 | 0.93131749 | -0.030902241 | FALSE | TRUE | FALSE |
| FDX1 | 0.5426549 | 0.71161018 | 1 | 0.542654862 | 0.542654862 | 0.542654862 | 0.57680415 | 0.721878529 | 1 | 0.576804146 | 2.307002287 | 2.307002287 | 0.940795702 | -0.026504675 | FALSE | TRUE | FALSE |
| PSME3 | 0.5425934 | 0.71165952 | 1 | 0.542593392 | 0.542593392 | 0.542593392 | 47.1083745 | 0.058368169 | 0.50595 | 47.10837454 | 466.9159967 | 466.9159967 | 0.011517982 | -1.938623619 | FALSE | FALSE | FALSE |
| MAFK | 0.5424964 | 0.71170884 | 0.0031 | 0.54249638 | 0.54249638 | 0.54249638 | 0.58328418 | 0.719393116 | 0.0032 | 0.583284179 | 0.583284179 | 0.583284179 | 0.930072167 | -0.031483352 | FALSE | TRUE | FALSE |
| OR7A17 | 0.5424785 | 0.71175815 | 0.74985 | 0.542478509 | 5.153827956 | 5.153827956 | 0.63552744 | 0.696917211 | 0.4972 | 0.068391167 | 0.635527438 | 0.635527438 | 0.853587865 | -0.068751768 | FALSE | TRUE | FALSE |
| KLF4 | 0.5423602 | 0.71180744 | 0.5126 | 5.126126477 | 5.126126477 | 18.37279671 | 0.43516024 | 0.779277714 | 1 | 40688.82116 | 52102982.85 | 591627585 | 1.246346014 | 0.095638629 | FALSE | TRUE | FALSE |
| PLEKHJ1 | 0.542357 | 0.71185671 | 1 | 5.107602349 | 20.42887358 | 20.42887358 | 2.40625757 | 0.454703103 | 0.4991 | 0.601604284 | 2.406257566 | 2.406257566 | 0.225394394 | -0.647056889 | FALSE | FALSE | FALSE |
| FAF2 | 0.5422727 | 0.71190596 | 1 | 0.5422727 | 0.5422727 | 10.73922969 | 0.58312969 | 0.719443282 | 1 | 0.583129689 | 0.583129689 | 11.56197153 | 0.929934987 | -0.031547412 | FALSE | TRUE | FALSE |
| AHCY | 0.542187 | 0.71195519 | 0.8917 | 5.000852625 | 80.00390574 | 320.1160797 | 0.21307771 | 0.848860391 | 0.87695 | 0.852014814 | 3.407727013 | 4.221096645 | 2.544550451 | 0.405611066 | FALSE | TRUE | FALSE |
| OSTM1 | 0.5420935 | 0.71200441 | 1 | 0.542093468 | 0.542093468 | 0.542093468 | 2.30430759 | 0.464401391 | 0.50765 | 2.304307587 | 2.304307587 | 2.304307587 | 0.235252217 | -0.628466275 | FALSE | FALSE | FALSE |
| FEZF2 | 0.5420448 | 0.71205361 | 1 | 0.542044753 | 0.542044753 | 10.70260698 | 0.58284401 | 0.719593663 | 1 | 0.582844012 | 0.582844012 | 11.52584764 | 0.929999694 | -0.031517194 | FALSE | TRUE | FALSE |
| HNRNPR | 0.5414723 | 0.71210279 | 1 | 0.541472324 | 0.541472324 | 10.69184791 | 0.58243228 | 0.719693825 | 1 | 0.582432275 | 0.582432275 | 11.51822138 | 0.929674311 | -0.031669169 | FALSE | TRUE | FALSE |
| SLC7A6OS | 0.5414663 | 0.71215196 | 1 | 0.54146626 | 0.54146626 | 10.73470962 | 0.58217273 | 0.71974388 | 0.0098 | 0.582172726 | 0.582172726 | 0.582172726 | 0.930078369 | -0.031480456 | FALSE | TRUE | FALSE |
| IFNGR2 | 0.5413353 | 0.71220111 | 1 | 0.541335287 | 0.541335287 | 0.541335287 | 0.58207843 | 0.719793916 | 0.0068 | 0.582078429 | 0.582078429 | 0.582078429 | 0.930004035 | -0.031515167 | FALSE | TRUE | FALSE |
| PTP4A1 | 0.5411944 | 0.71225024 | 1 | 0.541194448 | 0.541194448 | 0.541194448 | 0.58205094 | 0.719843933 | 1 | 0.58205094 | 0.58205094 | 0.58205094 | 0.929805986 | -0.031607662 | FALSE | TRUE | FALSE |
| PMEPA1 | 0.541191 | 0.71229935 | 1 | 0.541191013 | 0.541191013 | 0.541191013 | 0.58178264 | 0.719993869 | 1 | 0.581782641 | 0.581782641 | 0.581782641 | 0.930228878 | -0.031410182 | FALSE | TRUE | FALSE |
| PTPRR | 0.5411501 | 0.71234845 | 0.0169 | 0.541150098 | 0.541150098 | 4.576445672 | 0.17291153 | 0.858861145 | 1 | 0.172911531 | 0.172911531 | 3.420082084 | 3.129635689 | 0.495493786 | FALSE | TRUE | FALSE |
| HRH2 | 0.5410976 | 0.71239752 | 1 | 0.541097566 | 0.541097566 | 10.7307999 | 0.56385959 | 0.727329295 | 1 | 5.319781957 | 5.319781957 | 5.319781957 | 0.959631751 | -0.017895391 | FALSE | TRUE | FALSE |
| SPRN | 0.5410402 | 0.71244658 | 1 | 0.541040187 | 5.069851749 | 5.069851749 | 5.45346796 | 0.288861557 | 0.49955 | 0.581073908 | 5.453467961 | 5.453467961 | 0.099210299 | -1.003443241 | FALSE | FALSE | FALSE |
| RPS26 | 0.540974 | 0.71249562 | 1 | 0.540974018 | 0.540974018 | 0.540974018 | 0.58190116 | 0.719893931 | 1 | 0.581901157 | 0.581901157 | 0.581901157 | 0.92966651 | -0.031672814 | FALSE | TRUE | FALSE |
| NKAIN4 | 0.5407815 | 0.71254465 | 1 | 0.540781534 | 0.540781534 | 0.540781534 | 0.58111446 | 0.72029323 | 1 | 0.581114458 | 0.581114458 | 0.581114458 | 0.930593837 | -0.031239828 | FALSE | TRUE | FALSE |
| ARPC2 | 0.5407579 | 0.71259366 | 0.00965 | 0.540757924 | 0.540757924 | 0.540757924 | 2.30056518 | 0.464940296 | 0.5086 | 2.300565176 | 2.300565176 | 2.300565176 | 0.235054381 | -0.62883165 | FALSE | FALSE | FALSE |
| TMEM144 | 0.5406609 | 0.71264264 | 0.75395 | 1.226951676 | 4.907402293 | 4.907402293 | 0.17765196 | 0.857599547 | 0.0072 | 0.177651961 | 0.177651961 | 0.177651961 | 3.043371521 | 0.483354972 | FALSE | TRUE | FALSE |
| EIF4A2 | 0.5405645 | 0.71269162 | 1 | 0.540564507 | 0.540564507 | 10.72519219 | 0.58131578 | 0.720243383 | 1 | 0.581315781 | 0.581315781 | 11.54477304 | 0.929898214 | -0.031564586 | FALSE | TRUE | FALSE |
| TUBB3 | 0.5404561 | 0.71274057 | 1 | 2.161658545 | 2.161658545 | 10.67365839 | 0.58150306 | 0.720093731 | 1 | 2.325854982 | 2.325854982 | 11.50162797 | 0.929412236 | -0.031791614 | FALSE | TRUE | FALSE |
| TUBB6 | 0.5402856 | 0.71278951 | 0.755 | 0.540285604 | 2.160890166 | 10.65065245 | 0.6922016 | 0.669333317 | 0.51715 | 0.692201596 | 0.692201596 | 3.418327615 | 0.780532156 | -0.1076092 | FALSE | TRUE | FALSE |
| NSUN7 | 0.5401801 | 0.71283842 | 0.75155 | 0.5401801 | 4.911488416 | 10.71165828 | 0.08396192 | 0.879337753 | 1 | 0.749651886 | 6.869743828 | 6.869743828 | 6.433631928 | 0.808456211 | FALSE | TRUE | FALSE |
| RALY | 0.5401379 | 0.71288733 | 0.01 | 0.540137885 | 0.540137885 | 0.641894269 | 0.58099339 | 0.720343057 | 1 | 0.580993389 | 0.580993389 | 0.580993389 | 0.929679916 | -0.031666551 | FALSE | TRUE | FALSE |
| GPX2 | 0.5401279 | 0.71293621 | 1 | 0.540127858 | 0.540127858 | 0.540127858 | 0.55230709 | 0.732469054 | 1 | 0.552307092 | 0.552307092 | 0.552307092 | 0.977948439 | -0.009684042 | FALSE | TRUE | FALSE |
| MT1B | 0.5400512 | 0.71298507 | 1 | 0.540051175 | 0.540051175 | 0.540051175 | 0.58027928 | 0.720691326 | 1 | 0.580279279 | 0.580279279 | 0.580279279 | 0.930674582 | -0.031202147 | FALSE | TRUE | FALSE |
| SPIN1 | 0.5399327 | 0.71303392 | 1 | 489.6472936 | 4983.719727 | 51962.63912 | 0.49725712 | 0.756016636 | 0.99855 | 47137.43121 | 5463563.312 | 60853995.19 | 1.085821999 | 0.035758636 | FALSE | TRUE | FALSE |
| ZNF346 | 0.5399148 | 0.71308275 | 1 | 0.539914826 | 0.539914826 | 0.539914826 | 0.58049882 | 0.720542182 | 1 | 0.58049882 | 0.58049882 | 0.58049882 | 0.930087725 | -0.031476087 | FALSE | TRUE | FALSE |
| HLA-DMB | 0.5398919 | 0.71313156 | 1 | 0.539891907 | 0.539891907 | 0.539891907 | 0.58045596 | 0.72064163 | 1 | 0.580455956 | 0.580455956 | 0.580455956 | 0.930116922 | -0.031462454 | FALSE | TRUE | FALSE |
| RAB8A | 0.5397905 | 0.71318036 | 1 | 0.539790504 | 0.539790504 | 0.539790504 | 0.58076928 | 0.720442657 | 1 | 0.580769284 | 0.580769284 | 0.580769284 | 0.929440518 | -0.031778399 | FALSE | TRUE | FALSE |
| GGT1 | 0.5397524 | 0.71322913 | 1 | 52025766.57 | 59349094089 | 2.09538E+12 | 0.16709887 | 0.860380924 | 1 | 152096776.6 | 2.34316E+11 | 2.81973E+12 | 3.230138108 | 0.509221091 | FALSE | TRUE | FALSE |
| FGF18 | 0.5397033 | 0.71327789 | 1 | 0.53970328 | 0.53970328 | 0.53970328 | 0.58063239 | 0.720492429 | 1 | 0.580632387 | 0.580632387 | 0.580632387 | 0.929509432 | -0.031746199 | FALSE | TRUE | FALSE |
| DCTPP1 | 0.5394215 | 0.71332663 | 1 | 0.539421533 | 0.539421533 | 0.539421533 | 5.1209733 | 0.302755783 | 0.254 | 5.120973305 | 49.40946947 | 49.40946947 | 0.105335744 | -0.977424233 | FALSE | FALSE | FALSE |
| VPS29 | 0.5394067 | 0.71337536 | 1 | 0.539406701 | 0.539406701 | 0.539406701 | 0.57994003 | 0.720790664 | 1 | 0.579940034 | 0.579940034 | 0.579940034 | 0.930107717 | -0.031466752 | FALSE | TRUE | FALSE |
| LBH | 0.5393775 | 0.71342407 | 1 | 0.539377502 | 0.539377502 | 0.539377502 | 0.57972594 | 0.720939529 | 1 | 0.579725944 | 0.579725944 | 0.579725944 | 0.930400835 | -0.031329908 | FALSE | TRUE | FALSE |
| KCNMA1 | 0.5393568 | 0.71347275 | 0.26735 | 0.134887654 | 0.539356824 | 4.546592756 | 0.02571031 | 0.892216261 | 1 | 0.02571031 | 0.02571031 | 0.502531867 | 20.97823095 | 1.321768862 | FALSE | TRUE | FALSE |
| CHCHD1 | 0.5393002 | 0.71352142 | 1 | 0.539300202 | 0.539300202 | 0.539300202 | 0.57984753 | 0.720889926 | 1 | 0.579847534 | 0.579847534 | 0.579847534 | 0.930072425 | -0.031483232 | FALSE | TRUE | FALSE |
| ATP1B2 | 0.5392384 | 0.71357008 | 1 | 0.539238411 | 0.539238411 | 0.539238411 | 0.58003901 | 0.720741004 | 1 | 0.580039011 | 0.580039011 | 0.580039011 | 0.929658869 | -0.031676383 | FALSE | TRUE | FALSE |
| IVNS1ABP | 0.5391056 | 0.71361871 | 0.0157 | 0.539105644 | 0.539105644 | 4.557481474 | 14.6299266 | 0.159535447 | 0.01045 | 1.521132801 | 14.62992658 | 30.12535631 | 0.036849511 | -1.433568268 | FALSE | FALSE | FALSE |

| | | | | | | | | | | | | | | | | | |
|---|---|---|---|---|---|---|---|---|---|---|---|---|---|---|---|---|---|
| CSN3 | 0.5390875 | 0.71366733 | 1 | 0.539087509 | 0.539087509 | 0.539087509 | 0.5795351 | 0.721038678 | 1 | 0.579535103 | 0.579535103 | 0.579535103 | 0.930206826 | -0.031420478 | FALSE | TRUE | FALSE |
| MR1 | 0.5390332 | 0.71371593 | 0.50555 | 0.539033212 | 0.539033212 | 0.539033212 | 0.08576693 | 0.87901361 | 1 | 0.08576693 | 0.08576693 | 0.08576693 | 6.284860777 | 0.798295662 | FALSE | TRUE | FALSE |
| PTCRA | 0.5388408 | 0.71376451 | 1 | 5.052794675 | 48.62835444 | 48.62835444 | 4567.03676 | 0.000407085 | 0.1108 | 452.3526978 | 47234.91728 | 501753.088 | 0.000117985 | -3.928174001 | FALSE | FALSE | FALSE |
| PLA2G2A | 0.5388277 | 0.71381308 | 1 | 0.538827711 | 0.538827711 | 0.538827711 | 0.57921646 | 0.721088227 | 1 | 0.579216459 | 0.579216459 | 0.579216459 | 0.930270027 | -0.031390972 | FALSE | TRUE | FALSE |
| EIF4EBP3 | 0.5384428 | 0.71386163 | 1 | 0.538442843 | 0.538442843 | 0.538442843 | 0.54339049 | 0.736839607 | 1 | 5.080616415 | 48.7537532 | 48.7537532 | 0.990894864 | -0.003972423 | FALSE | TRUE | FALSE |
| RNF24 | 0.5383749 | 0.71391016 | 1 | 0.538374882 | 0.538374882 | 0.538374882 | 0.56416036 | 0.727140195 | 1 | 0.564160356 | 0.564160356 | 0.564160356 | 0.954294071 | -0.020317775 | FALSE | TRUE | FALSE |
| ST8SIA4 | 0.5381237 | 0.71395868 | 1 | 0.538123689 | 0.538123689 | 0.538123689 | 0.57895049 | 0.721187269 | 1 | 0.578950488 | 0.578950488 | 0.578950488 | 0.929481363 | -0.031759314 | FALSE | TRUE | FALSE |
| ACSM2B | 0.5381212 | 0.71400718 | 0.13945 | 0.538121177 | 2.153225514 | 4.547597313 | 1.6316485 | 0.522916897 | 0.78275 | 6.527352913 | 138.9927646 | 556.0357056 | 0.329802145 | -0.481746524 | FALSE | FALSE | FALSE |
| GRK4 | 0.5380458 | 0.71405566 | 0.01 | 0.237841213 | 0.237841213 | 0.541876951 | 0.35140602 | 0.808655074 | 0.2644 | 0.351406024 | 3.178967422 | 6.957119584 | 1.531122947 | 0.185010065 | FALSE | TRUE | FALSE |
| SLC25A18 | 0.5379375 | 0.71410412 | 0.7546 | 2.150982179 | 20.05989635 | 20.05989635 | 2.81711116 | 0.425602197 | 0.80165 | 45.07818451 | 1689.30511 | 4054.826949 | 0.190953601 | -0.719072148 | FALSE | FALSE | FALSE |
| GPR37L1 | 0.5375988 | 0.71415257 | 0.7593 | 0.537598808 | 2.15060493 | 10.6386476 | 0.68729366 | 0.672079214 | 0.50285 | 0.687293662 | 0.687293662 | 3.405003617 | 0.782196661 | -0.106684042 | FALSE | TRUE | FALSE |
| MT2A | 0.537543 | 0.714201 | 1 | 0.537543017 | 0.537543017 | 0.537543017 | 0.57790378 | 0.721533339 | 1 | 0.577903777 | 0.577903777 | 0.577903777 | 0.930160068 | -0.031442309 | FALSE | TRUE | FALSE |
| CBX4 | 0.5374836 | 0.71424942 | 1 | 0.537483634 | 0.537483634 | 10.57046212 | 0.57786829 | 0.721582704 | 1 | 0.577868289 | 0.577868289 | 11.38749869 | 0.930114429 | -0.031463618 | FALSE | TRUE | FALSE |
| PLK2 | 0.5374471 | 0.71429781 | 1 | 0.537447149 | 0.537447149 | 10.60462832 | 0.57865635 | 0.721335691 | 1 | 0.578656352 | 0.578656352 | 11.43619413 | 0.92878467 | -0.032084961 | FALSE | TRUE | FALSE |
| FRG1 | 0.5372722 | 0.71434619 | 1 | 5.090307425 | 49.49666086 | 49.49666086 | 5.24116328 | 0.296313352 | 0.2545 | 5.241163277 | 50.97939175 | 50.97939175 | 0.102510101 | -0.98923334 | FALSE | FALSE | FALSE |
| HECW1 | 0.5371992 | 0.71439456 | 0.66835 | 0.53719917 | 8.593998509 | 52.08358788 | 0.06252235 | 0.884178807 | 0.5304 | 0.062522353 | 0.301999044 | 1.207013304 | 8.592113702 | 0.934100016 | FALSE | TRUE | FALSE |
| SPINK7 | 0.5366067 | 0.71444291 | 1 | 0.536606744 | 0.536606744 | 0.536606744 | 0.57740425 | 0.721730696 | 1 | 0.577404249 | 0.577404249 | 0.577404249 | 0.929343255 | -0.031823849 | FALSE | TRUE | FALSE |
| TSN | 0.5365599 | 0.71449124 | 1 | 0.536559885 | 0.536559885 | 0.536559885 | 0.5774259 | 0.721681385 | 1 | 0.577425899 | 0.577425899 | 0.577425899 | 0.929227258 | -0.031878059 | FALSE | TRUE | FALSE |
| GFI1 | 0.5365001 | 0.71453955 | 1 | 0.536500132 | 0.536500132 | 10.6254861 | 0.57746567 | 0.721632054 | 1 | 0.577465672 | 0.577465672 | 11.45021628 | 0.929059783 | -0.031956339 | FALSE | TRUE | FALSE |
| SORCS1 | 0.5363684 | 0.71458785 | 0.5184 | 0.536368423 | 0.536368423 | 10.46712848 | 0.08522054 | 0.87910638 | 0.02785 | 0.085220543 | 0.085220543 | 1.667924205 | 6.293886454 | 0.798918904 | FALSE | TRUE | FALSE |
| SIAH2 | 0.5362731 | 0.71463613 | 1 | 0.536273093 | 0.536273093 | 0.536273093 | 0.55531749 | 0.731008469 | 1 | 0.555317495 | 0.555317495 | 0.555317495 | 0.965705381 | -0.015155349 | FALSE | TRUE | FALSE |
| IL2RA | 0.5362565 | 0.71468439 | 1 | 5.033435271 | 5.033435271 | 5.033435271 | 0.59543541 | 0.714174813 | 1 | 0.59543541 | 0.59543541 | 0.59543541 | 0.900612375 | -0.04546209 | FALSE | TRUE | FALSE |
| TSPO | 0.53613 | 0.71473264 | 1 | 0.536130035 | 0.536130035 | 0.536130035 | 0.57669073 | 0.721927769 | 1 | 0.576690732 | 0.576690732 | 0.576690732 | 0.929666467 | -0.031672834 | FALSE | TRUE | FALSE |
| FAM135B | 0.536099 | 0.71478087 | 0.75585 | 1.247798916 | 4.990474029 | 97.12345591 | 0.17724608 | 0.85773801 | 0.03285 | 0.17724608 | 0.17724608 | 3.460711795 | 3.02460306 | 0.480668387 | FALSE | TRUE | FALSE |
| ZNF43 | 0.5360016 | 0.71482908 | 0.82775 | 42.21055171 | 35181.05387 | 346282.1863 | 0.00250442 | 0.898855151 | 1 | 0.020826405 | 0.177764983 | 0.41204115 | 214.0221786 | 2.330458781 | FALSE | TRUE | FALSE |
| HAS2 | 0.5358464 | 0.71487727 | 1 | 0.53584643 | 0.53584643 | 10.61013134 | 0.57701483 | 0.721779992 | 1 | 0.577014827 | 0.577014827 | 11.43898981 | 0.928652791 | -0.032146632 | FALSE | TRUE | FALSE |
| C1orf198 | 0.5358384 | 0.71492545 | 1 | 0.535838445 | 0.535838445 | 0.535838445 | 5.25243643 | 0.295721758 | 0.5081 | 5.25243643 | 5.25243643 | 5.25243643 | 0.102017121 | -0.991326935 | FALSE | FALSE | FALSE |
| ADAMTSL5 | 0.5358339 | 0.7149736 | 0.01435 | 0.535833902 | 0.535833902 | 1.175840699 | 2.69409774 | 0.43507023 | 0.8125 | 24.99410419 | 580.0691031 | 2319.829642 | 0.198891782 | -0.70138316 | FALSE | FALSE | FALSE |
| WFDC10A | 0.535805 | 0.71502175 | 1 | 0.535804977 | 0.535804977 | 0.535804977 | 0.57632355 | 0.722026197 | 0.00215 | 0.576323547 | 0.576323547 | 0.576323547 | 0.929694751 | -0.031659621 | FALSE | TRUE | FALSE |
| LENEP | 0.5348786 | 0.71506988 | 1 | 0.534878576 | 0.534878576 | 0.534878576 | 0.57538383 | 0.722321042 | 1 | 0.575383832 | 0.575383832 | 0.575383832 | 0.929603068 | -0.031702452 | FALSE | TRUE | FALSE |
| ZNF681 | 0.5348648 | 0.71511799 | 1 | 48.83628939 | 484.8782061 | 484.8782061 | 5.6178355 | 0.281585259 | 0.75045 | 5.617835502 | 54.47738604 | 54.47738604 | 0.095208346 | -1.02132498 | FALSE | FALSE | FALSE |
| EFCAB14 | 0.5348598 | 0.71516608 | 0.505 | 0.534859758 | 0.534859758 | 2.651261693 | 0.17114607 | 0.859293331 | 1 | 0.171146065 | 0.171146065 | 3.396370004 | 3.125165379 | 0.494873004 | FALSE | TRUE | FALSE |
| ITGB7 | 0.5342785 | 0.71521417 | 0.74865 | 2.136366653 | 34.18356224 | 136.7888491 | 0.00521707 | 0.898078443 | 0.93895 | 0.626865094 | 23.42980952 | 93.67970672 | 102.4096973 | 2.010341082 | FALSE | TRUE | FALSE |
| ZBTB12 | 0.5340389 | 0.71526223 | 1 | 0.534038947 | 0.534038947 | 10.54873951 | 0.57525427 | 0.722370121 | 1 | 0.57525427 | 0.57525427 | 11.37975759 | 0.928352861 | -0.03228692 | FALSE | TRUE | FALSE |
| RASGEF1A | 0.5338697 | 0.71531029 | 1 | 0.533869738 | 0.533869738 | 10.56753173 | 0.57503137 | 0.722419183 | 1 | 0.575031366 | 0.575031366 | 11.39638205 | 0.928418465 | -0.032256231 | FALSE | TRUE | FALSE |
| ATP6V0D1 | 0.5337527 | 0.71535832 | 1 | 0.533752657 | 0.533752657 | 10.57170318 | 0.57490094 | 0.722517251 | 1 | 0.574900939 | 0.574900939 | 11.39996145 | 0.92842544 | -0.032252968 | FALSE | TRUE | FALSE |
| CHRM2 | 0.5337138 | 0.71540634 | 1 | 0.533713758 | 0.533713758 | 10.58751406 | 0.5749653 | 0.722468227 | 1 | 0.5749653 | 0.5749653 | 0.5749653 | 0.928253858 | -0.032333237 | FALSE | TRUE | FALSE |
| LRRC73 | 0.5336866 | 0.71545433 | 1 | 0.533686562 | 0.533686562 | 0.533686562 | 0.5746855 | 0.722566259 | 1 | 0.574685504 | 0.574685504 | 0.574685504 | 0.928658473 | -0.032143975 | FALSE | TRUE | FALSE |
| SLC44A2 | 0.5335107 | 0.71550232 | 0.7563 | 0.533510659 | 2.134456013 | 10.49100477 | 2.60951446 | 0.439801196 | 0.5057 | 10.442642 | 41.9271803 | 205.7603079 | 0.204448247 | -0.689416608 | FALSE | FALSE | FALSE |
| YY1 | 0.5334057 | 0.71555028 | 1 | 0.533405699 | 0.533405699 | 10.55529547 | 0.55722101 | 0.729901823 | 1 | 0.55722101 | 0.55722101 | 11.04059239 | 0.957260566 | -0.018969831 | FALSE | TRUE | FALSE |
| RIC8B | 0.5332206 | 0.71559823 | 1 | 0.533220606 | 0.533220606 | 10.56695623 | 0.57440757 | 0.722762104 | 0.01135 | 0.574407568 | 0.574407568 | 11.39567948 | 0.928296622 | -0.03231323 | FALSE | TRUE | FALSE |
| ARL4D | 0.533177 | 0.71564616 | 1 | 0.533176963 | 0.533176963 | 0.533176963 | 0.57444394 | 0.722664219 | 1 | 0.574443944 | 0.574443944 | 0.574443944 | 0.928161866 | -0.032376279 | FALSE | TRUE | FALSE |
| C11orf71 | 0.5331543 | 0.71569408 | 1 | 0.533154285 | 0.533154285 | 0.533154285 | 5.21018382 | 0.298081708 | 0.50685 | 5.210183818 | 5.210183818 | 5.210183818 | 0.102329266 | -0.990000142 | FALSE | FALSE | FALSE |
| CEBPG | 0.5331462 | 0.71574197 | 1 | 0.53314624 | 0.53314624 | 0.53314624 | 0.57396191 | 0.722859918 | 1 | 0.57396191 | 0.57396191 | 0.57396191 | 0.928887841 | -0.032036722 | FALSE | TRUE | FALSE |
| SULT4A1 | 0.5330326 | 0.71578985 | 1 | 2.131918411 | 2.131918411 | 2.131918411 | 0.58081343 | 0.720392867 | 1 | 0.580813432 | 0.580813432 | 0.580813432 | 0.917734655 | -0.037282868 | FALSE | TRUE | FALSE |
| ZNF581 | 0.5329046 | 0.71583772 | 1 | 0.532904619 | 0.532904619 | 0.532904619 | 0.57386903 | 0.722957658 | 1 | 0.573869035 | 0.573869035 | 0.573869035 | 0.928617135 | -0.032163307 | FALSE | TRUE | FALSE |
| UBE2W | 0.5327993 | 0.71588556 | 1 | 0.532799256 | 0.532799256 | 0.532799256 | 0.57357986 | 0.723104129 | 1 | 0.573579859 | 0.573579859 | 0.573579859 | 0.928901611 | -0.032030284 | FALSE | TRUE | FALSE |
| NIPA1 | 0.5327518 | 0.71593339 | 1 | 0.532751847 | 0.532751847 | 0.532751847 | 0.57385601 | 0.7230065 | 1 | 0.573856006 | 0.573856006 | 0.573856006 | 0.928371999 | -0.032277967 | FALSE | TRUE | FALSE |
| AK8 | 0.5325486 | 0.7159812 | 0.51275 | 0.532548606 | 0.532548606 | 1.160787592 | 0.08156369 | 0.879917223 | 0.5079 | 0.745359835 | 0.745359835 | 1.618344439 | 6.529236411 | 0.814862394 | FALSE | TRUE | FALSE |
| FZR1 | 0.5325138 | 0.716029 | 1 | 5.036211952 | 5.036211952 | 10.51369321 | 0.59250868 | 0.715006964 | 1 | 0.592508678 | 0.592508678 | 11.71625488 | 0.898744314 | -0.046363844 | FALSE | TRUE | FALSE |
| CD247 | 0.5323799 | 0.71607678 | 1 | 0.532379917 | 0.532379917 | 0.532379917 | 0.57328014 | 0.723201686 | 1 | 0.57328014 | 0.57328014 | 0.57328014 | 0.928655781 | -0.032145233 | FALSE | TRUE | FALSE |
| SREK1IP1 | 0.532218 | 0.71612454 | 1 | 0.532218027 | 0.532218027 | 0.532218027 | 0.5383184 | 0.739140755 | 1 | 5.051122533 | 48.64323351 | 48.64323351 | 0.988667718 | -0.004949646 | FALSE | TRUE | FALSE |
| SERPINE3 | 0.5320619 | 0.71617228 | 0.75075 | 0.532061897 | 4.931879013 | 4.931879013 | 0.08438185 | 0.8792638 | 1 | 0.767129127 | 0.767129127 | 0.767129127 | 6.305406933 | 0.79971312 | FALSE | TRUE | FALSE |
| PITPNC1 | 0.5320134 | 0.71622001 | 1 | 0.532013374 | 0.532013374 | 0.532013374 | 5.22214357 | 0.297198643 | 0.49995 | 0.55537517 | 5.222143571 | 5.222143571 | 0.101876436 | -0.991926258 | FALSE | FALSE | FALSE |
| OR2M7 | 0.5319168 | 0.71626773 | 1 | 0.531916801 | 0.531916801 | 0.531916801 | 5.20939566 | 0.298375454 | 0.503 | 5.209395656 | 5.209395656 | 5.209395656 | 0.102107199 | -0.990943636 | FALSE | FALSE | FALSE |
| ABR | 0.5316016 | 0.71631542 | 0.94035 | 11.43583731 | 440.8762847 | 440.8762847 | 0.1504921 | 0.864484011 | 1 | 1236.191227 | 129815.1627 | 423541.626 | 3.532422158 | 0.5480726 | FALSE | TRUE | FALSE |
| SERBP1 | 0.5315713 | 0.7163631 | 1 | 0.531571277 | 0.531571277 | 10.51192001 | 0.57295749 | 0.723299175 | 1 | 0.572957489 | 0.572957489 | 11.34566251 | 0.927767395 | -0.032560894 | FALSE | TRUE | FALSE |
| BANP | 0.5315029 | 0.71641076 | 1 | 0.531502875 | 0.531502875 | 10.4816379 | 0.57287598 | 0.723347892 | 1 | 0.572875983 | 0.572875983 | 11.31653017 | 0.927779992 | -0.032554997 | FALSE | TRUE | FALSE |
| TAF1D | 0.5309282 | 0.71645841 | 0.50575 | 0.530928232 | 0.530928232 | 0.530928232 | 0.71451836 | 0.658758309 | 0.7494 | 0.71451836 | 6.501139533 | 6.501139533 | 0.743057507 | -0.128977574 | FALSE | TRUE | FALSE |
| NANOG | 0.5308703 | 0.71650605 | 1 | 0.530870261 | 0.530870261 | 0.530870261 | 0.57232388 | 0.723591198 | 1 | 0.572323883 | 0.572323883 | 0.572323883 | 0.927569645 | -0.032653472 | FALSE | TRUE | FALSE |
| GPR12 | 0.5308606 | 0.71655366 | 1 | 0.530860631 | 0.530860631 | 10.52896689 | 0.57228166 | 0.723639805 | 1 | 0.57228166 | 0.57228166 | 0.57228166 | 0.927621255 | -0.032629309 | FALSE | TRUE | FALSE |
| PDXP | 0.5307314 | 0.71660126 | 1 | 0.530731403 | 0.530731403 | 0.530731403 | 0.5718098 | 0.723785525 | 1 | 0.571809801 | 0.571809801 | 0.571809801 | 0.928160731 | -0.03237681 | FALSE | TRUE | FALSE |
| IL6 | 0.530615 | 0.71664884 | 1 | 0.530615039 | 0.530615039 | 0.530615039 | 0.5469815 | 0.734851735 | 1 | 0.546981496 | 0.546981496 | 0.546981496 | 0.970078592 | -0.013193079 | FALSE | TRUE | FALSE |
| SDC1 | 0.5305277 | 0.71669641 | 1 | 0.530527676 | 0.530527676 | 0.530527676 | 0.57182312 | 0.72373697 | 1 | 0.571823124 | 0.571823124 | 0.571823124 | 0.92778283 | -0.032553669 | FALSE | TRUE | FALSE |
| CBLL1 | 0.5305014 | 0.71674396 | 1 | 0.530501366 | 0.530501366 | 10.50958321 | 0.5551297 | 0.731191958 | 1 | 0.555129702 | 0.555129702 | 11.01000959 | 0.955634989 | -0.019707958 | FALSE | TRUE | FALSE |
| CASP8 | 0.5304595 | 0.71679149 | 0.01575 | 0.058990127 | 0.530459465 | 1.997232545 | 0.08438779 | 0.879245299 | 1 | 0.08438779 | 0.08438779 | 1.674415755 | 6.285974116 | 0.798372588 | FALSE | TRUE | FALSE |
| ILKAP | 0.5304399 | 0.716839 | 1 | 0.53043995 | 0.53043995 | 10.51372627 | 0.57170962 | 0.723834062 | 0.0109 | 0.571709616 | 0.571709616 | 11.34393666 | 0.927813588 | -0.032539271 | FALSE | TRUE | FALSE |
| SFRP5 | 0.530388 | 0.7168865 | 0.7611 | 1.212557656 | 4.849513697 | 4.849513697 | 14.2243813 | 0.161850419 | 0.2517 | 1.510535287 | 14.22438134 | 14.22438134 | 0.037287245 | -1.428439704 | FALSE | FALSE | FALSE |
| GDF11 | 0.530355 | 0.71693398 | 1 | 5.01908165 | 5.01908165 | 10.49653181 | 0.50391006 | 0.75394282 | 1 | 456.9765548 | 4651.210957 | 48495.47059 | 1.052479475 | 0.022213635 | FALSE | TRUE | FALSE |
| ICMT | 0.5301683 | 0.71698144 | 1 | 0.530168349 | 0.530168349 | 0.530168349 | 0.57141308 | 0.723979564 | 1 | 0.571413079 | 0.571413079 | 0.571413079 | 0.927819767 | -0.032536379 | FALSE | TRUE | FALSE |
| SLC2A4RG | 0.5300547 | 0.71702889 | 1 | 0.530054745 | 0.530054745 | 0.629987057 | 0.57123501 | 0.724076475 | 1 | 0.571235014 | 0.571235014 | 0.571235014 | 0.927910111 | -0.032494093 | FALSE | TRUE | FALSE |
| RIPK2 | 0.5298487 | 0.71707632 | 1 | 0.529848664 | 0.529848664 | 10.49397689 | 0.57129239 | 0.724028029 | 1 | 0.571292394 | 0.571292394 | 11.32802345 | 0.927456183 | -0.032706599 | FALSE | TRUE | FALSE |
| CDO1 | 0.529768 | 0.71712373 | 1 | 0.529767982 | 0.529767982 | 0.529767982 | 0.57101681 | 0.724270075 | 1 | 0.571016814 | 0.571016814 | 0.571016814 | 0.927762493 | -0.032563189 | FALSE | TRUE | FALSE |

| | | | | | | | | | | | | | | | | | |
|---|---|---|---|---|---|---|---|---|---|---|---|---|---|---|---|---|---|
| CHRM4 | 0.529633 | 0.71717113 | 1 | 0.529632971 | 0.529632971 | 10.44839882 | 0.53618156 | 0.739912599 | 1 | 5.062881186 | 49.06445813 | 100.0422973 | 0.987786616 | -0.005336863 | FALSE | TRUE | FALSE |
| UFC1 | 0.5295453 | 0.71721851 | 1 | 4.9796663 | 4.9796663 | 4.9796663 | 0.58898948 | 0.717218057 | 1 | 0.588989485 | 0.588989485 | 0.588989485 | 0.899074305 | -0.046204414 | FALSE | TRUE | FALSE |
| LURAP1L | 0.5294764 | 0.71726587 | 1 | 0.529476365 | 0.529476365 | 0.529476365 | 0.57038312 | 0.724608177 | 1 | 0.570383116 | 0.570383116 | 0.570383116 | 0.928281975 | -0.032320082 | FALSE | TRUE | FALSE |
| PSMD7 | 0.5294639 | 0.71731322 | 1 | 0.529463944 | 0.529463944 | 0.529463944 | 0.57084765 | 0.724366765 | 1 | 0.570847652 | 0.570847652 | 0.570847652 | 0.927504811 | -0.032683829 | FALSE | TRUE | FALSE |
| TMEM225 | 0.5294287 | 0.71736055 | 1 | 0.529428699 | 0.529428699 | 0.529428699 | 0.57044395 | 0.724511667 | 1 | 0.570443948 | 0.570443948 | 0.570443948 | 0.928099422 | -0.032405498 | FALSE | TRUE | FALSE |
| SVIP | 0.5294273 | 0.71740786 | 1 | 0.529427253 | 0.529427253 | 0.529427253 | 5.15979214 | 0.299549572 | 0.49995 | 0.551669855 | 5.159792144 | 5.159792144 | 0.102606314 | -0.988825913 | FALSE | FALSE | FALSE |
| PRTG | 0.5293772 | 0.71745515 | 0.51535 | 0.529377167 | 4.802636926 | 10.36442262 | 0.02612119 | 0.892095604 | 1 | 0.026121192 | 0.026121192 | 0.512712476 | 20.26619503 | 1.306772218 | FALSE | TRUE | FALSE |
| ELL2 | 0.5292984 | 0.71750243 | 1 | 0.529298447 | 0.529298447 | 10.45283575 | 0.57083879 | 0.724415082 | 1 | 0.570838794 | 0.570838794 | 11.29025354 | 0.927229285 | -0.03281286 | FALSE | TRUE | FALSE |
| SUMO1 | 0.5289549 | 0.7175497 | 1 | 0.528954876 | 0.528954876 | 0.528954876 | 0.53783564 | 0.739441485 | 1 | 0.537835639 | 0.537835639 | 0.537835639 | 0.983487961 | -0.007230952 | FALSE | TRUE | FALSE |
| FBXO3 | 0.528944 | 0.71759694 | 1 | 0.528943995 | 0.528943995 | 10.47540641 | 0.57044396 | 0.724463384 | 1 | 0.570443956 | 0.570443956 | 11.31057892 | 0.927249714 | -0.032803292 | FALSE | TRUE | FALSE |
| RIPK4 | 0.5286336 | 0.71764417 | 0.5239 | 0.528633552 | 0.528633552 | 10.32040186 | 0.08026277 | 0.88022781 | 0.50965 | 0.742459479 | 0.742459479 | 14.5365526 | 6.586286135 | 0.818640594 | FALSE | TRUE | FALSE |
| PYY | 0.5285377 | 0.71769139 | 1 | 0.528537661 | 0.528537661 | 0.528537661 | 0.55196181 | 0.732650439 | 1 | 5.175751926 | 5.175751926 | 5.175751926 | 0.957562011 | -0.018833092 | FALSE | TRUE | FALSE |
| LRRTM2 | 0.528458 | 0.71773859 | 1 | 0.528458027 | 0.528458027 | 10.46351558 | 0.55225641 | 0.732559778 | 1 | 5.208354868 | 5.208354868 | 10.94787316 | 0.956907002 | -0.019130268 | FALSE | TRUE | FALSE |
| E2F1 | 0.528324 | 0.71778577 | 1 | 0.528324003 | 0.528324003 | 10.47903911 | 0.53495362 | 0.740211508 | 1 | 5.055540891 | 49.03455163 | 49.03455163 | 0.987607124 | -0.005415786 | FALSE | TRUE | FALSE |
| DENND2A | 0.5282193 | 0.71783293 | 0.5227 | 1.213921704 | 4.855361116 | 23.73593282 | 0.17455151 | 0.858451315 | 1 | 0.17455151 | 0.17455151 | 3.422210536 | 3.02615154 | 0.480890672 | FALSE | TRUE | FALSE |
| TM6SF1 | 0.5281366 | 0.71788008 | 0.01045 | 0.528136558 | 0.528136558 | 1.190567628 | 0.08255287 | 0.879660312 | 0.5057 | 0.733559957 | 0.733559957 | 0.733559957 | 6.397555448 | 0.806014059 | FALSE | TRUE | FALSE |
| RAB1B | 0.5281035 | 0.71792721 | 1 | 0.528103529 | 0.528103529 | 0.528103529 | 2.2527988 | 0.46797587 | 0.49845 | 0.563261612 | 2.252798799 | 2.252798799 | 0.234421081 | -0.630003337 | FALSE | FALSE | FALSE |
| IPMK | 0.5279156 | 0.71797433 | 1 | 0.527915627 | 0.527915627 | 10.4752767 | 0.5692331 | 0.724993512 | 1 | 0.569233098 | 0.569233098 | 11.30580989 | 0.927415551 | -0.032725626 | FALSE | TRUE | FALSE |
| MAP2K6 | 0.5278093 | 0.71802143 | 1 | 0.527809263 | 0.527809263 | 0.527809263 | 0.56922748 | 0.725041599 | 1 | 0.569227479 | 0.569227479 | 0.569227479 | 0.927237848 | -0.03280885 | FALSE | TRUE | FALSE |
| PPP4C | 0.5277884 | 0.71806851 | 1 | 0.527788429 | 0.527788429 | 10.46492739 | 5.15458141 | 0.300427659 | 0.50735 | 5.154581405 | 20.61694003 | 20.61694003 | 0.102392103 | -0.989733538 | FALSE | FALSE | FALSE |
| PTH2 | 0.5277289 | 0.71811558 | 0.00375 | 0.527728918 | 0.527728918 | 0.527728918 | 0.53365698 | 0.741103867 | 1 | 0.533656977 | 0.533656977 | 0.533656977 | 0.98889163 | -0.004851299 | FALSE | TRUE | FALSE |
| KCNT2 | 0.5275801 | 0.71816263 | 0.52285 | 0.527580149 | 0.527580149 | 10.34740246 | 2.92986259 | 0.416669126 | 0.2637 | 0.732684044 | 2.929862593 | 14.40358448 | 0.180069929 | -0.744558806 | FALSE | FALSE | FALSE |
| ID2 | 0.5275077 | 0.71820966 | 1 | 0.527507668 | 0.527507668 | 0.527507668 | 0.5690439 | 0.725137721 | 1 | 0.569043899 | 0.569043899 | 0.569043899 | 0.927006983 | -0.032916994 | FALSE | TRUE | FALSE |
| COQ10B | 0.5273297 | 0.71825668 | 1 | 4.949521535 | 4.949521535 | 4.949521535 | 0.58677629 | 0.717980749 | 1 | 0.586776286 | 0.586776286 | 0.586776286 | 0.898689576 | -0.046390296 | FALSE | TRUE | FALSE |
| HOXB7 | 0.5272195 | 0.71830368 | 0.9837 | 19.75257013 | 316.1492489 | 1265.287327 | 2.82725505 | 0.424782187 | 0.5025 | 2.827255051 | 11.31023082 | 11.31023082 | 0.1864775 | -0.729373561 | FALSE | FALSE | FALSE |
| HOMER1 | 0.5267973 | 0.71835066 | 1 | 0.526797282 | 0.526797282 | 0.526797282 | 19.27625 | 0.136343251 | 0.00345 | 0.568499811 | 0.568499811 | 0.568499811 | 0.027328826 | -1.563379026 | FALSE | FALSE | FALSE |
| GPIHBP1 | 0.5267066 | 0.71839764 | 1 | 0.526706557 | 0.526706557 | 0.526706557 | 0.56795215 | 0.725473595 | 1 | 0.567952151 | 0.567952151 | 0.567952151 | 0.9273784 | -0.032743024 | FALSE | TRUE | FALSE |
| NDUFA11 | 0.5265949 | 0.71844459 | 0.50575 | 0.526594912 | 0.526594912 | 0.526594912 | 5.48192641 | 0.287958852 | 0.00165 | 0.576890351 | 5.481926414 | 5.481926414 | 0.096060193 | -1.017456543 | FALSE | FALSE | FALSE |
| PF4 | 0.5264356 | 0.71849153 | 1 | 4.928403384 | 4.928403384 | 4.928403384 | 0.58554147 | 0.718385809 | 1 | 0.585541466 | 0.585541466 | 0.585541466 | 0.899057809 | -0.046212383 | FALSE | TRUE | FALSE |
| CYP4F11 | 0.5263456 | 0.71853845 | 0.88845 | 4.789708203 | 178.9360669 | 715.9091686 | 1478.04414 | 0.001579037 | 0.15475 | 62.56006771 | 5910.989815 | 29234.05917 | 0.00035611 | -3.4484164 | FALSE | FALSE | FALSE |
| SPAG7 | 0.5262857 | 0.71858536 | 1 | 0.526285736 | 0.526285736 | 0.526285736 | 0.55006548 | 0.733283278 | 1 | 5.16564505 | 5.16564505 | 5.16564505 | 0.95676925 | -0.019192791 | FALSE | TRUE | FALSE |
| P4HA2 | 0.5261201 | 0.71863225 | 0.2547 | 0.131613585 | 0.526120107 | 1.157540706 | 0.38629491 | 0.797130962 | 0.50405 | 0.386294905 | 1.545074947 | 1.912479695 | 1.361964913 | 0.13416592 | FALSE | TRUE | FALSE |
| ZNF384 | 0.5260974 | 0.71867912 | 1 | 4.966737535 | 48.12381938 | 98.12611308 | 0.56775368 | 0.725569404 | 1 | 5.363867083 | 52.00909022 | 106.1272047 | 0.926629623 | -0.03309382 | FALSE | TRUE | FALSE |
| SPIC | 0.5260553 | 0.71872597 | 1 | 0.526055335 | 4.932790368 | 4.932790368 | 0.56729746 | 0.725808613 | 1 | 0.567297462 | 5.327486451 | 5.327486451 | 0.927300703 | -0.032779411 | FALSE | TRUE | FALSE |
| RCAN2 | 0.5259192 | 0.71877281 | 1 | 0.525919159 | 0.525919159 | 0.525919159 | 0.56748354 | 0.725712984 | 1 | 0.567483537 | 0.567483537 | 0.567483537 | 0.92675668 | -0.033034275 | FALSE | TRUE | FALSE |
| GNAI3 | 0.5257231 | 0.71881964 | 0.5021 | 0.52572312 | 0.52572312 | 2.611275629 | 0.6680884 | 0.681287896 | 0.50345 | 0.668088402 | 0.668088402 | 0.668088402 | 0.78690652 | -0.104076856 | FALSE | TRUE | FALSE |
| ADORA1 | 0.5255101 | 0.71886645 | 1 | 0.525510111 | 0.525510111 | 10.41990766 | 0.56703 | 0.725856404 | 1 | 0.567029995 | 0.567029995 | 0.567029995 | 0.926776565 | -0.033024957 | FALSE | TRUE | FALSE |
| SNX3 | 0.5250234 | 0.71891324 | 0.00575 | 0.525023401 | 0.525023401 | 0.525023401 | 0.56625155 | 0.726095103 | 1 | 0.566251546 | 0.566251546 | 0.566251546 | 0.927191113 | -0.03283074 | FALSE | TRUE | FALSE |
| PIP4K2B | 0.5249558 | 0.71896002 | 1 | 0.524955834 | 0.524955834 | 10.38240061 | 0.56674598 | 0.725904178 | 1 | 0.566745981 | 0.566745981 | 11.22390297 | 0.926263003 | -0.033265682 | FALSE | TRUE | FALSE |
| CCDC106 | 0.5248846 | 0.71900679 | 1 | 0.524884569 | 0.524884569 | 10.40336347 | 0.56628075 | 0.726047398 | 1 | 0.56628075 | 0.56628075 | 0.56628075 | 0.926898132 | -0.032967993 | FALSE | TRUE | FALSE |
| GSK3B | 0.5245206 | 0.71905354 | 1 | 0.52452058 | 0.52452058 | 10.39964324 | 19.1756258 | 0.13696616 | 0.00405 | 0.566180516 | 0.566180516 | 11.23731446 | 0.027353505 | -1.562987013 | FALSE | FALSE | FALSE |
| CTR9 | 0.5244858 | 0.71910027 | 0.5167 | 0.524485777 | 0.524485777 | 10.24836226 | 0.66641382 | 0.682465504 | 0.50325 | 0.666413817 | 0.666413817 | 13.05770796 | 0.787027166 | -0.104010277 | FALSE | TRUE | FALSE |
| KRTAP26-1 | 0.5244013 | 0.71914698 | 1 | 0.524401338 | 0.524401338 | 0.524401338 | 0.56561842 | 0.726380971 | 1 | 0.565618425 | 0.565618425 | 0.565618425 | 0.927129165 | -0.032859757 | FALSE | TRUE | FALSE |
| LSM10 | 0.5243721 | 0.71919368 | 0.0046 | 0.52437212 | 0.52437212 | 0.52437212 | 0.56562731 | 0.726333371 | 1 | 0.565627311 | 0.565627311 | 0.565627311 | 0.927062944 | -0.032890778 | FALSE | TRUE | FALSE |
| MC3R | 0.5241579 | 0.71924037 | 0.7554 | 1.208360859 | 4.832210394 | 4.832210394 | 1.50461996 | 0.535119207 | 0.74985 | 6.017080948 | 96.30073002 | 96.30073002 | 0.34836561 | -0.457964724 | FALSE | FALSE | FALSE |
| RANBP17 | 0.5239283 | 0.71928703 | 0.4804 | 0.280963767 | 17.96434679 | 157.8707441 | 0.71093233 | 0.660870728 | 0.2083 | 0.326056133 | 6.352976883 | 25.39928937 | 0.736959514 | -0.13255637 | FALSE | TRUE | FALSE |
| CTDSP1 | 0.5236975 | 0.71933369 | 1 | 0.523697502 | 0.523697502 | 0.523697502 | 0.56516997 | 0.726476123 | 1 | 0.565169966 | 0.565169966 | 0.565169966 | 0.926619484 | -0.033098572 | FALSE | TRUE | FALSE |
| ASCL2 | 0.5236362 | 0.71938033 | 1 | 0.523636219 | 0.523636219 | 0.523636219 | 0.72548819 | 0.654214254 | 1 | 6.884312447 | 6.884312447 | 6.884312447 | 0.721770843 | -0.141600666 | FALSE | TRUE | FALSE |
| ZSCAN30 | 0.5236302 | 0.71942695 | 0.7554 | 0.523630215 | 4.772938024 | 4.772938024 | 0.08403126 | 0.879319272 | 0.5055 | 0.752599911 | 0.752599911 | 1.668164797 | 6.231374423 | 0.794583847 | FALSE | TRUE | FALSE |
| CDKN2D | 0.5234886 | 0.71947355 | 1 | 0.523488557 | 0.523488557 | 0.523488557 | 0.56498666 | 0.726618717 | 1 | 0.564986659 | 0.564986659 | 0.564986659 | 0.926550298 | -0.033131 | FALSE | TRUE | FALSE |
| GYPA | 0.5233602 | 0.71952014 | 1 | 0.523360216 | 0.523360216 | 0.523360216 | 0.56452582 | 0.726950817 | 1 | 0.564525823 | 0.564525823 | 0.564525823 | 0.927079318 | -0.032883107 | FALSE | TRUE | FALSE |
| MEGF9 | 0.5231885 | 0.71956671 | 1 | 0.523188474 | 0.523188474 | 10.345851 | 2.23493099 | 0.468686616 | 0.50585 | 2.234930989 | 2.234930989 | 11.06425337 | 0.234096031 | -0.630605949 | FALSE | FALSE | FALSE |
| GSPT1 | 0.523071 | 0.71961327 | 1 | 0.523070976 | 0.523070976 | 10.37377625 | 0.56471729 | 0.726713692 | 1 | 0.564717287 | 0.564717287 | 11.21100877 | 0.926252813 | -0.03327046 | FALSE | TRUE | FALSE |
| RPL5 | 0.5229726 | 0.71965981 | 1 | 0.522972556 | 0.522972556 | 0.522972556 | 0.56456673 | 0.726856021 | 1 | 0.564566727 | 0.564566727 | 0.564566727 | 0.926325501 | -0.03323638 | FALSE | TRUE | FALSE |
| SPSB1 | 0.5229165 | 0.71970633 | 1 | 0.522916542 | 0.522916542 | 0.522916542 | 0.5646743 | 0.726808596 | 1 | 0.564674296 | 0.564674296 | 0.564674296 | 0.926049842 | -0.033365638 | FALSE | TRUE | FALSE |
| RNPS1 | 0.5228845 | 0.71975284 | 1 | 0.522884543 | 0.522884543 | 10.3832356 | 0.56445578 | 0.726998188 | 0.00805 | 0.564455784 | 0.564455784 | 0.564455784 | 0.926351644 | -0.033224123 | FALSE | TRUE | FALSE |
| ZNF462 | 0.5228593 | 0.71979933 | 1 | 8226653694 | 1.45701E+13 | 2.35216E+15 | 0.07724323 | 0.881015749 | 1 | 10765319697 | 2.42748E+14 | 4.63671E+15 | 6.768998587 | 0.830524423 | FALSE | TRUE | FALSE |
| ETS1 | 0.5227117 | 0.7198458 | 1 | 0.522711659 | 0.522711659 | 10.35130639 | 0.56441114 | 0.727045541 | 1 | 0.564411144 | 0.564411144 | 11.19032187 | 0.926118601 | -0.033333393 | FALSE | TRUE | FALSE |
| TNFSF12 | 0.5226785 | 0.71989226 | 1 | 0.522678485 | 0.522678485 | 0.522678485 | 0.56454427 | 0.726903428 | 1 | 0.56454427 | 0.56454427 | 0.56454427 | 0.925841448 | -0.033463381 | FALSE | TRUE | FALSE |
| FBXL14 | 0.5224312 | 0.7199387 | 1 | 47.7195801 | 4823.619957 | 4823.619957 | 0.54652639 | 0.735029729 | 1 | 49.90852562 | 5043.717137 | 52585.68549 | 0.955912095 | -0.019582043 | FALSE | TRUE | FALSE |
| NKIRAS2 | 0.5223371 | 0.71998513 | 0.751 | 0.522337067 | 2.089174632 | 2.089174632 | 0.16968723 | 0.859784138 | 1 | 0.169687229 | 0.169687229 | 0.169687229 | 3.078234417 | 0.48830169 | FALSE | TRUE | FALSE |
| GJA1 | 0.5222822 | 0.72003154 | 0.9421 | 4.861026939 | 185.7190977 | 185.7190977 | 0.18072192 | 0.856954185 | 1 | 0.180721917 | 0.180721917 | 3.59028673 | 2.889977314 | 0.460894434 | FALSE | TRUE | FALSE |
| SPOCK2 | 0.5221826 | 0.72007794 | 0.01085 | 0.5221826 | 0.5221826 | 10.35379276 | 0.5639782 | 0.72723478 | 1 | 0.563978198 | 0.563978198 | 11.19408422 | 0.925891465 | -0.033439919 | FALSE | TRUE | FALSE |
| RPLP2 | 0.5221644 | 0.72012431 | 1 | 2.088313589 | 8.353967186 | 8.353967186 | 0.5768181 | 0.72182927 | 1 | 0.576818098 | 0.576818098 | 0.576818098 | 0.905249609 | -0.043231654 | FALSE | TRUE | FALSE |
| SLC30A1 | 0.522163 | 0.72017067 | 0.0144 | 0.522162979 | 0.522162979 | 10.3199033 | 0.5641085 | 0.727187496 | 1 | 0.564108496 | 0.564108496 | 11.16474888 | 0.92564282 | -0.033556563 | FALSE | TRUE | FALSE |
| HMGN1 | 0.5220161 | 0.72021702 | 1 | 0.522016066 | 0.522016066 | 0.522016066 | 0.56323962 | 0.727470943 | 1 | 0.563239616 | 0.563239616 | 0.563239616 | 0.926809924 | -0.033009325 | FALSE | TRUE | FALSE |
| EGLN3 | 0.5219559 | 0.72026335 | 1 | 0.521955896 | 0.521955896 | 0.521955896 | 0.56370363 | 0.727376527 | 1 | 0.563703627 | 0.563703627 | 0.563703627 | 0.925940284 | -0.033417021 | FALSE | TRUE | FALSE |
| TACR2 | 0.5219336 | 0.72030966 | 0.50365 | 0.521933561 | 0.521933561 | 1.153271124 | 11.4159309 | 0.184022691 | 0.007 | 1.265418707 | 11.41593092 | 11.41593092 | 0.045719755 | -1.339896109 | FALSE | FALSE | FALSE |
| FAM169A | 0.5216793 | 0.72035596 | 1 | 0.521679271 | 0.521679271 | 10.3164293 | 5.15165058 | 0.301011367 | 0.507 | 5.151650576 | 5.151650576 | 18.47941516 | 0.101264491 | -0.994542818 | FALSE | FALSE | FALSE |
| PGBD5 | 0.5215952 | 0.72040224 | 1 | 4.913343206 | 4.913343206 | 10.30829318 | 0.56320482 | 0.727518125 | 1 | 5.310473302 | 5.310473302 | 11.14649349 | 0.926119882 | -0.033332792 | FALSE | TRUE | FALSE |
| UQCRQ | 0.5215871 | 0.7204485 | 1 | 0.521587115 | 0.521587115 | 0.521587115 | 0.56332718 | 0.727423744 | 1 | 0.56332718 | 0.56332718 | 0.56332718 | 0.9259044 | -0.033433852 | FALSE | TRUE | FALSE |
| TUBB8 | 0.5213125 | 0.72049475 | 1 | 4797.76366 | 533211.7154 | 987996.8607 | 5.66975192 | 0.279127112 | 0.96875 | 547.2369523 | 5580.920171 | 58304.70937 | 0.091946267 | -1.036465896 | FALSE | FALSE | FALSE |
| SNCA | 0.521179 | 0.72054099 | 1 | 0.521178956 | 0.521178956 | 0.521178956 | 0.55589203 | 0.730594619 | 1 | 0.555892033 | 0.555892033 | 0.555892033 | 0.937554282 | -0.028003579 | FALSE | TRUE | FALSE |

| | | | | | | | | | | | | | | | | | |
|---|---|---|---|---|---|---|---|---|---|---|---|---|---|---|---|---|---|
| FUBP1 | 0.520904 | 0.7205872 | 1 | 0.520904025 | 0.520904025 | 10.29267265 | 0.56287701 | 0.727612439 | 1 | 0.562877008 | 0.562877008 | 11.13813573 | 0.925431343 | -0.033655796 | FALSE | TRUE | FALSE |
| RBM38 | 0.5208903 | 0.72063341 | 1 | 0.520890345 | 0.520890345 | 0.520890345 | 0.56249987 | 0.727988985 | 1 | 0.562499868 | 0.562499868 | 0.562499868 | 0.926027498 | -0.033376117 | FALSE | TRUE | FALSE |
| SIRPA | 0.5208235 | 0.72067959 | 1 | 0.520823455 | 0.520823455 | 10.29535628 | 0.56272081 | 0.727753777 | 1 | 0.562720807 | 0.562720807 | 11.13912101 | 0.925545045 | -0.03360244 | FALSE | TRUE | FALSE |
| CYC1 | 0.5207955 | 0.72072576 | 0.01005 | 0.520795489 | 0.520795489 | 10.33798261 | 0.54515753 | 0.735739186 | 0.50585 | 5.14203279 | 5.14203279 | 5.14203279 | 0.955311931 | -0.019854798 | FALSE | TRUE | FALSE |
| RGS19 | 0.5206839 | 0.72077192 | 1 | 97.77683317 | 4828.539324 | 50403.3323 | 0.56192908 | 0.728129906 | 1 | 51.60681066 | 5244.873189 | 5244.873189 | 0.926600804 | -0.033107327 | FALSE | TRUE | FALSE |
| ZNF391 | 0.5206743 | 0.72081806 | 0.75335 | 1.185031565 | 4.739612421 | 4.739612421 | 1.43314396 | 0.542359243 | 0.87165 | 13.43068264 | 129.097148 | 129.097148 | 0.363309121 | -0.439723699 | FALSE | FALSE | FALSE |
| CDC42EP3 | 0.5201449 | 0.72086418 | 1 | 0.520144918 | 0.520144918 | 0.520144918 | 0.5619696 | 0.728082949 | 1 | 0.561969603 | 0.561969603 | 0.561969603 | 0.925574827 | -0.033588466 | FALSE | TRUE | FALSE |
| AQP4 | 0.5201089 | 0.72091029 | 0.5046 | 0.520108946 | 0.520108946 | 0.520108946 | 0.15865555 | 0.862414388 | 1 | 2.537140442 | 23.76269197 | 23.76269197 | 3.27822731 | 0.515639064 | FALSE | TRUE | FALSE |
| RNF11 | 0.5200281 | 0.72095638 | 1 | 0.520028071 | 0.520028071 | 0.520028071 | 4.91439058 | 0.310241883 | 0.50075 | 4.914390577 | 4.914390577 | 4.914390577 | 0.105817408 | -0.975442882 | FALSE | FALSE | FALSE |
| ACTB | 0.5198465 | 0.72100246 | 1 | 0.519846533 | 0.519846533 | 10.27751033 | 0.56190786 | 0.728176845 | 1 | 0.561907856 | 0.561907856 | 11.12442601 | 0.925145515 | -0.033789952 | FALSE | TRUE | FALSE |
| CCNE1 | 0.5198335 | 0.72104852 | 1 | 0.519833547 | 0.519833547 | 10.30891142 | 0.56157767 | 0.728317559 | 1 | 0.56157767 | 0.56157767 | 11.14805238 | 0.925666342 | -0.033545528 | FALSE | TRUE | FALSE |
| SUSD3 | 0.5198243 | 0.72109456 | 1 | 0.519824292 | 0.519824292 | 0.519824292 | 0.56142651 | 0.728411283 | 1 | 0.56142651 | 0.56142651 | 0.56142651 | 0.925899085 | -0.033436345 | FALSE | TRUE | FALSE |
| SERTAD4 | 0.519535 | 0.72114059 | 1 | 0.519534959 | 0.519534959 | 10.32445272 | 0.54385554 | 0.736532481 | 1 | 5.106188044 | 5.106188044 | 5.106188044 | 0.955281173 | -0.019868782 | FALSE | TRUE | FALSE |
| RANBP3 | 0.5195166 | 0.72118661 | 1 | 0.519516578 | 0.519516578 | 10.24413581 | 0.56151421 | 0.72836443 | 0.01845 | 0.56151421 | 0.56151421 | 11.09101913 | 0.925206467 | -0.03376134 | FALSE | TRUE | FALSE |
| ZNF250 | 0.5193014 | 0.7212326 | 0.01725 | 0.519301408 | 0.519301408 | 10.26614066 | 0.56132023 | 0.728551737 | 1 | 0.561320227 | 0.561320227 | 11.11222738 | 0.925142875 | -0.033791191 | FALSE | TRUE | FALSE |
| MAU2 | 0.5190968 | 0.72127859 | 0.01965 | 0.519096784 | 0.519096784 | 10.22664803 | 0.54371516 | 0.736620311 | 1 | 5.148054136 | 5.148054136 | 18.41007559 | 0.954721918 | -0.020123107 | FALSE | TRUE | FALSE |
| ITGB5 | 0.5189427 | 0.72132456 | 0.12465 | 0.032496733 | 0.518942711 | 2.074540744 | 0.07766613 | 0.880916648 | 0.6473 | 0.310390395 | 4.961202243 | 24.43205931 | 6.681712159 | 0.824887763 | FALSE | TRUE | FALSE |
| IL1RL1 | 0.5187895 | 0.72137051 | 0.50465 | 0.518789503 | 0.518789503 | 0.518789503 | 0.08347671 | 0.879476184 | 1 | 0.083476708 | 0.083476708 | 0.083476708 | 6.214781523 | 0.793425866 | FALSE | TRUE | FALSE |
| AGA | 0.5183549 | 0.72141645 | 0.63025 | 0.924070326 | 8.288683022 | 33.15717124 | 0.38583238 | 0.797309205 | 0.8187 | 1.543174694 | 6.173621865 | 24.70435034 | 1.343471727 | 0.128228531 | FALSE | TRUE | FALSE |
| SPATA25 | 0.5183514 | 0.72146237 | 1 | 0.518351418 | 0.518351418 | 0.518351418 | 0.55986502 | 0.728832224 | 1 | 0.559865025 | 0.559865025 | 0.559865025 | 0.925850687 | -0.033459047 | FALSE | TRUE | FALSE |
| KLF7 | 0.5178784 | 0.72150828 | 1 | 0.517878386 | 0.517878386 | 10.27778779 | 0.54010304 | 0.738060678 | 1 | 0.540103038 | 0.540103038 | 0.540103038 | 0.95885109 | -0.018248834 | FALSE | TRUE | FALSE |
| MED26 | 0.517852 | 0.72155418 | 0.02095 | 0.517851987 | 0.517851987 | 10.18249759 | 0.56001155 | 0.728785518 | 1 | 0.560011546 | 0.560011546 | 11.03381947 | 0.924716625 | -0.033991334 | FALSE | TRUE | FALSE |
| C2orf69 | 0.5177371 | 0.72160006 | 1 | 2.070898803 | 2.070898803 | 2.070898803 | 0.56575414 | 0.726285753 | 1 | 0.565754137 | 0.565754137 | 0.565754137 | 0.915127432 | -0.038518426 | FALSE | TRUE | FALSE |
| PAFAH1B1 | 0.5174897 | 0.72164592 | 1 | 0.517489682 | 0.517489682 | 10.26334508 | 0.55951833 | 0.728878915 | 1 | 0.559518333 | 0.559518333 | 11.10804431 | 0.924884229 | -0.033912626 | FALSE | TRUE | FALSE |
| NFKBIA | 0.5173833 | 0.72169177 | 1 | 0.517383306 | 0.517383306 | 0.517383306 | 0.5594337 | 0.728972245 | 1 | 0.559433702 | 0.559433702 | 0.559433702 | 0.924833995 | -0.033936215 | FALSE | TRUE | FALSE |
| TRAF3 | 0.517079 | 0.7217376 | 1 | 0.517079045 | 0.517079045 | 10.19564899 | 0.55920628 | 0.729018886 | 1 | 0.559206281 | 0.559206281 | 11.04502995 | 0.924666018 | -0.034015103 | FALSE | TRUE | FALSE |
| SLC22A4 | 0.517043 | 0.72178342 | 0.50395 | 0.517043039 | 8.268661762 | 8.268661762 | 0.38090767 | 0.799049236 | 0.8145 | 1.523549948 | 6.095399595 | 24.39249061 | 1.357397289 | 0.132706977 | FALSE | TRUE | FALSE |
| KCMF1 | 0.5169036 | 0.72182922 | 1 | 0.51690356 | 0.51690356 | 10.24851679 | 0.55892118 | 0.729205276 | 1 | 0.558921184 | 0.558921184 | 11.09313496 | 0.924823704 | -0.033941047 | FALSE | TRUE | FALSE |
| GLRX2 | 0.5168559 | 0.72187501 | 1 | 0.516855904 | 0.516855904 | 0.516855904 | 5.06905989 | 0.304204405 | 0.50315 | 5.06905989 | 5.06905989 | 5.06905989 | 0.101962872 | -0.991557941 | FALSE | FALSE | FALSE |
| HAMP | 0.516654 | 0.72192078 | 1 | 0.516654008 | 0.516654008 | 0.516654008 | 0.55865867 | 0.729298369 | 1 | 0.55865867 | 0.55865867 | 0.55865867 | 0.924811581 | -0.033946741 | FALSE | TRUE | FALSE |
| GLRX | 0.5166139 | 0.72196654 | 0.00295 | 0.516613941 | 0.516613941 | 0.516613941 | 0.558104 | 0.729577236 | 1 | 0.558103995 | 0.558103995 | 0.558103995 | 0.925658919 | -0.03354901 | FALSE | TRUE | FALSE |
| ZNF274 | 0.5165754 | 0.72201228 | 1 | 0.516575394 | 0.516575394 | 10.1981528 | 0.54908362 | 0.733598549 | 1 | 0.549083617 | 0.549083617 | 10.85676164 | 0.940795496 | -0.026504771 | FALSE | TRUE | FALSE |
| RSL24D1 | 0.5165467 | 0.722058 | 1 | 0.51654671 | 0.51654671 | 0.51654671 | 0.55865645 | 0.72934489 | 1 | 0.558656447 | 0.558656447 | 0.558656447 | 0.924623197 | -0.034035215 | FALSE | TRUE | FALSE |
| RPS16 | 0.5162693 | 0.72210371 | 1 | 0.516269289 | 0.516269289 | 0.516269289 | 0.55560837 | 0.730732722 | 1 | 0.555608371 | 0.555608371 | 0.555608371 | 0.929196383 | -0.031892489 | FALSE | TRUE | FALSE |
| KCNMB2 | 0.5160845 | 0.7221494 | 1 | 0.516084529 | 0.516084529 | 0.516084529 | 0.55769947 | 0.729762812 | 1 | 0.557699468 | 0.557699468 | 0.557699468 | 0.925381066 | -0.033679391 | FALSE | TRUE | FALSE |
| SLC18A2 | 0.5157831 | 0.72219508 | 0.50955 | 0.515783104 | 0.515783104 | 1.12171638 | 0.0817588 | 0.879862257 | 0.012 | 0.081758798 | 0.081758798 | 1.618813758 | 6.308594484 | 0.799932612 | FALSE | TRUE | FALSE |
| IFI27 | 0.5154904 | 0.72224075 | 1 | 0.515490379 | 0.515490379 | 0.515490379 | 0.5568619 | 0.729994415 | 1 | 0.556861903 | 0.556861903 | 0.556861903 | 0.925705953 | -0.033526943 | FALSE | TRUE | FALSE |
| TIPRL | 0.5152246 | 0.7222864 | 1 | 4.840455784 | 4.840455784 | 4.840455784 | 5.24057484 | 0.296608625 | 0.4996 | 0.557100768 | 5.240574844 | 5.240574844 | 0.098314523 | -1.007382322 | FALSE | FALSE | FALSE |
| CXCR4 | 0.5151506 | 0.72233203 | 1 | 0.515150649 | 0.515150649 | 10.23279097 | 0.55724091 | 0.729855502 | 1 | 0.557240907 | 0.557240907 | 0.557240907 | 0.924466676 | -0.034108739 | FALSE | TRUE | FALSE |
| DCTN3 | 0.5149687 | 0.72237765 | 1 | 0.514968671 | 0.514968671 | 0.514968671 | 0.55653966 | 0.730179397 | 0.00405 | 0.556539662 | 0.556539662 | 0.556539662 | 0.925304531 | -0.033715311 | FALSE | TRUE | FALSE |
| USP18 | 0.514829 | 0.72242326 | 1 | 0.514828972 | 0.514828972 | 10.21950086 | 0.53878324 | 0.739054698 | 1 | 0.538783241 | 0.538783241 | 0.538783241 | 0.955540063 | -0.0197511 | FALSE | TRUE | FALSE |
| SPRR1A | 0.5147472 | 0.72246885 | 0.0025 | 0.514747198 | 0.514747198 | 0.514747198 | 0.53908146 | 0.738968585 | 1 | 0.539081461 | 5.036918842 | 5.036918842 | 0.954859766 | -0.020060406 | FALSE | TRUE | FALSE |
| TCFL5 | 0.5146363 | 0.72251442 | 1 | 0.514636287 | 0.514636287 | 10.21032792 | 0.55672697 | 0.730086939 | 1 | 0.556726967 | 0.556726967 | 11.05603623 | 0.92439619 | -0.034141853 | FALSE | TRUE | FALSE |
| HNRNPA0 | 0.5145585 | 0.72255998 | 0.0118 | 0.514558491 | 0.514558491 | 10.20351074 | 0.55397436 | 0.731695209 | 1 | 0.553974355 | 0.553974355 | 10.99636666 | 0.928848937 | -0.032054912 | FALSE | TRUE | FALSE |
| GPR22 | 0.5144918 | 0.72260553 | 1 | 0.514491817 | 0.514491817 | 10.21514512 | 0.55648865 | 0.7302256 | 1 | 0.556488649 | 0.556488649 | 11.05862909 | 0.924532455 | -0.034077838 | FALSE | TRUE | FALSE |
| RFXAP | 0.5144558 | 0.72265105 | 1 | 0.514455804 | 0.514455804 | 0.514455804 | 0.55605503 | 0.730456364 | 1 | 0.556055028 | 0.556055028 | 0.556055028 | 0.925188655 | -0.033769701 | FALSE | TRUE | FALSE |
| SLC35B4 | 0.5142838 | 0.72269656 | 0.75335 | 0.514283774 | 2.057477655 | 2.057477655 | 0.16672415 | 0.860526329 | 1 | 0.166724148 | 0.166724148 | 0.166724148 | 3.084638788 | 0.489204315 | FALSE | TRUE | FALSE |
| B4GALT6 | 0.5142386 | 0.72274206 | 1 | 0.514238637 | 0.514238637 | 10.20086954 | 0.55633009 | 0.730271787 | 1 | 0.556330095 | 0.556330095 | 0.556330095 | 0.924340857 | -0.03416785 | FALSE | TRUE | FALSE |
| CALN1 | 0.514069 | 0.72278754 | 0.5058 | 4.839796939 | 4.839796939 | 4.839796939 | 187.575506 | 0.014374574 | 0.1327 | 46.89656117 | 1857.479397 | 1857.479397 | 0.002740598 | -2.562154705 | FALSE | FALSE | FALSE |
| CBLN1 | 0.514013 | 0.722833 | 1 | 0.514013049 | 0.514013049 | 0.514013049 | 0.55622463 | 0.730364109 | 1 | 0.556224633 | 0.556224633 | 0.556224633 | 0.924110545 | -0.034276074 | FALSE | TRUE | FALSE |
| CTXN1 | 0.5139996 | 0.72287845 | 1 | 0.51399965 | 0.51399965 | 0.51399965 | 0.51947509 | 0.747575164 | 1 | 0.519475089 | 0.519475089 | 0.519475089 | 0.989459669 | -0.004601903 | FALSE | TRUE | FALSE |
| KRTAP17-1 | 0.5139286 | 0.72292388 | 0.00315 | 0.51392863 | 0.51392863 | 0.51392863 | 0.55544533 | 0.730870673 | 0.0032 | 0.555445333 | 0.555445333 | 0.555445333 | 0.925255104 | -0.03373851 | FALSE | TRUE | FALSE |
| TEX261 | 0.513909 | 0.7229693 | 1 | 0.51390901 | 0.51390901 | 0.51390901 | 0.55567305 | 0.730686705 | 1 | 0.55567305 | 0.55567305 | 0.55567305 | 0.924840623 | -0.033933102 | FALSE | TRUE | FALSE |
| MAP3K2 | 0.5137996 | 0.7230147 | 1 | 0.513799616 | 0.513799616 | 10.16665788 | 0.55609752 | 0.730410245 | 1 | 0.556097517 | 0.556097517 | 11.01769915 | 0.92393798 | -0.03435718 | FALSE | TRUE | FALSE |
| PNRC1 | 0.5137281 | 0.72306008 | 1 | 0.513728109 | 0.513728109 | 0.513728109 | 0.53369173 | 0.741019165 | 1 | 0.533691726 | 0.533691726 | 0.533691726 | 0.962593354 | -0.016557141 | FALSE | TRUE | FALSE |
| CST8 | 0.5135716 | 0.72310545 | 1 | 0.513571565 | 0.513571565 | 0.513571565 | 0.55521795 | 0.731146111 | 1 | 0.555217951 | 0.555217951 | 0.555217951 | 0.924990923 | -0.033862529 | FALSE | TRUE | FALSE |
| RBM4 | 0.513559 | 0.72315081 | 0.00415 | 0.513559029 | 0.513559029 | 0.513559029 | 0.55550788 | 0.730778723 | 1 | 0.555507878 | 0.555507878 | 0.555507878 | 0.924485592 | -0.034099853 | FALSE | TRUE | FALSE |
| MEX3C | 0.513342 | 0.72319614 | 0.0133 | 0.513341968 | 0.513341968 | 10.17152484 | 0.55547525 | 0.730824707 | 1 | 0.555475248 | 0.555475248 | 11.01865745 | 0.924149131 | -0.03425794 | FALSE | TRUE | FALSE |
| RSPRY1 | 0.5131893 | 0.72324147 | 0.5059 | 0.513189293 | 0.513189293 | 2.539038995 | 0.65433229 | 0.687589312 | 0.50705 | 0.654332295 | 0.654332295 | 3.241438933 | 0.784294612 | -0.105520769 | FALSE | TRUE | FALSE |
| RPL18 | 0.5131695 | 0.72328678 | 1 | 0.513169455 | 0.513169455 | 0.513169455 | 0.5553572 | 0.730962554 | 0.00715 | 0.555357201 | 0.555357201 | 0.555357201 | 0.924034936 | -0.034311609 | FALSE | TRUE | FALSE |
| TOP1 | 0.5128147 | 0.72333207 | 1 | 0.512814737 | 0.512814737 | 10.10880761 | 0.5552742 | 0.731100247 | 1 | 0.555274202 | 0.555274202 | 10.96472915 | 0.923534239 | -0.034546999 | FALSE | TRUE | FALSE |
| KRTAP13-3 | 0.5126458 | 0.72337735 | 1 | 0.512645822 | 0.512645822 | 0.512645822 | 0.55428596 | 0.731466711 | 0.0031 | 0.554285965 | 0.554285965 | 0.554285965 | 0.924876064 | -0.03391646 | FALSE | TRUE | FALSE |
| BGLAP | 0.5126361 | 0.72342261 | 1 | 4.848538111 | 4.848538111 | 4.848538111 | 0.57267687 | 0.723542571 | 1 | 0.572676866 | 0.572676866 | 0.572676866 | 0.895157724 | -0.048100436 | FALSE | TRUE | FALSE |
| STYX | 0.5125247 | 0.72346786 | 1 | 0.512524739 | 0.512524739 | 0.512524739 | 0.55458649 | 0.731237792 | 1 | 0.55458649 | 0.55458649 | 0.55458649 | 0.924156552 | -0.034254453 | FALSE | TRUE | FALSE |
| GDF7 | 0.5123717 | 0.72351309 | 1 | 0.512371691 | 0.512371691 | 10.15248135 | 0.5485033 | 0.733868156 | 1 | 2.193825023 | 2.193825023 | 10.88053037 | 0.934126914 | -0.029594115 | FALSE | TRUE | FALSE |
| CLEC4F | 0.512348 | 0.72355831 | 0.50915 | 0.51234798 | 0.51234798 | 1.961178923 | 0.07787146 | 0.880898614 | 1 | 0.693715979 | 6.342923083 | 6.342923083 | 6.579406378 | 0.818186711 | FALSE | TRUE | FALSE |
| SLC31A1 | 0.5123419 | 0.72360351 | 1 | 0.512341947 | 0.512341947 | 0.512341947 | 0.55450238 | 0.73128361 | 1 | 0.554502377 | 0.554502377 | 0.554502377 | 0.92396709 | -0.034343497 | FALSE | TRUE | FALSE |
| DMTN | 0.5123134 | 0.72364869 | 1 | 0.51231339 | 0.51231339 | 10.14684266 | 0.55434461 | 0.731420961 | 1 | 0.554344613 | 0.554344613 | 10.99212501 | 0.924178531 | -0.034244125 | FALSE | TRUE | FALSE |
| RPS9 | 0.5122483 | 0.72369386 | 1 | 0.512248292 | 0.512248292 | 0.512248292 | 0.5544451 | 0.73132941 | 1 | 0.554445103 | 0.554445103 | 0.554445103 | 0.923893617 | -0.034378033 | FALSE | TRUE | FALSE |
| SLC9A8 | 0.5121939 | 0.72373902 | 0.5125 | 0.512193891 | 0.512193891 | 1.944901499 | 0.07978315 | 0.88040072 | 1 | 0.716812848 | 0.716812848 | 1.577679791 | 6.419825529 | 0.807523225 | FALSE | TRUE | FALSE |
| RGS13 | 0.51219 | 0.72378415 | 1 | 2.048620524 | 2.048620524 | 2.048620524 | 0.54300439 | 0.736927218 | 1 | 0.543004389 | 5.105001523 | 5.105001523 | 0.943252003 | -0.025372264 | FALSE | TRUE | FALSE |
| ABHD2 | 0.512163 | 0.72382927 | 1 | 0.51216302 | 0.51216302 | 10.14106324 | 0.55427667 | 0.731512443 | 0.01145 | 0.55427667 | 0.55427667 | 10.98810413 | 0.924020526 | -0.034318381 | FALSE | TRUE | FALSE |
| ZNF705A | 0.5121166 | 0.72387438 | 1 | 0.512116552 | 0.512116552 | 0.512116552 | 0.55405562 | 0.731603861 | 1 | 0.554055617 | 0.554055617 | 0.554055617 | 0.924305315 | -0.034184549 | FALSE | TRUE | FALSE |

| | | | | | | | | | | | | | | | | | |
|---|---|---|---|---|---|---|---|---|---|---|---|---|---|---|---|---|---|
| CADM4 | 0.5118968 | 0.72391947 | 1 | 4.84960588 | 4.84960588 | 10.11832197 | 0.56578464 | 0.726238117 | 1 | 2.262953949 | 2.262953949 | 11.19920698 | 0.904755581 | -0.043468729 | FALSE | TRUE | FALSE |
| FUT9 | 0.5117195 | 0.72396454 | 1 | 4.802924923 | 46.2664312 | 46.2664312 | 50.2046884 | 0.054293801 | 0.25685 | 5.204537455 | 50.20468841 | 50.20468841 | 0.010192663 | -1.991712323 | FALSE | FALSE | FALSE |
| MYBL1 | 0.5116502 | 0.7240096 | 1 | 0.511650232 | 0.511650232 | 10.11891709 | 0.55350849 | 0.731786497 | 1 | 0.55350849 | 0.55350849 | 10.96143145 | 0.924376483 | -0.034151112 | FALSE | TRUE | FALSE |
| LIN7B | 0.5116329 | 0.72405465 | 1 | 0.511632915 | 0.511632915 | 0.511632915 | 0.55349518 | 0.731832116 | 1 | 0.553495182 | 0.553495182 | 0.553495182 | 0.924367424 | -0.034155368 | FALSE | TRUE | FALSE |
| GP5 | 0.5114119 | 0.72409968 | 0.75635 | 0.511411927 | 4.718161132 | 10.05953896 | 0.08423577 | 0.879291541 | 1 | 0.084235769 | 0.084235769 | 1.660218403 | 6.071196753 | 0.783274308 | FALSE | TRUE | FALSE |
| MAPRE3 | 0.5113709 | 0.72414469 | 1 | 0.511370914 | 0.511370914 | 0.511370914 | 0.55357061 | 0.731740862 | 1 | 0.553570614 | 0.553570614 | 0.553570614 | 0.923768172 | -0.034437005 | FALSE | TRUE | FALSE |
| ATP1B3 | 0.5112137 | 0.72418969 | 1 | 0.511213713 | 0.511213713 | 0.511213713 | 0.55336476 | 0.731923302 | 1 | 0.55336476 | 0.55336476 | 0.55336476 | 0.923827736 | -0.034409003 | FALSE | TRUE | FALSE |
| CCBE1 | 0.5110776 | 0.72423467 | 0.37795 | 0.223544572 | 2.043005646 | 2.043005646 | 0.36932193 | 0.802390972 | 0.501 | 0.369321931 | 3.390185294 | 3.390185294 | 1.383826742 | 0.141081719 | FALSE | TRUE | FALSE |
| TP53INP2 | 0.5109976 | 0.72427964 | 1 | 0.510997575 | 0.510997575 | 0.510997575 | 19.8439581 | 0.133233537 | 0.00435 | 4.961144471 | 19.84395812 | 19.84395812 | 0.025750789 | -1.589209462 | FALSE | FALSE | FALSE |
| CD8B | 0.5109358 | 0.7243246 | 1 | 4.786519576 | 4.786519576 | 4.786519576 | 0.57042679 | 0.724559932 | 1 | 0.57042679 | 0.57042679 | 0.57042679 | 0.895707943 | -0.047833574 | FALSE | TRUE | FALSE |
| PLIN1 | 0.5108826 | 0.72436953 | 0.94 | 4.624263655 | 408.5925511 | 408.5925511 | 0.78081271 | 0.628052122 | 0.4989 | 0.087575576 | 0.780812713 | 0.780812713 | 0.654295975 | -0.184225751 | FALSE | TRUE | FALSE |
| TPPP | 0.5106193 | 0.72441446 | 1 | 0.510619294 | 0.510619294 | 0.510619294 | 0.55267046 | 0.732150988 | 1 | 0.552670459 | 0.552670459 | 0.552670459 | 0.923912769 | -0.034369031 | FALSE | TRUE | FALSE |
| CD59 | 0.5106169 | 0.72445936 | 1 | 0.510616874 | 0.510616874 | 0.510616874 | 0.552608 | 0.732196477 | 1 | 0.552608004 | 0.552608004 | 0.552608004 | 0.924012809 | -0.034322008 | FALSE | TRUE | FALSE |
| MRPL27 | 0.5105598 | 0.72450425 | 1 | 0.510559807 | 0.510559807 | 0.510559807 | 0.55286763 | 0.732059963 | 1 | 0.552867635 | 0.552867635 | 0.552867635 | 0.923475665 | -0.034574544 | FALSE | TRUE | FALSE |
| CYP8B1 | 0.5104737 | 0.72454913 | 0.5084 | 0.51047372 | 0.51047372 | 1.125711205 | 0.07927557 | 0.880536893 | 1 | 0.715770037 | 0.715770037 | 1.570303007 | 6.43923106 | 0.808834009 | FALSE | TRUE | FALSE |
| NDRG3 | 0.5104033 | 0.72459399 | 1 | 0.510403329 | 0.510403329 | 0.510403329 | 0.55237136 | 0.732378262 | 1 | 0.552371365 | 0.552371365 | 0.552371365 | 0.924022064 | -0.034317659 | FALSE | TRUE | FALSE |
| PPM1L | 0.510348 | 0.72463884 | 1 | 0.510347999 | 0.510347999 | 10.12017671 | 0.55258873 | 0.732241948 | 1 | 0.552588734 | 0.552588734 | 0.552588734 | 0.923558458 | -0.03453561 | FALSE | TRUE | FALSE |
| TMEM129 | 0.5103367 | 0.72468367 | 1 | 0.510336733 | 0.510336733 | 0.510336733 | 0.54611875 | 0.735251871 | 1 | 0.546118751 | 2.18442067 | 2.18442067 | 0.934479419 | -0.029430259 | FALSE | TRUE | FALSE |
| ZKSCAN2 | 0.5102374 | 0.72472848 | 0.51735 | 0.510237431 | 2.040761495 | 17.19968179 | 0.08251802 | 0.879669505 | 1 | 0.08251802 | 0.08251802 | 1.621001584 | 6.183345525 | 0.791223516 | FALSE | TRUE | FALSE |
| TMEM51 | 0.5102294 | 0.72477328 | 1 | 0.510229371 | 0.510229371 | 0.510229371 | 0.5523059 | 0.732514425 | 1 | 0.552305897 | 0.552305897 | 0.552305897 | 0.923816628 | -0.034414225 | FALSE | TRUE | FALSE |
| WSB2 | 0.5101493 | 0.72481806 | 1 | 0.510149282 | 0.510149282 | 10.11267272 | 0.552444 | 0.732287403 | 1 | 0.552443996 | 0.552443996 | 10.96273805 | 0.923440721 | -0.034590978 | FALSE | TRUE | FALSE |
| NPS | 0.5101257 | 0.72486282 | 1 | 0.510125657 | 0.510125657 | 0.510125657 | 0.55166878 | 0.732786306 | 1 | 0.551668776 | 0.551668776 | 0.551668776 | 0.92469554 | -0.034001237 | FALSE | TRUE | FALSE |
| DRG1 | 0.5100334 | 0.72490758 | 1 | 0.510033367 | 0.510033367 | 0.510033367 | 0.55235057 | 0.732423667 | 1 | 0.552350573 | 0.552350573 | 0.552350573 | 0.923387052 | -0.034616219 | FALSE | TRUE | FALSE |
| EIF3J | 0.5098883 | 0.72495231 | 1 | 0.509888253 | 0.509888253 | 0.509888253 | 0.53326252 | 0.741357617 | 1 | 0.533262519 | 0.533262519 | 0.533262519 | 0.95616743 | -0.019466054 | FALSE | TRUE | FALSE |
| GPBP1L1 | 0.5097681 | 0.72499703 | 1 | 0.509768145 | 0.509768145 | 10.10377787 | 0.55183881 | 0.732741033 | 1 | 0.55183881 | 0.55183881 | 10.94945003 | 0.923762767 | -0.034439547 | FALSE | TRUE | FALSE |
| CDA | 0.5096281 | 0.72504174 | 1 | 0.509628139 | 0.509628139 | 0.509628139 | 0.55193851 | 0.732695744 | 1 | 0.551938514 | 0.551938514 | 0.551938514 | 0.923342231 | -0.034637301 | FALSE | TRUE | FALSE |
| PAX3 | 0.5096014 | 0.72508643 | 0.8586 | 2.037348346 | 32.58818303 | 160.8561791 | 2.09113104 | 0.481009842 | 0.5064 | 3.578266652 | 133.6872049 | 534.8224768 | 0.243696517 | -0.613150678 | FALSE | FALSE | FALSE |
| DCDC2B | 0.5095874 | 0.72513111 | 0.6901 | 1.145002644 | 18.3177335 | 18.3177335 | 34.5009223 | 0.080739683 | 0.50655 | 34.50092231 | 335.4777738 | 3341.809267 | 0.014770253 | -1.830612051 | FALSE | FALSE | FALSE |
| PRB2 | 0.5092464 | 0.72517577 | 0.97025 | 42.77039005 | 406.9280429 | 3966.318179 | 0.73249409 | 0.650101747 | 0.9375 | 6.689709595 | 601.7266727 | 601.7266727 | 0.695222559 | -0.157876144 | FALSE | TRUE | FALSE |
| FAM133B | 0.5092383 | 0.72522041 | 1 | 0.509238294 | 0.509238294 | 0.509238294 | 0.55106487 | 0.732922034 | 1 | 0.551064866 | 0.551064866 | 0.551064866 | 0.924098641 | -0.034281668 | FALSE | TRUE | FALSE |
| NANP | 0.5092016 | 0.72526504 | 1 | 0.509201599 | 0.509201599 | 0.509201599 | 0.53418107 | 0.740679756 | 1 | 5.003770402 | 5.003770402 | 5.003770402 | 0.953237819 | -0.020798736 | FALSE | TRUE | FALSE |
| EIF3G | 0.5090593 | 0.72530966 | 1 | 0.509059305 | 0.509059305 | 10.08795506 | 0.52833801 | 0.743742154 | 1 | 0.528338011 | 0.528338011 | 0.528338011 | 0.963510658 | -0.016143477 | FALSE | TRUE | FALSE |
| OTOL1 | 0.5089287 | 0.72535426 | 0.50755 | 0.508928728 | 4.555178118 | 18.21915691 | 0.23056063 | 0.844697836 | 0.6926 | 0.23056063 | 3.688144776 | 3.688144776 | 2.207353128 | 0.343871816 | FALSE | TRUE | FALSE |
| NOB1 | 0.5086839 | 0.72539885 | 0.75395 | 0.508683945 | 2.035033422 | 10.06044659 | 0.64651257 | 0.691809277 | 0.7622 | 0.646512574 | 2.586460209 | 12.8034595 | 0.786812145 | -0.104128945 | FALSE | TRUE | FALSE |
| ELF2 | 0.5085807 | 0.72544342 | 1 | 0.508580747 | 0.508580747 | 10.07421265 | 5.03443113 | 0.305360563 | 0.50095 | 5.034431129 | 5.034431129 | 10.58745665 | 0.1010205 | -0.995590488 | FALSE | FALSE | FALSE |
| GXYLT1 | 0.5085114 | 0.72548797 | 1 | 7.65727E+70 | 9.87315E+82 | 7.62159E+87 | 0.01139177 | 0.896288995 | 1 | 3.18266E+73 | 1.15318E+86 | 2.72084E+91 | 44.6384994 | 1.649709587 | FALSE | TRUE | FALSE |
| CYP27B1 | 0.5084322 | 0.72553251 | 0.5096 | 0.508432172 | 0.508432172 | 1.90005028 | 0.31990405 | 0.818778057 | 0.0139 | 0.31990405 | 0.31990405 | 1.581055483 | 1.589327088 | 0.201213285 | FALSE | TRUE | FALSE |
| BANF2 | 0.5082906 | 0.72557704 | 1 | 0.508290575 | 0.508290575 | 0.508290575 | 0.55015521 | 0.733238178 | 1 | 0.550155208 | 0.550155208 | 0.550155208 | 0.92390396 | -0.034373171 | FALSE | TRUE | FALSE |
| KRTAP5-9 | 0.5079718 | 0.72562155 | 1 | 0.507971775 | 0.507971775 | 0.507971775 | 4.98407686 | 0.306802529 | 0.4988 | 0.532951248 | 4.984076863 | 4.984076863 | 0.101918929 | -0.991745149 | FALSE | FALSE | FALSE |
| SORBS2 | 0.5079631 | 0.72566605 | 0.5236 | 0.507963059 | 0.507963059 | 9.907837086 | 0.08121763 | 0.88001787 | 1 | 0.081217633 | 0.081217633 | 1.588877064 | 6.254344544 | 0.796181802 | FALSE | TRUE | FALSE |
| NRBF2 | 0.5079251 | 0.72571053 | 1 | 4.757773178 | 4.757773178 | 4.757773178 | 0.56750751 | 0.725665142 | 1 | 0.567507511 | 0.567507511 | 0.567507511 | 0.895010326 | -0.048171954 | FALSE | TRUE | FALSE |
| HABP4 | 0.5078178 | 0.725755 | 1 | 0.507817772 | 0.507817772 | 10.0854941 | 46.6366464 | 0.059142783 | 0.25555 | 4.844474529 | 46.63664639 | 46.63664639 | 0.010888814 | -1.963019418 | FALSE | FALSE | FALSE |
| EHMT2 | 0.507752 | 0.72579945 | 0.5259 | 0.50775204 | 2.030406541 | 17.10129727 | 0.08086216 | 0.880090969 | 1 | 0.08086216 | 0.08086216 | 1.572544987 | 6.279229288 | 0.797906342 | FALSE | TRUE | FALSE |
| SAMD4B | 0.507741 | 0.72584389 | 1 | 0.507740977 | 0.507740977 | 9.981104948 | 0.55036424 | 0.733193061 | 1 | 0.550364235 | 0.550364235 | 10.84126901 | 0.922554455 | -0.03500799 | FALSE | TRUE | FALSE |
| DOLPP1 | 0.5075573 | 0.72588831 | 1 | 0.507557289 | 0.507557289 | 0.507557289 | 0.54996509 | 0.733328362 | 1 | 0.549965087 | 0.549965087 | 0.549965087 | 0.922890019 | -0.034850051 | FALSE | TRUE | FALSE |
| CLCN3 | 0.5075001 | 0.72593271 | 0.8164 | 2.841220213 | 45.40097156 | 181.554947 | 0.0519919 | 0.886237325 | 1 | 0.207828037 | 0.207828037 | 4.098636183 | 9.761137806 | 0.989500444 | FALSE | TRUE | FALSE |
| PLAC9 | 0.5073835 | 0.72597711 | 1 | 0.507383516 | 0.507383516 | 0.507383516 | 0.5491793 | 0.733463528 | 1 | 0.549179302 | 0.549179302 | 0.549179302 | 0.923894098 | -0.034377807 | FALSE | TRUE | FALSE |
| TADA2B | 0.507379 | 0.72602148 | 1 | 46.25165664 | 458.8448926 | 458.8448926 | 0.45530874 | 0.772709509 | 1 | 42618.12184 | 4915350.819 | 54612738.35 | 1.114362626 | 0.047026538 | FALSE | TRUE | FALSE |
| ZBTB16 | 0.5071786 | 0.72606584 | 1 | 2.028680009 | 2.028680009 | 17.15927732 | 0.54980091 | 0.733373431 | 1 | 2.199170821 | 2.199170821 | 18.61230641 | 0.922476797 | -0.035044549 | FALSE | TRUE | FALSE |
| RAC3 | 0.5067047 | 0.72611019 | 1 | 0.506704747 | 0.506704747 | 0.506704747 | 0.54913986 | 0.733553558 | 1 | 0.549139857 | 0.549139857 | 0.549139857 | 0.922724403 | -0.034927994 | FALSE | TRUE | FALSE |
| FBXW11 | 0.506598 | 0.72615453 | 1 | 0.506597972 | 0.506597972 | 10.01436758 | 0.54914986 | 0.733508551 | 1 | 0.549149859 | 0.549149859 | 10.87068624 | 0.922513161 | -0.035027429 | FALSE | TRUE | FALSE |
| COPS7A | 0.5065971 | 0.72619885 | 1 | 0.506597135 | 0.506597135 | 0.506597135 | 0.54876391 | 0.737733425 | 0.0088 | 0.548763913 | 0.548763913 | 0.548763913 | 0.923160438 | -0.034722815 | FALSE | TRUE | FALSE |
| SYCE2 | 0.5063725 | 0.72624315 | 1 | 0.506372472 | 0.506372472 | 0.506372472 | 0.54881627 | 0.733688483 | 1 | 0.54881627 | 0.54881627 | 0.54881627 | 0.922663011 | -0.03495689 | FALSE | TRUE | FALSE |
| ARSA | 0.5062802 | 0.72628744 | 0.2419 | 0.506280205 | 8.100289421 | 32.41290709 | 0.06543258 | 0.883471881 | 0.7733 | 0.261598826 | 16.7421777 | 67.00196732 | 7.737432938 | 0.888596898 | FALSE | TRUE | FALSE |
| RASA4 | 0.5062665 | 0.72633171 | 1 | 4.797947694 | 46.66743373 | 93.9863005 | 0.48668266 | 0.760345575 | 1 | 0.486682664 | 0.486682664 | 9.557160706 | 1.040239446 | 0.017133318 | FALSE | TRUE | FALSE |
| GABPA | 0.5059963 | 0.72637597 | 1 | 0.505996267 | 0.505996267 | 10.03455173 | 0.53614514 | 0.73995534 | 1 | 0.53614514 | 0.53614514 | 10.64322992 | 0.943767331 | -0.02513506 | FALSE | TRUE | FALSE |
| MB21D2 | 0.5058496 | 0.72642022 | 1 | 0.505849555 | 0.505849555 | 10.0091367 | 0.54816301 | 0.733957899 | 1 | 0.548163014 | 0.548163014 | 10.86029422 | 0.92280862 | -0.034888357 | FALSE | TRUE | FALSE |
| CNNM1 | 0.5058284 | 0.72646445 | 0.5172 | 0.505828433 | 2.023288212 | 9.94380257 | 0.04905841 | 0.886906391 | 0.02025 | 0.049058412 | 0.049058412 | 0.966395121 | 10.31073805 | 1.013289753 | FALSE | TRUE | FALSE |
| EIF1 | 0.5057483 | 0.72650867 | 1 | 0.505748334 | 0.505748334 | 0.505748334 | 0.51543474 | 0.749288512 | 1 | 0.515434738 | 0.515434738 | 0.515434738 | 0.981207312 | -0.008239224 | FALSE | TRUE | FALSE |
| MSANTD4 | 0.5057087 | 0.72655287 | 1 | 0.505708659 | 0.505708659 | 0.505708659 | 0.54800444 | 0.734092391 | 1 | 0.548004439 | 0.548004439 | 10.89734982 | 0.922818545 | -0.034883687 | FALSE | TRUE | FALSE |
| CDC25A | 0.5056541 | 0.72659705 | 1 | 0.505654147 | 0.505654147 | 10.02268559 | 0.54804411 | 0.734047576 | 1 | 0.548044107 | 0.548044107 | 10.87458156 | 0.922652283 | -0.034961939 | FALSE | TRUE | FALSE |
| CSNK2A1 | 0.5054856 | 0.72664122 | 0.01305 | 0.505485649 | 0.505485649 | 10.02793626 | 0.54798118 | 0.734137188 | 1 | 0.547981177 | 0.547981177 | 10.88154709 | 0.922450752 | -0.035056811 | FALSE | TRUE | FALSE |
| KRTAP13-4 | 0.5054439 | 0.72668538 | 1 | 0.505443852 | 0.505443852 | 0.505443852 | 0.54741945 | 0.734539633 | 1 | 0.54741945 | 0.54741945 | 0.54741945 | 0.923320961 | -0.034647305 | FALSE | TRUE | FALSE |
| ARL8B | 0.5054092 | 0.72672952 | 1 | 0.505409185 | 0.505409185 | 0.505409185 | 0.54780531 | 0.734316215 | 1 | 0.54780531 | 0.54780531 | 0.54780531 | 0.922607313 | -0.034983107 | FALSE | TRUE | FALSE |
| ANKRD2 | 0.5052998 | 0.72677364 | 0.00935 | 0.056460557 | 0.505299824 | 0.505299824 | 0.08106963 | 0.880072701 | 1 | 0.081069629 | 0.081069629 | 0.081069629 | 6.232911497 | 0.79469096 | FALSE | TRUE | FALSE |
| TM4SF19 | 0.5052841 | 0.72681775 | 1 | 0.505284058 | 0.505284058 | 0.505284058 | 0.5474407 | 0.734494981 | 1 | 0.547440701 | 0.547440701 | 0.547440701 | 0.922993224 | -0.034801487 | FALSE | TRUE | FALSE |
| DPPA5 | 0.5051085 | 0.72686185 | 1 | 0.505108521 | 0.505108521 | 0.505108521 | 0.54715299 | 0.734762644 | 1 | 0.547152987 | 0.547152987 | 0.547152987 | 0.923157751 | -0.034724079 | FALSE | TRUE | FALSE |
| EID2B | 0.5050127 | 0.72690593 | 1 | 0.505012669 | 0.505012669 | 0.505012669 | 0.54709677 | 0.734807197 | 1 | 0.54709677 | 0.54709677 | 0.54709677 | 0.923077409 | -0.034761878 | FALSE | TRUE | FALSE |
| UTP15 | 0.5049609 | 0.72694999 | 1 | 0.504960888 | 0.504960888 | 10.00699266 | 0.5476821 | 0.734360931 | 1 | 0.547682104 | 0.547682104 | 10.86553556 | 0.921996327 | -0.035270809 | FALSE | TRUE | FALSE |
| RELA | 0.5049369 | 0.72699404 | 1 | 0.504936862 | 0.504936862 | 9.963226811 | 0.5476248 | 0.734450314 | 1 | 0.547624795 | 0.547624795 | 10.8229777 | 0.92204894 | -0.035246027 | FALSE | TRUE | FALSE |
| GFRA4 | 0.5049331 | 0.72703808 | 1 | 0.504933088 | 4.737248185 | 4.737248185 | 0.56432818 | 0.727092877 | 1 | 0.564328179 | 0.564328179 | 0.564328179 | 0.894750797 | -0.048297906 | FALSE | TRUE | FALSE |
| SLC47A2 | 0.5049021 | 0.7270821 | 0.5033 | 0.504902128 | 0.504902128 | 1.122242152 | 0.08135674 | 0.879972141 | 0.01345 | 0.081356737 | 0.081356737 | 1.609249363 | 6.206027256 | 0.792813679 | FALSE | TRUE | FALSE |
| C3orf22 | 0.504861 | 0.7271261 | 1 | 0.504860995 | 0.504860995 | 0.504860995 | 0.54685423 | 0.734896258 | 1 | 0.546854227 | 0.546854227 | 0.546854227 | 0.923209458 | -0.034699755 | FALSE | TRUE | FALSE |

| | | | | | | | | | | | | | | | | | |
|---|---|---|---|---|---|---|---|---|---|---|---|---|---|---|---|---|---|
| LUM | 0.5048387 | 0.72717009 | 1 | 0.504838695 | 0.504838695 | 0.504838695 | 0.54730123 | 0.734718074 | 1 | 0.547301225 | 0.547301225 | 0.547301225 | 0.922414698 | -0.035073785 | FALSE | TRUE | FALSE |
| SRXN1 | 0.504834 | 0.72721406 | 1 | 0.504834016 | 0.504834016 | 0.504834016 | 0.54738284 | 0.734584268 | 1 | 0.547382838 | 0.547382838 | 0.547382838 | 0.922268622 | -0.035142567 | FALSE | TRUE | FALSE |
| RCOR3 | 0.5048182 | 0.72725802 | 1 | 0.504818173 | 0.504818173 | 9.994798144 | 0.54731257 | 0.734673488 | 1 | 0.547312573 | 0.547312573 | 10.84924908 | 0.922358078 | -0.035100444 | FALSE | TRUE | FALSE |
| PWWP2A | 0.5046478 | 0.72730196 | 1 | 0.504647796 | 0.504647796 | 9.973947921 | 2.14967938 | 0.476215052 | 0.0128 | 2.149679375 | 2.149679375 | 10.6371059 | 0.234754914 | -0.629385309 | FALSE | FALSE | FALSE |
| ZNF582 | 0.5046448 | 0.72734588 | 0.511 | 4.621315277 | 43.40768288 | 43.40768288 | 0.08359163 | 0.879457744 | 0.5117 | 0.752429059 | 0.752429059 | 1.658910721 | 6.03702501 | 0.780822975 | FALSE | TRUE | FALSE |
| PTPN11 | 0.5046447 | 0.7273898 | 0.5075 | 0.50464474 | 0.50464474 | 4.270189131 | 0.64515732 | 0.69241961 | 0.50695 | 0.645157319 | 0.645157319 | 5.46213086 | 0.782204163 | -0.106679877 | FALSE | TRUE | FALSE |
| TICAM2 | 0.5045167 | 0.72743369 | 1 | 0.504516713 | 0.504516713 | 0.504516713 | 0.50054664 | 0.754927692 | 1 | 4.7463248 | 4.7463248 | 4.7463248 | 1.007931475 | 0.003431007 | FALSE | TRUE | FALSE |
| ERI3 | 0.5043921 | 0.72747757 | 1 | 0.504392113 | 0.504392113 | 10.01405291 | 0.54678863 | 0.734985254 | 1 | 0.546788626 | 0.546788626 | 0.546788626 | 0.922462701 | -0.035051185 | FALSE | TRUE | FALSE |
| MPG | 0.5043419 | 0.72752144 | 0.5039 | 0.504341887 | 0.504341887 | 1.126322116 | 0.07660585 | 0.881150641 | 0.75145 | 0.684835794 | 6.283681723 | 6.283681723 | 6.583594649 | 0.818463084 | FALSE | TRUE | FALSE |
| ORMDL3 | 0.5040755 | 0.7275653 | 1 | 2.01606891 | 8.065359773 | 8.065359773 | 0.55906762 | 0.729065509 | 1 | 0.559067621 | 0.559067621 | 0.559067621 | 0.90163595 | -0.04496878 | FALSE | TRUE | FALSE |
| CPA1 | 0.5040172 | 0.72760913 | 0.7599 | 0.504017175 | 4.567175772 | 9.976449091 | 0.08302121 | 0.879549896 | 1 | 0.332079589 | 0.332079589 | 1.645348181 | 6.070944874 | 0.783256289 | FALSE | TRUE | FALSE |
| ZNF593 | 0.5039522 | 0.72765296 | 1 | 0.503952215 | 0.503952215 | 0.503952215 | 0.54614049 | 0.735207475 | 1 | 0.546140491 | 0.546140491 | 0.546140491 | 0.922751972 | -0.034915018 | FALSE | TRUE | FALSE |
| EHD1 | 0.5037123 | 0.72769677 | 0.89375 | 2.014522091 | 32.24604731 | 129.0599243 | 7.01897231 | 0.244729843 | 0.2572 | 1.755310104 | 28.07383472 | 28.07383472 | 0.071764395 | -1.144090974 | FALSE | FALSE | FALSE |
| CD37 | 0.5036638 | 0.72774056 | 0.75705 | 0.503663752 | 2.014610773 | 2.499614752 | 0.6494416 | 0.6899626 | 0.5057 | 0.649441601 | 0.649441601 | 0.649441601 | 0.775533552 | -0.110399408 | FALSE | TRUE | FALSE |
| VWC2L | 0.5036591 | 0.72778434 | 1 | 0.503659062 | 0.503659062 | 0.503659062 | 0.54584156 | 0.735384965 | 1 | 0.545841557 | 0.545841557 | 0.545841557 | 0.922720258 | -0.034929945 | FALSE | TRUE | FALSE |
| CTCF | 0.5034668 | 0.72782811 | 1 | 0.503466757 | 0.503466757 | 9.903833422 | 0.54627936 | 0.735118634 | 1 | 0.546279356 | 0.546279356 | 10.76727423 | 0.921628744 | -0.035443989 | FALSE | TRUE | FALSE |
| RALYL | 0.5034626 | 0.72787186 | 1 | 0.503462619 | 0.503462619 | 0.503462619 | 0.54564968 | 0.735473616 | 1 | 0.545649677 | 0.545649677 | 0.545649677 | 0.922684719 | -0.034946672 | FALSE | TRUE | FALSE |
| FEM1B | 0.5034622 | 0.72791559 | 1 | 0.503462154 | 0.503462154 | 9.934462176 | 0.54617983 | 0.735163063 | 1 | 0.546179826 | 0.546179826 | 10.79473966 | 0.921788265 | -0.035368825 | FALSE | TRUE | FALSE |
| ZBTB37 | 0.5033894 | 0.72795931 | 1 | 0.503389416 | 0.503389416 | 9.998623059 | 0.54597024 | 0.735340616 | 1 | 0.545970242 | 0.545970242 | 0.545970242 | 0.922008888 | -0.035264892 | FALSE | TRUE | FALSE |
| FANCD2OS | 0.5032943 | 0.72800302 | 1 | 4.701718841 | 4.701718841 | 4.701718841 | 0.56271242 | 0.727800854 | 1 | 0.562712419 | 0.562712419 | 0.562712419 | 0.894407718 | -0.048464462 | FALSE | TRUE | FALSE |
| SMIM14 | 0.5030766 | 0.72804671 | 1 | 0.503076627 | 0.503076627 | 0.503076627 | 0.50931088 | 0.75190978 | 1 | 0.509310881 | 0.509310881 | 0.509310881 | 0.987759434 | -0.005348814 | FALSE | TRUE | FALSE |
| AIF1L | 0.5030332 | 0.72809039 | 1 | 4.70813604 | 4.70813604 | 4.70813604 | 0.56254412 | 0.727941979 | 1 | 0.562544119 | 0.562544119 | 0.562544119 | 0.894211024 | -0.04855998 | FALSE | TRUE | FALSE |
| DEFB132 | 0.5028223 | 0.72813405 | 0.00205 | 0.502822262 | 0.502822262 | 0.502822262 | 0.54488609 | 0.735915917 | 1 | 0.544886092 | 0.544886092 | 0.544886092 | 0.922802527 | -0.034891225 | FALSE | TRUE | FALSE |
| OR6A2 | 0.5027726 | 0.7281777 | 1 | 4.725772359 | 4.725772359 | 4.725772359 | 0.56267923 | 0.727847913 | 1 | 0.562679234 | 0.562679234 | 0.562679234 | 0.893533241 | -0.048889287 | FALSE | TRUE | FALSE |
| HES1 | 0.5025503 | 0.72822133 | 0.0105 | 0.502550272 | 0.502550272 | 9.969533699 | 0.54531283 | 0.735694964 | 1 | 0.545312826 | 0.545312826 | 0.545312826 | 0.921581611 | -0.0354662 | FALSE | TRUE | FALSE |
| TCEA1 | 0.5023511 | 0.72826495 | 1 | 0.502351125 | 0.502351125 | 0.502351125 | 0.54490355 | 0.735871759 | 1 | 0.544903548 | 0.544903548 | 0.544903548 | 0.921908339 | -0.035312257 | FALSE | TRUE | FALSE |
| STX1B | 0.5023416 | 0.72830856 | 1 | 0.502341585 | 0.502341585 | 9.965462953 | 0.52009984 | 0.747374346 | 1 | 0.520099841 | 0.520099841 | 0.520099841 | 0.965856063 | -0.01508759 | FALSE | TRUE | FALSE |
| TNFSF8 | 0.5022079 | 0.72835215 | 1 | 0.502207923 | 0.502207923 | 0.502207923 | 0.54475892 | 0.736004187 | 1 | 0.544758924 | 0.544758924 | 0.544758924 | 0.921890218 | -0.035320793 | FALSE | TRUE | FALSE |
| HRH3 | 0.5021446 | 0.72839572 | 1 | 0.502144597 | 0.502144597 | 9.92736501 | 0.5449661 | 0.735827584 | 1 | 0.544966097 | 0.544966097 | 10.78885408 | 0.921423554 | -0.03554069 | FALSE | TRUE | FALSE |
| NOVA1 | 0.5021138 | 0.72843929 | 1 | 0.502113779 | 0.502113779 | 9.957004014 | 0.5448134 | 0.73596006 | 1 | 0.544813398 | 0.544813398 | 0.544813398 | 0.92162524 | -0.03544564 | FALSE | TRUE | FALSE |
| C19orf73 | 0.5020422 | 0.72848283 | 1 | 0.502042175 | 0.502042175 | 0.502042175 | 0.54417886 | 0.736400621 | 1 | 0.54417886 | 0.54417886 | 0.54417886 | 0.922568317 | -0.035001464 | FALSE | TRUE | FALSE |
| ZC3H15 | 0.5019553 | 0.72852636 | 1 | 0.501955257 | 0.501955257 | 0.501955257 | 0.5444063 | 0.736224589 | 1 | 0.544406302 | 0.544406302 | 0.544406302 | 0.92202323 | -0.035258137 | FALSE | TRUE | FALSE |
| LRRC3B | 0.5019523 | 0.72856988 | 1 | 4.718207039 | 4.718207039 | 4.718207039 | 0.54443546 | 0.736180541 | 1 | 5.123782668 | 5.123782668 | 5.123782668 | 0.92196846 | -0.035283936 | FALSE | TRUE | FALSE |
| F3 | 0.5019034 | 0.72861338 | 1 | 0.501903418 | 0.501903418 | 0.501903418 | 0.54444435 | 0.736092397 | 1 | 0.544444355 | 0.544444355 | 0.544444355 | 0.921863572 | -0.035333346 | FALSE | TRUE | FALSE |
| IGF2BP3 | 0.5018432 | 0.72865687 | 1 | 0.501843184 | 0.501843184 | 9.91561971 | 0.54456184 | 0.7360483 | 1 | 0.544561839 | 0.544561839 | 10.77531062 | 0.921554079 | -0.035479174 | FALSE | TRUE | FALSE |
| CALM2 | 0.5017727 | 0.72870034 | 1 | 0.501772723 | 0.501772723 | 0.501772723 | 0.54425249 | 0.736356636 | 1 | 0.544252488 | 0.544252488 | 0.544252488 | 0.921948424 | -0.035293374 | FALSE | TRUE | FALSE |
| ARID3B | 0.5017395 | 0.72874379 | 0.7519 | 4.727743768 | 45.72084241 | 93.41409467 | 0.54443867 | 0.736136477 | 1 | 5.134909226 | 49.70504192 | 101.6064368 | 0.921572121 | -0.035470672 | FALSE | TRUE | FALSE |
| PDLIM4 | 0.5015644 | 0.72878724 | 1 | 0.501564397 | 0.501564397 | 9.943465435 | 0.54440519 | 0.736268621 | 1 | 0.54440519 | 0.54440519 | 0.54440519 | 0.921307155 | -0.035595556 | FALSE | TRUE | FALSE |
| RELB | 0.5015185 | 0.72883066 | 1 | 0.501518537 | 0.501518537 | 9.866412012 | 0.51058262 | 0.751366812 | 1 | 4.814184445 | 46.58685955 | 94.89618126 | 0.982247578 | -0.007779033 | FALSE | TRUE | FALSE |
| FBXO32 | 0.5012235 | 0.72887408 | 1 | 0.501223483 | 0.501223483 | 0.595686291 | 0.54363091 | 0.736708076 | 1 | 0.54363091 | 0.54363091 | 0.54363091 | 0.921992245 | -0.035272732 | FALSE | TRUE | FALSE |
| MED18 | 0.5009621 | 0.72891748 | 1 | 0.500962128 | 0.500962128 | 0.500962128 | 0.54333015 | 0.736883419 | 1 | 0.543330147 | 0.543330147 | 0.543330147 | 0.922021593 | -0.035258908 | FALSE | TRUE | FALSE |
| TRAPPC4 | 0.5008606 | 0.72896087 | 0.50285 | 0.50086057 | 0.50086057 | 0.50086057 | 0.15962679 | 0.862130724 | 1 | 0.638378547 | 0.638378547 | 0.638378547 | 3.137697526 | 0.496611075 | FALSE | TRUE | FALSE |
| NFIB | 0.5007932 | 0.72900424 | 1 | 0.500793212 | 0.500793212 | 9.934071683 | 0.54365623 | 0.736664202 | 1 | 0.543656226 | 0.543656226 | 10.79492262 | 0.921157871 | -0.035665933 | FALSE | TRUE | FALSE |
| LEPROT | 0.5004262 | 0.7290476 | 1 | 0.500426214 | 0.500426214 | 0.500426214 | 2.08036242 | 0.482364769 | 0.74875 | 2.080362424 | 19.48925208 | 19.48925208 | 0.240547612 | -0.61879895 | FALSE | FALSE | FALSE |
| FAR1 | 0.5003531 | 0.72909095 | 1 | 0.500353141 | 0.500353141 | 9.910374806 | 4.95221239 | 0.307952982 | 0.5035 | 4.952212386 | 4.952212386 | 10.43433293 | 0.101036285 | -0.995522632 | FALSE | FALSE | FALSE |
| VGLL3 | 0.5001936 | 0.72913428 | 1 | 2.000747728 | 2.000747728 | 2.000747728 | 2.17066924 | 0.474133954 | 0.0075 | 0.542673709 | 2.17066924 | 2.17066924 | 0.230432905 | -0.637455506 | FALSE | FALSE | FALSE |
| TPPP3 | 0.5000936 | 0.7291776 | 1 | 0.500093618 | 0.500093618 | 0.500093618 | 0.54272809 | 0.737014771 | 1 | 0.54272809 | 0.54272809 | 0.54272809 | 0.921444139 | -0.035530988 | FALSE | TRUE | FALSE |
| UGT2B4 | 0.4999828 | 0.7292209 | 0.50165 | 0.499982818 | 4.469345179 | 4.469345179 | 0.01188419 | 0.89612026 | 1 | 0.901427279 | 8.158284955 | 8.158284955 | 42.0712539 | 1.623985456 | FALSE | TRUE | FALSE |
| F2RL2 | 0.4999613 | 0.72926419 | 0.50535 | 0.499961342 | 0.499961342 | 0.499961342 | 0.16097498 | 0.861810259 | 0.0084 | 0.160974977 | 0.160974977 | 0.160974977 | 3.105832664 | 0.492178053 | FALSE | TRUE | FALSE |
| LEP | 0.4998455 | 0.72930746 | 1 | 0.499845531 | 0.499845531 | 0.499845531 | 0.54238594 | 0.737145986 | 1 | 0.542385937 | 0.542385937 | 0.542385937 | 0.921568014 | -0.035472607 | FALSE | TRUE | FALSE |
| MTF2 | 0.4995866 | 0.72935072 | 0.7537 | 4.71922711 | 45.75246665 | 93.37753491 | 0.57795523 | 0.721483954 | 1 | 0.577955234 | 0.577955234 | 11.45045603 | 0.864403652 | -0.063283407 | FALSE | TRUE | FALSE |
| ARPC1A | 0.4995363 | 0.72939397 | 1 | 0.499536315 | 0.499536315 | 9.905856138 | 0.54242491 | 0.737102263 | 1 | 0.542424911 | 0.542424911 | 10.76733068 | 0.920931736 | -0.035772561 | FALSE | TRUE | FALSE |
| DAB2IP | 0.4995316 | 0.7294372 | 0.9918 | 466.910696 | 50409.03683 | 185301.5463 | 3.39025384 | 0.389654056 | 0.26285 | 0.847906525 | 3.390253836 | 16.54000641 | 0.147343431 | -0.831669222 | FALSE | FALSE | FALSE |
| ZNF281 | 0.4995266 | 0.72948041 | 1 | 0.499526635 | 0.499526635 | 9.859972573 | 0.52563904 | 0.744691281 | 1 | 4.970159705 | 4.970159705 | 17.77519542 | 0.950322554 | -0.022128964 | FALSE | TRUE | FALSE |
| HAPLN1 | 0.4993852 | 0.72952362 | 0.01125 | 0.499385203 | 0.499385203 | 9.914303193 | 0.54210247 | 0.737189696 | 1 | 0.542102473 | 0.542102473 | 0.542102473 | 0.921200747 | -0.035645718 | FALSE | TRUE | FALSE |
| GPR6 | 0.4991821 | 0.7295668 | 1 | 0.499182107 | 0.499182107 | 9.863900236 | 0.54197887 | 0.737277068 | 1 | 0.541978871 | 0.541978871 | 10.72502426 | 0.921036102 | -0.035723346 | FALSE | TRUE | FALSE |
| PHLDB3 | 0.4989949 | 0.72960998 | 0.81695 | 4.435962785 | 40.42484525 | 377.4149059 | 74.6137113 | 0.038342956 | 0.03995 | 8.082303207 | 74.61371128 | 705.6729419 | 0.006687711 | -2.174722508 | FALSE | FALSE | FALSE |
| CC2D2B | 0.4989897 | 0.72965314 | 0.93795 | 4.441540089 | 3626.167726 | 3626.167726 | 8.88238178 | 0.212811245 | 0.06655 | 0.982707836 | 82.29874425 | 82.29874425 | 0.056177461 | -1.250437891 | FALSE | FALSE | FALSE |
| PCOLCE2 | 0.498971 | 0.72969628 | 0.5083 | 0.498971014 | 0.498971014 | 1.118213513 | 0.08071637 | 0.880127489 | 0.0119 | 0.080716374 | 0.080716374 | 1.601619605 | 6.181781849 | 0.791113675 | FALSE | TRUE | FALSE |
| THY1 | 0.4989165 | 0.72973941 | 1 | 0.498916534 | 0.498916534 | 0.498916534 | 0.54124275 | 0.73749524 | 1 | 0.541242745 | 0.541242745 | 0.541242745 | 0.921798099 | -0.035364192 | FALSE | TRUE | FALSE |
| TMEM254 | 0.4987638 | 0.72978253 | 1 | 0.498763831 | 0.498763831 | 0.498763831 | 0.5410181 | 0.737800024 | 1 | 0.541018103 | 0.541018103 | 0.541018103 | 0.921898599 | -0.035316845 | FALSE | TRUE | FALSE |
| MAPKAP1 | 0.4986912 | 0.72982563 | 1 | 0.49869123 | 0.49869123 | 9.863518861 | 0.54154989 | 0.737408016 | 1 | 0.541549888 | 0.541549888 | 10.72546346 | 0.920859261 | -0.03580674 | FALSE | TRUE | FALSE |
| HIC2 | 0.4986405 | 0.72986871 | 1 | 0.49864046 | 0.49864046 | 9.804675907 | 0.54166141 | 0.737320733 | 1 | 0.541661415 | 0.541661415 | 10.67220601 | 0.920575928 | -0.035940386 | FALSE | TRUE | FALSE |
| C17orf67 | 0.4985049 | 0.72991179 | 0.508 | 4.651877723 | 4.651877723 | 4.651877723 | 0.55785106 | 0.729670058 | 1 | 0.557851059 | 0.557851059 | 0.557851059 | 0.893616524 | -0.048848809 | FALSE | TRUE | FALSE |
| CRHBP | 0.498439 | 0.72995484 | 0.0112 | 0.498439011 | 0.498439011 | 9.892267884 | 0.54117899 | 0.7375824 | 0.00915 | 0.541178992 | 0.541178992 | 0.541178992 | 0.921024316 | -0.035728904 | FALSE | TRUE | FALSE |
| SPPL3 | 0.4982921 | 0.72999789 | 1 | 4.705780581 | 4.705780581 | 9.884526553 | 0.55857918 | 0.729484345 | 1 | 0.558579184 | 0.558579184 | 11.09130346 | 0.892070615 | -0.049600766 | FALSE | TRUE | FALSE |
| SPATS2L | 0.498281 | 0.73004092 | 1 | 0.498280979 | 0.498280979 | 9.855394462 | 0.54102976 | 0.737756531 | 1 | 0.541029759 | 0.541029759 | 10.71515254 | 0.920986269 | -0.035746845 | FALSE | TRUE | FALSE |
| GDF5 | 0.4980944 | 0.73008393 | 1 | 0.498094356 | 0.498094356 | 9.828484027 | 0.54114136 | 0.737625956 | 1 | 0.541141359 | 0.541141359 | 10.69510226 | 0.920451464 | -0.035999107 | FALSE | TRUE | FALSE |
| SNRPB2 | 0.498052 | 0.73012693 | 1 | 0.498051976 | 0.498051976 | 0.498051976 | 0.5403289 | 0.737930418 | 0.0052 | 0.540328897 | 0.540328897 | 0.540328897 | 0.921757062 | -0.035383526 | FALSE | TRUE | FALSE |
| EI24 | 0.4980376 | 0.73016992 | 1 | 4.668420787 | 4.668420787 | 9.898421999 | 0.55784269 | 0.729716443 | 1 | 0.557842689 | 0.557842689 | 0.557842689 | 0.89279211 | -0.049249656 | FALSE | TRUE | FALSE |
| RILPL1 | 0.4980146 | 0.73021289 | 1 | 4.681091668 | 4.681091668 | 9.873935644 | 0.54074657 | 0.737843503 | 1 | 5.088968023 | 5.088968023 | 10.73234457 | 0.920975992 | -0.035751691 | FALSE | TRUE | FALSE |
| RNF138 | 0.4979458 | 0.73025585 | 0.00605 | 0.497945773 | 0.497945773 | 0.497945773 | 0.54071344 | 0.737886967 | 1 | 0.540713438 | 0.540713438 | 0.540713438 | 0.920905119 | -0.035785113 | FALSE | TRUE | FALSE |
| GRINA | 0.4978654 | 0.73029879 | 1 | 0.497865426 | 0.497865426 | 0.497865426 | 4.93177184 | 0.309671087 | 0.50375 | 4.931771843 | 4.931771843 | 4.931771843 | 0.10095062 | -0.995891009 | FALSE | FALSE | FALSE |

| | | | | | | | | | | | | | | | | | |
|---|---|---|---|---|---|---|---|---|---|---|---|---|---|---|---|---|---|
| CD47 | 0.4976369 | 0.73034172 | 0.0064 | 0.497636885 | 0.497636885 | 0.497636885 | 0.0565127 | 0.885384751 | 1 | 0.056512696 | 0.056512696 | 0.056512696 | 8.805753816 | 0.94476654 | FALSE | TRUE | FALSE |
| NRM | 0.4975433 | 0.73038463 | 1 | 0.497543319 | 0.497543319 | 0.497543319 | 0.53996272 | 0.738190798 | 1 | 0.539962722 | 0.539962722 | 0.539962722 | 0.921440128 | -0.035532879 | FALSE | TRUE | FALSE |
| GMNN | 0.4974724 | 0.73042753 | 0.00545 | 0.497472357 | 0.497472357 | 0.497472357 | 0.53979853 | 0.738407352 | 1 | 0.539798526 | 0.539798526 | 0.539798526 | 0.92158895 | -0.035462741 | FALSE | TRUE | FALSE |
| PRSS21 | 0.4974566 | 0.73047042 | 0.2511 | 0.220264573 | 1.989175534 | 1.989175534 | 0.0771499 | 0.88103375 | 1 | 0.700735379 | 2.802132738 | 2.802132738 | 6.447922655 | 0.809419819 | FALSE | TRUE | FALSE |
| PITHD1 | 0.4974547 | 0.73051329 | 1 | 0.497454747 | 0.497454747 | 0.497454747 | 0.54009475 | 0.738104067 | 1 | 0.540094755 | 0.540094755 | 0.540094755 | 0.921050877 | -0.03571638 | FALSE | TRUE | FALSE |
| NAMPT | 0.4973716 | 0.73055614 | 0.011 | 0.497371631 | 0.497371631 | 9.869228831 | 0.54030331 | 0.737973853 | 1 | 0.540303311 | 0.540303311 | 0.540303311 | 0.920541518 | -0.035956619 | FALSE | TRUE | FALSE |
| TCF15 | 0.4973632 | 0.73059898 | 1 | 18.75452739 | 181.4741763 | 181.4741763 | 0.54829128 | 0.733913036 | 1 | 5.173786049 | 50.10605988 | 50.10605988 | 0.907114995 | -0.042337654 | FALSE | TRUE | FALSE |
| SRP19 | 0.4972362 | 0.73064181 | 1 | 0.497236243 | 0.497236243 | 0.497236243 | 4.89368462 | 0.31109679 | 0.50555 | 4.893684618 | 4.893684618 | 4.893684618 | 0.101607742 | -0.993073201 | FALSE | FALSE | FALSE |
| HAVCR2 | 0.4972165 | 0.73068462 | 0.0089 | 0.497216483 | 0.497216483 | 0.497216483 | 0.53986324 | 0.738234141 | 1 | 0.539863242 | 0.539863242 | 0.539863242 | 0.921004515 | -0.035738241 | FALSE | TRUE | FALSE |
| PHYHIPL | 0.4971708 | 0.73072742 | 1 | 0.497170805 | 0.497170805 | 0.497170805 | 4.91294927 | 0.310526937 | 0.5039 | 4.912949268 | 4.912949268 | 4.912949268 | 0.101195998 | -0.994836661 | FALSE | FALSE | FALSE |
| JPH4 | 0.4971493 | 0.7307702 | 0.5046 | 4.701386625 | 4.701386625 | 9.836843231 | 4.94317609 | 0.30852682 | 0.7551 | 4.94317609 | 47.9566528 | 97.93374972 | 0.100572844 | -0.997519269 | FALSE | FALSE | FALSE |
| CBS | 0.4971417 | 0.73081297 | 0.01735 | 0.497141727 | 0.497141727 | 9.805554123 | 0.53999191 | 0.73814744 | 1 | 0.53999191 | 0.53999191 | 10.6684223 | 0.920646619 | -0.035907037 | FALSE | TRUE | FALSE |
| KLF10 | 0.4970976 | 0.73085572 | 1 | 0.497097599 | 0.497097599 | 9.851450417 | 0.53984104 | 0.738277468 | 1 | 0.539841039 | 0.539841039 | 10.7102509 | 0.920822173 | -0.035824232 | FALSE | TRUE | FALSE |
| PPME1 | 0.4970032 | 0.73089846 | 1 | 0.497003232 | 0.497003232 | 0.497003232 | 45.9987502 | 0.059920751 | 0.2521 | 4.766194371 | 45.99875015 | 45.99875015 | 0.010804712 | -1.966386818 | FALSE | FALSE | FALSE |
| LYNX1 | 0.4968183 | 0.73094118 | 0.0034 | 0.496818252 | 0.496818252 | 0.496818252 | 0.53925523 | 0.738796179 | 1 | 0.539255227 | 0.539255227 | 0.539255227 | 0.921304472 | -0.035596821 | FALSE | TRUE | FALSE |
| NAA35 | 0.4967641 | 0.73098389 | 1 | 0.496764092 | 0.496764092 | 9.804326643 | 0.53982365 | 0.738364073 | 1 | 0.539823654 | 0.539823654 | 10.67106541 | 0.920234022 | -0.036101715 | FALSE | TRUE | FALSE |
| CHST9 | 0.4967475 | 0.73102659 | 0.51025 | 0.496747455 | 0.496747455 | 1.088615515 | 0.67882969 | 0.675666248 | 0.2583 | 0.678829692 | 6.338646223 | 6.338646223 | 0.731770371 | -0.135625179 | FALSE | TRUE | FALSE |
| FAXC | 0.4967239 | 0.73106927 | 1 | 0.496723861 | 0.496723861 | 9.851607868 | 0.53942362 | 0.738580319 | 1 | 0.539423619 | 0.539423619 | 10.70921208 | 0.920841883 | -0.035814936 | FALSE | TRUE | FALSE |
| ZFP36L1 | 0.4966844 | 0.73111194 | 1 | 4.660607243 | 4.660607243 | 9.839476987 | 0.556307 | 0.730317957 | 1 | 0.556307003 | 0.556307003 | 11.03319109 | 0.892824328 | -0.049233984 | FALSE | TRUE | FALSE |
| CNOT8 | 0.4966721 | 0.73115459 | 1 | 0.496672099 | 0.496672099 | 0.496672099 | 0.53942468 | 0.7385371 | 1 | 0.539424681 | 0.539424681 | 0.539424681 | 0.920744114 | -0.035861049 | FALSE | TRUE | FALSE |
| PPM1B | 0.4965756 | 0.73119723 | 1 | 0.496575649 | 0.496575649 | 9.857022505 | 0.52261729 | 0.745755478 | 1 | 4.905733458 | 4.905733458 | 4.905733458 | 0.95017072 | -0.022198357 | FALSE | TRUE | FALSE |
| OSTF1 | 0.4965251 | 0.73123985 | 1 | 0.49652513 | 0.49652513 | 0.49652513 | 0.53911129 | 0.738925507 | 1 | 0.539111287 | 0.539111287 | 0.539111287 | 0.921006742 | -0.035737191 | FALSE | TRUE | FALSE |
| ARTN | 0.4964844 | 0.73128246 | 1 | 0.496484351 | 0.496484351 | 0.496484351 | 0.53932322 | 0.738623522 | 1 | 0.539323218 | 0.539323218 | 0.539323218 | 0.920569213 | -0.035943553 | FALSE | TRUE | FALSE |
| ARL6IP5 | 0.4964165 | 0.73132505 | 1 | 0.496416487 | 0.496416487 | 0.496416487 | 0.53925807 | 0.738753038 | 1 | 0.539258073 | 0.539258073 | 0.539258073 | 0.920554576 | -0.035950459 | FALSE | TRUE | FALSE |
| CELF1 | 0.4963124 | 0.73136763 | 1 | 0.496312351 | 0.496312351 | 9.834317938 | 0.53921528 | 0.738839304 | 1 | 0.539215275 | 0.539215275 | 10.69633139 | 0.920434517 | -0.036007103 | FALSE | TRUE | FALSE |
| ADRA2C | 0.4962484 | 0.7314102 | 1 | 0.496248419 | 0.496248419 | 9.817212852 | 0.53927823 | 0.738709882 | 1 | 0.539278234 | 0.539278234 | 10.68239899 | 0.920208508 | -0.036113756 | FALSE | TRUE | FALSE |
| NMT1 | 0.4962207 | 0.73145275 | 1 | 0.496220728 | 0.496220728 | 9.813486645 | 0.53929384 | 0.73866671 | 1 | 0.539293845 | 0.539293845 | 10.67966176 | 0.920130525 | -0.036150561 | FALSE | TRUE | FALSE |
| TVP23B | 0.4962026 | 0.73149529 | 1 | 0.496202615 | 0.496202615 | 0.496202615 | 0.52206439 | 0.746162294 | 0.50935 | 4.889296799 | 4.889296799 | 4.889296799 | 0.950462485 | -0.02206502 | FALSE | TRUE | FALSE |
| TAT | 0.4961364 | 0.73153781 | 0.5032 | 0.496136383 | 1.983914827 | 2.459981335 | 0.04786239 | 0.887160549 | 0.50705 | 0.191312927 | 0.191312927 | 0.949737406 | 10.36589275 | 1.015606711 | FALSE | TRUE | FALSE |
| CCL7 | 0.4957756 | 0.73158032 | 1 | 0.495775603 | 0.495775603 | 0.495775603 | 0.53801915 | 0.739269731 | 1 | 0.538019149 | 0.538019149 | 0.538019149 | 0.921483192 | -0.035512582 | FALSE | TRUE | FALSE |
| STAT5B | 0.4955321 | 0.73162282 | 0.51545 | 0.495532072 | 0.495532072 | 9.73093905 | 0.15872303 | 0.862390792 | 1 | 0.634670349 | 0.634670349 | 12.4904571 | 3.121992347 | 0.494431834 | FALSE | TRUE | FALSE |
| PBX1 | 0.4954231 | 0.7316653 | 1 | 0.495423089 | 0.495423089 | 9.82392511 | 0.53851028 | 0.739097733 | 1 | 0.538510279 | 0.538510279 | 10.6892737 | 0.919988176 | -0.036217754 | FALSE | TRUE | FALSE |
| KLF14 | 0.4953421 | 0.73170777 | 1 | 0.495342089 | 0.495342089 | 0.495342089 | 0.53821358 | 0.739183762 | 1 | 0.538213583 | 0.538213583 | 0.538213583 | 0.920344831 | -0.036049422 | FALSE | TRUE | FALSE |
| TAC3 | 0.4952857 | 0.73175023 | 0.0033 | 0.495285665 | 0.495285665 | 0.495285665 | 0.53789719 | 0.739355639 | 1 | 0.537897186 | 0.537897186 | 0.537897186 | 0.92078129 | -0.035843514 | FALSE | TRUE | FALSE |
| MAP1LC3B | 0.4952688 | 0.73179267 | 1 | 1.980921574 | 1.980921574 | 1.980921574 | 0.53799125 | 0.739312692 | 1 | 2.151817941 | 2.151817941 | 2.151817941 | 0.92058868 | -0.035934275 | FALSE | TRUE | FALSE |
| RAB5C | 0.4951812 | 0.73183509 | 1 | 0.495181199 | 0.495181199 | 0.495181199 | 4.94138097 | 0.309099509 | 0.0067 | 4.941380966 | 4.941380966 | 4.941380966 | 0.100211095 | -0.999084191 | FALSE | FALSE | FALSE |
| GTF2E1 | 0.4948143 | 0.73187751 | 1 | 0.494814323 | 0.494814323 | 9.803533539 | 0.53170388 | 0.742073727 | 1 | 2.126771472 | 2.126771472 | 10.54628534 | 0.930620116 | -0.031227564 | FALSE | TRUE | FALSE |
| CD96 | 0.4947825 | 0.73191991 | 0.504 | 0.494782454 | 0.494782454 | 1.102580828 | 0.07757378 | 0.880925663 | 1 | 0.694481158 | 0.694481158 | 1.537832747 | 6.378217307 | 0.804699312 | FALSE | TRUE | FALSE |
| VAMP3 | 0.4946713 | 0.7319623 | 1 | 0.494671319 | 0.494671319 | 0.494671319 | 4.87689544 | 0.311665662 | 0.4921 | 0.520667862 | 4.87689544 | 4.87689544 | 0.101431602 | -0.993826714 | FALSE | FALSE | FALSE |
| EZH2 | 0.4946148 | 0.73200467 | 0.516 | 0.494614768 | 0.494614768 | 9.740942804 | 3.16336715 | 0.402659276 | 0.0054 | 0.160330676 | 0.160330676 | 3.163367148 | 0.156357054 | -0.805882521 | FALSE | FALSE | FALSE |
| OMD | 0.4944389 | 0.73204703 | 0.87895 | 1.977834149 | 7.913627648 | 7.913627648 | 0.16322169 | 0.86128499 | 0.0092 | 0.163221685 | 0.163221685 | 0.163221685 | 3.029247828 | 0.481334805 | FALSE | TRUE | FALSE |
| SNX30 | 0.4941779 | 0.73208937 | 1 | 0.494177914 | 0.494177914 | 9.802342066 | 0.53707559 | 0.739698671 | 1 | 0.537075588 | 0.537075588 | 10.66377345 | 0.920127305 | -0.036152081 | FALSE | TRUE | FALSE |
| ADRB2 | 0.4940666 | 0.73213171 | 0.7514 | 4.662452 | 45.15722224 | 45.15722224 | 4.75357297 | 0.317305918 | 0.9357 | 45.99091251 | 4642.049808 | 4642.049808 | 0.103935845 | -0.98323465 | FALSE | FALSE | FALSE |
| PSENEN | 0.494034 | 0.73217402 | 1 | 0.49403397 | 0.49403397 | 0.49403397 | 0.53712472 | 0.739655844 | 1 | 0.537124716 | 0.537124716 | 0.537124716 | 0.919775158 | -0.036318324 | FALSE | TRUE | FALSE |
| SIGLECL1 | 0.4939454 | 0.73221633 | 1 | 0.493945424 | 0.493945424 | 0.493945424 | 4.87084232 | 0.313365583 | 0.50435 | 4.870842318 | 4.870842318 | 4.870842318 | 0.101408625 | -0.993925104 | FALSE | FALSE | FALSE |
| MIEN1 | 0.4938113 | 0.73225862 | 1 | 4.61263351 | 4.61263351 | 4.61263351 | 0.55328977 | 0.73196887 | 0.0028 | 0.553289775 | 0.553289775 | 0.553289775 | 0.892500234 | -0.049391661 | FALSE | TRUE | FALSE |
| AHRR | 0.4936758 | 0.7323009 | 0.2681 | 0.493675846 | 4.578250542 | 9.697644183 | 0.07897505 | 0.880645622 | 1 | 0.718280039 | 0.718280039 | 2.673873026 | 6.25103588 | 0.795951992 | FALSE | TRUE | FALSE |
| ISG20L2 | 0.4936711 | 0.73234316 | 1 | 0.493671108 | 0.493671108 | 0.493671108 | 0.53649362 | 0.73982707 | 1 | 0.536493623 | 0.536493623 | 0.536493623 | 0.920180758 | -0.036126853 | FALSE | TRUE | FALSE |
| SLURP1 | 0.4932693 | 0.73238541 | 1 | 0.493269277 | 0.493269277 | 0.493269277 | 0.5192618 | 0.747695495 | 1 | 0.519261804 | 4.859910082 | 4.859910082 | 0.949943309 | -0.022302312 | FALSE | TRUE | FALSE |
| PSAPL1 | 0.4930946 | 0.73242765 | 0.87875 | 4.548651359 | 43.03814102 | 43.03814102 | 59.4108735 | 0.047067277 | 0.073 | 6.245465259 | 579.3079265 | 579.3079265 | 0.008299736 | -2.080935698 | FALSE | FALSE | FALSE |
| PDGFB | 0.4929254 | 0.73246987 | 1 | 0.492925371 | 0.492925371 | 0.492925371 | 0.53589737 | 0.739998069 | 1 | 0.53589737 | 0.53589737 | 0.53589737 | 0.919813008 | -0.036300453 | FALSE | TRUE | FALSE |
| OR10G7 | 0.4926536 | 0.73251208 | 1 | 0.492653578 | 0.492653578 | 0.492653578 | 4.72492969 | 0.319261328 | 0.74775 | 4.724929693 | 45.55519618 | 45.55519618 | 0.104266859 | -0.981853709 | FALSE | FALSE | FALSE |
| SAMD5 | 0.4926206 | 0.73255428 | 1 | 0.492620592 | 0.492620592 | 0.492620592 | 0.53520362 | 0.740126177 | 1 | 0.535203625 | 0.535203625 | 0.535203625 | 0.920435828 | -0.036006485 | FALSE | TRUE | FALSE |
| CDCA4 | 0.4925872 | 0.73259647 | 1 | 0.492587173 | 0.492587173 | 0.492587173 | 0.53527419 | 0.740083489 | 0.0087 | 0.535274195 | 0.535274195 | 0.535274195 | 0.920252046 | -0.036093208 | FALSE | TRUE | FALSE |
| CHCHD7 | 0.4925114 | 0.73263863 | 1 | 0.492511397 | 0.492511397 | 0.492511397 | 0.53474771 | 0.74038199 | 1 | 0.534747709 | 0.534747709 | 0.534747709 | 0.921016377 | -0.035732647 | FALSE | TRUE | FALSE |
| SCGB1D4 | 0.4924076 | 0.73268079 | 1 | 0.492407645 | 0.492407645 | 0.492407645 | 0.5347969 | 0.740339392 | 0.00235 | 0.534796903 | 0.534796903 | 0.534796903 | 0.920737653 | -0.035864096 | FALSE | TRUE | FALSE |
| MEF2B | 0.4922239 | 0.73272293 | 1 | 0.492223902 | 0.492223902 | 0.492223902 | 0.53229033 | 0.741779375 | 1 | 0.532290332 | 0.532290332 | 0.532290332 | 0.924728241 | -0.033985879 | FALSE | TRUE | FALSE |
| NSFL1C | 0.4921921 | 0.73276506 | 1 | 0.492192102 | 0.492192102 | 0.492192102 | 0.5351979 | 0.740168849 | 1 | 0.535197896 | 0.535197896 | 0.535197896 | 0.919645061 | -0.036379757 | FALSE | TRUE | FALSE |
| PRG3 | 0.4920602 | 0.73280718 | 1 | 0.492060153 | 0.492060153 | 0.492060153 | 0.53485441 | 0.740296779 | 1 | 0.53485441 | 0.53485441 | 0.53485441 | 0.919988961 | -0.036217384 | FALSE | TRUE | FALSE |
| CRYBB3 | 0.4919506 | 0.73284928 | 0.6932 | 0.491950588 | 7.872370329 | 7.872370329 | 0.74723214 | 0.642815158 | 0.1328 | 0.747232145 | 2.988980899 | 2.988980899 | 0.658363792 | -0.181534062 | FALSE | TRUE | FALSE |
| HS3ST3B1 | 0.4917807 | 0.73289136 | 0.5108 | 1.96695406 | 1.96695406 | 9.706528534 | 0.52893177 | 0.743534856 | 1 | 2.115558952 | 8.463678685 | 41.82181976 | 0.929762072 | -0.031628174 | FALSE | TRUE | FALSE |
| ZMAT4 | 0.4917253 | 0.73293344 | 1 | 0.491725324 | 0.491725324 | 0.491725324 | 0.5344128 | 0.740594753 | 1 | 0.534412798 | 0.534412798 | 0.534412798 | 0.920122657 | -0.036154275 | FALSE | TRUE | FALSE |
| LRRC4C | 0.4916593 | 0.7329755 | 1 | 0.491659258 | 0.491659258 | 9.708572381 | 0.53487917 | 0.740254151 | 1 | 0.534879167 | 0.534879167 | 10.57812206 | 0.919196875 | -0.036591461 | FALSE | TRUE | FALSE |
| LTC4S | 0.4915653 | 0.73301754 | 1 | 0.491565283 | 1.966117539 | 1.966117539 | 32.3487785 | 0.085879585 | 0.34235 | 32.34877855 | 507.6707547 | 2033.052625 | 0.015195791 | -1.818276684 | FALSE | FALSE | FALSE |
| IQCF3 | 0.4914665 | 0.73305958 | 1 | 0.491466471 | 0.491466471 | 0.491466471 | 0.5344181 | 0.74055223 | 1 | 0.534418096 | 0.534418096 | 0.534418096 | 0.919629173 | -0.03638726 | FALSE | TRUE | FALSE |
| LSAMP | 0.4914602 | 0.73310159 | 1 | 4.65488788 | 45.24936214 | 45.24936214 | 0.53472913 | 0.740424573 | 1 | 5.066463194 | 49.26721426 | 49.26721426 | 0.919082513 | -0.036645497 | FALSE | TRUE | FALSE |
| RBM39 | 0.4914153 | 0.7331436 | 1 | 0.49141532 | 0.49141532 | 9.721082632 | 0.5346461 | 0.740509692 | 1 | 0.534646099 | 0.534646099 | 10.59014645 | 0.919141318 | -0.036617711 | FALSE | TRUE | FALSE |
| KRTAP12-4 | 0.4911626 | 0.73318559 | 0.50995 | 4.579462781 | 4.579462781 | 4.579462781 | 0.55056347 | 0.733102783 | 1 | 0.550563474 | 0.550563474 | 0.550563474 | 0.89210892 | -0.049582118 | FALSE | TRUE | FALSE |
| NANOS2 | 0.4910799 | 0.73322756 | 1 | 0.491079862 | 0.491079862 | 0.491079862 | 0.53367645 | 0.741061523 | 0.00525 | 0.533676448 | 0.533676448 | 0.533676448 | 0.920182751 | -0.036125912 | FALSE | TRUE | FALSE |
| TTL | 0.4910173 | 0.73326953 | 1 | 0.491017251 | 0.491017251 | 9.747163723 | 0.53378279 | 0.740976791 | 0.0104 | 0.533782789 | 0.533782789 | 10.60559609 | 0.919882133 | -0.036267816 | FALSE | TRUE | FALSE |
| SEC11A | 0.4908248 | 0.73331148 | 1 | 0.49082484 | 0.49082484 | 0.49082484 | 0.53365498 | 0.741146195 | 1 | 0.533654983 | 0.533654983 | 0.533654983 | 0.919741886 | -0.036334035 | FALSE | TRUE | FALSE |
| SSBP3 | 0.4908021 | 0.73335341 | 1 | 0.490802106 | 0.490802106 | 0.583345156 | 0.51299171 | 0.750430196 | 1 | 0.512991715 | 0.512991715 | 0.512991715 | 0.956744704 | -0.019203933 | FALSE | TRUE | FALSE |
| TRIB2 | 0.4907327 | 0.73339533 | 1 | 0.490732744 | 0.490732744 | 9.724969717 | 0.5340449 | 0.740807147 | 1 | 0.534044897 | 0.534044897 | 0.534044897 | 0.918897918 | -0.036732732 | FALSE | TRUE | FALSE |

| | | | | | | | | | | | | | | | | | |
|---|---|---|---|---|---|---|---|---|---|---|---|---|---|---|---|---|---|
| RAD51 | 0.4905076 | 0.73343724 | 1 | 4.607788648 | 4.607788648 | 4.607788648 | 0.55054659 | 0.733147929 | 0.0079 | 0.550546594 | 0.550546594 | 0.550546594 | 0.890946655 | -0.050148299 | FALSE | TRUE | FALSE |
| PLEKHH3 | 0.4904843 | 0.73347914 | 0.75445 | 0.49048429 | 4.615738667 | 9.628809319 | 0.08022509 | 0.880246034 | 1 | 0.080225086 | 0.080225086 | 1.57841367 | 6.113851828 | 0.786314909 | FALSE | TRUE | FALSE |
| GAS1 | 0.4903495 | 0.73352102 | 1 | 0.490349482 | 0.490349482 | 0.490349482 | 0.43778885 | 0.778148903 | 1 | 395.9334224 | 41941.75601 | 447312.7392 | 1.12005932 | 0.049241024 | FALSE | TRUE | FALSE |
| LHFPL4 | 0.4902914 | 0.73356289 | 1 | 0.490291374 | 0.490291374 | 0.490291374 | 0.53341555 | 0.74118851 | 1 | 0.533415552 | 0.533415552 | 0.533415552 | 0.91915463 | -0.036611421 | FALSE | TRUE | FALSE |
| SNAP91 | 0.4902518 | 0.73360474 | 1 | 0.490251796 | 0.490251796 | 9.700950866 | 0.53340702 | 0.741230809 | 1 | 0.533407019 | 0.533407019 | 10.56837187 | 0.919095135 | -0.036639533 | FALSE | TRUE | FALSE |
| INHBB | 0.4902175 | 0.73364658 | 1 | 0.49021753 | 0.49021753 | 9.698321185 | 18.0961548 | 0.143203897 | 0.0001 | 0.53356412 | 0.53356412 | 10.5696107 | 0.027089596 | -1.567197465 | FALSE | FALSE | FALSE |
| HSF5 | 0.4901962 | 0.7336884 | 1 | 4.628949255 | 44.86199263 | 91.37332756 | 0.56833773 | 0.725329753 | 0.0144 | 0.568337731 | 0.568337731 | 11.23630602 | 0.862508711 | -0.06423651 | FALSE | TRUE | FALSE |
| SGTB | 0.4901099 | 0.73373021 | 1 | 0.490109867 | 0.490109867 | 0.490109867 | 0.53317857 | 0.741399856 | 1 | 0.533178571 | 0.533178571 | 0.533178571 | 0.919222741 | -0.03657924 | FALSE | TRUE | FALSE |
| IKZF2 | 0.4900226 | 0.73377201 | 1 | 0.490022611 | 0.490022611 | 9.705550413 | 4.87680479 | 0.31194975 | 0.01295 | 4.876804789 | 4.876804789 | 10.24520465 | 0.10048026 | -0.997919252 | FALSE | FALSE | FALSE |
| DHX57 | 0.489772 | 0.7338138 | 0.33415 | 0.489772046 | 42.26277154 | 87.56057769 | 0.01194234 | 0.896085042 | 1 | 0.105262866 | 0.952289216 | 3.541485144 | 41.01140831 | 1.612904683 | FALSE | TRUE | FALSE |
| NEK7 | 0.4896071 | 0.73385557 | 1 | 0.489607109 | 0.489607109 | 0.489607109 | 0.53256741 | 0.741526495 | 0.0079 | 0.532567409 | 0.532567409 | 0.532567409 | 0.919333592 | -0.036526871 | FALSE | TRUE | FALSE |
| IL17F | 0.4895936 | 0.73389733 | 1 | 44.20237504 | 436.4102439 | 436.4102439 | 0.53247621 | 0.741610848 | 1 | 48.21548014 | 476.729677 | 476.729677 | 0.919465593 | -0.036464518 | FALSE | TRUE | FALSE |
| DCP2 | 0.4895182 | 0.73393907 | 1 | 0.489518176 | 0.489518176 | 9.715422395 | 0.53255438 | 0.741568679 | 1 | 0.532554383 | 0.532554383 | 0.532554383 | 0.919189085 | -0.036595141 | FALSE | TRUE | FALSE |
| SS18L1 | 0.489086 | 0.7339808 | 1 | 0.489085993 | 0.489085993 | 9.681944761 | 0.53223747 | 0.741821469 | 1 | 0.532237475 | 0.532237475 | 10.54909921 | 0.918924382 | -0.036720225 | FALSE | TRUE | FALSE |
| PDE6D | 0.488903 | 0.73402252 | 1 | 0.488902951 | 0.488902951 | 0.488902951 | 0.53194422 | 0.741989697 | 1 | 0.531944223 | 0.531944223 | 0.531944223 | 0.91908687 | -0.036643438 | FALSE | TRUE | FALSE |
| KRTAP5-6 | 0.4887082 | 0.73406423 | 0.4985 | 0.488708173 | 4.556605155 | 4.556605155 | 0.54813768 | 0.734002746 | 0.002 | 0.54813768 | 0.54813768 | 0.54813768 | 0.891579234 | -0.049840055 | FALSE | TRUE | FALSE |
| FSTL1 | 0.4883976 | 0.73410592 | 1 | 0.48839759 | 0.48839759 | 0.48839759 | 0.53141462 | 0.742115722 | 1 | 0.531414624 | 0.531414624 | 0.531414624 | 0.919051843 | -0.03665999 | FALSE | TRUE | FALSE |
| ITM2B | 0.4881825 | 0.73414761 | 1 | 0.488182508 | 0.488182508 | 0.488182508 | 0.53132809 | 0.742157702 | 1 | 0.531328094 | 0.531328094 | 0.531328094 | 0.918796716 | -0.036780566 | FALSE | TRUE | FALSE |
| KCTD11 | 0.4880736 | 0.73418928 | 1 | 0.488073584 | 0.488073584 | 0.488073584 | 0.53125035 | 0.742199667 | 1 | 0.531250345 | 0.531250345 | 0.531250345 | 0.91872615 | -0.036813922 | FALSE | TRUE | FALSE |
| MZB1 | 0.4880702 | 0.73423093 | 1 | 0.48807023 | 0.48807023 | 0.48807023 | 0.53064876 | 0.742618513 | 1 | 0.530648761 | 0.530648761 | 0.530648761 | 0.919761368 | -0.036324836 | FALSE | TRUE | FALSE |
| EVA1A | 0.4880557 | 0.73427257 | 1 | 0.48805569 | 0.48805569 | 0.48805569 | 0.5311559 | 0.742283553 | 1 | 0.531155896 | 0.531155896 | 0.531155896 | 0.918855827 | -0.036752626 | FALSE | TRUE | FALSE |
| PPP2R1A | 0.487984 | 0.7343142 | 0.01735 | 0.48798403 | 0.48798403 | 9.627903394 | 0.53124528 | 0.742241617 | 1 | 0.531245277 | 0.531245277 | 10.49847667 | 0.918566342 | -0.036889472 | FALSE | TRUE | FALSE |
| HS2ST1 | 0.4879254 | 0.73435581 | 1 | 1.951530235 | 1.951530235 | 9.689547298 | 0.53726595 | 0.739613001 | 1 | 0.537265954 | 0.537265954 | 0.537265954 | 0.908163672 | -0.041835875 | FALSE | TRUE | FALSE |
| DDIT4 | 0.4878638 | 0.73439741 | 1 | 0.487863841 | 0.487863841 | 0.487863841 | 0.53086509 | 0.742493013 | 1 | 0.53086509 | 0.53086509 | 0.53086509 | 0.918997785 | -0.036685536 | FALSE | TRUE | FALSE |
| PRPF18 | 0.4878588 | 0.734439 | 0.0082 | 0.487858762 | 0.487858762 | 0.487858762 | 0.53095111 | 0.742409273 | 1 | 0.530951106 | 0.530951106 | 0.530951106 | 0.918839338 | -0.03676042 | FALSE | TRUE | FALSE |
| RNASE12 | 0.4877508 | 0.73448057 | 1 | 0.487750806 | 0.487750806 | 0.487750806 | 0.53070035 | 0.742534861 | 1 | 0.530700347 | 0.530700347 | 0.530700347 | 0.919070072 | -0.036651376 | FALSE | TRUE | FALSE |
| OR10R2 | 0.4876755 | 0.73452213 | 1 | 0.48767551 | 0.48767551 | 0.48767551 | 0.53057861 | 0.742660318 | 1 | 0.530578609 | 0.530578609 | 0.530578609 | 0.919139033 | -0.03661879 | FALSE | TRUE | FALSE |
| RNF121 | 0.4875764 | 0.73456367 | 1 | 0.487576368 | 0.487576368 | 0.487576368 | 0.53091927 | 0.74245115 | 1 | 0.53091927 | 0.53091927 | 0.53091927 | 0.918362538 | -0.036985841 | FALSE | TRUE | FALSE |
| URB2 | 0.4874904 | 0.7346052 | 0.26875 | 0.056276998 | 0.487490441 | 9.465460254 | 0.10308533 | 0.875164064 | 0.03735 | 0.103085326 | 0.103085326 | 2.008615934 | 4.728999355 | 0.674769255 | FALSE | TRUE | FALSE |
| PAX7 | 0.4873959 | 0.73464672 | 0.9996 | 31985.57339 | 4775438.265 | 19135966.76 | 295.291202 | 0.008428737 | 0.21935 | 73.80198599 | 4730.794663 | 23343.21581 | 0.00165056 | -2.782368623 | FALSE | FALSE | FALSE |
| SLC39A10 | 0.4872667 | 0.73468822 | 1 | 0.487266685 | 0.487266685 | 9.601576366 | 0.53066764 | 0.742576695 | 1 | 0.530667643 | 0.530667643 | 10.47535652 | 0.918214425 | -0.037055889 | FALSE | TRUE | FALSE |
| HYAL1 | 0.4871774 | 0.73472971 | 0.75165 | 0.487177428 | 1.949009172 | 4.182946534 | 0.15907548 | 0.862284513 | 1 | 0.15907548 | 0.15907548 | 3.151447711 | 3.06255513 | 0.486083915 | FALSE | TRUE | FALSE |
| ATG4B | 0.4869728 | 0.73477119 | 1 | 0.486972823 | 0.486972823 | 9.657236088 | 0.53034067 | 0.742827387 | 1 | 0.530340667 | 0.530340667 | 10.52794882 | 0.91822644 | -0.037050206 | FALSE | TRUE | FALSE |
| KRTAP15-1 | 0.4869092 | 0.73481265 | 1 | 0.486909207 | 0.486909207 | 0.486909207 | 0.52954782 | 0.743244043 | 1 | 0.529547818 | 0.529547818 | 0.529547818 | 0.919481093 | -0.036457196 | FALSE | TRUE | FALSE |
| SYT11 | 0.4867346 | 0.7348541 | 1 | 0.486734636 | 0.486734636 | 9.645701746 | 0.53021691 | 0.742952535 | 1 | 0.530216914 | 0.530216914 | 10.51895047 | 0.91799153 | -0.037161326 | FALSE | TRUE | FALSE |
| CSNK1D | 0.4867157 | 0.73489554 | 1 | 0.486715675 | 0.486715675 | 9.643294784 | 0.53000166 | 0.743035896 | 1 | 0.530001661 | 0.530001661 | 10.51272963 | 0.918328584 | -0.037001897 | FALSE | TRUE | FALSE |
| BCAN | 0.4866702 | 0.73493697 | 0.7562 | 0.486670248 | 4.483347494 | 16.41962129 | 0.08070155 | 0.880136615 | 1 | 0.08070155 | 0.08070155 | 1.580169894 | 6.030494416 | 0.78035292 | FALSE | TRUE | FALSE |
| DDX19A | 0.4866656 | 0.73497837 | 1 | 0.486665603 | 0.486665603 | 9.634507692 | 0.53000932 | 0.742994223 | 1 | 0.530009317 | 0.530009317 | 10.50539463 | 0.918220846 | -0.037052852 | FALSE | TRUE | FALSE |
| MAT2B | 0.4865854 | 0.73501977 | 1 | 0.48658536 | 4.585236624 | 4.585236624 | 0.54684038 | 0.734940764 | 1 | 0.54684038 | 0.54684038 | 0.54684038 | 0.889812416 | -0.050701538 | FALSE | TRUE | FALSE |
| SAMD10 | 0.4865295 | 0.73506115 | 1 | 0.48652951 | 0.48652951 | 0.48652951 | 0.52977809 | 0.743202441 | 1 | 0.529778089 | 0.529778089 | 0.529778089 | 0.918364726 | -0.036984806 | FALSE | TRUE | FALSE |
| PON1 | 0.4864733 | 0.73510252 | 0.50665 | 4.394553186 | 40.71895842 | 40.71895842 | 0.0791323 | 0.88058222 | 1 | 0.703060298 | 6.411159936 | 6.411159936 | 6.147594208 | 0.788705193 | FALSE | TRUE | FALSE |
| ZBTB20 | 0.4864656 | 0.73514387 | 0.0231 | 0.48646558 | 0.48646558 | 9.540401552 | 0.5299486 | 0.743077554 | 1 | 0.529948599 | 0.529948599 | 10.41747795 | 0.917948611 | -0.037181631 | FALSE | TRUE | FALSE |
| IP6K1 | 0.4861782 | 0.73518521 | 1 | 0.486178157 | 0.486178157 | 9.606090584 | 0.52943255 | 0.743327202 | 1 | 0.529432551 | 0.529432551 | 10.47593288 | 0.918300464 | -0.037015196 | FALSE | TRUE | FALSE |
| NDNF | 0.4859038 | 0.73522654 | 1 | 0.485903826 | 0.485903826 | 9.620142656 | 0.52917623 | 0.743368762 | 1 | 0.529176231 | 0.529176231 | 10.48956952 | 0.918226855 | -0.03705001 | FALSE | TRUE | FALSE |
| AATF | 0.4857412 | 0.73526786 | 0.9905 | 88.26366836 | 40613.95667 | 79370.97344 | 0.09381666 | 0.877378602 | 1 | 0.093816661 | 0.093816661 | 1.85892913 | 5.177557765 | 0.714124953 | FALSE | TRUE | FALSE |
| DISP2 | 0.4856691 | 0.73530916 | 0.63975 | 0.847773654 | 9.409299581 | 37.61983673 | 1.93916137 | 0.494753753 | 0.26835 | 0.205597314 | 1.939161368 | 37.71093564 | 0.250453169 | -0.601273468 | FALSE | FALSE | FALSE |
| OR13C2 | 0.485606 | 0.73535045 | 1 | 0.485605999 | 0.485605999 | 0.485605999 | 0.52844043 | 0.743700722 | 1 | 0.528440434 | 0.528440434 | 0.528440434 | 0.918941791 | -0.036711997 | FALSE | TRUE | FALSE |
| HOXD11 | 0.4855631 | 0.73539173 | 0.74675 | 0.485563125 | 4.521578557 | 4.521578557 | 0.16244072 | 0.861476429 | 1 | 0.162440723 | 0.162440723 | 0.162440723 | 2.989171162 | 0.475550784 | FALSE | TRUE | FALSE |
| NFIX | 0.485519 | 0.73543299 | 1 | 0.485518963 | 0.485518963 | 9.592878675 | 0.5289191 | 0.743617814 | 1 | 0.528919101 | 0.528919101 | 10.46559315 | 0.917945602 | -0.037183054 | FALSE | TRUE | FALSE |
| CADM3 | 0.4854658 | 0.73547424 | 1 | 0.485465772 | 0.485465772 | 9.622475839 | 0.52895878 | 0.743493354 | 1 | 0.528958781 | 0.528958781 | 0.528958781 | 0.917776186 | -0.037263215 | FALSE | TRUE | FALSE |
| ING4 | 0.4852671 | 0.73551548 | 1 | 0.485267062 | 0.485267062 | 0.485267062 | 0.5284888 | 0.743659275 | 1 | 0.528488804 | 0.528488804 | 0.528488804 | 0.918216353 | -0.037054978 | FALSE | TRUE | FALSE |
| IGFBP6 | 0.4852147 | 0.7355567 | 1 | 4.540209967 | 4.540209967 | 4.540209967 | 4.95076095 | 0.308239996 | 0.50345 | 4.950760952 | 4.950760952 | 4.950760952 | 0.098008099 | -1.008738035 | FALSE | FALSE | FALSE |
| SLBP | 0.4852049 | 0.73559791 | 1 | 0.485204918 | 0.485204918 | 0.485204918 | 0.5281555 | 0.743783573 | 1 | 0.528155503 | 0.528155503 | 0.528155503 | 0.918678146 | -0.036836615 | FALSE | TRUE | FALSE |
| ZDHHC21 | 0.4850513 | 0.7356391 | 1 | 0.485051277 | 0.485051277 | 0.485051277 | 0.51175688 | 0.750899422 | 1 | 4.792290277 | 4.792290277 | 4.792290277 | 0.947815836 | -0.023276039 | FALSE | TRUE | FALSE |
| AKT3 | 0.4849877 | 0.73568028 | 1 | 4.571625933 | 4.571625933 | 9.61697597 | 0.5453597 | 0.735650726 | 1 | 0.545359697 | 0.545359697 | 10.82521176 | 0.889298671 | -0.050952357 | FALSE | TRUE | FALSE |
| IGFL4 | 0.4848407 | 0.73572145 | 1 | 0.484840653 | 0.484840653 | 0.484840653 | 0.52755946 | 0.743990456 | 1 | 0.527559465 | 0.527559465 | 0.527559465 | 0.9190256 | -0.036672391 | FALSE | TRUE | FALSE |
| MED20 | 0.4847804 | 0.73576261 | 1 | 1.939004268 | 1.939004268 | 1.939004268 | 0.53390215 | 0.740891998 | 1 | 0.533902153 | 0.533902153 | 0.533902153 | 0.907994922 | -0.04191658 | FALSE | TRUE | FALSE |
| TRAK1 | 0.4846635 | 0.73580375 | 0.8765 | 4.504524936 | 42.94110096 | 87.92121696 | 6.40455061 | 0.257715901 | 0.50135 | 6.404550608 | 61.38519883 | 125.3694066 | 0.075674867 | -1.121048333 | FALSE | FALSE | FALSE |
| UBE2J1 | 0.4845346 | 0.73584488 | 1 | 0.484534565 | 0.484534565 | 0.484534565 | 0.5278064 | 0.743949107 | 1 | 0.527806403 | 0.527806403 | 0.527806403 | 0.9180157 | -0.037149892 | FALSE | TRUE | FALSE |
| RPS6 | 0.4844853 | 0.73588599 | 1 | 0.484485327 | 0.484485327 | 0.484485327 | 0.52791219 | 0.743866368 | 1 | 0.527912187 | 0.527912187 | 0.527912187 | 0.917738478 | -0.037281059 | FALSE | TRUE | FALSE |
| TMEM218 | 0.4844826 | 0.73592709 | 1 | 0.484482569 | 0.484482569 | 0.484482569 | 0.52721894 | 0.744114421 | 1 | 0.527218942 | 0.527218942 | 0.527218942 | 0.918939991 | -0.036712848 | FALSE | TRUE | FALSE |
| HNRNPAB | 0.4843475 | 0.73596818 | 0.99975 | 67746.50202 | 54387111.07 | 870166295.3 | 25.2455827 | 0.109451865 | 1 | 8.44325E+12 | 2.72142E+17 | 1.42848E+19 | 0.019185435 | -1.717028343 | FALSE | FALSE | FALSE |
| ADORA3 | 0.4838022 | 0.73600926 | 1 | 0.483802183 | 0.483802183 | 0.483802183 | 0.5271672 | 0.744155714 | 1 | 0.527167202 | 0.527167202 | 0.527167202 | 0.917739534 | -0.037280559 | FALSE | TRUE | FALSE |
| COPZ1 | 0.4837822 | 0.73605032 | 1 | 0.483782214 | 0.483782214 | 0.483782214 | 0.52728943 | 0.744073113 | 1 | 0.527289428 | 0.527289428 | 0.527289428 | 0.917488933 | -0.037399166 | FALSE | TRUE | FALSE |
| TXNDC8 | 0.4837804 | 0.73609138 | 0.49805 | 0.483780435 | 4.523795959 | 4.523795959 | 0.54349177 | 0.736795779 | 1 | 0.543491771 | 0.543491771 | 0.543491771 | 0.890133872 | -0.050544672 | FALSE | TRUE | FALSE |
| TWIST1 | 0.4836688 | 0.73613241 | 1 | 1.934331413 | 7.737884722 | 7.737884722 | 0.52127405 | 0.74681027 | 1 | 8.339792365 | 33.37049265 | 33.37049265 | 0.927858967 | -0.032518031 | FALSE | TRUE | FALSE |
| TMEM33 | 0.483655 | 0.73617344 | 1 | 0.483655006 | 0.483655006 | 0.483655006 | 0.5267885 | 0.744238263 | 1 | 0.526788503 | 0.526788503 | 0.526788503 | 0.918119896 | -0.037100601 | FALSE | TRUE | FALSE |
| CHCHD3 | 0.4836549 | 0.73621445 | 1 | 0.483654864 | 0.483654864 | 0.483654864 | 0.52673128 | 0.744279515 | 1 | 0.526731283 | 0.526731283 | 0.526731283 | 0.918219364 | -0.037053553 | FALSE | TRUE | FALSE |
| ACOT7 | 0.4834666 | 0.73625544 | 1 | 0.483466633 | 0.483466633 | 9.562925647 | 0.5268379 | 0.744196996 | 1 | 0.526837901 | 0.526837901 | 10.43460137 | 0.917676256 | -0.037310505 | FALSE | TRUE | FALSE |
| CCL21 | 0.4833711 | 0.73629643 | 1 | 0.483371058 | 0.483371058 | 0.483371058 | 0.52627427 | 0.74452674 | 0.00235 | 0.526274272 | 0.526274272 | 0.526274272 | 0.918477462 | -0.036931496 | FALSE | TRUE | FALSE |
| FAM117A | 0.4831763 | 0.7363374 | 1 | 0.483176304 | 0.483176304 | 9.574092082 | 0.52643764 | 0.744320756 | 1 | 0.526437638 | 0.526437638 | 10.4429229 | 0.91782249 | -0.037241305 | FALSE | TRUE | FALSE |
| GRAMD4 | 0.4827304 | 0.73637836 | 1 | 0.482730434 | 0.482730434 | 9.499568721 | 0.52630993 | 0.74444439 | 1 | 0.526309929 | 0.526309929 | 10.37718337 | 0.917198038 | -0.037536883 | FALSE | TRUE | FALSE |
| RAB18 | 0.4826248 | 0.73641931 | 1 | 0.482624771 | 0.482624771 | 0.482624771 | 4.74153744 | 0.317586069 | 0.508 | 4.741537435 | 18.96461681 | 18.96461681 | 0.101786557 | -0.992309576 | FALSE | FALSE | FALSE |

| | | | | | | | | | | | | | | | | | |
|---|---|---|---|---|---|---|---|---|---|---|---|---|---|---|---|---|---|
| CLEC3B | 0.4825636 | 0.73646024 | 1 | 0.482563561 | 0.482563561 | 0.482563561 | 0.52570208 | 0.744609039 | 1 | 0.525702081 | 0.525702081 | 0.525702081 | 0.917941128 | -0.037185171 | FALSE | TRUE | FALSE |
| NMNAT3 | 0.4825635 | 0.73650116 | 0.1298 | 0.482563506 | 4.482414972 | 4.482414972 | 0.01125003 | 0.896338114 | 1 | 0.100173667 | 0.915499424 | 0.915499424 | 42.89442335 | 1.632400834 | FALSE | TRUE | FALSE |
| ATG16L2 | 0.4824908 | 0.73654207 | 0.0154 | 0.120622819 | 0.482490751 | 2.385718579 | 0.0456666 | 0.887649571 | 0.50385 | 0.182626623 | 0.182626623 | 0.904413603 | 10.56550679 | 1.023890333 | FALSE | TRUE | FALSE |
| RBM42 | 0.4824758 | 0.73658296 | 0.49815 | 0.482475821 | 4.536881241 | 16.35321798 | 0.54268574 | 0.737058524 | 1 | 0.542685738 | 0.542685738 | 10.7371075 | 0.889051964 | -0.051072854 | FALSE | TRUE | FALSE |
| BCL3 | 0.4823729 | 0.73662384 | 1 | 43.77440497 | 4396.140689 | 4396.140689 | 0.5971215 | 0.713547424 | 1 | 0.597121502 | 0.597121502 | 11.83488912 | 0.807830314 | -0.092679854 | FALSE | TRUE | FALSE |
| TSLP | 0.4822972 | 0.73666471 | 1 | 0.482297228 | 0.482297228 | 0.482297228 | 0.52519896 | 0.744855598 | 1 | 0.525198965 | 0.525198965 | 0.525198965 | 0.918313363 | -0.037009096 | FALSE | TRUE | FALSE |
| HID1 | 0.4822576 | 0.73670556 | 0.2705 | 0.482257561 | 1.928123074 | 9.47419381 | 0.29257029 | 0.827265675 | 0.87445 | 1.170083801 | 10.82942843 | 22.91954626 | 1.648347673 | 0.217048819 | FALSE | TRUE | FALSE |
| SRP54 | 0.4820722 | 0.7367464 | 1 | 0.482072191 | 0.482072191 | 9.558985882 | 0.52569081 | 0.744650167 | 1 | 0.525690809 | 0.525690809 | 10.43461922 | 0.917026096 | -0.037618305 | FALSE | TRUE | FALSE |
| OR2G2 | 0.481978 | 0.73678723 | 1 | 4.511819104 | 4.511819104 | 4.511819104 | 4.61734528 | 0.322314963 | 0.8782 | 4.617345281 | 437.0294506 | 437.0294506 | 0.104384224 | -0.981365133 | FALSE | FALSE | FALSE |
| USF2 | 0.4819156 | 0.73682804 | 0.50355 | 0.481915575 | 0.481915575 | 2.393456736 | 0.15664135 | 0.86281436 | 1 | 0.156641351 | 0.156641351 | 0.156641351 | 3.076553996 | 0.488064542 | FALSE | TRUE | FALSE |
| AHSA1 | 0.4819131 | 0.73686884 | 1 | 4.559379597 | 4.559379597 | 4.559379597 | 4.97416538 | 0.307090447 | 0.0074 | 4.974165377 | 4.974165377 | 4.974165377 | 0.096883203 | -1.01375151 | FALSE | FALSE | FALSE |
| CD40 | 0.4814329 | 0.73690963 | 0.0078 | 0.48143291 | 0.48143291 | 0.48143291 | 0.52485615 | 0.744978692 | 1 | 0.524856154 | 0.524856154 | 0.524856154 | 0.917266391 | -0.037504519 | FALSE | TRUE | FALSE |
| IFNW1 | 0.4813106 | 0.73695041 | 1 | 0.481310558 | 0.481310558 | 0.481310558 | 0.52413918 | 0.745183582 | 1 | 0.524139182 | 0.524139182 | 0.524139182 | 0.918287688 | -0.037021238 | FALSE | TRUE | FALSE |
| PRAP1 | 0.481265 | 0.73699117 | 0.0043 | 0.481265026 | 0.481265026 | 0.481265026 | 0.52420131 | 0.745142632 | 0.00375 | 0.524201314 | 0.524201314 | 0.524201314 | 0.918091987 | -0.037113803 | FALSE | TRUE | FALSE |
| TAPT1 | 0.4812436 | 0.73703192 | 1 | 4.513804811 | 4.513804811 | 9.556280391 | 0.54127557 | 0.737451636 | 1 | 0.541275572 | 0.541275572 | 0.541275572 | 0.889091666 | -0.05105346 | FALSE | TRUE | FALSE |
| TCTE1 | 0.4810477 | 0.73707266 | 0.7574 | 0.48104774 | 4.376226885 | 9.495884265 | 0.07806687 | 0.88085351 | 1 | 0.697963241 | 0.697963241 | 1.54345099 | 6.161995894 | 0.789721405 | FALSE | TRUE | FALSE |
| PACRGL | 0.480854 | 0.73711339 | 0.50965 | 0.48085403 | 0.48085403 | 0.48085403 | 1.35462823 | 0.550572375 | 0.5093 | 1.354628227 | 5.418000479 | 5.418000479 | 0.354971216 | -0.449806861 | FALSE | FALSE | FALSE |
| WEE1 | 0.4807736 | 0.7371541 | 1 | 0.480773643 | 0.480773643 | 9.533544612 | 0.52448753 | 0.745019698 | 1 | 0.524487532 | 0.524487532 | 10.41103078 | 0.916654093 | -0.037794518 | FALSE | TRUE | FALSE |
| ZNF710 | 0.4807528 | 0.7371948 | 1 | 0.480752783 | 0.480752783 | 9.43289059 | 0.52442689 | 0.74506069 | 1 | 0.524426892 | 0.524426892 | 10.31329944 | 0.91672031 | -0.037763147 | FALSE | TRUE | FALSE |
| ZNF557 | 0.4804281 | 0.73723549 | 0.49975 | 0.054192163 | 0.480428117 | 0.480428117 | 0.07796037 | 0.880880576 | 1 | 0.077960374 | 0.077960374 | 0.077960374 | 6.162465559 | 0.789754505 | FALSE | TRUE | FALSE |
| AANAT | 0.4800746 | 0.73727616 | 1 | 0.480074649 | 0.480074649 | 0.480074649 | 0.52350779 | 0.745306361 | 0.0061 | 0.523507791 | 0.523507791 | 0.523507791 | 0.917034393 | -0.037614376 | FALSE | TRUE | FALSE |
| NDUFS5 | 0.4800735 | 0.73731683 | 1 | 0.480073459 | 0.480073459 | 0.480073459 | 0.52354751 | 0.745265449 | 1 | 0.523547514 | 0.523547514 | 0.523547514 | 0.916962542 | -0.037648405 | FALSE | TRUE | FALSE |
| CDC14A | 0.4798599 | 0.73735748 | 0.509 | 0.479859864 | 0.479859864 | 4.059688011 | 0.1554096 | 0.863165467 | 1 | 0.155409597 | 0.155409597 | 3.077465943 | 3.087710636 | 0.489636594 | FALSE | TRUE | FALSE |
| TM2D1 | 0.4798578 | 0.73739811 | 0.0063 | 0.479857782 | 0.479857782 | 0.479857782 | 0.52308352 | 0.745510716 | 1 | 0.523083523 | 0.523083523 | 0.523083523 | 0.917363597 | -0.037458498 | FALSE | TRUE | FALSE |
| NPM1 | 0.4798324 | 0.73743874 | 1 | 4.506511603 | 4.506511603 | 4.506511603 | 0.52990559 | 0.743119197 | 1 | 0.529905589 | 0.529905589 | 0.529905589 | 0.905505464 | -0.043108925 | FALSE | TRUE | FALSE |
| LRTM1 | 0.4797195 | 0.73747935 | 0.75525 | 0.479719494 | 4.433773155 | 4.433773155 | 0.67824059 | 0.676142758 | 0.7491 | 0.678240593 | 6.306577766 | 6.306577766 | 0.707299886 | -0.150396412 | FALSE | TRUE | FALSE |
| ZSCAN5A | 0.4797031 | 0.73751994 | 1 | 0.479703084 | 0.479703084 | 9.494584571 | 0.52321412 | 0.745388145 | 1 | 0.523214115 | 0.523214115 | 10.36869884 | 0.916838957 | -0.037706942 | FALSE | TRUE | FALSE |
| SCGB1A1 | 0.4794917 | 0.73756053 | 1 | 0.479491671 | 0.479491671 | 0.479491671 | 0.52235409 | 0.745918374 | 1 | 0.522354089 | 0.522354089 | 0.522354089 | 0.917943749 | -0.037183931 | FALSE | TRUE | FALSE |
| BPNT1 | 0.479488 | 0.7376011 | 0.5084 | 0.479487992 | 0.479487992 | 0.479487992 | 0.15565451 | 0.86307201 | 1 | 0.155654507 | 0.155654507 | 0.155654507 | 3.080463271 | 0.488616035 | FALSE | TRUE | FALSE |
| BCL2A1 | 0.47939 | 0.73764165 | 1 | 0.479390038 | 0.479390038 | 0.479390038 | 0.52245991 | 0.745836954 | 1 | 0.522459913 | 0.522459913 | 0.522459913 | 0.917563292 | -0.037363969 | FALSE | TRUE | FALSE |
| NGF | 0.4792607 | 0.7376822 | 0.00855 | 0.479260743 | 0.479260743 | 0.479260743 | 2.06825903 | 0.483712893 | 0.5055 | 2.068259033 | 2.068259033 | 2.068259033 | 0.231721818 | -0.635033073 | FALSE | FALSE | FALSE |
| DYNAP | 0.4792429 | 0.73772273 | 0.00555 | 0.479242864 | 0.479242864 | 0.479242864 | 0.52227178 | 0.745959063 | 1 | 0.522271777 | 0.522271777 | 0.522271777 | 0.917612028 | -0.037340903 | FALSE | TRUE | FALSE |
| GNB2 | 0.4791372 | 0.73776325 | 1 | 0.479137204 | 0.479137204 | 9.490363665 | 0.5227656 | 0.745633159 | 1 | 0.522765598 | 0.522765598 | 10.36652652 | 0.916543106 | -0.037847105 | FALSE | TRUE | FALSE |
| RDH12 | 0.4790294 | 0.73780375 | 0.5025 | 0.479029382 | 0.479029382 | 0.479029382 | 0.66605445 | 0.682595772 | 0.6277 | 1.14653081 | 10.64790345 | 10.64790345 | 0.719204536 | -0.143147582 | FALSE | TRUE | FALSE |
| TMEM52B | 0.4789454 | 0.73784425 | 1 | 0.478945397 | 0.478945397 | 0.478945397 | 0.52198098 | 0.746243488 | 1 | 0.52198098 | 0.52198098 | 0.52198098 | 0.91755335 | -0.037368675 | FALSE | TRUE | FALSE |
| RAB3A | 0.4788672 | 0.73788473 | 0.00805 | 0.478867183 | 0.478867183 | 0.478867183 | 2.04725405 | 0.485221767 | 0.50475 | 2.047254048 | 2.047254048 | 2.047254048 | 0.233907064 | -0.630956663 | FALSE | FALSE | FALSE |
| ST6GAL1 | 0.4788316 | 0.73792519 | 1 | 0.478831577 | 0.478831577 | 0.478831577 | 0.52225735 | 0.745999738 | 1 | 0.522257351 | 0.522257351 | 0.522257351 | 0.916849857 | -0.037701779 | FALSE | TRUE | FALSE |
| HLA-DRB1 | 0.4787782 | 0.73796564 | 0.99745 | 44113.59241 | 5059173.025 | 56029216.46 | 4.58516805 | 0.324797293 | 0.99465 | 45176.02882 | 57569079.76 | 651746611.6 | 0.1044189 | -0.981220885 | FALSE | FALSE | FALSE |
| YWHAB | 0.4786943 | 0.73800608 | 1 | 43.64701054 | 4404.175532 | 4404.175532 | 0.5927642 | 0.714903218 | 1 | 0.592764195 | 0.592764195 | 0.592764195 | 0.807562787 | -0.092823702 | FALSE | TRUE | FALSE |
| C1orf54 | 0.4786436 | 0.73804651 | 1 | 0.478643586 | 0.478643586 | 0.478643586 | 0.52161058 | 0.746608163 | 1 | 0.521610585 | 0.521610585 | 0.521610585 | 0.91762629 | -0.037334152 | FALSE | TRUE | FALSE |
| GNL1 | 0.4786371 | 0.73808692 | 1 | 0.478637075 | 0.478637075 | 9.436898378 | 0.52235522 | 0.745877672 | 1 | 0.522355217 | 0.522355217 | 10.31644223 | 0.916305724 | -0.037959601 | FALSE | TRUE | FALSE |
| RABIF | 0.4785826 | 0.73812732 | 1 | 0.478582624 | 0.478582624 | 0.478582624 | 0.52185167 | 0.746365172 | 1 | 0.521851666 | 0.521851666 | 0.521851666 | 0.917085554 | -0.037590148 | FALSE | TRUE | FALSE |
| DBI | 0.4785673 | 0.7381677 | 1 | 0.478567276 | 0.478567276 | 0.478567276 | 0.50554016 | 0.75352369 | 1 | 4.722297594 | 4.722297594 | 4.722297594 | 0.946645426 | -0.023812659 | FALSE | TRUE | FALSE |
| IQCF2 | 0.4784094 | 0.73820808 | 1 | 0.478409367 | 0.478409367 | 0.478409367 | 0.5216436 | 0.746486732 | 1 | 0.521643605 | 0.521643605 | 0.521643605 | 0.917119202 | -0.037574214 | FALSE | TRUE | FALSE |
| MAGOHB | 0.4784081 | 0.73824844 | 1 | 0.478408101 | 0.478408101 | 0.478408101 | 0.52185728 | 0.746324625 | 1 | 0.521857278 | 0.521857278 | 0.521857278 | 0.916741266 | -0.037753219 | FALSE | TRUE | FALSE |
| APOC3 | 0.4783031 | 0.73828878 | 1 | 0.478303117 | 0.478303117 | 0.478303117 | 0.52178887 | 0.746405706 | 0.00235 | 0.521788872 | 0.521788872 | 0.521788872 | 0.916660247 | -0.037791602 | FALSE | TRUE | FALSE |
| SCRT1 | 0.4782505 | 0.73832912 | 1 | 0.478250471 | 0.478250471 | 9.487655005 | 0.50846336 | 0.752257507 | 1 | 0.508463363 | 0.508463363 | 0.508463363 | 0.940580002 | -0.026604259 | FALSE | TRUE | FALSE |
| OR1C1 | 0.4780747 | 0.73836944 | 1 | 0.478074673 | 0.478074673 | 0.478074673 | 0.52136267 | 0.74672947 | 1 | 0.521362668 | 0.521362668 | 0.521362668 | 0.916971433 | -0.037644194 | FALSE | TRUE | FALSE |
| PRDM12 | 0.4780281 | 0.73840974 | 1 | 0.478028132 | 0.478028132 | 9.463464275 | 4.60673402 | 0.323420783 | 0.7523 | 4.606734025 | 44.48188227 | 44.48188227 | 0.103767252 | -0.983939683 | FALSE | FALSE | FALSE |
| SRSF6 | 0.4780229 | 0.73845004 | 1 | 0.478022905 | 0.478022905 | 9.467428353 | 0.52162946 | 0.746527223 | 1 | 0.521629455 | 0.521629455 | 10.343165 | 0.916403206 | -0.0379134 | FALSE | TRUE | FALSE |
| PPM1H | 0.4780166 | 0.73849032 | 1 | 0.47801659 | 0.47801659 | 9.456115092 | 0.52166782 | 0.746446226 | 1 | 0.521667825 | 0.521667825 | 10.33315622 | 0.916323697 | -0.037951082 | FALSE | TRUE | FALSE |
| MAFG | 0.478003 | 0.73853058 | 1 | 0.478003002 | 0.478003002 | 0.478003002 | 0.50773074 | 0.752796348 | 1 | 0.507730735 | 0.507730735 | 0.507730735 | 0.941449807 | -0.026202829 | FALSE | TRUE | FALSE |
| NFYC | 0.477952 | 0.73857084 | 1 | 4.474543267 | 4.474543267 | 9.495877705 | 4.58160087 | 0.325346214 | 0.87635 | 44.01590764 | 433.7137716 | 433.7137716 | 0.104319871 | -0.981632957 | FALSE | FALSE | FALSE |
| ERP29 | 0.4779226 | 0.73861108 | 1 | 0.477922591 | 0.477922591 | 0.477922591 | 0.52120483 | 0.746891015 | 1 | 0.521204829 | 0.521204829 | 0.521204829 | 0.916957336 | -0.037650871 | FALSE | TRUE | FALSE |
| DUSP7 | 0.4778855 | 0.7386513 | 1 | 0.477885452 | 0.477885452 | 9.456847795 | 0.52161675 | 0.7465677 | 1 | 0.521616753 | 0.521616753 | 10.33534798 | 0.91616201 | -0.038027721 | FALSE | TRUE | FALSE |
| TMEM190 | 0.4778549 | 0.73869152 | 1 | 0.477854859 | 0.477854859 | 0.477854859 | 0.52099175 | 0.747052339 | 1 | 0.520991754 | 0.520991754 | 0.520991754 | 0.917202347 | -0.037534843 | FALSE | TRUE | FALSE |
| TRIM44 | 0.4776578 | 0.73873172 | 0.00975 | 0.477657846 | 0.477657846 | 0.477657846 | 0.52107436 | 0.746971705 | 1 | 0.521074361 | 0.521074361 | 0.521074361 | 0.916678851 | -0.037782788 | FALSE | TRUE | FALSE |
| SNX6 | 0.4776092 | 0.73877191 | 1 | 0.477609168 | 0.477609168 | 0.477609168 | 0.52123726 | 0.74685065 | 1 | 0.52123726 | 0.52123726 | 0.52123726 | 0.916298976 | -0.037962799 | FALSE | TRUE | FALSE |
| C12orf76 | 0.4774766 | 0.73881208 | 1 | 0.477476597 | 0.477476597 | 0.477476597 | 0.52040765 | 0.747173194 | 1 | 0.520407645 | 0.520407645 | 0.520407645 | 0.917504962 | -0.037391578 | FALSE | TRUE | FALSE |
| ADAD1 | 0.4772047 | 0.73885225 | 0.3179 | 0.477204736 | 7.633651774 | 7.633651774 | 2.88754617 | 0.420232844 | 0.31395 | 0.721991296 | 11.55139457 | 11.55139457 | 0.165263067 | -0.781824192 | FALSE | FALSE | FALSE |
| NXPH3 | 0.4770507 | 0.7388924 | 1 | 0.477050714 | 0.477050714 | 0.477050714 | 0.52073246 | 0.747092637 | 1 | 0.520732465 | 0.520732465 | 0.520732465 | 0.916114792 | -0.038050104 | FALSE | TRUE | FALSE |
| MAD2L2 | 0.4769348 | 0.73893254 | 1 | 0.476934808 | 0.476934808 | 0.476934808 | 0.52028837 | 0.747253696 | 1 | 0.520288372 | 0.520288372 | 0.520288372 | 0.91667397 | -0.037785101 | FALSE | TRUE | FALSE |
| MMP14 | 0.4766154 | 0.73897267 | 1 | 0.476615374 | 0.476615374 | 9.392911267 | 0.48861254 | 0.75936933 | 0.7595 | 4.610150854 | 44.64258936 | 91.01458843 | 0.975446464 | -0.010796561 | FALSE | TRUE | FALSE |
| PTBP1 | 0.4765633 | 0.73901278 | 1 | 0.47656327 | 0.47656327 | 9.379839596 | 0.50421614 | 0.75379058 | 1 | 4.767273526 | 4.767273526 | 17.0912273 | 0.945156713 | -0.024496177 | FALSE | TRUE | FALSE |
| FGF19 | 0.4765421 | 0.73905288 | 1 | 0.476542099 | 0.476542099 | 0.476542099 | 0.52027941 | 0.747293926 | 1 | 0.520279406 | 0.520279406 | 0.520279406 | 0.915934963 | -0.038135363 | FALSE | TRUE | FALSE |
| ADAMTS5 | 0.4765052 | 0.73909297 | 0.69825 | 0.476505158 | 7.625685093 | 16.08925582 | 1.64136248 | 0.522626108 | 0.3868 | 0.714240408 | 6.564130128 | 32.1348749 | 0.290310741 | -0.537136896 | FALSE | FALSE | FALSE |
| BBS10 | 0.4763913 | 0.73913305 | 0.9391 | 40.2211491 | 3740.854352 | 7570.207305 | 0.92466356 | 0.595350121 | 0.81235 | 8.49697074 | 80.03835982 | 772.3736324 | 0.515205017 | -0.288019916 | FALSE | TRUE | FALSE |
| RTN4RL2 | 0.4761283 | 0.73917311 | 0.01475 | 0.476128303 | 0.476128303 | 9.412328899 | 0.50333825 | 0.75420876 | 1 | 4.746199181 | 4.746199181 | 9.964086483 | 0.945941027 | -0.024135938 | FALSE | TRUE | FALSE |
| GPR160 | 0.4760997 | 0.73921316 | 0.12685 | 0.05041163 | 0.476099656 | 0.476099656 | 0.00705432 | 0.897518007 | 0.50475 | 0.065374406 | 0.621404634 | 0.621404634 | 67.49048945 | 1.829242578 | FALSE | TRUE | FALSE |
| PTGS1 | 0.476016 | 0.7392532 | 0.01785 | 0.119037128 | 0.47601601 | 2.346191376 | 0.17966805 | 0.857119215 | 0.7574 | 0.179668045 | 0.718516942 | 3.547396628 | 2.649419427 | 0.423150717 | FALSE | TRUE | FALSE |
| RAB35 | 0.4759633 | 0.73929323 | 1 | 0.475963291 | 0.475963291 | 0.475963291 | 0.51970374 | 0.747494877 | 1 | 0.519703737 | 0.519703737 | 0.519703737 | 0.915835807 | -0.038182381 | FALSE | TRUE | FALSE |
| DIRAS2 | 0.4759062 | 0.73933324 | 1 | 1.903419418 | 1.903419418 | 1.903419418 | 0.525447 | 0.744814539 | 1 | 0.525446996 | 0.525446996 | 0.525446996 | 0.905716911 | -0.043007523 | FALSE | TRUE | FALSE |
| PSMG1 | 0.4758745 | 0.73937324 | 1 | 0.475874547 | 0.475874547 | 0.475874547 | 0.51926327 | 0.747655399 | 0.00655 | 0.519263269 | 0.519263269 | 0.519263269 | 0.916441766 | -0.037895127 | FALSE | TRUE | FALSE |

| | | | | | | | | | | | | | | | | | |
|---|---|---|---|---|---|---|---|---|---|---|---|---|---|---|---|---|---|
| DYRK1A | 0.4758037 | 0.73941322 | 1 | 0.475803699 | 0.475803699 | 9.364508322 | 0.51967988 | 0.747535027 | 1 | 0.519679881 | 0.519679881 | 10.2476651 | 0.915570751 | -0.03830809 | FALSE | TRUE | FALSE |
| DNAJB13 | 0.4755842 | 0.7394532 | 0.936 | 4.408211766 | 70.53416145 | 70.53416145 | 0.16300436 | 0.861368807 | 1 | 0.163004361 | 0.163004361 | 0.163004361 | 2.917616624 | 0.465028225 | FALSE | TRUE | FALSE |
| CBLN4 | 0.4755759 | 0.73949316 | 1 | 0.475575924 | 0.475575924 | 0.475575924 | 0.51929496 | 0.747615289 | 1 | 0.51929496 | 0.51929496 | 0.51929496 | 0.915810784 | -0.038194247 | FALSE | TRUE | FALSE |
| CD1A | 0.4755617 | 0.7395331 | 0.2556 | 0.475561734 | 4.413007635 | 4.413007635 | 0.57901783 | 0.721137757 | 0.8739 | 5.398423476 | 51.62428854 | 51.62428854 | 0.821324846 | -0.085485039 | FALSE | TRUE | FALSE |
| OR3A2 | 0.4755323 | 0.73957304 | 1 | 0.475532274 | 0.475532274 | 0.475532274 | 0.50300659 | 0.754284633 | 1 | 4.719890898 | 4.719890898 | 4.719890898 | 0.945379808 | -0.024393678 | FALSE | TRUE | FALSE |
| ELOVL6 | 0.4754519 | 0.73961296 | 1 | 0.475451916 | 0.475451916 | 0.475451916 | 0.4983658 | 0.755754711 | 1 | 0.498365804 | 0.498365804 | 0.498365804 | 0.954021949 | -0.020441633 | FALSE | TRUE | FALSE |
| TXN2 | 0.4750522 | 0.73965287 | 1 | 0.475052243 | 0.475052243 | 0.475052243 | 0.51816453 | 0.748135696 | 1 | 0.518164527 | 0.518164527 | 0.518164527 | 0.91679808 | -0.037726305 | FALSE | TRUE | FALSE |
| MDK | 0.474946 | 0.73969277 | 1 | 0.474945982 | 0.474945982 | 0.474945982 | 0.51818488 | 0.748095746 | 1 | 0.518184883 | 0.518184883 | 0.518184883 | 0.916557 | -0.037840521 | FALSE | TRUE | FALSE |
| ABCA6 | 0.4748665 | 0.73973265 | 0.12425 | 0.014094233 | 1.897268644 | 9.247373105 | 5.313E-07 | 0.901343428 | 0.69675 | 2.12164E-06 | 0.008966534 | 0.008966534 | 893788.3736 | 5.951234701 | FALSE | TRUE | FALSE |
| FAM81A | 0.4748664 | 0.73977253 | 1 | 0.474866442 | 0.474866442 | 9.426361519 | 0.51861115 | 0.747775654 | 1 | 0.518611146 | 0.518611146 | 10.3039686 | 0.915650281 | -0.038270367 | FALSE | TRUE | FALSE |
| TMEM205 | 0.4747717 | 0.73981239 | 1 | 4.447186087 | 4.447186087 | 4.447186087 | 0.53471738 | 0.74046714 | 1 | 0.53471738 | 0.53471738 | 0.53471738 | 0.88789272 | -0.051639505 | FALSE | TRUE | FALSE |
| TIPARP | 0.4747132 | 0.73985223 | 0.01705 | 0.474713165 | 0.474713165 | 9.390104072 | 0.4948796 | 0.756947412 | 1 | 4.67013212 | 4.67013212 | 9.801844829 | 0.95924982 | -0.018068274 | FALSE | TRUE | FALSE |
| LMO3 | 0.4747079 | 0.73989207 | 1 | 0.474707939 | 0.474707939 | 0.474707939 | 0.5182235 | 0.748055782 | 1 | 0.518223504 | 0.518223504 | 0.518223504 | 0.916029349 | -0.038090612 | FALSE | TRUE | FALSE |
| LPAR1 | 0.4746996 | 0.73993189 | 0.0104 | 0.474699616 | 0.474699616 | 9.413190818 | 0.51849835 | 0.747935809 | 1 | 0.518498347 | 0.518498347 | 0.518498347 | 0.915527732 | -0.038328496 | FALSE | TRUE | FALSE |
| SLC30A10 | 0.4746393 | 0.73997169 | 1 | 0.47463926 | 0.47463926 | 9.39660264 | 0.51855821 | 0.747815713 | 1 | 0.51855821 | 0.51855821 | 10.27859415 | 0.915305651 | -0.038433856 | FALSE | TRUE | FALSE |
| FADD | 0.4746301 | 0.74001149 | 1 | 0.474630086 | 0.474630086 | 0.474630086 | 0.51852687 | 0.747855759 | 1 | 0.518526868 | 0.518526868 | 0.518526868 | 0.915343284 | -0.038416001 | FALSE | TRUE | FALSE |
| VDAC2 | 0.4745068 | 0.74005127 | 1 | 4.458097674 | 4.458097674 | 4.458097674 | 4.87378844 | 0.31251703 | 0.5044 | 4.87378844 | 4.87378844 | 4.87378844 | 0.097358934 | -1.01162419 | FALSE | FALSE | FALSE |
| PXDC1 | 0.4745053 | 0.74009103 | 1 | 0.47450527 | 0.47450527 | 0.47450527 | 0.51794952 | 0.748295358 | 1 | 0.517949517 | 0.517949517 | 0.517949517 | 0.916122622 | -0.038046393 | FALSE | TRUE | FALSE |
| HIGD1C | 0.4743812 | 0.74013079 | 0.002 | 0.474381213 | 0.474381213 | 0.474381213 | 0.48623568 | 0.760597304 | 1 | 4.538158199 | 43.47107044 | 43.47107044 | 0.975619914 | -0.010719344 | FALSE | TRUE | FALSE |
| HOXA7 | 0.4743796 | 0.74017053 | 0.5032 | 0.474379551 | 0.474379551 | 0.474379551 | 0.60940081 | 0.707876406 | 0.50035 | 0.60940081 | 0.60940081 | 0.60940081 | 0.778436036 | -0.108777068 | FALSE | TRUE | FALSE |
| CSNK1G2 | 0.474335 | 0.74021026 | 1 | 0.47433498 | 0.47433498 | 9.364005092 | 0.51802639 | 0.748215555 | 1 | 0.518026387 | 0.518026387 | 10.24244714 | 0.915657951 | -0.038266729 | FALSE | TRUE | FALSE |
| PI3 | 0.4743025 | 0.74024998 | 1 | 0.474302541 | 0.474302541 | 0.474302541 | 0.5177262 | 0.748335241 | 0.0031 | 0.517726202 | 0.517726202 | 0.517726202 | 0.916126205 | -0.038044694 | FALSE | TRUE | FALSE |
| LHFPL2 | 0.4742873 | 0.74028968 | 0.0079 | 0.474287293 | 0.474287293 | 0.474287293 | 0.51798708 | 0.748255463 | 1 | 0.517987082 | 0.517987082 | 0.517987082 | 0.91563537 | -0.038277439 | FALSE | TRUE | FALSE |
| ZNF256 | 0.4742709 | 0.74032937 | 0.87505 | 4.348205462 | 40.88974585 | 40.88974585 | 0.07675902 | 0.881114697 | 1 | 0.691037815 | 6.385208528 | 6.385208528 | 6.178699354 | 0.790897064 | FALSE | TRUE | FALSE |
| FAM107A | 0.4741768 | 0.74036904 | 1 | 0.474176799 | 0.474176799 | 0.474176799 | 0.51740054 | 0.748454813 | 1 | 0.517400536 | 0.517400536 | 0.517400536 | 0.916459813 | -0.037886574 | FALSE | TRUE | FALSE |
| LIN7A | 0.474037 | 0.74040871 | 1 | 4.43978113 | 4.43978113 | 4.43978113 | 4.85482781 | 0.313930192 | 0.5086 | 4.854827812 | 4.854827812 | 4.854827812 | 0.097642402 | -1.010361545 | FALSE | FALSE | FALSE |
| PSKH1 | 0.4740305 | 0.74044836 | 1 | 0.474030478 | 0.474030478 | 9.359203711 | 0.45649 | 0.772257345 | 0.93815 | 41.47146899 | 4169.447782 | 4169.447782 | 1.038424675 | 0.016374999 | FALSE | TRUE | FALSE |
| POLL | 0.4738679 | 0.740488 | 0.8932 | 4.251545469 | 68.05823782 | 272.4020482 | 214.754337 | 0.012110756 | 0.56695 | 214.7543371 | 32613.5188 | 130879.0572 | 0.002206558 | -2.656284647 | FALSE | FALSE | FALSE |
| IL10 | 0.4736496 | 0.74052763 | 1 | 0.473649578 | 0.473649578 | 0.473649578 | 0.51742053 | 0.74841497 | 1 | 0.517420526 | 0.517420526 | 0.517420526 | 0.915405466 | -0.038386499 | FALSE | TRUE | FALSE |
| BAK1 | 0.473638 | 0.74056724 | 1 | 0.473637984 | 0.473637984 | 0.473637984 | 0.51695336 | 0.748534463 | 1 | 0.516953357 | 0.516953357 | 0.516953357 | 0.916210288 | -0.038004836 | FALSE | TRUE | FALSE |
| RHOA | 0.4732397 | 0.74060684 | 0.00505 | 0.47323974 | 0.47323974 | 0.47323974 | 0.46491477 | 0.769005798 | 1 | 4.337685965 | 4.337685965 | 4.337685965 | 1.01790644 | 0.007707862 | FALSE | TRUE | FALSE |
| SLC35B2 | 0.4731442 | 0.74064644 | 0.50955 | 0.47314424 | 0.47314424 | 2.343698715 | 0.15318901 | 0.863758374 | 1 | 0.153189013 | 0.153189013 | 3.038458413 | 3.08863038 | 0.489765939 | FALSE | TRUE | FALSE |
| NDFIP2 | 0.4731218 | 0.74068602 | 1 | 0.473121768 | 0.473121768 | 0.473121768 | 0.51688325 | 0.748653837 | 1 | 0.516883251 | 0.516883251 | 0.516883251 | 0.915335845 | -0.03841953 | FALSE | TRUE | FALSE |
| FGFBP3 | 0.473117 | 0.74072558 | 1 | 4.417039561 | 4.417039561 | 4.417039561 | 0.5332721 | 0.741315362 | 1 | 0.533272101 | 0.533272101 | 0.533272101 | 0.887196317 | -0.05198027 | FALSE | TRUE | FALSE |
| NFKBIZ | 0.4730632 | 0.74076513 | 1 | 0.473063239 | 0.473063239 | 9.354062265 | 0.51695282 | 0.748574268 | 1 | 0.516952819 | 0.516952819 | 10.23584214 | 0.915099447 | -0.038531707 | FALSE | TRUE | FALSE |
| EIF3H | 0.4729979 | 0.74080467 | 1 | 0.472997854 | 0.472997854 | 0.472997854 | 419.294989 | 0.005710092 | 0.12975 | 42.46495434 | 419.2949888 | 419.2949888 | 0.001128079 | -2.947660502 | FALSE | FALSE | FALSE |
| FOSL2 | 0.4729418 | 0.7408442 | 1 | 0.472941838 | 0.472941838 | 9.375519947 | 0.51119263 | 0.751094391 | 1 | 4.848962944 | 4.848962944 | 10.144533 | 0.925173426 | -0.03377685 | FALSE | TRUE | FALSE |
| MAP3K8 | 0.4729159 | 0.74088371 | 1 | 0.472915888 | 0.472915888 | 9.3660175 | 0.51618741 | 0.748812824 | 1 | 0.516187411 | 0.516187411 | 10.23500254 | 0.916170906 | -0.038023504 | FALSE | TRUE | FALSE |
| USO1 | 0.4729064 | 0.74092321 | 0.0218 | 0.472906421 | 0.472906421 | 9.322484703 | 18.6886014 | 0.140082995 | 0.26035 | 4.672200498 | 18.68860141 | 66.91854789 | 0.025304538 | -1.596801591 | FALSE | FALSE | FALSE |
| GULP1 | 0.4727349 | 0.7409627 | 1 | 0.472734943 | 0.472734943 | 0.472734943 | 0.51638947 | 0.748733356 | 1 | 0.51638947 | 0.51638947 | 0.51638947 | 0.915462011 | -0.038359673 | FALSE | TRUE | FALSE |
| FBXW2 | 0.4726258 | 0.74100218 | 1 | 0.472625804 | 0.472625804 | 9.374284745 | 0.51629442 | 0.748773096 | 0.00965 | 0.516294424 | 0.516294424 | 0.516294424 | 0.915419152 | -0.038380006 | FALSE | TRUE | FALSE |
| BLNK | 0.4725135 | 0.74104164 | 1 | 4.444160299 | 4.444160299 | 9.365150225 | 0.53280053 | 0.741442084 | 1 | 0.532800534 | 0.532800534 | 10.57147123 | 0.886848717 | -0.052150458 | FALSE | TRUE | FALSE |
| PIP4K2A | 0.4724917 | 0.74108109 | 1 | 1.889877469 | 1.889877469 | 9.363255967 | 0.5220983 | 0.746121676 | 1 | 0.522098304 | 0.522098304 | 10.35769897 | 0.904986136 | -0.043358074 | FALSE | TRUE | FALSE |
| RAB12 | 0.4724774 | 0.74112053 | 1 | 4.448837594 | 4.448837594 | 4.448837594 | 0.53278283 | 0.741484296 | 1 | 0.532782828 | 0.532782828 | 0.532782828 | 0.886810572 | -0.052169138 | FALSE | TRUE | FALSE |
| ZNF177 | 0.4723819 | 0.74115996 | 1 | 4.420210734 | 4.420210734 | 4.420210734 | 0.53108243 | 0.742325474 | 1 | 0.531082429 | 0.531082429 | 0.531082429 | 0.889470031 | -0.05086868 | FALSE | TRUE | FALSE |
| NAA60 | 0.4723079 | 0.74119937 | 1 | 4.415285626 | 4.415285626 | 4.415285626 | 0.53214489 | 0.741905612 | 1 | 0.532144892 | 0.532144892 | 0.532144892 | 0.887555122 | -0.051804665 | FALSE | TRUE | FALSE |
| VIP | 0.4722562 | 0.74123877 | 1 | 0.472256179 | 0.472256179 | 0.472256179 | 0.51564934 | 0.749090546 | 0.00315 | 0.515649341 | 0.515649341 | 0.515649341 | 0.915847537 | -0.038176818 | FALSE | TRUE | FALSE |
| CORO1A | 0.4721024 | 0.74127816 | 1 | 0.472102448 | 0.472102448 | 9.323325342 | 0.51605017 | 0.748852538 | 1 | 0.51605017 | 0.51605017 | 10.20664963 | 0.914838277 | -0.038655673 | FALSE | TRUE | FALSE |
| S100Z | 0.4720855 | 0.74131753 | 0.00255 | 0.472085526 | 0.472085526 | 0.472085526 | 0.51524219 | 0.749407131 | 1 | 0.515242189 | 0.515242189 | 0.515242189 | 0.916240044 | -0.037990731 | FALSE | TRUE | FALSE |
| SMR3A | 0.4720827 | 0.74135689 | 0.00355 | 0.47208272 | 0.47208272 | 0.47208272 | 0.51524629 | 0.749367605 | 1 | 0.515246285 | 0.515246285 | 0.515246285 | 0.916227315 | -0.037996765 | FALSE | TRUE | FALSE |
| GNAQ | 0.4719089 | 0.74139624 | 1 | 0.471908873 | 0.471908873 | 9.361715481 | 0.51594307 | 0.748931927 | 1 | 0.515943073 | 0.515943073 | 0.515943073 | 0.914652986 | -0.038743643 | FALSE | TRUE | FALSE |
| UQCRH | 0.4718189 | 0.74143558 | 1 | 0.471818899 | 0.471818899 | 0.471818899 | 0.51504098 | 0.749565098 | 1 | 0.515040976 | 0.515040976 | 0.515040976 | 0.916080314 | -0.03806645 | FALSE | TRUE | FALSE |
| FBXL19 | 0.4717698 | 0.74147491 | 1 | 0.471769826 | 0.471769826 | 9.293783274 | 0.51572155 | 0.748971602 | 1 | 0.515721555 | 0.515721555 | 10.17797775 | 0.914776242 | -0.038685123 | FALSE | TRUE | FALSE |
| LY6D | 0.4716666 | 0.74151422 | 1 | 0.471666576 | 0.471666576 | 0.471666576 | 0.51491212 | 0.749604557 | 0.00295 | 0.514912124 | 0.514912124 | 0.514912124 | 0.91601373 | -0.038098017 | FALSE | TRUE | FALSE |
| TMEM107 | 0.4714464 | 0.74155352 | 1 | 0.471446382 | 0.471446382 | 0.471446382 | 0.51470753 | 0.749722856 | 1 | 0.514707533 | 0.514707533 | 0.514707533 | 0.915950032 | -0.038128218 | FALSE | TRUE | FALSE |
| CD2 | 0.4714306 | 0.74159281 | 1 | 0.47143063 | 0.47143063 | 0.47143063 | 0.51521103 | 0.749486141 | 1 | 0.515211028 | 0.515211028 | 0.515211028 | 0.915024339 | -0.038567354 | FALSE | TRUE | FALSE |
| TNNC1 | 0.4713923 | 0.74163208 | 1 | 0.471392295 | 0.471392295 | 0.471392295 | 2.03772739 | 0.486056654 | 0.5061 | 2.037727387 | 2.037727387 | 2.037727387 | 0.231332365 | -0.635763603 | FALSE | FALSE | FALSE |
| SLAMF8 | 0.4712569 | 0.74167135 | 0.68855 | 0.471256906 | 42.06888752 | 42.06888752 | 0.0088457 | 0.897076692 | 1 | 0.008845703 | 0.008845703 | 0.008845703 | 53.27523666 | 1.726525388 | FALSE | TRUE | FALSE |
| PTGER4 | 0.4712506 | 0.7417106 | 1 | 0.471250634 | 0.471250634 | 9.29084317 | 0.51521263 | 0.749446643 | 1 | 0.51521263 | 0.51521263 | 10.17496908 | 0.914672131 | -0.038734553 | FALSE | TRUE | FALSE |
| MTFP1 | 0.4710544 | 0.74174983 | 0.50695 | 0.471054417 | 0.471054417 | 0.471054417 | 0.15148273 | 0.864231464 | 1 | 0.151482727 | 0.605724604 | 0.605724604 | 3.109624622 | 0.492707966 | FALSE | TRUE | FALSE |
| DYNC1LI1 | 0.4709699 | 0.74178906 | 1 | 0.470969887 | 0.470969887 | 9.324532915 | 0.51465328 | 0.749762262 | 1 | 0.514653282 | 0.514653282 | 10.20174605 | 0.915120729 | -0.038521607 | FALSE | TRUE | FALSE |
| SENP1 | 0.4708148 | 0.74182828 | 1 | 0.470814783 | 0.470814783 | 9.314557102 | 0.51473327 | 0.749644004 | 1 | 0.514733272 | 0.514733272 | 10.19667287 | 0.91467719 | -0.038732151 | FALSE | TRUE | FALSE |
| RNASE4 | 0.4707665 | 0.74186748 | 1 | 0.470766525 | 0.470766525 | 0.470766525 | 0.49856464 | 0.755717245 | 1 | 4.691996001 | 4.691996001 | 4.691996001 | 0.944243718 | -0.024915896 | FALSE | TRUE | FALSE |
| ZER1 | 0.4706513 | 0.74190667 | 1 | 0.470651298 | 0.470651298 | 9.227762768 | 0.51471794 | 0.749683437 | 1 | 0.514717936 | 0.514717936 | 10.11566935 | 0.914386824 | -0.038870041 | FALSE | TRUE | FALSE |
| VPS37D | 0.4706066 | 0.74194584 | 1 | 0.470606592 | 0.470606592 | 0.470606592 | 0.51416492 | 0.749919754 | 1 | 0.51416492 | 0.51416492 | 0.51416492 | 0.915283352 | -0.038444437 | FALSE | TRUE | FALSE |
| ING2 | 0.4705175 | 0.74198501 | 1 | 0.470517518 | 0.470517518 | 0.470517518 | 0.51447633 | 0.7498804 | 1 | 0.514476326 | 0.514476326 | 0.514476326 | 0.914556208 | -0.038789598 | FALSE | TRUE | FALSE |
| HTR3D | 0.4704807 | 0.74202416 | 0.87935 | 4.291990729 | 40.16043089 | 40.16043089 | 0.08110949 | 0.880063566 | 0.0078 | 0.081109487 | 0.081109487 | 0.081109487 | 5.800562529 | 0.763470113 | FALSE | TRUE | FALSE |
| MOG | 0.470401 | 0.7420633 | 1 | 0.470401047 | 4.399012239 | 4.399012239 | 0.53027282 | 0.742910833 | 1 | 0.530272816 | 0.530272816 | 0.530272816 | 0.887092517 | -0.052031084 | FALSE | TRUE | FALSE |
| ZNF576 | 0.4701422 | 0.74210243 | 1 | 0.470142167 | 0.470142167 | 0.470142167 | 0.51358902 | 0.750037746 | 1 | 0.513589016 | 0.513589016 | 0.513589016 | 0.915405416 | -0.038386523 | FALSE | TRUE | FALSE |
| AIF1 | 0.4700159 | 0.74214154 | 1 | 0.470015925 | 0.470015925 | 0.470015925 | 0.51330599 | 0.750273376 | 1 | 0.513305995 | 0.513305995 | 0.513305995 | 0.915664204 | -0.038263763 | FALSE | TRUE | FALSE |
| STH | 0.4698626 | 0.74218065 | 0.00295 | 0.469862555 | 0.469862555 | 0.469862555 | 4.63865973 | 0.321760679 | 0.00275 | 4.638659729 | 4.638659729 | 4.638659729 | 0.10129274 | -0.99442168 | FALSE | FALSE | FALSE |
| BUD31 | 0.4696497 | 0.74221974 | 1 | 0.469649674 | 0.469649674 | 0.469649674 | 2.03067448 | 0.487220822 | 0.5027 | 2.030674479 | 2.030674479 | 2.030674479 | 0.231277676 | -0.635866285 | FALSE | FALSE | FALSE |
| EGLN1 | 0.4696076 | 0.74225882 | 0.01305 | 0.469607578 | 0.469607578 | 9.308244025 | 0.51355711 | 0.75007705 | 1 | 0.513557112 | 0.513557112 | 10.19031972 | 0.914421332 | -0.038853651 | FALSE | TRUE | FALSE |

| | | | | | | | | | | | | | | | | | |
|---|---|---|---|---|---|---|---|---|---|---|---|---|---|---|---|---|---|
| ZDHHC14 | 0.4695916 | 0.74229789 | 1 | 0.469591626 | 0.469591626 | 9.273839261 | 0.51346819 | 0.750116342 | 1 | 0.513468186 | 0.513468186 | 10.15567491 | 0.914548629 | -0.038793197 | FALSE | TRUE | FALSE |
| GSG1L | 0.4695433 | 0.74233694 | 1 | 4.415676567 | 4.415676567 | 4.415676567 | 0.52982286 | 0.743160827 | 1 | 0.529822862 | 0.529822862 | 0.529822862 | 0.886226867 | -0.052455088 | FALSE | TRUE | FALSE |
| VASH1 | 0.4694303 | 0.74237598 | 1 | 0.469430341 | 0.469430341 | 9.303752967 | 0.51315215 | 0.75039101 | 0.00955 | 0.51315215 | 0.51315215 | 0.51315215 | 0.914797571 | -0.038674997 | FALSE | TRUE | FALSE |
| GFRA3 | 0.4694127 | 0.74241501 | 1 | 0.469412725 | 0.469412725 | 9.297167145 | 0.51328754 | 0.750312601 | 1 | 0.513287538 | 0.513287538 | 0.513287538 | 0.914521959 | -0.038805862 | FALSE | TRUE | FALSE |
| RAB8B | 0.4694095 | 0.74245403 | 1 | 0.469409472 | 0.469409472 | 0.469409472 | 0.51334624 | 0.750155621 | 0.0049 | 0.513346241 | 0.513346241 | 0.513346241 | 0.914411043 | -0.038858538 | FALSE | TRUE | FALSE |
| GABRA1 | 0.4692297 | 0.74249303 | 0.5036 | 1.87668101 | 1.87668101 | 9.303206587 | 2.03037676 | 0.487552349 | 0.25755 | 2.030376757 | 8.122599424 | 8.122599424 | 0.231104744 | -0.636191139 | FALSE | FALSE | FALSE |
| C6orf118 | 0.4691961 | 0.74253203 | 0.50655 | 0.469196134 | 0.469196134 | 1.055164199 | 0.64104352 | 0.693753468 | 0.7516 | 0.641043524 | 5.836652747 | 5.836652747 | 0.73192555 | -0.135533092 | FALSE | TRUE | FALSE |
| SYNCRIP | 0.4690967 | 0.74257101 | 1 | 0.469096703 | 0.469096703 | 9.284621999 | 0.51298772 | 0.750469368 | 1 | 0.512987719 | 0.512987719 | 10.16600592 | 0.914440416 | -0.038844587 | FALSE | TRUE | FALSE |
| CD80 | 0.4690806 | 0.74260997 | 1 | 0.469080599 | 0.469080599 | 0.469080599 | 0.51274147 | 0.750547674 | 0.0058 | 0.512741472 | 0.512741472 | 0.512741472 | 0.914848174 | -0.038650974 | FALSE | TRUE | FALSE |
| PAWR | 0.4690118 | 0.74264893 | 1 | 0.469011752 | 0.469011752 | 0.469011752 | 0.5125954 | 0.750665035 | 1 | 0.512595397 | 0.512595397 | 0.512595397 | 0.914974569 | -0.038590977 | FALSE | TRUE | FALSE |
| FAU | 0.4689283 | 0.74268787 | 0.00435 | 0.468928303 | 0.468928303 | 0.468928303 | 0.49886433 | 0.755604778 | 1 | 0.498864327 | 0.498864327 | 0.498864327 | 0.939991652 | -0.026876003 | FALSE | TRUE | FALSE |
| ASB5 | 0.4688915 | 0.7427268 | 0.50175 | 0.468891516 | 0.468891516 | 0.474561223 | 0.07342817 | 0.88168751 | 1 | 0.07342817 | 0.664524967 | 0.664524967 | 6.385717057 | 0.805209672 | FALSE | TRUE | FALSE |
| MIDN | 0.4688137 | 0.74276572 | 1 | 0.468813665 | 0.468813665 | 9.248938106 | 0.51279841 | 0.750508528 | 1 | 0.512798414 | 0.512798414 | 10.13322002 | 0.914226042 | -0.038946412 | FALSE | TRUE | FALSE |
| NFIA | 0.4686704 | 0.74280462 | 1 | 0.468670393 | 0.468670393 | 9.282654247 | 0.51269697 | 0.750586807 | 1 | 0.512696974 | 0.512696974 | 10.16648664 | 0.914127479 | -0.038993236 | FALSE | TRUE | FALSE |
| PLA2G2E | 0.4685338 | 0.74284352 | 1 | 0.468533782 | 0.468533782 | 0.468533782 | 0.51213348 | 0.750704132 | 1 | 0.512133483 | 0.512133483 | 0.512133483 | 0.914866528 | -0.038642261 | FALSE | TRUE | FALSE |
| GATA3 | 0.4684651 | 0.7428824 | 0.69345 | 0.468465145 | 7.489204725 | 7.489204725 | 0.70409453 | 0.664793588 | 0.8149 | 2.81518919 | 11.25881572 | 45.03874868 | 0.665344107 | -0.176953685 | FALSE | TRUE | FALSE |
| FGF20 | 0.4684197 | 0.74292127 | 1 | 1.87339272 | 1.87339272 | 1.87339272 | 4.73965711 | 0.317865931 | 0.4944 | 0.501841967 | 4.739657114 | 4.739657114 | 0.098829861 | -1.005111816 | FALSE | FALSE | FALSE |
| NADK2 | 0.4681601 | 0.74296012 | 1 | 4.390431466 | 4.390431466 | 9.296408547 | 0.51196379 | 0.750821345 | 1 | 4.808353984 | 4.808353984 | 4.808353984 | 0.914439913 | -0.038844826 | FALSE | TRUE | FALSE |
| GLRX3 | 0.4680773 | 0.74299897 | 1 | 0.468077311 | 0.468077311 | 0.468077311 | 0.51199716 | 0.750782287 | 1 | 0.511997157 | 0.511997157 | 0.511997157 | 0.914218575 | -0.038949959 | FALSE | TRUE | FALSE |
| SLPI | 0.4679991 | 0.7430378 | 1 | 1.871842622 | 1.871842622 | 1.871842622 | 4.70642579 | 0.320095395 | 0.0028 | 4.706425791 | 4.706425791 | 4.706425791 | 0.099438329 | -1.002446182 | FALSE | FALSE | FALSE |
| PITPNA | 0.4678766 | 0.74307663 | 0.00845 | 0.467876616 | 0.467876616 | 0.467876616 | 0.51180276 | 0.75086039 | 1 | 0.511802761 | 0.511802761 | 0.511802761 | 0.914173684 | -0.038971285 | FALSE | TRUE | FALSE |
| ALOX5AP | 0.4678414 | 0.74311543 | 1 | 0.467841365 | 0.467841365 | 0.467841365 | 0.51152037 | 0.750977448 | 1 | 0.51152037 | 0.51152037 | 0.51152037 | 0.91460945 | -0.038764316 | FALSE | TRUE | FALSE |
| PAPOLA | 0.4678239 | 0.74315423 | 1 | 0.467823877 | 0.467823877 | 0.467823877 | 0.5116768 | 0.750938441 | 1 | 0.5116768 | 0.5116768 | 0.5116768 | 0.91429566 | -0.038913342 | FALSE | TRUE | FALSE |
| NOG | 0.4676937 | 0.74319301 | 0.7552 | 0.467693661 | 1.870167709 | 2.321481536 | 0.58311834 | 0.719493428 | 0.87445 | 2.331865813 | 9.327364386 | 9.327364386 | 0.802056167 | -0.095795218 | FALSE | TRUE | FALSE |
| SF3A3 | 0.4672288 | 0.74323179 | 1 | 0.467228768 | 0.467228768 | 9.259090633 | 0.51144062 | 0.751016441 | 1 | 0.511440616 | 0.511440616 | 10.14639033 | 0.913554289 | -0.039265639 | FALSE | TRUE | FALSE |
| RND3 | 0.4672206 | 0.74327055 | 1 | 0.467220644 | 0.467220644 | 0.467220644 | 0.50946331 | 0.751793641 | 0.00655 | 0.509463313 | 0.509463313 | 0.509463313 | 0.917083982 | -0.037590892 | FALSE | TRUE | FALSE |
| ZNF691 | 0.4670485 | 0.7433093 | 1 | 0.467048468 | 0.467048468 | 0.467048468 | 0.51095061 | 0.75117229 | 1 | 0.510950611 | 0.510950611 | 0.510950611 | 0.914077521 | -0.039016971 | FALSE | TRUE | FALSE |
| XBP1 | 0.4668882 | 0.74334804 | 1 | 0.466888185 | 0.466888185 | 0.466888185 | 0.47967757 | 0.763119563 | 1 | 4.507710671 | 43.47583428 | 43.47583428 | 0.9733337532 | -0.01173653 | FALSE | TRUE | FALSE |
| OR2AG1 | 0.4667918 | 0.74338677 | 0.0072 | 0.466791849 | 0.466791849 | 0.466791849 | 4.63079358 | 0.322037849 | 0.5127 | 4.630793584 | 4.630793584 | 4.630793584 | 0.100801696 | -0.996532159 | FALSE | FALSE | FALSE |
| DDI2 | 0.4667688 | 0.74342548 | 1 | 0.466768811 | 0.466768811 | 9.25048084 | 4.64948769 | 0.321205859 | 0.50655 | 4.649487695 | 4.649487695 | 4.649487695 | 0.10039145 | -0.998303273 | FALSE | FALSE | FALSE |
| TNNC2 | 0.4667399 | 0.74346419 | 1 | 0.466739925 | 0.466739925 | 0.466739925 | 0.51060723 | 0.751327933 | 1 | 0.510607235 | 0.510607235 | 0.510607235 | 0.914087958 | -0.039012012 | FALSE | TRUE | FALSE |
| EXOC8 | 0.4666239 | 0.74350288 | 0.9704 | 41.97080237 | 1044.832214 | 4177.813511 | 1.572002 | 0.529374338 | 0.02055 | 1.572002 | 1.572002 | 30.94947018 | 0.296834195 | -0.527486071 | FALSE | FALSE | FALSE |
| CST9 | 0.4664991 | 0.74354155 | 0.00425 | 0.466499091 | 0.466499091 | 0.466499091 | 0.49432289 | 0.757095648 | 1 | 0.494322892 | 4.618133388 | 4.618133388 | 0.943713306 | -0.025159921 | FALSE | TRUE | FALSE |
| CCDC92 | 0.4663254 | 0.74358022 | 1 | 0.466325437 | 0.466325437 | 9.244828394 | 0.51017029 | 0.751561017 | 1 | 0.510170292 | 0.510170292 | 10.12469904 | 0.914058394 | -0.039026059 | FALSE | TRUE | FALSE |
| NAB2 | 0.466238 | 0.74361888 | 1 | 0.466237989 | 0.466237989 | 9.229281545 | 0.51024194 | 0.751483375 | 1 | 0.510241937 | 0.510241937 | 10.11278979 | 0.913758661 | -0.039168493 | FALSE | TRUE | FALSE |
| YBX2 | 0.4662211 | 0.74365752 | 1 | 0.466221083 | 0.466221083 | 9.250051988 | 0.50212036 | 0.754398359 | 1 | 0.502120364 | 0.502120364 | 9.971866619 | 0.928504631 | -0.032215926 | FALSE | TRUE | FALSE |
| HERPUD2 | 0.4660684 | 0.74369615 | 0.0107 | 0.466068407 | 0.466068407 | 9.251570168 | 0.51017457 | 0.751522203 | 1 | 0.510174569 | 0.510174569 | 10.13620701 | 0.913546922 | -0.039269141 | FALSE | TRUE | FALSE |
| LDB1 | 0.4660517 | 0.74373476 | 1 | 0.466051723 | 0.466051723 | 9.232061661 | 0.51029719 | 0.75140568 | 1 | 0.510297188 | 0.510297188 | 10.12013019 | 0.913294711 | -0.039389058 | FALSE | TRUE | FALSE |
| RAE1 | 0.4660334 | 0.74377337 | 1 | 0.466033391 | 0.466033391 | 0.466033391 | 0.51025779 | 0.751444534 | 0.00875 | 0.510257788 | 0.510257788 | 0.510257788 | 0.913329304 | -0.039372608 | FALSE | TRUE | FALSE |
| RPP25 | 0.4657153 | 0.74381196 | 0.0082 | 0.465715297 | 0.465715297 | 0.465715297 | 0.49340969 | 0.757649826 | 1 | 4.629152916 | 4.629152916 | 4.629152916 | 0.943871403 | -0.025087172 | FALSE | TRUE | FALSE |
| CA4 | 0.4656814 | 0.74385055 | 0.50285 | 0.465681391 | 0.465681391 | 0.465681391 | 0.59341642 | 0.714643503 | 0.75315 | 0.593416416 | 2.373399141 | 2.373399141 | 0.784746391 | -0.105270673 | FALSE | TRUE | FALSE |
| IER3IP1 | 0.4656684 | 0.74388912 | 1 | 0.465668351 | 0.465668351 | 0.465668351 | 0.50972822 | 0.751677385 | 1 | 0.509728216 | 0.509728216 | 0.509728216 | 0.913562043 | -0.039261953 | FALSE | TRUE | FALSE |
| NAA40 | 0.4656285 | 0.74392767 | 1 | 0.46562849 | 0.46562849 | 0.46562849 | 0.50923851 | 0.751948468 | 1 | 0.509238513 | 0.509238513 | 0.509238513 | 0.914362285 | -0.038881696 | FALSE | TRUE | FALSE |
| COX6A1 | 0.4655992 | 0.74396622 | 1 | 0.465599161 | 0.465599161 | 0.465599161 | 0.50944587 | 0.751832367 | 1 | 0.509445871 | 0.509445871 | 0.509445871 | 0.913932545 | -0.039085857 | FALSE | TRUE | FALSE |
| ETV6 | 0.4655379 | 0.74400475 | 1 | 0.465537856 | 0.465537856 | 9.207974956 | 0.50955889 | 0.751754902 | 1 | 0.509558887 | 0.509558887 | 10.0920615 | 0.913609531 | -0.039239378 | FALSE | TRUE | FALSE |
| PDRG1 | 0.4652247 | 0.74404327 | 1 | 0.465224744 | 0.465224744 | 0.465224744 | 0.50876412 | 0.752180323 | 1 | 0.508764121 | 0.508764121 | 0.508764121 | 0.914421291 | -0.038853671 | FALSE | TRUE | FALSE |
| CRABP1 | 0.4651283 | 0.74408178 | 1 | 0.465128312 | 0.465128312 | 0.465128312 | 0.5086892 | 0.75221892 | 1 | 0.508689203 | 0.508689203 | 0.508689203 | 0.914366395 | -0.038879744 | FALSE | TRUE | FALSE |
| ARMC1 | 0.4649746 | 0.74412028 | 1 | 0.464974629 | 0.464974629 | 0.464974629 | 0.50897353 | 0.752103088 | 1 | 0.508973533 | 0.508973533 | 0.508973533 | 0.91355365 | -0.039265943 | FALSE | TRUE | FALSE |
| MED7 | 0.4648116 | 0.74415877 | 1 | 1.858994296 | 7.43682845 | 7.43682845 | 2.03559447 | 0.486389856 | 0.25605 | 2.03559447 | 8.143440095 | 8.143440095 | 0.22834192 | -0.641414351 | FALSE | FALSE | FALSE |
| SPATA9 | 0.4647907 | 0.74419724 | 1 | 0.464790659 | 0.464790659 | 0.464790659 | 0.50837604 | 0.752334641 | 1 | 0.508376045 | 0.508376045 | 0.508376045 | 0.914265461 | -0.038927686 | FALSE | TRUE | FALSE |
| TMEM121 | 0.4646433 | 0.7442357 | 1 | 0.464643308 | 0.464643308 | 0.464643308 | 0.50821755 | 0.752527243 | 1 | 0.508217555 | 0.508217555 | 0.508217555 | 0.914260643 | -0.038929975 | FALSE | TRUE | FALSE |
| KRCC1 | 0.4646134 | 0.74427416 | 1 | 4.339241622 | 4.339241622 | 4.339241622 | 44.1836637 | 0.062289423 | 0.0046 | 4.604657789 | 44.18366366 | 44.18366366 | 0.010515501 | -1.978170013 | FALSE | FALSE | FALSE |
| LACRT | 0.4645933 | 0.74431259 | 1 | 0.464593301 | 0.464593301 | 0.464593301 | 0.50802896 | 0.752604194 | 1 | 0.508028959 | 0.508028959 | 0.508028959 | 0.91450161 | -0.038815526 | FALSE | TRUE | FALSE |
| GPRC5A | 0.4644769 | 0.74435102 | 0.50465 | 0.464476884 | 0.464476884 | 0.464476884 | 2.56187115 | 0.44272448 | 0.49955 | 1.372380915 | 40.97651539 | 40.97651539 | 0.181303765 | -0.741593178 | FALSE | FALSE | FALSE |
| SOCS6 | 0.4644204 | 0.74438943 | 1 | 0.464420438 | 0.464420438 | 9.170306815 | 0.50877883 | 0.752141712 | 1 | 0.508778827 | 0.508778827 | 10.06156467 | 0.912814004 | -0.039617706 | FALSE | TRUE | FALSE |
| CLDN4 | 0.4643348 | 0.74442783 | 0.8153 | 1.856726698 | 7.426313099 | 29.71029807 | 0.04613829 | 0.887535809 | 1 | 0.184420884 | 0.184420884 | 0.184420884 | 10.06397886 | 1.002769716 | FALSE | TRUE | FALSE |
| ATP6V0B | 0.4642465 | 0.74446622 | 1 | 0.464246524 | 0.464246524 | 0.464246524 | 4.45722988 | 0.332137205 | 0.75385 | 4.457229877 | 42.73204546 | 42.73204546 | 0.10415584 | -0.982316372 | FALSE | FALSE | FALSE |
| RAB40C | 0.4641899 | 0.7445046 | 1 | 0.464189932 | 0.464189932 | 9.196652955 | 0.50839334 | 0.75229608 | 1 | 0.508393339 | 0.508393339 | 10.08379477 | 0.913052742 | -0.039504135 | FALSE | TRUE | FALSE |
| SIVA1 | 0.4641596 | 0.74454297 | 1 | 0.464159604 | 0.464159604 | 0.464159604 | 0.4871027 | 0.760129358 | 1 | 0.487102697 | 0.487102697 | 0.487102697 | 0.952898859 | -0.020953193 | FALSE | TRUE | FALSE |
| ZNF496 | 0.4640821 | 0.74458132 | 1 | 0.464082098 | 0.464082098 | 9.132992303 | 0.50837148 | 0.752373188 | 1 | 0.508371484 | 0.508371484 | 10.02388139 | 0.912879876 | -0.039586367 | FALSE | TRUE | FALSE |
| GTF2B | 0.4640064 | 0.74461966 | 1 | 0.46400644 | 0.46400644 | 0.46400644 | 0.50790649 | 0.752719525 | 1 | 0.507906492 | 0.507906492 | 0.507906492 | 0.913566665 | -0.039259756 | FALSE | TRUE | FALSE |
| UBA2 | 0.4640013 | 0.74465799 | 0.01865 | 0.464001291 | 0.464001291 | 9.165480856 | 0.50831367 | 0.752488749 | 1 | 0.508313675 | 0.508313675 | 10.05570168 | 0.912824726 | -0.039612605 | FALSE | TRUE | FALSE |
| RTF1 | 0.4639227 | 0.74469631 | 1 | 0.463922671 | 0.463922671 | 9.16927652 | 0.50820681 | 0.752565725 | 1 | 0.508206807 | 0.508206807 | 10.0587578 | 0.912861978 | -0.039594882 | FALSE | TRUE | FALSE |
| HOXC8 | 0.4637602 | 0.74473461 | 1 | 0.463760212 | 0.463760212 | 0.463760212 | 0.50793411 | 0.752681095 | 1 | 0.507934112 | 0.507934112 | 0.507934112 | 0.913032225 | -0.039513894 | FALSE | TRUE | FALSE |
| CCDC71 | 0.4636368 | 0.7447729 | 1 | 0.463636823 | 0.463636823 | 9.172644605 | 0.50769804 | 0.752834741 | 1 | 0.507698037 | 0.507698037 | 10.05739987 | 0.913213739 | -0.039427563 | FALSE | TRUE | FALSE |
| RPL8 | 0.4635054 | 0.74481119 | 1 | 0.46350543 | 0.46350543 | 0.550746341 | 0.5078457 | 0.752757943 | 1 | 0.507845699 | 0.507845699 | 10.07728514 | 0.912689486 | -0.039676953 | FALSE | TRUE | FALSE |
| STK39 | 0.463277 | 0.74484946 | 1 | 0.463276975 | 0.463276975 | 9.181067567 | 0.50731526 | 0.753026514 | 1 | 0.507315261 | 0.507315261 | 10.06482787 | 0.913193453 | -0.039437211 | FALSE | TRUE | FALSE |
| SYT2 | 0.4630946 | 0.74488772 | 1 | 0.463094577 | 0.463094577 | 9.180611499 | 0.50734388 | 0.752988185 | 1 | 0.507343877 | 0.507343877 | 10.06844684 | 0.912782429 | -0.039632728 | FALSE | TRUE | FALSE |
| MRPL53 | 0.4629453 | 0.74492597 | 0.00505 | 0.462945284 | 0.462945284 | 0.462945284 | 0.50664165 | 0.753179711 | 1 | 0.50664165 | 0.50664165 | 0.50664165 | 0.913752915 | -0.039171225 | FALSE | TRUE | FALSE |
| LEAP2 | 0.4625961 | 0.7449642 | 0.0021 | 0.462596126 | 0.462596126 | 0.462596126 | 0.50609199 | 0.753485517 | 1 | 0.506091986 | 0.506091986 | 0.506091986 | 0.914055426 | -0.039027469 | FALSE | TRUE | FALSE |
| INSIG2 | 0.4623795 | 0.74500243 | 1 | 0.462379478 | 0.462379478 | 0.462379478 | 0.50621028 | 0.753294485 | 1 | 0.506210278 | 0.506210278 | 0.506210278 | 0.913413849 | -0.039332408 | FALSE | TRUE | FALSE |
| PADI1 | 0.4621699 | 0.74504065 | 0.8773 | 4.215037531 | 39.42974903 | 39.42974903 | 4889.18814 | 0.000369945 | 0.11045 | 51.39993428 | 4889.188143 | 49415.07496 | 9.4529E-05 | -4.024435118 | FALSE | FALSE | FALSE |
| PLCXD3 | 0.4621447 | 0.74507885 | 1 | 0.462144749 | 0.462144749 | 0.462144749 | 0.50615978 | 0.753409143 | 1 | 0.506159783 | 0.506159783 | 0.506159783 | 0.913041227 | -0.039509612 | FALSE | TRUE | FALSE |

| | | | | | | | | | | | | | | | | | |
|---|---|---|---|---|---|---|---|---|---|---|---|---|---|---|---|---|---|
| DUSP6 | 0.4619931 | 0.74511705 | 0.01355 | 0.461993071 | 0.461993071 | 9.138654824 | 0.50635938 | 0.753256239 | 1 | 0.506359376 | 0.506359376 | 10.02946031 | 0.912381786 | -0.039823393 | FALSE | TRUE | FALSE |
| TMEM151A | 0.4619913 | 0.74515523 | 1 | 4.330422746 | 4.330422746 | 9.13712925 | 0.52264673 | 0.745714719 | 1 | 0.522646728 | 0.522646728 | 10.35057704 | 0.883945577 | -0.053574473 | FALSE | TRUE | FALSE |
| PNOC | 0.4618343 | 0.7451934 | 1 | 0.461834292 | 0.461834292 | 0.461834292 | 0.50610332 | 0.753447337 | 1 | 0.506103318 | 0.506103318 | 0.506103318 | 0.912529667 | -0.039753007 | FALSE | TRUE | FALSE |
| SP3 | 0.4615094 | 0.74523156 | 1 | 0.461509371 | 0.461509371 | 9.11373357 | 0.49914606 | 0.755417092 | 1 | 0.499146064 | 0.499146064 | 9.871934531 | 0.924597837 | -0.034047127 | FALSE | TRUE | FALSE |
| ZC3HAV1L | 0.461461 | 0.7452697 | 1 | 4.30111249 | 4.30111249 | 4.30111249 | 0.52108573 | 0.746931367 | 1 | 0.52108573 | 0.52108573 | 0.52108573 | 0.885576024 | -0.05277415 | FALSE | TRUE | FALSE |
| CYTH1 | 0.4610878 | 0.74530784 | 0.0124 | 0.461087831 | 0.461087831 | 9.136722395 | 0.48976507 | 0.758896274 | 1 | 4.634129087 | 4.634129087 | 9.715742201 | 0.941446945 | -0.02620415 | FALSE | TRUE | FALSE |
| RNF146 | 0.4610303 | 0.74534597 | 1 | 4.318044451 | 41.50802391 | 41.50802391 | 0.53783441 | 0.739484384 | 1 | 0.537834405 | 0.537834405 | 0.537834405 | 0.857197473 | -0.066919118 | FALSE | TRUE | FALSE |
| CACNG3 | 0.4609144 | 0.74538408 | 1 | 0.460914399 | 0.460914399 | 9.145017893 | 0.50535423 | 0.753599996 | 1 | 0.505354229 | 0.505354229 | 0.600664029 | 0.912062019 | -0.039975629 | FALSE | TRUE | FALSE |
| BLOC1S1 | 0.4606211 | 0.74542218 | 1 | 0.460621051 | 0.460621051 | 0.460621051 | 0.50426564 | 0.753714384 | 1 | 0.50426564 | 0.50426564 | 0.50426564 | 0.913449212 | -0.039315595 | FALSE | TRUE | FALSE |
| RCE1 | 0.460441 | 0.74546028 | 0.00945 | 0.460441031 | 0.460441031 | 0.460441031 | 0.50446813 | 0.753638138 | 1 | 0.50446813 | 0.50446813 | 0.50446813 | 0.912725706 | -0.039659718 | FALSE | TRUE | FALSE |
| PRELID1 | 0.4602811 | 0.74549836 | 0.00755 | 0.460281104 | 0.460281104 | 0.460281104 | 0.50414013 | 0.753828659 | 1 | 0.504140125 | 0.504140125 | 0.504140125 | 0.913002321 | -0.039528119 | FALSE | TRUE | FALSE |
| FAM153A | 0.4602465 | 0.74553643 | 1 | 0.46024652 | 0.46024652 | 0.46024652 | 0.50409535 | 0.753866725 | 1 | 0.504095345 | 0.504095345 | 0.504095345 | 0.913014818 | -0.039522174 | FALSE | TRUE | FALSE |
| BAZ2A | 0.4601918 | 0.74557448 | 0.40385 | 0.460191804 | 4.289141641 | 35.91793546 | 0.04617685 | 0.887527675 | 1 | 0.046176855 | 0.046176855 | 1.549121108 | 9.965854308 | 0.998514534 | FALSE | TRUE | FALSE |
| GTF3C4 | 0.459925 | 0.74561253 | 1 | 0.459925047 | 0.459925047 | 9.050192939 | 0.50422378 | 0.753752489 | 1 | 0.504223775 | 0.504223775 | 9.941139848 | 0.912144705 | -0.039936259 | FALSE | TRUE | FALSE |
| LYRM2 | 0.4597774 | 0.74565056 | 1 | 1.839029618 | 1.839029618 | 1.839029618 | 0.50354688 | 0.75413284 | 1 | 2.014110189 | 2.014110189 | 2.014110189 | 0.913077678 | -0.039492274 | FALSE | TRUE | FALSE |
| PTP4A2 | 0.4594626 | 0.74568859 | 0.0048 | 0.459462595 | 0.459462595 | 0.459462595 | 0.50189564 | 0.754436248 | 0.0046 | 0.501895643 | 0.501895643 | 0.501895643 | 0.915454441 | -0.038363264 | FALSE | TRUE | FALSE |
| MGARP | 0.4593321 | 0.7457266 | 1 | 4.286444009 | 4.286444009 | 4.286444009 | 0.5189641 | 0.74773558 | 0.0061 | 0.518964102 | 0.518964102 | 0.518964102 | 0.885094132 | -0.053010538 | FALSE | TRUE | FALSE |
| NRBP1 | 0.4592751 | 0.74576461 | 1 | 0.459275072 | 0.459275072 | 9.091895868 | 0.50369096 | 0.754018865 | 1 | 0.503690961 | 0.503690961 | 9.983384246 | 0.911819167 | -0.040091283 | FALSE | TRUE | FALSE |
| CCS | 0.4591984 | 0.7458026 | 0.1332 | 0.459198399 | 1.836531414 | 1.836531414 | 0.40000336 | 0.792111953 | 0.50345 | 0.400003363 | 0.400003363 | 0.400003363 | 1.147986344 | 0.059936722 | FALSE | TRUE | FALSE |
| SPINK4 | 0.4590663 | 0.74584057 | 1 | 0.459066312 | 0.459066312 | 0.459066312 | 0.50331956 | 0.754246702 | 1 | 0.503319563 | 0.503319563 | 0.503319563 | 0.912077227 | -0.039968388 | FALSE | TRUE | FALSE |
| PABPC1 | 0.459034 | 0.74587854 | 1 | 0.459033951 | 0.459033951 | 9.044685413 | 0.50351888 | 0.754170806 | 1 | 0.503518877 | 0.503518877 | 9.938893101 | 0.911651919 | -0.040170949 | FALSE | TRUE | FALSE |
| UBASH3A | 0.4589907 | 0.7459165 | 0.50495 | 1.015447381 | 16.25225758 | 19.99974435 | 0.04550313 | 0.887706372 | 1 | 0.181978874 | 0.181978874 | 1.540466529 | 10.08701499 | 1.003762666 | FALSE | TRUE | FALSE |
| SPAG8 | 0.4587063 | 0.74595444 | 0.51375 | 0.458706301 | 0.458706301 | 1.757558205 | 0.64850021 | 0.690519084 | 0.50945 | 0.648500207 | 0.648500207 | 1.442980317 | 0.707334086 | -0.150375413 | FALSE | TRUE | FALSE |
| UBTD1 | 0.4586618 | 0.74599237 | 1 | 41.74335209 | 4204.398504 | 4204.398504 | 0.50290879 | 0.754322552 | 1 | 45.8628943 | 4628.626271 | 4628.626271 | 0.91201778 | -0.039996695 | FALSE | TRUE | FALSE |
| PLAU | 0.4585625 | 0.74603029 | 0.75625 | 0.458562482 | 4.348151663 | 4.348151663 | 0.66882416 | 0.680893263 | 0.50875 | 0.668824162 | 2.673910143 | 2.673910143 | 0.685624874 | -0.163913435 | FALSE | TRUE | FALSE |
| ZNF512 | 0.4584302 | 0.7460682 | 1 | 0.458430193 | 0.458430193 | 9.066842613 | 0.50261672 | 0.75436046 | 1 | 0.50261672 | 0.50261672 | 9.954038142 | 0.912087033 | -0.039963718 | FALSE | TRUE | FALSE |
| SERTM1 | 0.4584168 | 0.7461061 | 0.00315 | 0.458416755 | 0.458416755 | 0.458416755 | 0.46215943 | 0.770232308 | 1 | 0.462159432 | 0.462159432 | 0.462159432 | 0.991901762 | -0.003531338 | FALSE | TRUE | FALSE |
| UGDH | 0.4578933 | 0.74614399 | 0.51025 | 0.457893288 | 0.457893288 | 2.268945582 | 1.34548151 | 0.551092981 | 0.50535 | 1.345481509 | 1.345481509 | 2.873779278 | 0.340319273 | -0.468113456 | FALSE | FALSE | FALSE |
| SERF2 | 0.4577491 | 0.74618186 | 0.50405 | 4.261458082 | 4.261458082 | 4.261458082 | 0.51727267 | 0.748494644 | 0.0025 | 0.517272673 | 0.517272673 | 0.517272673 | 0.884928029 | -0.053092049 | FALSE | TRUE | FALSE |
| TMED7 | 0.457587 | 0.74621973 | 1 | 0.457587033 | 0.457587033 | 0.457587033 | 0.4696573 | 0.76708923 | 1 | 0.4696573 | 0.4696573 | 0.4696573 | 0.974299841 | -0.011307368 | FALSE | TRUE | FALSE |
| RSU1 | 0.4575773 | 0.74625758 | 1 | 0.457577324 | 0.457577324 | 0.457577324 | 0.50159141 | 0.75454984 | 1 | 0.501591407 | 0.501591407 | 0.501591407 | 0.912251122 | -0.039885594 | FALSE | TRUE | FALSE |
| RASL10B | 0.4575304 | 0.74629542 | 0.50155 | 4.322119062 | 4.322119062 | 4.322119062 | 0.51808858 | 0.748175632 | 1 | 0.518088578 | 0.518088578 | 0.518088578 | 0.883112265 | -0.053984084 | FALSE | TRUE | FALSE |
| EEF1A1 | 0.4574755 | 0.74633325 | 1 | 0.457475499 | 0.457475499 | 9.059786006 | 0.50172399 | 0.754511988 | 1 | 0.501723993 | 0.501723993 | 9.947797874 | 0.9118071 | -0.04009703 | FALSE | TRUE | FALSE |
| PSMD13 | 0.4572746 | 0.74637107 | 1 | 4108.587636 | 450954.9147 | 4892907.573 | 0.37848435 | 0.79982102 | 1 | 41804497.61 | 7.66127E+11 | 9.38619E+12 | 1.208173109 | 0.082129165 | FALSE | TRUE | FALSE |
| VAMP1 | 0.4572605 | 0.74640888 | 1 | 0.457260484 | 0.457260484 | 0.457260484 | 0.50112889 | 0.754701125 | 1 | 0.501128886 | 0.501128886 | 0.501128886 | 0.91246084 | -0.039785765 | FALSE | TRUE | FALSE |
| PRDX2 | 0.4572049 | 0.74644667 | 1 | 0.457204899 | 0.457204899 | 0.457204899 | 0.50136453 | 0.754663321 | 1 | 0.50136453 | 0.50136453 | 0.50136453 | 0.91192111 | -0.040042731 | FALSE | TRUE | FALSE |
| CD3E | 0.4572033 | 0.74648446 | 1 | 0.457203262 | 0.457203262 | 0.457203262 | 0.50093239 | 0.754776696 | 1 | 0.500932387 | 0.500932387 | 0.500932387 | 0.912704535 | -0.039669792 | FALSE | TRUE | FALSE |
| PGGT1B | 0.4571436 | 0.74652223 | 1 | 0.457143592 | 0.457143592 | 0.457143592 | 0.50138121 | 0.754625507 | 1 | 0.501381209 | 0.501381209 | 0.501381209 | 0.911768497 | -0.040115418 | FALSE | TRUE | FALSE |
| LZTS1 | 0.4568946 | 0.74655999 | 0.02055 | 0.456894638 | 0.456894638 | 8.992963635 | 4.43668034 | 0.333746166 | 0.26335 | 4.436680339 | 42.91293088 | 87.4929906 | 0.102981194 | -0.987242078 | FALSE | FALSE | FALSE |
| NRTN | 0.456779 | 0.74659773 | 1 | 0.456779023 | 0.456779023 | 0.456779023 | 0.49543912 | 0.756687544 | 1 | 1.98142714 | 1.98142714 | 1.98142714 | 0.921968021 | -0.035284142 | FALSE | TRUE | FALSE |
| MIS18BP1 | 0.4566714 | 0.74663547 | 0.81745 | 3.98508788 | 81.96012723 | 327.9180494 | 14.0613362 | 0.162846438 | 0.1999 | 1.553780281 | 56.25873715 | 56.25873715 | 0.032477098 | -1.488422788 | FALSE | FALSE | FALSE |
| CAMK2A | 0.4566525 | 0.7466732 | 1 | 4.308960666 | 4.308960666 | 15.47124781 | 0.48552359 | 0.760920137 | 1 | 4.580757062 | 44.35554578 | 90.65342984 | 0.940536278 | -0.026624448 | FALSE | TRUE | FALSE |
| DCAF16 | 0.4565245 | 0.74671091 | 1 | 0.456524522 | 0.456524522 | 0.456524522 | 0.50033669 | 0.755040811 | 1 | 0.500336689 | 0.500336689 | 0.500336689 | 0.912434631 | -0.03979824 | FALSE | TRUE | FALSE |
| GPR26 | 0.4565199 | 0.74674861 | 1 | 0.45651988 | 0.45651988 | 9.030967383 | 0.48538166 | 0.760991754 | 1 | 4.575054814 | 4.575054814 | 9.614496308 | 0.940537963 | -0.026623671 | FALSE | TRUE | FALSE |
| ZDHHC18 | 0.4565106 | 0.7467863 | 1 | 0.456510557 | 0.456510557 | 0.456510557 | 0.50080581 | 0.754814463 | 1 | 0.500805807 | 0.500805807 | 0.500805807 | 0.911552042 | -0.040218532 | FALSE | TRUE | FALSE |
| LRRC57 | 0.4564895 | 0.74682398 | 0.0065 | 0.456489531 | 0.456489531 | 0.456489531 | 0.50067834 | 0.754852219 | 1 | 0.500678344 | 0.500678344 | 0.500678344 | 0.911742111 | -0.040127986 | FALSE | TRUE | FALSE |
| COMMD3 | 0.4563717 | 0.74686164 | 1 | 0.456371651 | 0.456371651 | 0.456371651 | 0.50002431 | 0.755153819 | 1 | 0.500024307 | 0.500024307 | 0.500024307 | 0.912698933 | -0.039672457 | FALSE | TRUE | FALSE |
| CEACAM19 | 0.4563216 | 0.74689929 | 0.6921 | 4.264086422 | 401.5175009 | 401.5175009 | 0.57283785 | 0.72339659 | 0.87255 | 0.572837849 | 51.89293477 | 51.89293477 | 0.796598221 | -0.098760668 | FALSE | TRUE | FALSE |
| MANEA | 0.4562539 | 0.74693693 | 0.69475 | 0.456253916 | 7.302469358 | 7.302469358 | 103035.07 | 1.86792E-05 | 0.13515 | 4084.583989 | 1071583.871 | 11398498.72 | 4.42814E-06 | -5.353778465 | FALSE | FALSE | FALSE |
| RHOU | 0.456169 | 0.74697456 | 1 | 0.456168973 | 0.456168973 | 0.456168973 | 0.50061539 | 0.754889962 | 1 | 0.500615392 | 0.500615392 | 0.500615392 | 0.911216437 | -0.040378455 | FALSE | TRUE | FALSE |
| IGSF21 | 0.456048 | 0.74701218 | 1 | 0.456047997 | 0.456047997 | 8.999943359 | 0.50048286 | 0.754965411 | 1 | 0.500482861 | 0.500482861 | 9.892618988 | 0.911216013 | -0.040378657 | FALSE | TRUE | FALSE |
| C3orf70 | 0.4559461 | 0.74704979 | 1 | 0.455946091 | 0.455946091 | 0.455946091 | 0.49986029 | 0.755229097 | 1 | 0.49986029 | 0.49986029 | 0.49986029 | 0.912147054 | -0.03993514 | FALSE | TRUE | FALSE |
| ZNF367 | 0.455946 | 0.74708738 | 1 | 0.455946003 | 0.455946003 | 9.054376763 | 0.47162011 | 0.766579348 | 1 | 0.47162011 | 0.47162011 | 0.47162011 | 0.966765397 | -0.014678902 | FALSE | TRUE | FALSE |
| DEFB112 | 0.4558703 | 0.74712497 | 1 | 0.45587034 | 0.45587034 | 0.45587034 | 4.52094096 | 0.328352161 | 0.5009 | 4.520940959 | 4.520940959 | 4.520940959 | 0.100835278 | -0.996387499 | FALSE | FALSE | FALSE |
| LCE1B | 0.4557155 | 0.74716254 | 0.00375 | 0.455715524 | 0.455715524 | 0.455715524 | 0.48410313 | 0.761563072 | 1 | 0.484103132 | 4.511134313 | 4.511134313 | 0.941360412 | -0.026244069 | FALSE | TRUE | FALSE |
| GDF2 | 0.4554986 | 0.7472001 | 0.504 | 4.271254541 | 4.271254541 | 9.018851354 | 0.51567264 | 0.749050912 | 1 | 0.515672637 | 0.515672637 | 10.22257925 | 0.883309615 | -0.053887042 | FALSE | TRUE | FALSE |
| ST8SIA3 | 0.4554159 | 0.74723765 | 1 | 4.270279351 | 4.270279351 | 9.034832684 | 0.51559075 | 0.749130166 | 1 | 0.515590751 | 0.515590751 | 0.515590751 | 0.883289589 | -0.053896888 | FALSE | TRUE | FALSE |
| CPVL | 0.4554097 | 0.74727519 | 0.6269 | 0.455409663 | 16.51431603 | 16.51431603 | 1286.91944 | 0.00187754 | 0.3679 | 131.3684733 | 12907.90246 | 132485.7326 | 0.000353876 | -3.45114912 | FALSE | FALSE | FALSE |
| GIMAP5 | 0.4553419 | 0.74731271 | 1 | 0.455341949 | 0.455341949 | 0.455341949 | 0.46674443 | 0.768236424 | 1 | 0.466744431 | 0.466744431 | 0.466744431 | 0.975570179 | -0.010741483 | FALSE | TRUE | FALSE |
| FNDC7 | 0.4552461 | 0.74735023 | 0.9992 | 37321.8033 | 43618411.7 | 479578870.1 | 0.01122783 | 0.896352141 | 1 | 7600.711348 | 8604381.979 | 93688426.4 | 40.54621963 | 1.607950368 | FALSE | TRUE | FALSE |
| TTC38 | 0.4551838 | 0.74738773 | 0.2627 | 0.113842677 | 0.455183774 | 1.026699415 | 0.02229537 | 0.893135596 | 1 | 0.022295368 | 0.022295368 | 0.441424037 | 20.41606947 | 1.309972135 | FALSE | TRUE | FALSE |
| OR8B4 | 0.455121 | 0.74742522 | 1 | 0.455120968 | 0.455120968 | 0.455120968 | 0.48375034 | 0.761705447 | 1 | 4.523539117 | 4.523539117 | 4.523539117 | 0.940817876 | -0.026494439 | FALSE | TRUE | FALSE |
| AGGF1 | 0.4550449 | 0.7474627 | 0.5121 | 0.455044894 | 0.455044894 | 8.974061917 | 0.64938122 | 0.690086449 | 0.51285 | 0.649381215 | 0.649381215 | 2.396879359 | 0.700736152 | -0.154445476 | FALSE | TRUE | FALSE |
| GOT1 | 0.4549318 | 0.74750017 | 0.75765 | 0.454931755 | 1.819412869 | 3.854992735 | 0.14872592 | 0.864918167 | 1 | 0.148725917 | 0.594684133 | 0.594684133 | 3.058859978 | 0.485559597 | FALSE | TRUE | FALSE |
| ASB7 | 0.4547257 | 0.74753763 | 1 | 0.454725698 | 0.454725698 | 9.027309417 | 0.49921609 | 0.755379517 | 0.01 | 0.499216086 | 0.499216086 | 0.593417409 | 0.910879499 | -0.040539073 | FALSE | TRUE | FALSE |
| LUC7L3 | 0.4546718 | 0.74757507 | 0.01645 | 0.454671773 | 0.454671773 | 9.001830259 | 0.49910265 | 0.755454654 | 1 | 0.499102647 | 0.499102647 | 9.893470165 | 0.910978484 | -0.04049188 | FALSE | TRUE | FALSE |
| ATP6V1C1 | 0.4546273 | 0.74761251 | 1 | 0.454627277 | 0.454627277 | 0.454627277 | 0.49905928 | 0.755492203 | 1 | 0.499059283 | 0.499059283 | 0.499059283 | 0.91096848 | -0.040496649 | FALSE | TRUE | FALSE |
| FAM83B | 0.4544867 | 0.74764993 | 0.5175 | 0.454486663 | 0.454486663 | 8.926083107 | 0.07202156 | 0.88201643 | 1 | 0.644298806 | 0.644298806 | 12.68140968 | 6.310424815 | 0.800058597 | FALSE | TRUE | FALSE |
| RBFOX2 | 0.4544688 | 0.74768734 | 1 | 0.454468833 | 0.454468833 | 9.008269455 | 0.49891491 | 0.755567265 | 1 | 0.498914912 | 0.498914912 | 9.899872447 | 0.910914509 | -0.04052238 | FALSE | TRUE | FALSE |
| CXCL6 | 0.4543203 | 0.74772474 | 1 | 0.454320311 | 0.454320311 | 0.454320311 | 0.49802458 | 0.755904452 | 0.0049 | 0.498024581 | 0.498024581 | 0.498024581 | 0.912244755 | -0.039888625 | FALSE | TRUE | FALSE |
| TEX30 | 0.4541024 | 0.74776213 | 1 | 1.816332362 | 7.266836022 | 7.266836022 | 0.50940981 | 0.75187108 | 1 | 0.509409813 | 0.509409813 | 0.509409813 | 0.891428541 | -0.049913465 | FALSE | TRUE | FALSE |
| AP1G1 | 0.4539669 | 0.7477995 | 0.02415 | 0.45396687 | 0.45396687 | 8.935452788 | 0.49862075 | 0.755679769 | 0.02015 | 0.498620747 | 0.498620747 | 9.833099398 | 0.910445209 | -0.040746185 | FALSE | TRUE | FALSE |
| UNC5D | 0.4537589 | 0.74783687 | 0.8779 | 1.815230991 | 7.263521274 | 35.61388634 | 143.558839 | 0.019488135 | 0.238 | 35.8588901 | 461.2884399 | 1847.65929 | 0.003160787 | -2.500204822 | FALSE | FALSE | FALSE |

| | | | | | | | | | | | | | | | | | |
|---|---|---|---|---|---|---|---|---|---|---|---|---|---|---|---|---|---|
| RPLP0 | 0.4536703 | 0.74787423 | 1 | 0.453670332 | 0.453670332 | 0.453670332 | 0.49823798 | 0.755792164 | 1 | 0.498237985 | 0.498237985 | 0.498237985 | 0.910549467 | -0.040696455 | FALSE | TRUE | FALSE |
| OAS1 | 0.4536191 | 0.74791157 | 0.01205 | 0.453619065 | 0.453619065 | 1.006336723 | 0.0715863 | 0.882087328 | 1 | 0.644408997 | 0.644408997 | 1.421258618 | 6.336674616 | 0.801861407 | FALSE | TRUE | FALSE |
| SKAP2 | 0.4534945 | 0.74794891 | 1 | 0.453494523 | 0.453494523 | 0.453494523 | 4.51079164 | 0.329166955 | 0.50485 | 4.510791639 | 4.510791639 | 4.510791639 | 0.100535462 | -0.99768072 | FALSE | FALSE | FALSE |
| BTG4 | 0.4534839 | 0.74798623 | 0.0063 | 0.453483929 | 0.453483929 | 0.453483929 | 0.49788906 | 0.755941858 | 1 | 0.497889063 | 0.497889063 | 0.497889063 | 0.910813198 | -0.040570685 | FALSE | TRUE | FALSE |
| TAF3 | 0.4533717 | 0.74802354 | 0.50435 | 4.294582492 | 4.294582492 | 84.36094456 | 0.5141546 | 0.749959095 | 1 | 0.514154603 | 0.514154603 | 10.12150501 | 0.881780817 | -0.054639353 | FALSE | TRUE | FALSE |
| CPEB3 | 0.4533481 | 0.74806083 | 1 | 0.453348127 | 0.453348127 | 8.946365975 | 0.49783436 | 0.755979251 | 1 | 0.497834358 | 0.497834358 | 9.839980969 | 0.910640497 | -0.04065304 | FALSE | TRUE | FALSE |
| RTP3 | 0.4528872 | 0.74809812 | 1 | 0.452887203 | 0.452887203 | 0.452887203 | 4.50334869 | 0.329979521 | 0.5093 | 4.503348689 | 4.503348689 | 4.503348689 | 0.100566764 | -0.997545526 | FALSE | FALSE | FALSE |
| EEF1A2 | 0.4526018 | 0.7481354 | 1 | 0.452601788 | 0.452601788 | 8.944982633 | 0.46659916 | 0.76830354 | 1 | 4.417399742 | 42.92119498 | 42.92119498 | 0.970001287 | -0.013227689 | FALSE | TRUE | FALSE |
| SCYL3 | 0.4525431 | 0.74817267 | 0.75445 | 1.810026039 | 16.50505969 | 16.50505969 | 0.07252211 | 0.881901056 | 1 | 0.649184896 | 5.964433471 | 12.8376811 | 6.240071585 | 0.795189572 | FALSE | TRUE | FALSE |
| HNRNPD | 0.4524436 | 0.74820993 | 0.0079 | 0.45244362 | 0.45244362 | 0.45244362 | 0.49701336 | 0.75612872 | 1 | 0.497013364 | 0.497013364 | 0.497013364 | 0.910324858 | -0.040803598 | FALSE | TRUE | FALSE |
| KLHDC10 | 0.4524106 | 0.74824717 | 1 | 0.452410601 | 0.452410601 | 8.970843626 | 0.49701301 | 0.756166056 | 0.0126 | 0.497013012 | 0.497013012 | 9.865399119 | 0.910259068 | -0.040834986 | FALSE | TRUE | FALSE |
| PRG2 | 0.4524001 | 0.7482844 | 1 | 0.45240009 | 0.45240009 | 0.45240009 | 0.49646813 | 0.756315286 | 1 | 0.496468127 | 0.496468127 | 0.496468127 | 0.911236925 | -0.04036869 | FALSE | TRUE | FALSE |
| NSG1 | 0.4523032 | 0.74832163 | 1 | 0.452303193 | 0.452303193 | 0.452303193 | 0.49626163 | 0.756352565 | 1 | 0.496261632 | 0.496261632 | 0.496261632 | 0.911420839 | -0.040281046 | FALSE | TRUE | FALSE |
| SPARC | 0.452156 | 0.74835884 | 0.00915 | 0.452155957 | 0.452155957 | 0.452155957 | 0.49654514 | 0.756277996 | 1 | 0.496545142 | 0.496545142 | 0.496545142 | 0.91060393 | -0.04067048 | FALSE | TRUE | FALSE |
| CDX2 | 0.4518715 | 0.74839604 | 1 | 0.451871521 | 0.451871521 | 0.451871521 | 0.49659041 | 0.756240694 | 0.0087 | 0.496590412 | 0.496590412 | 0.496590412 | 0.909948139 | -0.040983359 | FALSE | TRUE | FALSE |
| BMP3 | 0.4517167 | 0.74843323 | 0.93835 | 1.806467504 | 28.91237211 | 28.91237211 | 5.49714779 | 0.286752463 | 0.2561 | 1.374842772 | 5.497147791 | 11.61404868 | 0.082172924 | -1.085271258 | FALSE | FALSE | FALSE |
| ZNF564 | 0.4515209 | 0.7484704 | 0.50165 | 0.451520852 | 0.451520852 | 1.004565746 | 0.07364018 | 0.881669683 | 1 | 0.073640183 | 0.073640183 | 1.460512757 | 6.131446602 | 0.78756295 | FALSE | TRUE | FALSE |
| ZBTB1 | 0.4514696 | 0.74850757 | 1 | 0.451469639 | 0.451469639 | 8.92992969 | 0.49602899 | 0.756427088 | 1 | 0.496028992 | 0.496028992 | 9.824301876 | 0.910167845 | -0.040878512 | FALSE | TRUE | FALSE |
| ATPAF1 | 0.4514591 | 0.74854473 | 1 | 0.451459111 | 0.451459111 | 0.451459111 | 0.49561054 | 0.756575992 | 1 | 0.495610539 | 0.495610539 | 0.495610539 | 0.910915074 | -0.040522111 | FALSE | TRUE | FALSE |
| TMEM174 | 0.4514212 | 0.74858187 | 1 | 0.451421205 | 0.451421205 | 0.451421205 | 0.49541418 | 0.756724703 | 1 | 0.495414177 | 0.495414177 | 0.495414177 | 0.911199611 | -0.040386474 | FALSE | TRUE | FALSE |
| MBNL1 | 0.4513678 | 0.748619 | 1 | 0.451367836 | 0.451367836 | 8.952825834 | 0.49575096 | 0.756501564 | 1 | 0.495750961 | 0.495750961 | 0.495750961 | 0.910472943 | -0.040732956 | FALSE | TRUE | FALSE |
| HDAC2 | 0.4513456 | 0.74865612 | 0.0135 | 0.451345621 | 0.451345621 | 8.950110682 | 0.49604364 | 0.756389833 | 0.012 | 0.496043638 | 0.496043638 | 9.846530242 | 0.909890959 | -0.04101065 | FALSE | TRUE | FALSE |
| IL12B | 0.4513153 | 0.74869323 | 1 | 0.451315338 | 0.451315338 | 0.451315338 | 0.49554681 | 0.756613188 | 1 | 0.495546814 | 0.495546814 | 0.495546814 | 0.910742084 | -0.040604595 | FALSE | TRUE | FALSE |
| CLK2 | 0.4513025 | 0.74873033 | 1 | 0.451302531 | 0.451302531 | 8.894505534 | 0.49596305 | 0.756464331 | 1 | 0.495963046 | 0.495963046 | 9.791843555 | 0.90995193 | -0.040981549 | FALSE | TRUE | FALSE |
| CTAGE6 | 0.4512838 | 0.74876741 | 1 | 40.97209999 | 4116.613738 | 4116.613738 | 4.51978706 | 0.328623974 | 0.96925 | 432.1601693 | 4386.675379 | 45610.90185 | 0.099846257 | -1.000668211 | FALSE | FALSE | FALSE |
| NUP62 | 0.4511849 | 0.74880449 | 1 | 0.45118494 | 0.45118494 | 8.910751507 | 0.49051058 | 0.758713815 | 1 | 0.490510581 | 0.490510581 | 9.702009819 | 0.919827129 | -0.036293786 | FALSE | TRUE | FALSE |
| PLCL2 | 0.4511261 | 0.74884155 | 0.0268 | 0.451126128 | 0.451126128 | 8.856371993 | 0.48041886 | 0.762978935 | 0.5137 | 4.54922258 | 4.54922258 | 89.50699652 | 0.939026688 | -0.027322064 | FALSE | TRUE | FALSE |
| ATG12 | 0.4509241 | 0.7488786 | 1 | 0.45092407 | 0.45092407 | 0.45092407 | 0.49494651 | 0.756873225 | 1 | 0.494946509 | 0.494946509 | 0.494946509 | 0.911056169 | -0.040454847 | FALSE | TRUE | FALSE |
| FABP7 | 0.4508759 | 0.74891564 | 1 | 0.450875914 | 0.450875914 | 0.450875914 | 41.4310207 | 0.066297245 | 0.25205 | 4.332356206 | 41.43102071 | 41.43102071 | 0.010882568 | -1.963268597 | FALSE | FALSE | FALSE |
| CARD8 | 0.4508144 | 0.74895267 | 0.74945 | 0.450814424 | 4.055015627 | 4.055015627 | 5.78011838 | 0.275408102 | 0.2554 | 0.63683351 | 5.780118381 | 5.780118381 | 0.077993978 | -1.10793893 | FALSE | FALSE | FALSE |
| WNT3A | 0.4504734 | 0.74898968 | 0.7518 | 0.450473371 | 1.801378157 | 8.899093528 | 0.14925903 | 0.864769941 | 1 | 0.149259032 | 0.149259032 | 2.952955942 | 3.018064398 | 0.479728502 | FALSE | TRUE | FALSE |
| SLITRK1 | 0.4503207 | 0.74902669 | 0.02305 | 0.450320738 | 0.450320738 | 8.867480865 | 0.49507655 | 0.756798988 | 1 | 0.495076548 | 0.495076548 | 9.766890507 | 0.909598201 | -0.041150407 | FALSE | TRUE | FALSE |
| INTS12 | 0.4502453 | 0.74906369 | 1 | 0.450245348 | 0.450245348 | 8.934354422 | 1.95712486 | 0.492808426 | 0.01085 | 1.957124856 | 1.957124856 | 9.718752111 | 0.230054483 | -0.638169299 | FALSE | FALSE | FALSE |
| OR7D2 | 0.4502111 | 0.74910067 | 0.00785 | 0.450211054 | 0.450211054 | 0.450211054 | 0.49462818 | 0.757021552 | 1 | 0.494628179 | 0.494628179 | 0.494628179 | 0.91020098 | -0.040862701 | FALSE | TRUE | FALSE |
| SNPH | 0.4501324 | 0.74913765 | 1 | 0.450132393 | 0.450132393 | 8.8937351 | 0.49492815 | 0.756910324 | 1 | 0.494928154 | 0.494928154 | 9.793045768 | 0.909490377 | -0.041201892 | FALSE | TRUE | FALSE |
| OR2F2 | 0.4500103 | 0.74917461 | 1 | 0.450010289 | 0.450010289 | 0.450010289 | 0.49418391 | 0.7572067 | 1 | 0.494183908 | 0.494183908 | 0.494183908 | 0.910612995 | -0.040666156 | FALSE | TRUE | FALSE |
| VGLL4 | 0.4499506 | 0.74921156 | 1 | 0.449950576 | 0.449950576 | 0.449950576 | 0.4941604 | 0.757243693 | 1 | 0.494160399 | 0.494160399 | 0.494160399 | 0.91053548 | -0.040703127 | FALSE | TRUE | FALSE |
| CCR4 | 0.4498116 | 0.7492485 | 1 | 0.44981164 | 0.44981164 | 0.44981164 | 0.49424935 | 0.757132678 | 1 | 0.494249346 | 0.494249346 | 0.494249346 | 0.910090511 | -0.040915414 | FALSE | TRUE | FALSE |
| ID4 | 0.4497883 | 0.74928543 | 0.5009 | 1.798958673 | 1.798958673 | 1.798958673 | 0.49992312 | 0.755191464 | 1 | 0.499923121 | 0.499923121 | 0.499923121 | 0.899714999 | -0.04589504 | FALSE | TRUE | FALSE |
| COBLL1 | 0.4496389 | 0.74932235 | 0.2604 | 0.049859306 | 0.449638945 | 1.681298971 | 0.09436195 | 0.877254835 | 0.0246 | 0.09436195 | 0.09436195 | 1.855076792 | 4.765045077 | 0.678067013 | FALSE | TRUE | FALSE |
| TMEM184B | 0.4495582 | 0.74935925 | 0.01055 | 0.449558183 | 0.449558183 | 8.90833777 | 0.49409164 | 0.757317642 | 1 | 0.49409164 | 0.49409164 | 0.49409164 | 0.909868021 | -0.041021599 | FALSE | TRUE | FALSE |
| ABHD17A | 0.4495417 | 0.74939615 | 0.0142 | 0.449541717 | 0.449541717 | 8.881971932 | 0.49409833 | 0.757280674 | 1 | 0.494098327 | 0.494098327 | 9.776536386 | 0.909822381 | -0.041043384 | FALSE | TRUE | FALSE |
| SLC17A7 | 0.4495336 | 0.74943303 | 1 | 4.239668989 | 4.239668989 | 15.22886123 | 0.51006155 | 0.751599819 | 0.0175 | 0.510061548 | 0.510061548 | 10.04706177 | 0.88133203 | -0.054860446 | FALSE | TRUE | FALSE |
| HECA | 0.4493869 | 0.7494699 | 1 | 0.449386889 | 0.449386889 | 8.877654977 | 0.49383082 | 0.757428477 | 1 | 0.49383082 | 0.49383082 | 9.770023534 | 0.910001706 | -0.040957793 | FALSE | TRUE | FALSE |
| DIRAS3 | 0.4493301 | 0.74950676 | 1 | 4.202186428 | 4.202186428 | 4.202186428 | 17.2944528 | 0.146984138 | 0.0016 | 0.509272733 | 0.509272733 | 0.509272733 | 0.025981166 | -1.585341354 | FALSE | FALSE | FALSE |
| NMU | 0.4493106 | 0.74954361 | 1 | 4.199025853 | 4.199025853 | 4.199025853 | 0.50917209 | 0.751987142 | 1 | 0.509172088 | 0.509172088 | 0.509172088 | 0.882433742 | -0.054317894 | FALSE | TRUE | FALSE |
| GNPNAT1 | 0.4493017 | 0.74958045 | 1 | 0.449301729 | 0.449301729 | 0.449301729 | 0.49349332 | 0.757576092 | 1 | 0.493493325 | 0.493493325 | 0.493493325 | 0.910451483 | -0.040743193 | FALSE | TRUE | FALSE |
| MANF | 0.4492468 | 0.74961727 | 1 | 0.449246831 | 0.449246831 | 0.449246831 | 0.47825664 | 0.763645387 | 1 | 4.47999864 | 4.47999864 | 4.47999864 | 0.939342587 | -0.027175988 | FALSE | TRUE | FALSE |
| FBXL15 | 0.4490403 | 0.74965409 | 1 | 0.44904028 | 0.44904028 | 8.890102757 | 0.49323617 | 0.757686676 | 1 | 0.493236174 | 0.493236174 | 9.776956914 | 0.91039608 | -0.040769621 | FALSE | TRUE | FALSE |
| PER2 | 0.4490372 | 0.74969089 | 0.51735 | 0.449037182 | 0.449037182 | 8.726972116 | 0.07295012 | 0.881785473 | 1 | 0.072950118 | 0.072950118 | 1.422604588 | 6.155400338 | 0.789256304 | FALSE | TRUE | FALSE |
| TERF2 | 0.4489492 | 0.74972768 | 1 | 4.215854621 | 40.63125123 | 83.46471042 | 0.52549516 | 0.744773467 | 1 | 0.525495158 | 0.525495158 | 10.41630732 | 0.854335562 | -0.068371516 | FALSE | TRUE | FALSE |
| RAD23A | 0.4489314 | 0.74976446 | 1 | 0.448931427 | 0.448931427 | 8.893297744 | 0.4936086 | 0.757465399 | 1 | 0.493608598 | 0.493608598 | 9.789527073 | 0.909488669 | -0.041202707 | FALSE | TRUE | FALSE |
| MTF1 | 0.4488719 | 0.74980123 | 1 | 0.448871906 | 0.448871906 | 8.851295163 | 0.4935553 | 0.757502309 | 0.0174 | 0.493555301 | 0.493555301 | 9.748863716 | 0.909466285 | -0.041213396 | FALSE | TRUE | FALSE |
| XXYLT1 | 0.4487447 | 0.74983799 | 1 | 0.448744735 | 0.448744735 | 8.89775951 | 0.49307562 | 0.757760342 | 1 | 0.493075625 | 0.493075625 | 0.493075625 | 0.910093123 | -0.040914167 | FALSE | TRUE | FALSE |
| LYN | 0.4483113 | 0.74987474 | 1 | 0.448311286 | 0.448311286 | 8.879019596 | 0.49309154 | 0.757723515 | 1 | 0.493091538 | 0.493091538 | 9.77733907 | 0.909184708 | -0.041347877 | FALSE | TRUE | FALSE |
| CRYGC | 0.4482899 | 0.74991147 | 0.7481 | 0.448289885 | 1.792908818 | 1.792908818 | 0.5559522 | 0.730548551 | 0.93875 | 5.160676436 | 82.5753438 | 82.5753438 | 0.806346098 | -0.093478511 | FALSE | TRUE | FALSE |
| ZNF830 | 0.4482301 | 0.7499482 | 1 | 0.448230053 | 0.448230053 | 8.878172163 | 0.49269402 | 0.757833961 | 0.0112 | 0.492694023 | 0.492694023 | 9.770194999 | 0.909753381 | -0.041076322 | FALSE | TRUE | FALSE |
| CKMT1B | 0.4479584 | 0.74998492 | 1 | 0.447958369 | 0.447958369 | 8.865653465 | 4.49180412 | 0.330789879 | 0.01415 | 4.49180412 | 4.49180412 | 9.45986361 | 0.099727939 | -1.001183154 | FALSE | FALSE | FALSE |
| KCTD15 | 0.4476307 | 0.75002162 | 1 | 0.447630671 | 0.447630671 | 0.447630671 | 0.49219117 | 0.757944311 | 1 | 0.492191165 | 0.492191165 | 0.492191165 | 0.909465066 | -0.041213978 | FALSE | TRUE | FALSE |
| KCNAB2 | 0.4475804 | 0.75005832 | 1 | 0.44758037 | 0.44758037 | 8.875958693 | 0.49207472 | 0.758017818 | 1 | 0.492074719 | 0.492074719 | 0.492074719 | 0.909578063 | -0.041160022 | FALSE | TRUE | FALSE |
| DUSP1 | 0.4473908 | 0.750095 | 1 | 0.447390796 | 0.447390796 | 0.447390796 | 0.49206093 | 0.758054553 | 1 | 0.492060933 | 0.492060933 | 0.492060933 | 0.909218282 | -0.04133184 | FALSE | TRUE | FALSE |
| NHLRC4 | 0.4473639 | 0.75013168 | 1 | 0.447363927 | 0.447363927 | 0.447363927 | 0.49160619 | 0.758091279 | 1 | 0.491606186 | 0.491606186 | 0.491606186 | 0.910004673 | -0.040956377 | FALSE | TRUE | FALSE |
| NDUFB5 | 0.4473368 | 0.75016834 | 1 | 0.447336756 | 0.447336756 | 0.447336756 | 0.49155773 | 0.758201386 | 1 | 0.491557729 | 0.491557729 | 0.491557729 | 0.910039104 | -0.040939946 | FALSE | TRUE | FALSE |
| TMEM201 | 0.4472965 | 0.75020499 | 1 | 0.447296494 | 0.447296494 | 8.854630483 | 0.47650777 | 0.764481619 | 0.5088 | 4.468068392 | 4.468068392 | 9.444474053 | 0.938697162 | -0.027474495 | FALSE | TRUE | FALSE |
| CENPL | 0.4472062 | 0.75024163 | 1 | 0.447206231 | 0.447206231 | 0.447206231 | 0.49157051 | 0.758127994 | 0.00735 | 0.491570508 | 0.491570508 | 0.491570508 | 0.909749923 | -0.041077973 | FALSE | TRUE | FALSE |
| INO80E | 0.4471632 | 0.75027826 | 1 | 4.167086535 | 4.167086535 | 4.167086535 | 4.44358333 | 0.333478518 | 0.751 | 4.443583335 | 42.55522222 | 42.55522222 | 0.10063121 | -0.997267305 | FALSE | FALSE | FALSE |
| COMMD1 | 0.4471398 | 0.75031488 | 1 | 0.447139782 | 0.447139782 | 0.447139782 | 0.49119553 | 0.758311386 | 1 | 0.491195527 | 0.491195527 | 0.491195527 | 0.910309149 | -0.040811092 | FALSE | TRUE | FALSE |
| WFDC6 | 0.4470848 | 0.75035148 | 1 | 0.447084813 | 0.447084813 | 0.447084813 | 0.45425351 | 0.773031346 | 1 | 0.454253505 | 0.454253505 | 0.454253505 | 0.98421874 | -0.00690837 | FALSE | TRUE | FALSE |
| RHOH | 0.4469104 | 0.75038808 | 0.0063 | 0.446910391 | 0.446910391 | 0.446910391 | 0.4915681 | 0.758164696 | 1 | 0.491568097 | 0.491568097 | 0.491568097 | 0.909152556 | -0.041363236 | FALSE | TRUE | FALSE |
| SPRYD7 | 0.4467957 | 0.75042466 | 1 | 0.446795691 | 0.446795691 | 0.446795691 | 0.49108315 | 0.75842128 | 0.0051 | 0.491083151 | 0.491083151 | 0.491083151 | 0.909816778 | -0.041046058 | FALSE | TRUE | FALSE |
| ACTA2 | 0.4467342 | 0.75046123 | 0.7542 | 0.446734234 | 1.786386015 | 2.214988079 | 11.6231586 | 0.180632264 | 0.00145 | 0.585758941 | 0.585758941 | 2.907210339 | 0.03843484 | -1.415274927 | FALSE | FALSE | FALSE |
| DCTN4 | 0.4466823 | 0.75049779 | 0.0117 | 0.446682275 | 0.446682275 | 8.855069035 | 0.49115905 | 0.75838466 | 1 | 0.491159048 | 0.491159048 | 9.747080657 | 0.909445273 | -0.04122343 | FALSE | TRUE | FALSE |
| R3HDM2 | 0.446613 | 0.75053434 | 1 | 0.446612953 | 0.446612953 | 8.819458834 | 0.49141831 | 0.758238064 | 1 | 0.491418314 | 0.491418314 | 9.718995643 | 0.908824397 | -0.041520023 | FALSE | TRUE | FALSE |

| | | | | | | | | | | | | | | | | | |
|---|---|---|---|---|---|---|---|---|---|---|---|---|---|---|---|---|---|
| TP53INP1 | 0.4465961 | 0.75057088 | 1 | 0.446596148 | 0.446596148 | 0.446596148 | 0.4908083 | 0.758494485 | 1 | 0.490808296 | 0.490808296 | 0.490808296 | 0.909919721 | -0.040996922 | FALSE | TRUE | FALSE |
| INSIG1 | 0.4465784 | 0.75060741 | 1 | 0.446578404 | 0.446578404 | 0.446578404 | 0.48702359 | 0.760201477 | 1 | 0.487023592 | 0.487023592 | 0.487023592 | 0.916954356 | -0.037652282 | FALSE | TRUE | FALSE |
| ELOVL4 | 0.4464566 | 0.75064392 | 1 | 0.446456634 | 0.446456634 | 0.446456634 | 4.45418832 | 0.332674485 | 0.4946 | 0.475710577 | 4.45418832 | 4.45418832 | 0.100232995 | -0.998989294 | FALSE | FALSE | FALSE |
| COA5 | 0.4461409 | 0.75068043 | 1 | 0.446140875 | 0.446140875 | 0.446140875 | 0.49041726 | 0.758750329 | 1 | 0.490417264 | 0.490417264 | 0.490417264 | 0.909716904 | -0.041093735 | FALSE | TRUE | FALSE |
| DEFA4 | 0.4459425 | 0.75071693 | 1 | 0.445942549 | 0.445942549 | 0.445942549 | 0.49018532 | 0.758786832 | 1 | 0.490185321 | 0.490185321 | 0.490185321 | 0.909742765 | -0.04108139 | FALSE | TRUE | FALSE |
| TXLNA | 0.4458607 | 0.75075341 | 0.01815 | 0.44586075 | 0.44586075 | 8.810753269 | 0.49061918 | 0.758640753 | 1 | 0.490619176 | 0.490619176 | 9.709161226 | 0.908771551 | -0.041545277 | FALSE | TRUE | FALSE |
| SP2 | 0.4458163 | 0.75078989 | 1 | 0.445816266 | 0.445816266 | 8.789559555 | 0.47667796 | 0.764412172 | 1 | 0.476677955 | 0.476677955 | 9.414098433 | 0.935256731 | -0.029069157 | FALSE | TRUE | FALSE |
| PID1 | 0.4457696 | 0.75082635 | 1 | 0.445769613 | 0.445769613 | 0.445769613 | 0.48993624 | 0.758859804 | 1 | 0.48993624 | 0.48993624 | 0.48993624 | 0.909852297 | -0.041029104 | FALSE | TRUE | FALSE |
| TMEM220 | 0.4457104 | 0.7508628 | 1 | 0.445710385 | 0.445710385 | 0.445710385 | 0.48972663 | 0.758932732 | 1 | 0.489726626 | 0.489726626 | 0.489726626 | 0.910120793 | -0.040900963 | FALSE | TRUE | FALSE |
| INTS6 | 0.44562 | 0.75089924 | 1 | 0.445619975 | 0.445619975 | 8.770995966 | 0.49054004 | 0.75867729 | 1 | 0.490540039 | 0.490540039 | 9.673583098 | 0.908427325 | -0.041709811 | FALSE | TRUE | FALSE |
| FBXO46 | 0.4456199 | 0.75093567 | 1 | 0.445619858 | 0.445619858 | 8.744358251 | 4.19542358 | 0.344056358 | 0.87775 | 40.49424901 | 400.8761722 | 400.8761722 | 0.106215701 | -0.973811278 | FALSE | FALSE | FALSE |
| SERTAD3 | 0.4455445 | 0.75097209 | 0.00605 | 0.445544543 | 0.445544543 | 0.445544543 | 0.48966051 | 0.758969178 | 1 | 0.489660508 | 0.489660508 | 0.489660508 | 0.909904997 | -0.04100395 | FALSE | TRUE | FALSE |
| VNN1 | 0.4455186 | 0.75100849 | 0.38035 | 0.195427944 | 1.780941113 | 1.780941113 | 0.32424406 | 0.817410823 | 0.6924 | 0.74260754 | 11.87197009 | 11.87197009 | 1.374022471 | 0.137993835 | FALSE | TRUE | FALSE |
| EPHA3 | 0.445356 | 0.75104489 | 0.1497 | 0.445355972 | 1.781236227 | 8.740131355 | 0.04392415 | 0.888118596 | 1 | 0.043924155 | 0.043924155 | 0.863959558 | 10.13920419 | 1.006003869 | FALSE | TRUE | FALSE |
| MRPS24 | 0.4453162 | 0.75108127 | 1 | 0.445316159 | 0.445316159 | 0.445316159 | 0.48959114 | 0.759042034 | 1 | 0.489591145 | 0.489591145 | 0.489591145 | 0.90956743 | -0.041165099 | FALSE | TRUE | FALSE |
| ZC2HC1A | 0.4451515 | 0.75111764 | 1 | 0.445151541 | 0.445151541 | 0.445151541 | 0.48960264 | 0.759005612 | 1 | 0.489602644 | 0.489602644 | 0.489602644 | 0.909209838 | -0.041335873 | FALSE | TRUE | FALSE |
| FCAMR | 0.4448747 | 0.75115401 | 0.87795 | 4.022564891 | 37.30734295 | 37.30734295 | 5.74985128 | 0.27602993 | 0.50525 | 5.749851277 | 53.77871724 | 53.77871724 | 0.077371521 | -1.111418866 | FALSE | FALSE | FALSE |
| DUSP14 | 0.4446567 | 0.75119036 | 1 | 0.444656711 | 0.444656711 | 0.444656711 | 151.475705 | 0.018524859 | 0.00065 | 4.46163289 | 4.46163289 | 4.46163289 | 0.002935499 | -2.532318131 | FALSE | FALSE | FALSE |
| ERF | 0.4446361 | 0.75122671 | 1 | 0.444636143 | 0.444636143 | 8.754714197 | 0.48940374 | 0.759114845 | 1 | 0.489403742 | 0.489403742 | 9.654171708 | 0.908526243 | -0.041662523 | FALSE | TRUE | FALSE |
| STRN4 | 0.4445909 | 0.75126304 | 0.8805 | 1.778017917 | 7.112466674 | 34.94278413 | 0.14916894 | 0.864792766 | 1 | 0.149168941 | 0.149168941 | 2.937696322 | 2.98045211 | 0.474282148 | FALSE | TRUE | FALSE |
| TUBB4A | 0.4444896 | 0.75129936 | 1 | 0.444489553 | 0.444489553 | 8.758983397 | 9.66194741 | 0.203022991 | 0.0163 | 0.489445203 | 0.489445203 | 9.661947412 | 0.046004137 | -1.337203112 | FALSE | FALSE | FALSE |
| JDP2 | 0.4441768 | 0.75133567 | 1 | 0.444176787 | 0.444176787 | 0.444176787 | 0.4885437 | 0.759405639 | 0.00595 | 0.488543695 | 0.488543695 | 0.488543695 | 0.909185383 | -0.041347555 | FALSE | TRUE | FALSE |
| PGM5 | 0.444151 | 0.75137197 | 0.0158 | 0.444150993 | 0.444150993 | 2.196074114 | 0.57572572 | 0.722271945 | 0.51055 | 0.575725723 | 0.575725723 | 2.850443573 | 0.771462825 | -0.112684997 | FALSE | TRUE | FALSE |
| KHDRBS2 | 0.4440476 | 0.75140826 | 1 | 0.44404758 | 0.44404758 | 0.44404758 | 0.4885068 | 0.759441935 | 1 | 0.488506805 | 0.488506805 | 0.488506805 | 0.908989548 | -0.04144111 | FALSE | TRUE | FALSE |
| FKBP8 | 0.4439871 | 0.75144454 | 1 | 0.443987139 | 0.443987139 | 8.779918943 | 0.48895127 | 0.759187612 | 1 | 0.488951271 | 0.488951271 | 9.682169145 | 0.908039646 | -0.041895189 | FALSE | TRUE | FALSE |
| SNRPA | 0.4439417 | 0.7514808 | 1 | 0.443941682 | 0.443941682 | 0.443941682 | 0.48830345 | 0.759587002 | 1 | 0.488303449 | 0.488303449 | 0.488303449 | 0.909151232 | -0.041363868 | FALSE | TRUE | FALSE |
| EEF1B2 | 0.4438867 | 0.75151706 | 1 | 0.443886724 | 0.443886724 | 0.443886724 | 29.8942197 | 0.092821537 | 0.3191 | 7.470349776 | 119.6580751 | 119.6580751 | 0.01484858 | -1.828315065 | FALSE | FALSE | FALSE |
| HOXA11 | 0.4438798 | 0.7515533 | 1 | 0.443879766 | 0.443879766 | 8.807967965 | 0.43074532 | 0.780814192 | 1 | 39.35281707 | 3978.703004 | 3978.703004 | 1.030492359 | 0.013044776 | FALSE | TRUE | FALSE |
| C19orf33 | 0.4438733 | 0.75158953 | 0.0024 | 0.443873291 | 0.443873291 | 0.443873291 | 0.48786612 | 0.759804258 | 0.0022 | 0.487866117 | 0.487866117 | 0.487866117 | 0.909826029 | -0.041041643 | FALSE | TRUE | FALSE |
| NDUFA7 | 0.4438291 | 0.75162576 | 1 | 0.443829108 | 0.443829108 | 0.443829108 | 0.48808504 | 0.759659467 | 1 | 0.488085039 | 0.488085039 | 0.488085039 | 0.909327417 | -0.041279714 | FALSE | TRUE | FALSE |
| LIN9 | 0.4438216 | 0.75166196 | 1 | 0.44382162 | 0.44382162 | 8.779509289 | 0.48876903 | 0.759260335 | 1 | 0.488769031 | 0.488769031 | 9.681342485 | 0.908039568 | -0.041895227 | FALSE | TRUE | FALSE |
| BRD3 | 0.4437353 | 0.75169816 | 1 | 0.443735315 | 0.443735315 | 8.704896752 | 4.45366254 | 0.332942706 | 0.02 | 4.453662543 | 4.453662543 | 16.70684331 | 0.099633798 | -1.001593314 | FALSE | FALSE | FALSE |
| CNOT4 | 0.4436224 | 0.75173435 | 1 | 0.443622406 | 1.774431662 | 15.01058592 | 0.49393271 | 0.7573546 | 0.01445 | 0.493932705 | 0.493932705 | 9.783132978 | 0.898143414 | -0.04665431 | FALSE | TRUE | FALSE |
| MOB3B | 0.443604 | 0.75177053 | 1 | 0.44360397 | 0.44360397 | 0.44360397 | 0.48849482 | 0.759478219 | 1 | 0.48849482 | 0.48849482 | 0.48849482 | 0.908103735 | -0.041864538 | FALSE | TRUE | FALSE |
| RAB36 | 0.4434533 | 0.75180669 | 0.01275 | 0.443453274 | 0.443453274 | 0.974588036 | 0.07196379 | 0.882034162 | 0.0112 | 0.071963785 | 0.071963785 | 1.426840208 | 6.162172705 | 0.789733866 | FALSE | TRUE | FALSE |
| NFKBID | 0.44343 | 0.75184285 | 1 | 0.443430003 | 0.443430003 | 8.797986432 | 0.4878919 | 0.759731887 | 1 | 0.487891902 | 0.487891902 | 0.487891902 | 0.908869364 | -0.041498536 | FALSE | TRUE | FALSE |
| KRTAP11-1 | 0.443318 | 0.75187899 | 1 | 0.443317962 | 0.443317962 | 0.443317962 | 0.48748964 | 0.760021094 | 1 | 0.487489645 | 0.487489645 | 0.487489645 | 0.909389496 | -0.041250066 | FALSE | TRUE | FALSE |
| PLK1 | 0.4433013 | 0.75191512 | 1 | 0.443301335 | 0.443301335 | 8.71560328 | 0.48813879 | 0.759623241 | 0.0204 | 0.488138791 | 0.488138791 | 9.61674073 | 0.90814609 | -0.041844282 | FALSE | TRUE | FALSE |
| IGFL3 | 0.4431074 | 0.75195125 | 1 | 0.443107413 | 0.443107413 | 0.443107413 | 0.48720389 | 0.760093281 | 1 | 0.487203889 | 0.487203889 | 0.487203889 | 0.909490713 | -0.041201731 | FALSE | TRUE | FALSE |
| SESN3 | 0.4430884 | 0.75198736 | 1 | 0.443088408 | 0.443088408 | 8.774709952 | 1.93067612 | 0.495559986 | 0.50595 | 1.930676122 | 1.930676122 | 9.569917527 | 0.229499088 | -0.639219037 | FALSE | FALSE | FALSE |
| FTH1 | 0.4430585 | 0.75202346 | 1 | 4.158539067 | 16.63270961 | 16.63270961 | 2.01281466 | 0.488874511 | 0.50485 | 2.012814662 | 2.012814662 | 2.012814662 | 0.220118896 | -0.657342673 | FALSE | FALSE | FALSE |
| NDEL1 | 0.442996 | 0.75205955 | 0.0099 | 0.442995951 | 0.442995951 | 0.442995951 | 4.43085801 | 0.334013627 | 0.0089 | 4.43085801 | 4.43085801 | 4.43085801 | 0.099979722 | -1.000088076 | FALSE | FALSE | FALSE |
| DCP1A | 0.4429738 | 0.75209562 | 1 | 0.442973849 | 0.442973849 | 8.767650065 | 0.487603 | 0.759948862 | 1 | 0.487602995 | 0.487602995 | 9.663014644 | 0.908472372 | -0.041688276 | FALSE | TRUE | FALSE |
| ZNF605 | 0.4427015 | 0.75213169 | 1 | 0.442701459 | 0.442701459 | 8.759507483 | 0.48752091 | 0.759984984 | 1 | 0.487520914 | 0.487520914 | 9.658715492 | 0.908066601 | -0.041882298 | FALSE | TRUE | FALSE |
| ZNF800 | 0.4426787 | 0.75216775 | 1 | 0.442678659 | 0.442678659 | 8.748248882 | 0.48765349 | 0.759912728 | 1 | 0.48765349 | 0.48765349 | 9.650838513 | 0.907772974 | -0.042022751 | FALSE | TRUE | FALSE |
| PAGR1 | 0.4423231 | 0.75220379 | 1 | 0.442323116 | 0.442323116 | 0.442323116 | 0.48667296 | 0.760417555 | 1 | 0.486672956 | 0.486672956 | 0.486672956 | 0.908871371 | -0.041497577 | FALSE | TRUE | FALSE |
| POP1 | 0.4420955 | 0.75223983 | 0.02945 | 0.110506545 | 0.442095493 | 3.723050054 | 0.04265997 | 0.888415955 | 0.0276 | 0.042659969 | 0.042659969 | 0.837665201 | 10.36323984 | 1.015495549 | FALSE | TRUE | FALSE |
| PDPN | 0.4420431 | 0.75227586 | 1 | 0.442043079 | 0.442043079 | 0.442043079 | 0.48643237 | 0.760489489 | 0.00665 | 0.486432367 | 0.486432367 | 0.486432367 | 0.9087452 | -0.04155787 | FALSE | TRUE | FALSE |
| OPA3 | 0.4419694 | 0.75231187 | 0.49615 | 0.193527205 | 1.767005413 | 1.767005413 | 0.28128474 | 0.830865384 | 0.74865 | 0.281284738 | 1.124926312 | 1.124926312 | 1.571252574 | 0.196246002 | FALSE | TRUE | FALSE |
| EIF4H | 0.4418991 | 0.75234788 | 1 | 0.441899063 | 0.441899063 | 0.441899063 | 0.48636658 | 0.760525439 | 1 | 0.486366581 | 0.486366581 | 0.486366581 | 0.908572013 | -0.041640645 | FALSE | TRUE | FALSE |
| BPHL | 0.4418282 | 0.75238387 | 0.49725 | 0.441828201 | 4.014595841 | 4.014595841 | 11.6945767 | 0.179611075 | 0.3802 | 4.994441247 | 187.1054228 | 187.1054228 | 0.037780607 | -1.422731074 | FALSE | FALSE | FALSE |
| SLAMF6 | 0.4417688 | 0.75241986 | 1 | 0.441768799 | 0.441768799 | 0.441768799 | 0.48618098 | 0.760669124 | 1 | 0.48618098 | 0.48618098 | 0.48618098 | 0.908650927 | -0.041602926 | FALSE | TRUE | FALSE |
| APOBEC3A | 0.4416714 | 0.75245583 | 1 | 0.441671353 | 4.189552292 | 4.189552292 | 0.43071436 | 0.780844097 | 1 | 0.430714361 | 0.430714361 | 0.430714361 | 1.025439114 | 0.010909879 | FALSE | TRUE | FALSE |
| SLC36A3 | 0.4416588 | 0.75249179 | 0.49925 | 0.04885713 | 0.441658817 | 0.968296402 | 0.06336408 | 0.883929692 | 1 | 0.063364079 | 0.063364079 | 1.257176129 | 6.970176537 | 0.843243778 | FALSE | TRUE | FALSE |
| CEP68 | 0.4415751 | 0.75252774 | 0.7527 | 0.441575145 | 1.766580786 | 8.696183692 | 1.25359513 | 0.560786795 | 0.7544 | 1.253595131 | 11.7480925 | 24.73327398 | 0.352247017 | -0.453152676 | FALSE | FALSE | FALSE |
| SIAH3 | 0.4415719 | 0.75256368 | 1 | 0.441571938 | 0.441571938 | 0.441571938 | 0.48601164 | 0.760740896 | 0.0077 | 0.486011641 | 0.486011641 | 0.486011641 | 0.908562473 | -0.041645205 | FALSE | TRUE | FALSE |
| OTOP2 | 0.4411272 | 0.75259961 | 0.7261 | 2.175194216 | 34.80252762 | 139.2611489 | 0.17632916 | 0.858014134 | 0.87585 | 0.705410024 | 2.822718953 | 5.972348404 | 2.501725424 | 0.398239642 | FALSE | TRUE | FALSE |
| MRAS | 0.4411174 | 0.75263553 | 1 | 0.441117363 | 0.441117363 | 0.441117363 | 1.92154824 | 0.497004873 | 0.50195 | 1.921548242 | 1.921548242 | 1.921548242 | 0.229563512 | -0.639097139 | FALSE | FALSE | FALSE |
| CD226 | 0.4411036 | 0.75267144 | 1 | 0.441103555 | 0.441103555 | 0.441103555 | 0.4856544 | 0.760812627 | 1 | 0.485654404 | 0.485654404 | 0.485654404 | 0.908266354 | -0.041786773 | FALSE | TRUE | FALSE |
| AMIGO1 | 0.4410096 | 0.75270733 | 1 | 0.441009646 | 0.441009646 | 8.725454068 | 0.4649492 | 0.768972458 | 1 | 0.464949203 | 0.464949203 | 9.210989134 | 0.948511456 | -0.022957419 | FALSE | TRUE | FALSE |
| VRTN | 0.440962 | 0.75274322 | 1 | 0.440962018 | 0.440962018 | 8.659310699 | 0.48588148 | 0.760776767 | 1 | 0.485881483 | 0.485881483 | 9.562241926 | 0.907550572 | -0.042129165 | FALSE | TRUE | FALSE |
| NAP1L1 | 0.4409033 | 0.75277909 | 1 | 0.440903264 | 0.440903264 | 0.440903264 | 0.48543803 | 0.760955951 | 1 | 0.485438026 | 0.485438026 | 0.485438026 | 0.908258604 | -0.041790479 | FALSE | TRUE | FALSE |
| SATB2 | 0.4407381 | 0.75281496 | 0.74785 | 4.167312665 | 40.4403194 | 82.12655931 | 17.5118512 | 0.145404927 | 0.00485 | 0.517616928 | 0.517616928 | 10.21880855 | 0.025167989 | -1.599151478 | FALSE | FALSE | FALSE |
| MEPCE | 0.4406555 | 0.75285081 | 1 | 0.440655514 | 0.440655514 | 8.682609614 | 0.48558093 | 0.760848475 | 0.0186 | 0.485580933 | 0.485580933 | 9.584865207 | 0.907481089 | -0.042162416 | FALSE | TRUE | FALSE |
| FNTA | 0.4405753 | 0.75288666 | 1 | 0.440575287 | 0.440575287 | 0.440575287 | 0.48522587 | 0.761063324 | 0.0076 | 0.485225873 | 0.485225873 | 0.485225873 | 0.90797979 | -0.041923818 | FALSE | TRUE | FALSE |
| NHLH1 | 0.4405492 | 0.75292249 | 0.0067 | 0.440549182 | 0.440549182 | 0.440549182 | 0.48553975 | 0.760884312 | 1 | 0.485539749 | 0.485539749 | 0.485539749 | 0.907339065 | -0.04223039 | FALSE | TRUE | FALSE |
| MAPK11 | 0.4404775 | 0.75295831 | 0.01195 | 0.440477456 | 0.440477456 | 8.735405386 | 4.40818089 | 0.335081215 | 0.5057 | 4.408180891 | 4.408180891 | 4.457419725 | 0.099922727 | -1.000335722 | FALSE | FALSE | FALSE |
| SPECC1L | 0.440474 | 0.75299412 | 0.03305 | 0.440474001 | 0.440474001 | 8.606876822 | 8.93666646 | 0.21247362 | 0.1463 | 2.233381854 | 8.936666464 | 43.76127439 | 0.049288401 | -1.30725527 | FALSE | FALSE | FALSE |
| PPP2R1B | 0.4403395 | 0.75302992 | 0.9882 | 339.824729 | 324447.9223 | 3341228.971 | 0.11215427 | 0.872893566 | 0.87675 | 0.99648585 | 9.081429422 | 9.081429422 | 3.926194691 | 0.593971831 | FALSE | TRUE | FALSE |
| CENPB | 0.440307 | 0.75306571 | 1 | 0.44030705 | 0.44030705 | 8.649197895 | 3.20789245 | 0.400230107 | 0.9996 | 333116.6626 | 39843963.5 | 450484805.4 | 0.137257423 | -0.862464161 | FALSE | FALSE | FALSE |
| CPA2 | 0.4402183 | 0.75310148 | 0.511 | 0.440218288 | 0.440218288 | 0.992076175 | 0.62458889 | 0.701334814 | 0.50915 | 0.624588885 | 0.624588885 | 1.396956907 | 0.704812876 | -0.151926171 | FALSE | TRUE | FALSE |
| TREML4 | 0.4402039 | 0.75313725 | 1 | 0.44020393 | 0.44020393 | 0.44020393 | 0.4843436 | 0.761349168 | 1 | 0.484343599 | 0.484343599 | 0.484343599 | 0.908867033 | -0.041499649 | FALSE | TRUE | FALSE |
| PSMB4 | 0.4401085 | 0.75317301 | 1 | 0.44010851 | 0.44010851 | 8.736610944 | 0.48463402 | 0.761277773 | 1 | 0.484634025 | 0.484634025 | 0.484634025 | 0.908125486 | -0.041854136 | FALSE | TRUE | FALSE |

| | | | | | | | | | | | | | | | | | |
|---|---|---|---|---|---|---|---|---|---|---|---|---|---|---|---|---|---|
| SRRD | 0.4399819 | 0.75320875 | 0.9395 | 3.976284888 | 350.2431363 | 350.2431363 | 0.66346361 | 0.683763174 | 0.7492 | 0.663463613 | 6.057645813 | 6.057645813 | 0.663159002 | -0.178382331 | FALSE | TRUE | FALSE |
| CSNK1G3 | 0.4399801 | 0.75324449 | 1 | 0.439980143 | 0.439980143 | 8.727552086 | 0.48475339 | 0.761206336 | 0.01125 | 0.484753386 | 0.484753386 | 9.62515356 | 0.907637069 | -0.042087775 | FALSE | TRUE | FALSE |
| RELL2 | 0.4398958 | 0.75328021 | 1 | 0.439895786 | 0.439895786 | 0.439895786 | 0.48431338 | 0.761420516 | 1 | 0.484313376 | 0.484313376 | 0.484313376 | 0.908287502 | -0.041776662 | FALSE | TRUE | FALSE |
| C2orf88 | 0.4398504 | 0.75331592 | 1 | 0.439850351 | 0.439850351 | 0.439850351 | 0.48403698 | 0.761634282 | 1 | 0.484036985 | 0.484036985 | 0.484036985 | 0.908712277 | -0.041573605 | FALSE | TRUE | FALSE |
| PEBP4 | 0.4397592 | 0.75335162 | 0.5044 | 0.439759191 | 0.439759191 | 0.439759191 | 0.14326617 | 0.866014691 | 1 | 0.143266167 | 0.143266167 | 0.143266167 | 3.06952576 | 0.487071282 | FALSE | TRUE | FALSE |
| NUP50 | 0.4396768 | 0.75338731 | 1 | 0.439676787 | 0.439676787 | 8.71494801 | 0.48434184 | 0.761384848 | 1 | 0.484341844 | 0.484341844 | 9.610584895 | 0.907781958 | -0.042018453 | FALSE | TRUE | FALSE |
| MTHFD1L | 0.4395492 | 0.75342299 | 0.51425 | 0.439549227 | 0.439549227 | 8.624615588 | 0.07134816 | 0.882140453 | 1 | 0.071348157 | 0.071348157 | 1.403148313 | 6.160624816 | 0.789624761 | FALSE | TRUE | FALSE |
| TPD52 | 0.439273 | 0.75345866 | 1 | 0.439273001 | 0.439273001 | 0.439273001 | 0.48360601 | 0.761741014 | 0.0052 | 0.483606009 | 0.483606009 | 0.483606009 | 0.908328253 | -0.041757177 | FALSE | TRUE | FALSE |
| SRM | 0.4392609 | 0.75349432 | 1 | 0.43926086 | 0.43926086 | 8.716454214 | 0.4788227 | 0.763400305 | 1 | 1.915082538 | 1.915082538 | 1.915082538 | 0.917376857 | -0.03745222 | FALSE | TRUE | FALSE |
| NGB | 0.4392526 | 0.75352997 | 1 | 0.439252618 | 0.439252618 | 0.439252618 | 1.91460311 | 0.497644339 | 0.49765 | 0.478718403 | 1.914603114 | 1.914603114 | 0.229422283 | -0.639364403 | FALSE | FALSE | FALSE |
| DNAJB8 | 0.4392021 | 0.75356561 | 0.9359 | 39.91044233 | 4013.609762 | 4013.609762 | 0.515503 | 0.749248946 | 0.75 | 4.864858158 | 47.11864545 | 47.11864545 | 0.851987393 | -0.069566832 | FALSE | TRUE | FALSE |
| RPL3 | 0.4391718 | 0.75360124 | 1 | 0.439171787 | 0.439171787 | 8.695701033 | 0.4842934 | 0.761491817 | 1 | 0.484293395 | 0.484293395 | 9.600640675 | 0.906830015 | -0.042474114 | FALSE | TRUE | FALSE |
| CDC27 | 0.4391056 | 0.75363685 | 1 | 0.439105648 | 0.439105648 | 8.661358698 | 1.91462925 | 0.497484614 | 0.5068 | 1.91462925 | 1.91462925 | 37.82855346 | 0.22934239 | -0.639515666 | FALSE | FALSE | FALSE |
| NTMT1 | 0.4390962 | 0.75367246 | 1 | 0.439096231 | 0.439096231 | 0.439096231 | 0.48354077 | 0.76177657 | 0.0073 | 0.483540768 | 0.483540768 | 0.483540768 | 0.908085233 | -0.041873387 | FALSE | TRUE | FALSE |
| GALK2 | 0.4390192 | 0.75370805 | 0.8183 | 4.134742247 | 39.91737726 | 394.7896261 | 0.01110406 | 0.896429218 | 0.5048 | 0.100366175 | 0.100366175 | 0.220478141 | 39.5368128 | 1.597001656 | FALSE | TRUE | FALSE |
| HOOK3 | 0.438719 | 0.75374364 | 1 | 1.754851466 | 1.754851466 | 14.82074318 | 0.48910487 | 0.759151234 | 1 | 0.489104868 | 0.489104868 | 9.678738467 | 0.896983416 | -0.047215587 | FALSE | TRUE | FALSE |
| TTLL2 | 0.4386282 | 0.75377921 | 0.75115 | 0.438628175 | 3.964131169 | 8.665039498 | 5.33211394 | 0.292455481 | 0.69145 | 5.33211394 | 476.0823924 | 476.0823924 | 0.082261591 | -1.084802896 | FALSE | FALSE | FALSE |
| ALKBH6 | 0.438548 | 0.75381478 | 1 | 0.438548026 | 0.438548026 | 0.438548026 | 0.48298351 | 0.761918689 | 1 | 0.482983515 | 0.482983515 | 0.482983515 | 0.907997918 | -0.041915147 | FALSE | TRUE | FALSE |
| MLXIPL | 0.4384311 | 0.75385033 | 0.75465 | 0.438431105 | 1.753642043 | 8.596352679 | 0.14550105 | 0.865530625 | 1 | 0.145501055 | 0.145501055 | 2.859600197 | 3.013250349 | 0.479035216 | FALSE | TRUE | FALSE |
| COPS4 | 0.4383305 | 0.75388587 | 1 | 0.43833046 | 0.43833046 | 8.699432008 | 0.48242148 | 0.762166971 | 1 | 0.482421477 | 0.482421477 | 9.583317957 | 0.908604781 | -0.041624982 | FALSE | TRUE | FALSE |
| MAGEL2 | 0.4382043 | 0.7539214 | 1 | 4.124053163 | 4.124053163 | 14.78726292 | 0.49865935 | 0.75564228 | 1 | 0.498659354 | 0.498659354 | 9.857328243 | 0.878764921 | -0.056127288 | FALSE | TRUE | FALSE |
| VIM | 0.4381328 | 0.75395693 | 0.50535 | 0.438132848 | 1.751610754 | 2.163432447 | 0.04345154 | 0.888191058 | 1 | 0.043451543 | 0.043451543 | 0.858924692 | 10.08325167 | 1.003600607 | FALSE | TRUE | FALSE |
| SLC25A51 | 0.4381283 | 0.75399244 | 1 | 0.438128303 | 0.438128303 | 0.438128303 | 0.48299069 | 0.761883176 | 0.0077 | 0.48299069 | 0.48299069 | 0.48299069 | 0.907115421 | -0.04233745 | FALSE | TRUE | FALSE |
| VAPA | 0.4379397 | 0.75402794 | 1 | 0.437939694 | 0.437939694 | 0.437939694 | 0.48256762 | 0.762096089 | 0.00575 | 0.482567619 | 0.482567619 | 0.482567619 | 0.907519851 | -0.042143866 | FALSE | TRUE | FALSE |
| EGLN2 | 0.4377319 | 0.75406343 | 1 | 0.437731894 | 0.437731894 | 8.649437258 | 0.48271949 | 0.762025163 | 1 | 0.482719492 | 0.482719492 | 9.552167993 | 0.90680385 | -0.042486645 | FALSE | TRUE | FALSE |
| GATM | 0.43767 | 0.75409891 | 1 | 0.437670003 | 0.437670003 | 8.679090865 | 0.48248046 | 0.762131536 | 1 | 0.482480463 | 0.482480463 | 9.577460306 | 0.90712482 | -0.04233295 | FALSE | TRUE | FALSE |
| FASLG | 0.4376337 | 0.75413438 | 1 | 0.437633704 | 0.437633704 | 0.437633704 | 0.4826408 | 0.762060632 | 1 | 0.482640805 | 0.482640805 | 0.482640805 | 0.906748248 | -0.042513275 | FALSE | TRUE | FALSE |
| TMEM74 | 0.4375577 | 0.75416984 | 1 | 0.43755765 | 0.43755765 | 0.43755765 | 0.48194567 | 0.762414703 | 1 | 0.481945673 | 0.481945673 | 0.481945673 | 0.907898286 | -0.041962804 | FALSE | TRUE | FALSE |
| B4GALT1 | 0.4374736 | 0.75420528 | 1 | 1.749764688 | 1.749764688 | 8.655636633 | 0.48785181 | 0.759840426 | 0.0116 | 0.487851808 | 0.487851808 | 9.664848945 | 0.896734536 | -0.047336104 | FALSE | TRUE | FALSE |
| KCNK5 | 0.4372746 | 0.75424072 | 1 | 0.437274594 | 0.437274594 | 8.638748101 | 0.48220757 | 0.76227321 | 0.01355 | 0.482207565 | 0.482207565 | 9.540429268 | 0.906818196 | -0.042479774 | FALSE | TRUE | FALSE |
| PHACTR3 | 0.4372429 | 0.75427615 | 1 | 0.437242927 | 0.437242927 | 8.638946315 | 0.48212396 | 0.762343979 | 1 | 0.48212396 | 0.48212396 | 9.539576744 | 0.906909764 | -0.042435922 | FALSE | TRUE | FALSE |
| OR10G2 | 0.4371448 | 0.75431157 | 1 | 0.437144822 | 0.437144822 | 0.437144822 | 0.48171395 | 0.762485383 | 1 | 0.481713955 | 0.481713955 | 0.481713955 | 0.907478011 | -0.042163889 | FALSE | TRUE | FALSE |
| HTR1B | 0.4370632 | 0.75434697 | 1 | 0.437063197 | 0.437063197 | 8.648208251 | 0.48214127 | 0.7623086 | 0.01195 | 0.482141267 | 0.482141267 | 9.55238959 | 0.906504435 | -0.042630067 | FALSE | TRUE | FALSE |
| RALGPS1 | 0.4369271 | 0.75438237 | 1 | 0.436927096 | 0.436927096 | 8.641195159 | 0.4819262 | 0.762450048 | 1 | 0.4819262 | 0.4819262 | 9.543925109 | 0.906626566 | -0.042571559 | FALSE | TRUE | FALSE |
| RNF130 | 0.436896 | 0.75441776 | 1 | 0.436895954 | 0.436895954 | 8.653473499 | 0.47002051 | 0.766987434 | 1 | 0.470020514 | 0.470020514 | 0.470020514 | 0.929525289 | -0.03173879 | FALSE | TRUE | FALSE |
| SUN1 | 0.4368292 | 0.75445313 | 0.27205 | 0.436829223 | 4.000469989 | 8.561397289 | 5.56773268 | 0.284935978 | 0.50755 | 5.567732685 | 52.62345493 | 109.3852811 | 0.078457291 | -1.105366691 | FALSE | FALSE | FALSE |
| CCT4 | 0.4367594 | 0.75448849 | 1 | 1.79173E+11 | 4.31154E+15 | 2.25201E+17 | 22296.2494 | 8.32894E-05 | 0.9999 | 1.14882E+15 | 1.45898E+20 | 1.98485E+22 | 1.95889E-05 | -4.707989512 | FALSE | FALSE | FALSE |
| NBL1 | 0.4364921 | 0.75452385 | 1 | 390.3136925 | 435933.483 | 435933.483 | 0.37273832 | 0.801444282 | 1 | 3924765.971 | 5676602696 | 67000376324 | 1.171041564 | 0.06857231 | FALSE | TRUE | FALSE |
| SLC39A3 | 0.436361 | 0.75455919 | 1 | 0.436361009 | 0.436361009 | 0.436361009 | 0.48086951 | 0.762802917 | 1 | 0.480869509 | 0.480869509 | 0.480869509 | 0.907441626 | -0.042181303 | FALSE | TRUE | FALSE |
| GMDS | 0.4363192 | 0.75459453 | 0.50315 | 0.436319151 | 0.436319151 | 2.16572493 | 0.12101393 | 0.871048676 | 0.99585 | 173.1012249 | 17136.80435 | 68552.22251 | 3.605528351 | 0.556968915 | FALSE | TRUE | FALSE |
| PIK3IP1 | 0.4362792 | 0.75462985 | 1 | 0.436279177 | 0.436279177 | 0.436279177 | 0.48071221 | 0.76290856 | 1 | 0.480712206 | 0.480712206 | 0.480712206 | 0.907568337 | -0.042120664 | FALSE | TRUE | FALSE |
| PHYHIP | 0.4362559 | 0.75466516 | 1 | 0.436255926 | 0.436255926 | 8.647840506 | 0.45380574 | 0.773127716 | 1 | 0.453805743 | 0.453805743 | 0.453805743 | 0.961327467 | -0.017128649 | FALSE | TRUE | FALSE |
| OR2T6 | 0.4362552 | 0.75470046 | 0.8714 | 39.23581958 | 386.6334508 | 386.6334508 | 0.46612127 | 0.768538126 | 1 | 41.98742911 | 4185.593544 | 4185.593544 | 0.93592633 | -0.028758335 | FALSE | TRUE | FALSE |
| YIPF3 | 0.4362278 | 0.75473575 | 1 | 4.080806536 | 39.18018041 | 39.18018041 | 0.51200214 | 0.750743216 | 1 | 0.512002143 | 0.512002143 | 0.512002143 | 0.852003935 | -0.0695584 | FALSE | TRUE | FALSE |
| OR10A5 | 0.4360865 | 0.75477103 | 1 | 0.436086485 | 0.436086485 | 0.436086485 | 0.48072823 | 0.762873357 | 1 | 0.480728225 | 0.480728225 | 0.480728225 | 0.907137259 | -0.042326995 | FALSE | TRUE | FALSE |
| RBMS1 | 0.4359911 | 0.7548063 | 1 | 0.435991124 | 0.435991124 | 8.652633203 | 0.48087318 | 0.76276768 | 0.00915 | 0.48087318 | 0.48087318 | 0.48087318 | 0.906665503 | -0.042552908 | FALSE | TRUE | FALSE |
| BEGAIN | 0.4359505 | 0.75484156 | 1 | 0.43595048 | 0.43595048 | 8.567400376 | 0.48098931 | 0.762697172 | 1 | 0.480989314 | 0.480989314 | 9.472302762 | 0.906362091 | -0.042698267 | FALSE | TRUE | FALSE |
| MAZ | 0.4357032 | 0.75487681 | 1 | 4.087450078 | 4.087450078 | 8.607960874 | 42.0702742 | 0.065218676 | 0.2554 | 4.367242987 | 42.07027415 | 42.07027415 | 0.010356557 | -1.984784597 | FALSE | FALSE | FALSE |
| TAC4 | 0.4354114 | 0.75491205 | 1 | 0.435411394 | 0.435411394 | 0.435411394 | 0.47966943 | 0.763154693 | 1 | 0.47966943 | 0.47966943 | 0.47966943 | 0.907732215 | -0.042042251 | FALSE | TRUE | FALSE |
| DEGS1 | 0.4353652 | 0.75494728 | 0.75505 | 1.741380523 | 6.966942466 | 6.966942466 | 4.45661611 | 0.332405973 | 0.0068 | 4.45661611 | 4.45661611 | 4.45661611 | 0.097689632 | -1.010151524 | FALSE | FALSE | FALSE |
| MEIS1 | 0.4353558 | 0.7549825 | 1 | 0.435355751 | 0.435355751 | 8.62833735 | 0.48054249 | 0.762943753 | 1 | 0.480542487 | 0.480542487 | 0.480542487 | 0.90596724 | -0.042887506 | FALSE | TRUE | FALSE |
| UQCRB | 0.4348589 | 0.75501771 | 1 | 0.434858945 | 0.434858945 | 0.434858945 | 0.46461236 | 0.769139059 | 1 | 4.34011896 | 4.34011896 | 4.34011896 | 0.935960768 | -0.028742355 | FALSE | TRUE | FALSE |
| PLA2G3 | 0.4348501 | 0.75505291 | 0.7493 | 0.434850123 | 3.942434716 | 8.600724899 | 0.07370219 | 0.881642934 | 1 | 0.073702189 | 0.073702189 | 1.459667215 | 5.900097788 | 0.77085921 | FALSE | TRUE | FALSE |
| C1orf43 | 0.4347436 | 0.7550881 | 0.0083 | 0.434743573 | 0.434743573 | 0.434743573 | 0.47929406 | 0.763295108 | 1 | 0.479294061 | 0.479294061 | 0.479294061 | 0.907049782 | -0.042368877 | FALSE | TRUE | FALSE |
| DEGS2 | 0.4347252 | 0.75512328 | 1 | 4.072749387 | 39.16045239 | 39.16045239 | 0.51063067 | 0.751289041 | 1 | 0.510630673 | 0.510630673 | 10.1287319 | 0.851349623 | -0.069892052 | FALSE | TRUE | FALSE |
| TMC5 | 0.4346653 | 0.75515845 | 0.6324 | 0.434665308 | 15.67229413 | 15.67229413 | 0.18964743 | 0.854420712 | 0.2712 | 0.18964743 | 0.758351922 | 3.730425761 | 2.291965195 | 0.360208018 | FALSE | TRUE | FALSE |
| HMGB2 | 0.4346382 | 0.75519361 | 1 | 0.434638213 | 0.434638213 | 0.434638213 | 0.4797674 | 0.763084421 | 1 | 0.479767397 | 0.479767397 | 0.479767397 | 0.905935284 | -0.042902825 | FALSE | TRUE | FALSE |
| TMEM140 | 0.4344264 | 0.75522876 | 1 | 0.434426355 | 0.434426355 | 0.434426355 | 0.47881228 | 0.763435349 | 0.005 | 0.478812276 | 0.478812276 | 0.478812276 | 0.907299952 | -0.042249112 | FALSE | TRUE | FALSE |
| KRTAP27-1 | 0.4344044 | 0.75526389 | 1 | 4.041656786 | 38.59355779 | 38.59355779 | 0.49393115 | 0.757391544 | 1 | 4.61064211 | 4.61064211 | 4.61064211 | 0.879483626 | -0.055772242 | FALSE | TRUE | FALSE |
| INPP5A | 0.4341551 | 0.75529902 | 1 | 0.434155133 | 0.434155133 | 8.595235644 | 1.8954059 | 0.499553376 | 0.5101 | 1.895405904 | 1.895405904 | 9.393196094 | 0.229056548 | -0.640057289 | FALSE | FALSE | FALSE |
| BACE1 | 0.4341127 | 0.75533414 | 1 | 4.076611162 | 4.076611162 | 14.70525376 | 0.49440762 | 0.757058606 | 1 | 0.494407625 | 0.494407625 | 9.794333691 | 0.878046098 | -0.056482683 | FALSE | TRUE | FALSE |
| AMIGO2 | 0.4340583 | 0.75536925 | 1 | 0.434058311 | 0.434058311 | 8.585333435 | 0.47915985 | 0.763330184 | 1 | 0.479159848 | 0.479159848 | 9.489993035 | 0.905873714 | -0.042932342 | FALSE | TRUE | FALSE |
| ZKSCAN1 | 0.4338619 | 0.75540434 | 0.01675 | 0.43386189 | 0.43386189 | 8.572829315 | 0.4788032 | 0.763470382 | 1 | 0.478803195 | 0.478803195 | 9.474537048 | 0.906138251 | -0.042805536 | FALSE | TRUE | FALSE |
| GATAD1 | 0.4337546 | 0.75543943 | 1 | 0.433754561 | 0.433754561 | 0.433754561 | 0.4787371 | 0.763505404 | 1 | 0.4787371 | 0.4787371 | 0.4787371 | 0.906039163 | -0.04285303 | FALSE | TRUE | FALSE |
| POLR2J | 0.4336786 | 0.75547451 | 0.00405 | 0.433678644 | 0.433678644 | 0.433678644 | 0.47855671 | 0.763540416 | 1 | 0.478556708 | 0.478556708 | 0.478556708 | 0.906222056 | -0.042765372 | FALSE | TRUE | FALSE |
| ART1 | 0.4336177 | 0.75550957 | 0.5012 | 0.433617729 | 4.026691351 | 4.026691351 | 5.3265006 | 0.293051634 | 0.1902 | 2.120258283 | 51.03700792 | 500.9774955 | 0.081407618 | -1.08933495 | FALSE | FALSE | FALSE |
| VCPKMT | 0.4334718 | 0.75554463 | 1 | 0.43347178 | 0.43347178 | 0.43347178 | 0.47817064 | 0.763680356 | 1 | 0.478170642 | 0.478170642 | 0.478170642 | 0.906521108 | -0.042622079 | FALSE | TRUE | FALSE |
| YTHDF1 | 0.4334319 | 0.75557967 | 1 | 0.433431868 | 0.433431868 | 8.553325308 | 0.47843402 | 0.763575417 | 1 | 0.478434017 | 0.478434017 | 9.456509004 | 0.905938651 | -0.042901211 | FALSE | TRUE | FALSE |
| SYNPR | 0.4332136 | 0.75561471 | 1 | 0.433213579 | 0.433213579 | 0.433213579 | 0.47814823 | 0.763715313 | 1 | 0.478148233 | 0.478148233 | 0.478148233 | 0.906023591 | -0.042860494 | FALSE | TRUE | FALSE |
| CD74 | 0.4332096 | 0.75564973 | 1 | 0.433209561 | 0.433209561 | 0.433209561 | 0.47806671 | 0.763785194 | 1 | 0.478066706 | 0.478066706 | 0.478066706 | 0.906169694 | -0.042790466 | FALSE | TRUE | FALSE |
| EXOC5 | 0.43307 | 0.75568475 | 1 | 0.433070003 | 0.433070003 | 8.571595203 | 0.47809716 | 0.763750259 | 0.01415 | 0.478097161 | 0.478097161 | 9.474603756 | 0.905820067 | -0.042958062 | FALSE | TRUE | FALSE |
| CASP3 | 0.4329734 | 0.75571975 | 1 | 0.432973358 | 0.432973358 | 0.432973358 | 0.477737 | 0.763855033 | 1 | 0.477737 | 0.477737 | 0.477737 | 0.90630066 | -0.042727704 | FALSE | TRUE | FALSE |
| IKZF5 | 0.4329447 | 0.75575475 | 1 | 0.432944664 | 0.432944664 | 8.588830311 | 0.4730491 | 0.765861619 | 1 | 0.473049098 | 0.473049098 | 9.393560828 | 0.915221412 | -0.038473828 | FALSE | TRUE | FALSE |

| | | | | | | | | | | | | | | | | | |
|---|---|---|---|---|---|---|---|---|---|---|---|---|---|---|---|---|---|
| BIK | 0.4328802 | 0.75578973 | 1 | 0.432880199 | 0.432880199 | 0.432880199 | 0.4772225 | 0.764133952 | 0.00305 | 0.477222502 | 0.477222502 | 0.477222502 | 0.90708254 | -0.042353193 | FALSE | TRUE | FALSE |
| AP2M1 | 0.4327889 | 0.7558247 | 1 | 0.432788869 | 0.432788869 | 8.562310956 | 4.34496295 | 0.337738689 | 0.50775 | 4.344962947 | 4.344962947 | 9.171809748 | 0.099607033 | -1.001709994 | FALSE | FALSE | FALSE |
| USP22 | 0.4327048 | 0.75585967 | 1 | 0.432704802 | 0.432704802 | 8.554928619 | 0.47770502 | 0.763924828 | 1 | 0.477705017 | 0.477705017 | 9.457640464 | 0.905799156 | -0.042968088 | FALSE | TRUE | FALSE |
| PFN2 | 0.4326663 | 0.75589462 | 1 | 4.062314608 | 4.062314608 | 4.062314608 | 0.42114496 | 0.784112121 | 0.9689 | 254.0238695 | 3780.245004 | 39141.14062 | 1.027357175 | 0.011721458 | FALSE | TRUE | FALSE |
| ZBTB22 | 0.4326472 | 0.75592956 | 1 | 0.43264715 | 0.43264715 | 8.519283311 | 0.44858888 | 0.774816453 | 1 | 4.215798963 | 40.66273047 | 83.16791931 | 0.964462495 | -0.015714656 | FALSE | TRUE | FALSE |
| DUSP23 | 0.4324755 | 0.75596449 | 1 | 0.432475547 | 0.432475547 | 0.432475547 | 1.88779807 | 0.500028562 | 0.50385 | 1.887798074 | 1.887798074 | 1.887798074 | 0.22908994 | -0.639993982 | FALSE | FALSE | FALSE |
| KLHL14 | 0.4324676 | 0.75599941 | 0.5152 | 0.432467552 | 0.432467552 | 8.514283249 | 0.56255577 | 0.727894955 | 0.5015 | 0.562555766 | 0.562555766 | 4.76054323 | 0.768754989 | -0.114212053 | FALSE | TRUE | FALSE |
| NOS1AP | 0.4324438 | 0.75603432 | 1 | 0.432443805 | 0.432443805 | 0.432443805 | 0.47728937 | 0.764099125 | 1 | 0.477289366 | 0.477289366 | 0.477289366 | 0.906041147 | -0.042852079 | FALSE | TRUE | FALSE |
| POMZP3 | 0.4323176 | 0.75606922 | 1 | 0.432317644 | 0.432317644 | 0.432317644 | 38.9479217 | 0.069573989 | 0.2474 | 4.04064641 | 38.94792165 | 38.94792165 | 0.01109989 | -1.954681327 | FALSE | FALSE | FALSE |
| JKAMP | 0.4323028 | 0.75610411 | 1 | 0.432302826 | 0.432302826 | 0.432302826 | 0.47703947 | 0.76427315 | 1 | 0.477039466 | 0.477039466 | 0.477039466 | 0.906220252 | -0.042766236 | FALSE | TRUE | FALSE |
| ELMOD2 | 0.4322686 | 0.75613899 | 1 | 4.035359407 | 38.66334988 | 38.66334988 | 0.50768488 | 0.75287312 | 1 | 0.507684879 | 0.507684879 | 0.507684879 | 0.851450533 | -0.069840578 | FALSE | TRUE | FALSE |
| KBTBD2 | 0.4322458 | 0.75617386 | 1 | 0.432245791 | 0.432245791 | 8.546794191 | 0.47747645 | 0.764029439 | 1 | 0.477476453 | 0.477476453 | 9.454032019 | 0.905271429 | -0.043221186 | FALSE | TRUE | FALSE |
| SLC22A23 | 0.4322411 | 0.75620871 | 1 | 0.432241076 | 0.432241076 | 8.552095274 | 0.47743628 | 0.764064287 | 0.01345 | 0.477436284 | 0.477436284 | 9.458493079 | 0.905337719 | -0.043189385 | FALSE | TRUE | FALSE |
| PPP1R27 | 0.4322209 | 0.75624356 | 0.005 | 0.432220907 | 0.432220907 | 0.432220907 | 0.47694144 | 0.764307922 | 1 | 0.476941443 | 0.476941443 | 0.476941443 | 0.906234745 | -0.042759291 | FALSE | TRUE | FALSE |
| URM1 | 0.4321346 | 0.75627839 | 0.00275 | 0.432134646 | 0.432134646 | 0.432134646 | 1.88643863 | 0.500344892 | 0.50505 | 1.88643863 | 1.88643863 | 1.88643863 | 0.22907432 | -0.640023594 | FALSE | FALSE | FALSE |
| MED4 | 0.4321074 | 0.75631322 | 1 | 0.432107396 | 0.432107396 | 0.432107396 | 0.47689157 | 0.764342683 | 1 | 0.476891573 | 0.476891573 | 0.476891573 | 0.906091491 | -0.042827948 | FALSE | TRUE | FALSE |
| IRF2BP1 | 0.4318923 | 0.75634803 | 1 | 0.431892283 | 0.431892283 | 8.483539579 | 0.47715796 | 0.764168768 | 1 | 0.477157962 | 0.477157962 | 9.392840128 | 0.905134813 | -0.043286731 | FALSE | TRUE | FALSE |
| RPS6KB1 | 0.431874 | 0.75638284 | 1 | 0.431873974 | 0.431873974 | 8.551247421 | 0.47688871 | 0.764377432 | 1 | 0.47688871 | 0.47688871 | 9.453893645 | 0.905607461 | -0.043060008 | FALSE | TRUE | FALSE |
| C5orf58 | 0.4318359 | 0.75641763 | 1 | 0.431835924 | 0.431835924 | 0.431835924 | 0.47613104 | 0.764655051 | 1 | 0.476131037 | 0.476131037 | 0.476131037 | 0.906968651 | -0.042407724 | FALSE | TRUE | FALSE |
| PDCD1 | 0.4314968 | 0.75645242 | 1 | 0.431496814 | 0.431496814 | 0.431496814 | 4.28226562 | 0.339063928 | 0.50545 | 4.282265616 | 4.282265616 | 9.081456666 | 0.100763674 | -0.996696008 | FALSE | FALSE | FALSE |
| DIO3 | 0.4313202 | 0.75648719 | 1 | 0.431320186 | 0.431320186 | 8.538494326 | 0.4460375 | 0.775478501 | 1 | 0.446037498 | 0.446037498 | 8.840665142 | 0.967004315 | -0.014571588 | FALSE | TRUE | FALSE |
| AP4M1 | 0.4312823 | 0.75652196 | 0.25835 | 0.107853227 | 0.43128229 | 0.98346769 | 0.18185551 | 0.856559195 | 0.50545 | 0.181855513 | 0.181855513 | 0.411051826 | 2.371565657 | 0.375035153 | FALSE | TRUE | FALSE |
| SPRYD3 | 0.4312351 | 0.75655671 | 1 | 0.431235135 | 0.431235135 | 8.530164607 | 0.47627353 | 0.764551024 | 1 | 0.47627353 | 0.47627353 | 9.433482253 | 0.905435864 | -0.043142307 | FALSE | TRUE | FALSE |
| GATC | 0.4311923 | 0.75659146 | 1 | 4.019632051 | 4.019632051 | 4.019632051 | 0.47577873 | 0.764828213 | 0.50085 | 4.444887833 | 4.444887833 | 4.444887833 | 0.906287538 | -0.042733992 | FALSE | TRUE | FALSE |
| LHFPL3 | 0.4310513 | 0.75662619 | 0.50525 | 4.07163647 | 4.07163647 | 4.07163647 | 4.50135891 | 0.330249851 | 0.00675 | 4.501358908 | 4.501358908 | 4.501358908 | 0.095760255 | -1.018814706 | FALSE | FALSE | FALSE |
| TMEM230 | 0.4310012 | 0.75666092 | 1 | 0.431001248 | 0.431001248 | 0.431001248 | 0.47545207 | 0.764897404 | 1 | 0.47545207 | 0.47545207 | 0.47545207 | 0.906508302 | -0.042628214 | FALSE | TRUE | FALSE |
| LAIR2 | 0.4309414 | 0.75669563 | 0.2561 | 0.430941397 | 4.122874477 | 4.122874477 | 0.05375589 | 0.885929364 | 1 | 0.053755885 | 0.053755885 | 0.053755885 | 8.016636622 | 0.903992198 | FALSE | TRUE | FALSE |
| RGS7 | 0.4309328 | 0.75673033 | 1 | 0.43093282 | 0.43093282 | 8.533073892 | 0.47597276 | 0.764689705 | 0.0123 | 0.475972756 | 0.475972756 | 9.43618536 | 0.90537287 | -0.043172524 | FALSE | TRUE | FALSE |
| OR9I1 | 0.4309022 | 0.75676503 | 1 | 4.050405637 | 386.6426441 | 386.6426441 | 0.52312648 | 0.745429016 | 1 | 0.523126482 | 0.523126482 | 0.523126482 | 0.823705485 | -0.084228042 | FALSE | TRUE | FALSE |
| PLD6 | 0.4308913 | 0.75679971 | 1 | 0.430891324 | 0.430891324 | 0.430891324 | 38.7922137 | 0.069851792 | 0.50315 | 38.79221368 | 381.4731469 | 381.4731469 | 0.011107676 | -1.954376814 | FALSE | FALSE | FALSE |
| AVPR1A | 0.4307887 | 0.75683438 | 0.87565 | 1.723321736 | 6.895677558 | 14.59966802 | 0.56484724 | 0.726666213 | 0.25965 | 0.564847243 | 2.259656968 | 11.15975484 | 0.762664104 | -0.117666694 | FALSE | TRUE | FALSE |
| IL31 | 0.4306444 | 0.75686904 | 1 | 4.006237022 | 4.006237022 | 4.006237022 | 0.4901355 | 0.758823323 | 0.0042 | 0.490135498 | 0.490135498 | 0.490135498 | 0.878623234 | -0.056197317 | FALSE | TRUE | FALSE |
| XRCC6 | 0.4305577 | 0.75690369 | 1 | 0.430557652 | 0.430557652 | 8.511348554 | 0.47581568 | 0.764793603 | 1 | 0.475815682 | 0.475815682 | 9.419134276 | 0.904883275 | -0.043407439 | FALSE | TRUE | FALSE |
| PEX19 | 0.4305268 | 0.75693833 | 1 | 0.430526829 | 0.430526829 | 0.430526829 | 0.47520585 | 0.765035657 | 1 | 0.475205852 | 0.475205852 | 0.475205852 | 0.905979645 | -0.04288156 | FALSE | TRUE | FALSE |
| KRTAP4-2 | 0.4305169 | 0.75697296 | 1 | 0.430516919 | 0.430516919 | 0.430516919 | 0.47493772 | 0.765173737 | 0.0033 | 0.474937719 | 0.474937719 | 0.474937719 | 0.906470264 | -0.042646438 | FALSE | TRUE | FALSE |
| SPEM1 | 0.4304748 | 0.75700758 | 1 | 0.430474754 | 0.430474754 | 8.544462161 | 0.47509177 | 0.765104719 | 1 | 0.475091774 | 0.475091774 | 0.475091774 | 0.906087576 | -0.042829824 | FALSE | TRUE | FALSE |
| PERP | 0.4304218 | 0.75704219 | 1 | 1.721418824 | 6.886323 | 6.886323 | 0.48635594 | 0.760561377 | 1 | 0.486355938 | 0.486355938 | 0.486355938 | 0.884993425 | -0.053059956 | FALSE | TRUE | FALSE |
| STMN1 | 0.4303205 | 0.75707679 | 0.00495 | 0.430320543 | 0.430320543 | 0.430320543 | 0.47490726 | 0.76520823 | 1 | 0.474907256 | 0.474907256 | 0.474907256 | 0.906114904 | -0.042816726 | FALSE | TRUE | FALSE |
| MMP16 | 0.4302686 | 0.75711138 | 1 | 0.43026859 | 0.43026859 | 8.498391352 | 0.47539154 | 0.764931984 | 1 | 0.47539154 | 0.47539154 | 9.403676601 | 0.905082558 | -0.043311805 | FALSE | TRUE | FALSE |
| TEKT5 | 0.4302443 | 0.75714596 | 0.5113 | 0.430244264 | 0.430244264 | 1.657667333 | 0.61195033 | 0.706779593 | 0.5066 | 0.611950335 | 0.611950335 | 1.363892571 | 0.703070559 | -0.153001088 | FALSE | TRUE | FALSE |
| RGS18 | 0.4302162 | 0.75718052 | 1 | 4.030104598 | 4.030104598 | 4.030104598 | 0.48477588 | 0.761134852 | 1 | 1.93896948 | 1.93896948 | 1.93896948 | 0.887453766 | -0.051854263 | FALSE | TRUE | FALSE |
| FEZF1 | 0.4301377 | 0.75721508 | 1 | 0.430137717 | 0.430137717 | 8.493422988 | 0.47512645 | 0.765070194 | 1 | 0.47512645 | 0.47512645 | 9.396109461 | 0.905312085 | -0.043201682 | FALSE | TRUE | FALSE |
| E2F3 | 0.430087 | 0.75724963 | 1 | 0.430087048 | 0.430087048 | 8.515042109 | 0.47530843 | 0.764966553 | 1 | 0.475308431 | 0.475308431 | 9.421763264 | 0.904858865 | -0.043419154 | FALSE | TRUE | FALSE |
| LANCL2 | 0.430049 | 0.75728416 | 1 | 0.430049048 | 0.430049048 | 8.507850915 | 0.47524556 | 0.765001111 | 1 | 0.475245559 | 0.475245559 | 9.414240554 | 0.904898616 | -0.043400076 | FALSE | TRUE | FALSE |
| PHLDA2 | 0.4298386 | 0.75731869 | 1 | 0.429838628 | 0.429838628 | 0.429838628 | 0.47461606 | 0.765277185 | 1 | 0.474616061 | 0.474616061 | 0.474616061 | 0.905655463 | -0.043036989 | FALSE | TRUE | FALSE |
| CRY1 | 0.4297654 | 0.75735321 | 0.0175 | 0.429765442 | 0.429765442 | 1.606796907 | 0.06910012 | 0.8826254 | 1 | 0.276325664 | 0.276325664 | 1.367908452 | 6.219459955 | 0.793752676 | FALSE | TRUE | FALSE |
| HSPA8 | 0.4296751 | 0.75738772 | 0.01775 | 0.429675141 | 0.429675141 | 8.481707756 | 0.47299942 | 0.765930175 | 1 | 0.472999417 | 0.472999417 | 9.351537297 | 0.908405225 | -0.041720377 | FALSE | TRUE | FALSE |
| KRTAP4-12 | 0.4296688 | 0.75742221 | 1 | 0.429668753 | 0.429668753 | 0.429668753 | 0.47410788 | 0.765414492 | 0.00475 | 0.474107877 | 0.474107877 | 0.474107877 | 0.906267906 | -0.0427434 | FALSE | TRUE | FALSE |
| RANBP3L | 0.4295565 | 0.7574567 | 0.75045 | 0.429556504 | 3.894286824 | 3.894286824 | 0.63091952 | 0.698910286 | 0.50115 | 0.630919525 | 0.630919525 | 1.406167939 | 0.680841989 | -0.166953668 | FALSE | TRUE | FALSE |
| COX7A1 | 0.4293699 | 0.75749117 | 1 | 0.429369917 | 0.429369917 | 0.429369917 | 0.47373692 | 0.765552592 | 0.00265 | 0.473736916 | 0.473736916 | 0.473736916 | 0.906346757 | -0.042705615 | FALSE | TRUE | FALSE |
| C12orf54 | 0.4292666 | 0.75752564 | 1 | 3.988626192 | 38.03734961 | 38.03734961 | 4.556087 | 0.325894626 | 0.49525 | 0.488654529 | 4.556086995 | 4.556086995 | 0.094218264 | -1.0258649 | FALSE | FALSE | FALSE |
| MC4R | 0.429233 | 0.7575601 | 0.8763 | 1.716964236 | 6.869701977 | 6.869701977 | 0.13967884 | 0.866862239 | 0.49955 | 0.139678838 | 1.280700652 | 1.280700652 | 3.072999727 | 0.487562522 | FALSE | TRUE | FALSE |
| F2R | 0.4291872 | 0.75759454 | 0.0108 | 0.429187188 | 0.429187188 | 8.514200589 | 0.47427774 | 0.76538054 | 1 | 0.474277741 | 0.474277741 | 0.474277741 | 0.904927959 | -0.043385993 | FALSE | TRUE | FALSE |
| SS18L2 | 0.4291114 | 0.75762898 | 1 | 0.429111393 | 0.429111393 | 0.429111393 | 0.473488 | 0.765690043 | 1 | 0.473487999 | 0.473487999 | 0.473487999 | 0.906277231 | -0.042738931 | FALSE | TRUE | FALSE |
| HSPA5 | 0.4290925 | 0.7576634 | 1 | 4.030252238 | 4.030252238 | 79.42384862 | 0.47267483 | 0.76603293 | 1 | 4.445408378 | 4.445408378 | 16.01258328 | 0.907796467 | -0.042011512 | FALSE | TRUE | FALSE |
| GSC | 0.4288543 | 0.75769782 | 1 | 0.428854261 | 0.428854261 | 0.428854261 | 4.20337664 | 0.343534309 | 0.7463 | 4.203376644 | 40.74311285 | 40.74311285 | 0.102026132 | -0.991288577 | FALSE | FALSE | FALSE |
| ZNF624 | 0.4288391 | 0.75773223 | 0.7585 | 0.428839104 | 3.933743141 | 14.48551823 | 0.07236141 | 0.881936577 | 1 | 0.072361414 | 0.072361414 | 1.429023169 | 5.92635053 | 0.772787335 | FALSE | TRUE | FALSE |
| AKR1B1 | 0.4287137 | 0.75776662 | 1 | 0.428713745 | 0.428713745 | 0.428713745 | 0.46819227 | 0.767698039 | 0.50395 | 1.872731008 | 1.872731008 | 1.872731008 | 0.915678828 | -0.038256827 | FALSE | TRUE | FALSE |
| GRHL3 | 0.4285704 | 0.75780101 | 1 | 0.428570407 | 0.428570407 | 8.458607291 | 0.47363767 | 0.765586971 | 1 | 0.473637673 | 0.473637673 | 9.362894611 | 0.904848648 | -0.043424058 | FALSE | TRUE | FALSE |
| UBR2 | 0.4285261 | 0.75783538 | 0.5285 | 0.428526143 | 0.428526143 | 8.307310916 | 0.06826305 | 0.882835884 | 1 | 0.272988545 | 0.272988545 | 5.311693256 | 6.277571231 | 0.797791649 | FALSE | TRUE | FALSE |
| CDK8 | 0.4284035 | 0.75786975 | 1 | 0.428403497 | 0.428403497 | 8.492618512 | 0.47349626 | 0.765655697 | 1 | 0.473496258 | 0.473496258 | 9.39647342 | 0.904766384 | -0.043463544 | FALSE | TRUE | FALSE |
| TM4SF4 | 0.4283546 | 0.7579041 | 1 | 3.992391312 | 3.992391312 | 3.992391312 | 0.48798758 | 0.759695683 | 1 | 0.48798758 | 0.48798758 | 0.48798758 | 0.877798246 | -0.056605291 | FALSE | TRUE | FALSE |
| PRPF40B | 0.4282067 | 0.75793845 | 0.5021 | 0.428206659 | 0.428206659 | 8.394439255 | 2.06599817 | 0.484048778 | 0.89235 | 19.1763534 | 307.060859 | 1229.182621 | 0.207263814 | -0.683476515 | FALSE | FALSE | FALSE |
| LRRC58 | 0.4282015 | 0.75797279 | 1 | 0.42820151 | 0.42820151 | 0.42820151 | 0.47300131 | 0.765895903 | 1 | 0.473001313 | 0.473001313 | 0.473001313 | 0.905286092 | -0.043214152 | FALSE | TRUE | FALSE |
| MRPS25 | 0.4276523 | 0.75800712 | 1 | 0.427652274 | 0.427652274 | 0.427652274 | 0.47263254 | 0.76606716 | 0.00395 | 0.472632538 | 0.472632538 | 0.472632538 | 0.90483037 | -0.043432831 | FALSE | TRUE | FALSE |
| PSTPIP2 | 0.4275862 | 0.75804143 | 1 | 0.427586211 | 0.427586211 | 0.427586211 | 0.47243062 | 0.76613559 | 1 | 0.47243062 | 0.47243062 | 0.47243062 | 0.905077259 | -0.043314347 | FALSE | TRUE | FALSE |
| CEBPE | 0.4275166 | 0.75807574 | 0.01105 | 0.42751663 | 0.42751663 | 8.482737123 | 0.47235432 | 0.766169789 | 1 | 0.472354321 | 0.472354321 | 0.472354321 | 0.905076151 | -0.043314879 | FALSE | TRUE | FALSE |
| CENPF | 0.4273346 | 0.75811004 | 0.73165 | 3.563748494 | 265.5534246 | 8807.660215 | 0.02679887 | 0.891967139 | 0.67625 | 0.026798868 | 1.843077795 | 15.84186314 | 15.94599399 | 1.202651596 | FALSE | TRUE | FALSE |
| MEST | 0.4271001 | 0.75814433 | 1 | 0.427100139 | 0.427100139 | 0.427100139 | 0.47202319 | 0.766374759 | 1 | 0.472023187 | 0.472023187 | 0.472023187 | 0.904828725 | -0.043433621 | FALSE | TRUE | FALSE |
| B3GALT1 | 0.4270557 | 0.75817861 | 1 | 0.427055705 | 0.427055705 | 0.427055705 | 0.47218383 | 0.766272322 | 1 | 0.47218383 | 0.47218383 | 0.47218383 | 0.904426788 | -0.043626583 | FALSE | TRUE | FALSE |
| MS4A1 | 0.4270105 | 0.75821288 | 1 | 0.427010465 | 0.427010465 | 0.427010465 | 4.26283078 | 0.339857269 | 0.5026 | 4.26283078 | 4.26283078 | 4.26283078 | 0.100170635 | -0.999259574 | FALSE | FALSE | FALSE |
| PPARGC1A | 0.4269319 | 0.75824714 | 1 | 0.426931878 | 0.426931878 | 8.40199913 | 0.4722948 | 0.766203977 | 1 | 0.4722948 | 0.4722948 | 9.312643752 | 0.903952103 | -0.04385458 | FALSE | TRUE | FALSE |
| IPPK | 0.4269233 | 0.75828139 | 1 | 0.426923315 | 0.426923315 | 8.437536953 | 0.47198988 | 0.766408883 | 1 | 0.471989881 | 0.471989881 | 9.341480591 | 0.904517938 | -0.043582816 | FALSE | TRUE | FALSE |

| | | | | | | | | | | | | | | | | | |
|---|---|---|---|---|---|---|---|---|---|---|---|---|---|---|---|---|---|
| CCR6 | 0.426879 | 0.75831563 | 1 | 0.426879003 | 0.426879003 | 0.507317251 | 0.47189202 | 0.766442997 | 1 | 0.47189202 | 0.47189202 | 0.47189202 | 0.904611615 | -0.04353784 | FALSE | TRUE | FALSE |
| ZIC1 | 0.4267508 | 0.75834986 | 1 | 0.426750834 | 0.426750834 | 8.397536054 | 0.47221635 | 0.766238155 | 1 | 0.472216354 | 0.472216354 | 9.310217189 | 0.90371888 | -0.043966645 | FALSE | TRUE | FALSE |
| MAML1 | 0.4267233 | 0.75838408 | 1 | 0.426723299 | 0.426723299 | 8.37443411 | 0.47217356 | 0.766306479 | 0.02165 | 0.472173561 | 0.472173561 | 9.287317401 | 0.903742466 | -0.04395531 | FALSE | TRUE | FALSE |
| SMAP2 | 0.426625 | 0.75841829 | 1 | 4.001849995 | 4.001849995 | 8.461842512 | 0.47172593 | 0.766477101 | 1 | 4.431358822 | 4.431358822 | 9.365708985 | 0.904391732 | -0.043643417 | FALSE | TRUE | FALSE |
| GLTPD2 | 0.4265068 | 0.75845249 | 1 | 0.426506768 | 0.426506768 | 0.426506768 | 0.45674649 | 0.77216021 | 1 | 4.266128461 | 4.266128461 | 4.266128461 | 0.933793195 | -0.029749295 | FALSE | TRUE | FALSE |
| RPL26L1 | 0.4264901 | 0.75848668 | 1 | 0.426490096 | 0.426490096 | 0.426490096 | 0.47163065 | 0.766545277 | 1 | 0.471630654 | 0.471630654 | 0.471630654 | 0.904288329 | -0.043693074 | FALSE | TRUE | FALSE |
| ZNF414 | 0.4264695 | 0.75852085 | 1 | 0.426469547 | 0.426469547 | 0.426469547 | 0.47139907 | 0.766647461 | 1 | 0.471399072 | 0.471399072 | 0.471399072 | 0.904688983 | -0.043500698 | FALSE | TRUE | FALSE |
| OR5P3 | 0.4264161 | 0.75855502 | 1 | 3.979609521 | 3.979609521 | 3.979609521 | 0.48618077 | 0.760705015 | 1 | 0.486180768 | 0.486180768 | 0.486180768 | 0.877073143 | -0.056964187 | FALSE | TRUE | FALSE |
| COA3 | 0.4262016 | 0.75858918 | 1 | 0.426201598 | 1.704799091 | 1.704799091 | 0.02430468 | 0.892703866 | 1 | 0.024304677 | 0.024304677 | 0.024304677 | 17.53578513 | 1.243925215 | FALSE | TRUE | FALSE |
| XPO5 | 0.4259891 | 0.75862333 | 0.52135 | 0.42598907 | 0.42598907 | 8.325761445 | 0.14016366 | 0.866762391 | 0.0287 | 0.14016366 | 0.14016366 | 2.746950353 | 3.039226218 | 0.482763027 | FALSE | TRUE | FALSE |
| AGR2 | 0.4258283 | 0.75865747 | 1 | 0.425828284 | 0.425828284 | 0.425828284 | 0.4704235 | 0.766851566 | 0.00345 | 0.470423498 | 0.470423498 | 0.470423498 | 0.905201984 | -0.043254503 | FALSE | TRUE | FALSE |
| BAP1 | 0.4257548 | 0.7586916 | 1 | 0.425754812 | 0.425754812 | 8.371179782 | 0.47106673 | 0.766681503 | 1 | 0.471066727 | 0.471066727 | 9.280947365 | 0.903809986 | -0.043922865 | FALSE | TRUE | FALSE |
| NPAS4 | 0.4257091 | 0.75872572 | 1 | 0.425709138 | 0.425709138 | 8.373484307 | 0.46620649 | 0.768471153 | 1 | 0.466206493 | 0.466206493 | 9.188289406 | 0.913134297 | -0.039465345 | FALSE | TRUE | FALSE |
| NLRP9 | 0.4255776 | 0.75875983 | 0.63345 | 3.635871147 | 285.2378707 | 2616.343543 | 0.13690753 | 0.867392474 | 0.26515 | 0.016446153 | 0.136907528 | 0.323703185 | 3.108503887 | 0.492551415 | FALSE | TRUE | FALSE |
| PUF60 | 0.4255037 | 0.75879393 | 1 | 0.425503748 | 0.425503748 | 8.402594706 | 0.45570522 | 0.772612775 | 1 | 4.273041985 | 4.273041985 | 9.01267127 | 0.933725852 | -0.029780617 | FALSE | TRUE | FALSE |
| EDC3 | 0.4253921 | 0.75882802 | 1 | 0.425392088 | 0.425392088 | 8.418755082 | 1.86254498 | 0.502236057 | 0.5069 | 1.862544978 | 1.862544978 | 9.226890918 | 0.228392921 | -0.641317361 | FALSE | FALSE | FALSE |
| OR8D2 | 0.4253819 | 0.7588621 | 1 | 0.42538194 | 0.42538194 | 0.42538194 | 0.47020348 | 0.766953483 | 1 | 0.470203478 | 0.470203478 | 0.470203478 | 0.904676294 | -0.04350679 | FALSE | TRUE | FALSE |
| EDNRA | 0.425192 | 0.75889618 | 1 | 0.425192029 | 0.425192029 | 8.432312261 | 0.47053068 | 0.766817574 | 1 | 0.470530683 | 0.470530683 | 0.470530683 | 0.903643576 | -0.044002834 | FALSE | TRUE | FALSE |
| ANKRD13C | 0.4250964 | 0.75893024 | 1 | 0.425096409 | 0.425096409 | 8.40619066 | 0.4702948 | 0.766885549 | 1 | 0.470294802 | 0.470294802 | 9.312578289 | 0.903893489 | -0.043882742 | FALSE | TRUE | FALSE |
| OR2T11 | 0.4249849 | 0.75896429 | 0.50495 | 0.424984905 | 0.424984905 | 0.424984905 | 0.07042989 | 0.882343697 | 1 | 0.070429894 | 0.070429894 | 0.070429894 | 6.034155138 | 0.780616472 | FALSE | TRUE | FALSE |
| ANO1 | 0.4249272 | 0.75899833 | 0.2823 | 0.106274375 | 0.424927183 | 3.586589275 | 0.37451417 | 0.801004208 | 0.0265 | 0.374514169 | 0.374514169 | 7.329342493 | 1.134609097 | 0.054846261 | FALSE | TRUE | FALSE |
| MGST3 | 0.4248093 | 0.75903236 | 1 | 0.424809339 | 0.424809339 | 0.424809339 | 1.8580331 | 0.503176005 | 0.5052 | 1.858033103 | 1.858033103 | 1.858033103 | 0.228633892 | -0.640859391 | FALSE | FALSE | FALSE |
| ROPN1 | 0.4247339 | 0.75906639 | 0.74695 | 3.954808873 | 37.79409077 | 37.79409077 | 0.49963256 | 0.755266719 | 1 | 0.499632558 | 0.499632558 | 0.499632558 | 0.850092453 | -0.070533839 | FALSE | TRUE | FALSE |
| KRTAP10-2 | 0.4246605 | 0.7591004 | 1 | 3.954663936 | 37.7978445 | 37.7978445 | 0.48425173 | 0.761527449 | 1 | 4.524006425 | 4.524006425 | 4.524006425 | 0.876941624 | -0.057029316 | FALSE | TRUE | FALSE |
| KRTAP4-4 | 0.4245517 | 0.7591344 | 1 | 0.424551663 | 0.424551663 | 0.424551663 | 0.46913557 | 0.767394051 | 1 | 0.469135565 | 0.469135565 | 0.469135565 | 0.904965845 | -0.043367811 | FALSE | TRUE | FALSE |
| PPM1A | 0.4244297 | 0.75916839 | 1 | 38.04125387 | 3773.361661 | 3773.361661 | 0.52915266 | 0.743410307 | 1 | 0.52915266 | 0.52915266 | 0.52915266 | 0.802093141 | -0.095775197 | FALSE | TRUE | FALSE |
| UST | 0.4244265 | 0.75920237 | 1 | 0.424426459 | 0.424426459 | 8.417015791 | 0.46970544 | 0.767055309 | 0.01035 | 0.469705441 | 0.469705441 | 9.324404184 | 0.903601326 | -0.044023141 | FALSE | TRUE | FALSE |
| PRIM2 | 0.4243282 | 0.75923635 | 1 | 3.966950318 | 38.06271332 | 38.06271332 | 4.54073235 | 0.32671609 | 0.5069 | 4.540732347 | 4.540732347 | 9.616427334 | 0.093449289 | -1.029423996 | FALSE | FALSE | FALSE |
| IGF2BP1 | 0.4242627 | 0.75927031 | 1 | 0.42426273 | 0.42426273 | 8.36699856 | 0.46959459 | 0.767157041 | 1 | 0.469594587 | 0.469594587 | 9.276532961 | 0.903465972 | -0.0440882 | FALSE | TRUE | FALSE |
| CBY1 | 0.4241155 | 0.75930426 | 1 | 0.42411549 | 0.42411549 | 0.42411549 | 0.46892855 | 0.767461674 | 0.00365 | 0.468928551 | 0.468928551 | 0.468928551 | 0.904435203 | -0.043622542 | FALSE | TRUE | FALSE |
| ZNF331 | 0.424028 | 0.75933821 | 1 | 0.424027962 | 0.424027962 | 8.394930539 | 0.46922107 | 0.767326386 | 1 | 0.469221075 | 0.469221075 | 9.300949491 | 0.903684818 | -0.043983014 | FALSE | TRUE | FALSE |
| CENPM | 0.4238823 | 0.75937214 | 1 | 0.423882274 | 0.423882274 | 0.423882274 | 0.46853515 | 0.7675968 | 1 | 0.468535152 | 0.468535152 | 0.468535152 | 0.904696846 | -0.043496924 | FALSE | TRUE | FALSE |
| AQP12A | 0.4238517 | 0.75940606 | 1 | 0.423851675 | 0.423851675 | 8.409441271 | 0.46912257 | 0.767427867 | 1 | 0.46912257 | 0.46912257 | 9.316564841 | 0.903498792 | -0.044072424 | FALSE | TRUE | FALSE |
| GDF10 | 0.4238447 | 0.75943998 | 1 | 3.978544303 | 3.978544303 | 14.36326146 | 0.48403773 | 0.761598683 | 1 | 0.48403773 | 0.48403773 | 9.555647055 | 0.875643959 | -0.057672444 | FALSE | TRUE | FALSE |
| DUSP3 | 0.4238321 | 0.75947388 | 1 | 0.423832145 | 0.423832145 | 0.423832145 | 0.45454917 | 0.772934892 | 1 | 0.454549173 | 4.28901679 | 4.28901679 | 0.932423092 | -0.03038698 | FALSE | TRUE | FALSE |
| CADM2 | 0.4238317 | 0.75950777 | 1 | 0.423831692 | 0.423831692 | 8.411878462 | 0.46915978 | 0.767360224 | 1 | 0.469159776 | 0.469159776 | 9.320068773 | 0.903384549 | -0.044127342 | FALSE | TRUE | FALSE |
| TBC1D3B | 0.4236685 | 0.75954165 | 0.01925 | 0.423668516 | 0.423668516 | 8.350181979 | 0.44733445 | 0.775226762 | 1 | 0.447334445 | 0.447334445 | 8.832046221 | 0.94709567 | -0.023606149 | FALSE | TRUE | FALSE |
| DTX3L | 0.4235367 | 0.75957553 | 0.512 | 0.423536724 | 0.423536724 | 8.358877803 | 0.06520277 | 0.883541194 | 1 | 0.588917692 | 5.459375102 | 11.64128892 | 6.495686524 | 0.812625058 | FALSE | TRUE | FALSE |
| OMG | 0.4235316 | 0.75960939 | 1 | 0.423531648 | 0.423531648 | 8.412218425 | 0.46879802 | 0.767495471 | 1 | 0.468798024 | 0.468798024 | 0.468798024 | 0.903441623 | -0.044099905 | FALSE | TRUE | FALSE |
| MXD1 | 0.4235174 | 0.75964324 | 1 | 0.423517375 | 0.423517375 | 0.423517375 | 0.46869808 | 0.767529258 | 1 | 0.468698082 | 0.468698082 | 0.468698082 | 0.903603815 | -0.044021944 | FALSE | TRUE | FALSE |
| TMBIM4 | 0.4233927 | 0.75967709 | 0.75025 | 0.423392718 | 3.815584126 | 3.815584126 | 5.15060655 | 0.30130274 | 0.3143 | 5.150606548 | 457.5555352 | 457.5555352 | 0.082202497 | -1.085114991 | FALSE | FALSE | FALSE |
| HEY2 | 0.4233374 | 0.75971092 | 1 | 0.423337358 | 0.423337358 | 8.389832488 | 0.46856963 | 0.767563034 | 1 | 0.468569632 | 0.468569632 | 0.468569632 | 0.903467338 | -0.044087543 | FALSE | TRUE | FALSE |
| RASL10A | 0.423207 | 0.75974474 | 1 | 0.423206954 | 0.423206954 | 0.423206954 | 4.25833117 | 0.340121273 | 0.5004 | 4.258331172 | 4.258331172 | 4.258331172 | 0.099383288 | -1.002686638 | FALSE | FALSE | FALSE |
| IDNK | 0.4230385 | 0.75977856 | 1 | 0.423038497 | 0.423038497 | 0.423038497 | 0.46781955 | 0.767933894 | 1 | 0.467819551 | 0.467819551 | 0.467819551 | 0.90427708 | -0.043698477 | FALSE | TRUE | FALSE |
| ATG16L1 | 0.4227848 | 0.75981236 | 1 | 0.422784766 | 0.422784766 | 8.355299921 | 0.46810748 | 0.767765479 | 1 | 0.468107479 | 0.468107479 | 9.264207054 | 0.90317883 | -0.04422625 | FALSE | TRUE | FALSE |
| GNRHR | 0.4226211 | 0.75984616 | 1 | 0.422621059 | 0.422621059 | 0.422621059 | 4.26584386 | 0.339593061 | 0.0074 | 0.453400715 | 4.265843857 | 4.265843857 | 0.099070916 | -1.004053821 | FALSE | FALSE | FALSE |
| YIF1A | 0.4225739 | 0.75987995 | 0.503 | 1.690354952 | 6.763346996 | 6.763346996 | 0.55528306 | 0.731054367 | 0.2607 | 0.555283057 | 2.221274666 | 2.221274666 | 0.761006317 | -0.118611738 | FALSE | TRUE | FALSE |
| PSME2 | 0.4225277 | 0.75991372 | 1 | 0.422527651 | 0.422527651 | 0.422527651 | 0.4673351 | 0.76796755 | 1 | 0.467335096 | 0.467335096 | 0.467335096 | 0.904121377 | -0.043773262 | FALSE | TRUE | FALSE |
| QRICH1 | 0.4224582 | 0.75994749 | 1 | 0.422458214 | 0.422458214 | 8.3225813 | 0.43934288 | 0.777595394 | 1 | 4.133008874 | 39.90377177 | 81.56860737 | 0.961568362 | -0.017019834 | FALSE | TRUE | FALSE |
| WDR77 | 0.4223383 | 0.75998125 | 1 | 0.422338282 | 0.422338282 | 0.422338282 | 4.22898759 | 0.341963017 | 0.5017 | 4.228987588 | 4.228987588 | 4.228987588 | 0.099867468 | -1.000575962 | FALSE | FALSE | FALSE |
| CD55 | 0.4223347 | 0.76001499 | 0.87435 | 3.823225313 | 35.50000605 | 35.50000605 | 0.62116792 | 0.703154478 | 0.758 | 0.621167924 | 5.675070067 | 5.675070067 | 0.679904283 | -0.167552223 | FALSE | TRUE | FALSE |
| RPL12 | 0.4222292 | 0.76004873 | 1 | 0.422229229 | 0.422229229 | 0.422229229 | 0.46703062 | 0.768135672 | 1 | 0.467030624 | 0.467030624 | 0.467030624 | 0.904071827 | -0.043797064 | FALSE | TRUE | FALSE |
| THEM4 | 0.4221029 | 0.76008246 | 1 | 0.422102867 | 0.422102867 | 0.422102867 | 0.46672145 | 0.768269987 | 0.0055 | 0.466721448 | 0.466721448 | 0.466721448 | 0.904399978 | -0.043639457 | FALSE | TRUE | FALSE |
| SYNE3 | 0.4219336 | 0.76011618 | 1 | 9418885.431 | 36510014745 | 4.68719E+12 | 0.0079134 | 0.897325354 | 1 | 6.86551E+12 | 3.81875E+17 | 6.15516E+19 | 53.31889711 | 1.726881158 | FALSE | TRUE | FALSE |
| SAMD12 | 0.4219276 | 0.76014988 | 1 | 0.421927616 | 0.421927616 | 0.421927616 | 0.46716682 | 0.768068454 | 1 | 0.467166816 | 0.467166816 | 0.467166816 | 0.903162643 | -0.044234034 | FALSE | TRUE | FALSE |
| SP6 | 0.4217849 | 0.76018358 | 1 | 0.421784934 | 0.421784934 | 8.336915966 | 0.46698484 | 0.768169266 | 1 | 0.466984845 | 0.466984845 | 9.243332753 | 0.903209042 | -0.044211724 | FALSE | TRUE | FALSE |
| DDX47 | 0.4215269 | 0.76021727 | 0.5072 | 0.421526874 | 0.421526874 | 0.93117363 | 0.60451217 | 0.710205367 | 0.5046 | 0.604512165 | 0.604512165 | 1.328250776 | 0.697300896 | -0.156579777 | FALSE | TRUE | FALSE |
| EBF3 | 0.4214263 | 0.76025096 | 1 | 3.985728594 | 38.68803608 | 78.41911638 | 0.48202716 | 0.762379346 | 1 | 4.565410575 | 4.565410575 | 16.32791177 | 0.874279246 | -0.058349831 | FALSE | TRUE | FALSE |
| FNBP1L | 0.4210618 | 0.76028463 | 1 | 0.421061809 | 0.421061809 | 8.346279173 | 0.46623697 | 0.76843765 | 1 | 0.466236974 | 0.466236974 | 9.25163414 | 0.903106858 | -0.04426086 | FALSE | TRUE | FALSE |
| BDKRB2 | 0.4210262 | 0.76031829 | 1 | 0.421026227 | 0.421026227 | 8.336787342 | 0.46628088 | 0.768404138 | 1 | 0.466280884 | 0.466280884 | 0.466280884 | 0.902945501 | -0.044338461 | FALSE | TRUE | FALSE |
| KCNIP1 | 0.4209863 | 0.76035194 | 1 | 0.420986333 | 0.420986333 | 0.420986333 | 0.46599509 | 0.768605057 | 1 | 0.465995095 | 0.465995095 | 0.465995095 | 0.903413658 | -0.044113348 | FALSE | TRUE | FALSE |
| RNF13 | 0.4209377 | 0.76038559 | 1 | 0.420937675 | 0.420937675 | 0.420937675 | 0.46597626 | 0.768638507 | 1 | 0.465976256 | 0.465976256 | 0.465976256 | 0.90334576 | -0.04414599 | FALSE | TRUE | FALSE |
| PARP2 | 0.4208945 | 0.76041922 | 0.50525 | 0.420894506 | 3.691441812 | 3.691441812 | 0.01137611 | 0.896296015 | 0.0142 | 0.011376114 | 0.011376114 | 0.225305385 | 36.99809164 | 1.568179324 | FALSE | TRUE | FALSE |
| OR52J3 | 0.4208527 | 0.76045284 | 0.00985 | 0.420852695 | 0.420852695 | 0.420852695 | 0.46590388 | 0.768671947 | 1 | 0.465903884 | 0.465903884 | 0.465903884 | 0.903303685 | -0.044166218 | FALSE | TRUE | FALSE |
| S1PR3 | 0.4208191 | 0.76048646 | 0.0044 | 0.420819112 | 0.420819112 | 0.420819112 | 0.46572104 | 0.768705377 | 0.00365 | 0.465721039 | 0.465721039 | 0.465721039 | 0.903586218 | -0.044030402 | FALSE | TRUE | FALSE |
| TMEM64 | 0.4207552 | 0.76052006 | 1 | 0.420755249 | 0.420755249 | 0.420755249 | 4.08668744 | 0.351023102 | 0.75025 | 4.086687438 | 39.18036664 | 39.18036664 | 0.102957531 | -0.987341881 | FALSE | FALSE | FALSE |
| TAF2 | 0.4207337 | 0.76055365 | 1 | 1982603327 | 3.11124E+12 | 4.63809E+14 | 0.09348906 | 0.877426145 | 1 | 7268.716282 | 8334231.892 | 91218889.67 | 4.500352228 | 0.653246506 | FALSE | TRUE | FALSE |
| OR52A1 | 0.420491 | 0.76058724 | 1 | 37.66286601 | 370.3724116 | 370.3724116 | 0.51130378 | 0.751055423 | 1 | 0.511303777 | 0.511303777 | 0.511303777 | 0.822389817 | -0.084922276 | FALSE | TRUE | FALSE |
| ANXA2 | 0.4204602 | 0.76062081 | 1 | 0.420460226 | 0.420460226 | 0.420460226 | 0.4653984 | 0.768905746 | 1 | 0.4653984 | 0.4653984 | 0.4653984 | 0.903441495 | -0.044099966 | FALSE | TRUE | FALSE |
| BEND6 | 0.4203313 | 0.76065438 | 0.7531 | 3.912939467 | 37.38560427 | 37.38560427 | 0.49506683 | 0.756836112 | 1 | 0.495066831 | 0.495066831 | 0.495066831 | 0.849039531 | -0.071072089 | FALSE | TRUE | FALSE |
| IL1R1 | 0.4203086 | 0.76068793 | 1 | 0.420308636 | 0.420308636 | 8.321675733 | 0.46553832 | 0.768805608 | 0.01205 | 0.46553832 | 0.46553832 | 9.228321372 | 0.902844337 | -0.044387122 | FALSE | TRUE | FALSE |
| CNPY2 | 0.4202713 | 0.76072148 | 1 | 0.420271332 | 0.420271332 | 0.420271332 | 0.45082209 | 0.774182233 | 1 | 4.202959395 | 4.202959395 | 4.202959395 | 0.932233218 | -0.030475426 | FALSE | TRUE | FALSE |
| PPP6C | 0.4199268 | 0.76075501 | 1 | 3.924375469 | 37.64052707 | 37.64052707 | 0.46498714 | 0.768939107 | 1 | 4.353667454 | 41.83658679 | 41.83658679 | 0.903093461 | -0.044267302 | FALSE | TRUE | FALSE |

| | | | | | | | | | | | | | | | | | |
|---|---|---|---|---|---|---|---|---|---|---|---|---|---|---|---|---|---|
| ZNF32 | 0.4198622 | 0.76078854 | 0.0065 | 0.419862172 | 0.419862172 | 0.419862172 | 0.46481597 | 0.769039128 | 1 | 0.464815971 | 0.464815971 | 0.464815971 | 0.903286889 | -0.044174293 | FALSE | TRUE | FALSE |
| ZNF607 | 0.4195747 | 0.76082206 | 0.1413 | 0.419574699 | 3.725184579 | 8.286747288 | 59.954713 | 0.046835668 | 0.1945 | 6.512612109 | 59.95471296 | 565.4490967 | 0.006998194 | -2.155014037 | FALSE | FALSE | FALSE |
| CD81 | 0.4195646 | 0.76085557 | 1 | 0.419564568 | 0.419564568 | 0.419564568 | 4.12956976 | 0.34768734 | 0.2573 | 4.12956976 | 40.0937843 | 40.0937843 | 0.101600068 | -0.993106002 | FALSE | FALSE | FALSE |
| TSPAN3 | 0.4195598 | 0.76088907 | 1 | 0.419559816 | 0.419559816 | 0.419559816 | 0.45975931 | 0.771053343 | 1 | 1.838782816 | 1.838782816 | 1.838782816 | 0.91256405 | -0.039736644 | FALSE | TRUE | FALSE |
| TMED1 | 0.4194695 | 0.76092255 | 0.00635 | 0.419469454 | 0.419469454 | 0.419469454 | 0.4644732 | 0.769238896 | 1 | 0.464473199 | 0.464473199 | 0.464473199 | 0.903107983 | -0.044260319 | FALSE | TRUE | FALSE |
| HNF4A | 0.4193343 | 0.76095603 | 0.5107 | 0.419334339 | 1.676850294 | 6.707128622 | 0.0122579 | 0.896021592 | 0.75275 | 0.048987157 | 0.195820038 | 0.415396045 | 34.20932616 | 1.53414452 | FALSE | TRUE | FALSE |
| DCAF4L1 | 0.4192818 | 0.7609895 | 0.0142 | 0.419281813 | 0.419281813 | 8.299850612 | 0.46444752 | 0.769272155 | 1 | 0.464447515 | 0.464447515 | 9.205237721 | 0.902753917 | -0.044430619 | FALSE | TRUE | FALSE |
| LGALS13 | 0.4192521 | 0.76102296 | 1 | 0.419252149 | 0.419252149 | 0.419252149 | 0.46395334 | 0.769504677 | 1 | 0.463953341 | 0.463953341 | 0.463953341 | 0.903651536 | -0.043999009 | FALSE | TRUE | FALSE |
| DCUN1D4 | 0.4192192 | 0.76105641 | 1 | 0.419219177 | 0.419219177 | 0.419219177 | 0.4644081 | 0.769305403 | 1 | 0.464408099 | 0.464408099 | 0.464408099 | 0.902695664 | -0.044458644 | FALSE | TRUE | FALSE |
| COQ7 | 0.4191571 | 0.76108985 | 0.2536 | 0.419157147 | 1.676471139 | 1.676471139 | 0.13811186 | 0.867138898 | 1 | 0.138111859 | 0.552281929 | 0.552281929 | 3.034910627 | 0.482145906 | FALSE | TRUE | FALSE |
| ASPDH | 0.419068 | 0.76112328 | 1 | 0.419067955 | 0.419067955 | 0.419067955 | 0.435838 | 0.778882472 | 1 | 4.058546362 | 38.78865216 | 38.78865216 | 0.961522291 | -0.017040643 | FALSE | TRUE | FALSE |
| SPACA3 | 0.4190606 | 0.7611567 | 1 | 0.419060647 | 0.419060647 | 0.419060647 | 0.46430976 | 0.769405084 | 1 | 0.464309758 | 0.464309758 | 0.464309758 | 0.902545424 | -0.044530931 | FALSE | TRUE | FALSE |
| HNRNPM | 0.4190239 | 0.76119011 | 1 | 0.419023906 | 0.419023906 | 8.222478269 | 0.46455245 | 0.769172349 | 1 | 0.464552448 | 0.464552448 | 9.136577301 | 0.901994829 | -0.044795952 | FALSE | TRUE | FALSE |
| EBF2 | 0.4189832 | 0.76122351 | 1 | 0.418983154 | 0.418983154 | 8.275000805 | 0.46454934 | 0.769205628 | 1 | 0.464549335 | 0.464549335 | 9.188730668 | 0.901913149 | -0.044835282 | FALSE | TRUE | FALSE |
| SSBP1 | 0.418967 | 0.7612569 | 1 | 0.418966975 | 0.418966975 | 0.418966975 | 0.44966793 | 0.774594894 | 1 | 0.449667934 | 4.199097893 | 4.199097893 | 0.931725264 | -0.030712128 | FALSE | TRUE | FALSE |
| RXFP4 | 0.418895 | 0.76129028 | 0.50575 | 0.418894973 | 0.418894973 | 0.929622944 | 0.06881364 | 0.882678086 | 1 | 0.068813636 | 0.068813636 | 1.365012353 | 6.087383192 | 0.784430641 | FALSE | TRUE | FALSE |
| TCP11 | 0.4186706 | 0.76132366 | 0.0107 | 0.418670638 | 0.418670638 | 0.918182867 | 4.75672905 | 0.317025678 | 0.2547 | 0.509829943 | 4.756729046 | 4.756729046 | 0.088016499 | -1.055435909 | FALSE | FALSE | FALSE |
| TH | 0.4186162 | 0.76135702 | 0.03855 | 0.026224677 | 0.104763225 | 0.418616219 | 0.00109319 | 0.899355813 | 1 | 0.00109319 | 0.00109319 | 0.021603319 | 382.930998 | 2.583120524 | FALSE | TRUE | FALSE |
| CCM2 | 0.4185218 | 0.76139037 | 1 | 0.418521773 | 0.418521773 | 8.282057016 | 0.46369163 | 0.769537856 | 1 | 0.46369163 | 0.46369163 | 9.187570467 | 0.902586431 | -0.0445112 | FALSE | TRUE | FALSE |
| TLE3 | 0.4185032 | 0.76142371 | 1 | 0.418503159 | 0.418503159 | 8.219133867 | 0.46409663 | 0.769438291 | 1 | 0.464096634 | 0.464096634 | 9.134349624 | 0.90175866 | -0.044909678 | FALSE | TRUE | FALSE |
| WFDC12 | 0.4179566 | 0.76145705 | 0.0029 | 0.417956639 | 0.417956639 | 0.417956639 | 0.46296037 | 0.769836021 | 1 | 0.462960372 | 0.462960372 | 0.462960372 | 0.902791391 | -0.044412591 | FALSE | TRUE | FALSE |
| CADM1 | 0.417769 | 0.76149038 | 1 | 0.417768986 | 0.417768986 | 8.280885276 | 0.46327488 | 0.769670473 | 1 | 0.463274882 | 0.463274882 | 0.463274882 | 0.901773444 | -0.044902558 | FALSE | TRUE | FALSE |
| ITPK1 | 0.4177491 | 0.76152369 | 1 | 3.912052031 | 3.912052031 | 8.270360724 | 0.47770819 | 0.763889936 | 1 | 0.477708188 | 0.477708188 | 9.468918268 | 0.874486052 | -0.058247113 | FALSE | TRUE | FALSE |
| NIPSNAP1 | 0.4176596 | 0.761557 | 1 | 0.417659579 | 0.417659579 | 0.417659579 | 0.46283993 | 0.769869101 | 1 | 0.462839934 | 0.462839934 | 0.462839934 | 0.902384491 | -0.044608377 | FALSE | TRUE | FALSE |
| HAT1 | 0.4175729 | 0.7615903 | 1 | 0.417572897 | 0.417572897 | 8.284902834 | 0.46297477 | 0.769802932 | 1 | 0.462974765 | 0.462974765 | 9.194507215 | 0.901934465 | -0.044825017 | FALSE | TRUE | FALSE |
| C4orf33 | 0.4175388 | 0.76162359 | 1 | 0.417538814 | 0.417538814 | 0.417538814 | 0.45725366 | 0.772062993 | 1 | 1.828979275 | 1.828979275 | 1.828979275 | 0.913144829 | -0.039460336 | FALSE | TRUE | FALSE |
| KRTAP19-4 | 0.4175219 | 0.76165687 | 1 | 0.417521873 | 0.417521873 | 0.417521873 | 38.4365985 | 0.070686697 | 0.00145 | 4.033295242 | 38.43659854 | 38.43659854 | 0.010862612 | -1.964065716 | FALSE | FALSE | FALSE |
| IFNA2 | 0.4175032 | 0.76169013 | 1 | 0.417503159 | 0.417503159 | 0.417503159 | 0.46226255 | 0.770166361 | 1 | 0.462262548 | 0.462262548 | 0.462262548 | 0.903173231 | -0.044228943 | FALSE | TRUE | FALSE |
| RASSF5 | 0.4173489 | 0.76172339 | 1 | 0.417348888 | 0.417348888 | 8.284666428 | 0.46274414 | 0.769935229 | 1 | 0.462744144 | 0.462744144 | 9.194040005 | 0.90189988 | -0.044841671 | FALSE | TRUE | FALSE |
| UVRAG | 0.417231 | 0.76175664 | 1 | 0.417230986 | 0.417230986 | 8.238247851 | 0.46267367 | 0.769968278 | 0.01545 | 0.462673674 | 0.462673674 | 9.149528852 | 0.901782422 | -0.044898234 | FALSE | TRUE | FALSE |
| CNR1 | 0.4171377 | 0.76178988 | 0.0137 | 0.417137733 | 0.417137733 | 8.249988608 | 0.46265328 | 0.770034345 | 1 | 0.462653275 | 0.462653275 | 9.162417737 | 0.90162062 | -0.044976165 | FALSE | TRUE | FALSE |
| POLG2 | 0.4170015 | 0.76182311 | 0.13695 | 0.417001526 | 1.667893857 | 2.064603386 | 0.16341343 | 0.861249037 | 0.01285 | 0.163413428 | 0.163413428 | 0.810196487 | 2.55181922 | 0.406849904 | FALSE | TRUE | FALSE |
| CLRN1 | 0.4168673 | 0.76185634 | 0.2531 | 0.416867326 | 3.884469889 | 3.884469889 | 0.06990687 | 0.882475897 | 1 | 0.069906871 | 0.069906871 | 0.069906871 | 5.96318103 | 0.775477994 | FALSE | TRUE | FALSE |
| CSTF1 | 0.4167878 | 0.76188955 | 1 | 0.416787771 | 0.416787771 | 8.251073711 | 0.46245796 | 0.770067364 | 1 | 0.462457963 | 0.462457963 | 9.166386262 | 0.901244664 | -0.045157294 | FALSE | TRUE | FALSE |
| SLFN12 | 0.4167786 | 0.76192275 | 0.93785 | 3.758324915 | 329.5953211 | 329.5953211 | 0.65150413 | 0.689030343 | 0.5059 | 0.651504126 | 0.651504126 | 1.453956617 | 0.639717554 | -0.194011732 | FALSE | TRUE | FALSE |
| PLA2G2C | 0.4165675 | 0.76195594 | 0.50805 | 3.903063906 | 3.903063906 | 3.903063906 | 0.47644948 | 0.764516327 | 1 | 0.476449482 | 0.476449482 | 0.476449482 | 0.874316227 | -0.058331461 | FALSE | TRUE | FALSE |
| PHLDA1 | 0.4164527 | 0.76198913 | 1 | 0.416452666 | 0.416452666 | 8.255399288 | 0.46192287 | 0.770298217 | 1 | 0.461922874 | 0.461922874 | 0.461922874 | 0.901563204 | -0.045003822 | FALSE | TRUE | FALSE |
| RIOK1 | 0.4164069 | 0.7620223 | 0.50545 | 0.416406928 | 0.416406928 | 0.938527016 | 0.06907895 | 0.882634184 | 0.01275 | 0.069078952 | 0.069078952 | 1.369167207 | 6.02798557 | 0.780172204 | FALSE | TRUE | FALSE |
| TMEM62 | 0.4162612 | 0.76205547 | 0.7538 | 0.416261207 | 3.830385505 | 8.237437489 | 0.59788211 | 0.713074976 | 0.2659 | 0.597882114 | 5.537668875 | 11.84656124 | 0.696226225 | -0.157249622 | FALSE | TRUE | FALSE |
| IL3 | 0.4162516 | 0.76208863 | 1 | 0.416251578 | 0.416251578 | 0.416251578 | 0.46103828 | 0.770660004 | 1 | 0.461038276 | 0.461038276 | 0.461038276 | 0.902856876 | -0.04438109 | FALSE | TRUE | FALSE |
| CHMP7 | 0.4161303 | 0.76212177 | 1 | 0.416130326 | 0.416130326 | 8.247227861 | 0.46157041 | 0.770495703 | 1 | 0.461570414 | 0.461570414 | 0.461570414 | 0.901553291 | -0.045008597 | FALSE | TRUE | FALSE |
| TRAM1 | 0.4161112 | 0.76215491 | 1 | 0.41611119 | 0.41611119 | 0.41611119 | 0.46167067 | 0.770429914 | 1 | 0.461670675 | 0.461670675 | 0.461670675 | 0.901316052 | -0.045122894 | FALSE | TRUE | FALSE |
| B3GAT1 | 0.416077 | 0.76218803 | 1 | 0.416077039 | 0.416077039 | 8.217863477 | 0.4614475 | 0.770528582 | 1 | 0.461447501 | 0.461447501 | 9.127630261 | 0.901677955 | -0.044948548 | FALSE | TRUE | FALSE |
| TCEA2 | 0.4160117 | 0.76222115 | 0.01065 | 0.416011727 | 0.416011727 | 8.249546821 | 0.45625672 | 0.772322056 | 1 | 1.824888671 | 1.824888671 | 1.824888671 | 0.911793103 | -0.040103697 | FALSE | TRUE | FALSE |
| CREBZF | 0.4159034 | 0.76225426 | 1 | 3.892396313 | 3.892396313 | 8.21632603 | 0.47583492 | 0.764758982 | 1 | 0.475834915 | 0.475834915 | 9.414130049 | 0.874049748 | -0.058463848 | FALSE | TRUE | FALSE |
| FGF14 | 0.4157618 | 0.76228736 | 1 | 3.92695866 | 3.92695866 | 3.92695866 | 0.47613923 | 0.764620386 | 1 | 0.476139232 | 0.476139232 | 0.476139232 | 0.873193685 | -0.058889414 | FALSE | TRUE | FALSE |
| PCBD2 | 0.4157612 | 0.76232045 | 1 | 1.6626646 | 1.6626646 | 1.6626646 | 0.4665213 | 0.768370615 | 1 | 0.466521301 | 0.466521301 | 0.466521301 | 0.89119447 | -0.050027517 | FALSE | TRUE | FALSE |
| MXI1 | 0.4157223 | 0.76235352 | 1 | 0.415722258 | 0.415722258 | 0.415722258 | 0.46110374 | 0.770627163 | 0.0105 | 0.461103741 | 0.461103741 | 9.172282013 | 0.901580752 | -0.044995369 | FALSE | TRUE | FALSE |
| CTF1 | 0.4157214 | 0.76238659 | 1 | 0.415721375 | 0.415721375 | 0.415721375 | 0.43284367 | 0.780154099 | 1 | 4.051281133 | 38.91710483 | 38.91710483 | 0.960442317 | -0.017528713 | FALSE | TRUE | FALSE |
| ST13 | 0.415683 | 0.76241965 | 1 | 0.415683036 | 0.415683036 | 8.252653008 | 0.46129349 | 0.770594312 | 1 | 0.461293493 | 0.461293493 | 0.461293493 | 0.901124863 | -0.045215027 | FALSE | TRUE | FALSE |
| C1D | 0.4154126 | 0.7624527 | 0.00345 | 0.415412564 | 0.415412564 | 0.415412564 | 0.46031421 | 0.770922384 | 1 | 0.460314208 | 0.460314208 | 0.460314208 | 0.902454359 | -0.044574753 | FALSE | TRUE | FALSE |
| PIGS | 0.4154082 | 0.76248574 | 0.50695 | 0.415408199 | 0.415408199 | 3.518370195 | 0.54010714 | 0.738017273 | 0.50645 | 0.540107145 | 0.540107145 | 2.669501302 | 0.769121837 | -0.114004858 | FALSE | TRUE | FALSE |
| CALCR | 0.4154028 | 0.76251877 | 0.87385 | 0.415402794 | 35.4998761 | 35.4998761 | 0.0664577 | 0.883228687 | 1 | 5.475450707 | 51.64463665 | 51.64463665 | 6.250634503 | 0.795924105 | FALSE | TRUE | FALSE |
| MSI1 | 0.4153494 | 0.76255179 | 1 | 15.66389174 | 1504.619872 | 1504.619872 | 0.51260063 | 0.750625928 | 1 | 0.512600634 | 0.512600634 | 10.1788689 | 0.810278798 | -0.091365525 | FALSE | TRUE | FALSE |
| EHF | 0.4152535 | 0.76258481 | 1 | 0.415253529 | 0.415253529 | 0.415253529 | 0.44491128 | 0.77576105 | 1 | 0.444911282 | 0.444911282 | 0.444911282 | 0.933340075 | -0.029960086 | FALSE | TRUE | FALSE |
| HIF1AN | 0.4152027 | 0.76261781 | 1 | 0.415202675 | 0.415202675 | 8.250634084 | 0.46051889 | 0.770791271 | 1 | 0.460518892 | 0.460518892 | 0.460518892 | 0.901597486 | -0.044987308 | FALSE | TRUE | FALSE |
| NDUFA4 | 0.4151683 | 0.7626508 | 1 | 0.415168255 | 1.660479735 | 1.660479735 | 0.46556515 | 0.768738798 | 1 | 0.465565152 | 0.465565152 | 0.465565152 | 0.891751139 | -0.049756327 | FALSE | TRUE | FALSE |
| CSK | 0.4149825 | 0.76268378 | 1 | 0.414982498 | 0.414982498 | 8.197758544 | 0.46071483 | 0.770758468 | 1 | 0.460714832 | 0.460714832 | 9.114620224 | 0.900736137 | -0.045402413 | FALSE | TRUE | FALSE |
| KCNK2 | 0.4149478 | 0.76271676 | 1 | 0.414947841 | 0.414947841 | 8.225605731 | 0.46041563 | 0.770856848 | 1 | 0.460415625 | 0.460415625 | 9.136626249 | 0.901246219 | -0.045156544 | FALSE | TRUE | FALSE |
| TRAF6 | 0.4148709 | 0.76274972 | 1 | 0.414870872 | 0.414870872 | 8.210135207 | 0.46043343 | 0.770824065 | 1 | 0.460433427 | 0.460433427 | 9.123330959 | 0.901044207 | -0.045253901 | FALSE | TRUE | FALSE |
| ZNF777 | 0.4148394 | 0.76278268 | 1 | 38.00190707 | 377.9265661 | 741.5419875 | 0.50668608 | 0.753141429 | 1 | 0.506686084 | 0.506686084 | 9.916102086 | 0.818730545 | -0.086859007 | FALSE | TRUE | FALSE |
| N4BP1 | 0.4148283 | 0.76281562 | 0.02 | 0.414828337 | 0.414828337 | 8.183763732 | 0.46034199 | 0.770889621 | 1 | 0.460341995 | 0.460341995 | 9.096519902 | 0.901130772 | -0.04521218 | FALSE | TRUE | FALSE |
| SYT7 | 0.4146953 | 0.76284855 | 1 | 3.919996197 | 38.02992274 | 77.58407119 | 1.82068932 | 0.505511943 | 0.87795 | 17.22034754 | 167.1593809 | 167.1593809 | 0.227768292 | -0.642506735 | FALSE | FALSE | FALSE |
| OVOL2 | 0.4145407 | 0.76288148 | 1 | 0.41454073 | 0.41454073 | 0.41454073 | 0.4598996 | 0.771020618 | 1 | 0.459899598 | 0.459899598 | 0.459899598 | 0.901372237 | -0.045095823 | FALSE | TRUE | FALSE |
| SFT2D1 | 0.4143804 | 0.7629144 | 1 | 0.414380376 | 0.414380376 | 0.414380376 | 0.45924676 | 0.77128215 | 1 | 0.459246758 | 0.459246758 | 0.459246758 | 0.902304412 | -0.044646919 | FALSE | TRUE | FALSE |
| LPCAT3 | 0.4139878 | 0.76294731 | 1 | 0.413987791 | 0.413987791 | 8.201101222 | 0.45947808 | 0.771151463 | 1 | 0.459478077 | 0.459478077 | 9.112664659 | 0.900995743 | -0.045277261 | FALSE | TRUE | FALSE |
| MLEC | 0.4139285 | 0.76298021 | 0.00885 | 0.41392853 | 0.41392853 | 0.41392853 | 0.45912831 | 0.771314797 | 1 | 0.459128308 | 0.459128308 | 0.459128308 | 0.901553059 | -0.045008709 | FALSE | TRUE | FALSE |
| EFNA2 | 0.4139272 | 0.7630131 | 1 | 0.413927247 | 0.413927247 | 0.413927247 | 0.45951584 | 0.771118767 | 1 | 0.459515835 | 0.459515835 | 0.459515835 | 0.900789951 | -0.045376467 | FALSE | TRUE | FALSE |
| FIS1 | 0.4138912 | 0.76304598 | 0.00475 | 0.413891229 | 0.413891229 | 0.413891229 | 0.45910894 | 0.771347434 | 1 | 0.459108938 | 0.459108938 | 0.459108938 | 0.901509849 | -0.045029524 | FALSE | TRUE | FALSE |
| CRTC2 | 0.4138452 | 0.76307885 | 1 | 0.413845227 | 0.413845227 | 8.154570401 | 0.45943697 | 0.77118415 | 1 | 0.459436965 | 0.459436965 | 9.069032486 | 0.900766065 | -0.045387984 | FALSE | TRUE | FALSE |
| SP1 | 0.4137973 | 0.76311172 | 1 | 0.413797349 | 0.413797349 | 8.158893282 | 0.42775343 | 0.781855864 | 1 | 4.051588217 | 4.051588217 | 14.47095112 | 0.967373527 | -0.014405802 | FALSE | TRUE | FALSE |
| SLC7A8 | 0.4137526 | 0.76314457 | 1 | 9.22368E+27 | 6.63086E+32 | 1.78843E+35 | 0.41577068 | 0.786207057 | 1 | 2.01918E+14 | 8.59235E+17 | 5.62385E+19 | 0.995146225 | -0.0021131 | FALSE | TRUE | FALSE |
| DYDC2 | 0.4137053 | 0.76317741 | 1 | 0.413705336 | 0.413705336 | 0.413705336 | 0.4309511 | 0.780724422 | 1 | 4.007942392 | 38.25640575 | 38.25640575 | 0.959982079 | -0.017736874 | FALSE | TRUE | FALSE |

| | | | | | | | | | | | | | | | | | |
|---|---|---|---|---|---|---|---|---|---|---|---|---|---|---|---|---|---|
| FAM24A | 0.4136579 | 0.76321024 | 1 | 0.413657926 | 0.413657926 | 0.413657926 | 0.45836492 | 0.771738325 | 0.0021 | 0.458364917 | 0.458364917 | 0.458364917 | 0.902464195 | -0.04457002 | FALSE | TRUE | FALSE |
| DNAJC11 | 0.4135498 | 0.76324307 | 1 | 0.413549833 | 0.413549833 | 8.164481945 | 1.81568402 | 0.506131959 | 0.5058 | 1.815684022 | 1.815684022 | 15.36177997 | 0.227765309 | -0.642512422 | FALSE | FALSE | FALSE |
| COX20 | 0.4134112 | 0.76327588 | 1 | 0.413411207 | 0.413411207 | 0.413411207 | 0.45841171 | 0.771673277 | 1 | 0.45841171 | 0.45841171 | 0.45841171 | 0.901833873 | -0.044873457 | FALSE | TRUE | FALSE |
| TOX4 | 0.4133945 | 0.76330869 | 0.015 | 0.413394547 | 0.413394547 | 8.179502217 | 0.45883226 | 0.771380063 | 1 | 0.458832265 | 0.458832265 | 9.090217076 | 0.900970962 | -0.045289206 | FALSE | TRUE | FALSE |
| PDAP1 | 0.4132304 | 0.76334148 | 1 | 0.413230396 | 0.413230396 | 0.413230396 | 0.4584191 | 0.771640738 | 1 | 0.458419105 | 0.458419105 | 0.458419105 | 0.901424901 | -0.045070449 | FALSE | TRUE | FALSE |
| C17orf78 | 0.4131727 | 0.76337427 | 1 | 3.842671056 | 3.842671056 | 3.842671056 | 0.47259129 | 0.76610138 | 1 | 0.472591293 | 0.472591293 | 0.472591293 | 0.874270635 | -0.058354108 | FALSE | TRUE | FALSE |
| AHSP | 0.4129902 | 0.76340705 | 0.00265 | 0.412990151 | 0.412990151 | 0.412990151 | 0.45800386 | 0.771835824 | 0.00275 | 0.458003859 | 0.458003859 | 0.458003859 | 0.901717623 | -0.044929443 | FALSE | TRUE | FALSE |
| AGO1 | 0.4129561 | 0.76343982 | 0.02945 | 0.412956129 | 0.412956129 | 8.085969559 | 0.4587073 | 0.771412683 | 1 | 0.458707303 | 0.458707303 | 9.00449037 | 0.900260637 | -0.045631739 | FALSE | TRUE | FALSE |
| MAD2L1 | 0.412808 | 0.76347258 | 1 | 432.3287379 | 39102.1508 | 156586.3643 | 0.47706031 | 0.764238367 | 1 | 122.2592739 | 8371.046151 | 33539.22018 | 0.865316231 | -0.06282515 | FALSE | TRUE | FALSE |
| HNRNPLL | 0.4126109 | 0.76350533 | 1 | 3.865300406 | 37.16319132 | 76.58047074 | 0.48766449 | 0.759876583 | 1 | 0.487664494 | 0.487664494 | 9.672659438 | 0.846095849 | -0.072580436 | FALSE | TRUE | FALSE |
| RBM46 | 0.4125872 | 0.76353807 | 0.0126 | 0.41258716 | 0.41258716 | 8.176064767 | 0.45795008 | 0.771900775 | 1 | 0.457950081 | 0.457950081 | 9.085013028 | 0.900943526 | -0.045302431 | FALSE | TRUE | FALSE |
| LYPLA1 | 0.4124385 | 0.7635708 | 1 | 0.412438478 | 0.412438478 | 0.412438478 | 0.45735439 | 0.771998134 | 1 | 0.45735439 | 0.45735439 | 0.45735439 | 0.901791885 | -0.044893677 | FALSE | TRUE | FALSE |
| KRTAP12-2 | 0.4123585 | 0.76360352 | 1 | 0.412358523 | 3.828968679 | 3.828968679 | 0.45718068 | 0.772127812 | 0.5082 | 4.256271697 | 4.256271697 | 4.256271697 | 0.901959636 | -0.044812897 | FALSE | TRUE | FALSE |
| PHF21B | 0.4118363 | 0.76363624 | 0.50615 | 0.411836254 | 0.411836254 | 2.034598209 | 11.4692098 | 0.183007921 | 0.2567 | 1.19595754 | 11.46920983 | 23.65829339 | 0.035907988 | -1.444808923 | FALSE | FALSE | FALSE |
| TMEM156 | 0.4117387 | 0.76366895 | 1 | 0.411738711 | 0.411738711 | 0.411738711 | 0.45670866 | 0.772192598 | 1 | 0.456708664 | 0.456708664 | 0.456708664 | 0.901534705 | -0.04501755 | FALSE | TRUE | FALSE |
| SUMO4 | 0.4114357 | 0.76370165 | 1 | 0.411435673 | 0.411435673 | 0.411435673 | 3.98045063 | 0.357342312 | 0.7524 | 3.980450627 | 37.92925173 | 37.92925173 | 0.103364094 | -0.985630297 | FALSE | FALSE | FALSE |
| AASDH | 0.4111105 | 0.76373434 | 0.5047 | 0.411110538 | 7.997156765 | 31.97598085 | 0.02819704 | 0.891701808 | 0.02115 | 0.028197038 | 0.028197038 | 0.555137262 | 14.5799191 | 1.163755114 | FALSE | TRUE | FALSE |
| TDP2 | 0.4110299 | 0.76376702 | 0.80635 | 3.729497672 | 138.9420436 | 138.9420436 | 0.55805312 | 0.729623655 | 0.75035 | 1.273775548 | 5.094474358 | 5.094474358 | 0.736542634 | -0.132802109 | FALSE | TRUE | FALSE |
| SUOX | 0.4109806 | 0.7637997 | 0.6599 | 0.410980582 | 6.574507249 | 26.30551781 | 1.65686059 | 0.521314318 | 0.315 | 0.414457929 | 15.40537366 | 15.40537366 | 0.248047774 | -0.605464665 | FALSE | FALSE | FALSE |
| PNO1 | 0.4108644 | 0.76383236 | 1 | 0.410864365 | 0.410864365 | 0.410864365 | 0.45616715 | 0.772354396 | 1 | 0.456167147 | 0.456167147 | 0.456167147 | 0.900688196 | -0.045425529 | FALSE | TRUE | FALSE |
| IMPA1 | 0.4105316 | 0.76386502 | 1 | 0.410531592 | 0.410531592 | 0.410531592 | 0.45578898 | 0.772483665 | 1 | 0.45578898 | 0.45578898 | 0.45578898 | 0.900705392 | -0.045417237 | FALSE | TRUE | FALSE |
| DAOA | 0.4105013 | 0.76389767 | 1 | 3.806709359 | 3.806709359 | 3.806709359 | 0.46962883 | 0.767123141 | 1 | 0.469628835 | 0.469628835 | 0.469628835 | 0.874097339 | -0.058440202 | FALSE | TRUE | FALSE |
| C2orf76 | 0.4104829 | 0.76393031 | 1 | 0.410482877 | 0.410482877 | 0.410482877 | 0.45549704 | 0.772645029 | 1 | 0.455497044 | 0.455497044 | 0.455497044 | 0.901175721 | -0.045190517 | FALSE | TRUE | FALSE |
| DCAF8 | 0.4103031 | 0.76396294 | 0.0171 | 0.410303139 | 0.410303139 | 8.096946818 | 0.45604713 | 0.772386728 | 1 | 0.456047128 | 0.456047128 | 9.014061947 | 0.899694603 | -0.045904885 | FALSE | TRUE | FALSE |
| TMEM170A | 0.4101445 | 0.76399556 | 1 | 36.28544142 | 354.6345595 | 354.6345595 | 40.4665999 | 0.067650102 | 0.50355 | 40.46659989 | 396.5353864 | 396.5353864 | 0.010135383 | -1.994159836 | FALSE | FALSE | FALSE |
| KIF2A | 0.4100248 | 0.76402817 | 0.0176 | 0.410024837 | 0.410024837 | 8.104553086 | 0.45574876 | 0.772580512 | 1 | 0.455748763 | 0.455748763 | 9.021012914 | 0.899672957 | -0.045915334 | FALSE | TRUE | FALSE |
| SCAI | 0.4095132 | 0.76406078 | 1 | 0.409513206 | 0.409513206 | 8.094138313 | 0.4551033 | 0.772806156 | 1 | 0.455103296 | 0.455103296 | 9.007940658 | 0.899824741 | -0.04584207 | FALSE | TRUE | FALSE |
| CCDC167 | 0.4094185 | 0.76409337 | 0.00295 | 0.40941847 | 0.40941847 | 0.40941847 | 0.45422663 | 0.773063478 | 1 | 0.454226626 | 0.454226626 | 0.454226626 | 0.901352864 | -0.045105157 | FALSE | TRUE | FALSE |
| INO80C | 0.4093489 | 0.76412596 | 1 | 0.409348949 | 0.409348949 | 0.409348949 | 0.45436501 | 0.772999204 | 0.00525 | 0.454365008 | 0.454365008 | 0.454365008 | 0.900925338 | -0.045311198 | FALSE | TRUE | FALSE |
| HES5 | 0.4093158 | 0.76415854 | 1 | 0.40931578 | 0.40931578 | 0.40931578 | 1.7962857 | 0.507983564 | 0.50025 | 1.7962857 | 1.7962857 | 1.7962857 | 0.227867861 | -0.642316925 | FALSE | FALSE | FALSE |
| SULT2B1 | 0.4091678 | 0.76419111 | 1 | 0.409167759 | 0.409167759 | 8.106507873 | 1.7985639 | 0.507829795 | 0.5017 | 1.798563904 | 1.798563904 | 1.869771452 | 0.227496926 | -0.643024467 | FALSE | FALSE | FALSE |
| GSTP1 | 0.4089942 | 0.76422367 | 1 | 3566.60065 | 385878.4392 | 385878.4392 | 0.46828905 | 0.767664303 | 1 | 411.0444445 | 42684.26331 | 450468.632 | 0.873379818 | -0.058796848 | FALSE | TRUE | FALSE |
| OR7G3 | 0.408683 | 0.76425622 | 1 | 0.408682973 | 0.408682973 | 0.408682973 | 0.45379463 | 0.773159821 | 1 | 0.453794626 | 0.453794626 | 0.453794626 | 0.900590156 | -0.045472804 | FALSE | TRUE | FALSE |
| C17orf50 | 0.4086195 | 0.76428876 | 1 | 0.4086195 | 0.4086195 | 0.4086195 | 0.45340946 | 0.773352249 | 0.00195 | 0.453409462 | 0.453409462 | 0.453409462 | 0.901215203 | -0.045171491 | FALSE | TRUE | FALSE |
| PACS1 | 0.4085454 | 0.7643213 | 1 | 0.408545412 | 0.408545412 | 8.002178986 | 0.42651852 | 0.782328681 | 1 | 4.018429873 | 38.85609686 | 78.90474734 | 0.957860889 | -0.018697559 | FALSE | TRUE | FALSE |
| LRRN4 | 0.4084915 | 0.76435382 | 0.96995 | 35.15332478 | 338.5463564 | 3340.111196 | 0.07589348 | 0.881285259 | 1 | 0.075893479 | 0.075893479 | 1.491333834 | 5.382432578 | 0.730978598 | FALSE | TRUE | FALSE |
| TSPAN5 | 0.408491 | 0.76438634 | 1 | 1.633703502 | 1.633703502 | 1.633703502 | 0.45928526 | 0.771249493 | 0.00615 | 0.459285265 | 0.459285265 | 0.459285265 | 0.889405812 | -0.050900037 | FALSE | TRUE | FALSE |
| AGO2 | 0.4082571 | 0.76441885 | 1 | 0.408257108 | 0.408257108 | 7.968826163 | 0.45407355 | 0.773095601 | 1 | 0.45407355 | 0.45407355 | 8.888800255 | 0.899099071 | -0.046192451 | FALSE | TRUE | FALSE |
| TGFB2 | 0.4082183 | 0.76445134 | 1 | 0.408218255 | 0.408218255 | 8.084994033 | 1.79586189 | 0.508290761 | 0.0094 | 1.795861891 | 1.795861891 | 1.795861891 | 0.227310495 | -0.643380512 | FALSE | FALSE | FALSE |
| PPM1G | 0.4079933 | 0.76448383 | 0.01445 | 0.407993328 | 0.407993328 | 8.082908626 | 0.45126928 | 0.774023104 | 1 | 0.45126928 | 0.45126928 | 8.950406575 | 0.90410171 | -0.043782709 | FALSE | TRUE | FALSE |
| LY6G6C | 0.4079764 | 0.76451631 | 1 | 0.407976423 | 0.407976423 | 0.407976423 | 0.45316499 | 0.773448334 | 1 | 0.453164991 | 0.453164991 | 0.453164991 | 0.900282307 | -0.045621285 | FALSE | TRUE | FALSE |
| FAR2 | 0.4079761 | 0.76454878 | 1 | 1.631802627 | 1.631802627 | 8.07532441 | 0.4586622 | 0.77151048 | 0.01245 | 0.458662199 | 0.458662199 | 9.089856013 | 0.889491342 | -0.050858275 | FALSE | TRUE | FALSE |
| DIO2 | 0.4077422 | 0.76458124 | 1 | 0.407742238 | 0.407742238 | 0.407742238 | 0.44568412 | 0.775604166 | 1 | 0.445684117 | 0.445684117 | 0.445684117 | 0.914868226 | -0.038641456 | FALSE | TRUE | FALSE |
| TBC1D13 | 0.4076994 | 0.7646137 | 1 | 0.407699448 | 0.407699448 | 8.084178161 | 0.4531809 | 0.773416315 | 1 | 0.453180903 | 0.453180903 | 0.453180903 | 0.899639515 | -0.045931477 | FALSE | TRUE | FALSE |
| UPK3A | 0.4073799 | 0.76464614 | 0.25675 | 0.407379913 | 3.826383096 | 3.826383096 | 0.06411726 | 0.883774571 | 1 | 0.589269261 | 5.558354625 | 5.558354625 | 6.353670403 | 0.803024682 | FALSE | TRUE | FALSE |
| GSTA2 | 0.4073796 | 0.76467858 | 1 | 0.40737963 | 3.785649163 | 3.785649163 | 0.46655172 | 0.768337082 | 1 | 0.466551719 | 0.466551719 | 0.466551719 | 0.873171426 | -0.058900485 | FALSE | TRUE | FALSE |
| PHOX2A | 0.4073466 | 0.764711 | 1 | 0.407346623 | 0.407346623 | 0.407346623 | 0.45314572 | 0.773480343 | 1 | 0.453145717 | 0.453145717 | 0.453145717 | 0.89893076 | -0.046273759 | FALSE | TRUE | FALSE |
| OOEP | 0.4073372 | 0.76474342 | 1 | 0.407337228 | 0.407337228 | 0.407337228 | 0.45233155 | 0.773704156 | 1 | 0.452331554 | 0.452331554 | 0.452331554 | 0.900527996 | -0.045502781 | FALSE | TRUE | FALSE |
| PSMD8 | 0.4071441 | 0.76477582 | 1 | 0.407144115 | 0.407144115 | 0.407144115 | 4.11362031 | 0.349488235 | 0.505 | 4.113620311 | 4.113620311 | 4.113620311 | 0.098974646 | -1.004476042 | FALSE | FALSE | FALSE |
| S100PBP | 0.4071332 | 0.76480822 | 0.5064 | 3.797023281 | 3.797023281 | 8.080808687 | 0.4667604 | 0.76820285 | 1 | 0.4667604 | 0.4667604 | 0.4667604 | 0.872253053 | -0.059357502 | FALSE | TRUE | FALSE |
| ZDHHC7 | 0.4071138 | 0.76484061 | 1 | 0.407113824 | 0.407113824 | 0.407113824 | 0.4525622 | 0.773608292 | 0.0097 | 0.452562196 | 0.452562196 | 0.452562196 | 0.899575411 | -0.045962424 | FALSE | TRUE | FALSE |
| B4GALT2 | 0.4069967 | 0.76487299 | 0.0133 | 0.406996699 | 0.406996699 | 8.04870357 | 0.45267623 | 0.773576318 | 1 | 0.452676235 | 0.452676235 | 8.964125643 | 0.89909005 | -0.046196809 | FALSE | TRUE | FALSE |
| KIAA0930 | 0.406937 | 0.76490536 | 1 | 3.810968233 | 3.810968233 | 8.045859441 | 0.46202908 | 0.770265267 | 0.013 | 0.46202908 | 0.46202908 | 9.147668119 | 0.880760686 | -0.055142079 | FALSE | TRUE | FALSE |
| CRX | 0.4069137 | 0.76493772 | 0.93695 | 1.627048491 | 26.03288779 | 26.03288779 | 0.13345103 | 0.868104297 | 1 | 2.133405462 | 34.13964571 | 34.13964571 | 3.049161504 | 0.484180428 | FALSE | TRUE | FALSE |
| REEP2 | 0.406672 | 0.76497007 | 0.0086 | 0.406671976 | 0.406671976 | 0.406671976 | 0.45247506 | 0.773640256 | 1 | 0.452475059 | 0.452475059 | 0.452475059 | 0.898772139 | -0.046350399 | FALSE | TRUE | FALSE |
| MB | 0.4066456 | 0.76500241 | 1 | 0.406645568 | 0.406645568 | 0.406645568 | 0.43821999 | 0.778056853 | 1 | 4.127823357 | 4.127823357 | 4.127823357 | 0.92794847 | -0.03247614 | FALSE | TRUE | FALSE |
| PDCD10 | 0.406642 | 0.76503475 | 1 | 0.406642032 | 0.406642032 | 0.406642032 | 0.41768168 | 0.785494393 | 0.00555 | 0.417681682 | 0.417681682 | 0.417681682 | 0.973569227 | -0.011633162 | FALSE | TRUE | FALSE |
| TMEFF2 | 0.4065807 | 0.76506707 | 1 | 0.406580705 | 0.406580705 | 0.406580705 | 0.45198605 | 0.773799935 | 1 | 0.451986052 | 0.451986052 | 0.451986052 | 0.899542593 | -0.045978268 | FALSE | TRUE | FALSE |
| TM9SF2 | 0.4065665 | 0.76509938 | 1 | 0.40656655 | 0.40656655 | 8.022456788 | 4.14732867 | 0.346912033 | 0.50975 | 4.147328673 | 4.147328673 | 14.84117545 | 0.098030945 | -1.00863681 | FALSE | FALSE | FALSE |
| PRX | 0.4065538 | 0.76513169 | 0.5012 | 3.641660863 | 33.45878684 | 33.45878684 | 0.06985702 | 0.882493503 | 1 | 0.616511697 | 0.616511697 | 0.616511697 | 5.819799572 | 0.764908028 | FALSE | TRUE | FALSE |
| PKMYT1 | 0.4065416 | 0.76516398 | 1 | 0.40654163 | 0.40654163 | 8.042530529 | 1.78785348 | 0.509209918 | 0.50745 | 1.787853477 | 1.787853477 | 8.854064508 | 0.22739091 | -0.6432269 | FALSE | FALSE | FALSE |
| TECR | 0.4064535 | 0.76519627 | 1 | 3.79381665 | 3.79381665 | 3.79381665 | 0.46607956 | 0.768571596 | 1 | 0.466079556 | 0.466079556 | 9.253223793 | 0.872068994 | -0.059449155 | FALSE | TRUE | FALSE |
| OPCML | 0.4061267 | 0.76522855 | 1 | 0.406126687 | 0.406126687 | 8.058938653 | 0.45183166 | 0.773831844 | 0.0089 | 0.451831663 | 0.451831663 | 0.451831663 | 0.898845124 | -0.046315133 | FALSE | TRUE | FALSE |
| CCNG2 | 0.4060184 | 0.76526082 | 1 | 0.406018379 | 0.406018379 | 0.406018379 | 0.45128168 | 0.77399125 | 1 | 0.451281676 | 0.451281676 | 0.451281676 | 0.899700565 | -0.045902007 | FALSE | TRUE | FALSE |
| RNF38 | 0.4058863 | 0.76529308 | 1 | 0.405886254 | 0.405886254 | 8.048136413 | 0.45140632 | 0.773959387 | 1 | 0.451406319 | 0.451406319 | 8.959961501 | 0.899159442 | -0.046163291 | FALSE | TRUE | FALSE |
| PPP3CA | 0.4058218 | 0.76532533 | 1 | 0.405821798 | 0.405821798 | 8.034654129 | 0.45158423 | 0.773895634 | 1 | 0.451584226 | 0.451584226 | 8.951517168 | 0.898662473 | -0.046403394 | FALSE | TRUE | FALSE |
| GNG7 | 0.405793 | 0.76535757 | 1 | 0.405792967 | 0.405792967 | 0.405792967 | 0.40957309 | 0.788565976 | 1 | 0.409573086 | 0.409573086 | 0.409573086 | 0.990770587 | -0.004026895 | FALSE | TRUE | FALSE |
| FAM219B | 0.4056042 | 0.7653898 | 0.0068 | 0.405604244 | 0.405604244 | 0.405604244 | 0.45063476 | 0.774214031 | 0.005 | 0.450634762 | 0.450634762 | 0.450634762 | 0.900073138 | -0.045722199 | FALSE | TRUE | FALSE |
| MRPS10 | 0.4055849 | 0.76542203 | 1 | 0.405584861 | 0.405584861 | 0.405584861 | 0.45093102 | 0.77411861 | 1 | 0.450931016 | 0.450931016 | 0.450931016 | 0.899438821 | -0.046028372 | FALSE | TRUE | FALSE |
| AMT | 0.4053778 | 0.76545424 | 0.6897 | 0.405377826 | 6.483169697 | 6.483169697 | 2.4458458 | 0.451706539 | 0.6623 | 2.445845803 | 39.14295795 | 156.6493575 | 0.165741367 | -0.780569085 | FALSE | FALSE | FALSE |
| NPPB | 0.4052961 | 0.76548645 | 1 | 0.405296125 | 0.405296125 | 0.405296125 | 4.0756821 | 0.352296468 | 0.4994 | 0.436650081 | 4.075682097 | 4.075682097 | 0.099442527 | -1.00242785 | FALSE | FALSE | FALSE |
| ZNF699 | 0.4052718 | 0.76551865 | 0.5131 | 0.405271755 | 0.405271755 | 1.533912675 | 0.06679241 | 0.883159033 | 1 | 0.066792408 | 0.066792408 | 1.323247077 | 6.067632053 | 0.783019237 | FALSE | TRUE | FALSE |
| SHISA3 | 0.4052181 | 0.76555084 | 1 | 0.405218116 | 0.405218116 | 0.405218116 | 0.42317556 | 0.783384848 | 1 | 3.938704592 | 37.62488364 | 37.62488364 | 0.95756502 | -0.018831727 | FALSE | TRUE | FALSE |

| | | | | | | | | | | | | | | | | | |
|---|---|---|---|---|---|---|---|---|---|---|---|---|---|---|---|---|---|
| TOX3 | 0.4052028 | 0.76558301 | 1 | 3.817218206 | 36.90679186 | 75.58894365 | 0.48037097 | 0.763014106 | 1 | 0.480370971 | 0.480370971 | 9.523676814 | 0.843520573 | -0.073904321 | FALSE | TRUE | FALSE |
| AKT1 | 0.4051861 | 0.76561518 | 1 | 0.405186086 | 0.405186086 | 7.991828874 | 0.4509694 | 0.774086784 | 1 | 0.450969404 | 0.450969404 | 8.909632341 | 0.898477995 | -0.046492555 | FALSE | TRUE | FALSE |
| PHF1 | 0.4051114 | 0.76564734 | 1 | 0.405111429 | 0.405111429 | 7.999311967 | 0.45086927 | 0.774150426 | 1 | 0.450869272 | 0.450869272 | 8.91645173 | 0.89851195 | -0.046476142 | FALSE | TRUE | FALSE |
| PAFAH1B3 | 0.4050551 | 0.76567949 | 1 | 3.778770292 | 3.778770292 | 3.778770292 | 0.46463143 | 0.769105759 | 0.0071 | 0.464631427 | 0.464631427 | 0.464631427 | 0.871777275 | -0.059594456 | FALSE | TRUE | FALSE |
| SENP8 | 0.4050466 | 0.76571163 | 1 | 0.40504663 | 0.40504663 | 0.40504663 | 0.45009301 | 0.774341137 | 0.0044 | 0.450093012 | 0.450093012 | 0.450093012 | 0.899917615 | -0.045797247 | FALSE | TRUE | FALSE |
| OR8B12 | 0.4048552 | 0.76574377 | 1 | 0.404855186 | 0.404855186 | 0.404855186 | 0.45004351 | 0.77437289 | 0.00605 | 0.450043507 | 0.450043507 | 0.450043507 | 0.899591217 | -0.045954793 | FALSE | TRUE | FALSE |
| ANXA2R | 0.4047021 | 0.76577589 | 0.5036 | 3.761169864 | 3.761169864 | 3.761169864 | 0.46399986 | 0.769471489 | 1 | 0.463999862 | 0.463999862 | 0.463999862 | 0.872203014 | -0.059382417 | FALSE | TRUE | FALSE |
| NAPB | 0.4046696 | 0.765808 | 1 | 1.618461329 | 1.618461329 | 1.618461329 | 1.78088616 | 0.509973443 | 0.7483 | 1.780886164 | 7.124456097 | 7.124456097 | 0.227229352 | -0.643535571 | FALSE | FALSE | FALSE |
| VPREB1 | 0.4046545 | 0.76584011 | 1 | 0.404654456 | 0.404654456 | 0.404654456 | 0.44980884 | 0.774531512 | 0.0044 | 0.449808843 | 0.449808843 | 0.449808843 | 0.899614275 | -0.045943662 | FALSE | TRUE | FALSE |
| MS4A6E | 0.4045389 | 0.76587221 | 1 | 0.404538855 | 0.404538855 | 0.404538855 | 4.04928244 | 0.354323189 | 0.49865 | 0.435751597 | 4.049282438 | 4.049282438 | 0.099903838 | -1.000417829 | FALSE | FALSE | FALSE |
| M6PR | 0.4044753 | 0.76590429 | 1 | 0.404475317 | 0.404475317 | 0.404475317 | 0.44985978 | 0.774468093 | 0.0063 | 0.449859784 | 0.449859784 | 0.449859784 | 0.899114195 | -0.046185146 | FALSE | TRUE | FALSE |
| OR1K1 | 0.40447 | 0.76593637 | 0.01065 | 0.404470011 | 0.404470011 | 8.031300982 | 37.7047426 | 0.07291073 | 0.25405 | 3.940043782 | 37.70474265 | 37.70474265 | 0.010727298 | -1.969509654 | FALSE | FALSE | FALSE |
| MAP6 | 0.4042215 | 0.76596844 | 1 | 1.616833564 | 6.468741716 | 6.468741716 | 38.5698504 | 0.070130359 | 0.25815 | 4.028656899 | 38.56985044 | 38.56985044 | 0.010480245 | -1.979628573 | FALSE | FALSE | FALSE |
| EPC2 | 0.4041877 | 0.7660005 | 1 | 1.616634999 | 1.616634999 | 13.65127001 | 0.45520256 | 0.77277395 | 1 | 0.455202556 | 0.455202556 | 8.997884215 | 0.887929309 | -0.051621608 | FALSE | TRUE | FALSE |
| S1PR5 | 0.4041829 | 0.76603255 | 1 | 0.404182855 | 0.404182855 | 7.960732063 | 0.44990912 | 0.774436368 | 1 | 0.449909122 | 0.449909122 | 8.877569819 | 0.898365548 | -0.046546912 | FALSE | TRUE | FALSE |
| PIH1D1 | 0.4040386 | 0.76606459 | 1 | 0.40403855 | 0.40403855 | 0.40403855 | 3.93861272 | 0.358844096 | 0.7483 | 3.938612722 | 37.71392514 | 37.71392514 | 0.102583975 | -0.988920475 | FALSE | FALSE | FALSE |
| FAM110C | 0.4040051 | 0.76609663 | 1 | 0.404005075 | 0.404005075 | 0.404005075 | 0.44932641 | 0.774689901 | 1 | 0.449326406 | 0.449326406 | 0.449326406 | 0.89913495 | -0.046175121 | FALSE | TRUE | FALSE |
| LPGAT1 | 0.4039251 | 0.76612865 | 1 | 0.403925075 | 0.403925075 | 0.403925075 | 0.44976858 | 0.774563208 | 1 | 0.449768576 | 0.449768576 | 0.449768576 | 0.898073135 | -0.046688295 | FALSE | TRUE | FALSE |
| RXRB | 0.4038987 | 0.76616067 | 0.7508 | 0.946606089 | 3.784551748 | 7.989780583 | 0.13677507 | 0.867458475 | 1 | 0.546861591 | 0.546861591 | 2.709189246 | 2.953014351 | 0.470265558 | FALSE | TRUE | FALSE |
| OR52N1 | 0.403744 | 0.76619267 | 1 | 0.403743986 | 0.403743986 | 0.403743986 | 0.43521699 | 0.779186639 | 1 | 4.065593415 | 4.065593415 | 4.065593415 | 0.927684341 | -0.032599774 | FALSE | TRUE | FALSE |
| MIER3 | 0.4037032 | 0.76622467 | 1 | 0.403703167 | 0.403703167 | 7.99476218 | 0.43431972 | 0.779520201 | 1 | 0.43431972 | 0.43431972 | 8.611240575 | 0.929506879 | -0.031747392 | FALSE | TRUE | FALSE |
| MMP24 | 0.4036493 | 0.76625666 | 0.01835 | 0.403649317 | 0.403649317 | 7.961067045 | 4.10308375 | 0.350000758 | 0.5156 | 4.103083753 | 4.103083753 | 14.7495505 | 0.09837706 | -1.00710616 | FALSE | FALSE | FALSE |
| ADAR | 0.4035958 | 0.76628863 | 0.2692 | 0.403595812 | 3.807668349 | 13.62708515 | 0.60388779 | 0.710473627 | 0.51275 | 0.603887791 | 0.603887791 | 11.86653798 | 0.668329146 | -0.175009599 | FALSE | TRUE | FALSE |
| GPR45 | 0.4035275 | 0.7663206 | 1 | 0.403527468 | 0.403527468 | 7.977820663 | 0.44940697 | 0.774626573 | 1 | 0.449406972 | 0.449406972 | 8.897149774 | 0.897911008 | -0.046766704 | FALSE | TRUE | FALSE |
| FAM178B | 0.4034918 | 0.76635256 | 0.75265 | 0.403491844 | 3.646206084 | 3.646206084 | 0.06911252 | 0.882616615 | 1 | 0.06911252 | 0.06911252 | 0.06911252 | 5.838187419 | 0.766278033 | FALSE | TRUE | FALSE |
| SHB | 0.4034561 | 0.76638451 | 1 | 6.455266156 | 25.83068501 | 31.95924984 | 0.46465847 | 0.769072449 | 1 | 0.464658474 | 0.464658474 | 9.213742652 | 0.868285159 | -0.061337622 | FALSE | TRUE | FALSE |
| MBTD1 | 0.403277 | 0.76641646 | 1 | 36.44497816 | 3644.931787 | 3644.931787 | 0.47804453 | 0.763820118 | 1 | 4.500368678 | 43.48229098 | 43.48229098 | 0.843597053 | -0.073864946 | FALSE | TRUE | FALSE |
| BLVRA | 0.4031494 | 0.76644839 | 0.008 | 0.403149363 | 0.403149363 | 0.403149363 | 0.44841644 | 0.77484807 | 0.0077 | 0.448416443 | 0.448416443 | 0.448416443 | 0.899051249 | -0.046215551 | FALSE | TRUE | FALSE |
| FARS2 | 0.4030753 | 0.76648031 | 0.972 | 8.92584084 | 142.7344175 | 570.9936851 | 2.1796747 | 0.472912457 | 0.6889 | 5.079481201 | 81.27503353 | 81.27503353 | 0.184924501 | -0.733005544 | FALSE | FALSE | FALSE |
| NDUFB4 | 0.4030587 | 0.76651223 | 0.00435 | 0.403058719 | 0.403058719 | 0.403058719 | 0.44833568 | 0.774879677 | 1 | 0.448335678 | 0.448335678 | 0.448335678 | 0.899011029 | -0.04623498 | FALSE | TRUE | FALSE |
| PRH1 | 0.4026606 | 0.76654414 | 0.497 | 0.402660554 | 3.737657253 | 3.737657253 | 0.46182482 | 0.770364085 | 1 | 0.461824815 | 0.461824815 | 0.461824815 | 0.87189025 | -0.059538179 | FALSE | TRUE | FALSE |
| SOCS3 | 0.4026511 | 0.76657604 | 1 | 6.442540015 | 25.78006836 | 25.78006836 | 0.43303524 | 0.780093892 | 1 | 1.731971023 | 6.928936218 | 6.928936218 | 0.929834448 | -0.031594368 | FALSE | TRUE | FALSE |
| WFDC5 | 0.4024863 | 0.76660793 | 1 | 0.402486298 | 3.774965999 | 3.774965999 | 0.46230159 | 0.770133372 | 1 | 0.462301591 | 0.462301591 | 0.462301591 | 0.870614132 | -0.060174288 | FALSE | TRUE | FALSE |
| IGFL1 | 0.4024614 | 0.76663981 | 0.00385 | 0.402461414 | 0.402461414 | 0.402461414 | 0.44741606 | 0.775163739 | 1 | 0.447416063 | 0.447416063 | 0.447416063 | 0.899523837 | -0.045987323 | FALSE | TRUE | FALSE |
| KCNMB1 | 0.4023726 | 0.76667168 | 0.0051 | 0.402372555 | 0.402372555 | 0.402372555 | 0.44747569 | 0.775132213 | 1 | 0.447475693 | 0.447475693 | 0.447475693 | 0.89920539 | -0.046141099 | FALSE | TRUE | FALSE |
| BMP10 | 0.4023094 | 0.76670355 | 1 | 0.402309393 | 0.402309393 | 7.978082755 | 6.93469015 | 0.246038067 | 0.5005 | 6.934690145 | 27.74783106 | 27.74783106 | 0.05801404 | -1.236466888 | FALSE | FALSE | FALSE |
| GOLGA4 | 0.4021608 | 0.7667354 | 0.1629 | 0.402160847 | 3.597990273 | 13.40884202 | 0.0106702 | 0.896583052 | 1 | 0.0106702 | 0.0106702 | 0.356594818 | 37.69009573 | 1.576227241 | FALSE | TRUE | FALSE |
| ZNF558 | 0.4019182 | 0.76676725 | 1 | 0.401918157 | 0.401918157 | 7.978084018 | 0.44738387 | 0.775195255 | 1 | 0.447383871 | 0.447383871 | 0.447383871 | 0.898374266 | -0.046542697 | FALSE | TRUE | FALSE |
| NOA1 | 0.4017827 | 0.76679909 | 0.50585 | 0.401782674 | 0.401782674 | 7.884122428 | 0.06288279 | 0.884041488 | 0.881 | 1.006077748 | 9.061799794 | 19.78694539 | 6.389389824 | 0.805459386 | FALSE | TRUE | FALSE |
| TXNDC15 | 0.4016971 | 0.76683092 | 1 | 35.93717037 | 353.1944089 | 353.1944089 | 0.49118283 | 0.758348029 | 1 | 0.491182831 | 0.491182831 | 0.491182831 | 0.81781582 | -0.087344493 | FALSE | TRUE | FALSE |
| FAM53C | 0.4015862 | 0.76686274 | 1 | 0.401586188 | 0.401586188 | 7.954696444 | 0.44725129 | 0.775321225 | 0.01095 | 0.447251295 | 0.447251295 | 8.869459341 | 0.897898324 | -0.046772839 | FALSE | TRUE | FALSE |
| LARP6 | 0.4015274 | 0.76689455 | 1 | 3.757082701 | 144.3177 | 144.3177 | 0.48139831 | 0.762556023 | 0.0123 | 0.48139831 | 0.48139831 | 9.544622469 | 0.834085687 | -0.078789331 | FALSE | TRUE | FALSE |
| CCDC12 | 0.4013731 | 0.76692635 | 0.0052 | 0.401373126 | 0.401373126 | 0.401373126 | 0.44697264 | 0.775352696 | 1 | 0.446972635 | 0.446972635 | 0.446972635 | 0.89798143 | -0.046732644 | FALSE | TRUE | FALSE |
| MYL4 | 0.4012378 | 0.76695815 | 0.7478 | 0.401237775 | 1.604656178 | 1.604656178 | 0.52586798 | 0.744567896 | 0.2527 | 0.525867983 | 2.103192952 | 2.103192952 | 0.763000958 | -0.117474917 | FALSE | TRUE | FALSE |
| FN3KRP | 0.4009457 | 0.76698994 | 0.50675 | 0.400945711 | 1.603206036 | 1.603206036 | 0.06656578 | 0.883219985 | 1 | 0.066565778 | 0.066565778 | 0.066565778 | 6.023300859 | 0.779834556 | FALSE | TRUE | FALSE |
| ALCAM | 0.4008375 | 0.76702172 | 1 | 0.400837476 | 0.400837476 | 7.933801598 | 0.44659582 | 0.775384159 | 1 | 0.44659582 | 0.44659582 | 8.850501598 | 0.897539694 | -0.046946335 | FALSE | TRUE | FALSE |
| ASIC4 | 0.4008366 | 0.76705349 | 0.96895 | 34.74272634 | 335.7919489 | 3324.806773 | 462.050797 | 0.004989745 | 0.24055 | 462.050797 | 46831.69161 | 90752.28621 | 0.000867516 | -3.061722362 | FALSE | FALSE | FALSE |
| HLA-E | 0.4008168 | 0.76708525 | 1 | 0.400816818 | 0.400816818 | 7.935515563 | 0.44610153 | 0.775447063 | 1 | 0.446101533 | 0.446101533 | 8.842789894 | 0.898487875 | -0.046487779 | FALSE | TRUE | FALSE |
| PFN3 | 0.4008067 | 0.767117 | 1 | 0.400806681 | 0.400806681 | 0.400806681 | 0.44640914 | 0.775415615 | 1 | 0.446409143 | 0.446409143 | 0.446409143 | 0.89784604 | -0.046798129 | FALSE | TRUE | FALSE |
| GRN | 0.4007169 | 0.76714874 | 0.51085 | 0.400716906 | 1.602150164 | 7.895893406 | 0.26078428 | 0.836456571 | 0.51225 | 0.260784278 | 0.260784278 | 2.207912308 | 1.536583836 | 0.18655626 | FALSE | TRUE | FALSE |
| MED28 | 0.4006609 | 0.76718047 | 1 | 0.4006609 | 0.4006609 | 0.4006609 | 0.44572894 | 0.775541352 | 1 | 0.44572894 | 0.44572894 | 0.44572894 | 0.898889132 | -0.04629387 | FALSE | TRUE | FALSE |
| GYPE | 0.4002511 | 0.7672122 | 1 | 0.400251132 | 0.400251132 | 0.400251132 | 0.44516861 | 0.775698323 | 1 | 0.445168606 | 0.445168606 | 0.445168606 | 0.899100086 | -0.046191961 | FALSE | TRUE | FALSE |
| CDIP1 | 0.4001716 | 0.76724392 | 1 | 0.400171618 | 0.400171618 | 0.400171618 | 0.44600592 | 0.77550993 | 1 | 0.446005923 | 0.446005923 | 0.446005923 | 0.897233865 | -0.047094343 | FALSE | TRUE | FALSE |
| C17orf75 | 0.4000248 | 0.76727563 | 1 | 3.725866826 | 3.725866826 | 7.948734022 | 38.5420604 | 0.070408501 | 0.25425 | 4.026386609 | 38.54206037 | 38.54206037 | 0.010378915 | -1.983848057 | FALSE | FALSE | FALSE |
| NTS | 0.3998887 | 0.76730733 | 1 | 0.399888745 | 0.399888745 | 0.399888745 | 0.44489976 | 0.7757924 | 1 | 0.444899757 | 0.444899757 | 0.444899757 | 0.898828868 | -0.046322988 | FALSE | TRUE | FALSE |
| NUP98 | 0.3998366 | 0.76733902 | 0.764 | 0.399836635 | 1.599334748 | 30.9390489 | 0.13425009 | 0.86792973 | 1 | 0.134250093 | 0.134250093 | 4.511429381 | 2.978296888 | 0.473967988 | FALSE | TRUE | FALSE |
| CDK4 | 0.3997898 | 0.7673707 | 1 | 0.399789771 | 0.399789771 | 0.399789771 | 0.44532412 | 0.775635561 | 1 | 0.445324122 | 0.445324122 | 0.445324122 | 0.897750091 | -0.046844542 | FALSE | TRUE | FALSE |
| VTA1 | 0.3991642 | 0.76740238 | 1 | 0.399164249 | 0.399164249 | 0.399164249 | 0.44470683 | 0.775823741 | 1 | 0.444706833 | 0.444706833 | 0.444706833 | 0.897589646 | -0.046922166 | FALSE | TRUE | FALSE |
| PNLIPRP2 | 0.3991253 | 0.76743405 | 1 | 3.725521525 | 3.725521525 | 7.916023141 | 0.45868417 | 0.771445292 | 1 | 0.458684167 | 0.458684167 | 0.458684167 | 0.870152838 | -0.060404459 | FALSE | TRUE | FALSE |
| HNRNPA3 | 0.3988252 | 0.76746571 | 1 | 0.398825203 | 0.398825203 | 0.398825203 | 4.05613738 | 0.353311549 | 0.5107 | 4.056137377 | 4.056137377 | 4.056137377 | 0.098326355 | -1.007330061 | FALSE | FALSE | FALSE |
| GTF2F2 | 0.3987979 | 0.76749736 | 1 | 1.595105828 | 1.595105828 | 1.595105828 | 0.44924004 | 0.774721551 | 1 | 0.449240041 | 0.449240041 | 0.449240041 | 0.88771684 | -0.051725542 | FALSE | TRUE | FALSE |
| NCDN | 0.3987253 | 0.76752901 | 1 | 0.398725333 | 0.398725333 | 7.83009418 | 0.44454023 | 0.775886397 | 0.0203 | 0.444540233 | 0.444540233 | 8.748833789 | 0.896938687 | -0.047237243 | FALSE | TRUE | FALSE |
| SKP2 | 0.3985911 | 0.76756064 | 1 | 0.398591119 | 0.398591119 | 7.905436334 | 0.44405942 | 0.775980316 | 1 | 0.444059423 | 0.444059423 | 8.816050175 | 0.897607613 | -0.046913473 | FALSE | TRUE | FALSE |
| SLC35E1 | 0.3984424 | 0.76759227 | 0.0102 | 0.398442351 | 0.398442351 | 7.911236201 | 0.44405791 | 0.776011604 | 1 | 0.444057907 | 0.444057907 | 0.444057907 | 0.897275658 | -0.047074114 | FALSE | TRUE | FALSE |
| EMP2 | 0.3984302 | 0.76762389 | 0.5043 | 0.398430223 | 0.398430223 | 0.398430223 | 0.13083177 | 0.868755078 | 1 | 0.130831766 | 0.523086496 | 0.523086496 | 3.045363035 | 0.483639072 | FALSE | TRUE | FALSE |
| ATF4 | 0.3983555 | 0.76765549 | 1 | 0.398355528 | 0.398355528 | 0.398355528 | 4.01454272 | 0.355583665 | 0.501 | 4.014542717 | 4.014542717 | 4.014542717 | 0.09922812 | -1.003365235 | FALSE | FALSE | FALSE |
| NICN1 | 0.3982225 | 0.7676871 | 1 | 0.398222532 | 0.398222532 | 0.398222532 | 0.44341102 | 0.776323977 | 1 | 0.443411019 | 0.443411019 | 0.443411019 | 0.898088939 | -0.046680653 | FALSE | TRUE | FALSE |
| RHOV | 0.3981929 | 0.76771869 | 1 | 0.398192933 | 0.398192933 | 0.398192933 | 0.44409965 | 0.775949018 | 0.00775 | 0.444099649 | 0.444099649 | 0.444099649 | 0.896629693 | -0.047386883 | FALSE | TRUE | FALSE |
| MEF2A | 0.3980849 | 0.76775027 | 1 | 0.398084934 | 0.398084934 | 0.398084934 | 0.4439939 | 0.776074151 | 1 | 0.4439939 | 0.4439939 | 0.4439939 | 0.896600007 | -0.047401262 | FALSE | TRUE | FALSE |
| LGALSL | 0.3980248 | 0.76778184 | 1 | 0.398024786 | 0.398024786 | 0.398024786 | 0.44386836 | 0.776105412 | 1 | 0.443868364 | 0.443868364 | 0.443868364 | 0.896718076 | -0.047344076 | FALSE | TRUE | FALSE |
| RRAGA | 0.3979148 | 0.76781341 | 1 | 0.39791476 | 0.39791476 | 7.889449892 | 0.44368009 | 0.776167905 | 1 | 0.443680086 | 0.443680086 | 0.527303193 | 0.89685062 | -0.047279887 | FALSE | TRUE | FALSE |
| GNG8 | 0.397905 | 0.76784496 | 0.00325 | 0.39790502 | 0.39790502 | 0.39790502 | 0.40460715 | 0.790708526 | 1 | 0.404607155 | 0.404607155 | 0.404607155 | 0.983435452 | -0.00725414 | FALSE | TRUE | FALSE |
| IL7R | 0.3978687 | 0.76787651 | 0.50775 | 0.397868655 | 1.590883087 | 1.590883087 | 0.06531228 | 0.883515211 | 1 | 0.261163463 | 0.261163463 | 0.261163463 | 6.091789386 | 0.78474488 | FALSE | TRUE | FALSE |

| | | | | | | | | | | | | | | | | | |
|---|---|---|---|---|---|---|---|---|---|---|---|---|---|---|---|---|---|
| PRPF19 | 0.3978419 | 0.76790805 | 1 | 0.397841901 | 0.397841901 | 7.87232676 | 0.44380796 | 0.776136663 | 1 | 0.443807965 | 0.443807965 | 8.793104441 | 0.896428033 | -0.047484571 | FALSE | TRUE | FALSE |
| DOC2A | 0.3976017 | 0.76793957 | 0.7496 | 0.915087455 | 3.659233315 | 3.659233315 | 1.21746196 | 0.565730729 | 0.01305 | 1.217461962 | 1.217461962 | 2.611197762 | 0.32658241 | -0.48600721 | FALSE | FALSE | FALSE |
| ZNF92 | 0.3973084 | 0.7679711 | 1 | 0.397308389 | 0.397308389 | 7.871879611 | 3.88787834 | 0.362074786 | 0.2557 | 3.887878344 | 37.29708768 | 37.29708768 | 0.102191569 | -0.990584932 | FALSE | FALSE | FALSE |
| RIMKLA | 0.3972634 | 0.76800261 | 1 | 0.397263373 | 0.397263373 | 0.397263373 | 0.44290226 | 0.776417516 | 1 | 0.442902263 | 0.442902263 | 0.442902263 | 0.89695494 | -0.047229374 | FALSE | TRUE | FALSE |
| BLOC1S5 | 0.3971246 | 0.76803412 | 1 | 1.588485059 | 1.588485059 | 1.588485059 | 35.9370032 | 0.07709207 | 0.49625 | 3.772786919 | 351.0959456 | 351.0959456 | 0.011050578 | -1.956615014 | FALSE | FALSE | FALSE |
| PKDCC | 0.3970913 | 0.76806561 | 1 | 0.397091305 | 0.397091305 | 0.397091305 | 0.43776727 | 0.778179569 | 1 | 1.750986598 | 1.750986598 | 1.750986598 | 0.907083123 | -0.042352913 | FALSE | TRUE | FALSE |
| GPATCH3 | 0.397088 | 0.7680971 | 0.75955 | 0.397088049 | 3.583871247 | 7.830448679 | 0.06620498 | 0.883298268 | 1 | 0.587734444 | 0.587734444 | 1.307743192 | 5.997857293 | 0.777996128 | FALSE | TRUE | FALSE |
| PI15 | 0.3969852 | 0.76812858 | 1 | 0.396985212 | 0.396985212 | 0.396985212 | 0.44241819 | 0.77657324 | 1 | 0.442418189 | 0.442418189 | 0.442418189 | 0.897307618 | -0.047058645 | FALSE | TRUE | FALSE |
| TFCP2 | 0.3969446 | 0.76816004 | 1 | 0.396944614 | 0.396944614 | 7.866937229 | 0.44247733 | 0.776510978 | 1 | 0.442477332 | 0.442477332 | 8.778897483 | 0.897095932 | -0.047161113 | FALSE | TRUE | FALSE |
| LCE2D | 0.396751 | 0.7681915 | 1 | 0.396750991 | 0.396750991 | 0.396750991 | 0.44175975 | 0.776790881 | 1 | 0.441759752 | 0.441759752 | 0.441759752 | 0.898114843 | -0.046668126 | FALSE | TRUE | FALSE |
| MRPL51 | 0.3966152 | 0.76822296 | 0.0054 | 0.396615187 | 0.396615187 | 0.396615187 | 0.44202806 | 0.776635469 | 1 | 0.442028059 | 0.442028059 | 0.442028059 | 0.897262468 | -0.047080498 | FALSE | TRUE | FALSE |
| NCLN | 0.3965451 | 0.7682544 | 0.018 | 0.396545092 | 0.396545092 | 7.818461023 | 0.44243782 | 0.776542114 | 1 | 0.442437816 | 0.442437816 | 8.738186811 | 0.896273053 | -0.047559661 | FALSE | TRUE | FALSE |
| HP | 0.3965028 | 0.76828584 | 0.5007 | 0.396502789 | 0.396502789 | 1.967087288 | 0.52037398 | 0.747213451 | 0.49975 | 0.130136946 | 0.520373982 | 2.58404098 | 0.761957367 | -0.118069327 | FALSE | TRUE | FALSE |
| TMEM18 | 0.3964496 | 0.76831726 | 1 | 0.396449576 | 0.396449576 | 0.396449576 | 0.44180721 | 0.776759817 | 1 | 0.441807208 | 0.441807208 | 0.441807208 | 0.897336144 | -0.047044839 | FALSE | TRUE | FALSE |
| SLC39A13 | 0.396411 | 0.76834868 | 1 | 0.396411013 | 0.396411013 | 7.856877518 | 0.44201495 | 0.77666657 | 1 | 0.442014952 | 0.442014952 | 0.442014952 | 0.896827158 | -0.047291249 | FALSE | TRUE | FALSE |
| YAF2 | 0.3963405 | 0.76838009 | 1 | 0.396340524 | 0.396340524 | 0.396340524 | 0.42164067 | 0.783938036 | 0.00125 | 0.42164067 | 0.42164067 | 0.42164067 | 0.939995955 | -0.026874015 | FALSE | TRUE | FALSE |
| RPP40 | 0.3963096 | 0.76841148 | 1 | 3.70046545 | 3.70046545 | 3.70046545 | 0.45584257 | 0.772451362 | 0.00865 | 0.455842568 | 0.455842568 | 0.455842568 | 0.869400118 | -0.060780305 | FALSE | TRUE | FALSE |
| CRTC3 | 0.3962613 | 0.76844287 | 1 | 0.39626127 | 0.39626127 | 7.831168046 | 0.44186612 | 0.776728744 | 1 | 0.441866124 | 0.441866124 | 8.744909528 | 0.896790336 | -0.04730908 | FALSE | TRUE | FALSE |
| OR10H1 | 0.3962488 | 0.76847425 | 1 | 0.396248817 | 0.396248817 | 7.862053824 | 0.4416187 | 0.776884018 | 1 | 0.441618701 | 0.441618701 | 0.441618701 | 0.897264578 | -0.047079477 | FALSE | TRUE | FALSE |
| CCDC112 | 0.3961637 | 0.76850563 | 1 | 0.396163711 | 0.396163711 | 7.872554773 | 0.44161307 | 0.776946061 | 1 | 0.44161307 | 0.44161307 | 0.44161307 | 0.897083302 | -0.047167227 | FALSE | TRUE | FALSE |
| CAMK2B | 0.396086 | 0.76853699 | 1 | 0.396085964 | 0.396085964 | 7.810163448 | 0.42823202 | 0.781707673 | 0.5076 | 4.023898672 | 4.023898672 | 8.458341544 | 0.924933083 | -0.033889686 | FALSE | TRUE | FALSE |
| SH2D1B | 0.395969 | 0.76856834 | 1 | 0.395968984 | 0.395968984 | 0.395968984 | 0.44151983 | 0.776977069 | 1 | 0.441519832 | 0.441519832 | 0.441519832 | 0.896831706 | -0.047289046 | FALSE | TRUE | FALSE |
| FBXL7 | 0.395943 | 0.76859969 | 1 | 3.723153926 | 3.723153926 | 14.01118293 | 0.45602446 | 0.772419049 | 1 | 0.456024458 | 0.456024458 | 8.99311522 | 0.86824955 | -0.061355433 | FALSE | TRUE | FALSE |
| NKX2-3 | 0.3959254 | 0.76863103 | 1 | 1.583381601 | 1.583381601 | 1.583381601 | 0.44192616 | 0.776697662 | 1 | 1.767389679 | 1.767389679 | 1.767389679 | 0.89590843 | -0.047736377 | FALSE | TRUE | FALSE |
| CAP2 | 0.3958572 | 0.76866235 | 1 | 0.395857173 | 0.395857173 | 7.843104772 | 0.44161427 | 0.776915044 | 1 | 0.441614269 | 0.441614269 | 8.759530368 | 0.896386737 | -0.047504578 | FALSE | TRUE | FALSE |
| OR1L4 | 0.3958393 | 0.76869367 | 1 | 0.395839281 | 0.395839281 | 0.395839281 | 0.42763534 | 0.781944678 | 1 | 3.984675477 | 3.984675477 | 3.984675477 | 0.925646795 | -0.033554698 | FALSE | TRUE | FALSE |
| WASL | 0.395812 | 0.76872498 | 0.0131 | 0.395812045 | 0.395812045 | 0.833765647 | 0.06123078 | 0.884503727 | 1 | 0.576732544 | 0.576732544 | 1.214840775 | 6.464265635 | 0.810519195 | FALSE | TRUE | FALSE |
| BTBD9 | 0.3957846 | 0.76875628 | 0.39015 | 0.176558521 | 1.582440007 | 1.955352064 | 0.07527975 | 0.881365898 | 0.9402 | 0.301008476 | 4.81620978 | 5.953361447 | 5.257517095 | 0.720780693 | FALSE | TRUE | FALSE |
| AMZ2 | 0.3956585 | 0.76878757 | 0.8725 | 3.572079112 | 33.07866827 | 33.07866827 | 0.06406134 | 0.883783199 | 0.8792 | 5.175764043 | 48.34333495 | 48.34333495 | 6.176245113 | 0.790724523 | FALSE | TRUE | FALSE |
| PAK2 | 0.3956006 | 0.76881886 | 1 | 3.73731164 | 3.73731164 | 7.833595747 | 0.45578205 | 0.77254824 | 1 | 0.455782048 | 0.455782048 | 9.036054567 | 0.867960103 | -0.061500237 | FALSE | TRUE | FALSE |
| ZNF653 | 0.3954838 | 0.76885013 | 1 | 0.39548375 | 0.39548375 | 7.801025114 | 0.44117149 | 0.777070042 | 1 | 0.441171491 | 0.441171491 | 8.716613721 | 0.896439951 | -0.047478797 | FALSE | TRUE | FALSE |
| MAPKAPK5 | 0.3954472 | 0.7688814 | 0.01365 | 0.395447165 | 0.395447165 | 7.835921616 | 36.1106451 | 0.076811528 | 0.5027 | 36.1106451 | 355.8099483 | 355.8099483 | 0.010950986 | -1.960546781 | FALSE | FALSE | FALSE |
| DHX15 | 0.3954439 | 0.76891265 | 1 | 0.395443853 | 0.395443853 | 7.779944482 | 0.44141708 | 0.777008069 | 1 | 0.441417079 | 0.441417079 | 8.701462572 | 0.895850822 | -0.047764303 | FALSE | TRUE | FALSE |
| TMEM74B | 0.3953827 | 0.7689439 | 1 | 0.395382742 | 0.395382742 | 0.395382742 | 0.44080309 | 0.777193884 | 0.0066 | 0.440803085 | 0.440803085 | 0.440803085 | 0.896960017 | -0.047226916 | FALSE | TRUE | FALSE |
| MRPL12 | 0.3953727 | 0.76897514 | 1 | 0.395372665 | 0.395372665 | 0.395372665 | 0.44120302 | 0.77703906 | 0.00535 | 0.441203023 | 0.441203023 | 0.441203023 | 0.896124106 | -0.04763184 | FALSE | TRUE | FALSE |
| LPCAT1 | 0.3952116 | 0.76900637 | 0.5145 | 0.395211624 | 0.395211624 | 7.792061218 | 0.13147245 | 0.868571301 | 1 | 0.131472445 | 0.131472445 | 2.59656078 | 3.006041483 | 0.477994969 | FALSE | TRUE | FALSE |
| C10orf62 | 0.3951893 | 0.76903759 | 1 | 0.395189317 | 0.395189317 | 0.395189317 | 0.4403774 | 0.777317583 | 1 | 0.440377397 | 0.440377397 | 0.440377397 | 0.89738783 | -0.047019825 | FALSE | TRUE | FALSE |
| RRS1 | 0.3951589 | 0.7690688 | 0.01185 | 0.395158913 | 0.395158913 | 1.957147014 | 0.13090684 | 0.868744281 | 0.01115 | 0.130906836 | 0.130906836 | 2.595845914 | 3.018627022 | 0.479809455 | FALSE | TRUE | FALSE |
| FAM210A | 0.3951211 | 0.76910001 | 1 | 0.395121101 | 0.395121101 | 0.395121101 | 0.44053615 | 0.777255751 | 1 | 0.440536149 | 0.440536149 | 0.440536149 | 0.896909601 | -0.047251327 | FALSE | TRUE | FALSE |
| RAD21 | 0.3951042 | 0.7691312 | 1 | 0.3951042 | 0.3951042 | 7.811282802 | 0.44097129 | 0.777131981 | 1 | 0.440971288 | 0.440971288 | 8.730136025 | 0.895986225 | -0.047698667 | FALSE | TRUE | FALSE |
| TXK | 0.3950853 | 0.76916238 | 0.26295 | 0.098838555 | 0.39508528 | 0.875278104 | 0.17009003 | 0.859674002 | 0.01315 | 0.170090027 | 0.170090027 | 0.374685236 | 2.322800977 | 0.366012 | FALSE | TRUE | FALSE |
| RNASE10 | 0.3949356 | 0.76919356 | 1 | 0.394935575 | 0.394935575 | 0.394935575 | 0.42678443 | 0.782181154 | 1 | 3.980027471 | 3.980027471 | 3.980027471 | 0.925374847 | -0.033682309 | FALSE | TRUE | FALSE |
| TMEM169 | 0.3949312 | 0.76922473 | 1 | 0.39493123 | 0.39493123 | 0.39493123 | 0.44025398 | 0.777410262 | 0.0081 | 0.44025398 | 0.44025398 | 0.44025398 | 0.897053174 | -0.047181813 | FALSE | TRUE | FALSE |
| ERMN | 0.3947655 | 0.76925589 | 1 | 0.394765479 | 0.394765479 | 0.394765479 | 0.44012687 | 0.777441138 | 1 | 0.440126869 | 0.440126869 | 0.440126869 | 0.89693565 | -0.047238714 | FALSE | TRUE | FALSE |
| SLC24A3 | 0.3947631 | 0.76928704 | 1 | 0.39476309 | 0.39476309 | 7.786963275 | 0.44072865 | 0.777224821 | 1 | 0.440728648 | 0.440728648 | 8.708015237 | 0.895705536 | -0.047834741 | FALSE | TRUE | FALSE |
| PCSK4 | 0.3946532 | 0.76931818 | 0.85975 | 1.578301064 | 25.26176633 | 123.86366 | 0.04705825 | 0.887315819 | 0.75415 | 0.107016729 | 0.427720497 | 2.104354046 | 8.386481895 | 0.923579814 | FALSE | TRUE | FALSE |
| CIB1 | 0.3943904 | 0.76934932 | 1 | 0.394390421 | 0.394390421 | 0.394390421 | 0.42648109 | 0.782358161 | 0.5099 | 4.001555447 | 4.001555447 | 4.001555447 | 0.924754773 | -0.033973419 | FALSE | TRUE | FALSE |
| KLRC1 | 0.3943837 | 0.76938044 | 1 | 3.667710571 | 35.00753443 | 35.00753443 | 0.46793407 | 0.767900231 | 1 | 0.467934071 | 0.467934071 | 0.467934071 | 0.842818839 | -0.074265765 | FALSE | TRUE | FALSE |
| IPO11 | 0.3943635 | 0.76941156 | 0.87715 | 1.57760862 | 6.312630462 | 31.04521757 | 0.13375064 | 0.868049792 | 1 | 0.133750635 | 0.133750635 | 2.637123427 | 2.948498112 | 0.469600854 | FALSE | TRUE | FALSE |
| PAQR8 | 0.3941039 | 0.76944267 | 1 | 0.394103869 | 0.394103869 | 7.815466953 | 0.43967132 | 0.777502866 | 1 | 0.439671317 | 0.439671317 | 0.439671317 | 0.896360198 | -0.047517436 | FALSE | TRUE | FALSE |
| FBXL20 | 0.3939586 | 0.76947377 | 1 | 3.67919529 | 3.67919529 | 7.814409764 | 0.45349357 | 0.773288144 | 1 | 0.453493575 | 0.453493575 | 9.004213275 | 0.868719358 | -0.061120501 | FALSE | TRUE | FALSE |
| UBXN2A | 0.3939138 | 0.76950486 | 1 | 0.39391376 | 0.39391376 | 0.39391376 | 0.43439152 | 0.77948992 | 1 | 1.737512054 | 1.737512054 | 1.737512054 | 0.906817331 | -0.042480188 | FALSE | TRUE | FALSE |
| ISM1 | 0.3938546 | 0.76953594 | 1 | 0.393854583 | 0.393854583 | 7.795450654 | 0.43952837 | 0.777533717 | 1 | 0.43952837 | 0.43952837 | 8.710305069 | 0.896084552 | -0.04765101 | FALSE | TRUE | FALSE |
| FBXO21 | 0.3935777 | 0.76956702 | 1 | 0.39357771 | 0.39357771 | 7.76728752 | 0.43942817 | 0.77756456 | 1 | 0.439428175 | 0.439428175 | 8.685973275 | 0.895658795 | -0.047857405 | FALSE | TRUE | FALSE |
| GAP43 | 0.3933816 | 0.76959809 | 1 | 3.655005033 | 34.85400522 | 34.85400522 | 3.8316741 | 0.3650299 | 0.69185 | 36.56527429 | 3590.616256 | 3590.616256 | 0.10266572 | -0.988574542 | FALSE | FALSE | FALSE |
| THOC7 | 0.3933061 | 0.76962915 | 1 | 0.39330607 | 0.39330607 | 0.39330607 | 1.73488713 | 0.514358163 | 0.50135 | 1.734887131 | 1.734887131 | 1.734887131 | 0.226704126 | -0.644540576 | FALSE | FALSE | FALSE |
| CD28 | 0.393077 | 0.7696602 | 1 | 0.393076959 | 0.393076959 | 0.393076959 | 0.43891147 | 0.777718644 | 1 | 0.438911467 | 0.438911467 | 0.438911467 | 0.895572315 | -0.04789934 | FALSE | TRUE | FALSE |
| PPP3CB | 0.3928284 | 0.76969125 | 1 | 0.392828447 | 0.392828447 | 7.782489905 | 0.43867921 | 0.777780217 | 1 | 0.438679212 | 0.438679212 | 8.7007152 | 0.895479969 | -0.047944125 | FALSE | TRUE | FALSE |
| FPR2 | 0.392766 | 0.76972228 | 1 | 1.570918493 | 1.570918493 | 1.570918493 | 0.44346324 | 0.776292781 | 1 | 0.443463241 | 0.443463241 | 0.443463241 | 0.885678915 | -0.052723694 | FALSE | TRUE | FALSE |
| SEH1L | 0.3927313 | 0.76975331 | 1 | 0.392731314 | 0.392731314 | 0.392731314 | 0.42025697 | 0.784401616 | 0.75335 | 3.948696856 | 15.79332643 | 15.79332643 | 0.934502795 | -0.029419395 | FALSE | TRUE | FALSE |
| PABPC4 | 0.3925627 | 0.76978433 | 1 | 0.392562719 | 0.392562719 | 7.735374737 | 0.43854346 | 0.777810991 | 1 | 0.43854346 | 0.43854346 | 8.656767301 | 0.895151233 | -0.048103586 | FALSE | TRUE | FALSE |
| IKZF3 | 0.3925373 | 0.76981534 | 1 | 0.392537261 | 0.392537261 | 7.759672449 | 0.43832032 | 0.777933995 | 1 | 0.438320319 | 0.438320319 | 8.676804672 | 0.895548857 | -0.047910716 | FALSE | TRUE | FALSE |
| TPRKB | 0.3924747 | 0.76984634 | 1 | 0.392474743 | 0.392474743 | 0.392474743 | 0.43767908 | 0.778240874 | 0.0037 | 0.437679085 | 0.437679085 | 0.437679085 | 0.896718068 | -0.047344079 | FALSE | TRUE | FALSE |
| LTB4R | 0.3923889 | 0.76987734 | 1 | 0.392388877 | 0.392388877 | 7.778337684 | 0.4382406 | 0.777995442 | 0.01 | 0.438240603 | 0.438240603 | 0.520823228 | 0.895373166 | -0.047995925 | FALSE | TRUE | FALSE |
| TRIM28 | 0.3923396 | 0.76990832 | 1 | 0.392339641 | 0.392339641 | 7.708972455 | 0.43841418 | 0.777841756 | 1 | 0.438414177 | 0.438414177 | 8.63249455 | 0.894906373 | -0.048222399 | FALSE | TRUE | FALSE |
| MRTO4 | 0.3923136 | 0.7699393 | 1 | 0.392313579 | 0.392313579 | 0.392313579 | 1.73343305 | 0.514508065 | 0.5023 | 1.733433052 | 1.733433052 | 1.733433052 | 0.226321737 | -0.645273732 | FALSE | FALSE | FALSE |
| MCMBP | 0.3922839 | 0.76997027 | 0.01645 | 0.39228393 | 0.39228393 | 7.75343063 | 0.43814335 | 0.778087544 | 1 | 0.438143345 | 0.438143345 | 8.672085322 | 0.895332393 | -0.048015703 | FALSE | TRUE | FALSE |
| RBM18 | 0.3922231 | 0.77000122 | 1 | 0.392223086 | 0.392223086 | 0.392223086 | 0.43749303 | 0.778302144 | 1 | 0.437493031 | 0.437493031 | 0.437493031 | 0.896524193 | -0.047437986 | FALSE | TRUE | FALSE |
| TNFAIP3 | 0.3921662 | 0.77003217 | 1 | 0.392166184 | 0.392166184 | 7.702123785 | 0.43823011 | 0.778026152 | 0.02155 | 0.438230112 | 0.438230112 | 8.625473082 | 0.894886439 | -0.048232073 | FALSE | TRUE | FALSE |
| EXOSC7 | 0.3921483 | 0.77006311 | 0.5042 | 0.392148272 | 0.392148272 | 0.392148272 | 0.12611553 | 0.869868862 | 1 | 1.155778555 | 1.155778555 | 1.155778555 | 3.109436934 | 0.492681753 | FALSE | TRUE | FALSE |
| UBIAD1 | 0.392106 | 0.77009405 | 0.8734 | 1.568121267 | 6.272842589 | 6.272842589 | 0.13337168 | 0.868158759 | 1 | 0.133371677 | 0.133371677 | 0.133371677 | 2.939949166 | 0.468339821 | FALSE | TRUE | FALSE |
| KHDC3L | 0.3920418 | 0.77012497 | 1 | 0.392041836 | 0.392041836 | 0.392041836 | 0.43733599 | 0.778332766 | 1 | 0.437335986 | 0.437335986 | 0.437335986 | 0.896431687 | -0.047482801 | FALSE | TRUE | FALSE |
| HAPLN4 | 0.3919366 | 0.77015588 | 1 | 0.391936582 | 0.391936582 | 7.747977959 | 0.43762614 | 0.778271513 | 1 | 0.437626137 | 0.437626137 | 8.663239996 | 0.895596832 | -0.047887451 | FALSE | TRUE | FALSE |

| | | | | | | | | | | | | | | | | | |
|---|---|---|---|---|---|---|---|---|---|---|---|---|---|---|---|---|---|
| DOCK1 | 0.3919082 | 0.77018679 | 0.51795 | 0.391908216 | 3.55984219 | 13.11160645 | 0.01023064 | 0.896701635 | 1 | 0.010230639 | 0.010230639 | 0.342933759 | 38.30730609 | 1.583281612 | FALSE | TRUE | FALSE |
| NYAP2 | 0.3917813 | 0.77021769 | 1 | 0.39178132 | 0.39178132 | 7.706402513 | 0.43774734 | 0.778210226 | 1 | 0.437747339 | 0.437747339 | 8.627656494 | 0.894994177 | -0.04817979 | FALSE | TRUE | FALSE |
| ZNF787 | 0.391459 | 0.77024858 | 1 | 0.391458951 | 0.391458951 | 7.757711033 | 0.43644265 | 0.778638511 | 1 | 0.436442649 | 0.436442649 | 0.436442649 | 0.896931022 | -0.047240955 | FALSE | TRUE | FALSE |
| DDX39A | 0.3913741 | 0.77027946 | 0.01345 | 0.391374058 | 0.391374058 | 7.734680458 | 0.43732019 | 0.778363379 | 1 | 0.437320191 | 0.437320191 | 8.655036871 | 0.894937087 | -0.048207494 | FALSE | TRUE | FALSE |
| ZNF385B | 0.3913729 | 0.77031033 | 0.0123 | 0.391372934 | 0.391372934 | 7.755977169 | 0.43707537 | 0.77842458 | 0.01145 | 0.437075369 | 0.437075369 | 8.671192463 | 0.895435805 | -0.047965544 | FALSE | TRUE | FALSE |
| ASPHD2 | 0.3909977 | 0.7703412 | 1 | 3.658315955 | 35.12982554 | 35.12982554 | 0.43671131 | 0.778485747 | 1 | 4.093179192 | 39.37434556 | 39.37434556 | 0.895322923 | -0.048020296 | FALSE | TRUE | FALSE |
| IFNLR1 | 0.3909508 | 0.77037206 | 1 | 0.39095085 | 0.39095085 | 7.74317829 | 0.43657356 | 0.778607975 | 1 | 0.436573556 | 0.436573556 | 8.656870258 | 0.895498236 | -0.047935265 | FALSE | TRUE | FALSE |
| ZNF460 | 0.390873 | 0.77040291 | 0.0169 | 0.390872961 | 0.390872961 | 7.723861662 | 0.43669641 | 0.778546879 | 1 | 0.436696413 | 0.436696413 | 8.641788925 | 0.895067946 | -0.048143995 | FALSE | TRUE | FALSE |
| HBM | 0.3905067 | 0.77043375 | 1 | 0.390506713 | 0.390506713 | 0.390506713 | 0.43644139 | 0.778669036 | 1 | 0.436441393 | 0.436441393 | 0.436441393 | 0.894751779 | -0.048297429 | FALSE | TRUE | FALSE |
| PARD6B | 0.3904887 | 0.77046459 | 1 | 0.390488704 | 0.390488704 | 0.390488704 | 0.40656227 | 0.789780191 | 1 | 0.406562271 | 0.406562271 | 0.406562271 | 0.960464688 | -0.017518598 | FALSE | TRUE | FALSE |
| EGR2 | 0.3904812 | 0.77049541 | 0.01425 | 0.39048124 | 0.39048124 | 7.714573962 | 0.3570906 | 0.806712787 | 0.9703 | 320.4413263 | 3250.084299 | 33760.86528 | 1.093507478 | 0.038821757 | FALSE | TRUE | FALSE |
| CDKL2 | 0.3904591 | 0.77052623 | 0.7548 | 1.561535091 | 14.25775131 | 14.25775131 | 0.06752771 | 0.882993294 | 1 | 0.067527712 | 0.067527712 | 1.34099123 | 5.782204732 | 0.762093465 | FALSE | TRUE | FALSE |
| APOA1 | 0.390423 | 0.77055704 | 0.00985 | 0.390423022 | 0.390423022 | 0.390423022 | 1.72529923 | 0.515405514 | 0.50635 | 1.725299226 | 1.725299226 | 1.725299226 | 0.226292933 | -0.645329008 | FALSE | FALSE | FALSE |
| ARIH1 | 0.3903488 | 0.77058784 | 0.01625 | 0.390348838 | 0.390348838 | 7.724407999 | 0.43634793 | 0.778730061 | 1 | 0.43634793 | 0.43634793 | 8.64564572 | 0.89458162 | -0.048380029 | FALSE | TRUE | FALSE |
| GUCD1 | 0.3902977 | 0.77061863 | 1 | 0.390297738 | 0.390297738 | 0.390297738 | 0.43578069 | 0.778912928 | 1 | 0.435780687 | 0.435780687 | 0.435780687 | 0.895628808 | -0.047871946 | FALSE | TRUE | FALSE |
| TSPAN13 | 0.3902138 | 0.77064941 | 0.0059 | 0.390213827 | 0.390213827 | 0.390213827 | 0.4360441 | 0.778821534 | 1 | 0.436044096 | 0.436044096 | 0.436044096 | 0.894895333 | -0.048227757 | FALSE | TRUE | FALSE |
| CYB5A | 0.3901569 | 0.77068018 | 1 | 0.390156857 | 0.390156857 | 0.390156857 | 8.70725094 | 0.21483714 | 0.0004 | 0.436089102 | 0.436089102 | 0.436089102 | 0.044808271 | -1.348641817 | FALSE | FALSE | FALSE |
| DAB1 | 0.389999 | 0.77071095 | 1 | 0.389999001 | 0.389999001 | 7.71020494 | 0.4308791 | 0.780754354 | 1 | 1.723480124 | 1.723480124 | 8.530192342 | 0.905123969 | -0.043291934 | FALSE | TRUE | FALSE |
| CD7 | 0.3899616 | 0.7707417 | 1 | 3.634894725 | 34.77372532 | 34.77372532 | 0.46354499 | 0.769604183 | 1 | 0.463544992 | 0.463544992 | 0.463544992 | 0.841259372 | -0.075070084 | FALSE | TRUE | FALSE |
| SNTG2 | 0.3897965 | 0.77077245 | 0.49975 | 0.043484351 | 0.389796486 | 0.861332401 | 2.23097736 | 0.469218462 | 0.2589 | 0.55788323 | 2.23097736 | 2.23097736 | 0.174720054 | -0.757657244 | FALSE | FALSE | FALSE |
| KRT37 | 0.3897916 | 0.77080319 | 0.35115 | 0.38979158 | 6.23494095 | 24.94565916 | 0.04690683 | 0.887348459 | 0.5088 | 0.046906829 | 0.046906829 | 0.232570593 | 8.309911108 | 0.919596378 | FALSE | TRUE | FALSE |
| XPO6 | 0.3897087 | 0.77083393 | 1 | 3.68494249 | 3.68494249 | 71.96162243 | 0.44992484 | 0.774404634 | 1 | 0.449924839 | 0.449924839 | 8.811338542 | 0.866164028 | -0.062399856 | FALSE | TRUE | FALSE |
| RAB22A | 0.3896941 | 0.77086465 | 0.50145 | 1.558549556 | 1.558549556 | 1.558549556 | 0.4404876 | 0.777286671 | 1 | 0.440487597 | 0.440487597 | 0.440487597 | 0.88468791 | -0.053209908 | FALSE | TRUE | FALSE |
| NDRG1 | 0.3896257 | 0.77089536 | 1 | 0.389625727 | 0.389625727 | 7.722547895 | 0.43528324 | 0.779125877 | 1 | 0.435283238 | 0.435283238 | 8.636816099 | 0.895108503 | -0.048124317 | FALSE | TRUE | FALSE |
| MORN5 | 0.3896023 | 0.77092607 | 1 | 0.389602257 | 0.389602257 | 0.389602257 | 0.43513443 | 0.779308054 | 1 | 0.435134426 | 0.435134426 | 0.435134426 | 0.895360685 | -0.048001979 | FALSE | TRUE | FALSE |
| DESI1 | 0.3895921 | 0.77095676 | 0.0054 | 0.389592117 | 0.389592117 | 0.389592117 | 0.43530342 | 0.779095482 | 0.00455 | 0.435303416 | 0.435303416 | 0.435303416 | 0.894989799 | -0.048181915 | FALSE | TRUE | FALSE |
| IKZF1 | 0.3893978 | 0.77098745 | 1 | 35.53003958 | 352.6449785 | 352.6449785 | 0.43545449 | 0.779004244 | 1 | 39.80113106 | 395.377165 | 395.377165 | 0.894233158 | -0.048549231 | FALSE | TRUE | FALSE |
| TM9SF3 | 0.389123 | 0.77101813 | 1 | 0.389122974 | 0.389122974 | 7.70565665 | 0.43520116 | 0.779217006 | 1 | 0.435201161 | 0.435201161 | 8.628356128 | 0.894122097 | -0.048603172 | FALSE | TRUE | FALSE |
| ASCL1 | 0.3891067 | 0.77104881 | 1 | 0.389106716 | 0.389106716 | 0.389106716 | 0.4351975 | 0.779247365 | 1 | 0.435197496 | 0.435197496 | 0.435197496 | 0.894092266 | -0.048617662 | FALSE | TRUE | FALSE |
| TMEM196 | 0.3890944 | 0.77107947 | 1 | 0.38909442 | 3.604361539 | 3.604361539 | 0.4478916 | 0.775037579 | 1 | 0.447891604 | 0.447891604 | 0.447891604 | 0.868724523 | -0.061117919 | FALSE | TRUE | FALSE |
| NCK1 | 0.3890429 | 0.77111013 | 1 | 0.389042886 | 0.389042886 | 0.389042886 | 0.43485468 | 0.779399024 | 0.0083 | 0.434854676 | 0.434854676 | 0.434854676 | 0.894650346 | -0.048346666 | FALSE | TRUE | FALSE |
| PAN3 | 0.389028 | 0.77114077 | 0.0225 | 0.389028019 | 0.389028019 | 7.673307148 | 0.43492874 | 0.779338386 | 1 | 0.434928739 | 0.434928739 | 8.592898171 | 0.894463816 | -0.048437223 | FALSE | TRUE | FALSE |
| SCGB2B2 | 0.3890206 | 0.77117141 | 0.0023 | 0.389020634 | 0.389020634 | 0.389020634 | 0.43408759 | 0.779671476 | 1 | 0.434087586 | 0.434087586 | 0.434087586 | 0.89618005 | -0.047604728 | FALSE | TRUE | FALSE |
| TNFRSF17 | 0.3889563 | 0.77120204 | 1 | 0.38895626 | 0.38895626 | 0.38895626 | 0.43417905 | 0.779580738 | 1 | 0.434179048 | 0.434179048 | 0.434179048 | 0.895842999 | -0.047768096 | FALSE | TRUE | FALSE |
| DKKL1 | 0.3888529 | 0.77123266 | 1 | 3.612861276 | 3.612861276 | 3.612861276 | 37.4143241 | 0.073467834 | 0.24935 | 3.917147448 | 37.41432408 | 37.41432408 | 0.010393156 | -1.983252535 | FALSE | FALSE | FALSE |
| WDR74 | 0.3888306 | 0.77126327 | 0.5033 | 0.388830636 | 0.388830636 | 0.856612918 | 0.48619594 | 0.76063322 | 0.75545 | 0.486195941 | 4.511938202 | 4.511938202 | 0.799740606 | -0.097050852 | FALSE | TRUE | FALSE |
| XRCC5 | 0.3885911 | 0.77129388 | 1 | 0.388591111 | 0.388591111 | 7.671740468 | 0.42703366 | 0.782151623 | 1 | 0.427033661 | 0.427033661 | 8.443754447 | 0.909977705 | -0.040969248 | FALSE | TRUE | FALSE |
| CSRNP1 | 0.3884137 | 0.77132448 | 1 | 0.388413747 | 0.388413747 | 7.667275411 | 0.43413538 | 0.779641239 | 1 | 0.434135381 | 0.434135381 | 8.583226612 | 0.894683464 | -0.048330589 | FALSE | TRUE | FALSE |
| FBXL16 | 0.3883743 | 0.77135506 | 0.5133 | 3.67541658 | 3.67541658 | 7.66563584 | 0.42956477 | 0.781351185 | 1 | 0.429564767 | 0.429564767 | 8.492020423 | 0.90411126 | -0.043778122 | FALSE | TRUE | FALSE |
| LDB2 | 0.3883331 | 0.77138564 | 1 | 3.64336279 | 3.64336279 | 3.64336279 | 0.44303059 | 0.776386345 | 1 | 1.771957103 | 1.771957103 | 1.771957103 | 0.876537911 | -0.057229295 | FALSE | TRUE | FALSE |
| ZRANB1 | 0.3882946 | 0.77141621 | 1 | 3.640535331 | 3.640535331 | 71.79249631 | 0.44800967 | 0.775006017 | 1 | 0.448009674 | 0.448009674 | 8.849759866 | 0.866710297 | -0.062126044 | FALSE | TRUE | FALSE |
| PPP1R16B | 0.3881903 | 0.77144678 | 1 | 0.388190283 | 0.388190283 | 7.658639979 | 0.43440689 | 0.77945963 | 1 | 0.434406888 | 0.434406888 | 8.584421981 | 0.893609872 | -0.048852042 | FALSE | TRUE | FALSE |
| POLB | 0.3881689 | 0.77147733 | 1 | 0.388168874 | 0.388168874 | 0.388168874 | 0.43397319 | 0.779731924 | 1 | 0.433973189 | 0.433973189 | 0.433973189 | 0.894453583 | -0.048442192 | FALSE | TRUE | FALSE |
| LRFN3 | 0.3880582 | 0.77150788 | 1 | 0.388058249 | 0.388058249 | 7.599377061 | 3.96071605 | 0.35784354 | 0.5162 | 3.96071605 | 3.96071605 | 77.7577782 | 0.097976791 | -1.008876789 | FALSE | FALSE | FALSE |
| DNAJB11 | 0.3880539 | 0.77153842 | 1 | 1.551986152 | 1.551986152 | 7.704060791 | 0.43901611 | 0.777657037 | 1 | 0.439016114 | 0.439016114 | 0.439016114 | 0.883917307 | -0.053588362 | FALSE | TRUE | FALSE |
| ATG10 | 0.3880425 | 0.77156894 | 1 | 0.388042534 | 0.388042534 | 0.388042534 | 0.43365075 | 0.779822534 | 1 | 0.433650753 | 0.433650753 | 0.433650753 | 0.894827305 | -0.048260772 | FALSE | TRUE | FALSE |
| NPLOC4 | 0.3880169 | 0.77159946 | 1 | 0.388016906 | 0.388016906 | 7.66162262 | 0.43415147 | 0.779610993 | 1 | 0.43415147 | 0.43415147 | 8.585698334 | 0.893736246 | -0.048790628 | FALSE | TRUE | FALSE |
| TPST1 | 0.3879826 | 0.77162997 | 0.01005 | 0.38798264 | 0.38798264 | 7.703848422 | 0.43368034 | 0.779762136 | 1 | 0.433680343 | 0.433680343 | 0.433680343 | 0.894628143 | -0.048357444 | FALSE | TRUE | FALSE |
| ENSA | 0.3879574 | 0.77166048 | 1 | 0.387957419 | 0.387957419 | 0.387957419 | 0.43315674 | 0.780063776 | 1 | 0.433156736 | 0.433156736 | 0.433156736 | 0.895651359 | -0.047861011 | FALSE | TRUE | FALSE |
| TADA3 | 0.3879374 | 0.77169097 | 1 | 0.387937359 | 0.387937359 | 7.690060418 | 0.43341011 | 0.779913065 | 1 | 0.433410113 | 0.433410113 | 0.433410113 | 0.895081464 | -0.048137436 | FALSE | TRUE | FALSE |
| TSSK6 | 0.3875057 | 0.77172146 | 1 | 0.387505693 | 0.387505693 | 7.664629974 | 0.43322598 | 0.780003518 | 1 | 0.433225978 | 0.433225978 | 8.580320375 | 0.894465504 | -0.048436404 | FALSE | TRUE | FALSE |
| LAG3 | 0.3874992 | 0.77175194 | 1 | 0.387499182 | 0.387499182 | 7.680744265 | 0.43321154 | 0.780033652 | 1 | 0.433211544 | 0.433211544 | 8.596050386 | 0.894480278 | -0.048429231 | FALSE | TRUE | FALSE |
| PDCD6 | 0.3873194 | 0.77178241 | 1 | 0.387319352 | 0.387319352 | 0.387319352 | 0.43328624 | 0.779943225 | 1 | 0.433286242 | 0.433286242 | 0.433286242 | 0.893911032 | -0.048705703 | FALSE | TRUE | FALSE |
| BAALC | 0.3868637 | 0.77181287 | 1 | 0.38686374 | 0.38686374 | 0.38686374 | 0.41271215 | 0.787421388 | 1 | 0.412712152 | 0.412712152 | 0.412712152 | 0.937369395 | -0.02808923 | FALSE | TRUE | FALSE |
| PALD1 | 0.3867709 | 0.77184333 | 0.7532 | 0.386770882 | 3.52479935 | 13.0652295 | 0.06629239 | 0.883272184 | 0.0253 | 0.066292392 | 0.066292392 | 1.30080904 | 5.83431782 | 0.765990083 | FALSE | TRUE | FALSE |
| SERHL2 | 0.386768 | 0.77187378 | 0.98615 | 32.3472206 | 2987.140348 | 29748.36433 | 0.07065355 | 0.882290738 | 0.7525 | 0.629132902 | 5.749834912 | 5.749834912 | 5.474148772 | 0.738316596 | FALSE | TRUE | FALSE |
| DDIT4L | 0.386718 | 0.77190422 | 1 | 0.386717981 | 0.386717981 | 0.386717981 | 0.4321434 | 0.780274423 | 1 | 0.432143395 | 0.432143395 | 0.432143395 | 0.894883471 | -0.048233513 | FALSE | TRUE | FALSE |
| KRTAP9-3 | 0.386706 | 0.77193465 | 1 | 0.386705984 | 0.386705984 | 0.386705984 | 0.41867795 | 0.78509315 | 1 | 3.886964674 | 3.886964674 | 3.886964674 | 0.923635904 | -0.034499193 | FALSE | TRUE | FALSE |
| HGFAC | 0.3866141 | 0.77196507 | 0.75005 | 1.546302777 | 54.38298365 | 217.6189506 | 0.05545855 | 0.885519243 | 0.3245 | 0.055458553 | 0.886751747 | 1.090786688 | 6.971226643 | 0.843309202 | FALSE | TRUE | FALSE |
| CAP1 | 0.3865482 | 0.77199549 | 1 | 0.386548152 | 0.386548152 | 7.660620061 | 0.43239788 | 0.780184193 | 1 | 0.432397875 | 0.432397875 | 8.578648922 | 0.89396404 | -0.04867995 | FALSE | TRUE | FALSE |
| PPM1F | 0.3865377 | 0.77202589 | 1 | 0.386537744 | 0.386537744 | 7.655571278 | 3.93460533 | 0.359343079 | 0.5055 | 3.934605328 | 3.934605328 | 3.934605328 | 0.098240538 | -1.007709268 | FALSE | FALSE | FALSE |
| CER1 | 0.3864514 | 0.77205629 | 1 | 0.386451403 | 0.386451403 | 0.386451403 | 0.4318457 | 0.780394611 | 1 | 0.431845703 | 0.431845703 | 0.431845703 | 0.894883057 | -0.048233715 | FALSE | TRUE | FALSE |
| BAG4 | 0.3863695 | 0.77208668 | 0.50455 | 0.38636947 | 0.38636947 | 1.91621579 | 0.12877402 | 0.869260627 | 0.01245 | 0.128774018 | 0.128774018 | 2.556075831 | 3.000368219 | 0.477174557 | FALSE | TRUE | FALSE |
| UGT8 | 0.3863426 | 0.77211706 | 1 | 1.545217968 | 1.545217968 | 7.652522293 | 0.4371932 | 0.778393983 | 1 | 0.437193201 | 0.437193201 | 8.669771257 | 0.883688471 | -0.053700811 | FALSE | TRUE | FALSE |
| SLC6A2 | 0.3863247 | 0.77214743 | 0.93645 | 3.64098573 | 348.7468014 | 348.7468014 | 0.06906513 | 0.882642967 | 1 | 0.069065132 | 0.069065132 | 1.361978362 | 5.59362908 | 0.747693664 | FALSE | TRUE | FALSE |
| OLR1 | 0.3861081 | 0.7721778 | 1 | 0.386108059 | 0.386108059 | 0.386108059 | 0.43165643 | 0.78048466 | 1 | 0.431656426 | 0.431656426 | 0.431656426 | 0.894480045 | -0.048429343 | FALSE | TRUE | FALSE |
| LRRC8D | 0.3860524 | 0.77220815 | 1 | 0.386052396 | 0.386052396 | 7.602066013 | 0.43204624 | 0.780304482 | 1 | 0.432046235 | 0.432046235 | 8.523548229 | 0.893544174 | -0.048883973 | FALSE | TRUE | FALSE |
| FAM216A | 0.386048 | 0.7722385 | 1 | 0.386047978 | 0.386047978 | 0.386047978 | 0.43137063 | 0.780574632 | 1 | 0.431370629 | 0.431370629 | 0.431370629 | 0.894933387 | -0.04820929 | FALSE | TRUE | FALSE |
| TAS2R7 | 0.3860381 | 0.77226884 | 1 | 0.386038051 | 0.386038051 | 0.386038051 | 0.41827334 | 0.785293965 | 1 | 3.896713854 | 3.896713854 | 3.896713854 | 0.922932472 | -0.034830074 | FALSE | TRUE | FALSE |
| NELFA | 0.3859745 | 0.77229917 | 1 | 0.385974495 | 0.385974495 | 7.602783311 | 0.4318492 | 0.780364577 | 1 | 0.431849197 | 0.431849197 | 8.52188541 | 0.893771477 | -0.048773509 | FALSE | TRUE | FALSE |
| DCTN6 | 0.3858984 | 0.77232949 | 0.00525 | 0.38589837 | 0.38589837 | 0.38589837 | 0.43174536 | 0.780454652 | 1 | 0.431745356 | 0.431745356 | 0.431745356 | 0.893810123 | -0.048754731 | FALSE | TRUE | FALSE |
| PAEP | 0.3857802 | 0.7723598 | 1 | 0.385780206 | 3.573812676 | 3.573812676 | 0.43096292 | 0.780664533 | 1 | 4.003883275 | 4.003883275 | 4.003883275 | 0.895158703 | -0.048099962 | FALSE | TRUE | FALSE |
| ANKRD23 | 0.3857222 | 0.77239011 | 0.74895 | 0.385722245 | 3.475563432 | 3.475563432 | 0.5553876 | 0.730916622 | 0.7498 | 0.555387605 | 5.048712961 | 5.048712961 | 0.69451 | -0.158321497 | FALSE | TRUE | FALSE |

| | | | | | | | | | | | | | | | | | |
|---|---|---|---|---|---|---|---|---|---|---|---|---|---|---|---|---|---|
| IMPDH1 | 0.3857027 | 0.7724204 | 0.6649 | 0.385702732 | 6.168682738 | 24.67885397 | 0.01101633 | 0.89645722 | 1 | 0.703556434 | 11.25510198 | 45.03367357 | 35.01189739 | 1.544215647 | FALSE | TRUE | FALSE |
| TMED4 | 0.3856033 | 0.77245069 | 0.50345 | 3.582876484 | 3.582876484 | 3.582876484 | 3294.13515 | 0.000634664 | 0.1905 | 328.916148 | 3294.135146 | 33796.51911 | 0.000117058 | -3.931600654 | FALSE | FALSE | FALSE |
| USP3 | 0.3853586 | 0.77248097 | 1 | 0.385358562 | 0.385358562 | 7.627697326 | 0.43109833 | 0.780604607 | 1 | 0.431098331 | 0.431098331 | 8.543644269 | 0.893899454 | -0.048711328 | FALSE | TRUE | FALSE |
| AP2A1 | 0.3852839 | 0.77251125 | 1 | 0.385283926 | 0.385283926 | 7.531996968 | 0.43028914 | 0.781083034 | 1 | 0.430289145 | 0.430289145 | 8.434654462 | 0.895407031 | -0.0479795 | FALSE | TRUE | FALSE |
| MS4A5 | 0.3851202 | 0.77254151 | 1 | 0.385120238 | 0.385120238 | 0.385120238 | 0.43030314 | 0.781053197 | 0.00435 | 0.430303145 | 0.430303145 | 0.430303145 | 0.894997499 | -0.048178178 | FALSE | TRUE | FALSE |
| ZFP37 | 0.3850874 | 0.77257177 | 0.50245 | 0.385087442 | 0.385087442 | 1.46481472 | 0.55705254 | 0.729948127 | 0.50385 | 0.55705254 | 0.55705254 | 1.231502432 | 0.691294652 | -0.160336803 | FALSE | TRUE | FALSE |
| TEDDM1 | 0.3850082 | 0.77260202 | 1 | 0.385008213 | 0.385008213 | 0.385008213 | 0.43028727 | 0.781112861 | 1 | 0.430287272 | 0.430287272 | 0.430287272 | 0.894770165 | -0.048288505 | FALSE | TRUE | FALSE |
| CA1 | 0.3849508 | 0.77263226 | 0.49495 | 0.096278274 | 0.384950847 | 0.384950847 | 0.12535936 | 0.870059834 | 1 | 0.501249202 | 2.00474524 | 2.00474524 | 3.070778706 | 0.48724852 | FALSE | TRUE | FALSE |
| PRADC1 | 0.384827 | 0.77266249 | 1 | 0.384827036 | 0.384827036 | 0.384827036 | 3.88156221 | 0.362322034 | 0.50325 | 3.881562207 | 3.881562207 | 3.881562207 | 0.099142308 | -1.003740975 | FALSE | FALSE | FALSE |
| PTAR1 | 0.3847525 | 0.77269272 | 1 | 0.384752529 | 0.384752529 | 7.643607418 | 0.43024947 | 0.781172489 | 0.00695 | 0.430249472 | 0.430249472 | 0.430249472 | 0.894254508 | -0.048538862 | FALSE | TRUE | FALSE |
| VPS39 | 0.3847371 | 0.77272293 | 0.508 | 0.384737106 | 0.384737106 | 7.57268073 | 0.53919967 | 0.738882413 | 0.75585 | 0.539199666 | 4.937510014 | 10.6331925 | 0.713533651 | -0.14658554 | FALSE | TRUE | FALSE |
| BIRC2 | 0.3845889 | 0.77275314 | 1 | 0.384588892 | 0.384588892 | 7.612804827 | 0.43064808 | 0.780873995 | 1 | 0.430648081 | 0.430648081 | 8.535059031 | 0.893046803 | -0.04912578 | FALSE | TRUE | FALSE |
| MIER2 | 0.3845145 | 0.77278334 | 1 | 0.384514475 | 0.384514475 | 7.576561782 | 0.43036505 | 0.780993498 | 1 | 0.430365048 | 0.430365048 | 8.49510575 | 0.893461207 | -0.048924299 | FALSE | TRUE | FALSE |
| SCG3 | 0.3844875 | 0.77281353 | 1 | 3.578355757 | 3.578355757 | 7.632186023 | 0.44364017 | 0.776199138 | 1 | 0.443640167 | 0.443640167 | 0.443640167 | 0.866665137 | -0.062148673 | FALSE | TRUE | FALSE |
| TXNDC17 | 0.3844121 | 0.77284371 | 1 | 0.384412103 | 0.384412103 | 0.384412103 | 0.42990885 | 0.781202293 | 1 | 0.429908849 | 0.429908849 | 0.429908849 | 0.894171182 | -0.048579331 | FALSE | TRUE | FALSE |
| CABP7 | 0.3843934 | 0.77287389 | 1 | 1.537416553 | 1.537416553 | 1.537416553 | 3.95271063 | 0.358093911 | 0.5028 | 3.952710628 | 3.952710628 | 3.952710628 | 0.09724804 | -1.012119145 | FALSE | FALSE | FALSE |
| TMEM81 | 0.3843634 | 0.77290405 | 1 | 0.384363379 | 0.384363379 | 0.384363379 | 0.42962437 | 0.781291654 | 1 | 0.429624368 | 0.429624368 | 0.429624368 | 0.894649856 | -0.048346904 | FALSE | TRUE | FALSE |
| CFDP1 | 0.3843187 | 0.77293421 | 1 | 1.537220762 | 1.537220762 | 1.537220762 | 0.43474715 | 0.77942933 | 1 | 0.434747155 | 0.434747155 | 0.434747155 | 0.884005133 | -0.053545213 | FALSE | TRUE | FALSE |
| CPE | 0.3839547 | 0.77296436 | 0.0145 | 0.383954671 | 0.383954671 | 7.599535278 | 0.42965263 | 0.781261876 | 1 | 0.429652633 | 0.429652633 | 8.51462406 | 0.89363975 | -0.048837522 | FALSE | TRUE | FALSE |
| TCF23 | 0.383891 | 0.7729945 | 0.87565 | 1.535219565 | 6.141035384 | 6.141035384 | 1.18084367 | 0.569130104 | 0.00635 | 0.126865987 | 1.180843671 | 1.180843671 | 0.325098916 | -0.487984479 | FALSE | FALSE | FALSE |
| CLC | 0.3838182 | 0.77302464 | 1 | 0.383818151 | 0.383818151 | 0.383818151 | 0.42902763 | 0.781529575 | 1 | 0.429027625 | 0.429027625 | 0.429027625 | 0.894623395 | -0.048359749 | FALSE | TRUE | FALSE |
| ANGPTL2 | 0.3837701 | 0.77305476 | 0.8764 | 1.534761707 | 6.139306185 | 7.576510088 | 0.13093379 | 0.868722684 | 1 | 0.130933793 | 0.130933793 | 2.588943678 | 2.931024188 | 0.467019402 | FALSE | TRUE | FALSE |
| ZCCHC9 | 0.3837185 | 0.77308488 | 1 | 0.383718528 | 0.383718528 | 0.383718528 | 0.42925226 | 0.781440418 | 1 | 0.429252255 | 0.429252255 | 0.429252255 | 0.893923149 | -0.048699816 | FALSE | TRUE | FALSE |
| ARL3 | 0.3836911 | 0.77311499 | 1 | 0.383691099 | 0.383691099 | 0.383691099 | 1.69902812 | 0.516895017 | 0.5046 | 1.699028122 | 1.699028122 | 1.699028122 | 0.225829752 | -0.646218843 | FALSE | FALSE | FALSE |
| DDN | 0.3834805 | 0.77314509 | 0.02425 | 0.383480487 | 0.383480487 | 7.522387965 | 0.42951601 | 0.781380937 | 0.02045 | 0.429516012 | 0.429516012 | 8.444900878 | 0.892820004 | -0.049236088 | FALSE | TRUE | FALSE |
| CPEB4 | 0.383363 | 0.77317519 | 1 | 0.383363046 | 0.383363046 | 7.555075222 | 0.4200471 | 0.784488312 | 1 | 0.420047098 | 0.420047098 | 8.292598999 | 0.912666812 | -0.039687742 | FALSE | TRUE | FALSE |
| SF3A2 | 0.3833599 | 0.77320527 | 1 | 0.383359862 | 0.383359862 | 0.383359862 | 0.42913545 | 0.781499864 | 1 | 0.429135454 | 0.429135454 | 0.429135454 | 0.893330669 | -0.048987756 | FALSE | TRUE | FALSE |
| DPH1 | 0.3832818 | 0.77323535 | 0.5063 | 0.383281777 | 0.383281777 | 0.8523424 | 0.2449598 | 0.840634928 | 0.7539 | 0.548332158 | 2.193005315 | 4.84725117 | 1.564672147 | 0.194423352 | FALSE | TRUE | FALSE |
| FBXW7 | 0.3831239 | 0.77326542 | 1 | 0.383123861 | 0.383123861 | 7.568849284 | 0.42919014 | 0.781470145 | 1 | 0.42919014 | 0.42919014 | 8.49138294 | 0.892666967 | -0.049310536 | FALSE | TRUE | FALSE |
| AIMP2 | 0.3830236 | 0.77329548 | 1 | 0.383023579 | 0.383023579 | 0.383023579 | 0.42874767 | 0.781588974 | 0.0092 | 0.428747672 | 0.428747672 | 0.428747672 | 0.893354305 | -0.048976266 | FALSE | TRUE | FALSE |
| CD244 | 0.3829955 | 0.77332553 | 1 | 3.564345492 | 34.04520593 | 34.04520593 | 0.442094 | 0.776604359 | 1 | 4.12778182 | 4.12778182 | 4.12778182 | 0.866321333 | -0.062320991 | FALSE | TRUE | FALSE |
| ANP32E | 0.3829799 | 0.77335558 | 1 | 0.382979942 | 0.382979942 | 0.382979942 | 0.42864666 | 0.78161866 | 1 | 0.428646662 | 0.428646662 | 0.428646662 | 0.893463022 | -0.048923417 | FALSE | TRUE | FALSE |
| TMEM199 | 0.3828269 | 0.77338561 | 1 | 3.555794272 | 3.555794272 | 3.555794272 | 0.44172388 | 0.776821936 | 1 | 0.441723878 | 0.441723878 | 0.441723878 | 0.866665567 | -0.062148458 | FALSE | TRUE | FALSE |
| NR5A2 | 0.3828185 | 0.77341564 | 0.264 | 0.382818544 | 1.530055827 | 3.247135648 | 0.06146658 | 0.884443999 | 1 | 0.24569374 | 0.24569374 | 1.217594832 | 6.228076397 | 0.794353931 | FALSE | TRUE | FALSE |
| ARHGEF4 | 0.3826684 | 0.77344566 | 0.509 | 0.382668352 | 0.382668352 | 7.527495657 | 0.50737458 | 0.752949843 | 0.69205 | 4.769334081 | 454.4351821 | 454.4351821 | 0.754212696 | -0.122506161 | FALSE | TRUE | FALSE |
| CD200 | 0.3826165 | 0.77347567 | 1 | 0.382616528 | 0.382616528 | 0.382616528 | 0.42811594 | 0.781737328 | 0.00665 | 0.428115939 | 0.428115939 | 0.428115939 | 0.893721754 | -0.048797671 | FALSE | TRUE | FALSE |
| PODNL1 | 0.3826106 | 0.77350568 | 0.74875 | 0.38261059 | 3.463236998 | 7.579878473 | 0.06589721 | 0.883393813 | 1 | 0.065897206 | 0.065897206 | 1.306970405 | 5.806173222 | 0.763889988 | FALSE | TRUE | FALSE |
| STK24 | 0.3826078 | 0.77353567 | 1 | 0.3826078 | 0.3826078 | 7.585610547 | 3.89043747 | 0.361083747 | 0.00925 | 3.890437471 | 3.890437471 | 3.890437471 | 0.098345701 | -1.007244619 | FALSE | FALSE | FALSE |
| IQCF1 | 0.3825221 | 0.77356566 | 1 | 0.382522127 | 0.382522127 | 0.382522127 | 3.86182241 | 0.363801633 | 0.5074 | 3.861822414 | 3.861822414 | 3.861822414 | 0.099052231 | -1.004135737 | FALSE | FALSE | FALSE |
| FRRS1L | 0.3823861 | 0.77359564 | 1 | 0.382386146 | 0.382386146 | 0.382386146 | 0.41509438 | 0.786519066 | 1 | 0.415094384 | 3.90159167 | 3.90159167 | 0.921202887 | -0.035644709 | FALSE | TRUE | FALSE |
| ZNF704 | 0.3823779 | 0.77362561 | 1 | 0.382377901 | 0.382377901 | 7.569381052 | 0.42848384 | 0.78164834 | 1 | 0.428483838 | 0.428483838 | 8.492503733 | 0.892397489 | -0.04944166 | FALSE | TRUE | FALSE |
| DDT | 0.3823644 | 0.77365557 | 1 | 0.382364428 | 0.382364428 | 0.382364428 | 0.39299578 | 0.794443846 | 1 | 0.39299578 | 0.39299578 | 0.39299578 | 0.972947923 | -0.011910405 | FALSE | TRUE | FALSE |
| CSTF2T | 0.3821802 | 0.77368553 | 1 | 3.582577458 | 3.582577458 | 12.93357001 | 0.41492524 | 0.786575694 | 1 | 3.891958756 | 37.46731652 | 76.9681628 | 0.92108203 | -0.035701691 | FALSE | TRUE | FALSE |
| TBKBP1 | 0.3821637 | 0.77371547 | 1 | 3.562778322 | 34.08924689 | 34.08924689 | 0.41471138 | 0.786603997 | 1 | 37.08530766 | 364.3980049 | 364.3980049 | 0.921517362 | -0.035496478 | FALSE | TRUE | FALSE |
| TNFSF15 | 0.3821295 | 0.77374541 | 1 | 0.382129527 | 0.382129527 | 0.382129527 | 0.42801086 | 0.781796612 | 1 | 0.428010862 | 0.428010862 | 0.428010862 | 0.892803339 | -0.049244194 | FALSE | TRUE | FALSE |
| WBP2 | 0.382051 | 0.77377534 | 1 | 0.382050979 | 0.382050979 | 0.382050979 | 0.42807884 | 0.781766974 | 1 | 0.428078844 | 0.428078844 | 0.428078844 | 0.892478065 | -0.049402449 | FALSE | TRUE | FALSE |
| AICDA | 0.3819096 | 0.77380526 | 1 | 0.381909635 | 0.381909635 | 0.381909635 | 0.42766876 | 0.781915082 | 1 | 0.427668762 | 0.427668762 | 0.427668762 | 0.893003344 | -0.049146915 | FALSE | TRUE | FALSE |
| SCIMP | 0.3818717 | 0.77383517 | 1 | 0.381871675 | 0.381871675 | 0.381871675 | 0.42711899 | 0.782122085 | 1 | 0.427118986 | 0.427118986 | 0.427118986 | 0.894063921 | -0.04863143 | FALSE | TRUE | FALSE |
| OR1A2 | 0.3818032 | 0.77386508 | 1 | 0.381803193 | 0.381803193 | 0.381803193 | 0.42732775 | 0.782003848 | 1 | 0.427327746 | 0.427327746 | 0.427327746 | 0.893466891 | -0.048921536 | FALSE | TRUE | FALSE |
| TUSC1 | 0.3816706 | 0.77389498 | 1 | 0.381670574 | 0.381670574 | 0.381670574 | 0.42734088 | 0.781974268 | 1 | 0.427340882 | 0.427340882 | 0.427340882 | 0.893129092 | -0.049085764 | FALSE | TRUE | FALSE |
| FAF1 | 0.3816028 | 0.77392487 | 1 | 0.381602829 | 0.381602829 | 7.536512779 | 1.58831686 | 0.527526586 | 0.87825 | 14.89931802 | 143.4437412 | 143.4437412 | 0.24025611 | -0.619325559 | FALSE | FALSE | FALSE |
| ATP6V0E2 | 0.3812875 | 0.77395475 | 1 | 3.546677953 | 3.546677953 | 3.546677953 | 36.7826449 | 0.075416789 | 0.26085 | 3.847246953 | 36.7826449 | 36.7826449 | 0.010365962 | -1.984390373 | FALSE | FALSE | FALSE |
| IL15RA | 0.3812263 | 0.77398462 | 1 | 0.381226337 | 0.381226337 | 0.381226337 | 0.42654974 | 0.782299192 | 1 | 0.426549738 | 0.426549738 | 0.426549738 | 0.893744161 | -0.048786782 | FALSE | TRUE | FALSE |
| OR5AP2 | 0.3812057 | 0.77401449 | 1 | 0.381205698 | 0.381205698 | 0.381205698 | 3.86179062 | 0.36404744 | 0.00705 | 3.861790617 | 3.861790617 | 3.861790617 | 0.098712161 | -1.00562934 | FALSE | FALSE | FALSE |
| PJA2 | 0.3810908 | 0.77404435 | 1 | 0.381090787 | 0.381090787 | 7.536824095 | 0.4216709 | 0.783989994 | 0.01495 | 0.421670898 | 0.421670898 | 8.350563656 | 0.90376355 | -0.043945178 | FALSE | TRUE | FALSE |
| GPR62 | 0.3810478 | 0.7740742 | 1 | 3.547466684 | 3.547466684 | 3.547466684 | 0.44035094 | 0.777348485 | 1 | 0.440350936 | 0.440350936 | 0.440350936 | 0.8653275 | -0.062819494 | FALSE | TRUE | FALSE |
| SMARCD1 | 0.3809834 | 0.77410404 | 1 | 0.380983419 | 0.380983419 | 7.543932315 | 0.42716753 | 0.782092539 | 1 | 0.427167534 | 0.427167534 | 8.46854661 | 0.891882901 | -0.049692162 | FALSE | TRUE | FALSE |
| RABGAP1 | 0.3809715 | 0.77413387 | 0.0296 | 0.380971455 | 0.380971455 | 7.470892382 | 0.42719389 | 0.782062984 | 1 | 0.427193895 | 0.427193895 | 8.396986567 | 0.891799859 | -0.0497326 | FALSE | TRUE | FALSE |
| NFKBIL1 | 0.3809682 | 0.7741637 | 1 | 0.380968201 | 0.380968201 | 7.555922954 | 0.4134084 | 0.787112109 | 1 | 3.858647085 | 3.858647085 | 8.207535244 | 0.921529903 | -0.035490568 | FALSE | TRUE | FALSE |
| ARMC2 | 0.3809682 | 0.77419352 | 0.5126 | 0.380968168 | 0.380968168 | 7.497635501 | 0.55106376 | 0.732967244 | 0.5102 | 0.551063762 | 0.551063762 | 2.087907841 | 0.691332282 | -0.160313163 | FALSE | TRUE | FALSE |
| GTF2H2 | 0.3809495 | 0.77422332 | 1 | 0.380949459 | 0.380949459 | 0.380949459 | 0.39303407 | 0.794391445 | 1 | 0.393034067 | 0.393034067 | 0.393034067 | 0.969253028 | -0.013562834 | FALSE | TRUE | FALSE |
| FNBP1 | 0.3809302 | 0.77425312 | 0.0172 | 0.380930191 | 0.380930191 | 7.524210516 | 0.40100888 | 0.791870426 | 1 | 3.769019651 | 36.35699916 | 74.55742596 | 0.949929555 | -0.0223086 | FALSE | TRUE | FALSE |
| HCRTR2 | 0.3807991 | 0.77428291 | 1 | 0.380799068 | 0.380799068 | 7.538303801 | 0.42673997 | 0.782269695 | 1 | 0.426739966 | 0.426739966 | 8.458114945 | 0.892344516 | -0.049467441 | FALSE | TRUE | FALSE |
| MEX3B | 0.380783 | 0.7743127 | 1 | 338.1742443 | 3418.129051 | 35392.23886 | 0.45457984 | 0.772902722 | 1 | 41.22362903 | 407.9025573 | 407.9025573 | 0.837659163 | -0.076932657 | FALSE | TRUE | FALSE |
| PATZ1 | 0.3807409 | 0.77434247 | 1 | 0.38074094 | 0.38074094 | 7.476210555 | 1.68803708 | 0.518228972 | 0.50695 | 1.688037077 | 1.688037077 | 14.28719727 | 0.225552474 | -0.646752404 | FALSE | FALSE | FALSE |
| MUC7 | 0.3803528 | 0.77437224 | 1 | 0.380352848 | 0.380352848 | 0.380352848 | 0.42585231 | 0.78262311 | 0.00925 | 0.42585231 | 0.42585231 | 0.42585231 | 0.893156711 | -0.049072334 | FALSE | TRUE | FALSE |
| DUSP28 | 0.3803505 | 0.774402 | 1 | 0.380350524 | 0.380350524 | 0.380350524 | 0.4260146 | 0.782475999 | 1 | 0.426014599 | 0.426014599 | 0.426014599 | 0.89281101 | -0.049240463 | FALSE | TRUE | FALSE |
| PLEKHA2 | 0.3803168 | 0.77443175 | 1 | 0.380316754 | 0.380316754 | 7.544132922 | 0.42632071 | 0.782417096 | 1 | 0.426320707 | 0.426320707 | 0.426320707 | 0.892090737 | -0.04959097 | FALSE | TRUE | FALSE |
| C22orf15 | 0.3802297 | 0.7744615 | 1 | 3.518733386 | 3.518733386 | 3.518733386 | 36.4040183 | 0.076531539 | 0.00165 | 3.824802152 | 36.40401828 | 36.40401828 | 0.010444719 | -1.98110325 | FALSE | FALSE | FALSE |
| FGF17 | 0.3801539 | 0.77449123 | 1 | 0.380153917 | 0.380153917 | 0.380153917 | 1.68575385 | 0.518376785 | 0.5046 | 1.685753854 | 1.685753854 | 1.685753854 | 0.225509742 | -0.646834692 | FALSE | FALSE | FALSE |
| EHD3 | 0.3800839 | 0.77452096 | 0.75145 | 1.519230566 | 14.54285421 | 14.54285421 | 0.25029937 | 0.839371876 | 0.75905 | 0.250299368 | 1.000802013 | 4.948989186 | 1.51851713 | 0.181419695 | FALSE | TRUE | FALSE |
| CLINT1 | 0.3798635 | 0.77455068 | 1 | 0.37986346 | 0.37986346 | 7.512679017 | 0.42577246 | 0.782652507 | 1 | 0.425772458 | 0.425772458 | 8.431915261 | 0.892174807 | -0.049550044 | FALSE | TRUE | FALSE |
| KATNBL1 | 0.3798276 | 0.7745804 | 1 | 0.379827554 | 0.379827554 | 0.379827554 | 0.42529672 | 0.782740652 | 0.00585 | 0.425296721 | 0.425296721 | 0.425296721 | 0.893088367 | -0.049105568 | FALSE | TRUE | FALSE |

| | | | | | | | | | | | | | | | | | |
|---|---|---|---|---|---|---|---|---|---|---|---|---|---|---|---|---|---|
| IFITM2 | 0.379728 | 0.7746101 | 1 | 0.379727972 | 0.379727972 | 0.379727972 | 0.42538945 | 0.782711279 | 1 | 0.425389452 | 0.425389452 | 0.425389452 | 0.892659587 | -0.049314127 | FALSE | TRUE | FALSE |
| HNRNPA2B1 | 0.3796286 | 0.7746398 | 1 | 0.379628638 | 0.379628638 | 0.379628638 | 0.42589773 | 0.782593705 | 1 | 0.425897727 | 0.425897727 | 0.425897727 | 0.891361034 | -0.049946355 | FALSE | TRUE | FALSE |
| PDF | 0.3795034 | 0.77466949 | 1 | 0.379503383 | 1.517887819 | 1.517887819 | 3.89006789 | 0.361331879 | 0.50865 | 3.890067894 | 3.890067894 | 3.890067894 | 0.097557007 | -1.01074153 | FALSE | FALSE | FALSE |
| LAMTOR5 | 0.3794805 | 0.77469917 | 1 | 0.379480515 | 0.379480515 | 0.379480515 | 3.8271054 | 0.36527508 | 0.005 | 3.827105403 | 3.827105403 | 3.827105403 | 0.099156014 | -1.003680942 | FALSE | FALSE | FALSE |
| KCNG1 | 0.3794316 | 0.77472884 | 0.8786 | 1.517314694 | 14.28449618 | 29.78724775 | 4.34811764 | 0.337473725 | 0.0122 | 1.208910558 | 1.208910558 | 23.78543981 | 0.087263412 | -1.059167811 | FALSE | FALSE | FALSE |
| PPDPF | 0.3793743 | 0.77475851 | 1 | 0.379374334 | 0.379374334 | 0.379374334 | 0.42488178 | 0.782799378 | 0.00345 | 0.424881778 | 0.424881778 | 0.424881778 | 0.892893867 | -0.04920016 | FALSE | TRUE | FALSE |
| MRPL43 | 0.3791529 | 0.77478816 | 1 | 0.379152863 | 0.379152863 | 7.524176076 | 0.42472935 | 0.782858072 | 1 | 0.42472935 | 0.42472935 | 0.42472935 | 0.89269287 | -0.049297934 | FALSE | TRUE | FALSE |
| ZSCAN18 | 0.3787578 | 0.77481782 | 1 | 0.378757841 | 0.378757841 | 7.469358056 | 0.42454672 | 0.782916734 | 0.0156 | 0.42454672 | 0.42454672 | 8.386417824 | 0.892146432 | -0.049563857 | FALSE | TRUE | FALSE |
| LY9 | 0.3787361 | 0.77484746 | 0.87605 | 3.417017748 | 31.62164931 | 31.62164931 | 4.53267138 | 0.327807763 | 0.6652 | 42.20235825 | 3938.146736 | 39423.91978 | 0.083556918 | -1.078017588 | FALSE | FALSE | FALSE |
| DOK5 | 0.3786923 | 0.7748771 | 0.75165 | 0.378692305 | 1.514422798 | 1.880083639 | 0.12370165 | 0.870429674 | 1 | 1.978035309 | 7.91272609 | 7.91272609 | 3.061335991 | 0.485910997 | FALSE | TRUE | FALSE |
| ATF3 | 0.378684 | 0.77490672 | 1 | 0.378684016 | 0.378684016 | 0.378684016 | 0.42435188 | 0.783033955 | 1 | 0.424351882 | 0.424351882 | 0.424351882 | 0.892382084 | -0.049449158 | FALSE | TRUE | FALSE |
| COMMD8 | 0.378637 | 0.77493634 | 1 | 0.378636967 | 0.378636967 | 0.378636967 | 0.42398815 | 0.783092519 | 1 | 0.42398815 | 0.42398815 | 0.42398815 | 0.893036674 | -0.049130706 | FALSE | TRUE | FALSE |
| SLC1A2 | 0.3786313 | 0.77496595 | 1 | 0.378631334 | 0.378631334 | 7.485025751 | 0.4247913 | 0.78282873 | 1 | 0.424791297 | 0.424791297 | 8.409232859 | 0.891334962 | -0.049959058 | FALSE | TRUE | FALSE |
| NPY5R | 0.3784964 | 0.77499555 | 1 | 1.513839524 | 1.513839524 | 7.514546484 | 0.42925248 | 0.781410682 | 1 | 0.429252484 | 0.429252484 | 0.429252484 | 0.881757018 | -0.054651075 | FALSE | TRUE | FALSE |
| C1QBP | 0.3784891 | 0.77502515 | 1 | 0.378489119 | 0.378489119 | 0.378489119 | 0.41119331 | 0.78803716 | 1 | 0.41119331 | 3.844182873 | 3.844182873 | 0.920465168 | -0.035992641 | FALSE | TRUE | FALSE |
| PHTF1 | 0.3783565 | 0.77505474 | 0.6872 | 0.378356472 | 29.52074059 | 29.52074059 | 0.01071615 | 0.896569085 | 0.0178 | 0.010716152 | 0.010716152 | 0.211740528 | 35.30712142 | 1.547862311 | FALSE | TRUE | FALSE |
| PSMB6 | 0.3783427 | 0.77508431 | 1 | 0.378342692 | 0.378342692 | 0.378342692 | 0.4240512 | 0.783063241 | 1 | 0.424051204 | 0.424051204 | 0.424051204 | 0.892209923 | -0.049532951 | FALSE | TRUE | FALSE |
| RDH14 | 0.3781772 | 0.77511388 | 1 | 1.512421752 | 6.050050653 | 6.050050653 | 15.1984344 | 0.157248823 | 0.25095 | 3.800218677 | 15.19843444 | 15.19843444 | 0.024882644 | -1.604103475 | FALSE | FALSE | FALSE |
| MYBL2 | 0.3779917 | 0.77514345 | 0.0205 | 0.377991703 | 0.377991703 | 7.445031253 | 0.42397783 | 0.783121789 | 1 | 0.423977827 | 0.423977827 | 8.366050812 | 0.891536487 | -0.049860878 | FALSE | TRUE | FALSE |
| SPAST | 0.3779338 | 0.775173 | 0.5052 | 0.377933845 | 0.377933845 | 1.868737062 | 1.976757 | 0.491175964 | 0.25815 | 0.494315921 | 1.976757002 | 4.244023538 | 0.191188823 | -0.7185375 | FALSE | FALSE | FALSE |
| NECAP1 | 0.3778808 | 0.77520255 | 0.0078 | 0.377880828 | 0.377880828 | 0.377880828 | 0.42393641 | 0.783151049 | 0.00735 | 0.423936408 | 0.423936408 | 0.423936408 | 0.891362054 | -0.049945858 | FALSE | TRUE | FALSE |
| SLC38A7 | 0.3777329 | 0.77523209 | 1 | 0.377732929 | 0.377732929 | 7.477924791 | 0.42342693 | 0.783297234 | 1 | 0.423426928 | 0.423426928 | 8.39276427 | 0.892085278 | -0.049593628 | FALSE | TRUE | FALSE |
| ST6GALNAC! | 0.3776841 | 0.77526162 | 0.01 | 0.377684079 | 0.377684079 | 0.448838076 | 0.4186071 | 0.785150566 | 1 | 1.67440617 | 1.67440617 | 1.67440617 | 0.90224002 | -0.044677913 | FALSE | TRUE | FALSE |
| FDXACB1 | 0.3776133 | 0.77529115 | 0.5029 | 0.377613315 | 6.040864843 | 6.040864843 | 0.30250222 | 0.824634703 | 0.25745 | 0.075636616 | 0.302502218 | 1.496874815 | 1.248299327 | 0.096318736 | FALSE | TRUE | FALSE |
| RASGEF1B | 0.377608 | 0.77532066 | 1 | 0.377607983 | 0.377607983 | 7.481190918 | 0.42357855 | 0.783238785 | 1 | 0.423578548 | 0.423578548 | 8.401447737 | 0.89147098 | -0.04989279 | FALSE | TRUE | FALSE |
| RRM1 | 0.3775314 | 0.77535017 | 1 | 0.377531362 | 0.377531362 | 7.441775799 | 0.42383176 | 0.783180302 | 1 | 0.423831764 | 0.423831764 | 8.368929543 | 0.890757593 | -0.050240467 | FALSE | TRUE | FALSE |
| GPR21 | 0.37753 | 0.77537967 | 1 | 3.522714734 | 3.522714734 | 3.522714734 | 0.43670972 | 0.778516318 | 1 | 0.436709719 | 0.436709719 | 0.436709719 | 0.864487187 | -0.063241439 | FALSE | TRUE | FALSE |
| PPM1K | 0.3772415 | 0.77540916 | 1 | 0.377241526 | 0.377241526 | 0.377241526 | 0.42293415 | 0.783501551 | 1 | 0.422934147 | 0.422934147 | 0.422934147 | 0.8919628 | -0.049653258 | FALSE | TRUE | FALSE |
| S1PR2 | 0.3772068 | 0.77543865 | 1 | 0.377206823 | 0.377206823 | 7.462732607 | 0.42330821 | 0.783326447 | 1 | 0.423308205 | 0.423308205 | 8.385682475 | 0.891092631 | -0.050077148 | FALSE | TRUE | FALSE |
| NXF1 | 0.3771671 | 0.77546812 | 1 | 0.377167105 | 0.377167105 | 7.482533029 | 0.42308698 | 0.783443216 | 1 | 0.42308698 | 0.42308698 | 0.42308698 | 0.891464694 | -0.049895852 | FALSE | TRUE | FALSE |
| HSPA12A | 0.3770404 | 0.77549759 | 1 | 0.377040351 | 0.377040351 | 7.41438991 | 0.42311414 | 0.783414036 | 1 | 0.42311414 | 0.42311414 | 8.337210106 | 0.891107895 | -0.050069708 | FALSE | TRUE | FALSE |
| ANKRD40 | 0.3769983 | 0.77552705 | 0.0099 | 0.376998346 | 0.376998346 | 0.376998346 | 0.40967225 | 0.788510437 | 1 | 3.818870485 | 3.818870485 | 3.818870485 | 0.920243797 | -0.036097101 | FALSE | TRUE | FALSE |
| SH2D3A | 0.3769821 | 0.77555651 | 0.5054 | 0.376982144 | 0.376982144 | 1.429091085 | 4.89382656 | 0.31081203 | 0.0118 | 0.530714655 | 4.893826558 | 10.48883081 | 0.077032183 | -1.113327794 | FALSE | FALSE | FALSE |
| LYPD6B | 0.3769379 | 0.77558595 | 1 | 0.376937929 | 0.376937929 | 0.376937929 | 0.42237691 | 0.783676363 | 1 | 0.42237691 | 0.42237691 | 0.42237691 | 0.892420774 | -0.049430329 | FALSE | TRUE | FALSE |
| ING5 | 0.376895 | 0.77561539 | 1 | 3.515091763 | 33.64666618 | 33.64666618 | 4.07891377 | 0.352042242 | 0.5093 | 4.078913769 | 4.078913769 | 4.078913769 | 0.092400832 | -1.034324119 | FALSE | FALSE | FALSE |
| NPAS3 | 0.3767595 | 0.77564482 | 1 | 0.376759531 | 0.376759531 | 7.370399414 | 0.42302062 | 0.783472387 | 1 | 0.423020615 | 0.423020615 | 8.297187828 | 0.890641065 | -0.050297285 | FALSE | TRUE | FALSE |
| OR2A25 | 0.3767175 | 0.77567424 | 1 | 0.376717453 | 0.376717453 | 0.376717453 | 0.42235791 | 0.78370547 | 1 | 0.422357914 | 0.422357914 | 0.422357914 | 0.8919389 | -0.049664895 | FALSE | TRUE | FALSE |
| TARBP2 | 0.3765361 | 0.77570365 | 1 | 0.376536072 | 0.376536072 | 0.376536072 | 0.42232918 | 0.783734568 | 1 | 0.422329177 | 0.422329177 | 0.422329177 | 0.891570112 | -0.049844499 | FALSE | TRUE | FALSE |
| CSNK1G1 | 0.3765095 | 0.77573305 | 1 | 0.37650952 | 0.37650952 | 7.46814529 | 0.42263648 | 0.783588993 | 1 | 0.422636478 | 0.422636478 | 0.422636478 | 0.890859025 | -0.050191016 | FALSE | TRUE | FALSE |
| FANCL | 0.3764697 | 0.77576245 | 0.87355 | 0.376469732 | 31.51588686 | 31.51588686 | 0.06501982 | 0.883601781 | 1 | 0.578212892 | 0.578212892 | 0.578212892 | 5.790076261 | 0.762684284 | FALSE | TRUE | FALSE |
| HNMT | 0.3763526 | 0.77579184 | 1 | 3.50003823 | 3.50003823 | 3.50003823 | 3.93340616 | 0.359592226 | 0.4998 | 0.421922258 | 3.933406161 | 3.933406161 | 0.0956811 | -1.01917384 | FALSE | FALSE | FALSE |
| PGR | 0.3762523 | 0.77582122 | 0.02715 | 0.376252327 | 0.376252327 | 7.362764428 | 0.12301906 | 0.87059809 | 1 | 0.491914769 | 1.967505133 | 9.651132658 | 3.058488022 | 0.485506784 | FALSE | TRUE | FALSE |
| COQ2 | 0.3761922 | 0.7758506 | 0.7528 | 0.376192174 | 3.521491041 | 3.521491041 | 0.06369992 | 0.883852178 | 0.0061 | 0.063699923 | 0.063699923 | 0.063699923 | 5.905692784 | 0.771270851 | FALSE | TRUE | FALSE |
| C2orf68 | 0.3761607 | 0.77587996 | 1 | 0.376160678 | 0.376160678 | 0.376160678 | 0.42155773 | 0.783996096 | 1 | 0.421557731 | 0.421557731 | 0.421557731 | 0.892311183 | -0.049483664 | FALSE | TRUE | FALSE |
| PDE3A | 0.3761577 | 0.77590932 | 0.0291 | 0.376157696 | 0.376157696 | 7.358119845 | 0.06181951 | 0.884367124 | 1 | 0.061819509 | 0.061819509 | 1.212463266 | 6.084773271 | 0.7842444 | FALSE | TRUE | FALSE |
| LYRM1 | 0.3761407 | 0.77593867 | 1 | 0.376140731 | 0.376140731 | 0.376140731 | 3.70608796 | 0.370630733 | 0.7499 | 3.706087959 | 35.56211185 | 35.56211185 | 0.101492662 | -0.993565358 | FALSE | FALSE | FALSE |
| ZNF551 | 0.3761128 | 0.77596801 | 0.51305 | 0.376112802 | 0.376112802 | 1.430110054 | 0.54559397 | 0.735517917 | 0.50375 | 0.545593965 | 0.545593965 | 1.204902115 | 0.689363934 | -0.161551442 | FALSE | TRUE | FALSE |
| SMNDC1 | 0.3760738 | 0.77599734 | 1 | 0.376073878 | 0.376073878 | 0.376073878 | 0.42223435 | 0.783763658 | 1 | 0.422234354 | 0.422234354 | 0.422234354 | 0.890675531 | -0.050280479 | FALSE | TRUE | FALSE |
| SLC35A1 | 0.3756802 | 0.77602667 | 1 | 0.375680224 | 0.375680224 | 0.375680224 | 1.6666671 | 0.520729529 | 0.0061 | 1.666667097 | 1.666667097 | 1.666667097 | 0.225408076 | -0.647030528 | FALSE | FALSE | FALSE |
| MRPL3 | 0.3755799 | 0.77605599 | 0.74745 | 0.375579945 | 1.502429334 | 1.502429334 | 0.12638771 | 0.869783813 | 0.00875 | 0.12638771 | 0.12638771 | 0.12638771 | 2.97164926 | 0.472997549 | FALSE | TRUE | FALSE |
| KITLG | 0.3755075 | 0.7760853 | 1 | 0.375507485 | 0.375507485 | 0.375507485 | 0.42136905 | 0.784025114 | 1 | 0.421369052 | 0.421369052 | 0.421369052 | 0.891160572 | -0.050044036 | FALSE | TRUE | FALSE |
| ABCA1 | 0.375505 | 0.7761146 | 0.36445 | 0.203630981 | 51.8750092 | 248.5510006 | 0.00141987 | 0.899256047 | 0.92205 | 0.090293192 | 10.5172822 | 91.9391623 | 264.4645546 | 2.422367473 | FALSE | TRUE | FALSE |
| RBPJ | 0.375499 | 0.77614389 | 1 | 33.99358803 | 3405.657732 | 3405.657732 | 0.46332376 | 0.769637333 | 1 | 4.369127721 | 4.369127721 | 9.19180319 | 0.810446254 | -0.091275781 | FALSE | TRUE | FALSE |
| RCAN3 | 0.3754671 | 0.77617318 | 1 | 1.501705501 | 6.007672254 | 6.007672254 | 104.43222 | 0.028305809 | 0.0344 | 26.09127389 | 104.4322201 | 448.9559306 | 0.003595318 | -2.444262639 | FALSE | FALSE | FALSE |
| CDH7 | 0.3754579 | 0.77620246 | 0.75485 | 0.375457893 | 1.501740617 | 7.389331161 | 0.1268531 | 0.869645385 | 1 | 0.126853096 | 0.126853096 | 2.5013726 | 2.959785026 | 0.471260169 | FALSE | TRUE | FALSE |
| GCSH | 0.3753676 | 0.77623173 | 1 | 0.3753676 | 0.3753676 | 0.3753676 | 3.79366699 | 0.366742927 | 0.50785 | 3.793666993 | 3.793666993 | 3.793666993 | 0.098945849 | -1.004602423 | FALSE | FALSE | FALSE |
| ELOVL3 | 0.3751833 | 0.77626099 | 1 | 0.375183271 | 0.375183271 | 0.375183271 | 0.42088586 | 0.784256969 | 1 | 0.420885857 | 0.420885857 | 0.420885857 | 0.891413348 | -0.049920867 | FALSE | TRUE | FALSE |
| FSCN1 | 0.3751817 | 0.77629025 | 1 | 0.375181652 | 0.375181652 | 7.364546638 | 0.40621679 | 0.789889857 | 1 | 0.406216793 | 0.406216793 | 7.991541231 | 0.923599561 | -0.034516282 | FALSE | TRUE | FALSE |
| TLK2 | 0.3749288 | 0.7763195 | 1 | 0.374928781 | 0.374928781 | 7.391514296 | 0.42117949 | 0.784083127 | 1 | 0.421179494 | 0.421179494 | 8.317589352 | 0.890187644 | -0.050518438 | FALSE | TRUE | FALSE |
| DCUN1D3 | 0.3748086 | 0.77634874 | 0.00975 | 0.374808573 | 0.374808573 | 0.374808573 | 0.42106056 | 0.784170085 | 1 | 0.421060564 | 0.421060564 | 0.421060564 | 0.890153591 | -0.050535052 | FALSE | TRUE | FALSE |
| RNASE13 | 0.3747583 | 0.77637797 | 1 | 0.374758264 | 0.374758264 | 0.374758264 | 0.42048782 | 0.784372702 | 1 | 0.420487822 | 0.420487822 | 0.420487822 | 0.891246414 | -0.050002204 | FALSE | TRUE | FALSE |
| GPER1 | 0.3747437 | 0.77640719 | 1 | 0.374743705 | 0.374743705 | 7.405767736 | 0.41613992 | 0.786064917 | 1 | 1.664402511 | 1.664402511 | 8.235612676 | 0.900523315 | -0.045505038 | FALSE | TRUE | FALSE |
| RABL6 | 0.3747306 | 0.77643641 | 0.5115 | 0.374730595 | 0.374730595 | 1.381774395 | 41.1394177 | 0.067107403 | 0.5045 | 406.8475872 | 42753.29556 | 81492.393 | 0.009108797 | -2.040538988 | FALSE | FALSE | FALSE |
| ABCC9 | 0.3746741 | 0.77646562 | 0.2192 | 0.042058398 | 1.822822475 | 13.67991012 | 4.51449303 | 0.328895582 | 0.2276 | 0.501694302 | 41.66800192 | 166.5713596 | 0.082993605 | -1.080955368 | FALSE | FALSE | FALSE |
| SLC35C1 | 0.3746376 | 0.77649482 | 1 | 0.374637639 | 0.374637639 | 7.420386247 | 0.4208547 | 0.784285913 | 1 | 0.420854698 | 0.420854698 | 8.345483878 | 0.890182862 | -0.050520771 | FALSE | TRUE | FALSE |
| STAC | 0.3746309 | 0.77652401 | 0.507 | 0.374630932 | 0.374630932 | 0.374630932 | 0.06094386 | 0.884554878 | 1 | 0.243670541 | 0.243670541 | 1.210026994 | 6.147147894 | 0.788673662 | FALSE | TRUE | FALSE |
| RPL7L1 | 0.3746012 | 0.7765532 | 1 | 0.374601156 | 0.374601156 | 0.374601156 | 0.41562471 | 0.786292245 | 1 | 1.662427299 | 1.662427299 | 1.662427299 | 0.90129664 | -0.045132248 | FALSE | TRUE | FALSE |
| AKT2 | 0.374516 | 0.77658237 | 1 | 0.374516017 | 0.374516017 | 7.391334504 | 3.83697943 | 0.364538953 | 0.5084 | 3.836979435 | 3.836979435 | 13.82027356 | 0.097606991 | -1.010519075 | FALSE | FALSE | FALSE |
| STAM | 0.3743213 | 0.77661154 | 1 | 3.497014794 | 3.497014794 | 7.414046362 | 0.43353565 | 0.779882896 | 1 | 0.433535651 | 0.433535651 | 8.596870147 | 0.863415186 | -0.063780317 | FALSE | TRUE | FALSE |
| PIK3R5 | 0.3742923 | 0.7766407 | 0.75945 | 0.374292255 | 1.496848887 | 7.388618842 | 0.12666992 | 0.86970931 | 1 | 0.126669923 | 0.126669923 | 2.504402617 | 2.954862888 | 0.470537333 | FALSE | TRUE | FALSE |
| EYA4 | 0.374087 | 0.77666986 | 0.5086 | 0.374086988 | 3.454453461 | 13.81079469 | 0.01160065 | 0.896232806 | 0.0137 | 0.011600647 | 0.011600647 | 0.229750811 | 32.24707878 | 1.508490379 | FALSE | TRUE | FALSE |
| RAB5B | 0.3740306 | 0.776699 | 1 | 3.528731067 | 3.528731067 | 3.528731067 | 5.44135959 | 0.289161993 | 0.98 | 4867.611152 | 511646.0798 | 5436960.173 | 0.068738454 | -1.162800238 | FALSE | FALSE | FALSE |
| SPATA18 | 0.3739448 | 0.77672814 | 0.0178 | 0.373944782 | 0.373944782 | 1.411959788 | 0.06213014 | 0.884264476 | 1 | 0.062130141 | 0.062130141 | 1.228535735 | 6.018733872 | 0.779505141 | FALSE | TRUE | FALSE |

| | | | | | | | | | | | | | | | | | |
|---|---|---|---|---|---|---|---|---|---|---|---|---|---|---|---|---|---|
| PDP1 | 0.3739353 | 0.77675727 | 0.01455 | 0.373935255 | 0.373935255 | 7.402656731 | 0.4197913 | 0.784632639 | 1 | 0.419791299 | 0.419791299 | 8.320617079 | 0.890764665 | -0.050237019 | FALSE | TRUE | FALSE |
| OR5M11 | 0.3738795 | 0.77678639 | 1 | 0.373879503 | 0.373879503 | 0.373879503 | 0.40665584 | 0.789725314 | 1 | 3.7892071 | 3.7892071 | 3.7892071 | 0.91940031 | -0.036495354 | FALSE | TRUE | FALSE |
| MED8 | 0.3738506 | 0.77681551 | 1 | 0.373850605 | 0.373850605 | 0.373850605 | 0.4196545 | 0.784690313 | 0.00825 | 0.419654501 | 0.419654501 | 0.419654501 | 0.890853319 | -0.050193798 | FALSE | TRUE | FALSE |
| CXCL5 | 0.3738088 | 0.77684461 | 1 | 0.373808754 | 0.373808754 | 0.373808754 | 0.41922463 | 0.784776767 | 0.003 | 0.419224629 | 0.419224629 | 0.419224629 | 0.891666969 | -0.049797321 | FALSE | TRUE | FALSE |
| OXT | 0.3737283 | 0.77687371 | 1 | 0.373728314 | 0.373728314 | 0.373728314 | 0.41997673 | 0.784517194 | 1 | 0.419976728 | 0.419976728 | 0.419976728 | 0.889878627 | -0.050669224 | FALSE | TRUE | FALSE |
| TMEM61 | 0.3736484 | 0.7769028 | 0.0075 | 0.373648418 | 0.373648418 | 0.373648418 | 3.77444564 | 0.367961534 | 0.50185 | 3.774445636 | 3.774445636 | 3.774445636 | 0.098994251 | -1.004390026 | FALSE | FALSE | FALSE |
| NPC2 | 0.3736155 | 0.77693189 | 1 | 3.500014209 | 134.5869226 | 134.5869226 | 3.98093487 | 0.357091673 | 0.2545 | 3.98093487 | 38.33660378 | 38.33660378 | 0.093851205 | -1.027560147 | FALSE | FALSE | FALSE |
| CHMP6 | 0.3734056 | 0.77696096 | 1 | 0.373405625 | 0.373405625 | 0.373405625 | 0.41968143 | 0.78466148 | 1 | 0.419681435 | 0.419681435 | 0.419681435 | 0.889735867 | -0.050738902 | FALSE | TRUE | FALSE |
| SLC50A1 | 0.3733635 | 0.77699003 | 1 | 0.373363452 | 0.373363452 | 0.373363452 | 0.41897086 | 0.784834365 | 1 | 0.418970861 | 0.418970861 | 0.418970861 | 0.8911442 | -0.050052015 | FALSE | TRUE | FALSE |
| TESK1 | 0.3733277 | 0.77701909 | 1 | 0.373327674 | 0.373327674 | 7.354008174 | 3.82461336 | 0.365520051 | 0.0185 | 3.824613363 | 3.824613363 | 13.75635954 | 0.097611873 | -1.010497353 | FALSE | FALSE | FALSE |
| ERAL1 | 0.3733004 | 0.77704814 | 0.507 | 3.490458179 | 33.47492076 | 33.47492076 | 0.58047036 | 0.720591916 | 0.50125 | 0.580470359 | 0.580470359 | 1.255854033 | 0.64309981 | -0.191721619 | FALSE | TRUE | FALSE |
| PSMD11 | 0.3732172 | 0.77707719 | 1 | 33.89561309 | 335.6424364 | 335.6424364 | 0.44165355 | 0.776852981 | 1 | 4.166907257 | 16.6658192 | 16.6658192 | 0.845045124 | -0.0731201 | FALSE | TRUE | FALSE |
| FAM216B | 0.3731001 | 0.77710622 | 1 | 0.373100086 | 0.373100086 | 0.373100086 | 0.41845967 | 0.785236629 | 0.0041 | 0.418459671 | 0.418459671 | 0.418459671 | 0.891603449 | -0.04982826 | FALSE | TRUE | FALSE |
| NSMF | 0.3730672 | 0.77713525 | 1 | 0.373067172 | 0.373067172 | 7.381485397 | 0.41887441 | 0.784920701 | 0.01275 | 0.418874413 | 0.418874413 | 8.298495804 | 0.890642065 | -0.050296797 | FALSE | TRUE | FALSE |
| NPAS1 | 0.3729956 | 0.77716427 | 1 | 0.372995642 | 0.372995642 | 7.359573066 | 0.41894314 | 0.784863152 | 1 | 0.418943144 | 0.418943144 | 8.279542538 | 0.890325209 | -0.05045133 | FALSE | TRUE | FALSE |
| GPATCH11 | 0.3729745 | 0.77719329 | 1 | 3.459898013 | 3.459898013 | 3.459898013 | 0.43165008 | 0.780514659 | 1 | 0.431650079 | 0.431650079 | 0.431650079 | 0.864066808 | -0.063452677 | FALSE | TRUE | FALSE |
| OR1N1 | 0.3728848 | 0.77722229 | 1 | 0.372884836 | 0.372884836 | 0.372884836 | 0.41857657 | 0.785179261 | 1 | 0.418576575 | 0.418576575 | 0.418576575 | 0.890840192 | -0.050200197 | FALSE | TRUE | FALSE |
| CCR1 | 0.3727743 | 0.77725129 | 1 | 0.372774253 | 0.372774253 | 0.372774253 | 0.40594782 | 0.789944646 | 1 | 3.819351143 | 3.819351143 | 3.819351143 | 0.918281201 | -0.037024307 | FALSE | TRUE | FALSE |
| SSBP2 | 0.3726945 | 0.77728028 | 1 | 0.372694492 | 0.372694492 | 0.372694492 | 0.41871657 | 0.7850357 | 1 | 0.41871657 | 0.41871657 | 0.41871657 | 0.890087754 | -0.050567174 | FALSE | TRUE | FALSE |
| IMPA2 | 0.3725971 | 0.77730926 | 1 | 0.372597123 | 0.372597123 | 0.372597123 | 0.41847188 | 0.785207949 | 1 | 0.418471883 | 0.418471883 | 0.418471883 | 0.890375526 | -0.050426786 | FALSE | TRUE | FALSE |
| ZNF319 | 0.3724129 | 0.77733824 | 1 | 0.372412865 | 0.372412865 | 7.314818923 | 0.41870693 | 0.785064429 | 1 | 0.418706928 | 0.418706928 | 8.24185272 | 0.889435641 | -0.050885472 | FALSE | TRUE | FALSE |
| BACH2 | 0.3723824 | 0.77736721 | 1 | 0.372382378 | 0.372382378 | 7.303826442 | 0.41865586 | 0.785121862 | 1 | 0.418655861 | 0.418655861 | 8.230520065 | 0.889471313 | -0.050868054 | FALSE | TRUE | FALSE |
| FAM209A | 0.3722935 | 0.77739617 | 1 | 0.372293538 | 0.372293538 | 0.372293538 | 0.41768381 | 0.785465784 | 1 | 0.417683805 | 0.417683805 | 0.417683805 | 0.891328641 | -0.049962138 | FALSE | TRUE | FALSE |
| GID4 | 0.3722709 | 0.77742512 | 1 | 353764.4405 | 474259407.3 | 64266133605 | 0.47362847 | 0.765621339 | 1 | 4248.052811 | 5061906.336 | 56145008.46 | 0.785997762 | -0.104578691 | FALSE | TRUE | FALSE |
| HMOX1 | 0.3721054 | 0.77745406 | 1 | 0.372105398 | 0.372105398 | 0.372105398 | 3.67071838 | 0.37351832 | 0.25255 | 3.67071838 | 35.2203421 | 35.2203421 | 0.101371274 | -0.994085096 | FALSE | FALSE | FALSE |
| PLEKHA3 | 0.3720989 | 0.777483 | 1 | 0.372098882 | 0.372098882 | 0.372098882 | 0.41805459 | 0.78537991 | 1 | 0.418054589 | 0.418054589 | 0.418054589 | 0.890072472 | -0.050574631 | FALSE | TRUE | FALSE |
| GRPEL2 | 0.3720359 | 0.77751193 | 1 | 3.466711742 | 3.466711742 | 3.466711742 | 0.43099974 | 0.780634574 | 1 | 0.430999745 | 0.430999745 | 0.430999745 | 0.86319283 | -0.063892176 | FALSE | TRUE | FALSE |
| CXXC1 | 0.3717807 | 0.77754085 | 1 | 0.371780734 | 0.371780734 | 7.286756767 | 0.41796939 | 0.785408542 | 1 | 0.417969388 | 0.417969388 | 8.211787639 | 0.889492735 | -0.050857595 | FALSE | TRUE | FALSE |
| SLC39A14 | 0.3717611 | 0.77756977 | 1 | 0.371761139 | 0.371761139 | 7.369208067 | 0.41766309 | 0.785522994 | 1 | 0.417663088 | 0.417663088 | 8.287937019 | 0.890098143 | -0.050562105 | FALSE | TRUE | FALSE |
| RPL13A | 0.3717523 | 0.77759867 | 1 | 0.371752286 | 0.371752286 | 0.371752286 | 0.41725806 | 0.785665881 | 1 | 0.417258065 | 0.417258065 | 0.417258065 | 0.890940925 | -0.050151091 | FALSE | TRUE | FALSE |
| PEX11A | 0.3717501 | 0.77762757 | 1 | 0.37175014 | 0.37175014 | 0.37175014 | 0.41747443 | 0.785551587 | 1 | 0.417474433 | 0.417474433 | 0.417474433 | 0.890474027 | -0.050378743 | FALSE | TRUE | FALSE |
| DNASE2 | 0.371708 | 0.77765646 | 1 | 0.371707984 | 0.371707984 | 7.377696043 | 0.41729727 | 0.78560875 | 1 | 0.417297267 | 0.417297267 | 8.29012142 | 0.890751063 | -0.050243651 | FALSE | TRUE | FALSE |
| DENND2D | 0.3716205 | 0.77768534 | 0.5026 | 3.464518284 | 3.464518284 | 7.361287485 | 3117.28605 | 0.000710512 | 0.11505 | 310.621033 | 32047.63832 | 337320.4717 | 0.000119213 | -3.923676968 | FALSE | FALSE | FALSE |
| SH3BGRL3 | 0.3715604 | 0.77771422 | 1 | 3.467021124 | 3.467021124 | 3.467021124 | 0.43054339 | 0.780933763 | 1 | 0.430543388 | 0.430543388 | 0.430543388 | 0.863003495 | -0.063987445 | FALSE | TRUE | FALSE |
| C18orf21 | 0.3714954 | 0.77774308 | 0.49735 | 0.371495385 | 3.444795585 | 3.444795585 | 35.8031188 | 0.077652667 | 0.25255 | 3.754997681 | 35.80311876 | 35.80311876 | 0.010376062 | -1.983967437 | FALSE | FALSE | FALSE |
| RNASE11 | 0.3714002 | 0.77777194 | 1 | 0.371400161 | 0.371400161 | 0.371400161 | 0.41685719 | 0.785837082 | 1 | 0.416857191 | 0.416857191 | 0.416857191 | 0.890952991 | -0.05014521 | FALSE | TRUE | FALSE |
| SIGMAR1 | 0.37131 | 0.7778008 | 1 | 0.371310034 | 0.371310034 | 0.371310034 | 0.41268643 | 0.787449457 | 1 | 1.650533213 | 1.650533213 | 1.650533213 | 0.899738889 | -0.045883508 | FALSE | TRUE | FALSE |
| LRRC8A | 0.3712774 | 0.77782964 | 1 | 0.371277445 | 0.371277445 | 7.255362087 | 0.40457769 | 0.790762863 | 1 | 3.812150735 | 3.812150735 | 74.70129203 | 0.917691336 | -0.037303369 | FALSE | TRUE | FALSE |
| APOL3 | 0.3710631 | 0.77785848 | 0.00625 | 0.042634498 | 0.371063063 | 0.371063063 | 0.0608768 | 0.884571919 | 0.49665 | 0.060876796 | 0.535038531 | 0.535038531 | 6.095312011 | 0.784995941 | FALSE | TRUE | FALSE |
| APLNR | 0.3710271 | 0.77788731 | 1 | 3.494170973 | 3.494170973 | 7.345805501 | 0.43057371 | 0.780903883 | 1 | 0.43057371 | 0.43057371 | 8.535070628 | 0.861704065 | -0.064641858 | FALSE | TRUE | FALSE |
| ARL6IP1 | 0.3710244 | 0.77791613 | 1 | 0.37102438 | 0.37102438 | 0.37102438 | 0.41638124 | 0.785922579 | 1 | 0.416381241 | 0.416381241 | 0.416381241 | 0.891068914 | -0.050088707 | FALSE | TRUE | FALSE |
| B3GALT2 | 0.3707576 | 0.77794494 | 1 | 0.370757568 | 0.370757568 | 7.356736967 | 0.4170151 | 0.785751517 | 1 | 0.417015103 | 0.417015103 | 8.28243862 | 0.889074678 | -0.051061759 | FALSE | TRUE | FALSE |
| ATP2B1 | 0.3706517 | 0.77797375 | 0.7404 | 1.48121631 | 23.67276751 | 94.67319666 | 0.07270357 | 0.881865514 | 0.2734 | 0.072703575 | 0.290585476 | 2.452923151 | 5.098122121 | 0.707410234 | FALSE | TRUE | FALSE |
| CCDC25 | 0.3705858 | 0.77800255 | 1 | 1.482244217 | 1.482244217 | 1.482244217 | 1.64650176 | 0.521898078 | 0.2522 | 1.646501761 | 6.587240772 | 6.587240772 | 0.225074654 | -0.647673409 | FALSE | FALSE | FALSE |
| SNRPC | 0.3705822 | 0.77803134 | 1 | 0.370582195 | 0.370582195 | 0.370582195 | 0.38097594 | 0.798974227 | 0.00335 | 0.380975939 | 0.380975939 | 0.380975939 | 0.97271811 | -0.012012998 | FALSE | TRUE | FALSE |
| KRTAP4-5 | 0.3705065 | 0.77806013 | 1 | 3.432646371 | 32.6403075 | 32.6403075 | 0.44263369 | 0.776479833 | 1 | 0.44263369 | 0.44263369 | 0.44263369 | 0.837049938 | -0.077248632 | FALSE | TRUE | FALSE |
| NF2 | 0.3705002 | 0.7780889 | 0.5032 | 0.370500165 | 0.370500165 | 1.340114473 | 0.05920864 | 0.884852464 | 1 | 0.236674145 | 0.236674145 | 1.171659633 | 6.257535017 | 0.796403289 | FALSE | TRUE | FALSE |
| CLPX | 0.3704604 | 0.77811767 | 1 | 3.469904926 | 33.35651322 | 68.61619133 | 0.44347448 | 0.776261576 | 1 | 0.44347448 | 0.44347448 | 8.78144435 | 0.835358951 | -0.07812687 | FALSE | TRUE | FALSE |
| NIP7 | 0.3704287 | 0.77814643 | 0.0056 | 0.370428674 | 0.370428674 | 0.370428674 | 0.34487851 | 0.810735122 | 1 | 308.7191351 | 32465.86424 | 345072.1516 | 1.074084521 | 0.031038458 | FALSE | TRUE | FALSE |
| PMCH | 0.3703829 | 0.77817518 | 0.49955 | 0.039368234 | 0.370382917 | 0.370382917 | 0.04272707 | 0.888367833 | 1 | 0.042727074 | 0.042727074 | 0.042727074 | 8.668576716 | 0.937947797 | FALSE | TRUE | FALSE |
| ZBTB7C | 0.3702773 | 0.77820393 | 1 | 0.370277259 | 0.370277259 | 7.299570872 | 0.41643223 | 0.785894087 | 0.0166 | 0.416432234 | 0.416432234 | 8.223599406 | 0.8891657 | -0.051017299 | FALSE | TRUE | FALSE |
| REXO2 | 0.3701973 | 0.77823266 | 0.0072 | 0.370197319 | 0.370197319 | 0.370197319 | 0.4161999 | 0.786008006 | 1 | 0.416199904 | 0.416199904 | 0.416199904 | 0.889469976 | -0.050868707 | FALSE | TRUE | FALSE |
| KRTAP5-2 | 0.3701907 | 0.77826139 | 1 | 0.370190707 | 0.370190707 | 0.370190707 | 0.41554413 | 0.786320626 | 1 | 0.415544135 | 0.415544135 | 0.415544135 | 0.890857736 | -0.050191644 | FALSE | TRUE | FALSE |
| HMG20B | 0.3701631 | 0.77829011 | 1 | 0.370163142 | 0.370163142 | 0.370163142 | 0.41637823 | 0.785951062 | 1 | 0.416378228 | 0.416378228 | 0.416378228 | 0.889006958 | -0.05109484 | FALSE | TRUE | FALSE |
| TSC22D2 | 0.3700423 | 0.77831883 | 1 | 0.370042334 | 0.370042334 | 7.257959872 | 0.41619151 | 0.786036465 | 1 | 0.41619151 | 0.41619151 | 8.182102808 | 0.889115526 | -0.051041806 | FALSE | TRUE | FALSE |
| CLVS2 | 0.3700168 | 0.77834753 | 1 | 0.370016761 | 0.370016761 | 0.370016761 | 0.41625601 | 0.785979538 | 1 | 0.416256006 | 0.416256006 | 0.416256006 | 0.88891633 | -0.051139115 | FALSE | TRUE | FALSE |
| YTHDC1 | 0.3699928 | 0.77837623 | 1 | 0.36999285 | 0.36999285 | 7.288010207 | 0.40686573 | 0.78961547 | 0.0226 | 0.406865726 | 0.406865726 | 8.028901353 | 0.909373354 | -0.041257775 | FALSE | TRUE | FALSE |
| CA8 | 0.3699744 | 0.77840492 | 1 | 0.369974365 | 0.369974365 | 0.369974365 | 0.41586544 | 0.786150224 | 1 | 0.415865441 | 0.415865441 | 0.415865441 | 0.889649219 | -0.050781198 | FALSE | TRUE | FALSE |
| CTNNA3 | 0.36997 | 0.7784336 | 0.51075 | 0.369970035 | 0.369970035 | 7.272497707 | 0.23741136 | 0.842738081 | 0.75305 | 0.531579108 | 2.126097823 | 8.024529405 | 1.558350169 | 0.192665053 | FALSE | TRUE | FALSE |
| TMEM135 | 0.3698861 | 0.77846228 | 1 | 0.369886109 | 0.369886109 | 7.338121071 | 0.41564969 | 0.786263857 | 1 | 0.415649686 | 0.415649686 | 8.25400817 | 0.889898684 | -0.050659435 | FALSE | TRUE | FALSE |
| KPNA4 | 0.3697961 | 0.77849095 | 1 | 3.497131399 | 3.497131399 | 7.33451688 | 0.42957386 | 0.781321424 | 1 | 0.429573864 | 0.429573864 | 8.528653138 | 0.860844067 | -0.06507551 | FALSE | TRUE | FALSE |
| RNF26 | 0.3697405 | 0.77851961 | 1 | 0.369740465 | 0.369740465 | 7.318361363 | 1.60841383 | 0.524800298 | 0.8756 | 6.434539679 | 25.74813052 | 31.87531021 | 0.22987894 | -0.638500815 | FALSE | FALSE | FALSE |
| DUSP10 | 0.3697282 | 0.77854826 | 1 | 1.478752088 | 1.478752088 | 7.315812976 | 0.42058756 | 0.78434378 | 1 | 0.420587559 | 0.420587559 | 8.332829058 | 0.879075484 | -0.055973832 | FALSE | TRUE | FALSE |
| LZTS3 | 0.3696366 | 0.7785769 | 1 | 0.369636591 | 0.369636591 | 7.270829107 | 0.41578231 | 0.786178645 | 0.01805 | 0.41578231 | 0.41578231 | 8.194759931 | 0.889014712 | -0.051091052 | FALSE | TRUE | FALSE |
| KPRP | 0.3696258 | 0.77860554 | 0.7556 | 0.369625805 | 3.329586182 | 7.286177429 | 0.06221574 | 0.884255915 | 0.50405 | 0.55136972 | 0.55136972 | 2.105892609 | 5.941033329 | 0.773861989 | FALSE | TRUE | FALSE |
| TMEM38A | 0.3694515 | 0.77863417 | 1 | 0.36945152 | 0.36945152 | 0.36945152 | 0.41522115 | 0.786490741 | 1 | 0.415221151 | 0.415221151 | 0.415221151 | 0.889770474 | -0.05072201 | FALSE | TRUE | FALSE |
| TMEM200B | 0.3690832 | 0.77866279 | 1 | 0.36908317 | 0.36908317 | 0.36908317 | 0.41457058 | 0.786660579 | 1 | 0.414570577 | 0.414570577 | 0.414570577 | 0.890278256 | -0.050474234 | FALSE | TRUE | FALSE |
| BACE2 | 0.3690762 | 0.7786914 | 1 | 0.369076242 | 0.369076242 | 7.299558385 | 1.64207564 | 0.522480693 | 0.50915 | 1.642075638 | 1.642075638 | 8.130668092 | 0.224762023 | -0.648277069 | FALSE | FALSE | FALSE |
| ZBTB32 | 0.3690417 | 0.77872001 | 1 | 0.369041749 | 0.369041749 | 7.297896036 | 0.41493739 | 0.786547384 | 1 | 0.414937386 | 0.414937386 | 8.216587099 | 0.889391417 | -0.050907066 | FALSE | TRUE | FALSE |
| NUDT15 | 0.3690397 | 0.77874861 | 1 | 0.369039665 | 0.369039665 | 0.369039665 | 0.41450926 | 0.786688858 | 1 | 0.41450926 | 0.41450926 | 0.41450926 | 0.890304996 | -0.05046119 | FALSE | TRUE | FALSE |
| SUGT1 | 0.3688299 | 0.7787772 | 1 | 0.368829872 | 0.368829872 | 0.368829872 | 0.41448356 | 0.786773647 | 1 | 0.414483562 | 0.414483562 | 0.414483562 | 0.889854039 | -0.050681224 | FALSE | TRUE | FALSE |
| SOX7 | 0.3687005 | 0.77880579 | 0.814 | 1.474075362 | 5.894868635 | 23.57963379 | 0.03687763 | 0.889685643 | 0.8738 | 0.589165611 | 2.355794316 | 2.916792068 | 9.997943817 | 0.999910692 | FALSE | TRUE | FALSE |
| NKX6-1 | 0.3683888 | 0.77883437 | 0.0095 | 0.368388798 | 0.368388798 | 0.368388798 | 0.41467482 | 0.786632291 | 1 | 0.41467482 | 0.41467482 | 0.41467482 | 0.888379955 | -0.051401249 | FALSE | TRUE | FALSE |

| | | | | | | | | | | | | | | | | | |
|---|---|---|---|---|---|---|---|---|---|---|---|---|---|---|---|---|---|
| CD83 | 0.3683555 | 0.77886294 | 1 | 0.368355525 | 0.368355525 | 0.368355525 | 0.41374815 | 0.786999409 | 1 | 0.413748155 | 0.413748155 | 0.413748155 | 0.890289226 | -0.050468882 | FALSE | TRUE | FALSE |
| IGF2BP2 | 0.368295 | 0.77889151 | 0.0164 | 0.368294981 | 0.368294981 | 7.267582349 | 0.41426855 | 0.786858363 | 0.0146 | 0.414268551 | 0.414268551 | 8.187911498 | 0.889024715 | -0.051086166 | FALSE | TRUE | FALSE |
| SLC1A6 | 0.3681476 | 0.77892006 | 1 | 0.36814763 | 0.36814763 | 7.249183784 | 0.41449239 | 0.786745392 | 1 | 0.414492391 | 0.414492391 | 8.176936868 | 0.888189114 | -0.051494554 | FALSE | TRUE | FALSE |
| FBXO31 | 0.3681384 | 0.77894861 | 1 | 0.368138362 | 0.368138362 | 7.255484785 | 0.41436221 | 0.786830132 | 1 | 0.41436221 | 0.41436221 | 8.180812739 | 0.888445792 | -0.051369065 | FALSE | TRUE | FALSE |
| C1orf50 | 0.3680711 | 0.77897715 | 0.00565 | 0.368071113 | 0.368071113 | 0.368071113 | 3.73203629 | 0.369419898 | 0.50155 | 3.732036293 | 3.732036293 | 3.732036293 | 0.098624741 | -1.006014124 | FALSE | FALSE | FALSE |
| SOX8 | 0.3680309 | 0.77900569 | 1 | 1.87758E+19 | 3.22819E+23 | 2.11704E+25 | 3.08346912 | 0.407248874 | 1 | 1.85809E+11 | 4.6865E+14 | 7.57613E+15 | 0.119356109 | -0.923155347 | FALSE | FALSE | FALSE |
| CRIPT | 0.3679241 | 0.77903422 | 1 | 0.367924128 | 0.367924128 | 0.367924128 | 0.40917611 | 0.788732408 | 1 | 0.409176107 | 1.636472082 | 1.636472082 | 0.899182826 | -0.046151997 | FALSE | TRUE | FALSE |
| ZNF778 | 0.3679061 | 0.77906273 | 0.25825 | 0.04250668 | 0.367906127 | 0.837446061 | 0.00968239 | 0.89682693 | 0.02015 | 0.009682391 | 0.009682391 | 0.191100439 | 37.99744638 | 1.579754411 | FALSE | TRUE | FALSE |
| C16orf86 | 0.3678135 | 0.77909125 | 0.00955 | 0.367813498 | 0.367813498 | 0.367813498 | 0.36514995 | 0.804114755 | 0.93875 | 32.22165325 | 3146.714474 | 3146.714474 | 1.007294388 | 0.003156414 | FALSE | TRUE | FALSE |
| SUZ12 | 0.3677207 | 0.77911975 | 0.01825 | 0.367720703 | 0.367720703 | 7.268907658 | 0.41377664 | 0.786943014 | 1 | 0.413776645 | 0.413776645 | 8.190758456 | 0.888693713 | -0.051247892 | FALSE | TRUE | FALSE |
| CLUAP1 | 0.3676905 | 0.77914825 | 1 | 0.367690534 | 0.367690534 | 0.367690534 | 0.41338052 | 0.787140264 | 1 | 0.413380519 | 0.413380519 | 0.413380519 | 0.889472331 | -0.050867557 | FALSE | TRUE | FALSE |
| SCD5 | 0.3676322 | 0.77917673 | 1 | 0.367632197 | 0.367632197 | 7.286473083 | 0.41337609 | 0.787168411 | 1 | 0.413376091 | 0.413376091 | 0.413376091 | 0.889340736 | -0.050931814 | FALSE | TRUE | FALSE |
| RBP4 | 0.3675977 | 0.77920522 | 0.0089 | 0.367597716 | 0.367597716 | 0.367597716 | 0.40093955 | 0.791924148 | 1 | 0.400939549 | 3.775165186 | 3.775165186 | 0.916840747 | -0.037706094 | FALSE | TRUE | FALSE |
| TAF9 | 0.3673978 | 0.77923369 | 1 | 0.36739777 | 0.36739777 | 0.36739777 | 0.4130951 | 0.787252806 | 1 | 0.413095101 | 0.413095101 | 0.413095101 | 0.889378182 | -0.050913529 | FALSE | TRUE | FALSE |
| BZW1 | 0.3673691 | 0.77926216 | 0.008 | 0.367369052 | 0.367369052 | 0.367369052 | 0.41345405 | 0.787083946 | 1 | 0.413454051 | 0.413454051 | 0.413454051 | 0.888536588 | -0.051324684 | FALSE | TRUE | FALSE |
| CLEC1B | 0.3673358 | 0.77929061 | 1 | 0.367335792 | 0.367335792 | 0.367335792 | 0.41278643 | 0.787393311 | 1 | 0.41278643 | 0.41278643 | 0.41278643 | 0.889893089 | -0.050662166 | FALSE | TRUE | FALSE |
| ANAPC10 | 0.3672286 | 0.77931907 | 0.00395 | 0.367228597 | 0.367228597 | 0.367228597 | 0.41316555 | 0.787224682 | 1 | 0.413165552 | 0.413165552 | 0.413165552 | 0.888817075 | -0.051187611 | FALSE | TRUE | FALSE |
| POLR2G | 0.367193 | 0.77934751 | 1 | 0.367193 | 0.367193 | 0.367193 | 0.41284066 | 0.787337132 | 1 | 0.412840665 | 0.412840665 | 0.412840665 | 0.889430309 | -0.050888075 | FALSE | TRUE | FALSE |
| OLFML1 | 0.3669873 | 0.77937595 | 0.7256 | 0.853671006 | 13.63786445 | 54.53020169 | 0.66695037 | 0.682139332 | 0.25265 | 0.166867253 | 0.666950371 | 0.666950371 | 0.550246849 | -0.259442436 | FALSE | TRUE | FALSE |
| CDCA8 | 0.3669131 | 0.77940438 | 0.00915 | 0.366913115 | 0.366913115 | 0.366913115 | 0.4126377 | 0.787477519 | 1 | 0.412637704 | 0.412637704 | 0.412637704 | 0.889189504 | -0.051005673 | FALSE | TRUE | FALSE |
| AVPI1 | 0.3668949 | 0.7794328 | 1 | 0.366894948 | 0.366894948 | 0.366894948 | 0.41227638 | 0.787533622 | 1 | 0.412276381 | 0.412276381 | 0.412276381 | 0.889924733 | -0.050646723 | FALSE | TRUE | FALSE |
| OSBPL10 | 0.3666836 | 0.77946121 | 0.5153 | 0.366683615 | 0.366683615 | 7.221485834 | 0.53403346 | 0.74084958 | 0.50885 | 0.534033464 | 0.534033464 | 2.008956018 | 0.686630407 | -0.163276968 | FALSE | TRUE | FALSE |
| SLC2A14 | 0.3665175 | 0.77948962 | 0.9398 | 3.408153548 | 130.0795472 | 130.0795472 | 18.7034176 | 0.139771577 | 0.1342 | 2.004653978 | 18.70341759 | 18.70341759 | 0.019596285 | -1.707826257 | FALSE | FALSE | FALSE |
| SCRIB | 0.3664566 | 0.77951802 | 0.523 | 0.366456591 | 0.763284195 | 7.038119074 | 0.05989186 | 0.884750584 | 1 | 0.059891857 | 0.059891857 | 1.155789131 | 6.118638028 | 0.786654762 | FALSE | TRUE | FALSE |
| UPP1 | 0.366416 | 0.77954641 | 0.6871 | 0.366415954 | 5.861391865 | 5.861391865 | 9.03143021 | 0.211125371 | 0.19075 | 0.564492741 | 9.031430209 | 36.13835234 | 0.040571199 | -1.391782155 | FALSE | FALSE | FALSE |
| ST8SIA1 | 0.3663861 | 0.7795748 | 1 | 0.366386079 | 0.366386079 | 7.268976126 | 0.41208997 | 0.787589695 | 0.00965 | 0.412089973 | 0.412089973 | 0.412089973 | 0.889092439 | -0.051053083 | FALSE | TRUE | FALSE |
| ODF2 | 0.3663674 | 0.77960318 | 0.50895 | 0.366367376 | 0.366367376 | 7.18849426 | 0.06139911 | 0.884452535 | 0.02155 | 0.061399108 | 0.061399108 | 1.207460968 | 5.966982061 | 0.775754732 | FALSE | TRUE | FALSE |
| PNMA2 | 0.3662915 | 0.77963155 | 1 | 0.366291456 | 0.366291456 | 7.265076395 | 0.41198915 | 0.78761772 | 0.00945 | 0.411989149 | 0.411989149 | 0.411989149 | 0.889080348 | -0.051058989 | FALSE | TRUE | FALSE |
| SLC41A2 | 0.3661684 | 0.77965991 | 1 | 0.366168404 | 0.366168404 | 7.26601895 | 0.41197706 | 0.787645737 | 1 | 0.411977061 | 0.411977061 | 8.182716246 | 0.88880775 | -0.051192167 | FALSE | TRUE | FALSE |
| PHF19 | 0.3660854 | 0.77968826 | 1 | 0.366085443 | 0.366085443 | 7.223540995 | 0.39959118 | 0.792192343 | 1 | 3.762380745 | 3.762380745 | 13.52518491 | 0.916149954 | -0.038033436 | FALSE | TRUE | FALSE |
| BID | 0.3660046 | 0.77971661 | 1 | 3.394296864 | 3.394296864 | 3.394296864 | 0.4245016 | 0.782975361 | 1 | 0.424501604 | 0.424501604 | 0.424501604 | 0.862198363 | -0.064392806 | FALSE | TRUE | FALSE |
| UBR1 | 0.3659159 | 0.77974495 | 0.51665 | 0.365915851 | 0.365915851 | 7.097485305 | 0.05802243 | 0.885038845 | 0.759 | 0.529243425 | 2.116594122 | 17.78145909 | 6.306455319 | 0.799785323 | FALSE | TRUE | FALSE |
| RETN | 0.365794 | 0.77977329 | 1 | 0.365793951 | 0.365793951 | 0.365793951 | 0.4113441 | 0.787981336 | 1 | 0.411344104 | 0.411344104 | 0.411344104 | 0.889265088 | -0.050968758 | FALSE | TRUE | FALSE |
| IWS1 | 0.3656815 | 0.77980161 | 0.02395 | 0.365681527 | 0.365681527 | 7.201997324 | 3.743124 | 0.368691388 | 0.51195 | 3.743124002 | 3.743124002 | 13.48676308 | 0.097694206 | -1.010131193 | FALSE | FALSE | FALSE |
| MBLAC1 | 0.3655512 | 0.77982993 | 1 | 3.417278784 | 3.417278784 | 3.417278784 | 0.41143642 | 0.787897542 | 1 | 3.853314162 | 3.853314162 | 3.853314162 | 0.888475604 | -0.051354493 | FALSE | TRUE | FALSE |
| FKBP3 | 0.3655061 | 0.77985824 | 0.00675 | 0.365506116 | 0.365506116 | 0.365506116 | 0.41137303 | 0.787953413 | 1 | 0.41137303 | 0.41137303 | 0.41137303 | 0.888502866 | -0.051341167 | FALSE | TRUE | FALSE |
| RAB11FIP3 | 0.3654779 | 0.77988655 | 1 | 0.365477925 | 0.365477925 | 7.194090954 | 0.41157299 | 0.78784164 | 1 | 0.411572988 | 0.411572988 | 8.11683406 | 0.888002701 | -0.051585713 | FALSE | TRUE | FALSE |
| GPC5 | 0.3654004 | 0.77991484 | 0.01635 | 0.365400449 | 0.365400449 | 3.092787119 | 9.65838119 | 0.203360948 | 0.2577 | 1.038938191 | 9.658381188 | 20.56388354 | 0.037832473 | -1.422135265 | FALSE | FALSE | FALSE |
| REPS1 | 0.3653687 | 0.77994313 | 0.02 | 0.36536868 | 0.36536868 | 7.207159391 | 0.41137652 | 0.787925481 | 1 | 0.41137652 | 0.41137652 | 8.128089512 | 0.88816124 | -0.051508184 | FALSE | TRUE | FALSE |
| TSPYL4 | 0.3651592 | 0.77997142 | 1 | 0.365159169 | 0.365159169 | 7.249858971 | 0.4107864 | 0.788232296 | 1 | 0.410786404 | 0.410786404 | 0.410786404 | 0.888927106 | -0.051133851 | FALSE | TRUE | FALSE |
| RBM27 | 0.365044 | 0.77999969 | 1 | 0.365044015 | 0.365044015 | 12.28399367 | 0.41143738 | 0.787869595 | 0.02595 | 0.411437375 | 0.411437375 | 8.088935025 | 0.887240775 | -0.051958508 | FALSE | TRUE | FALSE |
| SNTN | 0.3649869 | 0.78002796 | 0.00335 | 0.364986907 | 0.364986907 | 0.364986907 | 0.41030791 | 0.788343643 | 1 | 0.410307914 | 0.410307914 | 0.410307914 | 0.88954391 | -0.050832609 | FALSE | TRUE | FALSE |
| TRIM11 | 0.3649105 | 0.78005622 | 1 | 0.364910513 | 0.364910513 | 7.208105362 | 0.41112244 | 0.788092952 | 1 | 0.411122444 | 0.411122444 | 8.13299704 | 0.8875957 | -0.05178481 | FALSE | TRUE | FALSE |
| TK1 | 0.3649016 | 0.78008447 | 1 | 0.364901613 | 0.364901613 | 0.364901613 | 0.41110298 | 0.788148712 | 1 | 0.41110298 | 0.41110298 | 0.41110298 | 0.887616074 | -0.051774841 | FALSE | TRUE | FALSE |
| ARHGEF38 | 0.3648469 | 0.78011272 | 0.8783 | 1.459473326 | 5.839694763 | 5.839694763 | 1.92712048 | 0.495881837 | 0.13305 | 1.927120484 | 7.711034563 | 7.711034563 | 0.189322331 | -0.722798158 | FALSE | FALSE | FALSE |
| LEO1 | 0.3648046 | 0.78014095 | 0.0178 | 0.364804552 | 0.364804552 | 7.201700033 | 0.41087438 | 0.788204442 | 1 | 0.41087438 | 0.41087438 | 8.123803396 | 0.887873691 | -0.051648813 | FALSE | TRUE | FALSE |
| USP15 | 0.3647882 | 0.78016918 | 1 | 0.364788244 | 0.364788244 | 7.184688293 | 0.41111105 | 0.788120836 | 1 | 0.411111052 | 0.411111052 | 8.111877388 | 0.887322885 | -0.051918318 | FALSE | TRUE | FALSE |
| ZFP36L2 | 0.3645328 | 0.78019741 | 1 | 0.364532817 | 0.364532817 | 7.195351036 | 0.41041869 | 0.788287985 | 1 | 0.41041869 | 0.41041869 | 8.113815101 | 0.888197408 | -0.051490499 | FALSE | TRUE | FALSE |
| MYPN | 0.3645319 | 0.78022562 | 0.1499 | 0.297563799 | 1.190288939 | 5.828469311 | 0.03028818 | 0.891190603 | 0.51415 | 0.121103228 | 0.121103228 | 2.377979957 | 12.03545178 | 1.080462397 | FALSE | TRUE | FALSE |
| TSPAN14 | 0.3642839 | 0.78025383 | 0.0093 | 0.3642839 | 0.3642839 | 0.3642839 | 0.41045731 | 0.788260145 | 1 | 0.41045731 | 0.41045731 | 0.41045731 | 0.887507401 | -0.051828017 | FALSE | TRUE | FALSE |
| GOLT1A | 0.3642479 | 0.78028203 | 1 | 0.364247946 | 0.364247946 | 0.364247946 | 1.62143849 | 0.523497855 | 0.4988 | 0.405388714 | 1.621438489 | 1.621438489 | 0.224644936 | -0.648503366 | FALSE | FALSE | FALSE |
| RHCE | 0.3642105 | 0.78031023 | 1 | 32.44399534 | 318.1808007 | 318.1808007 | 0.43640838 | 0.778699553 | 0.50895 | 4.082219251 | 4.082219251 | 4.082219251 | 0.834563429 | -0.07854065 | FALSE | TRUE | FALSE |
| NDUFB8 | 0.3641097 | 0.78033841 | 0.0062 | 0.364109704 | 0.364109704 | 0.364109704 | 0.41023383 | 0.78839927 | 1 | 0.410233832 | 0.410233832 | 0.410233832 | 0.887566252 | -0.051799219 | FALSE | TRUE | FALSE |
| KLF1 | 0.3640455 | 0.78036659 | 1 | 0.364045483 | 0.364045483 | 7.222241726 | 0.41008668 | 0.788427072 | 0.0102 | 0.410086685 | 0.410086685 | 8.143529697 | 0.887728124 | -0.051720021 | FALSE | TRUE | FALSE |
| HERPUD1 | 0.3639994 | 0.78039476 | 1 | 0.363999369 | 0.363999369 | 7.219055428 | 0.40987137 | 0.788454868 | 1 | 0.409871371 | 0.409871371 | 0.409871371 | 0.888081957 | -0.051546953 | FALSE | TRUE | FALSE |
| GCLM | 0.3637929 | 0.78042293 | 1 | 0.363792891 | 0.363792891 | 0.363792891 | 0.38011696 | 0.799248982 | 1 | 0.38011696 | 0.38011696 | 0.38011696 | 0.957055141 | -0.01906304 | FALSE | TRUE | FALSE |
| MORN4 | 0.363624 | 0.78045109 | 1 | 0.363624036 | 0.363624036 | 0.363624036 | 0.40962093 | 0.78853821 | 1 | 0.409620926 | 0.409620926 | 0.409620926 | 0.887708643 | -0.051729552 | FALSE | TRUE | FALSE |
| OXNAD1 | 0.3635573 | 0.78047924 | 0.75405 | 0.82279225 | 3.290580257 | 3.290580257 | 0.12514533 | 0.870112788 | 1 | 0.125145329 | 0.125145329 | 0.125145329 | 2.905080752 | 0.463158209 | FALSE | TRUE | FALSE |
| OR4N4 | 0.3634784 | 0.78050738 | 1 | 0.363478403 | 0.363478403 | 0.363478403 | 0.40907826 | 0.788787824 | 0.0064 | 0.40907826 | 0.40907826 | 0.40907826 | 0.888530236 | -0.051327789 | FALSE | TRUE | FALSE |
| POLR1D | 0.3633848 | 0.78053552 | 0.0036 | 0.363384798 | 0.363384798 | 0.363384798 | 0.39683227 | 0.793019335 | 1 | 0.396832268 | 3.729981476 | 3.729981476 | 0.915713836 | -0.038240224 | FALSE | TRUE | FALSE |
| TRIM26 | 0.3633546 | 0.78056365 | 1 | 0.363354633 | 0.363354633 | 7.172136746 | 0.4094181 | 0.788649225 | 0.01555 | 0.409418102 | 0.409418102 | 8.094075004 | 0.887490394 | -0.051836339 | FALSE | TRUE | FALSE |
| CXCL16 | 0.363331 | 0.78059177 | 1 | 0.363331018 | 0.363331018 | 0.363331018 | 0.40888807 | 0.788843212 | 1 | 0.408888074 | 0.408888074 | 0.408888074 | 0.888583064 | -0.051301969 | FALSE | TRUE | FALSE |
| CCDC60 | 0.3633267 | 0.78061988 | 0.0155 | 0.363326708 | 0.363326708 | 0.820466507 | 35.5395185 | 0.0787718 | 0.63735 | 336.8749267 | 32524.94014 | 330922.1814 | 0.010223175 | -1.990414216 | FALSE | FALSE | FALSE |
| MFAP1 | 0.3632826 | 0.78064799 | 1 | 0.363282578 | 0.363282578 | 7.188671436 | 8.11419395 | 0.225569647 | 0.00405 | 0.409532279 | 0.409532279 | 8.114193949 | 0.044771247 | -1.349000813 | FALSE | FALSE | FALSE |
| FYTTD1 | 0.3632487 | 0.78067609 | 0.0102 | 0.363248718 | 0.363248718 | 7.21186155 | 0.40891035 | 0.788815522 | 1 | 0.40891035 | 0.40891035 | 0.40891035 | 0.888333392 | -0.051424013 | FALSE | TRUE | FALSE |
| MYL12A | 0.3632324 | 0.78070418 | 1 | 0.363232442 | 1.452784247 | 1.452784247 | 6.47282105 | 0.255782586 | 0.25315 | 1.617949744 | 6.47282105 | 6.47282105 | 0.056116559 | -1.25090897 | FALSE | FALSE | FALSE |
| OR5AK2 | 0.3631599 | 0.78073226 | 0.87635 | 32.12624067 | 313.9726708 | 313.9726708 | 4.05684488 | 0.353058042 | 0.5052 | 4.056844883 | 4.056844883 | 4.056844883 | 0.089517818 | -1.048090511 | FALSE | FALSE | FALSE |
| DPCD | 0.3631151 | 0.78076033 | 0.00595 | 0.363115078 | 0.363115078 | 0.363115078 | 0.40864396 | 0.789036821 | 1 | 0.40864396 | 0.40864396 | 0.40864396 | 0.888585452 | -0.051300801 | FALSE | TRUE | FALSE |
| FAM220A | 0.3631059 | 0.7807884 | 1 | 3.367346925 | 3.367346925 | 3.367346925 | 35.0986766 | 0.079613682 | 0.2543 | 3.680790625 | 35.09867663 | 35.09867663 | 0.010345289 | -1.985257383 | FALSE | FALSE | FALSE |
| FADS2 | 0.3629933 | 0.78081646 | 1 | 0.362993257 | 0.362993257 | 7.185409338 | 0.40920219 | 0.788704688 | 1 | 0.409202191 | 0.409202191 | 8.110102915 | 0.887075547 | -0.052039393 | FALSE | TRUE | FALSE |
| ZNF768 | 0.3627433 | 0.78084452 | 1 | 0.362743265 | 0.362743265 | 7.14603669 | 0.40879387 | 0.788926233 | 1 | 0.408793875 | 0.408793875 | 8.067773223 | 0.887350049 | -0.051905022 | FALSE | TRUE | FALSE |
| CCDC159 | 0.3626723 | 0.78087256 | 0.50465 | 0.362672293 | 0.362672293 | 0.362672293 | 3.86761229 | 0.363062778 | 0.2547 | 0.408404939 | 3.867612291 | 3.867612291 | 0.093771626 | -1.027928554 | FALSE | FALSE | FALSE |
| CDSN | 0.3625979 | 0.7809006 | 1 | 0.36259786 | 0.36259786 | 7.183259684 | 0.40835864 | 0.78906445 | 1 | 0.408358643 | 0.408358643 | 8.099023229 | 0.887939723 | -0.051616515 | FALSE | TRUE | FALSE |

| | | | | | | | | | | | | | | | | | |
|---|---|---|---|---|---|---|---|---|---|---|---|---|---|---|---|---|---|
| DHFR | 0.3625236 | 0.78092864 | 1 | 0.362523642 | 0.362523642 | 0.362523642 | 0.4086467 | 0.788981543 | 1 | 0.408646704 | 0.408646704 | 0.408646704 | 0.887132181 | -0.052011666 | FALSE | TRUE | FALSE |
| OR6B2 | 0.3623514 | 0.78095666 | 1 | 32.18992394 | 315.256836 | 315.256836 | 0.44811323 | 0.774942866 | 1 | 0.44811323 | 0.44811323 | 0.44811323 | 0.808615798 | -0.092257778 | FALSE | TRUE | FALSE |
| VASH2 | 0.3623237 | 0.78098468 | 1 | 0.362323687 | 0.362323687 | 0.362323687 | 0.4080348 | 0.78920248 | 1 | 0.408034805 | 0.408034805 | 0.408034805 | 0.887972503 | -0.051600482 | FALSE | TRUE | FALSE |
| SLC30A4 | 0.362307 | 0.78101269 | 1 | 0.362306999 | 0.362306999 | 7.183374823 | 0.40833338 | 0.789092072 | 0.01085 | 0.408333381 | 0.408333381 | 8.104362983 | 0.887282344 | -0.05193816 | FALSE | TRUE | FALSE |
| BTBD17 | 0.3621527 | 0.78104069 | 0.99095 | 522.7990388 | 52196.86986 | 135145.1908 | 0.29766784 | 0.826062534 | 1 | 132616442.3 | 2.75868E+11 | 4.35294E+12 | 1.216633566 | 0.085159794 | FALSE | TRUE | FALSE |
| NDUFAF4 | 0.3620479 | 0.78106869 | 1 | 0.362047863 | 1.447877573 | 1.447877573 | 1.58212762 | 0.528381038 | 0.74835 | 3.73209305 | 14.92551811 | 14.92551811 | 0.228836068 | -0.640475523 | FALSE | FALSE | FALSE |
| DCAF12 | 0.3619358 | 0.78109668 | 1 | 0.361935826 | 0.361935826 | 7.170947746 | 0.4079201 | 0.78925764 | 1 | 0.407920096 | 0.407920096 | 8.091123903 | 0.887271377 | -0.051943528 | FALSE | TRUE | FALSE |
| CALCRL | 0.3619161 | 0.78112466 | 1 | 0.36191614 | 0.36191614 | 7.183165496 | 0.40797408 | 0.789230064 | 1 | 0.407974077 | 0.407974077 | 0.407974077 | 0.887105725 | -0.052024618 | FALSE | TRUE | FALSE |
| SUV39H2 | 0.3616963 | 0.78115264 | 1 | 3.397651991 | 3.397651991 | 7.184756549 | 0.42093029 | 0.784199055 | 1 | 0.420930291 | 0.420930291 | 0.420930291 | 0.85927831 | -0.065866151 | FALSE | TRUE | FALSE |
| SIAH1 | 0.3616838 | 0.78118061 | 0.00745 | 0.361683826 | 0.361683826 | 0.361683826 | 0.39316667 | 0.794365233 | 1 | 0.393166674 | 0.393166674 | 0.393166674 | 0.919924933 | -0.03624761 | FALSE | TRUE | FALSE |
| TMEM256 | 0.3616706 | 0.78120857 | 1 | 0.361670574 | 0.361670574 | 0.361670574 | 0.40707649 | 0.789477994 | 1 | 0.407076492 | 0.407076492 | 0.407076492 | 0.888458513 | -0.051362847 | FALSE | TRUE | FALSE |
| PLA2G12B | 0.3616548 | 0.78123652 | 1 | 0.361654789 | 0.361654789 | 0.361654789 | 0.40736531 | 0.789367877 | 1 | 0.407365313 | 0.407365313 | 0.407365313 | 0.887789846 | -0.051689826 | FALSE | TRUE | FALSE |
| PNMA1 | 0.3616387 | 0.78126446 | 0.01025 | 0.361638683 | 0.361638683 | 7.176005155 | 0.4073167 | 0.78942295 | 1 | 0.407316697 | 0.407316697 | 0.407316697 | 0.887856268 | -0.051657335 | FALSE | TRUE | FALSE |
| PYCR1 | 0.3616364 | 0.7812924 | 0.50385 | 0.361636413 | 0.361636413 | 0.361636413 | 1.02809614 | 0.585431769 | 0.7507 | 4.111503698 | 38.54097666 | 38.54097666 | 0.351753496 | -0.453761578 | FALSE | FALSE | FALSE |
| CNN3 | 0.3615872 | 0.78132033 | 1 | 0.361587231 | 0.361587231 | 0.361587231 | 0.40787775 | 0.789285209 | 1 | 0.407877755 | 0.407877755 | 0.407877755 | 0.886508828 | -0.052316935 | FALSE | TRUE | FALSE |
| OR4F17 | 0.3615814 | 0.78134825 | 1 | 0.361581379 | 3.43058681 | 3.43058681 | 0.32696218 | 0.816376765 | 1 | 298.5163328 | 3045.158995 | 31821.39196 | 1.105881354 | 0.043708536 | FALSE | TRUE | FALSE |
| LECT2 | 0.3615119 | 0.78137617 | 1 | 0.361511861 | 0.361511861 | 0.361511861 | 0.40704138 | 0.789533007 | 1 | 0.407041381 | 0.407041381 | 0.407041381 | 0.888145229 | -0.051516013 | FALSE | TRUE | FALSE |
| SENP5 | 0.3613904 | 0.78140408 | 1 | 0.36139043 | 0.36139043 | 7.133494012 | 0.40744657 | 0.789312772 | 1 | 0.407446575 | 0.407446575 | 8.055225866 | 0.886963968 | -0.052094023 | FALSE | TRUE | FALSE |
| NAT8L | 0.3611726 | 0.78143198 | 1 | 0.361172642 | 0.361172642 | 0.361172642 | 0.36880554 | 0.802608165 | 1 | 0.368805543 | 0.368805543 | 0.368805543 | 0.979303725 | -0.009082594 | FALSE | TRUE | FALSE |
| PPP2R5C | 0.3611665 | 0.78145987 | 0.0149 | 0.361166479 | 0.361166479 | 7.151133534 | 0.40741293 | 0.789340228 | 0.01255 | 0.407412932 | 0.407412932 | 8.07651656 | 0.886487519 | -0.052327375 | FALSE | TRUE | FALSE |
| SNX33 | 0.3610894 | 0.78148776 | 1 | 0.361089371 | 0.361089371 | 7.115922898 | 0.40715073 | 0.789450475 | 0.01685 | 0.407150732 | 0.407150732 | 8.03780202 | 0.88686902 | -0.052140515 | FALSE | TRUE | FALSE |
| SOD2 | 0.3610015 | 0.78151564 | 1 | 0.361001518 | 0.361001518 | 0.361001518 | 0.39420049 | 0.794076462 | 1 | 3.668836951 | 3.668836951 | 3.668836951 | 0.915781508 | -0.03820813 | FALSE | TRUE | FALSE |
| RBM15B | 0.361001 | 0.78154351 | 1 | 3.399932367 | 3.399932367 | 66.47442625 | 0.39183649 | 0.794992374 | 1 | 3.690656526 | 35.67787117 | 72.35342978 | 0.921305148 | -0.035596502 | FALSE | TRUE | FALSE |
| SAFB | 0.3609804 | 0.78157137 | 1 | 0.3609804 | 0.3609804 | 7.034129994 | 0.40733714 | 0.789395417 | 1 | 0.407337137 | 0.407337137 | 7.962021558 | 0.886195652 | -0.052470385 | FALSE | TRUE | FALSE |
| PRR4 | 0.3609235 | 0.78159923 | 1 | 0.36092351 | 0.36092351 | 0.36092351 | 0.40633636 | 0.789835038 | 1 | 0.406336364 | 0.406336364 | 0.406336364 | 0.888238273 | -0.051470517 | FALSE | TRUE | FALSE |
| PTX4 | 0.3607434 | 0.78162708 | 0.75195 | 0.360743382 | 3.26272005 | 7.113016279 | 0.06259395 | 0.884144507 | 1 | 0.062593949 | 0.062593949 | 1.23630321 | 5.763230943 | 0.760666023 | FALSE | TRUE | FALSE |
| OR13G1 | 0.3607247 | 0.78165492 | 1 | 0.360724744 | 0.360724744 | 0.360724744 | 0.40674211 | 0.789670407 | 1 | 0.40674211 | 0.40674211 | 0.40674211 | 0.886863532 | -0.052143203 | FALSE | TRUE | FALSE |
| EIF3B | 0.3606561 | 0.78168276 | 0.02185 | 0.360656071 | 0.360656071 | 7.105076658 | 0.40686751 | 0.78958799 | 1 | 0.406867509 | 0.406867509 | 8.029912383 | 0.88642141 | -0.052359763 | FALSE | TRUE | FALSE |
| P4HB | 0.3605496 | 0.78171058 | 1 | 3.383065291 | 3.383065291 | 12.22105187 | 3.70016587 | 0.371113979 | 0.2573 | 3.700165875 | 35.65125767 | 35.65125767 | 0.097441467 | -1.011256185 | FALSE | FALSE | FALSE |
| UBALD2 | 0.3604295 | 0.7817384 | 1 | 330973.6001 | 438171304.5 | 5018933142 | 0.38147591 | 0.79862339 | 1 | 352670.2155 | 469431242.1 | 63050612914 | 0.944828966 | -0.024646801 | FALSE | TRUE | FALSE |
| CHAF1A | 0.3603984 | 0.78176622 | 0.0302 | 0.360398448 | 0.360398448 | 7.038869473 | 0.40672304 | 0.789697864 | 1 | 0.406723043 | 0.406723043 | 7.966078987 | 0.886102852 | -0.052515866 | FALSE | TRUE | FALSE |
| ANKRD13B | 0.3603358 | 0.78179402 | 1 | 0.360335833 | 0.360335833 | 7.105966357 | 0.40657644 | 0.789752756 | 1 | 0.406576441 | 0.406576441 | 8.031339244 | 0.886268354 | -0.052434758 | FALSE | TRUE | FALSE |
| RAET1L | 0.3602148 | 0.78182182 | 1 | 0.360214809 | 0.360214809 | 0.360214809 | 0.40572269 | 0.790108833 | 0.0061 | 0.405722689 | 0.405722689 | 0.405722689 | 0.887835012 | -0.051667732 | FALSE | TRUE | FALSE |
| C12orf71 | 0.3601932 | 0.78184962 | 1 | 0.360193228 | 0.360193228 | 0.360193228 | 0.40570847 | 0.7901635 | 1 | 0.405708467 | 0.405708467 | 0.405708467 | 0.887812943 | -0.051678528 | FALSE | TRUE | FALSE |
| ARL2BP | 0.3601784 | 0.7818774 | 1 | 0.360178361 | 0.360178361 | 0.360178361 | 0.40551742 | 0.790245445 | 1 | 0.405517416 | 0.405517416 | 0.405517416 | 0.888194556 | -0.051491893 | FALSE | TRUE | FALSE |
| OR4P4 | 0.3601597 | 0.78190518 | 1 | 0.360159661 | 0.360159661 | 0.360159661 | 0.40571943 | 0.790136171 | 1 | 0.405719428 | 0.405719428 | 0.405719428 | 0.887706222 | -0.051730736 | FALSE | TRUE | FALSE |
| C1QL4 | 0.3601276 | 0.78193295 | 1 | 0.360127647 | 0.360127647 | 0.360127647 | 0.4064022 | 0.789807618 | 1 | 0.406402203 | 0.406402203 | 0.406402203 | 0.88613606 | -0.05249959 | FALSE | TRUE | FALSE |
| ISG20 | 0.360093 | 0.78196071 | 1 | 0.360093045 | 0.360093045 | 0.360093045 | 0.40582061 | 0.790054135 | 1 | 0.405820611 | 0.405820611 | 0.405820611 | 0.887320741 | -0.051919367 | FALSE | TRUE | FALSE |
| ALKBH5 | 0.3600636 | 0.78198846 | 1 | 0.360063561 | 0.360063561 | 7.116519488 | 0.38009755 | 0.799273922 | 0.0131 | 0.380097554 | 0.380097554 | 7.523121307 | 0.947292495 | -0.023515903 | FALSE | TRUE | FALSE |
| GRM5 | 0.3599676 | 0.78201621 | 0.69715 | 0.359967565 | 5.756638208 | 12.13756135 | 8.82354969 | 0.213486153 | 0.3658 | 2.205946154 | 35.30207585 | 172.998784 | 0.04079623 | -1.389379966 | FALSE | FALSE | FALSE |
| MED13L | 0.3599409 | 0.78204395 | 0.955 | 38.75184581 | 4095.478593 | 30145.01059 | 0.06420046 | 0.883757313 | 0.99945 | 6005.741684 | 1924813.636 | 21858440.42 | 5.606516762 | 0.748693125 | FALSE | TRUE | FALSE |
| ALG1 | 0.3598913 | 0.78207168 | 0.26175 | 0.090007343 | 0.359891321 | 1.779404755 | 0.03472441 | 0.890170658 | 1 | 0.555104565 | 2.220279182 | 2.748204125 | 10.36421757 | 1.015536521 | FALSE | TRUE | FALSE |
| GATAD2B | 0.359859 | 0.78209941 | 1 | 0.359858969 | 0.359858969 | 7.105283214 | 0.40591123 | 0.78997203 | 1 | 0.405911226 | 0.405911226 | 8.026881744 | 0.886545988 | -0.052298731 | FALSE | TRUE | FALSE |
| CKS2 | 0.3598473 | 0.78212712 | 1 | 0.359847262 | 0.359847262 | 0.359847262 | 0.36469301 | 0.804209604 | 1 | 0.364693011 | 0.364693011 | 0.364693011 | 0.9867128 | -0.005809238 | FALSE | TRUE | FALSE |
| PVALB | 0.3597264 | 0.78215483 | 1 | 0.359726393 | 0.359726393 | 0.359726393 | 0.40573945 | 0.790081488 | 1 | 0.405739454 | 0.405739454 | 0.405739454 | 0.886594561 | -0.052274937 | FALSE | TRUE | FALSE |
| TMCC1 | 0.3596884 | 0.78218254 | 1 | 32.23322987 | 3196.729983 | 3196.729983 | 0.4599592 | 0.770987883 | 1 | 0.459959198 | 0.459959198 | 9.094687614 | 0.782000645 | -0.106792889 | FALSE | TRUE | FALSE |
| RPS2 | 0.3596832 | 0.78221023 | 1 | 54.23892429 | 868.6070652 | 3477.300587 | 0.43327175 | 0.779973376 | 1 | 1.732841602 | 6.932117581 | 6.932117581 | 0.830156191 | -0.080840189 | FALSE | TRUE | FALSE |
| CCNH | 0.359672 | 0.78223792 | 1 | 0.359672035 | 0.359672035 | 0.359672035 | 0.4054361 | 0.790300037 | 1 | 0.405436096 | 0.405436096 | 0.405436096 | 0.887123862 | -0.052015739 | FALSE | TRUE | FALSE |
| ZBTB47 | 0.3596676 | 0.7822656 | 1 | 316.0251627 | 3182.914766 | 32839.87408 | 0.47447602 | 0.765346099 | 1 | 0.474476022 | 0.474476022 | 9.416042112 | 0.758031202 | -0.120312918 | FALSE | TRUE | FALSE |
| ERG | 0.3595499 | 0.78229327 | 1 | 0.359549895 | 0.359549895 | 7.109554959 | 0.40545897 | 0.790272745 | 1 | 0.405458973 | 0.405458973 | 8.028261701 | 0.886772568 | -0.05218775 | FALSE | TRUE | FALSE |
| SNX20 | 0.3595324 | 0.78232094 | 1 | 0.359532428 | 0.359532428 | 0.359532428 | 0.40493065 | 0.790545332 | 1 | 0.404930648 | 0.404930648 | 0.404930648 | 0.887886431 | -0.051642581 | FALSE | TRUE | FALSE |
| HORMAD1 | 0.3594594 | 0.78234859 | 1 | 3.344943314 | 31.94609661 | 31.94609661 | 0.39278765 | 0.794522395 | 0.8753 | 3.660430113 | 343.4476233 | 343.4476233 | 0.915149354 | -0.038508023 | FALSE | TRUE | FALSE |
| PAOX | 0.3594592 | 0.78237624 | 0.53835 | 0.834538094 | 13.33328913 | 53.31454138 | 0.01854931 | 0.89419381 | 1 | 0.07414226 | 0.07414226 | 0.367005935 | 19.37857014 | 1.287321729 | FALSE | TRUE | FALSE |
| TAS2R16 | 0.3594513 | 0.78240389 | 1 | 0.35945134 | 0.35945134 | 0.35945134 | 0.40497211 | 0.790490874 | 1 | 0.404972108 | 0.404972108 | 0.404972108 | 0.887595301 | -0.051785006 | FALSE | TRUE | FALSE |
| TMEM17 | 0.359445 | 0.78243152 | 0.0057 | 0.359445026 | 0.359445026 | 0.359445026 | 0.39296392 | 0.794470036 | 1 | 3.684519945 | 3.684519945 | 3.684519945 | 0.914702353 | -0.038720204 | FALSE | TRUE | FALSE |
| PLRG1 | 0.3594313 | 0.78245915 | 1 | 3.383562418 | 3.383562418 | 7.119235424 | 0.41871678 | 0.785006963 | 1 | 0.418716784 | 0.418716784 | 8.303128969 | 0.858411506 | -0.06630447 | FALSE | TRUE | FALSE |
| OR4D5 | 0.3592986 | 0.78248677 | 1 | 0.359298595 | 0.359298595 | 0.359298595 | 0.40510034 | 0.790436386 | 1 | 0.405100342 | 0.405100342 | 0.405100342 | 0.886937278 | -0.052107091 | FALSE | TRUE | FALSE |
| CENPP | 0.3592281 | 0.78251439 | 1 | 0.359228144 | 0.359228144 | 0.359228144 | 3.52972233 | 0.382262646 | 0.75505 | 3.52972233 | 33.61230333 | 33.61230333 | 0.101772352 | -0.992370188 | FALSE | FALSE | FALSE |
| OR4F6 | 0.3592192 | 0.78254199 | 1 | 4.49518E+18 | 1.92062E+25 | 4.10504E+27 | 0.30413728 | 0.824026337 | 1 | 7.21595E+28 | 1.27576E+36 | 3.71997E+38 | 1.181108818 | 0.072289912 | FALSE | TRUE | FALSE |
| PTGS2 | 0.3591917 | 0.78256959 | 0.0141 | 0.359191685 | 0.359191685 | 7.103702142 | 1.60421599 | 0.525376875 | 0.4999 | 0.401119132 | 1.604215994 | 7.943543054 | 0.223904815 | -0.649936568 | FALSE | FALSE | FALSE |
| OR8H2 | 0.3591523 | 0.78259718 | 1 | 0.359152332 | 0.359152332 | 0.359152332 | 3.6497218 | 0.373997406 | 0.50005 | 3.6497218 | 3.6497218 | 3.6497218 | 0.0984054 | -1.006981071 | FALSE | FALSE | FALSE |
| ARMC8 | 0.3591518 | 0.78262477 | 1 | 0.359151787 | 0.359151787 | 7.098379783 | 0.40542598 | 0.790327321 | 1 | 0.405425983 | 0.405425983 | 8.024318558 | 0.885862776 | -0.052633547 | FALSE | TRUE | FALSE |
| DEFB116 | 0.359096 | 0.78265234 | 1 | 0.359096015 | 0.359096015 | 0.359096015 | 0.40452015 | 0.79081717 | 1 | 0.404520149 | 0.404520149 | 0.404520149 | 0.887708599 | -0.051729573 | FALSE | TRUE | FALSE |
| SLC25A1 | 0.3590335 | 0.78267991 | 0.75135 | 0.359033468 | 1.435895566 | 1.435895566 | 0.11921396 | 0.871423709 | 1 | 0.476680922 | 1.906500779 | 1.906500779 | 3.011673026 | 0.478807819 | FALSE | TRUE | FALSE |
| LSM14A | 0.358853 | 0.78270747 | 1 | 0.358853006 | 0.358853006 | 7.105055812 | 0.4047155 | 0.790626962 | 1 | 0.404715502 | 0.404715502 | 8.022773674 | 0.886679666 | -0.052233251 | FALSE | TRUE | FALSE |
| GGA1 | 0.3588403 | 0.78273503 | 1 | 0.358840342 | 0.358840342 | 7.079250011 | 0.40492527 | 0.790572549 | 0.0168 | 0.404925265 | 0.404925265 | 8.001487266 | 0.886189065 | -0.052473613 | FALSE | TRUE | FALSE |
| NCK2 | 0.3587914 | 0.78276258 | 1 | 0.358791449 | 0.358791449 | 7.083720472 | 0.40493252 | 0.790518107 | 1 | 0.404932525 | 0.404932525 | 8.007044909 | 0.886052434 | -0.052540577 | FALSE | TRUE | FALSE |
| KPNA3 | 0.3587767 | 0.78279012 | 1 | 0.358776678 | 0.358776678 | 7.11177017 | 0.40502844 | 0.790463634 | 1 | 0.405028444 | 0.405028444 | 8.037166153 | 0.885806128 | -0.05266132 | FALSE | TRUE | FALSE |
| RNF150 | 0.3587735 | 0.78281765 | 1 | 3.342914213 | 3.342914213 | 7.095529907 | 0.41746183 | 0.785580172 | 1 | 0.417461833 | 0.417461833 | 8.267285503 | 0.859416313 | -0.065796407 | FALSE | TRUE | FALSE |
| TMEM243 | 0.3586682 | 0.78284518 | 1 | 0.358668237 | 0.358668237 | 0.358668237 | 0.40415497 | 0.790898575 | 1 | 0.404154973 | 0.404154973 | 0.404154973 | 0.887452243 | -0.051855008 | FALSE | TRUE | FALSE |
| DCAF15 | 0.3586509 | 0.78287269 | 1 | 0.358650852 | 0.358650852 | 7.061444744 | 0.40466478 | 0.790681346 | 1 | 0.404664783 | 0.404664783 | 7.982326181 | 0.886291237 | -0.052423545 | FALSE | TRUE | FALSE |
| B3GNT7 | 0.3585769 | 0.78290021 | 0.0155 | 0.358576935 | 0.358576935 | 7.082695351 | 1.60051074 | 0.526095614 | 0.5103 | 1.600510737 | 1.600510737 | 7.915629055 | 0.224039069 | -0.649676241 | FALSE | FALSE | FALSE |
| TIMP3 | 0.3585348 | 0.78292771 | 1 | 0.358534779 | 0.358534779 | 0.358534779 | 1.60125303 | 0.525808401 | 0.006 | 0.400409593 | 1.601253029 | 1.601253029 | 0.223908884 | -0.649928674 | FALSE | FALSE | FALSE |

| | | | | | | | | | | | | | | | | | |
|---|---|---|---|---|---|---|---|---|---|---|---|---|---|---|---|---|---|
| PLA2G5 | 0.3584504 | 0.78295521 | 1 | 1.433662139 | 1.433662139 | 1.433662139 | 0.40887971 | 0.788898567 | 1 | 0.408879713 | 0.408879713 | 0.408879713 | 0.876664643 | -0.057166509 | FALSE | TRUE | FALSE |
| NPY1R | 0.3584135 | 0.7829827 | 1 | 0.358413527 | 0.358413527 | 0.358413527 | 0.4045363 | 0.79079002 | 1 | 0.404536303 | 0.404536303 | 0.404536303 | 0.885986064 | -0.052573109 | FALSE | TRUE | FALSE |
| PEX10 | 0.3584031 | 0.78301018 | 0.2588 | 0.358403081 | 1.43294694 | 1.449918774 | 0.48391703 | 0.76166987 | 0.93855 | 4.519139576 | 424.9995571 | 424.9995571 | 0.740629198 | -0.130399171 | FALSE | TRUE | FALSE |
| PELI1 | 0.3583519 | 0.78303765 | 1 | 0.358351938 | 0.358351938 | 7.105889626 | 0.40440338 | 0.790871446 | 1 | 0.404403378 | 0.404403378 | 8.027288754 | 0.886124988 | -0.052505016 | FALSE | TRUE | FALSE |
| RING1 | 0.3583467 | 0.78306512 | 1 | 0.358346745 | 0.358346745 | 7.091586152 | 0.4046692 | 0.790654158 | 1 | 0.4046692 | 0.4046692 | 8.018423399 | 0.885530072 | -0.052796686 | FALSE | TRUE | FALSE |
| PSMB9 | 0.3582685 | 0.78309258 | 1 | 0.358268514 | 0.358268514 | 0.358268514 | 0.40386729 | 0.791061192 | 1 | 0.403867288 | 0.403867288 | 0.403867288 | 0.887094658 | -0.052030036 | FALSE | TRUE | FALSE |
| MFAP2 | 0.3582638 | 0.78312003 | 1 | 0.358263825 | 0.358263825 | 0.358263825 | 0.40370533 | 0.791115338 | 1 | 0.40370533 | 0.40370533 | 0.40370533 | 0.887438927 | -0.051861525 | FALSE | TRUE | FALSE |
| BATF2 | 0.3582563 | 0.78314748 | 1 | 0.358256251 | 0.358256251 | 0.358256251 | 0.39528149 | 0.793707744 | 1 | 0.39528149 | 0.39528149 | 0.39528149 | 0.906331968 | -0.042712701 | FALSE | TRUE | FALSE |
| METTL13 | 0.3581718 | 0.78317492 | 0.51395 | 0.358171823 | 1.432233327 | 7.053288837 | 0.05844631 | 0.884979599 | 1 | 0.530511982 | 0.530511982 | 1.976354999 | 6.128219693 | 0.787334326 | FALSE | TRUE | FALSE |
| ERAP1 | 0.3580436 | 0.78320235 | 0.9984 | 23604.52008 | 9396258.898 | 254930434.9 | 536.759063 | 0.00420275 | 0.62205 | 5141.19781 | 2020744.2 | 20711055.96 | 0.000667047 | -3.175843498 | FALSE | FALSE | FALSE |
| MRPL23 | 0.358014 | 0.78322977 | 1 | 0.35801403 | 0.35801403 | 0.35801403 | 0.40364352 | 0.791169454 | 0.0059 | 0.403643517 | 0.403643517 | 0.403643517 | 0.886955975 | -0.052097936 | FALSE | TRUE | FALSE |
| ZNF491 | 0.3579199 | 0.78325719 | 0.5057 | 0.357919883 | 0.357919883 | 0.803753538 | 0.50546947 | 0.753561849 | 0.75565 | 0.505469475 | 4.617436724 | 4.617436724 | 0.708093962 | -0.149909109 | FALSE | TRUE | FALSE |
| ANKRD49 | 0.3579148 | 0.7832846 | 1 | 0.357914779 | 0.357914779 | 0.357914779 | 0.40393059 | 0.791007016 | 1 | 0.40393059 | 0.40393059 | 0.40393059 | 0.886079905 | -0.052527112 | FALSE | TRUE | FALSE |
| TOE1 | 0.3579147 | 0.783312 | 0.5136 | 0.35791466 | 0.35791466 | 1.375668 | 0.05859526 | 0.884928774 | 0.49955 | 0.058595261 | 0.520641062 | 1.159906801 | 6.108252639 | 0.785916991 | FALSE | TRUE | FALSE |
| CYTH3 | 0.3578312 | 0.7833394 | 0.0119 | 0.357831189 | 0.357831189 | 7.090579241 | 14.0982919 | 0.162514306 | 0.5035 | 14.09829189 | 135.9430604 | 135.9430604 | 0.025381173 | -1.595488307 | FALSE | FALSE | FALSE |
| PCYT1A | 0.3577455 | 0.78336678 | 0.75205 | 0.357745538 | 1.430910819 | 1.774495528 | 1.06998178 | 0.581001761 | 0.5027 | 1.069981777 | 4.278811251 | 4.278811251 | 0.334347318 | -0.475802156 | FALSE | FALSE | FALSE |
| LSM6 | 0.3574972 | 0.78339417 | 1 | 0.357497168 | 0.357497168 | 0.357497168 | 0.36460938 | 0.804280682 | 1 | 0.36460938 | 0.36460938 | 0.36460938 | 0.980493611 | -0.008555232 | FALSE | TRUE | FALSE |
| DYNLT1 | 0.3573437 | 0.78342154 | 1 | 0.35734371 | 0.35734371 | 0.35734371 | 0.40264714 | 0.791358645 | 1 | 0.402647143 | 0.402647143 | 0.402647143 | 0.88748602 | -0.051838479 | FALSE | TRUE | FALSE |
| RNF114 | 0.3573091 | 0.78344891 | 1 | 0.357309129 | 0.357309129 | 0.357309129 | 0.40368839 | 0.7911424 | 1 | 0.403688391 | 0.403688391 | 0.403688391 | 0.885111232 | -0.053002148 | FALSE | TRUE | FALSE |
| MBLAC2 | 0.3573053 | 0.78347627 | 1 | 0.357305331 | 0.357305331 | 0.357305331 | 0.40321917 | 0.791277603 | 1 | 0.403219165 | 0.403219165 | 0.403219165 | 0.886131815 | -0.052501671 | FALSE | TRUE | FALSE |
| ANAPC7 | 0.3571137 | 0.78350363 | 0.01585 | 0.357113706 | 0.357113706 | 7.061903782 | 0.40329537 | 0.791223543 | 0.0127 | 0.403295368 | 0.403295368 | 7.985940247 | 0.885489233 | -0.052816715 | FALSE | TRUE | FALSE |
| CDC5L | 0.3571009 | 0.78353097 | 0.02125 | 0.357100933 | 0.357100933 | 7.038701051 | 0.40349262 | 0.791196502 | 1 | 0.403492625 | 0.403492625 | 7.966959506 | 0.885024685 | -0.053044616 | FALSE | TRUE | FALSE |
| PCDHB5 | 0.3570785 | 0.78355831 | 0.5075 | 0.357078523 | 3.240698825 | 6.989873694 | 6.18885012 | 0.263148543 | 0.1306 | 0.67461498 | 6.188850119 | 13.23949226 | 0.057697071 | -1.238846235 | FALSE | FALSE | FALSE |
| MTG1 | 0.3569115 | 0.78358565 | 1 | 0.356911485 | 0.356911485 | 0.356911485 | 3.53457176 | 0.381795806 | 0.75365 | 3.534571762 | 33.87710365 | 33.87710365 | 0.100977292 | -0.99577628 | FALSE | FALSE | FALSE |
| ELMO1 | 0.3569095 | 0.78361297 | 0.0223 | 0.356909469 | 0.356909469 | 7.020762201 | 0.35540696 | 0.807357026 | 1 | 31.85289975 | 3159.317826 | 3159.317826 | 1.004227575 | 0.001832142 | FALSE | TRUE | FALSE |
| GOLGA7B | 0.3569021 | 0.78364029 | 1 | 0.356902144 | 0.356902144 | 0.356902144 | 0.40263558 | 0.791385644 | 1 | 0.402635585 | 0.402635585 | 0.402635585 | 0.886414807 | -0.052362998 | FALSE | TRUE | FALSE |
| TAB2 | 0.356861 | 0.7836676 | 1 | 0.356861047 | 0.356861047 | 7.04922227 | 0.40325977 | 0.791250577 | 1 | 0.40325977 | 0.40325977 | 7.977572631 | 0.884940858 | -0.053085753 | FALSE | TRUE | FALSE |
| C3orf38 | 0.356824 | 0.78369491 | 1 | 0.356823968 | 0.356823968 | 0.356823968 | 0.40253844 | 0.791412637 | 1 | 0.402538441 | 0.402538441 | 0.402538441 | 0.886434516 | -0.052353342 | FALSE | TRUE | FALSE |
| UHMK1 | 0.3564622 | 0.7837222 | 1 | 3.338533651 | 3.338533651 | 7.064244605 | 0.41537809 | 0.786377363 | 1 | 0.415378088 | 0.415378088 | 8.240846267 | 0.858163225 | -0.0664301 | FALSE | TRUE | FALSE |
| EIF3I | 0.3564013 | 0.7837495 | 1 | 3.351989498 | 3.351989498 | 3.351989498 | 3.79114541 | 0.366987001 | 0.50195 | 3.791145414 | 3.791145414 | 3.791145414 | 0.094008859 | -1.026831216 | FALSE | FALSE | FALSE |
| RRAGC | 0.3562397 | 0.78377678 | 1 | 0.356239712 | 0.356239712 | 0.356239712 | 0.39798721 | 0.792779932 | 1 | 1.591715522 | 1.591715522 | 1.591715522 | 0.895103413 | -0.048126787 | FALSE | TRUE | FALSE |
| PRPF38A | 0.3561985 | 0.78380406 | 1 | 0.356198509 | 0.356198509 | 7.067268978 | 0.4024033 | 0.7914666 | 1 | 0.402403299 | 0.402403299 | 7.991651763 | 0.885177905 | -0.052969435 | FALSE | TRUE | FALSE |
| SLC25A11 | 0.3560965 | 0.78383133 | 1 | 0.356096459 | 0.356096459 | 7.058276941 | 1.5917819 | 0.527241101 | 0.5019 | 1.591781903 | 1.591781903 | 7.897705916 | 0.223709328 | -0.650315907 | FALSE | FALSE | FALSE |
| CDH11 | 0.3560446 | 0.7838586 | 0.50805 | 0.356044617 | 0.356044617 | 6.992591434 | 0.11989191 | 0.87124686 | 1 | 0.119891912 | 0.119891912 | 2.359743278 | 2.969713382 | 0.472714536 | FALSE | TRUE | FALSE |
| CCDC134 | 0.3560165 | 0.78388585 | 1 | 3.311214506 | 3.311214506 | 3.311214506 | 0.41447382 | 0.786801893 | 1 | 0.414473825 | 0.414473825 | 0.414473825 | 0.858960107 | -0.066027006 | FALSE | TRUE | FALSE |
| METTL1 | 0.3560109 | 0.7839131 | 1 | 0.356010944 | 0.356010944 | 0.356010944 | 0.40213371 | 0.791547492 | 1 | 0.402133706 | 0.402133706 | 0.402133706 | 0.885304909 | -0.052907128 | FALSE | TRUE | FALSE |
| MAPK8IP1 | 0.3559057 | 0.78394034 | 1 | 0.355905731 | 0.355905731 | 7.011691599 | 0.38959418 | 0.795822386 | 1 | 3.645475832 | 3.645475832 | 7.744161133 | 0.913529372 | -0.039277485 | FALSE | TRUE | FALSE |
| SPATA2L | 0.3557557 | 0.78396758 | 0.01295 | 0.355755681 | 0.355755681 | 7.041397459 | 0.38911408 | 0.79597724 | 1 | 3.623624646 | 3.623624646 | 7.711251882 | 0.914270902 | -0.038925102 | FALSE | TRUE | FALSE |
| TMEM101 | 0.355721 | 0.78399481 | 0.00775 | 0.355721049 | 0.355721049 | 0.355721049 | 0.40126171 | 0.791789787 | 1 | 0.401261707 | 0.401261707 | 0.401261707 | 0.886506346 | -0.052318151 | FALSE | TRUE | FALSE |
| NSMCE4A | 0.3557079 | 0.78402203 | 1 | 0.355707874 | 0.355707874 | 0.355707874 | 0.38916157 | 0.795951447 | 1 | 3.622238896 | 3.622238896 | 3.622238896 | 0.914036493 | -0.039036465 | FALSE | TRUE | FALSE |
| COX14 | 0.3555962 | 0.78404925 | 1 | 0.355596208 | 0.355596208 | 0.355596208 | 0.40140104 | 0.791762894 | 1 | 0.401401043 | 0.401401043 | 0.401401043 | 0.885887604 | -0.052621375 | FALSE | TRUE | FALSE |
| TRAF1 | 0.3555157 | 0.78407645 | 1 | 0.355515683 | 0.355515683 | 7.043609668 | 0.40140278 | 0.791735994 | 0.0108 | 0.401402785 | 0.401402785 | 7.96171583 | 0.885683151 | -0.052721617 | FALSE | TRUE | FALSE |
| BLVRB | 0.3554983 | 0.78410365 | 1 | 3.303121817 | 3.303121817 | 3.303121817 | 3.7372558 | 0.369177232 | 0.0059 | 3.737255805 | 3.737255805 | 3.737255805 | 0.095122823 | -1.021715268 | FALSE | FALSE | FALSE |
| SNIP1 | 0.3554423 | 0.78413085 | 1 | 0.355442341 | 0.355442341 | 7.023213264 | 0.40155733 | 0.79168217 | 1 | 0.401557327 | 0.401557327 | 7.945902758 | 0.885159645 | -0.052978394 | FALSE | TRUE | FALSE |
| RCN2 | 0.3553727 | 0.78415803 | 1 | 1246.667494 | 129393.4603 | 1372108.105 | 0.49510904 | 0.756761852 | 1 | 0.495109038 | 0.495109038 | 0.495109038 | 0.717766541 | -0.14401679 | FALSE | TRUE | FALSE |
| OTUD7A | 0.3553517 | 0.78418521 | 1 | 0.355351733 | 0.355351733 | 6.965246968 | 0.39437803 | 0.793997564 | 1 | 0.394378025 | 0.394378025 | 7.748774278 | 0.901043442 | -0.04525427 | FALSE | TRUE | FALSE |
| CHGA | 0.3553258 | 0.78421238 | 1 | 0.355325824 | 0.355325824 | 7.046895158 | 0.40110271 | 0.791816674 | 0.00945 | 0.401102707 | 0.401102707 | 0.401102707 | 0.885872417 | -0.052628821 | FALSE | TRUE | FALSE |
| RFT1 | 0.3552916 | 0.78423955 | 0.777 | 1.42096103 | 22.74584488 | 91.03841306 | 0.66596937 | 0.682660864 | 0.50735 | 0.665969366 | 2.663793088 | 10.6574966 | 0.533495388 | -0.272869331 | FALSE | TRUE | FALSE |
| GLCCI1 | 0.354942 | 0.78426671 | 1 | 0.354942035 | 0.354942035 | 7.036601851 | 1.48967169 | 0.537051465 | 0.87825 | 13.86950935 | 132.5319059 | 132.5319059 | 0.238268631 | -0.62293313 | FALSE | FALSE | FALSE |
| ELF5 | 0.3549349 | 0.78429386 | 1 | 3.296877918 | 3.296877918 | 3.296877918 | 0.4132687 | 0.78719655 | 1 | 0.413268702 | 0.413268702 | 0.413268702 | 0.858847676 | -0.066083855 | FALSE | TRUE | FALSE |
| SPG21 | 0.3548743 | 0.784321 | 1 | 0.354874324 | 0.354874324 | 0.354874324 | 0.4006803 | 0.792031503 | 1 | 0.400680299 | 0.400680299 | 0.400680299 | 0.885679493 | -0.052723411 | FALSE | TRUE | FALSE |
| ASPHD1 | 0.3548548 | 0.78434814 | 1 | 0.354854809 | 0.354854809 | 7.021306494 | 0.40093203 | 0.791950997 | 1 | 0.40093203 | 0.40093203 | 7.943201063 | 0.885074731 | -0.053020058 | FALSE | TRUE | FALSE |
| TIGIT | 0.3548287 | 0.78437527 | 0.00705 | 0.354828652 | 0.354828652 | 0.354828652 | 0.40047745 | 0.792058326 | 1 | 0.400477451 | 0.400477451 | 0.400477451 | 0.88601406 | -0.052559387 | FALSE | TRUE | FALSE |
| ZNF821 | 0.354824 | 0.7844024 | 1 | 0.354824044 | 0.354824044 | 7.024827596 | 0.4007521 | 0.792004675 | 1 | 0.400752095 | 0.400752095 | 7.943750464 | 0.885395357 | -0.05286276 | FALSE | TRUE | FALSE |
| FAM32A | 0.3548054 | 0.78442951 | 1 | 0.354805369 | 0.354805369 | 0.354805369 | 0.36708157 | 0.803280729 | 1 | 0.36708157 | 0.36708157 | 0.36708157 | 0.966557293 | -0.014772398 | FALSE | TRUE | FALSE |
| ESAM | 0.3547931 | 0.78445662 | 1 | 0.354793145 | 0.354793145 | 7.035519621 | 0.40085514 | 0.79197784 | 1 | 0.400855135 | 0.400855135 | 0.400855135 | 0.885090682 | -0.053012231 | FALSE | TRUE | FALSE |
| WDTC1 | 0.3545443 | 0.78448372 | 1 | 0.354544322 | 0.354544322 | 6.970745969 | 0.40097291 | 0.79189729 | 1 | 0.400972907 | 0.400972907 | 7.899665515 | 0.884210168 | -0.053444495 | FALSE | TRUE | FALSE |
| KLHL17 | 0.3544716 | 0.78451082 | 0.51055 | 0.354471607 | 1.417630259 | 6.965044812 | 0.01066186 | 0.896590034 | 0.5108 | 0.042613094 | 0.042613094 | 0.361054965 | 33.246689 | 1.521748401 | FALSE | TRUE | FALSE |
| ARL2 | 0.3542289 | 0.78453791 | 1 | 1.416622972 | 5.666738433 | 5.666738433 | 0.40956679 | 0.788593733 | 1 | 0.409566793 | 0.409566793 | 0.409566793 | 0.864886694 | -0.063040784 | FALSE | TRUE | FALSE |
| GPR63 | 0.3542211 | 0.78456499 | 1 | 1.416760212 | 5.667960879 | 7.028481709 | 0.40938308 | 0.78867696 | 1 | 0.409383082 | 0.409383082 | 0.409383082 | 0.865255818 | -0.062855472 | FALSE | TRUE | FALSE |
| KCNJ6 | 0.3541219 | 0.78459207 | 1 | 3.347738384 | 3.347738384 | 7.019210165 | 3.78862022 | 0.367230869 | 0.01195 | 3.788620219 | 3.788620219 | 7.947656847 | 0.093469897 | -1.029328234 | FALSE | FALSE | FALSE |
| PRSS55 | 0.3539965 | 0.78461913 | 0.13605 | 0.088523494 | 0.353996494 | 0.353996494 | 0.03925341 | 0.889125468 | 0.7513 | 0.348040729 | 5.565212261 | 5.565212261 | 9.018235766 | 0.955121585 | FALSE | TRUE | FALSE |
| NKG7 | 0.3539672 | 0.7846462 | 1 | 0.35396724 | 0.35396724 | 0.35396724 | 3.620514 | 0.377328823 | 0.508 | 3.620514002 | 3.620514002 | 3.620514002 | 0.097767124 | -1.009807162 | FALSE | FALSE | FALSE |
| AGTRAP | 0.3539582 | 0.78467325 | 0.49885 | 0.038598891 | 0.353958204 | 0.353958204 | 0.04369895 | 0.888158863 | 0.75255 | 0.401983359 | 3.795232484 | 3.795232484 | 8.09992401 | 0.908480945 | FALSE | TRUE | FALSE |
| GDF9 | 0.3539202 | 0.7847003 | 0.50295 | 0.353920212 | 0.353920212 | 0.777159755 | 0.05714683 | 0.885249989 | 1 | 0.519154753 | 0.519154753 | 1.134627325 | 6.193173417 | 0.791913241 | FALSE | TRUE | FALSE |
| NODAL | 0.3538915 | 0.78472734 | 0.0095 | 0.353891529 | 0.353891529 | 0.353891529 | 1.56468933 | 0.529798982 | 0.7475 | 1.564689331 | 6.259425211 | 6.259425211 | 0.22617367 | -0.645557954 | FALSE | FALSE | FALSE |
| TYW3 | 0.3538338 | 0.78475437 | 1 | 0.353833815 | 0.353833815 | 0.353833815 | 0.39954578 | 0.792219126 | 1 | 0.39954578 | 0.39954578 | 0.39954578 | 0.885590169 | -0.052767213 | FALSE | TRUE | FALSE |
| CASC3 | 0.3537637 | 0.7847814 | 1 | 0.353763666 | 0.353763666 | 6.962389192 | 0.39076825 | 0.795408251 | 1 | 0.390768252 | 0.390768252 | 7.705443316 | 0.905302987 | -0.043206047 | FALSE | TRUE | FALSE |
| TMEM52 | 0.3536441 | 0.78480842 | 0.75205 | 3.269805011 | 31.02916008 | 31.02916008 | 0.41159741 | 0.787813677 | 1 | 3.820874111 | 3.820874111 | 3.820874111 | 0.859199088 | -0.065906192 | FALSE | TRUE | FALSE |
| NPHS1 | 0.3535837 | 0.78483543 | 0.5143 | 0.353583708 | 11.90920367 | 30.02737211 | 0.00277921 | 0.898741023 | 1 | 0.024342782 | 0.024342782 | 0.472954289 | 127.2243608 | 2.104570278 | FALSE | TRUE | FALSE |
| ACHE | 0.3534229 | 0.78486244 | 0.0223 | 0.353422857 | 0.353422857 | 6.949101035 | 0.39985465 | 0.792138757 | 1 | 0.399854651 | 0.399854651 | 7.878038626 | 0.883878319 | -0.053607519 | FALSE | TRUE | FALSE |
| PSG6 | 0.3533474 | 0.78488944 | 0.2584 | 0.353347352 | 3.170270197 | 7.006718102 | 39.2500995 | 0.069296538 | 0.19485 | 9.309993903 | 372.7586905 | 3626.665039 | 0.009002457 | -2.04563892 | FALSE | FALSE | FALSE |
| RNF113B | 0.3532943 | 0.78491643 | 1 | 3.289852487 | 3.289852487 | 6.993876483 | 0.41178851 | 0.787701749 | 1 | 0.411788512 | 0.411788512 | 8.161830962 | 0.857950928 | -0.066537552 | FALSE | TRUE | FALSE |

| | | | | | | | | | | | | | | | | | |
|---|---|---|---|---|---|---|---|---|---|---|---|---|---|---|---|---|---|
| NCOA4 | 0.3532275 | 0.78494341 | 0.512 | 0.353227547 | 0.353227547 | 0.770845407 | 4.03630983 | 0.354827889 | 0.2511 | 0.431649678 | 4.036309831 | 4.036309831 | 0.087512496 | -1.05792993 | FALSE | FALSE | FALSE |
| PHACTR1 | 0.3532252 | 0.78497039 | 1 | 0.353225193 | 0.353225193 | 6.973433529 | 0.39925249 | 0.792299434 | 1 | 0.399252495 | 0.399252495 | 7.89433943 | 0.884716308 | -0.053195967 | FALSE | TRUE | FALSE |
| SCARB2 | 0.3531835 | 0.78499736 | 0.7565 | 0.810938428 | 3.242726186 | 3.242726186 | 355.276444 | 0.00663301 | 0.25985 | 36.71394485 | 3524.01798 | 14095.60385 | 0.000994109 | -3.002566029 | FALSE | FALSE | FALSE |
| HTR2A | 0.3530988 | 0.78502433 | 1 | 0.353098779 | 0.353098779 | 6.994867036 | 0.3991794 | 0.792352936 | 1 | 0.399179401 | 0.399179401 | 7.916760205 | 0.884561625 | -0.053271906 | FALSE | TRUE | FALSE |
| LCN15 | 0.3530981 | 0.78505128 | 0.00585 | 0.353098116 | 0.353098116 | 0.353098116 | 0.39847806 | 0.792593346 | 1 | 0.398478063 | 0.398478063 | 0.398478063 | 0.886116824 | -0.052509018 | FALSE | TRUE | FALSE |
| ATF1 | 0.3530601 | 0.78507823 | 1 | 0.353060127 | 0.353060127 | 0.353060127 | 0.39921687 | 0.792326188 | 1 | 0.399216872 | 0.399216872 | 0.399216872 | 0.884381776 | -0.053360215 | FALSE | TRUE | FALSE |
| NCOA5 | 0.353026 | 0.78510518 | 1 | 0.353026041 | 0.353026041 | 6.959488379 | 0.39288638 | 0.794496229 | 1 | 0.392886379 | 0.392886379 | 7.758626191 | 0.898544873 | -0.046460229 | FALSE | TRUE | FALSE |
| SUDS3 | 0.35294 | 0.78513211 | 0.0085 | 0.352940027 | 0.352940027 | 0.352940027 | 3.42107809 | 0.387822222 | 0.5057 | 3.421078086 | 3.421078086 | 3.421078086 | 0.103166317 | -0.986462073 | FALSE | FALSE | FALSE |
| MYT1L | 0.3529227 | 0.78515904 | 1 | 0.352922701 | 0.352922701 | 6.879372219 | 0.39933866 | 0.792245903 | 0.0287 | 0.399338662 | 0.399338662 | 7.8079391 | 0.883767927 | -0.053661763 | FALSE | TRUE | FALSE |
| KCTD9 | 0.3526866 | 0.78518596 | 1 | 0.352686649 | 0.352686649 | 7.003071468 | 0.39878033 | 0.792486568 | 1 | 0.39878033 | 0.39878033 | 0.39878033 | 0.884413353 | -0.053344709 | FALSE | TRUE | FALSE |
| IL17REL | 0.3526712 | 0.78521288 | 1 | 0.352671171 | 0.352671171 | 6.997238732 | 3.58961667 | 0.37827388 | 0.0087 | 3.589616674 | 3.589616674 | 3.589616674 | 0.098247585 | -1.007678115 | FALSE | FALSE | FALSE |
| TSKS | 0.3526676 | 0.78523979 | 1 | 0.352667571 | 0.352667571 | 6.94269518 | 122.472541 | 0.023323585 | 0.0026 | 3.617192706 | 3.617192706 | 71.34336409 | 0.002879564 | -2.540673201 | FALSE | FALSE | FALSE |
| UBQLN4 | 0.3526549 | 0.78526669 | 1 | 0.352654931 | 0.352654931 | 6.96169705 | 0.39900062 | 0.792379676 | 1 | 0.399000623 | 0.399000623 | 7.888881643 | 0.883845564 | -0.053623614 | FALSE | TRUE | FALSE |
| CATSPERD | 0.3525931 | 0.78529358 | 0.5179 | 0.352593129 | 0.352593129 | 6.926263967 | 0.05791694 | 0.885072675 | 0.5107 | 0.513369473 | 0.513369473 | 1.959213315 | 6.087910339 | 0.784468248 | FALSE | TRUE | FALSE |
| GAPDH | 0.3525331 | 0.78532047 | 0.00755 | 0.352533128 | 0.352533128 | 0.352533128 | 0.39853956 | 0.792539972 | 1 | 0.39853956 | 0.39853956 | 0.39853956 | 0.884562447 | -0.053271502 | FALSE | TRUE | FALSE |
| DNAJB6 | 0.3525163 | 0.78534735 | 1 | 0.352516343 | 0.352516343 | 0.352516343 | 0.39855102 | 0.792513274 | 1 | 0.398551022 | 0.398551022 | 0.398551022 | 0.884494893 | -0.05330467 | FALSE | TRUE | FALSE |
| EVI5L | 0.3524326 | 0.78537422 | 1 | 0.352432642 | 0.352432642 | 6.927727091 | 0.39882948 | 0.792459856 | 1 | 0.398829479 | 0.398829479 | 7.855936802 | 0.883667483 | -0.053711126 | FALSE | TRUE | FALSE |
| COX4I1 | 0.35226 | 0.78540109 | 1 | 0.352259955 | 0.352259955 | 0.352259955 | 0.39824431 | 0.792673354 | 1 | 0.398244309 | 0.398244309 | 0.398244309 | 0.884532301 | -0.053286303 | FALSE | TRUE | FALSE |
| STRAP | 0.3521823 | 0.78542795 | 1 | 0.352182297 | 0.352182297 | 0.352182297 | 0.39845335 | 0.792620022 | 0.0076 | 0.398453351 | 0.398453351 | 0.398453351 | 0.883873347 | -0.053609962 | FALSE | TRUE | FALSE |
| ZC3H18 | 0.3521418 | 0.7854548 | 1 | 0.352141751 | 0.352141751 | 6.876208636 | 0.39850876 | 0.792566663 | 1 | 0.398508758 | 0.398508758 | 7.803791109 | 0.883648713 | -0.053720351 | FALSE | TRUE | FALSE |
| IRX2 | 0.3521268 | 0.78548165 | 1 | 1.408280175 | 1.408280175 | 6.960859318 | 0.39048921 | 0.795511947 | 1 | 3.683472117 | 3.683472117 | 7.729979339 | 0.901758186 | -0.044909907 | FALSE | TRUE | FALSE |
| SLC35D3 | 0.3520979 | 0.78550848 | 1 | 0.352097878 | 0.352097878 | 6.965772433 | 0.38290958 | 0.798119967 | 1 | 0.382909579 | 0.382909579 | 7.585198925 | 0.919532696 | -0.036432824 | FALSE | TRUE | FALSE |
| OR5A2 | 0.3520595 | 0.78553532 | 1 | 0.352059505 | 0.352059505 | 0.352059505 | 31.9971579 | 0.087028758 | 0.5063 | 31.99715785 | 312.4221298 | 312.4221298 | 0.011002837 | -1.958495329 | FALSE | FALSE | FALSE |
| FRS2 | 0.3520336 | 0.78556214 | 1 | 1.407875702 | 1.407875702 | 6.969613885 | 0.40298528 | 0.791304624 | 0.014 | 0.402985284 | 0.402985284 | 7.988048001 | 0.873564523 | -0.058705012 | FALSE | TRUE | FALSE |
| PROX1 | 0.3519017 | 0.78558896 | 1 | 0.35190168 | 0.35190168 | 6.924888154 | 0.37957545 | 0.799348706 | 1 | 0.379575452 | 0.379575452 | 7.483920805 | 0.927092828 | -0.032876778 | FALSE | TRUE | FALSE |
| UBL4B | 0.3518898 | 0.78561577 | 1 | 0.351889756 | 0.351889756 | 0.351889756 | 0.3976515 | 0.79283318 | 1 | 0.397651498 | 0.397651498 | 0.397651498 | 0.884919981 | -0.053095998 | FALSE | TRUE | FALSE |
| KRT14 | 0.3516603 | 0.78564257 | 0.0148 | 0.087976034 | 0.35166026 | 1.73771478 | 0.10332951 | 0.875114878 | 0.99915 | 7036.64755 | 4206550.127 | 46166778.41 | 3.403289887 | 0.531898943 | FALSE | TRUE | FALSE |
| CFL2 | 0.3516289 | 0.78566937 | 1 | 0.351628941 | 0.351628941 | 0.351628941 | 0.39742209 | 0.792939593 | 1 | 0.397422088 | 0.397422088 | 0.397422088 | 0.884774529 | -0.053167388 | FALSE | TRUE | FALSE |
| LASP1 | 0.3514659 | 0.78569616 | 0.0091 | 0.351465916 | 0.351465916 | 0.351465916 | 0.39747018 | 0.792913 | 1 | 0.397470181 | 0.397470181 | 0.397470181 | 0.884257316 | -0.053421338 | FALSE | TRUE | FALSE |
| ZNF563 | 0.3513804 | 0.78572295 | 0.75375 | 0.351380377 | 3.192276305 | 6.961907631 | 0.52892894 | 0.743576343 | 0.4977 | 0.059232041 | 0.528928937 | 1.174885452 | 0.664324358 | -0.177619823 | FALSE | TRUE | FALSE |
| TMEM259 | 0.3513489 | 0.78574973 | 1 | 0.351348907 | 0.351348907 | 6.956855294 | 0.38501553 | 0.797639368 | 1 | 3.60076005 | 3.60076005 | 7.632619604 | 0.912557742 | -0.039739646 | FALSE | TRUE | FALSE |
| NHLRC3 | 0.3513273 | 0.7857765 | 0.8138 | 1.404609318 | 52.37875398 | 52.37875398 | 2.15473068 | 0.475523144 | 0.25205 | 0.538917424 | 2.154730676 | 2.154730676 | 0.163049287 | -0.787681095 | FALSE | FALSE | FALSE |
| PLVAP | 0.3513153 | 0.78580326 | 1 | 3.270098099 | 3.270098099 | 6.942486822 | 0.40976923 | 0.788482656 | 1 | 0.409769227 | 0.409769227 | 8.109240659 | 0.857349262 | -0.066842222 | FALSE | TRUE | FALSE |
| HHAT | 0.351262 | 0.78583002 | 0.50345 | 0.351262005 | 0.351262005 | 0.775087628 | 0.50916166 | 0.752025803 | 0.5013 | 0.509161661 | 2.036086787 | 2.036086787 | 0.689883062 | -0.161224518 | FALSE | TRUE | FALSE |
| WNT16 | 0.3511922 | 0.78585677 | 0.8775 | 1.404452212 | 5.617947341 | 6.964935833 | 4.18375187 | 0.3453574 | 0.5065 | 4.183751875 | 39.95337586 | 39.95337586 | 0.083941917 | -1.076021118 | FALSE | FALSE | FALSE |
| AGO3 | 0.3510587 | 0.78588351 | 1 | 0.351058669 | 0.351058669 | 6.894832807 | 1.572063 | 0.529232689 | 0.5084 | 1.572063 | 1.572063 | 30.9423935 | 0.223310814 | -0.651090244 | FALSE | FALSE | FALSE |
| GAST | 0.3510511 | 0.78591025 | 0.0034 | 0.351051088 | 0.351051088 | 0.351051088 | 0.36724101 | 0.803256791 | 1 | 0.367241012 | 0.367241012 | 0.367241012 | 0.955914717 | -0.019580852 | FALSE | TRUE | FALSE |
| TLDC2 | 0.3507755 | 0.78593698 | 1 | 0.350775488 | 0.350775488 | 0.350775488 | 0.39644216 | 0.79323167 | 0.00545 | 0.396442158 | 0.396442158 | 0.396442158 | 0.884808744 | -0.053150594 | FALSE | TRUE | FALSE |
| LMOD3 | 0.3507236 | 0.7859637 | 0.75745 | 0.3507236 | 3.228589401 | 6.938746433 | 0.48337878 | 0.761812115 | 0.9366 | 4.470869545 | 412.4523819 | 412.4523819 | 0.725566817 | -0.139322587 | FALSE | TRUE | FALSE |
| C10orf88 | 0.3506461 | 0.78599042 | 1 | 0.350646123 | 0.350646123 | 6.958521742 | 0.39644964 | 0.79317863 | 1 | 0.396449643 | 0.396449643 | 0.396449643 | 0.884465732 | -0.053318989 | FALSE | TRUE | FALSE |
| GTPBP4 | 0.3505981 | 0.78601713 | 0.50295 | 1.402259481 | 1.402259481 | 11.85416368 | 0.40142327 | 0.791709086 | 1 | 0.401423272 | 0.401423272 | 7.93226321 | 0.873387468 | -0.058793044 | FALSE | TRUE | FALSE |
| RPA3 | 0.3505515 | 0.78604383 | 1 | 0.350551475 | 0.350551475 | 0.350551475 | 0.39596998 | 0.793443544 | 1 | 0.395969975 | 0.395969975 | 0.395969975 | 0.88529812 | -0.052910458 | FALSE | TRUE | FALSE |
| C1orf162 | 0.3504868 | 0.78607053 | 1 | 0.350486808 | 0.350486808 | 0.350486808 | 0.3961802 | 0.793337664 | 1 | 0.396180197 | 0.396180197 | 0.396180197 | 0.884665137 | -0.053221087 | FALSE | TRUE | FALSE |
| TBC1D22A | 0.3504728 | 0.78609722 | 0.0162 | 0.35047279 | 0.35047279 | 6.916152037 | 0.39655933 | 0.793125561 | 1 | 0.396559332 | 0.396559332 | 7.838148847 | 0.883783992 | -0.053653869 | FALSE | TRUE | FALSE |
| PXT1 | 0.3504698 | 0.7861239 | 1 | 0.350469769 | 0.350469769 | 0.350469769 | 0.39574851 | 0.793628556 | 0.0018 | 0.395748507 | 0.395748507 | 0.395748507 | 0.885587089 | -0.052768724 | FALSE | TRUE | FALSE |
| ERGIC2 | 0.3503209 | 0.78615057 | 1 | 3.269990408 | 3.269990408 | 3.269990408 | 0.40888224 | 0.788870893 | 1 | 0.408882236 | 0.408882236 | 0.408882236 | 0.856776923 | -0.06713224 | FALSE | TRUE | FALSE |
| ADIPOR1 | 0.3502294 | 0.78617724 | 1 | 0.350229438 | 0.350229438 | 0.416201229 | 0.39661175 | 0.793072462 | 1 | 0.396611747 | 0.396611747 | 7.875205825 | 0.883053618 | -0.054012926 | FALSE | TRUE | FALSE |
| ZNF516 | 0.3502233 | 0.7862039 | 1 | 0.350223325 | 0.350223325 | 6.778597245 | 0.39657289 | 0.793099015 | 1 | 0.396572886 | 0.396572886 | 7.705549412 | 0.883124733 | -0.053977952 | FALSE | TRUE | FALSE |
| MGAT3 | 0.3501613 | 0.78623056 | 1 | 0.350161345 | 0.350161345 | 6.879817217 | 0.37219268 | 0.80176086 | 1 | 3.482279187 | 33.43846379 | 68.56346123 | 0.940806635 | -0.026499629 | FALSE | TRUE | FALSE |
| KANSL2 | 0.3499959 | 0.78625721 | 1 | 0.349995853 | 0.349995853 | 6.935655594 | 0.39610623 | 0.793364145 | 1 | 0.396106233 | 0.396106233 | 7.858064195 | 0.883590876 | -0.053748777 | FALSE | TRUE | FALSE |
| ICAM2 | 0.3499472 | 0.78628385 | 1 | 0.349947188 | 0.349947188 | 0.349947188 | 3.56170591 | 0.380391157 | 0.50395 | 3.561705908 | 3.561705908 | 3.561705908 | 0.09825269 | -1.007655549 | FALSE | FALSE | FALSE |
| ZBTB8A | 0.3498965 | 0.78631048 | 1 | 0.349896487 | 0.349896487 | 6.937943611 | 0.39590013 | 0.793496441 | 1 | 0.395900135 | 0.395900135 | 7.858222729 | 0.883799867 | -0.053646068 | FALSE | TRUE | FALSE |
| C18orf32 | 0.3498916 | 0.78633711 | 1 | 0.349891582 | 0.349891582 | 0.349891582 | 0.358192 | 0.806273095 | 1 | 0.358191999 | 0.358191999 | 0.358191999 | 0.976826905 | -0.010182387 | FALSE | TRUE | FALSE |
| VEGFC | 0.3498235 | 0.78636373 | 1 | 0.349823546 | 0.349823546 | 6.932179405 | 0.39582153 | 0.793549309 | 1 | 0.395821535 | 0.395821535 | 7.852354739 | 0.88379109 | -0.053650381 | FALSE | TRUE | FALSE |
| CYP26B1 | 0.3496591 | 0.78639035 | 1 | 0.349659084 | 0.349659084 | 6.885252683 | 0.39609059 | 0.793390618 | 1 | 0.396090594 | 0.396090594 | 7.814040058 | 0.88277553 | -0.054149714 | FALSE | TRUE | FALSE |
| KDM4D | 0.3495881 | 0.78641696 | 1 | 0.349588058 | 0.349588058 | 6.911387372 | 0.39577616 | 0.793575732 | 1 | 0.395776163 | 0.395776163 | 7.835351939 | 0.883297406 | -0.053893045 | FALSE | TRUE | FALSE |
| AZI2 | 0.3495755 | 0.78644356 | 0.008 | 0.349575464 | 0.349575464 | 0.349575464 | 0.39525011 | 0.793760501 | 1 | 0.395250107 | 0.395250107 | 0.395250107 | 0.884441162 | -0.053331054 | FALSE | TRUE | FALSE |
| VASP | 0.3495167 | 0.78647015 | 1 | 0.349516745 | 0.349516745 | 6.930596037 | 0.39576974 | 0.793602148 | 1 | 0.395769743 | 0.395769743 | 0.395769743 | 0.883131545 | -0.053974602 | FALSE | TRUE | FALSE |
| C1orf35 | 0.3495028 | 0.78649674 | 1 | 0.34950276 | 0.34950276 | 0.34950276 | 0.3951175 | 0.793786868 | 1 | 0.395117501 | 0.395117501 | 0.395117501 | 0.884553985 | -0.053275657 | FALSE | TRUE | FALSE |
| ICA1L | 0.3494373 | 0.78652332 | 0.5074 | 0.349437329 | 0.349437329 | 0.760288517 | 2.03372503 | 0.486556292 | 0.25945 | 0.508667446 | 2.033725033 | 2.033725033 | 0.171821325 | -0.764922937 | FALSE | FALSE | FALSE |
| TMEM179B | 0.3493623 | 0.78654989 | 1 | 0.349362345 | 0.349362345 | 0.349362345 | 0.39487216 | 0.79386593 | 1 | 0.39487216 | 0.39487216 | 0.39487216 | 0.884747976 | -0.053180422 | FALSE | TRUE | FALSE |
| GPR4 | 0.3492388 | 0.78657646 | 1 | 0.349238756 | 0.349238756 | 6.903886741 | 0.39525165 | 0.793734126 | 1 | 0.39525165 | 0.39525165 | 7.824373897 | 0.883585826 | -0.053751259 | FALSE | TRUE | FALSE |
| CDC40 | 0.3491571 | 0.78660302 | 1 | 0.349157084 | 0.349157084 | 6.911393962 | 0.39101811 | 0.795278477 | 1 | 1.563916748 | 1.563916748 | 7.749578666 | 0.892943518 | -0.049176011 | FALSE | TRUE | FALSE |
| COX7A2L | 0.3490982 | 0.78662957 | 1 | 0.349098204 | 0.349098204 | 0.349098204 | 0.39463576 | 0.79394493 | 1 | 0.394635762 | 0.394635762 | 0.394635762 | 0.884608637 | -0.053248825 | FALSE | TRUE | FALSE |
| OSR2 | 0.3490434 | 0.78665612 | 1 | 0.349043412 | 0.349043412 | 0.349043412 | 0.39184245 | 0.794940266 | 1 | 0.391842454 | 0.391842454 | 0.391842454 | 0.890774873 | -0.050232042 | FALSE | TRUE | FALSE |
| SRF | 0.3490059 | 0.78668266 | 1 | 0.349005875 | 0.349005875 | 6.912568839 | 0.39549622 | 0.793654959 | 1 | 0.395496224 | 0.395496224 | 7.842496068 | 0.882450587 | -0.054309604 | FALSE | TRUE | FALSE |
| SCAMP5 | 0.3489767 | 0.78670919 | 1 | 0.348976652 | 0.348976652 | 0.348976652 | 0.3689306 | 0.802584057 | 1 | 0.368930598 | 0.368930598 | 0.368930598 | 0.945914094 | -0.024148304 | FALSE | TRUE | FALSE |
| GGH | 0.348968 | 0.78673572 | 1 | 0.348967984 | 0.348967984 | 0.348967984 | 0.39474937 | 0.793918603 | 1 | 0.394749371 | 0.394749371 | 0.394749371 | 0.884024168 | -0.053535862 | FALSE | TRUE | FALSE |
| SOST | 0.3488057 | 0.78676224 | 1 | 0.348805746 | 0.348805746 | 0.348805746 | 0.39509193 | 0.793813229 | 1 | 0.395091933 | 0.395091933 | 0.395091933 | 0.882847047 | -0.054114532 | FALSE | TRUE | FALSE |
| ACBD3 | 0.3487755 | 0.78678875 | 1 | 0.348775459 | 0.348775459 | 6.916059045 | 0.39476935 | 0.79389227 | 1 | 0.394769351 | 0.394769351 | 7.836120893 | 0.883491734 | -0.053797509 | FALSE | TRUE | FALSE |
| CCDC71L | 0.3487415 | 0.78681526 | 0.93795 | 32.01859631 | 3253.153979 | 3253.153979 | 0.38396905 | 0.79791788 | 1 | 3.640172226 | 35.41844386 | 35.41844386 | 0.908254072 | -0.041792646 | FALSE | TRUE | FALSE |
| LIN7C | 0.3487057 | 0.78684175 | 1 | 0.348705656 | 0.348705656 | 0.348705656 | 0.39426372 | 0.79405017 | 1 | 0.394263719 | 0.394263719 | 0.394263719 | 0.884447742 | -0.053327822 | FALSE | TRUE | FALSE |
| C1QTNF2 | 0.3484694 | 0.78686825 | 0.0121 | 0.348469439 | 0.348469439 | 0.748414108 | 0.05729463 | 0.885216254 | 1 | 0.05729463 | 0.05729463 | 0.05729463 | 6.082061119 | 0.78405078 | FALSE | TRUE | FALSE |

| | | | | | | | | | | | | | | | | | |
|---|---|---|---|---|---|---|---|---|---|---|---|---|---|---|---|---|---|
| PCGF6 | 0.3483855 | 0.78689473 | 0.0101 | 0.348385455 | 0.348385455 | 6.922756296 | 0.39436105 | 0.79402387 | 1 | 0.394361051 | 0.394361051 | 0.394361051 | 0.883417504 | -0.053834 | FALSE | TRUE | FALSE |
| LCN8 | 0.3479526 | 0.78692122 | 1 | 0.347952556 | 0.347952556 | 0.347952556 | 0.39339482 | 0.794339015 | 1 | 0.393394817 | 0.393394817 | 0.393394817 | 0.884486884 | -0.053308603 | FALSE | TRUE | FALSE |
| SFPQ | 0.3479357 | 0.78694769 | 1 | 0.347935698 | 0.347935698 | 6.871282046 | 0.39442793 | 0.793971251 | 1 | 0.394427926 | 0.394427926 | 7.801219965 | 0.882127442 | -0.054468667 | FALSE | TRUE | FALSE |
| ICAM1 | 0.3478987 | 0.78697416 | 1 | 0.347898721 | 0.347898721 | 6.865238968 | 3.564995 | 0.380156531 | 0.50825 | 3.564994996 | 3.564994996 | 7.545975171 | 0.097587436 | -1.010606093 | FALSE | FALSE | FALSE |
| CAMLG | 0.3477617 | 0.78700062 | 1 | 0.347761732 | 0.347761732 | 0.347761732 | 0.39340824 | 0.794312792 | 1 | 0.393408237 | 0.393408237 | 0.393408237 | 0.883971658 | -0.053561659 | FALSE | TRUE | FALSE |
| PTGES3 | 0.3476489 | 0.78702708 | 1 | 0.34764892 | 0.34764892 | 0.34764892 | 0.35646161 | 0.806874262 | 1 | 0.356461609 | 0.356461609 | 0.356461609 | 0.975277314 | -0.010871878 | FALSE | TRUE | FALSE |
| TERF1 | 0.3476377 | 0.78705353 | 1 | 0.347637743 | 0.347637743 | 6.890885964 | 0.39347172 | 0.794286561 | 1 | 0.393471716 | 0.393471716 | 7.807755806 | 0.883513934 | -0.053786597 | FALSE | TRUE | FALSE |
| CORO6 | 0.3475648 | 0.78707997 | 0.5054 | 0.347564751 | 0.347564751 | 1.715671995 | 0.113341 | 0.872679634 | 1 | 1.036630693 | 1.036630693 | 2.240935614 | 3.066540325 | 0.48664868 | FALSE | TRUE | FALSE |
| PLEKHG7 | 0.3475441 | 0.7871064 | 0.93805 | 3.10904662 | 268.3363742 | 268.3363742 | 38.4312622 | 0.070964331 | 0.76885 | 364.0273806 | 35096.77358 | 35096.77358 | 0.009043266 | -2.043674702 | FALSE | FALSE | FALSE |
| PHC3 | 0.3474384 | 0.78713283 | 1 | 0.347438405 | 0.347438405 | 6.820501923 | 0.39382836 | 0.794129028 | 1 | 0.393828359 | 0.393828359 | 7.748358669 | 0.882207685 | -0.054429163 | FALSE | TRUE | FALSE |
| MDP1 | 0.3474203 | 0.78715925 | 1 | 0.347420283 | 0.347420283 | 0.347420283 | 0.39375389 | 0.794181566 | 1 | 0.39375389 | 0.39375389 | 0.39375389 | 0.88232851 | -0.054369688 | FALSE | TRUE | FALSE |
| ONECUT2 | 0.3473611 | 0.78718567 | 1 | 0.347361091 | 0.347361091 | 6.847913436 | 0.39380131 | 0.794155301 | 1 | 0.393801315 | 0.393801315 | 7.776797038 | 0.882071943 | -0.054495992 | FALSE | TRUE | FALSE |
| EAF1 | 0.3472815 | 0.78721207 | 1 | 0.34728155 | 0.34728155 | 0.34728155 | 0.38101959 | 0.798924188 | 1 | 3.549394128 | 3.549394128 | 3.549394128 | 0.911453269 | -0.040265593 | FALSE | TRUE | FALSE |
| CYB5R4 | 0.3472236 | 0.78723848 | 0.8762 | 1.388661242 | 5.555100654 | 6.882014284 | 0.11840295 | 0.871631178 | 0.50715 | 0.473440792 | 0.473440792 | 2.349299657 | 2.932558544 | 0.467246691 | FALSE | TRUE | FALSE |
| LRRC4B | 0.3472122 | 0.78726487 | 1 | 0.347212229 | 0.347212229 | 6.78306116 | 0.39366089 | 0.794234077 | 1 | 0.393660886 | 0.393660886 | 7.711978104 | 0.882008453 | -0.054527253 | FALSE | TRUE | FALSE |
| PLSCR1 | 0.3472112 | 0.78729126 | 1 | 3.235223281 | 3.235223281 | 3.235223281 | 0.40555011 | 0.790218138 | 1 | 0.405550112 | 0.405550112 | 0.405550112 | 0.856148778 | -0.067450759 | FALSE | TRUE | FALSE |
| KPNB1 | 0.3470386 | 0.78731764 | 1 | 0.34703865 | 0.34703865 | 6.831162847 | 0.3934792 | 0.794260322 | 1 | 0.393479196 | 0.393479196 | 7.760029275 | 0.881974582 | -0.054543931 | FALSE | TRUE | FALSE |
| PHLDB1 | 0.3470185 | 0.78734401 | 0.52415 | 0.347018471 | 0.347018471 | 6.698873399 | 0.05736619 | 0.885199379 | 1 | 0.057366186 | 0.057366186 | 1.932000451 | 6.049181471 | 0.781696613 | FALSE | TRUE | FALSE |
| ARPC5 | 0.346992 | 0.78737038 | 1 | 4782493278 | 1.01909E+14 | 1.67438E+16 | 0.271018 | 0.833571228 | 1 | 1.39836E+17 | 4.93866E+21 | 1.02982E+24 | 1.280328152 | 0.107321295 | FALSE | TRUE | FALSE |
| DUT | 0.3469009 | 0.78739674 | 1 | 0.346900895 | 0.346900895 | 0.346900895 | 0.38067558 | 0.799124186 | 1 | 0.380675584 | 3.554564721 | 3.554564721 | 0.911276975 | -0.040349603 | FALSE | TRUE | FALSE |
| SPRYD4 | 0.3466742 | 0.78742309 | 0.0061 | 0.346674191 | 0.346674191 | 0.346674191 | 0.3923614 | 0.794757665 | 1 | 0.392361399 | 0.392361399 | 0.392361399 | 0.883558351 | -0.053764764 | FALSE | TRUE | FALSE |
| ASAP3 | 0.3466659 | 0.78744944 | 0.5158 | 0.346665927 | 0.346665927 | 6.798151623 | 0.05821482 | 0.885004996 | 0.02365 | 0.058214817 | 0.058214817 | 1.144278251 | 5.95494319 | 0.774877623 | FALSE | TRUE | FALSE |
| SLAIN2 | 0.3464392 | 0.78747578 | 0.0142 | 0.346439207 | 0.346439207 | 6.86512578 | 13.2940168 | 0.168178105 | 0.00005 | 0.392489117 | 0.392489117 | 7.786256988 | 0.026059784 | -1.584029189 | FALSE | FALSE | FALSE |
| NDUFS3 | 0.3464143 | 0.78750212 | 0.75555 | 0.346414308 | 3.29267818 | 3.29267818 | 8.2572856 | 0.223570325 | 0.13105 | 0.888220073 | 8.257285599 | 8.257285599 | 0.041952565 | -1.377241485 | FALSE | FALSE | FALSE |
| ZNF483 | 0.3464028 | 0.78752845 | 0.0196 | 0.3464028 | 0.3464028 | 6.840212894 | 0.39255979 | 0.79462703 | 1 | 0.392559786 | 0.392559786 | 7.763474327 | 0.882420493 | -0.054324415 | FALSE | TRUE | FALSE |
| KRTAP3-3 | 0.3463371 | 0.78755477 | 0.5014 | 3.196379817 | 3.196379817 | 3.196379817 | 0.40394933 | 0.790979917 | 1 | 0.403949331 | 0.403949331 | 0.403949331 | 0.857377627 | -0.066827854 | FALSE | TRUE | FALSE |
| GOT2 | 0.3463256 | 0.78758108 | 0.26405 | 0.346325556 | 1.384859157 | 2.936053011 | 0.11702406 | 0.871951514 | 1 | 0.467862739 | 0.467862739 | 2.319780665 | 2.959438909 | 0.471209379 | FALSE | TRUE | FALSE |
| EMCN | 0.346308 | 0.78760739 | 1 | 0.346307964 | 0.346307964 | 0.346307964 | 0.39180436 | 0.795018417 | 1 | 0.391804358 | 0.391804358 | 0.391804358 | 0.883879818 | -0.053606782 | FALSE | TRUE | FALSE |
| MACROD2 | 0.3462912 | 0.78763369 | 0.01085 | 0.346291168 | 0.346291168 | 6.871071433 | 0.39202146 | 0.79486205 | 1 | 0.392021465 | 0.392021465 | 0.392021465 | 0.883347466 | -0.053868432 | FALSE | TRUE | FALSE |
| LMO1 | 0.3462635 | 0.78765999 | 1 | 0.346263541 | 0.346263541 | 0.346263541 | 0.39268087 | 0.794574727 | 1 | 0.392680866 | 0.392680866 | 0.392680866 | 0.88179377 | -0.054632974 | FALSE | TRUE | FALSE |
| ALKBH4 | 0.3462584 | 0.78768627 | 1 | 0.346258425 | 0.346258425 | 6.859333485 | 0.39252609 | 0.794679305 | 1 | 0.39252609 | 0.39252609 | 0.39252609 | 0.882128435 | -0.054468178 | FALSE | TRUE | FALSE |
| B3GNT5 | 0.3461987 | 0.78771255 | 0.0104 | 0.346198673 | 0.346198673 | 6.874952714 | 0.39215696 | 0.794809871 | 1 | 0.392156962 | 0.392156962 | 0.392156962 | 0.882806394 | -0.05413453 | FALSE | TRUE | FALSE |
| RAPGEF6 | 0.3461854 | 0.78773883 | 0.93855 | 3.215872598 | 299.2640512 | 595.7342379 | 0.47335558 | 0.76572438 | 0.98395 | 168.80403 | 16617.97599 | 81060.31907 | 0.731343248 | -0.135878744 | FALSE | TRUE | FALSE |
| FAM181A | 0.3461074 | 0.78776509 | 0.01005 | 0.346107393 | 0.346107393 | 6.86895744 | 0.39223331 | 0.794783772 | 0.0085 | 0.392233314 | 0.392233314 | 0.392233314 | 0.882401829 | -0.0543336 | FALSE | TRUE | FALSE |
| IFNA16 | 0.3460502 | 0.78779135 | 1 | 0.346050223 | 0.346050223 | 0.346050223 | 0.39144416 | 0.795122523 | 1 | 0.391444161 | 0.391444161 | 0.391444161 | 0.884034702 | -0.053530687 | FALSE | TRUE | FALSE |
| OR2K2 | 0.3460326 | 0.7878176 | 1 | 0.346032564 | 0.346032564 | 0.346032564 | 0.39184095 | 0.794966323 | 1 | 0.39184095 | 0.39184095 | 0.39184095 | 0.883094438 | -0.05399285 | FALSE | TRUE | FALSE |
| VAMP5 | 0.3459873 | 0.78784385 | 0.0033 | 0.345987266 | 0.345987266 | 0.345987266 | 0.36806911 | 0.802872956 | 1 | 0.368069106 | 0.368069106 | 0.368069106 | 0.940006268 | -0.026869251 | FALSE | TRUE | FALSE |
| OR52N5 | 0.3459711 | 0.78787009 | 1 | 3.211477305 | 30.59578881 | 30.59578881 | 0.40473532 | 0.79059976 | 1 | 3.770505703 | 3.770505703 | 3.770505703 | 0.854808175 | -0.068131333 | FALSE | TRUE | FALSE |
| CIDEC | 0.3457417 | 0.78789632 | 0.49725 | 0.345741684 | 1.382789687 | 1.382789687 | 3.47948196 | 0.384126486 | 0.25635 | 3.479481958 | 33.38710446 | 33.38710446 | 0.09936585 | -1.002762846 | FALSE | FALSE | FALSE |
| MED29 | 0.3456738 | 0.78792255 | 1 | 0.345673792 | 0.345673792 | 0.345673792 | 0.37928498 | 0.799423438 | 1 | 3.517517906 | 3.517517906 | 3.517517906 | 0.911382759 | -0.040299192 | FALSE | TRUE | FALSE |
| DTX1 | 0.3456525 | 0.78794877 | 1 | 3.242862323 | 31.22510352 | 63.79490679 | 0.41726921 | 0.785637319 | 1 | 0.417269206 | 0.417269206 | 8.224875515 | 0.828368059 | -0.081776655 | FALSE | TRUE | FALSE |
| UBXN2B | 0.3455774 | 0.78797498 | 1 | 0.345577379 | 0.345577379 | 0.345577379 | 1.54881928 | 0.530927984 | 0.5072 | 1.548819279 | 1.548819279 | 1.548819279 | 0.223123113 | -0.651455439 | FALSE | FALSE | FALSE |
| CPNE6 | 0.3455555 | 0.78800119 | 1 | 0.345555485 | 0.345555485 | 6.81219207 | 0.39168541 | 0.795044454 | 1 | 0.391685408 | 0.391685408 | 7.73488651 | 0.882227109 | -0.054419602 | FALSE | TRUE | FALSE |
| CLIC6 | 0.3453693 | 0.78802739 | 0.5117 | 0.796689313 | 3.185493052 | 3.185493052 | 9.9088413 | 0.199298033 | 0.25325 | 1.044153583 | 9.908841298 | 9.908841298 | 0.034854657 | -1.45773919 | FALSE | FALSE | FALSE |
| HCFC2 | 0.3452976 | 0.78805358 | 1 | 0.34529758 | 0.34529758 | 6.806880718 | 0.39159404 | 0.795070484 | 0.01525 | 0.391594045 | 0.391594045 | 7.73285492 | 0.88177434 | -0.054642543 | FALSE | TRUE | FALSE |
| OR56A1 | 0.3451975 | 0.78807977 | 1 | 0.345197488 | 0.345197488 | 0.345197488 | 0.39080096 | 0.795356362 | 1 | 0.390800961 | 0.390800961 | 0.390800961 | 0.883307676 | -0.053887995 | FALSE | TRUE | FALSE |
| ZNF207 | 0.3450919 | 0.78810595 | 1 | 0.345091928 | 0.345091928 | 6.837280394 | 0.38022824 | 0.799199082 | 1 | 0.380228238 | 0.380228238 | 7.541909074 | 0.907591531 | -0.042109566 | FALSE | TRUE | FALSE |
| IL20 | 0.3450522 | 0.78813213 | 1 | 0.34505221 | 0.34505221 | 0.34505221 | 0.37863837 | 0.799771413 | 1 | 3.511208717 | 3.511208717 | 3.511208717 | 0.911297528 | -0.040339808 | FALSE | TRUE | FALSE |
| DCAF6 | 0.3450262 | 0.78815829 | 0.2657 | 0.345026217 | 1.379376533 | 6.782787229 | 0.51551571 | 0.749209366 | 0.51545 | 0.515515709 | 0.515515709 | 10.15521636 | 0.669283614 | -0.174389807 | FALSE | TRUE | FALSE |
| PSME1 | 0.3450072 | 0.78818445 | 1 | 0.345007179 | 0.345007179 | 0.345007179 | 0.39056555 | 0.795486033 | 1 | 0.390565549 | 0.390565549 | 0.390565549 | 0.883352821 | -0.0538658 | FALSE | TRUE | FALSE |
| ZNF23 | 0.3449646 | 0.78821061 | 0.8742 | 3.147589007 | 29.45804897 | 29.45804897 | 0.05989792 | 0.884742087 | 1 | 0.537413143 | 0.537413143 | 1.186974196 | 5.759208732 | 0.760362819 | FALSE | TRUE | FALSE |
| SMARCD2 | 0.3448471 | 0.78823676 | 1 | 0.344847125 | 0.344847125 | 6.822769841 | 0.39089304 | 0.795330407 | 1 | 0.390893036 | 0.390893036 | 7.743789754 | 0.882203297 | -0.054431324 | FALSE | TRUE | FALSE |
| LENG8 | 0.3448346 | 0.7882629 | 1 | 0.344834603 | 0.344834603 | 6.746580938 | 0.39113491 | 0.795252501 | 0.02555 | 0.391134909 | 0.391134909 | 7.672495704 | 0.881625737 | -0.05471574 | FALSE | TRUE | FALSE |
| JAM3 | 0.3447969 | 0.78828903 | 1 | 0.344796877 | 0.344796877 | 6.844959172 | 0.39073951 | 0.795434185 | 1 | 0.390739513 | 0.390739513 | 0.390739513 | 0.882421322 | -0.054324007 | FALSE | TRUE | FALSE |
| TPSAB1 | 0.3446514 | 0.78831516 | 0.75295 | 0.7853564 | 3.140610128 | 3.140610128 | 0.11960696 | 0.87130933 | 1 | 0.119606963 | 0.119606963 | 2.374594843 | 2.88153316 | 0.459623622 | FALSE | TRUE | FALSE |
| ADAM10 | 0.3446 | 0.78834128 | 1 | 0.344599986 | 0.344599986 | 6.799727665 | 0.39099031 | 0.795304445 | 1 | 0.39099031 | 0.39099031 | 7.727542378 | 0.88135173 | -0.054850738 | FALSE | TRUE | FALSE |
| SLC7A2 | 0.3445399 | 0.78836739 | 0.715 | 2.478521851 | 158.6223561 | 1408.0147 | 8659.87157 | 0.000217276 | 0.072 | 58.99583999 | 8659.871568 | 60554.42063 | 3.97858E-05 | -4.400271941 | FALSE | FALSE | FALSE |
| FBXO33 | 0.3445092 | 0.7883935 | 1 | 3.237739866 | 31.22977765 | 64.10268465 | 0.39932695 | 0.792272672 | 1 | 0.399326954 | 0.399326954 | 7.915688015 | 0.862724593 | -0.064127822 | FALSE | TRUE | FALSE |
| SNRNP48 | 0.3444101 | 0.7884196 | 1 | 0.344410113 | 0.344410113 | 0.409400138 | 0.39007537 | 0.795641418 | 1 | 0.390075373 | 0.390075373 | 0.390075373 | 0.882932216 | -0.054072637 | FALSE | TRUE | FALSE |
| NTM | 0.3440981 | 0.7884457 | 1 | 0.34409813 | 0.34409813 | 6.824780797 | 0.38999922 | 0.795693157 | 1 | 0.389999215 | 0.389999215 | 7.742904251 | 0.882304673 | -0.054381421 | FALSE | TRUE | FALSE |
| HINT1 | 0.3440443 | 0.78847179 | 1 | 0.344044349 | 0.344044349 | 0.344044349 | 1.5434969 | 0.531209587 | 0.5064 | 1.543496895 | 1.543496895 | 1.543496895 | 0.222899282 | -0.651891331 | FALSE | FALSE | FALSE |
| SHOC2 | 0.3440315 | 0.78849787 | 1 | 0.344031498 | 0.344031498 | 6.8137962 | 0.38615603 | 0.797232854 | 1 | 1.544307192 | 1.544307192 | 7.657039219 | 0.890913186 | -0.050164613 | FALSE | TRUE | FALSE |
| CLK3 | 0.3439355 | 0.78852395 | 1 | 0.343935485 | 0.343935485 | 6.788699003 | 3.53310429 | 0.382029312 | 0.01505 | 3.533104291 | 3.533104291 | 7.525697581 | 0.097346542 | -1.011679471 | FALSE | FALSE | FALSE |
| XKR7 | 0.3439068 | 0.78855002 | 0.0174 | 0.343906759 | 0.343906759 | 6.776327909 | 0.39026832 | 0.795537855 | 1 | 0.390268324 | 0.390268324 | 7.703534136 | 0.881205924 | -0.054922592 | FALSE | TRUE | FALSE |
| VCPIP1 | 0.3438865 | 0.78857608 | 1 | 0.34388653 | 0.34388653 | 6.717495974 | 0.39025704 | 0.795563755 | 0.0287 | 0.390257042 | 0.390257042 | 7.644697887 | 0.881179565 | -0.054935583 | FALSE | TRUE | FALSE |
| PSMD9 | 0.3437302 | 0.78860214 | 1 | 0.3437302 | 0.3437302 | 0.3437302 | 0.37745095 | 0.800093424 | 0.5027 | 3.506548373 | 3.506548373 | 3.506548373 | 0.910661903 | -0.040642832 | FALSE | TRUE | FALSE |
| PLEKHO2 | 0.3436812 | 0.78862819 | 1 | 0.343681209 | 0.343681209 | 6.803123116 | 0.38985941 | 0.795770714 | 1 | 0.389859413 | 0.389859413 | 7.726736036 | 0.88155165 | -0.054752238 | FALSE | TRUE | FALSE |
| IL17C | 0.3436286 | 0.78865423 | 0.0067 | 0.343628584 | 0.343628584 | 0.343628584 | 0.38951435 | 0.795899842 | 0.00765 | 0.389514354 | 0.389514354 | 0.389514354 | 0.882197487 | -0.054434184 | FALSE | TRUE | FALSE |
| MRPS12 | 0.343579 | 0.78868027 | 1 | 0.343578972 | 0.343578972 | 0.343578972 | 0.37744275 | 0.80011815 | 1 | 0.377442746 | 3.519879982 | 3.519879982 | 0.910281033 | -0.040824506 | FALSE | TRUE | FALSE |
| CRTC1 | 0.3435135 | 0.7887063 | 0.02025 | 0.343513488 | 0.343513488 | 6.76359863 | 0.38952348 | 0.79587403 | 1 | 0.389523477 | 0.389523477 | 7.683844566 | 0.881881344 | -0.054589845 | FALSE | TRUE | FALSE |
| EMX2 | 0.3433585 | 0.78873232 | 1 | 0.343358475 | 0.343358475 | 0.343358475 | 1.54124389 | 0.531490937 | 0.4993 | 0.385399407 | 1.541243889 | 1.541243889 | 0.222780105 | -0.652123597 | FALSE | FALSE | FALSE |
| TSPYL1 | 0.3432681 | 0.78875834 | 1 | 0.343268067 | 0.343268067 | 6.785111255 | 0.38907586 | 0.796003026 | 1 | 0.389075857 | 0.389075857 | 7.701373682 | 0.882265144 | -0.054400878 | FALSE | TRUE | FALSE |
| THEM6 | 0.3432618 | 0.78878435 | 0.5016 | 3.174990715 | 3.174990715 | 3.174990715 | 0.4051209 | 0.790409131 | 1 | 0.405120903 | 0.405120903 | 0.405120903 | 0.847307075 | -0.071959167 | FALSE | TRUE | FALSE |

| | | | | | | | | | | | | | | | | | |
|---|---|---|---|---|---|---|---|---|---|---|---|---|---|---|---|---|---|
| DRAM2 | 0.3431269 | 0.78881036 | 1 | 0.343126859 | 0.343126859 | 0.343126859 | 0.38891031 | 0.796054577 | 1 | 0.38891031 | 0.38891031 | 0.38891031 | 0.882277611 | -0.054394742 | FALSE | TRUE | FALSE |
| SNRK | 0.3431192 | 0.78883636 | 1 | 0.343119198 | 0.343119198 | 6.751062699 | 0.38957653 | 0.795848211 | 1 | 0.38957653 | 0.38957653 | 7.680105136 | 0.88074915 | -0.055147767 | FALSE | TRUE | FALSE |
| HSPB1 | 0.3430985 | 0.78886235 | 0.7525 | 0.343098546 | 1.372172178 | 1.372172178 | 0.11574843 | 0.872229296 | 1 | 0.462827296 | 0.462827296 | 0.462827296 | 2.96417459 | 0.47190378 | FALSE | TRUE | FALSE |
| APOBEC2 | 0.3429679 | 0.78888833 | 1 | 0.342967857 | 0.342967857 | 0.342967857 | 0.38870411 | 0.796157599 | 1 | 0.388704111 | 0.388704111 | 0.388704111 | 0.882336584 | -0.054365714 | FALSE | TRUE | FALSE |
| GPR61 | 0.3428704 | 0.78891431 | 0.01245 | 0.342870383 | 0.342870383 | 6.785873996 | 3.41844108 | 0.388739254 | 0.7536 | 3.418441075 | 32.8259025 | 32.8259025 | 0.100300218 | -0.998698125 | FALSE | FALSE | FALSE |
| COLGALT2 | 0.3428551 | 0.78894028 | 0.2636 | 0.085742085 | 0.342855084 | 1.692300375 | 2.57189087 | 0.442336338 | 0.31785 | 2.571890875 | 240.2816943 | 240.2816943 | 0.133308566 | -0.875141944 | FALSE | FALSE | FALSE |
| NIPSNAP3B | 0.3428319 | 0.78896625 | 0.0072 | 0.342831883 | 0.342831883 | 0.342831883 | 0.38862245 | 0.796234792 | 1 | 0.388622446 | 0.388622446 | 0.388622446 | 0.88217211 | -0.054446677 | FALSE | TRUE | FALSE |
| DNAJA1 | 0.3427645 | 0.78899221 | 1 | 0.342764473 | 0.342764473 | 6.804984772 | 0.38880206 | 0.796131854 | 1 | 0.388802063 | 0.388802063 | 0.388802063 | 0.88159119 | -0.054732758 | FALSE | TRUE | FALSE |
| HTN3 | 0.3427432 | 0.78901816 | 1 | 0.34274325 | 0.34274325 | 0.34274325 | 0.38798015 | 0.796414677 | 1 | 0.387980153 | 0.387980153 | 0.387980153 | 0.883404079 | -0.0538406 | FALSE | TRUE | FALSE |
| E2F6 | 0.3426673 | 0.7890441 | 1 | 0.34266729 | 0.34266729 | 0.34266729 | 0.38887508 | 0.796080343 | 1 | 0.388875079 | 0.388875079 | 0.388875079 | 0.88117575 | -0.054937463 | FALSE | TRUE | FALSE |
| RAB27A | 0.3426264 | 0.78907004 | 0.74735 | 0.342626436 | 1.370458376 | 1.370458376 | 1.78451904 | 0.509668297 | 0.3207 | 1.784519044 | 28.57323608 | 28.57323608 | 0.191999316 | -0.716700319 | FALSE | FALSE | FALSE |
| DNAJC17 | 0.3425242 | 0.78909597 | 1 | 0.342524205 | 0.342524205 | 0.407107819 | 0.38813414 | 0.796388998 | 1 | 0.388134139 | 0.388134139 | 0.388134139 | 0.88248925 | -0.054290576 | FALSE | TRUE | FALSE |
| GTF3C6 | 0.3422996 | 0.7891219 | 1 | 0.342299649 | 0.342299649 | 0.342299649 | 1.53368799 | 0.532612764 | 0.5056 | 1.53368799 | 1.53368799 | 1.53368799 | 0.223187279 | -0.651330562 | FALSE | FALSE | FALSE |
| MLPH | 0.3422978 | 0.78914782 | 0.01825 | 0.342297776 | 0.342297776 | 1.294986892 | 0.05753642 | 0.885157173 | 1 | 0.057536422 | 0.057536422 | 1.137152652 | 5.949236352 | 0.774461223 | FALSE | TRUE | FALSE |
| TRIB1 | 0.3421702 | 0.78917373 | 0.00895 | 0.342170237 | 0.342170237 | 0.342170237 | 3.56132082 | 0.380625552 | 0.4979 | 0.376480644 | 3.561320817 | 3.561320817 | 0.096079588 | -1.017368868 | FALSE | FALSE | FALSE |
| S1PR4 | 0.342117 | 0.78919964 | 1 | 0.34211699 | 0.34211699 | 6.767609793 | 0.38839953 | 0.796286221 | 1 | 0.388399529 | 0.388399529 | 7.693245833 | 0.880837809 | -0.055104052 | FALSE | TRUE | FALSE |
| TMEM139 | 0.3420447 | 0.78922554 | 1 | 0.342044734 | 0.342044734 | 0.342044734 | 0.38752058 | 0.796542973 | 1 | 0.387520579 | 0.387520579 | 0.387520579 | 0.882649212 | -0.054211862 | FALSE | TRUE | FALSE |
| SLC10A2 | 0.3420183 | 0.78925144 | 0.9689 | 12.29882341 | 268.3825963 | 1073.682179 | 0.11703533 | 0.871941204 | 1 | 4.237657089 | 157.5223376 | 157.5223376 | 2.922350855 | 0.465732356 | FALSE | TRUE | FALSE |
| LARP4B | 0.3420064 | 0.78927732 | 1 | 3.2026101 | 30.77947748 | 62.98084944 | 0.38818194 | 0.796363314 | 1 | 3.641018791 | 35.05068603 | 71.74654412 | 0.881046732 | -0.055001055 | FALSE | TRUE | FALSE |
| UBN1 | 0.3419307 | 0.7893032 | 0.02975 | 0.341930667 | 0.341930667 | 6.683553746 | 0.38835964 | 0.796311925 | 1 | 0.388359638 | 0.388359638 | 7.611814349 | 0.880448514 | -0.055296035 | FALSE | TRUE | FALSE |
| ZNF827 | 0.3417614 | 0.78932908 | 0.03 | 0.341761393 | 0.341761393 | 6.681866208 | 0.38548267 | 0.797461727 | 1 | 0.385482671 | 0.385482671 | 7.557043669 | 0.886580433 | -0.052281858 | FALSE | TRUE | FALSE |
| TARDBP | 0.3415043 | 0.78935495 | 1 | 0.341504335 | 0.341504335 | 6.773569753 | 0.37584113 | 0.800488314 | 1 | 3.551721677 | 3.551721677 | 7.462041621 | 0.90864013 | -0.041608087 | FALSE | TRUE | FALSE |
| CCL27 | 0.3415027 | 0.78938081 | 0.00325 | 0.34150267 | 0.34150267 | 0.34150267 | 0.38688623 | 0.796824631 | 0.0025 | 0.386886234 | 0.386886234 | 0.386886234 | 0.882695325 | -0.054189174 | FALSE | TRUE | FALSE |
| ASH2L | 0.3413782 | 0.78940667 | 1 | 0.34137816 | 0.34137816 | 6.759452193 | 0.38776612 | 0.796440349 | 1 | 0.387766123 | 0.387766123 | 7.687169678 | 0.880371285 | -0.055334131 | FALSE | TRUE | FALSE |
| CA11 | 0.3413148 | 0.78943252 | 0.0118 | 0.34131476 | 0.34131476 | 6.772109347 | 0.387345 | 0.796645487 | 1 | 0.387345004 | 0.387345004 | 7.692743087 | 0.881164741 | -0.054942889 | FALSE | TRUE | FALSE |
| HOXC12 | 0.3413131 | 0.78945836 | 0.75195 | 0.341313146 | 3.245639944 | 3.245639944 | 0.9042649 | 0.597926245 | 0.25385 | 0.226160227 | 0.904264904 | 0.904264904 | 0.377448184 | -0.42314266 | FALSE | TRUE | FALSE |
| SNX10 | 0.3412365 | 0.7894842 | 1 | 0.341236548 | 0.341236548 | 0.341236548 | 0.38691311 | 0.796773481 | 1 | 0.386913113 | 0.386913113 | 0.386913113 | 0.881946197 | -0.054557908 | FALSE | TRUE | FALSE |
| FAM163B | 0.3411873 | 0.78951003 | 0.0062 | 0.34118731 | 0.34118731 | 0.34118731 | 0.34640921 | 0.810113707 | 1 | 0.346409214 | 0.346409214 | 0.346409214 | 0.984925619 | -0.006596566 | FALSE | TRUE | FALSE |
| CCL14 | 0.3410832 | 0.78953585 | 1 | 0.341083188 | 0.341083188 | 0.341083188 | 0.38634335 | 0.797079975 | 1 | 0.386343354 | 0.386343354 | 0.386343354 | 0.88284989 | -0.054113133 | FALSE | TRUE | FALSE |
| BNIP3 | 0.3410541 | 0.78956167 | 1 | 0.341054121 | 0.341054121 | 0.341054121 | 0.38697217 | 0.796747895 | 1 | 0.386972172 | 0.386972172 | 0.386972172 | 0.881340172 | -0.054856434 | FALSE | TRUE | FALSE |
| NUP35 | 0.3410481 | 0.78958748 | 0.0076 | 0.34104812 | 0.34104812 | 0.34104812 | 3.48531586 | 0.383661465 | 0.0069 | 3.485315862 | 3.485315862 | 3.485315862 | 0.097852858 | -1.009426483 | FALSE | FALSE | FALSE |
| EPGN | 0.3409983 | 0.78961329 | 1 | 1.363945633 | 1.363945633 | 1.363945633 | 32.3780109 | 0.085592286 | 0.2467 | 3.404396052 | 32.37801087 | 32.37801087 | 0.010531785 | -1.977498013 | FALSE | FALSE | FALSE |
| SREK1 | 0.3409443 | 0.78963908 | 1 | 0.340944276 | 0.340944276 | 6.744695081 | 0.38690395 | 0.796799059 | 0.0136 | 0.386903954 | 0.386903954 | 7.663905865 | 0.881211661 | -0.054919765 | FALSE | TRUE | FALSE |
| OR6N1 | 0.3409397 | 0.78966487 | 1 | 3.162368057 | 30.10500635 | 30.10500635 | 34.3239309 | 0.081022903 | 0.2529 | 3.595710635 | 34.3239309 | 34.3239309 | 0.009933002 | -2.002919495 | FALSE | FALSE | FALSE |
| PPIC | 0.3408828 | 0.78969066 | 1 | 0.340882802 | 0.340882802 | 0.340882802 | 0.38671848 | 0.796901305 | 1 | 0.386718483 | 0.386718483 | 0.386718483 | 0.881475328 | -0.054789839 | FALSE | TRUE | FALSE |
| ETV1 | 0.3406319 | 0.78971644 | 1 | 0.340631897 | 0.340631897 | 6.746671749 | 0.38649375 | 0.796977917 | 0.0115 | 0.386493751 | 0.386493751 | 7.663943137 | 0.881338693 | -0.054857163 | FALSE | TRUE | FALSE |
| OR52I2 | 0.3405883 | 0.78974221 | 1 | 3.194586558 | 30.75291193 | 30.75291193 | 3.63283692 | 0.375667955 | 0.7484 | 3.632836919 | 35.02292459 | 35.02292459 | 0.093752708 | -1.028016177 | FALSE | FALSE | FALSE |
| ABLIM3 | 0.3405048 | 0.78976797 | 0.26995 | 0.085168963 | 0.340504842 | 1.675861712 | 0.03429517 | 0.890295199 | 0.0187 | 0.034295168 | 0.034295168 | 0.676135116 | 9.928653574 | 0.996890358 | FALSE | TRUE | FALSE |
| SENP2 | 0.3404041 | 0.78979373 | 0.01695 | 0.340404082 | 0.340404082 | 6.732596891 | 0.38640101 | 0.79702896 | 1 | 0.386401008 | 0.386401008 | 7.652529562 | 0.880960645 | -0.055043492 | FALSE | TRUE | FALSE |
| ST3GAL4 | 0.3403317 | 0.78981949 | 1 | 0.340331712 | 0.340331712 | 6.75414892 | 0.38607402 | 0.79725831 | 1 | 0.386074022 | 0.386074022 | 0.386074022 | 0.881519327 | -0.054768161 | FALSE | TRUE | FALSE |
| ZNF512B | 0.3401912 | 0.78984524 | 0.02685 | 0.340191207 | 0.340191207 | 6.661139983 | 0.38645571 | 0.797003442 | 1 | 0.386455708 | 0.386455708 | 7.586139411 | 0.880285114 | -0.055376642 | FALSE | TRUE | FALSE |
| SPRY1 | 0.3400059 | 0.78987098 | 1 | 0.340005904 | 0.340005904 | 0.340005904 | 3.49835423 | 0.38296264 | 0.00675 | 3.498354226 | 3.498354226 | 3.498354226 | 0.097190245 | -1.012377324 | FALSE | FALSE | FALSE |
| B3GALT4 | 0.3399626 | 0.78989671 | 1 | 0.339962556 | 0.339962556 | 6.729922727 | 0.37400172 | 0.801077687 | 1 | 3.486190502 | 3.486190502 | 7.412845614 | 0.908986601 | -0.041442519 | FALSE | TRUE | FALSE |
| ZNF629 | 0.3399515 | 0.78992244 | 1 | 3.20217652 | 30.95696544 | 108.2065448 | 0.38630105 | 0.797105472 | 0.75555 | 3.642330107 | 35.24655598 | 71.42671916 | 0.880016982 | -0.055508947 | FALSE | TRUE | FALSE |
| SH3GL3 | 0.3398693 | 0.78994816 | 0.502 | 0.33986929 | 0.33986929 | 0.33986929 | 0.11435736 | 0.872536658 | 0.00775 | 0.11435736 | 0.11435736 | 0.11435736 | 2.971993148 | 0.473047804 | FALSE | TRUE | FALSE |
| MYO1D | 0.3398416 | 0.78997388 | 0.1898 | 0.085007217 | 5.435546581 | 21.74648881 | 0.01641887 | 0.89475189 | 0.7837 | 0.065621161 | 1.048988629 | 4.195626987 | 20.69823625 | 1.31593334 | FALSE | TRUE | FALSE |
| IL25 | 0.3397524 | 0.78999959 | 0.5068 | 3.160512528 | 3.160512528 | 3.160512528 | 33.2914916 | 0.083574644 | 0.25 | 3.481745388 | 33.29149161 | 33.29149161 | 0.010205384 | -1.99117066 | FALSE | FALSE | FALSE |
| PRKAA1 | 0.3397517 | 0.79002529 | 0.0142 | 0.339751732 | 0.339751732 | 6.72261758 | 0.38580854 | 0.797334642 | 1 | 0.385808536 | 0.385808536 | 7.643724 | 0.880622639 | -0.055210154 | FALSE | TRUE | FALSE |
| EFCAB7 | 0.3396984 | 0.79005099 | 0.7508 | 0.339698373 | 3.051991444 | 6.725073352 | 4.36089219 | 0.337208526 | 0.49995 | 1.845159835 | 40.52143878 | 40.52143878 | 0.077896531 | -1.108481884 | FALSE | FALSE | FALSE |
| APCDD1 | 0.3396491 | 0.79007667 | 1 | 0.339649096 | 0.339649096 | 6.70637795 | 0.38553749 | 0.797436324 | 0.01475 | 0.385537494 | 0.385537494 | 7.624123331 | 0.880975524 | -0.055036157 | FALSE | TRUE | FALSE |
| HMGXB4 | 0.3396172 | 0.79010236 | 0.01565 | 0.339617151 | 0.339617151 | 6.721686883 | 0.38571671 | 0.797360072 | 1 | 0.385716712 | 0.385716712 | 7.643640142 | 0.880483371 | -0.055278842 | FALSE | TRUE | FALSE |
| TMEM126A | 0.3396043 | 0.79012803 | 1 | 0.339604319 | 0.339604319 | 0.339604319 | 0.38500698 | 0.797664719 | 0.0042 | 0.385006983 | 0.385006983 | 0.385006983 | 0.882073141 | -0.054495402 | FALSE | TRUE | FALSE |
| UXS1 | 0.3395908 | 0.7901537 | 0.7506 | 0.789442587 | 3.15618929 | 3.15618929 | 0.11587807 | 0.872219027 | 1 | 0.463305856 | 1.852861116 | 1.852861116 | 2.93058721 | 0.46695465 | FALSE | TRUE | FALSE |
| GKAP1 | 0.3395826 | 0.79017936 | 0.5063 | 3.153153283 | 3.153153283 | 3.153153283 | 0.39752508 | 0.792886401 | 1 | 0.397525085 | 0.397525085 | 0.397525085 | 0.854241913 | -0.068419124 | FALSE | TRUE | FALSE |
| CTIF | 0.3395071 | 0.79020502 | 1 | 0.339507139 | 0.339507139 | 6.682449374 | 0.38565027 | 0.797385496 | 1 | 0.385650274 | 0.385650274 | 7.605167053 | 0.880349794 | -0.055344733 | FALSE | TRUE | FALSE |
| CLEC9A | 0.3394085 | 0.79023067 | 1 | 0.339408463 | 3.14073497 | 3.14073497 | 0.39705949 | 0.79296618 | 1 | 0.39705949 | 0.39705949 | 0.39705949 | 0.854805064 | -0.068132914 | FALSE | TRUE | FALSE |
| EME2 | 0.3393212 | 0.79025631 | 0.75245 | 0.339321226 | 3.050423554 | 3.050423554 | 32840.5769 | 5.81324E-05 | 0.0617 | 338.8258095 | 32840.57688 | 65342.4493 | 1.03324E-05 | -4.98579975 | FALSE | FALSE | FALSE |
| PARP6 | 0.3392825 | 0.79028195 | 0.0178 | 0.339282502 | 0.339282502 | 6.701333079 | 0.36619468 | 0.803686741 | 1 | 0.366194684 | 0.366194684 | 7.243702665 | 0.92650854 | -0.033150573 | FALSE | TRUE | FALSE |
| OR1G1 | 0.3391922 | 0.79030758 | 1 | 0.339192239 | 0.339192239 | 0.339192239 | 0.38470035 | 0.797766058 | 0.0064 | 0.384700352 | 0.384700352 | 0.384700352 | 0.881705038 | -0.054676677 | FALSE | TRUE | FALSE |
| HECTD2 | 0.3391472 | 0.79033321 | 1 | 3.175481368 | 3.175481368 | 11.46019746 | 0.39758728 | 0.792859794 | 1 | 0.397587282 | 0.397587282 | 7.863894876 | 0.853013221 | -0.069044237 | FALSE | TRUE | FALSE |
| ZNF324 | 0.3391376 | 0.79035882 | 1 | 0.339137573 | 0.339137573 | 6.657458588 | 0.38524546 | 0.797563276 | 0.02005 | 0.385245459 | 0.385245459 | 7.579403247 | 0.880315563 | -0.05536162 | FALSE | TRUE | FALSE |
| IL1B | 0.3391375 | 0.79038443 | 1 | 3316883.963 | 4702154219 | 55130873689 | 0.40948054 | 0.788621483 | 1 | 388156.6005 | 522210920.7 | 6022323941 | 0.828213947 | -0.08185746 | FALSE | TRUE | FALSE |
| SLC39A1 | 0.3389436 | 0.79041004 | 1 | 0.338943635 | 0.338943635 | 0.338943635 | 1.50488182 | 0.534841844 | 0.749 | 1.50488182 | 6.020960157 | 6.020960157 | 0.225229404 | -0.647374913 | FALSE | FALSE | FALSE |
| C1QTNF7 | 0.3389422 | 0.79043564 | 0.62895 | 1.355463851 | 12.55829174 | 12.55829174 | 0.47278966 | 0.765998689 | 0.5064 | 0.472789663 | 0.472789663 | 0.472789663 | 0.716898488 | -0.144542336 | FALSE | TRUE | FALSE |
| CDK2AP2 | 0.3388992 | 0.79046123 | 1 | 0.338899166 | 0.338899166 | 0.338899166 | 0.34248709 | 0.811374409 | 1 | 0.342487094 | 0.342487094 | 0.342487094 | 0.989523904 | -0.00457371 | FALSE | TRUE | FALSE |
| YBEY | 0.338589 | 0.79048682 | 1 | 0.338588974 | 0.338588974 | 0.338588974 | 0.37981686 | 0.799323783 | 1 | 1.51921277 | 1.51921277 | 1.51921277 | 0.891453245 | -0.04990143 | FALSE | TRUE | FALSE |
| CCDC127 | 0.3384956 | 0.7905124 | 1 | 0.338495551 | 0.338495551 | 0.338495551 | 3.27292007 | 0.396216948 | 0.50135 | 31.39962621 | 308.9692284 | 308.9692284 | 0.103423103 | -0.985382435 | FALSE | FALSE | FALSE |
| DYNLL2 | 0.3383718 | 0.79053797 | 1 | 0.338371817 | 0.338371817 | 0.338371817 | 0.35041031 | 0.808993677 | 1 | 0.35041031 | 0.35041031 | 0.35041031 | 0.965644579 | -0.015182693 | FALSE | TRUE | FALSE |
| GNAI1 | 0.3383518 | 0.79056354 | 1 | 0.338351767 | 0.338351767 | 6.717944329 | 0.38471615 | 0.797740734 | 1 | 0.384716147 | 0.384716147 | 0.384716147 | 0.879484184 | -0.055771966 | FALSE | TRUE | FALSE |
| OR6C74 | 0.3382286 | 0.7905891 | 1 | 3.137029261 | 29.86195486 | 29.86195486 | 3.6860834 | 0.372558706 | 0.50735 | 3.686083402 | 3.686083402 | 3.686083402 | 0.091758265 | -1.037354808 | FALSE | FALSE | FALSE |
| HSPB6 | 0.3382103 | 0.79061466 | 1 | 0.338210275 | 0.338210275 | 0.338210275 | 1.49492214 | 0.536500982 | 0.7557 | 1.494922139 | 5.980323712 | 5.980323712 | 0.226239391 | -0.645431776 | FALSE | FALSE | FALSE |
| GSTM4 | 0.3379609 | 0.79064021 | 1 | 0.337960921 | 0.337960921 | 0.337960921 | 0.38347967 | 0.79796844 | 1 | 0.38347967 | 0.38347967 | 0.38347967 | 0.88130075 | -0.05487586 | FALSE | TRUE | FALSE |
| GLI2 | 0.3379405 | 0.79066575 | 0.28285 | 0.084511045 | 0.337940494 | 6.502907136 | 0.53212095 | 0.741947662 | 0.2801 | 0.13306232 | 0.532120949 | 10.2862661 | 0.635082107 | -0.197170123 | FALSE | TRUE | FALSE |

| | | | | | | | | | | | | | | | | | |
|---|---|---|---|---|---|---|---|---|---|---|---|---|---|---|---|---|---|
| EPS8L1 | 0.3379395 | 0.79069129 | 0.5123 | 0.337939503 | 0.337939503 | 6.626491223 | 4.26822513 | 0.339328618 | 0.51055 | 4.268225132 | 40.05632412 | 83.89083933 | 0.079175651 | -1.101408358 | FALSE | FALSE | FALSE |
| LAT | 0.3378297 | 0.79071682 | 1 | 0.337829727 | 0.337829727 | 0.337829727 | 0.36037139 | 0.805574631 | 1 | 3.339851568 | 31.76834262 | 31.76834262 | 0.937448789 | -0.028052448 | FALSE | TRUE | FALSE |
| GNB4 | 0.3378211 | 0.79074234 | 1 | 3.147103284 | 3.147103284 | 6.708696213 | 0.39588983 | 0.793522878 | 1 | 0.395889829 | 0.395889829 | 0.395889829 | 0.85332108 | -0.068887526 | FALSE | TRUE | FALSE |
| RNASE9 | 0.3375127 | 0.79076786 | 1 | 0.337512721 | 0.337512721 | 0.337512721 | 3.44080991 | 0.386671467 | 0.50145 | 3.440809907 | 3.440809907 | 3.440809907 | 0.098091069 | -1.008370534 | FALSE | FALSE | FALSE |
| CHCHD2 | 0.3375054 | 0.79079337 | 1 | 0.337505383 | 0.337505383 | 0.337505383 | 0.38342049 | 0.797993711 | 1 | 0.383420495 | 0.383420495 | 0.383420495 | 0.880248676 | -0.055394619 | FALSE | TRUE | FALSE |
| SLC22A3 | 0.3374667 | 0.79081888 | 0.75285 | 0.337466666 | 3.130114515 | 6.668185136 | 0.05820607 | 0.88501346 | 0.01525 | 0.058206071 | 0.058206071 | 1.151788802 | 5.79779153 | 0.763262595 | FALSE | TRUE | FALSE |
| OR2T2 | 0.337254 | 0.79084438 | 1 | 1.611E+278 | 1.1018E+306 | 4.9472E+307 | 0.34535752 | 0.810558007 | 1 | 9.8922E+267 | 1.6379E+294 | 3.1446E+302 | 0.97653584 | -0.010311813 | FALSE | TRUE | FALSE |
| DR1 | 0.3371999 | 0.79086988 | 1 | 0.337199851 | 0.337199851 | 0.337199851 | 0.34241952 | 0.811418337 | 1 | 0.342419522 | 0.342419522 | 0.342419522 | 0.984756505 | -0.006671142 | FALSE | TRUE | FALSE |
| ZNF296 | 0.3370903 | 0.79089536 | 1 | 295.4966639 | 30659.00598 | 325061.0627 | 0.46240778 | 0.770100373 | 1 | 0.462407777 | 0.462407777 | 9.126250123 | 0.728989349 | -0.137278817 | FALSE | TRUE | FALSE |
| TCEANC2 | 0.3369728 | 0.79092085 | 1 | 3.124794068 | 3.124794068 | 3.124794068 | 0.38260696 | 0.798220858 | 1 | 3.557785829 | 3.557785829 | 3.557785829 | 0.880728413 | -0.055157993 | FALSE | TRUE | FALSE |
| ACOT13 | 0.3369046 | 0.79094632 | 0.0029 | 0.33690458 | 0.33690458 | 0.33690458 | 29.5684206 | 0.0936982 | 0.68865 | 29.56842065 | 2863.383738 | 2863.383738 | 0.011394067 | -1.943321213 | FALSE | FALSE | FALSE |
| MRPL14 | 0.3368872 | 0.79097179 | 1 | 0.336887235 | 1.347315188 | 1.347315188 | 0.38732309 | 0.796671098 | 1 | 0.387323088 | 0.387323088 | 0.387323088 | 0.86978351 | -0.06058883 | FALSE | TRUE | FALSE |
| PRKAB1 | 0.3368824 | 0.79099725 | 1 | 0.336882409 | 0.336882409 | 0.336882409 | 0.38290046 | 0.798145199 | 1 | 0.382900462 | 0.382900462 | 0.382900462 | 0.879817191 | -0.055607557 | FALSE | TRUE | FALSE |
| ELK3 | 0.3366337 | 0.79102271 | 1 | 0.33663372 | 0.33663372 | 6.668493831 | 0.38241173 | 0.798271265 | 1 | 0.382411731 | 0.382411731 | 7.584012288 | 0.880291299 | -0.055373591 | FALSE | TRUE | FALSE |
| SLC40A1 | 0.3366316 | 0.79104816 | 0.0155 | 0.336631606 | 0.336631606 | 2.849689908 | 0.4276839 | 0.781885478 | 0.93615 | 4.036061092 | 64.55299652 | 64.55299652 | 0.787103766 | -0.10396801 | FALSE | TRUE | FALSE |
| SIRT7 | 0.3365119 | 0.79107361 | 1 | 0.336511915 | 0.336511915 | 6.660206076 | 0.35933729 | 0.805831333 | 1 | 3.35260182 | 32.10330151 | 32.10330151 | 0.936479256 | -0.028501838 | FALSE | TRUE | FALSE |
| APBB1IP | 0.3364898 | 0.79109905 | 1 | 0.336489845 | 0.336489845 | 6.658544032 | 0.38266014 | 0.798195645 | 0.01415 | 0.382660136 | 0.382660136 | 7.581817594 | 0.87934387 | -0.05584126 | FALSE | TRUE | FALSE |
| UBAC1 | 0.3364218 | 0.79112448 | 1 | 0.336421777 | 0.336421777 | 6.656916106 | 0.3822814 | 0.798346827 | 1 | 0.382281398 | 0.382281398 | 7.574031842 | 0.880037007 | -0.055499065 | FALSE | TRUE | FALSE |
| BCL6 | 0.3364195 | 0.7911499 | 1 | 30.36886365 | 299.8103836 | 597.0570289 | 1.61160534 | 0.524077445 | 0.75725 | 3.79123512 | 15.16430147 | 54.52209954 | 0.208748055 | -0.680377562 | FALSE | FALSE | FALSE |
| PRIM1 | 0.336401 | 0.79117532 | 1 | 0.336401044 | 0.336401044 | 0.336401044 | 0.33757451 | 0.812791626 | 0.9383 | 29.76060577 | 2903.671487 | 2903.671487 | 0.99652382 | -0.001512316 | FALSE | TRUE | FALSE |
| RILPL2 | 0.3363096 | 0.79120073 | 0.0057 | 0.336309585 | 0.336309585 | 0.336309585 | 0.38208787 | 0.798397169 | 1 | 0.382087871 | 0.382087871 | 0.382087871 | 0.880189116 | -0.055424006 | FALSE | TRUE | FALSE |
| SEC13 | 0.3362517 | 0.79122614 | 0.5037 | 0.336251712 | 0.336251712 | 1.670292846 | 0.44939355 | 0.774658241 | 0.0075 | 0.449393554 | 0.449393554 | 0.449393554 | 0.748234391 | -0.125962335 | FALSE | TRUE | FALSE |
| ZNF688 | 0.3361 | 0.79125154 | 1 | 0.336100027 | 0.336100027 | 6.668841434 | 0.38175241 | 0.798522913 | 1 | 0.381752408 | 0.381752408 | 0.381752408 | 0.880413639 | -0.055313238 | FALSE | TRUE | FALSE |
| KCTD10 | 0.3360749 | 0.79127693 | 1 | 0.336074922 | 0.336074922 | 6.664948245 | 0.37795899 | 0.799944933 | 1 | 1.511651036 | 1.511651036 | 1.511651036 | 0.889183561 | -0.051008575 | FALSE | TRUE | FALSE |
| NR1H2 | 0.3360106 | 0.79130232 | 0.2543 | 0.143274775 | 1.342913775 | 2.833155152 | 0.05485042 | 0.885661836 | 1 | 0.21923912 | 0.21923912 | 1.086100109 | 6.125944938 | 0.787173089 | FALSE | TRUE | FALSE |
| SLC29A1 | 0.3359949 | 0.7913277 | 0.5043 | 0.335994939 | 0.335994939 | 1.664056167 | 0.43537147 | 0.779065078 | 0.75395 | 0.99946755 | 3.996817831 | 3.996817831 | 0.771743118 | -0.112527235 | FALSE | TRUE | FALSE |
| CSPG5 | 0.335945 | 0.79135308 | 1 | 0.335944977 | 0.335944977 | 6.659078726 | 0.35792385 | 0.806435324 | 0.00965 | 0.357923852 | 0.357923852 | 0.357923852 | 0.938593434 | -0.027522488 | FALSE | TRUE | FALSE |
| PPIL1 | 0.3359002 | 0.79137845 | 1 | 0.335900249 | 0.335900249 | 0.335900249 | 2896.6052 | 0.000789136 | 0.00165 | 29.70496694 | 2896.605202 | 2896.605202 | 0.000115963 | -3.93567898 | FALSE | FALSE | FALSE |
| ATN1 | 0.3357711 | 0.79140381 | 1 | 0.335771057 | 0.335771057 | 6.554073344 | 0.35882297 | 0.806064098 | 1 | 3.380437196 | 32.68509955 | 114.0373746 | 0.935756871 | -0.028836975 | FALSE | TRUE | FALSE |
| PEX16 | 0.3357671 | 0.79142916 | 1 | 0.335767076 | 0.335767076 | 6.660410084 | 0.38146311 | 0.798648493 | 1 | 0.381463115 | 0.381463115 | 0.381463115 | 0.880208498 | -0.055414443 | FALSE | TRUE | FALSE |
| STAT4 | 0.3356469 | 0.79145451 | 0.5144 | 0.33564694 | 0.33564694 | 6.625794729 | 0.11379356 | 0.872608181 | 1 | 0.113793557 | 0.113793557 | 2.249835877 | 2.949612861 | 0.469765018 | FALSE | TRUE | FALSE |
| CCDC28B | 0.3356097 | 0.79147986 | 1 | 0.335609735 | 0.335609735 | 0.335609735 | 0.38124079 | 0.79874884 | 1 | 0.381240795 | 0.381240795 | 0.381240795 | 0.880309086 | -0.055364816 | FALSE | TRUE | FALSE |
| AHR | 0.3355651 | 0.79150519 | 1 | 0.33556508 | 0.33556508 | 6.608973357 | 0.38185005 | 0.798472635 | 1 | 0.381850046 | 0.381850046 | 7.534360308 | 0.878787586 | -0.056116087 | FALSE | TRUE | FALSE |
| TXN | 0.3355524 | 0.79153052 | 1 | 0.335552369 | 0.335552369 | 0.335552369 | 0.34566539 | 0.810402745 | 1 | 0.345665386 | 0.345665386 | 0.345665386 | 0.970743335 | -0.012895583 | FALSE | TRUE | FALSE |
| MRC1 | 0.3354457 | 0.79155585 | 1 | 3.178302426 | 3.178302426 | 61.84605914 | 0.38124066 | 0.79877391 | 1 | 3.613307324 | 3.613307324 | 70.54006643 | 0.879879022 | -0.055577037 | FALSE | TRUE | FALSE |
| BCL11A | 0.3354141 | 0.79158116 | 1 | 3.149888208 | 30.35953228 | 106.513826 | 0.37903399 | 0.799547862 | 1 | 3.564245416 | 34.39870263 | 69.81657518 | 0.884918289 | -0.053096829 | FALSE | TRUE | FALSE |
| PRTN3 | 0.3353917 | 0.79160648 | 1 | 0.335391717 | 0.335391717 | 0.335391717 | 0.38098182 | 0.798949211 | 1 | 0.38098182 | 0.38098182 | 0.38098182 | 0.880335227 | -0.055351919 | FALSE | TRUE | FALSE |
| FBXO30 | 0.3353642 | 0.79163178 | 1 | 0.335364246 | 0.335364246 | 6.618485627 | 0.38137175 | 0.798673589 | 1 | 0.38137175 | 0.38137175 | 7.538444428 | 0.879363106 | -0.05583176 | FALSE | TRUE | FALSE |
| ZFP91 | 0.3352733 | 0.79165708 | 1 | 3.147198211 | 3.147198211 | 6.633663879 | 0.37944349 | 0.799373623 | 0.01395 | 0.379443487 | 0.379443487 | 7.517278652 | 0.883592255 | -0.0537481 | FALSE | TRUE | FALSE |
| MGLL | 0.3352531 | 0.79168237 | 1 | 1.340733149 | 1.340733149 | 1.340733149 | 0.38583798 | 0.797283761 | 1 | 0.385837979 | 0.385837979 | 0.385837979 | 0.868896067 | -0.061032168 | FALSE | TRUE | FALSE |
| PRMT5 | 0.3352384 | 0.79170766 | 1 | 0.335238412 | 0.335238412 | 6.614745072 | 0.38157237 | 0.79859828 | 1 | 0.381572372 | 0.381572372 | 7.541146971 | 0.87857098 | -0.056223146 | FALSE | TRUE | FALSE |
| ZNF540 | 0.3351998 | 0.79173293 | 0.51035 | 0.335199832 | 0.335199832 | 1.275825707 | 0.0566947 | 0.885351087 | 1 | 0.056694702 | 0.056694702 | 1.121925916 | 5.91236604 | 0.771761314 | FALSE | TRUE | FALSE |
| SRSF4 | 0.3350541 | 0.79175821 | 0.996 | 2971.605215 | 325173.3462 | 3516710.751 | 2.94176448 | 0.415824154 | 1 | 3160290.538 | 4519845589 | 53148711102 | 0.113895621 | -0.943492972 | FALSE | FALSE | FALSE |
| FGF4 | 0.3350168 | 0.79178347 | 1 | 0.335016828 | 0.335016828 | 0.335016828 | 0.38124592 | 0.798723763 | 1 | 0.381245918 | 0.381245918 | 0.381245918 | 0.878742074 | -0.056138579 | FALSE | TRUE | FALSE |
| TNIP2 | 0.3349973 | 0.79180873 | 1 | 0.33499728 | 0.33499728 | 6.632981209 | 0.36909849 | 0.802463426 | 0.50955 | 3.436241825 | 3.436241825 | 7.316975093 | 0.907609455 | -0.042100988 | FALSE | TRUE | FALSE |
| UBE2C | 0.3349767 | 0.79183399 | 1 | 0.334976656 | 0.334976656 | 0.334976656 | 0.38077634 | 0.799099208 | 1 | 0.380776335 | 0.380776335 | 0.380776335 | 0.879720259 | -0.055655407 | FALSE | TRUE | FALSE |
| MZT2A | 0.3349455 | 0.79185923 | 0.49425 | 0.036218524 | 0.334945486 | 0.334945486 | 0.3669991 | 0.803304662 | 0.74935 | 0.3669991 | 3.490940342 | 3.490940342 | 0.912660237 | -0.03969087 | FALSE | TRUE | FALSE |
| PTRH2 | 0.3348836 | 0.79188448 | 1 | 0.334883576 | 0.334883576 | 0.334883576 | 0.38095497 | 0.798999237 | 1 | 0.380954974 | 0.380954974 | 0.380954974 | 0.879063405 | -0.055979799 | FALSE | TRUE | FALSE |
| SOX14 | 0.3347582 | 0.79190971 | 0.8704 | 1.338447648 | 5.352787897 | 5.352787897 | 0.11240896 | 0.872832506 | 1 | 0.449366689 | 0.449366689 | 0.449366689 | 2.978038773 | 0.473930348 | FALSE | TRUE | FALSE |
| ZNF606 | 0.3346711 | 0.79193494 | 1 | 0.334671115 | 0.334671115 | 6.60249007 | 0.3809287 | 0.79902424 | 1 | 0.3809287 | 0.3809287 | 7.527359449 | 0.878566292 | -0.056225463 | FALSE | TRUE | FALSE |
| ICAM4 | 0.3346025 | 0.79196016 | 1 | 3.098884574 | 3.098884574 | 3.098884574 | 0.39204029 | 0.794835964 | 1 | 0.392040289 | 0.392040289 | 0.392040289 | 0.853490014 | -0.068801556 | FALSE | TRUE | FALSE |
| SLC30A8 | 0.3341643 | 0.79198538 | 0.00835 | 0.334164299 | 0.334164299 | 0.334164299 | 0.49141279 | 0.75827473 | 0.49785 | 0.054857849 | 0.491412785 | 0.491412785 | 0.680007337 | -0.167486401 | FALSE | TRUE | FALSE |
| SLC23A2 | 0.3341191 | 0.79201059 | 1 | 0.334119109 | 0.334119109 | 6.584782697 | 0.38016803 | 0.799224036 | 1 | 0.380168029 | 0.380168029 | 7.505480131 | 0.878872194 | -0.056074276 | FALSE | TRUE | FALSE |
| RAB42 | 0.3340933 | 0.7920358 | 0.0038 | 0.334093281 | 0.334093281 | 0.334093281 | 0.36817457 | 0.802800813 | 1 | 3.427708292 | 3.427708292 | 3.427708292 | 0.907431703 | -0.042186052 | FALSE | TRUE | FALSE |
| CRYL1 | 0.3340267 | 0.792061 | 0.6836 | 0.708713574 | 12.10051194 | 12.10051194 | 0.2999493 | 0.825482929 | 0.25055 | 0.299949295 | 2.801189373 | 2.801189373 | 1.113610631 | 0.046733368 | FALSE | TRUE | FALSE |
| MDM1 | 0.3338574 | 0.79208619 | 0.50415 | 0.333857363 | 0.333857363 | 1.341961117 | 0.47403479 | 0.765449393 | 0.75765 | 0.474034789 | 4.294591553 | 9.375628214 | 0.704288738 | -0.152249256 | FALSE | TRUE | FALSE |
| ZZZ3 | 0.3338566 | 0.79211138 | 1 | 3.144395561 | 3.144395561 | 61.91387496 | 0.39241726 | 0.794705432 | 1 | 0.392417263 | 0.392417263 | 7.74119177 | 0.850769391 | -0.070188144 | FALSE | TRUE | FALSE |
| TMEM128 | 0.333759 | 0.79213656 | 1 | 3.108109863 | 29.70636136 | 29.70636136 | 1.54868653 | 0.531068794 | 0.4988 | 1.54868653 | 14.46811219 | 14.46811219 | 0.215511022 | -0.666530513 | FALSE | FALSE | FALSE |
| RPF2 | 0.3336326 | 0.79216174 | 1 | 3.096031403 | 3.096031403 | 3.096031403 | 0.39127968 | 0.795200531 | 0.0062 | 0.391279681 | 0.391279681 | 0.391279681 | 0.852670322 | -0.069218853 | FALSE | TRUE | FALSE |
| PXMP4 | 0.3335903 | 0.79218691 | 1 | 0.333590325 | 0.333590325 | 0.333590325 | 0.36784081 | 0.802945045 | 1 | 3.432995467 | 3.432995467 | 3.432995467 | 0.90688774 | -0.042446469 | FALSE | TRUE | FALSE |
| RABGEF1 | 0.3335744 | 0.79221207 | 0.01485 | 0.333574424 | 0.333574424 | 6.603728103 | 0.37526456 | 0.800758889 | 0.51045 | 1.501021048 | 1.501021048 | 7.438138751 | 0.888904674 | -0.05114481 | FALSE | TRUE | FALSE |
| OR8B2 | 0.3335682 | 0.79223723 | 1 | 0.333568226 | 0.333568226 | 0.333568226 | 3.41957278 | 0.388281174 | 0.5018 | 3.41957278 | 3.41957278 | 3.41957278 | 0.097546754 | -1.010787176 | FALSE | FALSE | FALSE |
| CCNE2 | 0.33354 | 0.79226238 | 1 | 0.333540031 | 0.333540031 | 6.629003112 | 0.37922928 | 0.799448336 | 1 | 0.379229282 | 0.379229282 | 0.379229282 | 0.879520773 | -0.055753899 | FALSE | TRUE | FALSE |
| VAT1 | 0.3334151 | 0.79228752 | 1 | 3.104086105 | 3.104086105 | 6.606690452 | 0.39127069 | 0.795226519 | 1 | 0.391270688 | 0.391270688 | 0.391270688 | 0.852134139 | -0.069492035 | FALSE | TRUE | FALSE |
| METTL16 | 0.3332189 | 0.79231266 | 1 | 0.333218931 | 0.333218931 | 6.587543519 | 0.37685042 | 0.800315738 | 1 | 0.376850418 | 0.376850418 | 7.460446523 | 0.884220675 | -0.053439334 | FALSE | TRUE | FALSE |
| SGK2 | 0.3328728 | 0.7923378 | 0.9387 | 1.331508421 | 21.32065744 | 21.32065744 | 1.72551816 | 0.515256147 | 0.8126 | 6.904079161 | 63.03768532 | 252.2870712 | 0.192911818 | -0.714641167 | FALSE | FALSE | FALSE |
| DRD2 | 0.3328149 | 0.79236292 | 1 | 0.332814936 | 0.332814936 | 6.57661246 | 0.37921925 | 0.799473227 | 1 | 0.379219253 | 0.379219253 | 7.504381858 | 0.877631957 | -0.056687571 | FALSE | TRUE | FALSE |
| ADD3 | 0.3328129 | 0.79238805 | 1 | 0.332812875 | 0.332812875 | 6.572395895 | 0.37893997 | 0.799622438 | 1 | 0.378939966 | 0.378939966 | 7.494659271 | 0.878273356 | -0.056370292 | FALSE | TRUE | FALSE |
| CAMK2G | 0.332812 | 0.79241316 | 1 | 0.332812014 | 0.332812014 | 6.572364199 | 0.37894861 | 0.799597586 | 1 | 0.378948606 | 0.378948606 | 7.494815397 | 0.878251058 | -0.056381318 | FALSE | TRUE | FALSE |
| SGSM3 | 0.3327722 | 0.79243827 | 0.7568 | 0.332772207 | 3.0187487 | 6.520142921 | 0.50391902 | 0.753904779 | 0.51045 | 0.503919018 | 0.503919018 | 9.897667596 | 0.660368424 | -0.180213701 | FALSE | TRUE | FALSE |
| GPR3 | 0.3327518 | 0.79246337 | 1 | 0.332751804 | 0.332751804 | 6.604425197 | 0.37874269 | 0.7997466 | 1 | 0.378742695 | 0.378742695 | 0.378742695 | 0.878569564 | -0.056223846 | FALSE | TRUE | FALSE |
| OIP5 | 0.3327245 | 0.79248847 | 1 | 0.332724456 | 0.332724456 | 0.332724456 | 3.39799807 | 0.389425506 | 0.50085 | 3.397998066 | 3.397998066 | 3.397998066 | 0.097917789 | -1.009138403 | FALSE | FALSE | FALSE |
| ZNF202 | 0.332668 | 0.79251356 | 1 | 0.332667987 | 0.332667987 | 6.563720385 | 0.37876644 | 0.79972178 | 1 | 0.378766444 | 0.378766444 | 7.485384499 | 0.878293185 | -0.056360487 | FALSE | TRUE | FALSE |

| | | | | | | | | | | | | | | | | | |
|---|---|---|---|---|---|---|---|---|---|---|---|---|---|---|---|---|---|
| PRKAR1A | 0.332451 | 0.79253864 | 0.0117 | 0.332450994 | 0.332450994 | 6.593125999 | 3.47650785 | 0.384822413 | 0.51195 | 3.476507847 | 3.476507847 | 7.287634224 | 0.095627857 | -1.019415578 | FALSE | FALSE | FALSE |
| KIAA1143 | 0.3322295 | 0.79256372 | 1 | 0.332229489 | 0.332229489 | 0.332229489 | 0.37784479 | 0.799994454 | 1 | 0.377844792 | 0.377844792 | 0.377844792 | 0.879275027 | -0.055875262 | FALSE | TRUE | FALSE |
| DBNDD1 | 0.3322181 | 0.79258879 | 1 | 0.332218059 | 0.332218059 | 0.332218059 | 0.37766327 | 0.800019206 | 1 | 0.377663269 | 0.377663269 | 0.377663269 | 0.879667379 | -0.055681513 | FALSE | TRUE | FALSE |
| NOS1 | 0.3320767 | 0.79261386 | 0.7615 | 1.32763391 | 12.66707058 | 61.18174326 | 0.22526881 | 0.846085267 | 0.5196 | 0.225268807 | 0.225268807 | 4.368075412 | 1.474135374 | 0.168537368 | FALSE | TRUE | FALSE |
| SEL1L | 0.331987 | 0.79263892 | 1 | 0.331986984 | 0.331986984 | 6.540905018 | 0.37828153 | 0.799845815 | 0.0188 | 0.378281531 | 0.378281531 | 7.46636654 | 0.877618803 | -0.056694081 | FALSE | TRUE | FALSE |
| GLIPR2 | 0.3319041 | 0.79266398 | 0.00525 | 0.331904074 | 0.331904074 | 0.331904074 | 0.37730223 | 0.800142871 | 1 | 0.377302235 | 0.377302235 | 0.377302235 | 0.879676938 | -0.055676794 | FALSE | TRUE | FALSE |
| ZCCHC14 | 0.3318468 | 0.79268902 | 1 | 0.331846824 | 0.331846824 | 6.484779521 | 32.2703856 | 0.086453195 | 0.26345 | 3.341223966 | 32.27038562 | 65.47281476 | 0.010283324 | -1.98786649 | FALSE | FALSE | FALSE |
| RFX3 | 0.3317719 | 0.79271407 | 0.5018 | 3.112421996 | 3.112421996 | 61.37627928 | 0.39007589 | 0.795615538 | 1 | 0.390075891 | 0.390075891 | 7.705181491 | 0.850531735 | -0.070309477 | FALSE | TRUE | FALSE |
| NTRK1 | 0.3317531 | 0.7927391 | 0.7049 | 1.717513963 | 64.89322244 | 259.4617927 | 0.60523239 | 0.709828913 | 0.9164 | 12.70657596 | 813.0166901 | 3253.419344 | 0.548141743 | -0.261107123 | FALSE | TRUE | FALSE |
| LETMD1 | 0.3317007 | 0.79276413 | 0.0115 | 0.331700697 | 0.331700697 | 6.580301026 | 0.3772895 | 0.800167585 | 1 | 0.377289497 | 0.377289497 | 0.377289497 | 0.879167588 | -0.055928331 | FALSE | TRUE | FALSE |
| DLK2 | 0.3316942 | 0.79278916 | 1 | 0.331694193 | 0.331694193 | 6.573704402 | 0.37807789 | 0.799920163 | 1 | 0.378077888 | 0.378077888 | 7.501138348 | 0.877317089 | -0.056843411 | FALSE | TRUE | FALSE |
| TSSK1B | 0.3315631 | 0.79281417 | 1 | 3.083606281 | 3.083606281 | 6.566377543 | 0.38932048 | 0.795925648 | 1 | 0.389320482 | 0.389320482 | 7.719296429 | 0.851645742 | -0.06974102 | FALSE | TRUE | FALSE |
| TMEM63B | 0.3315357 | 0.79283919 | 1 | 0.331535677 | 0.331535677 | 6.493614674 | 0.35922646 | 0.805901224 | 1 | 3.379106292 | 3.379106292 | 66.34959741 | 0.922915513 | -0.034838054 | FALSE | TRUE | FALSE |
| ADAMTS3 | 0.3314398 | 0.79286419 | 0.0286 | 0.067849694 | 0.271273463 | 1.324815919 | 0.00824244 | 0.897228781 | 0.51795 | 0.032942037 | 0.032942037 | 0.645055589 | 40.21138 | 1.604348978 | FALSE | TRUE | FALSE |
| TMEM95 | 0.3314056 | 0.79288919 | 1 | 0.331405572 | 0.331405572 | 0.331405572 | 0.37678847 | 0.800340408 | 1 | 0.376788472 | 0.376788472 | 0.376788472 | 0.879553376 | -0.0557378 | FALSE | TRUE | FALSE |
| PPP1R12C | 0.3313859 | 0.79291418 | 0.02315 | 0.331385938 | 0.331385938 | 6.510779062 | 0.112058 | 0.872903737 | 1 | 0.112058003 | 0.112058003 | 2.206293944 | 2.957271479 | 0.470891195 | FALSE | TRUE | FALSE |
| NXNL2 | 0.3313665 | 0.79293917 | 1 | 0.331366517 | 0.331366517 | 0.331366517 | 0.36542838 | 0.803924764 | 1 | 3.425484605 | 3.425484605 | 3.425484605 | 0.906789234 | -0.042493645 | FALSE | TRUE | FALSE |
| OR5M1 | 0.3313242 | 0.79296415 | 1 | 0.331324243 | 0.331324243 | 0.331324243 | 0.37702269 | 0.80024169 | 0.0058 | 0.377022693 | 0.377022693 | 0.377022693 | 0.878791249 | -0.056114276 | FALSE | TRUE | FALSE |
| PPM1E | 0.3311375 | 0.79298912 | 0.0205 | 0.331137466 | 0.331137466 | 6.535792489 | 0.37704175 | 0.800216995 | 0.0163 | 0.377041746 | 0.377041746 | 7.453581638 | 0.878251465 | -0.056381117 | FALSE | TRUE | FALSE |
| ABHD14A | 0.3309281 | 0.79301409 | 0.7568 | 3.071355468 | 29.25618288 | 29.25618288 | 0.34314744 | 0.811154456 | 1 | 296.3531382 | 2967.68778 | 30443.9693 | 0.964390485 | -0.015747083 | FALSE | TRUE | FALSE |
| CYB5B | 0.3308723 | 0.79303906 | 1 | 0.330872344 | 0.330872344 | 0.330872344 | 0.37714841 | 0.800192293 | 1 | 0.377148408 | 0.377148408 | 0.377148408 | 0.877300121 | -0.056851811 | FALSE | TRUE | FALSE |
| YRDC | 0.3308339 | 0.79306401 | 1 | 0.330833855 | 0.330833855 | 0.330833855 | 0.35402574 | 0.807723209 | 1 | 3.310511246 | 31.77184632 | 31.77184632 | 0.93449096 | -0.029424895 | FALSE | TRUE | FALSE |
| BARHL2 | 0.330773 | 0.79308896 | 0.754 | 0.330773029 | 1.322550012 | 1.63875825 | 1.03934471 | 0.583850713 | 0.50255 | 1.039344705 | 1.039344705 | 2.184145977 | 0.318251516 | -0.497229518 | FALSE | FALSE | FALSE |
| TP53I11 | 0.330728 | 0.79311391 | 1 | 0.330728039 | 0.330728039 | 0.330728039 | 0.36050617 | 0.805527886 | 0.00575 | 0.360506169 | 0.360506169 | 0.360506169 | 0.91739911 | -0.037441685 | FALSE | TRUE | FALSE |
| SNTB2 | 0.3306434 | 0.79313885 | 1 | 0.330643408 | 0.330643408 | 6.556776903 | 0.37691895 | 0.800291062 | 0.0108 | 0.376918954 | 0.376918954 | 7.482062358 | 0.877226799 | -0.056888109 | FALSE | TRUE | FALSE |
| ZNF746 | 0.3302533 | 0.79316378 | 1 | 0.330253268 | 0.330253268 | 6.504489707 | 0.37298053 | 0.801346657 | 1 | 0.37298053 | 0.37298053 | 7.359486544 | 0.885443719 | -0.052839039 | FALSE | TRUE | FALSE |
| ZNF784 | 0.3301958 | 0.79318871 | 1 | 0.330195783 | 0.330195783 | 6.551289361 | 0.3757749 | 0.800537566 | 0.00985 | 0.375774903 | 0.375774903 | 0.375774903 | 0.878706323 | -0.056156248 | FALSE | TRUE | FALSE |
| SLC44A4 | 0.3301307 | 0.79321363 | 0.50545 | 0.330130698 | 0.330130698 | 6.509241494 | 0.05470611 | 0.885728827 | 0.50715 | 0.484113118 | 0.484113118 | 1.080485131 | 6.034622483 | 0.780650106 | FALSE | TRUE | FALSE |
| ADTRP | 0.3301213 | 0.79323855 | 1 | 0.330121264 | 0.330121264 | 0.330121264 | 0.37563092 | 0.800635994 | 1 | 0.375630921 | 0.375630921 | 0.375630921 | 0.878844755 | -0.056087835 | FALSE | TRUE | FALSE |
| UBA3 | 0.3300508 | 0.79326346 | 1 | 0.330050803 | 0.330050803 | 6.548181403 | 588488.168 | 2.92594E-06 | 0.000025 | 0.376300301 | 0.376300301 | 0.376300301 | 5.60845E-07 | -6.251156941 | FALSE | FALSE | FALSE |
| GBP2 | 0.3299837 | 0.79328836 | 0.50645 | 0.329983701 | 0.329983701 | 0.729823041 | 0.05405439 | 0.885887637 | 0.5051 | 0.486924099 | 0.486924099 | 1.071028788 | 6.104660592 | 0.785661523 | FALSE | TRUE | FALSE |
| TXNDC12 | 0.3299187 | 0.79331326 | 0.00435 | 0.329918704 | 0.329918704 | 0.329918704 | 0.37530744 | 0.80070975 | 1 | 0.375307439 | 0.375307439 | 0.375307439 | 0.879062521 | -0.055980236 | FALSE | TRUE | FALSE |
| CDKN1A | 0.3299104 | 0.79333815 | 1 | 1.319433208 | 1.319433208 | 1.319433208 | 1.5037028 | 0.535257757 | 0.50065 | 1.503702805 | 1.503702805 | 1.503702805 | 0.219398645 | -0.65876606 | FALSE | FALSE | FALSE |
| SLC6A1 | 0.3297683 | 0.79336303 | 1 | 0.329768269 | 0.329768269 | 6.502840015 | 154.126409 | 0.017848364 | 0.0001 | 0.376213278 | 0.376213278 | 7.431234059 | 0.002139596 | -2.669668195 | FALSE | FALSE | FALSE |
| TNFRSF9 | 0.3297472 | 0.79338791 | 1 | 0.329747198 | 0.329747198 | 0.329747198 | 0.37569468 | 0.800586793 | 1 | 0.375694679 | 0.375694679 | 0.375694679 | 0.877699944 | -0.056653929 | FALSE | TRUE | FALSE |
| TIFA | 0.3296978 | 0.79341279 | 1 | 0.329697847 | 0.329697847 | 0.329697847 | 0.37550184 | 0.800660585 | 1 | 0.375501844 | 0.375501844 | 0.375501844 | 0.878019247 | -0.056495964 | FALSE | TRUE | FALSE |
| ARFRP1 | 0.3295393 | 0.79343765 | 1 | 0.329539348 | 0.329539348 | 0.329539348 | 0.37569083 | 0.800611397 | 1 | 0.375690833 | 0.375690833 | 0.375690833 | 0.87715568 | -0.05692332 | FALSE | TRUE | FALSE |
| NCF1 | 0.3294504 | 0.79346251 | 1 | 3431216.52 | 428016172.4 | 58193024064 | 0.56514798 | 0.726523672 | 1 | 0.565147983 | 0.565147983 | 11.18020197 | 0.582945346 | -0.23437216 | FALSE | TRUE | FALSE |
| CCDC181 | 0.3293847 | 0.79348737 | 0.01375 | 0.329384746 | 0.329384746 | 0.748035516 | 4.10167379 | 0.350512321 | 0.5046 | 4.101673792 | 37.99204538 | 37.99204538 | 0.080304959 | -1.095257635 | FALSE | FALSE | FALSE |
| IFNGR1 | 0.3293209 | 0.79351222 | 1 | 0.329320858 | 0.329320858 | 6.529889959 | 0.37507308 | 0.800832548 | 1 | 0.375073081 | 0.375073081 | 7.444757637 | 0.878017845 | -0.056496657 | FALSE | TRUE | FALSE |
| PHKG1 | 0.3292444 | 0.79353706 | 0.75095 | 0.329244448 | 1.317054559 | 6.51545567 | 0.99506438 | 0.588894303 | 0.5064 | 0.995064376 | 0.995064376 | 2.16384367 | 0.330877535 | -0.480332719 | FALSE | TRUE | FALSE |
| DAPP1 | 0.3291996 | 0.7935619 | 1 | 3.061085271 | 3.061085271 | 3.061085271 | 0.38679296 | 0.796875753 | 1 | 0.386792964 | 0.386792964 | 0.386792964 | 0.851100263 | -0.070019275 | FALSE | TRUE | FALSE |
| TP53I13 | 0.3291852 | 0.79358673 | 1 | 0.329185189 | 0.329185189 | 6.527741938 | 0.37482962 | 0.800906154 | 1 | 0.374829621 | 0.374829621 | 0.374829621 | 0.878226188 | -0.056393617 | FALSE | TRUE | FALSE |
| DVL3 | 0.3290951 | 0.79361155 | 0.02655 | 0.329095112 | 0.329095112 | 6.443713736 | 0.37525834 | 0.800783448 | 1 | 0.375258341 | 0.375258341 | 7.366175063 | 0.876982804 | -0.057008922 | FALSE | TRUE | FALSE |
| FLRT3 | 0.3290697 | 0.79363637 | 1 | 0.329069659 | 0.329069659 | 6.501716387 | 0.37533546 | 0.800685171 | 1 | 0.37533546 | 0.37533546 | 7.426632332 | 0.876734799 | -0.057131755 | FALSE | TRUE | FALSE |
| VSTM4 | 0.3290658 | 0.79366118 | 1 | 0.329065798 | 0.329065798 | 0.329065798 | 0.37465146 | 0.800979703 | 0.00955 | 0.374651459 | 0.374651459 | 0.374651459 | 0.878325147 | -0.056344683 | FALSE | TRUE | FALSE |
| ZNF536 | 0.3288881 | 0.79368599 | 1 | 0.328888066 | 0.328888066 | 6.359312048 | 0.37375021 | 0.80110217 | 1 | 0.373750206 | 0.373750206 | 7.255707677 | 0.879967585 | -0.055533325 | FALSE | TRUE | FALSE |
| SCYL1 | 0.3288493 | 0.79371079 | 0.878 | 1.315383305 | 5.262791992 | 25.81844635 | 1.01817452 | 0.586442175 | 0.51265 | 1.018174518 | 1.018174518 | 20.02955967 | 0.322979349 | -0.490825245 | FALSE | FALSE | FALSE |
| LSM3 | 0.3288017 | 0.79373559 | 1 | 0.328801735 | 0.328801735 | 0.328801735 | 0.37417631 | 0.8010532 | 1 | 0.374176308 | 0.374176308 | 0.374176308 | 0.878734779 | -0.056142185 | FALSE | TRUE | FALSE |
| LYPD6 | 0.3285046 | 0.79376038 | 1 | 12.14610126 | 115.2314769 | 115.2314769 | 0.37817984 | 0.799870604 | 1 | 3.506686884 | 33.37228442 | 33.37228442 | 0.868646487 | -0.061156932 | FALSE | TRUE | FALSE |
| KIAA1958 | 0.3284504 | 0.79378516 | 1 | 3.060454233 | 3.060454233 | 60.29086488 | 0.38625341 | 0.797156445 | 1 | 0.386253413 | 0.386253413 | 7.622920727 | 0.850349538 | -0.07040252 | FALSE | TRUE | FALSE |
| PATL1 | 0.3284417 | 0.79380994 | 1 | 0.328441719 | 0.328441719 | 6.491129154 | 0.36293238 | 0.804682485 | 1 | 3.389941858 | 3.389941858 | 12.26888854 | 0.904966706 | -0.043367398 | FALSE | TRUE | FALSE |
| PTPN2 | 0.3280801 | 0.79383471 | 1 | 0.328080073 | 0.328080073 | 6.512488379 | 0.37436499 | 0.801028706 | 1 | 0.374364993 | 0.374364993 | 7.437946747 | 0.876364187 | -0.057315379 | FALSE | TRUE | FALSE |
| GFOD2 | 0.3280348 | 0.79385948 | 1 | 0.328034778 | 0.328034778 | 0.328034778 | 31.1061202 | 0.089618865 | 0.00155 | 3.262294472 | 31.10612024 | 31.10612024 | 0.010545667 | -1.976925956 | FALSE | FALSE | FALSE |
| TFDP2 | 0.3277809 | 0.79388424 | 1 | 0.327780889 | 0.327780889 | 0.327780889 | 3.17172581 | 0.401998627 | 0.8763 | 30.35743067 | 298.0142315 | 298.0142315 | 0.103344649 | -0.985712007 | FALSE | FALSE | FALSE |
| DPY19L1 | 0.3277044 | 0.79390899 | 1 | 0.327704357 | 0.327704357 | 6.481127895 | 3.28427049 | 0.395543651 | 0.2637 | 3.284270489 | 31.53196095 | 65.04119666 | 0.099779954 | -1.000956703 | FALSE | FALSE | FALSE |
| LXN | 0.3276975 | 0.79393374 | 0.0054 | 0.327697461 | 0.327697461 | 0.327697461 | 3.35174053 | 0.391931133 | 0.00515 | 3.35174053 | 3.35174053 | 3.35174053 | 0.097769341 | -1.009797314 | FALSE | FALSE | FALSE |
| TRIM4 | 0.327623 | 0.79395849 | 0.50655 | 0.327623031 | 0.327623031 | 0.744644653 | 4.07987385 | 0.351787803 | 0.5 | 2.91078166 | 37.77196811 | 37.77196811 | 0.080302245 | -1.095272312 | FALSE | FALSE | FALSE |
| TNFSF13 | 0.3274454 | 0.79398322 | 1 | 0.327445411 | 0.327445411 | 0.327445411 | 0.37365504 | 0.801151117 | 1 | 0.37365504 | 0.37365504 | 0.37365504 | 0.876330777 | -0.057331935 | FALSE | TRUE | FALSE |
| PPP5C | 0.3273592 | 0.79400796 | 1 | 0.327359185 | 0.327359185 | 6.460912906 | 0.37362983 | 0.801175581 | 1 | 0.373629829 | 0.373629829 | 7.38582445 | 0.876159128 | -0.05741701 | FALSE | TRUE | FALSE |
| DDTL | 0.3272933 | 0.79403268 | 1 | 3.020972182 | 3.020972182 | 3.020972182 | 0.38435348 | 0.797816691 | 1 | 0.384353478 | 0.384353478 | 0.384353478 | 0.851542318 | -0.069793764 | FALSE | TRUE | FALSE |
| DNAJC7 | 0.3272825 | 0.7940574 | 1 | 0.327282488 | 0.327282488 | 6.482291587 | 0.37357115 | 0.801200038 | 0.01195 | 0.373571151 | 0.373571151 | 7.407718545 | 0.876091442 | -0.057450562 | FALSE | TRUE | FALSE |
| KCTD7 | 0.3272271 | 0.79408212 | 1 | 0.327227144 | 0.327227144 | 6.492914771 | 0.37333291 | 0.801248935 | 0.0101 | 0.373332913 | 0.373332913 | 7.414783471 | 0.876502266 | -0.057246957 | FALSE | TRUE | FALSE |
| OR2C3 | 0.3272154 | 0.79410682 | 1 | 0.327215428 | 0.327215428 | 0.327215428 | 0.37267196 | 0.801468672 | 1 | 0.372671955 | 0.372671955 | 0.372671955 | 0.87802536 | -0.05649294 | FALSE | TRUE | FALSE |
| PARK7 | 0.3271895 | 0.79413153 | 1 | 1.308409565 | 1.308409565 | 1.308409565 | 1.47688139 | 0.538012309 | 0.75215 | 1.476881389 | 5.907592854 | 5.907592854 | 0.221540806 | -0.654546268 | FALSE | FALSE | FALSE |
| ADAM23 | 0.3271623 | 0.79415622 | 1 | 3.062709804 | 3.062709804 | 60.34108236 | 0.38518054 | 0.797588647 | 0.01975 | 0.385180542 | 0.385180542 | 7.602389243 | 0.8493738 | -0.070901139 | FALSE | TRUE | FALSE |
| TEX29 | 0.3271612 | 0.79418091 | 1 | 0.327161237 | 0.327161237 | 0.327161237 | 0.37248599 | 0.801590528 | 1 | 0.372485987 | 0.372485987 | 0.372485987 | 0.87831824 | -0.056348098 | FALSE | TRUE | FALSE |
| PSMD1 | 0.3271492 | 0.79420559 | 0.02415 | 0.327149188 | 0.327149188 | 6.431734163 | 0.37355647 | 0.801224489 | 1 | 0.373556469 | 0.373556469 | 7.359131878 | 0.875769036 | -0.057610414 | FALSE | TRUE | FALSE |
| OR4K15 | 0.3271333 | 0.79423027 | 1 | 0.327133312 | 0.327133312 | 0.327133312 | 0.37267069 | 0.801493056 | 1 | 0.372670693 | 0.372670693 | 0.372670693 | 0.87780799 | -0.056600471 | FALSE | TRUE | FALSE |
| NRROS | 0.3271267 | 0.79425494 | 1 | 0.327126667 | 0.327126667 | 6.425715533 | 0.37324242 | 0.801297807 | 1 | 0.373242417 | 0.373242417 | 7.347323921 | 0.876445582 | -0.057275044 | FALSE | TRUE | FALSE |
| DBF4 | 0.3270308 | 0.7942796 | 1 | 3.051447718 | 3.051447718 | 6.475191326 | 0.38486268 | 0.797690064 | 1 | 0.384862683 | 0.384862683 | 7.629427738 | 0.849733789 | -0.070717112 | FALSE | TRUE | FALSE |
| DAP | 0.3269664 | 0.79430426 | 0.9812 | 292.4023326 | 324984.2012 | 3002617.637 | 3.61929793 | 0.377565237 | 0.98055 | 3530.25638 | 391004.6622 | 3594066.701 | 0.09033972 | -1.04412126 | FALSE | FALSE | FALSE |
| TMEM163 | 0.3269186 | 0.79432891 | 1 | 0.326918587 | 0.326918587 | 0.326918587 | 0.33954125 | 0.812205536 | 1 | 0.339541249 | 0.339541249 | 0.339541249 | 0.962824364 | -0.016452928 | FALSE | TRUE | FALSE |

| | | | | | | | | | | | | | | | | | |
|---|---|---|---|---|---|---|---|---|---|---|---|---|---|---|---|---|---|
| SLC13A4 | 0.326802 | 0.79435355 | 1 | 0.326801987 | 0.326801987 | 6.455662142 | 0.37263579 | 0.801517433 | 1 | 0.372635792 | 0.372635792 | 7.371957324 | 0.877001065 | -0.056999879 | FALSE | TRUE | FALSE |
| GCNT1 | 0.3267698 | 0.79437819 | 0.0113 | 0.326769771 | 0.326769771 | 6.480561673 | 0.37235467 | 0.801712224 | 1 | 0.372354673 | 0.372354673 | 7.392061856 | 0.877576662 | -0.056714935 | FALSE | TRUE | FALSE |
| ISG15 | 0.3267224 | 0.79440283 | 1 | 0.326722445 | 0.326722445 | 0.326722445 | 3.35732123 | 0.391704119 | 0.0054 | 3.357321233 | 3.357321233 | 3.357321233 | 0.097316409 | -1.011813927 | FALSE | FALSE | FALSE |
| SPATS2 | 0.3266918 | 0.79442745 | 1 | 0.326691753 | 0.326691753 | 6.465463881 | 0.35013172 | 0.809061254 | 1 | 3.267771971 | 31.3011236 | 64.75295709 | 0.933053853 | -0.030093289 | FALSE | TRUE | FALSE |
| HDAC3 | 0.3266567 | 0.79445208 | 1 | 0.326656741 | 0.326656741 | 6.468907667 | 0.37287025 | 0.801371072 | 0.0126 | 0.372870248 | 0.372870248 | 7.392823277 | 0.876060084 | -0.057466107 | FALSE | TRUE | FALSE |
| SPSB3 | 0.3266459 | 0.79447669 | 0.0146 | 0.326645853 | 0.326645853 | 6.459757644 | 0.37252618 | 0.80156617 | 0.01145 | 0.372526179 | 0.372526179 | 7.377013658 | 0.876839996 | -0.057079649 | FALSE | TRUE | FALSE |
| PDGFD | 0.3266284 | 0.7945013 | 1 | 0.326628364 | 0.326628364 | 6.479729502 | 3.36749915 | 0.391022286 | 0.0112 | 3.367499149 | 3.367499149 | 7.169587415 | 0.096994342 | -1.013253599 | FALSE | FALSE | FALSE |
| HAPLN2 | 0.3264697 | 0.7945259 | 1 | 1.305847015 | 1.305847015 | 6.466413398 | 3.42311294 | 0.387592425 | 0.0108 | 3.423112937 | 3.423112937 | 7.295212049 | 0.09537217 | -1.020578336 | FALSE | FALSE | FALSE |
| TMEM178A | 0.3264368 | 0.7945505 | 0.0093 | 0.326436829 | 0.326436829 | 0.326436829 | 0.34368087 | 0.810956053 | 1 | 0.34368087 | 0.34368087 | 0.34368087 | 0.949825428 | -0.022356208 | FALSE | TRUE | FALSE |
| ESYT3 | 0.3263987 | 0.79457509 | 0.91535 | 20.93327688 | 957.9125217 | 8630.748758 | 345.795996 | 0.007020082 | 0.7516 | 5547.89505 | 3316894.935 | 13303663.1 | 0.000943905 | -3.025071583 | FALSE | FALSE | FALSE |
| MLANA | 0.3263118 | 0.79459967 | 1 | 0.326311832 | 0.326311832 | 0.326311832 | 0.37155925 | 0.801882348 | 1 | 0.371559249 | 0.371559249 | 0.371559249 | 0.878222877 | -0.056395254 | FALSE | TRUE | FALSE |
| SRD5A1 | 0.3262622 | 0.79462425 | 1 | 0.326262248 | 0.326262248 | 0.326262248 | 0.37182665 | 0.80183377 | 1 | 0.371826645 | 0.371826645 | 0.371826645 | 0.877457956 | -0.056773684 | FALSE | TRUE | FALSE |
| BLOC1S6 | 0.3261389 | 0.79464883 | 1 | 0.326138912 | 0.326138912 | 0.326138912 | 0.37150172 | 0.801906628 | 1 | 0.371501724 | 0.371501724 | 0.371501724 | 0.877893402 | -0.056558215 | FALSE | TRUE | FALSE |
| GCSAM | 0.3260953 | 0.79467339 | 1 | 0.32609526 | 0.32609526 | 0.32609526 | 0.37141218 | 0.801955167 | 1 | 0.371412184 | 0.371412184 | 0.371412184 | 0.877987514 | -0.05651166 | FALSE | TRUE | FALSE |
| OR4L1 | 0.3260687 | 0.79469795 | 1 | 0.326068732 | 0.326068732 | 0.326068732 | 0.37183547 | 0.801809473 | 0.0066 | 0.371835465 | 0.371835465 | 0.371835465 | 0.876916707 | -0.057041656 | FALSE | TRUE | FALSE |
| OR5H14 | 0.3256644 | 0.79472251 | 0.74805 | 0.325664406 | 3.090572092 | 3.090572092 | 3.42017366 | 0.388051807 | 0.1316 | 0.359609316 | 33.36340225 | 33.36340225 | 0.095218676 | -1.021277864 | FALSE | FALSE | FALSE |
| MEGF8 | 0.3256212 | 0.79474706 | 0.298 | 0.142947851 | 1.301961475 | 24.36436634 | 0.02932022 | 0.891435304 | 0.97165 | 1.0477201 | 80.1762575 | 320.6702723 | 11.1056891 | 1.045545512 | FALSE | TRUE | FALSE |
| MSMO1 | 0.3253673 | 0.79477161 | 0.0067 | 0.325367269 | 0.325367269 | 0.325367269 | 0.371577 | 0.801858062 | 1 | 0.371577002 | 0.371577002 | 0.371577002 | 0.875638878 | -0.057674964 | FALSE | TRUE | FALSE |
| YAP1 | 0.325167 | 0.79479615 | 1 | 3.031257761 | 29.00204198 | 60.02211563 | 0.39504694 | 0.793839583 | 1 | 0.395046938 | 0.395046938 | 7.831707485 | 0.823109808 | -0.084542224 | FALSE | TRUE | FALSE |
| SLC9A1 | 0.3250717 | 0.79482068 | 1 | 0.325071667 | 0.325071667 | 6.369978968 | 0.37115002 | 0.802076408 | 1 | 0.371150023 | 0.371150023 | 7.290623324 | 0.875849781 | -0.057570374 | FALSE | TRUE | FALSE |
| SSTR1 | 0.3250596 | 0.79484521 | 1 | 0.325059632 | 0.325059632 | 6.416551733 | 0.34856879 | 0.809555341 | 1 | 3.259181484 | 31.27633057 | 64.43577531 | 0.932555172 | -0.030325465 | FALSE | TRUE | FALSE |
| ZBTB5 | 0.3250398 | 0.79486974 | 1 | 0.32503978 | 0.32503978 | 6.410734904 | 0.36684789 | 0.80342423 | 1 | 1.467353738 | 1.467353738 | 12.42285591 | 0.886034216 | -0.052549506 | FALSE | TRUE | FALSE |
| SST | 0.3249965 | 0.79489425 | 1 | 0.324996542 | 0.324996542 | 0.324996542 | 0.33019884 | 0.815330579 | 1 | 0.330198838 | 0.330198838 | 0.330198838 | 0.98424496 | -0.0068968 | FALSE | TRUE | FALSE |
| NDUFAF5 | 0.3249339 | 0.79491876 | 0.2575 | 0.081258425 | 0.324933859 | 0.324933859 | 0.03163179 | 0.890837094 | 1 | 0.50573404 | 2.022945611 | 2.022945611 | 10.27238282 | 1.011671196 | FALSE | TRUE | FALSE |
| WNT5A | 0.3247954 | 0.79494327 | 1 | 1.298850999 | 1.298850999 | 6.431732046 | 1.48450988 | 0.537463606 | 0.5087 | 1.484509878 | 1.484509878 | 7.359604345 | 0.218789657 | -0.659973212 | FALSE | FALSE | FALSE |
| MRPL10 | 0.3246687 | 0.79496777 | 1 | 0.324668686 | 0.324668686 | 0.324668686 | 0.34519797 | 0.810668742 | 1 | 0.34519797 | 0.34519797 | 0.34519797 | 0.940528954 | -0.02662783 | FALSE | TRUE | FALSE |
| PRDM11 | 0.3246196 | 0.79499226 | 1 | 0.324619599 | 0.324619599 | 6.424900812 | 0.37091762 | 0.802124862 | 1 | 0.370917617 | 0.370917617 | 7.350415751 | 0.875179781 | -0.057902724 | FALSE | TRUE | FALSE |
| PAFAH1B2 | 0.3245498 | 0.79501675 | 1 | 0.324549836 | 0.324549836 | 0.324549836 | 3.33002395 | 0.393064062 | 0.00565 | 3.330023949 | 3.330023949 | 3.330023949 | 0.097461712 | -1.011165963 | FALSE | FALSE | FALSE |
| URGCP | 0.3243494 | 0.79504123 | 0.02525 | 0.324349427 | 0.324349427 | 6.355148758 | 0.37060808 | 0.802173294 | 1 | 0.370608076 | 0.370608076 | 7.279293241 | 0.875181757 | -0.057901744 | FALSE | TRUE | FALSE |
| COMMD7 | 0.3243114 | 0.79506571 | 1 | 2.991563991 | 2.991563991 | 2.991563991 | 0.38119379 | 0.798824029 | 0.00325 | 0.381193786 | 0.381193786 | 0.381193786 | 0.850778289 | -0.070183601 | FALSE | TRUE | FALSE |
| CNN1 | 0.3243016 | 0.79509018 | 1 | 0.32430165 | 0.32430165 | 0.32430165 | 1.46530309 | 0.538559868 | 0.00995 | 1.465303091 | 1.465303091 | 1.465303091 | 0.221320525 | -0.654978308 | FALSE | FALSE | FALSE |
| KCNS1 | 0.3241842 | 0.79511464 | 0.51495 | 0.324184226 | 1.296069227 | 1.600599878 | 4.67593249 | 0.320928098 | 0.25625 | 1.169737723 | 18.69633596 | 18.69633596 | 0.069330391 | -1.159076353 | FALSE | FALSE | FALSE |
| ZNF513 | 0.3239174 | 0.7951391 | 1 | 3.023611469 | 3.023611469 | 10.96527615 | 0.35852702 | 0.806180265 | 1 | 3.350390873 | 32.13338493 | 66.1575103 | 0.903467122 | -0.044087647 | FALSE | TRUE | FALSE |
| GEMIN2 | 0.3238528 | 0.79516356 | 1 | 0.323852789 | 0.323852789 | 0.323852789 | 0.3696817 | 0.802270094 | 1 | 0.369681696 | 0.369681696 | 0.369681696 | 0.87603144 | -0.057480307 | FALSE | TRUE | FALSE |
| ARRDC5 | 0.3236135 | 0.79518801 | 0.00965 | 0.323613549 | 0.323613549 | 0.323613549 | 0.36913799 | 0.802439281 | 1 | 0.369137995 | 0.369137995 | 7.331157716 | 0.87667364 | -0.057162052 | FALSE | TRUE | FALSE |
| OR14A16 | 0.3234341 | 0.79521245 | 1 | 0.323434125 | 0.323434125 | 0.323434125 | 0.32591633 | 0.816791832 | 1 | 28.57456703 | 2772.624073 | 2772.624073 | 0.992383915 | -0.003320283 | FALSE | TRUE | FALSE |
| CRLF3 | 0.3234078 | 0.79523689 | 1 | 3.002431096 | 3.002431096 | 6.409989907 | 30.735552 | 0.090489312 | 0.50645 | 30.73555195 | 300.6208366 | 300.6208366 | 0.010522269 | -1.977890582 | FALSE | FALSE | FALSE |
| KAT7 | 0.3233519 | 0.79526132 | 1 | 0.3233519 | 0.3233519 | 6.383689603 | 31.1713465 | 0.089329866 | 0.25255 | 3.24636074 | 31.17134647 | 64.18981507 | 0.01037337 | -1.984080145 | FALSE | FALSE | FALSE |
| MTHFSD | 0.3232986 | 0.79528575 | 0.50615 | 0.323298597 | 0.323298597 | 0.727135623 | 0.4217514 | 0.783879943 | 0.81525 | 35.79693661 | 342.3730225 | 3354.621776 | 0.766562001 | -0.115452713 | FALSE | TRUE | FALSE |
| NAA25 | 0.3232671 | 0.79531017 | 1 | 0.323267103 | 0.323267103 | 6.357774866 | 0.36940005 | 0.80234264 | 0.0211 | 0.369400053 | 0.369400053 | 7.279634432 | 0.87511385 | -0.057935443 | FALSE | TRUE | FALSE |
| HSD11B1 | 0.3232346 | 0.79533458 | 1 | 0.323234621 | 0.323234621 | 0.323234621 | 0.36933549 | 0.802366809 | 1 | 0.369335489 | 0.369335489 | 0.369335489 | 0.875178882 | -0.05790317 | FALSE | TRUE | FALSE |
| OR7G2 | 0.323016 | 0.79535899 | 0.0081 | 0.323016003 | 0.323016003 | 0.323016003 | 0.36858264 | 0.802656364 | 1 | 0.368582637 | 0.368582637 | 0.368582637 | 0.876373357 | -0.057310834 | FALSE | TRUE | FALSE |
| OR5C1 | 0.322879 | 0.79538339 | 1 | 3.000267338 | 3.000267338 | 6.405242829 | 3.32573717 | 0.393516148 | 0.753 | 3.325737169 | 31.76905248 | 31.76905248 | 0.097084941 | -1.01284813 | FALSE | FALSE | FALSE |
| BHLHE40 | 0.3228441 | 0.79540779 | 1 | 0.322844126 | 0.322844126 | 6.394905548 | 0.36907387 | 0.802487565 | 1 | 0.369073873 | 0.369073873 | 0.369073873 | 0.874741209 | -0.058120413 | FALSE | TRUE | FALSE |
| IPCEF1 | 0.3227542 | 0.79543218 | 1 | 0.322754224 | 0.322754224 | 6.404927528 | 0.34919855 | 0.809375972 | 1 | 0.349198548 | 0.349198548 | 6.936179488 | 0.92427138 | -0.034200495 | FALSE | TRUE | FALSE |
| MRPS2 | 0.3227226 | 0.79545657 | 1 | 0.322722617 | 0.322722617 | 6.396027991 | 0.36838741 | 0.802752688 | 1 | 0.368387408 | 0.368387408 | 7.309005569 | 0.876041389 | -0.057475375 | FALSE | TRUE | FALSE |
| DDX42 | 0.3225084 | 0.79548095 | 1 | 0.322508425 | 0.322508425 | 6.322505949 | 0.36870399 | 0.802632267 | 1 | 0.368703992 | 0.368703992 | 7.245359965 | 0.874708255 | -0.058136775 | FALSE | TRUE | FALSE |
| CCNO | 0.3224995 | 0.79550532 | 1 | 0.322499494 | 0.322499494 | 6.391368175 | 0.36816775 | 0.802824867 | 1 | 0.368167747 | 0.368167747 | 7.304408045 | 0.875958028 | -0.057516703 | FALSE | TRUE | FALSE |
| TRIM43 | 0.3224982 | 0.79552969 | 0.01085 | 0.322498212 | 0.322498212 | 6.399469269 | 0.33765465 | 0.812769985 | 1 | 0.337654651 | 0.337654651 | 0.337654651 | 0.9551126 | -0.019945425 | FALSE | TRUE | FALSE |
| CBFA2T2 | 0.3224805 | 0.79555405 | 0.01945 | 0.32248055 | 0.32248055 | 6.360617815 | 0.36838465 | 0.802776753 | 1 | 0.368384649 | 0.368384649 | 7.278097394 | 0.875390846 | -0.057797999 | FALSE | TRUE | FALSE |
| UNC45B | 0.3224477 | 0.79557841 | 0.14225 | 0.146800634 | 1.289522971 | 6.319283914 | 0.01662102 | 0.894708597 | 0.0247 | 0.016621025 | 0.016621025 | 0.326525311 | 19.3999867 | 1.287801432 | FALSE | TRUE | FALSE |
| PLD5 | 0.322326 | 0.79560276 | 0.0115 | 0.322325995 | 0.322325995 | 6.39280977 | 0.36794672 | 0.802896992 | 1 | 0.367946719 | 0.367946719 | 7.304934592 | 0.876012689 | -0.057489603 | FALSE | TRUE | FALSE |
| ASAH2B | 0.3222662 | 0.7956271 | 1 | 0.322266162 | 0.322266162 | 0.322266162 | 0.3311196 | 0.815056604 | 1 | 0.331119601 | 0.331119601 | 0.331119601 | 0.973262111 | -0.011770183 | FALSE | TRUE | FALSE |
| ENG | 0.3222575 | 0.79565144 | 1 | 0.322257482 | 0.322257482 | 6.352802724 | 0.36848545 | 0.802704538 | 1 | 0.368485448 | 0.368485448 | 7.276641197 | 0.874546019 | -0.058217333 | FALSE | TRUE | FALSE |
| CD3D | 0.3222363 | 0.79567577 | 1 | 0.322236318 | 0.322236318 | 0.322236318 | 0.36770209 | 0.802969063 | 1 | 0.367702087 | 0.367702087 | 0.367702087 | 0.876351615 | -0.057321609 | FALSE | TRUE | FALSE |
| NEK6 | 0.322175 | 0.7957001 | 1 | 0.322174988 | 0.322174988 | 6.390952626 | 0.3679152 | 0.802921021 | 1 | 0.367915204 | 0.367915204 | 0.367915204 | 0.875677287 | -0.057655915 | FALSE | TRUE | FALSE |
| OXTR | 0.322007 | 0.79572442 | 1 | 0.322007048 | 0.322007048 | 6.360699216 | 0.36811064 | 0.802848915 | 1 | 0.368110638 | 0.368110638 | 7.282189317 | 0.87475616 | -0.05811299 | FALSE | TRUE | FALSE |
| SYF2 | 0.321897 | 0.79574874 | 0.00805 | 0.321896966 | 0.321896966 | 0.321896966 | 0.36748252 | 0.803160981 | 1 | 0.367482515 | 0.367482515 | 0.367482515 | 0.875951787 | -0.057519797 | FALSE | TRUE | FALSE |
| WASF2 | 0.321813 | 0.79577305 | 1 | 0.321813026 | 0.321813026 | 6.366871784 | 0.35659184 | 0.80685121 | 1 | 3.350892862 | 3.350892862 | 7.064098923 | 0.902468857 | -0.044567776 | FALSE | TRUE | FALSE |
| FBXO47 | 0.3218053 | 0.79579735 | 1 | 0.321805316 | 0.321805316 | 6.383096586 | 0.36750991 | 0.803113039 | 1 | 0.367509911 | 0.367509911 | 7.296880356 | 0.875637109 | -0.057675842 | FALSE | TRUE | FALSE |
| LRRC20 | 0.3217472 | 0.79582165 | 0.50255 | 2.972032765 | 2.972032765 | 2.972032765 | 0.35593482 | 0.807150433 | 0.7543 | 3.295414676 | 31.31414718 | 31.31414718 | 0.903949904 | -0.043855637 | FALSE | TRUE | FALSE |
| TIMP2 | 0.3217223 | 0.79584594 | 1 | 1.286499212 | 1.286499212 | 1.286499212 | 0.37219397 | 0.801736545 | 1 | 0.372193973 | 0.372193973 | 0.372193973 | 0.864394158 | -0.063288177 | FALSE | TRUE | FALSE |
| TOMM34 | 0.3215136 | 0.79587022 | 1 | 0.321513636 | 0.321513636 | 0.321513636 | 0.36756963 | 0.803089058 | 1 | 0.367569628 | 0.367569628 | 0.367569628 | 0.874701311 | -0.058140222 | FALSE | TRUE | FALSE |
| ZCCHC8 | 0.3215059 | 0.7958945 | 1 | 3.005280877 | 3.005280877 | 10.87213921 | 3.44181994 | 0.38644089 | 0.5038 | 3.441819939 | 3.441819939 | 7.271878821 | 0.093411589 | -1.029599239 | FALSE | FALSE | FALSE |
| DEFB125 | 0.321486 | 0.79591878 | 1 | 0.321486031 | 0.321486031 | 0.321486031 | 0.3668951 | 0.803376422 | 1 | 0.366895105 | 0.366895105 | 0.366895105 | 0.87623418 | -0.05737981 | FALSE | TRUE | FALSE |
| DCTN5 | 0.3214769 | 0.79594304 | 1 | 2.989946703 | 28.54088261 | 28.54088261 | 0.39078799 | 0.79538231 | 1 | 0.390787992 | 0.390787992 | 0.390787992 | 0.82263762 | -0.084791433 | FALSE | TRUE | FALSE |
| STX12 | 0.3214391 | 0.7959673 | 1 | 3.004216866 | 28.81721961 | 28.81721961 | 0.35025855 | 0.809038734 | 0.75125 | 3.279704187 | 31.51862463 | 31.51862463 | 0.917719449 | -0.037290064 | FALSE | TRUE | FALSE |
| RHOC | 0.3214197 | 0.79599156 | 1 | 0.321419656 | 0.321419656 | 0.321419656 | 0.35603029 | 0.807081468 | 0.507 | 3.328691467 | 3.328691467 | 3.328691467 | 0.902787381 | -0.04441452 | FALSE | TRUE | FALSE |
| TNFSF14 | 0.3213666 | 0.79601581 | 1 | 0.321366603 | 0.321366603 | 0.321366603 | 0.36766558 | 0.80301708 | 1 | 0.367665578 | 0.367665578 | 0.367665578 | 0.874073132 | -0.058452229 | FALSE | TRUE | FALSE |
| IMMP1L | 0.3213141 | 0.79604005 | 1 | 0.321314141 | 0.321314141 | 0.321314141 | 1.45108596 | 0.540331565 | 0.50385 | 1.451085959 | 1.451085959 | 1.451085959 | 0.221430122 | -0.654763301 | FALSE | FALSE | FALSE |
| FCHSD2 | 0.3212947 | 0.79606429 | 1 | 2.995769795 | 2.995769795 | 59.12509426 | 0.33441734 | 0.814165886 | 0.9388 | 29.80496265 | 2939.77367 | 2939.77367 | 0.960759597 | -0.017385269 | FALSE | TRUE | FALSE |
| TNFRSF1B | 0.3212247 | 0.79608852 | 1 | 0.321224717 | 0.321224717 | 6.357407535 | 0.36747019 | 0.803184942 | 1 | 0.367470194 | 0.367470194 | 7.281794538 | 0.874151758 | -0.058413164 | FALSE | TRUE | FALSE |
| CAMK2D | 0.3211428 | 0.79611275 | 1 | 0.32114284 | 0.32114284 | 6.365966618 | 0.36696127 | 0.803328588 | 1 | 0.36696127 | 0.36696127 | 7.28196781 | 0.875140965 | -0.057921986 | FALSE | TRUE | FALSE |

| | | | | | | | | | | | | | | | | | |
|---|---|---|---|---|---|---|---|---|---|---|---|---|---|---|---|---|---|
| EIF3K | 0.3210979 | 0.79613697 | 1 | 2.979741589 | 2.979741589 | 2.979741589 | 1.49616365 | 0.536363171 | 0.5038 | 1.496163649 | 1.496163649 | 1.496163649 | 0.214614153 | -0.668341641 | FALSE | FALSE | FALSE |
| VPS9D1 | 0.3210398 | 0.79616118 | 0.75965 | 0.321039778 | 2.882361103 | 6.347246573 | 0.05667944 | 0.885359505 | 1 | 0.05667944 | 0.05667944 | 1.122148089 | 5.664131108 | 0.753133297 | FALSE | TRUE | FALSE |
| C6orf120 | 0.3210351 | 0.79618539 | 1 | 3.002003611 | 12.00636027 | 12.00636027 | 0.38297497 | 0.798094728 | 1 | 0.382974973 | 0.382974973 | 0.382974973 | 0.838266453 | -0.076617914 | FALSE | TRUE | FALSE |
| BECN1 | 0.3208095 | 0.79620959 | 1 | 0.320809519 | 0.320809519 | 6.357397703 | 0.36681311 | 0.803448126 | 0.0111 | 0.366813115 | 0.366813115 | 7.277050187 | 0.874585738 | -0.058197609 | FALSE | TRUE | FALSE |
| TRAF2 | 0.3208019 | 0.79623379 | 1 | 0.320801909 | 0.320801909 | 6.32794894 | 0.36686 | 0.803400329 | 1 | 0.366860002 | 0.366860002 | 7.248420142 | 0.874453219 | -0.058263419 | FALSE | TRUE | FALSE |
| SP4 | 0.3205917 | 0.79625798 | 1 | 0.320591659 | 0.320591659 | 6.330376838 | 0.35748919 | 0.806527902 | 1 | 0.357489192 | 0.357489192 | 7.069743248 | 0.896786996 | -0.047310698 | FALSE | TRUE | FALSE |
| POFUT1 | 0.3204501 | 0.79628217 | 1 | 0.320450094 | 0.320450094 | 6.349760642 | 0.36648761 | 0.803615219 | 1 | 0.366487605 | 0.366487605 | 7.270073733 | 0.874381806 | -0.058298887 | FALSE | TRUE | FALSE |
| CCNL1 | 0.3203775 | 0.79630635 | 1 | 0.320377461 | 0.320377461 | 6.339223123 | 0.36656268 | 0.803567509 | 1 | 0.366562685 | 0.366562685 | 7.262381958 | 0.874004569 | -0.058486297 | FALSE | TRUE | FALSE |
| PCIF1 | 0.3203719 | 0.79633053 | 1 | 0.320371893 | 0.320371893 | 6.289931435 | 0.36670009 | 0.803495897 | 1 | 0.366700091 | 0.366700091 | 7.215401035 | 0.873661887 | -0.058656609 | FALSE | TRUE | FALSE |
| NCBP1 | 0.3203267 | 0.79635469 | 1 | 0.320326659 | 0.320326659 | 6.308429539 | 0.36640241 | 0.803639066 | 1 | 0.366402411 | 0.366402411 | 7.229133395 | 0.874248228 | -0.058365239 | FALSE | TRUE | FALSE |
| HDGF | 0.3201625 | 0.79637886 | 0.00875 | 0.320162453 | 0.320162453 | 0.320162453 | 0.3656177 | 0.803900988 | 1 | 0.3656177 | 0.3656177 | 0.3656177 | 0.875675476 | -0.057656813 | FALSE | TRUE | FALSE |
| RNASEH1 | 0.3201539 | 0.79640301 | 1 | 0.32015385 | 0.32015385 | 0.32015385 | 0.3657868 | 0.803853419 | 1 | 0.3657868 | 0.3657868 | 0.3657868 | 0.87524714 | -0.0578693 | FALSE | TRUE | FALSE |
| SHARPIN | 0.320099 | 0.79642717 | 1 | 0.320099028 | 0.320099028 | 6.354698432 | 3.302385 | 0.394869077 | 0.50475 | 3.302384996 | 3.302384996 | 3.302384996 | 0.096929652 | -1.013543347 | FALSE | FALSE | FALSE |
| BRD7 | 0.3200372 | 0.79645131 | 1 | 0.320037152 | 0.320037152 | 6.321868076 | 0.36596608 | 0.803782022 | 1 | 0.365966079 | 0.365966079 | 7.239845879 | 0.874499496 | -0.058240436 | FALSE | TRUE | FALSE |
| FOLR2 | 0.3200245 | 0.79647545 | 0.74965 | 0.320024453 | 1.279996864 | 1.279996864 | 0.10992924 | 0.873359914 | 1 | 0.109929241 | 0.109929241 | 0.109929241 | 2.911185883 | 0.464069937 | FALSE | TRUE | FALSE |
| BAIAP2 | 0.3199728 | 0.79649958 | 0.0193 | 0.319972787 | 0.319972787 | 6.307183138 | 3.31024845 | 0.393967821 | 0.01385 | 3.310248451 | 3.310248451 | 7.002558123 | 0.09666126 | -1.014747547 | FALSE | FALSE | FALSE |
| INSL4 | 0.3198847 | 0.79652371 | 1 | 2.957806746 | 2.957806746 | 2.957806746 | 0.36527867 | 0.803996057 | 1 | 0.365278671 | 3.387993229 | 3.387993229 | 0.875727738 | -0.057630894 | FALSE | TRUE | FALSE |
| CPSF3 | 0.3198473 | 0.79654783 | 0.7594 | 0.319847299 | 2.91898951 | 6.315182605 | 0.05438425 | 0.885804102 | 0.51055 | 0.48760471 | 0.48760471 | 1.840685488 | 5.88124867 | 0.769469543 | FALSE | TRUE | FALSE |
| NUDCD2 | 0.3198005 | 0.79657195 | 1 | 0.319800467 | 0.319800467 | 0.319800467 | 0.36514677 | 0.804138475 | 1 | 0.365146768 | 0.365146768 | 0.365146768 | 0.875813495 | -0.057588367 | FALSE | TRUE | FALSE |
| ZNF217 | 0.3197316 | 0.79659606 | 1 | 285.7317884 | 2894.062547 | 30021.81252 | 0.36588608 | 0.803805827 | 0.96925 | 328.3253919 | 3330.019176 | 34590.81964 | 0.873855505 | -0.058560373 | FALSE | TRUE | FALSE |
| LYPD4 | 0.3197192 | 0.79662017 | 1 | 0.319719213 | 0.319719213 | 0.319719213 | 0.365183 | 0.804091027 | 0.00475 | 0.365182996 | 0.365182996 | 0.365182996 | 0.875504106 | -0.057741813 | FALSE | TRUE | FALSE |
| TMEM86B | 0.3196037 | 0.79664426 | 1 | 28.09527599 | 273.7139873 | 273.7139873 | 3.61281233 | 0.377801532 | 0.50785 | 3.61281233 | 3.61281233 | 3.61281233 | 0.088463975 | -1.053233551 | FALSE | FALSE | FALSE |
| TPGS1 | 0.3195242 | 0.79666836 | 1 | 0.319524231 | 0.319524231 | 0.319524231 | 0.33794614 | 0.812661703 | 1 | 3.147876816 | 3.147876816 | 3.147876816 | 0.945488637 | -0.024343686 | FALSE | TRUE | FALSE |
| NTF4 | 0.3195241 | 0.79669244 | 1 | 0.319524148 | 0.319524148 | 0.319524148 | 3.34037316 | 0.392384766 | 0.0055 | 3.340373164 | 3.340373164 | 3.340373164 | 0.095655225 | -1.019291301 | FALSE | FALSE | FALSE |
| GUCA1B | 0.3194529 | 0.79671652 | 0.00685 | 0.319452943 | 0.319452943 | 0.319452943 | 3.2942151 | 0.39509405 | 0.00525 | 3.294215097 | 3.294215097 | 3.294215097 | 0.096973917 | -1.01334506 | FALSE | FALSE | FALSE |
| TAGLN3 | 0.3193896 | 0.7967406 | 1 | 0.319389564 | 0.319389564 | 0.319389564 | 0.36541947 | 0.803948534 | 1 | 0.365419468 | 0.365419468 | 0.365419468 | 0.874035436 | -0.058470959 | FALSE | TRUE | FALSE |
| CDK5R1 | 0.3193802 | 0.79676467 | 1 | 0.319380194 | 0.319380194 | 6.329764881 | 0.36162442 | 0.805106122 | 1 | 1.446146514 | 1.446146514 | 1.446146514 | 0.883182052 | -0.053949765 | FALSE | TRUE | FALSE |
| F12 | 0.3193342 | 0.79678873 | 0.5076 | 0.319334184 | 1.277212389 | 6.267095822 | 0.03266587 | 0.890667299 | 1 | 0.03266587 | 0.03266587 | 0.642529792 | 9.775774577 | 0.990151178 | FALSE | TRUE | FALSE |
| SYT4 | 0.3191347 | 0.79681279 | 1 | 0.319134688 | 0.319134688 | 6.337404499 | 0.3650848 | 0.80416219 | 0.00875 | 0.3650848 | 0.3650848 | 0.3650848 | 0.874138521 | -0.058419741 | FALSE | TRUE | FALSE |
| PGC | 0.3190735 | 0.79683684 | 0.01275 | 0.319073544 | 0.319073544 | 0.713279242 | 0.05433136 | 0.885812461 | 1 | 0.054331359 | 0.054331359 | 1.078633682 | 5.872732568 | 0.768840224 | FALSE | TRUE | FALSE |
| APOLD1 | 0.3190725 | 0.79686089 | 0.7487 | 2.946652629 | 27.92940578 | 27.92940578 | 3.48560986 | 0.383428703 | 0.5016 | 3.485609858 | 3.485609858 | 3.485609858 | 0.091539949 | -1.038389333 | FALSE | FALSE | FALSE |
| RNF149 | 0.3190545 | 0.79688493 | 0.50845 | 1.276162179 | 1.276162179 | 6.311647214 | 0.36898028 | 0.802559943 | 1 | 0.368980282 | 0.368980282 | 7.308852566 | 0.864692555 | -0.06313828 | FALSE | TRUE | FALSE |
| GTF2H5 | 0.3189452 | 0.79690896 | 1 | 0.318945199 | 0.318945199 | 0.318945199 | 0.36466015 | 0.804256995 | 1 | 0.364660155 | 0.364660155 | 0.364660155 | 0.874636823 | -0.058172242 | FALSE | TRUE | FALSE |
| CCT7 | 0.318858 | 0.79693299 | 0.2597 | 0.318858021 | 1.274892956 | 2.702461536 | 0.10887317 | 0.873652261 | 0.50795 | 0.435235912 | 0.435235912 | 2.157508152 | 2.928710855 | 0.466676497 | FALSE | TRUE | FALSE |
| MFAP4 | 0.3185613 | 0.79695702 | 1 | 0.318561338 | 0.318561338 | 0.318561338 | 0.36435599 | 0.804304364 | 1 | 0.364355992 | 0.364355992 | 0.364355992 | 0.874313433 | -0.058332849 | FALSE | TRUE | FALSE |
| TMEM119 | 0.3184703 | 0.79698104 | 1 | 2.947274871 | 27.99399239 | 27.99399239 | 0.38741061 | 0.796594244 | 1 | 0.387410613 | 0.387410613 | 0.387410613 | 0.822048415 | -0.085102604 | FALSE | TRUE | FALSE |
| DHX30 | 0.3184656 | 0.79700505 | 1 | 0.318465555 | 0.318465555 | 10.73140472 | 0.36468411 | 0.804233303 | 1 | 0.364684105 | 0.364684105 | 7.09588025 | 0.873264149 | -0.058854369 | FALSE | TRUE | FALSE |
| PPFIA1 | 0.3184543 | 0.79702906 | 0.5139 | 0.318454293 | 0.318454293 | 6.204042905 | 0.10746771 | 0.873933355 | 0.51715 | 0.429712642 | 0.429712642 | 8.39769355 | 2.963255651 | 0.471769121 | FALSE | TRUE | FALSE |
| PCOLCE | 0.3184127 | 0.79705306 | 1 | 0.318412677 | 0.318412677 | 6.302810158 | 0.36414956 | 0.804375374 | 0.01115 | 0.364149559 | 0.364149559 | 7.217058148 | 0.874400831 | -0.058289438 | FALSE | TRUE | FALSE |
| CHAC1 | 0.3182967 | 0.79707705 | 1 | 0.318296704 | 0.318296704 | 0.318296704 | 0.3529734 | 0.808065219 | 0.5118 | 3.300049233 | 3.300049233 | 3.300049233 | 0.901758335 | -0.044909835 | FALSE | TRUE | FALSE |
| LAT2 | 0.318283 | 0.79710104 | 0.00725 | 0.318282953 | 0.318282953 | 0.318282953 | 0.36362257 | 0.80454086 | 1 | 0.363622569 | 0.363622569 | 0.363622569 | 0.875311326 | -0.057837452 | FALSE | TRUE | FALSE |
| LHB | 0.3181219 | 0.79712503 | 0.00495 | 0.318121858 | 0.318121858 | 0.318121858 | 0.36409915 | 0.804422684 | 1 | 0.364099151 | 0.364099151 | 0.364099151 | 0.873723152 | -0.058626156 | FALSE | TRUE | FALSE |
| KANSL3 | 0.3181071 | 0.79714901 | 0.01895 | 0.318107128 | 0.318107128 | 6.276688786 | 0.34317346 | 0.811132432 | 0.0175 | 0.343173461 | 0.343173461 | 6.782226546 | 0.926957249 | -0.032940295 | FALSE | TRUE | FALSE |
| SEMA4C | 0.3181008 | 0.79717298 | 1 | 2.982862814 | 2.982862814 | 58.3969873 | 0.37581908 | 0.800512943 | 1 | 0.375819078 | 0.375819078 | 7.376318431 | 0.846419931 | -0.072414119 | FALSE | TRUE | FALSE |
| IL24 | 0.3179999 | 0.79719695 | 0.005 | 0.317999921 | 0.317999921 | 0.317999921 | 0.363186 | 0.804635299 | 1 | 0.363185997 | 0.363185997 | 0.363185997 | 0.875584199 | -0.057702084 | FALSE | TRUE | FALSE |
| ARRDC3 | 0.3179678 | 0.79722091 | 1 | 2.995576395 | 2.995576395 | 6.308618569 | 0.35031772 | 0.809016208 | 1 | 0.350317723 | 0.350317723 | 6.957124149 | 0.907655432 | -0.042078989 | FALSE | TRUE | FALSE |
| SLC7A1 | 0.317897 | 0.79724486 | 0.51065 | 0.317896991 | 0.317896991 | 2.693238405 | 1.69677412 | 0.517636955 | 0.26365 | 0.424310804 | 1.696774123 | 8.371661259 | 0.187353748 | -0.727337615 | FALSE | FALSE | FALSE |
| UTF1 | 0.3177944 | 0.79726881 | 1 | 2.939741206 | 2.939741206 | 2.939741206 | 0.47168956 | 0.766511194 | 1 | 0.471689561 | 0.471689561 | 0.471689561 | 0.673736432 | -0.171509968 | FALSE | TRUE | FALSE |
| SEMA3A | 0.3177476 | 0.79729275 | 0.02175 | 0.317747592 | 0.317747592 | 6.259792018 | 0.36368262 | 0.804517236 | 1 | 0.363682622 | 0.363682622 | 7.177648903 | 0.873694734 | -0.058640282 | FALSE | TRUE | FALSE |
| VTI1A | 0.3176014 | 0.79731669 | 0.49825 | 0.317601428 | 1.270254679 | 1.270254679 | 1.45370492 | 0.540059872 | 0.51 | 1.453704919 | 1.453704919 | 1.453704919 | 0.218477233 | -0.660593813 | FALSE | FALSE | FALSE |
| STATH | 0.3176005 | 0.79734063 | 1 | 0.31760049 | 0.31760049 | 0.31760049 | 0.36266713 | 0.804753223 | 1 | 0.362667127 | 0.362667127 | 0.362667127 | 0.875735533 | -0.057627029 | FALSE | TRUE | FALSE |
| ZMYND11 | 0.3175819 | 0.79736455 | 1 | 2.967740055 | 28.46323599 | 58.56970293 | 0.38701324 | 0.796722303 | 1 | 0.387013243 | 0.387013243 | 7.650040044 | 0.820596889 | -0.085870134 | FALSE | TRUE | FALSE |
| MCU | 0.3173748 | 0.79738847 | 1 | 0.317374764 | 0.317374764 | 0.317374764 | 0.33470607 | 0.814059274 | 1 | 0.334706069 | 0.334706069 | 0.334706069 | 0.948219329 | -0.023091196 | FALSE | TRUE | FALSE |
| BPIFA1 | 0.3173443 | 0.79741239 | 1 | 0.31734428 | 0.31734428 | 0.31734428 | 0.36261293 | 0.80477679 | 1 | 0.362612926 | 0.362612926 | 0.362612926 | 0.875159866 | -0.057912607 | FALSE | TRUE | FALSE |
| DPPA2 | 0.317343 | 0.7974363 | 1 | 0.317343036 | 0.317343036 | 0.317343036 | 0.362798 | 0.80470607 | 1 | 0.362797996 | 0.362797996 | 0.362797996 | 0.874710002 | -0.058135907 | FALSE | TRUE | FALSE |
| RALB | 0.3173117 | 0.7974602 | 1 | 0.317311736 | 0.317311736 | 0.317311736 | 0.36321378 | 0.804611698 | 1 | 0.363213782 | 0.363213782 | 0.363213782 | 0.873622509 | -0.058676185 | FALSE | TRUE | FALSE |
| HSPA4 | 0.3172862 | 0.7974841 | 1 | 0.317286202 | 0.317286202 | 6.255787478 | 0.36327063 | 0.804588091 | 1 | 0.363270634 | 0.363270634 | 7.174658456 | 0.873415498 | -0.058779106 | FALSE | TRUE | FALSE |
| PDLIM5 | 0.3172644 | 0.79750799 | 0.01355 | 0.317264353 | 0.317264353 | 0.68878641 | 0.46726044 | 0.768034829 | 0.50575 | 0.46726044 | 1.868138017 | 1.868138017 | 0.67898826 | -0.168137735 | FALSE | TRUE | FALSE |
| NFKB1 | 0.3172419 | 0.79753188 | 0.02525 | 0.317241927 | 0.317241927 | 6.226349296 | 0.36351754 | 0.804564478 | 0.02285 | 0.363517536 | 0.363517536 | 7.150621386 | 0.872700477 | -0.059134787 | FALSE | TRUE | FALSE |
| SLAMF7 | 0.317117 | 0.79755576 | 1 | 0.317117033 | 0.317117033 | 0.317117033 | 0.36248451 | 0.804847456 | 1 | 0.362484515 | 0.362484515 | 0.362484515 | 0.874842979 | -0.058069889 | FALSE | TRUE | FALSE |
| SUGP1 | 0.3170727 | 0.79757963 | 1 | 0.317072717 | 0.317072717 | 6.242659803 | 0.3630925 | 0.804658895 | 1 | 0.363092497 | 0.363092497 | 7.162122125 | 0.873256042 | -0.058858401 | FALSE | TRUE | FALSE |
| USP10 | 0.317046 | 0.7976035 | 1 | 0.31704603 | 0.31704603 | 6.235721181 | 0.35193402 | 0.808519286 | 0.5077 | 3.301438281 | 3.301438281 | 65.06299933 | 0.900867816 | -0.045338928 | FALSE | TRUE | FALSE |
| IRX1 | 0.3168844 | 0.79762736 | 1 | 0.316884369 | 0.316884369 | 6.257177816 | 1.41917453 | 0.543701934 | 0.7546 | 1.419174534 | 5.677594129 | 28.06576146 | 0.223287807 | -0.651134992 | FALSE | FALSE | FALSE |
| CHKA | 0.316747 | 0.79765122 | 1 | 1.266801308 | 1.266801308 | 6.283687534 | 0.36692413 | 0.803352508 | 1 | 0.366924131 | 0.366924131 | 0.366924131 | 0.86324923 | -0.0638638 | FALSE | TRUE | FALSE |
| TRAPPC13 | 0.3167436 | 0.79767508 | 1 | 0.31674365 | 0.31674365 | 0.31674365 | 0.36261202 | 0.804800351 | 1 | 0.362612024 | 0.362612024 | 0.362612024 | 0.873505646 | -0.058734284 | FALSE | TRUE | FALSE |
| MRPL49 | 0.3167242 | 0.79769892 | 1 | 0.316724175 | 0.316724175 | 0.316724175 | 0.36231121 | 0.804894537 | 0.0051 | 0.362311208 | 0.362311208 | 0.362311208 | 0.874177138 | -0.058400556 | FALSE | TRUE | FALSE |
| PROK2 | 0.3166329 | 0.79772276 | 1 | 47.4156284 | 1822.662196 | 7294.719452 | 14.5186016 | 0.160195446 | 0.25295 | 3.630223395 | 14.51860164 | 14.51860164 | 0.021808776 | -1.6613687 | FALSE | FALSE | FALSE |
| NETO1 | 0.3165587 | 0.7977466 | 1 | 0.316558658 | 0.316558658 | 6.262473206 | 0.36240375 | 0.804870999 | 1 | 0.362403749 | 0.362403749 | 7.178784611 | 0.873497195 | -0.058738485 | FALSE | TRUE | FALSE |
| TRAPPC6B | 0.316547 | 0.79777043 | 0.0048 | 0.316546998 | 0.316546998 | 0.316546998 | 3.25437008 | 0.397336064 | 0.50435 | 3.254370076 | 3.254370076 | 3.254370076 | 0.097268286 | -1.012028739 | FALSE | FALSE | FALSE |
| KCNAB1 | 0.3165109 | 0.79779425 | 1 | 0.316510868 | 0.316510868 | 6.277649741 | 3.27446368 | 0.395992678 | 0.50595 | 3.274463679 | 3.274463679 | 6.972058328 | 0.096660369 | -1.01475155 | FALSE | FALSE | FALSE |
| YEATS4 | 0.3164508 | 0.79781807 | 1 | 1.265739486 | 1.265739486 | 1.265739486 | 0.36601395 | 0.803734394 | 1 | 0.36601395 | 0.36601395 | 0.36601395 | 0.864586692 | -0.063191453 | FALSE | TRUE | FALSE |
| USP49 | 0.3164225 | 0.79784188 | 1 | 0.316422497 | 0.316422497 | 6.210873142 | 0.36254942 | 0.804823906 | 1 | 0.36254942 | 0.36254942 | 7.132191999 | 0.872770659 | -0.059099862 | FALSE | TRUE | FALSE |
| TMEM242 | 0.3161928 | 0.79786568 | 1 | 0.31619279 | 0.31619279 | 0.31619279 | 0.36161481 | 0.805129602 | 1 | 0.361614812 | 0.361614812 | 0.361614812 | 0.874391148 | -0.058294248 | FALSE | TRUE | FALSE |

| | | | | | | | | | | | | | | | | | |
|---|---|---|---|---|---|---|---|---|---|---|---|---|---|---|---|---|---|
| B3GNT9 | 0.3158963 | 0.79788949 | 1 | 2.9469282 | 2.9469282 | 6.246548744 | 0.37325771 | 0.801273374 | 1 | 0.373257713 | 0.373257713 | 7.390847432 | 0.846322306 | -0.072464213 | FALSE | TRUE | FALSE |
| UNK | 0.315749 | 0.79791329 | 1 | 0.315749025 | 0.315749025 | 6.190359995 | 0.3619351 | 0.804965118 | 1 | 0.361935101 | 0.361935101 | 7.112716563 | 0.872391276 | -0.059288686 | FALSE | TRUE | FALSE |
| NAA15 | 0.3157312 | 0.79793708 | 1 | 0.315731181 | 0.315731181 | 6.224101341 | 0.36181174 | 0.805035647 | 1 | 0.36181174 | 0.36181174 | 7.144806501 | 0.872639404 | -0.05916518 | FALSE | TRUE | FALSE |
| XCL2 | 0.3157249 | 0.79796086 | 1 | 0.315724897 | 0.315724897 | 0.315724897 | 0.3609695 | 0.805317242 | 1 | 0.360969502 | 0.360969502 | 0.360969502 | 0.874658093 | -0.058161681 | FALSE | TRUE | FALSE |
| MAN2C1 | 0.3156859 | 0.79798464 | 0.12785 | 0.019751023 | 0.66130208 | 2.644155173 | 1.185E-06 | 0.901170627 | 1 | 0.477005691 | 37.31287802 | 149.1706004 | 266409.4886 | 5.425549689 | FALSE | TRUE | FALSE |
| RNPC3 | 0.3155545 | 0.79800842 | 1 | 0.31555449 | 0.31555449 | 0.31555449 | 0.36080857 | 0.805410919 | 1 | 0.360808573 | 0.360808573 | 0.360808573 | 0.874575921 | -0.058202484 | FALSE | TRUE | FALSE |
| ARHGAP26 | 0.3155052 | 0.79803218 | 1 | 2.951033189 | 2.951033189 | 58.01273828 | 0.37301304 | 0.801322235 | 1 | 0.373013039 | 0.373013039 | 7.347934422 | 0.845828945 | -0.072717457 | FALSE | TRUE | FALSE |
| MIER1 | 0.3155045 | 0.79805595 | 1 | 0.31550445 | 0.31550445 | 6.25491359 | 0.36118132 | 0.805200012 | 1 | 0.361181318 | 0.361181318 | 7.167993275 | 0.873534798 | -0.05871979 | FALSE | TRUE | FALSE |
| KCNJ5 | 0.3154476 | 0.7980797 | 0.69595 | 0.31544758 | 5.043899119 | 5.043899119 | 0.50726781 | 0.75306483 | 0.50435 | 0.507267806 | 2.02839619 | 2.515132943 | 0.6218561 | -0.206310101 | FALSE | TRUE | FALSE |
| SPAG11B | 0.3154355 | 0.79810346 | 1 | 0.315435475 | 0.315435475 | 0.315435475 | 0.35675305 | 0.806828153 | 1 | 1.426953075 | 1.426953075 | 1.426953075 | 0.884184393 | -0.053457155 | FALSE | TRUE | FALSE |
| RUVBL1 | 0.3154155 | 0.7981272 | 1 | 1.261372125 | 1.261372125 | 6.247824684 | 0.36577619 | 0.803877206 | 0.0117 | 0.365776194 | 0.365776194 | 7.253736597 | 0.862318377 | -0.064332359 | FALSE | TRUE | FALSE |
| GPR52 | 0.3152815 | 0.79815094 | 1 | 0.315281514 | 0.315281514 | 6.259511514 | 5.65092843 | 0.280050968 | 0.2602 | 1.412524402 | 5.65092843 | 5.65092843 | 0.05579287 | -1.2534213 | FALSE | FALSE | FALSE |
| STK40 | 0.3151266 | 0.79817467 | 1 | 0.315126624 | 0.315126624 | 6.225270533 | 0.36125834 | 0.805176548 | 1 | 0.361258341 | 0.361258341 | 7.147120962 | 0.872302693 | -0.059332787 | FALSE | TRUE | FALSE |
| XPO4 | 0.3151204 | 0.7981984 | 0.031 | 0.315120407 | 0.315120407 | 10.61042805 | 0.3613156 | 0.805153078 | 1 | 0.361315602 | 0.361315602 | 7.103618551 | 0.872147245 | -0.059410187 | FALSE | TRUE | FALSE |
| EPG5 | 0.3150777 | 0.79822213 | 0.37765 | 0.14287101 | 11.38372411 | 45.52017728 | 0.82372824 | 0.613534341 | 0.1523 | 0.092072452 | 0.823728238 | 3.074538151 | 0.38250195 | -0.417366346 | FALSE | TRUE | FALSE |
| DEFB110 | 0.3149943 | 0.79824584 | 1 | 0.314994268 | 0.314994268 | 0.314994268 | 0.34924977 | 0.809308621 | 1 | 0.349249772 | 3.230675056 | 3.230675056 | 0.901916889 | -0.04483348 | FALSE | TRUE | FALSE |
| CAPN5 | 0.3149723 | 0.79826956 | 0.01615 | 0.03417831 | 0.314972307 | 0.671736442 | 0.00753151 | 0.89742864 | 1 | 0.068115962 | 0.068115962 | 1.341532374 | 41.82061811 | 1.621390447 | FALSE | TRUE | FALSE |
| NSL1 | 0.3149536 | 0.79829326 | 1 | 0.314953603 | 0.314953603 | 0.314953603 | 0.36056919 | 0.805504505 | 1 | 0.360569189 | 0.360569189 | 0.360569189 | 0.873490062 | -0.058742032 | FALSE | TRUE | FALSE |
| ARID5B | 0.3148982 | 0.79831696 | 1 | 0.314898228 | 0.314898228 | 6.148225528 | 0.36114795 | 0.805246922 | 1 | 0.36114795 | 0.36114795 | 7.071427242 | 0.871936912 | -0.059514937 | FALSE | TRUE | FALSE |
| ZNF282 | 0.3146234 | 0.79834066 | 1 | 0.314623448 | 0.314623448 | 6.195748801 | 0.36046528 | 0.805551261 | 1 | 0.360465277 | 0.360465277 | 7.111626033 | 0.872825951 | -0.05907235 | FALSE | TRUE | FALSE |
| OR51I1 | 0.3145694 | 0.79836435 | 1 | 0.314569361 | 0.314569361 | 0.314569361 | 2.95791728 | 0.413913276 | 0.6911 | 28.23227614 | 2773.351901 | 2773.351901 | 0.106348262 | -0.973269605 | FALSE | FALSE | FALSE |
| ITPKB | 0.3144549 | 0.79838803 | 1 | 0.31445488 | 0.31445488 | 6.131458925 | 0.35125036 | 0.808722894 | 1 | 0.351250358 | 0.351250358 | 6.86961024 | 0.895244299 | -0.048058436 | FALSE | TRUE | FALSE |
| ZNF284 | 0.3141966 | 0.79841171 | 0.938 | 2.844171536 | 251.3444007 | 251.3444007 | 0.49349208 | 0.757612965 | 0.7515 | 0.493492078 | 4.512343668 | 4.512343668 | 0.636680228 | -0.196078637 | FALSE | TRUE | FALSE |
| DBNL | 0.3140844 | 0.79843539 | 1 | 0.314084402 | 0.314084402 | 6.227267501 | 0.35993987 | 0.805691397 | 1 | 0.359939875 | 0.359939875 | 7.143867436 | 0.87260241 | -0.059183592 | FALSE | TRUE | FALSE |
| SNX22 | 0.3140056 | 0.79845905 | 0.4995 | 0.314005643 | 1.255870234 | 1.255870234 | 0.36373743 | 0.804493607 | 0.00355 | 0.363737433 | 0.363737433 | 0.363737433 | 0.863275581 | -0.063850544 | FALSE | TRUE | FALSE |
| LY6G6D | 0.3139652 | 0.79848272 | 1 | 0.313965155 | 0.313965155 | 0.313965155 | 0.35916934 | 0.805924509 | 0.00415 | 0.359169343 | 0.359169343 | 0.359169343 | 0.874142409 | -0.058417809 | FALSE | TRUE | FALSE |
| SPCS1 | 0.3138941 | 0.79850638 | 0.004 | 0.313894063 | 0.313894063 | 0.313894063 | 0.35953371 | 0.80576139 | 1 | 0.359533713 | 0.359533713 | 0.359533713 | 0.873058776 | -0.058956518 | FALSE | TRUE | FALSE |
| C12orf60 | 0.3138391 | 0.79853003 | 0.00485 | 0.313839146 | 0.313839146 | 0.313839146 | 0.3590419 | 0.805947789 | 1 | 0.359041897 | 0.359041897 | 0.359041897 | 0.874101736 | -0.058438017 | FALSE | TRUE | FALSE |
| CREG2 | 0.3137894 | 0.79855367 | 1 | 0.313789419 | 0.313789419 | 0.313789419 | 0.35935052 | 0.805808025 | 1 | 0.359350521 | 0.359350521 | 0.359350521 | 0.873212646 | -0.058879983 | FALSE | TRUE | FALSE |
| EMB | 0.3137445 | 0.79857732 | 0.007 | 0.313744538 | 0.313744538 | 0.313744538 | 32.6313767 | 0.084149732 | 0.00295 | 3.546645261 | 32.63137666 | 32.63137666 | 0.009614812 | -2.017059223 | FALSE | FALSE | FALSE |
| CNEP1R1 | 0.3136599 | 0.79860095 | 1 | 0.313659893 | 0.313659893 | 0.313659893 | 0.31960091 | 0.818939919 | 0.00345 | 0.319600907 | 0.319600907 | 0.319600907 | 0.981411148 | -0.008149013 | FALSE | TRUE | FALSE |
| PCDHA1 | 0.3136209 | 0.79862458 | 0.0214 | 0.082270995 | 0.082270995 | 1.600891124 | 0.01646111 | 0.894744677 | 0.0297 | 0.016461109 | 0.016461109 | 0.321357534 | 19.05223681 | 1.279945971 | FALSE | TRUE | FALSE |
| RAVER1 | 0.3135805 | 0.7986482 | 1 | 0.31358048 | 0.31358048 | 6.153947244 | 0.35951517 | 0.80578471 | 1 | 0.359515169 | 0.359515169 | 7.071344429 | 0.87223157 | -0.059368198 | FALSE | TRUE | FALSE |
| NGRN | 0.313206 | 0.79867182 | 0.00895 | 0.313205969 | 0.313205969 | 0.313205969 | 0.35858357 | 0.806110582 | 1 | 0.358583572 | 0.358583572 | 0.358583572 | 0.873453201 | -0.058760359 | FALSE | TRUE | FALSE |
| MEA1 | 0.313174 | 0.79869544 | 1 | 0.313174049 | 0.313174049 | 0.313174049 | 0.35836042 | 0.806226691 | 1 | 0.358360416 | 0.358360416 | 0.358360416 | 0.873908042 | -0.058534264 | FALSE | TRUE | FALSE |
| TNFRSF8 | 0.3131654 | 0.79871905 | 1 | 0.313165413 | 0.313165413 | 6.175158731 | 0.35924837 | 0.805854636 | 1 | 0.359248367 | 0.359248367 | 7.095844832 | 0.871723973 | -0.05962101 | FALSE | TRUE | FALSE |
| AKTIP | 0.3130186 | 0.79874265 | 0.00885 | 0.31301856 | 0.31301856 | 0.31301856 | 0.35903099 | 0.805971062 | 1 | 0.359030995 | 0.359030995 | 0.359030995 | 0.871842721 | -0.059561854 | FALSE | TRUE | FALSE |
| CLDN11 | 0.312748 | 0.79876625 | 1 | 0.312748023 | 0.312748023 | 0.312748023 | 0.3589549 | 0.806017592 | 1 | 0.358954902 | 0.358954902 | 0.358954902 | 0.871273861 | -0.059845315 | FALSE | TRUE | FALSE |
| OR7A10 | 0.3127405 | 0.79878984 | 1 | 0.312740494 | 0.312740494 | 0.312740494 | 3.22089336 | 0.398452143 | 0.5035 | 3.220893356 | 3.220893356 | 3.220893356 | 0.097097438 | -1.012792228 | FALSE | FALSE | FALSE |
| AVEN | 0.3127032 | 0.79881343 | 0.0069 | 0.31270317 | 0.31270317 | 0.31270317 | 0.05230256 | 0.886187485 | 1 | 0.052302563 | 0.460480278 | 0.460480278 | 5.978735108 | 0.776609312 | FALSE | TRUE | FALSE |
| PCBP4 | 0.3126524 | 0.79883701 | 1 | 0.312652395 | 0.312652395 | 6.19481198 | 3.23759275 | 0.398005949 | 0.50785 | 3.23759275 | 3.23759275 | 6.890464288 | 0.096569402 | -1.01516046 | FALSE | FALSE | FALSE |
| ROGDI | 0.3126104 | 0.79886059 | 1 | 0.312610398 | 0.312610398 | 0.312610398 | 3.21069239 | 0.399564814 | 0.5011 | 3.210692385 | 3.210692385 | 3.210692385 | 0.097365416 | -1.011595278 | FALSE | FALSE | FALSE |
| C2CD4A | 0.3124292 | 0.79888416 | 0.7514 | 2.904960707 | 27.72166956 | 27.72166956 | 3.44763872 | 0.385979183 | 0.5027 | 3.447638725 | 3.447638725 | 3.447638725 | 0.090621225 | -1.04277007 | FALSE | FALSE | FALSE |
| ARHGAP25 | 0.3124013 | 0.79890772 | 0.0194 | 0.312401296 | 0.312401296 | 6.164414828 | 0.35833872 | 0.806249896 | 1 | 0.35833872 | 0.35833872 | 7.082255276 | 0.871804466 | -0.059580911 | FALSE | TRUE | FALSE |
| SPINT2 | 0.3122968 | 0.79893128 | 0.00845 | 0.312296756 | 0.312296756 | 0.312296756 | 0.35793631 | 0.806412166 | 1 | 0.357936311 | 0.357936311 | 0.357936311 | 0.872492526 | -0.059238285 | FALSE | TRUE | FALSE |
| ETNK1 | 0.3122693 | 0.79895484 | 1 | 0.312269349 | 0.312269349 | 6.189766661 | 0.35815414 | 0.806296288 | 1 | 0.35815414 | 0.35815414 | 7.106893933 | 0.871885352 | -0.059540619 | FALSE | TRUE | FALSE |
| REL | 0.3121727 | 0.79897839 | 1 | 0.31217267 | 0.31217267 | 6.176425582 | 0.35802765 | 0.806389001 | 0.0137 | 0.358027653 | 0.358027653 | 7.092828111 | 0.871923349 | -0.059521692 | FALSE | TRUE | FALSE |
| ZBTB26 | 0.3121593 | 0.79900193 | 1 | 0.312159263 | 0.312159263 | 6.197850839 | 0.3576984 | 0.806481623 | 1 | 0.357698403 | 0.357698403 | 0.357698403 | 0.872688447 | -0.059140773 | FALSE | TRUE | FALSE |
| ANKRD33 | 0.3121294 | 0.79902547 | 0.50625 | 0.312129412 | 0.312129412 | 0.697017486 | 0.05331756 | 0.885996073 | 1 | 0.053317563 | 0.053317563 | 1.056593582 | 5.854157562 | 0.767464407 | FALSE | TRUE | FALSE |
| TRAM2 | 0.3120564 | 0.799049 | 1 | 0.312056437 | 0.312056437 | 6.187603976 | 0.35810644 | 0.806365832 | 0.00925 | 0.358106439 | 0.358106439 | 0.358106439 | 0.871406942 | -0.059778984 | FALSE | TRUE | FALSE |
| DHX40 | 0.3120496 | 0.79907252 | 0.02145 | 0.312049617 | 0.312049617 | 6.148699064 | 0.34706007 | 0.80991307 | 1 | 3.258371023 | 3.258371023 | 64.31798467 | 0.899122773 | -0.046181003 | FALSE | TRUE | FALSE |
| RASD2 | 0.3120107 | 0.79909604 | 1 | 0.312010657 | 0.312010657 | 6.186684912 | 0.35428199 | 0.80760893 | 1 | 1.416764722 | 1.416764722 | 1.416764722 | 0.880684499 | -0.055179648 | FALSE | TRUE | FALSE |
| DRD1 | 0.3120065 | 0.79911956 | 1 | 2.916842141 | 27.98660535 | 27.98660535 | 0.38103527 | 0.798899158 | 0.0107 | 0.381035269 | 0.381035269 | 7.559125386 | 0.818839029 | -0.086801466 | FALSE | TRUE | FALSE |
| GSTA3 | 0.3120064 | 0.79914306 | 1 | 0.31200637 | 0.31200637 | 0.31200637 | 0.35724062 | 0.806597277 | 1 | 0.357240618 | 0.357240618 | 0.357240618 | 0.873378767 | -0.058797371 | FALSE | TRUE | FALSE |
| PLXDC2 | 0.3120059 | 0.79916657 | 1 | 0.312005858 | 0.312005858 | 6.178804345 | 0.35812828 | 0.806342657 | 0.01325 | 0.35812828 | 0.35812828 | 7.100571339 | 0.871212567 | -0.059875868 | FALSE | TRUE | FALSE |
| GBP3 | 0.3119003 | 0.79919006 | 0.01445 | 0.311900261 | 0.311900261 | 0.69505684 | 4.11665537 | 0.3492317 | 0.2539 | 0.44844898 | 4.116655369 | 4.116655369 | 0.075765454 | -1.120528772 | FALSE | FALSE | FALSE |
| PICALM | 0.3118758 | 0.79921355 | 1 | 0.311875844 | 0.311875844 | 6.169293281 | 0.35747149 | 0.806551032 | 1 | 0.35747149 | 0.35747149 | 7.080536542 | 0.87244956 | -0.059259672 | FALSE | TRUE | FALSE |
| XAB2 | 0.3118207 | 0.79923704 | 0.76375 | 0.311820733 | 1.247343138 | 10.52698465 | 0.1075364 | 0.873913315 | 1 | 0.107536399 | 0.107536399 | 2.098170046 | 2.899676166 | 0.462349499 | FALSE | TRUE | FALSE |
| SFXN4 | 0.3117567 | 0.79926052 | 1 | 2.883023435 | 2.883023435 | 6.190897151 | 0.35715377 | 0.806666601 | 1 | 3.313012939 | 3.313012939 | 3.313012939 | 0.872892157 | -0.059039409 | FALSE | TRUE | FALSE |
| CNOT11 | 0.3117259 | 0.79928399 | 0.01505 | 0.311725867 | 0.311725867 | 6.164099574 | 0.35753981 | 0.806504765 | 1 | 0.357539806 | 0.357539806 | 7.079636291 | 0.871863389 | -0.059551558 | FALSE | TRUE | FALSE |
| LYG1 | 0.3116522 | 0.79930746 | 0.6893 | 0.311652171 | 30.49743253 | 30.49743253 | 0.0041826 | 0.898261771 | 1 | 0.038995789 | 0.373144398 | 0.373144398 | 74.5115118 | 1.872223375 | FALSE | TRUE | FALSE |
| PCMTD2 | 0.3115445 | 0.79933092 | 1 | 0.31154446 | 0.31154446 | 6.181256521 | 0.35711267 | 0.806689697 | 1 | 0.35711267 | 0.35711267 | 0.424463029 | 0.872398225 | -0.059285227 | FALSE | TRUE | FALSE |
| GRIN2C | 0.3114952 | 0.79935438 | 0.9703 | 26.75596941 | 257.4340187 | 2537.479748 | 0.52291799 | 0.745592358 | 0.511 | 0.522917989 | 0.522917989 | 10.26209793 | 0.595686592 | -0.224982175 | FALSE | TRUE | FALSE |
| EVA1C | 0.3114733 | 0.79937783 | 1 | 0.311473256 | 0.311473256 | 6.17697107 | 0.35718653 | 0.806643499 | 1 | 0.357186528 | 0.357186528 | 0.357186528 | 0.872018486 | -0.059474308 | FALSE | TRUE | FALSE |
| KCTD4 | 0.3114028 | 0.79940128 | 1 | 0.311402826 | 0.311402826 | 0.311402826 | 0.34613199 | 0.81020274 | 1 | 3.227331335 | 3.227331335 | 3.227331335 | 0.899664976 | -0.045919186 | FALSE | TRUE | FALSE |
| PAX8 | 0.3112671 | 0.79942472 | 0.01345 | 0.077884995 | 0.311267104 | 1.541155991 | 0.03179174 | 0.890813966 | 1 | 0.031791737 | 0.031791737 | 0.629885461 | 9.790817771 | 0.990818967 | FALSE | TRUE | FALSE |
| CNOT6 | 0.3112561 | 0.79944815 | 1 | 0.311256147 | 0.311256147 | 6.162094276 | 0.35721484 | 0.806620391 | 1 | 0.357214841 | 0.357214841 | 7.080574802 | 0.871341587 | -0.059811558 | FALSE | TRUE | FALSE |
| PPID | 0.3111875 | 0.79947158 | 0.7496 | 0.311187499 | 2.838894571 | 2.838894571 | 0.05469354 | 0.885745562 | 1 | 0.054693544 | 0.054693544 | 0.054693544 | 5.689656921 | 0.75508608 | FALSE | TRUE | FALSE |
| PPIH | 0.3110781 | 0.79949501 | 1 | 0.31107807 | 0.31107807 | 0.31107807 | 0.35634163 | 0.806966411 | 0.00395 | 0.356341635 | 0.356341635 | 0.356341635 | 0.872977053 | -0.058997172 | FALSE | TRUE | FALSE |
| CHAMP1 | 0.3110439 | 0.79951843 | 1 | 0.311043902 | 0.311043902 | 6.130636592 | 0.33460031 | 0.814080607 | 1 | 0.334600312 | 0.334600312 | 6.606444509 | 0.929598361 | -0.031704651 | FALSE | TRUE | FALSE |
| KRTAP10-5 | 0.3109502 | 0.79954184 | 1 | 0.310950191 | 0.310950191 | 0.310950191 | 0.34542974 | 0.810447133 | 0.5056 | 3.197901115 | 3.197901115 | 3.197901115 | 0.900183618 | -0.045668895 | FALSE | TRUE | FALSE |
| IKZF4 | 0.3109335 | 0.79956525 | 1 | 0.310933521 | 0.310933521 | 6.128628071 | 0.35706226 | 0.80675895 | 1 | 0.357062257 | 0.357062257 | 7.050108984 | 0.870810383 | -0.060076401 | FALSE | TRUE | FALSE |
| PAK1IP1 | 0.3108512 | 0.79958865 | 0.0098 | 0.310851179 | 0.310851179 | 0.310851179 | 1.41053593 | 0.544504418 | 0.497 | 0.352671409 | 1.410535927 | 1.410535927 | 0.220378066 | -0.656831633 | FALSE | FALSE | FALSE |

| | | | | | | | | | | | | | | | | | |
|---|---|---|---|---|---|---|---|---|---|---|---|---|---|---|---|---|---|
| CHMP1A | 0.3107225 | 0.79961205 | 1 | 2.90055207 | 27.78928373 | 27.78928373 | 3.36713401 | 0.391249686 | 0.62165 | 3.36713401 | 53.87321084 | 53.87321084 | 0.092280999 | -1.034887711 | FALSE | FALSE | FALSE |
| C3orf33 | 0.3105756 | 0.79963544 | 1 | 0.310575645 | 0.310575645 | 0.310575645 | 266.844104 | 0.009334378 | 0.3148 | 27.46682932 | 2657.306631 | 2657.306631 | 0.001163884 | -2.934090216 | FALSE | FALSE | FALSE |
| CCR8 | 0.310538 | 0.79965882 | 1 | 0.310537956 | 0.310537956 | 0.310537956 | 0.35623217 | 0.806989434 | 0.00765 | 0.356232175 | 0.356232175 | 0.356232175 | 0.871729109 | -0.059618452 | FALSE | TRUE | FALSE |
| TAF5L | 0.3104652 | 0.7996822 | 0.01015 | 0.310465196 | 0.310465196 | 6.165336948 | 0.35642059 | 0.806897308 | 1 | 0.356420591 | 0.356420591 | 0.356420591 | 0.871064142 | -0.059949864 | FALSE | TRUE | FALSE |
| WDR53 | 0.3102692 | 0.79970558 | 1 | 0.310269191 | 0.310269191 | 0.310269191 | 0.35577856 | 0.807196382 | 1 | 0.355778563 | 0.355778563 | 0.355778563 | 0.872085121 | -0.059441123 | FALSE | TRUE | FALSE |
| FLAD1 | 0.3100767 | 0.79972895 | 0.8913 | 27.08567961 | 1050.841569 | 10445.19707 | 0.23722519 | 0.842814358 | 0.01545 | 0.23722519 | 0.23722519 | 1.172813205 | 1.307098595 | 0.116308348 | FALSE | TRUE | FALSE |
| ACTR2 | 0.3100602 | 0.79975232 | 1 | 1.239892925 | 1.239892925 | 6.156462557 | 0.36034521 | 0.805597995 | 1 | 0.360345212 | 0.360345212 | 0.360345212 | 0.860453242 | -0.065272725 | FALSE | TRUE | FALSE |
| GKN2 | 0.3099947 | 0.79977568 | 1 | 0.309994711 | 0.309994711 | 0.309994711 | 0.35563903 | 0.807219349 | 1 | 0.355639031 | 0.355639031 | 0.355639031 | 0.871655481 | -0.059655134 | FALSE | TRUE | FALSE |
| TTI2 | 0.3099095 | 0.79979903 | 0.5054 | 0.309909519 | 0.309909519 | 0.681346341 | 0.05250732 | 0.886145922 | 1 | 0.052507322 | 0.052507322 | 1.040595267 | 5.902215313 | 0.771015049 | FALSE | TRUE | FALSE |
| KRT40 | 0.3098583 | 0.79982238 | 0.78645 | 2.894900625 | 79.20644522 | 739.9987803 | 1816.60918 | 0.001338179 | 0.10475 | 46.12329517 | 2948.548924 | 29051.06355 | 0.00017057 | -3.768098325 | FALSE | FALSE | FALSE |
| OPRD1 | 0.3096955 | 0.79984572 | 1 | 0.309695468 | 0.309695468 | 6.118925015 | 0.35556754 | 0.80724231 | 1 | 0.355567544 | 0.355567544 | 7.035492877 | 0.870989138 | -0.059987261 | FALSE | TRUE | FALSE |
| MGST1 | 0.3096381 | 0.79986906 | 0.50905 | 1.238433419 | 1.238433419 | 1.238433419 | 3.23812803 | 0.397782769 | 0.50205 | 3.238128031 | 3.238128031 | 3.238128031 | 0.095622554 | -1.019439661 | FALSE | FALSE | FALSE |
| CASP10 | 0.3095947 | 0.7998924 | 0.818 | 2.761326404 | 25.24699057 | 236.4888303 | 0.06900791 | 0.882651748 | 0.93685 | 0.608186705 | 50.94858596 | 50.94858596 | 4.486365124 | 0.651894616 | FALSE | TRUE | FALSE |
| TRIM36 | 0.3095522 | 0.79991572 | 0.2672 | 0.309552249 | 1.238032404 | 6.112245029 | 0.10700623 | 0.874093455 | 0.0164 | 0.107006231 | 0.107006231 | 2.116120988 | 2.892843208 | 0.461324896 | FALSE | TRUE | FALSE |
| HCN1 | 0.3093137 | 0.79993905 | 0.5175 | 0.309313668 | 0.309313668 | 6.075081385 | 0.98777556 | 0.589890047 | 0.02195 | 0.2005846 | 0.2005846 | 3.948111762 | 0.313141648 | -0.504259167 | FALSE | TRUE | FALSE |
| LCE1C | 0.3092979 | 0.79996236 | 1 | 0.309297899 | 0.309297899 | 0.309297899 | 0.35432296 | 0.807586057 | 1 | 0.354322961 | 0.354322961 | 0.354322961 | 0.872926492 | -0.059022326 | FALSE | TRUE | FALSE |
| C11orf24 | 0.3092663 | 0.79998568 | 1 | 0.309266268 | 0.309266268 | 6.123479341 | 0.35478375 | 0.807471611 | 0.0106 | 0.354783754 | 0.354783754 | 7.033175619 | 0.871703578 | -0.059631171 | FALSE | TRUE | FALSE |
| MRGPRX4 | 0.3090742 | 0.80000898 | 0.0094 | 0.30907423 | 0.30907423 | 0.30907423 | 28.6348439 | 0.096063132 | 0.1314 | 28.63484389 | 280.089737 | 280.089737 | 0.010793641 | -1.966832023 | FALSE | FALSE | FALSE |
| KCNK7 | 0.309067 | 0.80003228 | 0.0075 | 0.309067044 | 0.309067044 | 0.309067044 | 3.18287266 | 0.401115545 | 0.50375 | 3.182872658 | 3.182872658 | 3.182872658 | 0.097103176 | -1.012766565 | FALSE | FALSE | FALSE |
| EPN1 | 0.3090667 | 0.80005558 | 0.01815 | 0.309066739 | 0.309066739 | 6.092380841 | 0.35484049 | 0.807425793 | 1 | 0.354840487 | 0.354840487 | 7.006795953 | 0.871001904 | -0.059980896 | FALSE | TRUE | FALSE |
| TRABD | 0.308861 | 0.80007887 | 1 | 0.308861022 | 0.308861022 | 6.103880215 | 1.40290332 | 0.545171202 | 0.50915 | 1.40290332 | 1.40290332 | 6.941764464 | 0.220158451 | -0.657264639 | FALSE | FALSE | FALSE |
| DEF6 | 0.3088017 | 0.80010215 | 1 | 1.23503409 | 1.23503409 | 10.44652739 | 0.35891891 | 0.806040848 | 1 | 0.358918907 | 0.358918907 | 7.08643496 | 0.86036621 | -0.065316654 | FALSE | TRUE | FALSE |
| GPN2 | 0.3086227 | 0.80012543 | 1 | 2.861429613 | 11.44535137 | 11.44535137 | 0.36941444 | 0.802318464 | 0.0093 | 0.369414445 | 0.369414445 | 0.369414445 | 0.835437632 | -0.078085966 | FALSE | TRUE | FALSE |
| TERF2IP | 0.3084849 | 0.80014871 | 0.0112 | 0.308484873 | 0.308484873 | 6.122452546 | 0.35387437 | 0.807768882 | 1 | 0.353874374 | 0.353874374 | 7.029758579 | 0.871735553 | -0.059615241 | FALSE | TRUE | FALSE |
| OR4K2 | 0.3084835 | 0.80017198 | 1 | 0.308483511 | 0.308483511 | 0.308483511 | 0.3541459 | 0.807677514 | 1 | 0.3541459 | 0.3541459 | 0.3541459 | 0.87106334 | -0.059950264 | FALSE | TRUE | FALSE |
| SFSWAP | 0.3084638 | 0.80019524 | 0.0328 | 0.308463825 | 0.308463825 | 6.013515502 | 0.354487 | 0.807540295 | 0.02815 | 0.354487002 | 0.354487002 | 6.931766072 | 0.870169635 | -0.060396076 | FALSE | TRUE | FALSE |
| HS6ST1 | 0.3084303 | 0.8002185 | 1 | 1.233417446 | 1.233417446 | 6.081700301 | 1.3869088 | 0.546367592 | 0.875 | 5.547775759 | 22.19721184 | 27.39375831 | 0.22238686 | -0.652890878 | FALSE | FALSE | FALSE |
| RGS8 | 0.3081413 | 0.80024175 | 1 | 0.308141317 | 0.308141317 | 0.308141317 | 0.33253064 | 0.81469708 | 1 | 0.332530644 | 0.332530644 | 0.332530644 | 0.9266554 | -0.033081739 | FALSE | TRUE | FALSE |
| PROKR2 | 0.3078805 | 0.800265 | 1 | 0.307880476 | 2.875751545 | 6.095341298 | 3.31004597 | 0.394193384 | 0.5054 | 3.310045974 | 3.310045974 | 7.010762685 | 0.093013957 | -1.031451877 | FALSE | FALSE | FALSE |
| MED6 | 0.3078664 | 0.80028825 | 1 | 0.307866364 | 0.307866364 | 0.307866364 | 0.35362543 | 0.807814533 | 1 | 0.353625428 | 0.353625428 | 0.353625428 | 0.870600188 | -0.060181244 | FALSE | TRUE | FALSE |
| ILDR2 | 0.3078653 | 0.80031149 | 1 | 1.231298316 | 1.231298316 | 10.42320361 | 0.35789815 | 0.806458476 | 1 | 0.357898147 | 0.357898147 | 7.078893578 | 0.860203615 | -0.065398737 | FALSE | TRUE | FALSE |
| VPREB3 | 0.3078464 | 0.80033472 | 0.0049 | 0.30784636 | 0.30784636 | 0.30784636 | 0.35314311 | 0.807996917 | 1 | 0.353143108 | 0.353143108 | 0.353143108 | 0.871732599 | -0.059616713 | FALSE | TRUE | FALSE |
| OR13C3 | 0.3078394 | 0.80035795 | 1 | 0.307839436 | 0.307839436 | 0.307839436 | 0.35336687 | 0.807905769 | 1 | 0.353366873 | 0.353366873 | 0.353366873 | 0.871160993 | -0.059901579 | FALSE | TRUE | FALSE |
| STX4 | 0.3076483 | 0.80038117 | 1 | 0.307648343 | 0.307648343 | 6.10384891 | 1.39631108 | 0.545437568 | 0.0085 | 1.396311075 | 1.396311075 | 1.396311075 | 0.220329373 | -0.656927602 | FALSE | FALSE | FALSE |
| MIXL1 | 0.3076245 | 0.80040438 | 1 | 0.307624533 | 0.307624533 | 0.307624533 | 0.35313521 | 0.80801969 | 1 | 0.353135207 | 0.353135207 | 0.353135207 | 0.87112394 | -0.059920051 | FALSE | TRUE | FALSE |
| GOSR1 | 0.3075857 | 0.80042759 | 1 | 0.307585727 | 0.307585727 | 0.307585727 | 0.35332389 | 0.807928565 | 1 | 0.353323888 | 0.353323888 | 0.353323888 | 0.870548914 | -0.060206822 | FALSE | TRUE | FALSE |
| HMX2 | 0.3075385 | 0.8004508 | 0.502 | 0.30753845 | 0.30753845 | 1.523811941 | 3.78415961 | 0.367474567 | 0.25195 | 0.946548779 | 3.784159606 | 3.784159606 | 0.081269947 | -1.090070024 | FALSE | FALSE | FALSE |
| ARHGEF19 | 0.3075068 | 0.800474 | 0.5061 | 0.307506781 | 0.307506781 | 6.042291199 | 0.04950912 | 0.88679127 | 1 | 0.442962815 | 4.067731407 | 8.722282488 | 6.211113684 | 0.793169478 | FALSE | TRUE | FALSE |
| CHP2 | 0.3074697 | 0.80049719 | 1 | 0.307469656 | 0.307469656 | 0.307469656 | 0.35301685 | 0.808042457 | 1 | 0.353016853 | 0.353016853 | 0.353016853 | 0.870977274 | -0.059993177 | FALSE | TRUE | FALSE |
| TNPO2 | 0.3074652 | 0.80052038 | 1 | 0.307465236 | 0.307465236 | 6.026747561 | 0.35361872 | 0.807837351 | 0.02215 | 0.35361872 | 0.35361872 | 6.948057441 | 0.869482349 | -0.06073923 | FALSE | TRUE | FALSE |
| PSMC5 | 0.3073859 | 0.80054356 | 0.9683 | 273.5676671 | 2767.056018 | 28656.74737 | 31.4546255 | 0.088752102 | 0.0054 | 3.733706557 | 14.93273666 | 14.93273666 | 0.009772358 | -2.010000639 | FALSE | FALSE | FALSE |
| FGF23 | 0.3073472 | 0.80056674 | 1 | 0.307347191 | 0.307347191 | 6.102713098 | 0.35330196 | 0.807951354 | 1 | 0.353301959 | 0.353301959 | 0.353301959 | 0.869927787 | -0.060516797 | FALSE | TRUE | FALSE |
| KCTD8 | 0.307238 | 0.80058991 | 1 | 0.307237977 | 0.307237977 | 6.063390178 | 0.34906718 | 0.809398412 | 0.5097 | 1.396170526 | 1.396170526 | 6.899873495 | 0.880168626 | -0.055434116 | FALSE | TRUE | FALSE |
| PRDX3 | 0.3071429 | 0.80061308 | 1 | 0.307142941 | 0.307142941 | 0.307142941 | 0.35272853 | 0.808178946 | 0.00575 | 0.35272853 | 0.35272853 | 0.35272853 | 0.870762967 | -0.060100049 | FALSE | TRUE | FALSE |
| CYP2D6 | 0.3071327 | 0.80063624 | 0.8086 | 2.800001178 | 26.16681039 | 250.5216363 | 0.06395985 | 0.883791825 | 0.98475 | 5.281366575 | 481.5165723 | 4764.743663 | 4.801960287 | 0.681418564 | FALSE | TRUE | FALSE |
| ULBP1 | 0.3070382 | 0.80065939 | 1 | 0.307038224 | 0.307038224 | 0.307038224 | 0.35217629 | 0.808405979 | 1 | 0.352176287 | 0.352176287 | 0.352176287 | 0.871831055 | -0.059567665 | FALSE | TRUE | FALSE |
| PTPRK | 0.3070037 | 0.80068254 | 0.52405 | 0.307003703 | 0.307003703 | 5.967193174 | 0.05002415 | 0.886626432 | 0.51365 | 0.45917989 | 0.45917989 | 8.95531712 | 6.137109909 | 0.787963901 | FALSE | TRUE | FALSE |
| TMEM63C | 0.3069957 | 0.80070569 | 0.0213 | 0.306995718 | 0.306995718 | 6.034469375 | 0.34894134 | 0.809420846 | 1 | 1.395695997 | 1.395695997 | 27.49125831 | 0.879791767 | -0.055620106 | FALSE | TRUE | FALSE |
| TUBE1 | 0.3068945 | 0.80072883 | 0.7546 | 0.306894452 | 1.227261152 | 1.523564823 | 0.42006894 | 0.784459421 | 0.50365 | 0.420068942 | 0.420068942 | 0.420068942 | 0.730581154 | -0.136331535 | FALSE | TRUE | FALSE |
| IMMT | 0.3068285 | 0.80075196 | 0.0221 | 0.306828469 | 0.306828469 | 6.051324673 | 3.19122175 | 0.400673109 | 0.50625 | 3.191221751 | 3.191221751 | 11.55241827 | 0.096147649 | -1.017061331 | FALSE | FALSE | FALSE |
| RRBP1 | 0.3067836 | 0.80077509 | 0.94075 | 2.825931707 | 258.5867061 | 258.5867061 | 0.50151249 | 0.754587679 | 0.516 | 0.50151249 | 0.50151249 | 9.828102081 | 0.611716695 | -0.213449667 | FALSE | TRUE | FALSE |
| PDCL3 | 0.3066606 | 0.80079821 | 1 | 0.306660566 | 0.306660566 | 0.306660566 | 0.35195405 | 0.808473981 | 1 | 0.351954055 | 0.351954055 | 0.351954055 | 0.871308518 | -0.05982804 | FALSE | TRUE | FALSE |
| NAB1 | 0.3065801 | 0.80082133 | 1 | 0.306580051 | 0.306580051 | 6.081236257 | 0.35235814 | 0.808269826 | 1 | 0.352358137 | 0.352358137 | 6.99618186 | 0.870080804 | -0.060440413 | FALSE | TRUE | FALSE |
| SYDE1 | 0.306557 | 0.80084444 | 1 | 0.306556953 | 0.306556953 | 6.055340424 | 0.35224636 | 0.808383301 | 1 | 0.352246359 | 0.352246359 | 6.968181557 | 0.870291332 | -0.060335342 | FALSE | TRUE | FALSE |
| PRNP | 0.3065453 | 0.80086755 | 0.5006 | 0.306545317 | 0.306545317 | 0.306545317 | 0.10390722 | 0.874937502 | 1 | 0.415393166 | 0.415393166 | 0.415393166 | 2.950183144 | 0.469848977 | FALSE | TRUE | FALSE |
| RNF141 | 0.3064727 | 0.80089065 | 1 | 0.306472749 | 0.306472749 | 0.306472749 | 0.35200235 | 0.808451319 | 1 | 0.352002347 | 0.352002347 | 0.352002347 | 0.870655416 | -0.060153694 | FALSE | TRUE | FALSE |
| ARFGAP2 | 0.3064681 | 0.80091374 | 1 | 0.306468149 | 0.306468149 | 6.055838623 | 0.35227711 | 0.808337928 | 1 | 0.352277108 | 0.352277108 | 6.971074462 | 0.86996328 | -0.060499078 | FALSE | TRUE | FALSE |
| CNOT6L | 0.3062992 | 0.80093683 | 1 | 0.306299227 | 0.306299227 | 6.062961378 | 0.35229289 | 0.808315233 | 1 | 0.352292894 | 0.352292894 | 6.982001153 | 0.869444807 | -0.060757982 | FALSE | TRUE | FALSE |
| SPINK5 | 0.3062314 | 0.80095992 | 0.6009 | 0.306231435 | 22.83256384 | 91.33371699 | 0.00271624 | 0.898794761 | 1 | 0.010862962 | 0.010862962 | 0.213358883 | 112.7410023 | 2.052081892 | FALSE | TRUE | FALSE |
| CKMT1A | 0.306198 | 0.800983 | 1 | 0.306197995 | 0.306197995 | 6.060183013 | 0.35168577 | 0.808587204 | 1 | 0.35168577 | 0.35168577 | 6.969184035 | 0.870657903 | -0.060152453 | FALSE | TRUE | FALSE |
| ZNF114 | 0.3061968 | 0.80100607 | 0.8735 | 2.751291258 | 25.35716221 | 25.35716221 | 0.48091996 | 0.762732432 | 0.50595 | 0.480919965 | 0.480919965 | 1.078755574 | 0.636689723 | -0.19607216 | FALSE | TRUE | FALSE |
| HNRNPH3 | 0.3061801 | 0.80102914 | 1 | 0.306180056 | 0.306180056 | 6.079761734 | 0.32305801 | 0.817718748 | 1 | 0.323058008 | 0.323058008 | 0.323058008 | 0.947755662 | -0.023303612 | FALSE | TRUE | FALSE |
| DAP3 | 0.3060898 | 0.8010522 | 1 | 0.306089762 | 0.306089762 | 6.074549768 | 0.3514882 | 0.808632456 | 1 | 0.351488205 | 0.351488205 | 0.351488205 | 0.870839357 | -0.060061952 | FALSE | TRUE | FALSE |
| ZNF770 | 0.3060384 | 0.80107526 | 1 | 0.306038408 | 0.306038408 | 6.052807096 | 0.33712655 | 0.813007746 | 0.75045 | 3.141595992 | 12.56607332 | 26.70563167 | 0.907784937 | -0.042017028 | FALSE | TRUE | FALSE |
| TMEM219 | 0.3060211 | 0.80109831 | 0.00555 | 0.306021053 | 0.306021053 | 0.306021053 | 0.35114444 | 0.808790664 | 1 | 0.351144437 | 0.351144437 | 0.351144437 | 0.871496229 | -0.059734488 | FALSE | TRUE | FALSE |
| NUP85 | 0.3059905 | 0.80112135 | 1 | 11.49625877 | 110.8352235 | 110.8352235 | 0.37912383 | 0.799498112 | 0.0182 | 0.379123834 | 0.379123834 | 7.486750429 | 0.807099085 | -0.093073145 | FALSE | TRUE | FALSE |
| PRPSAP1 | 0.3059314 | 0.80114439 | 0.81365 | 1.223457778 | 11.10555949 | 44.42361107 | 0.29030707 | 0.827979662 | 0.0087 | 0.290307074 | 0.290307074 | 0.290307074 | 1.053820057 | 0.02276646 | FALSE | TRUE | FALSE |
| QPRT | 0.3059061 | 0.80116743 | 1 | 0.305906071 | 0.305906071 | 0.305906071 | 0.34072666 | 0.811922036 | 0.50675 | 3.174133174 | 3.174133174 | 3.174133174 | 0.897804927 | -0.046818016 | FALSE | TRUE | FALSE |
| SLC25A4 | 0.3058757 | 0.80119046 | 1 | 0.305875745 | 0.305875745 | 0.305875745 | 0.34791478 | 0.809756728 | 0.5078 | 1.391396691 | 1.391396691 | 1.391396691 | 0.879168591 | -0.055927836 | FALSE | TRUE | FALSE |
| CMSS1 | 0.3058481 | 0.80121348 | 0.00725 | 0.305848107 | 0.305848107 | 0.305848107 | 3.15313617 | 0.403537771 | 0.5048 | 3.153136172 | 3.153136172 | 3.153136172 | 0.096998065 | -1.01323693 | FALSE | FALSE | FALSE |
| DUOXA2 | 0.305844 | 0.8012365 | 0.5051 | 0.305843956 | 0.305843956 | 1.5145248 | 0.88956229 | 0.599943863 | 0.75855 | 0.889562292 | 8.280136943 | 8.280136943 | 0.343813985 | -0.463676462 | FALSE | TRUE | FALSE |
| MYEF2 | 0.3057521 | 0.80125951 | 1 | 0.305752095 | 0.305752095 | 6.046454565 | 0.35173336 | 0.80856457 | 1 | 0.351733364 | 0.351733364 | 6.965151232 | 0.869272371 | -0.060844124 | FALSE | TRUE | FALSE |
| OR13C8 | 0.3057455 | 0.80128252 | 1 | 0.305745526 | 0.305745526 | 0.305745526 | 0.3512004 | 0.80874549 | 1 | 0.351200398 | 0.351200398 | 0.351200398 | 0.870572834 | -0.060194889 | FALSE | TRUE | FALSE |

| | | | | | | | | | | | | | | | | | |
|---|---|---|---|---|---|---|---|---|---|---|---|---|---|---|---|---|---|
| PLCL1 | 0.3057417 | 0.80130552 | 0.51805 | 0.305741677 | 0.305741677 | 5.999605434 | 0.1046383 | 0.87480911 | 1 | 0.1046383 | 0.1046383 | 2.057989153 | 2.921890719 | 0.465663969 | FALSE | TRUE | FALSE |
| EPM2AIP1 | 0.3056624 | 0.80132851 | 1 | 0.305662411 | 0.305662411 | 6.028855229 | 0.35137526 | 0.808677686 | 1 | 0.351375259 | 0.351375259 | 6.942004866 | 0.869903053 | -0.060529145 | FALSE | TRUE | FALSE |
| RWDD2A | 0.3056381 | 0.8013515 | 1 | 0.305638136 | 0.305638136 | 0.305638136 | 0.35098136 | 0.808880946 | 0.00575 | 0.350981361 | 0.350981361 | 0.350981361 | 0.870810162 | -0.060076512 | FALSE | TRUE | FALSE |
| C12orf50 | 0.3055603 | 0.80137449 | 0.01155 | 0.305560258 | 0.305560258 | 6.067232798 | 0.34016551 | 0.812052987 | 0.5043 | 3.155089339 | 3.155089339 | 6.760202003 | 0.898269365 | -0.046593411 | FALSE | TRUE | FALSE |
| STARD6 | 0.3055371 | 0.80139747 | 0.00675 | 0.305537137 | 0.305537137 | 0.305537137 | 5.04713378 | 0.304782796 | 0.89255 | 47.22532888 | 1814.664095 | 7262.880468 | 0.060536762 | -1.217980812 | FALSE | FALSE | FALSE |
| OR51L1 | 0.3054374 | 0.80142044 | 1 | 0.305437389 | 0.305437389 | 0.305437389 | 0.31967177 | 0.818859026 | 1 | 28.02317676 | 272.631234 | 272.631234 | 0.955471883 | -0.019782089 | FALSE | TRUE | FALSE |
| CCDC6 | 0.3054372 | 0.80144341 | 1 | 2.87391012 | 2.87391012 | 6.038443705 | 0.36272195 | 0.80472965 | 1 | 0.362721954 | 0.362721954 | 7.180881761 | 0.842069895 | -0.074651859 | FALSE | TRUE | FALSE |
| SIPA1L3 | 0.3053589 | 0.80146637 | 0.6563 | 0.696579106 | 23.38200744 | 178.1909974 | 10.4659397 | 0.193185659 | 0.3471 | 3.519969243 | 225.0031265 | 1077.227388 | 0.029176442 | -1.534967671 | FALSE | FALSE | FALSE |
| ELOVL5 | 0.3053238 | 0.80148933 | 1 | 0.305323831 | 0.305323831 | 0.305323831 | 0.32959029 | 0.815477771 | 1 | 3.053015183 | 29.02513535 | 29.02513535 | 0.926373858 | -0.033213709 | FALSE | TRUE | FALSE |
| SPRY4 | 0.3052901 | 0.80151228 | 0.75445 | 2.851675434 | 27.33857689 | 27.33857689 | 0.37369434 | 0.801126647 | 1 | 0.373694341 | 0.373694341 | 0.373694341 | 0.81695135 | -0.087803805 | FALSE | TRUE | FALSE |
| P2RY14 | 0.3052546 | 0.80153523 | 1 | 2.842535139 | 2.842535139 | 2.842535139 | 1.43164529 | 0.542628371 | 0.00815 | 1.43164529 | 1.43164529 | 1.43164529 | 0.213219454 | -0.671173173 | FALSE | FALSE | FALSE |
| ANKRD6 | 0.3050715 | 0.80155817 | 0.02055 | 0.305071511 | 0.305071511 | 6.016886058 | 0.05205211 | 0.886220716 | 1 | 0.052052108 | 0.052052108 | 1.028328946 | 5.860886787 | 0.767963332 | FALSE | TRUE | FALSE |
| CNKSR3 | 0.3049458 | 0.80158111 | 0.01805 | 0.304945824 | 0.304945824 | 6.022735438 | 3.16896202 | 0.402218993 | 0.50635 | 3.168962022 | 3.168962022 | 6.722654652 | 0.09622893 | -1.016694344 | FALSE | FALSE | FALSE |
| EFHD1 | 0.3048323 | 0.80160404 | 1 | 0.304832304 | 0.304832304 | 0.304832304 | 0.35005183 | 0.809128781 | 1 | 0.350051826 | 0.350051826 | 0.350051826 | 0.870820493 | -0.060071359 | FALSE | TRUE | FALSE |
| LRRC4 | 0.3048318 | 0.80162697 | 1 | 0.304831826 | 0.304831826 | 6.002311972 | 0.3509171 | 0.808903502 | 1 | 0.350917101 | 0.350917101 | 6.9226252 | 0.868671901 | -0.061144226 | FALSE | TRUE | FALSE |
| ZNF76 | 0.3047502 | 0.80164989 | 0.50765 | 0.304750216 | 0.304750216 | 1.177052542 | 0.05099491 | 0.886386627 | 1 | 0.451052809 | 0.451052809 | 1.727384313 | 5.976090584 | 0.776417172 | FALSE | TRUE | FALSE |
| IGFL2 | 0.304616 | 0.8016728 | 0.00335 | 0.304616016 | 0.304616016 | 0.304616016 | 0.34958296 | 0.80924122 | 0.00335 | 0.349582957 | 0.349582957 | 0.349582957 | 0.871369756 | -0.059797518 | FALSE | TRUE | FALSE |
| LTBP3 | 0.3045874 | 0.80169571 | 0.2108 | 0.304587352 | 1.482004023 | 5.92514652 | 0.00906725 | 0.897000493 | 1 | 0.082660812 | 0.330361248 | 1.613304122 | 33.59203224 | 1.526236278 | FALSE | TRUE | FALSE |
| DOCK5 | 0.3045241 | 0.80171862 | 0.52315 | 1.217634858 | 25.89891304 | 103.5563848 | 0.00849867 | 0.897180433 | 1 | 0.008498668 | 0.008498668 | 0.285335713 | 35.83198607 | 1.554270881 | FALSE | TRUE | FALSE |
| RETSAT | 0.3045104 | 0.80174152 | 0.7754 | 24.63844711 | 2196.23124 | 4538.655611 | 49.7595438 | 0.05530173 | 0.12085 | 5.388568146 | 470.7394555 | 4559.735923 | 0.006119639 | -2.213274196 | FALSE | FALSE | FALSE |
| HLA-DQA1 | 0.3043996 | 0.80176441 | 0.00645 | 0.304399567 | 0.304399567 | 0.304399567 | 0.29016484 | 0.828088976 | 1 | 2.73030779 | 261.1644992 | 261.1644992 | 1.049057397 | 0.02079925 | FALSE | TRUE | FALSE |
| BRINP2 | 0.3043785 | 0.8017873 | 1 | 0.304378536 | 0.304378536 | 5.982995062 | 1.3015159 | 0.554453491 | 0.8796 | 12.23591442 | 118.0615535 | 241.0118087 | 0.23386463 | -0.631035456 | FALSE | FALSE | FALSE |
| OAF | 0.3042675 | 0.80181018 | 1 | 0.304267548 | 0.304267548 | 0.304267548 | 0.34943671 | 0.809286159 | 1 | 0.349436708 | 0.349436708 | 0.349436708 | 0.87073722 | -0.060112891 | FALSE | TRUE | FALSE |
| APOBEC3C | 0.3042179 | 0.80183306 | 1 | 0.304217942 | 0.304217942 | 0.304217942 | 0.34962856 | 0.809218743 | 1 | 0.34962856 | 0.34962856 | 0.34962856 | 0.87011754 | -0.060422077 | FALSE | TRUE | FALSE |
| INO80D | 0.3041133 | 0.80185593 | 1 | 2.853099167 | 27.47166115 | 96.26180839 | 0.35007834 | 0.809106278 | 0.7556 | 3.289060303 | 31.71493247 | 64.6025473 | 0.868700608 | -0.061129875 | FALSE | TRUE | FALSE |
| AK1 | 0.3040613 | 0.8018788 | 0.49825 | 0.304061304 | 2.839838804 | 2.839838804 | 0.36087407 | 0.805387509 | 1 | 0.360874067 | 0.360874067 | 0.360874067 | 0.842569005 | -0.074394521 | FALSE | TRUE | FALSE |
| MPZ | 0.3040273 | 0.80190166 | 0.50225 | 2.859948485 | 2.859948485 | 2.859948485 | 0.36116144 | 0.80522347 | 1 | 0.361161439 | 0.361161439 | 0.361161439 | 0.841804522 | -0.074788746 | FALSE | TRUE | FALSE |
| LUC7L | 0.3040092 | 0.80192452 | 1 | 0.304009194 | 0.304009194 | 6.023506012 | 0.34985392 | 0.809173772 | 1 | 0.349853919 | 0.349853919 | 0.349853919 | 0.868960379 | -0.061000025 | FALSE | TRUE | FALSE |
| KCNJ9 | 0.3039406 | 0.80194737 | 1 | 1.215533911 | 4.862437436 | 10.30671786 | 64.6799177 | 0.043434845 | 0.0051 | 3.263122228 | 3.263122228 | 6.874676523 | 0.004699149 | -2.327980766 | FALSE | FALSE | FALSE |
| ARHGAP1 | 0.3039359 | 0.80197022 | 1 | 0.303935856 | 0.303935856 | 6.023814611 | 0.34599305 | 0.810269458 | 1 | 0.345993047 | 1.383752536 | 6.864810542 | 0.878444982 | -0.056285434 | FALSE | TRUE | FALSE |
| TSPAN31 | 0.3037842 | 0.80199306 | 1 | 2.837241621 | 2.837241621 | 2.837241621 | 0.36059813 | 0.805481117 | 1 | 0.360598132 | 0.360598132 | 0.360598132 | 0.842445381 | -0.074458247 | FALSE | TRUE | FALSE |
| DAB2 | 0.3037772 | 0.80201589 | 0.7569 | 2.849656704 | 27.43573147 | 56.20815843 | 0.37243333 | 0.801639226 | 1 | 0.372433335 | 0.372433335 | 7.358251218 | 0.815655196 | -0.088493393 | FALSE | TRUE | FALSE |
| ARMC12 | 0.3037288 | 0.80203872 | 1 | 2.809596833 | 2.809596833 | 6.023736207 | 0.36005653 | 0.805644708 | 0.01025 | 0.360056532 | 0.360056532 | 7.148049612 | 0.843558687 | -0.073884698 | FALSE | TRUE | FALSE |
| LDLRAD1 | 0.3037268 | 0.80206155 | 0.5014 | 0.685265786 | 2.740446064 | 2.740446064 | 0.93951598 | 0.594376251 | 0.50105 | 0.939515978 | 0.939515978 | 0.939515978 | 0.323280116 | -0.490421006 | FALSE | TRUE | FALSE |
| ADPRM | 0.3036709 | 0.80208437 | 0.50255 | 0.30367086 | 0.30367086 | 0.30367086 | 6.43181799 | 0.256750214 | 0.00775 | 1.607788551 | 6.431817985 | 6.431817985 | 0.047213845 | -1.325930627 | FALSE | FALSE | FALSE |
| LIF | 0.3034776 | 0.80210718 | 1 | 0.303477568 | 0.303477568 | 0.303477568 | 0.34922248 | 0.809331077 | 1 | 0.349222477 | 0.349222477 | 0.349222477 | 0.869009266 | -0.060975593 | FALSE | TRUE | FALSE |
| HPCAL4 | 0.3033714 | 0.80212999 | 1 | 0.303371402 | 0.303371402 | 0.303371402 | 0.338446 | 0.812553294 | 1 | 3.17622674 | 3.17622674 | 3.17622674 | 0.896365738 | -0.047514752 | FALSE | TRUE | FALSE |
| RBM25 | 0.3033665 | 0.80215279 | 1 | 0.303366495 | 0.303366495 | 10.20212566 | 3.09606117 | 0.405725289 | 0.02605 | 3.096061172 | 3.096061172 | 60.94778385 | 0.097984658 | -1.008841919 | FALSE | FALSE | FALSE |
| AMN | 0.3033537 | 0.80217559 | 0.5077 | 0.303353728 | 0.303353728 | 0.66191609 | 0.05126611 | 0.886345314 | 0.01045 | 0.051266107 | 0.051266107 | 1.016448223 | 5.917237393 | 0.772118993 | FALSE | TRUE | FALSE |
| WIPF2 | 0.303321 | 0.80219838 | 1 | 1.213193881 | 1.213193881 | 6.007021207 | 1.35312276 | 0.550702585 | 0.74985 | 3.158137351 | 12.63172281 | 12.63172281 | 0.224163675 | -0.649434763 | FALSE | FALSE | FALSE |
| KCNN4 | 0.3033198 | 0.80222117 | 0.0035 | 0.008949398 | 0.008949398 | 0.176744085 | 0.0155617 | 0.894975061 | 1 | 0.015561698 | 0.015561698 | 0.307795795 | 19.4914357 | 1.289843829 | FALSE | TRUE | FALSE |
| DDX46 | 0.303133 | 0.80224395 | 1 | 1.212409049 | 1.212409049 | 23.77487836 | 0.35322908 | 0.807974138 | 0.0265 | 0.353229084 | 0.353229084 | 6.942969472 | 0.858176893 | -0.066423183 | FALSE | TRUE | FALSE |
| GLMN | 0.3030486 | 0.80226673 | 0.87745 | 2.705335093 | 24.77194925 | 24.77194925 | 0.47508253 | 0.765139233 | 0.50415 | 0.475082534 | 0.475082534 | 0.475082534 | 0.637886251 | -0.195256759 | FALSE | TRUE | FALSE |
| TBC1D22B | 0.3030134 | 0.8022895 | 1 | 0.303013361 | 0.303013361 | 5.993131741 | 0.34874869 | 0.809488118 | 1 | 0.348748695 | 0.348748695 | 6.906898118 | 0.868858766 | -0.061050813 | FALSE | TRUE | FALSE |
| HAUS2 | 0.3029843 | 0.80231227 | 0.50065 | 2.792568275 | 2.792568275 | 2.792568275 | 0.35897376 | 0.80599433 | 0.00405 | 0.358973756 | 0.358973756 | 0.358973756 | 0.844029127 | -0.073642566 | FALSE | TRUE | FALSE |
| DLST | 0.3028693 | 0.80233503 | 1 | 0.302869302 | 0.302869302 | 6.000950143 | 0.34866255 | 0.809510531 | 1 | 0.348662547 | 0.348662547 | 6.916023637 | 0.868660271 | -0.061150041 | FALSE | TRUE | FALSE |
| DERA | 0.3027379 | 0.80235778 | 0.5064 | 0.302737903 | 0.302737903 | 0.302737903 | 0.10367923 | 0.874986821 | 0.00645 | 0.103679231 | 0.103679231 | 0.103679231 | 2.919947406 | 0.465375029 | FALSE | TRUE | FALSE |
| ERCC5 | 0.302718 | 0.80238054 | 1 | 2755.119146 | 304502.6391 | 3314498.843 | 0.37137969 | 0.801979428 | 1 | 336.3052471 | 35655.67262 | 380432.8378 | 0.815117338 | -0.088779869 | FALSE | TRUE | FALSE |
| ATL2 | 0.3026416 | 0.80240328 | 0.75375 | 0.302641605 | 2.867393534 | 5.984790977 | 0.05216965 | 0.886204102 | 1 | 0.052169652 | 0.052169652 | 1.033059255 | 5.80110459 | 0.763510696 | FALSE | TRUE | FALSE |
| SFMBT1 | 0.3025744 | 0.80242602 | 1 | 2.838832275 | 2.838832275 | 55.82967346 | 0.35964144 | 0.805738064 | 1 | 0.359641438 | 0.359641438 | 7.087061897 | 0.841322529 | -0.075037481 | FALSE | TRUE | FALSE |
| APOBEC4 | 0.3025094 | 0.80244876 | 1 | 0.302509371 | 0.302509371 | 0.302509371 | 0.33719559 | 0.812986157 | 1 | 3.124427138 | 3.124427138 | 3.124427138 | 0.897133228 | -0.047143058 | FALSE | TRUE | FALSE |
| TMEM222 | 0.3023764 | 0.80247149 | 1 | 0.302376431 | 0.302376431 | 0.302376431 | 0.34802664 | 0.809667277 | 1 | 0.348026637 | 0.348026637 | 0.348026637 | 0.868831288 | -0.061064548 | FALSE | TRUE | FALSE |
| RNF125 | 0.3022838 | 0.80249421 | 1 | 0.302283845 | 0.302283845 | 0.302283845 | 0.34794214 | 0.809712014 | 1 | 0.347942136 | 0.347942136 | 0.347942136 | 0.868776193 | -0.061092088 | FALSE | TRUE | FALSE |
| CLEC17A | 0.3022135 | 0.80251693 | 1 | 0.30221348 | 0.30221348 | 0.30221348 | 0.34733821 | 0.809846096 | 1 | 0.347338208 | 0.347338208 | 0.347338208 | 0.87008418 | -0.060438728 | FALSE | TRUE | FALSE |
| ARHGAP24 | 0.3021813 | 0.80253964 | 0.0205 | 0.302181274 | 0.302181274 | 5.959110806 | 0.10308317 | 0.875173897 | 1 | 0.103083172 | 0.103083172 | 2.036254068 | 2.931431657 | 0.467079774 | FALSE | TRUE | FALSE |
| NYAP1 | 0.3020544 | 0.80256235 | 1 | 0.302054439 | 0.302054439 | 5.92932567 | 0.3479701 | 0.809689649 | 1 | 0.347970101 | 0.347970101 | 6.845866187 | 0.868047107 | -0.061456706 | FALSE | TRUE | FALSE |
| SLC25A36 | 0.3020533 | 0.80258506 | 1 | 1.207905851 | 4.831601211 | 4.831601211 | 0.35596667 | 0.807127451 | 1 | 0.355966673 | 0.355966673 | 0.355966673 | 0.848543889 | -0.07132569 | FALSE | TRUE | FALSE |
| PAQR9 | 0.3019842 | 0.80260776 | 1 | 0.301984178 | 0.301984178 | 5.971977548 | 0.32700601 | 0.816314337 | 1 | 0.327006014 | 0.327006014 | 6.475525792 | 0.923482031 | -0.03457155 | FALSE | TRUE | FALSE |
| GAB1 | 0.3019254 | 0.80263045 | 1 | 2.839213959 | 27.40198744 | 56.01905021 | 0.37054916 | 0.802197501 | 1 | 0.370549164 | 0.370549164 | 7.322968786 | 0.814805362 | -0.088946122 | FALSE | TRUE | FALSE |
| HMG20A | 0.3018986 | 0.80265314 | 0.0106 | 0.301898605 | 0.301898605 | 5.991822872 | 0.34792553 | 0.809734374 | 1 | 0.347925528 | 0.347925528 | 0.347925528 | 0.867710419 | -0.061625188 | FALSE | TRUE | FALSE |
| PPFIBP2 | 0.3018736 | 0.80267582 | 0.6634 | 1.206971885 | 93.33546762 | 353.8776994 | 0.00139882 | 0.899262704 | 1 | 0.022369406 | 0.184047536 | 0.440728406 | 215.8064212 | 2.334064363 | FALSE | TRUE | FALSE |
| LRRN2 | 0.3018615 | 0.8026985 | 0.02235 | 0.301861465 | 0.301861465 | 5.931165584 | 0.33701676 | 0.813072481 | 0.5074 | 3.16128624 | 3.16128624 | 62.24634311 | 0.895686813 | -0.04784382 | FALSE | TRUE | FALSE |
| MAML3 | 0.3017389 | 0.80272117 | 1 | 2.828077227 | 27.20433912 | 95.30899336 | 0.37015233 | 0.802245901 | 1 | 0.370152327 | 0.370152327 | 7.267524557 | 0.815174975 | -0.088749161 | FALSE | TRUE | FALSE |
| UBR7 | 0.3014805 | 0.80274384 | 1 | 2.811602392 | 2.811602392 | 2.811602392 | 0.35813113 | 0.806319475 | 1 | 0.358131127 | 0.358131127 | 0.358131127 | 0.841815886 | -0.074782883 | FALSE | TRUE | FALSE |
| PCDHGA7 | 0.3014381 | 0.8027665 | 0.6079 | 0.301438143 | 10.775331 | 43.09771794 | 0.01800484 | 0.894346544 | 0.81735 | 0.158344606 | 5.714501205 | 7.008098321 | 16.74206029 | 1.223808901 | FALSE | TRUE | FALSE |
| TGFBRAP1 | 0.3014146 | 0.80278915 | 1 | 0.301414583 | 0.301414583 | 5.909774514 | 0.34335824 | 0.811110402 | 0.5101 | 1.373399762 | 1.373399762 | 26.99172631 | 0.877842867 | -0.056583216 | FALSE | TRUE | FALSE |
| KLHL2 | 0.3013402 | 0.80281181 | 1 | 0.301340203 | 0.301340203 | 5.959589699 | 1.37335675 | 0.548085036 | 0.50795 | 1.373356754 | 1.373356754 | 6.800175496 | 0.219418736 | -0.658726291 | FALSE | FALSE | FALSE |
| KLK12 | 0.3013238 | 0.80283445 | 0.50185 | 0.301323787 | 0.301323787 | 0.301323787 | 0.0509754 | 0.886394912 | 0.00775 | 0.050975396 | 0.050975396 | 0.050975396 | 5.911161327 | 0.771672812 | FALSE | TRUE | FALSE |
| CEACAM18 | 0.3012552 | 0.80285709 | 1 | 0.301255182 | 0.301255182 | 5.975588266 | 0.34655723 | 0.810046876 | 1 | 0.346557231 | 0.346557231 | 0.346557231 | 0.869279746 | -0.060840439 | FALSE | TRUE | FALSE |
| RGMA | 0.3012353 | 0.80287973 | 0.9362 | 11.37083736 | 440.6271802 | 2175.700224 | 0.13003159 | 0.868970652 | 0.93945 | 1.230974239 | 19.67121887 | 19.67121887 | 2.316631568 | 0.36485697 | FALSE | TRUE | FALSE |
| CD200R1 | 0.3011657 | 0.80290236 | 1 | 0.30116568 | 0.30116568 | 0.30116568 | 0.34250818 | 0.811352438 | 1 | 0.342508185 | 1.370004937 | 1.370004937 | 0.879294842 | -0.055865475 | FALSE | TRUE | FALSE |
| OR4S1 | 0.3011511 | 0.80292498 | 1 | 0.301151068 | 0.301151068 | 0.301151068 | 27.942337 | 0.099015863 | 0.50205 | 27.942337 | 273.1056825 | 273.1056825 | 0.010777591 | -1.967478318 | FALSE | FALSE | FALSE |
| TRMT61A | 0.3011004 | 0.8029476 | 0.012 | 0.301100376 | 0.301100376 | 5.962714567 | 0.34694226 | 0.809957692 | 0.01005 | 0.346942263 | 0.346942263 | 6.878662192 | 0.867868829 | -0.06154591 | FALSE | TRUE | FALSE |

| | | | | | | | | | | | | | | | | | |
|---|---|---|---|---|---|---|---|---|---|---|---|---|---|---|---|---|---|
| HP1BP3 | 0.3010289 | 0.80297022 | 1 | 0.301028896 | 0.301028896 | 5.962298722 | 0.33024172 | 0.815309533 | 1 | 0.330241723 | 0.330241723 | 6.548515585 | 0.911541076 | -0.040223756 | FALSE | TRUE | FALSE |
| WDFY3 | 0.3007091 | 0.80299283 | 0.54185 | 0.78092506 | 7.513424112 | 140.7474552 | 0.57348547 | 0.723152916 | 0.028 | 0.017311339 | 0.326777367 | 0.573485467 | 0.524353476 | -0.280375849 | FALSE | TRUE | FALSE |
| ALG5 | 0.3006762 | 0.80301543 | 1 | 0.300676162 | 0.300676162 | 5.970662888 | 0.34662406 | 0.810024588 | 1 | 0.346624062 | 0.346624062 | 6.888970263 | 0.86744169 | -0.061759709 | FALSE | TRUE | FALSE |
| SMARCB1 | 0.3006519 | 0.80303803 | 1 | 0.300651917 | 0.300651917 | 5.95492802 | 0.34673681 | 0.809979996 | 1 | 0.346736815 | 0.346736815 | 6.875701906 | 0.867089688 | -0.061935979 | FALSE | TRUE | FALSE |
| AZGP1 | 0.3005122 | 0.80306063 | 0.2608 | 0.300512221 | 1.20192367 | 1.20192367 | 0.92566103 | 0.595025936 | 0.4992 | 0.100895545 | 0.925661025 | 0.925661025 | 0.324646077 | -0.48858984 | FALSE | TRUE | FALSE |
| RUNDC1 | 0.3004345 | 0.80308322 | 1 | 0.300434497 | 0.300434497 | 5.940521963 | 0.34597934 | 0.810313909 | 1 | 0.345979338 | 0.345979338 | 6.850422064 | 0.868359651 | -0.061300365 | FALSE | TRUE | FALSE |
| BAG6 | 0.3004177 | 0.8031058 | 1 | 2.82202508 | 2.82202508 | 55.05323121 | 0.35741004 | 0.806574157 | 1 | 0.357410039 | 0.357410039 | 6.993154015 | 0.840540689 | -0.075441258 | FALSE | TRUE | FALSE |
| SSB | 0.3003266 | 0.80312838 | 1 | 0.300326578 | 0.300326578 | 5.96804844 | 0.34564683 | 0.810424941 | 0.00815 | 0.345646831 | 0.345646831 | 0.345646831 | 0.868882775 | -0.061038812 | FALSE | TRUE | FALSE |
| KIF13A | 0.300261 | 0.80315095 | 0.1049 | 0.075097002 | 1.200836075 | 5.805688264 | 33.790167 | 0.082720804 | 0.39725 | 8.444053951 | 541.4947694 | 2168.496877 | 0.008886046 | -2.05129143 | FALSE | FALSE | FALSE |
| RGS16 | 0.3002261 | 0.80317352 | 1 | 0.300226116 | 0.300226116 | 0.300226116 | 0.34537805 | 0.810513674 | 1 | 0.345378047 | 0.345378047 | 0.345378047 | 0.869268091 | -0.060846262 | FALSE | TRUE | FALSE |
| SLC1A3 | 0.300163 | 0.80319609 | 0.01585 | 0.300162994 | 0.300162994 | 5.935465671 | 0.34607147 | 0.810247224 | 1 | 0.346071473 | 0.346071473 | 6.852563888 | 0.867343936 | -0.061808654 | FALSE | TRUE | FALSE |
| SCML4 | 0.3000235 | 0.80321865 | 1 | 0.300023508 | 0.300023508 | 0.300023508 | 0.34532337 | 0.810602317 | 1 | 0.345323366 | 0.345323366 | 0.345323366 | 0.86881902 | -0.06107068 | FALSE | TRUE | FALSE |
| PDS5A | 0.299934 | 0.8032412 | 1 | 0.299933997 | 0.299933997 | 10.0931155 | 0.34586507 | 0.810358338 | 1 | 0.345865069 | 0.345865069 | 6.783954381 | 0.86719945 | -0.061881006 | FALSE | TRUE | FALSE |
| SARS2 | 0.2998561 | 0.80326375 | 0.37695 | 0.024744428 | 2.762017653 | 2.762017653 | 0.06206013 | 0.884298711 | 0.5087 | 0.062060127 | 0.062060127 | 0.306962729 | 4.831702404 | 0.684100177 | FALSE | TRUE | FALSE |
| ORC2 | 0.2998054 | 0.80328629 | 0.0149 | 0.299805405 | 0.299805405 | 5.939341073 | 0.3453583 | 0.810535844 | 1 | 0.345358299 | 0.345358299 | 6.849564691 | 0.868099611 | -0.061430438 | FALSE | TRUE | FALSE |
| IP6K2 | 0.2996532 | 0.80330883 | 0.49825 | 0.299653179 | 2.757401567 | 2.757401567 | 29.0860524 | 0.094287997 | 0.25135 | 3.071300886 | 29.08605245 | 29.08605245 | 0.010302298 | -1.987065893 | FALSE | FALSE | FALSE |
| ICA1 | 0.2996115 | 0.80333136 | 1 | 3.2669E+134 | 2.2017E+152 | 3.0847E+160 | 0.00355768 | 0.898491794 | 1 | 6.3274E+113 | 4.0439E+130 | 3.3307E+137 | 84.21546938 | 1.925391874 | FALSE | TRUE | FALSE |
| RPS6KA1 | 0.2995493 | 0.80335389 | 0.9981 | 76.80612268 | 1275.454851 | 5109.699617 | 0.43365127 | 0.779792339 | 0.8769 | 1.734726959 | 6.941129874 | 8.544886015 | 0.690760871 | -0.160672272 | FALSE | TRUE | FALSE |
| CBLN3 | 0.2995332 | 0.80337641 | 1 | 2.808038374 | 11.22990759 | 11.22990759 | 0.34920542 | 0.809353527 | 1 | 3.278695952 | 3.278695952 | 3.278695952 | 0.857756359 | -0.066636053 | FALSE | TRUE | FALSE |
| SPRTN | 0.2994813 | 0.80339893 | 1 | 2.792393683 | 2.792393683 | 2.792393683 | 0.3520701 | 0.808428652 | 1 | 1.408127678 | 1.408127678 | 1.408127678 | 0.850629734 | -0.07025944 | FALSE | TRUE | FALSE |
| SART1 | 0.299452 | 0.80342144 | 1 | 0.299451998 | 0.299451998 | 5.8689938 | 0.34107661 | 0.811790899 | 0.02095 | 0.341076609 | 0.341076609 | 6.70093658 | 0.877961109 | -0.056524721 | FALSE | TRUE | FALSE |
| OR10G8 | 0.2993571 | 0.80344395 | 0.505 | 2.796595048 | 2.796595048 | 2.796595048 | 0.35597675 | 0.807104463 | 0.00825 | 0.355976745 | 0.355976745 | 0.355976745 | 0.840945711 | -0.07523204 | FALSE | TRUE | FALSE |
| HIPK1 | 0.2993405 | 0.80346645 | 1 | 0.299340473 | 0.299340473 | 5.854858329 | 0.34525348 | 0.810624464 | 1 | 0.345253476 | 0.345253476 | 6.770808912 | 0.867016536 | -0.061972619 | FALSE | TRUE | FALSE |
| TRMT10A | 0.299314 | 0.80348895 | 0.5061 | 0.299313996 | 0.299313996 | 0.681677003 | 0.4431737 | 0.776355165 | 0.50395 | 0.443173701 | 0.443173701 | 0.443173701 | 0.675387542 | -0.170446955 | FALSE | TRUE | FALSE |
| PLXDC1 | 0.2992293 | 0.80351144 | 0.2579 | 0.299229265 | 1.196835229 | 5.918013839 | 0.09940714 | 0.876082052 | 1 | 0.912418731 | 3.648663778 | 3.648663778 | 3.010138556 | 0.478586487 | FALSE | TRUE | FALSE |
| RAB17 | 0.299228 | 0.80353392 | 1 | 2.797728757 | 2.797728757 | 2.797728757 | 0.35193315 | 0.80854193 | 1 | 1.407514965 | 1.407514965 | 1.407514965 | 0.850240974 | -0.07045797 | FALSE | TRUE | FALSE |
| TMEM87A | 0.2992148 | 0.8035564 | 0.0151 | 0.29921484 | 0.29921484 | 5.922997688 | 0.34483664 | 0.810757237 | 1 | 0.344836644 | 0.344836644 | 6.834498472 | 0.867700243 | -0.061630281 | FALSE | TRUE | FALSE |
| LRRC14 | 0.2991922 | 0.80357888 | 0.0164 | 0.299192167 | 0.299192167 | 5.90371987 | 0.34471888 | 0.810779348 | 1 | 0.344718878 | 0.344718878 | 6.813022388 | 0.867930903 | -0.061514848 | FALSE | TRUE | FALSE |
| ELAC1 | 0.2991355 | 0.80360135 | 1 | 1.196336983 | 1.196336983 | 5.943701129 | 0.32762383 | 0.816085056 | 1 | 3.068487751 | 29.49577907 | 29.49577907 | 0.913045641 | -0.039507512 | FALSE | TRUE | FALSE |
| TKT | 0.2991141 | 0.80362381 | 0.5003 | 0.299114062 | 4.780421601 | 5.886625628 | 0.23831409 | 0.8424935 | 0.1408 | 0.238314092 | 0.952942688 | 2.019380482 | 1.255125364 | 0.098687106 | FALSE | TRUE | FALSE |
| IL36A | 0.2990371 | 0.80364627 | 0.0045 | 0.299037099 | 0.299037099 | 0.299037099 | 264.679093 | 0.009429186 | 0.0019 | 27.30310657 | 264.6790933 | 264.6790933 | 0.00112981 | -2.946994567 | FALSE | FALSE | FALSE |
| DAGLA | 0.2987637 | 0.80366872 | 1 | 0.29876371 | 0.29876371 | 5.797937867 | 0.3445127 | 0.810823553 | 1 | 0.344512703 | 0.344512703 | 6.709687324 | 0.867206656 | -0.061877397 | FALSE | TRUE | FALSE |
| CHAD | 0.2987441 | 0.80369117 | 0.01295 | 0.298744066 | 0.298744066 | 5.914769134 | 0.344252 | 0.81086774 | 1 | 0.344252004 | 0.344252004 | 6.824019487 | 0.867806326 | -0.061577189 | FALSE | TRUE | FALSE |
| PTAFR | 0.2987372 | 0.80371362 | 1 | 0.298737239 | 0.298737239 | 5.923916981 | 6.83327432 | 0.247671183 | 0.00935 | 0.34424302 | 0.34424302 | 0.34424302 | 0.043718022 | -1.359339493 | FALSE | FALSE | FALSE |
| RARRES1 | 0.2987209 | 0.80373605 | 1 | 0.298720935 | 0.298720935 | 0.298720935 | 0.34364784 | 0.81100018 | 1 | 0.343647839 | 0.343647839 | 0.343647839 | 0.869264697 | -0.060847958 | FALSE | TRUE | FALSE |
| ZNF668 | 0.2986091 | 0.80375849 | 0.0252 | 0.298609126 | 0.298609126 | 5.842742444 | 0.34431658 | 0.810845649 | 1 | 0.344316576 | 0.344316576 | 6.75466744 | 0.867251671 | -0.061854855 | FALSE | TRUE | FALSE |
| LRRTM1 | 0.2985005 | 0.80378092 | 1 | 2.813162676 | 2.813162676 | 55.35321074 | 0.35549811 | 0.807288214 | 0.01775 | 0.355498108 | 0.355498108 | 7.008580267 | 0.839668344 | -0.075892219 | FALSE | TRUE | FALSE |
| PSMF1 | 0.298318 | 0.80380334 | 1 | 0.298318 | 0.298318 | 0.298318 | 0.34365773 | 0.810978119 | 0.00635 | 0.343657734 | 0.343657734 | 0.343657734 | 0.868067181 | -0.061446663 | FALSE | TRUE | FALSE |
| HLA-B | 0.2983088 | 0.80382576 | 0.01525 | 0.298308801 | 0.298308801 | 5.899699343 | 0.34355509 | 0.811066329 | 1 | 0.343555095 | 0.343555095 | 6.803650229 | 0.868299743 | -0.061330327 | FALSE | TRUE | FALSE |
| ATF7IP2 | 0.2981966 | 0.80384817 | 1 | 2.766246754 | 2.766246754 | 5.896692939 | 2.90386942 | 0.418980244 | 0.9347 | 27.64796386 | 2702.572667 | 2702.572667 | 0.102689393 | -0.988474414 | FALSE | FALSE | FALSE |
| DLG4 | 0.2981589 | 0.80387058 | 0.02635 | 0.298158947 | 0.298158947 | 5.847960508 | 3.13819852 | 0.40419518 | 0.50905 | 3.138198519 | 3.138198519 | 61.69448406 | 0.095009588 | -1.022232567 | FALSE | FALSE | FALSE |
| ZDHHC20 | 0.2981478 | 0.80389298 | 0.01 | 0.29814781 | 0.29814781 | 0.354380979 | 0.34356589 | 0.811044285 | 0.00925 | 0.343565887 | 0.343565887 | 0.343565887 | 0.867803882 | -0.061578412 | FALSE | TRUE | FALSE |
| RBM4B | 0.2981476 | 0.80391538 | 1 | 0.298147634 | 0.298147634 | 5.907386364 | 0.3435987 | 0.811022235 | 1 | 0.343598703 | 0.343598703 | 6.815567494 | 0.867720489 | -0.061620148 | FALSE | TRUE | FALSE |
| ZFR2 | 0.2981127 | 0.80393777 | 0.19945 | 0.298112696 | 2.509611842 | 21.64537288 | 0.08234705 | 0.879733818 | 0.6615 | 0.690067885 | 52.17430103 | 470.1894416 | 3.620198766 | 0.558732416 | FALSE | TRUE | FALSE |
| VTI1B | 0.2980087 | 0.80396016 | 0.00745 | 0.298008686 | 0.298008686 | 0.298008686 | 28.7341842 | 0.095767424 | 0.00515 | 3.006197489 | 28.73418421 | 28.73418421 | 0.010371225 | -1.984169949 | FALSE | FALSE | FALSE |
| GCNT4 | 0.2979823 | 0.80398254 | 1 | 2.757150588 | 2.757150588 | 5.914527906 | 0.35406336 | 0.807700364 | 1 | 0.354063365 | 0.354063365 | 0.354063365 | 0.841607211 | -0.074890552 | FALSE | TRUE | FALSE |
| SYPL2 | 0.2978597 | 0.80400492 | 1 | 0.297859658 | 0.297859658 | 0.297859658 | 0.34288126 | 0.811264499 | 1 | 0.342881263 | 0.342881263 | 0.342881263 | 0.868696222 | -0.061132067 | FALSE | TRUE | FALSE |
| SPNS1 | 0.2978545 | 0.80402729 | 1 | 2.786431453 | 11.14465117 | 54.9152803 | 0.35855009 | 0.806133816 | 0.01575 | 0.358550093 | 0.358550093 | 7.078754961 | 0.830719486 | -0.080545602 | FALSE | TRUE | FALSE |
| PARD6A | 0.2978458 | 0.80404966 | 1 | 0.297845755 | 0.297845755 | 5.890708181 | 0.32222358 | 0.818107262 | 1 | 0.322223577 | 0.322223577 | 6.381366941 | 0.92434501 | -0.034165899 | FALSE | TRUE | FALSE |
| OR8D1 | 0.2978346 | 0.80407202 | 1 | 0.297834635 | 2.756870235 | 2.756870235 | 0.35390025 | 0.807746048 | 1 | 0.353900254 | 0.353900254 | 0.353900254 | 0.841577908 | -0.074905674 | FALSE | TRUE | FALSE |
| CD4 | 0.2976923 | 0.80409437 | 1 | 0.297692349 | 0.297692349 | 0.297692349 | 0.34280193 | 0.811286491 | 1 | 0.34280193 | 0.34280193 | 0.34280193 | 0.868409179 | -0.061275585 | FALSE | TRUE | FALSE |
| RALGPS2 | 0.2975937 | 0.80411672 | 0.5077 | 2.764282924 | 2.764282924 | 10.06728448 | 1.23618977 | 0.563148474 | 0.96755 | 109.440252 | 2680.715447 | 10722.58606 | 0.240734642 | -0.618461409 | FALSE | FALSE | FALSE |
| FSTL5 | 0.2975736 | 0.80413907 | 0.5085 | 0.297573568 | 0.297573568 | 2.514161398 | 0.10202157 | 0.875487771 | 1 | 0.102021568 | 0.102021568 | 2.015392499 | 2.916771178 | 0.46490236 | FALSE | TRUE | FALSE |
| MTX2 | 0.2975377 | 0.80416141 | 1 | 0.297537701 | 0.297537701 | 0.297537701 | 0.33140832 | 0.814887593 | 1 | 0.331408319 | 0.331408319 | 0.331408319 | 0.897797925 | -0.046821403 | FALSE | TRUE | FALSE |
| VHL | 0.2975154 | 0.80418374 | 0.0071 | 0.297515381 | 0.297515381 | 0.297515381 | 0.40449287 | 0.790844312 | 0.49785 | 0.101180032 | 0.404492872 | 0.404492872 | 0.735526882 | -0.13340145 | FALSE | TRUE | FALSE |
| GPR171 | 0.2975115 | 0.80420607 | 1 | 0.297511452 | 0.297511452 | 0.297511452 | 0.3427474 | 0.811308478 | 0.00775 | 0.342747404 | 0.342747404 | 0.342747404 | 0.868019563 | -0.061470487 | FALSE | TRUE | FALSE |
| TMEM82 | 0.2975015 | 0.8042284 | 1 | 1.18999502 | 4.761126013 | 5.900590893 | 5.36131059 | 0.291260209 | 0.31705 | 5.361310588 | 85.88553399 | 85.88553399 | 0.055490445 | -1.255781794 | FALSE | FALSE | FALSE |
| PRKACB | 0.297425 | 0.80425072 | 1 | 0.297425011 | 0.297425011 | 0.297425011 | 0.34112355 | 0.811769025 | 1 | 0.341123548 | 0.341123548 | 0.341123548 | 0.871898211 | -0.059534213 | FALSE | TRUE | FALSE |
| DNAJB4 | 0.2973379 | 0.80427303 | 1 | 1.189192848 | 1.189192848 | 5.910703979 | 0.3429675 | 0.81119849 | 1 | 0.342967503 | 1.371708378 | 1.371708378 | 0.86695656 | -0.062002663 | FALSE | TRUE | FALSE |
| TMEM147 | 0.2973124 | 0.80429534 | 1 | 0.297312361 | 0.297312361 | 0.297312361 | 0.32606511 | 0.816750413 | 1 | 0.326065108 | 0.326065108 | 0.326065108 | 0.911819 | -0.040091363 | FALSE | TRUE | FALSE |
| ELL | 0.2971655 | 0.80431764 | 0.0227 | 0.297165456 | 0.297165456 | 5.838870568 | 0.32471217 | 0.817287367 | 1 | 0.324712168 | 0.324712168 | 6.393622068 | 0.915165753 | -0.03850024 | FALSE | TRUE | FALSE |
| APOBEC3B | 0.2971469 | 0.80433994 | 1 | 0.297146945 | 0.297146945 | 5.886759166 | 28.4636006 | 0.096949541 | 0.25855 | 2.985620548 | 28.46360057 | 28.46360057 | 0.010439542 | -1.981318567 | FALSE | FALSE | FALSE |
| SPATA22 | 0.2970843 | 0.80436224 | 1 | 0.297084283 | 0.297084283 | 0.297084283 | 0.34246726 | 0.811396376 | 1 | 0.342467265 | 0.342467265 | 0.342467265 | 0.867482278 | -0.061739389 | FALSE | TRUE | FALSE |
| CCDC142 | 0.2970638 | 0.80438452 | 0.7606 | 1.188144594 | 10.75812838 | 23.33397276 | 0.05175225 | 0.886270532 | 1 | 0.460755712 | 0.460755712 | 9.068447711 | 5.740114279 | 0.758920539 | FALSE | TRUE | FALSE |
| EDDM3B | 0.2970398 | 0.80440681 | 1 | 0.297039839 | 0.297039839 | 0.297039839 | 0.33158543 | 0.814845289 | 1 | 3.061573231 | 3.061573231 | 3.061573231 | 0.89581693 | -0.047780734 | FALSE | TRUE | FALSE |
| SLC23A1 | 0.2968972 | 0.80442908 | 1 | 0.296897218 | 0.296897218 | 5.861753624 | 0.3424166 | 0.811440292 | 1 | 0.342416602 | 0.342416602 | 6.770897954 | 0.867064319 | -0.061948685 | FALSE | TRUE | FALSE |
| OR1A1 | 0.2968607 | 0.80445136 | 0.75355 | 2.753205476 | 26.20692084 | 26.20692084 | 3.18233863 | 0.401336517 | 0.74585 | 3.182338638 | 30.37492464 | 30.37492464 | 0.093283824 | -1.030193657 | FALSE | FALSE | FALSE |
| UBQLN1 | 0.2967571 | 0.80447362 | 1 | 26.85558865 | 2689.579251 | 2689.579251 | 0.38854663 | 0.79626051 | 1 | 0.38854663 | 0.38854663 | 7.699251742 | 0.763761796 | -0.117042069 | FALSE | TRUE | FALSE |
| PTDSS1 | 0.296749 | 0.80449589 | 1 | 0.296748999 | 0.296748999 | 5.877983241 | 0.3422941 | 0.811462243 | 1 | 0.342294104 | 0.342294104 | 6.7879695 | 0.866941603 | -0.062010156 | FALSE | TRUE | FALSE |
| GPRIN2 | 0.2967384 | 0.80451814 | 1 | 2.745937231 | 2.745937231 | 5.872779429 | 0.35273946 | 0.808156212 | 1 | 0.352739457 | 0.352739457 | 6.989874253 | 0.841239513 | -0.075080337 | FALSE | TRUE | FALSE |
| SWAP70 | 0.2965881 | 0.8045404 | 1 | 0.296588132 | 0.296588132 | 5.872045265 | 0.33170932 | 0.814802967 | 0.50835 | 3.098548491 | 3.098548491 | 6.57535066 | 0.894120581 | -0.048603908 | FALSE | TRUE | FALSE |
| C4orf19 | 0.2965674 | 0.80456264 | 1 | 0.296567408 | 0.296567408 | 0.296567408 | 0.34158049 | 0.811637673 | 1 | 0.341580493 | 0.341580493 | 0.341580493 | 0.868221149 | -0.061369639 | FALSE | TRUE | FALSE |
| ZNF639 | 0.2965446 | 0.80458489 | 1 | 26.16100149 | 255.3220308 | 255.3220308 | 0.37525369 | 0.800808001 | 0.0097 | 0.375253692 | 0.375253692 | 0.375253692 | 0.790251059 | -0.102234914 | FALSE | TRUE | FALSE |

| | | | | | | | | | | | | | | | | | |
|---|---|---|---|---|---|---|---|---|---|---|---|---|---|---|---|---|---|
| ISOC1 | 0.2965398 | 0.80460712 | 0.8776 | 2.67837648 | 24.82918449 | 24.82918449 | 0.10772498 | 0.873833091 | 1 | 0.107724985 | 0.107724985 | 0.107724985 | 2.752748599 | 0.43976655 | FALSE | TRUE | FALSE |
| TMEM109 | 0.2965103 | 0.80462935 | 0.0077 | 0.296510327 | 0.296510327 | 0.296510327 | 0.34156616 | 0.811659578 | 0.0072 | 0.341566163 | 0.341566163 | 0.341566163 | 0.868090459 | -0.061435017 | FALSE | TRUE | FALSE |
| ZDHHC2 | 0.296506 | 0.80465158 | 1 | 0.296506038 | 0.296506038 | 0.296506038 | 0.33133042 | 0.814972139 | 1 | 3.073540857 | 3.073540857 | 3.073540857 | 0.894895307 | -0.04822777 | FALSE | TRUE | FALSE |
| SHISA2 | 0.2964753 | 0.8046738 | 1 | 0.296475301 | 0.296475301 | 0.296475301 | 0.31035331 | 0.821941164 | 1 | 0.310353311 | 0.310353311 | 0.310353311 | 0.95528319 | -0.019867864 | FALSE | TRUE | FALSE |
| ENOPH1 | 0.2964342 | 0.80469601 | 0.0076 | 0.296434156 | 0.296434156 | 0.296434156 | 0.34173718 | 0.81155 | 1 | 0.341737185 | 0.341737185 | 0.341737185 | 0.867433131 | -0.061763994 | FALSE | TRUE | FALSE |
| HSPA9 | 0.2964135 | 0.80471822 | 0.0177 | 0.296413506 | 0.296413506 | 5.848582662 | 0.33155492 | 0.814866443 | 1 | 3.099246489 | 3.099246489 | 11.21814284 | 0.894010283 | -0.048657486 | FALSE | TRUE | FALSE |
| LDLRAP1 | 0.296367 | 0.80474043 | 1 | 0.296366973 | 0.296366973 | 5.88748797 | 0.34181228 | 0.811506131 | 1 | 0.341812276 | 0.341812276 | 0.341812276 | 0.867046019 | -0.061957852 | FALSE | TRUE | FALSE |
| SCNN1B | 0.296115 | 0.80476263 | 0.51505 | 0.296115017 | 0.296115017 | 2.50612972 | 0.90282336 | 0.598459072 | 0.01765 | 0.902823362 | 0.902823362 | 3.334638255 | 0.327987765 | -0.484142357 | FALSE | TRUE | FALSE |
| MRPS18A | 0.2959219 | 0.80478483 | 1 | 1.183625419 | 4.735436777 | 4.735436777 | 0.34884693 | 0.809443276 | 1 | 0.348846926 | 0.348846926 | 0.348846926 | 0.848285773 | -0.071457817 | FALSE | TRUE | FALSE |
| RFFL | 0.2959166 | 0.80480702 | 1 | 0.295916559 | 0.295916559 | 0.295916559 | 0.34159975 | 0.811593848 | 1 | 0.341599752 | 0.341599752 | 0.341599752 | 0.866266901 | -0.062348279 | FALSE | TRUE | FALSE |
| UQCRC1 | 0.295847 | 0.8048292 | 0.01665 | 0.295847034 | 0.295847034 | 5.842814599 | 0.34154231 | 0.811681477 | 1 | 0.341542309 | 0.341542309 | 6.755438803 | 0.866209036 | -0.06237729 | FALSE | TRUE | FALSE |
| C8orf33 | 0.2956674 | 0.80485138 | 1 | 0.295667439 | 0.295667439 | 0.295667439 | 0.34065983 | 0.811943874 | 1 | 0.340659832 | 0.340659832 | 0.340659832 | 0.867925748 | -0.061517428 | FALSE | TRUE | FALSE |
| UBXN1 | 0.2956634 | 0.80487356 | 1 | 0.295663408 | 0.295663408 | 5.865439107 | 0.33046985 | 0.8152253 | 1 | 3.063765275 | 3.063765275 | 3.063765275 | 0.894675882 | -0.04833427 | FALSE | TRUE | FALSE |
| TSHZ2 | 0.2956225 | 0.80489573 | 1 | 1.182473127 | 1.182473127 | 23.14337294 | 0.34520891 | 0.810646606 | 0.02375 | 0.345208914 | 0.345208914 | 6.77384735 | 0.856358192 | -0.067344544 | FALSE | TRUE | FALSE |
| MCHR2 | 0.2956136 | 0.8049179 | 1 | 0.295613601 | 0.295613601 | 0.295613601 | 0.34091387 | 0.811812769 | 1 | 0.340913875 | 0.340913875 | 0.340913875 | 0.867121062 | -0.061920265 | FALSE | TRUE | FALSE |
| SAAL1 | 0.2955596 | 0.80494006 | 1 | 0.29555959 | 0.29555959 | 5.860017295 | 3.06161551 | 0.408548446 | 0.5035 | 3.061615508 | 3.061615508 | 6.555904437 | 0.096537135 | -1.015305593 | FALSE | FALSE | FALSE |
| TRIM14 | 0.2955526 | 0.80496221 | 1 | 1.182121454 | 1.182121454 | 5.83297754 | 0.34533333 | 0.810580165 | 1 | 0.345333329 | 0.345333329 | 6.826273746 | 0.855847402 | -0.067603663 | FALSE | TRUE | FALSE |
| OR11L1 | 0.2953792 | 0.80498436 | 1 | 0.295379152 | 0.295379152 | 0.295379152 | 0.340539 | 0.811965707 | 1 | 0.340538997 | 0.340538997 | 0.340538997 | 0.867387156 | -0.061787013 | FALSE | TRUE | FALSE |
| ADRA1B | 0.2952629 | 0.80500651 | 1 | 0.295262929 | 0.295262929 | 5.849711234 | 0.34080045 | 0.811878345 | 1 | 0.340800446 | 0.340800446 | 0.340800446 | 0.866380699 | -0.062291231 | FALSE | TRUE | FALSE |
| FAM181B | 0.2950998 | 0.80502865 | 1 | 262.7638271 | 27536.59295 | 292167.2654 | 0.38669902 | 0.796952386 | 1 | 0.386699025 | 0.386699025 | 0.386699025 | 0.763125161 | -0.117404227 | FALSE | TRUE | FALSE |
| FLRT2 | 0.2950864 | 0.80505078 | 1 | 0.295086406 | 0.295086406 | 5.811261612 | 3.10479188 | 0.40528848 | 0.5027 | 3.104791875 | 3.104791875 | 11.68614675 | 0.09504225 | -1.02208329 | FALSE | FALSE | FALSE |
| CTBP1 | 0.2950526 | 0.80507292 | 0.5063 | 1.179891706 | 1.179891706 | 5.836397825 | 0.34489399 | 0.810690874 | 1 | 0.344893994 | 0.344893994 | 6.831297188 | 0.855487683 | -0.067786239 | FALSE | TRUE | FALSE |
| MARK4 | 0.2950088 | 0.80509504 | 1 | 0.295008824 | 0.295008824 | 5.798271191 | 0.34078583 | 0.811900193 | 1 | 0.340785827 | 0.340785827 | 6.711926249 | 0.865672222 | -0.062646518 | FALSE | TRUE | FALSE |
| ILF3 | 0.2949839 | 0.80511716 | 1 | 0.29498392 | 0.29498392 | 5.76789796 | 0.34084183 | 0.811834633 | 1 | 0.34084183 | 0.34084183 | 6.682503234 | 0.865456919 | -0.062754546 | FALSE | TRUE | FALSE |
| SNX2 | 0.2949391 | 0.80513928 | 1 | 0.29493908 | 0.29493908 | 5.846112837 | 0.34049435 | 0.811987534 | 1 | 0.340494351 | 0.340494351 | 6.756331007 | 0.866208439 | -0.062377589 | FALSE | TRUE | FALSE |
| GADL1 | 0.2949121 | 0.80516139 | 0.752 | 1.179272106 | 10.77779077 | 10.77779077 | 0.20374315 | 0.851184855 | 0.5049 | 0.814940831 | 3.260450942 | 3.260450942 | 1.447470104 | 0.160609603 | FALSE | TRUE | FALSE |
| TSPAN8 | 0.2948085 | 0.80518349 | 0.50595 | 1.179057399 | 1.179057399 | 1.179057399 | 0.34418356 | 0.810911905 | 1 | 0.344183561 | 0.344183561 | 0.344183561 | 0.856544512 | -0.067250063 | FALSE | TRUE | FALSE |
| FBXO2 | 0.2947408 | 0.80520559 | 1 | 2.721706279 | 2.721706279 | 2.721706279 | 1.38641586 | 0.546500155 | 0.50105 | 1.38641586 | 1.38641586 | 1.38641586 | 0.212591924 | -0.672453238 | FALSE | FALSE | FALSE |
| TRIM8 | 0.2946743 | 0.80522769 | 1 | 2.773646388 | 26.79446642 | 54.57696825 | 0.3098647 | 0.822097352 | 1 | 277.7714761 | 2816.323311 | 29242.32221 | 0.950977171 | -0.021829909 | FALSE | TRUE | FALSE |
| UBE2T | 0.2946096 | 0.80524978 | 1 | 0.294609556 | 0.294609556 | 0.294609556 | 0.34007972 | 0.812096597 | 1 | 0.340079719 | 0.340079719 | 0.340079719 | 0.866295576 | -0.062333904 | FALSE | TRUE | FALSE |
| APBB1 | 0.2945454 | 0.80527186 | 1 | 2.745247198 | 2.745247198 | 54.01193978 | 0.3401071 | 0.812074795 | 1 | 3.17683997 | 3.17683997 | 11.51219451 | 0.8660371 | -0.062463503 | FALSE | TRUE | FALSE |
| TCHHL1 | 0.2944717 | 0.80529394 | 0.02075 | 0.294471656 | 0.294471656 | 1.186821078 | 0.051072 | 0.886353478 | 0.0175 | 0.051072 | 0.051072 | 1.009488597 | 5.765814105 | 0.760860636 | FALSE | TRUE | FALSE |
| OR2AK2 | 0.2943172 | 0.80531602 | 0.50815 | 2.715835977 | 2.715835977 | 2.715835977 | 27.9675697 | 0.09872018 | 0.50395 | 27.96756968 | 272.0843613 | 272.0843613 | 0.010523516 | -1.977839124 | FALSE | FALSE | FALSE |
| TNFAIP8L3 | 0.2943143 | 0.80533809 | 0.2623 | 0.294314313 | 2.798838685 | 2.798838685 | 0.03805859 | 0.889433807 | 0.00635 | 0.038058587 | 0.038058587 | 0.038058587 | 7.733190791 | 0.888358725 | FALSE | TRUE | FALSE |
| STXBP6 | 0.2942572 | 0.80536015 | 0.7621 | 0.294257156 | 2.826052666 | 2.826052666 | 0.03616823 | 0.889858184 | 1 | 0.036168226 | 0.036168226 | 0.036168226 | 8.135791872 | 0.91039983 | FALSE | TRUE | FALSE |
| ADCK1 | 0.2941391 | 0.80538221 | 0.01205 | 0.073585613 | 0.294139144 | 0.65404286 | 0.05718908 | 0.885241556 | 0.8777 | 0.228673239 | 2.045903737 | 2.045903737 | 5.143274661 | 0.711239717 | FALSE | TRUE | FALSE |
| PRND | 0.2941348 | 0.80540427 | 1 | 0.294134795 | 0.294134795 | 0.294134795 | 0.3399145 | 0.812140187 | 1 | 0.339914496 | 0.339914496 | 0.339914496 | 0.865319954 | -0.062823282 | FALSE | TRUE | FALSE |
| MYD88 | 0.2941056 | 0.80542632 | 0.01035 | 0.294105638 | 0.294105638 | 5.835596057 | 0.33960347 | 0.812183758 | 0.0097 | 0.339603471 | 0.339603471 | 0.339603471 | 0.866026597 | -0.06246877 | FALSE | TRUE | FALSE |
| LAMTOR1 | 0.2940718 | 0.80544836 | 0.00435 | 0.294071811 | 0.294071811 | 0.294071811 | 0.30064002 | 0.825332743 | 1 | 0.300640024 | 0.300640024 | 0.300640024 | 0.978152569 | -0.0095934 | FALSE | TRUE | FALSE |
| CLPSL1 | 0.2940459 | 0.8054704 | 1 | 0.294045903 | 0.294045903 | 0.294045903 | 0.33882038 | 0.812401311 | 1 | 0.338820378 | 0.338820378 | 0.338820378 | 0.867851881 | -0.061554391 | FALSE | TRUE | FALSE |
| ESCO1 | 0.294041 | 0.80549243 | 1 | 0.294041048 | 0.294041048 | 5.799035325 | 0.33606619 | 0.81348138 | 1 | 1.344133913 | 1.344133913 | 11.35923828 | 0.874949803 | -0.058016862 | FALSE | TRUE | FALSE |
| WDR45B | 0.2940228 | 0.80551446 | 1 | 1.175929386 | 1.175929386 | 5.83620284 | 0.34346422 | 0.811088368 | 1 | 0.343464216 | 0.343464216 | 0.343464216 | 0.856050885 | -0.067500419 | FALSE | TRUE | FALSE |
| MFAP3 | 0.2939873 | 0.80553649 | 1 | 0.293987313 | 0.293987313 | 0.293987313 | 0.32874637 | 0.8157086 | 1 | 0.328746375 | 3.043277921 | 3.043277921 | 0.894267847 | -0.048532384 | FALSE | TRUE | FALSE |
| ZNF398 | 0.2939515 | 0.80555851 | 1 | 0.293951541 | 0.293951541 | 5.817923457 | 0.33961089 | 0.812161975 | 0.01175 | 0.339610889 | 0.339610889 | 6.730023072 | 0.865553934 | -0.062705865 | FALSE | TRUE | FALSE |
| ALDH3A2 | 0.2939489 | 0.80558052 | 0.69455 | 0.29394892 | 4.703199059 | 5.824304438 | 1.87478327 | 0.500818876 | 0.32085 | 0.468719138 | 7.500633705 | 7.500633705 | 0.156790887 | -0.804679184 | FALSE | FALSE | FALSE |
| VPS11 | 0.2938502 | 0.80560253 | 0.5142 | 2.745168865 | 2.745168865 | 53.81009333 | 3.17777952 | 0.401557359 | 0.0256 | 3.177779521 | 3.177779521 | 62.43939748 | 0.092470297 | -1.033997747 | FALSE | FALSE | FALSE |
| TM4SF1 | 0.2938086 | 0.80562453 | 1 | 0.293808578 | 0.293808578 | 0.293808578 | 0.31848124 | 0.819202329 | 1 | 2.939979988 | 27.85459772 | 27.85459772 | 0.922530237 | -0.035019391 | FALSE | TRUE | FALSE |
| KPNA6 | 0.2937746 | 0.80564653 | 1 | 0.29377463 | 0.29377463 | 5.812620982 | 0.33595412 | 0.81350285 | 1 | 1.343538389 | 1.343538389 | 6.655740538 | 0.874448669 | -0.058265679 | FALSE | TRUE | FALSE |
| AVPR1B | 0.2936305 | 0.80566852 | 0.0117 | 0.293630496 | 0.293630496 | 5.809888897 | 0.33882794 | 0.812379578 | 0.01355 | 0.338827937 | 0.338827937 | 6.71279701 | 0.866606509 | -0.062178053 | FALSE | TRUE | FALSE |
| ELF1 | 0.2935334 | 0.80569051 | 1 | 0.293533444 | 0.293533444 | 5.806186765 | 0.32854909 | 0.815750507 | 1 | 3.055728308 | 3.055728308 | 6.507395069 | 0.893423391 | -0.048942681 | FALSE | TRUE | FALSE |
| PFDN1 | 0.2934492 | 0.80571249 | 1 | 0.293449175 | 0.293449175 | 0.293449175 | 0.29839622 | 0.825819934 | 1 | 0.298396224 | 0.298396224 | 0.298396224 | 0.983421207 | -0.007260431 | FALSE | TRUE | FALSE |
| EMC4 | 0.2934079 | 0.80573447 | 0.0051 | 0.293407857 | 0.293407857 | 0.293407857 | 0.32802705 | 0.815938845 | 0.50445 | 3.027878976 | 3.027878976 | 3.027878976 | 0.894462387 | -0.048437917 | FALSE | TRUE | FALSE |
| DACT1 | 0.2934039 | 0.80575644 | 1 | 2.752756949 | 2.752756949 | 54.02082171 | 0.34999713 | 0.809151279 | 1 | 0.349997132 | 0.349997132 | 6.883973318 | 0.838303624 | -0.076598656 | FALSE | TRUE | FALSE |
| ZNF530 | 0.2933997 | 0.80577841 | 0.96725 | 24.40890528 | 231.1330264 | 2242.192866 | 2997158.38 | 5.39763E-07 | 0.0377 | 28156.1849 | 2997158.382 | 31970034.27 | 9.78926E-08 | -7.009249996 | FALSE | FALSE | FALSE |
| DMTF1 | 0.2933546 | 0.80580037 | 1 | 0.293354578 | 0.293354578 | 5.787490924 | 0.33879234 | 0.812423038 | 1 | 0.338792336 | 0.338792336 | 6.694829532 | 0.865883161 | -0.062540706 | FALSE | TRUE | FALSE |
| PATE1 | 0.2932947 | 0.80582233 | 1 | 0.293294675 | 0.293294675 | 0.293294675 | 0.33815726 | 0.812618354 | 0.0024 | 0.338157264 | 0.338157264 | 0.338157264 | 0.867332172 | -0.061814544 | FALSE | TRUE | FALSE |
| LYZL6 | 0.2932152 | 0.80584428 | 0.0035 | 0.293215223 | 0.293215223 | 0.293215223 | 0.33848906 | 0.812509897 | 1 | 0.338489055 | 0.338489055 | 0.338489055 | 0.866247279 | -0.062358117 | FALSE | TRUE | FALSE |
| ANAPC11 | 0.2931029 | 0.80586623 | 1 | 0.293102872 | 0.293102872 | 0.293102872 | 0.33803771 | 0.812640031 | 1 | 0.338037707 | 0.338037707 | 0.338037707 | 0.867071531 | -0.061945073 | FALSE | TRUE | FALSE |
| SLC39A2 | 0.2931003 | 0.80588817 | 1 | 0.293100295 | 0.293100295 | 0.293100295 | 3.05331768 | 0.408980622 | 0.50465 | 3.053317681 | 3.053317681 | 3.053317681 | 0.095994039 | -1.017755736 | FALSE | FALSE | FALSE |
| ARNT | 0.2930765 | 0.80591011 | 1 | 0.293076471 | 0.293076471 | 5.773672097 | 0.33875626 | 0.81244476 | 1 | 0.338756262 | 0.338756262 | 6.685613818 | 0.865154401 | -0.062906379 | FALSE | TRUE | FALSE |
| GABRA4 | 0.2930599 | 0.80593204 | 1 | 0.293059893 | 0.293059893 | 5.795775159 | 0.33897732 | 0.812336097 | 0.01365 | 0.338977321 | 0.338977321 | 6.712875424 | 0.864541299 | -0.063214256 | FALSE | TRUE | FALSE |
| FPR3 | 0.2930244 | 0.80595397 | 1 | 0.293024404 | 0.293024404 | 5.817184743 | 0.3385119 | 0.81248819 | 1 | 0.338511898 | 0.338511898 | 0.338511898 | 0.865625127 | -0.062670145 | FALSE | TRUE | FALSE |
| CRLS1 | 0.2929707 | 0.80597589 | 0.00615 | 0.292970686 | 0.292970686 | 0.292970686 | 0.33819177 | 0.812596673 | 1 | 0.338191766 | 0.338191766 | 0.338191766 | 0.866285688 | -0.06233886 | FALSE | TRUE | FALSE |
| ITGB1BP1 | 0.2928568 | 0.80599781 | 1 | 0.292856823 | 0.292856823 | 0.292856823 | 0.32750541 | 0.816168498 | 1 | 3.023001732 | 3.023001732 | 3.023001732 | 0.894204545 | -0.048563127 | FALSE | TRUE | FALSE |
| ESR1 | 0.2927037 | 0.80601972 | 0.75585 | 0.292703669 | 1.170364921 | 5.776332905 | 0.10207269 | 0.875468198 | 1 | 0.10207269 | 0.10207269 | 2.017562843 | 2.86760024 | 0.457518608 | FALSE | TRUE | FALSE |
| ZNF239 | 0.2926279 | 0.80604163 | 0.75875 | 0.653675761 | 2.614507683 | 2.614507683 | 0.89912859 | 0.59909661 | 0.50065 | 0.899128585 | 0.899128585 | 1.984507132 | 0.325457284 | -0.487506004 | FALSE | TRUE | FALSE |
| KCNMB3 | 0.2925804 | 0.80606353 | 0.0055 | 0.292580383 | 0.292580383 | 0.292580383 | 0.33745057 | 0.812878136 | 1 | 0.337450574 | 0.337450574 | 0.337450574 | 0.867031813 | -0.061964967 | FALSE | TRUE | FALSE |
| GPR149 | 0.2923617 | 0.80608543 | 0.98515 | 10.5204808 | 168.5090659 | 669.2818051 | 0.90767515 | 0.597498919 | 0.75475 | 3.630477944 | 33.69371093 | 33.69371093 | 0.322099431 | -0.492010042 | FALSE | TRUE | FALSE |
| OR13A1 | 0.2920473 | 0.80610733 | 1 | 0.292047299 | 0.292047299 | 5.796199796 | 3.04458084 | 0.409627633 | 0.00805 | 3.04458084 | 3.04458084 | 3.04458084 | 0.095923647 | -1.018074316 | FALSE | FALSE | FALSE |
| ASB17 | 0.2920465 | 0.80612922 | 0.0061 | 0.292046534 | 0.292046534 | 0.292046534 | 0.33703398 | 0.813050908 | 1 | 0.337033977 | 0.337033977 | 0.337033977 | 0.866519564 | -0.062221627 | FALSE | TRUE | FALSE |
| GIP | 0.291996 | 0.8061511 | 1 | 0.291996018 | 0.291996018 | 0.291996018 | 0.33679311 | 0.813223345 | 1 | 0.336793109 | 0.336793109 | 0.336793109 | 0.866989287 | -0.061986269 | FALSE | TRUE | FALSE |
| OR2AE1 | 0.2919917 | 0.80617298 | 1 | 0.291991677 | 0.291991677 | 0.291991677 | 0.33699078 | 0.813094048 | 1 | 0.336990779 | 0.336990779 | 0.336990779 | 0.866467854 | -0.062247545 | FALSE | TRUE | FALSE |
| SEMA3F | 0.2919744 | 0.80619486 | 1 | 0.291974448 | 0.291974448 | 5.702778445 | 3.08018919 | 0.407465845 | 0.50905 | 3.080189186 | 3.080189186 | 60.3312507 | 0.094791076 | -1.023232546 | FALSE | FALSE | FALSE |

| | | | | | | | | | | | | | | | | | |
|---|---|---|---|---|---|---|---|---|---|---|---|---|---|---|---|---|---|
| ZNF689 | 0.2919526 | 0.80621673 | 1 | 0.291952566 | 0.291952566 | 5.743780113 | 0.33768995 | 0.812726688 | 1 | 0.337689946 | 0.337689946 | 6.656651554 | 0.864558065 | -0.063205834 | FALSE | TRUE | FALSE |
| ZNF423 | 0.2919305 | 0.80623859 | 1 | 0.291930539 | 0.291930539 | 5.645204197 | 0.33777442 | 0.812705031 | 1 | 0.337774421 | 0.337774421 | 6.557808895 | 0.864276632 | -0.063347229 | FALSE | TRUE | FALSE |
| HTR1E | 0.2918713 | 0.80626045 | 1 | 0.291871267 | 0.291871267 | 0.291871267 | 0.33749226 | 0.812856516 | 1 | 0.337492261 | 0.337492261 | 0.337492261 | 0.864823585 | -0.063072475 | FALSE | TRUE | FALSE |
| CTU1 | 0.2918068 | 0.8062823 | 0.49785 | 0.29180675 | 1.167148295 | 1.167148295 | 0.3429815 | 0.811176476 | 1 | 0.342981498 | 0.342981498 | 0.342981498 | 0.850794437 | -0.070175358 | FALSE | TRUE | FALSE |
| OR4N5 | 0.2916348 | 0.80630415 | 1 | 0.291634804 | 0.291634804 | 0.291634804 | 0.33682179 | 0.813201809 | 1 | 0.336821786 | 0.336821786 | 0.336821786 | 0.865843054 | -0.062560823 | FALSE | TRUE | FALSE |
| LRRC41 | 0.2916301 | 0.806326 | 1 | 0.291630096 | 0.291630096 | 5.72379939 | 0.33745004 | 0.81289975 | 1 | 0.337450037 | 0.337450037 | 6.637980584 | 0.864217108 | -0.063377141 | FALSE | TRUE | FALSE |
| GDPD1 | 0.2916286 | 0.80634784 | 1 | 0.29162861 | 0.29162861 | 0.29162861 | 0.33664205 | 0.813287922 | 1 | 0.33664205 | 0.33664205 | 0.33664205 | 0.866286935 | -0.062338235 | FALSE | TRUE | FALSE |
| C1orf146 | 0.2915344 | 0.80636967 | 1 | 0.291534363 | 0.291534363 | 0.291534363 | 0.33631195 | 0.813395448 | 1 | 0.336311947 | 0.336311947 | 0.336311947 | 0.866856991 | -0.062052544 | FALSE | TRUE | FALSE |
| CCR2 | 0.2913025 | 0.8063915 | 1 | 1.164997299 | 1.164997299 | 5.783464781 | 1.28866986 | 0.556116999 | 0.9681 | 20.62692923 | 82.5550805 | 330.4924244 | 0.226048966 | -0.645797475 | FALSE | FALSE | FALSE |
| POU4F3 | 0.2912373 | 0.80641333 | 0.01205 | 0.291237306 | 0.291237306 | 5.760231743 | 0.28786611 | 0.828760008 | 0.97185 | 261.1160517 | 2657.903014 | 27714.57489 | 1.011710992 | 0.005056468 | FALSE | TRUE | FALSE |
| RFX2 | 0.2911987 | 0.80643515 | 1 | 1.16464403 | 1.16464403 | 22.87771003 | 0.3408205 | 0.811856492 | 1 | 0.340820502 | 0.340820502 | 6.70928243 | 0.854405014 | -0.068336212 | FALSE | TRUE | FALSE |
| KCNN1 | 0.291116 | 0.80645696 | 1 | 0.291116003 | 0.291116003 | 5.737964607 | 0.33685829 | 0.813158721 | 1 | 0.336858293 | 0.336858293 | 6.651133417 | 0.864209101 | -0.063381164 | FALSE | TRUE | FALSE |
| TRMT44 | 0.2910225 | 0.80647877 | 0.85525 | 2.437797573 | 183.8630114 | 1653.630938 | 0.09519407 | 0.877092643 | 0.13055 | 0.095194067 | 0.797878031 | 0.797878031 | 3.05714919 | 0.485316633 | FALSE | TRUE | FALSE |
| MAP3K5 | 0.2909958 | 0.80650058 | 1 | 6719566.55 | 9021461375 | 2.88746E+11 | 0.05473132 | 0.885720457 | 1 | 72826.99 | 32056348.35 | 285792796.9 | 5.316805948 | 0.72565081 | FALSE | TRUE | FALSE |
| AATK | 0.290899 | 0.80652238 | 0.52245 | 0.290899037 | 0.290899037 | 5.683032879 | 0.04903423 | 0.886922819 | 1 | 0.049034233 | 0.049034233 | 0.960614696 | 5.932570405 | 0.773242901 | FALSE | TRUE | FALSE |
| SIRT1 | 0.2908037 | 0.80654418 | 1 | 0.29080368 | 0.29080368 | 5.75672748 | 0.32584222 | 0.816853925 | 1 | 3.03051208 | 3.03051208 | 6.458265124 | 0.892467774 | -0.049407457 | FALSE | TRUE | FALSE |
| COMMD5 | 0.2907616 | 0.80656597 | 1 | 0.290761612 | 0.290761612 | 0.290761612 | 0.33562065 | 0.813717269 | 0.0058 | 0.335620655 | 0.335620655 | 0.335620655 | 0.866340043 | -0.062311612 | FALSE | TRUE | FALSE |
| XPO1 | 0.2906905 | 0.80658775 | 1 | 0.290690537 | 0.290690537 | 5.710172827 | 0.33668069 | 0.813266401 | 1 | 0.336680694 | 0.336680694 | 6.62777423 | 0.863401263 | -0.06378732 | FALSE | TRUE | FALSE |
| OR2L13 | 0.2906584 | 0.80660954 | 1 | 0.29065839 | 0.29065839 | 0.29065839 | 0.33573541 | 0.813631565 | 1 | 0.335735409 | 0.335735409 | 0.335735409 | 0.865736477 | -0.062614283 | FALSE | TRUE | FALSE |
| GALNT14 | 0.2906323 | 0.80663131 | 0.8232 | 2.115965046 | 74.41704974 | 297.8693185 | 0.24069799 | 0.841847914 | 0.5048 | 0.240697989 | 2.133926609 | 2.133926609 | 1.207456491 | 0.081871491 | FALSE | TRUE | FALSE |
| FAIM2 | 0.2904412 | 0.80665308 | 0.00815 | 0.290441174 | 0.290441174 | 0.290441174 | 0.3356836 | 0.813674427 | 0.008 | 0.335683596 | 0.335683596 | 0.335683596 | 0.865223021 | -0.062871934 | FALSE | TRUE | FALSE |
| FITM2 | 0.2904273 | 0.80667485 | 1 | 0.290427262 | 0.290427262 | 0.290427262 | 0.33568245 | 0.81369585 | 1 | 0.335682446 | 0.335682446 | 0.335682446 | 0.865184538 | -0.062891251 | FALSE | TRUE | FALSE |
| RRP15 | 0.2903657 | 0.80669661 | 1 | 2.679113497 | 2.679113497 | 2.679113497 | 0.34584677 | 0.810380544 | 1 | 0.345846775 | 0.345846775 | 0.345846775 | 0.839579157 | -0.075938352 | FALSE | TRUE | FALSE |
| MRPS16 | 0.2903649 | 0.80671837 | 1 | 0.290364908 | 0.290364908 | 0.290364908 | 0.33530182 | 0.813824285 | 1 | 0.335301818 | 0.335301818 | 0.335301818 | 0.865980715 | -0.062491779 | FALSE | TRUE | FALSE |
| DDX21 | 0.2903219 | 0.80674012 | 1 | 2.70202217 | 2.70202217 | 9.810706123 | 0.33575972 | 0.813610126 | 0.50335 | 3.132265217 | 3.132265217 | 11.35448956 | 0.864671587 | -0.063148811 | FALSE | TRUE | FALSE |
| TMEM14B | 0.290293 | 0.80676187 | 0.00255 | 0.290292951 | 0.290292951 | 0.290292951 | 1.32571985 | 0.552907489 | 0.5022 | 1.325719851 | 1.325719851 | 1.325719851 | 0.218970057 | -0.659615269 | FALSE | FALSE | FALSE |
| SPEN | 0.2902471 | 0.80678361 | 0.5517 | 2.752321709 | 50.99521998 | 495.9846172 | 0.04852285 | 0.887013096 | 0.77675 | 0.450148219 | 8.414784616 | 80.12026437 | 5.981657538 | 0.776821545 | FALSE | TRUE | FALSE |
| PHF20L1 | 0.2902286 | 0.80680534 | 1 | 0.290228641 | 0.290228641 | 5.697242988 | 0.33584182 | 0.813524316 | 1 | 0.335841818 | 0.335841818 | 6.607316685 | 0.864182556 | -0.063394504 | FALSE | TRUE | FALSE |
| PALM | 0.2902158 | 0.80682708 | 1 | 0.290215755 | 0.290215755 | 5.748182512 | 0.27467408 | 0.832402372 | 0.87525 | 25.16671808 | 250.3054364 | 250.3054364 | 1.056582223 | 0.023903299 | FALSE | TRUE | FALSE |
| HIBADH | 0.2901133 | 0.8068488 | 1 | 0.290113327 | 0.290113327 | 0.290113327 | 0.33539582 | 0.81376009 | 1 | 0.335395815 | 0.335395815 | 0.335395815 | 0.864987916 | -0.062989959 | FALSE | TRUE | FALSE |
| NRG2 | 0.2900527 | 0.80687052 | 1 | 2.72021696 | 2.72021696 | 9.805114047 | 3.05320715 | 0.409196441 | 0.75605 | 3.053207149 | 29.40464555 | 60.22648784 | 0.094999353 | -1.022279352 | FALSE | FALSE | FALSE |
| STK35 | 0.2900454 | 0.80689224 | 1 | 0.290045445 | 0.290045445 | 5.743496396 | 0.33582013 | 0.813545776 | 1 | 0.335820135 | 0.335820135 | 6.657846183 | 0.863692837 | -0.063640682 | FALSE | TRUE | FALSE |
| TIMM10B | 0.290033 | 0.80691395 | 1 | 0.290032993 | 0.290032993 | 0.290032993 | 0.33533172 | 0.813802892 | 1 | 0.335331715 | 0.335331715 | 0.335331715 | 0.864913696 | -0.063027226 | FALSE | TRUE | FALSE |
| CALB1 | 0.2898706 | 0.80693566 | 1 | 2.7341798 | 2.7341798 | 2.7341798 | 0.3464377 | 0.810091436 | 1 | 0.346437704 | 0.346437704 | 0.346437704 | 0.836717743 | -0.077421021 | FALSE | TRUE | FALSE |
| SRPK2 | 0.2898173 | 0.80695736 | 1 | 0.28981734 | 0.28981734 | 5.717293318 | 0.33527942 | 0.813845673 | 1 | 0.335279415 | 0.335279415 | 6.624989792 | 0.864405408 | -0.063282525 | FALSE | TRUE | FALSE |
| ZNF501 | 0.2897439 | 0.80697905 | 1 | 2.692385869 | 25.67739225 | 25.67739225 | 0.32475501 | 0.817266775 | 1 | 3.022629774 | 282.894412 | 282.894412 | 0.892192381 | -0.04954149 | FALSE | TRUE | FALSE |
| GNPDA2 | 0.2897182 | 0.80700075 | 1 | 0.289718183 | 0.289718183 | 0.289718183 | 0.33484507 | 0.813973893 | 1 | 0.334845073 | 0.334845073 | 0.334845073 | 0.865230539 | -0.06286816 | FALSE | TRUE | FALSE |
| PACS2 | 0.2897015 | 0.80702243 | 1 | 0.289701539 | 0.289701539 | 5.67228911 | 0.33535944 | 0.813781494 | 1 | 0.335359439 | 0.335359439 | 6.582878212 | 0.86385384 | -0.063559732 | FALSE | TRUE | FALSE |
| MYCL | 0.2896729 | 0.80704411 | 1 | 0.28967287 | 0.28967287 | 5.745335415 | 0.33524807 | 0.813867056 | 1 | 0.335248072 | 0.335248072 | 0.335248072 | 0.864055291 | -0.063458466 | FALSE | TRUE | FALSE |
| EIF2AK1 | 0.2895987 | 0.80706579 | 0.50045 | 0.289598703 | 0.289598703 | 1.114792625 | 0.0501369 | 0.886568632 | 1 | 0.050136895 | 0.050136895 | 0.992597733 | 5.776159504 | 0.761639177 | FALSE | TRUE | FALSE |
| OLIG2 | 0.2895909 | 0.80708746 | 1 | 0.289590944 | 0.289590944 | 0.289590944 | 0.33093145 | 0.815098807 | 1 | 0.330931451 | 0.330931451 | 0.330931451 | 0.875078335 | -0.057953068 | FALSE | TRUE | FALSE |
| KCTD13 | 0.2895727 | 0.80710913 | 1 | 0.289572744 | 0.289572744 | 5.732487132 | 0.33510882 | 0.813931172 | 1 | 0.335108825 | 0.335108825 | 0.335108825 | 0.864115544 | -0.063428183 | FALSE | TRUE | FALSE |
| OR1N2 | 0.2895028 | 0.80713079 | 1 | 0.289502779 | 0.289502779 | 0.289502779 | 0.32322218 | 0.817657248 | 1 | 0.323222177 | 0.323222177 | 0.323222177 | 0.895677337 | -0.047848415 | FALSE | TRUE | FALSE |
| OR2S2 | 0.2893481 | 0.80715245 | 1 | 0.289348147 | 0.289348147 | 0.289348147 | 2.90242484 | 0.419398414 | 0.75385 | 2.902424842 | 27.50989325 | 27.50989325 | 0.099691865 | -1.001340277 | FALSE | FALSE | FALSE |
| MYOZ2 | 0.2892487 | 0.8071741 | 1 | 0.28924867 | 0.28924867 | 0.28924867 | 0.33471285 | 0.814037936 | 1 | 0.334712845 | 0.334712845 | 0.334712845 | 0.864169613 | -0.063401009 | FALSE | TRUE | FALSE |
| TMED5 | 0.2892457 | 0.80719575 | 1 | 0.289245714 | 0.289245714 | 0.289245714 | 0.33395415 | 0.814357471 | 1 | 0.333954152 | 0.333954152 | 0.333954152 | 0.866124023 | -0.062419915 | FALSE | TRUE | FALSE |
| CENPK | 0.289164 | 0.80721739 | 1 | 0.289163997 | 0.289163997 | 0.289163997 | 0.33397485 | 0.814336204 | 0.0055 | 0.333974855 | 0.333974855 | 0.333974855 | 0.865825653 | -0.062569551 | FALSE | TRUE | FALSE |
| PKN2 | 0.2890312 | 0.80723903 | 1 | 2.708076324 | 26.04139184 | 53.23298232 | 0.3243703 | 0.81736969 | 1 | 29.26504562 | 288.8330376 | 576.4538791 | 0.891053222 | -0.050096355 | FALSE | TRUE | FALSE |
| METTL9 | 0.2889428 | 0.80726066 | 0.00795 | 0.288942785 | 0.288942785 | 0.288942785 | 0.33387642 | 0.814399989 | 1 | 0.333876421 | 0.333876421 | 0.333876421 | 0.865418359 | -0.062773896 | FALSE | TRUE | FALSE |
| NPY2R | 0.2888974 | 0.80728229 | 1 | 0.288897365 | 0.288897365 | 0.288897365 | 0.33452094 | 0.814144573 | 0.0091 | 0.334520939 | 0.334520939 | 0.334520939 | 0.863615194 | -0.063679725 | FALSE | TRUE | FALSE |
| ZFP14 | 0.2888666 | 0.80730391 | 1 | 0.288866635 | 0.288866635 | 5.724395277 | 0.33415796 | 0.814272373 | 1 | 0.334157957 | 0.334157957 | 6.629219647 | 0.864461338 | -0.063254425 | FALSE | TRUE | FALSE |
| GJB7 | 0.2888027 | 0.80732553 | 0.89365 | 5.860922865 | 219.1238556 | 2099.285234 | 1.95169246 | 0.493458574 | 0.9051 | 70.14978469 | 6107.946023 | 59115.35942 | 0.147975528 | -0.829810101 | FALSE | FALSE | FALSE |
| RAB39A | 0.2887172 | 0.80734714 | 1 | 0.288717235 | 0.288717235 | 0.288717235 | 0.33421511 | 0.814208496 | 0.00635 | 0.334215111 | 0.334215111 | 0.334215111 | 0.863866489 | -0.063553373 | FALSE | TRUE | FALSE |
| TLX3 | 0.2886418 | 0.80736875 | 1 | 0.288641841 | 0.288641841 | 0.288641841 | 5.23129948 | 0.296903723 | 0.2626 | 1.307867612 | 5.231299476 | 5.231299476 | 0.055175935 | -1.258250297 | FALSE | FALSE | FALSE |
| GPN1 | 0.2885203 | 0.80739036 | 1 | 0.288520274 | 0.288520274 | 5.727402507 | 0.33367509 | 0.81446373 | 0.0088 | 0.333675088 | 0.333675088 | 0.333675088 | 0.864674302 | -0.063147448 | FALSE | TRUE | FALSE |
| RNASEH2A | 0.288509 | 0.80741195 | 0.5076 | 0.673225491 | 2.691308534 | 2.691308534 | 0.40605539 | 0.789917255 | 0.514 | 0.406055391 | 0.406055391 | 0.406055391 | 0.710516309 | -0.148425949 | FALSE | TRUE | FALSE |
| SCG2 | 0.2884202 | 0.80743355 | 0.0154 | 0.288420179 | 0.288420179 | 5.707215946 | 0.33357958 | 0.814506198 | 1 | 0.33357958 | 0.33357958 | 6.60924645 | 0.864621807 | -0.063173815 | FALSE | TRUE | FALSE |
| ZNF436 | 0.2883536 | 0.80745514 | 1 | 2.680952682 | 2.680952682 | 5.712865937 | 3.01215268 | 0.410489081 | 0.75735 | 3.012152676 | 28.78784409 | 28.78784409 | 0.095730082 | -1.01895157 | FALSE | FALSE | FALSE |
| STAG1 | 0.2883496 | 0.80747672 | 1 | 2.70934005 | 26.12723323 | 91.12069696 | 0.35580813 | 0.807173411 | 1 | 0.355808125 | 0.355808125 | 11.98214025 | 0.810407695 | -0.091296444 | FALSE | TRUE | FALSE |
| RPS24 | 0.2882583 | 0.8074983 | 1 | 0.288258309 | 0.288258309 | 0.288258309 | 0.3332442 | 0.814569865 | 1 | 0.3332442 | 0.3332442 | 0.3332442 | 0.865006229 | -0.062980765 | FALSE | TRUE | FALSE |
| XPOT | 0.2882108 | 0.80751988 | 1 | 0.288210844 | 0.288210844 | 5.669391398 | 0.33382783 | 0.814421241 | 1 | 0.333827832 | 0.333827832 | 6.579727107 | 0.863351753 | -0.063812225 | FALSE | TRUE | FALSE |
| SYT3 | 0.2878827 | 0.80754145 | 1 | 0.287882731 | 0.287882731 | 5.670462139 | 0.33346999 | 0.814548647 | 1 | 0.333469988 | 0.333469988 | 6.580372632 | 0.863294272 | -0.06384114 | FALSE | TRUE | FALSE |
| FNIP1 | 0.2878508 | 0.80756301 | 0.03115 | 0.287850845 | 0.287850845 | 9.692901323 | 0.33347971 | 0.814527425 | 1 | 0.333479715 | 0.333479715 | 6.545990539 | 0.863173476 | -0.063901913 | FALSE | TRUE | FALSE |
| CDC37 | 0.2877492 | 0.80758458 | 0.0132 | 0.287749235 | 0.287749235 | 5.692283533 | 0.31886019 | 0.81908131 | 1 | 0.318860187 | 0.318860187 | 6.315990983 | 0.902430742 | -0.044586119 | FALSE | TRUE | FALSE |
| KPNA1 | 0.2877093 | 0.80760613 | 1 | 1.150657767 | 1.150657767 | 5.700897524 | 0.33709226 | 0.813029329 | 1 | 0.337092256 | 0.337092256 | 6.68684975 | 0.853503201 | -0.068794846 | FALSE | TRUE | FALSE |
| ZYG11B | 0.2876718 | 0.80762768 | 1 | 0.287671847 | 0.287671847 | 5.676589102 | 0.32279658 | 0.817800685 | 1 | 3.003786261 | 3.003786261 | 10.91992669 | 0.891186164 | -0.050031565 | FALSE | TRUE | FALSE |
| AP1M1 | 0.2875166 | 0.80764923 | 1 | 0.287516614 | 0.287516614 | 5.678037146 | 27.9161078 | 0.099311193 | 0.00955 | 2.916990596 | 27.91610784 | 27.91610784 | 0.010299309 | -1.987191922 | FALSE | FALSE | FALSE |
| RBM47 | 0.287489 | 0.80767077 | 0.02585 | 0.287488989 | 0.287488989 | 5.638945792 | 0.32282365 | 0.817759726 | 1 | 3.030471455 | 3.030471455 | 59.57908989 | 0.890545003 | -0.050344129 | FALSE | TRUE | FALSE |
| PPP4R4 | 0.2873454 | 0.80769231 | 0.0237 | 0.287345431 | 0.287345431 | 5.653507972 | 0.09872279 | 0.876314403 | 1 | 0.098722793 | 0.098722793 | 1.946166488 | 2.910629077 | 0.463986864 | FALSE | TRUE | FALSE |
| FRMD4A | 0.2872808 | 0.80771385 | 0.08715 | 0.071882166 | 1.148419313 | 5.597507919 | 0.033809 | 0.8903962 | 0.75295 | 0.135108336 | 1.26528075 | 6.188313744 | 8.497169 | 0.929274256 | FALSE | TRUE | FALSE |
| DUSP4 | 0.2872311 | 0.80773537 | 1 | 0.287231109 | 0.287231109 | 0.287231109 | 0.33292318 | 0.814612288 | 1 | 0.332923181 | 0.332923181 | 0.332923181 | 0.86275491 | -0.06411256 | FALSE | TRUE | FALSE |
| OR10J3 | 0.2872184 | 0.8077569 | 1 | 0.287218444 | 0.287218444 | 0.287218444 | 0.33207788 | 0.814760624 | 1 | 0.332077881 | 0.332077881 | 0.332077881 | 0.864912903 | -0.063027624 | FALSE | TRUE | FALSE |
| HRH4 | 0.2871272 | 0.80777842 | 0.5032 | 0.287127189 | 0.287127189 | 0.287127189 | 0.86223315 | 0.604854544 | 0.5086 | 0.862233147 | 0.862233147 | 0.862233147 | 0.333004118 | -0.477550396 | FALSE | TRUE | FALSE |

| | | | | | | | | | | | | | | | | | |
|---|---|---|---|---|---|---|---|---|---|---|---|---|---|---|---|---|---|
| OR5D13 | 0.287105 | 0.80779993 | 1 | 0.28710503 | 0.28710503 | 0.28710503 | 27.3632674 | 0.101672112 | 0.0051 | 2.884766862 | 27.36326743 | 27.36326743 | 0.010492352 | -1.979127153 | FALSE | FALSE | FALSE |
| CD8A | 0.2870717 | 0.80782144 | 1 | 0.287071729 | 0.287071729 | 0.287071729 | 0.33255758 | 0.814675889 | 1 | 0.332557577 | 0.332557577 | 0.332557577 | 0.86322414 | -0.063876423 | FALSE | TRUE | FALSE |
| SH3GL2 | 0.2869915 | 0.80784294 | 1 | 25.6631024 | 252.1603042 | 252.1603042 | 0.36518495 | 0.804067294 | 1 | 0.365184948 | 0.365184948 | 0.365184948 | 0.785879871 | -0.104643835 | FALSE | TRUE | FALSE |
| ZNF775 | 0.2869131 | 0.80786444 | 0.01695 | 0.286913078 | 0.286913078 | 5.660871619 | 0.30127743 | 0.825125808 | 1 | 26.66609417 | 260.6786446 | 260.6786446 | 0.952321832 | -0.021216259 | FALSE | TRUE | FALSE |
| ZNF583 | 0.2868928 | 0.80788594 | 1 | 0.286892813 | 0.286892813 | 5.686773456 | 2.98633535 | 0.412847086 | 0.0122 | 2.986335348 | 2.986335348 | 6.386562162 | 0.096068519 | -1.017418906 | FALSE | FALSE | FALSE |
| MSL1 | 0.2868848 | 0.80790742 | 1 | 0.286884759 | 0.286884759 | 0.286884759 | 0.29898022 | 0.825745151 | 1 | 0.298980222 | 0.298980222 | 0.298980222 | 0.959544273 | -0.017934982 | FALSE | TRUE | FALSE |
| OR5B12 | 0.2867747 | 0.80792891 | 1 | 2.657114155 | 25.26803704 | 25.26803704 | 3.0837272 | 0.406814434 | 0.7482 | 3.083727199 | 29.40869519 | 29.40869519 | 0.092996127 | -1.03153514 | FALSE | FALSE | FALSE |
| FBXL8 | 0.2867519 | 0.80795039 | 1 | 0.286751918 | 0.286751918 | 0.286751918 | 2.86840776 | 0.421688498 | 0.7473 | 2.868407757 | 27.1693695 | 27.1693695 | 0.099969022 | -1.000134557 | FALSE | FALSE | FALSE |
| EBI3 | 0.2866519 | 0.80797186 | 1 | 25.2417599 | 246.1242727 | 246.1242727 | 0.36414579 | 0.804399032 | 1 | 0.364145791 | 0.364145791 | 0.364145791 | 0.787189811 | -0.103920536 | FALSE | TRUE | FALSE |
| ABHD8 | 0.2866284 | 0.80799333 | 1 | 2.657069914 | 2.657069914 | 5.653771038 | 2.98739564 | 0.412633491 | 0.2623 | 2.987395643 | 28.47680074 | 59.02412138 | 0.095945916 | -1.017973505 | FALSE | FALSE | FALSE |
| OR5B21 | 0.2864972 | 0.8080148 | 1 | 2.638547523 | 2.638547523 | 2.638547523 | 0.34165048 | 0.811571926 | 1 | 0.341650481 | 0.341650481 | 0.341650481 | 0.838568166 | -0.076461628 | FALSE | TRUE | FALSE |
| CTCFL | 0.2864944 | 0.80803626 | 0.7498 | 0.28649439 | 2.58501201 | 5.645099867 | 0.04965863 | 0.886725389 | 1 | 0.440736146 | 0.440736146 | 1.680881754 | 5.769276654 | 0.761121365 | FALSE | TRUE | FALSE |
| EPC1 | 0.2864276 | 0.80805771 | 0.97055 | 252.8204555 | 2550.194192 | 26351.68054 | 0.3882731 | 0.796337623 | 1 | 0.388273102 | 0.388273102 | 7.651060758 | 0.737696108 | -0.132122508 | FALSE | TRUE | FALSE |
| HNRNPC | 0.2863611 | 0.80807916 | 1 | 0.286361052 | 0.286361052 | 0.286361052 | 0.29616239 | 0.826360223 | 1 | 0.296162386 | 0.296162386 | 0.296162386 | 0.966905542 | -0.01461595 | FALSE | TRUE | FALSE |
| KLHDC8A | 0.2862044 | 0.80810061 | 0.7535 | 0.656152338 | 2.62365099 | 5.664463471 | 0.87933042 | 0.601628789 | 0.7487 | 0.879330422 | 8.299505859 | 8.299505859 | 0.325479871 | -0.487475864 | FALSE | TRUE | FALSE |
| SDF2L1 | 0.2860891 | 0.80812205 | 1 | 1.144059021 | 1.144059021 | 1.144059021 | 1.32652209 | 0.552778348 | 0.50245 | 1.326522086 | 1.326522086 | 1.326522086 | 0.21566858 | -0.666213121 | FALSE | FALSE | FALSE |
| GTSF1L | 0.2860629 | 0.80814349 | 1 | 0.286062908 | 0.286062908 | 0.286062908 | 0.33092161 | 0.815119901 | 1 | 0.330921614 | 0.330921614 | 0.330921614 | 0.864443109 | -0.063263583 | FALSE | TRUE | FALSE |
| ISCU | 0.2859064 | 0.80816492 | 0.0028 | 0.285906393 | 0.285906393 | 0.285906393 | 0.33123464 | 0.814993263 | 0.0036 | 0.331234636 | 0.331234636 | 0.331234636 | 0.863153675 | -0.063911876 | FALSE | TRUE | FALSE |
| OR52B6 | 0.2857545 | 0.80818635 | 1 | 0.285754539 | 0.285754539 | 0.285754539 | 0.32080574 | 0.818555015 | 0.5072 | 2.983637016 | 2.983637016 | 2.983637016 | 0.890740108 | -0.050248992 | FALSE | TRUE | FALSE |
| TRIM65 | 0.285678 | 0.80820777 | 0.9844 | 23.78311936 | 2186.2332 | 21722.49497 | 0.42623678 | 0.782446551 | 0.98385 | 36.11759889 | 3379.06394 | 33870.75334 | 0.670232999 | -0.173774194 | FALSE | TRUE | FALSE |
| RCVRN | 0.2856659 | 0.80822919 | 1 | 0.2856659 | 0.2856659 | 0.2856659 | 0.33098194 | 0.815077708 | 1 | 0.330981939 | 0.330981939 | 0.330981939 | 0.863086067 | -0.063945894 | FALSE | TRUE | FALSE |
| B9D1 | 0.2855982 | 0.8082506 | 0.50405 | 0.285598152 | 1.142141017 | 1.142141017 | 0.05005306 | 0.886609924 | 1 | 0.050053057 | 0.050053057 | 0.050053057 | 5.705908313 | 0.756324789 | FALSE | TRUE | FALSE |
| PDIA4 | 0.2855431 | 0.80827201 | 1 | 0.285543083 | 0.285543083 | 5.635207442 | 0.3311774 | 0.815035495 | 1 | 0.331177397 | 0.331177397 | 6.546221971 | 0.862205833 | -0.064389043 | FALSE | TRUE | FALSE |
| QTRT1 | 0.285436 | 0.80829342 | 0.87995 | 1.141439507 | 4.565681976 | 9.67704641 | 0.10081952 | 0.875741714 | 1 | 0.100819521 | 0.100819521 | 1.993751116 | 2.831157603 | 0.451964046 | FALSE | TRUE | FALSE |
| NPEPPS | 0.2851811 | 0.80831482 | 1 | 0.285181068 | 0.285181068 | 5.612497257 | 0.45546098 | 0.772677273 | 1 | 1.82183576 | 1.82183576 | 35.9235378 | 0.626137208 | -0.203330487 | FALSE | TRUE | FALSE |
| FAM118B | 0.285161 | 0.80833621 | 1 | 0.28516103 | 0.28516103 | 0.28516103 | 0.32012928 | 0.818717282 | 1 | 2.967857099 | 2.967857099 | 2.967857099 | 0.89076834 | -0.050235227 | FALSE | TRUE | FALSE |
| BCDIN3D | 0.285161 | 0.8083576 | 1 | 2.646433182 | 2.646433182 | 2.646433182 | 2.97716684 | 0.413486974 | 0.7515 | 2.977166841 | 28.40754038 | 28.40754038 | 0.095782661 | -1.018713101 | FALSE | FALSE | FALSE |
| OR5M9 | 0.2851365 | 0.80837899 | 1 | 0.285136476 | 0.285136476 | 0.285136476 | 0.31993198 | 0.818757804 | 1 | 2.956964006 | 2.956964006 | 2.956964006 | 0.891240946 | -0.050004869 | FALSE | TRUE | FALSE |
| FOXRED1 | 0.2851132 | 0.80840037 | 0.25665 | 0.031385538 | 0.285113164 | 0.620534688 | 0.24858115 | 0.839847782 | 0.25385 | 0.062197366 | 0.248581148 | 0.248581148 | 1.146962133 | 0.05954908 | FALSE | TRUE | FALSE |
| BMP2 | 0.2850657 | 0.80842174 | 1 | 1.139913954 | 1.139913954 | 5.64778175 | 0.33361261 | 0.814484967 | 1 | 0.333612608 | 0.333612608 | 0.333612608 | 0.85448108 | -0.068297549 | FALSE | TRUE | FALSE |
| ARL6IP6 | 0.2850112 | 0.80844311 | 0.00815 | 0.28501119 | 0.28501119 | 0.28501119 | 0.32976768 | 0.815414717 | 1 | 0.329767684 | 0.329767684 | 0.329767684 | 0.864278714 | -0.063346183 | FALSE | TRUE | FALSE |
| BCL2L14 | 0.2849353 | 0.80846448 | 1 | 0.284935261 | 0.284935261 | 0.284935261 | 0.32976809 | 0.81539369 | 1 | 0.329768086 | 0.329768086 | 0.329768086 | 0.864047411 | -0.063462427 | FALSE | TRUE | FALSE |
| SATB1 | 0.284854 | 0.80848584 | 0.0209 | 0.284854045 | 0.284854045 | 5.610759833 | 11.1701223 | 0.186058359 | 0.00615 | 0.330534758 | 0.330534758 | 6.522404609 | 0.025501426 | -1.593435539 | FALSE | FALSE | FALSE |
| DMRT2 | 0.2847051 | 0.8085072 | 1 | 0.284705066 | 0.284705066 | 0.284705066 | 0.32882587 | 0.815666675 | 1 | 0.328825867 | 0.328825867 | 0.328825867 | 0.865823206 | -0.062570778 | FALSE | TRUE | FALSE |
| MLLT6 | 0.2845711 | 0.80852855 | 1 | 0.284571097 | 0.284571097 | 5.562648033 | 0.33030144 | 0.815267427 | 1 | 0.330301437 | 0.330301437 | 6.474160235 | 0.86154968 | -0.064719675 | FALSE | TRUE | FALSE |
| KIF20B | 0.2843527 | 0.8085499 | 0.75185 | 2.468000627 | 199.4984149 | 1856.035306 | 1.9186E-06 | 0.901067941 | 0.5242 | 1.91858E-06 | 1.91858E-06 | 3.74109E-05 | 148210.2794 | 5.170878326 | FALSE | TRUE | FALSE |
| RAB13 | 0.2843315 | 0.80857124 | 1 | 2.671558465 | 10.68364342 | 10.68364342 | 0.34424431 | 0.810889825 | 1 | 0.344244312 | 0.344244312 | 0.344244312 | 0.825958335 | -0.08304186 | FALSE | TRUE | FALSE |
| SERINC1 | 0.2843209 | 0.80859258 | 1 | 0.284320923 | 0.284320923 | 5.640084028 | 0.32560224 | 0.816957315 | 0.50415 | 1.302394019 | 1.302394019 | 6.465312394 | 0.873215486 | -0.058878571 | FALSE | TRUE | FALSE |
| LRRC1 | 0.2842937 | 0.80861392 | 1 | 0.284293673 | 0.284293673 | 5.628148273 | 0.32956249 | 0.815484778 | 0.0133 | 0.329562486 | 0.329562486 | 6.532300598 | 0.862639667 | -0.064170576 | FALSE | TRUE | FALSE |
| ABCG4 | 0.2842935 | 0.80863525 | 0.51545 | 0.284293516 | 0.284293516 | 1.106274832 | 0.4152764 | 0.786462408 | 0.7554 | 0.415276402 | 3.883743808 | 8.19135817 | 0.684588661 | -0.164570298 | FALSE | TRUE | FALSE |
| LHPP | 0.2841899 | 0.80865657 | 1 | 24.82571617 | 241.1033539 | 241.1033539 | 0.36104654 | 0.805270368 | 1 | 0.361046539 | 0.361046539 | 0.361046539 | 0.787128111 | -0.103954577 | FALSE | TRUE | FALSE |
| JRKL | 0.2841742 | 0.80867789 | 1 | 0.284174164 | 0.284174164 | 5.641256212 | 10.8136629 | 0.189789103 | 0.0077 | 2.958312259 | 2.958312259 | 6.340964611 | 0.026279177 | -1.580388234 | FALSE | FALSE | FALSE |
| ECH1 | 0.2841672 | 0.8086992 | 0.75065 | 0.284167206 | 1.13667233 | 1.408254291 | 0.36655123 | 0.803591367 | 0.971 | 5.86657556 | 53.37414909 | 213.6085984 | 0.775245537 | -0.110560725 | FALSE | TRUE | FALSE |
| TRMT6 | 0.2841451 | 0.80872051 | 1 | 0.284145086 | 0.284145086 | 5.626851326 | 0.32919242 | 0.815540781 | 0.01245 | 0.329192421 | 0.329192421 | 6.526645966 | 0.863158044 | -0.063909678 | FALSE | TRUE | FALSE |
| GNAI2 | 0.2841324 | 0.80874182 | 1 | 0.284132411 | 0.284132411 | 5.625483314 | 0.32990132 | 0.815327657 | 1 | 0.329901318 | 0.329901318 | 6.539556875 | 0.861264855 | -0.064863274 | FALSE | TRUE | FALSE |
| GLE1 | 0.2841104 | 0.80876311 | 0.50745 | 0.284110351 | 0.284110351 | 1.070685169 | 0.18640142 | 0.855413302 | 0.9387 | 0.745563285 | 11.93660115 | 14.72081622 | 1.524185556 | 0.183037842 | FALSE | TRUE | FALSE |
| CP | 0.2841051 | 0.80878441 | 0.67295 | 1.13485223 | 133.2794875 | 1158.420011 | 0.03890152 | 0.889212597 | 0.81945 | 0.765365028 | 87.11442486 | 1393.463542 | 7.303187126 | 0.863512429 | FALSE | TRUE | FALSE |
| BPIFA2 | 0.2840855 | 0.8088057 | 1 | 0.284085529 | 0.284085529 | 0.284085529 | 0.32875619 | 0.81568764 | 1 | 0.328756188 | 0.328756188 | 0.328756188 | 0.864122226 | -0.063424824 | FALSE | TRUE | FALSE |
| TMX3 | 0.2840511 | 0.80882698 | 1 | 0.28405114 | 0.28405114 | 5.639690554 | 0.32893547 | 0.815603749 | 1 | 0.328935469 | 0.328935469 | 6.536559704 | 0.863546703 | -0.06371417 | FALSE | TRUE | FALSE |
| USP37 | 0.2840141 | 0.80884826 | 0.0253 | 0.28401413 | 0.28401413 | 5.586812205 | 0.32942429 | 0.815519781 | 0.02115 | 0.329424293 | 0.329424293 | 6.492907273 | 0.862152964 | -0.064415674 | FALSE | TRUE | FALSE |
| SQLE | 0.2839722 | 0.80886953 | 1 | 0.283972164 | 0.283972164 | 5.622665767 | 1.28863844 | 0.556244468 | 0.75085 | 1.288638443 | 5.155018084 | 10.9138191 | 0.22036605 | -0.656855313 | FALSE | FALSE | FALSE |
| LMNB2 | 0.283894 | 0.8088908 | 1 | 0.283894008 | 0.283894008 | 5.567670338 | 0.32602814 | 0.816771125 | 1 | 1.303843082 | 1.303843082 | 25.63047139 | 0.870765354 | -0.060098859 | FALSE | TRUE | FALSE |
| ZBTB6 | 0.2837968 | 0.80891207 | 0.008 | 0.283796769 | 0.283796769 | 0.283796769 | 0.32893137 | 0.815624729 | 1 | 0.328931374 | 0.328931374 | 0.328931374 | 0.862784128 | -0.064097853 | FALSE | TRUE | FALSE |
| TMEM69 | 0.2837734 | 0.80893333 | 1 | 0.283773366 | 0.283773366 | 0.283773366 | 2.87498739 | 0.420857682 | 0.762 | 2.874987387 | 27.44296117 | 27.44296117 | 0.098704212 | -1.005664312 | FALSE | FALSE | FALSE |
| CCNT2 | 0.2836157 | 0.80895458 | 1 | 2.633308262 | 25.09366547 | 51.94385979 | 0.33820561 | 0.812574986 | 1 | 3.151246063 | 3.151246063 | 11.44596641 | 0.838589622 | -0.076450517 | FALSE | TRUE | FALSE |
| E4F1 | 0.2835239 | 0.80897583 | 1 | 0.283523868 | 0.283523868 | 5.550156918 | 0.3288542 | 0.815645705 | 1 | 0.328854199 | 0.328854199 | 6.453979717 | 0.862156752 | -0.064413766 | FALSE | TRUE | FALSE |
| FAM110D | 0.2832782 | 0.80899708 | 1 | 0.283278236 | 0.283278236 | 0.283278236 | 0.28061071 | 0.830935841 | 1 | 2.647859231 | 2.647859231 | 2.647859231 | 1.009506154 | 0.004108971 | FALSE | TRUE | FALSE |
| USP1 | 0.2831899 | 0.80901833 | 0.0187 | 0.2831899 | 0.2831899 | 5.597480138 | 0.32845809 | 0.815771453 | 0.0153 | 0.328458089 | 0.328458089 | 6.501388967 | 0.862179712 | -0.064402201 | FALSE | TRUE | FALSE |
| METRN | 0.2831403 | 0.80903956 | 1 | 0.283140339 | 2.668809178 | 2.668809178 | 0.30700498 | 0.82312509 | 1 | 0.307004985 | 0.307004985 | 0.307004985 | 0.92226626 | -0.035143679 | FALSE | TRUE | FALSE |
| TNIP3 | 0.2830681 | 0.8090608 | 1 | 0.283068126 | 0.283068126 | 0.283068126 | 0.32844307 | 0.81581333 | 1 | 0.328443071 | 0.328443071 | 0.328443071 | 0.861848371 | -0.064569135 | FALSE | TRUE | FALSE |
| PRR11 | 0.2830619 | 0.80908203 | 1 | 2.610025544 | 24.70028334 | 24.70028334 | 0.3487645 | 0.8094657 | 1 | 0.348764504 | 0.348764504 | 0.348764504 | 0.811613383 | -0.090650801 | FALSE | TRUE | FALSE |
| SWT1 | 0.2829205 | 0.80910325 | 1 | 0.282920548 | 0.282920548 | 5.57764014 | 0.32828475 | 0.815855188 | 1 | 0.328284754 | 0.328284754 | 6.483091767 | 0.861814459 | -0.064586224 | FALSE | TRUE | FALSE |
| SIN3A | 0.2828902 | 0.80912447 | 1 | 0.282890237 | 0.282890237 | 9.515423529 | 0.32495225 | 0.817143121 | 1 | 1.299724443 | 1.299724443 | 25.40802636 | 0.870559415 | -0.060201583 | FALSE | TRUE | FALSE |
| PRKCE | 0.2828058 | 0.80914568 | 1 | 0.282805751 | 0.282805751 | 5.549127645 | 0.32845691 | 0.815792394 | 1 | 0.328456911 | 0.328456911 | 6.459560518 | 0.861013248 | -0.064990166 | FALSE | TRUE | FALSE |
| CAPZA3 | 0.2827039 | 0.80916689 | 0.50295 | 2.602865196 | 2.602865196 | 2.602865196 | 0.33766594 | 0.812748339 | 1 | 0.33766594 | 0.33766594 | 0.33766594 | 0.837229682 | -0.077155383 | FALSE | TRUE | FALSE |
| LUZP2 | 0.2826923 | 0.8091881 | 1 | 0.282692334 | 0.282692334 | 0.282692334 | 0.32743851 | 0.816189346 | 1 | 0.327438515 | 0.327438515 | 0.327438515 | 0.863344785 | -0.06381573 | FALSE | TRUE | FALSE |
| EGFL7 | 0.2826651 | 0.8092093 | 0.75375 | 0.64147965 | 2.565162172 | 2.565162172 | 0.10034009 | 0.875858597 | 0.00645 | 0.100340092 | 0.100340092 | 0.100340092 | 2.817070479 | 0.449797713 | FALSE | TRUE | FALSE |
| ACADS | 0.2826372 | 0.80923049 | 0.8943 | 8.145993578 | 357.0414867 | 4626.527132 | 0.73330716 | 0.649700765 | 0.498 | 0.328114103 | 11.72017102 | 46.87277388 | 0.385428116 | -0.414056608 | FALSE | TRUE | FALSE |
| STK25 | 0.282607 | 0.80925169 | 1 | 0.282606982 | 0.282606982 | 5.588806299 | 0.32799055 | 0.815980644 | 0.01225 | 0.32799055 | 0.32799055 | 6.495050925 | 0.861631479 | -0.064678443 | FALSE | TRUE | FALSE |
| DGCR8 | 0.2825071 | 0.80927287 | 1 | 0.282507059 | 0.282507059 | 5.541533154 | 0.32803831 | 0.815917938 | 1 | 0.328038309 | 0.328038309 | 6.4495488 | 0.861201425 | -0.06489526 | FALSE | TRUE | FALSE |
| CCSER1 | 0.2824545 | 0.80929405 | 0.01885 | 0.282454527 | 0.282454527 | 5.58231507 | 0.32760215 | 0.816105924 | 1 | 0.327602145 | 0.327602145 | 6.48380216 | 0.862187661 | -0.064398197 | FALSE | TRUE | FALSE |
| CD68 | 0.2823488 | 0.80931523 | 1 | 0.282348827 | 0.282348827 | 5.60722129 | 0.32720158 | 0.816251863 | 1 | 0.327201579 | 0.327201579 | 0.327201579 | 0.862920124 | -0.064029403 | FALSE | TRUE | FALSE |
| TBC1D21 | 0.282311 | 0.8093364 | 1 | 0.282310959 | 0.282310959 | 0.282310959 | 0.32705965 | 0.816293517 | 1 | 0.327059649 | 0.327059649 | 0.327059649 | 0.863178813 | -0.063899228 | FALSE | TRUE | FALSE |

| | | | | | | | | | | | | | | | | | |
|---|---|---|---|---|---|---|---|---|---|---|---|---|---|---|---|---|---|
| WLS | 0.282307 | 0.80935757 | 1 | 0.282306989 | 0.282306989 | 5.579986336 | 2.76417105 | 0.430072146 | 0.876 | 26.28882957 | 256.4380929 | 256.4380929 | 0.102130796 | -0.990843283 | FALSE | FALSE | FALSE |
| PROS1 | 0.2822939 | 0.80937873 | 0.3824 | 0.121761735 | 1.128107005 | 1.395308508 | 0.05467021 | 0.885762293 | 0.69525 | 0.218503073 | 3.492985198 | 4.321416845 | 5.163578136 | 0.712950753 | FALSE | TRUE | FALSE |
| CLP1 | 0.282262 | 0.80939989 | 1 | 0.282261971 | 0.282261971 | 5.591161332 | 0.32727842 | 0.816231029 | 1 | 0.327278416 | 0.327278416 | 6.490335433 | 0.862452142 | -0.064264995 | FALSE | TRUE | FALSE |
| PIGF | 0.282107 | 0.80942104 | 1 | 0.282106995 | 0.282106995 | 0.282106995 | 0.32686898 | 0.816418359 | 1 | 0.32686898 | 0.32686898 | 0.32686898 | 0.863058326 | -0.063959854 | FALSE | TRUE | FALSE |
| CAMK1G | 0.2820337 | 0.80944219 | 1 | 0.282033682 | 0.282033682 | 5.58658362 | 0.32712391 | 0.816272692 | 0.0108 | 0.327123909 | 0.327123909 | 6.487214416 | 0.862161629 | -0.06441131 | FALSE | TRUE | FALSE |
| KAT5 | 0.2820089 | 0.80946333 | 1 | 0.28200886 | 0.28200886 | 5.560430319 | 0.32734785 | 0.81621019 | 1 | 0.327347851 | 0.327347851 | 6.465372058 | 0.861495986 | -0.064746742 | FALSE | TRUE | FALSE |
| NPM2 | 0.2819522 | 0.80948447 | 1 | 0.28195216 | 0.28195216 | 0.28195216 | 0.32662134 | 0.816543023 | 1 | 0.326621339 | 0.326621339 | 0.326621339 | 0.863238638 | -0.063869129 | FALSE | TRUE | FALSE |
| WDFY1 | 0.2819483 | 0.8095056 | 1 | 0.281948276 | 0.281948276 | 5.580537895 | 0.32345475 | 0.81755465 | 1 | 0.323454747 | 1.293729325 | 6.410045658 | 0.871677657 | -0.059644086 | FALSE | TRUE | FALSE |
| LARP4 | 0.2818088 | 0.80952673 | 1 | 0.281808812 | 0.281808812 | 5.566228837 | 0.32698689 | 0.816335151 | 1 | 0.326986892 | 0.326986892 | 6.468221269 | 0.861835194 | -0.064575775 | FALSE | TRUE | FALSE |
| CTNNBL1 | 0.2817382 | 0.80954786 | 0.0164 | 0.281738208 | 0.281738208 | 5.565195378 | 0.32698032 | 0.816355961 | 1 | 0.326980324 | 0.326980324 | 6.468461159 | 0.861636581 | -0.064675871 | FALSE | TRUE | FALSE |
| CCDC43 | 0.2817179 | 0.80956898 | 1 | 2.607684209 | 24.77352743 | 24.77352743 | 2.93774156 | 0.416035645 | 0.49835 | 2.937741564 | 273.072405 | 273.072405 | 0.095896077 | -1.018199159 | FALSE | FALSE | FALSE |
| OR8B8 | 0.2816173 | 0.80959009 | 1 | 0.281617251 | 0.281617251 | 0.281617251 | 0.32641598 | 0.816605291 | 0.006 | 0.326415976 | 0.326415976 | 0.326415976 | 0.862755723 | -0.064112151 | FALSE | TRUE | FALSE |
| LRRC56 | 0.2815492 | 0.8096112 | 0.50765 | 0.28154921 | 0.28154921 | 0.640354636 | 0.0491799 | 0.886873522 | 0.0123 | 0.049179902 | 0.049179902 | 0.973417043 | 5.724883497 | 0.757766653 | FALSE | TRUE | FALSE |
| SLC44A1 | 0.2814074 | 0.80963231 | 0.7543 | 2.639123224 | 10.55499123 | 22.24392366 | 0.34114086 | 0.811747146 | 1 | 0.341140865 | 0.341140865 | 6.751918339 | 0.82490097 | -0.083598186 | FALSE | TRUE | FALSE |
| IDH3A | 0.2813572 | 0.80965341 | 1 | 1.125205692 | 1.125205692 | 5.582200963 | 0.32683479 | 0.816439149 | 1 | 1.307110686 | 1.307110686 | 1.307110686 | 0.860854396 | -0.065070299 | FALSE | TRUE | FALSE |
| RBM6 | 0.281271 | 0.80967451 | 1 | 0.281270968 | 0.281270968 | 9.456653731 | 0.32679717 | 0.816459934 | 1 | 0.326797174 | 0.326797174 | 11.00202586 | 0.860689719 | -0.065153385 | FALSE | TRUE | FALSE |
| LGR5 | 0.2811618 | 0.8096956 | 0.82155 | 1.124466738 | 10.21795963 | 40.87549446 | 0.03046195 | 0.891159947 | 1 | 0.030461952 | 0.030461952 | 0.600557524 | 9.229933052 | 0.965198551 | FALSE | TRUE | FALSE |
| MPLKIP | 0.28114 | 0.80971669 | 1 | 0.281139974 | 0.281139974 | 0.281139974 | 11.6165837 | 0.180971991 | 0.24875 | 1.250352266 | 11.61658367 | 11.61658367 | 0.024201605 | -1.616155825 | FALSE | FALSE | FALSE |
| PRDX5 | 0.2811236 | 0.80973777 | 1 | 0.281123591 | 0.281123591 | 0.281123591 | 0.31613083 | 0.819905031 | 1 | 2.934259409 | 2.934259409 | 2.934259409 | 0.88926344 | -0.050969562 | FALSE | TRUE | FALSE |
| PSMB11 | 0.2810817 | 0.80975885 | 1 | 0.281081738 | 0.281081738 | 0.281081738 | 0.3223561 | 0.81800519 | 1 | 1.289378436 | 1.289378436 | 1.289378436 | 0.871960337 | -0.05950327 | FALSE | TRUE | FALSE |
| TMBIM6 | 0.28103 | 0.80977992 | 1 | 0.281029995 | 0.281029995 | 0.281029995 | 0.32589212 | 0.816812535 | 1 | 0.325892116 | 0.325892116 | 0.325892116 | 0.862340577 | -0.064321178 | FALSE | TRUE | FALSE |
| TACO1 | 0.2810044 | 0.80980099 | 1 | 0.281004371 | 0.281004371 | 0.281004371 | 0.32613784 | 0.816708974 | 1 | 0.326137836 | 0.326137836 | 0.326137836 | 0.8616123 | -0.06468811 | FALSE | TRUE | FALSE |
| HOXB4 | 0.2807955 | 0.80982205 | 0.01055 | 0.280795503 | 0.280795503 | 5.570411434 | 0.32634615 | 0.816626037 | 1 | 0.32634615 | 0.32634615 | 0.32634615 | 0.860422293 | -0.065288346 | FALSE | TRUE | FALSE |
| BIRC3 | 0.2807479 | 0.80984311 | 0.0151 | 0.280747859 | 0.280747859 | 5.55494385 | 0.32623623 | 0.816646779 | 1 | 0.326236227 | 0.326236227 | 6.463287124 | 0.860566168 | -0.065215732 | FALSE | TRUE | FALSE |
| OR4D2 | 0.2807462 | 0.80986417 | 1 | 0.280746223 | 0.280746223 | 0.280746223 | 0.32611309 | 0.816729696 | 1 | 0.326113095 | 0.326113095 | 0.326113095 | 0.860886077 | -0.065054316 | FALSE | TRUE | FALSE |
| SPATA6L | 0.2807229 | 0.80988522 | 1 | 0.280722941 | 0.280722941 | 0.280722941 | 2.91441729 | 0.418141999 | 0.50775 | 2.914417289 | 2.914417289 | 2.914417289 | 0.09632215 | -1.01627383 | FALSE | FALSE | FALSE |
| ROBO4 | 0.2806932 | 0.80990626 | 0.5118 | 0.280693211 | 2.532718096 | 5.490244602 | 0.00726247 | 0.897476777 | 1 | 0.063010634 | 0.561120274 | 1.235731569 | 38.64980859 | 1.587147347 | FALSE | TRUE | FALSE |
| PAM | 0.28068 | 0.8099273 | 0.5159 | 0.280679663 | 1.122728033 | 5.515976712 | 0.02908119 | 0.891481073 | 0.02225 | 0.029081186 | 0.029081186 | 0.572705656 | 9.651599607 | 0.984599297 | FALSE | TRUE | FALSE |
| POLR2F | 0.2806673 | 0.80994834 | 1 | 0.280667281 | 0.280667281 | 0.280667281 | 0.3256357 | 0.816936647 | 1 | 0.325635704 | 0.325635704 | 0.325635704 | 0.861905733 | -0.06454023 | FALSE | TRUE | FALSE |
| SYVN1 | 0.2806624 | 0.80996937 | 0.01845 | 0.280662396 | 0.280662396 | 5.533399902 | 0.32577414 | 0.816895295 | 1 | 0.32577414 | 0.32577414 | 6.433796216 | 0.861524479 | -0.064732378 | FALSE | TRUE | FALSE |
| DGAT1 | 0.2804884 | 0.80999039 | 0.5109 | 0.280488369 | 0.280488369 | 0.617852175 | 0.04534652 | 0.887755015 | 1 | 0.409502886 | 3.795506266 | 3.795506266 | 6.18544351 | 0.791370845 | FALSE | TRUE | FALSE |
| PDCD6IP | 0.2802307 | 0.81001141 | 1 | 0.280230731 | 0.280230731 | 5.527752426 | 0.31597255 | 0.819984989 | 1 | 0.315972553 | 0.315972553 | 6.243072072 | 0.886883144 | -0.052133599 | FALSE | TRUE | FALSE |
| OR52E2 | 0.2802219 | 0.81003243 | 0.50415 | 2.590653567 | 2.590653567 | 2.590653567 | 0.32507894 | 0.817081228 | 1 | 0.325078936 | 3.014710985 | 3.014710985 | 0.862011776 | -0.064486801 | FALSE | TRUE | FALSE |
| PPP1CC | 0.2801897 | 0.81005344 | 0.0085 | 0.280189716 | 0.280189716 | 0.280189716 | 11.0286386 | 0.187753227 | 0.00285 | 0.325133367 | 0.325133367 | 0.325133367 | 0.025405649 | -1.595069715 | FALSE | FALSE | FALSE |
| ATXN10 | 0.2801272 | 0.81007445 | 1 | 0.280127168 | 0.280127168 | 5.557024652 | 0.32490004 | 0.817184359 | 1 | 0.324900043 | 0.324900043 | 0.324900043 | 0.862194923 | -0.064394539 | FALSE | TRUE | FALSE |
| CCR10 | 0.280101 | 0.81009546 | 1 | 1.120339744 | 1.120339744 | 5.541002218 | 0.32919005 | 0.815561775 | 1 | 0.329190054 | 0.329190054 | 6.520622929 | 0.850879257 | -0.070132063 | FALSE | TRUE | FALSE |
| CDH8 | 0.2800846 | 0.81011645 | 1 | 0.280084585 | 0.280084585 | 5.510601487 | 0.32570109 | 0.816915973 | 1 | 0.325701091 | 0.325701091 | 6.420639993 | 0.85994365 | -0.065530006 | FALSE | TRUE | FALSE |
| TPSG1 | 0.2800439 | 0.81013745 | 0.8417 | 12.77364556 | 4687.358511 | 18743.39512 | 0.30543448 | 0.823720583 | 0.94175 | 11.11125022 | 992.0605898 | 3967.333835 | 0.916870508 | -0.037691997 | FALSE | TRUE | FALSE |
| RAPGEFL1 | 0.2798359 | 0.81015844 | 1 | 0.279835901 | 0.279835901 | 5.531238062 | 0.32487836 | 0.817225576 | 1 | 0.324878365 | 0.324878365 | 6.430586859 | 0.861355914 | -0.06481736 | FALSE | TRUE | FALSE |
| HLA-DOA | 0.2796781 | 0.81017942 | 0.0092 | 0.279678065 | 0.279678065 | 0.279678065 | 0.3143979 | 0.820602238 | 1 | 2.903133404 | 2.903133404 | 2.903133404 | 0.889567223 | -0.050821227 | FALSE | TRUE | FALSE |
| SNX16 | 0.2795087 | 0.8102004 | 1 | 0.279508728 | 0.279508728 | 0.279508728 | 0.3241937 | 0.817431383 | 1 | 0.3241937 | 0.3241937 | 0.3241937 | 0.862165821 | -0.064409198 | FALSE | TRUE | FALSE |
| ERCC8 | 0.2794806 | 0.81022138 | 0.00975 | 0.279480605 | 0.279480605 | 0.279480605 | 0.04661996 | 0.887413686 | 1 | 0.420815838 | 0.420815838 | 0.420815838 | 5.994870163 | 0.777779782 | FALSE | TRUE | FALSE |
| DCAF5 | 0.2793766 | 0.81024235 | 1 | 0.27937659 | 0.27937659 | 5.47149444 | 0.32506128 | 0.817101864 | 1 | 0.325061284 | 0.325061284 | 6.38213731 | 0.859458212 | -0.065775235 | FALSE | TRUE | FALSE |
| REV1 | 0.2793326 | 0.81026332 | 0.51385 | 0.279332611 | 0.279332611 | 5.467554562 | 0.09538074 | 0.877054424 | 0.50955 | 0.381455858 | 0.381455858 | 7.485650497 | 2.928605984 | 0.466660945 | FALSE | TRUE | FALSE |
| U2SURP | 0.2793309 | 0.81028428 | 1 | 0.279330921 | 0.279330921 | 9.410580107 | 0.32497964 | 0.817122495 | 1 | 0.324979636 | 0.324979636 | 6.395240665 | 0.859533614 | -0.065737135 | FALSE | TRUE | FALSE |
| ODF2L | 0.2793124 | 0.81030524 | 0.25975 | 0.279312388 | 2.495541392 | 5.533916783 | 3.64443379 | 0.37495314 | 0.50255 | 3.644433788 | 33.69672301 | 33.69672301 | 0.076640818 | -1.115539867 | FALSE | FALSE | FALSE |
| HRH1 | 0.2792437 | 0.8103262 | 1 | 0.27924374 | 0.27924374 | 5.527625198 | 0.31731274 | 0.819564418 | 0.7545 | 1.269126419 | 5.077276628 | 25.15331475 | 0.880026889 | -0.055504058 | FALSE | TRUE | FALSE |
| LYL1 | 0.279235 | 0.81034714 | 1 | 0.279235036 | 0.279235036 | 0.279235036 | 0.32478713 | 0.817246178 | 1 | 0.324787126 | 0.324787126 | 0.324787126 | 0.85974786 | -0.065628897 | FALSE | TRUE | FALSE |
| C8orf82 | 0.2791985 | 0.81036809 | 0.5017 | 0.279198539 | 0.279198539 | 0.279198539 | 0.30746899 | 0.823009354 | 0.24915 | 0.307468987 | 2.918793208 | 2.918793208 | 0.908054311 | -0.041888175 | FALSE | TRUE | FALSE |
| FCF1 | 0.2791983 | 0.81038903 | 1 | 0.279198295 | 0.279198295 | 0.279198295 | 0.32453133 | 0.817328538 | 1 | 0.324531334 | 0.324531334 | 0.324531334 | 0.860312289 | -0.065343874 | FALSE | TRUE | FALSE |
| ZFYVE16 | 0.2790543 | 0.81040996 | 0.5139 | 0.279054267 | 9.849587096 | 21.78461324 | 0.00773035 | 0.897394231 | 0.5162 | 0.065805515 | 0.065805515 | 1.288194571 | 36.09852831 | 1.557489497 | FALSE | TRUE | FALSE |
| LEMD3 | 0.2790313 | 0.81043089 | 1 | 0.279031297 | 0.279031297 | 5.490618493 | 2.9371894 | 0.416246947 | 0.50615 | 2.937189404 | 2.937189404 | 10.62038325 | 0.094999423 | -1.022279034 | FALSE | FALSE | FALSE |
| SPC25 | 0.2788212 | 0.81045182 | 1 | 24.39098204 | 237.0475408 | 237.0475408 | 0.35505694 | 0.807402876 | 1 | 0.355056941 | 0.355056941 | 0.355056941 | 0.7852859 | -0.1049722 | FALSE | TRUE | FALSE |
| DDX19B | 0.2787794 | 0.81047274 | 1 | 2.58891986 | 2.58891986 | 5.520174557 | 0.33408065 | 0.814293655 | 1 | 0.33408065 | 0.33408065 | 6.623115057 | 0.834467455 | -0.078590597 | FALSE | TRUE | FALSE |
| PPP1R10 | 0.2787745 | 0.81049365 | 1 | 0.278774533 | 0.278774533 | 5.459485271 | 0.32413661 | 0.817451938 | 1 | 0.324136612 | 0.324136612 | 6.363758396 | 0.860052591 | -0.065474991 | FALSE | TRUE | FALSE |
| GPR19 | 0.2787657 | 0.81051456 | 1 | 0.278765727 | 0.278765727 | 5.531575628 | 0.32379111 | 0.817493035 | 1 | 0.323791113 | 0.323791113 | 6.431080353 | 0.860943109 | -0.065025546 | FALSE | TRUE | FALSE |
| ATP6V1G3 | 0.2785887 | 0.81053547 | 1 | 0.278588731 | 0.278588731 | 0.278588731 | 2.75495415 | 0.430676437 | 0.4964 | 0.295768692 | 2.754954149 | 2.754954149 | 0.10112282 | -0.995150829 | FALSE | FALSE | FALSE |
| ZNF625 | 0.2785218 | 0.81055637 | 0.00885 | 0.278521831 | 0.278521831 | 0.278521831 | 0.31353431 | 0.820800424 | 1 | 2.906656645 | 2.906656645 | 2.906656645 | 0.88832968 | -0.051425828 | FALSE | TRUE | FALSE |
| PAQR4 | 0.2784879 | 0.81057727 | 1 | 0.278487913 | 0.278487913 | 0.278487913 | 0.32321627 | 0.817677752 | 1 | 0.323216271 | 0.323216271 | 0.323216271 | 0.86161477 | -0.064686865 | FALSE | TRUE | FALSE |
| MRPS11 | 0.2784204 | 0.81059816 | 1 | 0.27842037 | 0.27842037 | 0.27842037 | 0.3233612 | 0.81757518 | 1 | 0.323361196 | 0.323361196 | 0.323361196 | 0.861019733 | -0.064986895 | FALSE | TRUE | FALSE |
| EN1 | 0.278226 | 0.81061905 | 1 | 2.631044525 | 25.53534287 | 25.53534287 | 0.34540883 | 0.810469319 | 0.011 | 0.345408826 | 0.345408826 | 6.852121774 | 0.805497616 | -0.09393574 | FALSE | TRUE | FALSE |
| EREG | 0.2781515 | 0.81063994 | 0.0053 | 0.278151465 | 0.278151465 | 0.278151465 | 0.32259506 | 0.817902998 | 0.005 | 0.322595057 | 0.322595057 | 0.322595057 | 0.862231021 | -0.064376357 | FALSE | TRUE | FALSE |
| FUT6 | 0.2781465 | 0.81066082 | 0.01365 | 0.278146482 | 0.278146482 | 5.504077165 | 0.31320965 | 0.820919123 | 0.50615 | 2.906722048 | 2.906722048 | 6.205809462 | 0.888052091 | -0.051561559 | FALSE | TRUE | FALSE |
| CD53 | 0.2780675 | 0.81068169 | 1 | 0.278067516 | 0.278067516 | 0.278067516 | 0.31329369 | 0.820899351 | 1 | 2.926391506 | 2.926391506 | 2.926391506 | 0.887561806 | -0.051801395 | FALSE | TRUE | FALSE |
| PI4K2B | 0.2779992 | 0.81070256 | 0.87345 | 2.532930384 | 23.68654105 | 23.68654105 | 0.10202267 | 0.875477985 | 0.0094 | 0.102022665 | 0.102022665 | 0.102022665 | 2.724877156 | 0.435346928 | FALSE | TRUE | FALSE |
| JAM2 | 0.2779727 | 0.81072343 | 0.0086 | 0.277972692 | 0.277972692 | 0.277972692 | 0.32281421 | 0.817780208 | 1 | 0.322814212 | 0.322814212 | 0.322814212 | 0.861091864 | -0.064950514 | FALSE | TRUE | FALSE |
| ZNF534 | 0.2778121 | 0.81074429 | 0.98145 | 197.9977478 | 17798.32212 | 174608.8676 | 3299.1783 | 0.000622255 | 0.70945 | 32015.38795 | 3224440.833 | 354066373.3 | 8.42064E-05 | -4.074654675 | FALSE | FALSE | FALSE |
| UROS | 0.2777794 | 0.81076515 | 0.7502 | 0.277779403 | 1.110790214 | 1.110790214 | 0.38027775 | 0.799174123 | 0.75655 | 0.380277747 | 1.520760432 | 1.520760432 | 0.730464521 | -0.136400873 | FALSE | TRUE | FALSE |
| OR3A3 | 0.2777787 | 0.810786 | 1 | 2.557979763 | 2.557979763 | 2.557979763 | 2.97896737 | 0.413273814 | 0.5017 | 2.978967366 | 2.978967366 | 2.978967366 | 0.093246657 | -1.03036673 | FALSE | FALSE | FALSE |
| B3GALT6 | 0.2775666 | 0.81080685 | 1 | 2.564886119 | 2.564886119 | 2.564886119 | 0.33558684 | 0.813738682 | 1 | 0.335586841 | 0.335586841 | 0.335586841 | 0.827108098 | -0.082437727 | FALSE | TRUE | FALSE |
| TCF19 | 0.2775367 | 0.81082769 | 1 | 2.576850487 | 2.576850487 | 5.502521074 | 0.32254402 | 0.817923446 | 1 | 3.002592822 | 3.002592822 | 6.401529729 | 0.860461399 | -0.065268608 | FALSE | TRUE | FALSE |
| GPR31 | 0.2775282 | 0.81084853 | 1 | 2.579219777 | 2.579219777 | 2.579219777 | 0.33274859 | 0.814633493 | 1 | 0.332748589 | 0.332748589 | 0.332748589 | 0.834047601 | -0.078809162 | FALSE | TRUE | FALSE |
| EIF3D | 0.2774514 | 0.81086937 | 1 | 0.277451387 | 0.277451387 | 5.476020815 | 0.32287998 | 0.817739239 | 1 | 0.322879981 | 0.322879981 | 6.382728106 | 0.859301918 | -0.065854219 | FALSE | TRUE | FALSE |

| | | | | | | | | | | | | | | | | | |
|---|---|---|---|---|---|---|---|---|---|---|---|---|---|---|---|---|---|
| DDX41 | 0.2774211 | 0.8108902 | 0.7581 | 0.277421085 | 2.539423686 | 5.45227595 | 0.04934274 | 0.886824184 | 0.02055 | 0.049342743 | 0.049342743 | 0.971777324 | 5.622327993 | 0.749916178 | FALSE | TRUE | FALSE |
| KCNA7 | 0.277403 | 0.81091102 | 0.5151 | 0.277402995 | 1.109009533 | 4.434737257 | 0.03373718 | 0.890419484 | 0.50565 | 0.134829621 | 0.538977132 | 0.665810209 | 8.222470465 | 0.915002322 | FALSE | TRUE | FALSE |
| DOCK2 | 0.2773879 | 0.81093184 | 0.5266 | 0.277387903 | 0.277387903 | 5.345272768 | 0.09586177 | 0.876939629 | 1 | 0.095861774 | 0.095861774 | 3.220399029 | 2.89362371 | 0.461442054 | FALSE | TRUE | FALSE |
| HHEX | 0.2773596 | 0.81095266 | 1 | 1.109083477 | 1.109083477 | 1.109083477 | 0.32651105 | 0.816563784 | 1 | 0.326511055 | 0.326511055 | 0.326511055 | 0.849464561 | -0.070854735 | FALSE | TRUE | FALSE |
| RANGRF | 0.2773304 | 0.81097347 | 1 | 0.277330425 | 0.277330425 | 0.277330425 | 0.32226889 | 0.818066447 | 1 | 0.322268888 | 0.322268888 | 0.322268888 | 0.860556 | -0.065220863 | FALSE | TRUE | FALSE |
| AOC3 | 0.2771945 | 0.81099427 | 0.936 | 2.493752945 | 217.6718993 | 217.6718993 | 35.0704905 | 0.079894156 | 0.32615 | 3.766552749 | 334.7257522 | 334.7257522 | 0.007903925 | -2.102157181 | FALSE | FALSE | FALSE |
| WNT3 | 0.2770577 | 0.81101508 | 1 | 0.27705774 | 0.27705774 | 5.478895285 | 0.32262008 | 0.817862087 | 1 | 0.322620076 | 0.322620076 | 6.388527419 | 0.858774021 | -0.066121102 | FALSE | TRUE | FALSE |
| OTOR | 0.2770546 | 0.81103587 | 1 | 0.277054577 | 0.277054577 | 0.277054577 | 0.312208 | 0.821293941 | 1 | 2.908982039 | 2.908982039 | 2.908982039 | 0.887403827 | -0.051878703 | FALSE | TRUE | FALSE |
| TGFB1 | 0.2769807 | 0.81105667 | 1 | 1.107695169 | 1.107695169 | 5.49245117 | 2.93537551 | 0.416458084 | 0.25495 | 1.249493278 | 11.73937478 | 11.73937478 | 0.094359539 | -1.025214188 | FALSE | FALSE | FALSE |
| SNRNP27 | 0.2769171 | 0.81107745 | 1 | 0.276917091 | 0.276917091 | 0.276917091 | 0.3214762 | 0.818311065 | 1 | 0.321476197 | 0.321476197 | 0.321476197 | 0.861392209 | -0.064799061 | FALSE | TRUE | FALSE |
| PAK1 | 0.2768783 | 0.81109824 | 1 | 0.27687833 | 0.27687833 | 5.478525955 | 6.38200778 | 0.25835853 | 0.0137 | 0.32212609 | 0.32212609 | 6.382007779 | 0.043384204 | -1.362668362 | FALSE | FALSE | FALSE |
| PSMD2 | 0.2768756 | 0.81111902 | 1 | 0.276875591 | 0.276875591 | 5.441121836 | 0.32226234 | 0.818086857 | 1 | 0.322262338 | 0.322262338 | 6.346328317 | 0.85916211 | -0.065924884 | FALSE | TRUE | FALSE |
| PWWP2B | 0.2768336 | 0.81113979 | 1 | 0.276833613 | 0.276833613 | 5.438078988 | 0.32227904 | 0.818046033 | 0.01855 | 0.322279039 | 0.322279039 | 6.344375785 | 0.858987334 | -0.06601324 | FALSE | TRUE | FALSE |
| SUCO | 0.2767775 | 0.81116056 | 0.97145 | 23.75827386 | 228.5166231 | 2251.709157 | 0.33167393 | 0.81482413 | 0.9391 | 2.987965876 | 47.84219193 | 58.6828743 | 0.834486805 | -0.078580526 | FALSE | TRUE | FALSE |
| MRPL11 | 0.2766699 | 0.81118132 | 1 | 0.276669866 | 0.276669866 | 0.276669866 | 2.90193286 | 0.419607223 | 0.5065 | 2.901932857 | 2.901932857 | 2.901932857 | 0.095339858 | -1.0207255 | FALSE | FALSE | FALSE |
| GPN3 | 0.2766462 | 0.81120208 | 1 | 0.276646154 | 0.276646154 | 0.276646154 | 2.90325029 | 0.419189418 | 0.00655 | 2.903250294 | 2.903250294 | 2.903250294 | 0.095288427 | -1.020959842 | FALSE | FALSE | FALSE |
| PDLIM2 | 0.2764844 | 0.81122284 | 0.01115 | 0.276484374 | 0.276484374 | 5.481251686 | 26.7884541 | 0.102859669 | 0.00665 | 2.808201105 | 26.78845415 | 26.78845415 | 0.010321028 | -1.986277061 | FALSE | FALSE | FALSE |
| NSA2 | 0.2764727 | 0.81124359 | 1 | 0.276472746 | 2.555220033 | 2.555220033 | 0.33134118 | 0.81495101 | 1 | 0.331341184 | 0.331341184 | 0.331341184 | 0.834405014 | -0.078623095 | FALSE | TRUE | FALSE |
| KLHL31 | 0.2763757 | 0.81126434 | 0.5052 | 0.276375656 | 0.276375656 | 5.437008627 | 0.0946401 | 0.877207172 | 1 | 0.378430781 | 0.378430781 | 3.204085155 | 2.920280824 | 0.465424617 | FALSE | TRUE | FALSE |
| NCOA2 | 0.2763671 | 0.81128508 | 1 | 2.596533717 | 25.03730184 | 87.3291159 | 0.34267243 | 0.81133046 | 1 | 0.342672425 | 0.342672425 | 11.54005553 | 0.806505214 | -0.093392821 | FALSE | TRUE | FALSE |
| TGFB1I1 | 0.276355 | 0.81130582 | 0.5083 | 0.27635505 | 0.27635505 | 1.364950046 | 0.09285467 | 0.877568572 | 1 | 0.863012228 | 0.863012228 | 1.83636433 | 2.976210527 | 0.473663648 | FALSE | TRUE | FALSE |
| SOSTDC1 | 0.276332 | 0.81132655 | 1 | 1.105012831 | 1.105012831 | 1.105012831 | 0.32538326 | 0.817039941 | 1 | 0.325383258 | 0.325383258 | 0.325383258 | 0.849250929 | -0.07096397 | FALSE | TRUE | FALSE |
| MYLK | 0.2763162 | 0.81134728 | 0.84265 | 4.420112706 | 636.5453147 | 2548.135162 | 81.7615701 | 0.035762275 | 0.22645 | 20.4381871 | 1309.448247 | 12178.5333 | 0.003379536 | -2.471142924 | FALSE | FALSE | FALSE |
| CCDC70 | 0.2762998 | 0.811368 | 1 | 2.541313817 | 2.541313817 | 2.541313817 | 0.32080673 | 0.818534712 | 1 | 2.961574145 | 2.961574145 | 2.961574145 | 0.861265499 | -0.064862949 | FALSE | TRUE | FALSE |
| RNF151 | 0.2762765 | 0.81138872 | 1 | 0.276276549 | 0.276276549 | 0.276276549 | 0.32107851 | 0.818412793 | 1 | 0.321078506 | 0.321078506 | 0.321078506 | 0.860464165 | -0.065267212 | FALSE | TRUE | FALSE |
| LONRF2 | 0.2762481 | 0.81140943 | 0.01765 | 0.276248124 | 0.276248124 | 5.451119107 | 0.32163424 | 0.8182296 | 1 | 0.321634241 | 0.321634241 | 6.356917181 | 0.858889039 | -0.06606294 | FALSE | TRUE | FALSE |
| TBC1D10A | 0.27622 | 0.81143014 | 1 | 0.276219984 | 0.276219984 | 5.445604433 | 0.32118242 | 0.818372115 | 1 | 0.321182424 | 0.321182424 | 6.342896543 | 0.860009651 | -0.065496675 | FALSE | TRUE | FALSE |
| CMPK2 | 0.2760483 | 0.81145084 | 0.5029 | 0.27604825 | 0.27604825 | 0.27604825 | 0.8501448 | 0.607417836 | 0.5087 | 0.850144798 | 0.850144798 | 0.850144798 | 0.324707333 | -0.488507903 | FALSE | TRUE | FALSE |
| PTBP2 | 0.276042 | 0.81147154 | 1 | 0.276041993 | 0.276041993 | 5.467802041 | 0.31148992 | 0.821529848 | 1 | 2.931339161 | 2.931339161 | 6.177079457 | 0.886198787 | -0.052468849 | FALSE | TRUE | FALSE |
| DLX1 | 0.2759773 | 0.81149224 | 0.0095 | 0.275977339 | 0.275977339 | 0.275977339 | 0.32150284 | 0.818290706 | 1 | 0.321502836 | 0.321502836 | 0.321502836 | 0.858397838 | -0.066311385 | FALSE | TRUE | FALSE |
| SOS2 | 0.2759054 | 0.81151293 | 0.5188 | 1.102909803 | 10.5250717 | 51.39166995 | 0.19132548 | 0.854011995 | 0.51605 | 0.191325481 | 0.191325481 | 3.745227589 | 1.442073284 | 0.158987331 | FALSE | TRUE | FALSE |
| NAAA | 0.275851 | 0.81153361 | 0.87355 | 2.508064696 | 23.40466363 | 23.40466363 | 0.10127104 | 0.875634393 | 1 | 0.101271045 | 0.101271045 | 0.101271045 | 2.723888646 | 0.435189349 | FALSE | TRUE | FALSE |
| CALML3 | 0.2758482 | 0.8115543 | 1 | 0.275848152 | 0.275848152 | 0.275848152 | 26.714137 | 0.103157826 | 0.13225 | 11.14171267 | 106.8385028 | 106.8385028 | 0.010325924 | -1.98607107 | FALSE | FALSE | FALSE |
| OR1E2 | 0.2757452 | 0.81157497 | 1 | 0.275745171 | 0.275745171 | 0.275745171 | 0.32050276 | 0.818595609 | 1 | 0.320502757 | 0.320502757 | 0.320502757 | 0.860351948 | -0.065323853 | FALSE | TRUE | FALSE |
| HEBP1 | 0.2757183 | 0.81159564 | 1 | 0.275718281 | 0.275718281 | 0.275718281 | 0.32025698 | 0.818697014 | 1 | 0.320256978 | 0.320256978 | 0.320256978 | 0.860928255 | -0.065033039 | FALSE | TRUE | FALSE |
| PRPF40A | 0.2756986 | 0.81161631 | 1 | 0.275698561 | 0.275698561 | 5.428826526 | 0.31726732 | 0.819584491 | 1 | 1.269045322 | 1.269045322 | 10.71013842 | 0.868978755 | -0.060990841 | FALSE | TRUE | FALSE |
| AP1S3 | 0.2756801 | 0.81163697 | 0.00385 | 0.275680121 | 0.275680121 | 0.275680121 | 0.32087883 | 0.818514404 | 1 | 0.32087883 | 0.32087883 | 0.32087883 | 0.859140883 | -0.065935614 | FALSE | TRUE | FALSE |
| OR10H2 | 0.2756619 | 0.81165763 | 1 | 0.27566193 | 0.27566193 | 0.27566193 | 0.32060767 | 0.818575314 | 1 | 0.320607668 | 0.320607668 | 0.320607668 | 0.859810783 | -0.065597112 | FALSE | TRUE | FALSE |
| HHIP | 0.2756176 | 0.81167828 | 1 | 0.275617598 | 0.275617598 | 5.438820023 | 0.31113236 | 0.821627958 | 1 | 2.930503158 | 2.930503158 | 6.149495789 | 0.885853217 | -0.052638233 | FALSE | TRUE | FALSE |
| RPS19BP1 | 0.2755994 | 0.81169893 | 1 | 0.275599444 | 0.275599444 | 0.275599444 | 0.31035822 | 0.821921622 | 1 | 0.31035822 | 2.862973679 | 2.862973679 | 0.888004334 | -0.051584915 | FALSE | TRUE | FALSE |
| SLC38A9 | 0.2755648 | 0.81171958 | 1 | 2.546203172 | 2.546203172 | 5.457687045 | 0.33038588 | 0.815246366 | 1 | 0.330385885 | 0.330385885 | 6.551100068 | 0.834069482 | -0.078797769 | FALSE | TRUE | FALSE |
| HNRNPA1 | 0.2755517 | 0.81174022 | 1 | 0.275551704 | 0.275551704 | 0.275551704 | 0.32101218 | 0.818453451 | 1 | 0.321012178 | 0.321012178 | 0.321012178 | 0.858383958 | -0.066318407 | FALSE | TRUE | FALSE |
| ORMDL2 | 0.2755332 | 0.81176085 | 1 | 0.275533158 | 0.275533158 | 0.275533158 | 1.22787368 | 0.564258858 | 0.5058 | 2.839532245 | 11.35749453 | 11.35749453 | 0.224398619 | -0.648979821 | FALSE | FALSE | FALSE |
| JMY | 0.2755252 | 0.81178148 | 0.99935 | 21216.63606 | 2254822.64 | 24032637.09 | 0.05559909 | 0.885485644 | 1 | 42.66236734 | 410.7373185 | 4051.087099 | 4.955570359 | 0.695093647 | FALSE | TRUE | FALSE |
| TPBG | 0.2755176 | 0.8118021 | 1 | 0.2755176 | 0.2755176 | 5.449214572 | 0.32047692 | 0.818615899 | 1 | 0.320476921 | 0.320476921 | 6.346884948 | 0.859711204 | -0.065647414 | FALSE | TRUE | FALSE |
| DNASE1 | 0.2755008 | 0.81182272 | 0.87685 | 2.444817558 | 110.5168111 | 830.4020425 | 84.5932377 | 0.034602826 | 0.63675 | 84.59323774 | 8026.780893 | 80962.4594 | 0.003256771 | -2.487212752 | FALSE | FALSE | FALSE |
| SLC2A1 | 0.2754453 | 0.81184334 | 1 | 0.275445272 | 0.275445272 | 5.436421092 | 0.32096984 | 0.818473773 | 1 | 0.320969842 | 0.320969842 | 6.344962818 | 0.858165582 | -0.066428907 | FALSE | TRUE | FALSE |
| SFXN1 | 0.2753538 | 0.81186395 | 1 | 0.275353763 | 0.275353763 | 0.275353763 | 0.32027132 | 0.818656466 | 1 | 0.320271325 | 0.320271325 | 0.320271325 | 0.859751534 | -0.065627041 | FALSE | TRUE | FALSE |
| ZFP1 | 0.2751098 | 0.81188456 | 1 | 0.275109751 | 0.275109751 | 0.275109751 | 0.3199916 | 0.818737545 | 1 | 0.319991602 | 0.319991602 | 0.319991602 | 0.859740535 | -0.065632597 | FALSE | TRUE | FALSE |
| AFTPH | 0.2750519 | 0.81190516 | 1 | 2.561192258 | 24.4770703 | 50.45271436 | 0.33030053 | 0.815288483 | 1 | 3.085499899 | 3.085499899 | 11.16582823 | 0.832732331 | -0.079494573 | FALSE | TRUE | FALSE |
| SUFU | 0.2748835 | 0.81192576 | 0.50545 | 1.099231176 | 1.099231176 | 5.433925818 | 0.32043004 | 0.818636185 | 1 | 1.281408642 | 1.281408642 | 6.343130213 | 0.857858183 | -0.066584502 | FALSE | TRUE | FALSE |
| RBP1 | 0.2746612 | 0.81194635 | 1 | 0.274661227 | 0.274661227 | 0.274661227 | 25.2269219 | 0.109750792 | 0.12455 | 25.22692187 | 244.1049162 | 244.1049162 | 0.010887624 | -1.963066906 | FALSE | FALSE | FALSE |
| FDPS | 0.2745972 | 0.81196694 | 1 | 0.274597191 | 0.274597191 | 5.440935577 | 0.31963551 | 0.818919703 | 1 | 0.319635514 | 0.319635514 | 6.34041103 | 0.859094746 | -0.065958937 | FALSE | TRUE | FALSE |
| EMX1 | 0.2744842 | 0.81198753 | 1 | 0.274484249 | 0.274484249 | 0.274484249 | 4.99312278 | 0.306514464 | 0.24855 | 1.248307129 | 4.993122781 | 4.993122781 | 0.054972461 | -1.259854818 | FALSE | FALSE | FALSE |
| GAB2 | 0.2744776 | 0.81200811 | 1 | 0.274477649 | 0.274477649 | 5.405745285 | 0.3099345 | 0.822019294 | 1 | 2.908075569 | 2.908075569 | 57.37881879 | 0.885598897 | -0.052762933 | FALSE | TRUE | FALSE |
| FAM111B | 0.2744415 | 0.81202869 | 0.6625 | 2.380921048 | 192.9323909 | 1799.970144 | 5.04274479 | 0.30507177 | 0.1315 | 0.570913961 | 5.042744786 | 5.042744786 | 0.054423048 | -1.264217139 | FALSE | FALSE | FALSE |
| PEX11G | 0.2744386 | 0.81204926 | 1 | 2307.771375 | 245197.8445 | 245197.8445 | 0.33931886 | 0.812249077 | 1 | 291.4970946 | 2913.913948 | 29839.6871 | 0.808792682 | -0.092162787 | FALSE | TRUE | FALSE |
| CCNT1 | 0.274432 | 0.81206983 | 1 | 2.553253008 | 2.553253008 | 9.278747656 | 0.32961086 | 0.815456758 | 1 | 0.329610858 | 0.329610858 | 6.510564385 | 0.832593981 | -0.079566733 | FALSE | TRUE | FALSE |
| CNBD1 | 0.2743702 | 0.81209039 | 0.50855 | 0.274370238 | 0.274370238 | 0.612781023 | 3.45672637 | 0.385748041 | 0.31925 | 3.456726373 | 313.7468876 | 313.7468876 | 0.079372854 | -1.100328004 | FALSE | FALSE | FALSE |
| ARV1 | 0.2742549 | 0.81211095 | 1 | 2.532585153 | 2.532585153 | 2.532585153 | 2.95453022 | 0.414338972 | 0.00655 | 2.954530219 | 2.954530219 | 2.954530219 | 0.092825218 | -1.032334024 | FALSE | FALSE | FALSE |
| HBE1 | 0.2742395 | 0.8121315 | 1 | 0.274239484 | 0.274239484 | 0.274239484 | 0.31959963 | 0.81896013 | 1 | 0.319599631 | 0.319599631 | 0.319599631 | 0.858071967 | -0.066476286 | FALSE | TRUE | FALSE |
| BOLL | 0.2741166 | 0.81215205 | 1 | 0.274116576 | 0.274116576 | 0.274116576 | 0.3187922 | 0.819101491 | 1 | 0.318792202 | 0.318792202 | 0.318792202 | 0.859859726 | -0.065572392 | FALSE | TRUE | FALSE |
| SLC35C2 | 0.274059 | 0.8121726 | 0.6322 | 0.48755419 | 4.382115278 | 4.382115278 | 0.19066437 | 0.854157275 | 0.0094 | 0.190664366 | 0.190664366 | 0.190664366 | 1.437389906 | 0.157574591 | FALSE | TRUE | FALSE |
| MYH8 | 0.2740506 | 0.81219314 | 0.69555 | 7.049988636 | 168415.389 | 9027625.53 | 3.0012296 | 0.411777618 | 0.35255 | 0.360177089 | 3614.641423 | 203606.0147 | 0.091312777 | -1.039468448 | FALSE | FALSE | FALSE |
| GALP | 0.2740436 | 0.81221368 | 1 | 0.274043551 | 0.274043551 | 0.274043551 | 0.31830564 | 0.819282917 | 1 | 0.318305643 | 0.318305643 | 0.318305643 | 0.860944682 | -0.065024752 | FALSE | TRUE | FALSE |
| IL1RAP | 0.2739691 | 0.81223421 | 1 | 0.273969105 | 0.273969105 | 5.418031728 | 0.31906767 | 0.81902074 | 1 | 0.319067668 | 0.319067668 | 6.318403555 | 0.858655178 | -0.066181207 | FALSE | TRUE | FALSE |
| LRRN1 | 0.273823 | 0.81225473 | 1 | 0.273822952 | 0.273822952 | 5.399985282 | 0.30927243 | 0.822194828 | 1 | 2.903760408 | 2.903760408 | 10.46889519 | 0.885377829 | -0.052871358 | FALSE | TRUE | FALSE |
| TMEM202 | 0.2737725 | 0.81227526 | 0.0076 | 0.273772461 | 0.273772461 | 0.273772461 | 0.31836755 | 0.819222484 | 1 | 0.318367546 | 0.318367546 | 0.318367546 | 0.859925781 | -0.06553903 | FALSE | TRUE | FALSE |
| ARHGAP44 | 0.2737567 | 0.81229578 | 1 | 0.273756728 | 0.273756728 | 5.393603865 | 0.31872505 | 0.819121668 | 1 | 0.318725048 | 0.318725048 | 6.290832532 | 0.858911875 | -0.066051393 | FALSE | TRUE | FALSE |
| MED21 | 0.2736945 | 0.81231629 | 0.004 | 0.273694526 | 0.273694526 | 0.273694526 | 0.3064239 | 0.823356091 | 1 | 0.306423903 | 0.306423903 | 0.306423903 | 0.893189218 | -0.049056528 | FALSE | TRUE | FALSE |
| CCT8 | 0.2736724 | 0.8123368 | 0.74605 | 0.273672397 | 1.094405063 | 5.419628872 | 0.09604467 | 0.87690132 | 1 | 0.096044671 | 0.096044671 | 1.90425534 | 2.849428235 | 0.454757723 | FALSE | TRUE | FALSE |
| DUSP2 | 0.2736549 | 0.8123573 | 1 | 0.27365491 | 0.27365491 | 0.27365491 | 0.31536857 | 0.820144697 | 0.5054 | 1.261202646 | 1.261202646 | 1.261202646 | 0.867730438 | -0.061615168 | FALSE | TRUE | FALSE |
| CDC73 | 0.2733124 | 0.8123778 | 1 | 0.27331235 | 0.27331235 | 5.407234664 | 0.31865103 | 0.81914184 | 1 | 0.318651031 | 0.318651031 | 6.312406477 | 0.857716825 | -0.066656071 | FALSE | TRUE | FALSE |

| | | | | | | | | | | | | | | | | | |
|---|---|---|---|---|---|---|---|---|---|---|---|---|---|---|---|---|---|
| SYDE2 | 0.2733032 | 0.8123983 | 0.5071 | 0.273303238 | 2.373438032 | 5.360811038 | 34.5375558 | 0.080457094 | 0.74905 | 318.3220599 | 29011.40293 | 286573.5782 | 0.007913219 | -2.101646823 | FALSE | FALSE | FALSE |
| CD300C | 0.2732884 | 0.81241879 | 1 | 2.513512196 | 2.513512196 | 2.513512196 | 0.32759744 | 0.816126787 | 1 | 0.327597443 | 0.327597443 | 0.327597443 | 0.834220324 | -0.078719234 | FALSE | TRUE | FALSE |
| ZNF287 | 0.2732619 | 0.81243928 | 1 | 0.273261922 | 0.273261922 | 5.394695988 | 0.3183318 | 0.819242633 | 1 | 0.318331803 | 0.318331803 | 6.294219688 | 0.85841854 | -0.066300911 | FALSE | TRUE | FALSE |
| STRA8 | 0.2732448 | 0.81245976 | 1 | 0.273244806 | 0.273244806 | 0.273244806 | 0.30856108 | 0.822428342 | 1 | 0.30856108 | 2.884030056 | 2.884030056 | 0.885545276 | -0.052789229 | FALSE | TRUE | FALSE |
| PXMP2 | 0.2729712 | 0.81248024 | 0.9358 | 2.487826172 | 223.2731513 | 223.2731513 | 0.09926783 | 0.876159593 | 1 | 0.910606672 | 3.641210705 | 3.641210705 | 2.749845929 | 0.439308361 | FALSE | TRUE | FALSE |
| P2RY12 | 0.2727511 | 0.81250072 | 1 | 0.272751094 | 0.272751094 | 0.272751094 | 0.31748349 | 0.819463986 | 1 | 0.317483486 | 0.317483486 | 0.317483486 | 0.859103249 | -0.065954638 | FALSE | TRUE | FALSE |
| ESCO2 | 0.2727097 | 0.81252119 | 0.75585 | 0.272709713 | 2.494730839 | 5.400777932 | 0.41115804 | 0.78806506 | 0.75135 | 0.41115804 | 3.788331318 | 3.788331318 | 0.663272236 | -0.178308181 | FALSE | TRUE | FALSE |
| HSCB | 0.272586 | 0.81254165 | 1 | 0.272585987 | 0.272585987 | 0.272585987 | 0.31719739 | 0.819644679 | 1 | 0.317197388 | 0.317197388 | 0.317197388 | 0.859357603 | -0.065826076 | FALSE | TRUE | FALSE |
| HES3 | 0.2725425 | 0.81256212 | 1 | 0.272542457 | 0.272542457 | 0.272542457 | 0.31752415 | 0.81942378 | 1 | 0.317524147 | 0.317524147 | 0.317524147 | 0.85833616 | -0.066342591 | FALSE | TRUE | FALSE |
| OR10G4 | 0.2724816 | 0.81258257 | 1 | 0.272481602 | 0.272481602 | 0.272481602 | 0.29796639 | 0.826006611 | 0.0071 | 0.297966387 | 0.297966387 | 0.297966387 | 0.914470941 | -0.03883009 | FALSE | TRUE | FALSE |
| ZNF641 | 0.2724694 | 0.81260303 | 1 | 0.272469391 | 0.272469391 | 5.406883078 | 0.31370681 | 0.820760822 | 0.5079 | 1.254773878 | 1.254773878 | 6.231037796 | 0.868547887 | -0.061206232 | FALSE | TRUE | FALSE |
| CDC25B | 0.2724599 | 0.81262348 | 0.9845 | 235.4589064 | 24199.22138 | 254220.9905 | 0.30767449 | 0.822932115 | 1 | 2681.844801 | 290085.3326 | 543907.7018 | 0.885545804 | -0.05278897 | FALSE | TRUE | FALSE |
| WARS2 | 0.2724445 | 0.81264392 | 1 | 0.27244451 | 0.27244451 | 0.27244451 | 0.31718007 | 0.819664733 | 1 | 0.317180072 | 0.317180072 | 0.317180072 | 0.858958472 | -0.066027833 | FALSE | TRUE | FALSE |
| TFAM | 0.272363 | 0.81266436 | 1 | 0.272363045 | 2.552866156 | 2.552866156 | 0.32766759 | 0.816064183 | 1 | 0.327667589 | 0.327667589 | 0.327667589 | 0.831217532 | -0.080285305 | FALSE | TRUE | FALSE |
| KRTAP5-7 | 0.272286 | 0.81268479 | 1 | 2.49777132 | 23.51668429 | 23.51668429 | 0.33659428 | 0.813309437 | 1 | 0.33659428 | 0.33659428 | 0.33659428 | 0.80894416 | -0.092081456 | FALSE | TRUE | FALSE |
| CSGALNACT | 0.2722624 | 0.81270522 | 0.0137 | 0.272262356 | 0.272262356 | 5.390122416 | 0.31736217 | 0.81952426 | 1 | 0.317362166 | 0.317362166 | 6.290638463 | 0.857891662 | -0.066567553 | FALSE | TRUE | FALSE |
| MYEOV | 0.2721724 | 0.81272565 | 0.5062 | 2.506092089 | 2.506092089 | 2.506092089 | 0.31665005 | 0.819804982 | 1 | 2.926040493 | 2.926040493 | 2.926040493 | 0.85953685 | -0.0657355 | FALSE | TRUE | FALSE |
| NR2F2 | 0.2721237 | 0.81274607 | 1 | 0.272123712 | 0.272123712 | 5.380296881 | 0.31760452 | 0.81940367 | 1 | 0.317604517 | 0.317604517 | 6.28815213 | 0.856800511 | -0.067120283 | FALSE | TRUE | FALSE |
| SH2D5 | 0.2719658 | 0.81276649 | 1 | 0.271965775 | 1.087815907 | 5.378444882 | 1.26699792 | 0.559408827 | 0.51255 | 1.266997923 | 1.266997923 | 6.272770203 | 0.214653687 | -0.668261648 | FALSE | FALSE | FALSE |
| ANKFY1 | 0.2719216 | 0.8127869 | 0.5169 | 0.271921601 | 0.271921601 | 5.300057654 | 1.47583772 | 0.538149264 | 0.2698 | 0.368968543 | 1.475837721 | 12.44612734 | 0.184248984 | -0.734594898 | FALSE | FALSE | FALSE |
| BUB3 | 0.2717992 | 0.81280731 | 0.0105 | 0.271799215 | 0.271799215 | 5.395068458 | 0.31352855 | 0.820820218 | 1 | 1.253864658 | 1.253864658 | 1.253864658 | 0.8669042 | -0.062028893 | FALSE | TRUE | FALSE |
| TAL2 | 0.2717176 | 0.81282772 | 1 | 4.34604018 | 17.38779456 | 17.38779456 | 46.7136197 | 0.058884565 | 0.00175 | 4.967690859 | 46.71361973 | 46.71361973 | 0.005816669 | -2.235325682 | FALSE | FALSE | FALSE |
| DCLK3 | 0.2715474 | 0.81284812 | 0.01725 | 0.271547427 | 0.271547427 | 5.357028398 | 0.316569 | 0.819825 | 1 | 0.316568998 | 0.316568998 | 6.255433486 | 0.857782755 | -0.066622689 | FALSE | TRUE | FALSE |
| DEFB4A | 0.2715345 | 0.81286851 | 1 | 0.271534494 | 0.271534494 | 0.271534494 | 0.27292636 | 0.832954471 | 1 | 0.272926361 | 0.272926361 | 0.272926361 | 0.994900211 | -0.002220477 | FALSE | TRUE | FALSE |
| RABGGTB | 0.2715045 | 0.81288891 | 0.0084 | 0.271504539 | 0.271504539 | 0.271504539 | 0.31613976 | 0.81988503 | 1 | 0.316139764 | 0.316139764 | 0.316139764 | 0.858811734 | -0.06610203 | FALSE | TRUE | FALSE |
| FGF3 | 0.2713736 | 0.81290929 | 1 | 0.271373565 | 0.271373565 | 0.271373565 | 0.3130586 | 0.820978412 | 1 | 1.25196497 | 1.25196497 | 1.25196497 | 0.866845892 | -0.062058105 | FALSE | TRUE | FALSE |
| ZHX2 | 0.2713442 | 0.81292968 | 0.51445 | 0.2713442 | 0.2713442 | 5.319227304 | 0.09330731 | 0.877473655 | 1 | 0.37305824 | 0.37305824 | 7.330624438 | 2.908070145 | 0.463604878 | FALSE | TRUE | FALSE |
| NPM3 | 0.2713371 | 0.81295005 | 1 | 0.271337101 | 0.271337101 | 0.271337101 | 0.31566171 | 0.820024943 | 0.0052 | 0.315661709 | 0.315661709 | 0.315661709 | 0.859581929 | -0.065712723 | FALSE | TRUE | FALSE |
| TMEM198 | 0.2713204 | 0.81297043 | 1 | 0.271320443 | 0.271320443 | 5.368978844 | 2.84047179 | 0.423959879 | 0.5035 | 2.840471786 | 2.840471786 | 6.070441554 | 0.095519499 | -1.019907962 | FALSE | FALSE | FALSE |
| CST5 | 0.2711149 | 0.8129908 | 0.50245 | 2.496749966 | 2.496749966 | 2.496749966 | 0.32540536 | 0.817019292 | 1 | 0.325405359 | 0.325405359 | 0.325405359 | 0.833160491 | -0.079271333 | FALSE | TRUE | FALSE |
| SLC17A6 | 0.2710733 | 0.81301116 | 1 | 0.271073309 | 0.271073309 | 5.35719911 | 0.31606502 | 0.819945019 | 1 | 0.316065024 | 0.316065024 | 6.255275468 | 0.857650447 | -0.066689682 | FALSE | TRUE | FALSE |
| MRPL40 | 0.271028 | 0.81303152 | 1 | 0.27102799 | 0.27102799 | 0.27102799 | 2.82922336 | 0.424576801 | 0.4962 | 0.305948409 | 2.82922336 | 2.82922336 | 0.095795897 | -1.018653091 | FALSE | FALSE | FALSE |
| MKS1 | 0.2708754 | 0.81305188 | 0.74845 | 1.08310373 | 17.32992363 | 17.32992363 | 1.53905589 | 0.532052513 | 0.8474 | 14.23375423 | 540.0416907 | 2160.546716 | 0.176001021 | -0.754484812 | FALSE | FALSE | FALSE |
| GTF2I | 0.2708518 | 0.81307223 | 1 | 0.270851801 | 0.270851801 | 5.310591254 | 2.85858799 | 0.423342062 | 0.5099 | 2.858587993 | 2.858587993 | 56.18096893 | 0.094750206 | -1.023419837 | FALSE | FALSE | FALSE |
| RNASET2 | 0.2708507 | 0.81309258 | 1 | 0.270850699 | 0.270850699 | 0.270850699 | 0.31567385 | 0.820004968 | 1 | 0.315673845 | 0.315673845 | 0.315673845 | 0.858008045 | -0.06650864 | FALSE | TRUE | FALSE |
| DDC | 0.2707022 | 0.81311292 | 0.5104 | 0.270702192 | 0.270702192 | 0.58940362 | 0.18325261 | 0.856187733 | 0.0149 | 0.183252608 | 0.183252608 | 0.907632806 | 1.477207855 | 0.169441608 | FALSE | TRUE | FALSE |
| DNAJC6 | 0.2706654 | 0.81313326 | 0.38935 | 0.115733962 | 1.081571753 | 3.906003659 | 0.01310501 | 0.89575291 | 1 | 0.2092649 | 3.344948773 | 4.123404994 | 20.65357529 | 1.314995242 | FALSE | TRUE | FALSE |
| MUCL1 | 0.2706187 | 0.8131536 | 1 | 0.270618694 | 0.270618694 | 0.270618694 | 0.31472382 | 0.820304115 | 1 | 0.314723817 | 0.314723817 | 0.314723817 | 0.859860868 | -0.065571815 | FALSE | TRUE | FALSE |
| ANG | 0.2706044 | 0.81317393 | 1 | 0.270604382 | 0.270604382 | 0.270604382 | 0.31546327 | 0.820084841 | 1 | 0.315463269 | 0.315463269 | 0.315463269 | 0.857799969 | -0.066613974 | FALSE | TRUE | FALSE |
| FUOM | 0.2704945 | 0.81319425 | 0.00285 | 0.270494483 | 0.270494483 | 0.270494483 | 0.31506788 | 0.820224441 | 1 | 0.315067875 | 0.315067875 | 0.315067875 | 0.858527652 | -0.066245712 | FALSE | TRUE | FALSE |
| PRKACG | 0.2704435 | 0.81321457 | 0.87575 | 23.72657356 | 230.9232894 | 230.9232894 | 27.8150133 | 0.099901528 | 0.50365 | 27.8150133 | 271.5525515 | 271.5525515 | 0.009722934 | -2.012202655 | FALSE | FALSE | FALSE |
| ATP6V1E2 | 0.2703325 | 0.81323489 | 1 | 0.270332513 | 0.270332513 | 0.270332513 | 0.31497915 | 0.820244367 | 1 | 0.314979146 | 0.314979146 | 0.314979146 | 0.85825527 | -0.066383521 | FALSE | TRUE | FALSE |
| HSPB9 | 0.2703131 | 0.8132552 | 1 | 2.499916028 | 23.72883821 | 23.72883821 | 3.01590096 | 0.410273729 | 0.499 | 0.324814661 | 3.015900963 | 3.015900963 | 0.089629286 | -1.047550062 | FALSE | FALSE | FALSE |
| TMEM41B | 0.2702372 | 0.81327551 | 1 | 0.270237236 | 0.270237236 | 0.270237236 | 0.31468939 | 0.820343926 | 1 | 0.314689392 | 0.314689392 | 0.314689392 | 0.858742758 | -0.066136912 | FALSE | TRUE | FALSE |
| CLDN5 | 0.2701706 | 0.81329582 | 1 | 0.270170573 | 0.270170573 | 0.270170573 | 2.85902827 | 0.423135922 | 0.5024 | 2.859028267 | 2.859028267 | 2.859028267 | 0.094497342 | -1.024580405 | FALSE | FALSE | FALSE |
| GLRA1 | 0.270116 | 0.81331612 | 0.5044 | 0.471496929 | 4.317343072 | 4.317343072 | 0.04705376 | 0.88732398 | 1 | 0.047053765 | 0.047053765 | 0.933254639 | 5.740582915 | 0.758955994 | FALSE | TRUE | FALSE |
| DNAJC9 | 0.2701049 | 0.81333641 | 1 | 0.270104876 | 0.270104876 | 0.270104876 | 0.31465543 | 0.820383718 | 1 | 0.314655431 | 0.314655431 | 0.314655431 | 0.858414791 | -0.066302808 | FALSE | TRUE | FALSE |
| ROBO3 | 0.2700803 | 0.8133567 | 0.2677 | 0.067547749 | 1.080148311 | 5.242488669 | 1.22841634 | 0.564135774 | 0.08495 | 0.137420244 | 4.912052099 | 10.70415284 | 0.219860538 | -0.657852714 | FALSE | FALSE | FALSE |
| ZDHHC19 | 0.2700782 | 0.81337699 | 0.00875 | 0.270078184 | 0.270078184 | 0.270078184 | 0.31452487 | 0.820423491 | 0.00765 | 0.314524869 | 0.314524869 | 0.314524869 | 0.858686259 | -0.066165487 | FALSE | TRUE | FALSE |
| HMGCS1 | 0.2700557 | 0.81339727 | 1 | 0.270055721 | 0.270055721 | 5.352863638 | 0.31542431 | 0.820124749 | 1 | 0.315424305 | 0.315424305 | 6.258847 | 0.856166494 | -0.067441772 | FALSE | TRUE | FALSE |
| CTNNB1 | 0.2700344 | 0.81341754 | 1 | 0.270034364 | 0.270034364 | 5.316374986 | 0.31545897 | 0.820104797 | 1 | 0.315458972 | 0.315458972 | 6.222355107 | 0.856004705 | -0.067523848 | FALSE | TRUE | FALSE |
| FFAR1 | 0.2700252 | 0.81343781 | 0.74445 | 1.080070073 | 4.321238187 | 4.321238187 | 0.32165289 | 0.818209222 | 1 | 0.321652895 | 0.321652895 | 0.321652895 | 0.839492379 | -0.075983242 | FALSE | TRUE | FALSE |
| RNF8 | 0.2699596 | 0.81345808 | 1 | 0.269959591 | 0.269959591 | 5.354872257 | 0.31532259 | 0.820164639 | 1 | 0.31532259 | 0.31532259 | 6.260859472 | 0.85613781 | -0.067456323 | FALSE | TRUE | FALSE |
| POFUT2 | 0.2699521 | 0.81347834 | 1 | 0.269952144 | 0.269952144 | 5.338897953 | 0.31470324 | 0.820324023 | 1 | 0.314703244 | 0.314703244 | 6.232290772 | 0.857799052 | -0.066614438 | FALSE | TRUE | FALSE |
| ZDHHC22 | 0.2699075 | 0.8134986 | 0.0088 | 0.269907523 | 0.269907523 | 0.269907523 | 0.28854479 | 0.828561061 | 1 | 0.288544786 | 0.288544786 | 0.288544786 | 0.935409461 | -0.028998242 | FALSE | TRUE | FALSE |
| INPP1 | 0.2699069 | 0.81351885 | 0.01035 | 0.26990692 | 0.26990692 | 5.358509189 | 0.31447336 | 0.820522843 | 1 | 0.314473358 | 0.314473358 | 0.314473358 | 0.858282307 | -0.06636984 | FALSE | TRUE | FALSE |
| NXPE4 | 0.269848 | 0.8135391 | 0.7494 | 0.269847965 | 2.410240987 | 2.410240987 | 0.04875059 | 0.886988486 | 0.01055 | 0.048750585 | 0.048750585 | 0.967183676 | 5.535276409 | 0.743139313 | FALSE | TRUE | FALSE |
| APBA1 | 0.2698273 | 0.81355935 | 1 | 0.269827284 | 0.269827284 | 5.264203723 | 2.86447963 | 0.422929599 | 0.5215 | 2.864479628 | 2.864479628 | 56.04979656 | 0.094197662 | -1.025959876 | FALSE | FALSE | FALSE |
| ACBD6 | 0.2698199 | 0.81357959 | 1 | 0.26981992 | 0.26981992 | 0.26981992 | 0.31449797 | 0.820463246 | 1 | 0.314497972 | 0.314497972 | 0.314497972 | 0.857938505 | -0.06654384 | FALSE | TRUE | FALSE |
| RTN1 | 0.2697702 | 0.81359982 | 0.0209 | 0.269770247 | 0.269770247 | 5.317013172 | 0.30506962 | 0.823892695 | 1 | 2.850255889 | 2.850255889 | 10.3303375 | 0.884290752 | -0.053404917 | FALSE | TRUE | FALSE |
| PPFIA3 | 0.2697406 | 0.81362005 | 1 | 0.269740627 | 0.269740627 | 9.087074913 | 0.31497045 | 0.820264287 | 1 | 0.314970452 | 0.314970452 | 6.114919246 | 0.856399785 | -0.06732345 | FALSE | TRUE | FALSE |
| ACVR2A | 0.2694237 | 0.81364028 | 1 | 0.269423713 | 0.269423713 | 5.340708497 | 0.31117573 | 0.821608344 | 1 | 1.24444689 | 1.24444689 | 6.174923002 | 0.865824964 | -0.062569897 | FALSE | TRUE | FALSE |
| MCFD2 | 0.2693478 | 0.8136605 | 1 | 0.269347783 | 0.269347783 | 0.269347783 | 0.31451594 | 0.820443371 | 1 | 0.314515945 | 0.314515945 | 0.314515945 | 0.856388324 | -0.067329263 | FALSE | TRUE | FALSE |
| GAS2L1 | 0.2692971 | 0.81368072 | 1 | 0.269297144 | 0.269297144 | 5.325779395 | 0.31439326 | 0.820622076 | 1 | 0.314393261 | 0.314393261 | 6.225982554 | 0.856561438 | -0.067241481 | FALSE | TRUE | FALSE |
| NRSN2 | 0.269244 | 0.81370093 | 1 | 0.269243961 | 0.269243961 | 0.269243961 | 0.3135277 | 0.820840008 | 1 | 0.313527699 | 0.313527699 | 0.313527699 | 0.858756539 | -0.066129943 | FALSE | TRUE | FALSE |
| CCNA1 | 0.2692432 | 0.81372114 | 1 | 0.269243209 | 0.269243209 | 5.339184975 | 0.31387334 | 0.820681565 | 1 | 0.313873343 | 0.313873343 | 6.230565821 | 0.857808461 | -0.066609674 | FALSE | TRUE | FALSE |
| PKIG | 0.2692375 | 0.81374135 | 1 | 0.269237465 | 0.269237465 | 0.269237465 | 0.272554 | 0.833091916 | 1 | 0.272553996 | 0.272553996 | 0.272553996 | 0.987831656 | -0.005317061 | FALSE | TRUE | FALSE |
| CAMTA2 | 0.2691849 | 0.81376155 | 0.0388 | 0.269184899 | 0.269184899 | 5.235952706 | 0.09340679 | 0.877454655 | 1 | 0.093406792 | 0.093406792 | 3.145576405 | 2.881855744 | 0.459672238 | FALSE | TRUE | FALSE |
| ZBTB4 | 0.2691092 | 0.81378174 | 1 | 0.269109204 | 0.269109204 | 5.245190219 | 0.31420301 | 0.820641909 | 1 | 0.31420301 | 0.31420301 | 6.14289607 | 0.856481941 | -0.06728179 | FALSE | TRUE | FALSE |
| MECOM | 0.2689428 | 0.81380194 | 1 | 24.28976902 | 2427.755206 | 2427.755206 | 0.33452575 | 0.814123256 | 1 | 3.148399736 | 30.41129227 | 61.90668512 | 0.803952343 | -0.094769695 | FALSE | TRUE | FALSE |
| HSD17B2 | 0.2687461 | 0.81382213 | 0.51045 | 2.491462746 | 23.69098768 | 23.69098768 | 3.67775409 | 0.372798906 | 0.32025 | 3.677754091 | 344.1610654 | 344.1610654 | 0.073073421 | -1.136240559 | FALSE | FALSE | FALSE |
| FBXL6 | 0.2687444 | 0.81384231 | 0.01605 | 0.26874437 | 0.26874437 | 0.606794314 | 0.04569032 | 0.887633333 | 0.5083 | 0.403471078 | 0.403471078 | 0.903971931 | 5.881866209 | 0.769515142 | FALSE | TRUE | FALSE |
| GHRL | 0.2685356 | 0.81386249 | 1 | 0.268535556 | 2.462508947 | 2.462508947 | 0.32235244 | 0.818025614 | 1 | 0.322352444 | 0.322352444 | 0.322352444 | 0.833049542 | -0.07932917 | FALSE | TRUE | FALSE |

| | | | | | | | | | | | | | | | | | |
|---|---|---|---|---|---|---|---|---|---|---|---|---|---|---|---|---|---|
| ZNF12 | 0.2685113 | 0.81388267 | 1 | 0.268511311 | 0.268511311 | 5.307528289 | 0.31332969 | 0.820879574 | 1 | 0.313329688 | 0.313329688 | 6.202130371 | 0.856960963 | -0.067038961 | FALSE | TRUE | FALSE |
| ZCCHC2 | 0.268334 | 0.81390284 | 1 | 24.09551195 | 2394.525447 | 2394.525447 | 26.4377829 | 0.103457822 | 0.89255 | 260.3066292 | 27160.44571 | 288593.6538 | 0.010149643 | -1.993549247 | FALSE | FALSE | FALSE |
| SLC25A28 | 0.2682864 | 0.81392301 | 1 | 0.268286369 | 0.268286369 | 5.323735388 | 0.31315119 | 0.820958653 | 1 | 0.313151192 | 0.313151192 | 0.313151192 | 0.85673111 | -0.067155462 | FALSE | TRUE | FALSE |
| CA10 | 0.2682654 | 0.81394317 | 1 | 1.072798004 | 4.291209779 | 5.330068711 | 0.31697781 | 0.819724867 | 1 | 1.267641024 | 1.267641024 | 1.267641024 | 0.846322475 | -0.072464126 | FALSE | TRUE | FALSE |
| C16orf90 | 0.2682562 | 0.81396333 | 1 | 0.268256182 | 0.268256182 | 0.268256182 | 0.31241216 | 0.821215164 | 1 | 0.312412159 | 0.312412159 | 0.312412159 | 0.858661143 | -0.06617819 | FALSE | TRUE | FALSE |
| SENP3 | 0.2681898 | 0.81398348 | 1 | 319724219.8 | 5.11517E+11 | 7.74757E+13 | 0.40912051 | 0.78876012 | 1 | 7196.719967 | 4369056.57 | 48452809.71 | 0.655527541 | -0.183409058 | FALSE | TRUE | FALSE |
| NEDD1 | 0.2681599 | 0.81400363 | 0.50995 | 2.481525458 | 2.481525458 | 5.303430698 | 0.32265675 | 0.817841624 | 1 | 0.322656753 | 0.322656753 | 6.389775751 | 0.831099749 | -0.080346849 | FALSE | TRUE | FALSE |
| SYCP2L | 0.268105 | 0.81402378 | 0.87445 | 2.395843178 | 21.96048363 | 21.96048363 | 0.04989587 | 0.886651187 | 0.0168 | 0.049895873 | 0.049895873 | 0.985461926 | 5.373289813 | 0.730240265 | FALSE | TRUE | FALSE |
| PTH1R | 0.2680768 | 0.81404392 | 1 | 0.268076752 | 0.268076752 | 5.285701914 | 0.31318361 | 0.82093889 | 1 | 0.313183609 | 0.313183609 | 6.185590567 | 0.855973123 | -0.067539872 | FALSE | TRUE | FALSE |
| PACSIN2 | 0.2680758 | 0.81406406 | 1 | 0.268075789 | 0.268075789 | 5.294126943 | 0.31277639 | 0.821136318 | 1 | 0.312776391 | 0.312776391 | 6.186243976 | 0.857084475 | -0.066976371 | FALSE | TRUE | FALSE |
| LRRC3 | 0.2680403 | 0.81408419 | 1 | 2.522163877 | 2.522163877 | 2.522163877 | 0.31964115 | 0.818899482 | 1 | 1.278265724 | 1.278265724 | 1.278265724 | 0.838566277 | -0.076462607 | FALSE | TRUE | FALSE |
| TSPAN16 | 0.2678787 | 0.81410431 | 0.0064 | 0.267878724 | 0.267878724 | 0.267878724 | 0.3028169 | 0.824501992 | 1 | 2.800906446 | 2.800906446 | 2.800906446 | 0.88462276 | -0.053241891 | FALSE | TRUE | FALSE |
| GTPBP1 | 0.2677871 | 0.81412444 | 1 | 0.267787114 | 0.267787114 | 5.274848195 | 0.31295145 | 0.821017916 | 1 | 0.312951446 | 0.312951446 | 6.175698368 | 0.855682624 | -0.067687287 | FALSE | TRUE | FALSE |
| DUSP5 | 0.267775 | 0.81414456 | 1 | 0.267774997 | 0.267774997 | 5.302029787 | 0.31284276 | 0.821096869 | 1 | 0.312842758 | 0.312842758 | 6.201825085 | 0.855941172 | -0.067556083 | FALSE | TRUE | FALSE |
| GPCPD1 | 0.2677731 | 0.81416467 | 1 | 0.267773133 | 0.267773133 | 5.291077593 | 0.31273729 | 0.821156036 | 1 | 0.312737294 | 0.312737294 | 6.188488085 | 0.856223861 | -0.067412673 | FALSE | TRUE | FALSE |
| INMT | 0.2677696 | 0.81418478 | 1 | 0.267769621 | 0.267769621 | 0.267769621 | 0.31206935 | 0.821333303 | 1 | 0.312069349 | 0.312069349 | 0.312069349 | 0.858045244 | -0.066489812 | FALSE | TRUE | FALSE |
| FBXL12 | 0.2677232 | 0.81420489 | 1 | 2.499704001 | 23.95409792 | 23.95409792 | 0.31382739 | 0.820721203 | 0.00885 | 0.313827393 | 0.313827393 | 0.313827393 | 0.853090564 | -0.069004862 | FALSE | TRUE | FALSE |
| LYAR | 0.2677022 | 0.81422499 | 1 | 0.267702158 | 0.267702158 | 0.267702158 | 0.3122862 | 0.821254562 | 1 | 0.312286202 | 0.312286202 | 0.312286202 | 0.857233387 | -0.066900923 | FALSE | TRUE | FALSE |
| TRAPPC8 | 0.2676904 | 0.81424508 | 0.03855 | 0.267690399 | 0.267690399 | 8.992989276 | 0.31291281 | 0.821057402 | 1 | 0.312912807 | 0.312912807 | 10.52729996 | 0.855479204 | -0.067790544 | FALSE | TRUE | FALSE |
| NOL9 | 0.2676244 | 0.81426518 | 0.017 | 0.267624357 | 0.267624357 | 5.283348582 | 0.31235722 | 0.821234865 | 1 | 0.312357221 | 0.312357221 | 6.176034191 | 0.856789403 | -0.067125913 | FALSE | TRUE | FALSE |
| TLR3 | 0.2675742 | 0.81428526 | 0.50945 | 0.267574177 | 0.267574177 | 5.270624853 | 0.17727327 | 0.857712855 | 0.7569 | 0.400717362 | 1.60236342 | 7.907608741 | 1.509388175 | 0.178800943 | FALSE | TRUE | FALSE |
| CPNE4 | 0.267329 | 0.81430535 | 1 | 0.267329039 | 0.267329039 | 5.288519947 | 0.3122404 | 0.821274254 | 1 | 0.312240397 | 0.312240397 | 6.185062529 | 0.856164166 | -0.067442953 | FALSE | TRUE | FALSE |
| FBL | 0.2672511 | 0.81432543 | 1 | 1.068696311 | 1.068696311 | 5.300676064 | 0.31603426 | 0.819965006 | 1 | 0.31603426 | 0.31603426 | 0.31603426 | 0.845639636 | -0.072814669 | FALSE | TRUE | FALSE |
| HTR1F | 0.2672068 | 0.8143455 | 1 | 2.48781731 | 2.48781731 | 2.48781731 | 0.32192511 | 0.818148058 | 0.00835 | 0.321925106 | 0.321925106 | 0.321925106 | 0.83002774 | -0.080907393 | FALSE | TRUE | FALSE |
| KCNJ4 | 0.2671794 | 0.81436557 | 1 | 0.267179424 | 0.267179424 | 5.263224209 | 1.22287903 | 0.564873277 | 0.5027 | 1.222879026 | 1.222879026 | 6.03482941 | 0.218483937 | -0.660580487 | FALSE | FALSE | FALSE |
| LMO2 | 0.2671738 | 0.81438564 | 0.50095 | 1.068329521 | 1.068329521 | 1.068329521 | 1.24923141 | 0.561161377 | 0.00455 | 1.249231409 | 1.249231409 | 1.249231409 | 0.213870566 | -0.669848981 | FALSE | FALSE | FALSE |
| TIPIN | 0.2671155 | 0.8144057 | 1 | 0.26711552 | 0.26711552 | 0.26711552 | 2.61413571 | 0.439605454 | 0.87495 | 24.69819134 | 239.3368267 | 239.3368267 | 0.102181198 | -0.990629008 | FALSE | FALSE | FALSE |
| NPBWR2 | 0.2670881 | 0.81442576 | 1 | 0.26708813 | 0.26708813 | 5.286427748 | 0.30230253 | 0.824729368 | 1 | 2.816366772 | 2.816366772 | 5.990876186 | 0.883512711 | -0.053787198 | FALSE | TRUE | FALSE |
| METAP2 | 0.267076 | 0.81444581 | 1 | 0.267076043 | 0.267076043 | 5.293627393 | 0.3119758 | 0.821372647 | 1 | 0.311975798 | 0.311975798 | 6.190239239 | 0.856079367 | -0.06748597 | FALSE | TRUE | FALSE |
| HAUS4 | 0.2669705 | 0.81446586 | 1 | 2.45710081 | 2.45710081 | 5.302380858 | 0.32103173 | 0.818433125 | 1 | 0.321031729 | 0.321031729 | 0.321031729 | 0.831601695 | -0.080084634 | FALSE | TRUE | FALSE |
| MRGPRE | 0.2669704 | 0.81448591 | 1 | 0.266970416 | 0.266970416 | 0.266970416 | 224.957524 | 0.011460855 | 0.31955 | 22.83581153 | 2271.508288 | 2271.508288 | 0.001186759 | -2.925637385 | FALSE | FALSE | FALSE |
| LEMD2 | 0.2669657 | 0.81450594 | 1 | 0.266965744 | 0.266965744 | 5.290249983 | 0.28563694 | 0.829443729 | 1 | 2.673733791 | 2.673733791 | 5.666498569 | 0.934633104 | -0.029358841 | FALSE | TRUE | FALSE |
| GTF2E2 | 0.266945 | 0.81452598 | 1 | 0.266945024 | 0.266945024 | 0.266945024 | 0.28647263 | 0.829192469 | 1 | 0.286472631 | 0.286472631 | 0.286472631 | 0.931834303 | -0.030661306 | FALSE | TRUE | FALSE |
| LYSMD1 | 0.2669155 | 0.81454601 | 1 | 0.266915494 | 0.266915494 | 0.266915494 | 2288.85244 | 0.001029593 | 0.002 | 23.85304991 | 2288.852435 | 2288.852435 | 0.000116615 | -3.93324401 | FALSE | FALSE | FALSE |
| RNFT2 | 0.2666458 | 0.81456604 | 0.0107 | 0.266645824 | 0.266645824 | 5.29244611 | 0.31109805 | 0.821647566 | 0.0092 | 0.311098052 | 0.311098052 | 0.311098052 | 0.857111842 | -0.066962505 | FALSE | TRUE | FALSE |
| EPS8 | 0.2665642 | 0.81458606 | 1 | 0.266564185 | 0.266564185 | 5.248111601 | 0.31141416 | 0.821549478 | 1 | 0.311414161 | 0.311414161 | 6.142630656 | 0.855979652 | -0.067536559 | FALSE | TRUE | FALSE |
| SCAF8 | 0.2665604 | 0.81460608 | 1 | 2.503034491 | 2.503034491 | 48.87108124 | 0.32171063 | 0.818188839 | 1 | 0.321710628 | 0.321710628 | 10.83056785 | 0.828571896 | -0.081669802 | FALSE | TRUE | FALSE |
| RAB3C | 0.2665454 | 0.81462609 | 1 | 1.065936516 | 4.263831482 | 4.263831482 | 1.25999966 | 0.560411667 | 0.50535 | 1.259999658 | 1.259999658 | 1.259999658 | 0.211544034 | -0.674599218 | FALSE | FALSE | FALSE |
| MRPL46 | 0.2665393 | 0.8146461 | 1 | 0.266539311 | 0.266539311 | 0.266539311 | 0.31108997 | 0.82168677 | 1 | 0.311089968 | 0.311089968 | 0.311089968 | 0.856791729 | -0.067124735 | FALSE | TRUE | FALSE |
| NR2C2AP | 0.2665116 | 0.81466611 | 1 | 0.266511629 | 0.266511629 | 0.266511629 | 0.31135532 | 0.821569104 | 0.00335 | 0.311355323 | 0.311355323 | 0.311355323 | 0.855972612 | -0.067540131 | FALSE | TRUE | FALSE |
| SAPCD2 | 0.2664517 | 0.81468611 | 1 | 0.266451676 | 0.266451676 | 0.266451676 | 0.29235927 | 0.827339181 | 0.74985 | 2.719786061 | 25.96819586 | 25.96819586 | 0.911384403 | -0.040298408 | FALSE | TRUE | FALSE |
| ADAM19 | 0.2663583 | 0.8147061 | 0.51045 | 0.266358288 | 0.266358288 | 5.22368049 | 0.04574116 | 0.887617089 | 1 | 0.045741158 | 0.045741158 | 0.899150389 | 5.823164553 | 0.765159063 | FALSE | TRUE | FALSE |
| ABT1 | 0.2662882 | 0.8147261 | 1 | 0.266288197 | 0.266288197 | 5.274218937 | 0.31075771 | 0.821804274 | 1 | 0.310757708 | 0.310757708 | 6.162166688 | 0.856899733 | -0.067069992 | FALSE | TRUE | FALSE |
| RNF43 | 0.2662801 | 0.81474608 | 1 | 0.266280142 | 0.266280142 | 5.241223487 | 0.31109396 | 0.82166717 | 1 | 0.311093957 | 0.311093957 | 6.134977515 | 0.855947654 | -0.067552794 | FALSE | TRUE | FALSE |
| DGCR6L | 0.2661921 | 0.81476607 | 0.009 | 0.266192089 | 0.266192089 | 0.266192089 | 0.31043674 | 0.821902075 | 1 | 0.310436737 | 0.310436737 | 0.310436737 | 0.857476121 | -0.066777965 | FALSE | TRUE | FALSE |
| PIK3C3 | 0.2661647 | 0.81478605 | 0.26895 | 0.066561703 | 0.266164713 | 2.247188607 | 3.76981824 | 0.368204743 | 0.13855 | 0.416689395 | 3.769818236 | 8.213744541 | 0.070604124 | -1.151169933 | FALSE | FALSE | FALSE |
| SPATA19 | 0.2660392 | 0.81480602 | 1 | 0.26603915 | 0.26603915 | 0.26603915 | 0.30076349 | 0.825276355 | 0.5034 | 2.768732959 | 2.768732959 | 2.768732959 | 0.884546018 | -0.053279568 | FALSE | TRUE | FALSE |
| ST3GAL5 | 0.2659862 | 0.81482599 | 1 | 0.265986172 | 0.265986172 | 5.279591745 | 0.30103952 | 0.825238743 | 1 | 2.787862191 | 2.787862191 | 2.787862191 | 0.883558976 | -0.053764457 | FALSE | TRUE | FALSE |
| MSGN1 | 0.265943 | 0.81484596 | 1 | 0.265943039 | 0.265943039 | 0.265943039 | 0.31079043 | 0.821784701 | 1 | 0.310790426 | 0.310790426 | 0.310790426 | 0.855698943 | -0.067679005 | FALSE | TRUE | FALSE |
| DRGX | 0.2659268 | 0.81486592 | 0.00845 | 0.265926833 | 0.265926833 | 0.265926833 | 0.36639347 | 0.803662906 | 0.49025 | 0.091651977 | 0.366393474 | 0.366393474 | 0.725795768 | -0.139185569 | FALSE | TRUE | FALSE |
| NCEH1 | 0.2659158 | 0.81488588 | 1 | 1.063628302 | 1.063628302 | 5.267110513 | 0.31394245 | 0.820661739 | 1 | 0.313942445 | 0.313942445 | 6.22559776 | 0.847020719 | -0.072105966 | FALSE | TRUE | FALSE |
| PTK7 | 0.2658903 | 0.81490583 | 0.51865 | 0.265890343 | 1.063438275 | 5.177648793 | 0.02751147 | 0.891830827 | 0.5078 | 0.109986942 | 0.109986942 | 2.148565314 | 9.664709847 | 0.98518882 | FALSE | TRUE | FALSE |
| HVCN1 | 0.2658763 | 0.81492578 | 1 | 0.265876278 | 0.265876278 | 0.265876278 | 1.21613447 | 0.565852948 | 0.74465 | 1.216134466 | 4.864570126 | 4.864570126 | 0.21862408 | -0.660302006 | FALSE | FALSE | FALSE |
| CSF2RB | 0.2656776 | 0.81494572 | 0.5099 | 0.265677581 | 0.265677581 | 5.208781251 | 0.04508259 | 0.887844091 | 1 | 0.045082593 | 0.045082593 | 0.885972305 | 5.893129968 | 0.770346019 | FALSE | TRUE | FALSE |
| CUL2 | 0.2656748 | 0.81496566 | 1 | 0.265674768 | 0.265674768 | 5.238363035 | 0.31081743 | 0.821745541 | 0.019 | 0.310817426 | 0.310817426 | 6.138878865 | 0.854761496 | -0.06815505 | FALSE | TRUE | FALSE |
| RNASE6 | 0.2655602 | 0.8149856 | 1 | 0.265560166 | 0.265560166 | 0.265560166 | 0.30982932 | 0.822116856 | 1 | 0.309829317 | 0.309829317 | 0.309829317 | 0.857117617 | -0.066959578 | FALSE | TRUE | FALSE |
| OSER1 | 0.2655447 | 0.81500553 | 1 | 0.265544745 | 0.265544745 | 0.265544745 | 10.0179486 | 0.197598698 | 0.00265 | 0.29539504 | 0.29539504 | 0.29539504 | 0.026506898 | -1.576641088 | FALSE | FALSE | FALSE |
| NOL3 | 0.2654749 | 0.81502546 | 0.7547 | 0.265474892 | 2.510453842 | 2.510453842 | 25.6718886 | 0.107052548 | 0.1908 | 25.67188859 | 255.4235501 | 2603.400404 | 0.010341074 | -1.985434366 | FALSE | FALSE | FALSE |
| PENK | 0.26537 | 0.81504538 | 1 | 2.453205304 | 2.453205304 | 2.453205304 | 2.87331073 | 0.421481006 | 0.0082 | 2.873310734 | 2.873310734 | 2.873310734 | 0.092356885 | -1.034530724 | FALSE | FALSE | FALSE |
| PPP1R26 | 0.2653481 | 0.8150653 | 1 | 0.26534815 | 0.26534815 | 5.156226707 | 0.31033685 | 0.821960702 | 1 | 0.310336846 | 0.310336846 | 6.051093418 | 0.855032694 | -0.068017279 | FALSE | TRUE | FALSE |
| APOA5 | 0.2652763 | 0.81508521 | 0.50615 | 0.265276289 | 0.265276289 | 1.312650909 | 0.0893352 | 0.878276231 | 1 | 0.089335199 | 0.817276788 | 1.769901042 | 2.969448687 | 0.472675825 | FALSE | TRUE | FALSE |
| KIF5B | 0.26497 | 0.81510512 | 0.75905 | 1.059256049 | 16.94084594 | 20.80624188 | 0.04476966 | 0.887924957 | 1 | 0.412849535 | 1.650552761 | 8.126782895 | 5.91851735 | 0.772212925 | FALSE | TRUE | FALSE |
| PUM1 | 0.2649389 | 0.81512503 | 1 | 0.264938936 | 0.264938936 | 5.166539127 | 0.31003495 | 0.821980237 | 1 | 0.310034945 | 0.310034945 | 6.064133135 | 0.8545454 | -0.068264859 | FALSE | TRUE | FALSE |
| ZBTB38 | 0.2647154 | 0.81514494 | 1 | 0.264715443 | 0.264715443 | 5.170089179 | 0.30989167 | 0.822058332 | 1 | 0.309891671 | 0.309891671 | 6.069524054 | 0.85421929 | -0.068430626 | FALSE | TRUE | FALSE |
| OR5V1 | 0.2646766 | 0.81516483 | 1 | 2.437241845 | 2.437241845 | 2.437241845 | 0.31863437 | 0.819162007 | 1 | 0.318634368 | 0.318634368 | 0.318634368 | 0.83065944 | -0.080576995 | FALSE | TRUE | FALSE |
| KRTAP1-5 | 0.264657 | 0.81518473 | 1 | 2.474935526 | 2.474935526 | 2.474935526 | 0.29512174 | 0.826693915 | 1 | 0.295121738 | 0.295121738 | 0.295121738 | 0.896772199 | -0.047317864 | FALSE | TRUE | FALSE |
| LACC1 | 0.2646266 | 0.81520462 | 1 | 0.264626582 | 0.264626582 | 0.264626582 | 0.30899105 | 0.822292201 | 1 | 0.308991049 | 0.308991049 | 0.308991049 | 0.856421511 | -0.067312433 | FALSE | TRUE | FALSE |
| BHLHA15 | 0.2645402 | 0.81522451 | 1 | 0.264540226 | 0.264540226 | 0.264540226 | 0.30919107 | 0.82223379 | 1 | 0.309191067 | 0.309191067 | 0.309191067 | 0.855588191 | -0.067735218 | FALSE | TRUE | FALSE |
| ZSWIM3 | 0.2645292 | 0.81524439 | 0.0214 | 0.26452923 | 0.26452923 | 5.21409851 | 0.29966529 | 0.825576671 | 1 | 2.782472494 | 2.782472494 | 10.13955352 | 0.882748985 | -0.054162773 | FALSE | TRUE | FALSE |
| OR8I2 | 0.264395 | 0.81526427 | 1 | 0.264395037 | 0.264395037 | 0.264395037 | 0.29926472 | 0.825689027 | 1 | 2.763317493 | 2.763317493 | 2.763317493 | 0.883482156 | -0.053802218 | FALSE | TRUE | FALSE |
| FBXO42 | 0.2642608 | 0.81528414 | 1 | 23.41388077 | 229.0045713 | 461.4332739 | 0.33927781 | 0.81227084 | 1 | 0.339277811 | 0.339277811 | 6.698140742 | 0.778892136 | -0.108522681 | FALSE | TRUE | FALSE |
| CYTH4 | 0.2642236 | 0.81530401 | 0.0151 | 0.264223644 | 0.264223644 | 5.226744644 | 0.3091743 | 0.822253265 | 1 | 0.309174301 | 0.309174301 | 6.123974414 | 0.854610629 | -0.06823171 | FALSE | TRUE | FALSE |
| RNF214 | 0.2642167 | 0.81532388 | 1 | 0.264216696 | 0.264216696 | 5.213984968 | 0.30921621 | 0.822214311 | 1 | 0.309216205 | 0.309216205 | 6.111769295 | 0.854472347 | -0.068301988 | FALSE | TRUE | FALSE |

| | | | | | | | | | | | | | | | | | |
|---|---|---|---|---|---|---|---|---|---|---|---|---|---|---|---|---|---|
| DAXX | 0.2641914 | 0.81534374 | 1 | 0.264191365 | 0.264191365 | 5.196010592 | 0.3088328 | 0.822370022 | 1 | 0.308832799 | 0.308832799 | 6.086153324 | 0.855451123 | -0.067804799 | FALSE | TRUE | FALSE |
| PLA2G2D | 0.2641794 | 0.81536359 | 1 | 0.264179378 | 0.264179378 | 0.264179378 | 0.308351 | 0.822525456 | 1 | 0.308350998 | 0.308350998 | 0.308350998 | 0.856748899 | -0.067146445 | FALSE | TRUE | FALSE |
| GBP1 | 0.2641601 | 0.81538345 | 0.75305 | 0.264160122 | 2.411068803 | 5.225012433 | 0.18717297 | 0.855192426 | 0.50805 | 0.18717297 | 0.18717297 | 0.927021673 | 1.411315543 | 0.149624125 | FALSE | TRUE | FALSE |
| PROM2 | 0.2640526 | 0.81540329 | 0.9407 | 2.378696329 | 208.1834031 | 208.1834031 | 0.04625519 | 0.887478848 | 0.8761 | 3.728973617 | 34.79182519 | 73.34584471 | 5.708605531 | 0.756530034 | FALSE | TRUE | FALSE |
| ZNF74 | 0.2639392 | 0.81542314 | 0.51395 | 0.263939208 | 0.263939208 | 5.177217633 | 3.50608363 | 0.382729419 | 0.26065 | 0.385803979 | 3.50608363 | 7.585152408 | 0.075280351 | -1.123318362 | FALSE | FALSE | FALSE |
| GPC6 | 0.2637575 | 0.81544298 | 1 | 0.263757544 | 0.263757544 | 5.209156273 | 0.30879794 | 0.822389466 | 1 | 0.308797945 | 0.308797945 | 6.107875286 | 0.85414281 | -0.068469511 | FALSE | TRUE | FALSE |
| CDKN1B | 0.2634836 | 0.81546281 | 1 | 0.263483634 | 0.263483634 | 0.263483634 | 1.21979772 | 0.565118609 | 0.4965 | 0.305021117 | 1.219797725 | 1.219797725 | 0.216006005 | -0.665534175 | FALSE | FALSE | FALSE |
| SELENBP1 | 0.2634408 | 0.81548265 | 0.50595 | 0.263440797 | 0.263440797 | 1.304645696 | 0.09169404 | 0.877795827 | 1 | 0.091694045 | 0.091694045 | 1.817806222 | 2.873041513 | 0.458341901 | FALSE | TRUE | FALSE |
| CRISP3 | 0.2633843 | 0.81550248 | 1 | 0.263384267 | 0.263384267 | 0.263384267 | 0.30768183 | 0.822912795 | 1 | 0.307681833 | 0.307681833 | 0.307681833 | 0.85602801 | -0.067512025 | FALSE | TRUE | FALSE |
| SFN | 0.26338 | 0.8155223 | 0.0093 | 0.263379966 | 0.263379966 | 0.263379966 | 0.30838128 | 0.822486623 | 0.0067 | 0.308381285 | 0.308381285 | 0.308381285 | 0.854072473 | -0.068505276 | FALSE | TRUE | FALSE |
| F11R | 0.2633585 | 0.81554212 | 0.0084 | 0.263358482 | 0.263358482 | 0.263358482 | 0.30797766 | 0.822796778 | 1 | 0.307977663 | 0.307977663 | 0.307977663 | 0.85512202 | -0.06797191 | FALSE | TRUE | FALSE |
| ITFG1 | 0.2632617 | 0.81556194 | 1 | 0.263261746 | 0.263261746 | 5.204543619 | 0.30808553 | 0.822719347 | 0.0164 | 0.30808553 | 0.30808553 | 6.099132382 | 0.854508636 | -0.068283544 | FALSE | TRUE | FALSE |
| SEMA4B | 0.2632302 | 0.81558175 | 0.9999 | 20912.1998 | 24133855.21 | 264473680.5 | 4.13206995 | 0.347429153 | 0.85765 | 376.5334345 | 37791.94049 | 392033.3283 | 0.06370419 | -1.195832003 | FALSE | FALSE | FALSE |
| EPN2 | 0.2632189 | 0.81560155 | 1 | 0.263218891 | 0.263218891 | 5.187796327 | 0.30802351 | 0.822777427 | 1 | 0.308023513 | 0.308023513 | 6.081482148 | 0.854541554 | -0.068266814 | FALSE | TRUE | FALSE |
| NSMCE1 | 0.2632172 | 0.81562136 | 1 | 0.263217199 | 0.263217199 | 0.263217199 | 0.30761973 | 0.822970743 | 1 | 0.307619731 | 0.307619731 | 0.307619731 | 0.855657724 | -0.067699925 | FALSE | TRUE | FALSE |
| HSD17B3 | 0.2631722 | 0.81564115 | 0.4955 | 0.065825285 | 0.263172204 | 0.263172204 | 0.09164279 | 0.87780528 | 0.0066 | 0.091642789 | 0.091642789 | 0.091642789 | 2.871717541 | 0.458141721 | FALSE | TRUE | FALSE |
| CAMK4 | 0.2631088 | 0.81566095 | 1 | 0.263108789 | 0.263108789 | 5.210549726 | 2.7751451 | 0.429062004 | 0.5035 | 2.7751451 | 2.7751451 | 5.914752962 | 0.094809021 | -1.02315034 | FALSE | FALSE | FALSE |
| GTF3C2 | 0.2630973 | 0.81568074 | 1 | 0.26309733 | 0.26309733 | 8.861463547 | 0.30815303 | 0.822699978 | 1 | 0.308153032 | 0.308153032 | 6.064812229 | 0.853787901 | -0.068650004 | FALSE | TRUE | FALSE |
| GLS | 0.2630418 | 0.81570052 | 0.50775 | 2.435392666 | 2.435392666 | 5.214243751 | 0.31724581 | 0.819604558 | 1 | 0.317245814 | 0.317245814 | 6.295421514 | 0.829141923 | -0.081371126 | FALSE | TRUE | FALSE |
| ISCA2 | 0.2629748 | 0.8157203 | 1 | 2.431328022 | 2.431328022 | 2.431328022 | 0.31700847 | 0.819684782 | 1 | 0.317008468 | 0.317008468 | 0.317008468 | 0.8295514 | -0.081156699 | FALSE | TRUE | FALSE |
| CDCP2 | 0.2628748 | 0.81574008 | 0.94115 | 2.348705093 | 202.2116893 | 202.2116893 | 3.6748905 | 0.373038912 | 0.1341 | 3.674890503 | 34.03683528 | 34.03683528 | 0.071532698 | -1.145495397 | FALSE | FALSE | FALSE |
| CNIH4 | 0.2628737 | 0.81575985 | 1 | 0.26287365 | 0.26287365 | 0.26287365 | 0.30694299 | 0.823144364 | 1 | 0.306942993 | 0.306942993 | 0.306942993 | 0.856424991 | -0.067310668 | FALSE | TRUE | FALSE |
| EEFSEC | 0.2628314 | 0.81577962 | 0.5043 | 0.262831413 | 1.050779275 | 5.179920335 | 0.04568519 | 0.887641453 | 1 | 0.045685193 | 0.045685193 | 0.901951855 | 5.753098467 | 0.759901807 | FALSE | TRUE | FALSE |
| OR2F1 | 0.2628189 | 0.81579938 | 1 | 2.420133885 | 2.420133885 | 2.420133885 | 2.7488606 | 0.431078667 | 0.7491 | 2.748860603 | 26.04852574 | 26.04852574 | 0.095610127 | -1.019496105 | FALSE | FALSE | FALSE |
| RLF | 0.2626795 | 0.81581914 | 1 | 0.262679481 | 0.262679481 | 8.797302581 | 0.30777974 | 0.822854807 | 1 | 0.307779743 | 0.307779743 | 10.32632559 | 0.853465789 | -0.068813883 | FALSE | TRUE | FALSE |
| GNAZ | 0.2626661 | 0.8158389 | 1 | 0.262666133 | 0.262666133 | 5.195441835 | 0.30783651 | 0.822816126 | 1 | 0.30783651 | 0.30783651 | 6.096966498 | 0.853265046 | -0.068916045 | FALSE | TRUE | FALSE |
| LRFN5 | 0.2626652 | 0.81585865 | 1 | 1.050511268 | 1.050511268 | 8.887692143 | 0.31105648 | 0.821725954 | 1 | 0.311056478 | 0.311056478 | 6.149065859 | 0.84442935 | -0.07343668 | FALSE | TRUE | FALSE |
| ZNF382 | 0.2626304 | 0.81587839 | 1 | 0.262630353 | 0.262630353 | 5.202974112 | 0.30713701 | 0.82304795 | 1 | 0.307137011 | 0.307137011 | 6.091611844 | 0.855091844 | -0.067987236 | FALSE | TRUE | FALSE |
| MEF2D | 0.2626054 | 0.81589813 | 1 | 0.262605372 | 0.262605372 | 5.191422408 | 0.30755412 | 0.82299005 | 1 | 0.307554117 | 0.307554117 | 6.088462181 | 0.85385094 | -0.068617939 | FALSE | TRUE | FALSE |
| OR10G3 | 0.262603 | 0.81591787 | 0.00775 | 0.262603017 | 0.262603017 | 0.262603017 | 25.296308 | 0.108852798 | 0.2522 | 2.666683004 | 25.29630801 | 25.29630801 | 0.010381081 | -1.983757429 | FALSE | FALSE | FALSE |
| ZNF521 | 0.2626028 | 0.8159376 | 1 | 2.476032617 | 23.96059909 | 83.33309072 | 0.32778151 | 0.816043305 | 0.0327 | 0.327781507 | 0.327781507 | 6.407733335 | 0.801152086 | -0.096285032 | FALSE | TRUE | FALSE |
| CASQ2 | 0.2625923 | 0.81595733 | 0.7513 | 0.262592319 | 1.050229428 | 1.052763839 | 1.39658798 | 0.545304429 | 0.8161 | 5.587194165 | 22.35773147 | 89.48910953 | 0.188024187 | -0.72578628 | FALSE | FALSE | FALSE |
| USE1 | 0.2625701 | 0.81597706 | 0.00845 | 0.262570055 | 0.262570055 | 0.262570055 | 0.30674732 | 0.823240668 | 1 | 0.306747316 | 0.306747316 | 0.306747316 | 0.855981589 | -0.067535576 | FALSE | TRUE | FALSE |
| POLD1 | 0.2624702 | 0.81599678 | 0.5208 | 2.374719805 | 22.03791176 | 46.11153747 | 0.04727429 | 0.887274994 | 1 | 0.420962813 | 0.420962813 | 8.202813842 | 5.552070057 | 0.744454937 | FALSE | TRUE | FALSE |
| TNP2 | 0.2624219 | 0.81601649 | 1 | 0.262421863 | 0.262421863 | 0.262421863 | 0.30633712 | 0.823375313 | 1 | 0.306337115 | 0.306337115 | 0.306337115 | 0.856644037 | -0.067199604 | FALSE | TRUE | FALSE |
| SLC22A17 | 0.2622515 | 0.8160362 | 1 | 0.262251465 | 0.262251465 | 5.191289427 | 0.30707906 | 0.823067242 | 1 | 0.307079061 | 0.307079061 | 6.086147356 | 0.854019365 | -0.068532281 | FALSE | TRUE | FALSE |
| SLC2A2 | 0.2622381 | 0.81605591 | 0.75855 | 0.26223809 | 2.427300232 | 2.427300232 | 0.41102044 | 0.788176581 | 0.502 | 0.411020442 | 0.411020442 | 0.898255637 | 0.638017147 | -0.195167649 | FALSE | TRUE | FALSE |
| OR3A1 | 0.2621946 | 0.81607561 | 0.9416 | 22.91113701 | 2215.723093 | 2215.723093 | 25.2595508 | 0.109152549 | 0.65915 | 245.7282886 | 25029.89051 | 261775.1842 | 0.010380016 | -1.983801959 | FALSE | FALSE | FALSE |
| TWF1 | 0.2621548 | 0.81609531 | 1 | 0.262154766 | 0.262154766 | 0.262154766 | 1.21341342 | 0.566219222 | 0.50005 | 1.213413422 | 1.213413422 | 1.213413422 | 0.21604736 | -0.665451037 | FALSE | FALSE | FALSE |
| ANKRD7 | 0.2621328 | 0.81611501 | 1 | 0.262132778 | 0.262132778 | 0.262132778 | 2.74398403 | 0.431480348 | 0.00615 | 2.743984027 | 2.743984027 | 2.743984027 | 0.095529994 | -1.019860249 | FALSE | FALSE | FALSE |
| CSNK1A1L | 0.2621017 | 0.8161347 | 0.25745 | 0.262101661 | 2.343847395 | 2.343847395 | 0.38235105 | 0.798321646 | 0.87595 | 0.382351052 | 31.91841854 | 31.91841854 | 0.685500038 | -0.163992517 | FALSE | TRUE | FALSE |
| IDI1 | 0.2620613 | 0.81615438 | 1 | 0.262061294 | 0.262061294 | 0.262061294 | 0.30633247 | 0.823394531 | 1 | 0.306332474 | 0.306332474 | 0.306332474 | 0.855479964 | -0.067790157 | FALSE | TRUE | FALSE |
| SHC3 | 0.2620456 | 0.81617406 | 1 | 0.262045631 | 0.262045631 | 5.180497719 | 0.3070266 | 0.823105812 | 1 | 0.307026605 | 0.307026605 | 6.078164867 | 0.853494868 | -0.068799086 | FALSE | TRUE | FALSE |
| ELF3 | 0.2620315 | 0.81619374 | 1 | 0.262031485 | 0.262031485 | 5.192233777 | 0.28694685 | 0.829048563 | 1 | 0.286946854 | 0.286946854 | 5.692254997 | 0.913170789 | -0.039447989 | FALSE | TRUE | FALSE |
| CDYL2 | 0.262018 | 0.81621341 | 1 | 0.262018028 | 0.262018028 | 5.176382073 | 0.30705321 | 0.823086529 | 0.016 | 0.307053212 | 0.307053212 | 6.075000177 | 0.853331012 | -0.068882471 | FALSE | TRUE | FALSE |
| NFE2 | 0.2618999 | 0.81623308 | 1 | 0.26189994 | 0.26189994 | 5.190430562 | 0.30678452 | 0.823221416 | 0.01085 | 0.306784518 | 0.306784518 | 6.086610799 | 0.853693473 | -0.068698039 | FALSE | TRUE | FALSE |
| SFRP1 | 0.2618371 | 0.81625274 | 1 | 0.261837102 | 2.433152752 | 5.194119238 | 0.31611319 | 0.819925028 | 1 | 0.316113194 | 0.316113194 | 0.316113194 | 0.82830172 | -0.081811437 | FALSE | TRUE | FALSE |
| SLA | 0.26176 | 0.8162724 | 1 | 0.26176003 | 0.26176003 | 0.26176003 | 0.30325638 | 0.824274003 | 1 | 1.21274649 | 1.21274649 | 1.21274649 | 0.863164122 | -0.06390662 | FALSE | TRUE | FALSE |
| PHOSPHO2 | 0.2617236 | 0.81629206 | 0.0062 | 0.261723586 | 0.261723586 | 0.261723586 | 0.30603269 | 0.823490557 | 1 | 0.306032689 | 0.306032689 | 0.306032689 | 0.855214478 | -0.067924955 | FALSE | TRUE | FALSE |
| SLC17A9 | 0.2616382 | 0.81631171 | 0.50435 | 0.592527046 | 2.369496023 | 2.369496023 | 658.046641 | 0.003254042 | 0.00405 | 7.02389158 | 658.0466405 | 658.0466405 | 0.000397598 | -3.400555547 | FALSE | FALSE | FALSE |
| ACP1 | 0.2616276 | 0.81633136 | 1 | 0.261627632 | 0.261627632 | 0.261627632 | 1.17228583 | 0.570092795 | 0.75115 | 2.698248224 | 10.79277652 | 10.79277652 | 0.223177339 | -0.651349905 | FALSE | FALSE | FALSE |
| PDSS2 | 0.2615931 | 0.816351 | 1 | 0.261593052 | 0.261593052 | 5.188747895 | 0.30613688 | 0.823452159 | 0.0102 | 0.30613688 | 0.30613688 | 6.078301024 | 0.85449702 | -0.068289448 | FALSE | TRUE | FALSE |
| LHX6 | 0.2615265 | 0.81637064 | 1 | 0.261526521 | 0.261526521 | 0.261526521 | 0.30657214 | 0.823317634 | 0.00635 | 0.306572142 | 0.306572142 | 0.306572142 | 0.853066817 | -0.069016951 | FALSE | TRUE | FALSE |
| CYBRD1 | 0.2614554 | 0.81639027 | 1 | 0.261455381 | 0.261455381 | 0.261455381 | 0.30610162 | 0.82347136 | 1 | 0.306101623 | 0.306101623 | 0.306101623 | 0.854145685 | -0.068468049 | FALSE | TRUE | FALSE |
| OTUD3 | 0.2614028 | 0.8164099 | 1 | 0.2614028 | 0.2614028 | 0.2614028 | 0.30555121 | 0.823682287 | 1 | 0.305551208 | 0.305551208 | 0.305551208 | 0.855512245 | -0.06777377 | FALSE | TRUE | FALSE |
| MEDAG | 0.2613354 | 0.81642953 | 1 | 0.261335389 | 0.261335389 | 0.261335389 | 0.30544018 | 0.823701437 | 1 | 0.305440184 | 0.305440184 | 0.305440184 | 0.855602514 | -0.067727948 | FALSE | TRUE | FALSE |
| GOLGA8A | 0.2612085 | 0.81644915 | 1 | 0.261208493 | 0.261208493 | 5.154367963 | 0.27474071 | 0.832367745 | 1 | 0.274740713 | 0.274740713 | 5.430113104 | 0.950745488 | -0.021935727 | FALSE | TRUE | FALSE |
| ZCCHC7 | 0.2611564 | 0.81646877 | 0.7571 | 0.261156411 | 1.044720414 | 5.172114143 | 0.08886321 | 0.878351279 | 1 | 0.803159126 | 0.803159126 | 1.761973278 | 2.93885869 | 0.468178704 | FALSE | TRUE | FALSE |
| ITPKC | 0.2611071 | 0.81648839 | 0.5118 | 1.044414164 | 1.044414164 | 20.58901603 | 0.30560979 | 0.823663133 | 1 | 1.222424661 | 1.222424661 | 10.34447752 | 0.854380523 | -0.068348661 | FALSE | TRUE | FALSE |
| TRMT12 | 0.2610547 | 0.816508 | 1 | 2.409712115 | 2.409712115 | 5.172272511 | 0.30537063 | 0.823816243 | 1 | 2.82804806 | 2.82804806 | 6.057108367 | 0.854878187 | -0.068095764 | FALSE | TRUE | FALSE |
| EFNA1 | 0.2610373 | 0.8165276 | 1 | 0.26103733 | 0.26103733 | 0.26103733 | 0.30599317 | 0.823528937 | 1 | 0.305993172 | 0.305993172 | 0.305993172 | 0.853082204 | -0.069009118 | FALSE | TRUE | FALSE |
| CCDC91 | 0.2609419 | 0.8165472 | 0.0114 | 0.260941883 | 0.260941883 | 5.183173229 | 0.30508257 | 0.823873589 | 1 | 0.305082571 | 0.305082571 | 0.305082571 | 0.855315602 | -0.067873606 | FALSE | TRUE | FALSE |
| MKRN3 | 0.2609264 | 0.8165668 | 1 | 0.260926422 | 0.260926422 | 5.158671025 | 2.7476483 | 0.431279565 | 0.50875 | 2.747648298 | 2.747648298 | 5.861370884 | 0.094963545 | -1.022443083 | FALSE | FALSE | FALSE |
| TMEM97 | 0.2609096 | 0.81658639 | 1 | 2.414772992 | 2.414772992 | 2.414772992 | 2.8338149 | 0.424371261 | 0.5046 | 2.833814905 | 2.833814905 | 2.833814905 | 0.09207011 | -1.035881339 | FALSE | FALSE | FALSE |
| OR10J1 | 0.260905 | 0.81660598 | 1 | 2.39796147 | 2.39796147 | 2.39796147 | 2.81347043 | 0.425806893 | 0.00745 | 2.813470429 | 2.813470429 | 2.813470429 | 0.092734235 | -1.032759907 | FALSE | FALSE | FALSE |
| FBXO9 | 0.2608421 | 0.81662556 | 1 | 1.04326358 | 1.04326358 | 5.173143534 | 0.30886735 | 0.822331121 | 1 | 0.30886735 | 0.30886735 | 6.131772932 | 0.844511872 | -0.073394241 | FALSE | TRUE | FALSE |
| ADAM2 | 0.260631 | 0.81664514 | 0.32445 | 0.260630983 | 21.00718344 | 21.00718344 | 4.54339471 | 0.326442516 | 0.31145 | 0.50564488 | 41.84809593 | 41.84809593 | 0.057364812 | -1.241354426 | FALSE | FALSE | FALSE |
| GFM2 | 0.2605933 | 0.81666472 | 0.25595 | 0.065179598 | 1.042133655 | 1.042133655 | 0.01625926 | 0.894802358 | 0.26145 | 0.016259259 | 0.065011672 | 0.32126183 | 16.02737952 | 1.204862521 | FALSE | TRUE | FALSE |
| IQGAP3 | 0.2605734 | 0.81668429 | 0.2156 | 0.035028064 | 1.258727915 | 5.033799124 | 0.03762819 | 0.889544113 | 0.883 | 0.327255662 | 26.69718304 | 106.7479252 | 6.924949925 | 0.840416637 | FALSE | TRUE | FALSE |
| C22orf31 | 0.2604712 | 0.81670386 | 1 | 2.392948108 | 22.56328066 | 22.56328066 | 2316.97388 | 0.001013185 | 0.0346 | 233.461468 | 2316.973877 | 23556.17335 | 0.000112419 | -3.949161429 | FALSE | FALSE | FALSE |
| OR4K5 | 0.2604138 | 0.81672342 | 1 | 0.26041377 | 0.26041377 | 0.26041377 | 0.30460212 | 0.823949994 | 0.0061 | 0.304602117 | 0.304602117 | 0.304602117 | 0.854930928 | -0.068068972 | FALSE | TRUE | FALSE |
| HNF4G | 0.2603975 | 0.81674298 | 0.6233 | 0.447931338 | 4.160426609 | 4.160426609 | 0.17432466 | 0.858501114 | 0.8752 | 0.696955271 | 2.787144665 | 2.787144665 | 1.493750074 | 0.17427794 | FALSE | TRUE | FALSE |
| PPP6R2 | 0.2600655 | 0.81676254 | 1 | 0.260065469 | 0.260065469 | 5.086721389 | 0.30468765 | 0.823930898 | 0.0262 | 0.304687649 | 0.304687649 | 5.975319746 | 0.853547786 | -0.06877216 | FALSE | TRUE | FALSE |

| | | | | | | | | | | | | | | | | | |
|---|---|---|---|---|---|---|---|---|---|---|---|---|---|---|---|---|---|
| CPXM2 | 0.2599778 | 0.81678209 | 0.26275 | 0.065029837 | 1.039605127 | 1.27888897 | 8.12546592 | 0.225236428 | 0.3451 | 2.031289369 | 72.56195188 | 290.3313368 | 0.031995433 | -1.494912003 | FALSE | FALSE | FALSE |
| PPP1R3B | 0.2599649 | 0.81680164 | 1 | 0.259964869 | 0.259964869 | 0.259964869 | 0.30110697 | 0.82516347 | 1 | 1.204284311 | 1.204284311 | 1.204284311 | 0.863363823 | -0.063806153 | FALSE | TRUE | FALSE |
| FJX1 | 0.2598771 | 0.81682119 | 0.5061 | 1.03922649 | 1.03922649 | 1.03922649 | 1.2189241 | 0.565363675 | 0.4996 | 0.30480393 | 1.218924098 | 6.054115235 | 0.213202025 | -0.671208675 | FALSE | FALSE | FALSE |
| LRPAP1 | 0.2598393 | 0.81684073 | 1 | 0.259839276 | 0.259839276 | 5.149735148 | 0.30400927 | 0.824083553 | 1 | 0.304009268 | 0.304009268 | 0.304009268 | 0.854708403 | -0.068182026 | FALSE | TRUE | FALSE |
| SLC25A17 | 0.2597553 | 0.81686026 | 0.75245 | 0.595578137 | 2.381288684 | 2.381288684 | 0.36904033 | 0.802511697 | 0.5051 | 0.369040333 | 0.369040333 | 0.369040333 | 0.703866882 | -0.152509469 | FALSE | TRUE | FALSE |
| CD27 | 0.2597479 | 0.8168798 | 1 | 0.25974789 | 0.25974789 | 0.25974789 | 2.76621894 | 0.429466544 | 0.507 | 2.766218945 | 2.766218945 | 2.766218945 | 0.093899975 | -1.027334522 | FALSE | FALSE | FALSE |
| CYP26A1 | 0.2597063 | 0.81689932 | 0.8796 | 2.36605923 | 22.12419725 | 22.12419725 | 0.09607815 | 0.87689174 | 0.0139 | 0.096078147 | 0.096078147 | 1.901106156 | 2.703073492 | 0.431857854 | FALSE | TRUE | FALSE |
| PKNOX2 | 0.2595002 | 0.81691885 | 1 | 0.25950017 | 0.25950017 | 5.139285885 | 0.30452737 | 0.823988173 | 1 | 0.304527367 | 0.304527367 | 6.038118743 | 0.852140722 | -0.06948868 | FALSE | TRUE | FALSE |
| AVL9 | 0.2593841 | 0.81693837 | 1 | 0.259384083 | 0.259384083 | 5.130739384 | 0.30381637 | 0.82414073 | 1 | 0.30381637 | 0.30381637 | 6.017566698 | 0.853752818 | -0.06866785 | FALSE | TRUE | FALSE |
| GEM | 0.2593329 | 0.81695788 | 0.84325 | 4.148429384 | 38.25426399 | 153.0578656 | 0.09650139 | 0.876815048 | 1 | 0.096501388 | 0.096501388 | 0.096501388 | 2.68734909 | 0.429324085 | FALSE | TRUE | FALSE |
| COPB1 | 0.2593039 | 0.8169774 | 0.0252 | 0.259303887 | 0.259303887 | 5.098448957 | 0.3043238 | 0.824007257 | 0.0213 | 0.304323797 | 0.304323797 | 5.995805114 | 0.852065759 | -0.069526887 | FALSE | TRUE | FALSE |
| COMTD1 | 0.259178 | 0.8169969 | 1 | 0.259178041 | 0.259178041 | 0.259178041 | 0.30313981 | 0.824293026 | 0.0063 | 0.303139806 | 0.303139806 | 0.303139806 | 0.854978581 | -0.068044765 | FALSE | TRUE | FALSE |
| HNRNPUL2 | 0.2591397 | 0.81701641 | 1 | 0.259139746 | 0.259139746 | 5.100563047 | 0.30404914 | 0.824045413 | 1 | 0.304049138 | 0.304049138 | 5.995929702 | 0.852295612 | -0.069409748 | FALSE | TRUE | FALSE |
| PRM2 | 0.2591386 | 0.81703591 | 1 | 0.259138643 | 0.259138643 | 0.259138643 | 0.30295435 | 0.824426065 | 1 | 0.302954353 | 0.302954353 | 0.302954353 | 0.855371907 | -0.067845018 | FALSE | TRUE | FALSE |
| SYT17 | 0.2590613 | 0.8170554 | 1 | 0.259061253 | 0.259061253 | 5.120930655 | 0.30340677 | 0.824235944 | 1 | 0.303406767 | 0.303406767 | 6.005916956 | 0.85384138 | -0.068622802 | FALSE | TRUE | FALSE |
| POU2F2 | 0.2590221 | 0.81707489 | 1 | 0.259022055 | 0.259022055 | 5.12573731 | 0.30399187 | 0.824102616 | 1 | 0.303991874 | 0.303991874 | 6.023277999 | 0.852069012 | -0.069525229 | FALSE | TRUE | FALSE |
| CPNE2 | 0.2589409 | 0.81709438 | 1 | 0.258940896 | 0.258940896 | 5.116441417 | 0.30371748 | 0.824178828 | 1 | 0.303717478 | 0.303717478 | 6.009883606 | 0.852571599 | -0.069269139 | FALSE | TRUE | FALSE |
| MED11 | 0.258837 | 0.81711386 | 1 | 0.25883703 | 0.25883703 | 0.25883703 | 0.30263669 | 0.824539931 | 0.00365 | 0.302636685 | 0.302636685 | 0.302636685 | 0.855273146 | -0.067895164 | FALSE | TRUE | FALSE |
| OR10K2 | 0.2586661 | 0.81713334 | 1 | 0.258666105 | 2.379972159 | 2.379972159 | 0.31217374 | 0.821313624 | 1 | 0.312173738 | 0.312173738 | 0.312173738 | 0.828596621 | -0.081656842 | FALSE | TRUE | FALSE |
| MED27 | 0.2585475 | 0.81715282 | 1 | 0.258547515 | 0.258547515 | 0.258547515 | 0.3026203 | 0.824577853 | 1 | 0.302620302 | 0.302620302 | 0.302620302 | 0.854362757 | -0.068357692 | FALSE | TRUE | FALSE |
| DSE | 0.2585287 | 0.81717229 | 1 | 0.258528744 | 0.258528744 | 5.084180813 | 0.30300989 | 0.824388076 | 1 | 0.303009894 | 0.303009894 | 5.970925738 | 0.853202319 | -0.068947973 | FALSE | TRUE | FALSE |
| RBM5 | 0.2584957 | 0.81719176 | 1 | 0.258495715 | 0.258495715 | 5.080590269 | 0.30307493 | 0.824331059 | 1 | 0.303074925 | 0.303074925 | 5.969160861 | 0.852910267 | -0.069096658 | FALSE | TRUE | FALSE |
| EDARADD | 0.2584894 | 0.81721122 | 0.00595 | 0.258489415 | 0.258489415 | 0.258489415 | 0.30308875 | 0.824312044 | 1 | 0.303088751 | 0.303088751 | 0.303088751 | 0.852850572 | -0.069127055 | FALSE | TRUE | FALSE |
| AGBL4 | 0.2584777 | 0.81723068 | 1 | 0.258477744 | 0.258477744 | 0.258477744 | 2.71114973 | 0.434276017 | 0.50585 | 2.711149732 | 2.711149732 | 2.711149732 | 0.095338793 | -1.020730348 | FALSE | FALSE | FALSE |
| BCL2L13 | 0.2583676 | 0.81725014 | 1 | 2.380638157 | 2.380638157 | 5.121207872 | 0.31191468 | 0.821392312 | 1 | 0.311914682 | 0.311914682 | 6.189226089 | 0.828327838 | -0.081797743 | FALSE | TRUE | FALSE |
| SERPINA12 | 0.2583326 | 0.81726959 | 0.5077 | 0.258332645 | 0.258332645 | 0.564961366 | 0.04451491 | 0.88798957 | 0.0092 | 0.044514908 | 0.044514908 | 0.044514908 | 5.803283838 | 0.763673812 | FALSE | TRUE | FALSE |
| SET | 0.2582591 | 0.81728903 | 1 | 0.258259089 | 0.258259089 | 0.258259089 | 0.28152441 | 0.830777236 | 1 | 2.612280547 | 2.612280547 | 2.612280547 | 0.917359502 | -0.037460436 | FALSE | TRUE | FALSE |
| HSPH1 | 0.2582091 | 0.81730847 | 1 | 0.258209091 | 0.258209091 | 5.086098395 | 0.3026652 | 0.824520964 | 1 | 0.302665196 | 0.302665196 | 5.972626734 | 0.85311788 | -0.068990956 | FALSE | TRUE | FALSE |
| CPNE8 | 0.2580063 | 0.81732791 | 1 | 0.258006278 | 0.258006278 | 5.110710385 | 2.73731237 | 0.432282011 | 0.01305 | 2.737312371 | 2.737312371 | 5.816975542 | 0.094255329 | -1.025694086 | FALSE | FALSE | FALSE |
| OR2G3 | 0.258004 | 0.81734735 | 1 | 0.258003962 | 0.258003962 | 0.258003962 | 0.3023005 | 0.824748288 | 1 | 0.302300504 | 0.302300504 | 0.302300504 | 0.853468515 | -0.068812496 | FALSE | TRUE | FALSE |
| ARMC7 | 0.2579321 | 0.81736678 | 1 | 0.257932139 | 0.257932139 | 0.257932139 | 2.62503741 | 0.438821321 | 0.7492 | 2.625037411 | 24.90073503 | 24.90073503 | 0.098258462 | -1.007630038 | FALSE | FALSE | FALSE |
| PNN | 0.2579265 | 0.8173862 | 1 | 0.257926452 | 0.257926452 | 5.083502983 | 0.30236664 | 0.824710443 | 0.01835 | 0.302366644 | 0.302366644 | 5.969811572 | 0.853025484 | -0.069037994 | FALSE | TRUE | FALSE |
| KCTD21 | 0.257907 | 0.81740563 | 1 | 0.257906984 | 0.257906984 | 0.257906984 | 0.30224303 | 0.824786115 | 1 | 0.302243027 | 0.302243027 | 0.302243027 | 0.853309957 | -0.068893187 | FALSE | TRUE | FALSE |
| NVL | 0.2578964 | 0.81742504 | 0.04185 | 0.016133399 | 0.064495786 | 0.317427187 | 0.16789102 | 0.860174433 | 0.1978 | 0.041990751 | 0.167891024 | 0.67144399 | 1.536094093 | 0.186417819 | FALSE | TRUE | FALSE |
| KIF17 | 0.2578401 | 0.81744446 | 0.04905 | 0.01615296 | 0.257840101 | 1.255208789 | 0.00238741 | 0.898908777 | 0.6368 | 0.009541105 | 0.082312212 | 0.401969465 | 107.9999258 | 2.033423457 | FALSE | TRUE | FALSE |
| PDIA3 | 0.2578327 | 0.81746387 | 1 | 0.257832659 | 0.257832659 | 5.10391347 | 0.30254501 | 0.824596807 | 1 | 0.302545006 | 0.302545006 | 5.996382727 | 0.852212576 | -0.069452061 | FALSE | TRUE | FALSE |
| RFPL2 | 0.2577948 | 0.81748327 | 1 | 0.257794795 | 0.257794795 | 5.111344569 | 2.70138415 | 0.434871786 | 0.5112 | 2.701384147 | 2.701384147 | 2.701384147 | 0.095430631 | -1.020312203 | FALSE | FALSE | FALSE |
| STK38 | 0.2577929 | 0.81750267 | 1 | 0.25779287 | 0.25779287 | 5.110579716 | 0.30212243 | 0.824805023 | 1 | 0.302122429 | 0.302122429 | 5.995711013 | 0.853272864 | -0.068912066 | FALSE | TRUE | FALSE |
| UCHL3 | 0.2576635 | 0.81752207 | 1 | 0.25766353 | 0.25766353 | 0.25766353 | 0.29265419 | 0.827210505 | 0.50465 | 2.7085131 | 2.7085131 | 2.7085131 | 0.880436851 | -0.055301788 | FALSE | TRUE | FALSE |
| ARL6 | 0.2576428 | 0.81754146 | 1 | 0.257642817 | 0.257642817 | 0.257642817 | 0.30241721 | 0.824672582 | 1 | 0.302417209 | 0.302417209 | 0.302417209 | 0.85194496 | -0.069588462 | FALSE | TRUE | FALSE |
| PLAUR | 0.2576191 | 0.81756085 | 1 | 0.257619138 | 0.257619138 | 5.110496933 | 0.30190047 | 0.82491838 | 0.00955 | 0.301900467 | 0.301900467 | 0.301900467 | 0.853324741 | -0.068885662 | FALSE | TRUE | FALSE |
| CD300LD | 0.2575572 | 0.81758023 | 1 | 0.257557245 | 0.257557245 | 0.257557245 | 0.30167648 | 0.825031583 | 1 | 0.301676481 | 0.301676481 | 0.301676481 | 0.853753144 | -0.068667684 | FALSE | TRUE | FALSE |
| TMEM88 | 0.2575035 | 0.81759961 | 1 | 0.257503474 | 0.257503474 | 0.257503474 | 0.30148621 | 0.825069285 | 1 | 0.301486208 | 0.301486208 | 0.301486208 | 0.854113612 | -0.068484357 | FALSE | TRUE | FALSE |
| CCDC3 | 0.2574686 | 0.81761899 | 0.9842 | 215.224527 | 21635.69461 | 224793.6894 | 3.2535299 | 0.397559381 | 0.50115 | 3.253529905 | 3.253529905 | 3.253529905 | 0.079135168 | -1.10163047 | FALSE | FALSE | FALSE |
| SH3GL1 | 0.257437 | 0.81763836 | 1 | 0.257436985 | 0.257436985 | 5.094343869 | 2.73317938 | 0.432681913 | 0.50435 | 2.733179376 | 2.733179376 | 5.800430106 | 0.094189568 | -1.025997195 | FALSE | FALSE | FALSE |
| VHLL | 0.2574105 | 0.81765772 | 1 | 2.358549478 | 2.358549478 | 2.358549478 | 0.31055988 | 0.821843407 | 1 | 0.310559884 | 0.310559884 | 0.310559884 | 0.828859421 | -0.081519122 | FALSE | TRUE | FALSE |
| MAP4K1 | 0.2573989 | 0.81767709 | 1 | 0.25739893 | 0.25739893 | 5.050005507 | 25.5023549 | 0.107952352 | 0.26085 | 2.655646005 | 25.50235488 | 52.22151366 | 0.010093144 | -1.995973547 | FALSE | FALSE | FALSE |
| TTPA | 0.2573701 | 0.81769644 | 1 | 1.30472E+17 | 6.137E+23 | 1.35264E+26 | 255.126405 | 0.009813 | 1 | 2.30746E+21 | 2.24587E+28 | 5.43857E+30 | 0.001008794 | -2.996197372 | FALSE | FALSE | FALSE |
| METTL5 | 0.257346 | 0.8177158 | 1 | 0.257345977 | 0.257345977 | 0.257345977 | 0.30179576 | 0.824975001 | 1 | 0.301795758 | 0.301795758 | 0.301795758 | 0.852715688 | -0.069195747 | FALSE | TRUE | FALSE |
| ZNF395 | 0.2573311 | 0.81773515 | 1 | 0.257331086 | 0.257331086 | 5.085701513 | 0.30203503 | 0.824842825 | 1 | 0.302035027 | 0.302035027 | 5.977698316 | 0.851990873 | -0.069565058 | FALSE | TRUE | FALSE |
| JAGN1 | 0.257304 | 0.81775449 | 1 | 0.257303987 | 0.257303987 | 0.257303987 | 0.30180153 | 0.824956131 | 1 | 0.30180153 | 0.30180153 | 0.30180153 | 0.852560245 | -0.069274923 | FALSE | TRUE | FALSE |
| DUSP16 | 0.2571855 | 0.81777383 | 1 | 0.257185507 | 0.257185507 | 5.074410048 | 2.73069766 | 0.432881667 | 0.01525 | 2.730697662 | 2.730697662 | 5.781291751 | 0.094183077 | -1.026027126 | FALSE | FALSE | FALSE |
| CASS4 | 0.2571272 | 0.81779317 | 1 | 0.257127165 | 0.257127165 | 5.056184693 | 0.30159155 | 0.825050436 | 0.02035 | 0.301591549 | 0.301591549 | 5.94252814 | 0.85256754 | -0.069271207 | FALSE | TRUE | FALSE |
| PLCD4 | 0.257055 | 0.81781251 | 0.78665 | 2.239418499 | 80.10377366 | 320.5194612 | 0.27641521 | 0.831968521 | 0.7274 | 0.624271794 | 9.97919744 | 39.91346033 | 0.929959692 | -0.031535875 | FALSE | TRUE | FALSE |
| ARID5A | 0.2570452 | 0.81783183 | 1 | 226.6572237 | 2285.526617 | 23608.92397 | 26.1878085 | 0.104955062 | 0.6589 | 257.8524296 | 26905.93394 | 285593.1721 | 0.009815453 | -2.008089665 | FALSE | FALSE | FALSE |
| ZNF549 | 0.257006 | 0.81785116 | 0.50625 | 0.257006039 | 0.257006039 | 0.991609048 | 0.04518802 | 0.887803618 | 1 | 0.045188019 | 0.045188019 | 0.894767057 | 5.687481869 | 0.754920025 | FALSE | TRUE | FALSE |
| CYB561A3 | 0.2569783 | 0.81787048 | 1 | 0.256978268 | 0.256978268 | 0.256978268 | 0.30104455 | 0.825201115 | 1 | 0.301044553 | 0.301044553 | 0.301044553 | 0.85362205 | -0.068734375 | FALSE | TRUE | FALSE |
| GPD1L | 0.2569322 | 0.8178898 | 1 | 0.256932193 | 0.256932193 | 0.256932193 | 0.30174238 | 0.825012727 | 1 | 0.301742383 | 0.301742383 | 0.301742383 | 0.851495209 | -0.069817791 | FALSE | TRUE | FALSE |
| MAP1LC3B2 | 0.2569221 | 0.81790911 | 1 | 2.402244154 | 23.05264893 | 23.05264893 | 0.2862987 | 0.829246372 | 1 | 0.286298699 | 2.684241786 | 2.684241786 | 0.897391793 | -0.047017907 | FALSE | TRUE | FALSE |
| ZC3H7A | 0.2569121 | 0.81792842 | 1 | 0.256912122 | 0.256912122 | 5.045700947 | 0.30174296 | 0.824993866 | 1 | 0.301742961 | 0.301742961 | 5.93902048 | 0.851427058 | -0.069852552 | FALSE | TRUE | FALSE |
| TNFRSF11A | 0.256903 | 0.81794772 | 1 | 2.405494819 | 2.405494819 | 8.70026541 | 0.31122069 | 0.821588727 | 1 | 0.311220688 | 0.311220688 | 6.156337393 | 0.825468764 | -0.083299356 | FALSE | TRUE | FALSE |
| CMAS | 0.256897 | 0.81796702 | 1 | 0.256897013 | 0.256897013 | 5.096613491 | 0.30131844 | 0.82508813 | 1 | 0.301318441 | 0.301318441 | 5.983678489 | 0.852576474 | -0.069266656 | FALSE | TRUE | FALSE |
| TUFM | 0.256789 | 0.81798631 | 0.7484 | 0.256789024 | 1.026870126 | 5.075697774 | 0.35101188 | 0.808858384 | 0.87605 | 1.403738843 | 5.61512401 | 6.947874549 | 0.731567897 | -0.135745361 | FALSE | TRUE | FALSE |
| C5AR2 | 0.2567727 | 0.8180056 | 1 | 0.256772722 | 0.256772722 | 5.086691301 | 0.30104392 | 0.825219931 | 1 | 0.301043924 | 0.301043924 | 5.970519173 | 0.852941055 | -0.069080981 | FALSE | TRUE | FALSE |
| ANKRD13D | 0.256652 | 0.81802489 | 1 | 0.256651992 | 0.256651992 | 5.070552287 | 0.30107056 | 0.825182295 | 1 | 0.30107056 | 0.30107056 | 5.956821358 | 0.852464593 | -0.06932365 | FALSE | TRUE | FALSE |
| CYTIP | 0.2566231 | 0.81804417 | 1 | 0.256623116 | 0.256623116 | 0.256623116 | 0.30067814 | 0.825313952 | 1 | 0.300678145 | 0.300678145 | 0.300678145 | 0.853481108 | -0.068806087 | FALSE | TRUE | FALSE |
| PLA2G2F | 0.256604 | 0.81806345 | 1 | 5.108470071 | 225.1937306 | 225.1937306 | 2.90770009 | 0.418770885 | 0.25665 | 2.907700094 | 27.92740775 | 27.92740775 | 0.088249834 | -1.054286104 | FALSE | FALSE | FALSE |
| TCP11L1 | 0.2564957 | 0.81808273 | 1 | 0.256495651 | 0.256495651 | 5.081705201 | 1.1893521 | 0.568647303 | 0.50915 | 1.189352099 | 1.189352099 | 5.897717584 | 0.215659981 | -0.666230437 | FALSE | FALSE | FALSE |
| SYCP3 | 0.2564365 | 0.818102 | 1 | 0.256436452 | 0.256436452 | 0.256436452 | 0.30031943 | 0.825445406 | 1 | 0.300319435 | 0.300319435 | 0.300319435 | 0.853878977 | -0.068603679 | FALSE | TRUE | FALSE |
| RNF19B | 0.2564177 | 0.81812127 | 1 | 2.385972513 | 22.78625144 | 22.78625144 | 2.7179765 | 0.434077167 | 0.87645 | 25.99500546 | 254.9981209 | 254.9981209 | 0.094341414 | -1.025297618 | FALSE | FALSE | FALSE |
| CXCR2 | 0.2563587 | 0.81814053 | 0.7522 | 2.38970913 | 22.86290441 | 22.86290441 | 0.3104385 | 0.821882524 | 1 | 2.90274561 | 2.90274561 | 6.165251745 | 0.825795346 | -0.083127569 | FALSE | TRUE | FALSE |
| GPR146 | 0.2563515 | 0.81815979 | 1 | 2.370107417 | 2.370107417 | 5.076118068 | 2.78750544 | 0.428048556 | 0.50585 | 2.787505438 | 2.787505438 | 5.958692303 | 0.091964487 | -1.036379847 | FALSE | FALSE | FALSE |
| VPS54 | 0.2563075 | 0.81817904 | 1 | 0.256307526 | 0.256307526 | 5.045691458 | 0.30083137 | 0.825257551 | 1 | 0.30083137 | 0.30083137 | 5.933380556 | 0.851997336 | -0.069561763 | FALSE | TRUE | FALSE |

| | | | | | | | | | | | | | | | | | |
|---|---|---|---|---|---|---|---|---|---|---|---|---|---|---|---|---|---|
| OR2B3 | 0.2561315 | 0.81819829 | 1 | 4454722.188 | 38504905733 | 4.56804E+11 | 0.33926947 | 0.812292597 | 1 | 310072.4872 | 39208913.72 | 439101311 | 0.754950193 | -0.1220817 | FALSE | TRUE | FALSE |
| GDAP1L1 | 0.256061 | 0.81821754 | 0.50525 | 0.256061028 | 0.256061028 | 1.270480811 | 0.08953511 | 0.878229284 | 0.00935 | 0.089535108 | 0.089535108 | 0.089535108 | 2.859895239 | 0.456350125 | FALSE | TRUE | FALSE |
| OXLD1 | 0.2560492 | 0.81823678 | 1 | 0.256049204 | 0.256049204 | 0.256049204 | 0.28751433 | 0.828850287 | 1 | 2.647555874 | 10.5899774 | 10.5899774 | 0.890561527 | -0.050336071 | FALSE | TRUE | FALSE |
| MGMT | 0.256022 | 0.81825602 | 1 | 0.256021962 | 0.256021962 | 0.256021962 | 2.68528569 | 0.435268655 | 0.50335 | 2.68528569 | 2.68528569 | 2.68528569 | 0.095342541 | -1.020713276 | FALSE | FALSE | FALSE |
| CORO2B | 0.2560102 | 0.81827525 | 0.01545 | 0.256010227 | 0.256010227 | 5.05449038 | 0.30069723 | 0.825295156 | 1 | 0.300697232 | 0.300697232 | 5.945928407 | 0.851388705 | -0.069872116 | FALSE | TRUE | FALSE |
| EDN1 | 0.255995 | 0.81829448 | 1 | 0.255994993 | 0.255994993 | 0.255994993 | 0.30051698 | 0.825389093 | 1 | 0.300516975 | 0.300516975 | 0.300516975 | 0.851848695 | -0.069637538 | FALSE | TRUE | FALSE |
| SLAIN1 | 0.2558492 | 0.81831371 | 1 | 0.255849247 | 0.255849247 | 5.08226599 | 0.30039484 | 0.825426639 | 1 | 0.300394839 | 0.300394839 | 0.300394839 | 0.851709863 | -0.069708323 | FALSE | TRUE | FALSE |
| AKIP1 | 0.2558088 | 0.81833293 | 1 | 0.255808787 | 0.255808787 | 0.255808787 | 0.29963016 | 0.825595406 | 1 | 0.299630158 | 0.299630158 | 0.299630158 | 0.853748466 | -0.068670064 | FALSE | TRUE | FALSE |
| CREB3L1 | 0.2555626 | 0.81835215 | 1 | 0.25556259 | 0.25556259 | 5.061514943 | 0.3002839 | 0.825464169 | 1 | 0.300283903 | 0.300283903 | 5.954198785 | 0.851069895 | -0.070034772 | FALSE | TRUE | FALSE |
| SLC9A3 | 0.2554246 | 0.81837136 | 1 | 3.14874E+23 | 2.44907E+29 | 7.56697E+31 | 0.03214058 | 0.890767687 | 1 | 6.78267E+29 | 1.20591E+37 | 3.52237E+39 | 7.947106884 | 0.900209054 | FALSE | TRUE | FALSE |
| NAA38 | 0.255305 | 0.81839057 | 1 | 0.255304978 | 0.255304978 | 0.255304978 | 2.42413065 | 0.453395992 | 0.93705 | 22.80151127 | 2175.375538 | 2175.375538 | 0.105318159 | -0.977496739 | FALSE | FALSE | FALSE |
| PHF20 | 0.2550134 | 0.81840978 | 1 | 0.255013418 | 0.255013418 | 5.002967184 | 0.29975968 | 0.825539187 | 1 | 0.299759677 | 0.299759677 | 5.894329543 | 0.850726223 | -0.07021018 | FALSE | TRUE | FALSE |
| FOSL1 | 0.2549915 | 0.81842899 | 1 | 0.25499149 | 0.25499149 | 0.25499149 | 0.29981284 | 0.825501686 | 1 | 0.29981284 | 0.29981284 | 0.29981284 | 0.850502234 | -0.070324541 | FALSE | TRUE | FALSE |
| PTX3 | 0.2548905 | 0.81844819 | 1 | 2.348899169 | 2.348899169 | 2.348899169 | 0.3082587 | 0.822603067 | 1 | 0.308258695 | 0.308258695 | 0.308258695 | 0.826872206 | -0.082561606 | FALSE | TRUE | FALSE |
| GHDC | 0.2548504 | 0.81846738 | 0.01055 | 0.254850365 | 0.254850365 | 0.553631114 | 0.03494386 | 0.890147278 | 1 | 0.034943859 | 0.034943859 | 0.693498107 | 7.293137496 | 0.862914401 | FALSE | TRUE | FALSE |
| TMEM59L | 0.2547823 | 0.81848658 | 1 | 2.347648952 | 2.347648952 | 2.347648952 | 0.30815746 | 0.822680605 | 1 | 0.308157458 | 0.308157458 | 0.308157458 | 0.826792498 | -0.082603473 | FALSE | TRUE | FALSE |
| DSEL | 0.2546268 | 0.81850577 | 0.76345 | 1.018323651 | 9.289911277 | 34.27793448 | 0.04635423 | 0.887462564 | 1 | 0.046354232 | 0.046354232 | 0.910583279 | 5.493064098 | 0.739814667 | FALSE | TRUE | FALSE |
| EYA1 | 0.2546022 | 0.81852495 | 0.26235 | 0.063698101 | 0.254602181 | 1.259159104 | 0.10506948 | 0.874700281 | 0.0156 | 0.105069481 | 0.105069481 | 0.88990484 | 2.423179205 | 0.384385533 | FALSE | TRUE | FALSE |
| CERS1 | 0.2544881 | 0.81854413 | 0.0078 | 0.254488071 | 0.254488071 | 0.254488071 | 0.29930791 | 0.825670311 | 0.00575 | 0.299307905 | 0.299307905 | 0.299307905 | 0.850255092 | -0.070450759 | FALSE | TRUE | FALSE |
| FLOT2 | 0.2543875 | 0.81856331 | 0.50555 | 0.254387453 | 0.254387453 | 1.258021661 | 0.35131863 | 0.808700293 | 0.505 | 0.351318632 | 0.351318632 | 1.739662592 | 0.724093258 | -0.140205496 | FALSE | TRUE | FALSE |
| GTPBP10 | 0.2543541 | 0.81858248 | 1 | 2.353143178 | 2.353143178 | 2.353143178 | 0.2985201 | 0.82578255 | 1 | 2.77012052 | 2.77012052 | 2.77012052 | 0.852050326 | -0.069534753 | FALSE | TRUE | FALSE |
| TBC1D19 | 0.2543247 | 0.81860165 | 1 | 0.254324738 | 0.254324738 | 5.041127522 | 24.5659033 | 0.112451293 | 0.0076 | 2.59125104 | 24.56590326 | 24.56590326 | 0.010352753 | -1.984944132 | FALSE | FALSE | FALSE |
| UTP23 | 0.2542916 | 0.81862082 | 0.00705 | 0.254291612 | 0.254291612 | 0.254291612 | 0.2890478 | 0.828361645 | 0.50195 | 2.661287111 | 2.661287111 | 2.661287111 | 0.87975627 | -0.05563763 | FALSE | TRUE | FALSE |
| MED22 | 0.2542881 | 0.81863998 | 1 | 0.254288118 | 0.254288118 | 0.254288118 | 0.29824342 | 0.825857302 | 1 | 0.298243424 | 0.298243424 | 0.298243424 | 0.852619362 | -0.069244809 | FALSE | TRUE | FALSE |
| SLC35F3 | 0.254284 | 0.81865913 | 1 | 1.017085286 | 1.017085286 | 8.613308591 | 0.30190538 | 0.824899498 | 1 | 0.301905381 | 0.301905381 | 5.971551692 | 0.842264012 | -0.074551755 | FALSE | TRUE | FALSE |
| LCMT1 | 0.2542813 | 0.81867829 | 0.01085 | 0.254281341 | 0.254281341 | 5.049589363 | 0.29820273 | 0.825875981 | 1 | 0.29820273 | 0.29820273 | 0.29820273 | 0.852712988 | -0.069197122 | FALSE | TRUE | FALSE |
| SRP68 | 0.2542789 | 0.81869743 | 1 | 0.254278863 | 0.254278863 | 5.014616276 | 0.29230108 | 0.827394267 | 1 | 1.169040508 | 4.676675755 | 23.093283 | 0.86992105 | -0.06052016 | FALSE | TRUE | FALSE |
| SERPINB12 | 0.2542787 | 0.81871658 | 0.5049 | 0.254278659 | 2.303473213 | 2.303473213 | 0.48465093 | 0.76124206 | 0.6902 | 0.484650933 | 41.60307975 | 41.60307975 | 0.524663508 | -0.280119141 | FALSE | TRUE | FALSE |
| MUC1 | 0.2542716 | 0.81873572 | 1 | 0.254271554 | 0.254271554 | 0.254271554 | 0.29810512 | 0.82593199 | 1 | 0.298105119 | 0.298105119 | 0.298105119 | 0.852959367 | -0.069071657 | FALSE | TRUE | FALSE |
| KIFC1 | 0.2541861 | 0.81875485 | 0.51475 | 0.254186069 | 0.254186069 | 5.001724659 | 0.08873115 | 0.87840751 | 0.0197 | 0.088731149 | 0.088731149 | 1.749389642 | 2.864676867 | 0.457075641 | FALSE | TRUE | FALSE |
| ZC2HC1C | 0.2541431 | 0.81877398 | 0.50475 | 0.254143137 | 0.254143137 | 1.259506042 | 0.08810873 | 0.878585245 | 1 | 0.088108726 | 0.088108726 | 1.748413557 | 2.884426421 | 0.460059465 | FALSE | TRUE | FALSE |
| ZNF597 | 0.2541347 | 0.81879311 | 1 | 2.37563485 | 2.37563485 | 5.041882404 | 0.30819759 | 0.822622458 | 1 | 0.308197595 | 0.308197595 | 6.120361971 | 0.824583761 | -0.083765222 | FALSE | TRUE | FALSE |
| KCND3 | 0.2539016 | 0.81881223 | 0.94015 | 5.776342819 | 369.0257688 | 1475.961183 | 1.76639394 | 0.511493648 | 0.51165 | 1.766393938 | 7.064044106 | 28.25710849 | 0.143740063 | -0.84242217 | FALSE | FALSE | FALSE |
| NINJ2 | 0.2538277 | 0.81883135 | 1 | 2.358945559 | 2.358945559 | 2.358945559 | 1.84868625 | 0.504268244 | 0.49625 | 0.462173581 | 1.848686253 | 1.848686253 | 0.13730168 | -0.862324149 | FALSE | FALSE | FALSE |
| GGPS1 | 0.2537567 | 0.81885047 | 1 | 0.253756684 | 0.253756684 | 0.253756684 | 0.29817593 | 0.825894655 | 0.0074 | 0.29817593 | 0.29817593 | 0.29817593 | 0.851030077 | -0.070055091 | FALSE | TRUE | FALSE |
| MZF1 | 0.2537431 | 0.81886958 | 0.97775 | 79.89520315 | 16774.27773 | 67099.64801 | 1.01475976 | 0.586666183 | 0.93705 | 85.99791934 | 8028.206929 | 16123.07553 | 0.250052404 | -0.601968965 | FALSE | FALSE | FALSE |
| CDPF1 | 0.253677 | 0.81888869 | 1 | 0.253677045 | 0.253677045 | 0.253677045 | 0.29736634 | 0.826081169 | 1 | 0.297366337 | 0.297366337 | 0.297366337 | 0.853079225 | -0.069010634 | FALSE | TRUE | FALSE |
| TMEM117 | 0.2536287 | 0.81890779 | 1 | 0.253628671 | 0.253628671 | 5.022673051 | 0.28861383 | 0.828542952 | 1 | 2.667509196 | 2.667509196 | 5.722256129 | 0.878782117 | -0.056118789 | FALSE | TRUE | FALSE |
| MAP4K5 | 0.2535722 | 0.81892689 | 1 | 0.25357215 | 0.25357215 | 5.01416673 | 0.29808419 | 0.825950652 | 1 | 0.298084194 | 0.298084194 | 5.90236224 | 0.850672914 | -0.070237395 | FALSE | TRUE | FALSE |
| FCER1A | 0.2533872 | 0.81894598 | 0.00545 | 0.253387248 | 0.253387248 | 0.253387248 | 0.2970157 | 0.826174285 | 1 | 0.297015703 | 0.297015703 | 0.297015703 | 0.853110613 | -0.068994655 | FALSE | TRUE | FALSE |
| KCNIP3 | 0.2533418 | 0.81896508 | 0.0084 | 0.253341842 | 0.253341842 | 0.253341842 | 0.27979473 | 0.831146882 | 1 | 2.61788367 | 25.13900639 | 25.13900639 | 0.905456081 | -0.04313261 | FALSE | TRUE | FALSE |
| ARHGAP35 | 0.2533129 | 0.81898416 | 1 | 0.253312883 | 0.253312883 | 8.513751472 | 0.2980684 | 0.825969309 | 1 | 0.2980684 | 0.2980684 | 5.798350223 | 0.849848165 | -0.070658659 | FALSE | TRUE | FALSE |
| ZC3H12C | 0.2532821 | 0.81900325 | 1 | 0.253282056 | 0.253282056 | 4.977271071 | 0.2978536 | 0.826043897 | 1 | 0.2978536 | 0.2978536 | 5.865444445 | 0.850357546 | -0.07039843 | FALSE | TRUE | FALSE |
| OR4K17 | 0.2531841 | 0.81902233 | 1 | 0.253184122 | 0.253184122 | 0.253184122 | 0.29718639 | 0.826118427 | 1 | 0.297186386 | 0.297186386 | 0.297186386 | 0.851937148 | -0.069592444 | FALSE | TRUE | FALSE |
| CREG1 | 0.2531199 | 0.8190414 | 1 | 0.253119907 | 2.331044411 | 2.331044411 | 0.296987 | 0.826192896 | 1 | 0.296987004 | 2.744740983 | 2.744740983 | 0.85229284 | -0.06941116 | FALSE | TRUE | FALSE |
| ANKRD39 | 0.253046 | 0.81906048 | 0.50695 | 2.351205975 | 2.351205975 | 2.351205975 | 0.30670979 | 0.823259916 | 1 | 0.306709788 | 0.306709788 | 0.306709788 | 0.825034099 | -0.083528101 | FALSE | TRUE | FALSE |
| BCL6B | 0.2530308 | 0.81907954 | 1 | 0.253030775 | 0.253030775 | 4.990400782 | 0.29733483 | 0.8260998 | 1 | 0.297334827 | 0.297334827 | 5.873979438 | 0.850996091 | -0.070072435 | FALSE | TRUE | FALSE |
| RCN1 | 0.2528337 | 0.81909861 | 1 | 0.252833667 | 0.252833667 | 0.252833667 | 1.16205779 | 0.571290471 | 0.75315 | 1.162057791 | 4.648670594 | 4.648670594 | 0.217574091 | -0.662392823 | FALSE | FALSE | FALSE |
| ADCK5 | 0.2527624 | 0.81911767 | 0.01535 | 0.252762377 | 0.252762377 | 4.992608545 | 0.2970779 | 0.82615567 | 1 | 0.297077897 | 0.297077897 | 5.876685374 | 0.85082862 | -0.07015791 | FALSE | TRUE | FALSE |
| TMEM177 | 0.2526311 | 0.81913673 | 1 | 2.321672748 | 21.89829918 | 21.89829918 | 0.30561325 | 0.823643975 | 1 | 2.821740888 | 2.821740888 | 2.821740888 | 0.826636736 | -0.082685298 | FALSE | TRUE | FALSE |
| DNAJC5G | 0.2526094 | 0.81915578 | 1 | 0.252609449 | 0.252609449 | 0.252609449 | 1.10308833 | 0.578119793 | 0.7491 | 10.22149121 | 97.2095246 | 97.2095246 | 0.229002013 | -0.640160699 | FALSE | FALSE | FALSE |
| PGPEP1L | 0.2523154 | 0.81917483 | 1 | 2.321207011 | 2.321207011 | 2.321207011 | 0.30538287 | 0.823777993 | 1 | 0.305382872 | 0.305382872 | 0.305382872 | 0.826226292 | -0.082900989 | FALSE | TRUE | FALSE |
| TXNRD1 | 0.2523017 | 0.81919388 | 1 | 22.08177937 | 214.6571124 | 214.6571124 | 0.32150357 | 0.818270342 | 1 | 1.28598352 | 1.28598352 | 6.372110556 | 0.784755416 | -0.105265678 | FALSE | TRUE | FALSE |
| KAT2A | 0.2523007 | 0.81921292 | 1 | 0.252300663 | 0.252300663 | 4.952152123 | 0.29660215 | 0.826285892 | 1 | 0.29660215 | 0.29660215 | 5.834740344 | 0.850636662 | -0.070255903 | FALSE | TRUE | FALSE |
| PILRA | 0.252194 | 0.81923196 | 1 | 0.252193988 | 0.252193988 | 0.252193988 | 0.29600475 | 0.826415928 | 1 | 0.296004754 | 0.296004754 | 0.296004754 | 0.851993034 | -0.069563956 | FALSE | TRUE | FALSE |
| GALR1 | 0.2521537 | 0.81925099 | 0.0115 | 0.252153696 | 0.252153696 | 4.995668429 | 0.29653415 | 0.82630448 | 1 | 0.296534151 | 0.296534151 | 5.881576395 | 0.850336108 | -0.070409379 | FALSE | TRUE | FALSE |
| KPTN | 0.2521079 | 0.81927002 | 1 | 2.326338064 | 2.326338064 | 4.99099861 | 0.30540131 | 0.823739724 | 1 | 0.305401312 | 0.305401312 | 6.053430966 | 0.82549719 | -0.083284401 | FALSE | TRUE | FALSE |
| ROPN1B | 0.2520955 | 0.81928905 | 1 | 2.325709204 | 2.325709204 | 2.325709204 | 0.30533239 | 0.823835362 | 0.0054 | 0.305332386 | 0.305332386 | 0.305332386 | 0.825642997 | -0.083207698 | FALSE | TRUE | FALSE |
| ALDH1A1 | 0.2520123 | 0.81930807 | 1 | 0.252012255 | 0.252012255 | 4.993380922 | 18.362691 | 0.141953142 | 0.0107 | 4.589488427 | 18.36269097 | 22.75931709 | 0.013724146 | -1.862514666 | FALSE | FALSE | FALSE |
| EPHX1 | 0.2519867 | 0.81932709 | 0.25915 | 0.251986709 | 1.008049454 | 1.247589562 | 0.77526019 | 0.630226615 | 0.5055 | 0.775260191 | 0.775260191 | 1.705719484 | 0.325035016 | -0.48806985 | FALSE | TRUE | FALSE |
| CEL | 0.2519356 | 0.8193461 | 0.32725 | 0.251935617 | 20.28839751 | 20.28839751 | 0.05540419 | 0.885552824 | 0.81285 | 1.939979485 | 40.11991353 | 160.4292377 | 4.547230304 | 0.657746951 | FALSE | TRUE | FALSE |
| KRT23 | 0.251787 | 0.81936511 | 0.98165 | 64.37174502 | 5659.506978 | 22644.63302 | 0.01352681 | 0.895610991 | 0.9983 | 13.82733142 | 221.4700215 | 886.677825 | 18.61392688 | 1.269838004 | FALSE | TRUE | FALSE |
| ZBTB9 | 0.251748 | 0.81938412 | 0.5005 | 1.006960724 | 1.006960724 | 4.98308013 | 0.29910347 | 0.825726447 | 1 | 0.299103465 | 0.299103465 | 5.927767829 | 0.841675466 | -0.074855332 | FALSE | TRUE | FALSE |
| OR4C12 | 0.2516492 | 0.81940312 | 1 | 0.251649212 | 0.251649212 | 0.251649212 | 0.29535285 | 0.826619873 | 1 | 0.295352851 | 0.295352851 | 0.295352851 | 0.852029059 | -0.069545593 | FALSE | TRUE | FALSE |
| ZNF675 | 0.251616 | 0.81942212 | 0.8734 | 2.268395014 | 20.97616477 | 20.97616477 | 0.35881566 | 0.806087342 | 0.969 | 30.33708761 | 289.5910837 | 2831.962708 | 0.701240323 | -0.154133119 | FALSE | TRUE | FALSE |
| ZNF609 | 0.2515763 | 0.81944111 | 1 | 2.364853024 | 22.81519974 | 443.5112546 | 0.31547284 | 0.82006488 | 1 | 0.315472836 | 0.315472836 | 6.153293804 | 0.797457906 | -0.098292232 | FALSE | TRUE | FALSE |
| ADAMTSL2 | 0.2515574 | 0.8194601 | 1 | 0.25155739 | 0.25155739 | 4.936924613 | 0.29621648 | 0.826323065 | 1 | 0.296216477 | 0.296216477 | 5.826488852 | 0.849234965 | -0.070972133 | FALSE | TRUE | FALSE |
| FNIP2 | 0.2515569 | 0.81947909 | 1 | 2.34569619 | 2.34569619 | 45.94537321 | 2.76585198 | 0.429668577 | 0.51825 | 2.765851978 | 2.765851978 | 54.30971509 | 0.090950951 | -1.041192754 | FALSE | FALSE | FALSE |
| GGNBP2 | 0.2514475 | 0.81949807 | 1 | 22.76020145 | 225.0007777 | 225.0007777 | 25.3020158 | 0.10855282 | 0.5072 | 25.30201578 | 250.4583926 | 250.4583926 | 0.009937846 | -2.002707758 | FALSE | FALSE | FALSE |
| CLEC6A | 0.2514286 | 0.81951705 | 0.0049 | 0.251428638 | 0.251428638 | 0.251428638 | 0.26918424 | 0.834047818 | 1 | 23.37777968 | 226.3780603 | 226.3780603 | 0.934039227 | -0.029634884 | FALSE | TRUE | FALSE |
| CKB | 0.2514112 | 0.81953602 | 1 | 2.344278874 | 22.43485132 | 46.39295171 | 2.76390031 | 0.430273662 | 0.75495 | 2.763900313 | 26.50709059 | 26.78968304 | 0.090962481 | -1.041137704 | FALSE | FALSE | FALSE |
| TET3 | 0.2512817 | 0.81955499 | 0.0469 | 0.251281702 | 0.251281702 | 8.446360987 | 0.29582282 | 0.826471598 | 1 | 0.295822819 | 0.295822819 | 9.957139307 | 0.849433126 | -0.070870807 | FALSE | TRUE | FALSE |
| IL2RB | 0.2512472 | 0.81957396 | 1 | 0.251247207 | 0.251247207 | 4.970714251 | 0.29589815 | 0.826434489 | 1 | 0.295898155 | 0.295898155 | 5.861696007 | 0.849100281 | -0.071041015 | FALSE | TRUE | FALSE |

| | | | | | | | | | | | | | | | | | |
|---|---|---|---|---|---|---|---|---|---|---|---|---|---|---|---|---|---|
| MITF | 0.2512193 | 0.81959292 | 1 | 0.251219302 | 0.251219302 | 4.965191171 | 0.29583069 | 0.826453046 | 1 | 0.295830686 | 0.295830686 | 5.855194987 | 0.849199608 | -0.070990215 | FALSE | TRUE | FALSE |
| ALG8 | 0.2511576 | 0.81961188 | 0.9384 | 5.167399551 | 330.6058285 | 1218.1772 | 8.60757746 | 0.217195562 | 0.3183 | 2.153017577 | 81.59921216 | 81.59921216 | 0.029178669 | -1.53493453 | FALSE | FALSE | FALSE |
| TLE4 | 0.2511549 | 0.81963083 | 1 | 0.251154867 | 0.251154867 | 4.938699101 | 0.29572058 | 0.826508691 | 1 | 0.295720579 | 0.295720579 | 5.826799868 | 0.849297898 | -0.070939951 | FALSE | TRUE | FALSE |
| MCRS1 | 0.2511462 | 0.81964978 | 1 | 0.251146246 | 0.251146246 | 4.971582542 | 0.29567437 | 0.826527231 | 1 | 0.295674375 | 0.295674375 | 5.860238927 | 0.84940146 | -0.070886997 | FALSE | TRUE | FALSE |
| OR6Y1 | 0.2511425 | 0.81966872 | 1 | 2.312733745 | 2.312733745 | 2.312733745 | 0.29497801 | 0.826712416 | 1 | 2.726068649 | 2.726068649 | 2.726068649 | 0.851394068 | -0.06986938 | FALSE | TRUE | FALSE |
| RTP1 | 0.2510967 | 0.81968767 | 1 | 0.251096701 | 2.307401792 | 2.307401792 | 0.24522028 | 0.840603586 | 0.99225 | 2025.827419 | 213341.8917 | 213341.8917 | 1.023963866 | 0.010284631 | FALSE | TRUE | FALSE |
| AGPAT4 | 0.2510928 | 0.8197066 | 0.5074 | 1.004115435 | 1.004115435 | 4.975746575 | 1.18261712 | 0.568888792 | 0.0117 | 1.182617123 | 1.182617123 | 5.866581149 | 0.212319598 | -0.673009918 | FALSE | FALSE | FALSE |
| RASL11B | 0.2509857 | 0.81972554 | 1 | 0.250985706 | 0.250985706 | 0.250985706 | 0.2955493 | 0.8265643 | 1 | 0.295549304 | 0.295549304 | 0.295549304 | 0.849217719 | -0.070980953 | FALSE | TRUE | FALSE |
| TLE1 | 0.2509849 | 0.81974446 | 1 | 0.250984928 | 0.250984928 | 4.924965881 | 0.2954833 | 0.826601353 | 1 | 0.295483299 | 0.295483299 | 5.81131418 | 0.849404786 | -0.070885296 | FALSE | TRUE | FALSE |
| TSNAX | 0.2507559 | 0.81976339 | 1 | 0.250755896 | 0.250755896 | 0.250755896 | 0.29482856 | 0.826749407 | 0.00605 | 0.294828559 | 0.294828559 | 0.294828559 | 0.850514268 | -0.070318396 | FALSE | TRUE | FALSE |
| CEACAM7 | 0.250754 | 0.81978231 | 1 | 0.250753974 | 0.250753974 | 0.250753974 | 0.29456223 | 0.826804864 | 1 | 0.294562227 | 0.294562227 | 0.294562227 | 0.851276747 | -0.069929229 | FALSE | TRUE | FALSE |
| OR52D1 | 0.2507372 | 0.81980123 | 0.00905 | 0.250737194 | 0.250737194 | 0.250737194 | 0.29482359 | 0.826767897 | 1 | 0.294823587 | 0.294823587 | 0.294823587 | 0.850465177 | -0.070343464 | FALSE | TRUE | FALSE |
| UBE3A | 0.2507155 | 0.81982014 | 0.99815 | 7422.131518 | 2610971.13 | 28841880.62 | 0.35608138 | 0.807058468 | 1 | 32.27418162 | 319.2644066 | 636.7538687 | 0.70409594 | -0.15236816 | FALSE | TRUE | FALSE |
| MED12L | 0.2505545 | 0.81983905 | 0.8826 | 2.382554344 | 23.23751522 | 79.52713393 | 0.04269099 | 0.888391899 | 1 | 0.397026799 | 3.78959406 | 73.02094115 | 5.869025621 | 0.768566005 | FALSE | TRUE | FALSE |
| CLPTM1L | 0.2505159 | 0.81985796 | 1 | 0.250515896 | 0.250515896 | 4.946266059 | 0.28579457 | 0.829354078 | 1 | 2.665241728 | 2.665241728 | 5.651499812 | 0.876559323 | -0.057218687 | FALSE | TRUE | FALSE |
| NHEJ1 | 0.2504218 | 0.81987686 | 1 | 0.250421813 | 0.250421813 | 0.250421813 | 1.1657548 | 0.570931804 | 0.50615 | 1.165754796 | 1.165754796 | 1.165754796 | 0.214815169 | -0.667935056 | FALSE | FALSE | FALSE |
| TXNDC5 | 0.2503913 | 0.81989576 | 0.5016 | 0.250391281 | 0.250391281 | 0.57044611 | 0.17021783 | 0.859624999 | 0.7509 | 0.374162614 | 1.496525452 | 1.496525452 | 1.471005054 | 0.167614165 | FALSE | TRUE | FALSE |
| DHX8 | 0.2503299 | 0.81991465 | 0.5197 | 0.250329922 | 0.250329922 | 4.87473883 | 3.37587819 | 0.390338931 | 0.26205 | 0.369041798 | 3.375878189 | 12.42370637 | 0.074152534 | -1.129874003 | FALSE | FALSE | FALSE |
| DTD2 | 0.2503211 | 0.81993354 | 1 | 0.250321101 | 0.250321101 | 0.250321101 | 0.29400592 | 0.826915678 | 0.00365 | 0.294005916 | 0.294005916 | 0.294005916 | 0.851415184 | -0.069858609 | FALSE | TRUE | FALSE |
| GPRC5B | 0.2502782 | 0.81995243 | 0.7529 | 0.250278189 | 1.000824244 | 2.122160599 | 0.08874734 | 0.878388772 | 1 | 0.088747345 | 0.088747345 | 1.758158244 | 2.820120303 | 0.450267635 | FALSE | TRUE | FALSE |
| ASB18 | 0.2502761 | 0.81997131 | 1 | 21.94738387 | 213.5590917 | 213.5590917 | 2235.73394 | 0.001062856 | 0.42415 | 896.1661245 | 91406.0741 | 956611.497 | 0.000111944 | -3.951000674 | FALSE | FALSE | FALSE |
| ZNF566 | 0.2502544 | 0.81999019 | 1 | 0.250254374 | 0.250254374 | 0.250254374 | 0.29428552 | 0.826841817 | 1 | 0.294285523 | 0.294285523 | 0.294285523 | 0.850379494 | -0.070387221 | FALSE | TRUE | FALSE |
| HYAL2 | 0.25022 | 0.82000906 | 1 | 0.250220033 | 0.250220033 | 4.938476969 | 0.29446829 | 0.826823342 | 1 | 0.294468291 | 0.294468291 | 5.820997142 | 0.849735066 | -0.070716459 | FALSE | TRUE | FALSE |
| PPCDC | 0.2502159 | 0.82002793 | 1 | 0.250215888 | 0.250215888 | 0.250215888 | 1.16463209 | 0.571170963 | 0.00495 | 0.291210311 | 1.164632086 | 1.164632086 | 0.214845436 | -0.667873868 | FALSE | FALSE | FALSE |
| SRCIN1 | 0.2501999 | 0.8200468 | 1 | 0.2501999 | 0.2501999 | 4.881748707 | 0.29482886 | 0.826730914 | 1 | 0.294828857 | 0.294828857 | 5.76945619 | 0.848627581 | -0.071282857 | FALSE | TRUE | FALSE |
| GLCE | 0.2498777 | 0.82006566 | 1 | 2.309061775 | 2.309061775 | 4.94253887 | 0.29387248 | 0.826952584 | 1 | 2.724201585 | 2.724201585 | 5.820445613 | 0.850293071 | -0.07043136 | FALSE | TRUE | FALSE |
| CALD1 | 0.2497104 | 0.82008452 | 1 | 0.249710372 | 0.249710372 | 4.917278279 | 0.29413232 | 0.826897218 | 1 | 0.294132323 | 0.294132323 | 5.802771058 | 0.848972905 | -0.07110617 | FALSE | TRUE | FALSE |
| PIGH | 0.2497097 | 0.82010337 | 1 | 0.249709738 | 0.249709738 | 0.249709738 | 2.45199719 | 0.45151831 | 0.49715 | 0.262254778 | 2.451997192 | 2.451997192 | 0.101839325 | -0.99208449 | FALSE | FALSE | FALSE |
| TMX2 | 0.2497067 | 0.82012222 | 1 | 0.249706712 | 0.249706712 | 0.249706712 | 0.29347809 | 0.827063215 | 0.00805 | 0.293478094 | 0.293478094 | 0.293478094 | 0.850852982 | -0.070145475 | FALSE | TRUE | FALSE |
| TNRC6C | 0.2496633 | 0.82014107 | 1 | 2.345226809 | 22.6100232 | 438.6451632 | 0.24370853 | 0.840932103 | 1 | 2175.462523 | 238988.9446 | 4653257.421 | 1.024433959 | 0.010483967 | FALSE | TRUE | FALSE |
| OR4X2 | 0.2496608 | 0.82015991 | 0.0065 | 0.249660796 | 0.249660796 | 0.249660796 | 0.29353018 | 0.827026354 | 1 | 0.293530179 | 0.293530179 | 0.293530179 | 0.85054558 | -0.070302408 | FALSE | TRUE | FALSE |
| NUP160 | 0.2496576 | 0.82017875 | 0.7594 | 0.249657645 | 2.27815845 | 8.383417676 | 0.38008326 | 0.799298855 | 0.76325 | 0.380083263 | 3.494240023 | 12.78205091 | 0.656849877 | -0.182533877 | FALSE | TRUE | FALSE |
| TTC5 | 0.2496103 | 0.82019759 | 1 | 0.998341605 | 3.993965695 | 4.950797253 | 0.2970904 | 0.826137051 | 1 | 1.1882613 | 1.1882613 | 5.898399961 | 0.840183112 | -0.075626052 | FALSE | TRUE | FALSE |
| CPSF7 | 0.2495396 | 0.82021642 | 0.01475 | 0.249539641 | 0.249539641 | 4.934595035 | 0.29418771 | 0.826860288 | 1 | 0.294187714 | 0.294187714 | 5.825382424 | 0.848232708 | -0.071484985 | FALSE | TRUE | FALSE |
| NELFB | 0.2495022 | 0.82023524 | 1 | 0.24950216 | 0.24950216 | 4.92356955 | 0.2935538 | 0.827007918 | 1 | 0.293553796 | 0.293553796 | 5.802152008 | 0.849936753 | -0.070613391 | FALSE | TRUE | FALSE |
| LBR | 0.2494329 | 0.82025407 | 0.125 | 0.062435378 | 0.996747395 | 3.984051907 | 2.21553175 | 0.470807692 | 0.3667 | 0.554304176 | 8.857603996 | 35.42117547 | 0.112583749 | -0.948524292 | FALSE | FALSE | FALSE |
| ZIM2 | 0.2493282 | 0.82027289 | 1 | 0.249328179 | 0.249328179 | 4.931464993 | 0.29328578 | 0.827118476 | 1 | 0.293285775 | 0.293285775 | 5.808613884 | 0.85012026 | -0.070519634 | FALSE | TRUE | FALSE |
| ING3 | 0.2493113 | 0.8202917 | 1 | 0.249311283 | 0.249311283 | 4.949268869 | 0.29398838 | 0.826934133 | 1 | 0.293988379 | 0.293988379 | 0.293988379 | 0.848031083 | -0.071588229 | FALSE | TRUE | FALSE |
| RLN2 | 0.2492203 | 0.82031051 | 1 | 0.249220334 | 0.249220334 | 0.249220334 | 2.60930924 | 0.439996768 | 0.5017 | 2.609309243 | 2.609309243 | 2.609309243 | 0.095511996 | -1.019942079 | FALSE | FALSE | FALSE |
| NUP43 | 0.2490913 | 0.82032932 | 0.00905 | 0.249091348 | 0.249091348 | 0.249091348 | 2.64689851 | 0.437444019 | 0.50325 | 2.646898514 | 2.646898514 | 2.646898514 | 0.094106875 | -1.026378648 | FALSE | FALSE | FALSE |
| SPP1 | 0.2490655 | 0.82034812 | 1 | 0.249065536 | 0.249065536 | 0.249065536 | 0.28380467 | 0.829926239 | 1 | 2.61299103 | 2.61299103 | 2.61299103 | 0.877594926 | -0.056705896 | FALSE | TRUE | FALSE |
| RCOR2 | 0.2490467 | 0.82036692 | 0.01265 | 0.249046706 | 0.249046706 | 4.931388363 | 0.28282266 | 0.830281871 | 1 | 0.282822657 | 0.282822657 | 5.606888651 | 0.880575514 | -0.055233395 | FALSE | TRUE | FALSE |
| SLC39A7 | 0.2490306 | 0.82038572 | 1 | 0.249030561 | 0.249030561 | 4.933028939 | 0.29304725 | 0.827155298 | 0.01015 | 0.293047255 | 0.293047255 | 5.811623128 | 0.849796601 | -0.07068501 | FALSE | TRUE | FALSE |
| CPPED1 | 0.2490216 | 0.82040451 | 0.50105 | 0.249021598 | 0.995592851 | 0.995592851 | 0.16234842 | 0.861512268 | 0.96945 | 2.597193036 | 23.53018951 | 94.14912457 | 1.53387144 | 0.185788961 | FALSE | TRUE | FALSE |
| STX17 | 0.2489138 | 0.8204233 | 0.00745 | 0.248913807 | 0.248913807 | 0.248913807 | 0.29244644 | 0.827284058 | 1 | 0.292446439 | 0.292446439 | 0.292446439 | 0.851143231 | -0.069997351 | FALSE | TRUE | FALSE |
| FGL1 | 0.2488611 | 0.82044208 | 0.00785 | 0.06222877 | 0.248861073 | 0.248861073 | 1.53869844 | 0.532192681 | 0.6614 | 1.538698441 | 24.64123455 | 98.64603391 | 0.161734792 | -0.791196544 | FALSE | FALSE | FALSE |
| TNPO1 | 0.2487623 | 0.82046086 | 1 | 0.248762294 | 0.248762294 | 4.89270256 | 0.29348636 | 0.827044787 | 1 | 0.293486363 | 0.293486363 | 5.783872738 | 0.847611083 | -0.071803373 | FALSE | TRUE | FALSE |
| TREM2 | 0.2487569 | 0.82047964 | 1 | 0.248756899 | 0.248756899 | 0.248756899 | 0.29244133 | 0.827302436 | 0.00535 | 0.292441332 | 0.292441332 | 0.292441332 | 0.850621551 | -0.070263619 | FALSE | TRUE | FALSE |
| ANKS6 | 0.2487273 | 0.82049841 | 0.5081 | 0.248727263 | 0.248727263 | 4.884743184 | 0.04317143 | 0.888271474 | 0.0203 | 0.043171431 | 0.043171431 | 0.849684606 | 5.761385618 | 0.760526944 | FALSE | TRUE | FALSE |
| RRP1 | 0.2486512 | 0.82051718 | 0.50385 | 2.306181798 | 2.306181798 | 4.926831713 | 0.30197428 | 0.82486172 | 1 | 0.301974282 | 0.301974282 | 5.990072564 | 0.823418408 | -0.084379428 | FALSE | TRUE | FALSE |
| SPATS1 | 0.2486095 | 0.82053594 | 1 | 0.248609509 | 0.248609509 | 0.248609509 | 0.29224177 | 0.82743097 | 1 | 0.292241775 | 0.292241775 | 0.292241775 | 0.850698053 | -0.070224561 | FALSE | TRUE | FALSE |
| FGD2 | 0.2485657 | 0.8205547 | 0.82545 | 0.994272543 | 3.978122829 | 15.9569238 | 0.02667041 | 0.892012509 | 0.0191 | 0.026670407 | 0.026670407 | 0.525547391 | 9.319906913 | 0.969411575 | FALSE | TRUE | FALSE |
| BICD2 | 0.2484598 | 0.82057346 | 1 | 0.248459848 | 0.248459848 | 4.846237467 | 0.29278785 | 0.827192106 | 1 | 0.292787854 | 0.292787854 | 5.727888656 | 0.848600258 | -0.07129684 | FALSE | TRUE | FALSE |
| MPHOSPH6 | 0.2484036 | 0.82059221 | 1 | 0.2484036 | 0.2484036 | 0.2484036 | 2.3910228 | 0.456374011 | 0.5048 | 2.391022803 | 2.391022803 | 2.391022803 | 0.103890101 | -0.983425833 | FALSE | FALSE | FALSE |
| LILRA6 | 0.2483973 | 0.82061096 | 1 | 22.65029872 | 2285.675597 | 2285.675597 | 0.26597459 | 0.834908976 | 1 | 24.29681749 | 2456.232725 | 2456.232725 | 0.933913639 | -0.029693282 | FALSE | TRUE | FALSE |
| MCM6 | 0.2483934 | 0.8206297 | 1 | 860.3376361 | 88538.37576 | 435235.6052 | 0.31766905 | 0.819383555 | 1 | 47.4724911 | 1117.842738 | 4472.380445 | 0.781925023 | -0.106834889 | FALSE | TRUE | FALSE |
| SH2B1 | 0.2483026 | 0.82064844 | 1 | 0.248302575 | 0.248302575 | 4.883055807 | 0.29260913 | 0.827247289 | 0.01855 | 0.292609134 | 0.292609134 | 5.765955043 | 0.848581081 | -0.071306655 | FALSE | TRUE | FALSE |
| TMTC3 | 0.2482882 | 0.82066718 | 0.86155 | 5.095893524 | 326.1180137 | 1305.098424 | 0.41547172 | 0.786348998 | 0.89455 | 3.743995604 | 59.92389076 | 239.8253423 | 0.59760562 | -0.223585327 | FALSE | TRUE | FALSE |
| CCDC85A | 0.2481195 | 0.82068591 | 1 | 0.248119486 | 0.248119486 | 4.909921443 | 0.29224422 | 0.82741262 | 1 | 0.292244222 | 0.292244222 | 5.790441042 | 0.849014171 | -0.071085061 | FALSE | TRUE | FALSE |
| AGO4 | 0.2480543 | 0.82070464 | 1 | 0.248054303 | 0.248054303 | 4.875366975 | 0.29234383 | 0.827357547 | 1 | 0.292343833 | 0.292343833 | 5.757805213 | 0.848501917 | -0.071347172 | FALSE | TRUE | FALSE |
| CDK14 | 0.2480395 | 0.82072336 | 0.509 | 2.293959037 | 2.293959037 | 4.919393964 | 0.3011658 | 0.825144641 | 0.01045 | 0.301165801 | 0.301165801 | 5.979046959 | 0.823597985 | -0.084284724 | FALSE | TRUE | FALSE |
| CDK17 | 0.248006 | 0.82074208 | 0.99955 | 2372.8653 | 223006.2518 | 867245.4155 | 2.34942537 | 0.460599187 | 0.9999 | 781911.3286 | 323954034.8 | 3356308591 | 0.105560264 | -0.976499533 | FALSE | FALSE | FALSE |
| FAM171A1 | 0.2479937 | 0.8207608 | 0.02715 | 0.247993745 | 0.247993745 | 4.853082497 | 0.24258187 | 0.841259279 | 1 | 208.9997563 | 21436.05008 | 224962.4277 | 1.022309489 | 0.009582392 | FALSE | TRUE | FALSE |
| ZNF10 | 0.2478875 | 0.82077951 | 1 | 0.247887475 | 0.247887475 | 4.910980003 | 0.28304259 | 0.830228621 | 1 | 2.631138429 | 2.631138429 | 5.61381227 | 0.875795658 | -0.057597212 | FALSE | TRUE | FALSE |
| GPR65 | 0.2478509 | 0.82079822 | 1 | 0.24785094 | 0.24785094 | 0.24785094 | 0.29161468 | 0.827577628 | 1 | 0.291614682 | 0.291614682 | 0.291614682 | 0.849926137 | -0.070618815 | FALSE | TRUE | FALSE |
| ST8SIA6 | 0.2477789 | 0.82081693 | 1 | 0.247778861 | 0.247778861 | 0.247778861 | 0.2917568 | 0.827540986 | 1 | 0.291756796 | 0.291756796 | 0.291756796 | 0.849265088 | -0.070956729 | FALSE | TRUE | FALSE |
| PAQR7 | 0.247719 | 0.82083563 | 1 | 0.247718987 | 0.247718987 | 4.913135556 | 0.28259552 | 0.830405988 | 1 | 2.608849605 | 2.608849605 | 2.608849605 | 0.876584971 | -0.05720598 | FALSE | TRUE | FALSE |
| RBMXL1 | 0.2475637 | 0.82085433 | 1 | 7.35414E+13 | 2.08764E+18 | 3.92472E+20 | 0.38272822 | 0.798170425 | 1 | 5698166974 | 9.91944E+12 | 1.58608E+15 | 0.64683932 | -0.189203588 | FALSE | TRUE | FALSE |
| CLK4 | 0.247563 | 0.82087302 | 0.0145 | 0.247562958 | 0.247562958 | 4.899194643 | 2.62659455 | 0.43823183 | 0.5074 | 2.626594555 | 2.626594555 | 5.601965019 | 0.094252445 | -1.025707377 | FALSE | FALSE | FALSE |
| ARHGDIB | 0.2475602 | 0.82089171 | 1 | 0.247560204 | 0.247560204 | 0.247560204 | 0.29173337 | 0.827559309 | 1 | 0.291733373 | 0.291733373 | 0.291733373 | 0.848583765 | -0.071305281 | FALSE | TRUE | FALSE |
| PRKCB | 0.2475128 | 0.8209104 | 0.02165 | 0.247512802 | 0.247512802 | 4.869984227 | 0.29204157 | 0.827485995 | 1 | 0.292041571 | 0.292041571 | 5.7573349 | 0.847525924 | -0.071847009 | FALSE | TRUE | FALSE |
| OR6K6 | 0.2475 | 0.82092908 | 1 | 0.24750001 | 0.24750001 | 0.24750001 | 0.29105892 | 0.827742324 | 1 | 0.291058921 | 0.291058921 | 0.291058921 | 0.850343322 | -0.070405694 | FALSE | TRUE | FALSE |
| RAET1G | 0.2474066 | 0.82094776 | 0.00675 | 0.247406604 | 0.247406604 | 0.247406604 | 0.29089913 | 0.827797153 | 0.0071 | 0.290899132 | 0.290899132 | 0.290899132 | 0.850489317 | -0.070331137 | FALSE | TRUE | FALSE |

| | | | | | | | | | | | | | | | | | |
|---|---|---|---|---|---|---|---|---|---|---|---|---|---|---|---|---|---|
| TRIM25 | 0.2474063 | 0.82096643 | 0.0166 | 0.247406333 | 0.247406333 | 4.877087629 | 0.28523233 | 0.829515385 | 1 | 1.140798589 | 4.563812175 | 22.5272655 | 0.867385311 | -0.061787937 | FALSE | TRUE | FALSE |
| MRGPRD | 0.2473769 | 0.8209851 | 0.01055 | 0.247376867 | 0.247376867 | 4.909738991 | 0.2823225 | 0.83049453 | 0.50955 | 2.611420107 | 2.611420107 | 2.611420107 | 0.876220884 | -0.0573864 | FALSE | TRUE | FALSE |
| POP5 | 0.2472407 | 0.82100377 | 1 | 0.247240691 | 0.247240691 | 0.247240691 | 0.28793218 | 0.828741941 | 1 | 1.151592978 | 1.151592978 | 1.151592978 | 0.858676815 | -0.066170263 | FALSE | TRUE | FALSE |
| VPS26A | 0.2468193 | 0.82102243 | 1 | 0.987131486 | 0.987131486 | 0.987131486 | 0.28514183 | 0.829569086 | 1 | 0.28514183 | 2.653265925 | 2.653265925 | 0.865601981 | -0.062681758 | FALSE | TRUE | FALSE |
| WDR93 | 0.2468046 | 0.8210411 | 0.50445 | 2.198719603 | 20.09174172 | 20.09174172 | 280.763547 | 0.008964166 | 0.06425 | 12.50047757 | 280.7635475 | 1123.036509 | 0.000879048 | -3.055987554 | FALSE | FALSE | FALSE |
| LRRTM3 | 0.2467061 | 0.82105975 | 0.87405 | 2.314557838 | 220.0990624 | 220.0990624 | 0.3198805 | 0.818798306 | 0.01595 | 0.319880496 | 0.319880496 | 6.329514556 | 0.771244713 | -0.1128078 | FALSE | TRUE | FALSE |
| C1orf159 | 0.2467008 | 0.8210784 | 1 | 0.246700811 | 0.246700811 | 0.246700811 | 0.2899552 | 0.828179962 | 1 | 0.289955198 | 0.289955198 | 0.289955198 | 0.850823895 | -0.070160322 | FALSE | TRUE | FALSE |
| MRPL1 | 0.2466883 | 0.82109705 | 1 | 0.246688346 | 0.246688346 | 0.246688346 | 0.29024972 | 0.828016117 | 0.00715 | 0.290249718 | 0.290249718 | 0.290249718 | 0.84991761 | -0.070623172 | FALSE | TRUE | FALSE |
| PEMT | 0.2466794 | 0.8211157 | 1 | 2.287275266 | 2.287275266 | 2.287275266 | 0.29979749 | 0.825520438 | 1 | 0.299797489 | 0.299797489 | 0.299797489 | 0.822820168 | -0.084695072 | FALSE | TRUE | FALSE |
| ZNF131 | 0.2466676 | 0.82113434 | 0.01725 | 0.246667649 | 0.246667649 | 4.880823105 | 0.29065494 | 0.827888454 | 1 | 0.290654945 | 0.290654945 | 5.758568421 | 0.84866146 | -0.07126552 | FALSE | TRUE | FALSE |
| FUT7 | 0.2466344 | 0.82115298 | 0.0109 | 0.24663442 | 0.24663442 | 4.895038176 | 0.29075283 | 0.827870202 | 1 | 0.290752833 | 0.290752833 | 0.290752833 | 0.848261451 | -0.071470269 | FALSE | TRUE | FALSE |
| MFAP5 | 0.2465987 | 0.82117161 | 0.0057 | 0.246598711 | 0.246598711 | 0.246598711 | 0.2899811 | 0.828143579 | 0.00505 | 0.289981096 | 0.289981096 | 0.289981096 | 0.850395816 | -0.070378885 | FALSE | TRUE | FALSE |
| PLCH2 | 0.2465334 | 0.82119024 | 0.81935 | 2.241927402 | 20.89914127 | 199.5896511 | 0.24213752 | 0.841368042 | 0.26855 | 0.060563803 | 0.242137515 | 1.18166167 | 1.018154631 | 0.007813741 | FALSE | TRUE | FALSE |
| GATAD2A | 0.24653 | 0.82120886 | 1 | 0.246529975 | 0.246529975 | 4.844744114 | 0.29101882 | 0.827760605 | 1 | 0.291018825 | 0.291018825 | 5.731017199 | 0.847127244 | -0.072051351 | FALSE | TRUE | FALSE |
| HNRNPF | 0.2464984 | 0.82122748 | 0.01105 | 0.246498407 | 0.246498407 | 4.884959012 | 0.27385014 | 0.832627109 | 1 | 0.273850137 | 0.273850137 | 5.432958668 | 0.900121541 | -0.045698845 | FALSE | TRUE | FALSE |
| TSEN34 | 0.2463719 | 0.8212461 | 1 | 0.24637189 | 0.24637189 | 0.24637189 | 1.14873012 | 0.57248253 | 0.00865 | 1.148730116 | 1.148730116 | 1.148730116 | 0.214473258 | -0.668626851 | FALSE | FALSE | FALSE |
| CIAPIN1 | 0.2463566 | 0.82126471 | 1 | 0.246356596 | 0.985302058 | 0.985302058 | 0.29359472 | 0.826989478 | 1 | 0.29359472 | 0.29359472 | 0.29359472 | 0.839104313 | -0.076184046 | FALSE | TRUE | FALSE |
| OR2L2 | 0.246185 | 0.82128332 | 0.00705 | 0.246184971 | 0.246184971 | 0.246184971 | 2.58359172 | 0.440973987 | 0.4962 | 0.28080547 | 2.583591718 | 2.583591718 | 0.095287878 | -1.020962347 | FALSE | FALSE | FALSE |
| SAV1 | 0.2460784 | 0.82130193 | 1 | 0.246078389 | 0.246078389 | 4.88655902 | 0.29031689 | 0.827961429 | 1 | 0.290316893 | 0.290316893 | 0.290316893 | 0.847619945 | -0.071798832 | FALSE | TRUE | FALSE |
| HYPK | 0.245851 | 0.82132053 | 1 | 2.28250686 | 2.28250686 | 2.28250686 | 0.28180574 | 0.830653671 | 1 | 0.281805738 | 0.281805738 | 0.281805738 | 0.8724132 | -0.059277772 | FALSE | TRUE | FALSE |
| FAM151B | 0.2457683 | 0.82133913 | 0.87595 | 21.17081326 | 204.1749708 | 204.1749708 | 0.29814136 | 0.825913325 | 1 | 2.745123591 | 25.94146454 | 25.94146454 | 0.824334848 | -0.08389634 | FALSE | TRUE | FALSE |
| CLDN3 | 0.2457282 | 0.82135773 | 0.0085 | 0.245728171 | 0.245728171 | 0.245728171 | 0.29024841 | 0.828034338 | 1 | 0.29024841 | 0.29024841 | 0.29024841 | 0.846613322 | -0.072314902 | FALSE | TRUE | FALSE |
| CTRB2 | 0.2457207 | 0.82137632 | 1 | 0.982515033 | 0.982515033 | 0.982515033 | 0.2902374 | 0.828052554 | 1 | 1.160566564 | 1.160566564 | 1.160566564 | 0.846619569 | -0.072311697 | FALSE | TRUE | FALSE |
| RNF112 | 0.2456104 | 0.82139491 | 1 | 0.245610356 | 0.245610356 | 4.840660794 | 0.28938868 | 0.828325337 | 1 | 0.289388681 | 0.289388681 | 5.713472699 | 0.848721364 | -0.071234865 | FALSE | TRUE | FALSE |
| RSAD1 | 0.2455845 | 0.82141349 | 0.93725 | 2.208064314 | 192.5065138 | 192.5065138 | 0.40825049 | 0.789147291 | 0.01125 | 0.408250489 | 0.408250489 | 0.913032698 | 0.601553523 | -0.220725726 | FALSE | TRUE | FALSE |
| TRIM72 | 0.2455332 | 0.82143207 | 1 | 4.865777514 | 86.66078964 | 86.66078964 | 24.4519759 | 0.112751767 | 0.68835 | 24.45197587 | 2392.679424 | 2392.679424 | 0.010041446 | -1.998203758 | FALSE | FALSE | FALSE |
| MAPK8 | 0.2454402 | 0.82145065 | 0.0104 | 0.245440163 | 0.245440163 | 4.8740793 | 0.28972411 | 0.828252679 | 1 | 0.289724109 | 0.289724109 | 0.289724109 | 0.847151324 | -0.072039006 | FALSE | TRUE | FALSE |
| FAM177A1 | 0.2454015 | 0.82146922 | 1 | 0.245401457 | 0.245401457 | 0.245401457 | 0.27176997 | 0.833263422 | 1 | 2.511208841 | 23.81528787 | 23.81528787 | 0.902974867 | -0.044324338 | FALSE | TRUE | FALSE |
| FBXO15 | 0.2453166 | 0.82148779 | 0.5062 | 2.255298768 | 2.255298768 | 4.867257417 | 23.6547747 | 0.115174258 | 0.1269 | 23.65477467 | 229.3733908 | 229.3733908 | 0.0103707 | -1.984191914 | FALSE | FALSE | FALSE |
| OR5L2 | 0.2452866 | 0.82150635 | 1 | 205.7937303 | 20738.13119 | 215730.4352 | 0.31683347 | 0.819764933 | 1 | 27.82925529 | 271.0116992 | 271.0116992 | 0.774181346 | -0.111157297 | FALSE | TRUE | FALSE |
| NECAB1 | 0.2451986 | 0.82152492 | 0.00625 | 0.245198558 | 0.245198558 | 0.245198558 | 2.24155153 | 0.468153726 | 0.0046 | 2.241551531 | 2.241551531 | 2.241551531 | 0.109387875 | -0.961030816 | FALSE | FALSE | FALSE |
| MRPS27 | 0.2451299 | 0.82154347 | 1 | 0.245129923 | 0.245129923 | 0.245129923 | 0.28872684 | 0.828488602 | 1 | 0.288726837 | 0.288726837 | 0.288726837 | 0.849002901 | -0.071090826 | FALSE | TRUE | FALSE |
| RXRA | 0.2450475 | 0.82156203 | 0.99865 | 62.71082498 | 1004.70709 | 4023.004843 | 0.09125746 | 0.877880856 | 1 | 1.458721594 | 5.834279544 | 7.211071151 | 2.685232302 | 0.428981863 | FALSE | TRUE | FALSE |
| CETN3 | 0.245041 | 0.82158058 | 1 | 0.245040993 | 0.245040993 | 0.245040993 | 1.1421159 | 0.573194935 | 0.50335 | 1.142115897 | 1.142115897 | 1.142115897 | 0.214550024 | -0.668471434 | FALSE | FALSE | FALSE |
| LNPEP | 0.2449302 | 0.82159912 | 1 | 0.244930175 | 0.244930175 | 4.81597561 | 0.28891516 | 0.828397938 | 1 | 0.288915161 | 0.288915161 | 5.692372789 | 0.847758122 | -0.071728041 | FALSE | TRUE | FALSE |
| EQTN | 0.2447038 | 0.82161767 | 0.0069 | 0.244703752 | 0.244703752 | 0.244703752 | 0.28803472 | 0.828687717 | 1 | 0.288034724 | 0.288034724 | 0.288034724 | 0.849563375 | -0.070804218 | FALSE | TRUE | FALSE |
| LNX1 | 0.2445878 | 0.82163621 | 0.75675 | 0.978209507 | 8.831903252 | 19.25695421 | 0.04240266 | 0.888448016 | 1 | 0.376225947 | 3.426179106 | 7.420354286 | 5.768218841 | 0.761041729 | FALSE | TRUE | FALSE |
| ERLIN1 | 0.2445805 | 0.82165474 | 1 | 0.2445805 | 0.2445805 | 0.2445805 | 1.12918679 | 0.575083761 | 0.75355 | 1.129186793 | 4.516993867 | 4.516993867 | 0.216598796 | -0.664343962 | FALSE | FALSE | FALSE |
| PIBF1 | 0.244571 | 0.82167327 | 0.3241 | 0.244570958 | 20.9336653 | 20.9336653 | 0.04182626 | 0.888576097 | 0.9404 | 0.376379959 | 32.86860858 | 32.86860858 | 5.847305836 | 0.76695581 | FALSE | TRUE | FALSE |
| IFT46 | 0.2445562 | 0.8216918 | 1 | 0.244556174 | 0.244556174 | 0.244556174 | 0.28843782 | 0.828597267 | 1 | 0.288437815 | 0.288437815 | 0.288437815 | 0.847864465 | -0.071673566 | FALSE | TRUE | FALSE |
| MAML2 | 0.2444639 | 0.82171033 | 1 | 0.244463889 | 0.244463889 | 8.239229882 | 0.28883552 | 0.828416078 | 1 | 0.288835516 | 0.288835516 | 5.687722413 | 0.846377522 | -0.072435879 | FALSE | TRUE | FALSE |
| ZC3H12A | 0.2444392 | 0.82172885 | 1 | 0.244439222 | 0.244439222 | 4.81464544 | 0.28853624 | 0.828579166 | 1 | 0.288536237 | 0.288536237 | 5.693584767 | 0.847169922 | -0.072029472 | FALSE | TRUE | FALSE |
| SLC30A2 | 0.2444369 | 0.82174736 | 1 | 0.244436947 | 0.244436947 | 4.848157486 | 0.28875328 | 0.828434215 | 1 | 0.288753275 | 0.288753275 | 0.288753275 | 0.846525279 | -0.072360068 | FALSE | TRUE | FALSE |
| LPAR3 | 0.2444298 | 0.82176587 | 1 | 0.244429783 | 0.244429783 | 0.244429783 | 0.27951142 | 0.831234661 | 1 | 2.59714746 | 2.59714746 | 2.59714746 | 0.874489448 | -0.058245427 | FALSE | TRUE | FALSE |
| ZNF219 | 0.244252 | 0.82178438 | 0.5045 | 2.258631872 | 2.258631872 | 8.264071897 | 24.5813363 | 0.112151321 | 0.25855 | 2.58450686 | 24.58133626 | 51.04022452 | 0.009936484 | -2.002767273 | FALSE | FALSE | FALSE |
| NPAS2 | 0.2441914 | 0.82180289 | 1 | 0.244191423 | 0.244191423 | 4.794397409 | 0.2883557 | 0.828615365 | 1 | 0.288355695 | 0.288355695 | 5.673991471 | 0.846840992 | -0.072198128 | FALSE | TRUE | FALSE |
| ANGPTL7 | 0.2441394 | 0.82182139 | 0.2528 | 0.24413939 | 0.975939856 | 0.975939856 | 0.16019449 | 0.862000336 | 0.75245 | 1.46555655 | 23.4399666 | 23.4399666 | 1.524018615 | 0.182990272 | FALSE | TRUE | FALSE |
| DPP8 | 0.2440028 | 0.82183989 | 1 | 21.56077513 | 2108.485213 | 2108.485213 | 0.32617102 | 0.816688247 | 1 | 0.326171025 | 0.326171025 | 6.428629183 | 0.748082484 | -0.126050514 | FALSE | TRUE | FALSE |
| DDRGK1 | 0.2439584 | 0.82185838 | 1 | 2.258629695 | 2.258629695 | 4.839039962 | 2.67219596 | 0.435863389 | 0.50475 | 2.672195961 | 2.672195961 | 2.672195961 | 0.091295101 | -1.039552524 | FALSE | FALSE | FALSE |
| ASCC2 | 0.2439325 | 0.82187687 | 1 | 0.243932517 | 0.243932517 | 4.793303018 | 23.094142 | 0.117001917 | 0.1421 | 23.09414198 | 225.2003461 | 454.7584583 | 0.010562528 | -1.976232134 | FALSE | FALSE | FALSE |
| TMEM184C | 0.2439182 | 0.82189536 | 0.7572 | 2.249226959 | 21.28708556 | 21.28708556 | 0.30588728 | 0.8235673 | 1 | 0.305887284 | 0.305887284 | 0.305887284 | 0.797412055 | -0.098317203 | FALSE | TRUE | FALSE |
| APOD | 0.2439101 | 0.82191384 | 0.00545 | 0.243910052 | 0.243910052 | 0.243910052 | 0.28740968 | 0.828886372 | 1 | 0.287409683 | 0.287409683 | 0.287409683 | 0.848649392 | -0.071271696 | FALSE | TRUE | FALSE |
| ZNF366 | 0.2438145 | 0.82193232 | 1 | 2.268885706 | 21.69984054 | 44.53172552 | 0.29696376 | 0.826211503 | 1 | 2.772531632 | 2.772531632 | 54.53901314 | 0.821024508 | -0.085643879 | FALSE | TRUE | FALSE |
| MXRA8 | 0.2437373 | 0.82195079 | 1 | 2.249626517 | 21.31044015 | 44.46269098 | 0.30576716 | 0.823605646 | 1 | 0.305767163 | 0.305767163 | 6.051890443 | 0.797133528 | -0.098468924 | FALSE | TRUE | FALSE |
| CNTF | 0.2437243 | 0.82196926 | 0.5019 | 2.239795245 | 2.239795245 | 2.239795245 | 0.29616754 | 0.826341646 | 1 | 0.296167539 | 0.296167539 | 0.296167539 | 0.822927152 | -0.084638608 | FALSE | TRUE | FALSE |
| CTU2 | 0.2437094 | 0.82198773 | 0.8147 | 2.100809371 | 18.56431604 | 168.0702021 | 4.23409563 | 0.341700479 | 0.31705 | 0.482914736 | 38.05556049 | 38.05556049 | 0.057558789 | -1.239888352 | FALSE | FALSE | FALSE |
| NDRG2 | 0.2436902 | 0.82200619 | 0.8721 | 21.13394771 | 204.5073734 | 204.5073734 | 0.30539791 | 0.823758861 | 1 | 2.822905999 | 2.822905999 | 6.064952255 | 0.797943268 | -0.098027985 | FALSE | TRUE | FALSE |
| VN1R1 | 0.243512 | 0.82202465 | 1 | 0.243511991 | 0.243511991 | 0.243511991 | 0.28686921 | 0.829102556 | 1 | 0.286869212 | 0.286869212 | 0.286869212 | 0.848860669 | -0.071163589 | FALSE | TRUE | FALSE |
| STK38L | 0.2434441 | 0.8220431 | 1 | 0.243444091 | 0.243444091 | 4.828504711 | 0.28716586 | 0.828958495 | 1 | 0.287165858 | 0.287165858 | 5.701368992 | 0.847747335 | -0.071733567 | FALSE | TRUE | FALSE |
| GINM1 | 0.2433733 | 0.82206156 | 0.0069 | 0.243373339 | 0.243373339 | 0.243373339 | 0.28670684 | 0.829156515 | 1 | 0.286706842 | 0.286706842 | 0.286706842 | 0.848857799 | -0.071165057 | FALSE | TRUE | FALSE |
| MAP2K5 | 0.2433217 | 0.82208 | 0.01115 | 0.243321735 | 0.243321735 | 4.829810905 | 0.27820801 | 0.831497476 | 1 | 2.570899378 | 2.570899378 | 2.570899378 | 0.874603632 | -0.058188724 | FALSE | TRUE | FALSE |
| HARBI1 | 0.2431522 | 0.82209845 | 1 | 0.243152229 | 0.243152229 | 4.826874392 | 2.58437141 | 0.440778818 | 0.00855 | 2.584371412 | 2.584371412 | 2.584371412 | 0.094085636 | -1.026476673 | FALSE | FALSE | FALSE |
| SPSB4 | 0.2429976 | 0.82211689 | 0.00635 | 0.242997636 | 0.242997636 | 0.242997636 | 0.28376464 | 0.829961872 | 1 | 0.283764637 | 1.134860973 | 1.134860973 | 0.856335162 | -0.067356223 | FALSE | TRUE | FALSE |
| PPP1R15B | 0.2429598 | 0.82213533 | 1 | 21084.03284 | 26525838.69 | 296554673.4 | 0.3676134 | 0.803065072 | 1 | 3.432103607 | 32.88646796 | 67.77901853 | 0.660911083 | -0.179856965 | FALSE | TRUE | FALSE |
| MRPS34 | 0.2429504 | 0.82215376 | 1 | 2.227828871 | 2.227828871 | 2.227828871 | 2.55329268 | 0.443499263 | 0.7531 | 2.553292682 | 24.10560669 | 24.10560669 | 0.095151792 | -1.021583027 | FALSE | FALSE | FALSE |
| FRMD4B | 0.2429403 | 0.82217219 | 0.61225 | 0.242940297 | 9.278844857 | 37.08108251 | 6.4041E-05 | 0.900277892 | 1 | 0.058805352 | 0.549472448 | 1.163554777 | 3793.506289 | 3.579040808 | FALSE | TRUE | FALSE |
| SDCBP | 0.2427356 | 0.82219062 | 0.00675 | 0.242735575 | 0.242735575 | 0.242735575 | 0.28633387 | 0.829228408 | 1 | 0.286333869 | 0.286333869 | 0.286333869 | 0.847736161 | -0.071739291 | FALSE | TRUE | FALSE |
| LINGO1 | 0.2424029 | 0.82220904 | 0.75265 | 2.296204575 | 9.182318917 | 45.07964071 | 0.29944725 | 0.825632867 | 1 | 0.299447247 | 0.299447247 | 5.891836728 | 0.809501241 | -0.091782481 | FALSE | TRUE | FALSE |
| CHERP | 0.2421834 | 0.82222746 | 0.02835 | 0.242183388 | 0.242183388 | 4.741814893 | 0.28627895 | 0.829264332 | 0.0249 | 0.286278949 | 0.286278949 | 5.619378076 | 0.845969948 | -0.072645065 | FALSE | TRUE | FALSE |
| UBQLN3 | 0.2420649 | 0.82224588 | 0.51045 | 0.548915269 | 2.195071514 | 4.779273905 | 64.3150743 | 0.043657542 | 0.07035 | 6.713725765 | 257.1993621 | 257.1993621 | 0.003763736 | -2.424380894 | FALSE | FALSE | FALSE |
| BTBD3 | 0.2420586 | 0.8222643 | 1 | 0.242058612 | 0.242058612 | 4.788518156 | 0.28310552 | 0.830210864 | 1 | 1.132187745 | 1.132187745 | 5.607855505 | 0.85501198 | -0.0680278 | FALSE | TRUE | FALSE |
| RAP1GDS1 | 0.2420174 | 0.82228271 | 1 | 0.242017372 | 0.242017372 | 4.793790584 | 0.27698606 | 0.831829226 | 0.50775 | 2.564177395 | 2.564177395 | 5.49278486 | 0.873752908 | -0.058611366 | FALSE | TRUE | FALSE |
| CASQ1 | 0.2419574 | 0.82230111 | 1 | 7.81225E+31 | 5.38983E+39 | 2.52282E+43 | 0.09501566 | 0.877140387 | 1 | 9.01528E+36 | 8.49072E+44 | 4.27509E+48 | 2.546500814 | 0.405943819 | FALSE | TRUE | FALSE |

| | | | | | | | | | | | | | | | | | |
|---|---|---|---|---|---|---|---|---|---|---|---|---|---|---|---|---|---|
| ZNF500 | 0.2419544 | 0.82231952 | 1 | 0.241954404 | 0.241954404 | 4.781642963 | 0.27689022 | 0.831864071 | 1 | 2.559960579 | 2.559960579 | 5.479880366 | 0.873827925 | -0.058574081 | FALSE | TRUE | FALSE |
| C19orf18 | 0.2416504 | 0.82233792 | 0.00665 | 0.241650383 | 0.241650383 | 0.241650383 | 0.28481049 | 0.829676388 | 0.0055 | 0.284810486 | 0.284810486 | 0.284810486 | 0.848460274 | -0.071368487 | FALSE | TRUE | FALSE |
| FADS3 | 0.2416007 | 0.82235631 | 0.01045 | 0.241600676 | 0.241600676 | 4.792558821 | 0.28573396 | 0.829372016 | 1 | 0.285733959 | 0.285733959 | 5.673583147 | 0.845544143 | -0.072863715 | FALSE | TRUE | FALSE |
| B3GALT5 | 0.2415283 | 0.8223747 | 0.00875 | 0.241528323 | 0.241528323 | 0.241528323 | 0.28572015 | 0.82938995 | 1 | 0.285720146 | 0.285720146 | 0.285720146 | 0.845331792 | -0.072972797 | FALSE | TRUE | FALSE |
| GPX8 | 0.2414116 | 0.82239309 | 1 | 0.241411585 | 0.241411585 | 0.241411585 | 0.28476569 | 0.829694258 | 1 | 0.284765694 | 0.284765694 | 0.284765694 | 0.847755155 | -0.071729561 | FALSE | TRUE | FALSE |
| ATG13 | 0.241266 | 0.82241148 | 1 | 0.241265984 | 0.241265984 | 4.77597513 | 0.26881229 | 0.834115679 | 1 | 0.268812293 | 0.268812293 | 5.327813285 | 0.89752586 | -0.046953029 | FALSE | TRUE | FALSE |
| DENND5B | 0.2412303 | 0.82242986 | 1 | 0.241230305 | 0.241230305 | 4.719260563 | 2.59208634 | 0.440388011 | 0.5136 | 2.592086336 | 2.592086336 | 50.84313976 | 0.093064147 | -1.031217597 | FALSE | FALSE | FALSE |
| IFNA7 | 0.2412297 | 0.82244824 | 0.12685 | 0.093705357 | 2.343595416 | 2.343595416 | 0.02998349 | 0.891244217 | 0.7483 | 0.029983486 | 0.285199227 | 0.285199227 | 8.045418693 | 0.90554865 | FALSE | TRUE | FALSE |
| SIL1 | 0.2410845 | 0.82246661 | 0.0115 | 0.241084451 | 0.241084451 | 4.774353099 | 0.28172473 | 0.830688994 | 0.5145 | 1.126777957 | 1.126777957 | 5.585767598 | 0.855744709 | -0.067655777 | FALSE | TRUE | FALSE |
| INTS7 | 0.2410029 | 0.82248498 | 1 | 0.241002873 | 0.241002873 | 4.741647292 | 0.28491075 | 0.829622755 | 0.01905 | 0.284910746 | 0.284910746 | 5.616496494 | 0.845889025 | -0.07268661 | FALSE | TRUE | FALSE |
| CREB5 | 0.2409772 | 0.82250335 | 1 | 0.240977213 | 0.240977213 | 4.765531614 | 0.28515728 | 0.82955119 | 1 | 0.285157279 | 0.285157279 | 5.646833659 | 0.845067723 | -0.073108486 | FALSE | TRUE | FALSE |
| WNT6 | 0.2407966 | 0.82252172 | 0.011 | 0.240796618 | 0.240796618 | 4.769166 | 1.10201841 | 0.57846747 | 0.8731 | 4.408043398 | 17.63645959 | 17.63645959 | 0.218505078 | -0.660538466 | FALSE | FALSE | FALSE |
| BRD9 | 0.2407468 | 0.82254008 | 1 | 0.240746757 | 0.240746757 | 4.761465637 | 0.28426195 | 0.829765703 | 1 | 0.284261947 | 0.284261947 | 5.629615786 | 0.846918695 | -0.072158281 | FALSE | TRUE | FALSE |
| TMCO1 | 0.2406957 | 0.82255843 | 1 | 0.240695723 | 0.240695723 | 0.240695723 | 0.28404358 | 0.82983709 | 1 | 0.284043584 | 0.284043584 | 0.284043584 | 0.847390105 | -0.071916612 | FALSE | TRUE | FALSE |
| CCL2 | 0.2406673 | 0.82257678 | 0.00245 | 0.240667347 | 0.240667347 | 0.240667347 | 0.24566067 | 0.840493799 | 0.0022 | 0.245660669 | 0.245660669 | 0.245660669 | 0.979673908 | -0.008918459 | FALSE | TRUE | FALSE |
| TRMT1L | 0.2405679 | 0.82259513 | 1 | 2.225838908 | 21.13695113 | 43.95344423 | 0.30246823 | 0.824653645 | 1 | 0.302468231 | 0.302468231 | 5.981908073 | 0.795349172 | -0.099442167 | FALSE | TRUE | FALSE |
| TRMT13 | 0.2405412 | 0.82261348 | 0.751 | 0.240541228 | 0.962299531 | 1.192539174 | 0.08005785 | 0.880300673 | 0.75505 | 0.717512547 | 6.600205088 | 6.600205088 | 3.004592589 | 0.477785592 | FALSE | TRUE | FALSE |
| VEZT | 0.2404842 | 0.82263182 | 0.01895 | 0.240484222 | 0.240484222 | 4.748288524 | 0.28418609 | 0.829819248 | 0.0176 | 0.284186091 | 0.284186091 | 5.619787559 | 0.846220944 | -0.07251623 | FALSE | TRUE | FALSE |
| GCA | 0.2404657 | 0.82265016 | 1 | 0.240465682 | 0.240465682 | 0.240465682 | 2.55615294 | 0.443112185 | 0.00635 | 0.27544966 | 2.556152944 | 2.556152944 | 0.094073276 | -1.02653373 | FALSE | FALSE | FALSE |
| TOLLIP | 0.2403851 | 0.82266849 | 1 | 0.240385141 | 0.240385141 | 4.769602704 | 0.28454527 | 0.829729987 | 1 | 0.284545271 | 0.284545271 | 0.284545271 | 0.844804555 | -0.073243753 | FALSE | TRUE | FALSE |
| C9orf78 | 0.2403774 | 0.82268682 | 1 | 0.24037744 | 0.24037744 | 0.24037744 | 0.28369273 | 0.829997489 | 0.00765 | 0.283692725 | 0.283692725 | 0.283692725 | 0.847316194 | -0.071954493 | FALSE | TRUE | FALSE |
| SMOC1 | 0.2402963 | 0.82270515 | 1 | 0.240296271 | 0.240296271 | 4.756081252 | 0.28389575 | 0.829872761 | 1 | 0.283895748 | 0.283895748 | 5.626037446 | 0.846424342 | -0.072411856 | FALSE | TRUE | FALSE |
| IFNK | 0.2402732 | 0.82272347 | 0.00485 | 0.240273177 | 0.240273177 | 0.240273177 | 0.27477536 | 0.832350426 | 1 | 0.274775359 | 2.523292619 | 2.523292619 | 0.874434949 | -0.058272493 | FALSE | TRUE | FALSE |
| ZNF184 | 0.2401897 | 0.82274179 | 1 | 0.240189702 | 0.240189702 | 4.744209752 | 2.41135855 | 0.454143594 | 0.8758 | 23.05791468 | 226.1419736 | 226.1419736 | 0.099607627 | -1.001707407 | FALSE | FALSE | FALSE |
| DNAJC22 | 0.2400676 | 0.8227601 | 0.50255 | 0.960191184 | 0.960191184 | 0.960191184 | 0.28699857 | 0.829030557 | 1 | 0.286998574 | 0.286998574 | 0.286998574 | 0.836476699 | -0.077546152 | FALSE | TRUE | FALSE |
| FAM171B | 0.2400483 | 0.82277841 | 1 | 2.21937153 | 2.21937153 | 43.72070995 | 0.29263172 | 0.827228899 | 1 | 0.292631715 | 0.292631715 | 5.775146912 | 0.820308453 | -0.086022813 | FALSE | TRUE | FALSE |
| TCF4 | 0.2400388 | 0.82279672 | 1 | 0.2400388 | 0.2400388 | 4.734343011 | 5.60567561 | 0.281891497 | 0.01675 | 0.283744237 | 0.283744237 | 5.605675605 | 0.042820673 | -1.368346515 | FALSE | FALSE | FALSE |
| TNFRSF19 | 0.240033 | 0.82281502 | 1 | 0.240033013 | 0.240033013 | 4.755286186 | 0.28385869 | 0.829908417 | 1 | 0.283858686 | 0.283858686 | 5.629908281 | 0.845607425 | -0.072831212 | FALSE | TRUE | FALSE |
| ZNF195 | 0.2400186 | 0.82283332 | 1 | 0.24001856 | 0.24001856 | 4.757119134 | 0.27500813 | 0.832315776 | 1 | 2.549275945 | 2.549275945 | 5.456512059 | 0.872768967 | -0.059100704 | FALSE | TRUE | FALSE |
| SMARCD3 | 0.2399936 | 0.82285162 | 1 | 0.239993574 | 0.239993574 | 4.752474033 | 0.28361279 | 0.830068676 | 1 | 0.283612786 | 0.283612786 | 5.622914722 | 0.84620153 | -0.072526194 | FALSE | TRUE | FALSE |
| WIPI2 | 0.2399505 | 0.82286991 | 0.01365 | 0.23995051 | 0.23995051 | 4.745982099 | 0.2838687 | 0.829890591 | 1 | 0.283868703 | 0.283868703 | 5.62210397 | 0.845286949 | -0.072995836 | FALSE | TRUE | FALSE |
| UBE3D | 0.2399262 | 0.8228882 | 1 | 2.208310721 | 2.208310721 | 4.760859686 | 0.29218787 | 0.827449316 | 1 | 0.292187873 | 0.292187873 | 0.292187873 | 0.821136603 | -0.085584588 | FALSE | TRUE | FALSE |
| LINGO4 | 0.239735 | 0.82290648 | 0.51145 | 0.239735025 | 0.958771302 | 2.158125678 | 0.67187032 | 0.679171024 | 0.0144 | 0.1679523 | 0.671870315 | 3.323935174 | 0.356817408 | -0.447553966 | FALSE | TRUE | FALSE |
| DRD3 | 0.2395827 | 0.82292476 | 1 | 0.239582725 | 0.239582725 | 4.750218449 | 0.28352569 | 0.830139799 | 1 | 0.283525687 | 0.283525687 | 0.283525687 | 0.845012414 | -0.073136911 | FALSE | TRUE | FALSE |
| GPR157 | 0.2395314 | 0.82294304 | 1 | 0.239531418 | 0.239531418 | 4.747663048 | 0.28293373 | 0.830264124 | 1 | 0.28293373 | 0.28293373 | 0.28293373 | 0.846599017 | -0.07232224 | FALSE | TRUE | FALSE |
| STAU1 | 0.2394181 | 0.82296131 | 0.0141 | 0.239418051 | 0.239418051 | 4.734550043 | 0.28041369 | 0.831023835 | 0.5118 | 1.12141022 | 1.12141022 | 5.552745968 | 0.853802993 | -0.068642327 | FALSE | TRUE | FALSE |
| OR10X1 | 0.2394026 | 0.82297958 | 1 | 2.198243047 | 2.198243047 | 2.198243047 | 0.29148098 | 0.827614255 | 1 | 0.291480985 | 0.291480985 | 0.291480985 | 0.821331964 | -0.085481275 | FALSE | TRUE | FALSE |
| NUDT9 | 0.2392794 | 0.82299785 | 1 | 0.239279428 | 0.239279428 | 0.239279428 | 0.28269571 | 0.830352818 | 1 | 0.282695706 | 0.282695706 | 0.282695706 | 0.846420456 | -0.07241385 | FALSE | TRUE | FALSE |
| LMOD1 | 0.2392072 | 0.82301611 | 1 | 0.239207233 | 0.239207233 | 4.72379067 | 0.26611013 | 0.834858565 | 1 | 2.5146778 | 24.3885735 | 49.7344629 | 0.898903155 | -0.046287095 | FALSE | TRUE | FALSE |
| RIC3 | 0.2391711 | 0.82303437 | 1 | 0.239171097 | 0.239171097 | 4.750067911 | 0.28249553 | 0.830423704 | 0.0079 | 0.282495533 | 0.282495533 | 0.282495533 | 0.846636738 | -0.07230289 | FALSE | TRUE | FALSE |
| ZFYVE21 | 0.2391699 | 0.82305263 | 1 | 2.226957983 | 2.226957983 | 2.226957983 | 0.29192319 | 0.827504329 | 0.0068 | 0.29192319 | 0.29192319 | 0.29192319 | 0.819290639 | -0.086562007 | FALSE | TRUE | FALSE |
| TSPAN4 | 0.2390806 | 0.82307088 | 0.93505 | 0.956223811 | 15.30789179 | 15.30789179 | 0.08492194 | 0.879152718 | 1 | 0.339584466 | 1.358264283 | 1.358264283 | 2.815298926 | 0.449524515 | FALSE | TRUE | FALSE |
| FAIM | 0.2388965 | 0.82308913 | 1 | 0.238896506 | 0.238896506 | 0.238896506 | 0.28186192 | 0.830618332 | 1 | 0.281861919 | 0.281861919 | 0.281861919 | 0.847565739 | -0.071826607 | FALSE | TRUE | FALSE |
| TIFAB | 0.2388952 | 0.82310737 | 0.49725 | 0.238895215 | 2.195736765 | 2.195736765 | 23.8598958 | 0.113961227 | 0.24855 | 2.522061335 | 23.85989576 | 23.85989576 | 0.010012417 | -1.999461092 | FALSE | FALSE | FALSE |
| MRPL48 | 0.238581 | 0.82312561 | 1 | 0.238581001 | 0.238581001 | 0.238581001 | 0.28165601 | 0.830724302 | 1 | 0.281656011 | 0.281656011 | 0.281656011 | 0.847065184 | -0.072083168 | FALSE | TRUE | FALSE |
| TANGO2 | 0.2385783 | 0.82314385 | 1 | 2.191236956 | 20.65585782 | 20.65585782 | 0.29972271 | 0.825557931 | 1 | 0.299722709 | 0.299722709 | 0.299722709 | 0.795996817 | -0.099088669 | FALSE | TRUE | FALSE |
| ZFP82 | 0.238571 | 0.82316209 | 1 | 0.238571031 | 0.238571031 | 4.727525508 | 0.28226891 | 0.83052992 | 1 | 0.282268908 | 0.282268908 | 5.599632157 | 0.845190612 | -0.073045335 | FALSE | TRUE | FALSE |
| DNAJC14 | 0.23857 | 0.82318032 | 0.02125 | 0.238569954 | 0.238569954 | 4.701653914 | 0.26578391 | 0.834992925 | 1 | 0.265783906 | 0.265783906 | 5.247192135 | 0.897608728 | -0.046912933 | FALSE | TRUE | FALSE |
| MAN1A2 | 0.2385324 | 0.82319854 | 1 | 0.238532437 | 0.238532437 | 4.71462879 | 0.28243898 | 0.830441416 | 1 | 0.282438983 | 0.282438983 | 5.590335077 | 0.844545019 | -0.073377195 | FALSE | TRUE | FALSE |
| IGFBP7 | 0.2384601 | 0.82321677 | 1 | 0.238460098 | 0.238460098 | 0.238460098 | 0.28162881 | 0.83074195 | 1 | 0.281628814 | 0.281628814 | 0.281628814 | 0.846717685 | -0.072261369 | FALSE | TRUE | FALSE |
| TGFB3 | 0.2384593 | 0.82323499 | 0.01285 | 0.238459259 | 0.238459259 | 4.724095352 | 0.2826872 | 0.830370545 | 1 | 0.282687203 | 0.282687203 | 5.606659847 | 0.843544581 | -0.07389196 | FALSE | TRUE | FALSE |
| CLEC7A | 0.2384296 | 0.8232532 | 1 | 0.238429554 | 0.238429554 | 0.238429554 | 0.28150523 | 0.830794873 | 1 | 0.281505231 | 0.281505231 | 0.281505231 | 0.8469809 | -0.072126383 | FALSE | TRUE | FALSE |
| SLC25A25 | 0.2384022 | 0.82327141 | 0.63365 | 0.41752084 | 3.810581802 | 4.706771248 | 5.80679293 | 0.274474611 | 0.07345 | 1.45214789 | 23.22578299 | 23.22578299 | 0.041055737 | -1.386626147 | FALSE | FALSE | FALSE |
| CASP1 | 0.2383705 | 0.82328962 | 1 | 0.238370457 | 0.238370457 | 4.729333177 | 0.28179694 | 0.830671334 | 1 | 0.281796942 | 0.281796942 | 0.281796942 | 0.845894408 | -0.072683846 | FALSE | TRUE | FALSE |
| PPFIBP1 | 0.2383587 | 0.82330782 | 0.8767 | 2.121028617 | 166.6682045 | 724.4247756 | 0.0136806 | 0.895568344 | 1 | 0.054702851 | 0.218787868 | 1.077068988 | 17.42311238 | 1.241125738 | FALSE | TRUE | FALSE |
| VAV3 | 0.2383324 | 0.82332602 | 0.6994 | 0.238332433 | 3.814315456 | 4.694791809 | 0.3886395 | 0.796209068 | 0.69245 | 0.388639495 | 6.220581654 | 7.669506488 | 0.613248102 | -0.212363787 | FALSE | TRUE | FALSE |
| KRT9 | 0.2383252 | 0.82334422 | 0.50735 | 0.238325229 | 0.238325229 | 0.872379409 | 0.04092333 | 0.888727865 | 1 | 0.04092333 | 0.04092333 | 0.810372029 | 5.823700802 | 0.765199055 | FALSE | TRUE | FALSE |
| CHMP5 | 0.2381547 | 0.82336241 | 1 | 0.238154672 | 0.238154672 | 0.238154672 | 0.27587486 | 0.83209022 | 1 | 1.103220648 | 4.412870606 | 4.412870606 | 0.863270647 | -0.063853026 | FALSE | TRUE | FALSE |
| DNAJC19 | 0.2381479 | 0.8233806 | 0.7523 | 2.201681247 | 20.89079105 | 20.89079105 | 23.6282173 | 0.115780172 | 0.2426 | 2.482465776 | 23.62821727 | 23.62821727 | 0.010078963 | -1.99658416 | FALSE | FALSE | FALSE |
| MDGA2 | 0.2381249 | 0.82339878 | 1 | 0.952417818 | 0.952417818 | 18.7817664 | 0.28517867 | 0.829533289 | 1 | 0.285178665 | 0.285178665 | 5.633221453 | 0.83500242 | -0.078312266 | FALSE | TRUE | FALSE |
| IPO7 | 0.2380966 | 0.82341696 | 1 | 0.238096615 | 0.238096615 | 4.678277292 | 0.27918022 | 0.831322348 | 1 | 1.116510232 | 1.116510232 | 21.98432074 | 0.852841978 | -0.069131431 | FALSE | TRUE | FALSE |
| AGMAT | 0.2379863 | 0.82343514 | 0.97055 | 3.806023486 | 15.2262857 | 60.92912832 | 3.00093508 | 0.411991752 | 0.62285 | 3.000935083 | 114.5288562 | 114.5288562 | 0.079304054 | -1.10070461 | FALSE | FALSE | FALSE |
| NSMCE2 | 0.2379185 | 0.82345331 | 0.0064 | 0.237918458 | 0.237918458 | 0.237918458 | 0.2809848 | 0.830900618 | 1 | 0.280984796 | 0.280984796 | 0.280984796 | 0.846730716 | -0.072254685 | FALSE | TRUE | FALSE |
| CRYGS | 0.237806 | 0.82347148 | 1 | 0.23780596 | 0.23780596 | 0.23780596 | 0.27862143 | 0.831357403 | 1 | 1.114200596 | 1.114200596 | 1.114200596 | 0.853509211 | -0.068791788 | FALSE | TRUE | FALSE |
| CDKN3 | 0.2377288 | 0.82348965 | 1 | 0.237728797 | 0.237728797 | 0.237728797 | 0.24877304 | 0.839784489 | 1 | 0.248773035 | 0.248773035 | 0.248773035 | 0.955605165 | -0.019721512 | FALSE | TRUE | FALSE |
| TMEM30A | 0.2376136 | 0.82350781 | 1 | 0.23761363 | 0.23761363 | 4.712660748 | 0.2812954 | 0.830847762 | 1 | 0.281295398 | 0.281295398 | 0.281295398 | 0.844712113 | -0.073291278 | FALSE | TRUE | FALSE |
| UQCRC2 | 0.2375204 | 0.82352597 | 1 | 0.237520374 | 0.237520374 | 4.709914828 | 0.28076918 | 0.830918231 | 1 | 0.28076918 | 0.28076918 | 5.573225486 | 0.84596313 | -0.072648565 | FALSE | TRUE | FALSE |
| GNG2 | 0.2374932 | 0.82354413 | 1 | 0.237493219 | 0.237493219 | 0.237493219 | 0.23900896 | 0.842309576 | 1 | 0.239008962 | 0.239008962 | 0.239008962 | 0.993658219 | -0.002762971 | FALSE | TRUE | FALSE |
| MTSS1 | 0.2374597 | 0.82356228 | 1 | 0.237459663 | 0.237459663 | 4.666859858 | 0.28135044 | 0.830830135 | 0.0196 | 0.281350437 | 0.281350437 | 5.540998396 | 0.843999624 | -0.073657747 | FALSE | TRUE | FALSE |
| POMK | 0.2372664 | 0.82358043 | 1 | 2.188740716 | 2.188740716 | 4.713060349 | 0.28945179 | 0.828289016 | 1 | 0.289451785 | 0.289451785 | 0.289451785 | 0.819709416 | -0.086340076 | FALSE | TRUE | FALSE |
| PEX3 | 0.2372567 | 0.82359857 | 1 | 0.237256681 | 0.237256681 | 0.237256681 | 2.50465003 | 0.446387081 | 0.0094 | 2.504650032 | 2.504650032 | 2.504650032 | 0.09472648 | -1.023528601 | FALSE | FALSE | FALSE |
| DEFB119 | 0.2372562 | 0.82361672 | 1 | 0.237256233 | 0.237256233 | 0.237256233 | 0.26367008 | 0.835644776 | 0.74535 | 2.425610231 | 22.90205344 | 22.90205344 | 0.899822362 | -0.045843218 | FALSE | TRUE | FALSE |
| ARHGEF37 | 0.2372453 | 0.82363485 | 0.01595 | 0.237245289 | 0.237245289 | 0.52985478 | 0.04196468 | 0.888536099 | 1 | 0.041964681 | 0.041964681 | 0.82865631 | 5.653451473 | 0.752313669 | FALSE | TRUE | FALSE |

| | | | | | | | | | | | | | | | | | |
|---|---|---|---|---|---|---|---|---|---|---|---|---|---|---|---|---|---|
| TRA2A | 0.2369928 | 0.82365299 | 1 | 0.236992778 | 0.236992778 | 4.706197656 | 0.28043073 | 0.830988649 | 0.01065 | 0.280430732 | 0.280430732 | 5.573542716 | 0.845102733 | -0.073090494 | FALSE | TRUE | FALSE |
| TUBA3C | 0.2368913 | 0.82367112 | 0.75985 | 0.236891322 | 2.18537788 | 4.679675889 | 0.37127111 | 0.802052172 | 0.50855 | 0.371271107 | 1.484474825 | 3.244199463 | 0.638054827 | -0.195142001 | FALSE | TRUE | FALSE |
| ZBTB3 | 0.2368903 | 0.82368925 | 1 | 0.236890293 | 0.236890293 | 4.679846879 | 0.28045158 | 0.83097105 | 1 | 0.28045158 | 0.28045158 | 5.548569746 | 0.844674481 | -0.073310626 | FALSE | TRUE | FALSE |
| ZNF70 | 0.2364747 | 0.82370737 | 0.01315 | 0.236474724 | 0.236474724 | 4.683304241 | 0.27996892 | 0.83105901 | 1 | 0.27996892 | 0.27996892 | 5.551206197 | 0.844646341 | -0.073325095 | FALSE | TRUE | FALSE |
| WIF1 | 0.2364106 | 0.82372549 | 1 | 0.945485508 | 0.945485508 | 0.945485508 | 0.28326799 | 0.830193103 | 1 | 0.283267986 | 0.283267986 | 0.283267986 | 0.834582866 | -0.078530536 | FALSE | TRUE | FALSE |
| SH3GLB2 | 0.2363729 | 0.82374361 | 1 | 0.236372887 | 0.236372887 | 4.693083196 | 0.27143022 | 0.833417469 | 1 | 2.525184069 | 2.525184069 | 2.525184069 | 0.870842176 | -0.060060545 | FALSE | TRUE | FALSE |
| KLK7 | 0.2361037 | 0.82376173 | 1 | 0.236103684 | 0.236103684 | 0.236103684 | 0.27996266 | 0.831076592 | 1 | 0.279962658 | 0.279962658 | 0.279962658 | 0.843339916 | -0.073997344 | FALSE | TRUE | FALSE |
| FBXO11 | 0.2360523 | 0.82377984 | 0.50865 | 2.223864693 | 2.223864693 | 43.80845697 | 1.14334141 | 0.573076351 | 0.5106 | 1.143341408 | 1.143341408 | 9.661779064 | 0.206458285 | -0.685167684 | FALSE | FALSE | FALSE |
| RGS5 | 0.2360172 | 0.82379795 | 1 | 0.236017247 | 0.236017247 | 0.236017247 | 0.27927411 | 0.831304818 | 1 | 0.27927411 | 0.27927411 | 0.27927411 | 0.845109655 | -0.073086937 | FALSE | TRUE | FALSE |
| TMEM40 | 0.2359238 | 0.82381605 | 1 | 2.158583111 | 2.158583111 | 2.158583111 | 0.28751992 | 0.828832239 | 1 | 0.287519918 | 0.287519918 | 0.287519918 | 0.820547527 | -0.085896259 | FALSE | TRUE | FALSE |
| STK4 | 0.2359071 | 0.82383415 | 0.013 | 0.235907061 | 0.235907061 | 4.676255259 | 0.2796854 | 0.831164445 | 1 | 0.2796854 | 0.2796854 | 5.54996867 | 0.84347292 | -0.073928856 | FALSE | TRUE | FALSE |
| SKAP1 | 0.2358944 | 0.82385225 | 1 | 0.235894357 | 0.235894357 | 4.687057994 | 0.27948764 | 0.831252206 | 1 | 0.279487637 | 0.279487637 | 0.279487637 | 0.844024301 | -0.073645049 | FALSE | TRUE | FALSE |
| MTMR2 | 0.2356126 | 0.82387034 | 0.5064 | 0.235612617 | 3.766636602 | 15.06586412 | 0.03230811 | 0.890721373 | 0.6384 | 0.058447705 | 0.516118229 | 0.516118229 | 7.292677522 | 0.86288701 | FALSE | TRUE | FALSE |
| ZNF618 | 0.2355978 | 0.82388843 | 1 | 2.195633819 | 2.195633819 | 42.94041682 | 0.25441113 | 0.838056612 | 1 | 22.65456872 | 2232.552332 | 2232.552332 | 0.92605153 | -0.033364847 | FALSE | TRUE | FALSE |
| TEX19 | 0.23558 | 0.82390652 | 1 | 0.23558003 | 0.23558003 | 0.23558003 | 0.27838357 | 0.831427469 | 1 | 0.278383574 | 0.278383574 | 0.278383574 | 0.846242567 | -0.072505133 | FALSE | TRUE | FALSE |
| PLSCR5 | 0.2354985 | 0.8239246 | 1 | 0.941928033 | 0.941928033 | 0.941928033 | 0.28184247 | 0.830636003 | 1 | 0.281842474 | 0.281842474 | 0.281842474 | 0.835567672 | -0.078018371 | FALSE | TRUE | FALSE |
| PIK3CA | 0.2354787 | 0.82394268 | 1 | 0.235478742 | 0.235478742 | 4.621367315 | 0.27952742 | 0.831199561 | 1 | 0.279527423 | 0.279527423 | 9.413082825 | 0.842417318 | -0.074472714 | FALSE | TRUE | FALSE |
| TMEM186 | 0.2354687 | 0.82396076 | 1 | 0.235468674 | 0.235468674 | 0.235468674 | 0.27843801 | 0.831392444 | 1 | 0.278438009 | 0.278438009 | 0.278438009 | 0.845677193 | -0.072795382 | FALSE | TRUE | FALSE |
| NAF1 | 0.235459 | 0.82397883 | 1 | 0.235459046 | 0.235459046 | 4.663222386 | 214.500995 | 0.012219826 | 0.0063 | 22.12333078 | 214.5009946 | 214.5009946 | 0.001097706 | -2.959513929 | FALSE | FALSE | FALSE |
| C1QTNF9B | 0.2354115 | 0.8239969 | 1 | 2.202424537 | 2.202424537 | 2.202424537 | 0.28810007 | 0.828669634 | 1 | 0.288100071 | 0.288100071 | 0.288100071 | 0.817116947 | -0.087715782 | FALSE | TRUE | FALSE |
| THRAP3 | 0.2353428 | 0.82401496 | 0.0327 | 0.235342775 | 0.235342775 | 7.922952929 | 0.26429055 | 0.835444748 | 1 | 0.264290555 | 0.264290555 | 5.185646574 | 0.890469867 | -0.050380772 | FALSE | TRUE | FALSE |
| FAM107B | 0.2352379 | 0.82403302 | 1 | 2.156906264 | 2.156906264 | 2.156906264 | 0.2868748 | 0.829066565 | 1 | 0.286874796 | 0.286874796 | 0.286874796 | 0.820001939 | -0.08618512 | FALSE | TRUE | FALSE |
| IPO13 | 0.2351019 | 0.82405108 | 1 | 0.235101876 | 0.235101876 | 4.612609969 | 2.5201165 | 0.446003325 | 0.5179 | 2.5201165 | 2.5201165 | 49.55697193 | 0.093290082 | -1.030164523 | FALSE | FALSE | FALSE |
| PCDHB12 | 0.2350823 | 0.82406913 | 0.6934 | 0.235082272 | 19.55188162 | 19.55188162 | 0.00659337 | 0.897668925 | 0.50775 | 0.057213132 | 0.057213132 | 1.124463682 | 35.6543437 | 1.552112447 | FALSE | TRUE | FALSE |
| INO80B | 0.2350651 | 0.82408718 | 1 | 0.235065141 | 0.235065141 | 4.663813103 | 0.27822677 | 0.83147998 | 1 | 0.278226775 | 0.278226775 | 0.278226775 | 0.84486887 | -0.073210692 | FALSE | TRUE | FALSE |
| OR56A4 | 0.2349368 | 0.82410523 | 1 | 0.234936844 | 0.234936844 | 0.234936844 | 0.27843276 | 0.831409958 | 1 | 0.278432765 | 0.278432765 | 0.278432765 | 0.843783038 | -0.073769209 | FALSE | TRUE | FALSE |
| RBCK1 | 0.2348918 | 0.82412327 | 0.5049 | 2.167490984 | 2.167490984 | 7.952636421 | 2.57582448 | 0.441753365 | 0.5096 | 2.57582448 | 2.57582448 | 5.507407024 | 0.09119092 | -1.040048404 | FALSE | FALSE | FALSE |
| NR4A1 | 0.2346856 | 0.82414131 | 0.81885 | 0.93824989 | 8.732136324 | 34.91894673 | 0.02611115 | 0.892103153 | 0.0178 | 0.026111154 | 0.026111154 | 0.51547024 | 8.98794349 | 0.953660333 | FALSE | TRUE | FALSE |
| PSMC3IP | 0.2346806 | 0.82415935 | 1 | 0.234680582 | 0.234680582 | 0.234680582 | 0.27758222 | 0.831619852 | 1 | 0.277582218 | 0.277582218 | 0.277582218 | 0.8454453 | -0.072914486 | FALSE | TRUE | FALSE |
| FOXN3 | 0.2346035 | 0.82417738 | 0.8777 | 2.201810085 | 21.20858269 | 21.20858269 | 0.08842079 | 0.878510468 | 1 | 0.088420794 | 0.088420794 | 1.751619607 | 2.653261556 | 0.423780064 | FALSE | TRUE | FALSE |
| OR2W3 | 0.2345232 | 0.82419541 | 0.00945 | 0.234523248 | 0.234523248 | 0.234523248 | 0.27751552 | 0.831672245 | 1 | 0.277515519 | 0.277515519 | 0.277515519 | 0.845081559 | -0.073101375 | FALSE | TRUE | FALSE |
| NEUROD2 | 0.2345213 | 0.82421343 | 1 | 0.937754686 | 3.750635252 | 18.54115678 | 0.28493085 | 0.829604869 | 1 | 0.284930846 | 0.284930846 | 5.641344848 | 0.823081324 | -0.084557253 | FALSE | TRUE | FALSE |
| KBTBD6 | 0.2344785 | 0.82423145 | 1 | 2.20025132 | 2.20025132 | 43.30458133 | 0.28725869 | 0.82894047 | 0.01805 | 0.287258693 | 0.287258693 | 5.664558526 | 0.816262488 | -0.088170161 | FALSE | TRUE | FALSE |
| PCDH9 | 0.234478 | 0.82424947 | 1 | 0.234477971 | 0.234477971 | 4.583353336 | 0.27835495 | 0.831444977 | 0.0275 | 0.278354951 | 0.278354951 | 5.455864146 | 0.8423704 | -0.074496902 | FALSE | TRUE | FALSE |
| ZFPM2 | 0.2343726 | 0.82426748 | 1 | 2.18519999 | 20.91051757 | 73.62498681 | 0.26953451 | 0.833945922 | 1 | 24.10729947 | 236.8981997 | 473.4284436 | 0.869545819 | -0.060707529 | FALSE | TRUE | FALSE |
| LHX8 | 0.2342977 | 0.8242855 | 1 | 0.234297696 | 0.234297696 | 4.653446064 | 5.52967944 | 0.2855421 | 0.0091 | 0.278183369 | 0.278183369 | 0.278183369 | 0.042370936 | -1.372931938 | FALSE | FALSE | FALSE |
| IPO9 | 0.2341789 | 0.8243035 | 1 | 0.234178875 | 0.234178875 | 4.591264418 | 0.27796916 | 0.831549944 | 1 | 0.27796916 | 0.27796916 | 5.462744044 | 0.842463513 | -0.074448899 | FALSE | TRUE | FALSE |
| TRAPPC6A | 0.2341339 | 0.82432151 | 1 | 0.234133932 | 0.234133932 | 0.234133932 | 0.27037614 | 0.833622423 | 1 | 0.270376142 | 0.270376142 | 0.270376142 | 0.865956333 | -0.062504007 | FALSE | TRUE | FALSE |
| PCSK2 | 0.2340732 | 0.82433951 | 0.0188 | 0.234073245 | 0.234073245 | 4.609078344 | 1.09966439 | 0.57858326 | 0.5097 | 1.099664392 | 1.099664392 | 9.305593793 | 0.212858802 | -0.671908387 | FALSE | FALSE | FALSE |
| ARHGEF3 | 0.2340472 | 0.8243575 | 1 | 2.164917657 | 2.164917657 | 7.917295166 | 0.28616851 | 0.829282288 | 1 | 0.286168509 | 0.286168509 | 5.665815403 | 0.817865062 | -0.087318344 | FALSE | TRUE | FALSE |
| CYP7A1 | 0.2340132 | 0.82437549 | 0.757 | 0.234013246 | 0.935934199 | 4.635848571 | 0.08248643 | 0.879678697 | 1 | 0.329837269 | 0.329837269 | 1.635937706 | 2.836990874 | 0.452857939 | FALSE | TRUE | FALSE |
| SMAD1 | 0.2339526 | 0.82439348 | 0.87445 | 0.935269286 | 3.739848712 | 3.739848712 | 0.08381548 | 0.879393162 | 0.51005 | 0.335019524 | 0.335019524 | 1.662580737 | 2.791281337 | 0.445803612 | FALSE | TRUE | FALSE |
| ITM2C | 0.2339282 | 0.82441147 | 1 | 2.193397641 | 2.193397641 | 2.193397641 | 0.28658318 | 0.829174493 | 1 | 0.286583176 | 0.286583176 | 0.286583176 | 0.816266165 | -0.088168205 | FALSE | TRUE | FALSE |
| NUF2 | 0.2339249 | 0.82442945 | 1 | 2.150310992 | 2.150310992 | 4.640518325 | 0.28570097 | 0.829425807 | 1 | 0.285700966 | 0.285700966 | 0.285700966 | 0.818775471 | -0.086835177 | FALSE | TRUE | FALSE |
| C7orf25 | 0.2338164 | 0.82444742 | 1 | 0.233816423 | 0.233816423 | 0.233816423 | 0.26856951 | 0.834183488 | 1 | 2.479079181 | 2.479079181 | 5.338213184 | 0.870599293 | -0.06018169 | FALSE | TRUE | FALSE |
| PARP1 | 0.2335045 | 0.8244654 | 0.51575 | 0.233504459 | 0.233504459 | 4.570886731 | 0.68576585 | 0.672497367 | 0.87985 | 6.424993161 | 61.74188042 | 126.09206 | 0.340501731 | -0.467880676 | FALSE | TRUE | FALSE |
| CCNB1 | 0.2334798 | 0.82448337 | 1 | 0.233479767 | 0.233479767 | 4.633910158 | 0.27665551 | 0.831933719 | 1 | 0.276655509 | 0.276655509 | 0.276655509 | 0.843936809 | -0.073690071 | FALSE | TRUE | FALSE |
| TRIP13 | 0.2334749 | 0.82450134 | 1 | 0.933616172 | 0.933616172 | 4.62692387 | 0.28053899 | 0.830953447 | 1 | 0.280538994 | 0.280538994 | 5.565720665 | 0.832236846 | -0.07975306 | FALSE | TRUE | FALSE |
| PCF11 | 0.2333626 | 0.8245193 | 1 | 0.233362579 | 0.233362579 | 7.824615144 | 0.27716785 | 0.831759492 | 1 | 0.277167849 | 0.277167849 | 9.308864703 | 0.841954 | -0.074711635 | FALSE | TRUE | FALSE |
| TEAD2 | 0.233362 | 0.82453726 | 1 | 0.233361995 | 0.233361995 | 4.614627444 | 0.27666539 | 0.831898902 | 1 | 0.276665388 | 0.276665388 | 5.478352003 | 0.843480988 | -0.073924702 | FALSE | TRUE | FALSE |
| CLPTM1 | 0.2332371 | 0.82455522 | 1 | 0.233237142 | 0.233237142 | 4.582440288 | 0.27699534 | 0.831811798 | 1 | 0.276995343 | 0.276995343 | 5.453723979 | 0.8420255 | -0.074674756 | FALSE | TRUE | FALSE |
| HTR1A | 0.2331762 | 0.82457317 | 1 | 0.233176151 | 0.233176151 | 4.604940069 | 0.26537127 | 0.835227514 | 1 | 2.483007937 | 9.930396911 | 20.99124545 | 0.878678964 | -0.056169771 | FALSE | TRUE | FALSE |
| DRG2 | 0.2331359 | 0.82459112 | 1 | 0.233135948 | 0.233135948 | 4.621626791 | 0.27637188 | 0.832003309 | 1 | 0.276371881 | 0.276371881 | 5.484531906 | 0.843558858 | -0.07388461 | FALSE | TRUE | FALSE |
| SERPING1 | 0.2330875 | 0.82460907 | 0.9958 | 2108.232267 | 232227.4732 | 2523877.799 | 0.28571174 | 0.82940788 | 1 | 2608.080117 | 288657.7726 | 537966.0077 | 0.815813668 | -0.088409023 | FALSE | TRUE | FALSE |
| TNFSF18 | 0.2330868 | 0.82462701 | 1 | 2.163896774 | 2.163896774 | 2.163896774 | 0.28525841 | 0.829497477 | 0.0036 | 0.285258414 | 0.285258414 | 0.285258414 | 0.817107444 | -0.087720833 | FALSE | TRUE | FALSE |
| SPINK6 | 0.2330517 | 0.82464495 | 0.00135 | 0.233051718 | 0.233051718 | 0.233051718 | 0.27637389 | 0.831985917 | 1 | 0.276373891 | 0.276373891 | 0.276373891 | 0.843247954 | -0.074044704 | FALSE | TRUE | FALSE |
| SNX7 | 0.2329305 | 0.82466288 | 0.0093 | 0.232930505 | 0.232930505 | 0.232930505 | 0.27598154 | 0.832072845 | 0.00885 | 0.27598154 | 0.27598154 | 0.27598154 | 0.844007556 | -0.073653665 | FALSE | TRUE | FALSE |
| CHST11 | 0.2329255 | 0.82468081 | 1 | 0.232925459 | 0.232925459 | 4.615445035 | 0.27678496 | 0.831881488 | 1 | 0.276784958 | 0.276784958 | 5.490624215 | 0.84153944 | -0.074925525 | FALSE | TRUE | FALSE |
| CHMP2A | 0.2328908 | 0.82469874 | 1 | 8.733865542 | 84.06392295 | 84.06392295 | 0.29789573 | 0.826025256 | 1 | 0.297895733 | 0.297895733 | 0.297895733 | 0.78178644 | -0.106911867 | FALSE | TRUE | FALSE |
| TTYH2 | 0.2326726 | 0.82471667 | 1 | 0.232672625 | 0.232672625 | 4.590615411 | 0.2758121 | 0.83212496 | 1 | 0.275812104 | 0.275812104 | 5.450597243 | 0.843591059 | -0.073868032 | FALSE | TRUE | FALSE |
| STXBP1 | 0.2326089 | 0.82473459 | 1 | 0.930165329 | 0.930165329 | 7.873940833 | 0.27349181 | 0.832747869 | 0.75615 | 1.093682795 | 4.374687115 | 21.5952647 | 0.850515124 | -0.070317959 | FALSE | TRUE | FALSE |
| USP7 | 0.2324633 | 0.8247525 | 1 | 2041.941657 | 222455.0597 | 2400313.314 | 0.28482069 | 0.829658514 | 1 | 2533.349971 | 278012.1214 | 882107.0031 | 0.816174123 | -0.088217179 | FALSE | TRUE | FALSE |
| TSPAN9 | 0.2324354 | 0.82477042 | 1 | 0.23243537 | 0.23243537 | 0.23243537 | 1.09166503 | 0.579622471 | 0.499 | 0.272978773 | 1.091665032 | 1.091665032 | 0.212918215 | -0.671787184 | FALSE | FALSE | FALSE |
| AP3M1 | 0.2324306 | 0.82478833 | 0.5083 | 2.165617112 | 2.165617112 | 4.611295885 | 0.28472711 | 0.829712124 | 1 | 0.284727112 | 0.284727112 | 0.284727112 | 0.816327687 | -0.088135473 | FALSE | TRUE | FALSE |
| APAF1 | 0.2323756 | 0.82480624 | 0.14655 | 0.23237563 | 2.119276139 | 4.549790704 | 0.09928539 | 0.876149906 | 0.15175 | 0.099285392 | 0.397112708 | 1.948878278 | 2.340481573 | 0.369305226 | FALSE | TRUE | FALSE |
| HUNK | 0.2323358 | 0.82482414 | 1 | 0.232335757 | 0.232335757 | 4.567624936 | 0.27586891 | 0.832107592 | 0.0191 | 0.275868906 | 0.275868906 | 5.43458205 | 0.842196245 | -0.074586699 | FALSE | TRUE | FALSE |
| NCAPG | 0.2323089 | 0.82484204 | 0.51215 | 0.232308922 | 0.232308922 | 4.572149539 | 0.04032563 | 0.888903126 | 1 | 0.354034715 | 0.354034715 | 1.362402978 | 5.760825138 | 0.760484693 | FALSE | TRUE | FALSE |
| NARF | 0.2322102 | 0.82485993 | 0.0134 | 0.232210181 | 0.232210181 | 1.149918987 | 0.08156954 | 0.879908065 | 1 | 0.081569535 | 0.081569535 | 1.617365464 | 2.84677582 | 0.454353268 | FALSE | TRUE | FALSE |
| TIA1 | 0.2321856 | 0.82487783 | 1 | 0.232185642 | 0.232185642 | 4.609686375 | 0.27545432 | 0.832194397 | 1 | 0.275454319 | 0.275454319 | 0.275454319 | 0.842918867 | -0.074214225 | FALSE | TRUE | FALSE |
| GRP | 0.2321836 | 0.82489571 | 1 | 0.232183643 | 0.232183643 | 0.232183643 | 0.24743848 | 0.84005314 | 0.00235 | 0.247438477 | 0.247438477 | 0.247438477 | 0.93834898 | -0.027635614 | FALSE | TRUE | FALSE |
| NDC80 | 0.2320757 | 0.8249136 | 1 | 0.232075656 | 0.232075656 | 4.591723517 | 0.27522273 | 0.832263777 | 1 | 0.275222725 | 0.275222725 | 5.45243518 | 0.843228536 | -0.074054705 | FALSE | TRUE | FALSE |
| OR5T1 | 0.2320355 | 0.82493148 | 1 | 2.124053625 | 2.124053625 | 2.124053625 | 215.745727 | 0.011892344 | 0.12955 | 22.33418197 | 215.7457266 | 215.7457266 | 0.001075505 | -2.968387713 | FALSE | FALSE | FALSE |
| RALBP1 | 0.2320026 | 0.82494936 | 1 | 2.173417064 | 20.89683357 | 42.89393566 | 2.50682177 | 0.446195268 | 0.50415 | 2.506821765 | 2.506821765 | 9.031366981 | 0.092548501 | -1.033630612 | FALSE | FALSE | FALSE |

| | | | | | | | | | | | | | | | | | |
|---|---|---|---|---|---|---|---|---|---|---|---|---|---|---|---|---|---|
| HYOU1 | 0.231977 | 0.82496723 | 0.03055 | 0.23197704 | 0.23197704 | 4.535997234 | 0.27544115 | 0.832211747 | 1 | 0.275441145 | 0.275441145 | 5.400359242 | 0.84220184 | -0.074583814 | FALSE | TRUE | FALSE |
| DMWD | 0.2319058 | 0.8249851 | 0.75975 | 2.169772137 | 20.83551898 | 42.69605856 | 2.66771433 | 0.436259209 | 0.5104 | 2.667714327 | 2.667714327 | 9.633398078 | 0.086930536 | -1.060827643 | FALSE | FALSE | FALSE |
| ANGEL1 | 0.2318829 | 0.82500297 | 0.9387 | 8.560986972 | 196.8855997 | 787.1935564 | 3.5782917 | 0.379686862 | 0.25555 | 0.384465926 | 3.578291699 | 7.601943907 | 0.064802669 | -1.188407103 | FALSE | FALSE | FALSE |
| MMP15 | 0.23177 | 0.82502083 | 1 | 0.231770035 | 0.231770035 | 4.552513896 | 0.26683838 | 0.83467346 | 1 | 2.485501749 | 2.485501749 | 48.92626692 | 0.868578342 | -0.061191004 | FALSE | TRUE | FALSE |
| SOX2 | 0.231757 | 0.82503869 | 1 | 21.25270004 | 2156.753504 | 2156.753504 | 1.15062359 | 0.572363588 | 0.75185 | 1.150623591 | 4.602257624 | 5.699475006 | 0.201418627 | -0.69590037 | FALSE | FALSE | FALSE |
| ZNF213 | 0.2315963 | 0.82505654 | 1 | 0.231596286 | 0.231596286 | 4.562852468 | 22.6662642 | 0.120356061 | 0.2536 | 2.388600015 | 22.66626416 | 47.14406938 | 0.010217665 | -1.990648355 | FALSE | FALSE | FALSE |
| LITAF | 0.231528 | 0.8250744 | 0.0055 | 0.231528037 | 0.231528037 | 0.231528037 | 0.27532171 | 0.832229094 | 0.0045 | 0.275321714 | 0.275321714 | 0.275321714 | 0.840936351 | -0.075236874 | FALSE | TRUE | FALSE |
| TRPC4AP | 0.2315183 | 0.82509225 | 1 | 2.138933397 | 2.138933397 | 42.13434049 | 0.28340526 | 0.83015757 | 1 | 0.283405259 | 0.283405259 | 5.592853672 | 0.816916209 | -0.087822487 | FALSE | TRUE | FALSE |
| ATXN7L2 | 0.2314699 | 0.82511009 | 1 | 0.231469879 | 0.231469879 | 4.548829432 | 0.2504905 | 0.839292286 | 1 | 0.250490497 | 0.250490497 | 4.932935755 | 0.924066509 | -0.03429677 | FALSE | TRUE | FALSE |
| DMAP1 | 0.2314322 | 0.82512793 | 0.01645 | 0.231432177 | 0.231432177 | 4.569198982 | 0.27480836 | 0.832333103 | 0.0167 | 0.274808358 | 0.274808358 | 5.433965734 | 0.842158438 | -0.074606196 | FALSE | TRUE | FALSE |
| EIF3L | 0.231387 | 0.82514577 | 1 | 0.231387008 | 0.231387008 | 4.572331884 | 2.47500234 | 0.448299293 | 0.5069 | 2.475002341 | 2.475002341 | 5.271093895 | 0.093489612 | -1.029236643 | FALSE | FALSE | FALSE |
| NGLY1 | 0.2311164 | 0.82516361 | 0.9383 | 2.086325256 | 183.3549341 | 183.3549341 | 0.37607427 | 0.800414387 | 0.75675 | 0.376074271 | 3.430219437 | 7.448906892 | 0.614549799 | -0.211442919 | FALSE | TRUE | FALSE |
| CEP19 | 0.2310947 | 0.82518144 | 0.005 | 0.231094735 | 0.231094735 | 0.231094735 | 0.27418068 | 0.832488881 | 0.0036 | 0.274180682 | 0.274180682 | 0.274180682 | 0.842855643 | -0.074246801 | FALSE | TRUE | FALSE |
| OR4C11 | 0.2310804 | 0.82519926 | 1 | 2.115226155 | 2.115226155 | 2.115226155 | 0.28303029 | 0.830246375 | 1 | 0.283030286 | 0.283030286 | 0.283030286 | 0.816451288 | -0.088069722 | FALSE | TRUE | FALSE |
| MTHFD2 | 0.23098 | 0.82521709 | 1 | 0.230979998 | 0.230979998 | 0.230979998 | 2.47440584 | 0.44848981 | 0.50605 | 2.474405837 | 2.474405837 | 2.474405837 | 0.093347661 | -1.029896558 | FALSE | FALSE | FALSE |
| DLGAP3 | 0.2309486 | 0.82523491 | 1 | 2.181275672 | 2.181275672 | 42.51910229 | 0.28360264 | 0.830086462 | 0.02905 | 0.283602641 | 0.283602641 | 5.5450969 | 0.814338806 | -0.089194869 | FALSE | TRUE | FALSE |
| SLC17A2 | 0.2308787 | 0.82525273 | 1 | 0.923378059 | 0.923378059 | 4.581358605 | 0.27739334 | 0.831742049 | 1 | 0.277393342 | 0.277393342 | 0.277393342 | 0.832315165 | -0.079712192 | FALSE | TRUE | FALSE |
| ARSG | 0.2307944 | 0.82527054 | 0.75405 | 0.230794389 | 2.132847097 | 4.560576533 | 0.04172615 | 0.888584093 | 0.0127 | 0.041726153 | 0.041726153 | 0.825711883 | 5.53116869 | 0.742816904 | FALSE | TRUE | FALSE |
| NCR3 | 0.2307849 | 0.82528835 | 1 | 0.230784886 | 0.230784886 | 0.230784886 | 0.27344125 | 0.832782339 | 1 | 0.273441254 | 0.273441254 | 0.273441254 | 0.844001709 | -0.073656674 | FALSE | TRUE | FALSE |
| UBR3 | 0.2307294 | 0.82530616 | 1 | 0.230729431 | 0.230729431 | 7.754727408 | 0.25990575 | 0.836785615 | 1 | 4.15931737 | 38.7082512 | 139.9861657 | 0.887742682 | -0.051712899 | FALSE | TRUE | FALSE |
| HTR7 | 0.2307192 | 0.82532396 | 1 | 0.230719231 | 0.230719231 | 4.558435701 | 0.27395798 | 0.832592575 | 0.0119 | 0.273957982 | 0.273957982 | 5.420621845 | 0.842170137 | -0.074600162 | FALSE | TRUE | FALSE |
| SFTPA1 | 0.2305114 | 0.82534176 | 0.7524 | 2.152708277 | 20.63319458 | 20.63319458 | 0.28273302 | 0.830317352 | 1 | 2.647985722 | 2.647985722 | 2.647985722 | 0.815297095 | -0.088684105 | FALSE | TRUE | FALSE |
| MRPS18C | 0.2304628 | 0.82535955 | 1 | 0.230462811 | 2.107331056 | 2.107331056 | 2.35496211 | 0.459503271 | 0.87825 | 2.354962108 | 213.8324314 | 213.8324314 | 0.097862641 | -1.009383068 | FALSE | FALSE | FALSE |
| ATP6V1H | 0.2304599 | 0.82537735 | 0.5086 | 2.137001646 | 2.137001646 | 4.563912182 | 0.28240692 | 0.830459125 | 1 | 0.282406923 | 0.282406923 | 5.599253703 | 0.816056258 | -0.088279901 | FALSE | TRUE | FALSE |
| FLRT1 | 0.2303443 | 0.82539514 | 0.0259 | 0.230344277 | 0.230344277 | 4.515436362 | 0.27411291 | 0.83252346 | 1 | 0.274112914 | 0.274112914 | 5.386278393 | 0.84032625 | -0.07555207 | FALSE | TRUE | FALSE |
| CAND1 | 0.2302937 | 0.82541292 | 1 | 0.230293669 | 0.230293669 | 4.543859739 | 0.27416231 | 0.832506173 | 0.0259 | 0.27416231 | 0.27416231 | 5.38399394 | 0.839990254 | -0.075725753 | FALSE | TRUE | FALSE |
| PGAP2 | 0.2302652 | 0.8254307 | 1 | 0.23026516 | 0.23026516 | 4.566320502 | 22.0517218 | 0.123407652 | 0.2483 | 9.344716526 | 88.20580956 | 88.20580956 | 0.010442049 | -1.981214273 | FALSE | FALSE | FALSE |
| OR10V1 | 0.230236 | 0.82544848 | 1 | 2.109645254 | 2.109645254 | 2.109645254 | 0.28150072 | 0.830812506 | 0.0068 | 0.28150072 | 0.28150072 | 0.28150072 | 0.817887825 | -0.087306257 | FALSE | TRUE | FALSE |
| WDR47 | 0.2302084 | 0.82546625 | 1 | 0.230208441 | 0.230208441 | 4.529824685 | 0.26537225 | 0.835210782 | 1 | 2.482324407 | 2.482324407 | 48.93978223 | 0.867492532 | -0.061734255 | FALSE | TRUE | FALSE |
| PSTPIP1 | 0.2301476 | 0.82548403 | 1 | 0.230147629 | 0.230147629 | 4.558962121 | 0.27319799 | 0.832885661 | 1 | 0.273197985 | 0.273197985 | 5.417961628 | 0.842420665 | -0.074470988 | FALSE | TRUE | FALSE |
| ZNF503 | 0.2300918 | 0.82550179 | 0.9996 | 217941.0066 | 25835884.98 | 291138103.8 | 0.28233765 | 0.830476829 | 1 | 25811.78423 | 32672589.97 | 369351444.4 | 0.814952709 | -0.088867592 | FALSE | TRUE | FALSE |
| DDX23 | 0.2298398 | 0.82551956 | 1 | 0.229839811 | 0.229839811 | 7.743814972 | 0.27324225 | 0.83286845 | 0.0259 | 0.273242249 | 0.273242249 | 5.362316042 | 0.84115766 | -0.075122596 | FALSE | TRUE | FALSE |
| ZNF750 | 0.229834 | 0.82553732 | 0.5104 | 2.123646805 | 2.123646805 | 41.75325267 | 0.28158561 | 0.830759595 | 1 | 0.281585608 | 0.281585608 | 5.547579232 | 0.816213608 | -0.088196169 | FALSE | TRUE | FALSE |
| CDKN2A | 0.2298157 | 0.82555507 | 0.0083 | 0.22981575 | 0.22981575 | 0.22981575 | 0.27009813 | 0.833758787 | 1 | 1.080209544 | 1.080209544 | 1.080209544 | 0.850860204 | -0.070141789 | FALSE | TRUE | FALSE |
| ANKIB1 | 0.229811 | 0.82557283 | 1 | 0.229810977 | 0.229810977 | 4.505025057 | 0.27029384 | 0.833690634 | 1 | 1.081103766 | 1.081103766 | 21.24370856 | 0.850226458 | -0.070465384 | FALSE | TRUE | FALSE |
| CREBBP | 0.2297448 | 0.82559058 | 0.02395 | 0.005970579 | 0.229744755 | 1.08969496 | 0.00752065 | 0.897442397 | 0.5371 | 0.007520652 | 0.035891405 | 0.251148019 | 30.54851695 | 1.484990131 | FALSE | TRUE | FALSE |
| ACAP2 | 0.2297292 | 0.82560832 | 1 | 0.22972923 | 0.22972923 | 4.531182998 | 0.27333613 | 0.832799569 | 1 | 0.273336132 | 0.273336132 | 5.400239281 | 0.840464187 | -0.075480788 | FALSE | TRUE | FALSE |
| CMTR1 | 0.2296597 | 0.82562607 | 0.0268 | 0.229659705 | 0.229659705 | 4.508506557 | 0.27317162 | 0.832902869 | 1 | 0.273171624 | 0.273171624 | 5.374605523 | 0.840715818 | -0.075350781 | FALSE | TRUE | FALSE |
| DLGAP1 | 0.2296489 | 0.8256438 | 1 | 0.229648889 | 0.229648889 | 4.470989731 | 0.27332924 | 0.832816795 | 1 | 0.273329238 | 0.273329238 | 5.338428902 | 0.840191453 | -0.075621741 | FALSE | TRUE | FALSE |
| OR13D1 | 0.2295632 | 0.82566154 | 1 | 0.229563217 | 0.229563217 | 0.229563217 | 0.27218229 | 0.833109083 | 1 | 0.272182291 | 0.272182291 | 0.272182291 | 0.843417168 | -0.073957563 | FALSE | TRUE | FALSE |
| GABRG2 | 0.2295479 | 0.82567927 | 1 | 0.229547898 | 0.229547898 | 4.547667681 | 2.48256397 | 0.447726795 | 0.50775 | 2.482563967 | 2.482563967 | 5.250158288 | 0.092464042 | -1.034027128 | FALSE | FALSE | FALSE |
| FGFBP1 | 0.2294842 | 0.825697 | 1 | 2.096519905 | 2.096519905 | 2.096519905 | 2.49634264 | 0.44677043 | 0.5004 | 2.496342638 | 2.496342638 | 2.496342638 | 0.091928159 | -1.036551435 | FALSE | FALSE | FALSE |
| TPX2 | 0.2293628 | 0.82571472 | 1 | 0.229362768 | 0.229362768 | 4.528035161 | 0.27274308 | 0.833074747 | 1 | 0.27274308 | 0.27274308 | 5.392805249 | 0.840948077 | -0.075230818 | FALSE | TRUE | FALSE |
| LIFR | 0.229319 | 0.82573244 | 0.0286 | 0.229318952 | 0.229318952 | 4.496866832 | 0.03946468 | 0.889077891 | 0.5086 | 0.352292459 | 0.352292459 | 6.924572698 | 5.810738432 | 0.764231326 | FALSE | TRUE | FALSE |
| MBD3L1 | 0.2292854 | 0.82575016 | 0.0049 | 0.229285402 | 0.229285402 | 0.229285402 | 0.27192705 | 0.833229149 | 1 | 0.27192705 | 0.27192705 | 0.27192705 | 0.843187177 | -0.074076007 | FALSE | TRUE | FALSE |
| MBOAT2 | 0.2292425 | 0.82576787 | 0.5072 | 0.916778305 | 0.916778305 | 4.54680963 | 4.1818278 | 0.345616836 | 0.50335 | 4.181827796 | 39.14202028 | 39.14202028 | 0.054818726 | -1.261071062 | FALSE | FALSE | FALSE |
| WDR18 | 0.2289227 | 0.82578559 | 0.01315 | 0.228922707 | 0.228922707 | 4.530605616 | 0.27193517 | 0.833212008 | 1 | 0.27193517 | 0.27193517 | 5.388630586 | 0.841828245 | -0.074776507 | FALSE | TRUE | FALSE |
| PPM1M | 0.2289213 | 0.82580329 | 1 | 0.228921278 | 0.228921278 | 0.228921278 | 22.2270593 | 0.121577974 | 0.25205 | 2.352610955 | 22.22705928 | 22.22705928 | 0.010299216 | -1.987195847 | FALSE | FALSE | FALSE |
| CNTNAP2 | 0.2288789 | 0.825821 | 0.38615 | 0.057271387 | 1.114572251 | 7.705416728 | 0.15278455 | 0.863885613 | 0.4065 | 0.152784551 | 5.532379422 | 22.12020072 | 1.498050221 | 0.175526373 | FALSE | TRUE | FALSE |
| KCNK1 | 0.228845 | 0.8258387 | 1 | 20.44575916 | 2021.586489 | 2021.586489 | 0.24035477 | 0.841955874 | 1 | 2110.130501 | 225800.9849 | 2436197.119 | 0.952113433 | -0.021311308 | FALSE | TRUE | FALSE |
| PNISR | 0.2288302 | 0.8258564 | 1 | 0.228830245 | 0.228830245 | 4.503235261 | 0.27203474 | 0.833177714 | 1 | 0.272034738 | 0.272034738 | 5.363803675 | 0.841180237 | -0.07511094 | FALSE | TRUE | FALSE |
| HNF1A | 0.2287487 | 0.82587409 | 0.721 | 0.913789506 | 14.59307577 | 58.33911932 | 0.01407494 | 0.895404562 | 0.93885 | 0.056224512 | 0.89785267 | 1.10955985 | 16.25219368 | 1.210911989 | FALSE | TRUE | FALSE |
| CXCR6 | 0.228713 | 0.82589178 | 1 | 0.228713025 | 0.228713025 | 0.228713025 | 0.27167331 | 0.833314803 | 1 | 0.271673312 | 0.271673312 | 0.271673312 | 0.841867843 | -0.074756079 | FALSE | TRUE | FALSE |
| SHISA4 | 0.2287124 | 0.82590946 | 0.75245 | 2.112482257 | 20.02579122 | 20.02579122 | 201.840492 | 0.013327909 | 0.1908 | 201.8404924 | 2012.656981 | 20559.24662 | 0.001133134 | -2.945718647 | FALSE | FALSE | FALSE |
| FURIN | 0.2286248 | 0.82592715 | 0.7519 | 0.228624762 | 0.914125342 | 4.508695275 | 1.25490384 | 0.560661813 | 0.69015 | 1.254903842 | 20.07991989 | 24.63175743 | 0.182185084 | -0.739487183 | FALSE | FALSE | FALSE |
| USP4 | 0.2286231 | 0.82594483 | 0.5055 | 0.228623146 | 0.228623146 | 4.485117778 | 0.34044014 | 0.812009356 | 0.26215 | 0.340440136 | 3.106720772 | 6.694101357 | 0.671551684 | -0.172920558 | FALSE | TRUE | FALSE |
| C5orf22 | 0.2285703 | 0.8259625 | 1 | 0.228570259 | 0.228570259 | 0.228570259 | 0.2714355 | 0.833400367 | 1 | 0.271435503 | 0.271435503 | 0.271435503 | 0.84207945 | -0.074646931 | FALSE | TRUE | FALSE |
| MON1B | 0.228552 | 0.82598017 | 1 | 0.228551979 | 0.228551979 | 4.499490242 | 0.27209031 | 0.833160561 | 1 | 0.272090307 | 0.272090307 | 5.366719523 | 0.839985744 | -0.075728085 | FALSE | TRUE | FALSE |
| GAS2 | 0.2284771 | 0.82599784 | 1 | 0.228477108 | 0.228477108 | 0.228477108 | 0.2719772 | 0.833194863 | 0.0082 | 0.271977196 | 0.271977196 | 0.271977196 | 0.840059798 | -0.075689798 | FALSE | TRUE | FALSE |
| CHID1 | 0.2284573 | 0.8260155 | 1 | 2.097921409 | 2.097921409 | 4.525413255 | 2.50040963 | 0.446578803 | 0.5057 | 2.500409627 | 2.500409627 | 5.378986554 | 0.091367964 | -1.039206054 | FALSE | FALSE | FALSE |
| UNC119B | 0.2283228 | 0.82603316 | 1 | 0.91301095 | 3.651836541 | 3.651836541 | 0.27802562 | 0.831532459 | 1 | 0.278025621 | 0.278025621 | 0.278025621 | 0.821229203 | -0.085535616 | FALSE | TRUE | FALSE |
| EXOSC4 | 0.2283024 | 0.82605082 | 1 | 0.228302437 | 0.228302437 | 0.228302437 | 0.27174475 | 0.83329768 | 1 | 0.271744753 | 0.271744753 | 0.271744753 | 0.840135584 | -0.07565062 | FALSE | TRUE | FALSE |
| ABHD16A | 0.2282964 | 0.82606848 | 1 | 0.913162019 | 3.65346746 | 18.05342992 | 0.27751099 | 0.831689702 | 1 | 0.277510989 | 0.277510989 | 5.494041211 | 0.822657025 | -0.084781189 | FALSE | TRUE | FALSE |
| SDHAF2 | 0.2282837 | 0.82608612 | 1 | 0.228283684 | 0.228283684 | 0.228283684 | 0.27140127 | 0.833434567 | 1 | 0.27140127 | 0.27140127 | 0.27140127 | 0.841129754 | -0.075137004 | FALSE | TRUE | FALSE |
| PIGR | 0.2282406 | 0.82610377 | 0.0227 | 0.228240623 | 0.228240623 | 4.487931637 | 0.27161339 | 0.833349039 | 1 | 0.27161339 | 0.27161339 | 5.351606105 | 0.840314329 | -0.075558231 | FALSE | TRUE | FALSE |
| IGF1 | 0.2282374 | 0.82612141 | 0.0042 | 0.228237377 | 0.228237377 | 0.228237377 | 0.27177938 | 0.833246287 | 1 | 0.271779385 | 0.271779385 | 0.271779385 | 0.839789143 | -0.075829744 | FALSE | TRUE | FALSE |
| TONSL | 0.2280643 | 0.82613905 | 0.27375 | 0.05704619 | 0.228064295 | 1.921469463 | 0.01173977 | 0.89619766 | 1 | 0.104321607 | 0.104321607 | 2.033985364 | 19.42664574 | 1.288397821 | FALSE | TRUE | FALSE |
| TMEM19 | 0.2278585 | 0.82615669 | 1 | 0.227858507 | 0.227858507 | 0.227858507 | 0.26803249 | 0.834318941 | 1 | 1.071977079 | 1.071977079 | 1.071977079 | 0.850115242 | -0.070522197 | FALSE | TRUE | FALSE |
| LRRC8B | 0.2278176 | 0.82617432 | 1 | 0.227817649 | 0.227817649 | 4.491439948 | 0.27104403 | 0.833520007 | 0.01705 | 0.271044027 | 0.271044027 | 5.352813048 | 0.84051898 | -0.075452475 | FALSE | TRUE | FALSE |
| RBPMS | 0.2277722 | 0.82619195 | 1 | 0.227772195 | 0.227772195 | 0.227772195 | 0.24387512 | 0.840885255 | 1 | 0.243875117 | 0.243875117 | 0.243875117 | 0.933970623 | -0.029666784 | FALSE | TRUE | FALSE |
| BNC1 | 0.2277524 | 0.82620958 | 1 | 0.227752389 | 0.227752389 | 4.472463603 | 0.27123694 | 0.833468753 | 0.0227 | 0.271236944 | 0.271236944 | 5.338014534 | 0.839680557 | -0.075885903 | FALSE | TRUE | FALSE |
| CDC123 | 0.2277403 | 0.8262272 | 1 | 0.227740254 | 0.227740254 | 4.523681021 | 1.0716462 | 0.580887054 | 0.5058 | 1.071646199 | 1.071646199 | 1.071646199 | 0.212514404 | -0.672611628 | FALSE | FALSE | FALSE |
| GFI1B | 0.227423 | 0.82624482 | 1 | 0.227422974 | 0.227422974 | 4.507826316 | 0.2702348 | 0.833724717 | 1 | 0.270234797 | 0.270234797 | 0.270234797 | 0.84157546 | -0.074906937 | FALSE | TRUE | FALSE |

| | | | | | | | | | | | | | | | | | |
|---|---|---|---|---|---|---|---|---|---|---|---|---|---|---|---|---|---|
| RAB40B | 0.2274035 | 0.82626244 | 1 | 0.227403477 | 0.227403477 | 4.507925097 | 0.27042752 | 0.833605361 | 1 | 0.270427524 | 0.270427524 | 5.366501887 | 0.840903596 | -0.07525379 | FALSE | TRUE | FALSE |
| UBASH3B | 0.2273868 | 0.82628005 | 1 | 0.227386803 | 0.227386803 | 4.483909762 | 0.26241924 | 0.836010246 | 0.50505 | 2.448861209 | 2.448861209 | 8.8807272 | 0.866502023 | -0.062230419 | FALSE | TRUE | FALSE |
| KIF5C | 0.227318 | 0.82629766 | 0.9396 | 0.908911563 | 14.54195204 | 30.68771311 | 1.29115141 | 0.555861824 | 0.50895 | 1.291151409 | 5.164260885 | 25.44238671 | 0.176058381 | -0.754343297 | FALSE | FALSE | FALSE |
| NEIL3 | 0.2271952 | 0.82631526 | 0.75715 | 0.227195204 | 2.029193022 | 4.487744582 | 3.13199485 | 0.40463276 | 0.2558 | 0.347255246 | 3.131994848 | 6.869468735 | 0.072540095 | -1.139421879 | FALSE | FALSE | FALSE |
| SYT5 | 0.2271406 | 0.82633287 | 1 | 0.227140621 | 0.227140621 | 4.502693759 | 1.06938199 | 0.581230975 | 0.01035 | 1.069381992 | 1.069381992 | 5.306138912 | 0.212403634 | -0.672838058 | FALSE | FALSE | FALSE |
| MRAP2 | 0.2270752 | 0.82635046 | 0.00515 | 0.227075186 | 0.227075186 | 0.227075186 | 0.27034571 | 0.833656536 | 1 | 0.270345711 | 0.270345711 | 0.270345711 | 0.839943735 | -0.075749805 | FALSE | TRUE | FALSE |
| OR14C36 | 0.2270541 | 0.82636806 | 1 | 0.227054071 | 0.227054071 | 0.227054071 | 0.26961271 | 0.833911929 | 1 | 0.269612711 | 0.269612711 | 0.269612711 | 0.842148984 | -0.074611071 | FALSE | TRUE | FALSE |
| ULK3 | 0.2269967 | 0.82638565 | 1 | 0.226996724 | 0.226996724 | 4.505122593 | 22.4525976 | 0.120965948 | 0.00775 | 2.356817192 | 22.45259763 | 22.45259763 | 0.010110043 | -1.995247004 | FALSE | FALSE | FALSE |
| RFTN2 | 0.2269812 | 0.82640324 | 1 | 0.226981205 | 0.226981205 | 4.49993703 | 0.26972451 | 0.833860913 | 1 | 0.269724509 | 0.269724509 | 5.352952006 | 0.841529775 | -0.074930513 | FALSE | TRUE | FALSE |
| HS3ST4 | 0.2269803 | 0.82642082 | 0.50765 | 2.141725664 | 2.141725664 | 4.503987321 | 0.27929957 | 0.831287284 | 1 | 0.279299574 | 0.279299574 | 0.279299574 | 0.812676817 | -0.090082129 | FALSE | TRUE | FALSE |
| EID3 | 0.2269771 | 0.8264384 | 1 | 2.089870238 | 19.74927432 | 19.74927432 | 2.57367105 | 0.441947848 | 0.5049 | 2.573671048 | 2.573671048 | 2.573671048 | 0.088191955 | -1.05457103 | FALSE | FALSE | FALSE |
| PLEK | 0.2269589 | 0.82645598 | 1 | 0.22695887 | 0.22695887 | 0.22695887 | 0.27006318 | 0.833775817 | 1 | 0.270063182 | 0.270063182 | 0.270063182 | 0.840391749 | -0.07551822 | FALSE | TRUE | FALSE |
| GML | 0.2269345 | 0.82647355 | 1 | 0.22693452 | 0.22693452 | 0.22693452 | 0.26935474 | 0.833962913 | 1 | 0.269354736 | 0.269354736 | 0.269354736 | 0.842511711 | -0.074424054 | FALSE | TRUE | FALSE |
| AIMP1 | 0.2268931 | 0.82649112 | 0.00825 | 0.226893148 | 0.226893148 | 0.226893148 | 0.26966578 | 0.833877922 | 1 | 0.269665777 | 0.269665777 | 0.269665777 | 0.841386513 | -0.075004454 | FALSE | TRUE | FALSE |
| POT1 | 0.2268232 | 0.82650869 | 1 | 2.090591909 | 19.77630031 | 41.35988021 | 1.13473332 | 0.574141387 | 0.50795 | 1.134733322 | 1.134733322 | 5.619780668 | 0.199891229 | -0.699206261 | FALSE | FALSE | FALSE |
| GPHA2 | 0.2268133 | 0.82652625 | 0.9995 | 18583.46192 | 21987190.57 | 192371879.9 | 0.2242936 | 0.846333609 | 1 | 21593545.28 | 4.12817E+11 | 4.94436E+12 | 1.011233781 | 0.004851569 | FALSE | TRUE | FALSE |
| BTBD2 | 0.2267616 | 0.82654381 | 1 | 0.226761551 | 0.226761551 | 4.474540807 | 0.27036874 | 0.833639481 | 1 | 0.270368736 | 0.270368736 | 5.343604799 | 0.838712177 | -0.076387052 | FALSE | TRUE | FALSE |
| REEP5 | 0.2266842 | 0.82656136 | 1 | 0.906556293 | 0.906556293 | 0.906556293 | 0.27287205 | 0.833023223 | 1 | 0.272872053 | 0.272872053 | 0.272872053 | 0.830734519 | -0.080537743 | FALSE | TRUE | FALSE |
| ZNF277 | 0.2266772 | 0.82657892 | 0.01215 | 0.226677203 | 0.226677203 | 0.516109553 | 0.04056997 | 0.888815559 | 0.01055 | 0.040569969 | 0.040569969 | 0.805903829 | 5.587315146 | 0.747203168 | FALSE | TRUE | FALSE |
| SESTD1 | 0.2266121 | 0.82659646 | 1 | 2.091906201 | 19.81959491 | 41.32704808 | 0.28679957 | 0.829138532 | 1 | 0.286799569 | 0.286799569 | 5.674250763 | 0.790140965 | -0.102295422 | FALSE | TRUE | FALSE |
| ASGR2 | 0.2265668 | 0.82661401 | 1 | 0.226566831 | 0.226566831 | 0.226566831 | 0.26918953 | 0.834030844 | 1 | 0.269189535 | 0.269189535 | 0.269189535 | 0.841662848 | -0.074861842 | FALSE | TRUE | FALSE |
| OR5AN1 | 0.2263927 | 0.82663155 | 1 | 187.0843657 | 18663.63469 | 193175.3378 | 0.30392925 | 0.824121675 | 1 | 2.801519053 | 26.5037421 | 26.5037421 | 0.744886208 | -0.127910067 | FALSE | TRUE | FALSE |
| SCN3B | 0.2263487 | 0.82664909 | 1 | 0.226348716 | 0.226348716 | 0.226348716 | 0.26985336 | 0.833809865 | 1 | 0.269853358 | 0.269853358 | 0.269853358 | 0.838784137 | -0.076349792 | FALSE | TRUE | FALSE |
| C1QC | 0.2262673 | 0.82666662 | 1 | 0.226267268 | 0.226267268 | 0.226267268 | 1.06666445 | 0.581802872 | 0.49905 | 0.266740609 | 1.06666445 | 1.06666445 | 0.212126005 | -0.673406088 | FALSE | FALSE | FALSE |
| OLIG3 | 0.2261943 | 0.82668415 | 1 | 0.226194271 | 0.226194271 | 4.485687713 | 0.26976837 | 0.833843901 | 1 | 0.269768365 | 0.269768365 | 5.320769864 | 0.838475893 | -0.076509419 | FALSE | TRUE | FALSE |
| CA2 | 0.226182 | 0.82670168 | 1 | 0.226182009 | 0.226182009 | 0.226182009 | 2.437066 | 0.451894763 | 0.5077 | 2.437066001 | 2.437066001 | 2.437066001 | 0.092809144 | -1.032409234 | FALSE | FALSE | FALSE |
| HOXA2 | 0.2261447 | 0.82671921 | 1 | 0.22614473 | 0.22614473 | 4.487132781 | 5.35441827 | 0.291858199 | 0.00855 | 0.26959826 | 0.26959826 | 0.26959826 | 0.042235163 | -1.374325822 | FALSE | FALSE | FALSE |
| PACSIN3 | 0.2260847 | 0.82673673 | 0.01655 | 0.226084713 | 0.226084713 | 4.465047652 | 0.26900813 | 0.834098719 | 1 | 0.269008132 | 0.269008132 | 5.320769864 | 0.840438211 | -0.075494211 | FALSE | TRUE | FALSE |
| PMF1 | 0.2260788 | 0.82675424 | 0.00565 | 0.226078772 | 0.226078772 | 0.226078772 | 0.24306537 | 0.841119227 | 1 | 0.243065365 | 0.243065365 | 0.243065365 | 0.930115124 | -0.031463294 | FALSE | TRUE | FALSE |
| RBM12 | 0.226025 | 0.82677176 | 1 | 2.091077613 | 2.091077613 | 41.07451195 | 0.27752132 | 0.831654785 | 1 | 0.277521321 | 0.277521321 | 5.463005655 | 0.814441896 | -0.089139894 | FALSE | TRUE | FALSE |
| OR8D4 | 0.2260189 | 0.82678927 | 1 | 0.226018914 | 0.226018914 | 0.226018914 | 0.26934415 | 0.833979901 | 1 | 0.26934415 | 0.26934415 | 0.26934415 | 0.839145436 | -0.076162763 | FALSE | TRUE | FALSE |
| CD2BP2 | 0.2259652 | 0.82680677 | 1 | 0.225965191 | 0.225965191 | 4.483000685 | 0.26879335 | 0.834132636 | 1 | 0.26879335 | 0.26879335 | 5.337834767 | 0.84066511 | -0.075376977 | FALSE | TRUE | FALSE |
| OPRM1 | 0.2257927 | 0.82682427 | 0.75615 | 0.225792687 | 2.03455931 | 2.03455931 | 0.35707716 | 0.806735872 | 0.513 | 0.357077165 | 0.357077165 | 0.357077165 | 0.632335833 | -0.199052207 | FALSE | TRUE | FALSE |
| GPX7 | 0.2257838 | 0.82684177 | 1 | 0.225783814 | 0.225783814 | 0.225783814 | 0.26825116 | 0.834268171 | 0.00565 | 0.268251162 | 0.268251162 | 0.268251162 | 0.841688112 | -0.074848807 | FALSE | TRUE | FALSE |
| MIA2 | 0.2257764 | 0.82685927 | 0.69445 | 0.22577635 | 18.2838467 | 18.2838467 | 0.05081463 | 0.886436318 | 0.94005 | 0.446075393 | 37.07403436 | 37.07403436 | 4.443136625 | 0.647689668 | FALSE | TRUE | FALSE |
| N4BP3 | 0.2257383 | 0.82687676 | 1 | 0.225738343 | 0.225738343 | 4.440725315 | 0.26073597 | 0.836473055 | 1 | 2.432306144 | 2.432306144 | 8.832257894 | 0.865773684 | -0.062595619 | FALSE | TRUE | FALSE |
| YIPF1 | 0.2257257 | 0.82689425 | 0.75495 | 0.902870932 | 3.612259266 | 3.612259266 | 0.27441581 | 0.832436987 | 1 | 0.274415811 | 0.274415811 | 0.274415811 | 0.82256809 | -0.084828142 | FALSE | TRUE | FALSE |
| OR5M8 | 0.225526 | 0.82691174 | 0.0082 | 0.225525979 | 0.225525979 | 0.225525979 | 0.26810872 | 0.834285098 | 1 | 0.26810872 | 0.26810872 | 0.26810872 | 0.841173607 | -0.075114362 | FALSE | TRUE | FALSE |
| C9orf50 | 0.2255073 | 0.82692922 | 1 | 0.225507278 | 0.225507278 | 4.476889123 | 0.26807439 | 0.834302021 | 0.00985 | 0.268074387 | 0.268074387 | 0.268074387 | 0.841211578 | -0.075094758 | FALSE | TRUE | FALSE |
| BPIFB2 | 0.2252643 | 0.8269467 | 1 | 0.225264348 | 0.225264348 | 4.457871568 | 2.39100967 | 0.456559037 | 0.505 | 2.391009673 | 2.391009673 | 5.147478017 | 0.094213064 | -1.025888871 | FALSE | FALSE | FALSE |
| TMEM183A | 0.2252569 | 0.82696417 | 1 | 0.225256902 | 0.225256902 | 4.471007273 | 0.23392817 | 0.84363358 | 1 | 0.233928166 | 0.233928166 | 4.647348759 | 0.962931936 | -0.016404409 | FALSE | TRUE | FALSE |
| OR4K13 | 0.2252159 | 0.82698164 | 1 | 2.061355652 | 2.061355652 | 2.061355652 | 0.27601637 | 0.832055466 | 0.00675 | 0.276016368 | 0.276016368 | 0.276016368 | 0.81595133 | -0.088335745 | FALSE | TRUE | FALSE |
| CUEDC2 | 0.2251796 | 0.82699911 | 0.0082 | 0.22517959 | 0.22517959 | 0.22517959 | 0.26867515 | 0.83414959 | 1 | 0.268675147 | 0.268675147 | 0.268675147 | 0.838110975 | -0.076698472 | FALSE | TRUE | FALSE |
| ADAT2 | 0.2250904 | 0.82701658 | 1 | 0.225090441 | 0.225090441 | 0.225090441 | 0.26758097 | 0.834437283 | 1 | 0.267580972 | 0.267580972 | 0.267580972 | 0.84120496 | -0.075098175 | FALSE | TRUE | FALSE |
| ISL1 | 0.2249501 | 0.82703404 | 0.01205 | 0.224950103 | 0.224950103 | 4.455729073 | 0.26853118 | 0.834200431 | 1 | 0.268531184 | 0.268531184 | 5.325120797 | 0.837705699 | -0.07690853 | FALSE | TRUE | FALSE |
| TNFRSF1A | 0.2248431 | 0.8270515 | 0.01455 | 0.224843085 | 0.224843085 | 4.444465644 | 0.26834484 | 0.834234308 | 1 | 0.268344843 | 0.268344843 | 5.311799349 | 0.837888599 | -0.076813719 | FALSE | TRUE | FALSE |
| HSPB8 | 0.2248376 | 0.82706896 | 1 | 0.89907659 | 0.89907659 | 0.89907659 | 2.4664255 | 0.450008647 | 0.5026 | 2.466425504 | 2.466425504 | 2.466425504 | 0.0911593 | -1.040199018 | FALSE | FALSE | FALSE |
| TTBK1 | 0.2248282 | 0.82708641 | 1 | 2.102169137 | 2.102169137 | 40.90523589 | 0.27665439 | 0.831951121 | 1 | 0.27665439 | 0.27665439 | 5.400868648 | 0.812667946 | -0.09008687 | FALSE | TRUE | FALSE |
| KDM2A | 0.2247758 | 0.82710386 | 0.04095 | 0.224775833 | 0.224775833 | 7.559474279 | 0.26798288 | 0.834335857 | 1 | 0.267982877 | 0.267982877 | 5.231919143 | 0.838769386 | -0.076357429 | FALSE | TRUE | FALSE |
| ZNF320 | 0.2247702 | 0.8271213 | 1 | 0.224770164 | 0.224770164 | 4.455080111 | 0.26790229 | 0.83435277 | 1 | 0.267902292 | 0.267902292 | 5.315713518 | 0.839000527 | -0.076237766 | FALSE | TRUE | FALSE |
| SEMA6D | 0.2247013 | 0.82713874 | 0.50965 | 0.898755784 | 0.898755784 | 17.59076058 | 2.34777747 | 0.46114528 | 0.5145 | 9.390655605 | 89.89692545 | 184.2705673 | 0.095708078 | -1.019051406 | FALSE | FALSE | FALSE |
| PLA2G4A | 0.2246975 | 0.82715618 | 1 | 0.224697458 | 0.224697458 | 4.435021451 | 0.26750248 | 0.834454176 | 1 | 0.267502477 | 0.267502477 | 5.288227232 | 0.839982718 | -0.075729649 | FALSE | TRUE | FALSE |
| SLC35G6 | 0.2246965 | 0.82717361 | 0.50435 | 2.058550299 | 2.058550299 | 4.45973638 | 0.27551229 | 0.832177042 | 1 | 0.27551229 | 0.27551229 | 0.27551229 | 0.815558909 | -0.088544664 | FALSE | TRUE | FALSE |
| LRRC2 | 0.2246254 | 0.82719104 | 1 | 0.224625419 | 0.224625419 | 0.224625419 | 0.26738291 | 0.834521711 | 1 | 0.267382911 | 0.267382911 | 0.267382911 | 0.840088913 | -0.075674747 | FALSE | TRUE | FALSE |
| DNAJC3 | 0.2246243 | 0.82720847 | 1 | 0.224624259 | 0.224624259 | 4.451089582 | 0.25947109 | 0.836884069 | 1 | 2.410987703 | 2.410987703 | 5.147402800 | 0.865700512 | -0.062632326 | FALSE | TRUE | FALSE |
| HINFP | 0.2245871 | 0.82722589 | 1 | 0.224587133 | 0.224587133 | 4.438837037 | 0.2674333 | 0.834504832 | 1 | 0.267433304 | 0.267433304 | 5.29315766 | 0.839787454 | -0.075830618 | FALSE | TRUE | FALSE |
| GPC1 | 0.2243858 | 0.82724331 | 1 | 0.224385761 | 0.224385761 | 4.419135983 | 0.26775332 | 0.834403488 | 1 | 0.267753319 | 0.267753319 | 5.282932094 | 0.838031672 | -0.076739567 | FALSE | TRUE | FALSE |
| EBLN2 | 0.2243553 | 0.82726073 | 1 | 0.224355313 | 0.224355313 | 0.224355313 | 0.26667212 | 0.834707145 | 1 | 0.266672122 | 0.266672122 | 0.266672122 | 0.841315214 | -0.075041257 | FALSE | TRUE | FALSE |
| BTN3A2 | 0.2243288 | 0.82727814 | 1 | 0.22432881 | 0.22432881 | 0.22432881 | 2.30579395 | 0.464041459 | 0.754 | 2.305793946 | 21.74319567 | 21.74319567 | 0.097289183 | -1.011935443 | FALSE | FALSE | FALSE |
| PDIA6 | 0.2242647 | 0.82729555 | 1 | 0.224264735 | 0.224264735 | 4.444375989 | 0.26704995 | 0.834656612 | 0.00975 | 0.267049945 | 0.267049945 | 0.267049945 | 0.839785735 | -0.075831507 | FALSE | TRUE | FALSE |
| LGALS1 | 0.2242257 | 0.82731296 | 1 | 0.224225682 | 0.224225682 | 0.224225682 | 0.26096608 | 0.836423593 | 1 | 1.043761172 | 4.175675738 | 4.175675738 | 0.859213881 | -0.065898715 | FALSE | TRUE | FALSE |
| OR52E6 | 0.2242253 | 0.82733036 | 1 | 0.224225319 | 0.224225319 | 0.224225319 | 0.26669405 | 0.834690304 | 1 | 0.26669405 | 0.26669405 | 0.26669405 | 0.84075861 | -0.075328677 | FALSE | TRUE | FALSE |
| ZSCAN25 | 0.224162 | 0.82734776 | 1 | 0.224161967 | 0.224161967 | 4.428338341 | 0.26711969 | 0.834622908 | 1 | 0.267119689 | 0.267119689 | 5.284747005 | 0.839181747 | -0.076143971 | FALSE | TRUE | FALSE |
| ZNF441 | 0.2241071 | 0.82736515 | 0.01725 | 0.224107054 | 0.224107054 | 4.433310212 | 0.2671341 | 0.83458919 | 0.0152 | 0.267134104 | 0.267134104 | 5.291402031 | 0.838930899 | -0.07627381 | FALSE | TRUE | FALSE |
| EFNA5 | 0.2236918 | 0.82738255 | 0.00695 | 0.223691774 | 0.223691774 | 0.223691774 | 0.26717698 | 0.834572325 | 1 | 0.267176981 | 0.267176981 | 0.267176981 | 0.837241942 | -0.077149024 | FALSE | TRUE | FALSE |
| OR52L1 | 0.2236126 | 0.82739994 | 1 | 0.223612637 | 0.223612637 | 0.223612637 | 0.26619246 | 0.83482494 | 1 | 0.266192461 | 0.266192461 | 0.266192461 | 0.84004121 | -0.075699408 | FALSE | TRUE | FALSE |
| TRERF1 | 0.2235837 | 0.82741733 | 1 | 0.223583665 | 0.223583665 | 4.350898758 | 0.25623808 | 0.837651335 | 1 | 0.256238085 | 0.256238085 | 5.002581192 | 0.872562193 | -0.059203609 | FALSE | TRUE | FALSE |
| FMOD | 0.2235214 | 0.82743471 | 1 | 0.893850667 | 3.575357005 | 4.430708282 | 0.2728558 | 0.833040401 | 1 | 0.272855804 | 0.272855804 | 0.272855804 | 0.819192351 | -0.086614112 | FALSE | TRUE | FALSE |
| LHX2 | 0.223314 | 0.82745209 | 1 | 46046518.3 | 4.7578E+11 | 5.84154E+12 | 0.2836514 | 0.83003309 | 1 | 33620179.42 | 52246286980 | 6.31028E+11 | 0.787283396 | -0.103868908 | FALSE | TRUE | FALSE |
| RAD51AP2 | 0.2233101 | 0.82746947 | 0.88805 | 1.924657228 | 153.9284329 | 1426.705851 | 3.69244812 | 0.372077923 | 0.7756 | 33.12880689 | 2865.958484 | 10275.4974 | 0.060477522 | -1.218406009 | FALSE | FALSE | FALSE |
| AMD1 | 0.223295 | 0.82748684 | 0.8731 | 8.188889183 | 77.05795976 | 77.05795976 | 0.28581899 | 0.829336136 | 0.00855 | 0.285818994 | 0.285818994 | 0.285818994 | 0.78124616 | -0.107212104 | FALSE | TRUE | FALSE |
| ZNF286A | 0.2232949 | 0.82750421 | 1 | 0.223294872 | 0.223294872 | 4.428555987 | 0.26602707 | 0.834892176 | 1 | 0.266027067 | 0.266027067 | 5.281359385 | 0.839368995 | -0.076047077 | FALSE | TRUE | FALSE |
| CTTN | 0.2232661 | 0.82752158 | 1 | 0.22326614 | 0.22326614 | 4.405879103 | 0.2664034 | 0.834757649 | 1 | 0.266403401 | 0.266403401 | 5.265560943 | 0.838075411 | -0.076716901 | FALSE | TRUE | FALSE |

| | | | | | | | | | | | | | | | | | |
|---|---|---|---|---|---|---|---|---|---|---|---|---|---|---|---|---|---|
| TOM1L2 | 0.2232016 | 0.82753894 | 1 | 0.223201617 | 0.223201617 | 4.411586621 | 0.26612861 | 0.834841755 | 1 | 0.266128612 | 0.266128612 | 5.267475091 | 0.838698311 | -0.076394232 | FALSE | TRUE | FALSE |
| SLC8A1 | 0.2230584 | 0.8275563 | 1 | 2.086850876 | 2.086850876 | 40.98004395 | 0.27175425 | 0.833280553 | 1 | 1.086884549 | 1.086884549 | 21.39004964 | 0.820809396 | -0.085757681 | FALSE | TRUE | FALSE |
| KIF22 | 0.2230398 | 0.82757366 | 0.26695 | 0.223039799 | 0.892037566 | 4.397533005 | 0.07673396 | 0.881123685 | 1 | 0.70458982 | 2.817409501 | 10.38829362 | 2.9066633 | 0.463394727 | FALSE | TRUE | FALSE |
| DDB1 | 0.2230331 | 0.82759101 | 1 | 0.223033088 | 0.223033088 | 4.351994181 | 0.26627534 | 0.834791302 | 0.027 | 0.266275336 | 0.266275336 | 5.211022425 | 0.837603252 | -0.076961645 | FALSE | TRUE | FALSE |
| CDKL1 | 0.2230153 | 0.82760836 | 0.8107 | 1.998242219 | 73.50924246 | 73.50924246 | 2.92861029 | 0.416879992 | 0.2565 | 0.732288905 | 2.928610293 | 2.928610293 | 0.076150549 | -1.118326963 | FALSE | FALSE | FALSE |
| MRGPRX2 | 0.2229571 | 0.8276257 | 0.0093 | 0.222957145 | 0.222957145 | 0.222957145 | 0.26555889 | 0.835043252 | 1 | 0.265558886 | 0.265558886 | 0.265558886 | 0.839577046 | -0.075939444 | FALSE | TRUE | FALSE |
| LAPTM4A | 0.2229544 | 0.82764305 | 1 | 2.040246885 | 2.040246885 | 2.040246885 | 0.27354658 | 0.832730628 | 0.00595 | 0.27354658 | 0.27354658 | 0.27354658 | 0.815051052 | -0.088815187 | FALSE | TRUE | FALSE |
| HMSD | 0.2229273 | 0.82766038 | 0.0035 | 0.222927327 | 0.222927327 | 0.222927327 | 2.37250502 | 0.458219025 | 0.50505 | 2.372505024 | 2.372505024 | 2.372505024 | 0.093962847 | -1.027043833 | FALSE | FALSE | FALSE |
| SNX17 | 0.2229266 | 0.82767772 | 1 | 0.222926614 | 0.222926614 | 4.409111247 | 2.39525308 | 0.456188827 | 0.50535 | 2.395253078 | 2.395253078 | 5.104954959 | 0.093070171 | -1.031189488 | FALSE | FALSE | FALSE |
| PTTG1IP | 0.2228965 | 0.82769505 | 1 | 2.04819733 | 2.04819733 | 2.04819733 | 0.27371524 | 0.832678885 | 1 | 0.273715238 | 0.273715238 | 0.273715238 | 0.814337367 | -0.089195636 | FALSE | TRUE | FALSE |
| TIGD2 | 0.2227928 | 0.82771238 | 1 | 0.222792822 | 0.222792822 | 4.414181849 | 0.26539249 | 0.835194045 | 1 | 0.26539249 | 0.26539249 | 5.264121908 | 0.839484277 | -0.075987433 | FALSE | TRUE | FALSE |
| CDCA7 | 0.2227712 | 0.8277297 | 1 | 0.222771213 | 0.222771213 | 4.408173368 | 0.26544507 | 0.835110307 | 0.0121 | 0.265445073 | 0.265445073 | 5.259300433 | 0.839236572 | -0.076115599 | FALSE | TRUE | FALSE |
| REEP1 | 0.2227323 | 0.82774702 | 1 | 0.222732258 | 0.222732258 | 0.222732258 | 0.26580798 | 0.834976143 | 0.0052 | 0.265807982 | 0.265807982 | 0.265807982 | 0.837944204 | -0.076784899 | FALSE | TRUE | FALSE |
| OR5AR1 | 0.2227111 | 0.82776434 | 1 | 0.222711138 | 0.222711138 | 0.222711138 | 0.25718425 | 0.837407225 | 1 | 2.368483691 | 2.368483691 | 2.368483691 | 0.865959485 | -0.062502426 | FALSE | TRUE | FALSE |
| ABHD14B | 0.222647 | 0.82778165 | 1 | 19.57495454 | 190.7196926 | 190.7196926 | 0.29132439 | 0.827650866 | 1 | 0.291324385 | 0.291324385 | 0.291324385 | 0.764258103 | -0.116759948 | FALSE | TRUE | FALSE |
| SMAD2 | 0.2225988 | 0.82779896 | 1 | 20.2647583 | 200.9057676 | 200.9057676 | 0.2924077 | 0.827320811 | 1 | 0.292407697 | 0.292407697 | 5.804018074 | 0.761261708 | -0.118466015 | FALSE | TRUE | FALSE |
| MARK1 | 0.2225289 | 0.82781627 | 0.5146 | 2.071376876 | 2.071376876 | 40.76890257 | 0.27402187 | 0.832540745 | 1 | 0.274021872 | 0.274021872 | 5.403628355 | 0.812084428 | -0.090398817 | FALSE | TRUE | FALSE |
| ZNHIT6 | 0.2224678 | 0.82783357 | 1 | 0.222467844 | 0.222467844 | 4.409204068 | 0.26499109 | 0.835311129 | 1 | 0.264991094 | 0.264991094 | 5.2576947 | 0.839529511 | -0.075964033 | FALSE | TRUE | FALSE |
| SENP7 | 0.2224668 | 0.82785087 | 0.0264 | 0.222466768 | 0.222466768 | 4.369935765 | 0.26213928 | 0.83609308 | 0.0237 | 0.262139281 | 0.262139281 | 5.160290423 | 0.848658647 | -0.071266959 | FALSE | TRUE | FALSE |
| RANBP10 | 0.222397 | 0.82786817 | 1 | 0.222397041 | 0.222397041 | 4.385714296 | 0.25753298 | 0.837342009 | 1 | 2.427475245 | 2.427475245 | 8.718416221 | 0.863567238 | -0.063703842 | FALSE | TRUE | FALSE |
| EFNB2 | 0.2223576 | 0.82788546 | 1 | 0.222357554 | 0.222357554 | 0.222357554 | 0.26578207 | 0.835009704 | 0.00865 | 0.265782067 | 0.265782067 | 0.265782067 | 0.836616093 | -0.077473786 | FALSE | TRUE | FALSE |
| PPAT | 0.2221985 | 0.82790275 | 1 | 0.22219851 | 0.22219851 | 4.399958983 | 0.26547616 | 0.835093549 | 1 | 0.265476163 | 0.265476163 | 5.263202439 | 0.836981023 | -0.077284389 | FALSE | TRUE | FALSE |
| RPL4 | 0.2221721 | 0.82792004 | 1 | 0.888480582 | 3.553979581 | 17.592045 | 21.039997 | 0.127985055 | 0.00925 | 1.061342544 | 4.245555614 | 5.261084456 | 0.010559512 | -1.97635616 | FALSE | FALSE | FALSE |
| LY6G5B | 0.2221599 | 0.82793732 | 1 | 19.19216591 | 1835.007039 | 1835.007039 | 0.29017249 | 0.828070767 | 1 | 2.679961506 | 2.679961506 | 2.679961506 | 0.765613172 | -0.115990603 | FALSE | TRUE | FALSE |
| MED1 | 0.2221474 | 0.8279546 | 0.9694 | 196.1704819 | 1979.062654 | 20453.11784 | 0.29143928 | 0.827632562 | 1 | 2.737343394 | 26.38736006 | 92.19179646 | 0.762242639 | -0.117906761 | FALSE | TRUE | FALSE |
| IPO5 | 0.2220943 | 0.82797188 | 1 | 0.222094343 | 0.222094343 | 4.352101299 | 0.26542223 | 0.835143813 | 1 | 0.265422234 | 0.265422234 | 5.213804933 | 0.836758621 | -0.077399804 | FALSE | TRUE | FALSE |
| RAB4A | 0.2220241 | 0.82798915 | 1 | 0.22202411 | 0.22202411 | 0.22202411 | 0.26503969 | 0.835294413 | 1 | 0.265039689 | 0.265039689 | 0.265039689 | 0.837701367 | -0.076910776 | FALSE | TRUE | FALSE |
| TREM1 | 0.222012 | 0.82800642 | 1 | 0.222011959 | 0.222011959 | 0.222011959 | 2.34836445 | 0.460781367 | 0.5017 | 2.348364451 | 2.348364451 | 2.348364451 | 0.094538971 | -1.024389129 | FALSE | FALSE | FALSE |
| RTBDN | 0.2219087 | 0.82802368 | 0.00965 | 0.22190871 | 0.22190871 | 0.22190871 | 0.26448406 | 0.835394666 | 1 | 0.264484061 | 0.264484061 | 0.264484061 | 0.83902489 | -0.076225156 | FALSE | TRUE | FALSE |
| NOSIP | 0.2218105 | 0.82804094 | 1 | 0.22181053 | 0.22181053 | 4.39133771 | 0.2647647 | 0.835344553 | 1 | 0.264764703 | 0.264764703 | 0.264764703 | 0.837764729 | -0.076877928 | FALSE | TRUE | FALSE |
| HLA-F | 0.2218066 | 0.8280582 | 1 | 0.221806579 | 0.221806579 | 4.382477477 | 0.24853929 | 0.839911022 | 0.7491 | 2.289024656 | 21.6370828 | 21.6370828 | 0.892440687 | -0.049420638 | FALSE | TRUE | FALSE |
| POLD3 | 0.2217347 | 0.82807546 | 0.01155 | 0.22173471 | 0.22173471 | 4.399765971 | 0.25063754 | 0.839196672 | 1 | 0.250637539 | 0.250637539 | 4.977980231 | 0.884682762 | -0.053212435 | FALSE | TRUE | FALSE |
| PHACTR2 | 0.2214787 | 0.82809271 | 0.01765 | 0.2214787 | 0.2214787 | 4.37633728 | 0.26428085 | 0.835461436 | 1 | 0.264280849 | 0.264280849 | 5.229643281 | 0.838042943 | -0.076733726 | FALSE | TRUE | FALSE |
| PTBP3 | 0.2214741 | 0.82810996 | 1 | 0.22147409 | 0.22147409 | 4.383584661 | 0.26465888 | 0.835361261 | 1 | 0.26465888 | 0.26465888 | 5.24486043 | 0.836828487 | -0.077363544 | FALSE | TRUE | FALSE |
| MXD3 | 0.2214637 | 0.82812721 | 1 | 0.221463702 | 0.221463702 | 0.221463702 | 2.30515001 | 0.464221493 | 0.74865 | 2.305150008 | 21.95667392 | 21.95667392 | 0.096073445 | -1.017396637 | FALSE | FALSE | FALSE |
| OR51B4 | 0.2213633 | 0.82814445 | 1 | 0.221363332 | 0.221363332 | 0.221363332 | 0.26378656 | 0.835611472 | 1 | 0.263786561 | 0.263786561 | 0.263786561 | 0.839175929 | -0.076146982 | FALSE | TRUE | FALSE |
| RFX7 | 0.2211646 | 0.82816169 | 0.0354 | 0.221164554 | 0.221164554 | 7.444222167 | 0.26447247 | 0.835411363 | 1 | 0.264472469 | 0.264472469 | 5.170528181 | 0.836247928 | -0.077664945 | FALSE | TRUE | FALSE |
| ARL10 | 0.2211243 | 0.82817893 | 0.0064 | 0.221124273 | 0.221124273 | 0.221124273 | 9.3775356 | 0.20707722 | 0.25855 | 2.344690729 | 9.377535603 | 9.377535603 | 0.023580211 | -1.627452304 | FALSE | FALSE | FALSE |
| GLIS2 | 0.2210067 | 0.82819616 | 1 | 0.221006734 | 0.221006734 | 4.368512754 | 0.26397642 | 0.835544823 | 1 | 0.263976416 | 0.263976416 | 5.225201398 | 0.837221514 | -0.07715962 | FALSE | TRUE | FALSE |
| MPV17L2 | 0.220956 | 0.82821339 | 1 | 0.220956008 | 0.220956008 | 0.220956008 | 0.263789 | 0.835594815 | 1 | 0.263789002 | 0.263789002 | 0.263789002 | 0.837624036 | -0.076950869 | FALSE | TRUE | FALSE |
| NEUROG2 | 0.2208766 | 0.82823062 | 0.0117 | 0.220876585 | 0.220876585 | 4.37439341 | 0.26407169 | 0.835511478 | 1 | 0.264071689 | 0.264071689 | 5.236002364 | 0.8364266 | -0.077572164 | FALSE | TRUE | FALSE |
| SELL | 0.2208468 | 0.82824784 | 0.4949 | 0.220846765 | 2.053167894 | 2.053167894 | 0.27210561 | 0.833143406 | 1 | 0.272105614 | 0.272105614 | 0.272105614 | 0.811621494 | -0.09064646 | FALSE | TRUE | FALSE |
| MED16 | 0.220698 | 0.82826506 | 0.94075 | 1.973646619 | 170.2273034 | 170.2273034 | 226.739025 | 0.011249038 | 0.75155 | 2195.837691 | 220931.0194 | 2292871.856 | 0.000973357 | -3.011727795 | FALSE | FALSE | FALSE |
| SFRP2 | 0.2206607 | 0.82828228 | 1 | 0.220660736 | 0.220660736 | 0.220660736 | 0.26326938 | 0.835727981 | 1 | 0.263269381 | 0.263269381 | 0.263269381 | 0.838155712 | -0.076675291 | FALSE | TRUE | FALSE |
| YDJC | 0.2206453 | 0.82829949 | 1 | 8.172656576 | 77.68319357 | 77.68319357 | 0.28365238 | 0.830015292 | 1 | 0.28365238 | 0.28365238 | 0.28365238 | 0.777872104 | -0.109091803 | FALSE | TRUE | FALSE |
| SETD7 | 0.2205921 | 0.8283167 | 0.0103 | 0.220592065 | 0.220592065 | 4.378764445 | 0.2638114 | 0.835578154 | 0.009 | 0.263811404 | 0.263811404 | 0.263811404 | 0.83617335 | -0.077703678 | FALSE | TRUE | FALSE |
| HBQ1 | 0.2205446 | 0.82833391 | 1 | 0.220544592 | 0.220544592 | 0.220544592 | 0.26310154 | 0.835761239 | 1 | 0.263101536 | 0.263101536 | 0.263101536 | 0.83824897 | -0.076626971 | FALSE | TRUE | FALSE |
| ADORA2B | 0.2204765 | 0.82835111 | 1 | 0.220476523 | 0.220476523 | 0.220476523 | 2.29892427 | 0.465119685 | 0.75325 | 2.29892427 | 21.91985007 | 21.91985007 | 0.095904213 | -1.018162314 | FALSE | FALSE | FALSE |
| CDC42EP2 | 0.2204318 | 0.82836831 | 1 | 0.220431799 | 0.220431799 | 0.220431799 | 0.26309582 | 0.835777863 | 1 | 0.263095822 | 0.263095822 | 0.263095822 | 0.837838463 | -0.076839706 | FALSE | TRUE | FALSE |
| PPP1R35 | 0.2203935 | 0.82838551 | 1 | 0.220393521 | 0.220393521 | 0.220393521 | 0.2625648 | 0.835943918 | 1 | 0.262564799 | 0.262564799 | 0.262564799 | 0.839387159 | -0.076037679 | FALSE | TRUE | FALSE |
| LRRC24 | 0.22032 | 0.8284027 | 0.50575 | 2.044287498 | 2.044287498 | 4.360358389 | 0.27147341 | 0.833383261 | 1 | 0.271473407 | 0.271473407 | 5.379490851 | 0.811571046 | -0.090673455 | FALSE | TRUE | FALSE |
| PLSCR4 | 0.2202898 | 0.82841989 | 1 | 0.220289839 | 0.220289839 | 0.220289839 | 0.26260053 | 0.835910734 | 0.00695 | 0.262600528 | 0.262600528 | 0.262600528 | 0.838878129 | -0.076301128 | FALSE | TRUE | FALSE |
| ITLN2 | 0.2202882 | 0.82843707 | 1 | 0.220288162 | 0.220288162 | 0.220288162 | 0.26290601 | 0.835844325 | 1 | 0.262906005 | 0.262906005 | 0.262906005 | 0.837897035 | -0.076809346 | FALSE | TRUE | FALSE |
| CHRDL2 | 0.2202168 | 0.82845426 | 0.81525 | 1.98628155 | 73.55081716 | 73.55081716 | 0.08378598 | 0.879402392 | 1 | 0.083785977 | 0.083785977 | 1.660190427 | 2.628325635 | 0.419679171 | FALSE | TRUE | FALSE |
| RAB15 | 0.2201479 | 0.82847143 | 1 | 0.220147911 | 0.220147911 | 0.220147911 | 0.26306644 | 0.835811101 | 1 | 0.263066442 | 0.263066442 | 0.263066442 | 0.836852886 | -0.077350882 | FALSE | TRUE | FALSE |
| UBP1 | 0.2201353 | 0.82848861 | 0.01655 | 0.220135332 | 0.220135332 | 4.354156406 | 0.23494794 | 0.843421992 | 1 | 0.234947944 | 0.234947944 | 4.653306824 | 0.936953643 | -0.028281896 | FALSE | TRUE | FALSE |
| DUSP11 | 0.2199835 | 0.82850578 | 1 | 2.034684216 | 2.034684216 | 2.034684216 | 2.43682818 | 0.452082828 | 0.5029 | 2.436828176 | 2.436828176 | 2.436828176 | 0.090274526 | -1.044434785 | FALSE | FALSE | FALSE |
| CFLAR | 0.2199776 | 0.82852295 | 1 | 0.219977601 | 0.219977601 | 0.219977601 | 0.25439893 | 0.838072782 | 0.50525 | 2.34239061 | 2.34239061 | 2.34239061 | 0.864695457 | -0.063136823 | FALSE | TRUE | FALSE |
| GTF2IRD1 | 0.2199053 | 0.82854012 | 1 | 0.21990532 | 0.21990532 | 4.288474424 | 0.26279312 | 0.835877536 | 1 | 0.262793123 | 0.262793123 | 5.140243381 | 0.836800135 | -0.077378258 | FALSE | TRUE | FALSE |
| MAP2K1 | 0.2198256 | 0.82855728 | 1 | 0.219825643 | 0.219825643 | 0.219825643 | 0.26291256 | 0.835827715 | 1 | 0.262912562 | 0.262912562 | 0.262912562 | 0.836116927 | -0.077732984 | FALSE | TRUE | FALSE |
| ZWINT | 0.2198067 | 0.82857444 | 1 | 0.219806672 | 0.219806672 | 0.219806672 | 2.3299474 | 0.462053492 | 0.50355 | 2.329947402 | 2.329947402 | 2.329947402 | 0.094339757 | -1.025305245 | FALSE | FALSE | FALSE |
| PRKG1 | 0.2197982 | 0.82859159 | 1 | 0.219798176 | 0.219798176 | 4.341025906 | 0.25485997 | 0.837959526 | 1 | 2.405971982 | 2.405971982 | 5.041067282 | 0.862427226 | -0.064277542 | FALSE | TRUE | FALSE |
| FUZ | 0.2197754 | 0.82860875 | 1 | 2.020410366 | 2.020410366 | 2.020410366 | 2.27100018 | 0.467264211 | 0.87775 | 21.4693756 | 208.175112 | 208.175112 | 0.096774728 | -1.01423804 | FALSE | FALSE | FALSE |
| AP4S1 | 0.2197187 | 0.82862589 | 1 | 18.88283727 | 181.8977144 | 181.8977144 | 21.9744854 | 0.124016364 | 0.6918 | 21.9744854 | 2104.224761 | 2104.224761 | 0.00999881 | -2.000051699 | FALSE | FALSE | FALSE |
| IFI35 | 0.2196184 | 0.82864304 | 1 | 2.017640677 | 19.02454706 | 19.02454706 | 22.868026 | 0.118223157 | 0.00575 | 2.41603573 | 22.86802595 | 22.86802595 | 0.009603733 | -2.017559943 | FALSE | FALSE | FALSE |
| SCEL | 0.2195795 | 0.82866018 | 0.50395 | 0.219579498 | 0.219579498 | 0.848163306 | 0.03821565 | 0.889394364 | 1 | 0.337789331 | 0.337789331 | 0.756560287 | 5.745800681 | 0.759350556 | FALSE | TRUE | FALSE |
| GSX2 | 0.2195794 | 0.82867732 | 0.00985 | 0.219579376 | 0.219579376 | 0.219579376 | 0.26286008 | 0.835860932 | 1 | 0.262860082 | 0.262860082 | 0.262860082 | 0.835346981 | -0.078133092 | FALSE | TRUE | FALSE |
| GPRIN1 | 0.2195632 | 0.82869445 | 0.0262 | 0.219563201 | 0.219563201 | 4.303167556 | 0.26225372 | 0.83602682 | 1 | 0.262253723 | 0.262253723 | 5.152267615 | 0.837216719 | -0.077162108 | FALSE | TRUE | FALSE |
| MAG | 0.2193436 | 0.82871159 | 0.51615 | 0.877267341 | 0.877267341 | 17.26250972 | 0.26519771 | 0.835277693 | 1 | 0.265197709 | 0.265197709 | 5.228579153 | 0.827094808 | -0.082444705 | FALSE | TRUE | FALSE |
| ACTC1 | 0.2192914 | 0.82872871 | 1 | 0.219291449 | 0.219291449 | 4.343891059 | 0.2624277 | 0.835993669 | 1 | 0.262427705 | 0.262427705 | 5.204345631 | 0.835626136 | -0.077987985 | FALSE | TRUE | FALSE |
| CYP7B1 | 0.2191635 | 0.82874584 | 0.26145 | 0.219163538 | 0.876170109 | 1.925103045 | 0.03862018 | 0.889299596 | 1 | 0.15440548 | 0.15440548 | 0.76689651 | 5.674844947 | 0.753954 | FALSE | TRUE | FALSE |
| TEX101 | 0.2191629 | 0.82876296 | 0.00685 | 0.21916294 | 0.21916294 | 0.21916294 | 0.26150486 | 0.836241975 | 1 | 0.261504865 | 0.261504865 | 0.261504865 | 0.838083608 | -0.076712654 | FALSE | TRUE | FALSE |
| PSG8 | 0.2190731 | 0.82878008 | 0.94145 | 1.941918298 | 164.5664503 | 164.5664503 | 0.34539769 | 0.8104915 | 0.87585 | 3.096923723 | 28.48174114 | 28.48174114 | 0.634263497 | -0.197730282 | FALSE | TRUE | FALSE |

| | | | | | | | | | | | | | | | | | |
|---|---|---|---|---|---|---|---|---|---|---|---|---|---|---|---|---|---|
| SRP72 | 0.2190712 | 0.8287972 | 0.2643 | 0.219071229 | 2.024520439 | 4.327471197 | 0.35063171 | 0.808971141 | 0.0141 | 0.350631715 | 0.350631715 | 0.765056964 | 0.624790115 | -0.20426585 | FALSE | TRUE | FALSE |
| PRKCI | 0.2188929 | 0.82881431 | 1 | 2.037345646 | 2.037345646 | 7.403779424 | 0.27004345 | 0.833792842 | 1 | 0.270043449 | 0.270043449 | 5.343208257 | 0.81058409 | -0.091201925 | FALSE | TRUE | FALSE |
| OR13C5 | 0.2188565 | 0.82883142 | 1 | 0.218856475 | 0.218856475 | 0.218856475 | 1.97715862 | 0.491012225 | 0.87845 | 19.07997921 | 188.8485733 | 188.8485733 | 0.110692421 | -0.955882112 | FALSE | FALSE | FALSE |
| GIMAP4 | 0.2188479 | 0.82884852 | 1 | 0.218847891 | 0.218847891 | 0.218847891 | 0.26114523 | 0.836341089 | 0.007 | 0.261145226 | 0.261145226 | 0.261145226 | 0.838031366 | -0.076739726 | FALSE | TRUE | FALSE |
| ACO2 | 0.2187395 | 0.82886562 | 0.76055 | 0.218739503 | 0.87473214 | 4.293971433 | 0.31166049 | 0.821470931 | 0.51335 | 0.311660495 | 0.311660495 | 6.131668679 | 0.701851877 | -0.153754534 | FALSE | TRUE | FALSE |
| ATXN2L | 0.2186936 | 0.82888272 | 0.0307 | 0.218693562 | 0.218693562 | 4.271227107 | 0.26155686 | 0.836225444 | 1 | 0.261556857 | 0.261556857 | 5.122818851 | 0.836122457 | -0.077730112 | FALSE | TRUE | FALSE |
| TOMM40L | 0.2186746 | 0.82889982 | 1 | 0.218674571 | 0.218674571 | 0.218674571 | 0.23359689 | 0.843754234 | 1 | 0.233596892 | 0.233596892 | 0.233596892 | 0.93611935 | -0.028668778 | FALSE | TRUE | FALSE |
| TRIM13 | 0.2185696 | 0.82891691 | 1 | 0.218569637 | 0.218569637 | 0.218569637 | 0.26103391 | 0.836374101 | 1 | 0.261033907 | 0.261033907 | 0.261033907 | 0.83732278 | -0.077107093 | FALSE | TRUE | FALSE |
| FAM78A | 0.2185046 | 0.828934 | 1 | 2.045111716 | 2.045111716 | 4.334682914 | 0.24558669 | 0.840509492 | 1 | 22.07544172 | 217.4735093 | 217.4735093 | 0.88972506 | -0.050744177 | FALSE | TRUE | FALSE |
| RHOD | 0.2183434 | 0.82895109 | 1 | 0.218343423 | 0.218343423 | 0.218343423 | 1.03274379 | 0.584981232 | 0.49615 | 0.258238494 | 1.032743792 | 1.032743792 | 0.211420707 | -0.674852479 | FALSE | FALSE | FALSE |
| ST6GALNAC4 | 0.2183376 | 0.82896817 | 1 | 0.218337615 | 0.218337615 | 4.325944506 | 0.25275082 | 0.838732933 | 1 | 2.328838312 | 2.328838312 | 2.328838312 | 0.863845342 | -0.063564004 | FALSE | TRUE | FALSE |
| PTGR1 | 0.2183099 | 0.82898525 | 1 | 0.218309902 | 0.218309902 | 0.218309902 | 0.26097039 | 0.836407099 | 0.00615 | 0.260970395 | 0.260970395 | 0.260970395 | 0.836531296 | -0.077517807 | FALSE | TRUE | FALSE |
| SLC31A2 | 0.2183068 | 0.82900232 | 0.0035 | 0.21830678 | 0.21830678 | 0.21830678 | 0.26067528 | 0.836489536 | 1 | 0.260675279 | 0.260675279 | 0.260675279 | 0.837466371 | -0.077032623 | FALSE | TRUE | FALSE |
| PRCC | 0.2183028 | 0.8290194 | 1 | 6.57997E+14 | 1.98484E+19 | 3.83933E+21 | 0.20397208 | 0.851129985 | 1 | 1.78224E+19 | 1.01067E+25 | 2.00547E+26 | 1.070258121 | 0.029488532 | FALSE | TRUE | FALSE |
| NEIL2 | 0.218034 | 0.82903646 | 0.502 | 1.997163189 | 1.997163189 | 1.997163189 | 2.31901442 | 0.46295872 | 0.7486 | 2.319014419 | 21.8726764 | 21.8726764 | 0.094020131 | -1.026779149 | FALSE | FALSE | FALSE |
| MAT2A | 0.2179525 | 0.82905353 | 1 | 81328.70832 | 37474568.84 | 416120398.9 | 1.4308737 | 0.542762816 | 0.75035 | 1.430873696 | 5.723539756 | 5.723539756 | 0.152321283 | -0.81723941 | FALSE | FALSE | FALSE |
| MGP | 0.2178414 | 0.82907059 | 1 | 0.217841407 | 0.217841407 | 0.217841407 | 0.26009575 | 0.836687041 | 1 | 0.260095753 | 0.260095753 | 0.260095753 | 0.837543111 | -0.076992829 | FALSE | TRUE | FALSE |
| SMAD6 | 0.217729 | 0.82908766 | 0.8618 | 2.00596568 | 55.66396062 | 275.97091 | 135.819644 | 0.020776108 | 0.1384 | 10.93683987 | 543.090145 | 2172.149751 | 0.001603075 | -2.795046236 | FALSE | FALSE | FALSE |
| PHAX | 0.2177199 | 0.82910471 | 0.01075 | 0.217719864 | 0.217719864 | 4.32254314 | 0.25999549 | 0.836719913 | 0.01015 | 0.259995494 | 0.259995494 | 5.166411261 | 0.837398606 | -0.077067767 | FALSE | TRUE | FALSE |
| TRMT112 | 0.2177039 | 0.82912177 | 0.0044 | 0.217703872 | 0.217703872 | 0.217703872 | 0.25971983 | 0.836834856 | 1 | 0.25971983 | 0.25971983 | 0.25971983 | 0.838225837 | -0.076638957 | FALSE | TRUE | FALSE |
| RDH11 | 0.2175999 | 0.82913882 | 0.25505 | 0.217599932 | 0.870067443 | 0.870067443 | 0.6916188 | 0.66961721 | 0.6334 | 1.189156522 | 11.05633643 | 11.05633643 | 0.314624085 | -0.502208035 | FALSE | TRUE | FALSE |
| ZNF562 | 0.2175822 | 0.82915586 | 1 | 0.217582208 | 0.217582208 | 0.217582208 | 0.25984928 | 0.836802032 | 1 | 0.259849282 | 0.259849282 | 0.259849282 | 0.837340039 | -0.077098142 | FALSE | TRUE | FALSE |
| OR6C4 | 0.2175566 | 0.8291729 | 1 | 1.993417306 | 180.82325 | 180.82325 | 2.46616235 | 0.450197823 | 0.256 | 2.466162347 | 23.30861987 | 23.30861987 | 0.08821664 | -1.054449488 | FALSE | FALSE | FALSE |
| SF3B3 | 0.2174957 | 0.82918994 | 1 | 0.217495739 | 0.217495739 | 4.244529282 | 0.26043728 | 0.836604806 | 1 | 0.260437278 | 0.260437278 | 5.097390346 | 0.835117541 | -0.078252394 | FALSE | TRUE | FALSE |
| TSSC4 | 0.2174463 | 0.82920698 | 1 | 0.217446316 | 0.217446316 | 4.314674235 | 20.5297239 | 0.129833417 | 0.4997 | 4.702883661 | 198.6327541 | 198.6327541 | 0.01059178 | -1.975031055 | FALSE | FALSE | FALSE |
| LRRC7 | 0.2174126 | 0.82922401 | 1 | 0.217412636 | 0.217412636 | 7.304394446 | 0.25231257 | 0.838813067 | 1 | 2.361447799 | 2.361447799 | 46.0648792 | 0.861679782 | -0.064654097 | FALSE | TRUE | FALSE |
| EYA3 | 0.2174088 | 0.82924104 | 0.2655 | 0.054381625 | 0.217408826 | 1.077440056 | 0.02277073 | 0.892987075 | 0.50665 | 0.091019727 | 0.091019727 | 0.451655369 | 9.547730993 | 0.979900174 | FALSE | TRUE | FALSE |
| COMMD4 | 0.2172986 | 0.82925807 | 0.5082 | 2.000804514 | 2.000804514 | 2.000804514 | 0.25005759 | 0.839435491 | 1 | 2.31066057 | 2.31066057 | 2.31066057 | 0.868994342 | -0.060983051 | FALSE | TRUE | FALSE |
| XYLT2 | 0.2172973 | 0.82927509 | 1 | 0.217297317 | 0.217297317 | 4.255102617 | 0.25991805 | 0.836769195 | 1 | 0.259918055 | 0.259918055 | 5.102513679 | 0.836022404 | -0.077782084 | FALSE | TRUE | FALSE |
| CST4 | 0.2172893 | 0.82929211 | 1 | 1.982943631 | 1.982943631 | 1.982943631 | 0.26724572 | 0.834555457 | 1 | 0.267245725 | 0.267245725 | 0.267245725 | 0.813069242 | -0.089872468 | FALSE | TRUE | FALSE |
| CALU | 0.217177 | 0.82930913 | 0.00775 | 0.217177024 | 0.217177024 | 0.217177024 | 0.26012031 | 0.836670601 | 1 | 0.260120312 | 0.260120312 | 0.260120312 | 0.834909902 | -0.078360388 | FALSE | TRUE | FALSE |
| DPEP1 | 0.2171444 | 0.82932614 | 0.7525 | 0.217144416 | 1.948823271 | 4.295229572 | 0.0378731 | 0.889465344 | 0.75195 | 0.334247435 | 3.027699247 | 3.027699247 | 5.733472836 | 0.758417759 | FALSE | TRUE | FALSE |
| WASF3 | 0.2171247 | 0.82934315 | 1 | 0.217124732 | 0.217124732 | 4.288856345 | 2.34807476 | 0.460963398 | 0.4995 | 0.251925137 | 2.348074761 | 4.983663243 | 0.092469258 | -1.034002626 | FALSE | FALSE | FALSE |
| C16orf78 | 0.217067 | 0.82936016 | 1 | 1.984010359 | 1.984010359 | 1.984010359 | 0.26712039 | 0.834606051 | 0.0071 | 0.26712039 | 0.26712039 | 0.26712039 | 0.812618579 | -0.090113253 | FALSE | TRUE | FALSE |
| TSHZ3 | 0.2169803 | 0.82937716 | 1 | 193.6317439 | 7840.186646 | 81283.59555 | 0.30775277 | 0.822874141 | 1 | 0.307752766 | 0.307752766 | 6.017184418 | 0.705047349 | -0.151781716 | FALSE | TRUE | FALSE |
| SNTG1 | 0.2169737 | 0.82939416 | 1 | 0.216973724 | 0.216973724 | 4.297991188 | 0.25963896 | 0.836867667 | 1 | 0.259638957 | 0.259638957 | 5.149052481 | 0.835674763 | -0.077962713 | FALSE | TRUE | FALSE |
| RPTOR | 0.2167717 | 0.82941115 | 1 | 0.216771725 | 0.216771725 | 7.295526026 | 0.25976179 | 0.836818446 | 1 | 0.259761792 | 0.259761792 | 5.028516208 | 0.834501961 | -0.078572638 | FALSE | TRUE | FALSE |
| OSBPL11 | 0.2166523 | 0.82942815 | 1 | 0.216652259 | 0.216652259 | 4.274450231 | 0.25928305 | 0.836933251 | 1 | 0.25928305 | 0.25928305 | 5.123860973 | 0.835582039 | -0.078010904 | FALSE | TRUE | FALSE |
| RHD | 0.2165944 | 0.82944514 | 0.74895 | 0.216594381 | 1.921505208 | 1.921505208 | 0.04028159 | 0.888911081 | 1 | 0.040281588 | 0.040281588 | 0.040281588 | 5.377006956 | 0.730540598 | FALSE | TRUE | FALSE |
| OR6C1 | 0.2165874 | 0.82946212 | 1 | 18.70022961 | 180.556146 | 180.556146 | 0.25871326 | 0.837031538 | 1 | 22.5167614 | 218.2746231 | 218.2746231 | 0.837171712 | -0.077185455 | FALSE | TRUE | FALSE |
| NMRK1 | 0.2165328 | 0.82947911 | 1 | 0.216532848 | 0.216532848 | 0.216532848 | 0.25884862 | 0.836982409 | 1 | 0.258848622 | 0.258848622 | 0.258848622 | 0.836523084 | -0.07752207 | FALSE | TRUE | FALSE |
| OR2AT4 | 0.21642 | 0.82949609 | 1 | 0.216420002 | 0.216420002 | 0.216420002 | 0.25854407 | 0.837113352 | 1 | 0.258544073 | 0.258544073 | 0.258544073 | 0.837071992 | -0.077237189 | FALSE | TRUE | FALSE |
| PRKRIP1 | 0.2163028 | 0.82951307 | 1 | 0.216302816 | 0.216302816 | 0.216302816 | 0.25834843 | 0.837146056 | 1 | 0.258348427 | 0.258348427 | 0.258348427 | 0.837252304 | -0.077143648 | FALSE | TRUE | FALSE |
| AKAP8 | 0.2161955 | 0.82953004 | 1 | 0.216195517 | 0.216195517 | 4.241575197 | 0.25879494 | 0.837015165 | 1 | 0.258794941 | 0.258794941 | 5.088982681 | 0.835393136 | -0.078109097 | FALSE | TRUE | FALSE |
| ADCY5 | 0.2161784 | 0.82954701 | 0.0415 | 0.216178393 | 0.216178393 | 4.186785571 | 0.1480093 | 0.865088819 | 0.51355 | 0.148009304 | 0.148009304 | 2.877473451 | 1.460573 | 0.164523268 | FALSE | TRUE | FALSE |
| SLC6A6 | 0.2161766 | 0.82956398 | 1 | 0.216176637 | 0.216176637 | 4.261818304 | 0.25919857 | 0.83694964 | 1 | 0.259198566 | 0.259198566 | 5.118753329 | 0.834019419 | -0.078823838 | FALSE | TRUE | FALSE |
| CDCP1 | 0.2161746 | 0.82958094 | 1 | 0.216174629 | 0.216174629 | 4.241847485 | 0.25082196 | 0.839084971 | 1 | 2.326104101 | 2.326104101 | 45.74734859 | 0.861864831 | -0.064560841 | FALSE | TRUE | FALSE |
| PTGFR | 0.2161348 | 0.8295979 | 1 | 0.216134769 | 0.216134769 | 0.216134769 | 0.25863722 | 0.837064274 | 1 | 0.258637217 | 0.258637217 | 0.258637217 | 0.835667703 | -0.077966382 | FALSE | TRUE | FALSE |
| BRPF1 | 0.2161309 | 0.82961486 | 1 | 0.216130947 | 0.216130947 | 4.189353235 | 0.25105332 | 0.839053027 | 1 | 2.360025227 | 2.360025227 | 45.9153923 | 0.860896586 | -0.065049014 | FALSE | TRUE | FALSE |
| CD84 | 0.2160779 | 0.82963181 | 0.5124 | 1.977134799 | 1.977134799 | 1.977134799 | 20.2962859 | 0.131686051 | 0.68735 | 20.29628592 | 1936.424717 | 1936.424717 | 0.010646182 | -1.972806129 | FALSE | FALSE | FALSE |
| RPRD2 | 0.2160229 | 0.82964876 | 1 | 0.216022856 | 0.216022856 | 4.206593756 | 0.24143257 | 0.841631555 | 0.0316 | 0.241432575 | 0.241432575 | 4.716328463 | 0.894754389 | -0.048296162 | FALSE | TRUE | FALSE |
| ADAM11 | 0.2160164 | 0.82966571 | 1 | 0.216016422 | 0.216016422 | 4.238349853 | 0.25861552 | 0.837080637 | 1 | 0.258615522 | 0.258615522 | 5.085759702 | 0.835280188 | -0.078167819 | FALSE | TRUE | FALSE |
| SCRN1 | 0.2158849 | 0.82968265 | 1 | 0.215884901 | 0.215884901 | 4.279353608 | 0.25811589 | 0.837211421 | 1 | 0.258115893 | 0.258115893 | 5.121937551 | 0.836387481 | -0.077592476 | FALSE | TRUE | FALSE |
| TRIAP1 | 0.215877 | 0.82969959 | 1 | 0.215877025 | 0.215877025 | 0.215877025 | 0.21944702 | 0.847290576 | 1 | 0.219447021 | 0.219447021 | 0.219447021 | 0.983731855 | -0.007123265 | FALSE | TRUE | FALSE |
| LILRB3 | 0.21582 | 0.82971653 | 1 | 181.9229503 | 18389.91902 | 191601.1129 | 0.2019681 | 0.851649652 | 0.99995 | 1864680.26 | 2024447827 | 23451687872 | 1.068584738 | 0.028808967 | FALSE | TRUE | FALSE |
| NMNAT1 | 0.2158185 | 0.82973346 | 0.87425 | 0.862807503 | 3.450226517 | 3.450226517 | 0.31180887 | 0.821411974 | 0.51085 | 0.311808874 | 0.311808874 | 0.311808874 | 0.692150092 | -0.159799719 | FALSE | TRUE | FALSE |
| BTNL9 | 0.2157751 | 0.82975039 | 0.7584 | 0.215775081 | 1.92482366 | 4.260576815 | 0.03629124 | 0.889842519 | 0.9406 | 2.865112191 | 105.8070619 | 105.8070619 | 5.945651347 | 0.774199439 | FALSE | TRUE | FALSE |
| CTSL | 0.2156424 | 0.82976732 | 1 | 0.215642424 | 0.215642424 | 0.215642424 | 0.25838599 | 0.837129706 | 1 | 0.258385993 | 0.258385993 | 0.258385993 | 0.834574742 | -0.078534763 | FALSE | TRUE | FALSE |
| CD1D | 0.2156391 | 0.82978424 | 0.7505 | 1.974669015 | 18.55914222 | 18.55914222 | 0.27398414 | 0.832575302 | 0.00925 | 0.273984138 | 0.273984138 | 0.273984138 | 0.787049486 | -0.103997961 | FALSE | TRUE | FALSE |
| LRRC71 | 0.2156337 | 0.82980116 | 0.50625 | 1.979145534 | 1.979145534 | 4.277851726 | 2.37519748 | 0.458035091 | 0.01095 | 2.375197477 | 2.375197477 | 5.118759738 | 0.090785583 | -1.041983112 | FALSE | FALSE | FALSE |
| OR51B6 | 0.2156238 | 0.82981808 | 0.007 | 0.215623777 | 0.215623777 | 0.215623777 | 2.30424963 | 0.464581139 | 0.5022 | 2.304249632 | 2.304249632 | 2.304249632 | 0.09357657 | -1.028832877 | FALSE | FALSE | FALSE |
| CITED2 | 0.2155785 | 0.82983499 | 0.01 | 0.21557851 | 0.21557851 | 0.256183861 | 0.22169116 | 0.84685697 | 1 | 0.221691159 | 0.221691159 | 0.221691159 | 0.972427187 | -0.012142908 | FALSE | TRUE | FALSE |
| RSL1D1 | 0.2155447 | 0.8298519 | 0.01365 | 0.215544725 | 0.215544725 | 4.271831298 | 0.25771335 | 0.837260418 | 1 | 0.257713355 | 0.257713355 | 5.113128296 | 0.836373903 | -0.077599527 | FALSE | TRUE | FALSE |
| RPL15 | 0.215493 | 0.8298688 | 1 | 0.215492951 | 0.215492951 | 0.215492951 | 0.24161773 | 0.841616078 | 0.50545 | 2.224158957 | 2.224158957 | 2.224158957 | 0.89187556 | -0.049695737 | FALSE | TRUE | FALSE |
| SAE1 | 0.215439 | 0.82988571 | 1 | 0.215439 | 0.215439 | 0.215439 | 0.25775116 | 0.837227757 | 1 | 0.257751164 | 0.257751164 | 0.257751164 | 0.835841036 | -0.077876311 | FALSE | TRUE | FALSE |
| SCMH1 | 0.2153972 | 0.82990261 | 1 | 1.987293982 | 1.987293982 | 4.252904259 | 0.26584199 | 0.834942566 | 1 | 0.26584199 | 0.26584199 | 5.256979362 | 0.81024525 | -0.091383506 | FALSE | TRUE | FALSE |
| PFDN2 | 0.215329 | 0.8299195 | 1 | 0.215329021 | 0.215329021 | 0.215329021 | 0.22545219 | 0.84598278 | 1 | 0.225452195 | 0.225452195 | 0.225452195 | 0.955098356 | -0.019951903 | FALSE | TRUE | FALSE |
| THOP1 | 0.2152859 | 0.8299364 | 1 | 0.215285879 | 0.215285879 | 4.221936461 | 0.25772222 | 0.837244089 | 1 | 0.25772222 | 0.25772222 | 5.065991856 | 0.835340776 | -0.078136318 | FALSE | TRUE | FALSE |
| CASP2 | 0.2152787 | 0.82995328 | 1 | 0.215278715 | 0.215278715 | 4.261211394 | 0.25767727 | 0.837309382 | 0.01245 | 0.257677269 | 0.257677269 | 5.106767089 | 0.835458696 | -0.078075016 | FALSE | TRUE | FALSE |
| CGRRF1 | 0.21527 | 0.82997017 | 1 | 18.69000063 | 180.958517 | 180.958517 | 0.24209148 | 0.841383568 | 1 | 204.6654531 | 2035.028789 | 20728.77232 | 0.889209127 | -0.050996088 | FALSE | TRUE | FALSE |
| ZNF93 | 0.2151498 | 0.82998705 | 0.9696 | 17.77964542 | 167.7974549 | 1622.355847 | 0.32171206 | 0.818168451 | 0.983 | 27.03506995 | 2508.434006 | 25039.8649 | 0.668765202 | -0.174726333 | FALSE | TRUE | FALSE |
| OR5B2 | 0.2151012 | 0.83000393 | 1 | 0.215101182 | 0.215101182 | 0.215101182 | 0.24929979 | 0.839641895 | 1 | 2.289843311 | 2.289843311 | 2.289843311 | 0.862821342 | -0.064079121 | FALSE | TRUE | FALSE |
| UBOX5 | 0.2150867 | 0.83002081 | 0.0147 | 0.215086714 | 0.215086714 | 4.254985935 | 0.24197751 | 0.84141461 | 1 | 2.233450152 | 21.15775428 | 21.15775428 | 0.88887067 | -0.051161424 | FALSE | TRUE | FALSE |

| | | | | | | | | | | | | | | | | | |
|---|---|---|---|---|---|---|---|---|---|---|---|---|---|---|---|---|---|
| OR5T3 | 0.2150556 | 0.83003768 | 1 | 0.215055562 | 0.215055562 | 0.215055562 | 0.24175231 | 0.841569633 | 1 | 2.218484119 | 20.89470088 | 20.89470088 | 0.889569825 | -0.050819957 | FALSE | TRUE | FALSE |
| CRYAB | 0.2150459 | 0.83005455 | 0.5105 | 2.015081865 | 2.015081865 | 2.015081865 | 0.26604769 | 0.834875372 | 1 | 0.266047686 | 0.266047686 | 0.266047686 | 0.80829841 | -0.092428276 | FALSE | TRUE | FALSE |
| HCAR2 | 0.214987 | 0.83007142 | 1 | 0.214986972 | 0.214986972 | 4.262135625 | 0.25738083 | 0.837374623 | 1 | 0.257380828 | 0.257380828 | 5.107964664 | 0.835287438 | -0.07816405 | FALSE | TRUE | FALSE |
| IBA57 | 0.2149383 | 0.83008828 | 1 | 0.859588899 | 0.859588899 | 0.859588899 | 0.26042357 | 0.836621259 | 1 | 0.260423572 | 0.260423572 | 0.260423572 | 0.825341007 | -0.083366576 | FALSE | TRUE | FALSE |
| LY96 | 0.21493 | 0.83010514 | 0.0046 | 0.214930039 | 0.214930039 | 0.214930039 | 0.25707719 | 0.837423522 | 1 | 0.257077186 | 0.257077186 | 0.257077186 | 0.836052558 | -0.07776642 | FALSE | TRUE | FALSE |
| UTP3 | 0.214909 | 0.83012199 | 1 | 1.985944009 | 18.83529324 | 39.31653532 | 0.27374088 | 0.83266163 | 1 | 0.273740884 | 0.273740884 | 5.425962414 | 0.785081862 | -0.105085056 | FALSE | TRUE | FALSE |
| DNM1L | 0.2148933 | 0.83013884 | 0.9989 | 220.3151142 | 3532.904079 | 7260.043078 | 122.285597 | 0.023477968 | 0.35145 | 52.2983909 | 3353.144316 | 31364.87849 | 0.001757307 | -2.755152449 | FALSE | FALSE | FALSE |
| NOTUM | 0.2147128 | 0.83015569 | 1 | 0.214712827 | 0.214712827 | 4.249913628 | 2.30969589 | 0.463680961 | 0.0123 | 2.309695892 | 2.309695892 | 4.940269361 | 0.092961514 | -1.031696811 | FALSE | FALSE | FALSE |
| TTC17 | 0.214665 | 0.83017254 | 1 | 0.214665035 | 0.214665035 | 7.229556164 | 0.24184214 | 0.841507662 | 1 | 2.260290835 | 21.68123611 | 76.20941119 | 0.887624607 | -0.051770667 | FALSE | TRUE | FALSE |
| SPAM1 | 0.2145585 | 0.83018938 | 1 | 0.214558469 | 0.214558469 | 4.25451929 | 2.09704444 | 0.480159074 | 0.694 | 19.87951768 | 1926.412046 | 1926.412046 | 0.102314698 | -0.990061972 | FALSE | FALSE | FALSE |
| GHRHR | 0.2145136 | 0.83020622 | 1 | 1.981953535 | 1.981953535 | 2.004707807 | 2.38069712 | 0.457298168 | 0.50515 | 2.380697117 | 2.380697117 | 2.380697117 | 0.090105361 | -1.045249369 | FALSE | FALSE | FALSE |
| ALKBH7 | 0.214455 | 0.83022306 | 0.0073 | 0.214455019 | 0.214455019 | 0.214455019 | 0.25645072 | 0.837586311 | 1 | 0.256450717 | 0.256450717 | 0.256450717 | 0.836242618 | -0.077667703 | FALSE | TRUE | FALSE |
| OR2D2 | 0.2144255 | 0.83023989 | 1 | 0.214425542 | 0.214425542 | 0.214425542 | 2.29722984 | 0.465298952 | 0.50575 | 2.297229841 | 2.297229841 | 2.297229841 | 0.093340918 | -1.029927932 | FALSE | FALSE | FALSE |
| GSTO1 | 0.2143406 | 0.83025672 | 1 | 0.2143406 | 0.2143406 | 0.2143406 | 2.27482172 | 0.466551046 | 0.4988 | 0.248386801 | 2.274821715 | 2.274821715 | 0.094223032 | -1.025842924 | FALSE | FALSE | FALSE |
| INPP5D | 0.2140857 | 0.83027355 | 0.0289 | 0.214085732 | 0.214085732 | 4.18586467 | 0.25685535 | 0.837472392 | 1 | 0.25685535 | 0.25685535 | 5.035651906 | 0.833487534 | -0.079100891 | FALSE | TRUE | FALSE |
| PURG | 0.2140284 | 0.83029037 | 1 | 0.214028392 | 0.214028392 | 0.214028392 | 0.2268157 | 0.845586206 | 0.00785 | 0.226815697 | 0.226815697 | 0.226815697 | 0.943622484 | -0.025201719 | FALSE | TRUE | FALSE |
| CH25H | 0.2140282 | 0.83030719 | 0.00955 | 0.214028201 | 0.214028201 | 0.214028201 | 2.31480785 | 0.463320059 | 0.0084 | 2.314807847 | 2.314807847 | 2.314807847 | 0.092460461 | -1.034043944 | FALSE | FALSE | FALSE |
| ITPRIPL2 | 0.2139733 | 0.83032401 | 0.02165 | 0.213973332 | 0.213973332 | 4.205624691 | 0.25615463 | 0.837667583 | 1 | 0.256154626 | 0.256154626 | 5.045152906 | 0.835328781 | -0.078142555 | FALSE | TRUE | FALSE |
| DHPS | 0.2138317 | 0.83034083 | 0.5052 | 0.213831694 | 0.213831694 | 0.471970162 | 0.03794043 | 0.889449577 | 0.01145 | 0.037940426 | 0.037940426 | 0.752030528 | 5.635985548 | 0.750969871 | FALSE | TRUE | FALSE |
| FZD7 | 0.2136791 | 0.83035764 | 0.5162 | 0.213679135 | 0.213679135 | 4.186089122 | 0.03668644 | 0.889740595 | 0.5125 | 0.146641971 | 0.146641971 | 2.879878772 | 5.824471888 | 0.765256553 | FALSE | TRUE | FALSE |
| SPPL2A | 0.2136261 | 0.83037445 | 0.01225 | 0.213626131 | 0.213626131 | 4.237471027 | 15.8291965 | 0.154993221 | 0.5066 | 15.82919647 | 151.6484534 | 151.6484534 | 0.013495703 | -1.869804494 | FALSE | FALSE | FALSE |
| MBTPS1 | 0.2135336 | 0.83039125 | 0.51845 | 0.213533638 | 0.853810069 | 7.199381091 | 0.1424152 | 0.866305778 | 0.88135 | 1.288206515 | 11.95952294 | 25.23704987 | 1.49937394 | 0.175909958 | FALSE | TRUE | FALSE |
| OR4S2 | 0.2134296 | 0.83040806 | 1 | 18.47914463 | 1770.792969 | 1770.792969 | 21.5033515 | 0.126149895 | 0.8082 | 21.50335147 | 2072.334943 | 21100.04515 | 0.00992541 | -2.003251534 | FALSE | FALSE | FALSE |
| ZNF574 | 0.2133823 | 0.83042486 | 1 | 0.213382275 | 0.213382275 | 4.165876614 | 0.255923 | 0.837716309 | 1 | 0.255922998 | 0.255922998 | 5.010758052 | 0.833775303 | -0.078950973 | FALSE | TRUE | FALSE |
| CCDC9 | 0.2133674 | 0.83044165 | 1 | 0.213367417 | 0.213367417 | 4.207940465 | 0.25573812 | 0.837748778 | 1 | 0.255738121 | 0.255738121 | 5.052007207 | 0.834319955 | -0.078667369 | FALSE | TRUE | FALSE |
| PPIB | 0.213362 | 0.83045844 | 1 | 0.213361956 | 0.213361956 | 0.213361956 | 1.01028911 | 0.587225374 | 0.0062 | 1.010289111 | 1.010289111 | 1.010289111 | 0.211189008 | -0.675328689 | FALSE | FALSE | FALSE |
| ARNT2 | 0.2132633 | 0.83047523 | 0.02275 | 0.213263257 | 0.213263257 | 4.195948894 | 0.25591832 | 0.837732545 | 1 | 0.255918315 | 0.255918315 | 5.04503651 | 0.833325495 | -0.079185331 | FALSE | TRUE | FALSE |
| RAB11FIP4 | 0.2131493 | 0.83049202 | 1 | 0.213149264 | 0.213149264 | 4.208722731 | 0.23307351 | 0.844055036 | 1 | 0.233073509 | 0.233073509 | 4.609184195 | 0.914515189 | -0.038809077 | FALSE | TRUE | FALSE |
| DPYSL5 | 0.2130891 | 0.8305088 | 0.261 | 0.213089091 | 0.851911758 | 4.197646561 | 0.07718122 | 0.88102475 | 1 | 0.077181223 | 0.077181223 | 1.523148487 | 2.760892903 | 0.44104956 | FALSE | TRUE | FALSE |
| TFG | 0.2130428 | 0.83052558 | 1 | 0.213042796 | 0.213042796 | 4.228847835 | 0.22981265 | 0.844950546 | 1 | 0.229812653 | 0.229812653 | 4.565846291 | 0.927028139 | -0.032907083 | FALSE | TRUE | FALSE |
| RPL37A | 0.2130322 | 0.83054236 | 1 | 0.21303221 | 0.21303221 | 0.21303221 | 0.21273933 | 0.848916889 | 1 | 1.994659214 | 19.19445494 | 19.19445494 | 1.001376729 | 0.000597495 | FALSE | TRUE | FALSE |
| SLC46A1 | 0.2129906 | 0.83055913 | 1 | 1.98702565 | 1.98702565 | 4.214932286 | 0.26369542 | 0.835628126 | 1 | 0.263695421 | 0.263695421 | 5.22495917 | 0.807714292 | -0.092742232 | FALSE | TRUE | FALSE |
| RCSD1 | 0.212987 | 0.8305759 | 1 | 0.212986982 | 0.212986982 | 4.226394848 | 0.25543416 | 0.837781236 | 1 | 0.255434164 | 0.255434164 | 0.255434164 | 0.833823397 | -0.078925923 | FALSE | TRUE | FALSE |
| TRAPPC10 | 0.2129838 | 0.83059267 | 1 | 1.984822771 | 18.98393606 | 369.7329684 | 0.19928342 | 0.852274084 | 0.9992 | 17602.01041 | 22242124.91 | 249555374.6 | 1.0687484 | 0.028875478 | FALSE | TRUE | FALSE |
| CYLC2 | 0.212935 | 0.83060943 | 1 | 0.212934958 | 0.212934958 | 0.212934958 | 0.2546756 | 0.838008083 | 1 | 0.254675604 | 0.254675604 | 0.254675604 | 0.836102691 | -0.077740379 | FALSE | TRUE | FALSE |
| DEK | 0.2128505 | 0.83062619 | 1 | 0.212850528 | 0.212850528 | 0.212850528 | 0.23304928 | 0.844070044 | 1 | 0.233049283 | 0.233049283 | 0.233049283 | 0.913328397 | -0.039373039 | FALSE | TRUE | FALSE |
| MEF2C | 0.2127909 | 0.83064295 | 1 | 0.212790869 | 0.212790869 | 4.213956656 | 0.25534952 | 0.837797461 | 1 | 0.255349521 | 0.255349521 | 5.062740371 | 0.833331774 | -0.079182059 | FALSE | TRUE | FALSE |
| ZNF304 | 0.2127142 | 0.8306597 | 1 | 1.958142967 | 18.50064722 | 38.69044247 | 0.27103724 | 0.833537085 | 1 | 0.271037239 | 0.271037239 | 5.363127707 | 0.784815326 | -0.105232525 | FALSE | TRUE | FALSE |
| KLHL10 | 0.2125955 | 0.83067645 | 1 | 0.850106337 | 0.850106337 | 7.198073063 | 0.25833471 | 0.837178745 | 1 | 0.258334715 | 0.258334715 | 5.106226608 | 0.822945776 | -0.084628779 | FALSE | TRUE | FALSE |
| SRRM1 | 0.2125235 | 0.8306932 | 1 | 0.212523509 | 0.212523509 | 7.152187599 | 0.25507534 | 0.837894738 | 0.02595 | 0.255075338 | 0.255075338 | 5.004684679 | 0.833179369 | -0.079261493 | FALSE | TRUE | FALSE |
| UBB | 0.212498 | 0.83070994 | 0.00695 | 0.212498044 | 0.212498044 | 0.212498044 | 0.25449248 | 0.838040439 | 1 | 0.254492482 | 0.254492482 | 0.254492482 | 0.834987508 | -0.078320022 | FALSE | TRUE | FALSE |
| OR1D2 | 0.2124907 | 0.83072668 | 1 | 1.960824677 | 1.960824677 | 1.960824677 | 0.25148163 | 0.838925135 | 1 | 0.251481631 | 0.251481631 | 0.251481631 | 0.84495514 | -0.073166348 | FALSE | TRUE | FALSE |
| MYBBP1A | 0.2124677 | 0.83074342 | 0.32775 | 0.025796776 | 1.793951721 | 4.115988822 | 0.06277685 | 0.884075847 | 0.6947 | 0.062776851 | 4.550625836 | 10.26522178 | 3.384491858 | 0.529493474 | FALSE | TRUE | FALSE |
| MGRN1 | 0.2123984 | 0.83076015 | 1 | 0.849520515 | 0.849520515 | 7.194036786 | 1.00749825 | 0.587783052 | 0.75285 | 1.007498251 | 4.030679133 | 19.87756813 | 0.210817665 | -0.676093 | FALSE | FALSE | FALSE |
| ZNF596 | 0.2123753 | 0.83077688 | 0.01355 | 0.212375336 | 0.212375336 | 0.480434 | 0.03497817 | 0.890131686 | 1 | 0.308862659 | 26.02184817 | 26.02184817 | 6.071653465 | 0.783306977 | FALSE | TRUE | FALSE |
| FKBP5 | 0.2122161 | 0.83079361 | 1 | 0.212216052 | 0.212216052 | 0.212216052 | 0.25471463 | 0.8379919 | 1 | 0.254714627 | 0.254714627 | 0.254714627 | 0.833152199 | -0.079275655 | FALSE | TRUE | FALSE |
| GIMAP6 | 0.2121946 | 0.83081034 | 1 | 0.212194647 | 0.212194647 | 4.216914392 | 0.23892911 | 0.842355596 | 1 | 2.19181007 | 20.63626182 | 20.63626182 | 0.888107126 | -0.051534645 | FALSE | TRUE | FALSE |
| KLHDC8B | 0.212189 | 0.83082706 | 1 | 0.212189018 | 0.212189018 | 4.20479695 | 0.25480466 | 0.837975715 | 1 | 0.254804661 | 0.254804661 | 0.254804661 | 0.83275171 | -0.079484467 | FALSE | TRUE | FALSE |
| GRHL2 | 0.2121537 | 0.83084377 | 1 | 0.212153671 | 0.212153671 | 4.187628914 | 0.23226988 | 0.844369612 | 1 | 0.232269882 | 0.232269882 | 4.591904923 | 0.913392945 | -0.039342347 | FALSE | TRUE | FALSE |
| FAM120AOS | 0.212085 | 0.83086049 | 1 | 0.212084955 | 0.212084955 | 0.212084955 | 0.25378453 | 0.838266569 | 1 | 0.253784531 | 0.253784531 | 0.253784531 | 0.835689054 | -0.077955286 | FALSE | TRUE | FALSE |
| CABS1 | 0.21205 | 0.8308772 | 1 | 0.212049967 | 0.212049967 | 0.212049967 | 0.24610935 | 0.840273763 | 1 | 0.246109354 | 2.256384128 | 2.256384128 | 0.861608729 | -0.06468991 | FALSE | TRUE | FALSE |
| FBXL2 | 0.2119603 | 0.83089391 | 1 | 0.211960261 | 0.211960261 | 4.202092686 | 0.25394691 | 0.838202025 | 1 | 0.253946912 | 0.253946912 | 5.039775102 | 0.834663668 | -0.07848849 | FALSE | TRUE | FALSE |
| RPS6KA4 | 0.2118311 | 0.83091061 | 1 | 0.211831133 | 0.211831133 | 4.152929591 | 0.25430417 | 0.838105112 | 1 | 0.254304168 | 0.254304168 | 4.997470539 | 0.832983333 | -0.079363688 | FALSE | TRUE | FALSE |
| ACP5 | 0.211789 | 0.83092731 | 1 | 0.211788952 | 0.211788952 | 4.201486706 | 0.25429621 | 0.838137429 | 1 | 0.254296206 | 0.254296206 | 0.254296206 | 0.832843537 | -0.07943658 | FALSE | TRUE | FALSE |
| VEGFB | 0.2117097 | 0.83094401 | 0.0055 | 0.21170971 | 0.21170971 | 0.21170971 | 0.25051279 | 0.839276358 | 1 | 1.002027419 | 1.002027419 | 1.002027419 | 0.845105383 | -0.073089132 | FALSE | TRUE | FALSE |
| DAPK3 | 0.2116626 | 0.83096071 | 1 | 0.211662625 | 0.211662625 | 4.170947906 | 0.2537903 | 0.838250438 | 0.0153 | 0.253790303 | 0.253790303 | 5.009959836 | 0.834005959 | -0.078830846 | FALSE | TRUE | FALSE |
| OLFML3 | 0.2116473 | 0.8309774 | 0.50645 | 0.211647273 | 1.974919707 | 1.974919707 | 0.0224193 | 0.893091079 | 0.259 | 0.0224193 | 0.089613603 | 0.111286824 | 9.440405023 | 0.974990627 | FALSE | TRUE | FALSE |
| POLR2C | 0.2115714 | 0.83099409 | 1 | 0.211571383 | 0.211571383 | 0.211571383 | 0.25410574 | 0.838153583 | 1 | 0.254105736 | 0.254105736 | 0.254105736 | 0.832611597 | -0.079557544 | FALSE | TRUE | FALSE |
| C4orf51 | 0.2115712 | 0.83101077 | 1 | 0.211571239 | 0.211571239 | 0.211571239 | 0.25316392 | 0.838572428 | 1 | 0.253163916 | 0.253163916 | 0.253163916 | 0.835708509 | -0.077945176 | FALSE | TRUE | FALSE |
| KIF11 | 0.2115326 | 0.83102746 | 1 | 0.21153257 | 0.21153257 | 4.156264256 | 0.25437382 | 0.838088949 | 1 | 0.254373823 | 0.254373823 | 5.008604472 | 0.831581518 | -0.080095172 | FALSE | TRUE | FALSE |
| SMYD2 | 0.2115214 | 0.83104413 | 0.0109 | 0.211521398 | 0.211521398 | 4.194616673 | 0.25366269 | 0.838363285 | 0.01115 | 0.253662685 | 0.253662685 | 5.035429002 | 0.833868797 | -0.078902277 | FALSE | TRUE | FALSE |
| CMTM1 | 0.2114607 | 0.83106081 | 1 | 1.933415435 | 1.933415435 | 1.933415435 | 0.26106081 | 0.836357597 | 1 | 0.26106081 | 0.26106081 | 0.26106081 | 0.810005595 | -0.091511981 | FALSE | TRUE | FALSE |
| SNX1 | 0.2113588 | 0.83107748 | 1 | 1.950213994 | 1.950213994 | 4.183987748 | 0.26142963 | 0.836258502 | 1 | 0.261429628 | 0.261429628 | 5.181532982 | 0.808473078 | -0.092334438 | FALSE | TRUE | FALSE |
| LRG1 | 0.2113341 | 0.83109415 | 0.7529 | 1.943856595 | 18.35074952 | 18.35074952 | 2.41390921 | 0.453956855 | 0.50905 | 2.413909207 | 2.413909207 | 2.413909207 | 0.08754848 | -1.057751388 | FALSE | FALSE | FALSE |
| KCNJ16 | 0.2113203 | 0.83111082 | 0.9896 | 714.7690646 | 288779.6789 | 3005854.141 | 1.1156957 | 0.576840246 | 0.38465 | 0.477884994 | 17.82525194 | 17.82525194 | 0.189406763 | -0.722604518 | FALSE | FALSE | FALSE |
| SLC35B1 | 0.2112948 | 0.83112748 | 1 | 0.211294819 | 0.211294819 | 0.211294819 | 0.25353269 | 0.838427695 | 1 | 0.253532693 | 0.253532693 | 0.253532693 | 0.83340265 | -0.079145123 | FALSE | TRUE | FALSE |
| C4BPA | 0.2112373 | 0.83114414 | 1 | 73.68581998 | 1778.146363 | 7112.235985 | 0.28997028 | 0.828161773 | 1 | 0.289970284 | 0.289970284 | 5.742814883 | 0.728479225 | -0.137582829 | FALSE | TRUE | FALSE |
| EBF1 | 0.2112056 | 0.83116079 | 1 | 0.844586881 | 0.844586881 | 7.152214778 | 0.25393259 | 0.838218166 | 1 | 1.015480622 | 1.015480622 | 8.603253482 | 0.831738805 | -0.080013036 | FALSE | TRUE | FALSE |
| BNC2 | 0.211144 | 0.83117745 | 1 | 0.211144047 | 0.211144047 | 4.129413876 | 0.25373494 | 0.838314943 | 1 | 0.253734936 | 0.253734936 | 4.975609185 | 0.832144167 | -0.079801427 | FALSE | TRUE | FALSE |
| CCDC149 | 0.2110966 | 0.8311941 | 1 | 0.211096576 | 0.211096576 | 4.198065714 | 0.25323783 | 0.838524213 | 1 | 0.253237833 | 0.253237833 | 0.253237833 | 0.833590199 | -0.0790474 | FALSE | TRUE | FALSE |
| ZMYM4 | 0.2109737 | 0.83121074 | 0.03985 | 0.210973706 | 0.210973706 | 7.083370558 | 0.24570072 | 0.840446701 | 1 | 2.296364327 | 2.296364327 | 44.8563731 | 0.858661319 | -0.066178101 | FALSE | TRUE | FALSE |
| ZFAND2B | 0.2109017 | 0.83122738 | 0.5011 | 1.943096651 | 1.943096651 | 1.943096651 | 0.26085129 | 0.836440084 | 1 | 0.260851288 | 0.260851288 | 0.260851288 | 0.808513302 | -0.09231283 | FALSE | TRUE | FALSE |
| HOXB8 | 0.210879 | 0.83124402 | 1 | 11549.86688 | 3085072.701 | 18892974.05 | 0.23787214 | 0.842615884 | 1 | 130700.3435 | 56147627.24 | 632694419.6 | 0.886522662 | -0.052310158 | FALSE | TRUE | FALSE |

| | | | | | | | | | | | | | | | | | |
|---|---|---|---|---|---|---|---|---|---|---|---|---|---|---|---|---|---|
| SPOCK3 | 0.2108676 | 0.83126066 | 1 | 0.210867551 | 0.210867551 | 0.210867551 | 0.25317108 | 0.838556359 | 1 | 0.253171081 | 0.253171081 | 0.253171081 | 0.832905362 | -0.079404342 | FALSE | TRUE | FALSE |
| SCAF4 | 0.2108488 | 0.83127729 | 0.0357 | 0.210848782 | 0.210848782 | 7.09557798 | 8.53249674 | 0.217868223 | 0.0098 | 0.253236181 | 0.253236181 | 4.96002599 | 0.024711264 | -1.607105034 | FALSE | FALSE | FALSE |
| GALNT1 | 0.2108106 | 0.83129392 | 0.01445 | 0.210810611 | 0.210810611 | 4.174771004 | 0.25323099 | 0.838540288 | 1 | 0.253230989 | 0.253230989 | 5.020768579 | 0.832483466 | -0.079624383 | FALSE | TRUE | FALSE |
| GPR37 | 0.2108029 | 0.83131055 | 1 | 0.210802854 | 0.210802854 | 4.148797645 | 0.25303882 | 0.838604554 | 1 | 0.25303882 | 0.25303882 | 4.989604513 | 0.833085036 | -0.079310666 | FALSE | TRUE | FALSE |
| OR6Q1 | 0.2107916 | 0.83132717 | 0.0093 | 0.210791581 | 0.210791581 | 0.210791581 | 0.25256076 | 0.838764996 | 1 | 0.25256076 | 0.25256076 | 0.25256076 | 0.834617308 | -0.078512613 | FALSE | TRUE | FALSE |
| KAZN | 0.2107659 | 0.83134379 | 0.02395 | 0.210765932 | 0.210765932 | 4.136412242 | 0.2534007 | 0.838475968 | 0.02215 | 0.253400704 | 0.253400704 | 4.984355829 | 0.831749592 | -0.080007404 | FALSE | TRUE | FALSE |
| CABLES1 | 0.2107091 | 0.83136041 | 1 | 0.21070913 | 0.21070913 | 4.16896911 | 0.25264708 | 0.838748966 | 1 | 0.252647081 | 0.252647081 | 5.005180928 | 0.834005799 | -0.07883093 | FALSE | TRUE | FALSE |
| SNX13 | 0.2106414 | 0.83137702 | 1 | 1.946832339 | 18.46742761 | 38.29926176 | 0.26906871 | 0.834064788 | 1 | 0.269068706 | 0.269068706 | 5.303713151 | 0.782853494 | -0.106319506 | FALSE | TRUE | FALSE |
| CACNG5 | 0.2106398 | 0.83139363 | 0.9681 | 4.641143664 | 74.1842246 | 296.7000716 | 1.26795176 | 0.559283137 | 0.50425 | 1.267951758 | 5.071550646 | 5.071550646 | 0.166126037 | -0.779562296 | FALSE | FALSE | FALSE |
| RBFOX1 | 0.2106184 | 0.83141024 | 1 | 1.953050823 | 1.953050823 | 4.164292472 | 0.24180155 | 0.841538653 | 1 | 2.248421992 | 2.248421992 | 4.787403105 | 0.871038056 | -0.05996287 | FALSE | TRUE | FALSE |
| EIF2B3 | 0.2105761 | 0.83142684 | 0.8764 | 0.841890642 | 3.366749784 | 3.366749784 | 1.19300413 | 0.568284694 | 0.50515 | 1.193004127 | 4.771281638 | 4.771281638 | 0.176509104 | -0.753232889 | FALSE | FALSE | FALSE |
| FFAR4 | 0.2105182 | 0.83144344 | 1 | 0.210518182 | 0.210518182 | 4.162417428 | 0.24473261 | 0.840666259 | 1 | 2.252956024 | 2.252956024 | 4.845543105 | 0.860196683 | -0.065402236 | FALSE | TRUE | FALSE |
| USP8 | 0.2104769 | 0.83146004 | 0.0284 | 0.210476856 | 0.210476856 | 4.127564903 | 0.25282795 | 0.838700858 | 1 | 0.252827949 | 0.252827949 | 4.969719745 | 0.832490461 | -0.079620734 | FALSE | TRUE | FALSE |
| IL4R | 0.2104434 | 0.83147663 | 1 | 0.210443355 | 0.210443355 | 4.138769978 | 0.25253737 | 0.838781022 | 1 | 0.25253737 | 0.25253737 | 4.976587032 | 0.8333157 | -0.079190436 | FALSE | TRUE | FALSE |
| ASIC2 | 0.2104348 | 0.83149322 | 1 | 0.21043485 | 0.21043485 | 4.15207299 | 0.2529508 | 0.838636669 | 1 | 0.2529508 | 0.2529508 | 4.999032348 | 0.83192008 | -0.079918393 | FALSE | TRUE | FALSE |
| NDUFA6 | 0.2104337 | 0.83150981 | 1 | 0.21043367 | 0.21043367 | 0.21043367 | 20.9251231 | 0.128292392 | 0.2551 | 2.200114228 | 20.9251231 | 20.9251231 | 0.010056508 | -1.997552792 | FALSE | FALSE | FALSE |
| BCKDK | 0.210137 | 0.83152639 | 1 | 1.949858366 | 7.79824847 | 16.62168896 | 0.263274 | 0.835711347 | 0.0105 | 0.263273997 | 0.263273997 | 5.21394768 | 0.798168309 | -0.09790552 | FALSE | TRUE | FALSE |
| PDE4B | 0.2100958 | 0.83154297 | 0.75395 | 0.210095766 | 0.840127675 | 4.142934061 | 0.07607319 | 0.881222471 | 1 | 0.076073194 | 0.076073194 | 1.502641569 | 2.761758197 | 0.441185652 | FALSE | TRUE | FALSE |
| GPR17 | 0.2100831 | 0.83155955 | 1 | 0.210083076 | 0.210083076 | 4.1591331 | 2.1990147 | 0.472212428 | 0.75375 | 2.199014703 | 20.93073105 | 20.93073105 | 0.095535094 | -1.019837066 | FALSE | FALSE | FALSE |
| ZNF75A | 0.20997 | 0.83157612 | 1 | 0.209969964 | 0.209969964 | 0.209969964 | 2.25730298 | 0.467798064 | 0.0057 | 2.257302982 | 2.257302982 | 2.257302982 | 0.093018069 | -1.031432681 | FALSE | FALSE | FALSE |
| SH3BP5 | 0.2097451 | 0.8315927 | 0.0119 | 0.209745069 | 0.209745069 | 4.155144425 | 0.25219058 | 0.838829085 | 1 | 0.252190575 | 0.252190575 | 5.001704304 | 0.831692734 | -0.080037092 | FALSE | TRUE | FALSE |
| SRSF12 | 0.209736 | 0.83160926 | 1 | 1.974118155 | 1.974118155 | 1.974118155 | 0.22349891 | 0.846508444 | 1 | 2.105903093 | 2.105903093 | 2.105903093 | 0.938420467 | -0.027602529 | FALSE | TRUE | FALSE |
| GPR148 | 0.2097303 | 0.83162583 | 1 | 1.917639183 | 1.917639183 | 1.917639183 | 0.25135239 | 0.839005092 | 1 | 2.308229665 | 2.308229665 | 2.308229665 | 0.834407431 | -0.078621837 | FALSE | TRUE | FALSE |
| HS3ST1 | 0.2096597 | 0.83164239 | 1 | 0.209659702 | 0.209659702 | 4.153514117 | 0.25212769 | 0.8388451 | 1 | 0.252127693 | 0.252127693 | 5.000521695 | 0.831561577 | -0.080105586 | FALSE | TRUE | FALSE |
| DPP9 | 0.2095852 | 0.83165895 | 1 | 0.209585181 | 0.209585181 | 4.101487103 | 2.19719722 | 0.472387545 | 0.75815 | 2.197197223 | 20.9368203 | 43.10962894 | 0.095387514 | -1.02050847 | FALSE | FALSE | FALSE |
| TYMP | 0.2095803 | 0.83167551 | 0.3797 | 0.092301722 | 0.837622242 | 0.837622242 | 0.63220064 | 0.698268464 | 0.9102 | 10.11310393 | 369.1101348 | 1477.208703 | 0.331509227 | -0.47950438 | FALSE | TRUE | FALSE |
| CCR9 | 0.2095776 | 0.83169206 | 1 | 0.209577557 | 0.209577557 | 4.163272804 | 0.24857342 | 0.839879408 | 0.4959 | 0.248573424 | 0.99423867 | 0.99423867 | 0.843121336 | -0.07410992 | FALSE | TRUE | FALSE |
| ATF7IP | 0.2095571 | 0.83170861 | 1 | 1.955147647 | 18.72171806 | 65.80796765 | 0.2467976 | 0.840147761 | 1 | 2.307371288 | 22.14018654 | 77.75714516 | 0.849105058 | -0.071038572 | FALSE | TRUE | FALSE |
| ZFP3 | 0.2094398 | 0.83172516 | 0.50985 | 1.937875055 | 1.937875055 | 4.153990799 | 0.2594657 | 0.836900466 | 1 | 0.259465704 | 0.259465704 | 5.15133797 | 0.807196476 | -0.093020743 | FALSE | TRUE | FALSE |
| CTSC | 0.2093861 | 0.8317417 | 0.31225 | 0.209386081 | 3.349185214 | 3.349185214 | 0.76134532 | 0.635809966 | 0.6253 | 1.347664761 | 12.17405135 | 48.6995139 | 0.27502117 | -0.560633875 | FALSE | TRUE | FALSE |
| MRPL45 | 0.2093744 | 0.83175824 | 1 | 1.942643341 | 18.49923814 | 18.49923814 | 0.26767943 | 0.834420387 | 0.00715 | 0.267679428 | 0.267679428 | 0.267679428 | 0.7821834 | -0.106691405 | FALSE | TRUE | FALSE |
| SH3RF3 | 0.2093384 | 0.83177478 | 1 | 0.837268284 | 0.837268284 | 16.48491818 | 307.098193 | 0.007998953 | 0.0002 | 2.268488488 | 9.073140323 | 19.24209943 | 0.000681666 | -3.166428464 | FALSE | FALSE | FALSE |
| RFX4 | 0.2092959 | 0.83179131 | 1 | 0.209295915 | 0.209295915 | 4.122478976 | 0.25194263 | 0.838877121 | 1 | 0.251942632 | 0.251942632 | 4.971539134 | 0.830728459 | -0.080540911 | FALSE | TRUE | FALSE |
| MYSM1 | 0.2092384 | 0.83180784 | 1 | 0.209238417 | 0.209238417 | 4.123620655 | 3.88965253 | 0.361579757 | 0.5105 | 3.889652535 | 15.56563831 | 32.85254519 | 0.053793601 | -1.269269382 | FALSE | FALSE | FALSE |
| PHKG2 | 0.2091899 | 0.83182437 | 1 | 0.209189922 | 0.209189922 | 4.141880717 | 0.25138456 | 0.838973119 | 1 | 0.251384559 | 0.251384559 | 4.98331704 | 0.832151039 | -0.07979784 | FALSE | TRUE | FALSE |
| SMARCC1 | 0.2091859 | 0.83184089 | 0.02995 | 0.209185949 | 0.209185949 | 4.09410961 | 0.24392967 | 0.840854008 | 1 | 2.285518798 | 2.285518798 | 44.84672935 | 0.857566661 | -0.066732111 | FALSE | TRUE | FALSE |
| DRAXIN | 0.2090626 | 0.83185741 | 0.0121 | 0.209062642 | 0.209062642 | 4.144268743 | 0.25095252 | 0.839069 | 1 | 0.25095252 | 0.25095252 | 0.25095252 | 0.833076479 | -0.079315127 | FALSE | TRUE | FALSE |
| NUP153 | 0.2090314 | 0.83187393 | 0.0374 | 0.209031422 | 0.209031422 | 7.032663713 | 0.25159004 | 0.838893129 | 1 | 0.251590039 | 0.251590039 | 8.475168074 | 0.830841408 | -0.080481867 | FALSE | TRUE | FALSE |
| CACNG4 | 0.2089927 | 0.83189044 | 0.0117 | 0.208992676 | 0.208992676 | 4.138207429 | 0.24877386 | 0.839768658 | 0.50535 | 0.994788226 | 0.994788226 | 4.931807499 | 0.840090994 | -0.075673671 | FALSE | TRUE | FALSE |
| AZU1 | 0.2089735 | 0.83190695 | 0.74955 | 0.208973508 | 1.924750155 | 1.924750155 | 0.33607994 | 0.813459905 | 0.00895 | 0.336079942 | 0.336079942 | 0.336079942 | 0.621797025 | -0.20635136 | FALSE | TRUE | FALSE |
| CTPS1 | 0.2089548 | 0.83192346 | 1 | 0.208954833 | 0.208954833 | 4.131624284 | 0.25137941 | 0.838989107 | 0.0154 | 0.251379407 | 0.251379407 | 4.977270226 | 0.831232896 | -0.080277278 | FALSE | TRUE | FALSE |
| USP5 | 0.2088842 | 0.83193996 | 1 | 0.208884198 | 0.208884198 | 4.093227776 | 0.25142504 | 0.838957128 | 0.0217 | 0.251425038 | 0.251425038 | 4.938840108 | 0.830801101 | -0.080502936 | FALSE | TRUE | FALSE |
| LAIR1 | 0.2087767 | 0.83195646 | 1 | 0.208776702 | 0.208776702 | 0.208776702 | 0.25032621 | 0.839355964 | 1 | 0.250326214 | 0.250326214 | 0.250326214 | 0.834018533 | -0.078824299 | FALSE | TRUE | FALSE |
| MGAT4A | 0.2087468 | 0.83197296 | 1 | 0.208746794 | 0.208746794 | 4.136247492 | 0.25063926 | 0.839180724 | 1 | 0.25063926 | 0.25063926 | 4.971868079 | 0.832857526 | -0.079429285 | FALSE | TRUE | FALSE |
| ETNK2 | 0.2086145 | 0.83198946 | 1 | 0.208614519 | 0.208614519 | 0.208614519 | 0.2507287 | 0.839132862 | 1 | 0.250728698 | 0.250728698 | 0.250728698 | 0.832032873 | -0.079859515 | FALSE | TRUE | FALSE |
| SEZ6L | 0.2086018 | 0.83200595 | 1 | 0.208601803 | 0.208601803 | 4.078343247 | 0.24052398 | 0.841894201 | 1 | 2.245690339 | 8.982074533 | 44.02394444 | 0.867280687 | -0.061840325 | FALSE | TRUE | FALSE |
| MAP7 | 0.2084722 | 0.83202244 | 0.0231 | 0.208472154 | 0.208472154 | 4.098188754 | 0.25058271 | 0.839228556 | 1 | 0.250582709 | 0.250582709 | 4.936141783 | 0.831949479 | -0.079903046 | FALSE | TRUE | FALSE |
| OR7D4 | 0.2083285 | 0.83203892 | 1 | 1.903381923 | 1.903381923 | 1.903381923 | 0.25757997 | 0.837325697 | 1 | 0.257579972 | 0.257579972 | 0.257579972 | 0.808791523 | -0.092163409 | FALSE | TRUE | FALSE |
| E2F4 | 0.2082288 | 0.83205541 | 1 | 0.208228815 | 0.208228815 | 4.13079337 | 0.25075727 | 0.839116902 | 1 | 0.250757271 | 0.250757271 | 0.250757271 | 0.830399909 | -0.080712707 | FALSE | TRUE | FALSE |
| HDHD2 | 0.2082192 | 0.83207189 | 1 | 0.208219238 | 0.208219238 | 0.208219238 | 0.24978398 | 0.839514938 | 1 | 0.249783984 | 0.249783984 | 0.249783984 | 0.833597234 | -0.079043735 | FALSE | TRUE | FALSE |
| ATF2 | 0.2082129 | 0.83208836 | 0.014 | 0.208212856 | 0.208212856 | 4.126217691 | 0.25070997 | 0.83914882 | 1 | 0.250709965 | 0.250709965 | 4.97385463 | 0.830492938 | -0.080664056 | FALSE | TRUE | FALSE |
| ASAP1 | 0.2080818 | 0.83210484 | 1 | 0.208081811 | 0.208081811 | 4.076355972 | 0.25040595 | 0.83930821 | 1 | 0.250405955 | 0.250405955 | 4.917599859 | 0.830977886 | -0.080410533 | FALSE | TRUE | FALSE |
| CPM | 0.2080245 | 0.83212131 | 1 | 0.208024464 | 0.208024464 | 4.12644048 | 0.24977003 | 0.839562568 | 1 | 0.24977003 | 0.24977003 | 0.24977003 | 0.83286399 | -0.079425915 | FALSE | TRUE | FALSE |
| IFI27L1 | 0.2079274 | 0.83213778 | 1 | 0.207927399 | 0.207927399 | 0.207927399 | 0.21541228 | 0.848278904 | 1 | 0.215412283 | 0.215412283 | 0.215412283 | 0.965253216 | -0.015358743 | FALSE | TRUE | FALSE |
| MUC13 | 0.2079056 | 0.83215424 | 1 | 0.20790565 | 0.20790565 | 4.119449586 | 0.24992388 | 0.839499055 | 0.01065 | 0.249923879 | 0.249923879 | 4.957538473 | 0.831875892 | -0.079941462 | FALSE | TRUE | FALSE |
| RAB4B | 0.2078334 | 0.8321707 | 1 | 0.20783342 | 0.20783342 | 0.20783342 | 0.25014106 | 0.839387784 | 1 | 0.250141063 | 0.250141063 | 0.250141063 | 0.830864865 | -0.080469606 | FALSE | TRUE | FALSE |
| NKX3-2 | 0.2078302 | 0.83218716 | 1 | 0.207830189 | 0.207830189 | 4.118601032 | 0.2503741 | 0.83934005 | 1 | 0.250374103 | 0.250374103 | 0.250374103 | 0.830078618 | -0.080880773 | FALSE | TRUE | FALSE |
| GPR75 | 0.2078012 | 0.83220362 | 1 | 0.207801206 | 0.207801206 | 4.113670069 | 2.23671504 | 0.468509088 | 0.5116 | 2.236715037 | 2.236715037 | 4.798261658 | 0.09290464 | -1.031962594 | FALSE | FALSE | FALSE |
| MPV17L | 0.2077516 | 0.83222007 | 1 | 1.926577406 | 1.926577406 | 1.926577406 | 0.24977991 | 0.839530818 | 0.50055 | 2.322727511 | 2.322727511 | 2.322727511 | 0.831738601 | -0.080013142 | FALSE | TRUE | FALSE |
| TIGD5 | 0.2077079 | 0.83223652 | 1 | 0.20770788 | 0.20770788 | 4.093884974 | 0.22677627 | 0.845615651 | 1 | 0.226776265 | 0.226776265 | 4.477524531 | 0.915915428 | -0.038144625 | FALSE | TRUE | FALSE |
| NXPE3 | 0.2076127 | 0.83225296 | 0.01485 | 0.207612664 | 0.207612664 | 4.106323424 | 0.24930195 | 0.839626036 | 0.0126 | 0.249301953 | 0.249301953 | 4.937448075 | 0.832775921 | -0.079471841 | FALSE | TRUE | FALSE |
| GNA11 | 0.2074946 | 0.8322694 | 1 | 0.207494555 | 0.207494555 | 4.098991857 | 0.24726027 | 0.840100463 | 1 | 0.988721836 | 0.988721836 | 4.891759917 | 0.839174674 | -0.076147632 | FALSE | TRUE | FALSE |
| SAMD8 | 0.2074539 | 0.83228584 | 1 | 0.207453869 | 0.207453869 | 0.207453869 | 0.2494321 | 0.83957844 | 1 | 0.249432101 | 0.249432101 | 0.249432101 | 0.831704774 | -0.080030806 | FALSE | TRUE | FALSE |
| ENO1 | 0.2073532 | 0.83230228 | 0.50645 | 0.207353217 | 0.207353217 | 0.207353217 | 0.07403214 | 0.881607252 | 1 | 0.074032136 | 0.074032136 | 0.074032136 | 2.800854159 | 0.447290496 | FALSE | TRUE | FALSE |
| PLD3 | 0.2073434 | 0.83231871 | 1 | 1.929455474 | 1.929455474 | 4.095790987 | 0.25742603 | 0.837358318 | 1 | 0.257426033 | 0.257426033 | 5.092642365 | 0.805448474 | -0.093962237 | FALSE | TRUE | FALSE |
| CCIN | 0.2073408 | 0.83233514 | 1 | 0.207340816 | 0.207340816 | 4.089922048 | 0.24931314 | 0.839610174 | 1 | 0.249313142 | 0.249313142 | 4.925971251 | 0.831648161 | -0.080060368 | FALSE | TRUE | FALSE |
| HMX3 | 0.2072312 | 0.83235157 | 0.75695 | 1.961484978 | 19.05449281 | 19.05449281 | 0.24194944 | 0.84144564 | 0.87745 | 22.22461826 | 221.3235369 | 221.3235369 | 0.856506396 | -0.067269389 | FALSE | TRUE | FALSE |
| SPDYC | 0.207216 | 0.83236799 | 0.00855 | 0.207216001 | 0.207216001 | 0.207216001 | 0.248683 | 0.839816141 | 1 | 0.248682999 | 0.248682999 | 0.248682999 | 0.833253586 | -0.079222809 | FALSE | TRUE | FALSE |
| LYPD1 | 0.2071388 | 0.83238441 | 1 | 0.207138795 | 0.207138795 | 0.207138795 | 2.13732879 | 0.477593237 | 0.74995 | 2.137328789 | 81.06833335 | 81.06833335 | 0.096914801 | -1.01360989 | FALSE | FALSE | FALSE |
| CCNB1IP1 | 0.2070889 | 0.83240083 | 1 | 0.207088934 | 0.207088934 | 0.207088934 | 0.24896924 | 0.839689455 | 1 | 0.248969235 | 0.248969235 | 0.248969235 | 0.831785233 | -0.079988794 | FALSE | TRUE | FALSE |
| EFCAB11 | 0.2070359 | 0.83241724 | 0.00505 | 0.207035931 | 0.207035931 | 0.207035931 | 0.24896767 | 0.839705302 | 1 | 0.248967668 | 0.248967668 | 0.248967668 | 0.831577582 | -0.080097227 | FALSE | TRUE | FALSE |
| LBX2 | 0.2069925 | 0.83243365 | 0.75175 | 1.927186402 | 7.707049527 | 7.707049527 | 0.25706722 | 0.837439815 | 1 | 1.028069318 | 1.028069318 | 5.106047064 | 0.805207543 | -0.094092166 | FALSE | TRUE | FALSE |
| OPN3 | 0.2069319 | 0.83245006 | 0.0123 | 0.206931857 | 0.206931857 | 4.103573616 | 0.24902833 | 0.839673604 | 1 | 0.249028333 | 0.249028333 | 4.943401963 | 0.830957082 | -0.080421406 | FALSE | TRUE | FALSE |

| | | | | | | | | | | | | | | | | | |
|---|---|---|---|---|---|---|---|---|---|---|---|---|---|---|---|---|---|
| LIPH | 0.2067629 | 0.83246647 | 0.12695 | 0.092339087 | 0.826480333 | 0.826480333 | 0.0439502 | 0.888102482 | 0.50205 | 0.043950197 | 0.043950197 | 0.218388602 | 4.704482414 | 0.672511849 | FALSE | TRUE | FALSE |
| FGF11 | 0.2067375 | 0.83248287 | 1 | 0.206737506 | 0.206737506 | 0.206737506 | 0.24855341 | 0.839895217 | 1 | 0.248553407 | 0.248553407 | 0.248553407 | 0.831762912 | -0.080000448 | FALSE | TRUE | FALSE |
| REP15 | 0.2067177 | 0.83249927 | 1 | 0.206717676 | 0.206717676 | 0.206717676 | 2.19930384 | 0.472037199 | 0.50345 | 2.199303845 | 2.199303845 | 2.199303845 | 0.093992322 | -1.02690762 | FALSE | FALSE | FALSE |
| KIAA0232 | 0.2066222 | 0.83251566 | 1 | 18.31239932 | 179.1347112 | 357.5854814 | 0.27309014 | 0.832920073 | 1 | 0.273090138 | 0.273090138 | 5.347816761 | 0.756607895 | -0.121129131 | FALSE | TRUE | FALSE |
| ULBP2 | 0.2066012 | 0.83253206 | 1 | 1.88335829 | 169.0997607 | 169.0997607 | 0.27166631 | 0.833331923 | 1 | 0.271666311 | 0.271666311 | 0.271666311 | 0.760496306 | -0.118902891 | FALSE | TRUE | FALSE |
| ISLR | 0.2064876 | 0.83254845 | 1 | 0.206487583 | 0.206487583 | 4.083137307 | 0.95290592 | 0.592526244 | 0.7566 | 2.217662983 | 8.86929593 | 8.86929593 | 0.216692518 | -0.664156084 | FALSE | TRUE | FALSE |
| GRHL1 | 0.2064833 | 0.83256483 | 1 | 0.206483277 | 0.206483277 | 4.082571135 | 0.22977458 | 0.844980223 | 1 | 0.229774585 | 0.229774585 | 4.549318151 | 0.898634099 | -0.046417106 | FALSE | TRUE | FALSE |
| PPARGC1B | 0.2064222 | 0.83258122 | 1 | 0.206422181 | 0.206422181 | 4.035196023 | 0.24847306 | 0.839926824 | 1 | 0.248473059 | 0.248473059 | 4.870434529 | 0.830762829 | -0.080522943 | FALSE | TRUE | FALSE |
| PHF21A | 0.2063797 | 0.8325976 | 1 | 0.206379748 | 0.206379748 | 4.073002198 | 0.24891213 | 0.839721145 | 1 | 0.248912135 | 0.248912135 | 4.920217572 | 0.829126906 | -0.081378992 | FALSE | TRUE | FALSE |
| SNX24 | 0.206361 | 0.83261397 | 0.508 | 1.880686777 | 1.880686777 | 1.880686777 | 0.25527583 | 0.837846114 | 1 | 0.255275834 | 0.255275834 | 0.255275834 | 0.808384447 | -0.092382051 | FALSE | TRUE | FALSE |
| CEP70 | 0.2063407 | 0.83263034 | 0.5068 | 0.20634073 | 0.20634073 | 0.474871332 | 0.03648309 | 0.889779817 | 1 | 0.036483085 | 0.317638056 | 0.723861645 | 5.655791713 | 0.752493407 | FALSE | TRUE | FALSE |
| GPX1 | 0.2063372 | 0.83264671 | 1 | 1.909963724 | 18.14533864 | 18.14533864 | 0.21661185 | 0.848065107 | 1 | 19.09341276 | 1862.595877 | 1862.595877 | 0.95256633 | -0.021104773 | FALSE | TRUE | FALSE |
| ZFP64 | 0.2062046 | 0.83266308 | 0.0223 | 0.206204643 | 0.206204643 | 4.051302199 | 0.24818677 | 0.839974212 | 0.0191 | 0.248186769 | 0.248186769 | 4.886487048 | 0.830844625 | -0.080480185 | FALSE | TRUE | FALSE |
| TBRG1 | 0.2060408 | 0.83267945 | 1 | 0.206040754 | 0.206040754 | 0.206040754 | 0.2405384 | 0.841878775 | 1 | 0.2405384 | 2.243982959 | 2.243982959 | 0.856581541 | -0.067231289 | FALSE | TRUE | FALSE |
| RASL11A | 0.205952 | 0.83269581 | 1 | 0.205951985 | 0.205951985 | 0.205951985 | 0.248034 | 0.84000579 | 1 | 0.248033998 | 0.248033998 | 0.248033998 | 0.830337723 | -0.080745231 | FALSE | TRUE | FALSE |
| TBC1D10B | 0.2059344 | 0.83271217 | 1 | 1.938169899 | 18.7215031 | 38.34721056 | 0.26744104 | 0.83448795 | 1 | 0.267441044 | 0.267441044 | 5.298227103 | 0.770017873 | -0.113499194 | FALSE | TRUE | FALSE |
| PTGER2 | 0.2059151 | 0.83272852 | 0.5053 | 1.905225721 | 1.905225721 | 4.07253597 | 0.22584448 | 0.845880156 | 1 | 19.85608107 | 1932.027372 | 1932.027372 | 0.911755957 | -0.040121391 | FALSE | TRUE | FALSE |
| POC5 | 0.2058779 | 0.83274487 | 1 | 1.890499319 | 1.890499319 | 4.080012016 | 0.25523125 | 0.837862325 | 1 | 0.255231249 | 0.255231249 | 5.063625246 | 0.806632915 | -0.09332406 | FALSE | TRUE | FALSE |
| MKRN2 | 0.2058027 | 0.83276122 | 0.01245 | 0.205802709 | 0.205802709 | 4.078690806 | 0.2477967 | 0.840021576 | 1 | 0.247796704 | 0.247796704 | 4.916308481 | 0.830530455 | -0.080644438 | FALSE | TRUE | FALSE |
| MORC2 | 0.2057821 | 0.83277757 | 1 | 0.205782128 | 0.205782128 | 4.021416427 | 0.24813102 | 0.839990003 | 1 | 0.248131025 | 0.248131025 | 4.862377897 | 0.829328488 | -0.081273416 | FALSE | TRUE | FALSE |
| AQP2 | 0.2057698 | 0.83279391 | 0.25535 | 0.205769805 | 0.822800151 | 0.822800151 | 0.06939493 | 0.882590251 | 1 | 2.565862241 | 24.35534621 | 24.35534621 | 2.965199502 | 0.472053919 | FALSE | TRUE | FALSE |
| RIOK3 | 0.2056494 | 0.83281025 | 1 | 1.888153685 | 1.888153685 | 4.07094841 | 0.25497512 | 0.837927138 | 1 | 0.254975122 | 0.254975122 | 5.053586232 | 0.806546773 | -0.093370442 | FALSE | TRUE | FALSE |
| RWDD2B | 0.2056132 | 0.83282658 | 0.751 | 1.889044415 | 7.555581072 | 7.555581072 | 0.249987 | 0.83946728 | 0.50345 | 2.305498167 | 2.305498167 | 2.305498167 | 0.822495499 | -0.08486647 | FALSE | TRUE | FALSE |
| PNLIPRP1 | 0.2055376 | 0.83284292 | 0.5057 | 0.360891937 | 7.661189088 | 30.61898856 | 0.18823699 | 0.854983951 | 0.37985 | 0.188236994 | 6.902723958 | 6.902723958 | 1.091908865 | 0.038186392 | FALSE | TRUE | FALSE |
| FES | 0.205505 | 0.83285925 | 0.9627 | 40.7105195 | 3948.448469 | 40296.82187 | 0.02368743 | 0.892793445 | 1 | 2.010685133 | 76.95449825 | 76.95449825 | 8.675697742 | 0.938304413 | FALSE | TRUE | FALSE |
| RASSF2 | 0.2053957 | 0.83287557 | 1 | 0.205395724 | 0.205395724 | 4.072736058 | 0.2467675 | 0.840163521 | 1 | 0.246767501 | 0.246767501 | 0.246767501 | 0.832345115 | -0.079696565 | FALSE | TRUE | FALSE |
| ZMYND12 | 0.2053187 | 0.8328919 | 1 | 1.885945042 | 17.77972371 | 17.77972371 | 0.26257295 | 0.835927328 | 1 | 0.262572952 | 0.262572952 | 0.262572952 | 0.781949214 | -0.106821453 | FALSE | TRUE | FALSE |
| AP2B1 | 0.2052925 | 0.83290822 | 1 | 38923108.41 | 4.02057E+11 | 4.88243E+12 | 0.17097149 | 0.859342564 | 1 | 4.39293E+14 | 1.30358E+19 | 2.50143E+21 | 1.200740978 | 0.079449332 | FALSE | TRUE | FALSE |
| ZNF835 | 0.2051479 | 0.83292454 | 0.9997 | 15905.5779 | 18090451.37 | 197287600.1 | 0.0454829 | 0.887714482 | 1 | 3.737969775 | 340.0217325 | 340.0217325 | 4.510440199 | 0.654218929 | FALSE | TRUE | FALSE |
| CRYM | 0.2050029 | 0.83294085 | 1 | 0.205002926 | 0.205002926 | 0.205002926 | 0.2395876 | 0.842125225 | 0.50225 | 2.250141924 | 2.250141924 | 2.250141924 | 0.855649141 | -0.067704281 | FALSE | TRUE | FALSE |
| CEP120 | 0.2047333 | 0.83295717 | 0.753 | 0.204733252 | 1.843739382 | 6.903646001 | 0.03807718 | 0.88942592 | 1 | 0.038077184 | 0.038077184 | 0.749523554 | 5.376796095 | 0.730523567 | FALSE | TRUE | FALSE |
| HJURP | 0.204687 | 0.83297348 | 0.51075 | 0.204687037 | 0.204687037 | 4.029472338 | 0.02941263 | 0.891397136 | 1 | 0.267036639 | 0.267036639 | 0.995143752 | 6.959153913 | 0.842556442 | FALSE | TRUE | FALSE |
| DNAJC15 | 0.2046089 | 0.83298979 | 1 | 0.20460893 | 0.20460893 | 0.20460893 | 0.24594843 | 0.840336694 | 1 | 0.245948435 | 0.245948435 | 0.245948435 | 0.831917999 | -0.079919479 | FALSE | TRUE | FALSE |
| PAQR5 | 0.2046079 | 0.83300609 | 1 | 0.204607859 | 0.204607859 | 0.204607859 | 0.24587752 | 0.840383859 | 1 | 0.245877519 | 0.245877519 | 0.245877519 | 0.832153587 | -0.07979651 | FALSE | TRUE | FALSE |
| TUBA8 | 0.2046063 | 0.83302239 | 0.85975 | 0.817988616 | 13.08365889 | 52.34592077 | 0.1040135 | 0.874907893 | 0.5098 | 0.415795707 | 1.662565582 | 6.649454998 | 1.967112814 | 0.293829267 | FALSE | TRUE | FALSE |
| PRSS8 | 0.2045964 | 0.83303869 | 1 | 0.204596364 | 0.204596364 | 0.204596364 | 0.24671738 | 0.840195033 | 0.00855 | 0.24671738 | 0.24671738 | 0.24671738 | 0.829274226 | -0.081301832 | FALSE | TRUE | FALSE |
| CLIC4 | 0.2045448 | 0.83305498 | 1 | 0.204544761 | 0.204544761 | 0.204544761 | 0.24669498 | 0.840210784 | 0.00525 | 0.24669498 | 0.24669498 | 0.24669498 | 0.829140347 | -0.081371951 | FALSE | TRUE | FALSE |
| HERC5 | 0.2044994 | 0.83307127 | 0.75065 | 0.20449943 | 1.871400821 | 6.906214774 | 0.03730179 | 0.889607055 | 0.51175 | 0.149160631 | 0.149160631 | 2.940262805 | 5.482295397 | 0.738962432 | FALSE | TRUE | FALSE |
| NOP16 | 0.2043204 | 0.83308756 | 1 | 0.204320386 | 0.204320386 | 0.204320386 | 0.23831342 | 0.842508808 | 0.50005 | 2.192003642 | 2.192003642 | 2.192003642 | 0.857359967 | -0.066836799 | FALSE | TRUE | FALSE |
| PIP5KL1 | 0.2042692 | 0.83310385 | 0.50365 | 0.817022368 | 0.817022368 | 0.817022368 | 171.711626 | 0.015599124 | 0.65805 | 686.8009308 | 27188.43787 | 275598.388 | 0.001189606 | -2.924596757 | FALSE | FALSE | FALSE |
| SYT12 | 0.2042427 | 0.83312013 | 1 | 0.816920274 | 0.816920274 | 4.038020853 | 0.24862024 | 0.839831963 | 1 | 0.248620243 | 0.248620243 | 4.922175468 | 0.821504809 | -0.08538989 | FALSE | TRUE | FALSE |
| TAF4B | 0.2042406 | 0.83313641 | 0.01965 | 0.204240581 | 0.204240581 | 4.022329425 | 0.2459587 | 0.840305235 | 0.01795 | 0.245958701 | 0.245958701 | 4.85284758 | 0.830385673 | -0.080720153 | FALSE | TRUE | FALSE |
| LCT | 0.2042316 | 0.83315268 | 0.9941 | 1590.324167 | 423368.4402 | 5428655.498 | 0.01197926 | 0.896077995 | 1 | 9.189037995 | 872.3793156 | 8808.64419 | 17.04876499 | 1.231692924 | FALSE | TRUE | FALSE |
| LINGO2 | 0.2041942 | 0.83316896 | 1 | 1.916484475 | 18.4609304 | 37.86343128 | 0.26243811 | 0.835977089 | 0.0141 | 0.262438107 | 0.262438107 | 5.192502796 | 0.778066156 | -0.108983475 | FALSE | TRUE | FALSE |
| SIGLEC14 | 0.2041923 | 0.83318522 | 1 | 1.867721148 | 1.867721148 | 4.049046331 | 0.25364961 | 0.838395497 | 1 | 0.253649614 | 0.253649614 | 5.03491919 | 0.805017064 | -0.094194914 | FALSE | TRUE | FALSE |
| SEC61A2 | 0.2040583 | 0.83320149 | 1 | 0.204058331 | 0.204058331 | 4.043844131 | 2.21733373 | 0.470278986 | 0.5036 | 2.217333729 | 2.217333729 | 4.730581058 | 0.092028696 | -1.036076733 | FALSE | FALSE | FALSE |
| EPHA8 | 0.2040178 | 0.83321775 | 0.60705 | 0.204017761 | 7.411146585 | 29.63426212 | 0.05209022 | 0.88621241 | 0.5146 | 0.052090217 | 0.465244404 | 2.268525777 | 3.916623391 | 0.592911813 | FALSE | TRUE | FALSE |
| DDX18 | 0.2040126 | 0.83323401 | 1 | 0.204012616 | 0.204012616 | 4.02982965 | 0.24590103 | 0.840368141 | 1 | 0.24590103 | 0.24590103 | 4.864467361 | 0.82965336 | -0.081103323 | FALSE | TRUE | FALSE |
| SLC16A1 | 0.2040029 | 0.83325027 | 1 | 0.20400288 | 0.20400288 | 4.042403114 | 0.24604503 | 0.8402895 | 1 | 0.246045026 | 0.246045026 | 4.880897925 | 0.829128243 | -0.081378291 | FALSE | TRUE | FALSE |
| SPOPL | 0.2038401 | 0.83326652 | 1 | 1.890660369 | 1.890660369 | 4.048203576 | 0.25345162 | 0.838459881 | 0.00945 | 0.253451619 | 0.253451619 | 0.253451619 | 0.804256345 | -0.094605504 | FALSE | TRUE | FALSE |
| RERG | 0.2038389 | 0.83328277 | 1 | 0.815129478 | 3.260428622 | 3.260428622 | 3.93296422 | 0.359841116 | 0.25275 | 0.983235828 | 3.932964217 | 3.932964217 | 0.051828312 | -1.285432933 | FALSE | FALSE | FALSE |
| EHD2 | 0.2037696 | 0.83329902 | 0.5035 | 0.814666273 | 3.257832128 | 6.903488296 | 0.06884854 | 0.882669308 | 1 | 2.59331306 | 100.278523 | 100.278523 | 2.959678519 | 0.47124454 | FALSE | TRUE | FALSE |
| CDK20 | 0.2037273 | 0.83331527 | 0.9683 | 16.62421314 | 375.4536445 | 1501.525833 | 0.068386 | 0.882783329 | 1 | 53.23539825 | 5103.755364 | 51786.68748 | 2.979079346 | 0.474082071 | FALSE | TRUE | FALSE |
| SPTY2D1 | 0.2037265 | 0.83333151 | 0.02025 | 0.203726492 | 0.203726492 | 4.021591544 | 0.22687542 | 0.845542016 | 1 | 0.226875418 | 0.226875418 | 4.485556677 | 0.897966353 | -0.046739936 | FALSE | TRUE | FALSE |
| SCTR | 0.2035838 | 0.83334775 | 0.75205 | 0.203583764 | 1.846308786 | 1.846308786 | 0.32670783 | 0.816501488 | 0.5079 | 0.326707828 | 0.326707828 | 0.727092306 | 0.623137085 | -0.205416401 | FALSE | TRUE | FALSE |
| HESX1 | 0.2035406 | 0.83336398 | 0.7534 | 0.203540561 | 0.813994547 | 0.813994547 | 0.65325666 | 0.688342923 | 0.50595 | 0.653256663 | 0.653256663 | 0.653256663 | 0.31157824 | -0.50643288 | FALSE | TRUE | FALSE |
| VN1R4 | 0.2035084 | 0.83338021 | 0.68655 | 0.203508374 | 19.8862305 | 19.8862305 | 0.00275593 | 0.898747743 | 1 | 0.025678143 | 0.24555414 | 0.24555414 | 73.84375242 | 1.868313758 | FALSE | TRUE | FALSE |
| TNFSF9 | 0.2034424 | 0.83339644 | 0.5116 | 1.903983322 | 1.903983322 | 1.903983322 | 2.30135107 | 0.464760786 | 0.00825 | 2.301351073 | 2.301351073 | 2.301351073 | 0.088401292 | -1.053541387 | FALSE | FALSE | FALSE |
| FTMT | 0.2033778 | 0.83341267 | 1 | 0.203377761 | 0.203377761 | 4.037551111 | 0.23766028 | 0.842646451 | 1 | 2.20634882 | 2.20634882 | 2.20634882 | 0.855749891 | -0.067653147 | FALSE | TRUE | FALSE |
| HDAC7 | 0.203349 | 0.83342889 | 1 | 0.203349008 | 0.203349008 | 3.990676987 | 0.23780816 | 0.842631168 | 1 | 2.220360122 | 2.220360122 | 43.67249343 | 0.855096833 | -0.067984702 | FALSE | TRUE | FALSE |
| TMCC2 | 0.2033008 | 0.83344511 | 1 | 174.4400723 | 17842.97877 | 187002.0197 | 0.25279457 | 0.838716897 | 1 | 219.4020699 | 2200.71041 | 22613.03463 | 0.804213278 | -0.094628761 | FALSE | TRUE | FALSE |
| FGB | 0.2032557 | 0.83346133 | 0.50565 | 0.812637706 | 3.249822545 | 3.257612534 | 10.3836551 | 0.194542721 | 0.38525 | 4.43181745 | 41.52870491 | 166.1326637 | 0.019574585 | -1.708307428 | FALSE | FALSE | FALSE |
| ZNF599 | 0.2031933 | 0.83347754 | 0.8748 | 1.819545851 | 16.71267742 | 16.71267742 | 0.03791914 | 0.889457461 | 1 | 0.334560394 | 0.334560394 | 0.751283942 | 5.358592798 | 0.729050756 | FALSE | TRUE | FALSE |
| PSD | 0.203167 | 0.83349375 | 0.52055 | 1.892287616 | 1.892287616 | 36.893234 | 0.24512338 | 0.840619258 | 0.5196 | 2.288246021 | 2.288246021 | 44.74840149 | 0.828835545 | -0.081531632 | FALSE | TRUE | FALSE |
| SLC25A15 | 0.203067 | 0.83350996 | 0.753 | 0.203066973 | 0.811985161 | 0.811985161 | 17.4979727 | 0.146035878 | 0.18975 | 4.373554129 | 40.48152658 | 161.9608736 | 0.011605171 | -1.93534844 | FALSE | FALSE | FALSE |
| AP3D1 | 0.2029892 | 0.83352616 | 1 | 0.202989182 | 0.202989182 | 3.941281584 | 0.23740054 | 0.842753342 | 0.51675 | 2.215576595 | 2.215576595 | 43.16950375 | 0.855049352 | -0.068008818 | FALSE | TRUE | FALSE |
| CRISP1 | 0.2029045 | 0.83354236 | 1 | 1.849342003 | 17.29981584 | 17.29981584 | 2.23361829 | 0.468864018 | 0.74885 | 2.233618287 | 20.99299351 | 20.99299351 | 0.090841153 | -1.04171736 | FALSE | FALSE | FALSE |
| PTGIR | 0.2029006 | 0.83355856 | 0.50755 | 1.858405643 | 1.858405643 | 4.029207694 | 20.5230485 | 0.130141764 | 0.25485 | 2.17607316 | 20.52304846 | 20.52304846 | 0.009886474 | -2.004958573 | FALSE | FALSE | FALSE |
| CHORDC1 | 0.2028362 | 0.83357476 | 1 | 0.202836178 | 0.202836178 | 0.202836178 | 0.24437061 | 0.840713236 | 1 | 0.244370615 | 0.244370615 | 0.244370615 | 0.830035062 | -0.080903562 | FALSE | TRUE | FALSE |
| SAT2 | 0.2028283 | 0.83359095 | 1 | 1.854510586 | 1.854510586 | 1.854510586 | 2.24088669 | 0.468331444 | 0.00425 | 2.240886688 | 2.240886688 | 2.240886688 | 0.090512505 | -1.043291415 | FALSE | FALSE | FALSE |
| ANKRD10 | 0.2028234 | 0.83360714 | 1 | 174.1129456 | 17815.12928 | 183442.181 | 0.29472869 | 0.826786382 | 1 | 0.294728689 | 0.294728689 | 5.847395007 | 0.688169985 | -0.162304273 | FALSE | TRUE | FALSE |
| FCGR3A | 0.2028221 | 0.83362332 | 0.8756 | 17.32379868 | 166.3675985 | 166.3675985 | 0.25149284 | 0.838909134 | 1 | 2.306896551 | 21.71825418 | 21.71825418 | 0.806472795 | -0.093410278 | FALSE | TRUE | FALSE |
| ING1 | 0.2027993 | 0.83363951 | 0.01055 | 0.202799282 | 0.202799282 | 4.017833886 | 0.23666642 | 0.842951475 | 1 | 2.171552644 | 2.171552644 | 4.694110195 | 0.856899277 | -0.067070224 | FALSE | TRUE | FALSE |

| | | | | | | | | | | | | | | | | | |
|---|---|---|---|---|---|---|---|---|---|---|---|---|---|---|---|---|---|
| CALR3 | 0.2027311 | 0.83365568 | 0.0117 | 0.202731067 | 0.202731067 | 1.005615059 | 0.0721295 | 0.881989823 | 0.0087 | 0.072129502 | 0.072129502 | 0.072129502 | 2.810653926 | 0.448807375 | FALSE | TRUE | FALSE |
| CHMP2B | 0.2027065 | 0.83367186 | 1 | 0.810673081 | 3.242891005 | 3.242891005 | 8.79404857 | 0.214161055 | 0.1292 | 3.790792176 | 35.1791322 | 35.1791322 | 0.023050421 | -1.637321147 | FALSE | FALSE | FALSE |
| PSMB10 | 0.2026636 | 0.83368803 | 0.7557 | 1.863958619 | 17.59509846 | 17.59509846 | 0.25965919 | 0.836851264 | 1 | 0.259659194 | 0.259659194 | 5.154195937 | 0.780498524 | -0.107627914 | FALSE | TRUE | FALSE |
| PTPN18 | 0.2025857 | 0.8337042 | 0.75105 | 0.202585654 | 0.810245912 | 4.009178088 | 1.07669535 | 0.580657482 | 0.73375 | 9.819578259 | 157.2037426 | 629.2317617 | 0.188155037 | -0.72548415 | FALSE | FALSE | FALSE |
| VAT1L | 0.202558 | 0.83372037 | 1 | 0.202558006 | 0.202558006 | 4.022721996 | 0.24379199 | 0.84091649 | 0.00775 | 0.243791994 | 0.243791994 | 0.243791994 | 0.83086406 | -0.080470027 | FALSE | TRUE | FALSE |
| NCAM2 | 0.2025176 | 0.83373653 | 0.0209 | 0.202517563 | 0.202517563 | 3.988909872 | 0.24449151 | 0.840697579 | 0.0189 | 0.244491512 | 0.244491512 | 4.824447119 | 0.828321446 | -0.081801094 | FALSE | TRUE | FALSE |
| MMP11 | 0.2024768 | 0.83375269 | 0.0149 | 0.202476783 | 0.202476783 | 4.000318744 | 0.24389533 | 0.840869633 | 1 | 0.243895332 | 0.243895332 | 4.825668405 | 0.830178998 | -0.080828258 | FALSE | TRUE | FALSE |
| HPN | 0.2024214 | 0.83376885 | 1 | 0.202421398 | 0.202421398 | 4.001202908 | 0.24465838 | 0.840681921 | 1 | 0.244658385 | 0.244658385 | 4.842875968 | 0.82736342 | -0.082303684 | FALSE | TRUE | FALSE |
| ZNF649 | 0.2023721 | 0.833785 | 1 | 0.202372064 | 0.202372064 | 4.011256786 | 0.24394557 | 0.840807112 | 1 | 0.243945569 | 0.243945569 | 4.840492645 | 0.829578768 | -0.081142372 | FALSE | TRUE | FALSE |
| SIM1 | 0.2023606 | 0.83380115 | 1 | 0.202360612 | 0.202360612 | 3.973866866 | 0.2443512 | 0.840728889 | 1 | 0.244351205 | 0.244351205 | 4.808929819 | 0.828154755 | -0.0818885 | FALSE | TRUE | FALSE |
| CA14 | 0.2023424 | 0.8338173 | 0.7527 | 0.202342361 | 1.813977966 | 1.813977966 | 0.1493045 | 0.86474711 | 0.5004 | 0.149304502 | 0.149304502 | 0.149304502 | 1.355232812 | 0.132013908 | FALSE | TRUE | FALSE |
| IDO1 | 0.2022693 | 0.83383345 | 0.0107 | 0.202269315 | 0.202269315 | 1.003880445 | 0.0701146 | 0.882449478 | 0.5083 | 0.63239551 | 0.63239551 | 1.392864612 | 2.884838826 | 0.460121554 | FALSE | TRUE | FALSE |
| OSMR | 0.2022041 | 0.83384959 | 0.14535 | 0.202204126 | 0.809123478 | 3.976771134 | 0.70790142 | 0.662436748 | 0.3856 | 0.326193355 | 2.832023288 | 6.428135717 | 0.285638821 | -0.544182768 | FALSE | TRUE | FALSE |
| ANKRD45 | 0.2021972 | 0.83386573 | 1 | 0.202197247 | 0.202197247 | 0.202197247 | 0.24344731 | 0.840963321 | 1 | 0.243447307 | 0.243447307 | 0.243447307 | 0.830558569 | -0.080629737 | FALSE | TRUE | FALSE |
| CCDC82 | 0.2020494 | 0.83388186 | 1 | 1.851501449 | 1.851501449 | 4.001873958 | 0.24329335 | 0.841025721 | 1 | 2.238614518 | 2.238614518 | 4.824378892 | 0.830476491 | -0.080672657 | FALSE | TRUE | FALSE |
| ITLN1 | 0.2020439 | 0.83389799 | 0.50035 | 0.808076904 | 0.808076904 | 0.808076904 | 0.24632116 | 0.840242279 | 1 | 0.24632116 | 0.24632116 | 0.24632116 | 0.820245674 | -0.086056052 | FALSE | TRUE | FALSE |
| CNOT1 | 0.2020253 | 0.83391412 | 1 | 0.202025316 | 3.864880735 | 7.8743E+302 | 0.24411476 | 0.840744539 | 0.59965 | 1.2822E+279 | 3.3058E+305 | 1.5283E+307 | 0.827583358 | -0.082188251 | FALSE | TRUE | FALSE |
| CTSS | 0.2019331 | 0.83393025 | 1 | 0.201933097 | 0.201933097 | 0.201933097 | 0.24405382 | 0.840760187 | 1 | 0.244053815 | 0.244053815 | 0.244053815 | 0.827412173 | -0.082278094 | FALSE | TRUE | FALSE |
| LRRC8C | 0.2018674 | 0.83394637 | 1 | 1.859129297 | 17.57306207 | 36.64489704 | 0.25884683 | 0.836998789 | 1 | 0.258846834 | 0.258846834 | 5.110953872 | 0.779871873 | -0.107976743 | FALSE | TRUE | FALSE |
| REST | 0.2018671 | 0.83396249 | 1 | 0.201867134 | 0.201867134 | 3.959278196 | 0.24097347 | 0.841739806 | 1 | 0.240973471 | 0.240973471 | 4.737287071 | 0.837715179 | -0.076903615 | FALSE | TRUE | FALSE |
| OR1L8 | 0.2018368 | 0.83397861 | 0.99935 | 17142.64102 | 22251707.75 | 248059958.4 | 0.26541465 | 0.835160561 | 1 | 228.1937815 | 23372.1376 | 245110.4816 | 0.76045828 | -0.118924607 | FALSE | TRUE | FALSE |
| FBXO38 | 0.2018287 | 0.83399472 | 0.7631 | 1.872698865 | 17.8338107 | 63.01743716 | 0.22894596 | 0.84514325 | 0.9396 | 20.26604253 | 1985.342862 | 1985.342862 | 0.881556266 | -0.054749963 | FALSE | TRUE | FALSE |
| TMEM165 | 0.2017919 | 0.83401083 | 1 | 0.201791869 | 0.201791869 | 0.201791869 | 0.24366738 | 0.840947713 | 1 | 0.243667383 | 0.243667383 | 0.243667383 | 0.828144771 | -0.081893736 | FALSE | TRUE | FALSE |
| GRIA2 | 0.2017592 | 0.83402694 | 1 | 0.201759199 | 0.201759199 | 3.96850049 | 0.24395887 | 0.840791474 | 1 | 0.243958871 | 0.243958871 | 4.808092728 | 0.827021368 | -0.082483269 | FALSE | TRUE | FALSE |
| REEP3 | 0.2016597 | 0.83404304 | 1 | 0.201659744 | 0.201659744 | 0.201659744 | 0.24093512 | 0.841755259 | 1 | 0.963463198 | 0.963463198 | 0.963463198 | 0.836987739 | -0.077280904 | FALSE | TRUE | FALSE |
| LGALS3 | 0.2015566 | 0.83405915 | 1 | 0.201556628 | 0.201556628 | 0.201556628 | 0.24272675 | 0.841228178 | 1 | 0.242726751 | 0.242726751 | 0.242726751 | 0.830384897 | -0.080720558 | FALSE | TRUE | FALSE |
| C15orf40 | 0.2015399 | 0.83407524 | 1 | 1.837283779 | 1.837283779 | 1.837283779 | 0.25002507 | 0.839451387 | 1 | 0.25002507 | 0.25002507 | 0.25002507 | 0.80607867 | -0.093622571 | FALSE | TRUE | FALSE |
| GK2 | 0.2015199 | 0.83409134 | 0.77305 | 0.805933602 | 12.89992396 | 51.62900395 | 0.40241441 | 0.791439622 | 0.19645 | 0.40241441 | 1.609554322 | 6.439412309 | 0.500777149 | -0.300355497 | FALSE | TRUE | FALSE |
| NSUN3 | 0.2014754 | 0.83410743 | 1 | 0.201475397 | 0.201475397 | 0.201475397 | 0.24272836 | 0.841212622 | 0.00785 | 0.242728356 | 0.242728356 | 0.242728356 | 0.830044746 | -0.080898495 | FALSE | TRUE | FALSE |
| GOLGA3 | 0.2013612 | 0.83412352 | 0.5286 | 0.20136122 | 0.20136122 | 3.885687689 | 0.30582793 | 0.823586475 | 0.7614 | 0.305827925 | 2.839765096 | 55.03160204 | 0.658413452 | -0.181501305 | FALSE | TRUE | FALSE |
| PI4KB | 0.2013524 | 0.83413961 | 0.0251 | 0.201352355 | 0.201352355 | 3.956751934 | 0.24335431 | 0.841010126 | 1 | 0.24335431 | 0.24335431 | 4.792177733 | 0.827404106 | -0.082282328 | FALSE | TRUE | FALSE |
| ADAMTS16 | 0.2012946 | 0.83415569 | 0.52555 | 0.201294552 | 1.694887899 | 6.777428411 | 0.09774406 | 0.876545939 | 0.7024 | 0.217811605 | 3.482652008 | 7.628342796 | 2.059404461 | 0.313741649 | FALSE | TRUE | FALSE |
| PELO | 0.201274 | 0.83417177 | 0.0126 | 0.20127402 | 0.20127402 | 3.987464615 | 0.2429595 | 0.841165938 | 1 | 0.242959503 | 0.242959503 | 4.818768365 | 0.828426209 | -0.08174617 | FALSE | TRUE | FALSE |
| EXT1 | 0.2012602 | 0.83418784 | 0.2646 | 0.050363323 | 0.201260163 | 0.990441848 | 0.02164843 | 0.893335495 | 0.0189 | 0.021648431 | 0.021648431 | 0.42659273 | 9.296755215 | 0.968331396 | FALSE | TRUE | FALSE |
| LGI2 | 0.2012503 | 0.83420392 | 0.5102 | 0.201250326 | 0.201250326 | 0.995681301 | 0.07154937 | 0.882105041 | 0.503 | 0.286064744 | 0.286064744 | 1.417087955 | 2.812747575 | 0.449130759 | FALSE | TRUE | FALSE |
| SUPT16H | 0.2012449 | 0.83421999 | 1 | 0.201244895 | 0.201244895 | 3.945223179 | 0.24322458 | 0.841072488 | 1 | 0.243224579 | 0.243224579 | 4.779564014 | 0.827403614 | -0.082282586 | FALSE | TRUE | FALSE |
| YJEFN3 | 0.2010787 | 0.83423605 | 0.50745 | 0.804176611 | 0.804176611 | 3.985237051 | 0.23800539 | 0.842585305 | 0.50535 | 2.210244234 | 2.210244234 | 4.722224599 | 0.844849257 | -0.073220773 | FALSE | TRUE | FALSE |
| CA3 | 0.2010506 | 0.83425212 | 1 | 0.201050558 | 0.201050558 | 0.201050558 | 0.24018139 | 0.842048292 | 0.501 | 0.960499677 | 0.960499677 | 0.960499677 | 0.83707799 | -0.077234077 | FALSE | TRUE | FALSE |
| RAPGEF1 | 0.2009694 | 0.83426818 | 1 | 0.200969361 | 0.200969361 | 3.91780539 | 0.2427088 | 0.84124373 | 1 | 0.242708801 | 0.242708801 | 4.745907109 | 0.828026674 | -0.081955673 | FALSE | TRUE | FALSE |
| SV2A | 0.2009639 | 0.83428424 | 0.0251 | 0.200963948 | 0.200963948 | 3.948211794 | 0.2400776 | 0.842094459 | 1 | 0.960212201 | 0.960212201 | 18.90484324 | 0.837079138 | -0.077233482 | FALSE | TRUE | FALSE |
| WNT11 | 0.2009509 | 0.83430029 | 1 | 0.200950859 | 0.200950859 | 3.987883135 | 0.24295555 | 0.841181503 | 0.00815 | 0.242955555 | 0.242955555 | 0.242955555 | 0.827109547 | -0.082436966 | FALSE | TRUE | FALSE |
| CTH | 0.2009295 | 0.83431635 | 0.51145 | 0.200929484 | 0.200929484 | 0.439984558 | 0.03528073 | 0.890053668 | 1 | 0.035280728 | 0.141042119 | 0.700926199 | 5.695162647 | 0.755506131 | FALSE | TRUE | FALSE |
| TEKT1 | 0.2008591 | 0.83433239 | 1 | 0.200859072 | 0.200859072 | 3.981074842 | 0.24195192 | 0.841430126 | 0.0097 | 0.241951916 | 0.241951916 | 0.241951916 | 0.830161113 | -0.080837614 | FALSE | TRUE | FALSE |
| FEN1 | 0.2008177 | 0.83434844 | 0.7489 | 1.88305181 | 18.12213276 | 18.12213276 | 20.1120038 | 0.132303786 | 0.65975 | 80.42643183 | 8026.859658 | 32106.84935 | 0.009984968 | -2.000653339 | FALSE | FALSE | FALSE |
| DSCC1 | 0.2006869 | 0.83436448 | 1 | 0.200686949 | 0.200686949 | 0.200686949 | 0.23935614 | 0.842171355 | 1 | 0.957340587 | 0.957340587 | 0.957340587 | 0.838444942 | -0.076525451 | FALSE | TRUE | FALSE |
| SLCO2A1 | 0.2006828 | 0.83438052 | 0.75325 | 0.456140327 | 1.82393289 | 6.789712389 | 0.07422023 | 0.881553695 | 1 | 0.074220233 | 0.074220233 | 1.466442769 | 2.703881966 | 0.431987729 | FALSE | TRUE | FALSE |
| ANXA5 | 0.2006364 | 0.83439656 | 0.87675 | 1.837542589 | 17.26181032 | 17.26181032 | 0.03124812 | 0.890929518 | 1 | 0.281300059 | 0.281300059 | 0.281300059 | 6.420749291 | 0.807585713 | FALSE | TRUE | FALSE |
| N6AMT1 | 0.2006334 | 0.83441259 | 1 | 0.200633402 | 0.200633402 | 0.200633402 | 0.22569327 | 0.845924154 | 0.0066 | 0.22569327 | 0.22569327 | 0.22569327 | 0.888964931 | -0.051115371 | FALSE | TRUE | FALSE |
| IMP3 | 0.2005552 | 0.83442862 | 0.00915 | 0.200555181 | 0.200555181 | 0.200555181 | 0.24185147 | 0.841492161 | 1 | 0.241851471 | 0.241851471 | 0.241851471 | 0.829249374 | -0.081314848 | FALSE | TRUE | FALSE |
| DPF1 | 0.2004442 | 0.83444465 | 0.0128 | 0.200444151 | 0.200444151 | 3.970241179 | 0.24251153 | 0.841290369 | 1 | 0.242511532 | 0.242511532 | 4.80904826 | 0.826534514 | -0.082739007 | FALSE | TRUE | FALSE |
| THEMIS2 | 0.2004384 | 0.83446067 | 1 | 0.200438356 | 0.200438356 | 3.953657443 | 0.24182324 | 0.841523159 | 1 | 0.241823243 | 0.241823243 | 4.777868004 | 0.828863072 | -0.081517209 | FALSE | TRUE | FALSE |
| UPF2 | 0.2004288 | 0.8344767 | 1 | 1.874884825 | 18.00017726 | 63.08055011 | 2.26832908 | 0.46744219 | 0.51385 | 2.268329077 | 2.268329077 | 44.44962301 | 0.088359686 | -1.053745838 | FALSE | FALSE | FALSE |
| PKN3 | 0.2004041 | 0.83449271 | 0.76395 | 0.801496915 | 7.233213099 | 15.66765081 | 0.03558338 | 0.889991181 | 1 | 0.315595388 | 2.872896518 | 10.66294243 | 5.631957954 | 0.750659404 | FALSE | TRUE | FALSE |
| BTBD10 | 0.2003909 | 0.83450873 | 1 | 0.200390931 | 0.200390931 | 3.969413294 | 20.375938 | 0.130451146 | 0.2553 | 2.125591464 | 20.375938 | 42.15298596 | 0.009834685 | -2.007239547 | FALSE | FALSE | FALSE |
| ACOX3 | 0.2003515 | 0.83452474 | 0.20405 | 0.050111743 | 0.200351462 | 0.801224101 | 0.53899919 | 0.739011648 | 0.4111 | 0.239186952 | 4.979271964 | 19.90074801 | 0.371710133 | -0.429795599 | FALSE | TRUE | FALSE |
| RNF44 | 0.2002301 | 0.83454075 | 1 | 17.21856677 | 165.9168569 | 165.9168569 | 2.36314729 | 0.459136893 | 0.50515 | 2.363147294 | 2.363147294 | 5.09186041 | 0.084730258 | -1.071961469 | FALSE | FALSE | FALSE |
| GBX1 | 0.2000962 | 0.83455675 | 0.009 | 0.200096181 | 0.200096181 | 0.200096181 | 0.24213874 | 0.841352513 | 1 | 0.242138736 | 0.242138736 | 0.242138736 | 0.826369973 | -0.082825472 | FALSE | TRUE | FALSE |
| PRDM14 | 0.2000053 | 0.83457276 | 1 | 0.200005276 | 0.200005276 | 3.953005734 | 0.24167521 | 0.8416006 | 0.01495 | 0.241675211 | 0.241675211 | 4.783358279 | 0.82757878 | -0.082190654 | FALSE | TRUE | FALSE |
| MSRB2 | 0.1999005 | 0.83458876 | 1 | 0.19990054 | 0.19990054 | 0.19990054 | 0.2340156 | 0.843588285 | 0.5002 | 2.17041107 | 2.17041107 | 2.17041107 | 0.854218855 | -0.068430847 | FALSE | TRUE | FALSE |
| OR4C6 | 0.1998933 | 0.83460476 | 1 | 0.1998933 | 0.1998933 | 0.1998933 | 7.93395982 | 0.229222941 | 0.00275 | 2.148593667 | 2.148593667 | 2.148593667 | 0.025194645 | -1.59869176 | FALSE | FALSE | FALSE |
| AKAP1 | 0.1998309 | 0.83462075 | 0.51245 | 1.843773461 | 1.843773461 | 36.17502107 | 0.24884749 | 0.839736986 | 0.0224 | 0.248847491 | 0.248847491 | 4.893580695 | 0.803025458 | -0.095270686 | FALSE | TRUE | FALSE |
| SGMS2 | 0.1998082 | 0.83463674 | 1 | 1.851872647 | 1.851872647 | 3.967190753 | 0.24138224 | 0.841677966 | 0.5089 | 2.243516922 | 2.243516922 | 4.796821482 | 0.827766789 | -0.082092002 | FALSE | TRUE | FALSE |
| SMC2 | 0.1997381 | 0.83465273 | 0.3384 | 0.199738102 | 3.193983286 | 15.66755992 | 0.02169005 | 0.893328103 | 1 | 0.086691503 | 0.086691503 | 1.704639836 | 9.208742955 | 0.964200351 | FALSE | TRUE | FALSE |
| TMPPE | 0.1996129 | 0.83466872 | 1 | 0.199612924 | 0.199612924 | 3.951257478 | 0.24143113 | 0.841647028 | 1 | 0.24143113 | 0.24143113 | 4.784936178 | 0.826790329 | -0.082604612 | FALSE | TRUE | FALSE |
| SFR1 | 0.1995916 | 0.8346847 | 1 | 0.199591585 | 0.199591585 | 0.199591585 | 0.24043967 | 0.841909624 | 1 | 0.240439669 | 0.240439669 | 0.240439669 | 0.830110879 | -0.080863895 | FALSE | TRUE | FALSE |
| FOXA3 | 0.1995699 | 0.83470068 | 1 | 0.199569927 | 0.199569927 | 3.957712627 | 0.94414693 | 0.593616379 | 0.7529 | 0.944146926 | 3.776393131 | 3.776393131 | 0.211375922 | -0.674944486 | FALSE | TRUE | FALSE |
| PPP6R1 | 0.1995688 | 0.83471666 | 1 | 0.199568783 | 0.199568783 | 3.911712228 | 0.24141918 | 0.841662499 | 1 | 0.241419181 | 0.241419181 | 4.743388849 | 0.826648415 | -0.082679163 | FALSE | TRUE | FALSE |
| TIGD7 | 0.1995658 | 0.83473263 | 0.50285 | 0.199565772 | 0.199565772 | 0.444937626 | 0.02951041 | 0.891351301 | 0.74985 | 0.264007696 | 2.42415769 | 2.42415769 | 6.762554684 | 0.83011079 | FALSE | TRUE | FALSE |
| STK32C | 0.1994946 | 0.8347486 | 1 | 0.199494643 | 0.199494643 | 3.941357217 | 0.23357893 | 0.843784368 | 1 | 2.160686809 | 2.160686809 | 4.621504917 | 0.854078071 | -0.068502429 | FALSE | TRUE | FALSE |
| TVP23A | 0.1994913 | 0.83476456 | 1 | 0.199491319 | 0.199491319 | 0.199491319 | 0.24042067 | 0.841925044 | 1 | 0.240420673 | 0.240420673 | 0.240420673 | 0.829759422 | -0.081047808 | FALSE | TRUE | FALSE |
| CEP57 | 0.1994139 | 0.83478053 | 1 | 0.19941392 | 0.19941392 | 3.955237486 | 0.23826088 | 0.842524113 | 1 | 0.238260875 | 0.95292305 | 4.730448422 | 0.836956215 | -0.077297261 | FALSE | TRUE | FALSE |
| EPAS1 | 0.199403 | 0.83479649 | 0.5167 | 1.860282275 | 1.860282275 | 36.5212176 | 0.24883607 | 0.839752824 | 1 | 0.248836072 | 0.248836072 | 4.89599153 | 0.801342881 | -0.096181617 | FALSE | TRUE | FALSE |
| PPT1 | 0.1993376 | 0.83481245 | 0.74635 | 0.199337628 | 1.890747882 | 1.890747882 | 26.3947542 | 0.103757625 | 0.1929 | 26.39475422 | 263.3289563 | 2691.258326 | 0.007552168 | -2.121928335 | FALSE | FALSE | FALSE |

| | | | | | | | | | | | | | | | | | |
|---|---|---|---|---|---|---|---|---|---|---|---|---|---|---|---|---|---|
| CRYZL1 | 0.1992364 | 0.8348284 | 1 | 0.199236374 | 0.199236374 | 0.199236374 | 0.24024941 | 0.842002097 | 0.0076 | 0.24024941 | 0.24024941 | 0.24024941 | 0.829289754 | -0.0812937 | FALSE | TRUE | FALSE |
| FOLH1 | 0.1991777 | 0.83484435 | 1 | 0.199177662 | 0.199177662 | 3.936991113 | 2.17403048 | 0.473611183 | 0.5032 | 2.174030485 | 2.174030485 | 7.898377191 | 0.091616775 | -1.038024999 | FALSE | FALSE | FALSE |
| TRIM22 | 0.1991135 | 0.8348603 | 0.7582 | 0.45142042 | 1.805035406 | 3.946405336 | 23.0630425 | 0.117307301 | 0.1945 | 2.467815694 | 92.24575657 | 92.24575657 | 0.008633445 | -2.063815872 | FALSE | FALSE | FALSE |
| VPS18 | 0.1990698 | 0.83487625 | 1 | 0.199069824 | 0.199069824 | 6.712005424 | 0.24077123 | 0.841786156 | 1 | 0.240771225 | 0.240771225 | 4.70802614 | 0.826800728 | -0.08259915 | FALSE | TRUE | FALSE |
| MAPK4 | 0.1990623 | 0.83489219 | 1 | 0.199062271 | 0.199062271 | 3.924490892 | 0.23347541 | 0.8438446 | 1 | 2.190080661 | 2.190080661 | 7.910107417 | 0.852604864 | -0.069252194 | FALSE | TRUE | FALSE |
| TTBK2 | 0.1990244 | 0.83490813 | 1 | 0.199024417 | 0.199024417 | 3.891218462 | 0.24073359 | 0.841832479 | 1 | 0.240733588 | 0.240733588 | 4.719410043 | 0.826741374 | -0.082630328 | FALSE | TRUE | FALSE |
| CABLES2 | 0.1988264 | 0.83492407 | 0.0138 | 0.198826399 | 0.198826399 | 3.937796494 | 0.24020786 | 0.842032897 | 1 | 0.240207859 | 0.240207859 | 4.762938196 | 0.827726454 | -0.082113164 | FALSE | TRUE | FALSE |
| IL6ST | 0.1987599 | 0.83494 | 1 | 0.198759871 | 0.198759871 | 3.914324227 | 0.24059549 | 0.841863346 | 1 | 0.240595486 | 0.240595486 | 4.746957211 | 0.826116376 | -0.082958769 | FALSE | TRUE | FALSE |
| GADD45GIP1 | 0.1985919 | 0.83495593 | 0.96755 | 163.9461429 | 1617.315985 | 16344.44842 | 2.49508541 | 0.446961911 | 0.50325 | 2.495085413 | 2.495085413 | 2.495085413 | 0.07959323 | -1.09912387 | FALSE | FALSE | FALSE |
| PPP6R3 | 0.1985498 | 0.83497186 | 1 | 0.198549762 | 0.198549762 | 3.912070103 | 2.1682511 | 0.474481865 | 0.5123 | 2.168251097 | 2.168251097 | 7.866396258 | 0.091571388 | -1.038240204 | FALSE | FALSE | FALSE |
| TMEM138 | 0.1983917 | 0.83498779 | 1 | 0.198391665 | 0.198391665 | 0.198391665 | 0.23478258 | 0.843482498 | 1 | 0.93885754 | 3.755277941 | 3.755277941 | 0.845001641 | -0.073142448 | FALSE | TRUE | FALSE |
| COA6 | 0.1983911 | 0.83500371 | 1 | 0.198391076 | 0.198391076 | 0.198391076 | 0.23933608 | 0.842186725 | 0.00155 | 0.239336077 | 0.239336077 | 0.239336077 | 0.828922568 | -0.081486036 | FALSE | TRUE | FALSE |
| ZNF354A | 0.1983305 | 0.83501963 | 1 | 17.26149484 | 167.3319254 | 167.3319254 | 2.35934349 | 0.459320121 | 0.5082 | 2.359343486 | 2.359343486 | 5.048475285 | 0.084061743 | -1.07540161 | FALSE | FALSE | FALSE |
| RBMXL2 | 0.1983229 | 0.83503555 | 0.01375 | 0.198322921 | 0.198322921 | 3.923839601 | 0.240274 | 0.841986693 | 1 | 0.240273997 | 0.240273997 | 4.759990815 | 0.825403179 | -0.083333863 | FALSE | TRUE | FALSE |
| TRIM56 | 0.1983131 | 0.83505146 | 0.5121 | 1.832598646 | 1.832598646 | 35.94804959 | 0.24724819 | 0.840116231 | 1 | 0.24724819 | 0.24724819 | 4.861209763 | 0.802081213 | -0.095781656 | FALSE | TRUE | FALSE |
| SMCHD1 | 0.1982829 | 0.83506738 | 0.942 | 0.793106839 | 12.69843482 | 61.373155 | 0.64134272 | 0.693632697 | 0.7568 | 0.641342716 | 6.080933751 | 118.0818678 | 0.309168376 | -0.509804935 | FALSE | TRUE | FALSE |
| DPH5 | 0.1981587 | 0.83508328 | 1 | 0.19815869 | 1.831577191 | 1.831577191 | 0.24703422 | 0.840131997 | 1 | 0.247034221 | 0.247034221 | 0.247034221 | 0.802150767 | -0.095743997 | FALSE | TRUE | FALSE |
| SAP30BP | 0.1981093 | 0.83509919 | 1 | 0.198109286 | 0.198109286 | 0.198109286 | 0.23914718 | 0.84229423 | 1 | 0.239147177 | 0.239147177 | 0.239147177 | 0.828399016 | -0.081760426 | FALSE | TRUE | FALSE |
| SMAP1 | 0.1980134 | 0.83511509 | 1 | 0.198013399 | 0.198013399 | 3.927371413 | 0.23174505 | 0.844489113 | 1 | 2.126820033 | 2.126820033 | 4.600992306 | 0.854444993 | -0.068315891 | FALSE | TRUE | FALSE |
| SNRPE | 0.1979503 | 0.83513099 | 0.003 | 0.197950298 | 0.197950298 | 0.197950298 | 0.20334935 | 0.851280783 | 0.00205 | 0.20334935 | 0.20334935 | 0.20334935 | 0.973449379 | -0.011686628 | FALSE | TRUE | FALSE |
| C2CD2L | 0.1979461 | 0.83514689 | 0.0217 | 0.197946139 | 0.197946139 | 3.887463652 | 0.20502059 | 0.850799924 | 0.9673 | 180.5577097 | 1819.909338 | 18791.23914 | 0.965493959 | -0.015250439 | FALSE | TRUE | FALSE |
| LGALS9 | 0.1978258 | 0.83516278 | 1 | 0.197825816 | 0.197825816 | 3.923094443 | 0.23872505 | 0.842432237 | 1 | 0.238725053 | 0.238725053 | 0.283727592 | 0.828676396 | -0.081615032 | FALSE | TRUE | FALSE |
| RPA1 | 0.1977652 | 0.83517867 | 0.01945 | 0.197765165 | 0.197765165 | 3.904250456 | 0.23919936 | 0.842278882 | 1 | 0.239199363 | 0.239199363 | 4.729572645 | 0.826779649 | -0.082610222 | FALSE | TRUE | FALSE |
| SOHLH1 | 0.1977623 | 0.83519456 | 1 | 0.197762264 | 0.197762264 | 3.917289546 | 0.2392968 | 0.842232817 | 1 | 0.239296798 | 0.239296798 | 4.745479942 | 0.826430882 | -0.082793463 | FALSE | TRUE | FALSE |
| MRRF | 0.1976994 | 0.83521044 | 1 | 0.197699357 | 0.197699357 | 0.197699357 | 0.23875339 | 0.842386262 | 1 | 0.238753394 | 0.238753394 | 0.238753394 | 0.828048364 | -0.081944297 | FALSE | TRUE | FALSE |
| VPS45 | 0.1976522 | 0.83522632 | 0.2586 | 0.197652152 | 1.814202833 | 3.910589997 | 0.03654549 | 0.889764131 | 0.01425 | 0.036545489 | 0.036545489 | 0.723994446 | 5.408387127 | 0.73306777 | FALSE | TRUE | FALSE |
| PVRIG | 0.1976303 | 0.8352422 | 0.8778 | 16.87937419 | 162.094968 | 162.094968 | 15559.8128 | 0.000126167 | 0.62615 | 15559.81281 | 18251395.23 | 264801219.3 | 1.27013E-05 | -4.896150902 | FALSE | FALSE | FALSE |
| THOC5 | 0.197564 | 0.83525808 | 1 | 0.197564003 | 0.197564003 | 3.896439844 | 0.23895912 | 0.84234026 | 1 | 0.238959119 | 0.238959119 | 4.72072342 | 0.826769047 | -0.082615791 | FALSE | TRUE | FALSE |
| PLCD1 | 0.1975353 | 0.83527395 | 0.60255 | 0.197535341 | 6.871885622 | 27.48965472 | 7.13533935 | 0.242106712 | 0.80675 | 65.75907173 | 23999.9666 | 237426.5374 | 0.027684085 | -1.557769827 | FALSE | FALSE | FALSE |
| TGFBR1 | 0.1974792 | 0.83528982 | 1 | 0.197479171 | 0.197479171 | 3.913296484 | 0.2393724 | 0.842155981 | 1 | 0.2393724 | 0.2393724 | 4.748704913 | 0.824987223 | -0.083552777 | FALSE | TRUE | FALSE |
| FAM204A | 0.1974044 | 0.83530568 | 1 | 0.197404384 | 0.197404384 | 0.197404384 | 1.9187808 | 0.497324766 | 0.5052 | 1.918780798 | 1.918780798 | 1.918780798 | 0.102880112 | -0.987668571 | FALSE | FALSE | FALSE |
| BUD13 | 0.1974036 | 0.83532155 | 1 | 0.19740357 | 0.19740357 | 3.892391515 | 2.13651129 | 0.477764996 | 0.01635 | 2.136511288 | 2.136511288 | 7.833227203 | 0.092395285 | -1.03435019 | FALSE | FALSE | FALSE |
| TICAM1 | 0.1973079 | 0.83533741 | 1 | 0.197307921 | 0.197307921 | 3.878276208 | 0.22277676 | 0.846653847 | 1 | 0.222776757 | 0.222776757 | 4.387959342 | 0.885675524 | -0.052725357 | FALSE | TRUE | FALSE |
| NCAPH2 | 0.1972593 | 0.83535327 | 1 | 1.808657377 | 1.808657377 | 6.677241929 | 0.24569287 | 0.840462404 | 0.0157 | 0.245692868 | 0.245692868 | 4.860456452 | 0.802869386 | -0.095355102 | FALSE | TRUE | FALSE |
| TMEM217 | 0.1972318 | 0.83536912 | 1 | 0.197231792 | 0.197231792 | 0.197231792 | 0.23071886 | 0.844668051 | 1 | 0.230718856 | 2.10752075 | 2.10752075 | 0.854857706 | -0.068106169 | FALSE | TRUE | FALSE |
| WFDC3 | 0.1971973 | 0.83538497 | 0.0064 | 0.197197327 | 0.197197327 | 0.197197327 | 0.2310208 | 0.844608448 | 1 | 0.231020795 | 2.127068877 | 2.127068877 | 0.853591238 | -0.068750052 | FALSE | TRUE | FALSE |
| KMO | 0.1970693 | 0.83540082 | 1 | 1.81305251 | 1.81305251 | 3.906797325 | 0.24567856 | 0.840478103 | 1 | 0.245678558 | 0.245678558 | 4.87559046 | 0.802143039 | -0.095748181 | FALSE | TRUE | FALSE |
| NELFE | 0.1969313 | 0.83541667 | 1 | 1.8048253 | 16.97664525 | 35.73626228 | 0.25296656 | 0.838620613 | 1 | 0.252966558 | 0.252966558 | 5.014851983 | 0.778487652 | -0.108748271 | FALSE | TRUE | FALSE |
| OR51S1 | 0.196909 | 0.83543251 | 1 | 0.196908981 | 0.196908981 | 0.196908981 | 0.88555729 | 0.600471898 | 0.74815 | 8.19814309 | 77.89422623 | 77.89422623 | 0.222356005 | -0.652951137 | FALSE | TRUE | FALSE |
| ARL13B | 0.1969088 | 0.83544835 | 0.50785 | 0.441898581 | 1.767139941 | 1.767139941 | 0.07273076 | 0.881856626 | 1 | 0.07273076 | 0.07273076 | 0.07273076 | 2.707366596 | 0.432547066 | FALSE | TRUE | FALSE |
| RAD17 | 0.1969031 | 0.83546418 | 1 | 1.806883119 | 1.806883119 | 6.657055533 | 0.24536094 | 0.840572234 | 0.0149 | 0.245360942 | 0.245360942 | 4.858744507 | 0.802503873 | -0.095552863 | FALSE | TRUE | FALSE |
| TGM3 | 0.1968798 | 0.83548002 | 0.2681 | 0.04924456 | 0.196879772 | 0.968497297 | 0.08245109 | 0.879706263 | 0.26385 | 0.082451086 | 0.329668624 | 1.624822531 | 2.387837206 | 0.378004715 | FALSE | TRUE | FALSE |
| MIA3 | 0.1968525 | 0.83549585 | 0.5251 | 0.196852481 | 0.196852481 | 3.812319719 | 0.03554356 | 0.89001462 | 1 | 0.035543561 | 0.035543561 | 1.193988122 | 5.538344418 | 0.74337996 | FALSE | TRUE | FALSE |
| BPIFB1 | 0.1968015 | 0.83551168 | 0.0137 | 0.196801515 | 0.196801515 | 3.89773309 | 0.23762553 | 0.842692279 | 1 | 0.237625532 | 0.237625532 | 4.711777208 | 0.828200206 | -0.081864666 | FALSE | TRUE | FALSE |
| FOS | 0.1967513 | 0.8355275 | 1 | 0.196751298 | 0.786712824 | 3.900138636 | 0.24124795 | 0.841708891 | 0.01105 | 0.241247954 | 0.241247954 | 4.787281085 | 0.815556337 | -0.088546033 | FALSE | TRUE | FALSE |
| TBL2 | 0.1967375 | 0.83554332 | 0.0137 | 0.196737544 | 0.196737544 | 3.892112924 | 0.23794767 | 0.842600596 | 0.01355 | 0.237947673 | 0.237947673 | 4.713520842 | 0.826810121 | -0.082594216 | FALSE | TRUE | FALSE |
| LMBRD2 | 0.1966926 | 0.83555914 | 1 | 0.196692636 | 0.196692636 | 3.884951502 | 2.13081326 | 0.478450926 | 0.01555 | 2.130813258 | 2.130813258 | 4.594933909 | 0.092308716 | -1.03475729 | FALSE | FALSE | FALSE |
| KREMEN1 | 0.1966153 | 0.83557495 | 0.0133 | 0.196615298 | 0.196615298 | 3.89426593 | 0.23533941 | 0.843255354 | 0.513 | 0.941217465 | 0.941217465 | 4.66667985 | 0.835454201 | -0.078077353 | FALSE | TRUE | FALSE |
| ANXA6 | 0.1965427 | 0.83559077 | 1 | 0.196542695 | 0.196542695 | 3.870728552 | 0.23764321 | 0.84266173 | 0.01945 | 0.237643212 | 0.237643212 | 4.688782202 | 0.827049481 | -0.082468506 | FALSE | TRUE | FALSE |
| PIPOX | 0.1964014 | 0.83560658 | 0.01115 | 0.049127033 | 0.196401406 | 0.243670375 | 0.02103463 | 0.893556792 | 1 | 0.021034627 | 0.021034627 | 0.417437192 | 9.337052183 | 0.970209786 | FALSE | TRUE | FALSE |
| POLR3F | 0.196352 | 0.83562238 | 1 | 0.196352029 | 0.196352029 | 0.196352029 | 0.23010238 | 0.84490601 | 1 | 2.116289524 | 2.116289524 | 2.116289524 | 0.853324621 | -0.068885724 | FALSE | TRUE | FALSE |
| GSN | 0.1963407 | 0.83563819 | 0.8814 | 1.79513831 | 16.83477562 | 35.24485396 | 0.03751263 | 0.889559855 | 1 | 0.037512627 | 0.037512627 | 0.738129381 | 5.23398912 | 0.718832815 | FALSE | TRUE | FALSE |
| ARFIP2 | 0.1961114 | 0.83565399 | 0.50475 | 1.840331545 | 1.840331545 | 3.893247698 | 0.24536636 | 0.840556553 | 1 | 0.245366355 | 0.245366355 | 0.245366355 | 0.799259731 | -0.097312067 | FALSE | TRUE | FALSE |
| SH3BP5L | 0.1960335 | 0.83566979 | 0.0144 | 0.196033505 | 0.196033505 | 3.877794091 | 0.23760317 | 0.842707549 | 0.01355 | 0.237603173 | 0.237603173 | 4.706278936 | 0.825045823 | -0.08352193 | FALSE | TRUE | FALSE |
| UGT2A1 | 0.1959835 | 0.83568558 | 0.01585 | 0.195983501 | 0.195983501 | 3.874680811 | 2.1406702 | 0.477421352 | 0.0141 | 2.140670201 | 2.140670201 | 4.555979031 | 0.091552403 | -1.038330251 | FALSE | FALSE | FALSE |
| OR2T1 | 0.1959009 | 0.83570138 | 1 | 0.195900907 | 0.195900907 | 0.195900907 | 0.23668435 | 0.842936252 | 1 | 0.236684351 | 0.236684351 | 0.236684351 | 0.827688464 | -0.082133098 | FALSE | TRUE | FALSE |
| PDIA2 | 0.1957898 | 0.83571717 | 0.13615 | 0.195789755 | 1.676073633 | 1.676073633 | 0.3963636 | 0.793258179 | 0.2503 | 0.046660431 | 0.396363599 | 0.92137913 | 0.493965024 | -0.306303801 | FALSE | TRUE | FALSE |
| CD300A | 0.1957848 | 0.83573295 | 0.007 | 0.195784768 | 0.195784768 | 0.195784768 | 0.23663011 | 0.842997127 | 1 | 0.236630114 | 0.236630114 | 0.236630114 | 0.827387371 | -0.082291112 | FALSE | TRUE | FALSE |
| TBK1 | 0.1956238 | 0.83574874 | 1 | 0.195623798 | 0.195623798 | 3.861261708 | 0.23698174 | 0.842875327 | 1 | 0.236981738 | 0.236981738 | 4.684960995 | 0.825480478 | -0.083293193 | FALSE | TRUE | FALSE |
| KLHL22 | 0.1955822 | 0.83576452 | 1 | 0.195582159 | 0.195582159 | 3.847364558 | 0.23720256 | 0.842829604 | 1 | 0.237202561 | 0.237202561 | 4.67532438 | 0.824536458 | -0.083790137 | FALSE | TRUE | FALSE |
| RNASE3 | 0.1955416 | 0.8357803 | 0.0059 | 0.195541593 | 0.195541593 | 0.195541593 | 0.23604923 | 0.843088354 | 1 | 0.236049231 | 0.236049231 | 0.236049231 | 0.828393267 | -0.08176344 | FALSE | TRUE | FALSE |
| RFC3 | 0.1955023 | 0.83579608 | 0.00965 | 0.19550228 | 0.19550228 | 0.19550228 | 0.23672044 | 0.842921025 | 0.0083 | 0.236720444 | 0.236720444 | 0.236720444 | 0.825878311 | -0.083083939 | FALSE | TRUE | FALSE |
| CAV2 | 0.1954866 | 0.83581185 | 0.0052 | 0.195486619 | 0.195486619 | 0.195486619 | 0.23440524 | 0.843497619 | 0.5075 | 0.937335366 | 0.937335366 | 0.937335366 | 0.833968644 | -0.078850278 | FALSE | TRUE | FALSE |
| EMC2 | 0.1954563 | 0.83582762 | 1 | 0.19545634 | 0.19545634 | 0.19545634 | 0.23635295 | 0.843057956 | 1 | 0.236352955 | 0.236352955 | 0.236352955 | 0.826968043 | -0.082511273 | FALSE | TRUE | FALSE |
| RXFP2 | 0.1953811 | 0.83584338 | 0.26695 | 0.195381058 | 1.770858772 | 3.855605297 | 2.62313659 | 0.439017515 | 0.69055 | 2.623136593 | 235.4075298 | 235.4075298 | 0.074483753 | -1.12793845 | FALSE | FALSE | FALSE |
| CHST5 | 0.1952111 | 0.83585915 | 0.75325 | 0.195211068 | 1.831295331 | 3.84472799 | 0.30806511 | 0.822738712 | 0.7535 | 0.308065111 | 2.902685164 | 6.078543475 | 0.633668214 | -0.198138078 | FALSE | TRUE | FALSE |
| DPPA3 | 0.1950571 | 0.83587491 | 1 | 0.1950571 | 0.1950571 | 0.1950571 | 2.08471998 | 0.48219577 | 0.50405 | 2.084719985 | 2.084719985 | 2.084719985 | 0.093565132 | -1.028885968 | FALSE | FALSE | FALSE |
| ST18 | 0.1949917 | 0.83589067 | 1 | 0.194991739 | 0.194991739 | 3.824075158 | 0.23655778 | 0.843012338 | 0.02395 | 0.236557779 | 0.236557779 | 4.650099619 | 0.824288001 | -0.083921022 | FALSE | TRUE | FALSE |
| CCL28 | 0.1949802 | 0.83590643 | 1 | 0.194980162 | 0.194980162 | 0.194980162 | 0.20466079 | 0.850896341 | 1 | 0.204660793 | 0.204660793 | 0.204660793 | 0.952699144 | -0.021044225 | FALSE | TRUE | FALSE |
| NUDC | 0.1949595 | 0.83592218 | 0.5096 | 0.779544796 | 0.779544796 | 0.779544796 | 0.23926672 | 0.842248175 | 1 | 0.23926672 | 0.23926672 | 0.23926672 | 0.814820779 | -0.088937904 | FALSE | TRUE | FALSE |
| G3BP1 | 0.1949538 | 0.83593793 | 1 | 0.19495379 | 0.19495379 | 3.863018657 | 0.23630338 | 0.843073156 | 1 | 0.236303385 | 0.236303385 | 4.687570986 | 0.8250148 | -0.083538261 | FALSE | TRUE | FALSE |
| KCNE2 | 0.1949165 | 0.83595368 | 1 | 0.194916459 | 0.194916459 | 0.194916459 | 0.23354604 | 0.84381449 | 0.50655 | 0.933973175 | 0.933973175 | 0.933973175 | 0.834595427 | -0.078523999 | FALSE | TRUE | FALSE |
| COMMD10 | 0.1948577 | 0.83596942 | 1 | 0.194857738 | 0.194857738 | 0.194857738 | 0.23540831 | 0.843225017 | 1 | 0.235408315 | 0.235408315 | 0.235408315 | 0.827743653 | -0.082104141 | FALSE | TRUE | FALSE |

| | | | | | | | | | | | | | | | | | |
|---|---|---|---|---|---|---|---|---|---|---|---|---|---|---|---|---|---|
| PTGDS | 0.1947554 | 0.83598516 | 1 | 0.194755417 | 0.194755417 | 0.194755417 | 0.23519781 | 0.843300837 | 1 | 0.235197805 | 0.235197805 | 0.235197805 | 0.828049467 | -0.081943718 | FALSE | TRUE | FALSE |
| ZNF592 | 0.1947336 | 0.8360009 | 1 | 0.194733555 | 0.194733555 | 3.789703108 | 0.2363582 | 0.843042753 | 1 | 0.236358197 | 0.236358197 | 4.614701714 | 0.823891693 | -0.084129876 | FALSE | TRUE | FALSE |
| ASF1B | 0.1945068 | 0.83601663 | 1 | 0.19450677 | 0.19450677 | 0.19450677 | 0.22843683 | 0.845291169 | 0.5087 | 2.116706877 | 2.116706877 | 2.116706877 | 0.851468535 | -0.069831396 | FALSE | TRUE | FALSE |
| SNX9 | 0.1944716 | 0.83603237 | 1 | 0.194471551 | 0.194471551 | 3.843843332 | 0.22825197 | 0.845335492 | 1 | 2.10114436 | 2.10114436 | 4.517895106 | 0.852003818 | -0.069558459 | FALSE | TRUE | FALSE |
| KLRG2 | 0.1943839 | 0.8360481 | 1 | 1.773150533 | 1.773150533 | 1.773150533 | 0.23493134 | 0.843437123 | 1 | 2.152934594 | 2.152934594 | 2.152934594 | 0.827407425 | -0.082280586 | FALSE | TRUE | FALSE |
| SLC25A37 | 0.1943749 | 0.83606382 | 1 | 0.194374916 | 0.194374916 | 3.849942407 | 0.92283597 | 0.595673786 | 0.7534 | 0.922835972 | 3.691038511 | 4.576436049 | 0.210627806 | -0.676484297 | FALSE | TRUE | FALSE |
| TOR1A | 0.1943183 | 0.83607955 | 1 | 1.785239262 | 16.8335035 | 16.8335035 | 0.24254509 | 0.841274826 | 1 | 2.238826645 | 2.238826645 | 2.238826645 | 0.801163665 | -0.096278755 | FALSE | TRUE | FALSE |
| PRR14L | 0.1943173 | 0.83609527 | 1 | 0.194317268 | 0.194317268 | 3.870294814 | 0.23513444 | 0.843346293 | 1 | 0.235134444 | 0.235134444 | 4.685591822 | 0.8264092 | -0.082804857 | FALSE | TRUE | FALSE |
| RNF34 | 0.1942816 | 0.83611099 | 0.7568 | 1.808077027 | 7.230704294 | 7.230704294 | 2.22256503 | 0.469925872 | 0.0099 | 2.222565031 | 2.222565031 | 2.222565031 | 0.087413249 | -1.058422737 | FALSE | FALSE | FALSE |
| GHSR | 0.1942362 | 0.8361267 | 1 | 7.254663407 | 69.54116859 | 69.54116859 | 0.2537234 | 0.83833106 | 1 | 0.253723404 | 0.253723404 | 5.025818208 | 0.765543224 | -0.116030283 | FALSE | TRUE | FALSE |
| CD200R1L | 0.1942197 | 0.83614241 | 1 | 1.779347314 | 16.73110764 | 16.73110764 | 0.23496167 | 0.843406858 | 1 | 2.161514581 | 20.40856926 | 20.40856926 | 0.826601642 | -0.082703737 | FALSE | TRUE | FALSE |
| ACSBG2 | 0.1941702 | 0.83615812 | 0.5079 | 0.194170187 | 0.194170187 | 0.749611333 | 0.03530123 | 0.890038052 | 1 | 0.035301233 | 0.035301233 | 0.698241963 | 5.500379714 | 0.740392672 | FALSE | TRUE | FALSE |
| SETD3 | 0.1941108 | 0.83617383 | 1 | 0.194110769 | 0.194110769 | 3.834845362 | 0.2350904 | 0.843376581 | 0.0147 | 0.235090396 | 0.235090396 | 4.651256053 | 0.825685659 | -0.083185258 | FALSE | TRUE | FALSE |
| GRID2 | 0.194095 | 0.83618953 | 0.6969 | 0.194095 | 3.102415947 | 6.537253003 | 0.0837782 | 0.87941162 | 0.764 | 0.083778203 | 0.334866652 | 2.824894912 | 2.316772059 | 0.364883307 | FALSE | TRUE | FALSE |
| ZNF317 | 0.1940898 | 0.83620523 | 1 | 0.194089782 | 0.194089782 | 3.823597903 | 0.23516208 | 0.843331144 | 0.01745 | 0.235162079 | 0.235162079 | 4.641088161 | 0.825344727 | -0.083364619 | FALSE | TRUE | FALSE |
| ASXL2 | 0.1939057 | 0.83622093 | 1 | 0.193905685 | 0.193905685 | 6.5258819 | 0.23541538 | 0.843209844 | 0.0295 | 0.23541538 | 0.23541538 | 4.6072152 | 0.823674667 | -0.084244291 | FALSE | TRUE | FALSE |
| ACSL3 | 0.1938781 | 0.83623662 | 1 | 0.193878064 | 0.193878064 | 3.824599809 | 0.23527509 | 0.843285679 | 1 | 0.23527509 | 0.23527509 | 4.648860674 | 0.82404841 | -0.084047274 | FALSE | TRUE | FALSE |
| FAM193B | 0.1938069 | 0.83625231 | 1 | 1637.379243 | 174375.8564 | 1863234.212 | 0.26577586 | 0.835026479 | 1 | 23.54486192 | 2308.377041 | 2308.377041 | 0.729211945 | -0.137146226 | FALSE | TRUE | FALSE |
| NEUROD6 | 0.1937977 | 0.836268 | 0.9392 | 17.63394904 | 1775.714992 | 1775.714992 | 1.05616015 | 0.582828832 | 0.50575 | 1.056160149 | 1.056160149 | 1.056160149 | 0.183492732 | -0.736381133 | FALSE | FALSE | FALSE |
| AHCYL1 | 0.1937694 | 0.83628369 | 0.51105 | 0.774815914 | 0.774815914 | 6.557977691 | 0.23802064 | 0.842554715 | 0.0128 | 0.238020638 | 0.238020638 | 4.716913162 | 0.81408666 | -0.089329362 | FALSE | TRUE | FALSE |
| ZNF708 | 0.1937067 | 0.83629937 | 0.50765 | 1.733905772 | 15.9197365 | 15.9197365 | 0.03635833 | 0.889826849 | 0.50965 | 0.320675162 | 0.320675162 | 0.721502174 | 5.327711275 | 0.726540681 | FALSE | TRUE | FALSE |
| OR51V1 | 0.193651 | 0.83631505 | 1 | 0.193651042 | 0.193651042 | 0.193651042 | 2.0934519 | 0.480669896 | 0.00705 | 2.093451897 | 2.093451897 | 2.093451897 | 0.092503221 | -1.033843147 | FALSE | FALSE | FALSE |
| BMP8B | 0.1934701 | 0.83633073 | 0.7536 | 0.77364129 | 3.094381782 | 3.830729735 | 3.716415 | 0.370388828 | 0.1281 | 3.716414996 | 14.86898142 | 14.86898142 | 0.052058271 | -1.283510258 | FALSE | FALSE | FALSE |
| ZNF248 | 0.1934522 | 0.8363464 | 1 | 0.193452219 | 0.193452219 | 3.833232645 | 0.23433033 | 0.843527852 | 0.0127 | 0.234330334 | 0.234330334 | 4.64839688 | 0.825553465 | -0.083254795 | FALSE | TRUE | FALSE |
| ATP4B | 0.1933617 | 0.83636207 | 0.0092 | 0.193361657 | 0.193361657 | 0.193361657 | 0.23436731 | 0.843512737 | 1 | 0.234367315 | 0.234367315 | 0.234367315 | 0.825036789 | -0.083526685 | FALSE | TRUE | FALSE |
| IFT27 | 0.1933363 | 0.83637774 | 1 | 0.193336303 | 0.773098263 | 0.773098263 | 2.15247107 | 0.475869343 | 0.00455 | 2.152471073 | 2.152471073 | 2.152471073 | 0.089820628 | -1.046623914 | FALSE | FALSE | FALSE |
| PRMT6 | 0.1933298 | 0.8363934 | 1 | 0.1933298 | 0.1933298 | 3.824031763 | 0.23399409 | 0.843618484 | 1 | 0.233994092 | 0.233994092 | 4.634494268 | 0.826216585 | -0.082906091 | FALSE | TRUE | FALSE |
| OR8K1 | 0.1932767 | 0.83640907 | 1 | 0.193276676 | 0.193276676 | 0.193276676 | 0.23388357 | 0.843648672 | 0.00535 | 0.233883572 | 0.233883572 | 0.233883572 | 0.826379872 | -0.082820269 | FALSE | TRUE | FALSE |
| TFPT | 0.1932731 | 0.83642473 | 1 | 0.193273089 | 0.193273089 | 0.193273089 | 0.23373782 | 0.843724088 | 1 | 0.233737819 | 0.233737819 | 0.233737819 | 0.826879831 | -0.082557601 | FALSE | TRUE | FALSE |
| FANCE | 0.1931667 | 0.83644038 | 1 | 0.193166656 | 0.193166656 | 3.830840317 | 0.23380224 | 0.84370901 | 1 | 0.233802241 | 0.233802241 | 0.233802241 | 0.826196767 | -0.082916509 | FALSE | TRUE | FALSE |
| ZNF532 | 0.1931326 | 0.83645603 | 1 | 0.193132553 | 0.193132553 | 3.75611635 | 0.23430274 | 0.843542964 | 1 | 0.234302744 | 0.234302744 | 4.571960074 | 0.824286348 | -0.083921893 | FALSE | TRUE | FALSE |
| NSUN2 | 0.1930966 | 0.83647169 | 0.5031 | 0.193096613 | 0.193096613 | 3.803277857 | 0.06948451 | 0.882563879 | 1 | 0.069484506 | 0.069484506 | 1.371085606 | 2.778988054 | 0.44388668 | FALSE | TRUE | FALSE |
| TTC1 | 0.1929991 | 0.83648733 | 1 | 0.192999119 | 0.192999119 | 0.192999119 | 2.0865905 | 0.48202671 | 0.509 | 2.086590502 | 2.086590502 | 2.086590502 | 0.092494967 | -1.033881899 | FALSE | FALSE | FALSE |
| IL36RN | 0.1928571 | 0.83650298 | 1 | 0.192857113 | 0.192857113 | 0.192857113 | 0.23384339 | 0.843678847 | 0.00485 | 0.233843392 | 0.233843392 | 0.233843392 | 0.824727659 | -0.08368944 | FALSE | TRUE | FALSE |
| CRISP2 | 0.1928315 | 0.83651862 | 1 | 0.192831526 | 0.192831526 | 0.192831526 | 0.23319964 | 0.843979947 | 1 | 0.233199635 | 0.233199635 | 0.233199635 | 0.826894631 | -0.082549828 | FALSE | TRUE | FALSE |
| C12orf43 | 0.1927218 | 0.83653426 | 1 | 0.192721796 | 0.192721796 | 0.192721796 | 0.23311958 | 0.844009992 | 0.00865 | 0.23311958 | 0.23311958 | 0.23311958 | 0.82670789 | -0.082647917 | FALSE | TRUE | FALSE |
| BICC1 | 0.192708 | 0.8365499 | 1 | 0.19270797 | 0.19270797 | 3.780616405 | 0.2338292 | 0.84369393 | 1 | 0.233829203 | 0.233829203 | 4.597888831 | 0.824139871 | -0.083999075 | FALSE | TRUE | FALSE |
| MAEL | 0.1925733 | 0.83656553 | 1 | 0.192573253 | 0.192573253 | 3.819219494 | 0.2333233 | 0.843919823 | 1 | 0.233323295 | 0.233323295 | 4.632056244 | 0.825349446 | -0.083362136 | FALSE | TRUE | FALSE |
| LEFTY2 | 0.1925589 | 0.83658116 | 1 | 0.192558862 | 0.192558862 | 3.813922812 | 0.23385979 | 0.843663761 | 1 | 0.233859791 | 0.233859791 | 4.637340494 | 0.823394485 | -0.084392046 | FALSE | TRUE | FALSE |
| PAICS | 0.1924286 | 0.83659679 | 1 | 0.769670169 | 0.769670169 | 3.826110536 | 0.23040863 | 0.84472761 | 1 | 0.921584516 | 3.687059549 | 3.687059549 | 0.835162078 | -0.078229234 | FALSE | TRUE | FALSE |
| TTLL11 | 0.1922966 | 0.83661242 | 0.01355 | 0.192296607 | 0.192296607 | 3.811678138 | 177.011384 | 0.015097532 | 0.12805 | 18.33700529 | 177.0113836 | 177.0113836 | 0.001086352 | -2.964029575 | FALSE | FALSE | FALSE |
| POGLUT1 | 0.1922866 | 0.83662804 | 1 | 0.192286622 | 0.192286622 | 3.817392801 | 0.23289425 | 0.844115053 | 1 | 0.232894247 | 0.232894247 | 4.627657593 | 0.825639209 | -0.083209691 | FALSE | TRUE | FALSE |
| DTX4 | 0.1921769 | 0.83664366 | 1 | 0.192176893 | 0.192176893 | 3.782874069 | 2.10381831 | 0.479818039 | 0.5126 | 2.103818309 | 2.103818309 | 7.997099184 | 0.091346716 | -1.039307062 | FALSE | FALSE | FALSE |
| ANAPC2 | 0.1921377 | 0.83665928 | 0.02965 | 0.192137665 | 0.192137665 | 3.750383329 | 0.23363412 | 0.843739162 | 1 | 0.233634123 | 0.233634123 | 4.573571435 | 0.822386999 | -0.084923764 | FALSE | TRUE | FALSE |
| UNC5A | 0.1920907 | 0.83667489 | 0.9853 | 170.6596582 | 17865.10334 | 189314.0805 | 0.24090891 | 0.841770709 | 1 | 215.6451096 | 2185.371437 | 22677.54113 | 0.797358348 | -0.098346454 | FALSE | TRUE | FALSE |
| SLC25A20 | 0.1919702 | 0.8366905 | 0.939 | 0.767659429 | 12.28468102 | 12.28468102 | 1.12138941 | 0.575905204 | 0.0088 | 0.280408268 | 1.121389412 | 1.121389412 | 0.171189561 | -0.766522723 | FALSE | FALSE | FALSE |
| MBOAT1 | 0.1919159 | 0.83670611 | 0.68985 | 1.730013135 | 63.96756374 | 63.96756374 | 0.1437319 | 0.865936108 | 0.75305 | 0.31899856 | 1.275696245 | 1.275696245 | 1.335235188 | 0.125557769 | FALSE | TRUE | FALSE |
| ERGIC1 | 0.1919143 | 0.83672172 | 1 | 0.191914288 | 0.191914288 | 0.191914288 | 18.1527459 | 0.142890938 | 0.6874 | 18.15274587 | 1771.77729 | 1771.77729 | 0.010572191 | -1.975835018 | FALSE | FALSE | FALSE |
| COX4I2 | 0.191908 | 0.83673732 | 1 | 0.191907989 | 0.191907989 | 0.191907989 | 0.23222272 | 0.844384559 | 1 | 0.23222272 | 0.23222272 | 0.23222272 | 0.826396265 | -0.082811654 | FALSE | TRUE | FALSE |
| TESPA1 | 0.191892 | 0.83675292 | 1 | 0.191891976 | 0.191891976 | 3.800254899 | 0.23257145 | 0.844190012 | 1 | 0.232571453 | 0.232571453 | 4.611302472 | 0.825088259 | -0.083499593 | FALSE | TRUE | FALSE |
| RPIA | 0.191851 | 0.83676851 | 0.51115 | 0.767197107 | 0.767197107 | 3.8067737 | 2.06776741 | 0.483880888 | 0.7541 | 2.067767408 | 19.81659661 | 19.81659661 | 0.092781689 | -1.032537725 | FALSE | FALSE | FALSE |
| AADACL4 | 0.1917387 | 0.83678411 | 1 | 0.191738653 | 0.191738653 | 3.800457791 | 0.23274995 | 0.844145045 | 1 | 0.232749952 | 0.232749952 | 0.232749952 | 0.823796749 | -0.084179926 | FALSE | TRUE | FALSE |
| C21orf58 | 0.1916141 | 0.8367997 | 0.0078 | 0.191614058 | 0.191614058 | 0.191614058 | 0.23194356 | 0.844459254 | 1 | 0.231943562 | 0.231943562 | 0.231943562 | 0.826123633 | -0.082954954 | FALSE | TRUE | FALSE |
| SAC3D1 | 0.1916105 | 0.83681529 | 1 | 1.762229735 | 1.762229735 | 1.762229735 | 2.05414851 | 0.484719613 | 0.51015 | 2.05414851 | 2.05414851 | 2.05414851 | 0.09327977 | -1.030212531 | FALSE | FALSE | FALSE |
| FGF8 | 0.1914028 | 0.83683087 | 1 | 0.191402848 | 0.191402848 | 0.191402848 | 0.23249404 | 0.844264898 | 1 | 0.232494041 | 0.232494041 | 0.232494041 | 0.823259156 | -0.084463431 | FALSE | TRUE | FALSE |
| MTA2 | 0.1913737 | 0.83684646 | 1 | 0.191373748 | 0.191373748 | 3.769873865 | 0.23250732 | 0.844234953 | 1 | 0.232507325 | 0.232507325 | 4.588461099 | 0.823086965 | -0.084554276 | FALSE | TRUE | FALSE |
| GABRA5 | 0.1913725 | 0.83686204 | 1 | 0.19137249 | 0.19137249 | 3.786702425 | 0.23253616 | 0.844219976 | 1 | 0.232536161 | 0.232536161 | 4.607104008 | 0.822979485 | -0.084610991 | FALSE | TRUE | FALSE |
| GUCA1C | 0.1913136 | 0.83687761 | 1 | 0.191313579 | 0.191313579 | 0.191313579 | 0.23193573 | 0.844474185 | 1 | 0.231935729 | 0.231935729 | 0.231935729 | 0.824856006 | -0.083621859 | FALSE | TRUE | FALSE |
| ZBTB17 | 0.1913066 | 0.83689319 | 1 | 0.191306615 | 0.191306615 | 3.745506515 | 0.23240326 | 0.844324752 | 1 | 0.232403261 | 0.232403261 | 4.561668258 | 0.823166656 | -0.08451223 | FALSE | TRUE | FALSE |
| PTPRC | 0.1912567 | 0.83690876 | 0.51495 | 0.191256665 | 0.191256665 | 3.738005359 | 0.2622061 | 0.836059957 | 0.88275 | 1.048724537 | 9.668951629 | 47.37772651 | 0.72941349 | -0.137026209 | FALSE | TRUE | FALSE |
| ADCY8 | 0.1912465 | 0.83692433 | 0.6965 | 0.19124655 | 3.058617929 | 14.89304876 | 0.02109674 | 0.893527336 | 0.0307 | 0.021096742 | 0.021096742 | 0.41215974 | 9.065217159 | 0.957378212 | FALSE | TRUE | FALSE |
| PRSS22 | 0.1912256 | 0.83693989 | 1 | 0.191225573 | 0.191225573 | 3.796407422 | 0.2324955 | 0.844249927 | 0.00885 | 0.232495496 | 0.232495496 | 0.232495496 | 0.822491516 | -0.084868573 | FALSE | TRUE | FALSE |
| BIN1 | 0.1912076 | 0.83695545 | 0.0166 | 0.19120755 | 0.19120755 | 3.777143604 | 0.23245565 | 0.844279866 | 1 | 0.232455648 | 0.232455648 | 4.598756577 | 0.822554978 | -0.084835065 | FALSE | TRUE | FALSE |
| L3MBTL3 | 0.1911731 | 0.83697101 | 1 | 0.191173076 | 0.191173076 | 3.766772993 | 3.63648828 | 0.375429854 | 0.2611 | 0.909084984 | 3.636488284 | 17.94409302 | 0.052570794 | -1.279255464 | FALSE | FALSE | FALSE |
| KHDC1 | 0.1910155 | 0.83698657 | 1 | 1.736118892 | 16.19533863 | 16.19533863 | 0.24570474 | 0.840430995 | 1 | 0.24570474 | 0.24570474 | 0.24570474 | 0.777418897 | -0.109344907 | FALSE | TRUE | FALSE |
| SAMM50 | 0.1909345 | 0.83700212 | 1 | 0.1909345 | 0.1909345 | 3.777311258 | 0.22458472 | 0.846275248 | 1 | 2.068229479 | 2.068229479 | 4.448814876 | 0.850166917 | -0.070495799 | FALSE | TRUE | FALSE |
| STX7 | 0.1909108 | 0.83701767 | 1 | 0.190910803 | 0.190910803 | 0.190910803 | 0.20239699 | 0.851554187 | 0.5054 | 1.891810652 | 1.891810652 | 1.891810652 | 0.943249212 | -0.025373549 | FALSE | TRUE | FALSE |
| CCNB2 | 0.1909034 | 0.83703322 | 1 | 0.190903394 | 0.190903394 | 0.190903394 | 0.23137205 | 0.844548801 | 0.00975 | 0.231372049 | 0.231372049 | 0.231372049 | 0.825092725 | -0.083497242 | FALSE | TRUE | FALSE |
| MBD6 | 0.1908399 | 0.83704877 | 1 | 0.190839865 | 0.190839865 | 3.7384931 | 1.96473871 | 0.491993405 | 0.87825 | 18.75261032 | 183.578695 | 368.2654735 | 0.09713244 | -1.012635702 | FALSE | FALSE | FALSE |
| ETS2 | 0.1908129 | 0.83706431 | 0.0141 | 0.190812897 | 0.190812897 | 3.775093967 | 0.23015451 | 0.844876306 | 1 | 0.230154507 | 0.230154507 | 4.559347636 | 0.829064353 | -0.081411758 | FALSE | TRUE | FALSE |
| ZGLP1 | 0.1907602 | 0.83707985 | 1 | 0.190760174 | 0.190760174 | 0.190760174 | 0.23217055 | 0.844399504 | 1 | 0.232170554 | 0.232170554 | 0.232170554 | 0.821638104 | -0.085319428 | FALSE | TRUE | FALSE |
| TAF1C | 0.1907555 | 0.83709538 | 0.75315 | 0.190755521 | 1.70188533 | 3.733889813 | 0.03593029 | 0.889905157 | 1 | 0.035930291 | 0.035930291 | 0.705110794 | 5.309044771 | 0.725016388 | FALSE | TRUE | FALSE |
| OR10K1 | 0.1906029 | 0.83711092 | 1 | 1.74998275 | 16.49049862 | 16.49049862 | 0.23841916 | 0.842447558 | 1 | 0.238419157 | 2.199492452 | 2.199492452 | 0.799444405 | -0.097211732 | FALSE | TRUE | FALSE |

| | | | | | | | | | | | | | | | | | |
|---|---|---|---|---|---|---|---|---|---|---|---|---|---|---|---|---|---|
| TMEM70 | 0.1905548 | 0.83712645 | 1 | 1.74011369 | 1.74011369 | 1.74011369 | 0.23807217 | 0.842539415 | 1 | 0.238072165 | 0.238072165 | 0.238072165 | 0.800407548 | -0.096688825 | FALSE | TRUE | FALSE |
| CEACAM16 | 0.1904103 | 0.83714197 | 1 | 0.190410255 | 0.190410255 | 3.768981176 | 0.23141702 | 0.844533883 | 1 | 0.231417022 | 0.231417022 | 4.586344715 | 0.822801423 | -0.084704966 | FALSE | TRUE | FALSE |
| KLHL6 | 0.1903318 | 0.8371575 | 0.5043 | 0.190331777 | 0.190331777 | 1.611024293 | 0.06881273 | 0.882686863 | 1 | 0.068812733 | 0.068812733 | 1.357608599 | 2.765938342 | 0.441842495 | FALSE | TRUE | FALSE |
| ZNF260 | 0.1902817 | 0.83717302 | 1 | 0.190281741 | 0.190281741 | 3.781205415 | 0.23108274 | 0.84457863 | 0.0092 | 0.23108274 | 0.23108274 | 0.23108274 | 0.823435541 | -0.084370392 | FALSE | TRUE | FALSE |
| LATS2 | 0.190232 | 0.83718854 | 1 | 0.760832417 | 0.760832417 | 14.82888131 | 0.23404941 | 0.843573181 | 1 | 0.234049413 | 0.234049413 | 4.575711511 | 0.812785645 | -0.090023975 | FALSE | TRUE | FALSE |
| ZNF189 | 0.1900124 | 0.83720406 | 1 | 0.759958725 | 0.759958725 | 3.761790469 | 2.03046265 | 0.487386651 | 0.7544 | 2.030462653 | 19.28289748 | 40.24722771 | 0.09358085 | -1.028813013 | FALSE | FALSE | FALSE |
| ATP6V1D | 0.1900063 | 0.83721957 | 0.00725 | 0.190006256 | 0.190006256 | 0.190006256 | 0.23076137 | 0.844653154 | 1 | 0.230761371 | 0.230761371 | 0.230761371 | 0.82338849 | -0.084395208 | FALSE | TRUE | FALSE |
| RAB38 | 0.1899345 | 0.83723508 | 1 | 0.189934515 | 0.189934515 | 0.189934515 | 0.23091854 | 0.844623353 | 1 | 0.23091854 | 0.23091854 | 0.23091854 | 0.822517392 | -0.08485491 | FALSE | TRUE | FALSE |
| ZNHIT2 | 0.1899291 | 0.83725059 | 1 | 0.189929127 | 0.189929127 | 3.751373464 | 0.23036261 | 0.844757373 | 0.01225 | 0.230362611 | 0.230362611 | 4.556796981 | 0.824478966 | -0.08382042 | FALSE | TRUE | FALSE |
| DLX2 | 0.1897658 | 0.8372661 | 1 | 0.189765804 | 0.189765804 | 3.76359395 | 0.22392108 | 0.846421073 | 0.5065 | 2.118337044 | 2.118337044 | 2.118337044 | 0.847467348 | -0.071877026 | FALSE | TRUE | FALSE |
| TBC1D2 | 0.189761 | 0.8372816 | 0.0287 | 0.189761003 | 0.189761003 | 3.710996119 | 0.03454166 | 0.890225177 | 1 | 0.034541657 | 0.034541657 | 0.677307976 | 5.493685572 | 0.739863799 | FALSE | TRUE | FALSE |
| GAPT | 0.1895842 | 0.8372971 | 1 | 0.189584168 | 0.189584168 | 0.189584168 | 0.22959456 | 0.844995057 | 1 | 0.229594558 | 0.229594558 | 0.229594558 | 0.825734589 | -0.083159523 | FALSE | TRUE | FALSE |
| MNAT1 | 0.1895267 | 0.8373126 | 1 | 0.189526696 | 0.189526696 | 0.189526696 | 0.22990289 | 0.844935704 | 1 | 0.229902894 | 0.229902894 | 0.229902894 | 0.824377165 | -0.083874046 | FALSE | TRUE | FALSE |
| USP2 | 0.1895138 | 0.8373281 | 1 | 0.189513776 | 0.189513776 | 3.736330744 | 0.23017424 | 0.84486145 | 1 | 0.230174239 | 0.230174239 | 4.545739028 | 0.823349205 | -0.084415929 | FALSE | TRUE | FALSE |
| CD209 | 0.1894842 | 0.83734359 | 1 | 0.189484199 | 0.189484199 | 3.760630687 | 0.22978071 | 0.844965386 | 1 | 0.229780715 | 0.229780715 | 0.229780715 | 0.824630556 | -0.083740577 | FALSE | TRUE | FALSE |
| TOX2 | 0.1894595 | 0.83735908 | 1 | 0.189459455 | 0.189459455 | 3.748021021 | 2.07181604 | 0.483208212 | 0.50995 | 2.071816038 | 2.071816038 | 4.422710655 | 0.09144608 | -1.038834907 | FALSE | FALSE | FALSE |
| RAI14 | 0.1894518 | 0.83737456 | 1 | 0.189451832 | 0.189451832 | 3.728086797 | 0.22350184 | 0.846493889 | 0.50865 | 2.091803259 | 2.091803259 | 41.2428037 | 0.847652211 | -0.071782301 | FALSE | TRUE | FALSE |
| SLC15A4 | 0.189444 | 0.83739005 | 0.50535 | 0.757710216 | 0.757710216 | 3.755263064 | 0.22559996 | 0.845953472 | 0.5054 | 2.081832212 | 2.081832212 | 4.47673795 | 0.83973431 | -0.075858102 | FALSE | TRUE | FALSE |
| OSBPL2 | 0.189428 | 0.83740553 | 1 | 0.189428005 | 0.189428005 | 3.746732549 | 18.9774705 | 0.137590185 | 0.2581 | 2.000334394 | 18.97747051 | 18.97747051 | 0.009981731 | -2.000794139 | FALSE | FALSE | FALSE |
| TCF3 | 0.1892909 | 0.837421 | 0.2649 | 0.189290921 | 1.788286893 | 3.723516247 | 0.30991021 | 0.822038815 | 0.50555 | 0.309910207 | 0.309910207 | 1.135278492 | 0.610792792 | -0.214106096 | FALSE | TRUE | FALSE |
| NFKB2 | 0.1892263 | 0.83743648 | 1 | 0.189226255 | 0.189226255 | 3.717830113 | 0.23037405 | 0.844742493 | 0.0244 | 0.23037405 | 0.23037405 | 4.535886914 | 0.821387021 | -0.085452164 | FALSE | TRUE | FALSE |
| MKI67 | 0.1892225 | 0.83745195 | 0.8416 | 14.58054335 | 11762.58692 | 114527.6869 | 0.00093846 | 0.899501799 | 1 | 0.562141257 | 18.7334568 | 94.60470058 | 201.6306922 | 2.304556641 | FALSE | TRUE | FALSE |
| FANCF | 0.1892008 | 0.83746742 | 1 | 1.728316278 | 16.2039611 | 16.2039611 | 2.17260713 | 0.473959814 | 0.0113 | 2.172607132 | 2.172607132 | 4.684573269 | 0.087084678 | -1.060058251 | FALSE | FALSE | FALSE |
| DUOXA1 | 0.1891882 | 0.83748288 | 0.01475 | 0.189188215 | 0.189188215 | 3.742260489 | 0.22991449 | 0.844920858 | 0.01275 | 0.229914493 | 0.229914493 | 4.553852889 | 0.822863375 | -0.084672267 | FALSE | TRUE | FALSE |
| XRCC4 | 0.1890275 | 0.83749835 | 1 | 0.189027516 | 0.189027516 | 0.189027516 | 2.03323384 | 0.48688878 | 0.50565 | 2.033233837 | 2.033233837 | 2.033233837 | 0.092968901 | -1.031662301 | FALSE | FALSE | FALSE |
| UHRF2 | 0.188962 | 0.83751381 | 1 | 0.188961999 | 0.188961999 | 3.718738317 | 0.230194 | 0.84484659 | 1 | 0.230193999 | 0.230193999 | 4.53890676 | 0.820881519 | -0.085719522 | FALSE | TRUE | FALSE |
| CCER1 | 0.1886634 | 0.83752926 | 1 | 1.722665269 | 16.14403666 | 16.14403666 | 0.24339805 | 0.840978926 | 1 | 0.243398047 | 0.243398047 | 4.826966932 | 0.775122776 | -0.110629502 | FALSE | TRUE | FALSE |
| LRRTM4 | 0.1886488 | 0.83754472 | 1 | 0.188648825 | 0.188648825 | 3.73584154 | 2.05538907 | 0.484552058 | 0.01265 | 2.055389072 | 2.055389072 | 4.408128013 | 0.091782538 | -1.037239938 | FALSE | FALSE | FALSE |
| C11orf42 | 0.1886406 | 0.83756017 | 1 | 0.18864062 | 0.18864062 | 3.745406359 | 0.22879634 | 0.845217243 | 0.0084 | 0.228796341 | 0.228796341 | 0.228796341 | 0.82449142 | -0.08381386 | FALSE | TRUE | FALSE |
| KCNB1 | 0.1885836 | 0.83757562 | 0.99805 | 1715.892651 | 189618.528 | 2063852.905 | 0.30717419 | 0.823028654 | 0.96995 | 28.12565784 | 712.905658 | 2849.872354 | 0.613930641 | -0.211880691 | FALSE | TRUE | FALSE |
| CDKN2B | 0.1883894 | 0.83759107 | 1 | 0.188389383 | 0.188389383 | 0.188389383 | 0.22943337 | 0.845054368 | 0.00395 | 0.229433374 | 0.229433374 | 0.229433374 | 0.821107147 | -0.085600168 | FALSE | TRUE | FALSE |
| FAM162B | 0.1883625 | 0.83760651 | 1 | 0.188362511 | 0.188362511 | 0.188362511 | 0.20454656 | 0.850910106 | 0.004 | 0.204546557 | 0.204546557 | 0.204546557 | 0.920878425 | -0.035797702 | FALSE | TRUE | FALSE |
| SLC35A3 | 0.1883273 | 0.83762196 | 1 | 16.14986593 | 155.4022384 | 155.4022384 | 0.25063585 | 0.839212615 | 1 | 0.250635849 | 0.250635849 | 0.250635849 | 0.751398257 | -0.124129817 | FALSE | TRUE | FALSE |
| TMOD3 | 0.1882935 | 0.83763739 | 0.00895 | 0.188293475 | 0.188293475 | 0.188293475 | 0.2292961 | 0.845084006 | 1 | 0.229296096 | 0.229296096 | 0.229296096 | 0.821180464 | -0.085561391 | FALSE | TRUE | FALSE |
| EZR | 0.1882714 | 0.83765283 | 0.696 | 0.752460782 | 28.51864085 | 28.51864085 | 0.03364615 | 0.890435003 | 1 | 0.312075627 | 1.247393212 | 2.65642362 | 5.595630783 | 0.747849051 | FALSE | TRUE | FALSE |
| STARD5 | 0.1882443 | 0.83766826 | 1 | 0.18824429 | 0.18824429 | 0.18824429 | 0.22927595 | 0.845098821 | 0.0054 | 0.229275946 | 0.229275946 | 0.229275946 | 0.821038112 | -0.085636683 | FALSE | TRUE | FALSE |
| TTC21B | 0.1882175 | 0.83768369 | 0.67155 | 0.86091424 | 55.06022307 | 488.8704259 | 0.12198245 | 0.870818501 | 0.6144 | 0.487860656 | 20.50744786 | 147.5207499 | 1.542988583 | 0.188362713 | FALSE | TRUE | FALSE |
| LRRC30 | 0.1881965 | 0.83769912 | 0.50645 | 1.720953461 | 1.720953461 | 1.720953461 | 0.22156771 | 0.846900432 | 1 | 2.033110977 | 19.14740401 | 19.14740401 | 0.849386134 | -0.070894833 | FALSE | TRUE | FALSE |
| PCYOX1L | 0.1881212 | 0.83771454 | 1 | 0.188121236 | 0.188121236 | 3.72148495 | 0.22847023 | 0.845261607 | 1 | 0.228470227 | 0.228470227 | 4.525600257 | 0.82339497 | -0.084391791 | FALSE | TRUE | FALSE |
| ALOX15B | 0.1880465 | 0.83772996 | 0.9134 | 14.46038605 | 1230.940212 | 11780.06798 | 8.28230017 | 0.222234552 | 0.663 | 44.89092025 | 6923.87183 | 68429.78952 | 0.022704628 | -1.643885608 | FALSE | FALSE | FALSE |
| BBS4 | 0.1879741 | 0.83774538 | 0.50505 | 0.18797405 | 0.18797405 | 0.419152553 | 0.03403244 | 0.890349605 | 1 | 0.034032441 | 0.034032441 | 0.034032441 | 5.523378463 | 0.742204802 | FALSE | TRUE | FALSE |
| CHRNB1 | 0.1878939 | 0.8377608 | 0.70075 | 0.187893913 | 3.005598472 | 3.706651264 | 4.99876872 | 0.306226234 | 0.04275 | 1.249577777 | 4.99876872 | 20.00190103 | 0.037588039 | -1.424950331 | FALSE | FALSE | FALSE |
| TNFSF11 | 0.1878805 | 0.83777621 | 1 | 0.187880467 | 0.187880467 | 0.187880467 | 0.22893428 | 0.845158055 | 1 | 0.228934283 | 0.228934283 | 0.228934283 | 0.820674234 | -0.085829201 | FALSE | TRUE | FALSE |
| SLA2 | 0.1878069 | 0.83779162 | 0.5068 | 0.751076494 | 0.751076494 | 0.751076494 | 3.57417261 | 0.379921729 | 0.13175 | 3.574172611 | 14.30119226 | 14.30119226 | 0.052545569 | -1.279463904 | FALSE | FALSE | FALSE |
| ZNF324B | 0.1877673 | 0.83780703 | 1 | 0.187767302 | 0.187767302 | 3.689327078 | 1.98092484 | 0.490520326 | 0.2628 | 1.980924838 | 18.76086181 | 39.00438662 | 0.094787696 | -1.023248032 | FALSE | FALSE | FALSE |
| PAQR3 | 0.1877509 | 0.83782243 | 1 | 16.0308694 | 1514.945045 | 1514.945045 | 0.2278765 | 0.84539455 | 1 | 19.63769753 | 1872.97434 | 1872.97434 | 0.823915243 | -0.084117462 | FALSE | TRUE | FALSE |
| AMOTL2 | 0.1877462 | 0.83783783 | 1 | 0.18774615 | 0.18774615 | 3.68262583 | 0.22829403 | 0.84532072 | 1 | 0.228294035 | 0.228294035 | 4.488352085 | 0.822387456 | -0.084923523 | FALSE | TRUE | FALSE |
| MTPAP | 0.187737 | 0.83785323 | 1 | 0.18773696 | 0.18773696 | 3.710620226 | 0.22832442 | 0.845305946 | 1 | 0.228324424 | 0.228324424 | 4.519211165 | 0.822237746 | -0.08500259 | FALSE | TRUE | FALSE |
| GJD2 | 0.1876991 | 0.83786863 | 0.00975 | 0.187699067 | 0.187699067 | 0.187699067 | 0.2288801 | 0.845172856 | 0.0094 | 0.228880105 | 0.228880105 | 0.228880105 | 0.820075939 | -0.08614593 | FALSE | TRUE | FALSE |
| NKX2-2 | 0.1876042 | 0.83788402 | 1 | 0.187604236 | 0.187604236 | 3.717391017 | 0.22621209 | 0.845806771 | 0.51045 | 0.9045234 | 0.9045234 | 0.9045234 | 0.829328965 | -0.081273166 | FALSE | TRUE | FALSE |
| FEZ2 | 0.187563 | 0.83789941 | 1 | 1507.872141 | 156657.2853 | 1653345.974 | 2.12657063 | 0.478793222 | 0.9935 | 1907.904171 | 200441.5456 | 801748.1449 | 0.088199749 | -1.054532652 | FALSE | FALSE | FALSE |
| SLC35E4 | 0.1875243 | 0.8379148 | 1 | 0.187524342 | 0.187524342 | 3.717042681 | 2.03096982 | 0.487054864 | 0.50225 | 2.030969821 | 2.030969821 | 2.030969821 | 0.092332412 | -1.03464582 | FALSE | FALSE | FALSE |
| GAL | 0.1874952 | 0.83793018 | 1 | 0.187495179 | 0.187495179 | 0.187495179 | 0.22735588 | 0.845497802 | 1 | 0.227355884 | 0.227355884 | 0.227355884 | 0.824677047 | -0.083716093 | FALSE | TRUE | FALSE |
| ZNF181 | 0.1874351 | 0.83794556 | 0.0123 | 0.187435072 | 0.187435072 | 0.430967569 | 0.03349596 | 0.890481533 | 1 | 0.292087371 | 0.292087371 | 0.665084286 | 5.595750873 | 0.747858371 | FALSE | TRUE | FALSE |
| ZNF707 | 0.1873785 | 0.83796094 | 1 | 0.187378452 | 0.187378452 | 3.705182447 | 17.8935402 | 0.144144918 | 0.5079 | 17.89354023 | 172.6172452 | 172.6172452 | 0.010471849 | -1.979976627 | FALSE | FALSE | FALSE |
| LARP7 | 0.1873219 | 0.83797631 | 0.5076 | 1.653338986 | 14.96820187 | 14.96820187 | 0.03656737 | 0.889756287 | 1 | 0.036567374 | 0.036567374 | 0.72511222 | 5.122650834 | 0.709494755 | FALSE | TRUE | FALSE |
| YME1L1 | 0.1872448 | 0.83799169 | 0.508 | 1.794103203 | 17.63145396 | 35.37178023 | 0.13828525 | 0.867072597 | 0.51335 | 0.138285245 | 0.138285245 | 1.169882691 | 1.35404756 | 0.131633919 | FALSE | TRUE | FALSE |
| ZNF836 | 0.1872341 | 0.83800706 | 0.51195 | 0.187234082 | 0.187234082 | 3.680835124 | 0.03414829 | 0.890326294 | 1 | 0.034148285 | 0.034148285 | 0.67269919 | 5.482971707 | 0.739016005 | FALSE | TRUE | FALSE |
| DDX4 | 0.1871756 | 0.83802242 | 1 | 0.748609399 | 0.748609399 | 14.77264447 | 0.23051567 | 0.844712724 | 1 | 0.230515672 | 0.230515672 | 4.556164489 | 0.811986259 | -0.09045132 | FALSE | TRUE | FALSE |
| RSBN1 | 0.1871169 | 0.83803779 | 0.5148 | 1.725470817 | 1.725470817 | 33.85901154 | 2.04092776 | 0.485889887 | 0.7567 | 2.040927763 | 19.36422158 | 40.14013374 | 0.091682267 | -1.037714658 | FALSE | FALSE | FALSE |
| DCAF11 | 0.1870547 | 0.83805315 | 0.01965 | 0.187054657 | 0.187054657 | 3.691282514 | 0.22075045 | 0.847045141 | 1 | 2.044499504 | 2.044499504 | 4.394258559 | 0.847357972 | -0.07193308 | FALSE | TRUE | FALSE |
| FUT8 | 0.1870289 | 0.83806851 | 0.01635 | 0.187028885 | 0.187028885 | 3.698996022 | 0.22784997 | 0.845424063 | 0.0145 | 0.227849966 | 0.227849966 | 4.51236858 | 0.820842278 | -0.085740283 | FALSE | TRUE | FALSE |
| CHSY1 | 0.1870122 | 0.83808386 | 0.5113 | 1.758003805 | 1.758003805 | 34.58847101 | 0.23539078 | 0.843240187 | 1 | 0.235390777 | 0.235390777 | 4.64034315 | 0.794475604 | -0.099919434 | FALSE | TRUE | FALSE |
| PINK1 | 0.1870075 | 0.83809921 | 0.01725 | 0.187007529 | 0.187007529 | 0.705463527 | 0.03333169 | 0.890504785 | 1 | 0.133266824 | 0.133266824 | 1.128930065 | 5.610501961 | 0.749001718 | FALSE | TRUE | FALSE |
| OR8S1 | 0.1868708 | 0.83811456 | 1 | 0.186870798 | 0.186870798 | 0.186870798 | 0.21344781 | 0.848789714 | 1 | 1.950066085 | 18.28532409 | 18.28532409 | 0.875487063 | -0.057750267 | FALSE | TRUE | FALSE |
| ZNF565 | 0.1868495 | 0.83812991 | 0.01735 | 0.186849535 | 0.186849535 | 3.698657506 | 0.21366226 | 0.848662329 | 1 | 1.964425812 | 18.53689177 | 18.53689177 | 0.874508845 | -0.058235793 | FALSE | TRUE | FALSE |
| TGM4 | 0.186787 | 0.83814525 | 0.8215 | 1.665473108 | 15.22286129 | 142.5489951 | 3.37538116 | 0.390566853 | 0.50655 | 3.375381156 | 124.917789 | 1184.595218 | 0.055338049 | -1.256976157 | FALSE | FALSE | FALSE |
| ARMC10 | 0.1866883 | 0.83816059 | 1 | 1.723928928 | 1.723928928 | 1.723928928 | 0.21441651 | 0.848449544 | 0.50155 | 1.986403288 | 1.986403288 | 1.986403288 | 0.870680508 | -0.060141178 | FALSE | TRUE | FALSE |
| CHPT1 | 0.1866493 | 0.83817593 | 1 | 1.711113076 | 1.711113076 | 1.711113076 | 0.2340011 | 0.843603386 | 1 | 0.234001101 | 0.234001101 | 0.234001101 | 0.797642707 | -0.098191601 | FALSE | TRUE | FALSE |
| NPHS2 | 0.1866176 | 0.83819127 | 0.5031 | 0.186617633 | 0.746205023 | 0.746205023 | 0.07778044 | 0.880907631 | 0.68965 | 0.310963598 | 4.974087567 | 4.974087567 | 2.399287403 | 0.380082274 | FALSE | TRUE | FALSE |
| RIMBP2 | 0.1866106 | 0.8382066 | 0.03665 | 0.186610551 | 0.186610551 | 3.626877232 | 0.22735844 | 0.845483059 | 1 | 0.22735844 | 0.22735844 | 4.433869028 | 0.82077688 | -0.085774886 | FALSE | TRUE | FALSE |
| ZNF48 | 0.1865677 | 0.83822193 | 1 | 0.186567712 | 0.186567712 | 3.672786146 | 0.2270794 | 0.84552728 | 0.01755 | 0.227079403 | 0.227079403 | 4.478748448 | 0.821596806 | -0.085341258 | FALSE | TRUE | FALSE |
| NTAN1 | 0.1865433 | 0.83823726 | 1 | 0.186543313 | 0.186543313 | 0.186543313 | 0.22649272 | 0.845733319 | 1 | 0.226492724 | 0.226492724 | 0.226492724 | 0.823617243 | -0.08427457 | FALSE | TRUE | FALSE |

| | | | | | | | | | | | | | | | | | |
|---|---|---|---|---|---|---|---|---|---|---|---|---|---|---|---|---|---|
| SHCBP1L | 0.1865432 | 0.83825258 | 0.75825 | 0.186543196 | 1.651834671 | 3.689631011 | 0.29088001 | 0.827815421 | 0.75395 | 0.290880005 | 2.603459531 | 2.603459531 | 0.641306355 | -0.192934456 | FALSE | TRUE | FALSE |
| PXK | 0.1864996 | 0.8382679 | 1 | 0.745954549 | 0.745954549 | 3.691582888 | 0.22947172 | 0.845024718 | 1 | 0.229471722 | 0.229471722 | 4.547801714 | 0.812734571 | -0.090051266 | FALSE | TRUE | FALSE |
| OPTC | 0.1864871 | 0.83828322 | 1 | 0.186487117 | 0.186487117 | 3.700639411 | 0.22654427 | 0.845703919 | 0.00825 | 0.226544267 | 0.226544267 | 0.226544267 | 0.823181798 | -0.084504241 | FALSE | TRUE | FALSE |
| MND1 | 0.1863866 | 0.83829853 | 1 | 0.186386567 | 0.186386567 | 0.186386567 | 0.22656813 | 0.845689215 | 0.00505 | 0.226568126 | 0.226568126 | 0.226568126 | 0.822651318 | -0.084784202 | FALSE | TRUE | FALSE |
| AIDA | 0.1863629 | 0.83831385 | 1 | 0.186362871 | 0.186362871 | 0.186362871 | 0.2007286 | 0.851935346 | 1 | 0.2007286 | 0.2007286 | 0.2007286 | 0.928432073 | -0.032249865 | FALSE | TRUE | FALSE |
| PPIG | 0.1863131 | 0.83832915 | 1 | 1.714402398 | 1.714402398 | 33.82941156 | 1.96562728 | 0.491830106 | 0.87635 | 18.59798336 | 180.4834464 | 367.5750524 | 0.094785579 | -1.023257732 | FALSE | FALSE | FALSE |
| GPALPP1 | 0.1862964 | 0.83834446 | 1 | 0.186296423 | 0.186296423 | 0.186296423 | 0.22661553 | 0.845674508 | 1 | 0.226615529 | 0.226615529 | 0.226615529 | 0.822081452 | -0.08508515 | FALSE | TRUE | FALSE |
| TDRD6 | 0.1862841 | 0.83835976 | 0.6458 | 0.186284134 | 12.73164448 | 109.0055668 | 0.07982699 | 0.880391632 | 0.08875 | 0.009967234 | 0.655410029 | 1.548797925 | 2.333598454 | 0.368026128 | FALSE | TRUE | FALSE |
| EDEM1 | 0.1862228 | 0.83837506 | 0.51185 | 0.186222825 | 0.186222825 | 0.698773951 | 0.13239971 | 0.868365336 | 0.51345 | 0.132399709 | 0.132399709 | 0.654552191 | 1.4065199 | 0.148145881 | FALSE | TRUE | FALSE |
| PGM2L1 | 0.1859809 | 0.83839036 | 1 | 0.185980901 | 0.185980901 | 3.67767297 | 0.22672594 | 0.845645085 | 1 | 0.22672594 | 0.22672594 | 4.489466842 | 0.820289468 | -0.086032864 | FALSE | TRUE | FALSE |
| RNF39 | 0.185901 | 0.83840566 | 1 | 1.697839277 | 1.697839277 | 3.682683379 | 0.22595958 | 0.84585081 | 1 | 2.0729606 | 2.0729606 | 4.48135814 | 0.822717837 | -0.084749087 | FALSE | TRUE | FALSE |
| ZSCAN16 | 0.1858642 | 0.83842095 | 1 | 0.185864181 | 0.185864181 | 3.693822219 | 0.21931972 | 0.847348212 | 1 | 2.020784055 | 2.020784055 | 2.020784055 | 0.847457681 | -0.07188198 | FALSE | TRUE | FALSE |
| BCL2L2 | 0.1858131 | 0.83843624 | 1 | 1.757428944 | 1.757428944 | 1.757428944 | 0.20435865 | 0.850978889 | 0.00575 | 0.204358651 | 0.204358651 | 0.204358651 | 0.909249854 | -0.04131676 | FALSE | TRUE | FALSE |
| TBC1D20 | 0.1858049 | 0.83845153 | 0.0111 | 0.185804898 | 0.185804898 | 3.685258496 | 0.19726268 | 0.852772692 | 1 | 0.197262681 | 0.197262681 | 0.197262681 | 0.941916114 | -0.025987773 | FALSE | TRUE | FALSE |
| UBE4A | 0.1857993 | 0.83846681 | 1 | 16.3286091 | 1588.164349 | 1588.164349 | 0.25629139 | 0.837635084 | 0.025 | 0.256291389 | 0.256291389 | 5.035428559 | 0.724953414 | -0.139689901 | FALSE | TRUE | FALSE |
| PABPC1L | 0.1857192 | 0.8384821 | 1 | 0.742756752 | 0.742756752 | 6.289140862 | 0.2263881 | 0.845748015 | 1 | 0.905423375 | 0.905423375 | 7.670035079 | 0.820357548 | -0.085996821 | FALSE | TRUE | FALSE |
| COG2 | 0.1856755 | 0.83849738 | 1 | 1.714406158 | 1.714406158 | 33.79641383 | 0.23328846 | 0.843934858 | 1 | 0.23328846 | 0.23328846 | 4.606796877 | 0.795905073 | -0.099138727 | FALSE | TRUE | FALSE |
| SLITRK5 | 0.185634 | 0.83851265 | 1 | 16.70096213 | 1662.826198 | 1662.826198 | 0.21952155 | 0.847261741 | 0.96905 | 195.0924822 | 1972.363134 | 20427.05121 | 0.84562976 | -0.072819741 | FALSE | TRUE | FALSE |
| EIF2AK2 | 0.1855656 | 0.83852792 | 1 | 0.185565557 | 0.185565557 | 3.676135111 | 18.5867599 | 0.140706288 | 0.2586 | 1.961505155 | 18.58675989 | 18.58675989 | 0.00998375 | -2.000706318 | FALSE | FALSE | FALSE |
| NT5DC1 | 0.1855606 | 0.83854319 | 1 | 0.185560589 | 0.185560589 | 3.68145392 | 0.21873699 | 0.847477741 | 1 | 2.002353742 | 2.002353742 | 2.002353742 | 0.848327414 | -0.071436498 | FALSE | TRUE | FALSE |
| KRTAP5-3 | 0.1855365 | 0.83855846 | 1 | 0.185536486 | 0.185536486 | 0.185536486 | 0.22528668 | 0.846070634 | 1 | 0.225286677 | 0.225286677 | 0.225286677 | 0.823557291 | -0.084306184 | FALSE | TRUE | FALSE |
| SPTSSA | 0.1854932 | 0.83857373 | 1 | 0.185493189 | 0.185493189 | 0.185493189 | 0.19905197 | 0.852341651 | 0.00185 | 0.199051972 | 0.199051972 | 0.199051972 | 0.931883204 | -0.030638516 | FALSE | TRUE | FALSE |
| OR4N2 | 0.1854314 | 0.83858899 | 1 | 1.694027784 | 1.694027784 | 1.694027784 | 1572.10819 | 0.001555241 | 0.1111 | 159.4764957 | 15876.26305 | 164152.6505 | 0.000117951 | -3.928299215 | FALSE | FALSE | FALSE |
| RAB3IP | 0.1853996 | 0.83860425 | 1 | 1.697112609 | 1.697112609 | 3.677583377 | 0.23256247 | 0.844204996 | 1 | 0.232562471 | 0.232562471 | 4.617654432 | 0.797203545 | -0.098430779 | FALSE | TRUE | FALSE |
| SH3BP1 | 0.1853922 | 0.8386195 | 1 | 0.185392178 | 0.185392178 | 3.66356646 | 0.21878355 | 0.847448975 | 1 | 2.012914357 | 2.012914357 | 4.329630882 | 0.847377118 | -0.071923268 | FALSE | TRUE | FALSE |
| ZSCAN10 | 0.1853554 | 0.83863475 | 1 | 1.719209753 | 1.719209753 | 33.62144868 | 0.23311713 | 0.844025009 | 1 | 0.233117131 | 0.233117131 | 4.571100412 | 0.795117002 | -0.09956896 | FALSE | TRUE | FALSE |
| BRAF | 0.1853147 | 0.83865 | 1 | 0.185314715 | 0.185314715 | 3.657330253 | 0.22626609 | 0.845777398 | 1 | 0.226266091 | 0.226266091 | 4.472636702 | 0.819012312 | -0.08670957 | FALSE | TRUE | FALSE |
| GPATCH2L | 0.1852273 | 0.83866525 | 1 | 0.185227317 | 0.185227317 | 3.666995257 | 0.22546523 | 0.845968127 | 1 | 0.225465234 | 0.225465234 | 4.469038777 | 0.821533827 | -0.08537455 | FALSE | TRUE | FALSE |
| SF3B2 | 0.1851658 | 0.8386805 | 1 | 0.185165803 | 0.185165803 | 3.628045508 | 0.22340768 | 0.846522996 | 1 | 0.893526199 | 0.893526199 | 17.55015919 | 0.828824684 | -0.081537323 | FALSE | TRUE | FALSE |
| TRIM29 | 0.1851544 | 0.83869574 | 1 | 1.698842279 | 1.698842279 | 6.268870507 | 0.23241353 | 0.844309793 | 0.01585 | 0.232413525 | 0.232413525 | 4.585893062 | 0.796659224 | -0.098727411 | FALSE | TRUE | FALSE |
| SYN3 | 0.1851258 | 0.83871098 | 1 | 0.740444946 | 0.740444946 | 6.268130657 | 0.2280274 | 0.845379789 | 0.01525 | 0.228027396 | 0.228027396 | 4.509855699 | 0.811857751 | -0.090520058 | FALSE | TRUE | FALSE |
| SFTPB | 0.1851152 | 0.83872621 | 1 | 1.698360305 | 15.99239724 | 15.99239724 | 2.07532778 | 0.482702473 | 0.7504 | 2.075327779 | 19.61964774 | 19.61964774 | 0.089198048 | -1.049644648 | FALSE | FALSE | FALSE |
| OR7C1 | 0.1850725 | 0.83874144 | 1 | 1.696017157 | 1.696017157 | 1.696017157 | 2.07221392 | 0.483039746 | 0.5022 | 2.072213925 | 2.072213925 | 2.072213925 | 0.089311486 | -1.049092683 | FALSE | FALSE | FALSE |
| MS4A10 | 0.1850554 | 0.83875667 | 0.006 | 0.185055436 | 0.185055436 | 0.185055436 | 0.2248144 | 0.846173009 | 0.0052 | 0.224814404 | 0.224814404 | 0.224814404 | 0.823147592 | -0.084522288 | FALSE | TRUE | FALSE |
| KANSL1 | 0.1850185 | 0.8387719 | 1 | 0.185018545 | 0.185018545 | 3.611338108 | 0.21724782 | 0.847865036 | 1 | 0.217247815 | 0.217247815 | 4.252707931 | 0.851647436 | -0.069740157 | FALSE | TRUE | FALSE |
| ARHGAP31 | 0.1849867 | 0.83878713 | 1 | 0.184986681 | 0.184986681 | 6.221201751 | 0.21871763 | 0.84749212 | 1 | 2.035573739 | 2.035573739 | 39.72148696 | 0.845778546 | -0.072743336 | FALSE | TRUE | FALSE |
| CCL8 | 0.1849709 | 0.83880235 | 0.5039 | 0.184970892 | 0.184970892 | 0.184970892 | 0.02045942 | 0.893725873 | 0.7505 | 0.191675417 | 1.843002971 | 1.843002971 | 9.040867878 | 0.956210123 | FALSE | TRUE | FALSE |
| NUAK1 | 0.1849701 | 0.83881756 | 1 | 0.184970143 | 0.184970143 | 3.639564017 | 0.21888336 | 0.847420198 | 1 | 2.054046351 | 2.054046351 | 7.406145774 | 0.845062615 | -0.073111111 | FALSE | TRUE | FALSE |
| TBC1D23 | 0.1848838 | 0.83883278 | 1 | 15.92753066 | 153.6145724 | 153.6145724 | 0.24674433 | 0.840179279 | 0.015 | 0.246744328 | 0.246744328 | 4.882093827 | 0.749292874 | -0.125348398 | FALSE | TRUE | FALSE |
| JPH3 | 0.1848733 | 0.83884799 | 0.51995 | 0.739326956 | 0.739326956 | 14.45914296 | 0.00022898 | 0.899963603 | 1 | 0.00022898 | 0.00022898 | 0.004490151 | 807.3764868 | 2.907076097 | FALSE | TRUE | FALSE |
| OR2T10 | 0.1848508 | 0.8388632 | 1 | 0.184850835 | 0.184850835 | 0.184850835 | 0.2246298 | 0.846231448 | 1 | 0.224629801 | 0.224629801 | 0.224629801 | 0.822913228 | -0.084645956 | FALSE | TRUE | FALSE |
| MLX | 0.1848457 | 0.83887841 | 0.9965 | 1625.002863 | 177193.3747 | 1912299.338 | 1.14565004 | 0.572720238 | 0.506 | 1.145650036 | 1.145650036 | 1.145650036 | 0.16134566 | -0.792242712 | FALSE | FALSE | FALSE |
| TRPV6 | 0.1846911 | 0.83889361 | 0.93135 | 11.86438761 | 3063.099998 | 12286.00761 | 207.514201 | 0.012877601 | 0.25295 | 19.15560991 | 207.5142012 | 1233.1902 | 0.000890017 | -3.050601927 | FALSE | FALSE | FALSE |
| FLCN | 0.1846742 | 0.83890881 | 0.5835 | 0.434834397 | 41.47794167 | 165.6783699 | 3.7219398 | 0.369904726 | 0.66505 | 9.023881428 | 575.8397309 | 5717.29055 | 0.049617733 | -1.304363082 | FALSE | FALSE | FALSE |
| CHST12 | 0.1846654 | 0.83892401 | 1 | 0.184665426 | 0.184665426 | 3.635401616 | 0.22542311 | 0.846012076 | 1 | 0.225423112 | 0.225423112 | 4.446149993 | 0.819194733 | -0.086612849 | FALSE | TRUE | FALSE |
| RREB1 | 0.1846362 | 0.83893921 | 1 | 0.184636202 | 3.539758076 | 6.20782398 | 0.22537766 | 0.846041361 | 0.04655 | 0.225377663 | 0.225377663 | 7.587767458 | 0.819230263 | -0.086594013 | FALSE | TRUE | FALSE |
| PRR5L | 0.1846073 | 0.8389544 | 1 | 0.18460728 | 0.18460728 | 3.659157853 | 0.2254336 | 0.84599743 | 1 | 0.225433602 | 0.225433602 | 4.473190671 | 0.818898686 | -0.086769826 | FALSE | TRUE | FALSE |
| SPP2 | 0.184597 | 0.83896959 | 0.00615 | 0.184596966 | 0.184596966 | 0.184596966 | 0.2246234 | 0.846260651 | 1 | 0.224623404 | 0.224623404 | 0.224623404 | 0.821806465 | -0.085230446 | FALSE | TRUE | FALSE |
| GLI1 | 0.1845904 | 0.83898478 | 0.51275 | 0.184590414 | 0.184590414 | 3.604848096 | 0.03337242 | 0.890497035 | 0.028 | 0.033372419 | 0.033372419 | 0.653576323 | 5.531226599 | 0.742821451 | FALSE | TRUE | FALSE |
| DSN1 | 0.1845276 | 0.83899996 | 1 | 15.69255806 | 150.3740795 | 150.3740795 | 0.24580648 | 0.840415286 | 0.0079 | 0.245806476 | 0.245806476 | 0.245806476 | 0.750702599 | -0.124532081 | FALSE | TRUE | FALSE |
| UQCC2 | 0.1844836 | 0.83901514 | 1 | 0.184483592 | 0.184483592 | 0.184483592 | 0.20099472 | 0.85186742 | 1 | 0.20099472 | 0.20099472 | 0.20099472 | 0.917852924 | -0.037226904 | FALSE | TRUE | FALSE |
| TOR1AIP1 | 0.1844551 | 0.83903032 | 1 | 0.184455091 | 0.184455091 | 3.642664653 | 0.22456082 | 0.846289842 | 0.01585 | 0.224560816 | 0.224560816 | 4.44139931 | 0.821403726 | -0.085443332 | FALSE | TRUE | FALSE |
| CDKAL1 | 0.1844348 | 0.83904549 | 0.01445 | 0.184434812 | 0.184434812 | 3.649898218 | 0.22480728 | 0.846187623 | 1 | 0.22480728 | 0.22480728 | 4.454483837 | 0.820412988 | -0.085967473 | FALSE | TRUE | FALSE |
| CCDC47 | 0.1843914 | 0.83906067 | 1 | 0.184391409 | 0.184391409 | 3.655405442 | 0.22465064 | 0.846216843 | 0.01065 | 0.224650642 | 0.224650642 | 4.458220233 | 0.820791818 | -0.085766981 | FALSE | TRUE | FALSE |
| CDK13 | 0.1843181 | 0.83907583 | 1 | 0.18431808 | 0.18431808 | 6.191472732 | 0.22504611 | 0.846099897 | 1 | 0.22504611 | 0.22504611 | 7.570512754 | 0.819023621 | -0.086703573 | FALSE | TRUE | FALSE |
| ELSPBP1 | 0.1842035 | 0.839091 | 1 | 0.184203494 | 0.184203494 | 0.184203494 | 0.22402877 | 0.846377353 | 0.0057 | 0.224028768 | 0.224028768 | 0.224028768 | 0.822231433 | -0.085005925 | FALSE | TRUE | FALSE |
| GPR55 | 0.1841394 | 0.83910617 | 1 | 0.184139441 | 0.184139441 | 0.184139441 | 0.22431745 | 0.846319022 | 1 | 0.224317452 | 0.224317452 | 0.224317452 | 0.820887715 | -0.085716244 | FALSE | TRUE | FALSE |
| BCKDHB | 0.1839199 | 0.83912133 | 0.2583 | 0.046023047 | 0.183919878 | 0.183919878 | 0.14905383 | 0.864815583 | 0.6913 | 0.334764293 | 5.351317997 | 5.351317997 | 1.233915789 | 0.091285521 | FALSE | TRUE | FALSE |
| SLC35E3 | 0.1838923 | 0.83913649 | 0.0071 | 0.183892344 | 0.183892344 | 0.183892344 | 0.22410948 | 0.846362775 | 0.0072 | 0.22410948 | 0.22410948 | 0.22410948 | 0.820546924 | -0.085896578 | FALSE | TRUE | FALSE |
| WDR20 | 0.183759 | 0.83915165 | 1 | 0.183758987 | 0.183758987 | 0.183758987 | 6.72760596 | 0.251250952 | 0.68275 | 17.81434835 | 1773.192459 | 1773.192459 | 0.027314172 | -1.563611958 | FALSE | FALSE | FALSE |
| IMPG2 | 0.1836818 | 0.8391668 | 0.93675 | 1.646419912 | 142.6609563 | 142.6609563 | 0.03637246 | 0.889819013 | 1 | 0.145477571 | 0.145477571 | 2.857030679 | 5.050023952 | 0.703293438 | FALSE | TRUE | FALSE |
| CXCL13 | 0.1835699 | 0.83918195 | 0.00305 | 0.183569915 | 0.183569915 | 0.183569915 | 0.18856207 | 0.854774884 | 1 | 0.188562074 | 0.188562074 | 0.188562074 | 0.973525115 | -0.01165284 | FALSE | TRUE | FALSE |
| ARID1A | 0.183477 | 0.8391971 | 1 | 16.65512674 | 164.8821849 | 556.7246808 | 0.2387506 | 0.84240159 | 1 | 2.252073337 | 4.607752057 | 43.46374587 | 0.768488026 | -0.114362895 | FALSE | TRUE | FALSE |
| CPSF4 | 0.1833024 | 0.83921225 | 1 | 0.1833024 | 0.1833024 | 0.1833024 | 0.22302108 | 0.846610253 | 0.0086 | 0.223021081 | 0.223021081 | 0.223021081 | 0.821906162 | -0.085177764 | FALSE | TRUE | FALSE |
| HINT3 | 0.1832614 | 0.83922739 | 1 | 0.183261421 | 1.708400189 | 1.708400189 | 0.23086967 | 0.844638255 | 1 | 0.230869673 | 0.230869673 | 0.230869673 | 0.793787328 | -0.100295838 | FALSE | TRUE | FALSE |
| NHP2 | 0.1831584 | 0.83924254 | 1 | 0.183158435 | 0.183158435 | 0.183158435 | 0.22271789 | 0.846682896 | 1 | 0.222717895 | 0.222717895 | 0.222717895 | 0.822378618 | -0.08492819 | FALSE | TRUE | FALSE |
| SLCO4C1 | 0.1831266 | 0.83925768 | 0.51295 | 0.183126553 | 0.183126553 | 0.695061553 | 0.28944137 | 0.828307178 | 0.5062 | 0.289441375 | 0.289441375 | 0.637743428 | 0.632689617 | -0.198809292 | FALSE | TRUE | FALSE |
| CPN2 | 0.1831253 | 0.83927281 | 1 | 1.686004587 | 15.9317599 | 33.32972742 | 0.23752948 | 0.842722816 | 1 | 0.237529484 | 0.237529484 | 4.702133993 | 0.770958048 | -0.112969254 | FALSE | TRUE | FALSE |
| KCNJ11 | 0.1830988 | 0.83928795 | 0.50545 | 0.183098802 | 0.183098802 | 0.183098802 | 0.06268417 | 0.884110187 | 1 | 0.589897768 | 5.697448004 | 5.697448004 | 2.920973678 | 0.465527643 | FALSE | TRUE | FALSE |
| ASPA | 0.1830516 | 0.83930308 | 0.50835 | 0.183051634 | 0.183051634 | 0.183051634 | 0.26344666 | 0.835005291 | 0.5012 | 0.263446662 | 0.263446662 | 0.263446662 | 0.694833759 | -0.158119089 | FALSE | TRUE | FALSE |
| REG1A | 0.1829669 | 0.8393182 | 1 | 0.182966867 | 0.182966867 | 0.182966867 | 0.19593292 | 0.853067629 | 1 | 0.195932924 | 0.195932924 | 0.195932924 | 0.933824001 | -0.029734968 | FALSE | TRUE | FALSE |
| ARHGAP17 | 0.1826971 | 0.83933333 | 1 | 0.182697145 | 0.182697145 | 3.588572154 | 0.22303191 | 0.846595717 | 1 | 0.223031906 | 0.223031906 | 4.390271842 | 0.81915251 | -0.086635234 | FALSE | TRUE | FALSE |
| SESN2 | 0.1825256 | 0.83934845 | 1 | 0.182525636 | 0.182525636 | 3.608988227 | 0.22234202 | 0.846784483 | 1 | 0.222342023 | 0.222342023 | 4.402274074 | 0.820922802 | -0.085697681 | FALSE | TRUE | FALSE |

| | | | | | | | | | | | | | | | | | |
|---|---|---|---|---|---|---|---|---|---|---|---|---|---|---|---|---|---|
| ARHGAP30 | 0.1823173 | 0.83936358 | 1 | 0.182317271 | 0.182317271 | 3.56493055 | 0.22259491 | 0.846697416 | 1 | 0.222594906 | 0.222594906 | 4.36420429 | 0.819054101 | -0.086687411 | FALSE | TRUE | FALSE |
| ZNF155 | 0.1822095 | 0.8393787 | 0.7578 | 0.182209461 | 1.641717308 | 3.61086067 | 0.29558491 | 0.826545768 | 0.50615 | 0.295584909 | 0.295584909 | 0.661558104 | 0.616436955 | -0.210111334 | FALSE | TRUE | FALSE |
| PPIP5K1 | 0.1821744 | 0.83939381 | 1 | 0.182174445 | 0.182174445 | 6.116362001 | 0.22247517 | 0.846755472 | 1 | 0.222475171 | 0.222475171 | 4.334596993 | 0.818852926 | -0.086794095 | FALSE | TRUE | FALSE |
| LEMD1 | 0.1820154 | 0.83940893 | 0.0032 | 0.182015372 | 0.182015372 | 0.182015372 | 1.96361377 | 0.492156595 | 0.4967 | 0.214946769 | 1.963613767 | 1.963613767 | 0.09269408 | -1.032948 | FALSE | FALSE | FALSE |
| TRIP12 | 0.18184 | 0.83942404 | 1 | 1.706292958 | 16.43252218 | 316.7163245 | 0.23682514 | 0.842890563 | 0.04425 | 0.236825136 | 0.236825136 | 7.938571989 | 0.767823939 | -0.114738352 | FALSE | TRUE | FALSE |
| SLC43A3 | 0.1817765 | 0.83943915 | 0.50245 | 0.410399505 | 1.641039068 | 1.641039068 | 0.06789272 | 0.882905892 | 1 | 0.067892725 | 0.067892725 | 1.345659351 | 2.677407774 | 0.42771452 | FALSE | TRUE | FALSE |
| DNAJC18 | 0.1817509 | 0.83945426 | 1 | 0.181750916 | 0.181750916 | 3.609722371 | 0.22180251 | 0.846842477 | 1 | 0.221802511 | 0.221802511 | 0.221802511 | 0.819426772 | -0.086489851 | FALSE | TRUE | FALSE |
| OR1I1 | 0.1817445 | 0.83946936 | 0.00925 | 0.181744487 | 0.181744487 | 0.181744487 | 1.98401445 | 0.490027434 | 0.5096 | 1.984014445 | 1.984014445 | 1.984014445 | 0.091604417 | -1.038083583 | FALSE | FALSE | FALSE |
| OTP | 0.1817378 | 0.83948446 | 1 | 0.181737842 | 0.181737842 | 0.181737842 | 0.22249114 | 0.846711935 | 0.0081 | 0.222491139 | 0.222491139 | 0.222491139 | 0.816831819 | -0.087867353 | FALSE | TRUE | FALSE |
| PLCXD2 | 0.1816893 | 0.83949956 | 1 | 0.181689344 | 0.181689344 | 0.181689344 | 0.2215968 | 0.846885947 | 1 | 0.221596804 | 0.221596804 | 0.221596804 | 0.819909587 | -0.086234035 | FALSE | TRUE | FALSE |
| NR2F6 | 0.1814392 | 0.83951466 | 1 | 2.901320407 | 11.60621947 | 11.60621947 | 0.88807515 | 0.600049575 | 0.87405 | 3.5518754 | 14.20935164 | 14.20935164 | 0.204306147 | -0.689718567 | FALSE | TRUE | FALSE |
| OR6K3 | 0.1814017 | 0.83952975 | 0.00735 | 0.181401711 | 0.181401711 | 0.181401711 | 1.95522721 | 0.492971118 | 0.5061 | 1.95522721 | 1.95522721 | 1.95522721 | 0.092777816 | -1.032555853 | FALSE | FALSE | FALSE |
| RAB11FIP2 | 0.1813174 | 0.83954484 | 1 | 0.181317447 | 0.181317447 | 3.594279058 | 0.22139726 | 0.846914914 | 0.01085 | 0.221397257 | 0.221397257 | 4.393454363 | 0.818968804 | -0.086732641 | FALSE | TRUE | FALSE |
| GLYR1 | 0.1812719 | 0.83955993 | 1 | 0.181271946 | 0.181271946 | 3.579023017 | 39.6572799 | 0.068743755 | 0.00195 | 2.005489935 | 2.005489935 | 4.249567813 | 0.004570963 | -2.339992325 | FALSE | FALSE | FALSE |
| G2E3 | 0.18099 | 0.83957502 | 1 | 0.180990007 | 0.180990007 | 3.57829204 | 0.22126316 | 0.846958346 | 1 | 0.221263158 | 0.221263158 | 4.380553899 | 0.817985285 | -0.087254509 | FALSE | TRUE | FALSE |
| ADAT3 | 0.1809821 | 0.8395901 | 0.0092 | 0.180982083 | 0.180982083 | 0.180982083 | 0.21812566 | 0.847664459 | 1 | 0.872454981 | 0.872454981 | 0.872454981 | 0.829714779 | -0.081071174 | FALSE | TRUE | FALSE |
| CYP21A2 | 0.1809455 | 0.83960518 | 1 | 1.669137086 | 15.80265524 | 15.80265524 | 1.98150714 | 0.49035614 | 0.87245 | 18.80607217 | 183.0654202 | 183.0654202 | 0.091317096 | -1.039447906 | FALSE | FALSE | FALSE |
| HCLS1 | 0.1807366 | 0.83962026 | 1 | 1.650875431 | 1.650875431 | 3.577830901 | 0.21380255 | 0.848619819 | 1 | 1.959901259 | 18.43953781 | 38.84551554 | 0.845343507 | -0.072966779 | FALSE | TRUE | FALSE |
| ERLIN2 | 0.1807312 | 0.83963534 | 1 | 0.722729772 | 0.722729772 | 0.722729772 | 0.88401167 | 0.600788059 | 0.5178 | 0.884011669 | 0.884011669 | 0.884011669 | 0.204444305 | -0.689424982 | FALSE | TRUE | FALSE |
| TOPORS | 0.1806698 | 0.83965041 | 1 | 0.180669767 | 0.180669767 | 3.538162644 | 0.21417122 | 0.848534728 | 0.50925 | 1.988425976 | 1.988425976 | 39.03817178 | 0.843576316 | -0.073875622 | FALSE | TRUE | FALSE |
| SLC16A9 | 0.1806005 | 0.83966548 | 1 | 1.675908265 | 15.96146466 | 15.96146466 | 0.22085067 | 0.84701622 | 1 | 2.054924694 | 19.6237565 | 19.6237565 | 0.817749433 | -0.087379748 | FALSE | TRUE | FALSE |
| SLC35A4 | 0.1805105 | 0.83968055 | 1 | 1.643850788 | 1.643850788 | 1.66323499 | 170.477612 | 0.015726457 | 0.00375 | 17.72910235 | 170.4776122 | 170.4776122 | 0.001058851 | -2.97516499 | FALSE | FALSE | FALSE |
| TM4SF20 | 0.1804075 | 0.83969562 | 1 | 1.642979966 | 1.642979966 | 1.642979966 | 0.22672775 | 0.84563037 | 1 | 0.226727748 | 0.226727748 | 0.226727748 | 0.795701235 | -0.099249968 | FALSE | TRUE | FALSE |
| INSL3 | 0.1803719 | 0.83971068 | 1 | 0.180371933 | 1.642303248 | 1.642303248 | 0.21984297 | 0.847146291 | 1 | 2.011363878 | 2.011363878 | 2.011363878 | 0.820458031 | -0.08594363 | FALSE | TRUE | FALSE |
| HSPBP1 | 0.1803467 | 0.83972574 | 1 | 0.18034671 | 0.18034671 | 3.571834031 | 0.2200234 | 0.847131847 | 0.0114 | 0.220023403 | 0.220023403 | 4.362746536 | 0.819670578 | -0.086360654 | FALSE | TRUE | FALSE |
| TDRD5 | 0.1802821 | 0.8397408 | 1 | 0.180282106 | 0.180282106 | 3.542529432 | 0.21329515 | 0.848832128 | 1 | 0.213295146 | 0.213295146 | 4.200065829 | 0.845223669 | -0.07302835 | FALSE | TRUE | FALSE |
| GSTA4 | 0.1802279 | 0.83975585 | 1 | 0.180227948 | 0.180227948 | 0.180227948 | 0.19458802 | 0.853308126 | 0.0061 | 0.19458802 | 0.19458802 | 0.19458802 | 0.926202696 | -0.033293959 | FALSE | TRUE | FALSE |
| ERCC6L2 | 0.1801976 | 0.8397709 | 0.93815 | 15.33570514 | 1437.734819 | 4959.241573 | 0.00544804 | 0.898003586 | 1 | 0.047846246 | 0.431285314 | 0.946404837 | 33.07569234 | 1.519508944 | FALSE | TRUE | FALSE |
| MC2R | 0.180162 | 0.83978595 | 1 | 0.180162038 | 0.180162038 | 0.180162038 | 0.21365705 | 0.848676494 | 1 | 1.988984125 | 1.988984125 | 1.988984125 | 0.84323001 | -0.074053946 | FALSE | TRUE | FALSE |
| RGS2 | 0.1800854 | 0.839801 | 0.0066 | 0.180085412 | 0.180085412 | 0.180085412 | 0.21973615 | 0.847160732 | 1 | 0.21973615 | 0.21973615 | 0.21973615 | 0.819552959 | -0.086422977 | FALSE | TRUE | FALSE |
| PMVK | 0.1799984 | 0.83981604 | 1 | 0.179998355 | 0.179998355 | 0.179998355 | 1.8658868 | 0.501764573 | 0.00655 | 1.865886804 | 1.865886804 | 1.865886804 | 0.096467993 | -1.015616757 | FALSE | FALSE | FALSE |
| MVB12A | 0.1799891 | 0.83983108 | 1 | 0.17998912 | 0.17998912 | 0.17998912 | 0.21959717 | 0.847218469 | 1 | 0.219597166 | 0.219597166 | 0.219597166 | 0.819633161 | -0.086380479 | FALSE | TRUE | FALSE |
| ADPGK | 0.179886 | 0.83984612 | 1 | 0.179886048 | 0.179886048 | 3.570751749 | 1.95895517 | 0.492482729 | 0.5005 | 1.958955173 | 1.958955173 | 1.958955173 | 0.091827547 | -1.037027018 | FALSE | FALSE | FALSE |
| CLEC12B | 0.1798735 | 0.83986116 | 0.50115 | 1.634326057 | 1.634326057 | 1.634326057 | 0.21923953 | 0.847377014 | 0.50245 | 2.002103187 | 2.002103187 | 2.002103187 | 0.820442833 | -0.085951674 | FALSE | TRUE | FALSE |
| RPP30 | 0.1798498 | 0.83987619 | 0.0072 | 0.179849753 | 0.179849753 | 0.179849753 | 0.21943359 | 0.8473194 | 1 | 0.219433594 | 0.219433594 | 0.219433594 | 0.819609021 | -0.08639327 | FALSE | TRUE | FALSE |
| INSL5 | 0.1798036 | 0.83989122 | 1 | 0.17980363 | 0.17980363 | 0.17980363 | 0.21966202 | 0.84717517 | 1 | 0.219662019 | 0.219662019 | 0.219662019 | 0.818546743 | -0.086956515 | FALSE | TRUE | FALSE |
| OBP2A | 0.1797655 | 0.83990625 | 0.5054 | 1.630608848 | 1.630608848 | 1.630608848 | 0.22579762 | 0.845894824 | 1 | 0.225797619 | 0.225797619 | 0.225797619 | 0.796135367 | -0.099013083 | FALSE | TRUE | FALSE |
| MYL7 | 0.1797526 | 0.83992127 | 1 | 0.179752588 | 0.179752588 | 0.179752588 | 0.21762173 | 0.847807781 | 1 | 0.217621733 | 0.87022507 | 0.87022507 | 0.825986383 | -0.083027112 | FALSE | TRUE | FALSE |
| CAMK2N1 | 0.1797454 | 0.83993629 | 1 | 0.179745447 | 0.179745447 | 0.179745447 | 0.18174592 | 0.856623048 | 1 | 0.181745924 | 0.181745924 | 0.181745924 | 0.988993005 | -0.00480678 | FALSE | TRUE | FALSE |
| FUT3 | 0.1797358 | 0.83995131 | 1 | 0.179735833 | 0.179735833 | 3.557054975 | 0.21942984 | 0.847333807 | 1 | 0.219429837 | 0.219429837 | 4.348090761 | 0.81910389 | -0.086661011 | FALSE | TRUE | FALSE |
| OR13F1 | 0.1796625 | 0.83996633 | 1 | 0.17966248 | 0.17966248 | 0.17966248 | 1.9392993 | 0.494592188 | 0.5024 | 1.939299299 | 1.939299299 | 1.939299299 | 0.092642987 | -1.033187451 | FALSE | FALSE | FALSE |
| SLC30A6 | 0.1796372 | 0.83998134 | 0.01245 | 0.179637159 | 0.179637159 | 3.563743366 | 0.21959736 | 0.847204039 | 1 | 0.21959736 | 0.21959736 | 4.360725321 | 0.818029686 | -0.087230935 | FALSE | TRUE | FALSE |
| LILRB5 | 0.1794936 | 0.83999635 | 0.8753 | 1.584327948 | 14.34417525 | 14.34417525 | 0.03521306 | 0.890077084 | 1 | 0.035213057 | 0.035213057 | 0.696231414 | 5.097357923 | 0.70734513 | FALSE | TRUE | FALSE |
| UBAC2 | 0.1793683 | 0.84001136 | 1 | 0.179368262 | 0.179368262 | 3.563460066 | 1.94353366 | 0.494430499 | 0.5058 | 1.943533662 | 1.943533662 | 1.943533662 | 0.092289763 | -1.034846468 | FALSE | FALSE | FALSE |
| ARPC1B | 0.1793682 | 0.84002636 | 1 | 15.71588559 | 152.8584575 | 152.8584575 | 0.24073603 | 0.841817042 | 0.0102 | 0.240736031 | 0.240736031 | 4.777275675 | 0.745082537 | -0.127795615 | FALSE | TRUE | FALSE |
| KIAA1586 | 0.1793508 | 0.84004136 | 0.8808 | 1.581664991 | 14.30738281 | 14.30738281 | 18.3906099 | 0.141641163 | 0.9465 | 1636.978619 | 1638868.372 | 17145757.47 | 0.009752304 | -2.010892768 | FALSE | FALSE | FALSE |
| AK5 | 0.1789808 | 0.84005637 | 1 | 0.178980789 | 0.178980789 | 3.540229911 | 0.21676328 | 0.848007998 | 0.5024 | 0.866878459 | 0.866878459 | 4.293240887 | 0.82569698 | -0.083179304 | FALSE | TRUE | FALSE |
| SAFB2 | 0.1789369 | 0.84007136 | 1 | 1.671829556 | 6.686547015 | 56.4147541 | 2.07527713 | 0.482871151 | 0.03155 | 2.075277132 | 2.075277132 | 40.46357764 | 0.086223119 | -1.064376273 | FALSE | FALSE | FALSE |
| TRIM33 | 0.1789225 | 0.84008636 | 1 | 0.178922513 | 0.178922513 | 3.516029381 | 0.21896607 | 0.847405806 | 0.0223 | 0.218966066 | 0.218966066 | 4.311967197 | 0.81712439 | -0.087711826 | FALSE | TRUE | FALSE |
| SEC16B | 0.178869 | 0.84010135 | 0.93775 | 1.600829595 | 138.2871844 | 138.2871844 | 0.29009278 | 0.828107181 | 0.5082 | 2.623438571 | 24.33476605 | 51.57944047 | 0.616592538 | -0.210001735 | FALSE | TRUE | FALSE |
| GNL3 | 0.1788289 | 0.84011634 | 0.5079 | 0.178828932 | 0.178828932 | 0.411783064 | 0.03217268 | 0.890752253 | 1 | 0.279972856 | 0.279972856 | 0.63834118 | 5.558410293 | 0.744950601 | FALSE | TRUE | FALSE |
| PRR12 | 0.1787835 | 0.84013133 | 1 | 173971.0328 | 20838380.27 | 404684558.2 | 0.30851967 | 0.8224672 | 1 | 2.917476589 | 2.917476589 | 56.48513503 | 0.57948813 | -0.236955455 | FALSE | TRUE | FALSE |
| HNRNPA1L2 | 0.1787797 | 0.84014631 | 0.50675 | 1.664742748 | 1.664742748 | 1.664742748 | 0.22589779 | 0.845865484 | 1 | 0.225897792 | 0.225897792 | 0.225897792 | 0.791418395 | -0.10159386 | FALSE | TRUE | FALSE |
| RGS12 | 0.1787482 | 0.8401613 | 0.98615 | 15.06362385 | 1401.663386 | 14011.89473 | 0.31905406 | 0.819040934 | 0.28275 | 0.319054063 | 2.925844023 | 10.78556039 | 0.56024432 | -0.251622537 | FALSE | TRUE | FALSE |
| RDM1 | 0.1787384 | 0.84017627 | 1 | 0.178738402 | 0.178738402 | 0.178738402 | 0.21803062 | 0.847693145 | 1 | 0.218030619 | 0.218030619 | 0.218030619 | 0.819785785 | -0.086299617 | FALSE | TRUE | FALSE |
| PTPRN | 0.1786444 | 0.84019125 | 0.93955 | 1.61344022 | 25.82165674 | 31.57733484 | 0.25944847 | 0.83691686 | 0.8728 | 1.03783118 | 9.427063557 | 20.36426988 | 0.688554585 | -0.162061625 | FALSE | TRUE | FALSE |
| OR5H15 | 0.178536 | 0.84020622 | 1 | 0.178536035 | 0.178536035 | 0.178536035 | 0.21782612 | 0.847750485 | 1 | 0.217826124 | 0.217826124 | 0.217826124 | 0.819626369 | -0.086384078 | FALSE | TRUE | FALSE |
| SPIRE1 | 0.1783829 | 0.84022119 | 1 | 0.178382922 | 0.178382922 | 3.523005604 | 0.21184585 | 0.849114317 | 1 | 1.972787723 | 1.972787723 | 7.166913478 | 0.84204116 | -0.074666679 | FALSE | TRUE | FALSE |
| SAP18 | 0.1783643 | 0.84023616 | 0.9996 | 149281.2045 | 154942133.2 | 1743303342 | 0.13825627 | 0.867094704 | 1 | 2.95243E+13 | 7.91304E+17 | 1.4454E+20 | 1.290099363 | 0.110623161 | FALSE | TRUE | FALSE |
| CCDC107 | 0.1783277 | 0.84025113 | 1 | 0.178327681 | 0.178327681 | 0.178327681 | 1.92585183 | 0.496203259 | 0.50815 | 1.925851826 | 1.925851826 | 1.925851826 | 0.092596782 | -1.033404106 | FALSE | FALSE | FALSE |
| TST | 0.1783143 | 0.84026609 | 1 | 0.178314301 | 0.178314301 | 3.535230133 | 0.21818885 | 0.847650113 | 1 | 0.218188846 | 0.218188846 | 0.218188846 | 0.817247555 | -0.08764637 | FALSE | TRUE | FALSE |
| CHSY3 | 0.17826 | 0.84028105 | 1 | 0.178260028 | 0.178260028 | 3.512825539 | 0.20938069 | 0.84988506 | 1 | 1.952053967 | 7.806964683 | 28.31314534 | 0.85136806 | -0.069882647 | FALSE | TRUE | FALSE |
| GNB1L | 0.1781779 | 0.84029601 | 1 | 0.17817792 | 0.17817792 | 3.526180938 | 0.21786134 | 0.847736154 | 1 | 0.217861342 | 0.217861342 | 0.217861342 | 0.817850099 | -0.08732629 | FALSE | TRUE | FALSE |
| CRYBB2 | 0.1781319 | 0.84031096 | 1 | 0.178131896 | 0.178131896 | 0.178131896 | 0.21829414 | 0.847607056 | 1 | 0.218294144 | 0.218294144 | 0.218294144 | 0.816017748 | -0.088300396 | FALSE | TRUE | FALSE |
| BSX | 0.1781192 | 0.84032592 | 1 | 0.178119161 | 0.178119161 | 0.178119161 | 0.21848042 | 0.84754961 | 1 | 0.218480418 | 0.218480418 | 0.218480418 | 0.815263734 | -0.088701876 | FALSE | TRUE | FALSE |
| EN2 | 0.1780843 | 0.84034087 | 1 | 0.178084256 | 0.178084256 | 0.178084256 | 0.21849091 | 0.847535242 | 0.0051 | 0.218490905 | 0.218490905 | 0.218490905 | 0.815064848 | -0.088807837 | FALSE | TRUE | FALSE |
| UTP6 | 0.1780706 | 0.84035581 | 0.0163 | 0.178070617 | 0.178070617 | 3.52236525 | 0.21769221 | 0.847793461 | 0.01425 | 0.217692211 | 0.217692211 | 4.311788694 | 0.817992597 | -0.087250627 | FALSE | TRUE | FALSE |
| PIGC | 0.1779718 | 0.84037076 | 0.5001 | 1.620758017 | 1.620758017 | 1.620758017 | 67.4718804 | 0.042338809 | 0.0009 | 1.988532605 | 1.988532605 | 1.988532605 | 0.002637717 | -2.578771734 | FALSE | FALSE | FALSE |
| JMJD6 | 0.1779294 | 0.8403857 | 1 | 0.177929398 | 0.177929398 | 3.522291275 | 0.21827311 | 0.847621411 | 0.01225 | 0.218273109 | 0.218273109 | 4.326237158 | 0.815168658 | -0.088752526 | FALSE | TRUE | FALSE |
| TRHDE | 0.1778945 | 0.84040063 | 1 | 0.177894481 | 0.177894481 | 3.48617252 | 0.21777885 | 0.847764813 | 1 | 0.217778846 | 0.217778846 | 4.278141385 | 0.816858406 | -0.087853218 | FALSE | TRUE | FALSE |
| VPS36 | 0.1778277 | 0.84041557 | 1 | 0.17782767 | 0.17782767 | 3.530126505 | 0.21750366 | 0.847850725 | 0.00955 | 0.217503663 | 0.217503663 | 0.217503663 | 0.817584716 | -0.087467236 | FALSE | TRUE | FALSE |
| ZSCAN4 | 0.1778236 | 0.8404305 | 1 | 1.620748108 | 1.620748108 | 3.530692692 | 0.2171996 | 0.847879344 | 1 | 1.988978006 | 1.988978006 | 1.988978006 | 0.818710571 | -0.086869602 | FALSE | TRUE | FALSE |
| TBX21 | 0.177702 | 0.84044543 | 1 | 0.177701999 | 0.177701999 | 3.516198141 | 0.21809685 | 0.847678803 | 0.012 | 0.218096852 | 0.218096852 | 4.321020717 | 0.814784796 | -0.088957084 | FALSE | TRUE | FALSE |

| | | | | | | | | | | | | | | | | | |
|---|---|---|---|---|---|---|---|---|---|---|---|---|---|---|---|---|---|
| OR13C4 | 0.1775834 | 0.84046036 | 1 | 0.177583379 | 0.177583379 | 0.177583379 | 0.21037861 | 0.849591705 | 1 | 1.923440125 | 1.923440125 | 1.923440125 | 0.84411329 | -0.073599262 | FALSE | TRUE | FALSE |
| DLD | 0.1775472 | 0.84047529 | 0.2567 | 0.177547209 | 0.710003485 | 0.880420419 | 0.06544048 | 0.883463211 | 0.0102 | 0.065440482 | 0.065440482 | 1.298725832 | 2.713109744 | 0.433467361 | FALSE | TRUE | FALSE |
| DCAKD | 0.1774869 | 0.84049021 | 1 | 0.177486926 | 0.177486926 | 0.177486926 | 1.94480085 | 0.494268755 | 0.50795 | 1.944800853 | 1.944800853 | 1.944800853 | 0.091262262 | -1.03970877 | FALSE | FALSE | FALSE |
| TOR1B | 0.1773405 | 0.84050513 | 0.0111 | 0.177340484 | 0.177340484 | 3.516303293 | 0.2168055 | 0.847965137 | 1 | 0.216805504 | 0.216805504 | 4.303261722 | 0.817970398 | -0.087262413 | FALSE | TRUE | FALSE |
| HTRA3 | 0.1773272 | 0.84052005 | 0.01595 | 0.044362131 | 0.177327168 | 0.87653695 | 0.29116274 | 0.82772404 | 0.81465 | 1.164260626 | 4.656645009 | 18.629643 | 0.609031117 | -0.215360517 | FALSE | TRUE | FALSE |
| OR5AS1 | 0.1773089 | 0.84053496 | 1 | 1.613336429 | 1.613336429 | 1.613336429 | 0.22326233 | 0.846566636 | 1 | 0.223262332 | 0.223262332 | 0.223262332 | 0.794173002 | -0.100084881 | FALSE | TRUE | FALSE |
| PPIF | 0.1772725 | 0.84054987 | 1 | 0.177272503 | 0.177272503 | 0.177272503 | 0.21061277 | 0.849451611 | 1 | 1.960522856 | 1.960522856 | 1.960522856 | 0.84169876 | -0.074843313 | FALSE | TRUE | FALSE |
| RORB | 0.177176 | 0.84056478 | 1 | 0.177176045 | 0.177176045 | 3.511203744 | 0.21757484 | 0.847836413 | 1 | 0.217574837 | 0.217574837 | 4.316540517 | 0.814322319 | -0.089203662 | FALSE | TRUE | FALSE |
| PLAA | 0.1770448 | 0.84057969 | 1 | 0.177044833 | 0.177044833 | 3.491578699 | 0.2170756 | 0.847922252 | 1 | 0.217075597 | 0.217075597 | 4.288218768 | 0.815590677 | -0.088527747 | FALSE | TRUE | FALSE |
| MYRF | 0.1770421 | 0.84059459 | 1 | 15.49831808 | 602.686863 | 1211.562572 | 0.23333504 | 0.843904784 | 1 | 2.165804096 | 2.165804096 | 42.44465148 | 0.758746161 | -0.119903493 | FALSE | TRUE | FALSE |
| DLGAP2 | 0.1769769 | 0.84060949 | 1 | 0.17697691 | 0.17697691 | 3.436612545 | 0.21053311 | 0.849521692 | 1 | 1.985852955 | 1.985852955 | 38.69732857 | 0.840613185 | -0.075403802 | FALSE | TRUE | FALSE |
| TBCK | 0.1769104 | 0.84062439 | 0.74915 | 0.391147942 | 1.564501041 | 7.711508438 | 0.5701484 | 0.724752806 | 0.5099 | 0.570148397 | 0.570148397 | 2.162509271 | 0.31028838 | -0.508234488 | FALSE | TRUE | FALSE |
| OSBPL9 | 0.1769073 | 0.84063929 | 1 | 0.176907305 | 0.176907305 | 3.491765658 | 0.21029002 | 0.849619693 | 1 | 1.958235143 | 1.958235143 | 7.114639354 | 0.841253932 | -0.075072893 | FALSE | TRUE | FALSE |
| PSMD6 | 0.1768964 | 0.84065418 | 1 | 0.176896403 | 0.176896403 | 3.507354667 | 0.21687233 | 0.847950845 | 0.01005 | 0.216872327 | 0.216872327 | 4.304433071 | 0.815670699 | -0.088485138 | FALSE | TRUE | FALSE |
| SGK1 | 0.1768831 | 0.84066907 | 0.0126 | 0.176883144 | 0.176883144 | 3.50082702 | 0.21012837 | 0.849689615 | 1 | 1.946545638 | 1.946545638 | 4.164017462 | 0.841786107 | -0.074798246 | FALSE | TRUE | FALSE |
| DDB2 | 0.1768675 | 0.84068396 | 0.0131 | 0.176867514 | 0.176867514 | 3.503914341 | 0.21651837 | 0.848107911 | 1 | 0.216518366 | 0.216518366 | 4.294312359 | 0.816870723 | -0.087846669 | FALSE | TRUE | FALSE |
| ADAM29 | 0.1767515 | 0.84069884 | 0.97075 | 14.29227969 | 133.4281946 | 1276.133084 | 2.6494051 | 0.437049546 | 0.511 | 2.649405098 | 24.42249012 | 24.42249012 | 0.066713658 | -1.175785245 | FALSE | FALSE | FALSE |
| IFNE | 0.1767309 | 0.84071373 | 1 | 0.176730921 | 0.176730921 | 0.176730921 | 0.21572708 | 0.848236189 | 1 | 0.215727078 | 0.215727078 | 0.215727078 | 0.819233834 | -0.08659212 | FALSE | TRUE | FALSE |
| GPATCH8 | 0.1766882 | 0.84072861 | 1 | 1.646258732 | 15.74266 | 305.455019 | 2.08988938 | 0.481349284 | 0.5161 | 2.089889383 | 2.089889383 | 40.69609461 | 0.084544292 | -1.072915708 | FALSE | FALSE | FALSE |
| P2RY13 | 0.1766874 | 0.84074348 | 1 | 0.176687414 | 0.176687414 | 0.176687414 | 0.21628707 | 0.848136434 | 0.00615 | 0.216287075 | 0.216287075 | 0.216287075 | 0.816911569 | -0.087824953 | FALSE | TRUE | FALSE |
| PRCP | 0.1766707 | 0.84075836 | 0.51215 | 0.176670725 | 0.176670725 | 0.875671494 | 0.06364088 | 0.883860795 | 1 | 0.254465012 | 0.254465012 | 1.262580138 | 2.776057352 | 0.443428434 | FALSE | TRUE | FALSE |
| TAGAP | 0.1766158 | 0.84077323 | 0.51425 | 1.619771018 | 1.619771018 | 31.88092795 | 0.22291667 | 0.846639318 | 1 | 0.222916672 | 0.222916672 | 4.395915609 | 0.792294964 | -0.101113105 | FALSE | TRUE | FALSE |
| ARL6IP4 | 0.1765824 | 0.84078809 | 1 | 0.176582381 | 0.176582381 | 0.176582381 | 1.92430407 | 0.496363818 | 0.5083 | 1.924304067 | 1.924304067 | 1.924304067 | 0.091764282 | -1.03732633 | FALSE | FALSE | FALSE |
| TEC | 0.1765633 | 0.84080296 | 0.5065 | 0.176563305 | 0.176563305 | 0.647218934 | 0.03077967 | 0.891083236 | 0.75145 | 0.123026203 | 0.491858112 | 2.436593008 | 5.736361131 | 0.758636484 | FALSE | TRUE | FALSE |
| PI16 | 0.1765573 | 0.84081782 | 1 | 0.176557302 | 0.176557302 | 3.493429536 | 1.86403335 | 0.50192184 | 0.25605 | 1.864033348 | 17.54971487 | 36.92985985 | 0.094717888 | -1.023567995 | FALSE | FALSE | FALSE |
| KCTD1 | 0.1765505 | 0.84083268 | 0.9926 | 1586.653105 | 174167.4338 | 174167.4338 | 0.17418139 | 0.858600607 | 1 | 1796402.332 | 2585490283 | 30466448824 | 1.01360123 | 0.005867129 | FALSE | TRUE | FALSE |
| ZMIZ2 | 0.1764684 | 0.84084754 | 1 | 1.652880315 | 1.652880315 | 32.32518851 | 0.22354185 | 0.846464771 | 1 | 0.223541854 | 0.223541854 | 4.383450394 | 0.789419737 | -0.10269202 | FALSE | TRUE | FALSE |
| SAMD14 | 0.1763878 | 0.84086239 | 1 | 0.176387842 | 0.176387842 | 3.489710679 | 7.64425534 | 0.233519913 | 0.26515 | 1.911241621 | 7.644255337 | 7.644255337 | 0.023074562 | -1.636866539 | FALSE | FALSE | FALSE |
| USF1 | 0.176378 | 0.84087724 | 1 | 0.176378012 | 0.176378012 | 0.176378012 | 0.21660021 | 0.848079378 | 0.00845 | 0.21660021 | 0.21660021 | 0.21660021 | 0.814302127 | -0.089214431 | FALSE | TRUE | FALSE |
| OR5L1 | 0.1762062 | 0.84089209 | 1 | 0.176206229 | 0.176206229 | 0.176206229 | 0.18860656 | 0.854735615 | 1 | 1.759040333 | 1.759040333 | 1.759040333 | 0.934252921 | -0.029535535 | FALSE | TRUE | FALSE |
| TBCC | 0.1761432 | 0.84090694 | 0.0135 | 0.176143226 | 0.176143226 | 3.485471862 | 0.21601286 | 0.848179201 | 1 | 0.216012859 | 0.216012859 | 4.279858664 | 0.815429352 | -0.08861366 | FALSE | TRUE | FALSE |
| MAF | 0.1761354 | 0.84092178 | 0.87685 | 2.816355296 | 26.63065348 | 26.63065348 | 0.2233784 | 0.846537545 | 1 | 0.893161816 | 3.572133549 | 3.572133549 | 0.788506846 | -0.103194532 | FALSE | TRUE | FALSE |
| SULT1E1 | 0.1760803 | 0.84093663 | 1 | 0.176080309 | 0.176080309 | 0.176080309 | 0.21594705 | 0.848193451 | 1 | 0.215947053 | 0.215947053 | 0.215947053 | 0.81538649 | -0.088636489 | FALSE | TRUE | FALSE |
| TLR6 | 0.1760632 | 0.84095146 | 0.8813 | 1.574528688 | 14.45234929 | 14.45234929 | 0.06783258 | 0.882932125 | 1 | 0.067832583 | 0.067832583 | 1.340613132 | 2.595555507 | 0.414230321 | FALSE | TRUE | FALSE |
| RNF40 | 0.175997 | 0.8409663 | 1 | 0.175997008 | 0.175997008 | 5.927457113 | 0.21352853 | 0.848761426 | 0.5188 | 0.854062891 | 0.854062891 | 16.70073467 | 0.824231827 | -0.08395062 | FALSE | TRUE | FALSE |
| RBMS2 | 0.1759252 | 0.84098113 | 1 | 0.175925205 | 0.175925205 | 3.491866971 | 0.21562818 | 0.84825043 | 1 | 0.215628177 | 0.215628177 | 0.215628177 | 0.815872987 | -0.088377446 | FALSE | TRUE | FALSE |
| OR56B4 | 0.1758325 | 0.84099596 | 0.8806 | 14.94345674 | 143.1496558 | 143.1496558 | 0.22161582 | 0.84687146 | 1 | 2.028315879 | 19.05309471 | 19.05309471 | 0.793411119 | -0.100501718 | FALSE | TRUE | FALSE |
| FAM228A | 0.1757154 | 0.84101079 | 0.4964 | 0.018987883 | 0.175715412 | 0.175715412 | 0.0220956 | 0.893194898 | 1 | 0.022095603 | 0.022095603 | 0.022095603 | 7.952505979 | 0.900504004 | FALSE | TRUE | FALSE |
| CHUK | 0.175707 | 0.84102562 | 1 | 0.175707 | 0.175707 | 3.468591 | 0.2154655 | 0.848264668 | 1 | 0.215465504 | 0.215465504 | 4.260088486 | 0.815476244 | -0.088588686 | FALSE | TRUE | FALSE |
| FNDC3B | 0.1756767 | 0.84104044 | 0.87765 | 15.96428839 | 158.1282805 | 311.6580095 | 0.2372706 | 0.842783856 | 1 | 0.237270605 | 0.237270605 | 4.64537733 | 0.740406371 | -0.130529853 | FALSE | TRUE | FALSE |
| CTNNA1 | 0.1756694 | 0.84105526 | 1 | 1.619790075 | 1.619790075 | 31.76981469 | 0.21963449 | 0.847189606 | 1 | 0.878507041 | 0.878507041 | 17.27072659 | 0.799826006 | -0.097004479 | FALSE | TRUE | FALSE |
| ARRB2 | 0.1756543 | 0.84107008 | 1 | 19499335124 | 4.17886E+14 | 5.30881E+15 | 0.23553751 | 0.843164308 | 1 | 2469560048 | 4.11821E+12 | 6.40012E+14 | 0.745759544 | -0.12740118 | FALSE | TRUE | FALSE |
| OR52K1 | 0.1756083 | 0.84108489 | 1 | 0.17560833 | 0.17560833 | 0.17560833 | 0.21485184 | 0.848406917 | 0.00675 | 0.214851837 | 0.214851837 | 0.214851837 | 0.817346189 | -0.087593958 | FALSE | TRUE | FALSE |
| RNGTT | 0.1755505 | 0.8410997 | 0.75485 | 1.639192653 | 6.555133185 | 13.87242234 | 0.22493407 | 0.846114525 | 0.0157 | 0.224934072 | 0.224934072 | 4.451221934 | 0.780452918 | -0.107653291 | FALSE | TRUE | FALSE |
| CDH5 | 0.1755025 | 0.84111451 | 0.51525 | 0.175502506 | 0.175502506 | 3.43567874 | 0.25392853 | 0.838234304 | 0.51315 | 0.253928526 | 0.253928526 | 4.98365171 | 0.691149232 | -0.16042817 | FALSE | TRUE | FALSE |
| SIRT2 | 0.1754472 | 0.84112932 | 0.01025 | 0.175447198 | 0.175447198 | 3.476671262 | 0.21504481 | 0.848335825 | 1 | 0.215044811 | 0.215044811 | 4.266053723 | 0.815863434 | -0.088382531 | FALSE | TRUE | FALSE |
| FBXL5 | 0.1753237 | 0.84114412 | 1 | 0.175323662 | 0.175323662 | 3.465833738 | 0.21466434 | 0.848421128 | 1 | 0.214664343 | 0.214664343 | 4.249445484 | 0.816733973 | -0.087919379 | FALSE | TRUE | FALSE |
| CIB2 | 0.1748538 | 0.84115892 | 1 | 0.174853784 | 0.174853784 | 0.174853784 | 0.2142535 | 0.848520537 | 1 | 0.214253497 | 0.214253497 | 0.214253497 | 0.816107027 | -0.088252883 | FALSE | TRUE | FALSE |
| C21orf91 | 0.1746618 | 0.84117373 | 0.0075 | 0.174661819 | 0.174661819 | 0.174661819 | 1.77436316 | 0.510582629 | 0.87545 | 16.57263587 | 158.7640492 | 158.7640492 | 0.098436342 | -1.006844533 | FALSE | FALSE | FALSE |
| DUSP22 | 0.1745693 | 0.84118853 | 1 | 1.613863399 | 1.613863399 | 1.613863399 | 1.92413313 | 0.496524254 | 0.7571 | 1.924133133 | 18.29724109 | 18.29724109 | 0.090726192 | -1.042267317 | FALSE | FALSE | FALSE |
| CALCOCO2 | 0.1745532 | 0.84120332 | 1 | 0.174553207 | 0.174553207 | 3.465356386 | 0.21365394 | 0.848690656 | 1 | 0.213653939 | 0.213653939 | 0.213653939 | 0.816990355 | -0.087783071 | FALSE | TRUE | FALSE |
| CTRB1 | 0.1745317 | 0.84121811 | 0.74815 | 0.697990977 | 2.792117872 | 2.792117872 | 0.86604193 | 0.604130772 | 0.49975 | 0.216546889 | 0.866041932 | 0.866041932 | 0.201528053 | -0.695664491 | FALSE | TRUE | FALSE |
| MUL1 | 0.1745268 | 0.84123291 | 0.5021 | 0.697972812 | 0.697972812 | 3.461965924 | 0.21661364 | 0.848050834 | 1 | 0.216613645 | 0.216613645 | 4.301043786 | 0.805705261 | -0.0938238 | FALSE | TRUE | FALSE |
| HPDL | 0.1743762 | 0.84124769 | 1 | 0.174376177 | 0.174376177 | 3.449018173 | 0.21407331 | 0.848548916 | 0.01205 | 0.214073312 | 0.214073312 | 4.239819165 | 0.814562897 | -0.089075376 | FALSE | TRUE | FALSE |
| TEFM | 0.1742449 | 0.84126248 | 1 | 0.174244909 | 0.174244909 | 0.174244909 | 1.90688302 | 0.498600498 | 0.00955 | 1.906883016 | 1.906883016 | 1.906883016 | 0.091376822 | -1.039163952 | FALSE | FALSE | FALSE |
| P4HA1 | 0.1742034 | 0.84127726 | 1 | 0.174203363 | 0.174203363 | 3.448920683 | 1.92688808 | 0.496042602 | 0.0124 | 1.926888076 | 1.926888076 | 4.110880546 | 0.090406581 | -1.043799955 | FALSE | FALSE | FALSE |
| C8G | 0.1741731 | 0.84129204 | 0.4996 | 0.174173085 | 1.580785696 | 1.580785696 | 0.20662057 | 0.850523805 | 1 | 1.883343274 | 17.61903058 | 17.61903058 | 0.842961008 | -0.074192514 | FALSE | TRUE | FALSE |
| RAMP1 | 0.1740246 | 0.84130682 | 0.751 | 1.581877294 | 14.75824175 | 14.75824175 | 0.21295044 | 0.848888645 | 1 | 1.945429086 | 18.24097494 | 18.24097494 | 0.817207104 | -0.087667867 | FALSE | TRUE | FALSE |
| OR1S1 | 0.174015 | 0.8413216 | 1 | 0.17401499 | 0.17401499 | 0.17401499 | 0.21332948 | 0.848817992 | 0.00725 | 0.21332948 | 0.21332948 | 0.21332948 | 0.815709998 | -0.088464215 | FALSE | TRUE | FALSE |
| SRGAP1 | 0.1739801 | 0.84133637 | 0.02875 | 0.173980107 | 0.173980107 | 3.403961093 | 0.2137895 | 0.848633992 | 1 | 0.213789498 | 0.213789498 | 4.193794209 | 0.813791642 | -0.089486775 | FALSE | TRUE | FALSE |
| MAP3K10 | 0.1739453 | 0.84135114 | 1 | 218172139.6 | 1.04467E+12 | 5.16772E+13 | 0.24393889 | 0.840838379 | 1 | 897296606.4 | 6.078E+12 | 1.19155E+14 | 0.713068974 | -0.146868459 | FALSE | TRUE | FALSE |
| MRO | 0.1739238 | 0.84136591 | 1 | 0.173923829 | 0.173923829 | 0.173923829 | 0.20679319 | 0.850468469 | 1 | 0.206793191 | 1.903336304 | 1.903336304 | 0.841052009 | -0.075177147 | FALSE | TRUE | FALSE |
| BNIP2 | 0.1738798 | 0.84138067 | 1 | 0.173879788 | 0.173879788 | 0.173879788 | 0.21353543 | 0.848747277 | 0.0069 | 0.21353543 | 0.21353543 | 0.21353543 | 0.814290105 | -0.089220843 | FALSE | TRUE | FALSE |
| TSPAN33 | 0.1738355 | 0.84139543 | 1 | 0.173835461 | 0.173835461 | 0.173835461 | 0.21357574 | 0.848733126 | 1 | 0.213575742 | 0.213575742 | 0.213575742 | 0.813928865 | -0.089413549 | FALSE | TRUE | FALSE |
| BMP4 | 0.1736492 | 0.84141019 | 0.8751 | 6.563263364 | 63.67519601 | 63.67519601 | 0.23030084 | 0.844787124 | 1 | 0.230300839 | 0.230300839 | 4.560918254 | 0.754010409 | -0.122622659 | FALSE | TRUE | FALSE |
| DIXDC1 | 0.1736276 | 0.84142495 | 0.01975 | 0.173627606 | 0.173627606 | 3.425827074 | 0.21321367 | 0.84884626 | 0.01475 | 0.213213675 | 0.213213675 | 4.21370733 | 0.814336165 | -0.089196278 | FALSE | TRUE | FALSE |
| SLIRP | 0.1736098 | 0.84143971 | 1 | 0.173609806 | 0.173609806 | 0.173609806 | 0.21262403 | 0.848945123 | 1 | 0.212624034 | 0.212624034 | 0.212624034 | 0.816510732 | -0.088038103 | FALSE | TRUE | FALSE |
| KIF3A | 0.1735762 | 0.84145446 | 1 | 0.173576189 | 0.173576189 | 3.426278684 | 0.21103277 | 0.849339344 | 1 | 0.843956751 | 0.843956751 | 7.141801238 | 0.822508224 | -0.084859751 | FALSE | TRUE | FALSE |
| TMA16 | 0.1733917 | 0.84146921 | 1 | 0.173391686 | 0.173391686 | 0.173391686 | 0.19048119 | 0.854210037 | 1 | 0.190481187 | 0.190481187 | 0.190481187 | 0.910282473 | -0.040823819 | FALSE | TRUE | FALSE |
| MYL5 | 0.1733171 | 0.84148395 | 1 | 0.173317108 | 0.173317108 | 0.173317108 | 0.21259093 | 0.848959237 | 1 | 0.212590929 | 0.212590929 | 0.212590929 | 0.815261066 | -0.088703298 | FALSE | TRUE | FALSE |
| ZDHHC23 | 0.1729993 | 0.8414987 | 0.5052 | 1.582532089 | 1.582532089 | 3.430905428 | 0.21874128 | 0.84746336 | 0.00955 | 0.218741282 | 0.218741282 | 0.218741282 | 0.790885523 | -0.101886374 | FALSE | TRUE | FALSE |
| SIRPB2 | 0.1729706 | 0.84151344 | 1 | 0.1729706 | 1.578196423 | 1.578196423 | 0.21859222 | 0.847506497 | 1 | 0.218592219 | 0.218592219 | 0.218592219 | 0.791293489 | -0.101662408 | FALSE | TRUE | FALSE |
| SUPV3L1 | 0.1728983 | 0.84152818 | 1 | 0.172898347 | 0.172898347 | 3.404453749 | 0.21237085 | 0.849015665 | 1 | 0.212370846 | 0.212370846 | 4.189476113 | 0.814134099 | -0.089304055 | FALSE | TRUE | FALSE |

| | | | | | | | | | | | | | | | | | |
|---|---|---|---|---|---|---|---|---|---|---|---|---|---|---|---|---|---|
| CMBL | 0.1728341 | 0.84154292 | 1 | 0.172834117 | 0.172834117 | 0.172834117 | 0.21174977 | 0.849170634 | 1 | 0.211749771 | 0.211749771 | 0.211749771 | 0.81621867 | -0.088193475 | FALSE | TRUE | FALSE |
| FKBPL | 0.1728155 | 0.84155766 | 1 | 0.172815547 | 0.172815547 | 3.430687364 | 0.83819289 | 0.610550415 | 0.50695 | 0.838192893 | 0.838192893 | 0.838192893 | 0.206176345 | -0.685761163 | FALSE | TRUE | FALSE |
| MNS1 | 0.1728111 | 0.84157239 | 0.873 | 0.17281107 | 13.67692827 | 13.67692827 | 2.33601726 | 0.461508885 | 0.68655 | 2.336017258 | 198.6018376 | 198.6018376 | 0.073976795 | -1.130904487 | FALSE | FALSE | FALSE |
| RANBP9 | 0.1727532 | 0.84158712 | 1 | 0.172753214 | 0.172753214 | 3.41591967 | 0.21270932 | 0.848931007 | 1 | 0.212709319 | 0.212709319 | 4.211720991 | 0.8121563 | -0.090360383 | FALSE | TRUE | FALSE |
| GLO1 | 0.1726729 | 0.84160185 | 1 | 1.584782732 | 144.2323364 | 144.2323364 | 0.2323842 | 0.844339708 | 0.005 | 0.232384197 | 0.232384197 | 0.232384197 | 0.743049206 | -0.128982425 | FALSE | TRUE | FALSE |
| TCTN3 | 0.17265 | 0.84161657 | 0.68865 | 0.379731014 | 13.69345128 | 13.69345128 | 0.15430544 | 0.863503218 | 0.37985 | 0.154305442 | 1.339463211 | 1.339463211 | 1.118884973 | 0.048785441 | FALSE | TRUE | FALSE |
| VPS37C | 0.1726304 | 0.84163129 | 0.87255 | 2.761102431 | 11.04658122 | 11.04658122 | 0.21956134 | 0.84724732 | 1 | 0.21956134 | 0.21956134 | 0.21956134 | 0.786251578 | -0.10443847 | FALSE | TRUE | FALSE |
| PICK1 | 0.1725088 | 0.84164601 | 0.0139 | 0.172508771 | 0.172508771 | 3.411946983 | 0.21238144 | 0.849001562 | 0.01215 | 0.212381445 | 0.212381445 | 4.206163871 | 0.812259149 | -0.090305388 | FALSE | TRUE | FALSE |
| APOL2 | 0.17246 | 0.84166073 | 1 | 1.56705737 | 14.61440825 | 14.61440825 | 147.388942 | 0.019208649 | 0.50285 | 1446.041676 | 149558.7222 | 288935.3246 | 0.001170101 | -2.931776485 | FALSE | FALSE | FALSE |
| CDS2 | 0.1721528 | 0.84167544 | 1 | 0.172152842 | 0.172152842 | 3.41284764 | 0.21106574 | 0.849311252 | 1 | 0.211065745 | 0.211065745 | 4.18869277 | 0.815636107 | -0.088503557 | FALSE | TRUE | FALSE |
| GAD1 | 0.1721392 | 0.84169016 | 1 | 15.25486822 | 1496.136369 | 1496.136369 | 0.23982788 | 0.842109843 | 1 | 0.239827876 | 0.239827876 | 4.741941087 | 0.717761618 | -0.144019769 | FALSE | TRUE | FALSE |
| HOXB9 | 0.1721094 | 0.84170486 | 1 | 0.688115902 | 2.751865637 | 2.751865637 | 51.3085597 | 0.053291206 | 0.5003 | 51.30855973 | 822.0765753 | 3291.82053 | 0.003354399 | -2.4743853 | FALSE | FALSE | FALSE |
| PANX3 | 0.1719626 | 0.84171957 | 1 | 1.565567039 | 14.62879271 | 14.62879271 | 0.22411024 | 0.846348193 | 1 | 0.22411024 | 0.22411024 | 0.22411024 | 0.767312429 | -0.115027767 | FALSE | TRUE | FALSE |
| RBM33 | 0.1718843 | 0.84173428 | 1 | 15.06446351 | 1461.2164 | 2861.896767 | 0.23922484 | 0.84226353 | 0.0296 | 0.239224839 | 0.239224839 | 4.685151689 | 0.718505377 | -0.143569977 | FALSE | TRUE | FALSE |
| OR8J3 | 0.1718716 | 0.84174898 | 0.7461 | 1.578125915 | 14.87218732 | 14.87218732 | 0.2137805 | 0.848648162 | 1 | 0.213780496 | 0.213780496 | 0.213780496 | 0.803962857 | -0.094764015 | FALSE | TRUE | FALSE |
| TAF6L | 0.1718194 | 0.84176368 | 1 | 1.584018057 | 1.584018057 | 31.17293111 | 0.21117277 | 0.849269093 | 1 | 1.953499348 | 1.953499348 | 7.150990828 | 0.813643947 | -0.089565602 | FALSE | TRUE | FALSE |
| ABI1 | 0.1717795 | 0.84177838 | 1 | 1.617902703 | 1.617902703 | 3.403255151 | 0.21841796 | 0.847578338 | 1 | 0.218417957 | 0.218417957 | 4.332144341 | 0.786471431 | -0.104317049 | FALSE | TRUE | FALSE |
| NOL6 | 0.1717686 | 0.84179307 | 1 | 0.171768557 | 0.171768557 | 3.339791351 | 0.21156493 | 0.849198777 | 1 | 0.211564929 | 0.211564929 | 4.12737083 | 0.81189523 | -0.09050001 | FALSE | TRUE | FALSE |
| SASS6 | 0.1717378 | 0.84180776 | 0.01625 | 0.17173776 | 0.17173776 | 3.398969444 | 0.21085504 | 0.849395498 | 0.0136 | 0.210855044 | 0.210855044 | 4.178394394 | 0.81448258 | -0.0891182 | FALSE | TRUE | FALSE |
| ANKRD13A | 0.1716554 | 0.84182245 | 1 | 0.686515064 | 0.686515064 | 5.806983144 | 0.20467127 | 0.850882575 | 0.7547 | 1.897951239 | 7.590765452 | 16.20538334 | 0.838688345 | -0.076399392 | FALSE | TRUE | FALSE |
| LRIF1 | 0.1715648 | 0.84183714 | 1 | 0.171564827 | 0.171564827 | 3.387584655 | 0.2105822 | 0.849479651 | 1 | 0.210582196 | 0.210582196 | 4.164364785 | 0.814716678 | -0.088993393 | FALSE | TRUE | FALSE |
| CTBS | 0.1714965 | 0.84185182 | 1 | 1.559651726 | 1.559651726 | 1.559651726 | 0.2167802 | 0.847979427 | 1 | 0.216780203 | 0.216780203 | 0.216780203 | 0.791107865 | -0.101764298 | FALSE | TRUE | FALSE |
| ANKRA2 | 0.1714926 | 0.8418665 | 0.00795 | 0.171492587 | 0.171492587 | 0.171492587 | 1.84959407 | 0.503956691 | 0.5067 | 1.849594071 | 7.39789086 | 7.39789086 | 0.09271904 | -1.032831074 | FALSE | FALSE | FALSE |
| SCAND3 | 0.1714738 | 0.84188118 | 0.2661 | 0.019289884 | 0.171473846 | 0.649258906 | 0.00369206 | 0.898437757 | 1 | 0.003692062 | 0.003692062 | 0.072410647 | 46.44392849 | 1.666928948 | FALSE | TRUE | FALSE |
| AIFM2 | 0.1714734 | 0.84189586 | 1 | 0.171473409 | 0.171473409 | 3.396346025 | 0.21058307 | 0.849465632 | 1 | 0.210583067 | 0.210583067 | 4.175830182 | 0.81427919 | -0.089226664 | FALSE | TRUE | FALSE |
| RNF139 | 0.17142 | 0.84191053 | 1 | 0.171419965 | 0.171419965 | 3.382570412 | 0.21053684 | 0.849507681 | 1 | 0.210536837 | 0.210536837 | 4.161132879 | 0.814204145 | -0.089266691 | FALSE | TRUE | FALSE |
| CYP2C18 | 0.1713241 | 0.8419252 | 0.51385 | 0.171324106 | 0.171324106 | 0.380089255 | 0.12367618 | 0.870440212 | 0.5128 | 0.123676183 | 0.123676183 | 0.613763067 | 1.385263529 | 0.1415324 | FALSE | TRUE | FALSE |
| MSMB | 0.1712824 | 0.84193987 | 1 | 0.1712824 | 0.1712824 | 0.1712824 | 0.18936252 | 0.85448643 | 1 | 0.189362517 | 0.189362517 | 0.189362517 | 0.904521139 | -0.043581279 | FALSE | TRUE | FALSE |
| SUPT5H | 0.171278 | 0.84195453 | 1 | 0.171278017 | 0.171278017 | 3.332613812 | 0.21108838 | 0.849297201 | 0.03135 | 0.211088381 | 0.211088381 | 4.120650358 | 0.811404286 | -0.090762702 | FALSE | TRUE | FALSE |
| SHROOM1 | 0.1712537 | 0.84196919 | 1 | 0.171253732 | 0.171253732 | 3.361199864 | 16.6169745 | 0.149216295 | 0.50265 | 16.61697448 | 160.6748512 | 326.8740277 | 0.010305951 | -1.986911907 | FALSE | FALSE | FALSE |
| MEP1A | 0.1711753 | 0.84198385 | 0.02275 | 0.171175276 | 0.171175276 | 3.373418963 | 2.09020044 | 0.481179629 | 0.6382 | 3.704889739 | 33.45300284 | 133.8817764 | 0.081894192 | -1.0867469 | FALSE | FALSE | FALSE |
| TMEM41A | 0.171165 | 0.84199851 | 1 | 2.41493E+28 | 1.93895E+35 | 1.86863E+38 | 4.73340427 | 0.318424893 | 1 | 3.47321E+25 | 7.77492E+31 | 2.78863E+34 | 0.036161078 | -1.441758631 | FALSE | FALSE | FALSE |
| OTUD6B | 0.1711332 | 0.84201316 | 1 | 0.171133182 | 0.171133182 | 0.171133182 | 0.21054061 | 0.849493668 | 1 | 0.210540614 | 0.210540614 | 0.210540614 | 0.812827408 | -0.090001661 | FALSE | TRUE | FALSE |
| ZSWIM4 | 0.1710443 | 0.84202781 | 1 | 1.585907361 | 1.585907361 | 30.85109141 | 0.21712449 | 0.847893649 | 1 | 0.21712449 | 0.21712449 | 4.237700224 | 0.787770568 | -0.103600249 | FALSE | TRUE | FALSE |
| ALDH1A2 | 0.171026 | 0.84204246 | 1 | 0.171025964 | 0.171025964 | 3.388734928 | 0.21064381 | 0.849437586 | 1 | 0.210643807 | 0.210643807 | 4.178393821 | 0.81192021 | -0.090486648 | FALSE | TRUE | FALSE |
| HSD17B7 | 0.1709954 | 0.84205711 | 0.00855 | 0.170995389 | 0.170995389 | 0.170995389 | 0.21052799 | 0.8495357 | 0.00645 | 0.21052799 | 0.21052799 | 0.21052799 | 0.812221639 | -0.090325444 | FALSE | TRUE | FALSE |
| ABHD10 | 0.1709937 | 0.84207175 | 1 | 1.570665905 | 1.570665905 | 1.570665905 | 0.21668476 | 0.848036558 | 1 | 0.21668476 | 0.21668476 | 0.21668476 | 0.789135926 | -0.102848185 | FALSE | TRUE | FALSE |
| KCND2 | 0.1709477 | 0.84208639 | 0.8747 | 3.80548413 | 149.1067138 | 149.1067138 | 0.06276228 | 0.884084434 | 0.87495 | 1.237711997 | 55.12124941 | 55.12124941 | 2.72373267 | 0.43516448 | FALSE | TRUE | FALSE |
| HIF3A | 0.1705955 | 0.84210103 | 1 | 0.170595521 | 0.170595521 | 3.357512004 | 0.2096661 | 0.84980136 | 0.01695 | 0.209666104 | 0.209666104 | 4.134384695 | 0.813653316 | -0.089560601 | FALSE | TRUE | FALSE |
| SLC35G1 | 0.1704724 | 0.84211567 | 1 | 0.170472388 | 0.170472388 | 0.170472388 | 0.2096382 | 0.849815317 | 0.00645 | 0.209638197 | 0.209638197 | 0.209638197 | 0.813174268 | -0.089816372 | FALSE | TRUE | FALSE |
| LIPJ | 0.1704655 | 0.8421303 | 1 | 3728.175768 | 239307.6777 | 2208578.505 | 16701.302 | 0.000116686 | 0.23065 | 4168.936125 | 247776.7674 | 3983042.256 | 1.02067E-05 | -4.991113941 | FALSE | FALSE | FALSE |
| KRT27 | 0.1704565 | 0.84214494 | 0.32135 | 0.170456524 | 2.724087935 | 2.724087935 | 8.8778308 | 0.213148463 | 0.15365 | 2.219169612 | 35.52480343 | 142.1886495 | 0.019200245 | -1.716693236 | FALSE | FALSE | FALSE |
| ZNF141 | 0.1704304 | 0.84215956 | 1 | 0.170430414 | 0.170430414 | 3.38147106 | 0.20949181 | 0.84985717 | 1 | 0.209491814 | 0.209491814 | 0.209491814 | 0.813542117 | -0.089619959 | FALSE | TRUE | FALSE |
| PSMG2 | 0.1703539 | 0.84217419 | 0.0067 | 0.170353896 | 0.170353896 | 0.170353896 | 0.2094638 | 0.849871117 | 1 | 0.209463796 | 0.209463796 | 0.209463796 | 0.813285633 | -0.0897569 | FALSE | TRUE | FALSE |
| EIF5 | 0.1703106 | 0.84218881 | 1 | 0.170310582 | 0.170310582 | 3.376679997 | 1.90659361 | 0.498759492 | 0.0113 | 1.90659361 | 1.90659361 | 4.038257826 | 0.089327154 | -1.049016501 | FALSE | FALSE | FALSE |
| MCOLN2 | 0.1702941 | 0.84220343 | 0.5084 | 0.170294071 | 0.170294071 | 0.379503793 | 0.03122667 | 0.890937214 | 0.01435 | 0.03122667 | 0.03122667 | 0.618632253 | 5.453481633 | 0.736673855 | FALSE | TRUE | FALSE |
| KANK2 | 0.1702503 | 0.84221805 | 1 | 0.170250306 | 0.170250306 | 3.319404247 | 0.20978786 | 0.849731539 | 1 | 0.209787861 | 0.209787861 | 4.102667696 | 0.811535545 | -0.090692454 | FALSE | TRUE | FALSE |
| CAMKK1 | 0.1701986 | 0.84223267 | 1 | 0.170198634 | 0.170198634 | 3.362585646 | 0.20961838 | 0.849829271 | 0.0128 | 0.209618381 | 0.209618381 | 4.147453406 | 0.81194518 | -0.090473292 | FALSE | TRUE | FALSE |
| STK32A | 0.1701747 | 0.84224728 | 1 | 0.170174727 | 0.170174727 | 3.377427663 | 0.81796644 | 0.615009211 | 0.2623 | 0.817966444 | 3.272391402 | 4.062609646 | 0.208046098 | -0.681840425 | FALSE | TRUE | FALSE |
| PEG3 | 0.1701387 | 0.84226189 | 0.99205 | 34.19679064 | 2189.097951 | 21404.66339 | 2.38474339 | 0.456744015 | 0.72675 | 5.585024086 | 89.35540316 | 434.2684972 | 0.071344666 | -1.146638493 | FALSE | FALSE | FALSE |
| RGP1 | 0.1700332 | 0.8422765 | 1 | 0.170033183 | 0.170033183 | 3.371768642 | 0.20893157 | 0.849996518 | 1 | 0.208931567 | 0.208931567 | 4.147359869 | 0.813822371 | -0.089470376 | FALSE | TRUE | FALSE |
| MAVS | 0.1699584 | 0.8422911 | 1 | 0.169958415 | 0.169958415 | 3.358693129 | 0.20871418 | 0.850052188 | 1 | 0.208714179 | 0.208714179 | 4.130487922 | 0.814311782 | -0.089209282 | FALSE | TRUE | FALSE |
| HDAC5 | 0.1697919 | 0.84230571 | 1 | 0.169791913 | 0.169791913 | 3.308790021 | 0.20930865 | 0.84991294 | 0.0309 | 0.209308647 | 0.209308647 | 4.091466145 | 0.811203531 | -0.090870168 | FALSE | TRUE | FALSE |
| SPRED2 | 0.1697186 | 0.84232031 | 0.5075 | 0.16971859 | 0.16971859 | 0.837940012 | 0.05851556 | 0.884954191 | 0.75355 | 0.538062144 | 5.077932174 | 10.63726024 | 2.900400791 | 0.462458015 | FALSE | TRUE | FALSE |
| ZEB2 | 0.1696848 | 0.84233491 | 1 | 15.51851603 | 154.2033957 | 302.8036677 | 0.22316422 | 0.846581178 | 1 | 2.1120485 | 2.1120485 | 41.33222343 | 0.760358618 | -0.118981527 | FALSE | TRUE | FALSE |
| BCAP29 | 0.1696734 | 0.8423495 | 1 | 0.169673395 | 1.547560647 | 1.547560647 | 0.21487676 | 0.848392704 | 0.00495 | 0.214876757 | 0.214876757 | 0.214876757 | 0.789631217 | -0.102575691 | FALSE | TRUE | FALSE |
| PSMC2 | 0.1695494 | 0.8423641 | 1 | 0.169549443 | 0.169549443 | 3.360307863 | 0.20927338 | 0.849940809 | 1 | 0.209273384 | 0.209273384 | 4.152111928 | 0.810181589 | -0.09141763 | FALSE | TRUE | FALSE |
| DNAJC21 | 0.1695413 | 0.84237869 | 0.7574 | 0.1695413 | 1.519006303 | 3.351954284 | 0.26860519 | 0.834166541 | 0.75325 | 0.26860519 | 2.429403634 | 5.31784347 | 0.631191451 | -0.199838892 | FALSE | TRUE | FALSE |
| CYYR1 | 0.1695321 | 0.84239327 | 1 | 0.169532062 | 0.169532062 | 0.169532062 | 0.20169765 | 0.85170415 | 0.50335 | 1.834168333 | 1.834168333 | 1.834168333 | 0.840525721 | -0.075448992 | FALSE | TRUE | FALSE |
| LRRC46 | 0.1694095 | 0.84240786 | 0.008 | 0.169409492 | 0.169409492 | 0.169409492 | 0.2080993 | 0.850177305 | 1 | 0.208099298 | 0.208099298 | 0.208099298 | 0.814080076 | -0.089332874 | FALSE | TRUE | FALSE |
| SLC35D1 | 0.1693894 | 0.84242244 | 1 | 0.169389381 | 0.169389381 | 0.169389381 | 0.20845496 | 0.850093916 | 1 | 0.208454962 | 0.208454962 | 0.208454962 | 0.812594621 | -0.090126057 | FALSE | TRUE | FALSE |
| BTRC | 0.1693882 | 0.84243702 | 1 | 0.169388173 | 0.169388173 | 3.349260762 | 0.20226113 | 0.851595114 | 1 | 1.872968597 | 1.872968597 | 6.837623849 | 0.837472684 | -0.077029349 | FALSE | TRUE | FALSE |
| PPIL6 | 0.1693578 | 0.8424516 | 1 | 0.677375466 | 0.677375466 | 0.677375466 | 0.21045909 | 0.849563708 | 1 | 0.210459086 | 0.210459086 | 0.210459086 | 0.804706506 | -0.094362487 | FALSE | TRUE | FALSE |
| BMP7 | 0.1693193 | 0.84246617 | 0.75175 | 0.677018412 | 2.707715686 | 3.350034509 | 0.21130548 | 0.849255034 | 1 | 0.844940161 | 0.844940161 | 4.18614288 | 0.801301038 | -0.096204295 | FALSE | TRUE | FALSE |
| HSF4 | 0.1692786 | 0.84248074 | 0.63125 | 0.677099945 | 6.064917038 | 6.064917038 | 0.06433919 | 0.883731416 | 0.01235 | 0.064339192 | 0.064339192 | 1.273952851 | 2.63103331 | 0.420126347 | FALSE | TRUE | FALSE |
| NMS | 0.1690324 | 0.84249531 | 1 | 0.169032391 | 0.169032391 | 0.169032391 | 0.20736374 | 0.850343816 | 0.00425 | 0.207363742 | 0.207363742 | 0.207363742 | 0.815149213 | -0.088762887 | FALSE | TRUE | FALSE |
| NFAT5 | 0.1690055 | 0.84250988 | 1 | 1.572180209 | 15.01046069 | 52.77693957 | 6.86176349 | 0.247344448 | 0.26705 | 1.903172799 | 18.21429394 | 63.98550846 | 0.024630038 | -1.60853491 | FALSE | FALSE | FALSE |
| PRKCD | 0.1689599 | 0.84252445 | 0.5082 | 0.675629271 | 0.675629271 | 5.79430262 | 0.21089298 | 0.849367426 | 1 | 0.210892976 | 0.210892976 | 4.156894317 | 0.80116412 | -0.096278509 | FALSE | TRUE | FALSE |
| GLI4 | 0.1688888 | 0.84253901 | 1 | 1.551754505 | 1.551754505 | 3.348245996 | 0.21434812 | 0.848492148 | 1 | 0.214348117 | 0.214348117 | 0.214348117 | 0.787918504 | -0.1035187 | FALSE | TRUE | FALSE |
| COPG1 | 0.1688582 | 0.84255357 | 1 | 0.168858231 | 0.168858231 | 3.310897636 | 0.20791208 | 0.850232847 | 1 | 0.207912077 | 0.207912077 | 4.086281565 | 0.812161722 | -0.090357483 | FALSE | TRUE | FALSE |
| NIPA2 | 0.1688487 | 0.84256812 | 1 | 0.168848709 | 0.168848709 | 3.354143553 | 0.20741386 | 0.850329954 | 1 | 0.207413859 | 0.207413859 | 0.207413859 | 0.814066667 | -0.089340027 | FALSE | TRUE | FALSE |
| CDK3 | 0.1688423 | 0.84258268 | 1 | 0.168842254 | 0.168842254 | 0.168842254 | 0.20768284 | 0.850274478 | 1 | 0.207682836 | 0.207682836 | 0.207682836 | 0.81298126 | -0.089919465 | FALSE | TRUE | FALSE |
| EDNRB | 0.1687977 | 0.84259723 | 1 | 0.168797733 | 0.168797733 | 3.33713702 | 1.85959558 | 0.503019601 | 0.0131 | 1.859595583 | 1.859595583 | 3.989545646 | 0.090771205 | -1.042051897 | FALSE | FALSE | FALSE |

| | | | | | | | | | | | | | | | | | |
|---|---|---|---|---|---|---|---|---|---|---|---|---|---|---|---|---|---|
| SIGLEC6 | 0.1687444 | 0.84261178 | 1 | 0.168744445 | 0.168744445 | 3.344045966 | 0.20747069 | 0.850288351 | 0.01175 | 0.207470693 | 0.207470693 | 4.116011936 | 0.813341117 | -0.089727272 | FALSE | TRUE | FALSE |
| BYSL | 0.1687318 | 0.84262632 | 1 | 0.674777232 | 2.699183163 | 13.33411425 | 0.82284036 | 0.614125656 | 0.87785 | 3.291526103 | 13.17005403 | 16.28335629 | 0.20506024 | -0.688118538 | FALSE | TRUE | FALSE |
| TRIM9 | 0.168687 | 0.84264087 | 0.5175 | 1.593753019 | 1.593753019 | 31.28763077 | 0.21502146 | 0.848350049 | 0.02 | 0.21502146 | 0.21502146 | 4.230543134 | 0.784512326 | -0.105400229 | FALSE | TRUE | FALSE |
| LATS1 | 0.1686539 | 0.84265541 | 1 | 0.168653932 | 0.168653932 | 5.674393145 | 17.24575 | 0.147615621 | 0.0206 | 1.81211072 | 17.24574997 | 61.04534276 | 0.009779449 | -2.009685615 | FALSE | FALSE | FALSE |
| MTUS2 | 0.1685238 | 0.84266995 | 1 | 0.168523812 | 0.168523812 | 5.680707521 | 0.19531506 | 0.85318797 | 1 | 1.818666751 | 17.38038116 | 64.15423046 | 0.862830587 | -0.064074468 | FALSE | TRUE | FALSE |
| F2RL3 | 0.1685062 | 0.84268448 | 1 | 0.168506185 | 0.168506185 | 3.330418127 | 0.19503595 | 0.853228041 | 1 | 1.798760289 | 17.02645871 | 35.60175325 | 0.86397502 | -0.063498814 | FALSE | TRUE | FALSE |
| KLHL21 | 0.1684156 | 0.84269901 | 1 | 0.168415552 | 0.168415552 | 3.32984194 | 0.20542957 | 0.850717188 | 0.5131 | 0.821540819 | 0.821540819 | 4.067248489 | 0.819821384 | -0.086280758 | FALSE | TRUE | FALSE |
| OAS2 | 0.1682354 | 0.84271355 | 0.50805 | 0.168235376 | 0.168235376 | 0.659799402 | 0.26551325 | 0.835076787 | 0.5077 | 0.265513253 | 0.265513253 | 1.030814099 | 0.633623271 | -0.198168881 | FALSE | TRUE | FALSE |
| LHX5 | 0.1681053 | 0.84272807 | 1 | 0.672109407 | 2.687863606 | 13.30266416 | 4.20956814 | 0.342749616 | 0.0118 | 0.212420764 | 0.212420764 | 4.209568137 | 0.039934085 | -1.398656262 | FALSE | FALSE | FALSE |
| MRPS31 | 0.168081 | 0.8427426 | 0.5027 | 1.536058908 | 1.536058908 | 1.536058908 | 1.83924355 | 0.504734962 | 0.75115 | 1.839243553 | 17.31212191 | 17.31212191 | 0.091385957 | -1.039120537 | FALSE | FALSE | FALSE |
| PRPSAP2 | 0.1680753 | 0.84275712 | 0.0094 | 0.168075339 | 0.168075339 | 0.168075339 | 0.81140621 | 0.616958151 | 0.74885 | 0.811406206 | 3.245391555 | 3.245391555 | 0.20714081 | -0.68373433 | FALSE | TRUE | FALSE |
| ASIC5 | 0.168059 | 0.84277164 | 0.87375 | 0.168058975 | 13.88130891 | 13.88130891 | 0.2832854 | 0.830175338 | 0.50875 | 0.283285403 | 0.283285403 | 0.636132765 | 0.593249682 | -0.226762486 | FALSE | TRUE | FALSE |
| UGT1A6 | 0.1680538 | 0.84278616 | 1 | 0.168053834 | 0.168053834 | 3.324927549 | 0.20702526 | 0.850413092 | 0.01395 | 0.207025257 | 0.207025257 | 4.101272663 | 0.811755226 | -0.090574907 | FALSE | TRUE | FALSE |
| NPEPL1 | 0.167839 | 0.84280068 | 1 | 0.167839048 | 0.167839048 | 3.321401323 | 0.20670957 | 0.850509975 | 0.0123 | 0.206709573 | 0.206709573 | 4.095804603 | 0.811955854 | -0.090467583 | FALSE | TRUE | FALSE |
| PDCL | 0.1677919 | 0.84281519 | 1 | 0.167791873 | 0.167791873 | 0.167791873 | 0.18101213 | 0.856839732 | 0.00655 | 0.181012131 | 0.181012131 | 0.181012131 | 0.926964798 | -0.032936758 | FALSE | TRUE | FALSE |
| TBC1D2B | 0.1677846 | 0.8428297 | 1 | 0.167784562 | 0.167784562 | 3.300975426 | 0.20043029 | 0.852016777 | 0.5146 | 1.846398797 | 1.846398797 | 6.771841057 | 0.837121773 | -0.077211362 | FALSE | TRUE | FALSE |
| ACTG1 | 0.1674232 | 0.84284421 | 0.0158 | 0.167423207 | 0.167423207 | 3.30974354 | 0.20687576 | 0.850440786 | 1 | 0.206875758 | 0.206875758 | 4.095359952 | 0.809293505 | -0.091893945 | FALSE | TRUE | FALSE |
| OR9Q2 | 0.1673727 | 0.84285872 | 1 | 1.527255356 | 1.527255356 | 3.323678909 | 0.21234077 | 0.849029765 | 1 | 0.212340771 | 0.212340771 | 0.212340771 | 0.788227001 | -0.103348692 | FALSE | TRUE | FALSE |
| NKX3-1 | 0.1673569 | 0.84287323 | 1 | 0.167356943 | 0.167356943 | 0.167356943 | 0.20675236 | 0.850496142 | 1 | 0.206752361 | 0.206752361 | 0.206752361 | 0.809456022 | -0.091806742 | FALSE | TRUE | FALSE |
| PLIN5 | 0.1672994 | 0.84288773 | 0.7523 | 0.16729939 | 1.572359427 | 3.313305483 | 0.20130085 | 0.851799434 | 0.26505 | 0.201300848 | 1.899837727 | 1.899837727 | 0.831091331 | -0.080351248 | FALSE | TRUE | FALSE |
| WNT1 | 0.1672332 | 0.84290223 | 1 | 0.167233154 | 0.167233154 | 3.30827765 | 0.20207534 | 0.851608752 | 0.7495 | 0.808017177 | 3.231740131 | 4.031285251 | 0.827578243 | -0.082190936 | FALSE | TRUE | FALSE |
| LRIT1 | 0.167206 | 0.84291672 | 0.87355 | 1.487682307 | 13.5768863 | 13.5768863 | 0.27473761 | 0.83238506 | 0.635 | 0.501094677 | 4.39578216 | 4.39578216 | 0.608602383 | -0.215666351 | FALSE | TRUE | FALSE |
| GPNMB | 0.1671027 | 0.84293122 | 0.7515 | 0.167102671 | 1.476150107 | 3.304131883 | 0.03030804 | 0.89118294 | 1 | 0.263726617 | 2.35538494 | 2.35538494 | 5.513475772 | 0.741425471 | FALSE | TRUE | FALSE |
| DYTN | 0.1670928 | 0.84294571 | 0.9421 | 1.487539222 | 54.37164511 | 54.37164511 | 43.5837317 | 0.063346765 | 0.19865 | 43.58373171 | 420.6744266 | 4159.634285 | 0.003833834 | -2.416366697 | FALSE | FALSE | FALSE |
| TBC1D12 | 0.1670641 | 0.8429602 | 1 | 0.167064123 | 0.167064123 | 3.301925013 | 0.20578427 | 0.850661982 | 1 | 0.205784266 | 0.205784266 | 4.072970132 | 0.811841092 | -0.09052897 | FALSE | TRUE | FALSE |
| HPR | 0.1668388 | 0.84297469 | 0.8732 | 1.473714168 | 24.59955453 | 98.38711149 | 90.3482812 | 0.032732409 | 0.2354 | 16.38088474 | 607.7953256 | 3019.455913 | 0.001846619 | -2.733622752 | FALSE | FALSE | FALSE |
| COQ9 | 0.1668375 | 0.84298917 | 0.5051 | 0.667219707 | 0.667219707 | 0.667219707 | 0.82297458 | 0.614027238 | 0.50245 | 0.822974578 | 0.822974578 | 0.822974578 | 0.202724984 | -0.693092725 | FALSE | TRUE | FALSE |
| POGZ | 0.1668096 | 0.84300365 | 1 | 0.166809642 | 0.166809642 | 5.603606968 | 0.20622163 | 0.850592923 | 1 | 0.206221628 | 0.206221628 | 6.937548772 | 0.808885294 | -0.09211306 | FALSE | TRUE | FALSE |
| PSMC6 | 0.166794 | 0.84301813 | 0.8782 | 2.667030087 | 25.17792457 | 25.17792457 | 7.87828952 | 0.230546178 | 0.2552 | 1.970327452 | 7.878289524 | 7.878289524 | 0.021171348 | -1.674251493 | FALSE | FALSE | FALSE |
| PDS5B | 0.1667732 | 0.84303261 | 1 | 0.166773215 | 0.166773215 | 5.603221127 | 0.2059191 | 0.850634365 | 1 | 0.2059191 | 0.2059191 | 6.928284211 | 0.809896773 | -0.091570331 | FALSE | TRUE | FALSE |
| HIPK2 | 0.1667328 | 0.84304708 | 1 | 0.166732752 | 0.166732752 | 3.242070612 | 0.81433219 | 0.616083684 | 0.51125 | 0.814332188 | 0.814332188 | 15.8874525 | 0.204747834 | -0.688780685 | FALSE | TRUE | FALSE |
| GSDMB | 0.166728 | 0.84306155 | 1 | 1.514548695 | 14.12074951 | 14.12074951 | 0.21791861 | 0.847707484 | 1 | 0.21791861 | 0.21791861 | 0.21791861 | 0.765093169 | -0.116285676 | FALSE | TRUE | FALSE |
| MRPL20 | 0.1666401 | 0.84307602 | 1 | 0.166640058 | 0.166640058 | 0.166640058 | 0.20538684 | 0.850744776 | 1 | 0.205386837 | 0.205386837 | 0.205386837 | 0.811347313 | -0.090793198 | FALSE | TRUE | FALSE |
| GABRG3 | 0.1666071 | 0.84309049 | 1 | 0.16660707 | 0.16660707 | 3.300336653 | 0.20596311 | 0.850620554 | 1 | 0.205963106 | 0.205963106 | 4.084628529 | 0.808917058 | -0.092096007 | FALSE | TRUE | FALSE |
| RLN1 | 0.1665747 | 0.84310495 | 1 | 14.12410814 | 135.1453271 | 135.1453271 | 152.810073 | 0.018118793 | 0.3496 | 152.8100732 | 15129.6036 | 156005.774 | 0.001090076 | -2.962543054 | FALSE | FALSE | FALSE |
| RTL1 | 0.1664612 | 0.84311941 | 1 | 1.520709339 | 1.520709339 | 3.316789022 | 0.21136619 | 0.849226911 | 1 | 0.211366193 | 0.211366193 | 0.211366193 | 0.787549028 | -0.1037224 | FALSE | TRUE | FALSE |
| RHOT1 | 0.1664342 | 0.84313387 | 1 | 6.173189665 | 58.76074411 | 111.3169818 | 0.21438043 | 0.848463748 | 1 | 1.995520745 | 1.995520745 | 4.273089317 | 0.776349904 | -0.109942496 | FALSE | TRUE | FALSE |
| THSD7B | 0.166415 | 0.84314832 | 0.2796 | 0.16641504 | 1.481518019 | 7.180363466 | 0.08004606 | 0.880318875 | 0.27895 | 0.080046056 | 0.320078857 | 3.649709366 | 2.078991134 | 0.317852637 | FALSE | TRUE | FALSE |
| PPP2R5B | 0.1662662 | 0.84316278 | 1 | 0.166266171 | 0.166266171 | 3.289573389 | 0.20539605 | 0.850730983 | 0.0119 | 0.205396047 | 0.205396047 | 4.069013072 | 0.809490611 | -0.091788184 | FALSE | TRUE | FALSE |
| SACM1L | 0.1662378 | 0.84317723 | 1 | 0.664850404 | 0.664850404 | 3.290630857 | 0.20746907 | 0.850302221 | 1 | 0.207469069 | 0.207469069 | 4.111862826 | 0.80126548 | -0.096223567 | FALSE | TRUE | FALSE |
| XKR4 | 0.1661813 | 0.84319168 | 1 | 15.16437356 | 150.5173163 | 298.1510152 | 0.22600471 | 0.845836133 | 1 | 0.226004713 | 0.226004713 | 4.45260222 | 0.735300312 | -0.13353525 | FALSE | TRUE | FALSE |
| CHODL | 0.1661733 | 0.84320612 | 1 | 0.166173294 | 0.166173294 | 0.166173294 | 0.19246599 | 0.853760413 | 1 | 1.759137192 | 16.50216868 | 16.50216868 | 0.863390421 | -0.063792774 | FALSE | TRUE | FALSE |
| RBBP9 | 0.1660765 | 0.84322057 | 1 | 14.10518602 | 135.0758101 | 135.0758101 | 0.22396739 | 0.846406503 | 1 | 0.22396739 | 0.22396739 | 0.22396739 | 0.741520749 | -0.129876692 | FALSE | TRUE | FALSE |
| TMED10 | 0.1660152 | 0.84323501 | 0.0065 | 0.166015181 | 0.166015181 | 0.166015181 | 0.17255527 | 0.858947787 | 1 | 0.172555274 | 0.172555274 | 0.172555274 | 0.962098563 | -0.016780434 | FALSE | TRUE | FALSE |
| RANBP6 | 0.1660005 | 0.84324944 | 0.5156 | 0.166000467 | 0.166000467 | 3.256398453 | 0.02897251 | 0.891496322 | 1 | 0.463388625 | 4.148257906 | 9.11117685 | 5.729584836 | 0.758123154 | FALSE | TRUE | FALSE |
| LRGUK | 0.1659636 | 0.84326388 | 0.50955 | 0.165963592 | 0.165963592 | 3.266268936 | 0.02917964 | 0.89146582 | 0.75615 | 0.255115478 | 2.289307102 | 5.03048567 | 5.687650516 | 0.754932903 | FALSE | TRUE | FALSE |
| PANX1 | 0.1659185 | 0.84327831 | 0.0117 | 0.165918453 | 0.165918453 | 3.288662516 | 0.20428313 | 0.851006384 | 1 | 0.204283128 | 0.204283128 | 4.053446782 | 0.812198514 | -0.090337809 | FALSE | TRUE | FALSE |
| EIF3A | 0.1658912 | 0.84329274 | 1 | 0.165891235 | 3.204113322 | 5.547336037 | 0.20507063 | 0.850786141 | 1 | 0.205070634 | 0.205070634 | 6.871085987 | 0.80894681 | -0.092080034 | FALSE | TRUE | FALSE |
| TAB1 | 0.1658038 | 0.84330717 | 1 | 0.165803762 | 0.165803762 | 3.271273249 | 0.20436011 | 0.850965137 | 1 | 0.204360106 | 0.204360106 | 4.038537212 | 0.811331356 | -0.090801739 | FALSE | TRUE | FALSE |
| UCK1 | 0.1658016 | 0.84332159 | 1 | 1.530828882 | 1.530828882 | 1.530828882 | 0.21108968 | 0.849283148 | 1 | 0.211089682 | 0.211089682 | 0.211089682 | 0.785455846 | -0.104878224 | FALSE | TRUE | FALSE |
| LETM1 | 0.1656416 | 0.84333602 | 1 | 1.527070425 | 1.527070425 | 30.00469201 | 16.8230495 | 0.148254405 | 0.50645 | 16.82304952 | 163.5311393 | 331.2476054 | 0.00984611 | -2.006735309 | FALSE | FALSE | FALSE |
| BRPF3 | 0.1656156 | 0.84335044 | 1 | 0.165615641 | 0.165615641 | 3.223138836 | 0.20493985 | 0.850827484 | 0.03185 | 0.204939848 | 0.204939848 | 4.001391874 | 0.8081183 | -0.092525059 | FALSE | TRUE | FALSE |
| TTC7B | 0.1655723 | 0.84336485 | 0.5139 | 0.662131955 | 0.662131955 | 12.98163816 | 0.20713005 | 0.850385389 | 1 | 0.207130053 | 0.207130053 | 4.070592922 | 0.79936408 | -0.097255371 | FALSE | TRUE | FALSE |
| XRCC2 | 0.165571 | 0.84337927 | 1 | 0.165570978 | 0.165570978 | 0.165570978 | 1.81348391 | 0.50628673 | 0.5006 | 1.813483911 | 1.813483911 | 1.813483911 | 0.091299943 | -1.039529492 | FALSE | FALSE | FALSE |
| PGLYRP3 | 0.165529 | 0.84339368 | 1 | 1.511507979 | 1.511507979 | 1.511507979 | 0.21030211 | 0.849605701 | 1 | 0.210302109 | 0.210302109 | 0.210302109 | 0.78710118 | -0.103969436 | FALSE | TRUE | FALSE |
| ZC3H6 | 0.1654434 | 0.84340809 | 1 | 1.519814914 | 1.519814914 | 29.84788802 | 0.20631066 | 0.850579105 | 1 | 0.825229667 | 0.825229667 | 16.24202434 | 0.801914048 | -0.095872179 | FALSE | TRUE | FALSE |
| LILRB1 | 0.1653534 | 0.8434225 | 0.94 | 1.528582191 | 140.8709355 | 140.8709355 | 158.812856 | 0.016770752 | 0.5059 | 1583.855391 | 168987.5489 | 1805945.431 | 0.001041184 | -2.982472585 | FALSE | FALSE | FALSE |
| RTN2 | 0.1653404 | 0.8434369 | 0.0194 | 0.165340414 | 0.165340414 | 3.259115346 | 0.20380171 | 0.851157425 | 0.01755 | 0.203801711 | 0.203801711 | 4.0242202 | 0.811280792 | -0.090828806 | FALSE | TRUE | FALSE |
| USP33 | 0.1653158 | 0.84345131 | 1 | 1.520677888 | 1.520677888 | 29.96268578 | 0.2104365 | 0.849577708 | 1 | 0.2104365 | 0.2104365 | 4.153735075 | 0.785585066 | -0.104806781 | FALSE | TRUE | FALSE |
| MAP1B | 0.1652622 | 0.8434657 | 1 | 0.165262249 | 3.156726954 | 5.516022341 | 0.204391 | 0.850937627 | 1 | 0.204391004 | 3.925328992 | 6.837080258 | 0.808559309 | -0.092288118 | FALSE | TRUE | FALSE |
| DPH3 | 0.1652247 | 0.8434801 | 1 | 0.165224704 | 0.165224704 | 0.165224704 | 0.17572011 | 0.858151812 | 0.0022 | 0.175720108 | 0.175720108 | 0.175720108 | 0.940272034 | -0.026746481 | FALSE | TRUE | FALSE |
| ASNS | 0.1651745 | 0.8434945 | 1 | 0.165174512 | 0.165174512 | 3.267445878 | 0.20401591 | 0.851116262 | 1 | 0.204015913 | 0.204015913 | 4.041095403 | 0.809615824 | -0.091721012 | FALSE | TRUE | FALSE |
| TUSC3 | 0.1651575 | 0.84350889 | 1 | 0.16515746 | 0.16515746 | 3.277693674 | 7.22927744 | 0.239804331 | 0.25365 | 1.807607878 | 7.22927744 | 7.22927744 | 0.022845639 | -1.641196696 | FALSE | FALSE | FALSE |
| ATG9A | 0.1651161 | 0.84352328 | 0.02735 | 0.165116086 | 0.165116086 | 3.234279763 | 0.19781467 | 0.852584407 | 1 | 1.83581761 | 1.83581761 | 36.04907743 | 0.834700917 | -0.07846911 | FALSE | TRUE | FALSE |
| HMGCL | 0.1650258 | 0.84353766 | 0.81905 | 0.659875178 | 2.639247994 | 10.55861826 | 0.28171395 | 0.83070665 | 0.81785 | 1.126578694 | 4.506332396 | 18.02990308 | 0.58579214 | -0.23225646 | FALSE | TRUE | FALSE |
| NLN | 0.1650153 | 0.84355205 | 0.7583 | 0.165015278 | 1.486791163 | 3.256726423 | 0.49232922 | 0.757907539 | 0.2598 | 0.123081257 | 0.492329218 | 2.432980242 | 0.335172628 | -0.474731456 | FALSE | TRUE | FALSE |
| PLIN3 | 0.1650067 | 0.84356643 | 0.01565 | 0.165006674 | 0.165006674 | 3.259544452 | 0.20334174 | 0.851294477 | 1 | 0.203341739 | 0.203341739 | 4.022760286 | 0.811474689 | -0.090725022 | FALSE | TRUE | FALSE |
| PKNOX1 | 0.1649607 | 0.84358081 | 1 | 0.164960719 | 0.164960719 | 3.268728113 | 0.20424761 | 0.851020128 | 0.0107 | 0.204247611 | 0.204247611 | 4.051703525 | 0.807650668 | -0.092776443 | FALSE | TRUE | FALSE |
| AJUBA | 0.1649492 | 0.84359518 | 1 | 0.164949201 | 0.164949201 | 3.261074078 | 0.20408464 | 0.851075076 | 0.0115 | 0.204084644 | 0.204084644 | 4.040366762 | 0.808239159 | -0.092460112 | FALSE | TRUE | FALSE |
| UNC5C | 0.1649459 | 0.84360956 | 0.508 | 0.164945854 | 0.164945854 | 3.233873209 | 0.06070045 | 0.884597473 | 1 | 0.060700449 | 0.060700449 | 1.192898499 | 2.717374519 | 0.434149499 | FALSE | TRUE | FALSE |
| HERC3 | 0.1649214 | 0.84362393 | 0.76175 | 0.374885042 | 1.499010342 | 7.365063989 | 0.06243205 | 0.88418738 | 0.0218 | 0.062432046 | 0.062432046 | 1.229164501 | 2.641615343 | 0.421869578 | FALSE | TRUE | FALSE |
| NME6 | 0.1648772 | 0.84363829 | 1 | 1.503844219 | 6.014900266 | 6.014900266 | 0.21187068 | 0.849100231 | 0.00635 | 0.211870684 | 0.211870684 | 0.211870684 | 0.778197275 | -0.108910294 | FALSE | TRUE | FALSE |
| HSPA12B | 0.1648696 | 0.84365266 | 1 | 0.164869635 | 0.164869635 | 3.238342727 | 0.20349631 | 0.851225982 | 0.02065 | 0.203496315 | 0.203496315 | 4.005660153 | 0.810184865 | -0.091415874 | FALSE | TRUE | FALSE |

| | | | | | | | | | | | | | | | | | |
|---|---|---|---|---|---|---|---|---|---|---|---|---|---|---|---|---|---|
| OR5AC2 | 0.1648504 | 0.84366702 | 0.0083 | 0.164850373 | 0.164850373 | 0.164850373 | 0.20295331 | 0.851417609 | 1 | 0.202953307 | 0.202953307 | 0.202953307 | 0.812257635 | -0.090306198 | FALSE | TRUE | FALSE |
| HEYL | 0.1648313 | 0.84368138 | 1 | 0.164831307 | 0.164831307 | 0.164831307 | 0.1952994 | 0.853201329 | 1 | 1.812273397 | 7.24758558 | 7.24758558 | 0.843992922 | -0.073661196 | FALSE | TRUE | FALSE |
| SSTR3 | 0.1648265 | 0.84369574 | 0.0164 | 0.164826485 | 0.164826485 | 3.251474533 | 0.20384339 | 0.851143706 | 1 | 0.20384339 | 0.20384339 | 4.027761264 | 0.80859372 | -0.092269636 | FALSE | TRUE | FALSE |
| SMUG1 | 0.164802 | 0.84371009 | 1 | 1.513920966 | 1.513920966 | 1.513920966 | 16.6325111 | 0.148895563 | 0.508 | 16.63251107 | 161.1694139 | 161.1694139 | 0.009908428 | -2.003995248 | FALSE | FALSE | FALSE |
| MROH7 | 0.1647846 | 0.84372444 | 0.2642 | 0.019541035 | 0.164784611 | 0.658079448 | 0.0388584 | 0.889244248 | 0.76075 | 0.038858404 | 0.332914666 | 1.310122843 | 4.240642734 | 0.627431685 | FALSE | TRUE | FALSE |
| HHATL | 0.1647799 | 0.84373879 | 0.509 | 1.522022603 | 14.41969175 | 14.41969175 | 9.60576337 | 0.203699304 | 0.4028 | 2.402718477 | 91.56210057 | 366.1451656 | 0.017154271 | -1.765627742 | FALSE | FALSE | FALSE |
| TFAP2C | 0.1647248 | 0.84375314 | 1 | 0.164724786 | 0.164724786 | 3.262035036 | 0.20404095 | 0.851102536 | 1 | 0.204040949 | 0.204040949 | 4.045399522 | 0.807312391 | -0.092958382 | FALSE | TRUE | FALSE |
| INTS2 | 0.1646837 | 0.84376748 | 1 | 0.164683695 | 0.164683695 | 5.544778154 | 0.18713711 | 0.855218445 | 1 | 1.748901632 | 1.748901632 | 34.30798303 | 0.880016253 | -0.055509307 | FALSE | TRUE | FALSE |
| PLEKHA6 | 0.1646138 | 0.84378182 | 1 | 0.164613816 | 0.164613816 | 3.217214497 | 0.20365399 | 0.851198566 | 1 | 0.203653994 | 0.203653994 | 3.991148863 | 0.808301437 | -0.092426649 | FALSE | TRUE | FALSE |
| CD207 | 0.164565 | 0.84379616 | 1 | 0.164565012 | 0.164565012 | 3.269241941 | 1.79976748 | 0.507367927 | 0.50305 | 1.799767475 | 1.799767475 | 1.799767475 | 0.091436818 | -1.038878894 | FALSE | FALSE | FALSE |
| MRPL24 | 0.1645181 | 0.8438105 | 1 | 0.164518149 | 0.164518149 | 0.164518149 | 0.20331568 | 0.851308169 | 1 | 0.203315682 | 0.203315682 | 0.203315682 | 0.809175894 | -0.091957063 | FALSE | TRUE | FALSE |
| OR9Q1 | 0.1644146 | 0.84382483 | 1 | 0.164414621 | 1.491621106 | 1.491621106 | 1.84664414 | 0.504423882 | 0.0068 | 1.846644139 | 1.846644139 | 1.846644139 | 0.089034274 | -1.050442777 | FALSE | FALSE | FALSE |
| BFAR | 0.1643899 | 0.84383916 | 1 | 1.496195045 | 1.496195045 | 3.260044589 | 1.85313743 | 0.503488544 | 0.50515 | 1.853137431 | 1.853137431 | 4.022262298 | 0.088708982 | -1.052032402 | FALSE | FALSE | FALSE |
| GDI2 | 0.1643798 | 0.84385349 | 0.0117 | 0.164379754 | 0.164379754 | 3.259358137 | 0.20130696 | 0.851785829 | 0.50055 | 0.804940465 | 0.804940465 | 3.995682132 | 0.8165627 | -0.088010462 | FALSE | TRUE | FALSE |
| WNT7A | 0.1643661 | 0.84386782 | 1 | 0.65722101 | 0.65722101 | 3.249798102 | 0.81383541 | 0.616375629 | 0.5064 | 0.813835413 | 0.813835413 | 4.029568805 | 0.201964789 | -0.694724339 | FALSE | TRUE | FALSE |
| CLMP | 0.1643433 | 0.84388214 | 1 | 0.164343321 | 0.164343321 | 3.262335357 | 0.20303974 | 0.851390264 | 0.0096 | 0.203039745 | 0.203039745 | 0.203039745 | 0.809414535 | -0.091829001 | FALSE | TRUE | FALSE |
| DDX1 | 0.1642665 | 0.84389646 | 0.0169 | 0.164266469 | 0.164266469 | 3.245401646 | 0.20331185 | 0.851321857 | 1 | 0.203311847 | 0.203311847 | 4.022692336 | 0.807953257 | -0.092613764 | FALSE | TRUE | FALSE |
| NKAIN1 | 0.1641342 | 0.84391078 | 1 | 0.164134187 | 0.164134187 | 0.164134187 | 0.17006089 | 0.859686248 | 0.00415 | 0.170060891 | 0.170060891 | 0.170060891 | 0.965149516 | -0.015405403 | FALSE | TRUE | FALSE |
| HIVEP2 | 0.1641152 | 0.8439251 | 1 | 1.545440412 | 3.13294568 | 51.61265279 | 0.20961836 | 0.849843222 | 1 | 0.20961836 | 4.02359365 | 7.015522732 | 0.782923981 | -0.106280404 | FALSE | TRUE | FALSE |
| TBC1D1 | 0.1641049 | 0.84393941 | 0.9998 | 108097.4299 | 119068354.9 | 14096936883 | 0.03434895 | 0.890278422 | 0.99995 | 22309742.78 | 3.26191E+11 | 3.76984E+12 | 4.77758117 | 0.679208074 | FALSE | TRUE | FALSE |
| GSX1 | 0.1640826 | 0.84395372 | 1 | 0.164082556 | 0.164082556 | 0.164082556 | 0.20102223 | 0.851853827 | 1 | 0.803737679 | 0.803737679 | 0.803737679 | 0.816240858 | -0.08818167 | FALSE | TRUE | FALSE |
| GNAT3 | 0.1640805 | 0.84396803 | 0.97105 | 2.623648917 | 10.49525283 | 41.99412829 | 0.96670285 | 0.591431066 | 0.68945 | 0.966702851 | 15.46793899 | 15.46793899 | 0.169732066 | -0.770236103 | FALSE | TRUE | FALSE |
| SPAG9 | 0.163993 | 0.84398233 | 0.03405 | 0.163992989 | 0.163992989 | 3.196704245 | 0.15816155 | 0.862520475 | 1 | 1389.62503 | 149027.419 | 1608204.48 | 1.036870134 | 0.015724365 | FALSE | TRUE | FALSE |
| TNFRSF11B | 0.1639306 | 0.84399664 | 0.01005 | 0.163930616 | 0.163930616 | 3.251494857 | 0.20298953 | 0.851403938 | 0.0107 | 0.202989529 | 0.202989529 | 4.03021687 | 0.807581638 | -0.092813564 | FALSE | TRUE | FALSE |
| NUBPL | 0.1639101 | 0.84401094 | 1 | 0.163910068 | 0.163910068 | 0.163910068 | 0.19621441 | 0.853027476 | 1 | 1.80247498 | 1.80247498 | 1.80247498 | 0.83536202 | -0.078125274 | FALSE | TRUE | FALSE |
| ADH5 | 0.1637238 | 0.84402523 | 1 | 1.492058463 | 1.492058463 | 1.492058463 | 0.20193497 | 0.85166328 | 1 | 1.848997089 | 1.848997089 | 1.848997089 | 0.81077482 | -0.091099747 | FALSE | TRUE | FALSE |
| LRRC49 | 0.163656 | 0.84403953 | 1 | 0.16365599 | 0.16365599 | 3.230905247 | 1.79532807 | 0.508444196 | 0.5089 | 1.795328072 | 1.795328072 | 6.630353683 | 0.091156593 | -1.040211917 | FALSE | FALSE | FALSE |
| HK1 | 0.1636498 | 0.84405382 | 0.51535 | 0.163649835 | 2.614153686 | 10.45205061 | 0.0076095 | 0.897407997 | 0.3286 | 0.0076095 | 0.12143968 | 0.193254274 | 21.50599029 | 1.332559445 | FALSE | TRUE | FALSE |
| USP43 | 0.1635374 | 0.84406811 | 1 | 1.503207238 | 1.503207238 | 29.47381552 | 0.20840616 | 0.850121721 | 1 | 0.208406156 | 0.208406156 | 4.095942091 | 0.784705182 | -0.10529348 | FALSE | TRUE | FALSE |
| MAP3K7 | 0.1635361 | 0.8440824 | 0.0159 | 0.163536074 | 0.163536074 | 3.23364795 | 0.20026442 | 0.852057461 | 1 | 0.800857294 | 0.800857294 | 3.965288611 | 0.816600749 | -0.087990226 | FALSE | TRUE | FALSE |
| TM4SF5 | 0.1634851 | 0.84409669 | 0.00725 | 0.163485074 | 0.163485074 | 0.163485074 | 0.20145797 | 0.851731386 | 1 | 0.201457967 | 0.201457967 | 0.201457967 | 0.811509598 | -0.090706339 | FALSE | TRUE | FALSE |
| DOCK7 | 0.1634309 | 0.84411097 | 0.7604 | 0.163430942 | 1.497680695 | 28.81606228 | 0.03000752 | 0.891228904 | 0.52095 | 0.270038615 | 0.580004809 | 5.219481742 | 5.446333122 | 0.736104201 | FALSE | TRUE | FALSE |
| CPA5 | 0.1633338 | 0.84412525 | 0.0114 | 0.163333761 | 0.163333761 | 3.235980648 | 0.19543486 | 0.853147877 | 1 | 1.785915754 | 1.785915754 | 3.87634613 | 0.835745261 | -0.077926077 | FALSE | TRUE | FALSE |
| PRPF4 | 0.1633188 | 0.84413953 | 1 | 0.163318832 | 0.163318832 | 3.229616776 | 0.20232172 | 0.851581474 | 1 | 0.202321715 | 0.202321715 | 4.006312635 | 0.807223443 | -0.093006234 | FALSE | TRUE | FALSE |
| TFEB | 0.1630763 | 0.84415381 | 1 | 0.163076268 | 0.163076268 | 3.227036307 | 0.20199311 | 0.851636021 | 1 | 0.201993112 | 0.201993112 | 4.002227362 | 0.807335787 | -0.092945796 | FALSE | TRUE | FALSE |
| PUS10 | 0.1630441 | 0.84416808 | 0.50995 | 0.163044111 | 0.163044111 | 0.363773039 | 2.11451981 | 0.479647384 | 0.5058 | 19.85807858 | 191.1654644 | 1885.254234 | 0.077106921 | -1.112906641 | FALSE | FALSE | FALSE |
| SNF8 | 0.1629628 | 0.84418235 | 1 | 5.957901047 | 55.89579074 | 55.89579074 | 0.85565245 | 0.606498853 | 0.50355 | 0.855652451 | 0.855652451 | 0.855652451 | 0.190454411 | -0.720208964 | FALSE | TRUE | FALSE |
| INCA1 | 0.1627726 | 0.84419662 | 1 | 0.162772645 | 1.473128811 | 1.473128811 | 0.20676383 | 0.850482307 | 1 | 0.206763832 | 0.206763832 | 0.206763832 | 0.787239447 | -0.103893152 | FALSE | TRUE | FALSE |
| OR8H1 | 0.1627174 | 0.84421089 | 1 | 1.47617769 | 1.47617769 | 1.47617769 | 1.77341 | 0.510734699 | 0.74735 | 1.773409997 | 16.58426276 | 16.58426276 | 0.091753974 | -1.037375118 | FALSE | FALSE | FALSE |
| POLR3H | 0.1626052 | 0.84422516 | 1 | 0.162605193 | 0.162605193 | 0.162605193 | 0.20131222 | 0.851772222 | 1 | 0.201312216 | 0.201312216 | 0.201312216 | 0.807726406 | -0.092735719 | FALSE | TRUE | FALSE |
| ARID2 | 0.1625545 | 0.84423942 | 1 | 0.162554506 | 0.162554506 | 5.449769752 | 0.20157567 | 0.851717769 | 0.0407 | 0.201575671 | 0.201575671 | 6.76932863 | 0.806419273 | -0.093439102 | FALSE | TRUE | FALSE |
| SNW1 | 0.1624721 | 0.84425368 | 1 | 0.162472075 | 0.162472075 | 3.21331528 | 0.20118397 | 0.851826635 | 1 | 0.201183971 | 0.201183971 | 3.984267386 | 0.807579618 | -0.092814651 | FALSE | TRUE | FALSE |
| SART3 | 0.162456 | 0.84426794 | 0.0302 | 0.162456036 | 0.162456036 | 3.178168663 | 16.4951922 | 0.15081734 | 0.2636 | 1.742735113 | 16.49519222 | 58.83346989 | 0.00984869 | -2.00662153 | FALSE | FALSE | FALSE |
| AMPH | 0.1623698 | 0.84428219 | 1 | 0.162369754 | 0.162369754 | 3.204058651 | 0.20091601 | 0.851894598 | 0.01655 | 0.200916014 | 0.200916014 | 3.971062775 | 0.808147399 | -0.09250942 | FALSE | TRUE | FALSE |
| BDKRB1 | 0.1623449 | 0.84429645 | 0.01085 | 0.162344879 | 0.162344879 | 3.220725941 | 0.19889554 | 0.852382163 | 0.49675 | 0.198895542 | 0.795368769 | 0.795368769 | 0.816231867 | -0.088186454 | FALSE | TRUE | FALSE |
| EEF2 | 0.1623359 | 0.8443107 | 1 | 0.162335877 | 0.162335877 | 3.168422215 | 0.20141459 | 0.851745001 | 1 | 0.201414587 | 0.201414587 | 3.942571429 | 0.805978752 | -0.093676407 | FALSE | TRUE | FALSE |
| OS9 | 0.1622667 | 0.84432494 | 0.0184 | 0.162266682 | 0.162266682 | 3.194953432 | 0.20050433 | 0.851989643 | 0.0173 | 0.200504332 | 0.200504332 | 3.955211411 | 0.809292646 | -0.091894406 | FALSE | TRUE | FALSE |
| ZFYVE1 | 0.162265 | 0.84433919 | 1 | 0.162264971 | 0.162264971 | 3.185634886 | 0.19497407 | 0.853241394 | 1 | 1.824588274 | 1.824588274 | 35.90016561 | 0.832238728 | -0.079752078 | FALSE | TRUE | FALSE |
| PEX13 | 0.1622393 | 0.84435343 | 1 | 0.648712817 | 0.648712817 | 3.222811711 | 1.84940942 | 0.504112521 | 0.0078 | 1.849409421 | 1.849409421 | 1.849409421 | 0.087724931 | -1.056876964 | FALSE | FALSE | FALSE |
| ANKHD1 | 0.1621982 | 0.84436767 | 1 | 0.162198187 | 3.10473935 | 5.412581092 | 0.20107581 | 0.851840232 | 1 | 0.201075808 | 3.868870068 | 6.724900883 | 0.806651924 | -0.093313826 | FALSE | TRUE | FALSE |
| TDG | 0.162187 | 0.84438191 | 0.01025 | 0.16218699 | 0.16218699 | 3.220088864 | 0.2002165 | 0.852071017 | 0.0085 | 0.200216499 | 0.200216499 | 0.200216499 | 0.810058069 | -0.091483848 | FALSE | TRUE | FALSE |
| TEX10 | 0.1619777 | 0.84439614 | 1 | 0.161977718 | 0.161977718 | 3.187794008 | 0.20066398 | 0.851962499 | 1 | 0.200663976 | 0.200663976 | 3.956752364 | 0.807208753 | -0.093014138 | FALSE | TRUE | FALSE |
| FYN | 0.1619674 | 0.84441038 | 1 | 0.647726097 | 0.647726097 | 5.4802913 | 0.20293749 | 0.851431278 | 1 | 0.20293749 | 0.20293749 | 4.017913165 | 0.798114924 | -0.097934568 | FALSE | TRUE | FALSE |
| OR6C3 | 0.1619641 | 0.84442461 | 1 | 0.161964054 | 0.161964054 | 0.161964054 | 0.18805997 | 0.855023082 | 1 | 1.713467803 | 16.02331061 | 16.02331061 | 0.861236179 | -0.064877735 | FALSE | TRUE | FALSE |
| RASSF8 | 0.1618885 | 0.84443883 | 1 | 0.161888515 | 0.161888515 | 3.215866969 | 0.20077111 | 0.851921766 | 0.00905 | 0.200771111 | 0.200771111 | 0.200771111 | 0.806333711 | -0.093485183 | FALSE | TRUE | FALSE |
| CIB4 | 0.1618747 | 0.84445306 | 1 | 0.161874654 | 0.161874654 | 0.161874654 | 0.20033103 | 0.852030341 | 1 | 0.200331034 | 0.200331034 | 0.200331034 | 0.808035832 | -0.09256938 | FALSE | TRUE | FALSE |
| B3GAT2 | 0.1618624 | 0.84446728 | 0.0124 | 0.161862425 | 0.161862425 | 3.20599039 | 0.19992063 | 0.85212522 | 0.0092 | 0.199920629 | 0.199920629 | 0.199920629 | 0.809633432 | -0.091711567 | FALSE | TRUE | FALSE |
| NDE1 | 0.1618088 | 0.8444815 | 0.50405 | 0.647069071 | 0.647069071 | 3.20820552 | 0.19655956 | 0.85296051 | 1 | 1.828640611 | 1.828640611 | 1.828640611 | 0.82320486 | -0.084492074 | FALSE | TRUE | FALSE |
| DFFA | 0.1618084 | 0.84449572 | 1 | 0.161808367 | 0.161808367 | 0.161808367 | 1.76975033 | 0.511342033 | 0.49905 | 0.193781031 | 1.769750331 | 1.769750331 | 0.091430053 | -1.038911028 | FALSE | FALSE | FALSE |
| MRPL35 | 0.1617561 | 0.84450993 | 0.00595 | 0.017658122 | 0.161756056 | 0.161756056 | 0.02140057 | 0.893416738 | 1 | 0.021400573 | 0.021400573 | 0.021400573 | 7.558491665 | 0.878435139 | FALSE | TRUE | FALSE |
| NKIRAS1 | 0.1616641 | 0.84452414 | 1 | 1.488618983 | 1.488618983 | 1.488618983 | 0.20631149 | 0.850565284 | 0.00485 | 0.206311488 | 0.206311488 | 0.206311488 | 0.783592403 | -0.105909783 | FALSE | TRUE | FALSE |
| MDN1 | 0.1616074 | 0.84453835 | 0.17455 | 0.010098661 | 0.730080744 | 6.140024465 | 0.0010658 | 0.899382387 | 0.12745 | 0.001065799 | 0.019489064 | 0.034841489 | 151.6303119 | 2.180786028 | FALSE | TRUE | FALSE |
| TRIM71 | 0.1615874 | 0.84455256 | 1 | 0.16158736 | 0.16158736 | 3.165423547 | 0.20062025 | 0.851976072 | 1 | 0.200620246 | 0.200620246 | 3.939774379 | 0.805438948 | -0.093967373 | FALSE | TRUE | FALSE |
| CLEC12A | 0.1614839 | 0.84456677 | 1 | 0.161483874 | 0.161483874 | 0.161483874 | 1.70351828 | 0.516746325 | 0.75105 | 1.703518284 | 15.88550203 | 15.88550203 | 0.09479433 | -1.023217638 | FALSE | FALSE | FALSE |
| KCNG3 | 0.1614412 | 0.84458097 | 1 | 0.645500708 | 0.645500708 | 3.191186039 | 0.19023965 | 0.854275937 | 1 | 1.791554853 | 17.31584249 | 35.46307939 | 0.848620284 | -0.071286592 | FALSE | TRUE | FALSE |
| RAB32 | 0.1614188 | 0.84459517 | 0.00625 | 0.161418837 | 0.161418837 | 0.161418837 | 0.7819479 | 0.627412784 | 0.75215 | 0.781947897 | 3.127790207 | 3.127790207 | 0.206431704 | -0.685223603 | FALSE | TRUE | FALSE |
| NT5C2 | 0.1613933 | 0.84460936 | 1 | 0.16139325 | 0.16139325 | 3.192627134 | 0.19947045 | 0.852192921 | 1 | 0.199470449 | 0.199470449 | 3.951039657 | 0.809108572 | -0.091993198 | FALSE | TRUE | FALSE |
| FOLR1 | 0.1613583 | 0.84462356 | 0.00785 | 0.161358315 | 0.161358315 | 0.161358315 | 0.20000461 | 0.852098123 | 1 | 0.200004605 | 0.200004605 | 0.200004605 | 0.806772998 | -0.093248646 | FALSE | TRUE | FALSE |
| ITPA | 0.1611387 | 0.84463775 | 1 | 0.161138679 | 0.161138679 | 0.161138679 | 0.00019234 | 0.900048915 | 1 | 0.000192342 | 0.000192342 | 0.000192342 | 837.7718661 | 2.923125772 | FALSE | TRUE | FALSE |
| KLHDC2 | 0.1611332 | 0.84465194 | 1 | 0.161133212 | 0.161133212 | 3.197764904 | 0.19926404 | 0.852301118 | 1 | 0.199264037 | 0.199264037 | 0.199264037 | 0.808641708 | -0.092243863 | FALSE | TRUE | FALSE |
| MYLIP | 0.161132 | 0.84466613 | 1 | 0.161131953 | 0.161131953 | 3.192337957 | 0.19987139 | 0.852138765 | 1 | 0.199871394 | 0.199871394 | 3.964320129 | 0.806178162 | -0.09356897 | FALSE | TRUE | FALSE |
| OAT | 0.1611099 | 0.84468031 | 0.3227 | 0.161109928 | 2.576083646 | 2.576083646 | 0.01810526 | 0.894281136 | 0.01125 | 0.018105262 | 0.018105262 | 0.359431279 | 8.898513956 | 0.949317486 | FALSE | TRUE | FALSE |
| ADAMTSL4 | 0.1610894 | 0.8446945 | 0.88635 | 6.462411509 | 2009.522263 | 10534.01714 | 1475.79784 | 0.001602772 | 0.30245 | 627.4246563 | 139565.577 | 5605889.52 | 0.000109154 | -3.961959981 | FALSE | FALSE | FALSE |

| | | | | | | | | | | | | | | | | | |
|---|---|---|---|---|---|---|---|---|---|---|---|---|---|---|---|---|---|
| LETM2 | 0.1610589 | 0.84470868 | 0.4988 | 0.07453606 | 0.644135709 | 0.644135709 | 0.03048075 | 0.89115228 | 1 | 0.030480748 | 0.030480748 | 0.030480748 | 5.283956221 | 0.722959211 | FALSE | TRUE | FALSE |
| IL22RA1 | 0.1609726 | 0.84472285 | 1 | 0.160972612 | 0.160972612 | 3.179178821 | 0.19931708 | 0.852260563 | 1 | 0.199317082 | 0.199317082 | 3.942400079 | 0.807620752 | -0.09279253 | FALSE | TRUE | FALSE |
| SFRP4 | 0.1609444 | 0.84473703 | 0.0129 | 0.160944372 | 0.160944372 | 3.187408689 | 0.19940521 | 0.852233514 | 1 | 0.199405214 | 0.199405214 | 3.953748461 | 0.807122188 | -0.093060714 | FALSE | TRUE | FALSE |
| TNFRSF4 | 0.1609342 | 0.8447512 | 0.00975 | 0.160934205 | 0.160934205 | 0.160934205 | 0.19886551 | 0.852395662 | 1 | 0.198865508 | 0.198865508 | 0.198865508 | 0.809261529 | -0.091911105 | FALSE | TRUE | FALSE |
| HS1BP3 | 0.1608775 | 0.84476537 | 1 | 0.160877491 | 0.160877491 | 3.189484947 | 0.18705283 | 0.855231452 | 1 | 1.708109047 | 16.00894895 | 16.00894895 | 0.860064476 | -0.06546899 | FALSE | TRUE | FALSE |
| SOX17 | 0.1608141 | 0.84477954 | 1 | 0.642936198 | 0.642936198 | 3.184136003 | 0.20139262 | 0.851758613 | 0.0114 | 0.201392622 | 0.201392622 | 3.992390186 | 0.798510141 | -0.097719564 | FALSE | TRUE | FALSE |
| NPSR1 | 0.1606905 | 0.8447937 | 0.0098 | 0.160690514 | 0.160690514 | 0.160690514 | 1.72308254 | 0.516001965 | 0.7522 | 1.723082544 | 16.28648633 | 16.28648633 | 0.093257583 | -1.030315842 | FALSE | FALSE | FALSE |
| CARNS1 | 0.1606369 | 0.84480786 | 0.7595 | 0.160636851 | 1.472084596 | 3.167227067 | 0.02546624 | 0.892314111 | 1 | 19.79227204 | 1895.472853 | 19222.61303 | 6.30783468 | 0.799880303 | FALSE | TRUE | FALSE |
| SRGAP3 | 0.1606037 | 0.84482202 | 1 | 1372.708118 | 147037.9629 | 1568077.975 | 0.22538644 | 0.84602672 | 1 | 20.07047471 | 196.7976488 | 391.82069 | 0.712570329 | -0.147172266 | FALSE | TRUE | FALSE |
| IL17RA | 0.160561 | 0.84483618 | 0.0246 | 0.160560993 | 0.160560993 | 3.15027428 | 0.19866819 | 0.852436146 | 0.0221 | 0.198668186 | 0.198668186 | 3.906861959 | 0.808186734 | -0.092488283 | FALSE | TRUE | FALSE |
| ZFYVE9 | 0.1605385 | 0.84485034 | 1 | 0.160538486 | 0.160538486 | 3.135406073 | 0.19894248 | 0.852368661 | 1 | 0.198942483 | 0.198942483 | 3.896445707 | 0.806959295 | -0.093148372 | FALSE | TRUE | FALSE |
| PI4K2A | 0.1605053 | 0.84486449 | 1 | 0.160505276 | 0.160505276 | 3.175144937 | 0.19928077 | 0.852287602 | 1 | 0.199280768 | 0.199280768 | 3.94737389 | 0.805422807 | -0.093976077 | FALSE | TRUE | FALSE |
| GLT1D1 | 0.1603187 | 0.84487864 | 1 | 0.160318707 | 0.160318707 | 0.160318707 | 16.0132924 | 0.153386695 | 0.13005 | 16.01329241 | 156.426826 | 156.426826 | 0.010011602 | -1.999496433 | FALSE | FALSE | FALSE |
| CTDP1 | 0.1602896 | 0.84489279 | 0.0282 | 0.160289624 | 0.160289624 | 3.139902779 | 0.19876659 | 0.852409159 | 1 | 0.198766585 | 0.198766585 | 3.903264016 | 0.80642138 | -0.093437967 | FALSE | TRUE | FALSE |
| ARHGEF39 | 0.1602788 | 0.84490693 | 1 | 0.160278828 | 0.160278828 | 0.160278828 | 0.19817586 | 0.852543999 | 1 | 0.198175864 | 0.198175864 | 0.198175864 | 0.808770681 | -0.092174601 | FALSE | TRUE | FALSE |
| KRTAP10-9 | 0.1601872 | 0.84492107 | 1 | 0.160187241 | 0.160187241 | 0.160187241 | 1.747256 | 0.513155633 | 0.5057 | 1.747255998 | 1.747255998 | 1.747255998 | 0.091679319 | -1.037728619 | FALSE | FALSE | FALSE |
| P2RX1 | 0.1601562 | 0.84493521 | 0.5012 | 1.454815714 | 1.454815714 | 3.171295279 | 0.2040448 | 0.851088808 | 1 | 0.204044803 | 0.204044803 | 4.045174255 | 0.78490707 | -0.105181759 | FALSE | TRUE | FALSE |
| SNX5 | 0.1601169 | 0.84494935 | 1 | 0.160116857 | 0.160116857 | 3.179564887 | 1.76167469 | 0.51179662 | 0.50155 | 1.761674688 | 1.761674688 | 1.761674688 | 0.090889004 | -1.041488658 | FALSE | FALSE | FALSE |
| GDF3 | 0.160072 | 0.84496349 | 1 | 0.640117993 | 0.640117993 | 0.640117993 | 6.93231184 | 0.24636467 | 0.51335 | 6.932311842 | 66.09612501 | 66.09612501 | 0.023090706 | -1.636562796 | FALSE | FALSE | FALSE |
| ZMIZ1 | 0.1600706 | 0.84497762 | 1 | 0.160070554 | 0.160070554 | 3.114259723 | 6.71399312 | 0.251574754 | 0.00725 | 0.198800545 | 0.198800545 | 3.880464498 | 0.023841334 | -1.622669445 | FALSE | FALSE | FALSE |
| KLK13 | 0.1600543 | 0.84499175 | 1 | 1.48100222 | 1.48100222 | 1.48100222 | 0.20469726 | 0.850855035 | 1 | 0.204697264 | 0.204697264 | 0.204697264 | 0.781907233 | -0.106844769 | FALSE | TRUE | FALSE |
| CCDC88A | 0.1599664 | 0.84500588 | 1 | 0.159966359 | 0.159966359 | 5.351941306 | 0.19859832 | 0.852449636 | 1 | 0.198598317 | 0.198598317 | 6.657219438 | 0.805476911 | -0.093946904 | FALSE | TRUE | FALSE |
| RGL2 | 0.1599585 | 0.84502 | 0.51165 | 1.464623668 | 1.464623668 | 28.78088967 | 0.20421093 | 0.851061343 | 1 | 0.204210929 | 0.204210929 | 4.021326908 | 0.783300437 | -0.106071631 | FALSE | TRUE | FALSE |
| SMC3 | 0.1599435 | 0.84503413 | 1 | 0.15994352 | 0.15994352 | 5.37727214 | 0.19873461 | 0.852422654 | 1 | 0.198734606 | 0.198734606 | 6.690409538 | 0.804809606 | -0.094306849 | FALSE | TRUE | FALSE |
| TWSG1 | 0.1598925 | 0.84504825 | 0.00695 | 0.159892515 | 0.159892515 | 0.159892515 | 0.19759369 | 0.852651706 | 1 | 0.197593687 | 0.197593687 | 0.197593687 | 0.809198503 | -0.091944929 | FALSE | TRUE | FALSE |
| MKNK1 | 0.1598597 | 0.84506236 | 1 | 0.15985968 | 0.15985968 | 3.168420953 | 1.78131047 | 0.509820924 | 0.51005 | 1.781310471 | 1.781310471 | 3.813621757 | 0.089742739 | -1.047000681 | FALSE | FALSE | FALSE |
| SIT1 | 0.1598244 | 0.84507648 | 1 | 0.15982441 | 0.15982441 | 0.15982441 | 0.16679177 | 0.860490006 | 1 | 0.166791766 | 0.166791766 | 0.166791766 | 0.958227219 | -0.018531497 | FALSE | TRUE | FALSE |
| LRRN3 | 0.1598214 | 0.84509059 | 1 | 0.159821428 | 0.159821428 | 3.159047822 | 1.79401357 | 0.508750742 | 0.0149 | 1.794013571 | 1.794013571 | 3.806704181 | 0.089085964 | -1.050190716 | FALSE | FALSE | FALSE |
| CNTNAP3 | 0.1598008 | 0.8451047 | 0.999 | 2095.849543 | 534142.0008 | 10424695.91 | 0.26454132 | 0.835377965 | 0.99695 | 889.2968655 | 357867.3764 | 3716728.544 | 0.604067399 | -0.218914602 | FALSE | TRUE | FALSE |
| ATP6V1B2 | 0.1597796 | 0.84511881 | 0.01585 | 0.159779626 | 0.159779626 | 3.15980002 | 0.19857365 | 0.852463123 | 1 | 0.198573648 | 0.198573648 | 3.932281779 | 0.804636604 | -0.094400215 | FALSE | TRUE | FALSE |
| MAP3K7CL | 0.1597341 | 0.84513291 | 1 | 0.159734083 | 0.159734083 | 0.159734083 | 0.19659704 | 0.85294711 | 1 | 0.196597039 | 0.196597039 | 0.196597039 | 0.812494859 | -0.090179379 | FALSE | TRUE | FALSE |
| HSPA13 | 0.1596263 | 0.84514701 | 1 | 0.159626332 | 0.159626332 | 3.164877059 | 0.18348002 | 0.85611065 | 1 | 0.183480019 | 0.183480019 | 3.641608211 | 0.869993003 | -0.06048424 | FALSE | TRUE | FALSE |
| ATF6 | 0.1595971 | 0.84516111 | 1 | 1.460444268 | 5.84151137 | 12.61346433 | 0.20377538 | 0.851171141 | 1 | 0.203775382 | 0.815068826 | 4.032269015 | 0.783201229 | -0.106126639 | FALSE | TRUE | FALSE |
| NDUFB9 | 0.1595749 | 0.84517521 | 1 | 0.159574914 | 0.159574914 | 0.159574914 | 0.17843653 | 0.857385036 | 1 | 15.85643802 | 621.0824014 | 621.0824014 | 0.894295109 | -0.048519144 | FALSE | TRUE | FALSE |
| ZNF232 | 0.1595211 | 0.8451893 | 1 | 0.1595211 | 0.1595211 | 3.165841617 | 1.75309253 | 0.512854237 | 0.01055 | 1.753092527 | 1.753092527 | 3.803541618 | 0.090994113 | -1.040986702 | FALSE | FALSE | FALSE |
| PDGFRL | 0.1594888 | 0.8452034 | 1 | 1.459399714 | 1.459399714 | 3.163352485 | 0.19748257 | 0.852692057 | 1 | 1.814785443 | 1.814785443 | 1.814785443 | 0.807609332 | -0.092798671 | FALSE | TRUE | FALSE |
| GOPC | 0.1593266 | 0.84521749 | 1 | 0.159326609 | 0.159326609 | 3.158342492 | 0.19740412 | 0.852745824 | 0.01115 | 0.197404121 | 0.197404121 | 3.91731922 | 0.807108832 | -0.0930679 | FALSE | TRUE | FALSE |
| OSBPL8 | 0.1593007 | 0.84523157 | 1 | 0.159300734 | 0.159300734 | 3.135985321 | 0.19756155 | 0.852665159 | 1 | 0.197561547 | 0.197561547 | 3.896536758 | 0.806334719 | -0.09348464 | FALSE | TRUE | FALSE |
| CLASRP | 0.1592959 | 0.84524566 | 0.0231 | 0.159295906 | 0.159295906 | 3.134263906 | 1.75707521 | 0.512552503 | 0.0201 | 1.757075211 | 1.757075211 | 6.468726661 | 0.090659697 | -1.042585739 | FALSE | FALSE | FALSE |
| RRP7A | 0.1590792 | 0.84525974 | 0.50105 | 1.442217692 | 1.442217692 | 1.442217692 | 0.2027065 | 0.851499584 | 0.0089 | 0.202706505 | 0.202706505 | 0.202706505 | 0.784775814 | -0.10525439 | FALSE | TRUE | FALSE |
| GPR137C | 0.159005 | 0.84527382 | 1 | 1.493652853 | 1.493652853 | 1.493652853 | 0.17695065 | 0.857851097 | 1 | 0.176950653 | 0.176950653 | 0.176950653 | 0.898583806 | -0.046441412 | FALSE | TRUE | FALSE |
| TMEM116 | 0.1589071 | 0.8452879 | 0.00655 | 0.158907064 | 0.158907064 | 0.158907064 | 0.19643508 | 0.853000696 | 1 | 0.196435078 | 0.196435078 | 0.196435078 | 0.808954621 | -0.09207584 | FALSE | TRUE | FALSE |
| PRPF3 | 0.1588279 | 0.84530198 | 1 | 0.158827916 | 0.158827916 | 3.135387235 | 0.19740891 | 0.852732386 | 1 | 0.197408913 | 0.197408913 | 3.903084088 | 0.804563045 | -0.094439919 | FALSE | TRUE | FALSE |
| ERVFRD-1 | 0.1588233 | 0.84531605 | 1 | 1.44124941 | 1.44124941 | 1.44124941 | 0.20247726 | 0.85154054 | 0.008 | 0.202477256 | 0.202477256 | 0.202477256 | 0.784400881 | -0.105461927 | FALSE | TRUE | FALSE |
| GPR107 | 0.1587376 | 0.84533012 | 0.01165 | 0.158737571 | 0.158737571 | 3.146259799 | 0.19714954 | 0.852812978 | 1 | 0.19714954 | 0.19714954 | 3.911822502 | 0.805163284 | -0.094116037 | FALSE | TRUE | FALSE |
| BMP6 | 0.1586336 | 0.84534419 | 0.01655 | 0.158633621 | 0.158633621 | 3.133109693 | 0.78020805 | 0.628416397 | 0.5069 | 0.780208047 | 0.780208047 | 3.859519413 | 0.203322206 | -0.691815187 | FALSE | TRUE | FALSE |
| RHOF | 0.1586306 | 0.84535826 | 1 | 0.158630556 | 0.158630556 | 0.158630556 | 1.77013529 | 0.51119036 | 0.50185 | 1.770135291 | 1.770135291 | 1.770135291 | 0.089614933 | -1.047619615 | FALSE | FALSE | FALSE |
| HAND2 | 0.1585376 | 0.84537232 | 1 | 0.158537553 | 0.158537553 | 0.158537553 | 0.19717869 | 0.852786123 | 1 | 0.197178688 | 0.197178688 | 0.197178688 | 0.804029862 | -0.094727821 | FALSE | TRUE | FALSE |
| THAP4 | 0.1585299 | 0.84538638 | 0.5183 | 0.158529938 | 0.158529938 | 1.343226873 | 0.23243622 | 0.844294831 | 0.0149 | 0.232436219 | 0.232436219 | 1.147737631 | 0.682036295 | -0.166192513 | FALSE | TRUE | FALSE |
| CLEC2B | 0.1585032 | 0.84540044 | 1 | 0.158503154 | 0.158503154 | 0.158503154 | 0.17192813 | 0.859170111 | 1 | 0.17192813 | 0.17192813 | 0.17192813 | 0.921915187 | -0.035309031 | FALSE | TRUE | FALSE |
| WNT4 | 0.1584854 | 0.8454145 | 1 | 0.158485384 | 0.158485384 | 3.136705994 | 0.77943843 | 0.628870663 | 0.4972 | 0.194931003 | 0.779438435 | 3.862861281 | 0.20333278 | -0.691792602 | FALSE | TRUE | FALSE |
| IDO2 | 0.1584013 | 0.84542855 | 0.0111 | 0.158401295 | 0.158401295 | 3.14437882 | 16.2446945 | 0.151780356 | 0.25235 | 1.710404474 | 16.24469452 | 16.24469452 | 0.009750956 | -2.010952821 | FALSE | FALSE | FALSE |
| DHRS3 | 0.1583351 | 0.8454426 | 1 | 0.158335127 | 0.158335127 | 0.158335127 | 0.19701403 | 0.852839823 | 0.00865 | 0.197014032 | 0.197014032 | 0.197014032 | 0.803674366 | -0.094919884 | FALSE | TRUE | FALSE |
| PIGU | 0.1583153 | 0.84545665 | 1 | 0.15831533 | 0.15831533 | 3.140844126 | 0.19418306 | 0.853374801 | 1 | 0.776621536 | 0.776621536 | 0.776621536 | 0.815289091 | -0.088688369 | FALSE | TRUE | FALSE |
| THUMPD1 | 0.1583112 | 0.8454707 | 0.756 | 1.441295811 | 5.764905393 | 5.764905393 | 0.80825467 | 0.618117697 | 0.50085 | 0.808254666 | 0.808254666 | 0.808254666 | 0.195867923 | -0.708036682 | FALSE | TRUE | FALSE |
| TBX5 | 0.1582521 | 0.84548474 | 0.50405 | 0.632717987 | 0.632717987 | 3.126746749 | 0.19297552 | 0.853640949 | 1 | 1.82651951 | 1.82651951 | 3.818363994 | 0.820062879 | -0.086152847 | FALSE | TRUE | FALSE |
| RHEBL1 | 0.1582035 | 0.84549879 | 1 | 0.632708258 | 0.632708258 | 0.632708258 | 0.19833795 | 0.85251705 | 1 | 0.19833795 | 0.19833795 | 0.19833795 | 0.797646215 | -0.098189691 | FALSE | TRUE | FALSE |
| TCF20 | 0.1581155 | 0.84551283 | 1 | 0.158115526 | 3.046020273 | 5.295907162 | 0.19667402 | 0.852906895 | 1 | 0.196674023 | 0.196674023 | 6.599183627 | 0.803947182 | -0.094772483 | FALSE | TRUE | FALSE |
| CAPS2 | 0.1581113 | 0.84552686 | 0.8752 | 1.410671088 | 12.90979002 | 12.90979002 | 0.03044851 | 0.891167612 | 0.50695 | 0.26776928 | 0.26776928 | 0.26776928 | 5.192744143 | 0.715396925 | FALSE | TRUE | FALSE |
| ZFP41 | 0.1580315 | 0.8455409 | 1 | 0.158031465 | 1.446806762 | 1.446806762 | 1.80095046 | 0.507213749 | 0.50335 | 1.800950465 | 1.800950465 | 1.800950465 | 0.087748924 | -1.056758202 | FALSE | FALSE | FALSE |
| BCAR1 | 0.1580154 | 0.84555493 | 1 | 1.452920173 | 1.452920173 | 28.38383529 | 1.80926808 | 0.506596057 | 0.0287 | 1.809268076 | 1.809268076 | 35.44420264 | 0.087336653 | -1.058803457 | FALSE | FALSE | FALSE |
| NDUFS4 | 0.1580054 | 0.84556896 | 1 | 0.158005356 | 0.158005356 | 0.158005356 | 0.19008924 | 0.854328614 | 1 | 1.754811677 | 1.754811677 | 1.754811677 | 0.831216739 | -0.08028572 | FALSE | TRUE | FALSE |
| RNF144B | 0.1579739 | 0.84558298 | 1 | 0.157973948 | 0.631810344 | 0.631810344 | 0.19808573 | 0.85255747 | 1 | 0.198085731 | 0.198085731 | 0.198085731 | 0.797502918 | -0.098267719 | FALSE | TRUE | FALSE |
| LGALS4 | 0.1579314 | 0.84559701 | 1 | 0.157931417 | 0.157931417 | 3.133048879 | 0.19546009 | 0.853134508 | 1 | 0.195460087 | 0.195460087 | 0.195460087 | 0.807998296 | -0.092589555 | FALSE | TRUE | FALSE |
| FCER1G | 0.1578522 | 0.84561103 | 0.00245 | 0.157852193 | 0.157852193 | 0.157852193 | 0.1641087 | 0.861105053 | 1 | 0.164108704 | 0.164108704 | 0.164108704 | 0.961875807 | -0.016880998 | FALSE | TRUE | FALSE |
| CD33 | 0.1578396 | 0.84562505 | 1 | 1.437194977 | 13.43118901 | 13.43118901 | 1.73337621 | 0.514657856 | 0.8802 | 16.26279 | 156.4974129 | 156.4974129 | 0.091059047 | -1.040676901 | FALSE | FALSE | FALSE |
| PUM2 | 0.1578339 | 0.84563907 | 1 | 131.9057232 | 13257.86116 | 137737.4902 | 0.23550952 | 0.843179489 | 1 | 0.235509517 | 0.235509517 | 4.630443176 | 0.670180667 | -0.173808105 | FALSE | TRUE | FALSE |
| CEACAM5 | 0.1578021 | 0.84565308 | 1 | 1.441724867 | 1.441724867 | 5.339972889 | 16.3695901 | 0.151137976 | 0.2616 | 1.739529797 | 16.36959013 | 34.36910409 | 0.009639955 | -2.015925005 | FALSE | FALSE | FALSE |
| PCDHB7 | 0.1577646 | 0.84566709 | 0.3327 | 0.157764588 | 5.647893695 | 5.647893695 | 2.78080156 | 0.428657004 | 0.50655 | 2.780801556 | 103.1345062 | 980.1721387 | 0.056733494 | -1.24616047 | FALSE | FALSE | FALSE |
| CENPO | 0.1576588 | 0.8456811 | 1 | 0.157658751 | 0.157658751 | 0.157658751 | 0.19519842 | 0.853214686 | 1 | 0.195198419 | 0.195198419 | 0.195198419 | 0.807684569 | -0.092758215 | FALSE | TRUE | FALSE |
| RASL12 | 0.1575933 | 0.84569511 | 1 | 0.157593278 | 0.157593278 | 0.157593278 | 0.18582628 | 0.855555876 | 1 | 2.971674961 | 27.93695673 | 27.93695673 | 0.848067761 | -0.071569446 | FALSE | TRUE | FALSE |
| DAW1 | 0.1575542 | 0.84570911 | 1 | 0.157554239 | 0.157554239 | 3.126843728 | 0.19565698 | 0.853094387 | 1 | 0.195656976 | 0.195656976 | 3.886628309 | 0.805257456 | -0.094065245 | FALSE | TRUE | FALSE |
| OR2A4 | 0.1575366 | 0.84572311 | 0.5131 | 0.157536632 | 0.157536632 | 0.157536632 | 0.01707331 | 0.894571287 | 0.4998 | 0.017073311 | 0.162016607 | 0.162016607 | 9.227069991 | 0.965063815 | FALSE | TRUE | FALSE |

| | | | | | | | | | | | | | | | | | |
|---|---|---|---|---|---|---|---|---|---|---|---|---|---|---|---|---|---|
| MVP | 0.1574839 | 0.84573711 | 0.52045 | 1.413846918 | 13.01958441 | 27.61678418 | 0.03115575 | 0.890967986 | 1 | 0.031155746 | 0.031155746 | 0.610279429 | 5.054729677 | 0.703697935 | FALSE | TRUE | FALSE |
| CCAR2 | 0.1574234 | 0.84575111 | 1 | 0.157423407 | 0.157423407 | 3.087754135 | 0.19562434 | 0.853107763 | 1 | 0.195624341 | 0.195624341 | 3.845950119 | 0.804723006 | -0.094353583 | FALSE | TRUE | FALSE |
| DSG1 | 0.1573822 | 0.8457651 | 0.87945 | 0.629282285 | 2.516771934 | 12.34040719 | 0.92260541 | 0.595996903 | 0.51265 | 0.922605408 | 3.690177108 | 18.13501388 | 0.170584547 | -0.768060313 | FALSE | TRUE | FALSE |
| MRPL9 | 0.1571028 | 0.8457791 | 1 | 0.157102831 | 0.157102831 | 0.157102831 | 0.19450792 | 0.853348137 | 1 | 0.194507919 | 0.194507919 | 0.194507919 | 0.807693752 | -0.092753276 | FALSE | TRUE | FALSE |
| CCNK | 0.1570571 | 0.84579309 | 0.5165 | 1.400951302 | 12.81799505 | 12.81799505 | 0.02771537 | 0.891792907 | 1 | 2.187725627 | 191.1511137 | 191.1511137 | 5.666784893 | 0.753336727 | FALSE | TRUE | FALSE |
| PSPC1 | 0.1570066 | 0.84580708 | 1 | 1.48514785 | 1.48514785 | 5.316718203 | 0.1730837 | 0.858786796 | 1 | 0.173083704 | 0.173083704 | 3.419635315 | 0.907113633 | -0.042338306 | FALSE | TRUE | FALSE |
| MINPP1 | 0.1569852 | 0.84582106 | 1 | 0.156985244 | 0.156985244 | 3.101154557 | 0.19315483 | 0.853561203 | 1 | 0.772419337 | 0.772419337 | 3.821301339 | 0.812743037 | -0.090046742 | FALSE | TRUE | FALSE |
| GABPB2 | 0.1569683 | 0.84583505 | 0.01295 | 0.156968331 | 0.156968331 | 3.11402759 | 0.19468727 | 0.85328144 | 1 | 0.194687268 | 0.194687268 | 0.194687268 | 0.806258839 | -0.093525511 | FALSE | TRUE | FALSE |
| USP32 | 0.1569506 | 0.84584903 | 1 | 0.156950587 | 0.156950587 | 5.259698855 | 0.19531749 | 0.853174608 | 1 | 0.195317489 | 0.195317489 | 6.556754286 | 0.80356648 | -0.094978188 | FALSE | TRUE | FALSE |
| ALKBH1 | 0.1568512 | 0.84586301 | 0.75625 | 1.430713466 | 5.722531106 | 5.722531106 | 0.19076762 | 0.85413088 | 1 | 1.746589392 | 16.41239416 | 16.41239416 | 0.822210768 | -0.08501684 | FALSE | TRUE | FALSE |
| GPR158 | 0.1568393 | 0.84587698 | 0.33575 | 0.156839332 | 2.506252294 | 12.19918997 | 0.03140177 | 0.890891025 | 0.985 | 1.145449472 | 42.89458487 | 171.5537179 | 4.994601901 | 0.698500878 | FALSE | TRUE | FALSE |
| SCG5 | 0.1566924 | 0.84589096 | 1 | 0.156692364 | 0.156692364 | 0.156692364 | 0.19405417 | 0.853401455 | 0.0054 | 0.194054171 | 0.194054171 | 0.194054171 | 0.807467127 | -0.092875149 | FALSE | TRUE | FALSE |
| GSTM5 | 0.1565006 | 0.84590493 | 1 | 0.156500644 | 0.156500644 | 0.156500644 | 1.65520949 | 0.521606291 | 0.75145 | 1.655209491 | 15.42106825 | 15.42106825 | 0.094550354 | -1.024336839 | FALSE | FALSE | FALSE |
| RUVBL2 | 0.1564963 | 0.8459189 | 1 | 0.156496349 | 0.156496349 | 3.082768334 | 0.19497195 | 0.853254745 | 0.01455 | 0.19497195 | 0.19497195 | 3.847651191 | 0.802660841 | -0.095467924 | FALSE | TRUE | FALSE |
| CANT1 | 0.1563165 | 0.84593286 | 0.13795 | 0.156316543 | 0.624897561 | 0.77287701 | 202.246397 | 0.01321501 | 0.18495 | 12.66155654 | 808.6125095 | 2012.749809 | 0.000772901 | -3.111875852 | FALSE | FALSE | FALSE |
| RBBP6 | 0.1562978 | 0.84594683 | 0.0463 | 0.156297753 | 0.156297753 | 5.235488718 | 0.19454252 | 0.853334803 | 0.04375 | 0.194542518 | 0.194542518 | 6.528175355 | 0.80341179 | -0.095061799 | FALSE | TRUE | FALSE |
| LHX3 | 0.1561861 | 0.84596079 | 0.75215 | 0.156186147 | 0.624420798 | 3.088092849 | 0.22916162 | 0.845113633 | 0.75165 | 0.229161617 | 0.916254371 | 1.135062203 | 0.681554569 | -0.166499367 | FALSE | TRUE | FALSE |
| IRF2BPL | 0.156144 | 0.84597475 | 1 | 1.435485041 | 1.435485041 | 28.10720859 | 0.18202812 | 0.856520856 | 1 | 15.84954074 | 153.6773797 | 311.1229895 | 0.857801593 | -0.066613152 | FALSE | TRUE | FALSE |
| SNX11 | 0.1560672 | 0.84598871 | 0.0076 | 0.156067189 | 0.156067189 | 0.156067189 | 0.19365056 | 0.853454742 | 1 | 0.193650556 | 0.193650556 | 0.193650556 | 0.805921719 | -0.09370714 | FALSE | TRUE | FALSE |
| LARP1 | 0.1560104 | 0.84600267 | 1 | 0.156010415 | 0.156010415 | 3.046870382 | 0.17684342 | 0.857888753 | 1 | 15.46263532 | 150.2391869 | 302.8391643 | 0.882195183 | -0.054435318 | FALSE | TRUE | FALSE |
| SPIB | 0.1559522 | 0.84601662 | 1 | 0.155952228 | 0.155952228 | 0.155952228 | 0.17940843 | 0.85719526 | 1 | 0.179408427 | 0.179408427 | 0.179408427 | 0.869258097 | -0.060851255 | FALSE | TRUE | FALSE |
| MGAT5B | 0.155884 | 0.84603057 | 1 | 1.442138889 | 13.69320727 | 28.22406893 | 0.17670306 | 0.85792639 | 0.9713 | 151.3136658 | 1510.975467 | 15456.53458 | 0.882180541 | -0.054442526 | FALSE | TRUE | FALSE |
| CREM | 0.1558664 | 0.84604452 | 1 | 0.155866436 | 0.155866436 | 0.155866436 | 0.19377362 | 0.853414781 | 1 | 0.193773622 | 0.193773622 | 0.193773622 | 0.80437386 | -0.094542051 | FALSE | TRUE | FALSE |
| DGKB | 0.155842 | 0.84605847 | 1 | 0.155842038 | 0.155842038 | 3.073320069 | 1.75903672 | 0.512099277 | 0.5063 | 1.759036716 | 1.759036716 | 6.359768814 | 0.088595103 | -1.052590284 | FALSE | FALSE | FALSE |
| ZNF772 | 0.1557756 | 0.84607241 | 1 | 0.15577558 | 0.15577558 | 3.0884382 | 0.19346577 | 0.85350799 | 0.01125 | 0.193465771 | 0.193465771 | 3.839700373 | 0.805184188 | -0.094104763 | FALSE | TRUE | FALSE |
| DUSP12 | 0.1557284 | 0.84608635 | 1 | 0.155728358 | 0.155728358 | 3.090405734 | 0.19336712 | 0.853521297 | 0.0096 | 0.193367125 | 0.193367125 | 0.193367125 | 0.805350745 | -0.094014935 | FALSE | TRUE | FALSE |
| RHBDD1 | 0.155717 | 0.84610029 | 0.0073 | 0.155716973 | 0.155716973 | 0.155716973 | 0.1930812 | 0.853614377 | 1 | 0.193081196 | 0.193081196 | 0.193081196 | 0.806484401 | -0.093404028 | FALSE | TRUE | FALSE |
| LMTK2 | 0.1556653 | 0.84611423 | 0.69115 | 0.155665316 | 2.488664309 | 12.04217502 | 9.87014348 | 0.200653863 | 0.07665 | 0.617530109 | 9.870143475 | 35.55920204 | 0.015771333 | -1.802131608 | FALSE | FALSE | FALSE |
| GXYLT2 | 0.1556507 | 0.84612816 | 1 | 0.15565072 | 0.15565072 | 0.15565072 | 0.19309802 | 0.853587795 | 0.00885 | 0.193098023 | 0.193098023 | 0.193098023 | 0.806071018 | -0.093626693 | FALSE | TRUE | FALSE |
| FBXL22 | 0.1556235 | 0.84614209 | 0.9927 | 137.0008418 | 151089.7765 | 283080.0045 | 13.8874448 | 0.164836008 | 0.98915 | 14173.85626 | 17898562.1 | 202262234.2 | 0.011206054 | -1.950547294 | FALSE | FALSE | FALSE |
| CA5A | 0.1554947 | 0.84615602 | 1 | 0.15549469 | 0.15549469 | 0.15549469 | 0.19368198 | 0.853441424 | 1 | 0.193681984 | 0.193681984 | 0.193681984 | 0.802835075 | -0.095373662 | FALSE | TRUE | FALSE |
| STX11 | 0.1554669 | 0.84616995 | 1 | 0.155466877 | 0.155466877 | 3.07591061 | 0.18573417 | 0.855607652 | 1 | 0.185734171 | 0.185734171 | 3.679501211 | 0.837039713 | -0.077253937 | FALSE | TRUE | FALSE |
| SYNRG | 0.1554013 | 0.84618387 | 0.03355 | 0.155401328 | 0.155401328 | 3.039126366 | 1.73042624 | 0.514957154 | 0.51675 | 1.730426242 | 1.730426242 | 33.93168104 | 0.089805231 | -1.046698366 | FALSE | FALSE | FALSE |
| CHRM5 | 0.1553478 | 0.84619779 | 0.0156 | 0.155347804 | 0.155347804 | 3.075342268 | 0.19095385 | 0.854117679 | 1 | 0.763734914 | 0.763734914 | 3.784844125 | 0.813535868 | -0.089623295 | FALSE | TRUE | FALSE |
| LMBR1 | 0.1553218 | 0.84621171 | 1 | 0.155321778 | 0.155321778 | 3.078446068 | 0.19264692 | 0.853694067 | 1 | 0.192646918 | 0.192646918 | 3.822360548 | 0.806251039 | -0.093529713 | FALSE | TRUE | FALSE |
| PPFIA4 | 0.1552713 | 0.84622563 | 0.75565 | 0.155271341 | 1.417087854 | 3.055049011 | 0.25010211 | 0.83940369 | 0.26655 | 0.250102113 | 2.299591285 | 4.930506378 | 0.620831783 | -0.207026057 | FALSE | TRUE | FALSE |
| TIMM50 | 0.155267 | 0.84623954 | 1 | 0.155267002 | 0.155267002 | 3.062288563 | 0.19271752 | 0.853680791 | 1 | 0.19271752 | 0.19271752 | 3.807256414 | 0.805671442 | -0.09384203 | FALSE | TRUE | FALSE |
| TFRC | 0.1549931 | 0.84625346 | 1 | 0.619809905 | 2.479209196 | 12.21612024 | 0.19752179 | 0.85267861 | 0.01815 | 0.197521793 | 0.197521793 | 3.899854922 | 0.784688689 | -0.105302607 | FALSE | TRUE | FALSE |
| ADH1B | 0.1549132 | 0.84626737 | 1 | 0.15491316 | 0.15491316 | 3.074768145 | 0.1924417 | 0.853773675 | 1 | 0.192441701 | 0.192441701 | 0.192441701 | 0.80498748 | -0.094210874 | FALSE | TRUE | FALSE |
| TNIP1 | 0.1548729 | 0.84628128 | 1 | 0.154872852 | 0.154872852 | 3.052492148 | 0.19256223 | 0.853720613 | 1 | 0.192562234 | 0.192562234 | 3.801968313 | 0.804274276 | -0.094595822 | FALSE | TRUE | FALSE |
| GNA13 | 0.1548355 | 0.84629518 | 0.01165 | 0.154835491 | 0.154835491 | 3.065811639 | 0.16833619 | 0.860028312 | 0.9696 | 19.11224447 | 1559.243766 | 6234.258828 | 0.919799176 | -0.036306984 | FALSE | TRUE | FALSE |
| PGAM5 | 0.1547174 | 0.84630909 | 1 | 0.154717446 | 0.154717446 | 0.154717446 | 0.1922413 | 0.853839949 | 0.00585 | 0.192241299 | 0.192241299 | 0.192241299 | 0.804808574 | -0.094307405 | FALSE | TRUE | FALSE |
| B4GALT3 | 0.1547022 | 0.84632299 | 1 | 1.414850562 | 5.658971336 | 5.658971336 | 0.20045973 | 0.852003212 | 1 | 0.200459732 | 0.200459732 | 3.973717256 | 0.771736964 | -0.112530698 | FALSE | TRUE | FALSE |
| LAMB2 | 0.1546987 | 0.84633689 | 0.3588 | 0.085665232 | 5.468421097 | 49.56934106 | 0.10237602 | 0.875380048 | 0.89675 | 3.658640825 | 757.2799228 | 6048.220861 | 1.511083632 | 0.179288501 | FALSE | TRUE | FALSE |
| AIG1 | 0.1546599 | 0.84635078 | 1 | 0.154659877 | 0.154659877 | 0.154659877 | 0.19179501 | 0.853919402 | 1 | 0.191795011 | 0.191795011 | 0.191795011 | 0.806381126 | -0.093459646 | FALSE | TRUE | FALSE |
| DENND1A | 0.1546507 | 0.84636468 | 0.51545 | 1.422708726 | 1.422708726 | 27.8424802 | 0.19841217 | 0.852490091 | 1 | 0.198412171 | 0.198412171 | 3.892990871 | 0.779441807 | -0.108216304 | FALSE | TRUE | FALSE |
| C19orf47 | 0.1546268 | 0.84637857 | 1 | 0.154626815 | 0.154626815 | 3.064553969 | 0.19281533 | 0.853667513 | 1 | 0.192815333 | 0.192815333 | 3.82557229 | 0.801942524 | -0.095856757 | FALSE | TRUE | FALSE |
| FAM20B | 0.1545198 | 0.84639245 | 0.5048 | 0.154519761 | 0.154519761 | 0.766704283 | 0.86951615 | 0.603404302 | 0.4974 | 0.505624901 | 8.081785516 | 8.081785516 | 0.177707752 | -0.750293626 | FALSE | TRUE | FALSE |
| TRAF7 | 0.1545179 | 0.84640634 | 1 | 0.154517877 | 0.154517877 | 3.037423764 | 0.18441414 | 0.855866013 | 0.7539 | 1.705147281 | 6.819845209 | 25.05812086 | 0.837885203 | -0.076815479 | FALSE | TRUE | FALSE |
| GINS3 | 0.1545164 | 0.84642022 | 0.0083 | 0.154516415 | 0.154516415 | 0.154516415 | 0.19156269 | 0.853972326 | 0.0058 | 0.191562695 | 0.191562695 | 0.191562695 | 0.806610154 | -0.093336315 | FALSE | TRUE | FALSE |
| MME | 0.1544528 | 0.84643411 | 0.60265 | 0.154452841 | 5.619599745 | 22.46557768 | 0.35226825 | 0.808360617 | 0.5091 | 0.352268252 | 5.630167755 | 22.51686815 | 0.438452344 | -0.358077604 | FALSE | TRUE | FALSE |
| HDLBP | 0.1544203 | 0.84644798 | 1 | 0.154420287 | 0.154420287 | 5.20140269 | 0.19254367 | 0.853733882 | 1 | 0.192543674 | 0.192543674 | 3.750329456 | 0.802001353 | -0.095824899 | FALSE | TRUE | FALSE |
| IFRD1 | 0.1543972 | 0.84646186 | 1 | 1.429465786 | 1.429465786 | 3.059828647 | 0.19834866 | 0.852503572 | 0.01135 | 0.198348663 | 0.198348663 | 3.935160133 | 0.778412886 | -0.108789983 | FALSE | TRUE | FALSE |
| G6PC3 | 0.1543607 | 0.84647573 | 1 | 0.154360669 | 0.154360669 | 0.154360669 | 0.19213406 | 0.853866442 | 1 | 0.192134056 | 0.192134056 | 0.192134056 | 0.803400878 | -0.095067698 | FALSE | TRUE | FALSE |
| SH2B3 | 0.1543285 | 0.84648961 | 1 | 0.154328516 | 0.154328516 | 3.056053005 | 0.19207287 | 0.853879685 | 1 | 0.192072871 | 0.192072871 | 3.807999903 | 0.803489401 | -0.095019848 | FALSE | TRUE | FALSE |
| GNG13 | 0.1543023 | 0.84650347 | 1 | 0.154302294 | 0.154302294 | 0.154302294 | 1.50005763 | 0.535949241 | 0.5041 | 1.500057634 | 1.500057634 | 1.500057634 | 0.102864244 | -0.987735561 | FALSE | FALSE | FALSE |
| ENGASE | 0.1542605 | 0.84651734 | 0.51025 | 0.154260503 | 1.361273424 | 3.028884292 | 2.75686067 | 0.430475124 | 0.31805 | 0.308399953 | 25.26254916 | 25.26254916 | 0.055955132 | -1.252160075 | FALSE | FALSE | FALSE |
| ZFAND2A | 0.1542602 | 0.8465312 | 1 | 1.402180413 | 1.402180413 | 1.402180413 | 0.19740386 | 0.852759259 | 1 | 0.197403859 | 0.197403859 | 0.197403859 | 0.78144452 | -0.10710185 | FALSE | TRUE | FALSE |
| CLDN7 | 0.1542445 | 0.84654506 | 1 | 0.154244494 | 0.154244494 | 0.154244494 | 0.19234206 | 0.853813446 | 0.00585 | 0.192342061 | 0.192342061 | 0.192342061 | 0.801928051 | -0.095864595 | FALSE | TRUE | FALSE |
| CLSTN1 | 0.1542378 | 0.84655892 | 0.52385 | 0.61681036 | 0.61681036 | 12.04500248 | 0.19238859 | 0.853800192 | 0.5102 | 0.769399591 | 0.769399591 | 15.06748041 | 0.801699078 | -0.095988616 | FALSE | TRUE | FALSE |
| PHF7 | 0.154229 | 0.84657278 | 1 | 0.15422896 | 0.15422896 | 0.15422896 | 0.75964116 | 0.636766678 | 0.5018 | 0.759641157 | 0.759641157 | 0.759641157 | 0.203028705 | -0.692442556 | FALSE | TRUE | FALSE |
| TMEM80 | 0.1541912 | 0.84658663 | 1 | 0.154191249 | 0.154191249 | 0.154191249 | 0.18968567 | 0.854407561 | 1 | 0.758594228 | 0.758594228 | 0.758594228 | 0.812877704 | -0.089974789 | FALSE | TRUE | FALSE |
| STAT3 | 0.1541576 | 0.84660048 | 1 | 0.154157591 | 0.154157591 | 3.033660329 | 1.69876352 | 0.517043603 | 0.7521 | 1.698763522 | 16.39338644 | 33.49448843 | 0.09074694 | -1.042168013 | FALSE | FALSE | FALSE |
| SIK2 | 0.1541342 | 0.84661433 | 0.0236 | 0.154134218 | 0.154134218 | 3.027214332 | 0.76034329 | 0.636158493 | 0.50835 | 0.760343291 | 0.760343291 | 14.96558635 | 0.202716615 | -0.693110653 | FALSE | TRUE | FALSE |
| CMC2 | 0.1540459 | 0.84662818 | 1 | 0.154045869 | 0.154045869 | 0.154045869 | 1.44732657 | 0.54087401 | 0.50015 | 4.213211728 | 134.2855522 | 134.2855522 | 0.106434769 | -0.972916479 | FALSE | FALSE | FALSE |
| ADAMTSL3 | 0.1540246 | 0.84664202 | 0.89405 | 2.865506259 | 183.4798678 | 734.4028032 | 0.20729646 | 0.850371533 | 0.5199 | 0.384445284 | 13.26774486 | 53.09852515 | 0.743015811 | -0.129001945 | FALSE | TRUE | FALSE |
| RNF148 | 0.1539832 | 0.84665586 | 1 | 0.153983216 | 0.153983216 | 0.153983216 | 0.19127244 | 0.85403843 | 1 | 0.191272436 | 0.191272436 | 0.191272436 | 0.805046558 | -0.094179002 | FALSE | TRUE | FALSE |
| LYPLAL1 | 0.1539413 | 0.8466697 | 1 | 0.153941346 | 0.153941346 | 0.153941346 | 0.19153354 | 0.853985551 | 0.0051 | 0.191533537 | 0.191533537 | 0.191533537 | 0.803730501 | -0.09488955 | FALSE | TRUE | FALSE |
| PROCR | 0.1538742 | 0.84668354 | 1 | 0.153874203 | 0.153874203 | 0.153874203 | 0.18934195 | 0.8545127 | 1 | 0.757245288 | 0.757245288 | 0.757245288 | 0.812678869 | -0.090081032 | FALSE | TRUE | FALSE |
| CARD11 | 0.1538721 | 0.84669737 | 1 | 0.153872111 | 0.153872111 | 2.983284703 | 1.6977295 | 0.517192096 | 0.0342 | 1.697729505 | 1.697729505 | 33.03715719 | 0.090634056 | -1.042708584 | FALSE | FALSE | FALSE |
| TEX26 | 0.1538045 | 0.8467112 | 0.0067 | 0.153804491 | 0.153804491 | 0.153804491 | 1.58796451 | 0.527669206 | 0.87525 | 1.587964507 | 142.3290002 | 142.3290002 | 0.096856378 | -1.013871774 | FALSE | FALSE | FALSE |
| TRIM47 | 0.1537726 | 0.84672503 | 0.50975 | 1.402390667 | 1.402390667 | 3.041081026 | 1.69693306 | 0.517488776 | 0.75465 | 1.696933064 | 15.94331317 | 15.94331317 | 0.09061795 | -1.042785766 | FALSE | FALSE | FALSE |
| SETD9 | 0.153724 | 0.84673886 | 1 | 1.392459209 | 1.392459209 | 1.392459209 | 0.1966528 | 0.852920303 | 1 | 0.196652796 | 0.196652796 | 0.196652796 | 0.781702346 | -0.106958585 | FALSE | TRUE | FALSE |

| | | | | | | | | | | | | | | | | | |
|---|---|---|---|---|---|---|---|---|---|---|---|---|---|---|---|---|---|
| NKX6-2 | 0.1536257 | 0.84675268 | 1 | 5.74356453 | 220.442705 | 220.442705 | 0.20876112 | 0.850038274 | 1 | 0.208761117 | 0.208761117 | 0.208761117 | 0.735892314 | -0.133185733 | FALSE | TRUE | FALSE |
| SNRNP35 | 0.1535818 | 0.84676651 | 1 | 1.406368691 | 5.624776449 | 5.624776449 | 0.19920837 | 0.852328142 | 1 | 0.199208372 | 0.199208372 | 0.199208372 | 0.770960652 | -0.112967787 | FALSE | TRUE | FALSE |
| STAC3 | 0.1535339 | 0.84678033 | 0.505 | 0.153533875 | 0.153533875 | 0.761471454 | 0.50615998 | 0.753370937 | 0.502 | 0.506159979 | 0.506159979 | 0.506159979 | 0.303330727 | -0.518083594 | FALSE | TRUE | FALSE |
| RPL36AL | 0.1535202 | 0.84679414 | 1 | 0.153520223 | 0.153520223 | 0.153520223 | 0.1623024 | 0.861524211 | 1 | 0.162302402 | 0.162302402 | 0.162302402 | 0.945890022 | -0.024159356 | FALSE | TRUE | FALSE |
| FBXO27 | 0.1534701 | 0.84680796 | 0.00905 | 0.153470135 | 0.153470135 | 0.153470135 | 0.19049372 | 0.85419685 | 1 | 0.190493719 | 0.190493719 | 0.190493719 | 0.805644068 | -0.093856786 | FALSE | TRUE | FALSE |
| CACNA2D3 | 0.1534127 | 0.84682177 | 1 | 0.153412692 | 0.153412692 | 3.004438045 | 1.72331036 | 0.515853005 | 0.5126 | 1.723310356 | 1.723310356 | 33.83389958 | 0.089022092 | -1.050502206 | FALSE | FALSE | FALSE |
| POM121L12 | 0.1533935 | 0.84683558 | 1 | 0.153393458 | 0.153393458 | 3.040643898 | 0.19043247 | 0.854223222 | 0.01025 | 0.190432469 | 0.190432469 | 3.778879248 | 0.805500546 | -0.093934161 | FALSE | TRUE | FALSE |
| CKM | 0.1533828 | 0.84684939 | 0.01215 | 0.153382818 | 0.153382818 | 3.035657096 | 0.1910976 | 0.854064856 | 1 | 0.191097596 | 0.191097596 | 3.786830314 | 0.802641273 | -0.095478512 | FALSE | TRUE | FALSE |
| PRDM16 | 0.1533803 | 0.84686319 | 1 | 0.61339313 | 0.61339313 | 11.84125049 | 0.19363326 | 0.853468057 | 0.0375 | 0.19363326 | 0.19363326 | 3.753677009 | 0.792117378 | -0.101210459 | FALSE | TRUE | FALSE |
| SNRPG | 0.1533514 | 0.84687699 | 1 | 0.153351426 | 0.153351426 | 0.153351426 | 0.16088737 | 0.861834046 | 0.00195 | 0.160887366 | 0.160887366 | 0.160887366 | 0.953160155 | -0.020834121 | FALSE | TRUE | FALSE |
| TPI1 | 0.1533478 | 0.84689079 | 0.75545 | 0.153347792 | 0.613083188 | 0.613083188 | 0.89054488 | 0.59983811 | 0.1309 | 0.890544877 | 3.561583425 | 3.561583425 | 0.172195468 | -0.763978282 | FALSE | TRUE | FALSE |
| BNIPL | 0.1531692 | 0.84690459 | 1 | 0.153169161 | 0.153169161 | 0.153169161 | 0.1845227 | 0.855840216 | 1 | 1.680161885 | 1.680161885 | 1.680161885 | 0.830083022 | -0.080878469 | FALSE | TRUE | FALSE |
| SLC38A6 | 0.1531137 | 0.84691839 | 0.8772 | 1.338915561 | 12.00963536 | 12.00963536 | 2.15124043 | 0.476042255 | 0.49985 | 0.955614193 | 19.52932772 | 19.52932772 | 0.071174593 | -1.14767501 | FALSE | FALSE | FALSE |
| UGT1A1 | 0.1530645 | 0.84693218 | 1 | 0.153064473 | 0.153064473 | 3.026731854 | 0.17943939 | 0.857182591 | 1 | 1.671179278 | 15.97412235 | 33.09116097 | 0.853014912 | -0.069043377 | FALSE | TRUE | FALSE |
| WNT5B | 0.1530556 | 0.84694597 | 0.62665 | 0.611956507 | 5.667485934 | 5.667485934 | 0.23198667 | 0.844444321 | 0.7566 | 0.231986673 | 0.927618313 | 1.148876395 | 0.659760473 | -0.180613707 | FALSE | TRUE | FALSE |
| NFS1 | 0.1530207 | 0.84695976 | 0.50925 | 0.153020664 | 0.153020664 | 0.758223729 | 3.48061525 | 0.383894082 | 0.32405 | 0.870342927 | 13.922914 | 13.922914 | 0.043963682 | -1.356905938 | FALSE | FALSE | FALSE |
| RNMT | 0.1528684 | 0.84697354 | 1 | 1.403833738 | 1.403833738 | 3.032314252 | 16.0741648 | 0.153065255 | 0.25375 | 1.700387008 | 16.07416483 | 16.07416483 | 0.009510195 | -2.021810573 | FALSE | FALSE | FALSE |
| ZFPM1 | 0.1527939 | 0.84698733 | 1 | 14467749.8 | 17973316372 | 2.51575E+12 | 0.16309366 | 0.861344869 | 1 | 25580840478 | 4.8989E+13 | 7.73724E+15 | 0.936847512 | -0.028331092 | FALSE | TRUE | FALSE |
| CCL4 | 0.1527868 | 0.84700111 | 1 | 0.152786765 | 0.152786765 | 0.152786765 | 0.15925974 | 0.862249053 | 1 | 0.159259738 | 0.159259738 | 0.159259738 | 0.959355876 | -0.01802026 | FALSE | TRUE | FALSE |
| RIF1 | 0.1527776 | 0.84701489 | 0.9694 | 13.56813662 | 260.7062391 | 2550.543787 | 2.55206782 | 0.443885737 | 0.26975 | 0.272092638 | 2.55206782 | 49.26940603 | 0.059864235 | -1.222832562 | FALSE | FALSE | FALSE |
| OVCA2 | 0.1526314 | 0.84702867 | 1 | 0.152631385 | 0.152631385 | 0.152631385 | 0.18984307 | 0.854381254 | 1 | 0.189843073 | 0.189843073 | 0.189843073 | 0.80398712 | -0.094750909 | FALSE | TRUE | FALSE |
| DOT1L | 0.1525166 | 0.84704244 | 1 | 0.152516555 | 2.922777499 | 5.132302412 | 0.19022346 | 0.85428911 | 1 | 0.190223456 | 0.190223456 | 6.40907813 | 0.801775757 | -0.09594708 | FALSE | TRUE | FALSE |
| OLA1 | 0.1525126 | 0.84705621 | 1 | 2.90724E+13 | 7.86177E+17 | 1.44312E+20 | 0.04945168 | 0.886807729 | 1 | 7.97635E+39 | 2.91092E+47 | 1.88478E+51 | 3.084074204 | 0.489124819 | FALSE | TRUE | FALSE |
| LDLRAD2 | 0.1523965 | 0.84706998 | 1 | 0.152396521 | 0.152396521 | 0.152396521 | 0.18381556 | 0.856020614 | 1 | 1.679072085 | 1.679072085 | 1.679072085 | 0.829073007 | -0.081407224 | FALSE | TRUE | FALSE |
| CNOT10 | 0.1522754 | 0.84708375 | 1 | 0.152275385 | 0.152275385 | 3.008026461 | 6.50992977 | 0.255137055 | 0.25725 | 6.509929774 | 61.61639883 | 61.61639883 | 0.023391248 | -1.630946599 | FALSE | FALSE | FALSE |
| SLC10A4 | 0.1522719 | 0.84709752 | 1 | 0.152271941 | 0.152271941 | 0.152271941 | 0.1893665 | 0.854473291 | 1 | 0.189366498 | 0.189366498 | 0.189366498 | 0.804112356 | -0.094683265 | FALSE | TRUE | FALSE |
| IFI44L | 0.1521838 | 0.84711128 | 1 | 0.152183848 | 0.152183848 | 0.152183848 | 0.18352214 | 0.856097795 | 1 | 1.67287781 | 1.67287781 | 1.67287781 | 0.829239706 | -0.081319911 | FALSE | TRUE | FALSE |
| HAS3 | 0.1521568 | 0.84712504 | 1 | 1.409982132 | 13.40996396 | 27.78789076 | 0.20204401 | 0.851622388 | 1 | 0.202044008 | 0.202044008 | 3.988875551 | 0.753087558 | -0.123154527 | FALSE | TRUE | FALSE |
| DKK3 | 0.1520538 | 0.8471388 | 0.0121 | 0.152053797 | 0.152053797 | 3.017834999 | 0.1889932 | 0.854643908 | 0.00885 | 0.188993203 | 0.188993203 | 0.188993203 | 0.804546378 | -0.094448916 | FALSE | TRUE | FALSE |
| CCL22 | 0.1520309 | 0.84715256 | 1 | 0.152030948 | 0.152030948 | 0.152030948 | 0.16150977 | 0.861667376 | 1 | 0.161509771 | 0.161509771 | 0.161509771 | 0.941311145 | -0.026266799 | FALSE | TRUE | FALSE |
| NR5A1 | 0.1519611 | 0.84716631 | 1 | 0.151961147 | 0.151961147 | 3.003290361 | 0.19003974 | 0.854341777 | 1 | 0.190039738 | 0.190039738 | 3.761200813 | 0.799628268 | -0.097111862 | FALSE | TRUE | FALSE |
| CLCA2 | 0.1519354 | 0.84718006 | 0.5124 | 0.151935389 | 0.151935389 | 2.990758927 | 0.02630806 | 0.8920654 | 0.8806 | 2.039186087 | 18.65621861 | 40.21642335 | 5.775242144 | 0.761570198 | FALSE | TRUE | FALSE |
| STX6 | 0.151885 | 0.84719381 | 0.00755 | 0.151885015 | 0.151885015 | 0.151885015 | 0.18933757 | 0.854525832 | 1 | 0.189337575 | 0.189337575 | 0.189337575 | 0.802191618 | -0.09572188 | FALSE | TRUE | FALSE |
| MYB | 0.1518697 | 0.84720756 | 1 | 0.151869725 | 0.151869725 | 2.993885156 | 0.18913423 | 0.854617685 | 0.017 | 0.189134227 | 0.189134227 | 3.73492712 | 0.80297325 | -0.095298922 | FALSE | TRUE | FALSE |
| USP21 | 0.1518676 | 0.8472213 | 1 | 1.414307463 | 1.414307463 | 27.87651805 | 0.18036782 | 0.856979596 | 0.021 | 0.180367818 | 0.180367818 | 3.561312061 | 0.841988242 | -0.074693973 | FALSE | TRUE | FALSE |
| KCNF1 | 0.1518359 | 0.84723504 | 0.7444 | 0.151835882 | 0.606950619 | 2.989697601 | 0.22629078 | 0.845762708 | 0.51455 | 0.226290781 | 0.226290781 | 1.116890157 | 0.67097688 | -0.173292444 | FALSE | TRUE | FALSE |
| SPRED1 | 0.1518031 | 0.84724878 | 1 | 0.151803124 | 0.151803124 | 3.009072858 | 0.18980926 | 0.854394409 | 1 | 0.18980926 | 0.18980926 | 3.766461088 | 0.799766691 | -0.097036687 | FALSE | TRUE | FALSE |
| POMT1 | 0.151774 | 0.84726252 | 0.14705 | 0.018019068 | 1.282935274 | 5.245369862 | 1.842E-05 | 0.900570765 | 0.758 | 0.001447707 | 2.262651658 | 9.0422036 | 8239.805847 | 3.915916979 | FALSE | TRUE | FALSE |
| ITGBL1 | 0.1516006 | 0.84727625 | 0.63175 | 0.151600633 | 5.619317339 | 5.619317339 | 0.13391929 | 0.867973414 | 0.50245 | 0.133919292 | 0.535404681 | 0.535404681 | 1.132029825 | 0.053857869 | FALSE | TRUE | FALSE |
| ZNF80 | 0.1515113 | 0.84728999 | 1 | 0.151511268 | 0.151511268 | 0.151511268 | 1.68259131 | 0.518672158 | 0.00735 | 1.682591311 | 1.682591311 | 1.682591311 | 0.090046387 | -1.04553371 | FALSE | FALSE | FALSE |
| CCDC83 | 0.1514794 | 0.84730372 | 1 | 0.151479379 | 0.151479379 | 3.007274332 | 0.18831879 | 0.854879496 | 1 | 0.188318789 | 0.188318789 | 0.188318789 | 0.804377407 | -0.094540136 | FALSE | TRUE | FALSE |
| CAMKMT | 0.1514779 | 0.84731745 | 0.00675 | 0.151477942 | 0.151477942 | 0.151477942 | 0.18825854 | 0.854944799 | 0.0059 | 0.188258541 | 0.188258541 | 0.188258541 | 0.804627196 | -0.094405293 | FALSE | TRUE | FALSE |
| SIX1 | 0.1514367 | 0.84733117 | 1 | 0.151436739 | 0.151436739 | 3.003624949 | 0.18939867 | 0.85446015 | 1 | 0.189398667 | 0.189398667 | 3.760314528 | 0.799566022 | -0.09714567 | FALSE | TRUE | FALSE |
| METTL14 | 0.1513427 | 0.84734489 | 1 | 0.151342709 | 0.151342709 | 3.000074837 | 0.18836025 | 0.854853357 | 0.0124 | 0.188360248 | 0.188360248 | 3.737848678 | 0.803474784 | -0.095027749 | FALSE | TRUE | FALSE |
| GOLGA1 | 0.1513397 | 0.84735862 | 1 | 0.15133973 | 0.15133973 | 2.986035963 | 0.18857565 | 0.854748707 | 1 | 0.188575648 | 0.188575648 | 3.726755343 | 0.802541219 | -0.095532652 | FALSE | TRUE | FALSE |
| NUDT13 | 0.1513057 | 0.84737233 | 0.499 | 0.037829873 | 0.151305724 | 0.151305724 | 0.46223623 | 0.77019934 | 0.7526 | 0.462236226 | 4.187019203 | 4.187019203 | 0.327334198 | -0.48500862 | FALSE | TRUE | FALSE |
| KATNB1 | 0.1511674 | 0.84738605 | 0.02205 | 0.151167444 | 0.151167444 | 2.970491357 | 0.18883284 | 0.854696326 | 1 | 0.188832843 | 0.188832843 | 3.718439753 | 0.800535763 | -0.096619262 | FALSE | TRUE | FALSE |
| GGN | 0.15115 | 0.84739976 | 1 | 0.151150004 | 0.151150004 | 2.970255106 | 0.18824751 | 0.854957852 | 1 | 0.188247509 | 0.188247509 | 3.707030677 | 0.802932292 | -0.095321076 | FALSE | TRUE | FALSE |
| ZC3H7B | 0.151134 | 0.84741347 | 1 | 0.604524054 | 0.604524054 | 11.78118884 | 0.19066493 | 0.854144079 | 1 | 0.190664927 | 0.190664927 | 3.727204241 | 0.792668182 | -0.100908574 | FALSE | TRUE | FALSE |
| CRIP2 | 0.1511149 | 0.84742718 | 0.0046 | 0.151114911 | 0.151114911 | 0.151114911 | 0.18661496 | 0.855374371 | 0.50295 | 0.74623643 | 0.74623643 | 0.74623643 | 0.80976845 | -0.091639148 | FALSE | TRUE | FALSE |
| HSPB2 | 0.1511067 | 0.84744089 | 1 | 0.151106663 | 0.151106663 | 0.151106663 | 0.18681652 | 0.855309443 | 0.5025 | 0.746982697 | 0.746982697 | 0.746982697 | 0.808850656 | -0.092131658 | FALSE | TRUE | FALSE |
| SDHD | 0.1510944 | 0.84745459 | 1 | 1.425681007 | 1.425681007 | 1.425681007 | 16.1814802 | 0.152101777 | 0.5036 | 16.18148022 | 160.5542274 | 160.5542274 | 0.009337489 | -2.029769914 | FALSE | FALSE | FALSE |
| DLG1 | 0.1510761 | 0.84746829 | 0.0242 | 0.15107611 | 0.15107611 | 2.969501385 | 0.17745903 | 0.857649928 | 1 | 1.659341619 | 15.9243127 | 32.6830677 | 0.85132952 | -0.069902307 | FALSE | TRUE | FALSE |
| PRICKLE4 | 0.1510599 | 0.84748199 | 0.49885 | 0.151059935 | 5.531124619 | 5.531124619 | 0.2491692 | 0.839657751 | 0.63315 | 0.442925039 | 3.983876858 | 3.983876858 | 0.606254444 | -0.217345065 | FALSE | TRUE | FALSE |
| ZNF425 | 0.1510035 | 0.84749569 | 0.2653 | 0.037224313 | 0.318283215 | 1.257893456 | 0.00864809 | 0.897132045 | 1 | 0.00864809 | 0.00864809 | 0.170043695 | 17.46090377 | 1.242066719 | FALSE | TRUE | FALSE |
| SYK | 0.1509611 | 0.84750938 | 1 | 0.150961144 | 0.150961144 | 2.976240361 | 0.18885951 | 0.854670122 | 1 | 0.188859507 | 0.188859507 | 3.729796484 | 0.79933039 | -0.097273675 | FALSE | TRUE | FALSE |
| TMEM131 | 0.1508797 | 0.84752308 | 1 | 0.15087965 | 0.15087965 | 5.058917398 | 0.18848879 | 0.854787969 | 1 | 0.188488794 | 0.188488794 | 6.33045794 | 0.800470135 | -0.096654867 | FALSE | TRUE | FALSE |
| ORC5 | 0.1508633 | 0.84753676 | 1 | 0.150863317 | 0.150863317 | 2.994194715 | 0.18768102 | 0.855088258 | 1 | 0.187681025 | 0.187681025 | 0.187681025 | 0.803828288 | -0.094836714 | FALSE | TRUE | FALSE |
| GRM4 | 0.1508489 | 0.84755045 | 0.2064 | 0.037748689 | 0.318994074 | 0.734494659 | 0.00148661 | 0.899229412 | 1 | 0.005937546 | 0.005937546 | 0.11590748 | 101.4715127 | 2.006344135 | FALSE | TRUE | FALSE |
| GNG10 | 0.1508126 | 0.84756414 | 1 | 0.150812645 | 0.150812645 | 0.150812645 | 1.60061146 | 0.525952059 | 0.00115 | 0.168915757 | 1.600611456 | 1.600611456 | 0.094221895 | -1.025848163 | FALSE | FALSE | FALSE |
| CLDN10 | 0.1507928 | 0.84757782 | 1 | 0.150792846 | 0.150792846 | 0.150792846 | 0.18639724 | 0.855426274 | 0.50705 | 0.745337238 | 0.745337238 | 0.745337238 | 0.808986482 | -0.092058735 | FALSE | TRUE | FALSE |
| DBN1 | 0.150776 | 0.8475915 | 1 | 0.150776023 | 0.150776023 | 2.972472547 | 0.1883046 | 0.854892561 | 0.0164 | 0.188304602 | 0.188304602 | 3.718707406 | 0.800702804 | -0.096528651 | FALSE | TRUE | FALSE |
| GUK1 | 0.1507524 | 0.84760517 | 1 | 0.150752439 | 0.150752439 | 0.150752439 | 0.74242201 | 0.644977074 | 0.5032 | 0.742422006 | 0.742422006 | 0.742422006 | 0.203054917 | -0.692386489 | FALSE | TRUE | FALSE |
| ZC3H4 | 0.1507262 | 0.84761885 | 1 | 0.150726229 | 0.150726229 | 5.070235504 | 0.18824652 | 0.854970903 | 1 | 0.188246521 | 0.188246521 | 6.340464417 | 0.800685335 | -0.096538126 | FALSE | TRUE | FALSE |
| MYT1 | 0.1506044 | 0.84763252 | 0.03565 | 0.150604442 | 0.150604442 | 2.934569173 | 0.18829241 | 0.854918685 | 1 | 0.188292414 | 0.188292414 | 3.680309472 | 0.799843385 | -0.096995043 | FALSE | TRUE | FALSE |
| TSC22D1 | 0.1505783 | 0.84764619 | 0.75045 | 1.392214067 | 13.21120239 | 27.30667452 | 0.18807804 | 0.855010041 | 1 | 1.744324695 | 16.60377444 | 34.29246712 | 0.800616174 | -0.096575641 | FALSE | TRUE | FALSE |
| CCNI2 | 0.1504889 | 0.84765986 | 1 | 0.150488873 | 0.150488873 | 0.150488873 | 0.1871482 | 0.855205437 | 1 | 0.187148197 | 0.187148197 | 0.187148197 | 0.804116071 | -0.094681258 | FALSE | TRUE | FALSE |
| IMPG1 | 0.150203 | 0.84767352 | 0.39175 | 0.071024196 | 0.600771852 | 1.268945085 | 0.27764459 | 0.83160238 | 0.5041 | 0.277644594 | 4.443129491 | 4.443129491 | 0.540990137 | -0.266810652 | FALSE | TRUE | FALSE |
| AGFG2 | 0.1501287 | 0.84768719 | 1 | 0.150128691 | 0.150128691 | 2.976113321 | 0.18692812 | 0.855257458 | 1 | 0.186928122 | 0.186928122 | 0.186928122 | 0.803135927 | -0.095210946 | FALSE | TRUE | FALSE |
| HOXB1 | 0.150109 | 0.84770085 | 1 | 1.405961692 | 1.405961692 | 2.978479869 | 0.1916451 | 0.853959098 | 0.50235 | 0.766316789 | 0.766316789 | 0.766316789 | 0.783265273 | -0.106091128 | FALSE | TRUE | FALSE |
| ZNF280B | 0.1500764 | 0.84771451 | 1 | 171299755.3 | 2.55907E+11 | 3.04792E+12 | 0.16554274 | 0.860816245 | 0.99995 | 30670636298 | 5.25229E+13 | 8.29013E+15 | 0.906571706 | -0.042597839 | FALSE | TRUE | FALSE |
| CIRBP | 0.1498561 | 0.84772817 | 0.0066 | 0.14985606 | 0.14985606 | 0.14985606 | 0.18676365 | 0.855335421 | 1 | 0.186763654 | 0.186763654 | 0.186763654 | 0.80238342 | -0.095618054 | FALSE | TRUE | FALSE |
| PPP2R5A | 0.1495382 | 0.84774182 | 0.01075 | 0.149538185 | 0.149538185 | 2.964309545 | 0.71494125 | 0.658606345 | 0.9396 | 2.859818637 | 45.79313061 | 45.79313061 | 0.2091615 | -0.679518253 | FALSE | TRUE | FALSE |

| | | | | | | | | | | | | | | | | | |
|---|---|---|---|---|---|---|---|---|---|---|---|---|---|---|---|---|---|
| PPP1R13L | 0.1494989 | 0.84775548 | 1 | 0.149498876 | 0.149498876 | 2.923159509 | 0.18114854 | 0.856788805 | 0.50865 | 1.676606532 | 1.676606532 | 32.87103275 | 0.825283361 | -0.083396911 | FALSE | TRUE | FALSE |
| HDAC11 | 0.1494385 | 0.84776913 | 1 | 0.597578144 | 2.390207431 | 2.390207431 | 0.18903376 | 0.854630798 | 0.50765 | 0.755941784 | 0.755941784 | 3.749176102 | 0.790538483 | -0.102076984 | FALSE | TRUE | FALSE |
| PAF1 | 0.1493963 | 0.84778278 | 0.01785 | 0.149396328 | 0.149396328 | 2.951707295 | 0.17790019 | 0.85748606 | 1 | 0.177900191 | 0.177900191 | 3.520000597 | 0.839776097 | -0.075836491 | FALSE | TRUE | FALSE |
| POLR2K | 0.1493765 | 0.84779643 | 1 | 0.14937648 | 0.14937648 | 0.14937648 | 0.15483582 | 0.863305424 | 1 | 0.154835824 | 0.154835824 | 0.154835824 | 0.964741078 | -0.015589229 | FALSE | TRUE | FALSE |
| BCAR3 | 0.1493749 | 0.84781007 | 0.7595 | 0.149374883 | 0.597523607 | 2.931690424 | 0.05527899 | 0.885594774 | 0.5141 | 0.22107591 | 0.22107591 | 4.348682968 | 2.702199712 | 0.431717443 | FALSE | TRUE | FALSE |
| WBP1L | 0.1493708 | 0.84782372 | 1 | 0.149370786 | 0.149370786 | 0.149370786 | 0.17126109 | 0.859268702 | 1 | 0.171261089 | 0.171261089 | 0.171261089 | 0.872181687 | -0.059393036 | FALSE | TRUE | FALSE |
| ZNF507 | 0.1493664 | 0.84783736 | 0.02535 | 0.149366425 | 0.149366425 | 2.9349574 | 0.18646022 | 0.855400327 | 1 | 0.186460216 | 0.186460216 | 3.6715606 | 0.801063243 | -0.096333195 | FALSE | TRUE | FALSE |
| USP35 | 0.1491487 | 0.84785099 | 0.87755 | 1.320261283 | 11.98764018 | 25.83779078 | 0.02987486 | 0.891274832 | 1 | 0.029874858 | 0.029874858 | 0.586170659 | 4.992447636 | 0.698313518 | FALSE | TRUE | FALSE |
| RFC2 | 0.1490719 | 0.84786463 | 1 | 0.149071921 | 0.59618034 | 0.59618034 | 0.18832312 | 0.854866428 | 0.00985 | 0.188323118 | 0.188323118 | 0.188323118 | 0.791575259 | -0.101507788 | FALSE | TRUE | FALSE |
| SALL4 | 0.1490697 | 0.84787826 | 0.0341 | 0.149069695 | 0.149069695 | 2.906498858 | 0.18639532 | 0.855439244 | 1 | 0.186395322 | 0.186395322 | 3.645260254 | 0.799750196 | -0.097045645 | FALSE | TRUE | FALSE |
| ARHGAP27 | 0.1490172 | 0.84789189 | 0.02115 | 0.149017225 | 0.149017225 | 2.934190201 | 0.18621542 | 0.85547814 | 1 | 0.186215422 | 0.186215422 | 3.673467744 | 0.800241052 | -0.096779173 | FALSE | TRUE | FALSE |
| SLC4A8 | 0.148998 | 0.84790552 | 0.6936 | 0.148997996 | 2.381728297 | 5.025693 | 0.06626938 | 0.88328088 | 0.2659 | 0.066269382 | 0.264889828 | 1.301783077 | 2.248368571 | 0.351867506 | FALSE | TRUE | FALSE |
| RASGRF1 | 0.148855 | 0.84791915 | 1 | 0.148854961 | 0.148854961 | 2.883364773 | 0.18637005 | 0.855452211 | 1 | 0.186370049 | 0.186370049 | 3.623750992 | 0.798706451 | -0.097612808 | FALSE | TRUE | FALSE |
| TRNAU1AP | 0.1488416 | 0.84793277 | 0.0075 | 0.148841624 | 0.148841624 | 0.148841624 | 0.16418175 | 0.861069014 | 1 | 0.164181753 | 0.164181753 | 0.164181753 | 0.906566176 | -0.042600488 | FALSE | TRUE | FALSE |
| TNRC6B | 0.1488184 | 0.8479464 | 0.75815 | 1.383809256 | 13.20645335 | 255.7986449 | 0.169649 | 0.85980859 | 0.96995 | 147.2506439 | 1477.033576 | 15177.3976 | 0.877213584 | -0.056894652 | FALSE | TRUE | FALSE |
| PTPRT | 0.1488006 | 0.84796001 | 0.28165 | 0.148800553 | 0.595034408 | 11.49885097 | 0.05442542 | 0.885787382 | 1 | 0.508273526 | 0.508273526 | 9.862233445 | 2.734026878 | 0.43680278 | FALSE | TRUE | FALSE |
| LNX2 | 0.1487993 | 0.84797363 | 1 | 0.148799318 | 0.148799318 | 2.936477002 | 0.18596304 | 0.855504061 | 1 | 0.185963039 | 0.185963039 | 3.67574665 | 0.800155337 | -0.096825694 | FALSE | TRUE | FALSE |
| ZNF18 | 0.148794 | 0.84798725 | 1 | 0.148794022 | 0.148794022 | 2.947975822 | 0.18562249 | 0.85564646 | 1 | 0.185622493 | 0.185622493 | 3.681786344 | 0.801594781 | -0.096045119 | FALSE | TRUE | FALSE |
| SHC1 | 0.1487672 | 0.84800086 | 1 | 1.37459164 | 1.37459164 | 2.934001413 | 0.1918885 | 0.853906165 | 1 | 0.191888499 | 0.191888499 | 3.790772548 | 0.775279342 | -0.110541788 | FALSE | TRUE | FALSE |
| CSE1L | 0.1487465 | 0.84801447 | 1 | 0.594766307 | 0.594766307 | 11.69868042 | 0.18856445 | 0.854761797 | 1 | 0.188564446 | 0.188564446 | 3.716325637 | 0.788836648 | -0.103012921 | FALSE | TRUE | FALSE |
| SUGP2 | 0.1487284 | 0.84802807 | 0.9999 | 1404934.213 | 556871742.9 | 24419369663 | 0.19774345 | 0.852638251 | 1 | 33118.82307 | 22808232.98 | 255473580.7 | 0.752128238 | -0.123708106 | FALSE | TRUE | FALSE |
| IQGAP1 | 0.1487267 | 0.84804168 | 0.51535 | 0.148726703 | 0.594712728 | 4.987673494 | 0.14932415 | 0.864735691 | 0.51805 | 0.149324149 | 0.149324149 | 2.905842452 | 0.995999001 | -0.001741097 | FALSE | TRUE | FALSE |
| WNT2 | 0.1487251 | 0.84805528 | 1 | 1.398614171 | 1.398614171 | 2.949516635 | 0.19232476 | 0.853826699 | 0.01005 | 0.192324762 | 0.192324762 | 3.818037855 | 0.773302008 | -0.111650862 | FALSE | TRUE | FALSE |
| RTCB | 0.1485995 | 0.84806888 | 0.75585 | 1.353572852 | 12.6544973 | 26.79855379 | 0.19707971 | 0.852826401 | 1 | 0.197079705 | 0.197079705 | 3.906590994 | 0.754006935 | -0.12262466 | FALSE | TRUE | FALSE |
| HAND1 | 0.1484875 | 0.84808248 | 0.00855 | 0.148487515 | 0.148487515 | 0.148487515 | 1.70510721 | 0.516597596 | 0.0072 | 1.705107207 | 1.705107207 | 1.705107207 | 0.087083976 | -1.060061751 | FALSE | FALSE | FALSE |
| TRAFD1 | 0.1482004 | 0.84809607 | 1 | 0.148200385 | 0.148200385 | 2.932241634 | 0.1849253 | 0.855762771 | 1 | 0.184925298 | 0.184925298 | 3.663585396 | 0.801406764 | -0.096146996 | FALSE | TRUE | FALSE |
| RHBDL3 | 0.1481851 | 0.84810967 | 1 | 0.148185111 | 0.148185111 | 2.937303672 | 0.18515732 | 0.855711098 | 0.01215 | 0.185157316 | 0.185157316 | 3.674093199 | 0.800320044 | -0.096736306 | FALSE | TRUE | FALSE |
| THRB | 0.1481755 | 0.84812326 | 1 | 0.148175473 | 0.148175473 | 2.933553478 | 0.18583074 | 0.855542926 | 1 | 0.185830743 | 0.185830743 | 3.683519741 | 0.797367921 | -0.09834124 | FALSE | TRUE | FALSE |
| ZNF490 | 0.1481403 | 0.84813685 | 1 | 0.148140303 | 0.148140303 | 2.932988659 | 0.18517656 | 0.855698175 | 1 | 0.185176558 | 0.185176558 | 3.670697496 | 0.799994906 | -0.096912779 | FALSE | TRUE | FALSE |
| ZBTB24 | 0.1479712 | 0.84815044 | 1 | 0.147971216 | 0.147971216 | 2.921128275 | 0.18475476 | 0.855788594 | 1 | 0.184754765 | 0.184754765 | 3.652958701 | 0.80090609 | -0.096418404 | FALSE | TRUE | FALSE |
| PAIP2B | 0.1479607 | 0.84816402 | 0.00335 | 0.147960735 | 0.147960735 | 0.147960735 | 0.15229392 | 0.863989546 | 1 | 0.152293917 | 0.152293917 | 0.152293917 | 0.971547239 | -0.012536078 | FALSE | TRUE | FALSE |
| OR10S1 | 0.1478966 | 0.8481776 | 0.75545 | 1.340322659 | 12.46698644 | 12.46698644 | 0.1844058 | 0.855878908 | 1 | 1.680144636 | 15.71142201 | 15.71142201 | 0.802016975 | -0.09581644 | FALSE | TRUE | FALSE |
| RPP38 | 0.1478721 | 0.84819118 | 1 | 0.147872134 | 0.147872134 | 0.147872134 | 14.5688551 | 0.15986537 | 0.2562 | 6.242419206 | 58.2732175 | 58.2732175 | 0.01014988 | -1.993539085 | FALSE | FALSE | FALSE |
| SOCS5 | 0.1477687 | 0.84820476 | 1 | 0.147768691 | 0.147768691 | 2.923884454 | 0.17917795 | 0.857245913 | 0.5064 | 1.650390743 | 1.650390743 | 3.549920691 | 0.824703556 | -0.083702133 | FALSE | TRUE | FALSE |
| SWSAP1 | 0.1476685 | 0.84821834 | 1 | 1.36083484 | 1.36083484 | 1.36083484 | 0.19053428 | 0.854170469 | 1 | 0.190534282 | 0.190534282 | 0.190534282 | 0.775023166 | -0.110685316 | FALSE | TRUE | FALSE |
| DCAF17 | 0.1474698 | 0.84823191 | 1 | 0.147469838 | 0.147469838 | 2.925930749 | 0.18403713 | 0.855943354 | 0.0098 | 0.184037125 | 0.184037125 | 0.184037125 | 0.801304834 | -0.096202237 | FALSE | TRUE | FALSE |
| TNFRSF25 | 0.1474589 | 0.84824548 | 1 | 0.147458858 | 0.147458858 | 2.916895412 | 0.18491587 | 0.855775683 | 1 | 0.184915872 | 0.184915872 | 3.662652352 | 0.797437541 | -0.098303323 | FALSE | TRUE | FALSE |
| GFER | 0.1474459 | 0.84825905 | 1 | 0.147445911 | 0.589653338 | 0.589653338 | 0.1808161 | 0.85690334 | 1 | 0.180816097 | 1.666519274 | 1.666519274 | 0.815446817 | -0.088604358 | FALSE | TRUE | FALSE |
| RASGRP4 | 0.1473873 | 0.84827262 | 1 | 0.147387324 | 0.147387324 | 2.902564968 | 0.17890529 | 0.857271228 | 1 | 1.655505259 | 1.655505259 | 6.055419234 | 0.823828786 | -0.084163037 | FALSE | TRUE | FALSE |
| GSTM3 | 0.1473785 | 0.84828619 | 0.005 | 0.147378453 | 0.147378453 | 0.147378453 | 0.17029371 | 0.859575962 | 1 | 1.560580995 | 1.560580995 | 1.560580995 | 0.865436863 | -0.06276461 | FALSE | TRUE | FALSE |
| MRM1 | 0.1473545 | 0.84829975 | 1 | 0.147354484 | 0.147354484 | 2.916812689 | 0.17328845 | 0.858774397 | 0.7503 | 1.591366095 | 14.99907362 | 14.99907362 | 0.850342187 | -0.070406274 | FALSE | TRUE | FALSE |
| RRP1B | 0.1473325 | 0.84831331 | 1 | 0.147332525 | 0.147332525 | 2.905131627 | 0.18417671 | 0.855904693 | 0.0182 | 0.184176714 | 0.184176714 | 3.637790841 | 0.799951969 | -0.096936088 | FALSE | TRUE | FALSE |
| SLC13A3 | 0.1472559 | 0.84832687 | 0.01525 | 0.147255906 | 0.147255906 | 2.90876677 | 0.1838444 | 0.856007743 | 1 | 0.183844403 | 0.183844403 | 3.636897814 | 0.800981175 | -0.096377691 | FALSE | TRUE | FALSE |
| TMEM71 | 0.1472232 | 0.84834042 | 1 | 1.330901087 | 1.330901087 | 1.330901087 | 0.18920679 | 0.854552087 | 0.00605 | 0.189206795 | 0.189206795 | 0.189206795 | 0.778107496 | -0.108960401 | FALSE | TRUE | FALSE |
| FER | 0.1472079 | 0.84835397 | 0.50895 | 0.147207927 | 0.147207927 | 1.243491643 | 0.05323901 | 0.886021071 | 1 | 0.490434476 | 0.490434476 | 1.799351843 | 2.765039007 | 0.441701262 | FALSE | TRUE | FALSE |
| PGLYRP4 | 0.1471722 | 0.84836752 | 0.00985 | 0.147172154 | 0.147172154 | 0.147172154 | 0.178057 | 0.857460817 | 1 | 1.618829871 | 1.618829871 | 1.618829871 | 0.826545183 | -0.082733401 | FALSE | TRUE | FALSE |
| PLEKHA5 | 0.147136 | 0.84838107 | 1 | 0.147136014 | 0.147136014 | 4.949406746 | 0.18459996 | 0.85581441 | 1 | 0.184599959 | 0.184599959 | 3.625549873 | 0.797053341 | -0.098512613 | FALSE | TRUE | FALSE |
| SEC14L5 | 0.147019 | 0.84839462 | 0.50865 | 0.147018976 | 0.147018976 | 2.894858988 | 0.1066781 | 0.874153393 | 0.2673 | 0.1066781 | 0.426612551 | 2.104446182 | 1.378155175 | 0.13929812 | FALSE | TRUE | FALSE |
| CETP | 0.1470041 | 0.84840816 | 0.51545 | 0.147004093 | 0.147004093 | 0.584530382 | 0.22771634 | 0.845438815 | 0.7533 | 0.227716337 | 2.013476247 | 2.013476247 | 0.645557959 | -0.19006476 | FALSE | TRUE | FALSE |
| SF3B1 | 0.1469688 | 0.8484217 | 1 | 0.146968826 | 0.146968826 | 4.940677776 | 0.72899516 | 0.652171031 | 0.51455 | 0.728995163 | 0.728995163 | 14.26587271 | 0.201604665 | -0.695499422 | FALSE | TRUE | FALSE |
| HSF2BP | 0.1469523 | 0.84843524 | 1 | 1.336606936 | 1.336606936 | 2.9190188 | 0.18919462 | 0.854565212 | 0.00785 | 0.189194624 | 0.189194624 | 0.189194624 | 0.776725509 | -0.109732432 | FALSE | TRUE | FALSE |
| ALDH2 | 0.1469106 | 0.84844878 | 0.81685 | 0.587414328 | 2.349332044 | 9.398374562 | 9.07917119 | 0.210112648 | 0.0381 | 0.99879761 | 9.079171191 | 36.31559345 | 0.01618106 | -1.790993028 | FALSE | FALSE | FALSE |
| RHOJ | 0.1468844 | 0.84846231 | 1 | 0.146884426 | 0.146884426 | 0.146884426 | 0.1840543 | 0.855917582 | 1 | 0.184054299 | 0.184054299 | 0.184054299 | 0.798049415 | -0.097970216 | FALSE | TRUE | FALSE |
| CCSER2 | 0.1468831 | 0.84847584 | 1 | 1.341463974 | 12.57431057 | 26.4573081 | 1.73855968 | 0.51375758 | 0.5045 | 1.738559676 | 1.738559676 | 6.401272859 | 0.084485484 | -1.073217903 | FALSE | FALSE | FALSE |
| CHI3L2 | 0.1467929 | 0.84848937 | 1 | 0.146792925 | 0.146792925 | 0.146792925 | 0.18320708 | 0.856200573 | 1 | 0.183207084 | 0.183207084 | 0.183207084 | 0.801240439 | -0.09623714 | FALSE | TRUE | FALSE |
| POU6F1 | 0.1467858 | 0.8485029 | 1 | 0.146785839 | 0.146785839 | 0.146785839 | 0.72047794 | 0.655768907 | 0.75425 | 0.720477943 | 2.881270231 | 2.881270231 | 0.20373398 | -0.690936531 | FALSE | TRUE | FALSE |
| HOXB6 | 0.1467664 | 0.84851642 | 1 | 0.146766401 | 0.146766401 | 0.146766401 | 0.72826872 | 0.652487065 | 0.5035 | 0.728268722 | 0.728268722 | 0.728268722 | 0.201527811 | -0.695665012 | FALSE | TRUE | FALSE |
| NEUROG1 | 0.1467179 | 0.84852995 | 1 | 0.146717866 | 0.146717866 | 2.906647157 | 0.18417963 | 0.855891802 | 0.00935 | 0.184179634 | 0.184179634 | 0.184179634 | 0.796602006 | -0.098758604 | FALSE | TRUE | FALSE |
| TMIE | 0.1466907 | 0.84854347 | 1 | 0.146690721 | 0.146690721 | 0.146690721 | 0.17818711 | 0.857422935 | 1 | 1.655644654 | 1.655644654 | 1.655644654 | 0.823239818 | -0.084473632 | FALSE | TRUE | FALSE |
| DHRS2 | 0.1466685 | 0.84855698 | 0.01 | 0.146668521 | 0.146668521 | 0.174314591 | 1.57475519 | 0.528949096 | 0.75175 | 1.574755195 | 14.76126752 | 14.76126752 | 0.093137347 | -1.030876137 | FALSE | FALSE | FALSE |
| KCNJ3 | 0.1466232 | 0.8485705 | 0.75025 | 0.586323326 | 2.345201455 | 11.5864116 | 0.18596465 | 0.855491101 | 1 | 0.743671681 | 0.743671681 | 3.680152482 | 0.788446724 | -0.103227647 | FALSE | TRUE | FALSE |
| ZHX1 | 0.1465988 | 0.84858401 | 0.01985 | 0.14659884 | 0.14659884 | 2.887345785 | 0.18405295 | 0.855930469 | 1 | 0.184052954 | 0.184052954 | 3.631667214 | 0.7965036 | -0.098812257 | FALSE | TRUE | FALSE |
| CD151 | 0.1465834 | 0.84859752 | 1 | 0.146583356 | 0.146583356 | 0.146583356 | 0.7265136 | 0.653666702 | 0.5081 | 0.726513602 | 0.726513602 | 0.726513602 | 0.201762714 | -0.695159088 | FALSE | TRUE | FALSE |
| NEU2 | 0.1465251 | 0.84861103 | 0.51345 | 0.146525116 | 0.585695128 | 0.585695128 | 0.10584397 | 0.874412497 | 0.5081 | 0.42322014 | 1.692680865 | 2.099243182 | 1.384350496 | 0.141246061 | FALSE | TRUE | FALSE |
| LRIT3 | 0.1464637 | 0.84862453 | 1 | 0.146463667 | 0.146463667 | 2.905290949 | 0.18336153 | 0.856162048 | 0.01105 | 0.183361529 | 0.183361529 | 3.640787648 | 0.798769884 | -0.097578318 | FALSE | TRUE | FALSE |
| OR52B4 | 0.146459 | 0.84863804 | 0.50265 | 1.32725567 | 1.32725567 | 1.32725567 | 0.18845902 | 0.854801051 | 1 | 0.188459018 | 0.188459018 | 0.188459018 | 0.77713965 | -0.109500933 | FALSE | TRUE | FALSE |
| DAND5 | 0.1464153 | 0.84865154 | 1 | 0.146415264 | 0.146415264 | 0.146415264 | 0.18267837 | 0.856316032 | 1 | 0.18267837 | 0.18267837 | 0.18267837 | 0.801492064 | -0.096100774 | FALSE | TRUE | FALSE |
| CHRNB2 | 0.14641 | 0.84866504 | 0.9854 | 2.341274774 | 37.47727917 | 149.9990748 | 3.41897359 | 0.388510324 | 0.77765 | 13.67826373 | 219.0921609 | 877.1773792 | 0.042822794 | -1.368325 | FALSE | FALSE | FALSE |
| CTHRC1 | 0.1463805 | 0.84867853 | 0.5002 | 0.1463805 | 0.1463805 | 0.1463805 | 0.20781946 | 0.850246726 | 0.75485 | 0.4635318 | 1.853710933 | 1.853710933 | 0.704363781 | -0.152202984 | FALSE | TRUE | FALSE |
| RSPO2 | 0.146306 | 0.84869203 | 1 | 0.146306015 | 0.146306015 | 0.146306015 | 0.70326883 | 0.665085704 | 0.74695 | 0.70326883 | 6.555011306 | 6.555011306 | 0.20803711 | -0.681859187 | FALSE | TRUE | FALSE |
| ELP6 | 0.1460274 | 0.84870552 | 1 | 1.321884825 | 1.321884825 | 1.321884825 | 0.18790653 | 0.855062194 | 1 | 0.187906529 | 0.187906529 | 0.187906529 | 0.777127721 | -0.109507599 | FALSE | TRUE | FALSE |
| VMO1 | 0.1459869 | 0.84871901 | 1 | 0.58389433 | 0.58389433 | 0.58389433 | 0.17889482 | 0.857283882 | 1 | 1.631317764 | 1.631317764 | 1.631317764 | 0.816049052 | -0.088283735 | FALSE | TRUE | FALSE |
| CALML6 | 0.1459492 | 0.8487325 | 1 | 0.145949171 | 0.145949171 | 0.145949171 | 0.18239655 | 0.85635448 | 0.00515 | 0.182396546 | 0.182396546 | 0.182396546 | 0.800175082 | -0.096814977 | FALSE | TRUE | FALSE |

| | | | | | | | | | | | | | | | | | |
|---|---|---|---|---|---|---|---|---|---|---|---|---|---|---|---|---|---|
| NUDT16 | 0.1459072 | 0.84874598 | 1 | 0.145907218 | 0.145907218 | 0.145907218 | 0.16986598 | 0.859722973 | 1 | 1.554523438 | 1.554523438 | 1.554523438 | 0.858954935 | -0.066029621 | FALSE | TRUE | FALSE |
| SLC7A13 | 0.1458983 | 0.84875946 | 0.0127 | 0.145898329 | 0.145898329 | 0.321785145 | 0.23022002 | 0.844831728 | 0.25745 | 0.230220022 | 2.136834553 | 2.136834553 | 0.633734319 | -0.198092774 | FALSE | TRUE | FALSE |
| BPIFB6 | 0.1457971 | 0.84877294 | 1 | 0.145797065 | 0.145797065 | 2.889156216 | 0.18207789 | 0.856495285 | 1 | 0.182077891 | 0.182077891 | 3.612093782 | 0.800740079 | -0.096508434 | FALSE | TRUE | FALSE |
| FXN | 0.1457949 | 0.84878642 | 1 | 0.145794919 | 0.145794919 | 0.145794919 | 0.18307688 | 0.856226245 | 1 | 0.183076882 | 0.183076882 | 0.183076882 | 0.796358982 | -0.098891117 | FALSE | TRUE | FALSE |
| WDR4 | 0.1457857 | 0.8487999 | 1 | 0.145785686 | 0.145785686 | 2.887959247 | 0.18222621 | 0.856392909 | 1 | 0.182226209 | 0.182226209 | 3.613965329 | 0.800025892 | -0.096895957 | FALSE | TRUE | FALSE |
| TAS2R41 | 0.1456782 | 0.84881337 | 0.495 | 0.145678233 | 1.318543078 | 1.318543078 | 0.18751433 | 0.855114313 | 1 | 0.187514333 | 0.187514333 | 0.187514333 | 0.776891192 | -0.109639802 | FALSE | TRUE | FALSE |
| RCHY1 | 0.14562 | 0.84882684 | 1 | 0.145619974 | 0.145619974 | 0.145619974 | 0.17643965 | 0.857989075 | 0.5076 | 1.605370378 | 1.605370378 | 1.605370378 | 0.825324567 | -0.083375227 | FALSE | TRUE | FALSE |
| EXOC6 | 0.1455993 | 0.84884031 | 0.02005 | 0.145599287 | 0.145599287 | 2.872429425 | 0.18214995 | 0.856418517 | 1 | 0.182149946 | 0.182149946 | 3.599386881 | 0.799337526 | -0.097269798 | FALSE | TRUE | FALSE |
| PBLD | 0.1455847 | 0.84885378 | 1 | 0.145584688 | 0.145584688 | 0.145584688 | 0.18221153 | 0.856405714 | 1 | 0.182211529 | 0.182211529 | 0.182211529 | 0.798987248 | -0.097460152 | FALSE | TRUE | FALSE |
| GPR18 | 0.145565 | 0.84886724 | 1 | 0.145564982 | 0.145564982 | 0.145564982 | 0.1821016 | 0.856456912 | 1 | 0.182101598 | 0.182101598 | 0.182101598 | 0.799361362 | -0.097256848 | FALSE | TRUE | FALSE |
| MSX2 | 0.1455158 | 0.84888071 | 1 | 0.581735159 | 2.326210924 | 2.326210924 | 0.73944267 | 0.646621195 | 0.49925 | 0.184951913 | 0.739442672 | 0.739442672 | 0.19679116 | -0.705994414 | FALSE | TRUE | FALSE |
| SV2B | 0.1455049 | 0.84889417 | 1 | 0.145504878 | 0.145504878 | 2.870819326 | 2.70716348 | 0.434673295 | 0.8153 | 10.83172264 | 100.7188462 | 403.090252 | 0.053748094 | -1.26963693 | FALSE | FALSE | FALSE |
| OR2M4 | 0.1454816 | 0.84890762 | 1 | 1.346561325 | 1.346561325 | 1.346561325 | 0.17016286 | 0.859637253 | 1 | 0.170162861 | 0.170162861 | 0.170162861 | 0.854955313 | -0.068056584 | FALSE | TRUE | FALSE |
| NR1I3 | 0.1454662 | 0.84892108 | 0.69065 | 0.327208222 | 12.07232509 | 12.07232509 | 0.06569171 | 0.883437195 | 0.7577 | 0.148838504 | 0.594911061 | 0.594911061 | 2.214376259 | 0.345251417 | FALSE | TRUE | FALSE |
| DPP6 | 0.14545 | 0.84893453 | 0.7528 | 1.337695532 | 12.62687204 | 26.28453011 | 0.76670733 | 0.633350374 | 0.507 | 0.766707332 | 0.766707332 | 15.09694093 | 0.189707334 | -0.721915879 | FALSE | TRUE | FALSE |
| RPP21 | 0.1454338 | 0.84894798 | 1 | 0.145433794 | 0.145433794 | 0.145433794 | 1.38978074 | 0.545969436 | 0.49895 | 0.147407631 | 1.389780739 | 1.389780739 | 0.104645136 | -0.980280955 | FALSE | FALSE | FALSE |
| KLHL28 | 0.145412 | 0.84896143 | 1 | 0.145411983 | 0.145411983 | 2.875740353 | 0.17701938 | 0.857825982 | 0.5111 | 1.660324969 | 1.660324969 | 3.505530544 | 0.821446708 | -0.085420606 | FALSE | TRUE | FALSE |
| ZNF205 | 0.1454059 | 0.84897487 | 1 | 0.14540593 | 0.14540593 | 2.857394395 | 0.1821008 | 0.856469706 | 1 | 0.182100796 | 0.182100796 | 3.586005676 | 0.79849146 | -0.097729724 | FALSE | TRUE | FALSE |
| TIMM23 | 0.1453496 | 0.84898832 | 1 | 0.145349587 | 0.145349587 | 0.145349587 | 0.14994261 | 0.864598521 | 0.0053 | 0.149942606 | 0.149942606 | 0.149942606 | 0.969368157 | -0.013511251 | FALSE | TRUE | FALSE |
| CEACAM6 | 0.1453052 | 0.84900176 | 1 | 0.145305225 | 0.145305225 | 0.172704626 | 0.18150363 | 0.856712347 | 0.0095 | 0.181503632 | 0.181503632 | 0.181503632 | 0.800563729 | -0.09660409 | FALSE | TRUE | FALSE |
| ZNF365 | 0.1453027 | 0.84901519 | 1 | 0.145302671 | 0.145302671 | 2.881500692 | 0.18160142 | 0.856648574 | 0.0095 | 0.181601418 | 0.181601418 | 0.181601418 | 0.800118594 | -0.096845637 | FALSE | TRUE | FALSE |
| LRRC18 | 0.1452422 | 0.84902863 | 1 | 17848157869 | 3.03517E+13 | 4.76962E+15 | 0.34631389 | 0.810135974 | 1 | 0.346313885 | 0.346313885 | 0.346313885 | 0.41939461 | -0.377377156 | FALSE | TRUE | FALSE |
| RAB34 | 0.1452413 | 0.84904206 | 0.5039 | 0.145241341 | 0.145241341 | 0.145241341 | 0.20031971 | 0.852043902 | 0.87465 | 0.446208612 | 16.31431627 | 16.31431627 | 0.725047675 | -0.139633436 | FALSE | TRUE | FALSE |
| UGT2B11 | 0.1452109 | 0.84905549 | 0.51005 | 0.145210912 | 0.145210912 | 0.333095233 | 0.02751033 | 0.891838408 | 1 | 0.027510327 | 0.027510327 | 0.545961746 | 5.278414563 | 0.722503496 | FALSE | TRUE | FALSE |
| TUBA1C | 0.1449433 | 0.84906892 | 0.01345 | 0.144943338 | 0.144943338 | 2.869285512 | 3.61068225 | 0.378037627 | 0.0123 | 0.182171733 | 0.182171733 | 3.610682255 | 0.040142923 | -1.396391013 | FALSE | FALSE | FALSE |
| ZNF253 | 0.1449423 | 0.84908235 | 0.50965 | 1.312986689 | 1.312986689 | 2.87567628 | 1.6494653 | 0.521752214 | 0.50655 | 1.649465305 | 1.649465305 | 3.596653937 | 0.087872296 | -1.056148025 | FALSE | FALSE | FALSE |
| NAPEPLD | 0.1449368 | 0.84909577 | 0.5008 | 0.144936774 | 0.144936774 | 0.719630162 | 1.82656265 | 0.505045989 | 0.50035 | 1.826562651 | 17.04280031 | 17.04280031 | 0.079349467 | -1.100455984 | FALSE | FALSE | FALSE |
| COG3 | 0.1448955 | 0.8491092 | 0.0202 | 0.144895517 | 0.144895517 | 2.855358457 | 0.1819152 | 0.856546417 | 1 | 0.181915196 | 0.181915196 | 3.591215847 | 0.796500346 | -0.098814031 | FALSE | TRUE | FALSE |
| DTYMK | 0.144895 | 0.84912262 | 1 | 0.144894976 | 0.144894976 | 0.144894976 | 0.18165012 | 0.856635812 | 1 | 0.181650116 | 0.181650116 | 0.181650116 | 0.797659695 | -0.098182352 | FALSE | TRUE | FALSE |
| MCM5 | 0.1448917 | 0.84913603 | 0.02425 | 0.144891652 | 0.144891652 | 2.841891868 | 0.18158008 | 0.85667409 | 0.0216 | 0.181580083 | 0.181580083 | 3.569774935 | 0.797949037 | -0.098024845 | FALSE | TRUE | FALSE |
| ZFAND1 | 0.1448499 | 0.84914945 | 1 | 0.144849905 | 1.330414656 | 1.330414656 | 0.67504539 | 0.677496057 | 0.8742 | 14.67476088 | 568.6104254 | 568.6104254 | 0.214578023 | -0.66841476 | FALSE | TRUE | FALSE |
| ABHD1 | 0.1448382 | 0.84916286 | 0.25975 | 0.036208212 | 0.144838205 | 0.179518944 | 0.12412141 | 0.870324208 | 0.87655 | 1.09416334 | 9.893451128 | 9.893451128 | 1.166907549 | 0.067036449 | FALSE | TRUE | FALSE |
| SMARCAD1 | 0.1447883 | 0.84917627 | 1 | 0.144788267 | 0.144788267 | 2.845498581 | 0.18208878 | 0.856482497 | 0.024 | 0.182088776 | 0.182088776 | 3.586037844 | 0.795152073 | -0.099549804 | FALSE | TRUE | FALSE |
| SEC24B | 0.1446403 | 0.84918968 | 0.51335 | 0.57836236 | 2.313232822 | 11.32288117 | 28.8594559 | 0.094879613 | 0.00425 | 0.856007534 | 3.423949084 | 16.80111929 | 0.005011885 | -2.299998908 | FALSE | FALSE | FALSE |
| KLHL24 | 0.1446002 | 0.84920309 | 1 | 0.57822746 | 0.57822746 | 4.893722097 | 0.17806442 | 0.857448192 | 1 | 1.662427773 | 1.662427773 | 3.52300332 | 0.812066523 | -0.090408392 | FALSE | TRUE | FALSE |
| PTGES2 | 0.1445981 | 0.84921649 | 1 | 0.144598116 | 0.144598116 | 2.867002254 | 0.17359122 | 0.858699964 | 1 | 1.587880905 | 6.351061834 | 6.351061834 | 0.832980596 | -0.079365115 | FALSE | TRUE | FALSE |
| CNST | 0.144598 | 0.84922989 | 0.50905 | 0.144597965 | 0.144597965 | 2.85350526 | 0.17388002 | 0.858650301 | 0.50705 | 0.173880015 | 0.173880015 | 0.637197609 | 0.831596228 | -0.080087489 | FALSE | TRUE | FALSE |
| DHRS12 | 0.1445853 | 0.84924329 | 0.49925 | 0.016931698 | 0.144585311 | 0.144585311 | 0.02676905 | 0.891989828 | 0.5073 | 0.231306714 | 0.231306714 | 0.231306714 | 5.401211154 | 0.732491156 | FALSE | TRUE | FALSE |
| GPX6 | 0.1445371 | 0.84925668 | 1 | 0.144537059 | 0.144537059 | 0.144537059 | 0.14885911 | 0.864883989 | 0.0035 | 0.148859106 | 0.148859106 | 0.148859106 | 0.97096552 | -0.012796192 | FALSE | TRUE | FALSE |
| NOBOX | 0.1445175 | 0.84927008 | 0.9385 | 1.313525196 | 21.01198083 | 21.01198083 | 1.94679606 | 0.494106905 | 0.5129 | 1.946796062 | 31.14570796 | 31.14570796 | 0.074233508 | -1.129400014 | FALSE | FALSE | FALSE |
| MTA3 | 0.1444308 | 0.84928347 | 1 | 15420248.26 | 23100922110 | 2.75633E+11 | 44.7424733 | 0.061235173 | 1 | 1.00122E+13 | 2.63304E+17 | 1.40557E+19 | 0.003228048 | -2.491060082 | FALSE | FALSE | FALSE |
| LMX1A | 0.14443 | 0.84929686 | 1 | 0.14442997 | 0.14442997 | 2.864660034 | 0.17599931 | 0.858076747 | 1 | 1.656608227 | 1.656608227 | 3.494293415 | 0.820628059 | -0.085853637 | FALSE | TRUE | FALSE |
| THG1L | 0.1444063 | 0.84931025 | 0.00885 | 0.144406262 | 0.144406262 | 0.144406262 | 0.17908869 | 0.857258571 | 1 | 0.716216256 | 0.716216256 | 0.716216256 | 0.806339379 | -0.09348213 | FALSE | TRUE | FALSE |
| KAT2B | 0.1443162 | 0.84932363 | 1 | 0.577255999 | 0.577255999 | 11.37154803 | 0.16683987 | 0.860465781 | 1 | 14.33728244 | 138.1043773 | 282.9457289 | 0.864998533 | -0.062984629 | FALSE | TRUE | FALSE |
| CIC | 0.1442901 | 0.84933701 | 1 | 1.63352E+39 | 9.81819E+47 | 6.85072E+51 | 0.05061382 | 0.886502508 | 1 | 1.35034E+26 | 1.96044E+33 | 5.38695E+35 | 2.850804236 | 0.454967395 | FALSE | TRUE | FALSE |
| CD164 | 0.1442086 | 0.84935039 | 0.00775 | 0.144208608 | 0.144208608 | 0.144208608 | 0.15106722 | 0.864380805 | 1 | 0.151067219 | 0.151067219 | 0.151067219 | 0.954598946 | -0.020179049 | FALSE | TRUE | FALSE |
| ST6GAL2 | 0.1441826 | 0.84936377 | 1 | 0.144182596 | 0.144182596 | 2.841442 | 0.18084623 | 0.856890622 | 1 | 0.180846234 | 0.180846234 | 3.57026013 | 0.797266238 | -0.098396626 | FALSE | TRUE | FALSE |
| MTIF2 | 0.1441081 | 0.84937715 | 0.26035 | 0.036048125 | 0.144108054 | 0.329866506 | 0.06831879 | 0.882809612 | 0.50395 | 0.068318794 | 0.273133995 | 0.619476243 | 2.109347147 | 0.32414806 | FALSE | TRUE | FALSE |
| GMCL1 | 0.1440733 | 0.84939052 | 1 | 0.144073259 | 0.144073259 | 2.856757282 | 1.60492554 | 0.525232859 | 0.50355 | 1.604925536 | 1.604925536 | 3.474938094 | 0.089769435 | -1.046871508 | FALSE | FALSE | FALSE |
| SLITRK3 | 0.1440581 | 0.84940389 | 1 | 0.144058132 | 0.144058132 | 2.828597007 | 0.17550947 | 0.858226799 | 1 | 1.636338016 | 1.636338016 | 32.20025664 | 0.820799775 | -0.085762771 | FALSE | TRUE | FALSE |
| CSN2 | 0.144053 | 0.84941726 | 1 | 0.144053031 | 0.144053031 | 0.144053031 | 0.1745329 | 0.858463767 | 1 | 1.581465071 | 1.581465071 | 1.581465071 | 0.825363171 | -0.083354914 | FALSE | TRUE | FALSE |
| ARFIP1 | 0.1440139 | 0.84943063 | 0.0093 | 0.144013868 | 0.144013868 | 0.144013868 | 0.18095664 | 0.856865181 | 0.00745 | 0.180956639 | 0.180956639 | 0.180956639 | 0.795847386 | -0.099170206 | FALSE | TRUE | FALSE |
| PRKCH | 0.1439905 | 0.84944399 | 1 | 0.14399053 | 0.14399053 | 2.833508334 | 0.18114735 | 0.85680154 | 0.0197 | 0.181147345 | 0.181147345 | 3.571595609 | 0.794880709 | -0.099698043 | FALSE | TRUE | FALSE |
| SGPL1 | 0.1439059 | 0.84945735 | 1 | 0.143905931 | 0.143905931 | 2.846076592 | 6.38123157 | 0.258679359 | 0.25175 | 1.595520286 | 6.381231569 | 13.70988227 | 0.022551435 | -1.646825811 | FALSE | FALSE | FALSE |
| SLC17A8 | 0.1438634 | 0.84947071 | 0.7555 | 1.296283339 | 11.98816455 | 11.98816455 | 0.50103057 | 0.754738916 | 0.50525 | 0.501030568 | 0.501030568 | 1.093878666 | 0.287134969 | -0.541913913 | FALSE | TRUE | FALSE |
| GSTM1 | 0.1437824 | 0.84948407 | 0.00555 | 0.14378244 | 0.14378244 | 0.14378244 | 0.15434159 | 0.863468354 | 1 | 0.154341586 | 0.154341586 | 0.154341586 | 0.931585866 | -0.030777109 | FALSE | TRUE | FALSE |
| PRRX1 | 0.1437449 | 0.84949743 | 0.0075 | 0.143744908 | 0.143744908 | 0.143744908 | 0.18087989 | 0.856877903 | 1 | 0.180879893 | 0.180879893 | 0.180879893 | 0.794698102 | -0.099797824 | FALSE | TRUE | FALSE |
| TBX15 | 0.1436811 | 0.84951078 | 0.51325 | 0.143681077 | 0.143681077 | 0.7125376 | 0.20007057 | 0.852084571 | 0.87875 | 1.874773235 | 18.03020443 | 18.03020443 | 0.718151985 | -0.143783635 | FALSE | TRUE | FALSE |
| POU5F2 | 0.1436392 | 0.84952413 | 1 | 0.143639168 | 0.143639168 | 2.849628275 | 1.58462443 | 0.528096558 | 0.504 | 1.584624432 | 1.584624432 | 1.584624432 | 0.090645559 | -1.042653467 | FALSE | FALSE | FALSE |
| PTP4A3 | 0.143623 | 0.84953748 | 1 | 0.14362299 | 0.14362299 | 0.14362299 | 0.18073614 | 0.856941477 | 1 | 0.180736142 | 0.180736142 | 0.180736142 | 0.794655615 | -0.099821044 | FALSE | TRUE | FALSE |
| RHPN2 | 0.1435552 | 0.84955082 | 1 | 117.8785284 | 1160.79434 | 11710.03843 | 0.20338117 | 0.851267087 | 1 | 1.871170897 | 1.871170897 | 36.89455067 | 0.705842987 | -0.151291896 | FALSE | TRUE | FALSE |
| AMOTL1 | 0.1435548 | 0.84956417 | 1 | 0.143554792 | 0.143554792 | 2.810261572 | 0.18000593 | 0.857030396 | 1 | 0.180005928 | 0.180005928 | 3.532830858 | 0.797500357 | -0.098269114 | FALSE | TRUE | FALSE |
| OR51F2 | 0.1435452 | 0.84957751 | 0.0088 | 0.143545159 | 0.143545159 | 0.143545159 | 1.59270576 | 0.526955266 | 0.4996 | 0.174369727 | 1.592705761 | 1.592705761 | 0.090126602 | -1.045147001 | FALSE | FALSE | FALSE |
| ZP4 | 0.1435399 | 0.84959085 | 0.5021 | 0.143539912 | 1.259595409 | 1.259595409 | 0.07061505 | 0.882299567 | 0.01285 | 0.070615046 | 0.070615046 | 0.161648204 | 2.032710037 | 0.308075432 | FALSE | TRUE | FALSE |
| TUBD1 | 0.1434862 | 0.84960418 | 0.5098 | 0.143486167 | 0.143486167 | 0.314719228 | 0.02654973 | 0.892027625 | 0.0109 | 0.026549728 | 0.026549728 | 0.527348126 | 5.40443075 | 0.732749956 | FALSE | TRUE | FALSE |
| RNF180 | 0.1434264 | 0.84961752 | 0.0106 | 0.143426351 | 0.143426351 | 2.849784035 | 0.17961634 | 0.857131894 | 1 | 0.179616335 | 0.179616335 | 0.179616335 | 0.798515073 | -0.097716881 | FALSE | TRUE | FALSE |
| ATXN1 | 0.1433084 | 0.84963085 | 0.02705 | 0.143308424 | 0.143308424 | 2.805315895 | 1.52869034 | 0.532892481 | 0.6957 | 14.78224206 | 586.2377002 | 586.2377002 | 0.093745882 | -1.028047801 | FALSE | FALSE | FALSE |
| IRGC | 0.1432966 | 0.84964418 | 0.01665 | 0.143296556 | 0.143296556 | 2.824076462 | 0.15618385 | 0.862943301 | 1 | 0.156183845 | 0.156183845 | 3.083468387 | 0.91748641 | -0.03740036 | FALSE | TRUE | FALSE |
| KTI12 | 0.1432956 | 0.84965751 | 1 | 0.143295575 | 0.143295575 | 2.828990391 | 14.0329562 | 0.163178432 | 0.50685 | 14.03295621 | 134.9355333 | 134.9355333 | 0.010211361 | -1.990916391 | FALSE | FALSE | FALSE |
| UBN2 | 0.1432794 | 0.84967083 | 0.515 | 1.314483664 | 1.314483664 | 25.68631209 | 0.17978627 | 0.857068475 | 1 | 1.655974431 | 1.655974431 | 32.44885348 | 0.796943119 | -0.098572675 | FALSE | TRUE | FALSE |
| RSPH9 | 0.1431733 | 0.84968416 | 1 | 0.143173261 | 0.143173261 | 0.143173261 | 0.17385091 | 0.85866272 | 1 | 1.582276782 | 1.582276782 | 1.582276782 | 0.823540462 | -0.084315058 | FALSE | TRUE | FALSE |
| EXD1 | 0.1430762 | 0.84969748 | 0.50755 | 0.143076182 | 0.143076182 | 0.328887237 | 0.02564982 | 0.892253915 | 0.7474 | 0.223449633 | 1.997958786 | 1.997958786 | 5.578057821 | 0.746483012 | FALSE | TRUE | FALSE |
| OPRK1 | 0.1429701 | 0.8497108 | 1 | 1.321557298 | 1.321557298 | 2.830389953 | 135.259758 | 0.020921759 | 0.5027 | 135.2597584 | 1348.456555 | 13771.50396 | 0.001057004 | -2.975923488 | FALSE | FALSE | FALSE |

| | | | | | | | | | | | | | | | | | |
|---|---|---|---|---|---|---|---|---|---|---|---|---|---|---|---|---|---|
| HAL | 0.1428795 | 0.84972411 | 0.93665 | 147.0631555 | 82242.76428 | 1322870.879 | 53.2878685 | 0.052056533 | 0.77335 | 855.404921 | 886998.6053 | 14288279.29 | 0.002681276 | -2.571658502 | FALSE | FALSE | FALSE |
| BRINP3 | 0.1428583 | 0.84973743 | 1 | 0.142858344 | 0.142858344 | 2.810709399 | 0.17771584 | 0.857549131 | 1 | 0.710707388 | 0.710707388 | 14.01039591 | 0.803858266 | -0.094820518 | FALSE | TRUE | FALSE |
| E2F5 | 0.142855 | 0.84975074 | 0.5056 | 0.571242689 | 0.571242689 | 0.571242689 | 0.18158991 | 0.856661333 | 0.0069 | 0.181589911 | 0.181589911 | 0.181589911 | 0.786689939 | -0.104196404 | FALSE | TRUE | FALSE |
| KLK2 | 0.1427888 | 0.84976405 | 1 | 1.304196051 | 1.304196051 | 1.304196051 | 0.18462213 | 0.855801503 | 1 | 0.18462213 | 0.18462213 | 0.18462213 | 0.773411022 | -0.111589643 | FALSE | TRUE | FALSE |
| HIRA | 0.142722 | 0.84977736 | 0.0313 | 0.142721998 | 0.142721998 | 2.787031077 | 0.17974321 | 0.857093849 | 1 | 0.179743208 | 0.179743208 | 3.519956888 | 0.794032773 | -0.100161572 | FALSE | TRUE | FALSE |
| EIF4E1B | 0.1426428 | 0.84979066 | 0.00755 | 0.142642811 | 0.142642811 | 0.142642811 | 0.17889112 | 0.857296534 | 0.00565 | 0.178891119 | 0.178891119 | 0.178891119 | 0.797372232 | -0.098338893 | FALSE | TRUE | FALSE |
| VPS52 | 0.1424972 | 0.84980397 | 0.5115 | 1.30754611 | 1.30754611 | 25.70410181 | 0.18451221 | 0.855853115 | 1 | 0.184512206 | 0.184512206 | 3.634653067 | 0.772291448 | -0.112218775 | FALSE | TRUE | FALSE |
| OTUD7B | 0.1424485 | 0.84981727 | 0.02435 | 0.142448465 | 0.142448465 | 2.800620465 | 0.16981405 | 0.859747445 | 1 | 0.169814046 | 0.169814046 | 3.345465009 | 0.838849724 | -0.076315834 | FALSE | TRUE | FALSE |
| OLAH | 0.1424462 | 0.84983057 | 1 | 12.26442491 | 118.2509776 | 118.2509776 | 0.19602649 | 0.853040863 | 1 | 0.196026489 | 0.196026489 | 0.196026489 | 0.726667875 | -0.138664039 | FALSE | TRUE | FALSE |
| GALNT13 | 0.1423492 | 0.84984387 | 1 | 0.142349171 | 0.142349171 | 2.818360854 | 0.17924307 | 0.857220591 | 1 | 0.17924307 | 0.17924307 | 3.553112239 | 0.794168338 | -0.100087431 | FALSE | TRUE | FALSE |
| SLC35F5 | 0.1422932 | 0.84985716 | 1 | 0.142293213 | 0.142293213 | 2.819881414 | 0.17832618 | 0.857410304 | 0.0112 | 0.17832618 | 0.17832618 | 3.537841855 | 0.797937873 | -0.098030921 | FALSE | TRUE | FALSE |
| CHFR | 0.1421601 | 0.84987046 | 1 | 0.142160074 | 0.142160074 | 2.805882314 | 1.59897555 | 0.52623909 | 0.5089 | 1.598975553 | 1.598975553 | 3.424468089 | 0.088906971 | -1.051064184 | FALSE | FALSE | FALSE |
| AMPD2 | 0.1421514 | 0.84988375 | 0.8189 | 0.738538477 | 11.80253856 | 47.1997169 | 0.06509617 | 0.883567167 | 0.7613 | 0.147501644 | 0.589635831 | 2.887625424 | 2.183714119 | 0.339195782 | FALSE | TRUE | FALSE |
| KPNA2 | 0.1421459 | 0.84989703 | 0.01355 | 0.142145915 | 0.142145915 | 2.814740342 | 0.17882645 | 0.857347118 | 0.0101 | 0.178826452 | 0.178826452 | 3.545301654 | 0.794881927 | -0.099697377 | FALSE | TRUE | FALSE |
| GPATCH4 | 0.1420845 | 0.84991032 | 1 | 1.289347893 | 1.289347893 | 1.289347893 | 0.17274025 | 0.858910667 | 1 | 1.574310462 | 14.72596687 | 14.72596687 | 0.822532413 | -0.084846979 | FALSE | TRUE | FALSE |
| CREB3 | 0.1420317 | 0.8499236 | 0.00915 | 0.142031728 | 0.142031728 | 0.142031728 | 0.70556067 | 0.663840425 | 0.4992 | 0.176420015 | 0.705560668 | 0.705560668 | 0.201303353 | -0.696148991 | FALSE | TRUE | FALSE |
| TCF21 | 0.1419693 | 0.84993689 | 1 | 2.73089E+11 | 6.71701E+15 | 1.18041E+18 | 0.32348551 | 0.817534117 | 1 | 29.55691683 | 2988.639025 | 31168.08858 | 0.438873845 | -0.3576603 | FALSE | TRUE | FALSE |
| TFAP2B | 0.1419114 | 0.84995017 | 0.5072 | 0.141911362 | 0.141911362 | 1.203172701 | 0.21085084 | 0.849409529 | 0.51075 | 0.210850839 | 0.210850839 | 1.043515547 | 0.673041483 | -0.171958167 | FALSE | TRUE | FALSE |
| BAIAP2L1 | 0.1418934 | 0.84996344 | 1 | 0.141893409 | 0.141893409 | 2.805938852 | 0.17806553 | 0.857435565 | 1 | 0.178065535 | 0.178065535 | 3.526008144 | 0.796860598 | -0.098617647 | FALSE | TRUE | FALSE |
| LAMP3 | 0.1418293 | 0.84997672 | 1 | 0.141829282 | 0.141829282 | 2.814064693 | 0.1776768 | 0.857574344 | 1 | 0.177676805 | 0.177676805 | 0.177676805 | 0.79824309 | -0.097864832 | FALSE | TRUE | FALSE |
| SLIT3 | 0.1418278 | 0.84998999 | 0.6354 | 0.141827803 | 9.066200178 | 36.26853924 | 3.47645626 | 0.385053939 | 0.5096 | 3.476456261 | 222.3753924 | 889.7885829 | 0.04079666 | -1.389375395 | FALSE | FALSE | FALSE |
| HOXB3 | 0.1418262 | 0.85000326 | 0.7547 | 0.566640943 | 5.476284152 | 5.476284152 | 0.23841192 | 0.842462875 | 0.6297 | 0.403574833 | 3.807642135 | 3.807642135 | 0.594878817 | -0.225571495 | FALSE | TRUE | FALSE |
| RIT2 | 0.1417716 | 0.85001653 | 1 | 0.14177161 | 0.14177161 | 0.14177161 | 0.1780274 | 0.857473439 | 1 | 0.178027397 | 0.178027397 | 0.178027397 | 0.796347151 | -0.098897569 | FALSE | TRUE | FALSE |
| TRIM3 | 0.1417531 | 0.85002979 | 1 | 0.141753088 | 0.141753088 | 2.775543836 | 0.1767207 | 0.857901301 | 0.51095 | 0.706645663 | 0.706645663 | 13.87120038 | 0.802130645 | -0.095754891 | FALSE | TRUE | FALSE |
| EVI2A | 0.1417491 | 0.85004306 | 1 | 0.14174909 | 1.278038688 | 1.278038688 | 1.60908233 | 0.52436689 | 0.49815 | 0.17741334 | 1.609082329 | 1.609082329 | 0.088093125 | -1.055057986 | FALSE | FALSE | FALSE |
| FERD3L | 0.1416461 | 0.85005632 | 1 | 1.332763635 | 5.328447136 | 5.328447136 | 0.70571591 | 0.663619632 | 0.87765 | 2.822340596 | 26.58508431 | 26.58508431 | 0.200712625 | -0.69742531 | FALSE | TRUE | FALSE |
| ACY3 | 0.1416139 | 0.85006958 | 0.2615 | 0.035428574 | 0.141613918 | 0.175869693 | 0.06237513 | 0.88420452 | 0.5 | 0.038991024 | 0.062375132 | 0.062375132 | 2.270358597 | 0.356094458 | FALSE | TRUE | FALSE |
| SLC25A27 | 0.1415593 | 0.85008283 | 1 | 0.141559288 | 0.141559288 | 0.141559288 | 0.1725341 | 0.858960157 | 0.50735 | 1.59025372 | 1.59025372 | 1.59025372 | 0.820471347 | -0.085936581 | FALSE | TRUE | FALSE |
| FBXW12 | 0.1414989 | 0.85009609 | 0.5139 | 1.281290912 | 1.281290912 | 2.80616593 | 0.17729928 | 0.857687691 | 0.50405 | 1.614219367 | 1.614219367 | 3.519707771 | 0.798079193 | -0.097954012 | FALSE | TRUE | FALSE |
| ASB10 | 0.141479 | 0.85010934 | 1 | 0.141479015 | 0.141479015 | 2.795062194 | 0.17600381 | 0.858064228 | 1 | 0.703871093 | 0.703871093 | 3.48223843 | 0.803840642 | -0.094830039 | FALSE | TRUE | FALSE |
| B9D2 | 0.1414645 | 0.85012259 | 1 | 0.565727049 | 0.565727049 | 0.565727049 | 1.60868299 | 0.524511463 | 0.5046 | 1.608682986 | 1.608682986 | 1.608682986 | 0.087938058 | -1.055823129 | FALSE | FALSE | FALSE |
| LRCH3 | 0.1414436 | 0.85013584 | 0.6364 | 0.565766423 | 5.063321382 | 11.14559184 | 0.19995761 | 0.852111673 | 0.87755 | 4.158807051 | 66.52603146 | 266.174136 | 0.707368034 | -0.15035457 | FALSE | TRUE | FALSE |
| PTPN12 | 0.14144 | 0.85014908 | 1 | 0.141440031 | 0.141440031 | 2.79196957 | 0.17773016 | 0.857523909 | 0.01815 | 0.177730164 | 0.177730164 | 3.51381889 | 0.795813312 | -0.099188801 | FALSE | TRUE | FALSE |
| PIGT | 0.1412985 | 0.85016233 | 0.5109 | 0.141298462 | 0.141298462 | 1.195392405 | 0.05251877 | 0.886137606 | 1 | 0.052518774 | 0.052518774 | 1.037706667 | 2.690437205 | 0.42982286 | FALSE | TRUE | FALSE |
| STARD10 | 0.1412883 | 0.85017557 | 1 | 0.141288311 | 0.141288311 | 0.141288311 | 0.1723073 | 0.859009613 | 1 | 1.593271861 | 1.593271861 | 1.593271861 | 0.819978689 | -0.086197434 | FALSE | TRUE | FALSE |
| LGALS14 | 0.1412669 | 0.8501888 | 0.0063 | 0.141266944 | 0.141266944 | 0.141266944 | 1.55320666 | 0.530646122 | 0.5008 | 1.553206657 | 1.553206657 | 1.553206657 | 0.090951802 | -1.041188691 | FALSE | FALSE | FALSE |
| GJA4 | 0.1412293 | 0.85020204 | 0.50705 | 0.564578782 | 2.257526255 | 2.799267864 | 3.31620857 | 0.39374204 | 0.07045 | 0.829261906 | 13.26479262 | 13.26479262 | 0.042587583 | -1.370717007 | FALSE | FALSE | FALSE |
| FAM13C | 0.1410925 | 0.85021527 | 1 | 0.141092541 | 0.141092541 | 2.794946568 | 0.17769528 | 0.857561739 | 0.012 | 0.177695276 | 0.177695276 | 3.524058436 | 0.794014024 | -0.100171827 | FALSE | TRUE | FALSE |
| THOC6 | 0.141083 | 0.85022851 | 1 | 0.141082994 | 0.141082994 | 2.798577587 | 0.17720223 | 0.857763156 | 1 | 0.177202233 | 0.177202233 | 0.177202233 | 0.796169393 | -0.098994522 | FALSE | TRUE | FALSE |
| TREML1 | 0.1410824 | 0.85024174 | 1 | 0.141082404 | 0.141082404 | 0.141082404 | 0.17692321 | 0.857863651 | 1 | 0.176923212 | 0.176923212 | 0.176923212 | 0.797421673 | -0.098311965 | FALSE | TRUE | FALSE |
| MCM3AP | 0.1410561 | 0.85025496 | 0.9987 | 9913.535976 | 10566480.67 | 202640418.9 | 0.03557403 | 0.889998995 | 0.99975 | 2384.404261 | 2485439.168 | 26329265.79 | 3.965141756 | 0.598258718 | FALSE | TRUE | FALSE |
| SPDL1 | 0.1410425 | 0.85026819 | 0.5057 | 0.141042511 | 0.141042511 | 0.558336902 | 0.02695614 | 0.891952008 | 1 | 0.026956136 | 0.026956136 | 0.533960636 | 5.232297131 | 0.718692399 | FALSE | TRUE | FALSE |
| SH3BP4 | 0.1410242 | 0.85028141 | 1 | 0.141024235 | 0.141024235 | 2.754235089 | 0.17196886 | 0.859133106 | 1 | 1.585186804 | 1.585186804 | 31.04667063 | 0.820056811 | -0.08615606 | FALSE | TRUE | FALSE |
| DEF8 | 0.1409571 | 0.85029463 | 1 | 0.140957132 | 0.140957132 | 2.78446302 | 0.17721717 | 0.857750584 | 1 | 0.177217169 | 0.177217169 | 3.505924322 | 0.795392076 | -0.09941874 | FALSE | TRUE | FALSE |
| LRCH1 | 0.1409493 | 0.85030785 | 0.0188 | 0.14094926 | 0.14094926 | 2.782474109 | 0.17215514 | 0.859059036 | 1 | 1.606048445 | 1.606048445 | 5.823423388 | 0.818733956 | -0.086857198 | FALSE | TRUE | FALSE |
| WIPI1 | 0.1409085 | 0.85032106 | 1 | 0.140908517 | 0.140908517 | 2.793031285 | 0.17726098 | 0.857725434 | 1 | 0.177260975 | 0.177260975 | 3.517364588 | 0.794921255 | -0.09967589 | FALSE | TRUE | FALSE |
| TSPAN1 | 0.140818 | 0.85033428 | 1 | 1.299161301 | 1.299161301 | 1.299161301 | 0.72265565 | 0.654915662 | 0.00545 | 0.722655649 | 0.722655649 | 0.722655649 | 0.194861785 | -0.710273323 | FALSE | TRUE | FALSE |
| AKAP11 | 0.1407306 | 0.85034749 | 1 | 1.30202309 | 1.30202309 | 25.25984718 | 0.18275755 | 0.856272566 | 1 | 0.182757549 | 0.182757549 | 6.141065518 | 0.770039751 | -0.113486855 | FALSE | TRUE | FALSE |
| BHLHE23 | 0.1407294 | 0.8503607 | 1 | 0.140729389 | 0.140729389 | 0.140729389 | 0.17705727 | 0.857800858 | 1 | 0.177057271 | 0.177057271 | 0.177057271 | 0.794824117 | -0.099728964 | FALSE | TRUE | FALSE |
| ZNF214 | 0.1407096 | 0.8503739 | 0.50605 | 0.140709601 | 0.140709601 | 0.551443272 | 1.91189372 | 0.498122843 | 0.50845 | 1.911893718 | 17.53506701 | 17.53506701 | 0.073596978 | -1.133140016 | FALSE | FALSE | FALSE |
| TMEM45B | 0.1406628 | 0.85038711 | 1 | 1.273273755 | 1.273273755 | 1.273273755 | 0.18180454 | 0.856597514 | 1 | 0.181804537 | 0.181804537 | 0.181804537 | 0.773703588 | -0.111425389 | FALSE | TRUE | FALSE |
| TMEM216 | 0.1406519 | 0.85040031 | 0.5028 | 0.562454962 | 0.562454962 | 0.562454962 | 0.70754408 | 0.662659208 | 0.4975 | 0.176928337 | 0.707544081 | 0.707544081 | 0.198788854 | -0.701607971 | FALSE | TRUE | FALSE |
| AQP11 | 0.1405758 | 0.85041351 | 0.0097 | 0.140575772 | 0.140575772 | 0.140575772 | 0.17671885 | 0.857913846 | 1 | 0.176718846 | 0.176718846 | 0.176718846 | 0.79547697 | -0.099372389 | FALSE | TRUE | FALSE |
| SLU7 | 0.1405713 | 0.85042671 | 1 | 0.14057126 | 0.14057126 | 2.783779576 | 1.56645541 | 0.529515976 | 0.50825 | 1.566455415 | 1.566455415 | 3.395562188 | 0.089738437 | -1.047021499 | FALSE | FALSE | FALSE |
| FANCG | 0.1405413 | 0.8504399 | 0.51225 | 1.259469375 | 11.57711022 | 11.57711022 | 0.02738959 | 0.891868721 | 1 | 0.241822351 | 0.241822351 | 0.927341181 | 5.131196412 | 0.710218639 | FALSE | TRUE | FALSE |
| BMP5 | 0.1405181 | 0.8504531 | 1 | 2.247084617 | 21.05968419 | 21.05968419 | 0.18122818 | 0.856763328 | 0.50255 | 1.701118385 | 1.701118385 | 3.59508035 | 0.775365774 | -0.110493373 | FALSE | TRUE | FALSE |
| ZC3HC1 | 0.1402498 | 0.85046629 | 1 | 0.140249762 | 0.140249762 | 2.775344469 | 0.17624711 | 0.858026661 | 0.01615 | 0.176247106 | 0.176247106 | 3.492116046 | 0.795756394 | -0.099219863 | FALSE | TRUE | FALSE |
| PRR5 | 0.140244 | 0.85047948 | 1 | 0.140243968 | 0.140243968 | 2.773907022 | 0.17113901 | 0.859305642 | 1 | 1.578385218 | 1.578385218 | 3.38948163 | 0.819473973 | -0.086464835 | FALSE | TRUE | FALSE |
| RFX6 | 0.1401966 | 0.85049266 | 1 | 0.140196586 | 0.140196586 | 2.756386188 | 1.5836862 | 0.528238837 | 0.5125 | 1.583686198 | 1.583686198 | 31.20020378 | 0.088525484 | -1.052931692 | FALSE | FALSE | FALSE |
| KCNK18 | 0.1401257 | 0.85050585 | 1 | 1.277285813 | 11.94969543 | 11.94969543 | 0.18152062 | 0.856699597 | 1 | 1.663881542 | 1.663881542 | 3.607348207 | 0.77195495 | -0.112408044 | FALSE | TRUE | FALSE |
| CAMK1 | 0.1401066 | 0.85051903 | 1 | 0.14010658 | 0.14010658 | 2.779484495 | 0.17246474 | 0.858972524 | 1 | 0.689715869 | 2.758980858 | 2.758980858 | 0.812378125 | -0.09024178 | FALSE | TRUE | FALSE |
| BAZ1B | 0.1400394 | 0.85053221 | 1 | 1.298677854 | 12.36073397 | 240.3667491 | 0.18778899 | 0.855075227 | 0.03455 | 0.187788993 | 0.187788993 | 6.315218082 | 0.745727188 | -0.127420023 | FALSE | TRUE | FALSE |
| GRIN3B | 0.1400147 | 0.85054539 | 1 | 1.02888E+19 | 4.20838E+24 | 8.26953E+26 | 3.0928742 | 0.406161417 | 1 | 2.42074E+15 | 4.9291E+20 | 8.79557E+22 | 0.045270079 | -1.344188746 | FALSE | FALSE | FALSE |
| PARP8 | 0.1400113 | 0.85055857 | 0.0236 | 0.140011284 | 0.140011284 | 2.757228082 | 0.16624158 | 0.860683513 | 1 | 0.166241579 | 0.166241579 | 3.27982935 | 0.842215796 | -0.074576618 | FALSE | TRUE | FALSE |
| PIK3R1 | 0.1399683 | 0.85057174 | 0.0211 | 0.139968261 | 0.139968261 | 2.760382738 | 0.69806439 | 0.667115634 | 0.5164 | 0.698064395 | 0.698064395 | 13.78994193 | 0.200509096 | -0.697865921 | FALSE | TRUE | FALSE |
| TLX2 | 0.1399122 | 0.85058491 | 1 | 1.317559424 | 5.267676698 | 5.267676698 | 0.18226378 | 0.856380102 | 0.50095 | 0.728742602 | 0.728742602 | 0.728742602 | 0.767636027 | -0.114844651 | FALSE | TRUE | FALSE |
| MRPL28 | 0.1398373 | 0.85059808 | 1 | 0.139837317 | 0.139837317 | 0.139837317 | 0.17562141 | 0.858214307 | 1 | 0.175621408 | 0.175621408 | 0.175621408 | 0.796243001 | -0.098954372 | FALSE | TRUE | FALSE |
| OR6X1 | 0.1398144 | 0.85061125 | 1 | 0.139814407 | 1.263258033 | 1.263258033 | 13.5022098 | 0.166505389 | 0.31285 | 13.50220975 | 1259.756662 | 1259.756662 | 0.010354928 | -1.984852926 | FALSE | FALSE | FALSE |
| RUFY3 | 0.1398075 | 0.85062441 | 1 | 1.282796624 | 12.08039188 | 25.37323298 | 1.67296566 | 0.520143459 | 0.5109 | 1.672965664 | 1.672965664 | 3.593723487 | 0.083568675 | -1.077956483 | FALSE | FALSE | FALSE |
| GDPD5 | 0.1398058 | 0.85063757 | 1 | 1.278389786 | 1.278389786 | 4.732667727 | 0.18129405 | 0.856725096 | 1 | 0.181294048 | 0.181294048 | 3.576545901 | 0.771154673 | -0.112858506 | FALSE | TRUE | FALSE |
| FCHO1 | 0.1397934 | 0.85065073 | 1 | 1.278624584 | 12.00319045 | 43.21042585 | 0.18694853 | 0.855244456 | 1 | 0.186948534 | 0.186948534 | 3.665142444 | 0.747764004 | -0.126235445 | FALSE | TRUE | FALSE |
| ANKRD29 | 0.1397885 | 0.85066389 | 1 | 0.139788476 | 0.139788476 | 0.139788476 | 0.17576688 | 0.858139307 | 1 | 0.17576688 | 0.17576688 | 0.17576688 | 0.795306126 | -0.099465672 | FALSE | TRUE | FALSE |
| SLC16A14 | 0.1397714 | 0.85067704 | 1 | 0.139771445 | 0.139771445 | 2.76304428 | 0.17612621 | 0.858051708 | 0.01255 | 0.176126207 | 0.176126207 | 3.486569044 | 0.793586866 | -0.100405528 | FALSE | TRUE | FALSE |

| | | | | | | | | | | | | | | | | | |
|---|---|---|---|---|---|---|---|---|---|---|---|---|---|---|---|---|---|
| RPAP3 | 0.1397537 | 0.8506902 | 1 | 0.13975373 | 0.13975373 | 2.764741673 | 0.17592441 | 0.858101777 | 1 | 0.175924414 | 0.175924414 | 3.484848361 | 0.794396449 | -0.099962706 | FALSE | TRUE | FALSE |
| RCL1 | 0.1396853 | 0.85070335 | 1 | 0.139685336 | 0.139685336 | 2.770076249 | 0.69728214 | 0.667403538 | 0.5054 | 0.697282137 | 0.697282137 | 0.724927642 | 0.200328287 | -0.698257723 | FALSE | TRUE | FALSE |
| DUOX1 | 0.1396366 | 0.8507165 | 0.4306 | 0.139636561 | 19.2595996 | 171.3690797 | 0.71848019 | 0.657078861 | 0.384 | 0.179843582 | 56.92513452 | 479.5967794 | 0.19434991 | -0.711415655 | FALSE | TRUE | FALSE |
| PDGFRB | 0.1396179 | 0.85072964 | 0.2797 | 0.03492949 | 0.13961789 | 2.714227619 | 0.01563597 | 0.894960689 | 1 | 0.015635966 | 0.015635966 | 0.304994714 | 8.929278225 | 0.950816355 | FALSE | TRUE | FALSE |
| MBD5 | 0.1395335 | 0.85074279 | 1 | 22673066560 | 4.06919E+13 | 6.6275E+15 | 0.24742175 | 0.840068917 | 1 | 22928.40289 | 29260625.32 | 331637358.7 | 0.563949912 | -0.248759467 | FALSE | TRUE | FALSE |
| ACSL1 | 0.1394952 | 0.85075593 | 1 | 0.139495169 | 0.139495169 | 2.748076375 | 0.17571546 | 0.858164315 | 0.01825 | 0.175715465 | 0.175715465 | 3.46786825 | 0.793869621 | -0.100250817 | FALSE | TRUE | FALSE |
| MYCN | 0.1394273 | 0.85076907 | 1 | 15666457.07 | 23201267443 | 3.34459E+12 | 0.17563507 | 0.858201812 | 1 | 20286300.99 | 30359807238 | 4.40716E+12 | 0.793846571 | -0.100263427 | FALSE | TRUE | FALSE |
| PSMD5 | 0.1392248 | 0.8507822 | 1 | 1.262222566 | 1.262222566 | 2.756638925 | 0.17487426 | 0.858339143 | 1 | 1.593850564 | 1.593850564 | 3.466662223 | 0.796142253 | -0.099009327 | FALSE | TRUE | FALSE |
| USP39 | 0.1392204 | 0.85079534 | 0.01935 | 0.139220373 | 0.139220373 | 2.746573301 | 0.17507865 | 0.858289233 | 1 | 0.175078647 | 0.175078647 | 3.459641932 | 0.79518762 | -0.09953039 | FALSE | TRUE | FALSE |
| SCAMP2 | 0.139214 | 0.85080847 | 1 | 0.139213965 | 0.139213965 | 0.139213965 | 0.17473264 | 0.858401482 | 1 | 0.174732639 | 0.174732639 | 0.174732639 | 0.796725588 | -0.098691235 | FALSE | TRUE | FALSE |
| IL18R1 | 0.1392128 | 0.8508216 | 0.0134 | 0.139212797 | 0.139212797 | 2.758307014 | 0.17478035 | 0.858389019 | 1 | 0.174780351 | 0.174780351 | 3.466910416 | 0.796501412 | -0.09881345 | FALSE | TRUE | FALSE |
| CDK5RAP3 | 0.1391606 | 0.85083473 | 0.5116 | 0.306426593 | 1.225539281 | 2.754862688 | 0.20819356 | 0.850135621 | 0.7541 | 0.20819356 | 0.83287917 | 4.126509431 | 0.668419184 | -0.174951094 | FALSE | TRUE | FALSE |
| ZNF416 | 0.1391334 | 0.85084786 | 1 | 0.139133407 | 0.139133407 | 2.755037667 | 1.55234111 | 0.530787075 | 0.5078 | 1.552341108 | 1.552341108 | 3.364359579 | 0.089628115 | -1.047555739 | FALSE | FALSE | FALSE |
| ESPN | 0.1391044 | 0.85086098 | 0.9807 | 1199.582671 | 1370442.489 | 14967651.58 | 0.03054453 | 0.891129274 | 0.9991 | 25359.16014 | 31556620.33 | 354276933.7 | 4.554150594 | 0.658407388 | FALSE | TRUE | FALSE |
| SEC63 | 0.1390612 | 0.8508741 | 1 | 1.271583926 | 1.271583926 | 25.09461469 | 1.55753025 | 0.530505082 | 0.75215 | 1.557530249 | 14.66796093 | 30.78676781 | 0.089283139 | -1.049230551 | FALSE | FALSE | FALSE |
| REC8 | 0.1389631 | 0.85088722 | 0.01795 | 0.138963076 | 0.138963076 | 2.741252703 | 0.17478978 | 0.858376553 | 1 | 0.174789779 | 0.174789779 | 3.453662187 | 0.795029759 | -0.099616615 | FALSE | TRUE | FALSE |
| FAM76A | 0.1389426 | 0.85090034 | 1 | 0.138942571 | 0.138942571 | 0.138942571 | 0.15528933 | 0.863212149 | 1 | 0.155289334 | 0.155289334 | 0.155289334 | 0.894733509 | -0.048306297 | FALSE | TRUE | FALSE |
| RNF133 | 0.1388535 | 0.85091346 | 1 | 0.138853545 | 0.138853545 | 2.758428989 | 121.116045 | 0.023941849 | 0.1903 | 121.116045 | 1186.227893 | 11901.91899 | 0.00114645 | -2.940644708 | FALSE | FALSE | FALSE |
| PLCG1 | 0.1388344 | 0.85092657 | 0.8806 | 1.292219263 | 12.33644887 | 43.43250263 | 0.02146653 | 0.893394592 | 1 | 169.8445975 | 17505.33143 | 184013.1414 | 6.46748299 | 0.810735295 | FALSE | TRUE | FALSE |
| MYBPC3 | 0.138825 | 0.85093968 | 0.86375 | 0.555272922 | 10.76542299 | 43.07036015 | 0.07067434 | 0.882281907 | 0.96515 | 1.130610065 | 18.10497097 | 88.02910615 | 1.96429083 | 0.293205789 | FALSE | TRUE | FALSE |
| AKAP8L | 0.1387797 | 0.85095279 | 1 | 0.138779666 | 0.138779666 | 2.735864495 | 0.15745252 | 0.862685197 | 1 | 0.157452523 | 0.157452523 | 3.109323226 | 0.881406428 | -0.054823786 | FALSE | TRUE | FALSE |
| BPIFB3 | 0.1387689 | 0.8509659 | 1 | 0.138768916 | 0.138768916 | 2.746583994 | 0.17427693 | 0.858526 | 1 | 0.174276934 | 0.174276934 | 3.453681488 | 0.796255203 | -0.098947717 | FALSE | TRUE | FALSE |
| LGMN | 0.1385017 | 0.850979 | 1 | 1.274504603 | 1.274504603 | 2.743016395 | 0.18002376 | 0.857017699 | 1 | 0.180023759 | 0.180023759 | 3.569540304 | 0.769352447 | -0.11387466 | FALSE | TRUE | FALSE |
| ACMSD | 0.1384836 | 0.8509921 | 0.93895 | 1.260572361 | 20.16193987 | 20.16193987 | 0.05475021 | 0.885712086 | 0.0083 | 0.054750212 | 0.054750212 | 0.054750212 | 2.529371106 | 0.403012553 | FALSE | TRUE | FALSE |
| APOM | 0.1384452 | 0.85100521 | 1 | 0.138445205 | 0.138445205 | 0.138445205 | 0.14765161 | 0.865179663 | 0.0043 | 0.147651612 | 0.147651612 | 0.147651612 | 0.937647773 | -0.027960273 | FALSE | TRUE | FALSE |
| ACRBP | 0.1384165 | 0.8510183 | 0.26205 | 0.138416523 | 1.214689795 | 2.736334671 | 138.828687 | 0.020195799 | 0.36785 | 138.8286866 | 12871.15011 | 128330.8799 | 0.000997031 | -3.001291281 | FALSE | FALSE | FALSE |
| ARHGAP10 | 0.1383125 | 0.8510314 | 0.75315 | 0.138312508 | 1.221071436 | 2.723123865 | 0.22071232 | 0.847059598 | 0.7516 | 0.882793916 | 7.880034936 | 17.41323653 | 0.626664204 | -0.202965112 | FALSE | TRUE | FALSE |
| TUBG1 | 0.1383021 | 0.85104449 | 1 | 0.553066011 | 2.212248024 | 10.93787001 | 2.72938239 | 0.433081265 | 0.69255 | 2.729382385 | 43.70423322 | 43.70423322 | 0.050671563 | -1.2952357 | FALSE | FALSE | FALSE |
| SLMAP | 0.1382692 | 0.85105759 | 1 | 0.138269236 | 0.138269236 | 2.723178551 | 0.17419873 | 0.858550878 | 0.01975 | 0.174198731 | 0.174198731 | 3.437103659 | 0.793744223 | -0.100319423 | FALSE | TRUE | FALSE |
| CALCA | 0.1382459 | 0.85107067 | 1 | 0.138245927 | 1.244158867 | 1.244158867 | 0.17876388 | 0.857359759 | 1 | 0.178763877 | 0.178763877 | 0.178763877 | 0.773343749 | -0.111627421 | FALSE | TRUE | FALSE |
| CCNJ | 0.1380928 | 0.85108376 | 1 | 0.138092812 | 0.138092812 | 2.740941057 | 0.14589552 | 0.865474127 | 0.01085 | 0.145895519 | 0.145895519 | 2.898450552 | 0.946518526 | -0.023870881 | FALSE | TRUE | FALSE |
| TSPYL6 | 0.1380009 | 0.85109685 | 0.75595 | 0.138000856 | 1.264573477 | 2.732794369 | 1.69366781 | 0.517785076 | 0.25455 | 0.183540133 | 1.69366781 | 1.69366781 | 0.081480474 | -1.088946454 | FALSE | FALSE | FALSE |
| CTSD | 0.1379554 | 0.85110993 | 1 | 0.137955385 | 0.137955385 | 2.73073681 | 0.17418883 | 0.858563314 | 1 | 0.17418883 | 0.17418883 | 3.452219095 | 0.791987549 | -0.101281646 | FALSE | TRUE | FALSE |
| TOP2A | 0.137926 | 0.85112301 | 1 | 0.137925977 | 0.137925977 | 4.634106111 | 0.17401454 | 0.858625458 | 1 | 0.174014539 | 0.174014539 | 3.404964388 | 0.792611801 | -0.100939466 | FALSE | TRUE | FALSE |
| NIPAL2 | 0.1379015 | 0.85113609 | 0.9704 | 2.205331271 | 8.822447711 | 35.30309898 | 0.20703301 | 0.850399242 | 0.93645 | 0.827887455 | 13.24830356 | 13.24830356 | 0.666084403 | -0.176470736 | FALSE | TRUE | FALSE |
| GPANK1 | 0.1378293 | 0.85114917 | 0.5053 | 0.551279283 | 0.551279283 | 2.73537298 | 1.50477111 | 0.534980573 | 0.25205 | 1.504771109 | 14.08314052 | 14.08314052 | 0.09159487 | -1.038128851 | FALSE | FALSE | FALSE |
| TBX2 | 0.137764 | 0.85116224 | 0.9916 | 8.806985053 | 140.9821076 | 564.2804722 | 0.85884743 | 0.605883936 | 0.5139 | 0.858847426 | 3.434507276 | 7.274488727 | 0.160405702 | -0.794780198 | FALSE | TRUE | FALSE |
| NCAPH | 0.1376745 | 0.85117532 | 1 | 0.137674517 | 0.137674517 | 2.716183847 | 1.54165105 | 0.531350309 | 0.50635 | 1.541651055 | 1.541651055 | 5.693087848 | 0.089303294 | -1.049132522 | FALSE | FALSE | FALSE |
| ZPBP2 | 0.1375864 | 0.85118839 | 0.50615 | 0.550305217 | 0.550305217 | 0.550305217 | 0.17507692 | 0.858301714 | 1 | 0.17507692 | 0.17507692 | 0.17507692 | 0.785862795 | -0.104653271 | FALSE | TRUE | FALSE |
| NUMB | 0.1374891 | 0.85120146 | 1 | 0.137489143 | 0.137489143 | 2.712639953 | 0.17350931 | 0.858724783 | 1 | 0.173509313 | 0.173509313 | 3.428877057 | 0.79240209 | -0.101054388 | FALSE | TRUE | FALSE |
| SUSD4 | 0.1374569 | 0.85121452 | 1 | 4506.612694 | 472775.8927 | 5013892.63 | 0.19589379 | 0.853081009 | 1 | 35.79556631 | 1656.324617 | 6624.554046 | 0.701690922 | -0.153854142 | FALSE | TRUE | FALSE |
| OR4D6 | 0.1374347 | 0.85122759 | 1 | 1085.293204 | 111753.2773 | 1174193.259 | 0.2206144 | 0.847074052 | 0.00795 | 0.220614403 | 0.220614403 | 0.220614403 | 0.622963561 | -0.205537356 | FALSE | TRUE | FALSE |
| KIF2B | 0.1374168 | 0.85124065 | 0.69195 | 0.306453212 | 4.900002109 | 4.900002109 | 0.01558967 | 0.894967876 | 0.7552 | 0.062320079 | 0.249188245 | 1.230284834 | 8.814602792 | 0.945202747 | FALSE | TRUE | FALSE |
| PSMC3 | 0.1373614 | 0.85125371 | 1 | 0.137361413 | 0.137361413 | 2.722904881 | 0.17371967 | 0.858687551 | 1 | 0.173719665 | 0.173719665 | 0.173719665 | 0.790707329 | -0.101984236 | FALSE | TRUE | FALSE |
| HOXD8 | 0.1372281 | 0.85126677 | 0.00695 | 0.137228098 | 0.137228098 | 0.137228098 | 0.16813176 | 0.860077052 | 0.5 | 0.869232224 | 1.570332683 | 1.570332683 | 0.816193768 | -0.088206726 | FALSE | TRUE | FALSE |
| MIP | 0.1371958 | 0.85127982 | 0.00875 | 0.137195759 | 0.137195759 | 0.137195759 | 0.17350992 | 0.858712375 | 1 | 0.173509922 | 0.173509922 | 0.173509922 | 0.790708433 | -0.10198363 | FALSE | TRUE | FALSE |
| LRRC61 | 0.1371564 | 0.85129288 | 1 | 0.137156389 | 0.137156389 | 0.137156389 | 0.16727339 | 0.860356661 | 1 | 1.518331982 | 1.518331982 | 1.518331982 | 0.819953419 | -0.086210819 | FALSE | TRUE | FALSE |
| PLEKHM2 | 0.1370894 | 0.85130593 | 0.02725 | 0.137089365 | 0.137089365 | 2.682118718 | 0.16776594 | 0.860223072 | 1 | 1.547905171 | 1.547905171 | 30.36376384 | 0.817146596 | -0.087700024 | FALSE | TRUE | FALSE |
| MLXIP | 0.1370648 | 0.85131898 | 1 | 0.137064844 | 0.137064844 | 2.689799999 | 0.17341725 | 0.85873719 | 0.0219 | 0.173417249 | 0.173417249 | 3.410888391 | 0.790376073 | -0.102166216 | FALSE | TRUE | FALSE |
| RAG1 | 0.1370537 | 0.85133202 | 0.01045 | 0.001667817 | 0.137053669 | 0.292505714 | 0.00102933 | 0.899408947 | 0.9404 | 0.037035662 | 1.36802185 | 1.36802185 | 133.1478828 | 2.124334265 | FALSE | TRUE | FALSE |
| FAM221B | 0.1370392 | 0.85134507 | 0.00705 | 0.137039197 | 0.137039197 | 0.137039197 | 0.17220067 | 0.859046683 | 1 | 0.172200667 | 0.172200667 | 0.172200667 | 0.79581107 | -0.099190024 | FALSE | TRUE | FALSE |
| CNNM2 | 0.1369705 | 0.85135811 | 1 | 0.547707875 | 0.547707875 | 10.69511866 | 0.17528057 | 0.858264265 | 1 | 0.175280571 | 0.175280571 | 3.432464401 | 0.781435818 | -0.107106686 | FALSE | TRUE | FALSE |
| SAMD7 | 0.1369331 | 0.85137115 | 1 | 1.24149851 | 1.24149851 | 2.716223064 | 0.17765264 | 0.857586947 | 1 | 0.177652642 | 0.177652642 | 0.177652642 | 0.770791423 | -0.113063126 | FALSE | TRUE | FALSE |
| VKORC1 | 0.136909 | 0.85138419 | 0.94235 | 5.128648588 | 197.2284718 | 197.2284718 | 1.60862322 | 0.524655931 | 0.87335 | 15.47897691 | 152.7674201 | 152.7674201 | 0.08510943 | -1.070022317 | FALSE | FALSE | FALSE |
| ENOX1 | 0.1369052 | 0.85139722 | 0.0199 | 0.136905232 | 0.136905232 | 2.700055137 | 0.17287458 | 0.85888591 | 1 | 0.172874581 | 0.172874581 | 3.415147119 | 0.791933845 | -0.101311096 | FALSE | TRUE | FALSE |
| PRSS33 | 0.1368914 | 0.85141026 | 1 | 0.136891362 | 0.136891362 | 0.136891362 | 0.17284726 | 0.858898289 | 0.0065 | 0.172847258 | 0.172847258 | 0.172847258 | 0.791978788 | -0.10128645 | FALSE | TRUE | FALSE |
| HBD | 0.1368813 | 0.85142329 | 1 | 0.136881327 | 0.136881327 | 0.136881327 | 0.16763511 | 0.860271678 | 1 | 1.55644047 | 1.55644047 | 1.55644047 | 0.816543322 | -0.088020769 | FALSE | TRUE | FALSE |
| GOLGA2 | 0.1368294 | 0.85143632 | 1 | 0.13682939 | 0.13682939 | 2.675262107 | 0.1725859 | 0.858935416 | 1 | 0.172585898 | 0.172585898 | 3.383476158 | 0.79281906 | -0.100825918 | FALSE | TRUE | FALSE |
| HHIPL2 | 0.1367987 | 0.85144934 | 0.75505 | 0.136798705 | 1.209525645 | 2.692437307 | 0.20844358 | 0.85010782 | 0.94165 | 1.859973457 | 159.7169432 | 159.7169432 | 0.656286472 | -0.182906548 | FALSE | TRUE | FALSE |
| TMUB2 | 0.1367656 | 0.85146237 | 1 | 0.136765577 | 0.136765577 | 0.136765577 | 0.17225364 | 0.859021972 | 1 | 0.172253637 | 0.172253637 | 0.172253637 | 0.793977877 | -0.100191599 | FALSE | TRUE | FALSE |
| ZNF430 | 0.1367003 | 0.85147539 | 1 | 0.136700348 | 0.136700348 | 2.709692405 | 13.9717209 | 0.163841874 | 0.2518 | 1.485681771 | 13.97172089 | 13.97172089 | 0.009784074 | -2.009480281 | FALSE | FALSE | FALSE |
| NCOA1 | 0.1366739 | 0.85148841 | 0.03735 | 0.136673904 | 0.136673904 | 4.594980426 | 1.5659637 | 0.529657519 | 0.03175 | 1.565963696 | 1.565963696 | 30.62260369 | 0.087277824 | -1.059096089 | FALSE | FALSE | FALSE |
| OR1J1 | 0.1366385 | 0.85150143 | 1 | 0.136638484 | 0.136638484 | 0.136638484 | 0.17178369 | 0.859219421 | 0.00685 | 0.171783693 | 0.171783693 | 0.171783693 | 0.795410096 | -0.099408901 | FALSE | TRUE | FALSE |
| SYNGR3 | 0.1366288 | 0.85151444 | 1 | 0.136628795 | 0.136628795 | 0.136628795 | 0.14163547 | 0.866417412 | 0.00645 | 0.141635466 | 0.141635466 | 0.141635466 | 0.964651003 | -0.01562978 | FALSE | TRUE | FALSE |
| IFIT1B | 0.1366089 | 0.85152746 | 1 | 0.546336433 | 0.546336433 | 2.709798621 | 0.17435189 | 0.858488667 | 1 | 0.174351895 | 0.174351895 | 0.174351895 | 0.783523871 | -0.105947768 | FALSE | TRUE | FALSE |
| CWC25 | 0.136602 | 0.85154047 | 1 | 0.136602021 | 0.136602021 | 2.707796981 | 0.17236308 | 0.858984889 | 1 | 0.172363079 | 0.172363079 | 3.420317966 | 0.792524835 | -0.10098712 | FALSE | TRUE | FALSE |
| FABP6 | 0.1365914 | 0.85155347 | 1 | 0.13659143 | 0.13659143 | 0.13659143 | 0.15451908 | 0.863433472 | 1 | 0.154519079 | 0.154519079 | 0.154519079 | 0.88397776 | -0.053558661 | FALSE | TRUE | FALSE |
| TINF2 | 0.1365672 | 0.85156648 | 1 | 0.136567171 | 0.136567171 | 2.710068805 | 0.17235105 | 0.858997252 | 1 | 0.172351045 | 0.172351045 | 0.172351045 | 0.792377969 | -0.101067608 | FALSE | TRUE | FALSE |
| UBE3C | 0.1363621 | 0.85157949 | 0.0288 | 0.136362077 | 0.136362077 | 2.668847685 | 0.17225006 | 0.859034329 | 0.02575 | 0.172250064 | 0.172250064 | 3.37993397 | 0.79165182 | -0.101465786 | FALSE | TRUE | FALSE |
| GFOD1 | 0.1363433 | 0.85159249 | 1 | 11.48974771 | 109.6010137 | 109.6010137 | 0.16156384 | 0.861643535 | 1 | 90.55760927 | 1288.234958 | 12950.15137 | 0.84389734 | -0.073710382 | FALSE | TRUE | FALSE |
| POLD2 | 0.1362431 | 0.85160549 | 1 | 1.250008776 | 11.77083897 | 24.72658202 | 0.17733604 | 0.857662518 | 1 | 1.634957197 | 1.634957197 | 3.512525084 | 0.768276657 | -0.114482362 | FALSE | TRUE | FALSE |
| FOXH1 | 0.1361832 | 0.85161849 | 0.5058 | 0.136183236 | 0.136183236 | 0.141583326 | 0.04979224 | 0.886700665 | 1 | 0.049792242 | 0.464999332 | 0.464999332 | 2.735029215 | 0.43696197 | FALSE | TRUE | FALSE |
| CIDEA | 0.1361584 | 0.85163148 | 1 | 0.136158412 | 0.136158412 | 0.136158412 | 0.17213411 | 0.859071386 | 1 | 0.172134109 | 0.172134109 | 0.172134109 | 0.791001926 | -0.101822459 | FALSE | TRUE | FALSE |

| | | | | | | | | | | | | | | | | | |
|---|---|---|---|---|---|---|---|---|---|---|---|---|---|---|---|---|---|
| TMEM130 | 0.1360729 | 0.85164448 | 1 | 1.233920821 | 1.233920821 | 2.695659574 | 0.1766624 | 0.857938931 | 1 | 0.176662398 | 0.176662398 | 3.503745084 | 0.770242444 | -0.113372553 | FALSE | TRUE | FALSE |
| IL21R | 0.1360717 | 0.85165747 | 1 | 0.136071731 | 0.136071731 | 2.689933687 | 0.16693347 | 0.860429426 | 1 | 1.563014086 | 1.563014086 | 3.304619581 | 0.81512552 | -0.08877551 | FALSE | TRUE | FALSE |
| ARRDC2 | 0.1359815 | 0.85167046 | 0.01235 | 0.135981542 | 0.135981542 | 2.692064692 | 0.16122126 | 0.861762662 | 1 | 1.469086536 | 13.7394436 | 13.7394436 | 0.843446738 | -0.073942337 | FALSE | TRUE | FALSE |
| YBX3 | 0.1359492 | 0.85168345 | 1 | 0.135949168 | 0.135949168 | 2.691989192 | 0.17116295 | 0.859281018 | 1 | 0.171162945 | 0.171162945 | 3.393320228 | 0.794267516 | -0.100033199 | FALSE | TRUE | FALSE |
| IFT43 | 0.1359027 | 0.85169643 | 1 | 0.135902738 | 0.135902738 | 0.135902738 | 1.41463876 | 0.544103475 | 0.87425 | 3.118008769 | 125.4079216 | 125.4079216 | 0.096068864 | -1.017417346 | FALSE | FALSE | FALSE |
| NEK2 | 0.1358962 | 0.85170941 | 0.5047 | 0.543514811 | 0.543514811 | 2.693162898 | 0.68597691 | 0.672358103 | 0.5064 | 0.685976911 | 0.685976911 | 3.402747807 | 0.198106084 | -0.703102186 | FALSE | TRUE | FALSE |
| PPP1R3C | 0.1358748 | 0.85172239 | 0.50235 | 1.236986461 | 1.236986461 | 1.251620199 | 0.17660345 | 0.85795147 | 1 | 0.176603451 | 0.176603451 | 0.176603451 | 0.769377702 | -0.113860405 | FALSE | TRUE | FALSE |
| PRDX1 | 0.1357016 | 0.85173537 | 0.00535 | 0.135701609 | 0.135701609 | 0.135701609 | 0.17084497 | 0.859379466 | 1 | 0.17084497 | 0.17084497 | 0.17084497 | 0.794296773 | -0.100017202 | FALSE | TRUE | FALSE |
| NTSR1 | 0.1356725 | 0.85174835 | 1 | 0.135672535 | 0.135672535 | 2.67626916 | 0.17137047 | 0.859256384 | 1 | 0.171370475 | 0.171370475 | 3.386019208 | 0.791691425 | -0.101444059 | FALSE | TRUE | FALSE |
| CARD9 | 0.1356078 | 0.85176132 | 1 | 0.135607798 | 0.135607798 | 2.674013511 | 0.17085548 | 0.859367168 | 1 | 0.170855484 | 0.170855484 | 3.374755279 | 0.79369883 | -0.10034426 | FALSE | TRUE | FALSE |
| ODAM | 0.1355786 | 0.8517743 | 0.505 | 0.135578587 | 0.135578587 | 0.135578587 | 1.37977831 | 0.547425972 | 0.8133 | 1.379778307 | 129.032317 | 1294.184339 | 0.098261138 | -1.007618211 | FALSE | FALSE | FALSE |
| ZNF669 | 0.1355267 | 0.85178727 | 1 | 1.232452986 | 11.50311193 | 24.38263928 | 12.4759285 | 0.175565221 | 0.78515 | 1173.247059 | 121519.0324 | 233762.0277 | 0.010863056 | -1.964048 | FALSE | FALSE | FALSE |
| FCER2 | 0.135439 | 0.85180024 | 0.01015 | 0.135438993 | 0.135438993 | 2.685929755 | 1.50279035 | 0.535396224 | 0.49535 | 0.165468358 | 1.502790352 | 1.520610247 | 0.090125008 | -1.045154682 | FALSE | FALSE | FALSE |
| TCERG1L | 0.1352346 | 0.8518132 | 1 | 1.224441662 | 1.224441662 | 2.678240517 | 0.17564421 | 0.858189316 | 1 | 0.175644207 | 0.175644207 | 3.482620275 | 0.769934984 | -0.113545947 | FALSE | TRUE | FALSE |
| ZNF510 | 0.1352183 | 0.85182617 | 0.50665 | 0.135218269 | 0.135218269 | 0.532794179 | 0.02591244 | 0.892178582 | 1 | 0.025912442 | 0.025912442 | 0.513232222 | 5.21827576 | 0.717527026 | FALSE | TRUE | FALSE |
| KLHL20 | 0.1351824 | 0.85183913 | 1 | 0.540611401 | 0.540611401 | 10.67335807 | 0.17296236 | 0.858823981 | 1 | 0.172962364 | 0.172962364 | 3.420095032 | 0.781571309 | -0.107031392 | FALSE | TRUE | FALSE |
| NAT10 | 0.1351317 | 0.85185209 | 0.21195 | 0.135131724 | 0.661405894 | 2.645784034 | 0.06529427 | 0.883523873 | 0.9376 | 1.04519654 | 35.64034202 | 142.683748 | 2.069580275 | 0.315882277 | FALSE | TRUE | FALSE |
| TMEM245 | 0.1350528 | 0.85186505 | 1 | 0.13505283 | 0.13505283 | 2.668026048 | 0.17034139 | 0.859539161 | 0.01545 | 0.170341388 | 0.170341388 | 3.370117951 | 0.792836264 | -0.100816494 | FALSE | TRUE | FALSE |
| SPNS2 | 0.1350385 | 0.851878 | 0.5118 | 0.540052039 | 0.540052039 | 2.6669266 | 0.17074267 | 0.85942864 | 1 | 0.682856972 | 0.682856972 | 3.377146284 | 0.790888983 | -0.101884474 | FALSE | TRUE | FALSE |
| SPSB2 | 0.1349718 | 0.85189096 | 0.75295 | 1.244141792 | 11.77038739 | 11.77038739 | 0.18155325 | 0.856686845 | 1 | 0.181553253 | 0.181553253 | 0.181553253 | 0.743428158 | -0.128760994 | FALSE | TRUE | FALSE |
| PYCR2 | 0.134944 | 0.85190391 | 0.00915 | 0.13494399 | 0.13494399 | 0.13494399 | 0.17078623 | 0.859404058 | 1 | 0.17078623 | 0.17078623 | 0.17078623 | 0.790133902 | -0.102299303 | FALSE | TRUE | FALSE |
| CRTAM | 0.1348599 | 0.85191686 | 0.0117 | 0.134859938 | 0.134859938 | 2.678299932 | 0.17039397 | 0.859514615 | 1 | 0.17039397 | 0.17039397 | 0.17039397 | 0.791459572 | -0.101571264 | FALSE | TRUE | FALSE |
| PAK4 | 0.1347912 | 0.85192981 | 1 | 0.539019345 | 0.539019345 | 4.56203285 | 0.17261679 | 0.858923043 | 0.01715 | 0.172616792 | 0.172616792 | 3.408445707 | 0.780869397 | -0.107421597 | FALSE | TRUE | FALSE |
| SYBU | 0.1347565 | 0.85194275 | 1 | 0.134756493 | 0.134756493 | 2.65559031 | 0.17049378 | 0.859490061 | 1 | 0.17049378 | 0.17049378 | 3.365782764 | 0.790389496 | -0.10215884 | FALSE | TRUE | FALSE |
| CLEC3A | 0.134709 | 0.8519557 | 1 | 0.134708955 | 0.134708955 | 0.134708955 | 0.1703368 | 0.85955143 | 1 | 0.170336797 | 0.170336797 | 0.170336797 | 0.790838838 | -0.101912011 | FALSE | TRUE | FALSE |
| MLST8 | 0.1346826 | 0.85196864 | 1 | 0.13468255 | 0.13468255 | 2.667694866 | 0.17022337 | 0.859612743 | 1 | 0.17022337 | 0.17022337 | 0.17022337 | 0.791210692 | -0.101707853 | FALSE | TRUE | FALSE |
| ZNF208 | 0.1346413 | 0.85198158 | 0.77455 | 1.140882849 | 88.0465713 | 333.8383173 | 1.9239E-05 | 0.900557784 | 0.98535 | 0.36451265 | 27.97332835 | 253.9750929 | 6998.447002 | 3.845001678 | FALSE | TRUE | FALSE |
| CENPH | 0.1346347 | 0.85199451 | 1 | 1.211975662 | 1.211975662 | 1.211975662 | 0.17467889 | 0.858413943 | 0.0046 | 0.174678889 | 0.174678889 | 0.174678889 | 0.770755497 | -0.113083369 | FALSE | TRUE | FALSE |
| HMMR | 0.1346265 | 0.85200745 | 0.8181 | 10.83103198 | 100.8596701 | 962.2064847 | 0.0289185 | 0.891503945 | 1 | 0.028918499 | 0.028918499 | 0.572779462 | 4.655376739 | 0.667954832 | FALSE | TRUE | FALSE |
| CCNF | 0.1345592 | 0.85202038 | 0.0245 | 0.134559205 | 0.134559205 | 2.641927643 | 0.15892621 | 0.862319957 | 1 | 0.158926207 | 0.158926207 | 3.127224694 | 0.846677257 | -0.072282106 | FALSE | TRUE | FALSE |
| ELMO2 | 0.1345512 | 0.85203331 | 0.0207 | 0.134551206 | 0.134551206 | 2.650076393 | 0.17000929 | 0.859698491 | 1 | 0.170009288 | 0.170009288 | 3.354579877 | 0.791434442 | -0.101585054 | FALSE | TRUE | FALSE |
| PDC | 0.1345109 | 0.85204624 | 0.0063 | 0.134510919 | 0.134510919 | 0.134510919 | 0.16947903 | 0.859833033 | 1 | 0.16947903 | 0.16947903 | 0.16947903 | 0.793672932 | -0.100358431 | FALSE | TRUE | FALSE |
| POLH | 0.1344161 | 0.85205916 | 0.81545 | 0.537683644 | 4.690076327 | 18.76566759 | 0.01523362 | 0.895075564 | 1 | 0.134152452 | 0.134152452 | 0.301011814 | 8.823643946 | 0.945647975 | FALSE | TRUE | FALSE |
| BCL2L1 | 0.1343908 | 0.85207209 | 1 | 1.274828711 | 1.274828711 | 1.274828711 | 0.12958156 | 0.869045946 | 1 | 0.129581565 | 0.129581565 | 0.129581565 | 1.037113827 | 0.015826424 | FALSE | TRUE | FALSE |
| OR11H12 | 0.1343906 | 0.85208501 | 1 | 12.17961582 | 1221.555618 | 1221.555618 | 13.9455527 | 0.164173494 | 0.2495 | 1.441531538 | 13.94555268 | 13.94555268 | 0.009636806 | -2.016066876 | FALSE | FALSE | FALSE |
| NSRP1 | 0.1343474 | 0.85209793 | 0.0131 | 0.13434744 | 0.13434744 | 2.661867648 | 0.16960284 | 0.859820812 | 0.0122 | 0.169602838 | 0.169602838 | 3.36416668 | 0.792129668 | -0.101203721 | FALSE | TRUE | FALSE |
| CCR3 | 0.134331 | 0.85211084 | 0.00865 | 0.134331048 | 0.134331048 | 0.134331048 | 0.16965407 | 0.859796365 | 1 | 0.169654069 | 0.169654069 | 0.169654069 | 0.791793847 | -0.101387877 | FALSE | TRUE | FALSE |
| LMX1B | 0.1342828 | 0.85212376 | 1 | 0.134282849 | 0.134282849 | 2.655830402 | 0.17039429 | 0.859502339 | 1 | 0.170394291 | 0.170394291 | 3.37454095 | 0.78807129 | -0.103434494 | FALSE | TRUE | FALSE |
| DEPDC1 | 0.1341731 | 0.85213667 | 0.7539 | 0.134173108 | 1.198352537 | 2.648037829 | 0.8797278 | 0.601418931 | 0.37575 | 0.400125887 | 3.519064764 | 4.348364921 | 0.152516618 | -0.816682834 | FALSE | TRUE | FALSE |
| FNDC3A | 0.1341552 | 0.85214958 | 1 | 0.13415525 | 0.13415525 | 4.516652722 | 0.17014295 | 0.859649505 | 0.0251 | 0.170142951 | 0.170142951 | 3.339813125 | 0.7884855 | -0.103206289 | FALSE | TRUE | FALSE |
| RGR | 0.1341542 | 0.85216249 | 1 | 0.134154204 | 0.134154204 | 0.134154204 | 1.50646616 | 0.534703018 | 0.50175 | 1.506466161 | 1.506466161 | 1.506466161 | 0.089052252 | -1.050355093 | FALSE | FALSE | FALSE |
| SERPINC1 | 0.1341328 | 0.85217539 | 1 | 0.536239736 | 0.536239736 | 2.656980821 | 0.68065592 | 0.674708724 | 0.5118 | 0.680655923 | 0.680655923 | 3.376205774 | 0.197064047 | -0.705392604 | FALSE | TRUE | FALSE |
| OR10P1 | 0.134093 | 0.8521883 | 1 | 0.134092988 | 0.134092988 | 0.134092988 | 1.49058121 | 0.536913958 | 0.49755 | 0.164035508 | 1.490581207 | 1.490581207 | 0.089960203 | -1.045949573 | FALSE | FALSE | FALSE |
| KRT79 | 0.1340144 | 0.8522012 | 0.66245 | 0.134014447 | 2.143696508 | 8.576929385 | 0.01774139 | 0.894382851 | 0.8758 | 0.070921535 | 0.283581056 | 0.350923318 | 7.553774972 | 0.878164043 | FALSE | TRUE | FALSE |
| SHKBP1 | 0.1339716 | 0.8522141 | 0.5157 | 0.133971566 | 0.133971566 | 2.627177123 | 0.02486715 | 0.892479326 | 1 | 0.217614907 | 0.217614907 | 4.277442023 | 5.387491421 | 0.731386592 | FALSE | TRUE | FALSE |
| SF3B4 | 0.1339405 | 0.85222699 | 1 | 0.133940548 | 0.133940548 | 2.655661349 | 0.13922042 | 0.866917653 | 1 | 0.139220419 | 0.139220419 | 2.763215145 | 0.962075459 | -0.016790863 | FALSE | TRUE | FALSE |
| MRPS35 | 0.1338813 | 0.85223989 | 1 | 0.133881324 | 0.133881324 | 0.133881324 | 0.16907256 | 0.859894111 | 0.00925 | 0.169072558 | 0.169072558 | 0.169072558 | 0.791857213 | -0.101353123 | FALSE | TRUE | FALSE |
| ZNF431 | 0.133818 | 0.85225278 | 1 | 0.133818027 | 0.133818027 | 2.652748544 | 0.1689478 | 0.85990632 | 1 | 0.168947802 | 0.168947802 | 3.352699191 | 0.792067287 | -0.101237923 | FALSE | TRUE | FALSE |
| CORIN | 0.1338042 | 0.85226567 | 0.1189 | 4.76708E-05 | 0.618814668 | 2.471507253 | 156.551438 | 0.017173384 | 0.1932 | 4.425166265 | 1418.903732 | 13160.91521 | 0.000854698 | -3.068187175 | FALSE | FALSE | FALSE |
| SFT2D2 | 0.1337565 | 0.85227856 | 1 | 0.13375654 | 0.13375654 | 0.13375654 | 0.16855251 | 0.859979538 | 1 | 0.168552506 | 0.168552506 | 0.168552506 | 0.793560078 | -0.100420189 | FALSE | TRUE | FALSE |
| SH2D4B | 0.1337268 | 0.85229145 | 0.01055 | 0.13372685 | 0.13372685 | 2.654388087 | 0.16362031 | 0.861201073 | 0.5118 | 1.485490777 | 1.485490777 | 1.485490777 | 0.817299797 | -0.087618609 | FALSE | TRUE | FALSE |
| PHF2 | 0.1337081 | 0.85230433 | 1 | 1.238854749 | 11.78080318 | 41.80245248 | 0.16425256 | 0.861056997 | 1 | 14.52878363 | 141.9832185 | 284.766708 | 0.814039768 | -0.089354378 | FALSE | TRUE | FALSE |
| STAMBP | 0.1336931 | 0.85231721 | 1 | 0.133693056 | 0.133693056 | 2.651420813 | 0.16926355 | 0.859857469 | 1 | 0.169263549 | 0.169263549 | 3.360248357 | 0.789851428 | -0.102454592 | FALSE | TRUE | FALSE |
| OR2L8 | 0.1336883 | 0.85233009 | 0.0086 | 0.133688347 | 0.133688347 | 0.133688347 | 0.16348265 | 0.861213067 | 0.5095 | 1.481428543 | 1.481428543 | 1.481428543 | 0.81775249 | -0.087378125 | FALSE | TRUE | FALSE |
| PRPF38B | 0.1336812 | 0.85234297 | 1 | 1.216477163 | 11.36153282 | 24.01852827 | 0.17950907 | 0.857157247 | 1 | 0.179509075 | 0.179509075 | 3.549738561 | 0.744704289 | -0.128016145 | FALSE | TRUE | FALSE |
| RNF157 | 0.1335648 | 0.85235585 | 0.0213 | 0.133564789 | 0.133564789 | 2.631268867 | 0.16894603 | 0.859918528 | 1 | 0.168946035 | 0.168946035 | 3.334293252 | 0.790576643 | -0.102056021 | FALSE | TRUE | FALSE |
| RAB3GAP1 | 0.1335426 | 0.85236872 | 0.51435 | 0.133542572 | 0.133542572 | 2.623551374 | 1.85241174 | 0.503644682 | 0.5052 | 1.852411744 | 17.16825416 | 36.46971819 | 0.072091193 | -1.14211779 | FALSE | FALSE | FALSE |
| RFC5 | 0.1334968 | 0.85238159 | 1 | 0.133496824 | 0.133496824 | 0.133496824 | 1.51676821 | 0.534147069 | 0.5029 | 1.51676821 | 1.51676821 | 1.51676821 | 0.088013991 | -1.055448285 | FALSE | FALSE | FALSE |
| SPO11 | 0.1334376 | 0.85239446 | 1 | 1.212250117 | 1.212250117 | 2.650070574 | 0.16662919 | 0.860550534 | 1 | 1.521484168 | 6.085467822 | 6.085467822 | 0.8008058 | -0.09647279 | FALSE | TRUE | FALSE |
| EAPP | 0.133408 | 0.85240733 | 0.0068 | 0.133408031 | 0.133408031 | 0.133408031 | 0.16836945 | 0.860003929 | 1 | 0.16836945 | 0.16836945 | 0.16836945 | 0.792352954 | -0.101081319 | FALSE | TRUE | FALSE |
| SNTA1 | 0.1334069 | 0.85242019 | 0.01265 | 0.133406909 | 0.133406909 | 2.641862643 | 0.16909754 | 0.859869685 | 1 | 0.169097543 | 0.169097543 | 3.352612476 | 0.788934641 | -0.102958974 | FALSE | TRUE | FALSE |
| E2F7 | 0.1333788 | 0.85243305 | 0.02275 | 0.133378806 | 0.133378806 | 2.622328019 | 0.16907334 | 0.859881899 | 1 | 0.169073344 | 0.169073344 | 3.330897697 | 0.788881338 | -0.102988318 | FALSE | TRUE | FALSE |
| FBXO39 | 0.1333767 | 0.85244591 | 1 | 0.533490558 | 0.533490558 | 2.642173431 | 0.17026027 | 0.859600485 | 0.01195 | 0.170260271 | 0.170260271 | 3.376689091 | 0.783369369 | -0.106033414 | FALSE | TRUE | FALSE |
| FH | 0.1332123 | 0.85245877 | 0.6968 | 0.133212346 | 2.128550268 | 2.128550268 | 0.29836232 | 0.82583862 | 0.0103 | 0.060060313 | 0.240029116 | 0.298362316 | 0.446478455 | -0.350199493 | FALSE | TRUE | FALSE |
| STX2 | 0.1331782 | 0.85247163 | 1 | 0.133178168 | 0.133178168 | 0.133178168 | 0.16803057 | 0.86010141 | 1 | 0.168030575 | 0.168030575 | 0.168030575 | 0.792582944 | -0.100955277 | FALSE | TRUE | FALSE |
| CABP2 | 0.1331252 | 0.85248448 | 1 | 0.13312525 | 0.13312525 | 0.13312525 | 0.66646591 | 0.682400327 | 0.50195 | 0.666465908 | 0.666465908 | 0.666465908 | 0.199748027 | -0.699517503 | FALSE | TRUE | FALSE |
| ICAM3 | 0.1331165 | 0.85249733 | 0.7548 | 0.133116504 | 1.167415168 | 2.6207172 | 0.02635512 | 0.892057847 | 0.01765 | 0.026355124 | 0.026355124 | 0.519839943 | 5.0508775 | 0.703366836 | FALSE | TRUE | FALSE |
| PDCD4 | 0.1330822 | 0.85251018 | 0.0125 | 0.133082209 | 0.133082209 | 2.640326083 | 5.89026249 | 0.272916315 | 0.2573 | 1.472676323 | 5.89026249 | 5.89026249 | 0.022593596 | -1.646014647 | FALSE | FALSE | FALSE |
| IL20RB | 0.1329832 | 0.85252303 | 1 | 0.132983156 | 0.132983156 | 0.132983156 | 0.16789922 | 0.860162268 | 1 | 0.167899216 | 0.167899216 | 0.167899216 | 0.792041556 | -0.101252032 | FALSE | TRUE | FALSE |
| CACFD1 | 0.1327562 | 0.85253588 | 1 | 0.132756206 | 0.132756206 | 0.132756206 | 0.16818172 | 0.860052686 | 0.0047 | 0.168181723 | 0.168181723 | 0.168181723 | 0.789361675 | -0.102723963 | FALSE | TRUE | FALSE |
| ADNP2 | 0.1327477 | 0.85254872 | 1 | 0.132747702 | 0.132747702 | 2.598201254 | 0.16325758 | 0.861261023 | 1 | 1.519799897 | 1.519799897 | 29.82283946 | 0.813118144 | -0.089846348 | FALSE | TRUE | FALSE |
| ROR1 | 0.1327103 | 0.85256156 | 0.0254 | 0.132710306 | 0.132710306 | 2.605908988 | 0.16667626 | 0.860538433 | 1 | 0.66649673 | 0.66649673 | 13.11605149 | 0.79621602 | -0.098969089 | FALSE | TRUE | FALSE |
| HDC | 0.1326636 | 0.8525744 | 1 | 0.132663635 | 0.132663635 | 2.616473929 | 0.16312758 | 0.861308948 | 1 | 1.515516588 | 1.515516588 | 3.222672543 | 0.813250811 | -0.089775495 | FALSE | TRUE | FALSE |

| | | | | | | | | | | | | | | | | | |
|---|---|---|---|---|---|---|---|---|---|---|---|---|---|---|---|---|---|
| NCR2 | 0.1326107 | 0.85258724 | 1 | 0.132610697 | 0.132610697 | 0.132610697 | 0.00015786 | 0.900094793 | 1 | 0.000157859 | 0.000157859 | 0.000157859 | 840.0604984 | 2.924310564 | FALSE | TRUE | FALSE |
| APBB3 | 0.1325547 | 0.85260007 | 1 | 0.132554702 | 0.132554702 | 2.621704814 | 0.16763789 | 0.860259529 | 1 | 0.167637892 | 0.167637892 | 3.320012048 | 0.790720409 | -0.101977052 | FALSE | TRUE | FALSE |
| ZNF236 | 0.1325295 | 0.85261291 | 1 | 0.132529532 | 2.540773756 | 4.444578434 | 0.16815299 | 0.86006487 | 0.0485 | 0.16815299 | 0.16815299 | 5.648516176 | 0.788148531 | -0.103391929 | FALSE | TRUE | FALSE |
| SLC15A2 | 0.1324804 | 0.85262574 | 0.50835 | 0.132480433 | 0.132480433 | 0.51788198 | 0.02463671 | 0.892554272 | 0.50615 | 0.215388975 | 0.215388975 | 0.833782749 | 5.377358677 | 0.730569005 | FALSE | TRUE | FALSE |
| KRT16 | 0.132408 | 0.85263857 | 0.50735 | 0.299453743 | 4.786135419 | 4.786135419 | 0.52814903 | 0.743824977 | 0.50075 | 0.528149032 | 8.44315746 | 8.44315746 | 0.250701871 | -0.600842425 | FALSE | TRUE | FALSE |
| EPCAM | 0.1324007 | 0.85265139 | 1 | 0.132400729 | 0.132400729 | 0.132400729 | 0.16802356 | 0.860113586 | 0.0074 | 0.168023556 | 0.168023556 | 0.168023556 | 0.78798909 | -0.103479795 | FALSE | TRUE | FALSE |
| PCDHGB6 | 0.1323828 | 0.85266422 | 0.9698 | 18.1631099 | 2558.740413 | 23070.70648 | 0.31165743 | 0.821490574 | 0.8928 | 4.987449842 | 319.8848311 | 2786.486094 | 0.424770294 | -0.371845863 | FALSE | TRUE | FALSE |
| TMEM14A | 0.1323745 | 0.85267704 | 1 | 0.132374488 | 0.132374488 | 0.132374488 | 0.13280488 | 0.868245812 | 1 | 0.132804877 | 1.253220465 | 1.253220465 | 0.996759241 | -0.001409729 | FALSE | TRUE | FALSE |
| SERPINA1 | 0.1322591 | 0.85268986 | 1 | 1.61861E+61 | 3.0215E+72 | 1.85992E+77 | 0.02332181 | 0.892897783 | 1 | 4.47664E+57 | 1.81031E+68 | 7.56197E+72 | 5.671046572 | 0.753663214 | FALSE | TRUE | FALSE |
| ZNF19 | 0.1321808 | 0.85270268 | 1 | 107.7880541 | 10658.34162 | 109831.7397 | 133.82164 | 0.021505469 | 0.77505 | 1319.912586 | 137967.3202 | 264876.8465 | 0.000987739 | -3.005357978 | FALSE | FALSE | FALSE |
| AKR1B15 | 0.1321443 | 0.8527155 | 1 | 0.132144288 | 0.132144288 | 0.132144288 | 0.16688736 | 0.860453665 | 1 | 0.166887358 | 0.166887358 | 0.166887358 | 0.791817245 | -0.101375044 | FALSE | TRUE | FALSE |
| COQ6 | 0.1321028 | 0.85272831 | 1 | 0.132102786 | 0.132102786 | 2.619031086 | 0.16722228 | 0.860368793 | 1 | 0.167222278 | 0.167222278 | 3.318774661 | 0.789983173 | -0.102382159 | FALSE | TRUE | FALSE |
| COG7 | 0.1320719 | 0.85274112 | 1 | 0.132071882 | 0.132071882 | 2.5980999 | 0.16703828 | 0.860393053 | 1 | 0.167038279 | 0.167038279 | 3.29244409 | 0.790668364 | -0.102005638 | FALSE | TRUE | FALSE |
| CCDC116 | 0.132012 | 0.85275393 | 1 | 1.1970151 | 1.1970151 | 4.470060897 | 0.1720287 | 0.859096081 | 1 | 0.1720287 | 0.1720287 | 3.395780607 | 0.767383353 | -0.114987626 | FALSE | TRUE | FALSE |
| MITD1 | 0.1319121 | 0.85276674 | 1 | 0.527475837 | 0.527475837 | 0.527475837 | 1.52275566 | 0.533868781 | 0.49735 | 0.164088071 | 1.522755656 | 1.522755656 | 0.086627254 | -1.062345451 | FALSE | FALSE | FALSE |
| AKR7A3 | 0.1319084 | 0.85277955 | 0.49425 | 0.015501414 | 0.131908421 | 0.131908421 | 0.0247301 | 0.892531798 | 1 | 0.024730095 | 0.213020095 | 0.213020095 | 5.33392285 | 0.72704673 | FALSE | TRUE | FALSE |
| SYNPO | 0.1318879 | 0.85279235 | 1 | 14640354.68 | 21638822777 | 3.11522E+12 | 0.13466928 | 0.867842288 | 1 | 1.88331E+11 | 3.52599E+14 | 5.89699E+16 | 0.979346512 | -0.009063619 | FALSE | TRUE | FALSE |
| PRPF39 | 0.1318589 | 0.85280515 | 0.01495 | 0.13185889 | 0.13185889 | 0.472611769 | 0.01551598 | 0.894996613 | 1 | 0.015515982 | 0.015515982 | 0.307538439 | 8.498262559 | 0.929330145 | FALSE | TRUE | FALSE |
| OR52E8 | 0.131856 | 0.85281795 | 1 | 1.190033135 | 1.190033135 | 1.190033135 | 0.16649707 | 0.860598919 | 1 | 0.166497065 | 1.511422102 | 1.511422102 | 0.79194174 | -0.101306766 | FALSE | TRUE | FALSE |
| TMBIM1 | 0.1318492 | 0.85283075 | 1 | 0.131849187 | 0.131849187 | 2.618911266 | 0.16659288 | 0.860562633 | 1 | 0.166592876 | 0.166592876 | 0.166592876 | 0.791445531 | -0.101578969 | FALSE | TRUE | FALSE |
| PCDHGA3 | 0.1318252 | 0.85284354 | 0.89415 | 2.108499324 | 89.97611964 | 680.9268095 | 9.15875864 | 0.209436097 | 0.31165 | 2.290203153 | 84.21692299 | 336.8763395 | 0.014393347 | -1.841838198 | FALSE | FALSE | FALSE |
| NQO1 | 0.1318112 | 0.85285633 | 0.5112 | 0.527242631 | 0.527242631 | 0.527242631 | 0.03453118 | 0.890232961 | 1 | 0.312594291 | 0.312594291 | 0.312594291 | 3.817165795 | 0.581741023 | FALSE | TRUE | FALSE |
| NCOA7 | 0.1317345 | 0.85286912 | 1 | 0.131734519 | 0.131734519 | 2.591255282 | 0.16683675 | 0.860477895 | 1 | 0.166836751 | 0.166836751 | 3.288239085 | 0.789601322 | -0.102592133 | FALSE | TRUE | FALSE |
| LRRC66 | 0.1316734 | 0.85288191 | 0.9378 | 1.157128313 | 96.37930335 | 96.37930335 | 0.23308041 | 0.844040024 | 0.512 | 0.233080406 | 0.233080406 | 0.907988721 | 0.564927125 | -0.248007572 | FALSE | TRUE | FALSE |
| PLXNA1 | 0.1316635 | 0.8528947 | 0.98235 | 1189.801907 | 130592.1456 | 1185951.296 | 0.02225335 | 0.893143012 | 1 | 19341.92586 | 1773786.677 | 33766204.71 | 5.916570605 | 0.772070051 | FALSE | TRUE | FALSE |
| DHRS7 | 0.1316322 | 0.85290748 | 0.0104 | 0.131632151 | 0.131632151 | 2.61557288 | 1.49221832 | 0.536776368 | 0.5011 | 1.492218323 | 1.492218323 | 1.492218323 | 0.088212394 | -1.05447039 | FALSE | FALSE | FALSE |
| GBX2 | 0.1316023 | 0.85292026 | 1 | 0.131602316 | 0.131602316 | 2.604661536 | 0.16734637 | 0.860332391 | 0.011 | 0.167346367 | 0.167346367 | 3.316250077 | 0.78640677 | -0.104352756 | FALSE | TRUE | FALSE |
| MRPL21 | 0.1315299 | 0.85293304 | 0.0038 | 0.131529919 | 0.131529919 | 0.131529919 | 0.16626073 | 0.860671434 | 1 | 0.166260726 | 0.166260726 | 0.166260726 | 0.791106366 | -0.101765121 | FALSE | TRUE | FALSE |
| ESPL1 | 0.1314419 | 0.85294582 | 1 | 0.131441915 | 2.518560766 | 4.391951722 | 0.16691132 | 0.860441546 | 1 | 0.166911318 | 3.214391015 | 5.588693234 | 0.787495516 | -0.10375191 | FALSE | TRUE | FALSE |
| PSG11 | 0.1314253 | 0.8529586 | 1 | 0.131425253 | 0.131425253 | 0.131425253 | 0.16600635 | 0.860755942 | 1 | 0.166006348 | 0.166006348 | 0.166006348 | 0.791688119 | -0.101445873 | FALSE | TRUE | FALSE |
| FLOT1 | 0.1314235 | 0.85297137 | 0.0138 | 0.131423492 | 0.131423492 | 2.602664497 | 0.16674882 | 0.860502116 | 1 | 0.166748823 | 0.166748823 | 3.30613459 | 0.788152444 | -0.103389774 | FALSE | TRUE | FALSE |
| STRN | 0.1313811 | 0.85298414 | 1 | 0.131381106 | 0.131381106 | 2.59359589 | 0.16697443 | 0.860405179 | 0.01795 | 0.16697443 | 0.16697443 | 3.301380416 | 0.786833681 | -0.104117058 | FALSE | TRUE | FALSE |
| PRSS12 | 0.1313663 | 0.85299691 | 0.66085 | 0.524935644 | 42.69746373 | 170.660374 | 0.00257311 | 0.898848444 | 0.7557 | 0.005415176 | 0.021641606 | 0.086888889 | 51.05343842 | 1.708024997 | FALSE | TRUE | FALSE |
| NMBR | 0.1312948 | 0.85300968 | 1 | 1.20414763 | 1.20414763 | 2.601322768 | 13.1395892 | 0.169183343 | 0.3221 | 13.13958917 | 1258.761698 | 1258.761698 | 0.009992309 | -2.000334135 | FALSE | FALSE | FALSE |
| SHANK1 | 0.1312833 | 0.85302244 | 0.5312 | 1.22006528 | 2.530641019 | 23.51820952 | 0.17198292 | 0.859120767 | 1 | 0.171982919 | 0.171982919 | 5.772974317 | 0.763350583 | -0.117275959 | FALSE | TRUE | FALSE |
| SRRM5 | 0.1312738 | 0.8530352 | 0.6891 | 0.131273774 | 9.444273996 | 9.444273996 | 0.26433249 | 0.835428057 | 0.6921 | 0.264332494 | 19.74204688 | 19.74204688 | 0.496623674 | -0.303972582 | FALSE | TRUE | FALSE |
| CXCL9 | 0.1312366 | 0.85304796 | 1 | 0.131236589 | 0.131236589 | 0.131236589 | 0.1429927 | 0.866115594 | 1 | 0.142992701 | 0.142992701 | 0.142992701 | 0.917785229 | -0.037258936 | FALSE | TRUE | FALSE |
| RFPL1 | 0.1311622 | 0.85306072 | 0.0091 | 0.131162232 | 0.131162232 | 0.131162232 | 1.35575587 | 0.550311738 | 0.50795 | 1.355755874 | 1.355755874 | 1.355755874 | 0.096744727 | -1.014372695 | FALSE | FALSE | FALSE |
| SLC39A6 | 0.1311085 | 0.85307348 | 0.0213 | 0.131108515 | 0.131108515 | 2.585893683 | 0.16595901 | 0.860768006 | 1 | 0.165959008 | 0.165959008 | 3.278710415 | 0.790005414 | -0.102369932 | FALSE | TRUE | FALSE |
| CTSK | 0.1310844 | 0.85308623 | 0.50665 | 0.131084352 | 0.131084352 | 0.131084352 | 0.04806069 | 0.887111438 | 0.5067 | 0.442790599 | 0.442790599 | 0.442790599 | 2.727475335 | 0.435760832 | FALSE | TRUE | FALSE |
| PHYH | 0.1310576 | 0.85309898 | 0.50615 | 0.131057642 | 0.523985379 | 0.523985379 | 0.01472963 | 0.895197374 | 0.5025 | 0.058868669 | 0.058868669 | 0.058868669 | 8.897553644 | 0.949270615 | FALSE | TRUE | FALSE |
| SLC38A11 | 0.1310013 | 0.85311173 | 0.5096 | 0.13100131 | 0.13100131 | 0.13100131 | 0.0194668 | 0.893960308 | 0.00845 | 0.0194668 | 0.0194668 | 0.0194668 | 6.729473288 | 0.827981074 | FALSE | TRUE | FALSE |
| ZAR1 | 0.1309975 | 0.85312448 | 1 | 0.130997469 | 0.130997469 | 0.130997469 | 0.14276984 | 0.866216346 | 0.00545 | 0.14276984 | 0.14276984 | 0.14276984 | 0.917543014 | -0.037373567 | FALSE | TRUE | FALSE |
| DUSP26 | 0.1309939 | 0.85313722 | 0.00735 | 0.130993907 | 0.130993907 | 0.130993907 | 0.16629604 | 0.860659353 | 1 | 0.16629604 | 0.16629604 | 0.16629604 | 0.787715132 | -0.103630812 | FALSE | TRUE | FALSE |
| PPP1R12A | 0.1309766 | 0.85314997 | 1 | 0.130976604 | 0.130976604 | 2.576499498 | 0.1660715 | 0.860731807 | 0.02155 | 0.166071502 | 0.166071502 | 3.273327468 | 0.788675981 | -0.103101385 | FALSE | TRUE | FALSE |
| PSD4 | 0.1309601 | 0.85316271 | 0.0296 | 0.130960108 | 0.130960108 | 2.56170106 | 0.16633113 | 0.86064727 | 1 | 0.166331125 | 0.166331125 | 3.262195343 | 0.787345771 | -0.103834501 | FALSE | TRUE | FALSE |
| TBCB | 0.1309414 | 0.85317545 | 1 | 0.13094138 | 0.13094138 | 0.13094138 | 10.1870692 | 0.196237749 | 0.32195 | 2.546653205 | 40.76028503 | 40.76028503 | 0.012853685 | -1.890972344 | FALSE | FALSE | FALSE |
| C1QTNF1 | 0.1308626 | 0.85318818 | 1 | 1.220863345 | 1.220863345 | 1.220863345 | 0.17162328 | 0.859231744 | 1 | 0.171623279 | 0.171623279 | 0.171623279 | 0.762499217 | -0.117760598 | FALSE | TRUE | FALSE |
| PLCH1 | 0.1308584 | 0.85320092 | 0.7571 | 0.130858387 | 0.523411955 | 4.408013141 | 3.88840081 | 0.361827397 | 0.2673 | 1.676571971 | 15.54966607 | 32.97885554 | 0.033653523 | -1.472969463 | FALSE | FALSE | FALSE |
| ANGPT2 | 0.1308292 | 0.85321365 | 1 | 0.130829201 | 0.130829201 | 2.591464718 | 0.16642393 | 0.860623098 | 1 | 0.166423926 | 0.166423926 | 3.300331579 | 0.786120146 | -0.104511074 | FALSE | TRUE | FALSE |
| FERMT2 | 0.1308282 | 0.85322638 | 1 | 0.523089849 | 0.523089849 | 4.425105816 | 0.16834317 | 0.860016122 | 1 | 0.168343166 | 0.168343166 | 3.330642128 | 0.777152008 | -0.109494026 | FALSE | TRUE | FALSE |
| PCDHGC4 | 0.130818 | 0.8532391 | 1 | 0.130817968 | 0.130817968 | 2.560245484 | 0.16451223 | 0.86099688 | 0.5172 | 0.657889017 | 0.657889017 | 12.9088445 | 0.79518687 | -0.099530799 | FALSE | TRUE | FALSE |
| WNT7B | 0.1307568 | 0.85325183 | 1 | 0.130756819 | 0.130756819 | 2.58248901 | 0.15138733 | 0.864277449 | 1 | 13.67240757 | 135.0100319 | 135.0100319 | 0.86372364 | -0.063625194 | FALSE | TRUE | FALSE |
| ACBD4 | 0.1307357 | 0.85326455 | 1 | 0.1307357 | 0.1307357 | 2.595813177 | 1.45648726 | 0.539515473 | 0.5044 | 1.456487255 | 1.456487255 | 1.456487255 | 0.089760964 | -1.046912493 | FALSE | FALSE | FALSE |
| ECHS1 | 0.130714 | 0.85327727 | 1 | 1.199754881 | 1.199754881 | 1.199754881 | 0.17096981 | 0.859354866 | 1 | 0.170969814 | 0.170969814 | 0.170969814 | 0.764544405 | -0.116597286 | FALSE | TRUE | FALSE |
| ST8SIA5 | 0.1306779 | 0.85328999 | 1 | 0.13067792 | 0.13067792 | 2.586407851 | 0.16544875 | 0.860828301 | 1 | 0.165448754 | 0.165448754 | 3.278692453 | 0.78983925 | -0.102461288 | FALSE | TRUE | FALSE |
| NECAB3 | 0.1306529 | 0.85330271 | 1 | 0.130652867 | 0.130652867 | 0.130652867 | 0.14926972 | 0.864758526 | 1 | 0.149269723 | 0.149269723 | 0.149269723 | 0.875280427 | -0.057852783 | FALSE | TRUE | FALSE |
| POLR2E | 0.1306503 | 0.85331542 | 1 | 0.130650318 | 0.130650318 | 0.130650318 | 0.16585771 | 0.860792129 | 1 | 0.165857712 | 0.165857712 | 0.165857712 | 0.78772531 | -0.1036252 | FALSE | TRUE | FALSE |
| QSER1 | 0.1306286 | 0.85332813 | 1 | 0.130628588 | 0.130628588 | 4.383310746 | 0.16605326 | 0.860743875 | 1 | 0.16605326 | 0.16605326 | 5.580769525 | 0.786666803 | -0.104209176 | FALSE | TRUE | FALSE |
| RABEP2 | 0.1306144 | 0.85334084 | 1 | 1.192113027 | 11.16713682 | 23.49268456 | 0.17068664 | 0.859440929 | 1 | 1.566355589 | 1.566355589 | 5.78023624 | 0.765229285 | -0.116208418 | FALSE | TRUE | FALSE |
| RNF169 | 0.1305574 | 0.85335355 | 1 | 0.130557375 | 0.130557375 | 2.582109127 | 0.15873843 | 0.862378991 | 1 | 1.455236046 | 5.82041481 | 12.5735065 | 0.822468611 | -0.084880668 | FALSE | TRUE | FALSE |
| HTR5A | 0.1304907 | 0.85336626 | 1 | 0.130490711 | 0.130490711 | 2.580335894 | 0.64812613 | 0.690765728 | 0.75235 | 0.648126125 | 2.59254966 | 3.209127264 | 0.201335366 | -0.696079931 | FALSE | TRUE | FALSE |
| GLI3 | 0.1304438 | 0.85337896 | 0.7639 | 0.306615822 | 1.225687376 | 23.54352514 | 0.05105067 | 0.886370055 | 1 | 0.051050666 | 0.051050666 | 0.985288141 | 2.555183105 | 0.407422027 | FALSE | TRUE | FALSE |
| C10orf120 | 0.1304142 | 0.85339166 | 1 | 0.130414166 | 0.130414166 | 0.130414166 | 0.16492837 | 0.860912637 | 1 | 0.164928374 | 0.164928374 | 3.278312534 | 0.790732139 | -0.101970609 | FALSE | TRUE | FALSE |
| C4BPB | 0.1303994 | 0.85340436 | 1 | 0.13039939 | 0.13039939 | 0.13039939 | 0.16478805 | 0.860948753 | 1 | 0.164788054 | 0.164788054 | 0.164788054 | 0.791315797 | -0.101650165 | FALSE | TRUE | FALSE |
| DCTD | 0.1303866 | 0.85341706 | 1 | 0.130386551 | 0.130386551 | 0.130386551 | 1.47260615 | 0.538286174 | 0.00445 | 0.160343778 | 1.472606149 | 1.472606149 | 0.08854136 | -1.052853812 | FALSE | FALSE | FALSE |
| ADCYAP1R1 | 0.1303715 | 0.85342975 | 0.50935 | 0.521369718 | 0.521369718 | 4.41439937 | 0.16741989 | 0.860295969 | 0.0137 | 0.167419889 | 0.167419889 | 3.319671598 | 0.778710002 | -0.108624247 | FALSE | TRUE | FALSE |
| GRIN1 | 0.1303575 | 0.85344244 | 1 | 0.130357533 | 0.130357533 | 2.540308476 | 1.50159845 | 0.535534612 | 0.0288 | 1.501598455 | 1.501598455 | 29.3524221 | 0.086812512 | -1.061417679 | FALSE | FALSE | FALSE |
| TCN2 | 0.1303344 | 0.85345513 | 1 | 0.130334355 | 0.130334355 | 2.584487863 | 0.15998829 | 0.862047778 | 1 | 1.45417516 | 1.45417516 | 3.175765233 | 0.814649316 | -0.089029303 | FALSE | TRUE | FALSE |
| OSCAR | 0.1303127 | 0.85346782 | 1 | 0.13031269 | 0.13031269 | 0.13031269 | 1.44662172 | 0.541009426 | 0.50265 | 1.446621719 | 1.446621719 | 1.446621719 | 0.090080695 | -1.045368272 | FALSE | FALSE | FALSE |
| CAPN7 | 0.1302853 | 0.85348051 | 0.7531 | 0.13028529 | 1.183448557 | 2.571292736 | 0.02545081 | 0.89232915 | 0.01725 | 0.025450814 | 0.025450814 | 0.503092066 | 5.11910112 | 0.709193708 | FALSE | TRUE | FALSE |
| LAMA5 | 0.1302207 | 0.85349319 | 0.3039 | 0.032572779 | 2.366656309 | 20.2971821 | 0.01793874 | 0.894353807 | 0.17035 | 0.001122727 | 0.071732082 | 0.329637665 | 7.259187622 | 0.860888021 | FALSE | TRUE | FALSE |
| KCNA1 | 0.1301609 | 0.85350587 | 1 | 0.130160914 | 0.130160914 | 2.566286922 | 0.63496335 | 0.697388671 | 0.75085 | 1.496907074 | 5.985179245 | 12.5404798 | 0.204989648 | -0.68826807 | FALSE | TRUE | FALSE |

| | | | | | | | | | | | | | | | | | |
|---|---|---|---|---|---|---|---|---|---|---|---|---|---|---|---|---|---|
| RAB28 | 0.1301293 | 0.85351855 | 1 | 0.520406441 | 0.520406441 | 0.520406441 | 0.16187804 | 0.861595828 | 1 | 0.161878036 | 1.487415581 | 1.487415581 | 0.80387221 | -0.094812985 | FALSE | TRUE | FALSE |
| MTCH2 | 0.1301207 | 0.85353123 | 1 | 22289662297 | 2.33899E+13 | 3.78576E+14 | 0.15480378 | 0.863352015 | 1 | 5.29178E+11 | 2.24426E+15 | 3.63992E+16 | 0.840552439 | -0.075435187 | FALSE | TRUE | FALSE |
| FBXL4 | 0.1301098 | 0.85354391 | 1 | 0.130109761 | 0.130109761 | 2.574613001 | 0.16477696 | 0.860960787 | 0.01355 | 0.164776962 | 0.164776962 | 3.264765486 | 0.789611358 | -0.102586613 | FALSE | TRUE | FALSE |
| TSPEAR | 0.1300974 | 0.85355658 | 0.8148 | 1.153581371 | 41.91929583 | 390.3679999 | 0.27713226 | 0.831776931 | 0.8167 | 2.490093683 | 22.93531762 | 216.4192124 | 0.469441472 | -0.328418546 | FALSE | TRUE | FALSE |
| SLCO5A1 | 0.1300743 | 0.85356925 | 0.76 | 0.520185469 | 4.648921645 | 10.20527353 | 0.02590425 | 0.89218612 | 0.0205 | 0.025904252 | 0.025904252 | 0.509370577 | 5.021349002 | 0.700820407 | FALSE | TRUE | FALSE |
| AS3MT | 0.1300545 | 0.85358192 | 1 | 1.172363859 | 1.172363859 | 1.172363859 | 0.16947494 | 0.859845252 | 1 | 0.169474939 | 0.169474939 | 0.169474939 | 0.767396656 | -0.114980097 | FALSE | TRUE | FALSE |
| TNPO3 | 0.1300225 | 0.85359459 | 0.02845 | 0.130022459 | 0.130022459 | 2.551422611 | 0.16006988 | 0.862012199 | 1 | 1.473285609 | 1.473285609 | 28.97582471 | 0.812285588 | -0.090291252 | FALSE | TRUE | FALSE |
| TNFAIP1 | 0.1299354 | 0.85360725 | 1 | 0.129935366 | 0.129935366 | 2.576234564 | 0.16493276 | 0.860900594 | 0.0108 | 0.164932757 | 0.164932757 | 3.27353055 | 0.787808129 | -0.103579542 | FALSE | TRUE | FALSE |
| CDNF | 0.1298904 | 0.85361991 | 1 | 0.129890359 | 0.129890359 | 0.129890359 | 0.15955945 | 0.862166244 | 1 | 1.453619197 | 1.453619197 | 1.453619197 | 0.814056193 | -0.089345615 | FALSE | TRUE | FALSE |
| GJC2 | 0.1298497 | 0.85363257 | 0.50815 | 0.129849713 | 0.129849713 | 0.643176313 | 0.19239106 | 0.853786935 | 0.7534 | 0.192391057 | 0.76923464 | 0.954034878 | 0.674925928 | -0.170743888 | FALSE | TRUE | FALSE |
| MYOCD | 0.1298478 | 0.85364523 | 0.0264 | 0.129847841 | 0.129847841 | 2.551446665 | 0.16002022 | 0.86203592 | 0.50545 | 1.481842591 | 1.481842591 | 5.642234807 | 0.811446458 | -0.090740131 | FALSE | TRUE | FALSE |
| GPRIN3 | 0.129784 | 0.85365789 | 1 | 0.129784022 | 0.129784022 | 2.555538363 | 1.45412886 | 0.539923905 | 0.514 | 1.45412886 | 1.45412886 | 28.68611621 | 0.089252077 | -1.049381666 | FALSE | FALSE | FALSE |
| ARFGAP1 | 0.1297554 | 0.85367054 | 1 | 0.129755397 | 0.129755397 | 2.569326026 | 0.15955593 | 0.86217808 | 1 | 1.458756411 | 1.458756411 | 3.163191155 | 0.813228271 | -0.089787532 | FALSE | TRUE | FALSE |
| SLC45A3 | 0.1296982 | 0.85368319 | 0.5136 | 0.12969818 | 0.12969818 | 2.55767822 | 0.0490391 | 0.886914606 | 1 | 0.049039095 | 0.049039095 | 0.968690357 | 2.644791438 | 0.42239143 | FALSE | TRUE | FALSE |
| CDC16 | 0.1296703 | 0.85369584 | 0.0171 | 0.129670255 | 0.129670255 | 2.564013266 | 0.15991993 | 0.862071487 | 1 | 1.489740951 | 1.489740951 | 5.414247081 | 0.810844878 | -0.091062222 | FALSE | TRUE | FALSE |
| TRIP10 | 0.129632 | 0.85370849 | 1 | 0.129631951 | 0.129631951 | 2.554571921 | 0.16480174 | 0.860936716 | 1 | 0.164801743 | 0.164801743 | 3.253379101 | 0.786593325 | -0.104249743 | FALSE | TRUE | FALSE |
| PRSS37 | 0.1295261 | 0.85372113 | 1 | 0.12952608 | 0.12952608 | 0.12952608 | 0.16440217 | 0.861020933 | 1 | 0.164402166 | 0.164402166 | 0.164402166 | 0.787861153 | -0.103550313 | FALSE | TRUE | FALSE |
| RTN4 | 0.1293224 | 0.85373378 | 1 | 0.129322442 | 0.129322442 | 2.538223722 | 0.1642842 | 0.861044977 | 1 | 0.164284203 | 0.164284203 | 3.231660824 | 0.787187326 | -0.103921907 | FALSE | TRUE | FALSE |
| PLEK2 | 0.1293183 | 0.85374642 | 1 | 0.129318328 | 0.129318328 | 2.565391253 | 0.16444862 | 0.861008908 | 1 | 0.16444862 | 0.16444862 | 0.16444862 | 0.786375271 | -0.104370152 | FALSE | TRUE | FALSE |
| MATN3 | 0.1292274 | 0.85375906 | 0.5062 | 0.129227353 | 0.129227353 | 0.28599164 | 0.02381465 | 0.892771063 | 1 | 0.095203969 | 0.38069255 | 0.38069255 | 5.426379413 | 0.734510156 | FALSE | TRUE | FALSE |
| NPHP4 | 0.129208 | 0.8537717 | 0.10615 | 0.004019984 | 0.12951098 | 2.065363757 | 7.5709E-07 | 0.901266703 | 0.6722 | 7.57089E-07 | 0.005255956 | 0.02545544 | 170664.2006 | 5.232142431 | FALSE | TRUE | FALSE |
| SEZ6L2 | 0.1291329 | 0.85378434 | 1 | 0.129132863 | 0.129132863 | 2.521365393 | 1.42887747 | 0.542897199 | 0.7562 | 1.428877474 | 13.57342857 | 48.30131895 | 0.090373644 | -1.043958208 | FALSE | FALSE | FALSE |
| DCLK1 | 0.1291262 | 0.85379697 | 0.0256 | 0.12912623 | 0.12912623 | 2.539398367 | 0.162604 | 0.861464478 | 1 | 0.65024684 | 0.65024684 | 5.497457442 | 0.79411473 | -0.100116748 | FALSE | TRUE | FALSE |
| RCN3 | 0.1290454 | 0.8538096 | 0.5017 | 1.179499516 | 1.179499516 | 2.561462424 | 1.45721516 | 0.539379168 | 0.75125 | 1.457215155 | 13.71808143 | 13.71808143 | 0.08855617 | -1.052781173 | FALSE | FALSE | FALSE |
| TMEM43 | 0.1290006 | 0.85382223 | 1 | 1.168348311 | 10.86060864 | 10.86060864 | 0.16346445 | 0.861237049 | 1 | 1.488522133 | 13.91182661 | 13.91182661 | 0.789165843 | -0.102831721 | FALSE | TRUE | FALSE |
| B4GALT7 | 0.1289886 | 0.85383486 | 1 | 1.184021705 | 11.15484676 | 11.15484676 | 1.51015382 | 0.534564119 | 0.25265 | 1.510153824 | 14.28305602 | 14.28305602 | 0.085414242 | -1.06846971 | FALSE | FALSE | FALSE |
| PITX2 | 0.1288908 | 0.85384749 | 1 | 0.128890783 | 0.128890783 | 2.553380058 | 0.16430925 | 0.861032956 | 1 | 0.164309247 | 0.164309247 | 0.164309247 | 0.784440226 | -0.105440144 | FALSE | TRUE | FALSE |
| CIAO1 | 0.1288475 | 0.85386011 | 1 | 0.128847518 | 0.128847518 | 2.556481719 | 0.16376952 | 0.86117708 | 1 | 0.163769524 | 0.163769524 | 0.163769524 | 0.786761267 | -0.104157029 | FALSE | TRUE | FALSE |
| CYSLTR2 | 0.1288314 | 0.85387273 | 1 | 0.128831448 | 0.128831448 | 0.128831448 | 0.16385721 | 0.861141076 | 1 | 0.163857212 | 0.163857212 | 0.163857212 | 0.78624216 | -0.104443672 | FALSE | TRUE | FALSE |
| INIP | 0.1287483 | 0.85388535 | 1 | 0.128748336 | 0.128748336 | 0.128748336 | 0.13319097 | 0.868169646 | 1 | 0.133190966 | 0.133190966 | 0.133190966 | 0.966644664 | -0.014733142 | FALSE | TRUE | FALSE |
| EXOC3L4 | 0.1287154 | 0.85389797 | 1 | 1.166088794 | 1.166088794 | 4.356545222 | 1.44003215 | 0.541820243 | 0.74895 | 1.440032147 | 13.45003457 | 28.4716211 | 0.089383721 | -1.048741572 | FALSE | FALSE | FALSE |
| HSP90B1 | 0.1286947 | 0.85391058 | 1 | 1.197742944 | 1.197742944 | 23.59377553 | 14.1661806 | 0.162182156 | 0.01455 | 1.480920119 | 14.16618064 | 29.22487583 | 0.009084642 | -2.041692176 | FALSE | FALSE | FALSE |
| ZNF471 | 0.1286456 | 0.8539232 | 0.9354 | 1.136998568 | 95.78998559 | 95.78998559 | 0.02549266 | 0.892299068 | 1 | 0.222301699 | 1.989673056 | 1.989673056 | 5.046377323 | 0.70297972 | FALSE | TRUE | FALSE |
| DCLRE1B | 0.1286319 | 0.85393581 | 1 | 1.188691208 | 1.188691208 | 2.544981667 | 1.46972683 | 0.538423034 | 0.75155 | 1.469726832 | 13.97102586 | 29.11613981 | 0.087520978 | -1.057887839 | FALSE | FALSE | FALSE |
| TCERG1 | 0.1286218 | 0.85394842 | 1 | 0.128621794 | 0.128621794 | 4.326262199 | 117.734639 | 0.024719201 | 0.5079 | 117.7346395 | 1168.017576 | 11870.59139 | 0.001092472 | -2.961589697 | FALSE | FALSE | FALSE |
| GRIN2B | 0.128561 | 0.85396102 | 1 | 11.58742618 | 114.3064445 | 223.8078579 | 6.06356627 | 0.267269369 | 0.0087 | 0.180021741 | 0.180021741 | 3.491442035 | 0.021202216 | -1.673618753 | FALSE | FALSE | FALSE |
| NME9 | 0.128495 | 0.85397363 | 1 | 0.128494996 | 0.128494996 | 0.128494996 | 12.9073283 | 0.169858528 | 0.2526 | 1.388136641 | 12.90732829 | 12.90732829 | 0.009955197 | -2.00195014 | FALSE | FALSE | FALSE |
| COX18 | 0.1284857 | 0.85398623 | 1 | 0.128485701 | 0.128485701 | 0.128485701 | 0.15633865 | 0.862872999 | 1 | 1.426387163 | 5.705016805 | 5.705016805 | 0.82184221 | -0.085211557 | FALSE | TRUE | FALSE |
| GSAP | 0.1284725 | 0.85399883 | 0.51045 | 0.12847252 | 0.12847252 | 2.533794755 | 0.02484339 | 0.892486825 | 1 | 0.024843392 | 0.024843392 | 0.490791181 | 5.171295439 | 0.71359935 | FALSE | TRUE | FALSE |
| ARHGAP12 | 0.1284547 | 0.85401143 | 1 | 0.128454668 | 0.128454668 | 2.529869655 | 2.52997325 | 0.445042479 | 0.5107 | 2.529973249 | 10.12265245 | 21.371894 | 0.050773133 | -1.294366038 | FALSE | FALSE | FALSE |
| POMT2 | 0.1283886 | 0.85402403 | 0.2695 | 0.056553491 | 0.512961695 | 1.114558881 | 0.01406682 | 0.895418823 | 0.87565 | 0.056223649 | 0.224776306 | 0.475936584 | 9.127048625 | 0.960330364 | FALSE | TRUE | FALSE |
| CS | 0.1282504 | 0.85403662 | 1 | 108691531.9 | 1.65864E+11 | 1.98977E+12 | 0.0603339 | 0.884674075 | 1 | 2.15322E+25 | 2.68646E+32 | 7.02049E+34 | 2.125677956 | 0.327497469 | FALSE | TRUE | FALSE |
| FSTL4 | 0.1282398 | 0.85404921 | 0.75815 | 0.512827626 | 2.051296463 | 10.06383461 | 23.6309386 | 0.11547748 | 0.01375 | 2.533940959 | 23.63093863 | 49.83908851 | 0.005426777 | -2.265458018 | FALSE | FALSE | FALSE |
| B4GALNT4 | 0.1282307 | 0.8540618 | 0.03005 | 0.12823072 | 0.12823072 | 2.504838034 | 0.16312448 | 0.861320924 | 0.02795 | 0.163124483 | 0.163124483 | 3.195395331 | 0.786091198 | -0.104527067 | FALSE | TRUE | FALSE |
| OCIAD2 | 0.1282242 | 0.85407439 | 0.00425 | 0.128224172 | 0.128224172 | 0.128224172 | 1.28728336 | 0.556499182 | 0.0036 | 1.287283361 | 1.287283361 | 1.287283361 | 0.09960835 | -1.001704252 | FALSE | FALSE | FALSE |
| GZF1 | 0.1281085 | 0.85408698 | 1 | 0.128108542 | 0.128108542 | 2.518739263 | 0.643837 | 0.693027384 | 0.0201 | 0.643836997 | 0.643836997 | 12.6842978 | 0.198976671 | -0.70119784 | FALSE | TRUE | FALSE |
| SLC25A22 | 0.1280076 | 0.85409956 | 1 | 0.128007587 | 0.128007587 | 2.537486521 | 0.16323249 | 0.861273008 | 1 | 0.163232492 | 0.163232492 | 0.163232492 | 0.784204082 | -0.105570901 | FALSE | TRUE | FALSE |
| CHST8 | 0.1280071 | 0.85411214 | 1 | 0.128007104 | 0.128007104 | 2.521357889 | 0.16317812 | 0.86129697 | 1 | 0.163178119 | 0.163178119 | 3.219982588 | 0.784462433 | -0.10542785 | FALSE | TRUE | FALSE |
| AP3B1 | 0.1280007 | 0.85412472 | 1 | 0.128000711 | 0.128000711 | 2.512424051 | 0.16285604 | 0.861404698 | 1 | 0.162856043 | 0.162856043 | 3.20372701 | 0.785974589 | -0.104591495 | FALSE | TRUE | FALSE |
| MAP4K3 | 0.1279849 | 0.8541373 | 1 | 0.127984881 | 0.127984881 | 2.519896837 | 0.15319105 | 0.863746796 | 1 | 1.416534661 | 13.44350084 | 27.94237882 | 0.835459258 | -0.078074724 | FALSE | TRUE | FALSE |
| MAD1L1 | 0.1279469 | 0.85414988 | 0.75795 | 0.127946872 | 1.152873166 | 2.510885947 | 0.02514921 | 0.892411794 | 1 | 0.025149207 | 0.025149207 | 0.494655114 | 5.08751128 | 0.706505385 | FALSE | TRUE | FALSE |
| OR51F1 | 0.1278273 | 0.85416245 | 1 | 0.127827306 | 0.127827306 | 0.127827306 | 0.16229656 | 0.861536151 | 1 | 0.162296564 | 0.162296564 | 0.162296564 | 0.787615601 | -0.10368569 | FALSE | TRUE | FALSE |
| SMARCA5 | 0.1277754 | 0.85417502 | 1 | 0.127775411 | 0.127775411 | 2.506720565 | 0.16311169 | 0.861332898 | 1 | 0.163111695 | 0.163111695 | 3.207310635 | 0.783361431 | -0.106037815 | FALSE | TRUE | FALSE |
| PITX1 | 0.127769 | 0.85418759 | 0.75615 | 1.208040834 | 11.72250885 | 11.72250885 | 0.68706809 | 0.672218714 | 0.5044 | 0.687068086 | 0.687068086 | 3.40506446 | 0.185962718 | -0.730574115 | FALSE | TRUE | FALSE |
| CD300LG | 0.1277679 | 0.85420016 | 1 | 0.127767906 | 0.127767906 | 0.127767906 | 0.15226594 | 0.864001085 | 1 | 0.152265938 | 0.152265938 | 0.152265938 | 0.839110229 | -0.076180985 | FALSE | TRUE | FALSE |
| TACC1 | 0.1277651 | 0.85421272 | 1 | 0.127765058 | 0.127765058 | 2.520462542 | 5.40389161 | 0.29066127 | 0.00615 | 0.159752963 | 0.159752963 | 3.156668986 | 0.023643157 | -1.626294531 | FALSE | FALSE | FALSE |
| PPP1R14B | 0.127757 | 0.85422529 | 0.00345 | 0.127757042 | 0.127757042 | 0.127757042 | 0.13281667 | 0.868234936 | 0.00425 | 0.132816671 | 0.132816671 | 0.132816671 | 0.961905167 | -0.016867742 | FALSE | TRUE | FALSE |
| TNNI2 | 0.1275801 | 0.85423785 | 1 | 0.510024587 | 0.510024587 | 0.510024587 | 0.64379327 | 0.69314863 | 0.75585 | 0.643793272 | 2.574489665 | 2.574489665 | 0.198169408 | -0.702963387 | FALSE | TRUE | FALSE |
| BICD1 | 0.1275781 | 0.85425041 | 1 | 0.127578108 | 0.127578108 | 2.501119856 | 1.4458631 | 0.54114476 | 0.5166 | 1.445863099 | 1.445863099 | 28.4130768 | 0.088236645 | -1.054351015 | FALSE | FALSE | FALSE |
| SETDB2 | 0.1275637 | 0.85426296 | 1 | 1484472492 | 2.46813E+12 | 3.82778E+14 | 0.0784182 | 0.880736089 | 1 | 9.84111E+21 | 7.02079E+27 | 1.72074E+30 | 1.626710253 | 0.211310204 | FALSE | TRUE | FALSE |
| MRPL44 | 0.1274589 | 0.85427552 | 1 | 0.127458899 | 0.127458899 | 0.127458899 | 0.16188151 | 0.861583897 | 0.0084 | 0.161881509 | 0.161881509 | 0.161881509 | 0.787359222 | -0.103827081 | FALSE | TRUE | FALSE |
| ANKRD28 | 0.1274436 | 0.85428807 | 0.51085 | 1.191647587 | 1.191647587 | 23.44964499 | 0.16776005 | 0.860235227 | 1 | 0.167760048 | 0.167760048 | 3.307630378 | 0.759677724 | -0.119370608 | FALSE | TRUE | FALSE |
| WBP4 | 0.1274134 | 0.85430062 | 0.75155 | 0.509555054 | 2.038335126 | 2.038335126 | 0.16569989 | 0.860804188 | 1 | 0.165699888 | 0.165699888 | 0.165699888 | 0.768940941 | -0.114107015 | FALSE | TRUE | FALSE |
| CDCA2 | 0.1272709 | 0.85431317 | 0.75875 | 0.127270889 | 1.121747019 | 4.29480271 | 1.79208784 | 0.508903883 | 0.14565 | 1.792087838 | 16.37009931 | 35.26632579 | 0.07101822 | -1.148630216 | FALSE | FALSE | FALSE |
| EPS15L1 | 0.1272645 | 0.85432572 | 0.02565 | 0.127264512 | 0.127264512 | 2.497990765 | 0.16239009 | 0.861500324 | 1 | 0.16239009 | 0.16239009 | 3.194579876 | 0.783696296 | -0.105852206 | FALSE | TRUE | FALSE |
| SLC22A13 | 0.1271397 | 0.85433826 | 0.2623 | 0.031799685 | 0.127139706 | 0.62797568 | 0.46951512 | 0.767190931 | 0.6271 | 0.469515124 | 17.21084687 | 17.21084687 | 0.270789373 | -0.567368384 | FALSE | TRUE | FALSE |
| CLTC | 0.127123 | 0.85435081 | 1 | 1.175954442 | 1.175954442 | 22.78866637 | 0.16198189 | 0.861560028 | 1 | 1.502980226 | 1.502980226 | 29.23514046 | 0.784797677 | -0.105242291 | FALSE | TRUE | FALSE |
| FAM83G | 0.1270561 | 0.85436335 | 1 | 1.158495176 | 1.158495176 | 22.64356573 | 0.16150372 | 0.861679294 | 1 | 1.479472278 | 1.479472278 | 28.99641726 | 0.786706645 | -0.104187182 | FALSE | TRUE | FALSE |
| ZNF808 | 0.1270529 | 0.85437589 | 0.75895 | 0.127052922 | 1.136642368 | 2.496251608 | 15.8403412 | 0.154672065 | 0.1996 | 15.84034121 | 150.050128 | 1456.135108 | 0.008020845 | -2.095779874 | FALSE | FALSE | FALSE |
| MED24 | 0.1270367 | 0.85438842 | 0.5096 | 0.127036702 | 0.127036702 | 2.483916722 | 0.02450794 | 0.892614157 | 1 | 0.024507937 | 0.024507937 | 0.480488484 | 5.183492261 | 0.714622454 | FALSE | TRUE | FALSE |
| LENG1 | 0.1268628 | 0.85440096 | 1 | 12696015531 | 2.2911E+13 | 3.4759E+15 | 0.02512034 | 0.892426807 | 1 | 628033018.5 | 4.5044E+12 | 6.70748E+14 | 5.05020291 | 0.703308828 | FALSE | TRUE | FALSE |
| KLRG1 | 0.1268395 | 0.85441349 | 0.0035 | 0.126839536 | 0.126839536 | 0.126839536 | 0.14383371 | 0.865913641 | 1 | 0.143833711 | 0.143833711 | 0.143833711 | 0.881848457 | -0.054606041 | FALSE | TRUE | FALSE |
| LYSMD3 | 0.126706 | 0.85442602 | 1 | 0.126706043 | 0.126706043 | 0.126706043 | 0.16078989 | 0.86186971 | 1 | 0.16078989 | 0.16078989 | 0.16078989 | 0.788022449 | -0.10346141 | FALSE | TRUE | FALSE |

| | | | | | | | | | | | | | | | | | |
|---|---|---|---|---|---|---|---|---|---|---|---|---|---|---|---|---|---|
| ZNF804B | 0.1266784 | 0.85443855 | 0.75815 | 0.126678448 | 1.116590268 | 4.265503481 | 0.2063885 | 0.850537634 | 0.26675 | 0.206388499 | 1.839668743 | 6.957558652 | 0.613786373 | -0.211982758 | FALSE | TRUE | FALSE |
| MNT | 0.1266502 | 0.85445108 | 0.0157 | 0.126650199 | 0.126650199 | 2.501298035 | 0.16184069 | 0.861607757 | 1 | 0.16184069 | 0.16184069 | 3.201112208 | 0.78256092 | -0.106481844 | FALSE | TRUE | FALSE |
| ZSWIM5 | 0.1266056 | 0.85446361 | 1 | 0.126605593 | 0.126605593 | 2.469867343 | 1.44237871 | 0.541685287 | 0.51745 | 1.44237871 | 1.44237871 | 28.22179174 | 0.087775556 | -1.056626412 | FALSE | FALSE | FALSE |
| EXOSC2 | 0.126587 | 0.85447613 | 0.5084 | 1.160549405 | 1.160549405 | 1.160549405 | 0.16616841 | 0.860707664 | 0.0063 | 0.166168406 | 0.166168406 | 0.166168406 | 0.761799665 | -0.118159223 | FALSE | TRUE | FALSE |
| UBE2O | 0.1265798 | 0.85448865 | 1 | 0.126579841 | 0.126579841 | 4.263553512 | 0.16170454 | 0.861631611 | 0.0357 | 0.161704543 | 0.161704543 | 3.152079275 | 0.782784695 | -0.106357674 | FALSE | TRUE | FALSE |
| GSTA1 | 0.1265278 | 0.85450117 | 1 | 0.126527776 | 0.126527776 | 0.126527776 | 0.16031987 | 0.861952863 | 1 | 0.160319868 | 0.160319868 | 0.160319868 | 0.789220813 | -0.10280147 | FALSE | TRUE | FALSE |
| USH1G | 0.1265181 | 0.85451369 | 0.0203 | 0.126518111 | 0.126518111 | 2.493264919 | 0.1612942 | 0.861750758 | 0.0164 | 0.161294197 | 0.161294197 | 3.184198422 | 0.784393443 | -0.105466046 | FALSE | TRUE | FALSE |
| GHITM | 0.1264338 | 0.8545262 | 0.4998 | 0.126433824 | 1.142906202 | 1.142906202 | 0.16544591 | 0.860840355 | 1 | 0.165445907 | 0.165445907 | 0.165445907 | 0.764200375 | -0.116792754 | FALSE | TRUE | FALSE |
| PPP1R13B | 0.1263775 | 0.85453871 | 1 | 0.126377495 | 0.126377495 | 2.462741959 | 0.16134349 | 0.861738852 | 1 | 0.161343486 | 0.161343486 | 3.153846471 | 0.783282287 | -0.106081694 | FALSE | TRUE | FALSE |
| ATE1 | 0.126363 | 0.85455122 | 0.75075 | 1.164417683 | 4.656621677 | 4.656621677 | 0.16797668 | 0.860137931 | 1 | 0.167976679 | 0.167976679 | 3.332197758 | 0.752265061 | -0.123629109 | FALSE | TRUE | FALSE |
| BHMT2 | 0.1262906 | 0.85456373 | 0.4987 | 0.126290558 | 0.505133496 | 0.505133496 | 0.16039486 | 0.861929114 | 0.50955 | 0.64154724 | 0.64154724 | 0.64154724 | 0.787372858 | -0.10381956 | FALSE | TRUE | FALSE |
| FSD2 | 0.1262658 | 0.85457624 | 0.01975 | 0.126265789 | 0.126265789 | 2.49094318 | 0.02362247 | 0.892815819 | 1 | 0.206417413 | 0.206417413 | 0.799070014 | 5.34515662 | 0.727960435 | FALSE | TRUE | FALSE |
| GPR153 | 0.1262195 | 0.85458874 | 1 | 51564584764 | 7.3044E+14 | 7.4046E+16 | 0.25349057 | 0.838443789 | 0.99655 | 2129.656211 | 226164.7968 | 2413203.679 | 0.497925728 | -0.302835433 | FALSE | TRUE | FALSE |
| C12orf57 | 0.1261743 | 0.85460125 | 0.00455 | 0.12617426 | 0.12617426 | 0.12617426 | 0.16046475 | 0.861917237 | 1 | 0.160464754 | 0.160464754 | 0.160464754 | 0.786305133 | -0.104408889 | FALSE | TRUE | FALSE |
| HIPK4 | 0.1261583 | 0.85461375 | 0.023 | 0.126158282 | 0.126158282 | 2.480203262 | 0.16086775 | 0.861857824 | 1 | 0.160867747 | 0.160867747 | 3.1690555 | 0.784236018 | -0.105553215 | FALSE | TRUE | FALSE |
| UGT1A9 | 0.1261148 | 0.85462624 | 1 | 1.158289733 | 10.91851931 | 22.90784725 | 14.0189406 | 0.163510088 | 0.2569 | 1.481493979 | 14.01894056 | 14.01894056 | 0.008996032 | -2.045949003 | FALSE | FALSE | FALSE |
| DGCR2 | 0.1260757 | 0.85463874 | 0.3802 | 0.055736974 | 0.503838018 | 0.621601208 | 0.06304535 | 0.883998513 | 0.50465 | 0.063045345 | 0.063045345 | 0.136861205 | 1.999762803 | 0.300978486 | FALSE | TRUE | FALSE |
| LGI1 | 0.1260218 | 0.85465123 | 1 | 4.722276726 | 45.42198102 | 45.42198102 | 0.16799834 | 0.86012576 | 0.5052 | 1.578352592 | 1.578352592 | 3.329712472 | 0.750137092 | -0.12485936 | FALSE | TRUE | FALSE |
| MFSD2B | 0.1259267 | 0.85466373 | 0.50635 | 0.125926666 | 1.129237762 | 1.129237762 | 0.22852369 | 0.845246822 | 0.01225 | 0.057159343 | 0.228523688 | 0.283424196 | 0.551044259 | -0.258813518 | FALSE | TRUE | FALSE |
| WDR48 | 0.1258593 | 0.85467622 | 1 | 0.125859334 | 0.125859334 | 2.486815986 | 0.16075673 | 0.861881595 | 0.01625 | 0.160756726 | 0.160756726 | 3.180953916 | 0.782917997 | -0.106283724 | FALSE | TRUE | FALSE |
| IGSF8 | 0.1258188 | 0.85468871 | 0.88265 | 1.11897048 | 10.20759729 | 22.02995536 | 0.02394936 | 0.892733741 | 1 | 0.023949364 | 0.023949364 | 0.472394132 | 5.253533517 | 0.720451507 | FALSE | TRUE | FALSE |
| ADRB3 | 0.1256932 | 0.85470119 | 1 | 1.142315983 | 1.142315983 | 1.142315983 | 0.15986443 | 0.862107034 | 0.50735 | 1.460194357 | 1.460194357 | 1.460194357 | 0.786248771 | -0.10444002 | FALSE | TRUE | FALSE |
| DNALI1 | 0.1255425 | 0.85471368 | 1 | 0.125542465 | 0.125542465 | 0.125542465 | 1.38822118 | 0.546234946 | 0.7497 | 1.388221183 | 13.13986162 | 13.13986162 | 0.090434051 | -1.043668016 | FALSE | FALSE | FALSE |
| TMEM168 | 0.1254145 | 0.85472616 | 0.50445 | 1.134723365 | 1.134723365 | 2.47646394 | 1.4504645 | 0.540602933 | 0.50795 | 1.450464496 | 1.450464496 | 5.398641188 | 0.086465082 | -1.063159243 | FALSE | FALSE | FALSE |
| OR5K4 | 0.1253773 | 0.85473864 | 1 | 1.127412487 | 1.127412487 | 1.127412487 | 1.43966513 | 0.542224553 | 0.5072 | 1.439665135 | 1.439665135 | 1.439665135 | 0.087087805 | -1.060042657 | FALSE | FALSE | FALSE |
| SLC46A3 | 0.1253348 | 0.85475112 | 0.8147 | 9.964486894 | 92.24178657 | 874.7934787 | 1.79941179 | 0.507521993 | 0.8158 | 16.43424788 | 615.3580526 | 5902.008625 | 0.069653206 | -1.157058887 | FALSE | FALSE | FALSE |
| TADA2A | 0.1253063 | 0.8547636 | 1 | 0.125306297 | 0.125306297 | 2.484187786 | 0.15935027 | 0.862225403 | 0.0121 | 0.159350269 | 0.159350269 | 3.162432943 | 0.78635761 | -0.104379906 | FALSE | TRUE | FALSE |
| LRP3 | 0.1251546 | 0.85477607 | 0.5147 | 0.125154619 | 0.125154619 | 2.451298942 | 0.04769848 | 0.887209622 | 0.0229 | 0.047698482 | 0.047698482 | 0.936558599 | 2.623870063 | 0.418942325 | FALSE | TRUE | FALSE |
| PLTP | 0.1251331 | 0.85478855 | 1 | 0.125133141 | 0.125133141 | 2.472567851 | 0.15903132 | 0.86229633 | 1 | 0.159031321 | 0.159031321 | 3.146925372 | 0.786845887 | -0.104110321 | FALSE | TRUE | FALSE |
| BAG3 | 0.1251199 | 0.85480102 | 0.01825 | 0.125119926 | 0.125119926 | 2.468118257 | 0.15830628 | 0.862473345 | 1 | 0.63295501 | 0.63295501 | 3.127901039 | 0.790366155 | -0.102171665 | FALSE | TRUE | FALSE |
| RASSF4 | 0.1251147 | 0.85481349 | 1 | 0.125114708 | 0.125114708 | 2.48223683 | 0.15893002 | 0.862308145 | 1 | 0.158930023 | 0.158930023 | 0.158930023 | 0.787231421 | -0.10389758 | FALSE | TRUE | FALSE |
| KRT3 | 0.1250886 | 0.85482596 | 0.012 | 0.03130988 | 0.12508857 | 0.273147599 | 0.00701584 | 0.897538611 | 1 | 0.007015843 | 0.007015843 | 0.138835883 | 17.82944231 | 1.251137759 | FALSE | TRUE | FALSE |
| PDP2 | 0.1250657 | 0.85483842 | 0.93805 | 10.597225 | 993.8409958 | 993.8409958 | 0.17979351 | 0.857055784 | 1 | 0.179793512 | 0.179793512 | 3.562439416 | 0.695607368 | -0.157635826 | FALSE | TRUE | FALSE |
| TTLL1 | 0.1250166 | 0.85485088 | 0.01295 | 0.125016645 | 0.125016645 | 2.474252703 | 0.1573697 | 0.862708697 | 1 | 0.629431114 | 0.629431114 | 3.118472462 | 0.794413718 | -0.099953265 | FALSE | TRUE | FALSE |
| NUP93 | 0.1249197 | 0.85486334 | 1 | 0.12491969 | 0.12491969 | 2.459003918 | 5.52479175 | 0.285845 | 0.38265 | 2.415299691 | 22.10348681 | 27.24152038 | 0.022610751 | -1.645685014 | FALSE | FALSE | FALSE |
| PKD2L1 | 0.1249147 | 0.8548758 | 0.87765 | 1.997699413 | 273.2844531 | 1093.837209 | 0.02138375 | 0.893431497 | 0.8021 | 0.341742581 | 11.83936298 | 100.1223776 | 5.841573666 | 0.766529858 | FALSE | TRUE | FALSE |
| ZBTB21 | 0.1248905 | 0.85488826 | 0.0272 | 0.124890488 | 0.124890488 | 2.445543761 | 0.15925577 | 0.862260875 | 1 | 0.159255765 | 0.159255765 | 3.126327177 | 0.784213294 | -0.1055658 | FALSE | TRUE | FALSE |
| MAP3K1 | 0.1248645 | 0.85490071 | 1 | 0.124864454 | 0.124864454 | 4.199736641 | 0.15954366 | 0.862189913 | 1 | 0.159543661 | 0.159543661 | 3.123021689 | 0.782635009 | -0.106440729 | FALSE | TRUE | FALSE |
| IL1A | 0.1247855 | 0.85491317 | 1 | 0.124785548 | 0.124785548 | 0.124785548 | 0.13230392 | 0.868387045 | 1 | 0.132303916 | 0.132303916 | 0.132303916 | 0.943173498 | -0.025408411 | FALSE | TRUE | FALSE |
| UNC50 | 0.1247148 | 0.85492562 | 0.00765 | 0.124714797 | 0.124714797 | 0.124714797 | 0.15394686 | 0.863584501 | 1 | 1.402048922 | 1.402048922 | 1.402048922 | 0.81011586 | -0.091452866 | FALSE | TRUE | FALSE |
| SLC30A7 | 0.1246755 | 0.85493807 | 1 | 1.137322302 | 1.137322302 | 1.137322302 | 0.1637841 | 0.861165081 | 1 | 0.163784104 | 0.163784104 | 0.163784104 | 0.761218483 | -0.118490675 | FALSE | TRUE | FALSE |
| OGFR | 0.1246544 | 0.85495051 | 0.75875 | 1.134027619 | 10.58861395 | 22.36177872 | 1.4978234 | 0.536087299 | 0.5109 | 1.497823405 | 1.497823405 | 5.539337013 | 0.083223685 | -1.07975306 | FALSE | FALSE | FALSE |
| TMEM72 | 0.1245698 | 0.85496296 | 1 | 0.124569802 | 0.124569802 | 0.124569802 | 1.38588479 | 0.546765019 | 0.5035 | 1.385884792 | 1.385884792 | 1.385884792 | 0.089884673 | -1.046314356 | FALSE | FALSE | FALSE |
| ZDHHC4 | 0.1244782 | 0.8549754 | 1 | 0.124478152 | 0.124478152 | 0.124478152 | 0.60838872 | 0.708258243 | 0.75635 | 1.390500151 | 5.561329921 | 5.561329921 | 0.204602989 | -0.689088026 | FALSE | TRUE | FALSE |
| TAF8 | 0.124459 | 0.85498784 | 1 | 0.497813898 | 0.497813898 | 0.497813898 | 0.16005037 | 0.862024061 | 1 | 0.160050368 | 0.160050368 | 0.160050368 | 0.777623924 | -0.109230386 | FALSE | TRUE | FALSE |
| CFHR4 | 0.1244052 | 0.85500028 | 0.50535 | 0.124405189 | 0.124405189 | 0.124405189 | 0.20286433 | 0.851472269 | 0.00865 | 0.202864329 | 0.202864329 | 0.202864329 | 0.613243294 | -0.212367192 | FALSE | TRUE | FALSE |
| BST1 | 0.1243852 | 0.85501272 | 0.00795 | 0.124385216 | 0.124385216 | 0.124385216 | 1.38621621 | 0.54663263 | 0.50405 | 1.386216212 | 1.386216212 | 1.386216212 | 0.089730026 | -1.047062208 | FALSE | FALSE | FALSE |
| GRAP2 | 0.1243704 | 0.85502515 | 0.0107 | 0.124370406 | 0.124370406 | 2.468934215 | 0.15879075 | 0.862355384 | 1 | 0.158790752 | 0.158790752 | 0.158790752 | 0.783234569 | -0.106108153 | FALSE | TRUE | FALSE |
| AMFR | 0.1243544 | 0.85503758 | 0.01935 | 0.124354398 | 0.124354398 | 2.452168393 | 0.15829237 | 0.86248513 | 1 | 0.158292369 | 0.158292369 | 3.126672098 | 0.785599448 | -0.104798831 | FALSE | TRUE | FALSE |
| KCNA4 | 0.1242079 | 0.85505002 | 0.7571 | 0.124207907 | 0.496457669 | 2.450691884 | 3.65793456 | 0.373757908 | 0.0133 | 0.739904504 | 2.958468296 | 3.657934557 | 0.033955749 | -1.469086687 | FALSE | FALSE | FALSE |
| SPPL2C | 0.124116 | 0.85506244 | 0.0211 | 0.12411599 | 0.12411599 | 2.440915121 | 0.15830701 | 0.862461557 | 1 | 0.158307012 | 0.158307012 | 3.119581172 | 0.784020796 | -0.105672417 | FALSE | TRUE | FALSE |
| FAAH | 0.1240772 | 0.85507487 | 0.2586 | 0.124077199 | 0.496236388 | 2.453502723 | 25.1944736 | 0.110049456 | 0.27315 | 6.297739428 | 100.8171454 | 403.5264281 | 0.004924778 | -2.307613307 | FALSE | FALSE | FALSE |
| SCYL2 | 0.1240662 | 0.8550873 | 1 | 0.124066236 | 0.124066236 | 2.444400798 | 0.15866319 | 0.862402591 | 0.0198 | 0.158663192 | 0.158663192 | 3.131642221 | 0.781947185 | -0.106822579 | FALSE | TRUE | FALSE |
| NLE1 | 0.1240215 | 0.85509972 | 1 | 0.124021481 | 0.124021481 | 0.124021481 | 0.14891566 | 0.864849795 | 1 | 1.37520204 | 13.03420391 | 13.03420391 | 0.832830353 | -0.079443455 | FALSE | TRUE | FALSE |
| DIP2C | 0.123966 | 0.85511214 | 0.05675 | 0.123965955 | 2.378036285 | 4.161270564 | 5.32887824 | 0.292753716 | 0.00165 | 0.158507044 | 3.055634412 | 5.328878238 | 0.023263049 | -1.633333366 | FALSE | FALSE | FALSE |
| TRDN | 0.1238931 | 0.85512456 | 0.96855 | 9.94814442 | 370.1212657 | 3527.553256 | 0.86046321 | 0.605266977 | 0.688 | 1.927533471 | 70.83832765 | 70.83832765 | 0.143984181 | -0.84168522 | FALSE | TRUE | FALSE |
| ANLN | 0.1238183 | 0.85513698 | 1 | 0.123818285 | 0.123818285 | 2.42442284 | 12.5211692 | 0.174895233 | 0.51135 | 12.52116916 | 121.2297945 | 245.7900546 | 0.009888716 | -2.004860099 | FALSE | FALSE | FALSE |
| SYNGR4 | 0.1237729 | 0.85514939 | 0.00645 | 0.123772882 | 0.123772882 | 0.123772882 | 0.15727922 | 0.862743933 | 1 | 0.15727922 | 0.15727922 | 0.15727922 | 0.786962716 | -0.104045843 | FALSE | TRUE | FALSE |
| RAVER2 | 0.1237725 | 0.8551618 | 0.5109 | 1.12486237 | 1.12486237 | 4.1852877 | 0.15774076 | 0.862591116 | 1 | 1.440799708 | 1.440799708 | 3.126517689 | 0.784657583 | -0.105319824 | FALSE | TRUE | FALSE |
| ZNF561 | 0.1237614 | 0.85517422 | 0.50955 | 1.123224307 | 1.123224307 | 2.457410467 | 12.6303877 | 0.174224107 | 0.13535 | 12.63038773 | 121.0845926 | 121.0845926 | 0.009798702 | -2.00883145 | FALSE | FALSE | FALSE |
| CHST10 | 0.1236497 | 0.85518662 | 1 | 0.123649699 | 0.123649699 | 2.453115868 | 0.15818592 | 0.862508695 | 1 | 0.158185922 | 0.158185922 | 0.158185922 | 0.781673222 | -0.106974766 | FALSE | TRUE | FALSE |
| RTP4 | 0.1236423 | 0.85519903 | 1 | 0.123642305 | 0.123642305 | 0.123642305 | 0.1412216 | 0.866573417 | 0.00605 | 0.141221604 | 0.141221604 | 0.141221604 | 0.875519763 | -0.057734046 | FALSE | TRUE | FALSE |
| SOX5 | 0.123563 | 0.85521144 | 0.51035 | 0.12356304 | 0.12356304 | 2.435080014 | 0.08923931 | 0.878304383 | 0.5104 | 0.089239312 | 0.089239312 | 0.755155879 | 1.384625638 | 0.141332369 | FALSE | TRUE | FALSE |
| BAIAP2L2 | 0.1233863 | 0.85522384 | 0.8768 | 0.493597417 | 1.975092368 | 2.437288445 | 0.38429991 | 0.797841997 | 0.87815 | 3.485878224 | 32.43213355 | 32.43213355 | 0.321067682 | -0.493403408 | FALSE | TRUE | FALSE |
| MTA1 | 0.1233571 | 0.85523624 | 0.02445 | 0.123357075 | 0.123357075 | 2.417118811 | 0.15769316 | 0.862602883 | 0.02405 | 0.157693164 | 0.157693164 | 3.097462336 | 0.782260129 | -0.106648805 | FALSE | TRUE | FALSE |
| TRIM24 | 0.1233063 | 0.85524864 | 1 | 11.10968285 | 1107.740389 | 1107.740389 | 0.1630003 | 0.861380773 | 1 | 14.75955974 | 145.9346618 | 290.2235994 | 0.756479183 | -0.121203019 | FALSE | TRUE | FALSE |
| RAX | 0.1232863 | 0.85526104 | 1 | 0.49292221 | 0.49292221 | 0.49292221 | 0.15473 | 0.863375298 | 1 | 1.449156271 | 1.449156271 | 1.449156271 | 0.796783708 | -0.098659554 | FALSE | TRUE | FALSE |
| DBR1 | 0.1232418 | 0.85527343 | 1 | 2.439203642 | 42.15800912 | 42.15800912 | 0.66870641 | 0.680959108 | 0.50235 | 0.668706411 | 0.668706411 | 3.313271701 | 0.18429886 | -0.734477351 | FALSE | TRUE | FALSE |
| TACSTD2 | 0.1231925 | 0.85528583 | 1 | 1.145876131 | 10.9390969 | 10.9390969 | 0.16792689 | 0.860150101 | 1 | 0.167926893 | 0.167926893 | 3.325258007 | 0.733607845 | -0.134536033 | FALSE | TRUE | FALSE |
| MLF1 | 0.1231816 | 0.85529822 | 1 | 0.123181625 | 0.123181625 | 0.123181625 | 0.15674473 | 0.862790891 | 1 | 0.156744729 | 0.156744729 | 0.156744729 | 0.785874114 | -0.104647016 | FALSE | TRUE | FALSE |
| RTN4IP1 | 0.1230465 | 0.85531061 | 1 | 0.123046494 | 0.123046494 | 2.442467461 | 0.15668294 | 0.862802626 | 1 | 0.156682936 | 0.156682936 | 0.156682936 | 0.785321596 | -0.104952459 | FALSE | TRUE | FALSE |
| KBTBD8 | 0.1229358 | 0.85532299 | 1 | 0.122935779 | 0.122935779 | 2.432423312 | 0.122955 | 0.870619112 | 1 | 1024.338669 | 108327.3911 | 1153447.82 | 0.999843654 | -6.79057E-05 | FALSE | TRUE | FALSE |
| QRICH2 | 0.1229298 | 0.85533538 | 0.9962 | 6202.47583 | 5389413.478 | 53641721.16 | 0.38118422 | 0.798849079 | 0.91595 | 28.50786465 | 2343.72006 | 21989.31905 | 0.322494438 | -0.491477771 | FALSE | TRUE | FALSE |

| | | | | | | | | | | | | | | | | | |
|---|---|---|---|---|---|---|---|---|---|---|---|---|---|---|---|---|---|
| HTR3E | 0.1228435 | 0.85534776 | 0.50875 | 0.122843464 | 0.122843464 | 0.276574878 | 0.0235293 | 0.89285309 | 0.0107 | 0.023529296 | 0.023529296 | 0.466577233 | 5.22087289 | 0.71774312 | FALSE | TRUE | FALSE |
| OTX2 | 0.1228074 | 0.85536015 | 1 | 0.122807414 | 0.122807414 | 0.122807414 | 0.1574762 | 0.862673444 | 1 | 0.157476204 | 0.157476204 | 0.157476204 | 0.779847435 | -0.107990352 | FALSE | TRUE | FALSE |
| SCAF11 | 0.1227938 | 0.85537253 | 1 | 1.130269107 | 1.130269107 | 22.0623749 | 0.16193581 | 0.861571963 | 0.03235 | 0.161935806 | 0.161935806 | 5.449739949 | 0.758286821 | -0.120166492 | FALSE | TRUE | FALSE |
| PPP3CC | 0.1227295 | 0.8553849 | 1 | 0.122729484 | 0.122729484 | 2.431317534 | 0.15514541 | 0.863235478 | 1 | 0.620469711 | 0.620469711 | 3.076994227 | 0.791061024 | -0.101790013 | FALSE | TRUE | FALSE |
| ACTL10 | 0.1227227 | 0.85539728 | 0.0093 | 0.122722723 | 0.122722723 | 0.122722723 | 1.36105509 | 0.549789611 | 0.74685 | 1.361055093 | 12.89469941 | 12.89469941 | 0.090167344 | -1.044950721 | FALSE | FALSE | FALSE |
| TSPAN2 | 0.1227095 | 0.85540965 | 1 | 0.490689342 | 0.490689342 | 0.490689342 | 1.42282631 | 0.543567956 | 0.5052 | 1.422826312 | 1.422826312 | 1.422826312 | 0.086243465 | -1.064273803 | FALSE | FALSE | FALSE |
| SIGLEC11 | 0.1227049 | 0.85542202 | 0.98565 | 10.14683011 | 926.4871811 | 9174.770603 | 0.22820177 | 0.845365025 | 0.7524 | 0.228201766 | 2.075082763 | 4.50126703 | 0.537703708 | -0.269456968 | FALSE | TRUE | FALSE |
| CDH19 | 0.1226581 | 0.85543439 | 0.69035 | 0.271274335 | 9.844724363 | 9.844724363 | 1.11379548 | 0.57707337 | 0.1371 | 0.486652694 | 4.452643772 | 4.452643772 | 0.110126252 | -0.958109142 | FALSE | FALSE | FALSE |
| CXCL2 | 0.1226564 | 0.85544676 | 1 | 0.122656374 | 0.122656374 | 0.122656374 | 0.13264109 | 0.868278431 | 0.0019 | 0.132641092 | 0.132641092 | 0.132641092 | 0.924723796 | -0.033987967 | FALSE | TRUE | FALSE |
| TBR1 | 0.1226514 | 0.85545912 | 0.01945 | 0.12265139 | 0.12265139 | 2.414029548 | 0.1572908 | 0.862720445 | 1 | 0.157290796 | 0.157290796 | 3.101728193 | 0.779774745 | -0.108030835 | FALSE | TRUE | FALSE |
| AKAP10 | 0.1226198 | 0.85547149 | 0.0159 | 0.122619753 | 0.122619753 | 2.425272478 | 0.13468261 | 0.86783135 | 1 | 1.256679757 | 1.256679757 | 4.557132749 | 0.910434948 | -0.04075108 | FALSE | TRUE | FALSE |
| EXOC7 | 0.122579 | 0.85548385 | 1 | 0.122578984 | 0.122578984 | 2.411642018 | 0.15183747 | 0.864150917 | 0.5134 | 1.39119731 | 1.39119731 | 5.138560385 | 0.807303934 | -0.092962931 | FALSE | TRUE | FALSE |
| OR51B2 | 0.1225314 | 0.85549621 | 0.00705 | 0.122531442 | 0.122531442 | 0.122531442 | 0.15593442 | 0.863001834 | 1 | 0.155934421 | 0.155934421 | 0.155934421 | 0.785788287 | -0.104694449 | FALSE | TRUE | FALSE |
| KLHL7 | 0.1225124 | 0.85550856 | 0.01525 | 0.12251244 | 0.12251244 | 2.425880225 | 0.15489758 | 0.863258801 | 1 | 0.619479452 | 0.619479452 | 3.070810717 | 0.790925443 | -0.101864454 | FALSE | TRUE | FALSE |
| SLX4IP | 0.1224774 | 0.85552092 | 1 | 0.122477448 | 0.122477448 | 2.431133615 | 0.15597598 | 0.862978427 | 0.0089 | 0.155975985 | 0.155975985 | 0.155975985 | 0.785232731 | -0.105001606 | FALSE | TRUE | FALSE |
| PHIP | 0.1224709 | 0.85553327 | 1 | 10.81075421 | 1056.128009 | 2041.475909 | 5.94853227 | 0.271354903 | 0.01405 | 0.177566838 | 0.177566838 | 5.948532269 | 0.020588425 | -1.686376877 | FALSE | FALSE | FALSE |
| DUS1L | 0.1224444 | 0.85554562 | 1 | 0.122444436 | 0.122444436 | 2.416950227 | 0.14716957 | 0.865270392 | 0.75535 | 1.350051936 | 12.71092655 | 26.69071272 | 0.831995625 | -0.079878957 | FALSE | TRUE | FALSE |
| GPA33 | 0.1223602 | 0.85555797 | 1 | 0.122360247 | 0.122360247 | 2.42865178 | 0.15594713 | 0.862990132 | 1 | 0.155947126 | 0.155947126 | 0.155947126 | 0.784626493 | -0.105337032 | FALSE | TRUE | FALSE |
| HERC4 | 0.1223064 | 0.85557031 | 0.02465 | 0.122306391 | 0.122306391 | 2.404358019 | 0.15683508 | 0.862779154 | 0.0207 | 0.15683508 | 0.15683508 | 3.089477943 | 0.779840777 | -0.10799406 | FALSE | TRUE | FALSE |
| HLA-DQA2 | 0.1222969 | 0.85558266 | 1 | 4.56445E+16 | 1.48638E+21 | 2.97578E+23 | 0.15558868 | 0.863118754 | 1 | 6.20398E+16 | 2.06776E+21 | 4.18809E+23 | 0.78602689 | -0.104562597 | FALSE | TRUE | FALSE |
| ATF6B | 0.122268 | 0.855595 | 1 | 0.122268008 | 0.122268008 | 2.406650291 | 0.15620157 | 0.862919875 | 1 | 0.156201569 | 0.156201569 | 3.080436814 | 0.782757873 | -0.106372555 | FALSE | TRUE | FALSE |
| FBXO40 | 0.1222252 | 0.85560734 | 0.8809 | 0.488970565 | 4.291127634 | 5.292416171 | 1677.90669 | 0.001401836 | 0.0097 | 24.40550725 | 418.9994262 | 1677.906688 | 7.28439E-05 | -4.137607015 | FALSE | FALSE | FALSE |
| PFKFB2 | 0.1221688 | 0.85561968 | 0.50225 | 0.122168836 | 0.122168836 | 1.034519927 | 0.0440051 | 0.888086364 | 0.87595 | 0.703732323 | 6.36651665 | 6.36651665 | 2.776242915 | 0.443457463 | FALSE | TRUE | FALSE |
| CHPF | 0.1221545 | 0.85563202 | 1 | 0.122154537 | 0.122154537 | 2.385056791 | 1.29744898 | 0.555222818 | 0.5143 | 1.297448983 | 1.297448983 | 25.40508419 | 0.094149781 | -1.026180686 | FALSE | FALSE | FALSE |
| FN3K | 0.1221023 | 0.85564435 | 1 | 0.122102309 | 0.122102309 | 0.122102309 | 0.14953515 | 0.86466714 | 0.7549 | 1.366531829 | 5.465465808 | 5.465465808 | 0.816545862 | -0.088019418 | FALSE | TRUE | FALSE |
| GPR156 | 0.1220305 | 0.85565668 | 0.51135 | 0.122030455 | 0.487965365 | 2.403794452 | 0.02304214 | 0.892942446 | 0.5068 | 0.203472962 | 0.203472962 | 0.779532959 | 5.295968786 | 0.723945417 | FALSE | TRUE | FALSE |
| SYMPK | 0.1220243 | 0.85566901 | 1 | 10.81008691 | 1059.855572 | 2055.142754 | 1.60321499 | 0.525520803 | 0.519 | 1.603214991 | 1.603214991 | 31.27008281 | 0.076112225 | -1.118545583 | FALSE | FALSE | FALSE |
| TPP2 | 0.122021 | 0.85568134 | 1 | 0.122021025 | 0.122021025 | 4.10907152 | 0.15624606 | 0.862908159 | 1 | 0.156246059 | 0.156246059 | 3.062576511 | 0.780954262 | -0.107374401 | FALSE | TRUE | FALSE |
| TRIM67 | 0.1220154 | 0.85569367 | 0.01885 | 0.122015449 | 0.122015449 | 2.407707995 | 0.15653735 | 0.862837821 | 1 | 0.156537352 | 0.156537352 | 3.093888096 | 0.779465397 | -0.10820316 | FALSE | TRUE | FALSE |
| LDHC | 0.1220073 | 0.85570599 | 0.5097 | 0.122007295 | 0.122007295 | 0.122007295 | 0.72635921 | 0.653901594 | 0.0083 | 0.181646734 | 0.72635921 | 0.72635921 | 0.167971017 | -0.774765649 | FALSE | TRUE | FALSE |
| CPA6 | 0.1219748 | 0.85571831 | 0.51065 | 0.121974813 | 0.121974813 | 0.605054206 | 0.39189093 | 0.794914201 | 0.49805 | 0.044543423 | 0.391890926 | 0.884603391 | 0.311246841 | -0.506895047 | FALSE | TRUE | FALSE |
| DPH2 | 0.1219681 | 0.85573063 | 1 | 1.107620143 | 1.107620143 | 4.127172012 | 0.16049914 | 0.861905358 | 1 | 0.160499143 | 0.160499143 | 3.179486212 | 0.759930055 | -0.119226379 | FALSE | TRUE | FALSE |
| TDRD9 | 0.1219553 | 0.85574295 | 0.8756 | 1.082772635 | 9.86066811 | 21.29261205 | 0.02358245 | 0.892838185 | 1 | 0.206305721 | 1.852438806 | 6.957836772 | 5.171441392 | 0.713611607 | FALSE | TRUE | FALSE |
| VSIG8 | 0.1219259 | 0.85575526 | 1 | 104886.1024 | 10514841.38 | 116807343.5 | 0.19175786 | 0.853932636 | 1 | 1.777667065 | 16.91368021 | 16.91368021 | 0.635832647 | -0.196657177 | FALSE | TRUE | FALSE |
| HCAR1 | 0.1219059 | 0.85576758 | 1 | 1.950143407 | 7.802797285 | 7.802797285 | 0.62346528 | 0.701791972 | 0.87785 | 2.494011871 | 9.979144068 | 9.979144068 | 0.195529528 | -0.708787647 | FALSE | TRUE | FALSE |
| L3MBTL2 | 0.1217293 | 0.85577989 | 1 | 0.121729281 | 0.121729281 | 2.396040429 | 0.15567981 | 0.863060319 | 0.01855 | 0.155679813 | 0.155679813 | 3.070140658 | 0.781920784 | -0.106837243 | FALSE | TRUE | FALSE |
| ZNF235 | 0.1217127 | 0.8557922 | 0.7495 | 0.121712737 | 0.486877411 | 2.403717114 | 0.04651173 | 0.887446275 | 1 | 0.046511728 | 0.046511728 | 0.919951367 | 2.616818205 | 0.417773552 | FALSE | TRUE | FALSE |
| USP29 | 0.1215719 | 0.85580451 | 0.8789 | 1.066347503 | 9.594020469 | 21.01658931 | 0.02433112 | 0.892673979 | 1 | 0.210734571 | 0.210734571 | 0.822849232 | 4.996561645 | 0.69867125 | FALSE | TRUE | FALSE |
| KLHL3 | 0.1215655 | 0.85581681 | 0.50715 | 1.148584798 | 1.148584798 | 4.117417959 | 0.16108731 | 0.861774564 | 1 | 0.161087308 | 0.161087308 | 3.18326917 | 0.754655894 | -0.122251032 | FALSE | TRUE | FALSE |
| ANXA10 | 0.1215542 | 0.85582912 | 1 | 1.09294755 | 1.09294755 | 1.09294755 | 0.15957887 | 0.862142566 | 1 | 0.15957887 | 0.15957887 | 0.15957887 | 0.761718772 | -0.118205342 | FALSE | TRUE | FALSE |
| GAPVD1 | 0.1214962 | 0.85584142 | 0.5165 | 0.485875359 | 0.485875359 | 9.452015575 | 0.15588638 | 0.863025234 | 0.5184 | 0.623421795 | 0.623421795 | 12.1677264 | 0.779389253 | -0.108245587 | FALSE | TRUE | FALSE |
| PPP2R3C | 0.1214523 | 0.85585372 | 0.012 | 0.121452308 | 0.121452308 | 2.410280732 | 1.37242327 | 0.548348152 | 0.50805 | 1.37242327 | 1.37242327 | 1.37242327 | 0.088494789 | -1.053082301 | FALSE | FALSE | FALSE |
| STPG1 | 0.1213486 | 0.85586602 | 1 | 0.121348566 | 0.121348566 | 0.121348566 | 0.15000589 | 0.864575634 | 1 | 1.355370504 | 1.355370504 | 1.355370504 | 0.808958693 | -0.092073654 | FALSE | TRUE | FALSE |
| PDK2 | 0.1213464 | 0.85587831 | 0.7552 | 1.106594801 | 4.425725175 | 4.425725175 | 0.15991945 | 0.862083338 | 1 | 0.639594423 | 0.639594423 | 0.639594423 | 0.758797244 | -0.119874255 | FALSE | TRUE | FALSE |
| SH3PXD2A | 0.1212593 | 0.85589061 | 0.034 | 0.121259263 | 0.121259263 | 2.361357887 | 0.15549249 | 0.863153792 | 1 | 0.155492487 | 0.155492487 | 3.037607998 | 0.779840012 | -0.107994486 | FALSE | TRUE | FALSE |
| MOCS3 | 0.1210715 | 0.8559029 | 0.0176 | 0.121071459 | 0.121071459 | 2.38812736 | 2.42470064 | 0.453021519 | 0.25985 | 0.606130386 | 2.42470064 | 11.97567386 | 0.049932539 | -1.30161635 | FALSE | FALSE | FALSE |
| ABCD2 | 0.1210574 | 0.85591519 | 0.89445 | 0.483896169 | 7.73747771 | 30.95178635 | 0.12590278 | 0.869911345 | 0.26175 | 0.125902783 | 1.198592249 | 2.488401449 | 0.961514832 | -0.017044012 | FALSE | TRUE | FALSE |
| CCDC54 | 0.1210532 | 0.85592748 | 1 | 0.121053194 | 0.121053194 | 0.121053194 | 0.15452405 | 0.863421841 | 1 | 0.154524053 | 0.154524053 | 0.154524053 | 0.783393857 | -0.106019838 | FALSE | TRUE | FALSE |
| LUZP1 | 0.1209905 | 0.85593976 | 1 | 0.120990456 | 0.120990456 | 2.369138163 | 0.15476462 | 0.863363657 | 1 | 0.154764621 | 0.154764621 | 3.038120197 | 0.781770767 | -0.106920573 | FALSE | TRUE | FALSE |
| TRIM68 | 0.1209589 | 0.85595205 | 0.01455 | 0.120958938 | 0.120958938 | 0.598790251 | 0.04261617 | 0.888423972 | 0.8787 | 1.514177345 | 13.80714528 | 13.80714528 | 2.838334039 | 0.453063506 | FALSE | TRUE | FALSE |
| FCRLB | 0.1209563 | 0.85596433 | 1 | 0.483775179 | 0.483775179 | 2.386138241 | 1.37235197 | 0.548479577 | 0.265 | 1.37235197 | 5.488908648 | 11.8559022 | 0.088137933 | -1.054837141 | FALSE | FALSE | FALSE |
| HIP1 | 0.1209165 | 0.85597661 | 1 | 0.120916522 | 0.120916522 | 2.365718447 | 1.38181622 | 0.547029642 | 0.5154 | 1.381816219 | 1.381816219 | 27.10593181 | 0.087505502 | -1.057964638 | FALSE | FALSE | FALSE |
| TACR1 | 0.1208969 | 0.85598889 | 1 | 1.100713134 | 1.100713134 | 2.395836736 | 0.15933428 | 0.862237228 | 0.01175 | 0.15933428 | 0.15933428 | 3.161030024 | 0.758762358 | -0.119894222 | FALSE | TRUE | FALSE |
| A1BG | 0.1208542 | 0.85600117 | 1 | 1.090363855 | 1.090363855 | 2.390705326 | 0.15889653 | 0.862331768 | 1 | 0.158896535 | 0.158896535 | 3.147375442 | 0.760584221 | -0.118852688 | FALSE | TRUE | FALSE |
| CPN1 | 0.1208517 | 0.85601344 | 1 | 0.120851729 | 0.120851729 | 2.389060408 | 1.37830142 | 0.547557536 | 0.5072 | 1.378301424 | 1.378301424 | 2.969691575 | 0.08768164 | -1.057091335 | FALSE | FALSE | FALSE |
| CENPC | 0.1208249 | 0.85602571 | 0.0188 | 0.120824858 | 0.120824858 | 2.386831064 | 0.13987192 | 0.866806785 | 0.01635 | 0.139871922 | 0.139871922 | 2.767177186 | 0.863824962 | -0.06357425 | FALSE | TRUE | FALSE |
| ZNF575 | 0.1207467 | 0.85603798 | 1 | 118124539.6 | 2.44679E+11 | 3.53199E+13 | 0.18501236 | 0.855736939 | 1 | 157362.6932 | 196710411.9 | 2217252003 | 0.652641129 | -0.185325561 | FALSE | TRUE | FALSE |
| EIF4B | 0.1207021 | 0.85605025 | 1 | 0.12070211 | 0.12070211 | 2.378037837 | 0.15510345 | 0.86324714 | 1 | 0.155103447 | 0.155103447 | 3.061288496 | 0.778203916 | -0.108906588 | FALSE | TRUE | FALSE |
| ZNF180 | 0.1206342 | 0.85606252 | 0.5029 | 0.12063419 | 0.12063419 | 0.47672324 | 0.19744319 | 0.852718946 | 0.50825 | 0.19744319 | 0.19744319 | 0.451294504 | 0.610981773 | -0.213971746 | FALSE | TRUE | FALSE |
| OR51Q1 | 0.120589 | 0.85607478 | 0.00865 | 0.120588988 | 0.120588988 | 0.120588988 | 0.15386381 | 0.863607708 | 1 | 0.153863813 | 0.153863813 | 0.153863813 | 0.78373846 | -0.105828841 | FALSE | TRUE | FALSE |
| THAP10 | 0.1205409 | 0.85608704 | 1 | 0.120540945 | 0.120540945 | 0.120540945 | 1.37694965 | 0.54768983 | 0.50275 | 1.376949646 | 1.376949646 | 1.376949646 | 0.087542014 | -1.057783467 | FALSE | FALSE | FALSE |
| JPH2 | 0.120395 | 0.8560993 | 1 | 1.106873719 | 10.44438483 | 21.77532394 | 0.13910605 | 0.866939807 | 0.9854 | 117.1595638 | 4653.211015 | 47338.30866 | 0.865490671 | -0.062737609 | FALSE | TRUE | FALSE |
| PCED1A | 0.1203707 | 0.85611156 | 1 | 1.086104291 | 1.086104291 | 2.380704564 | 1.27073457 | 0.558653598 | 0.93795 | 11.80571228 | 1098.842004 | 1098.842004 | 0.094725323 | -1.023533903 | FALSE | FALSE | FALSE |
| LRRC28 | 0.1203665 | 0.85612382 | 1 | 0.120366492 | 0.120366492 | 2.389309916 | 2.380417 | 0.457482591 | 0.5039 | 2.380416997 | 9.525527829 | 9.525527829 | 0.050565297 | -1.296147439 | FALSE | FALSE | FALSE |
| PPBP | 0.1203522 | 0.85613607 | 0.00275 | 0.120352213 | 0.120352213 | 0.120352213 | 0.12771638 | 0.869474636 | 1 | 0.12771638 | 0.12771638 | 0.12771638 | 0.942339685 | -0.025792519 | FALSE | TRUE | FALSE |
| TBX20 | 0.1203149 | 0.85614833 | 1 | 0.12031493 | 0.480991248 | 2.383168074 | 0.15466854 | 0.863398573 | 1 | 0.61836972 | 0.61836972 | 3.067178106 | 0.777888818 | -0.109082472 | FALSE | TRUE | FALSE |
| SLC22A14 | 0.1200973 | 0.85616058 | 0.7554 | 0.265363603 | 1.061225933 | 2.373232648 | 3.47738496 | 0.384590654 | 0.50285 | 3.47738496 | 32.556096 | 32.556096 | 0.034536679 | -1.461719421 | FALSE | FALSE | FALSE |
| ASTN1 | 0.1200753 | 0.85617283 | 0.04015 | 0.120075283 | 0.120075283 | 4.042481844 | 0.15431334 | 0.863479977 | 0.0361 | 0.154313337 | 0.154313337 | 3.004764295 | 0.778126405 | -0.108949847 | FALSE | TRUE | FALSE |
| SMURF2 | 0.1200732 | 0.85618507 | 0.0233 | 0.120073232 | 0.120073232 | 2.368151652 | 0.15254867 | 0.863943371 | 1 | 0.610008098 | 0.610008098 | 12.05100668 | 0.787114245 | -0.103962228 | FALSE | TRUE | FALSE |
| REEP6 | 0.1200465 | 0.85619732 | 0.50115 | 0.479997999 | 0.479997999 | 0.479997999 | 0.15579971 | 0.863036931 | 1 | 0.155799712 | 0.155799712 | 0.155799712 | 0.770518324 | -0.113217029 | FALSE | TRUE | FALSE |
| ZFP69 | 0.1200041 | 0.85620956 | 1 | 0.120004074 | 0.120004074 | 2.377976806 | 0.14888694 | 0.864872593 | 0.5029 | 1.358185693 | 1.358185693 | 2.953582244 | 0.806008033 | -0.09366063 | FALSE | TRUE | FALSE |
| AASDHPPT | 0.1200001 | 0.8562218 | 1 | 0.120000126 | 0.479937333 | 2.384095548 | 0.14889662 | 0.864861195 | 0.7477 | 1.359636594 | 5.437915012 | 5.437915012 | 0.805929142 | -0.09370314 | FALSE | TRUE | FALSE |
| DLG5 | 0.1199805 | 0.85623404 | 1 | 0.119980525 | 2.290527917 | 4.025936536 | 0.15394941 | 0.863572895 | 1 | 0.153949406 | 2.955161944 | 5.173896873 | 0.779350361 | -0.108267259 | FALSE | TRUE | FALSE |

| | | | | | | | | | | | | | | | | | |
|---|---|---|---|---|---|---|---|---|---|---|---|---|---|---|---|---|---|
| DTL | 0.1198403 | 0.85624628 | 0.0192 | 0.119840298 | 0.119840298 | 2.365390302 | 0.15361191 | 0.863665693 | 1 | 0.153611905 | 0.153611905 | 3.036751531 | 0.780149808 | -0.107821994 | FALSE | TRUE | FALSE |
| CEP57L1 | 0.119837 | 0.85625851 | 0.01185 | 0.119836978 | 0.119836978 | 2.379086284 | 0.14418638 | 0.865834952 | 1 | 1.311357453 | 12.24095484 | 12.24095484 | 0.831125522 | -0.080333382 | FALSE | TRUE | FALSE |
| ACVR1B | 0.1198211 | 0.85627075 | 1 | 0.119821052 | 0.119821052 | 2.36830275 | 0.154094 | 0.863561286 | 1 | 0.154094 | 0.154094 | 3.050018557 | 0.777584149 | -0.109252601 | FALSE | TRUE | FALSE |
| ZNF134 | 0.1197707 | 0.85628298 | 0.49755 | 0.119770668 | 1.083836457 | 2.377450041 | 0.15779463 | 0.862567577 | 1 | 0.157794635 | 0.157794635 | 3.135051099 | 0.759028771 | -0.119741762 | FALSE | TRUE | FALSE |
| MARK3 | 0.1197624 | 0.85629521 | 0.02315 | 0.119762445 | 0.119762445 | 2.357062386 | 12.5244864 | 0.174559908 | 0.25845 | 1.327361219 | 12.52448638 | 26.17412677 | 0.009562264 | -2.019439271 | FALSE | FALSE | FALSE |
| SLC35F6 | 0.1197568 | 0.85630743 | 1 | 0.119756825 | 0.119756825 | 2.375598184 | 5.52321469 | 0.286147557 | 0.2574 | 1.381362113 | 5.523214694 | 5.523214694 | 0.02168245 | -1.663891651 | FALSE | FALSE | FALSE |
| CDK18 | 0.1196898 | 0.85631966 | 0.2645 | 0.119689811 | 0.478743155 | 2.364698974 | 0.04590436 | 0.887568334 | 0.0138 | 0.045904361 | 0.045904361 | 0.908254035 | 2.607373393 | 0.41620323 | FALSE | TRUE | FALSE |
| IK | 0.1196888 | 0.85633188 | 1 | 0.119688809 | 0.119688809 | 2.364824182 | 0.15314957 | 0.863769951 | 1 | 0.153149565 | 0.153149565 | 3.030348801 | 0.781515823 | -0.107062225 | FALSE | TRUE | FALSE |
| PRKCA | 0.1196605 | 0.8563441 | 1 | 0.119660512 | 0.119660512 | 2.357772646 | 0.60093678 | 0.711966216 | 0.75265 | 0.60093678 | 2.403743635 | 11.86187034 | 0.199123295 | -0.70087793 | FALSE | TRUE | FALSE |
| ERMAP | 0.1196543 | 0.85635632 | 1 | 1.089189827 | 1.089189827 | 2.369982146 | 0.1578907 | 0.862555804 | 1 | 0.157890703 | 0.157890703 | 3.130958485 | 0.757830036 | -0.120428186 | FALSE | TRUE | FALSE |
| CCT6A | 0.1196475 | 0.85636854 | 1 | 0.478376952 | 1.913133749 | 2.371930197 | 0.15728367 | 0.86273219 | 0.0117 | 0.157283665 | 0.157283665 | 3.121335069 | 0.760711709 | -0.118779899 | FALSE | TRUE | FALSE |
| PLEKHB1 | 0.1195695 | 0.85638075 | 0.49615 | 0.119569453 | 1.098473659 | 1.098473659 | 0.15632285 | 0.862896441 | 1 | 0.625146824 | 0.625146824 | 0.625146824 | 0.764887891 | -0.116402214 | FALSE | TRUE | FALSE |
| IGSF5 | 0.1195639 | 0.85639296 | 0.0121 | 0.119563865 | 0.119563865 | 2.370502145 | 1.34076409 | 0.551352967 | 0.50375 | 1.340764086 | 1.340764086 | 2.939987552 | 0.089175916 | -1.049752422 | FALSE | FALSE | FALSE |
| RRP9 | 0.1195602 | 0.85640518 | 1 | 0.11956024 | 0.11956024 | 2.361068588 | 0.15367635 | 0.863642505 | 1 | 0.153676349 | 0.153676349 | 3.039393676 | 0.778000263 | -0.109020256 | FALSE | TRUE | FALSE |
| ARPP21 | 0.1195271 | 0.85641738 | 0.8749 | 10.18359383 | 97.67444256 | 200.608244 | 0.16775542 | 0.860247379 | 1 | 0.167755421 | 0.167755421 | 3.310623878 | 0.712508065 | -0.147210216 | FALSE | TRUE | FALSE |
| PPIL4 | 0.1195032 | 0.85642959 | 0.0124 | 0.119503198 | 0.119503198 | 2.37120703 | 0.1528595 | 0.863862496 | 1 | 0.152859505 | 0.152859505 | 3.035947518 | 0.781784544 | -0.10691292 | FALSE | TRUE | FALSE |
| APOE | 0.119468 | 0.85644179 | 1 | 0.119467986 | 0.119467986 | 0.119467986 | 1.34221014 | 0.551223007 | 0.26135 | 1.342210138 | 12.8205415 | 12.8205415 | 0.089008407 | -1.050568971 | FALSE | FALSE | FALSE |
| ROCK2 | 0.119446 | 0.856454 | 0.03785 | 0.119446038 | 0.119446038 | 4.010066356 | 5.15460055 | 0.300135313 | 0.01 | 0.15330566 | 0.15330566 | 3.022343257 | 0.023172705 | -1.635023265 | FALSE | FALSE | FALSE |
| PTTG1 | 0.1194136 | 0.8564662 | 1 | 0.119413602 | 0.119413602 | 0.119413602 | 0.12248296 | 0.870766084 | 1 | 1.151530083 | 1.151530083 | 1.151530083 | 0.974940544 | -0.011021868 | FALSE | TRUE | FALSE |
| PDIA5 | 0.1193631 | 0.85647839 | 1 | 0.119363105 | 0.119363105 | 2.362146742 | 0.14809308 | 0.865054722 | 1 | 0.148093078 | 1.346663807 | 2.934410134 | 0.806000573 | -0.09366465 | FALSE | TRUE | FALSE |
| SEL1L3 | 0.1193459 | 0.85649059 | 1 | 0.119345937 | 0.119345937 | 2.339886352 | 0.15286716 | 0.863839372 | 0.02525 | 0.152867159 | 0.152867159 | 3.004212645 | 0.780716652 | -0.107506557 | FALSE | TRUE | FALSE |
| PCDHA5 | 0.1192689 | 0.85650278 | 0.96645 | 4.242942864 | 82.61079388 | 330.4814479 | 0.05813016 | 0.885030384 | 0.5145 | 0.058130163 | 0.058130163 | 1.135417664 | 2.051755882 | 0.312125687 | FALSE | TRUE | FALSE |
| MRPS28 | 0.1192008 | 0.85651498 | 0.00605 | 0.11920079 | 0.11920079 | 0.11920079 | 0.15210285 | 0.864070279 | 1 | 0.15210285 | 0.15210285 | 0.15210285 | 0.78368545 | -0.105858216 | FALSE | TRUE | FALSE |
| DNTTIP1 | 0.1191669 | 0.85652717 | 0.0113 | 0.119166939 | 0.119166939 | 2.364072628 | 0.13075611 | 0.868776665 | 1 | 0.13075611 | 0.13075611 | 0.13075611 | 0.911368036 | -0.040306207 | FALSE | TRUE | FALSE |
| MYOG | 0.1190856 | 0.85653936 | 0.00745 | 0.119085641 | 0.119085641 | 0.119085641 | 2.36915423 | 0.458402861 | 0.50105 | 2.369154231 | 9.476243466 | 9.476243466 | 0.050265044 | -1.298733935 | FALSE | FALSE | FALSE |
| CD2AP | 0.1190663 | 0.85655154 | 1 | 1.094072166 | 1.094072166 | 2.357086343 | 0.15753767 | 0.862649932 | 1 | 0.157537666 | 0.157537666 | 3.122500163 | 0.755795799 | -0.121595526 | FALSE | TRUE | FALSE |
| STRIP1 | 0.1190213 | 0.85656373 | 1 | 1.089784741 | 10.24126228 | 21.44150851 | 0.15262085 | 0.863931823 | 1 | 1.403304006 | 13.24289982 | 27.66400899 | 0.779849598 | -0.107989148 | FALSE | TRUE | FALSE |
| HTR4 | 0.119001 | 0.85657591 | 1 | 0.119001022 | 0.119001022 | 2.359118164 | 111.234061 | 0.026155444 | 0.31685 | 11.56366124 | 1096.778029 | 1096.778029 | 0.001069825 | -2.970687103 | FALSE | FALSE | FALSE |
| SPC24 | 0.1189708 | 0.85658809 | 0.0032 | 0.012610274 | 0.118970765 | 0.118970765 | 0.0135219 | 0.895625199 | 0.0028 | 0.013521899 | 0.013521899 | 0.013521899 | 8.798376965 | 0.944402565 | FALSE | TRUE | FALSE |
| TNFRSF21 | 0.1189637 | 0.85660027 | 1 | 0.118963709 | 0.118963709 | 2.345338878 | 0.15302492 | 0.863793099 | 1 | 0.153024919 | 0.153024919 | 3.022025425 | 0.777413964 | -0.109347663 | FALSE | TRUE | FALSE |
| GRIA1 | 0.1189568 | 0.85661245 | 1 | 0.118956833 | 0.118956833 | 2.336845065 | 0.1482813 | 0.864986477 | 1 | 1.387030531 | 1.387030531 | 5.014184585 | 0.802237576 | -0.095697 | FALSE | TRUE | FALSE |
| HIPK3 | 0.1189554 | 0.85662462 | 1 | 0.118955437 | 0.118955437 | 2.32958275 | 0.60420009 | 0.710366387 | 0.51645 | 0.604200091 | 0.604200091 | 11.86211648 | 0.196880866 | -0.705796488 | FALSE | TRUE | FALSE |
| PSG3 | 0.1189072 | 0.85663679 | 0.8763 | 1.037196586 | 9.280090541 | 9.280090541 | 0.02390701 | 0.892756137 | 1 | 0.023907011 | 0.206033029 | 0.474625049 | 4.973737311 | 0.696682844 | FALSE | TRUE | FALSE |
| KRTAP1-1 | 0.1187835 | 0.85664896 | 1 | 0.118783503 | 0.118783503 | 0.118783503 | 1.19133173 | 0.568405626 | 0.5064 | 1.191331735 | 1.191331735 | 1.191331735 | 0.099706488 | -1.001276581 | FALSE | FALSE | FALSE |
| GPR141 | 0.1187568 | 0.85666113 | 1 | 0.118756761 | 0.118756761 | 0.118756761 | 0.1521311 | 0.864035691 | 0.00615 | 0.152131101 | 0.152131101 | 0.152131101 | 0.780621188 | -0.107559665 | FALSE | TRUE | FALSE |
| TMEM176A | 0.1187521 | 0.8566733 | 0.006 | 0.118752092 | 0.118752092 | 0.118752092 | 0.15159484 | 0.864196954 | 1 | 0.151594844 | 0.151594844 | 0.151594844 | 0.783351791 | -0.10604316 | FALSE | TRUE | FALSE |
| LIMK1 | 0.1186288 | 0.85668546 | 1 | 4.471140377 | 43.2584593 | 87.93062267 | 0.16270586 | 0.861440573 | 0.51025 | 0.65055878 | 0.65055878 | 12.81976158 | 0.729099984 | -0.137212911 | FALSE | TRUE | FALSE |
| ST6GALNAC: | 0.1185864 | 0.85669762 | 0.5109 | 0.118586432 | 0.118586432 | 0.470135826 | 0.02313641 | 0.892912674 | 1 | 0.023136407 | 0.023136407 | 0.457377698 | 5.12553367 | 0.709739091 | FALSE | TRUE | FALSE |
| SUPT6H | 0.1184775 | 0.85670979 | 1 | 0.473814004 | 2.281170498 | 9.12283499 | 0.15421078 | 0.863538065 | 1 | 0.154210784 | 0.154210784 | 5.173851029 | 0.768282737 | -0.114478925 | FALSE | TRUE | FALSE |
| GPR180 | 0.1184368 | 0.85672195 | 1 | 1.06641847 | 1.06641847 | 1.06641847 | 0.15602965 | 0.86296672 | 1 | 0.156029651 | 0.156029651 | 0.156029651 | 0.759065773 | -0.119720591 | FALSE | TRUE | FALSE |
| AKR1A1 | 0.1183318 | 0.8567341 | 1 | 1.076165939 | 1.076165939 | 1.076165939 | 5.31162589 | 0.294240611 | 0.50055 | 5.311625894 | 49.78141904 | 49.78141904 | 0.022277893 | -1.652125886 | FALSE | FALSE | FALSE |
| GLUD1 | 0.1183245 | 0.85674626 | 0.5046 | 0.118324482 | 0.118324482 | 1.003341763 | 0.17946483 | 0.85716992 | 0.5038 | 0.179464829 | 0.179464829 | 0.887249185 | 0.6593185 | -0.180904738 | FALSE | TRUE | FALSE |
| MAST3 | 0.1183106 | 0.85675841 | 1 | 0.118310624 | 0.118310624 | 2.286139547 | 12.7275443 | 0.172883803 | 0.2709 | 1.331538604 | 12.7275443 | 246.9408401 | 0.009295636 | -2.031720872 | FALSE | FALSE | FALSE |
| HNRNPCL1 | 0.1183003 | 0.85677056 | 0.75625 | 0.11830026 | 1.150337798 | 1.150337798 | 0.0133182 | 0.895710371 | 1 | 0.013318196 | 0.013318196 | 0.013318196 | 8.882603981 | 0.9485403 | FALSE | TRUE | FALSE |
| PHLPP1 | 0.1182396 | 0.85678271 | 0.03475 | 0.118239619 | 0.118239619 | 3.976409976 | 0.1521035 | 0.864058752 | 1 | 0.152103504 | 0.152103504 | 2.975999672 | 0.777362887 | -0.109376197 | FALSE | TRUE | FALSE |
| ZNF792 | 0.1182096 | 0.85679486 | 1 | 0.118209592 | 0.118209592 | 2.332492359 | 1.29766118 | 0.554966697 | 0.7487 | 1.297661181 | 12.12861885 | 25.64655284 | 0.091094343 | -1.040508593 | FALSE | FALSE | FALSE |
| GGA3 | 0.1181821 | 0.856807 | 1 | 0.118182137 | 0.118182137 | 2.332705666 | 0.1468765 | 0.865338365 | 1 | 1.339394459 | 1.339394459 | 2.903647484 | 0.804636143 | -0.094400463 | FALSE | TRUE | FALSE |
| SLC25A41 | 0.1181408 | 0.85681915 | 0.89125 | 1.889406666 | 30.24718258 | 121.0674409 | 24.0074004 | 0.113657404 | 0.76555 | 96.05015688 | 6160.382453 | 52236.48602 | 0.004921017 | -2.307945149 | FALSE | FALSE | FALSE |
| MAP4K4 | 0.1181153 | 0.85683129 | 1 | 0.118115294 | 0.118115294 | 3.97399373 | 0.15199201 | 0.864116369 | 1 | 0.151992006 | 0.151992006 | 2.978673789 | 0.777115175 | -0.10951461 | FALSE | TRUE | FALSE |
| CEP41 | 0.1181058 | 0.85684343 | 0.9883 | 94.50549924 | 9228.07245 | 94497.52651 | 0.18574538 | 0.855594712 | 0.9834 | 37.89570273 | 1488.690518 | 14984.49052 | 0.635847995 | -0.196646694 | FALSE | TRUE | FALSE |
| PSMD3 | 0.1180847 | 0.85685557 | 0.9961 | 172.9234019 | 17528.49655 | 70112.38161 | 0.18051947 | 0.856966892 | 1 | 0.180519475 | 0.180519475 | 3.572314104 | 0.654138327 | -0.184330404 | FALSE | TRUE | FALSE |
| HOXC6 | 0.1180784 | 0.8568677 | 0.50455 | 0.472056016 | 0.472056016 | 0.472056016 | 0.15376215 | 0.863630908 | 1 | 0.153762152 | 0.153762152 | 0.153762152 | 0.767929128 | -0.114678859 | FALSE | TRUE | FALSE |
| CALCB | 0.1180747 | 0.85687983 | 1 | 0.118074731 | 0.118074731 | 0.118074731 | 0.13022692 | 0.868895278 | 1 | 0.130226918 | 0.130226918 | 0.130226918 | 0.906684527 | -0.042543796 | FALSE | TRUE | FALSE |
| KLK3 | 0.1180681 | 0.85689196 | 1 | 0.118068094 | 0.118068094 | 0.118068094 | 0.15145886 | 0.86425446 | 1 | 0.151458855 | 0.151458855 | 0.151458855 | 0.77953906 | -0.108162119 | FALSE | TRUE | FALSE |
| CASP6 | 0.1180632 | 0.85690409 | 1 | 0.118063207 | 0.118063207 | 0.118063207 | 0.15137239 | 0.86428894 | 1 | 0.151372393 | 0.151372393 | 0.151372393 | 0.779952046 | -0.107932098 | FALSE | TRUE | FALSE |
| HLA-DRB5 | 0.1180399 | 0.85691622 | 0.999 | 10502.86154 | 12883958.82 | 144863568.4 | 4.81169642 | 0.314775912 | 0.96655 | 1251.99689 | 135813.0219 | 1467352.195 | 0.024531859 | -1.610269537 | FALSE | FALSE | FALSE |
| MYL6B | 0.1179948 | 0.85692834 | 0.00555 | 0.117994761 | 0.117994761 | 0.117994761 | 1.36375401 | 0.549528124 | 0.5014 | 1.363754007 | 1.363754007 | 1.363754007 | 0.086522027 | -1.062873313 | FALSE | FALSE | FALSE |
| TUBA4A | 0.1179864 | 0.85694047 | 1 | 7.545949711 | 120.8308253 | 483.6958392 | 0.15539196 | 0.863188812 | 1 | 2.484830182 | 39.77397791 | 39.77397791 | 0.759282672 | -0.119596511 | FALSE | TRUE | FALSE |
| MAPK7 | 0.1179783 | 0.85695259 | 1 | 0.117978262 | 0.117978262 | 2.31433268 | 0.15178387 | 0.864173939 | 0.0211 | 0.151783872 | 0.151783872 | 2.984357343 | 0.777277981 | -0.109423635 | FALSE | TRUE | FALSE |
| HLX | 0.1178978 | 0.85696471 | 0.01855 | 0.117897815 | 0.117897815 | 2.32412846 | 0.15190164 | 0.864139403 | 1 | 0.151901638 | 0.151901638 | 2.999618576 | 0.77614578 | -0.110056699 | FALSE | TRUE | FALSE |
| SLCO1C1 | 0.1178893 | 0.85697682 | 0.26645 | 0.117889322 | 1.066687992 | 2.332298294 | 0.09232926 | 0.877691766 | 0.5071 | 0.092329258 | 0.092329258 | 0.457434385 | 1.276836013 | 0.106135123 | FALSE | TRUE | FALSE |
| CUL4A | 0.1178835 | 0.85698894 | 1 | 0.117883538 | 0.117883538 | 2.325297402 | 0.151744 | 0.864185447 | 1 | 0.151744005 | 0.151744005 | 2.998154032 | 0.776857957 | -0.109658382 | FALSE | TRUE | FALSE |
| OR56A3 | 0.1178736 | 0.85700105 | 1 | 0.117873624 | 0.117873624 | 0.117873624 | 1.33022652 | 0.552261042 | 0.5035 | 1.330226515 | 1.330226515 | 1.330226515 | 0.088611693 | -1.052508965 | FALSE | FALSE | FALSE |
| DCP1B | 0.1177783 | 0.85701316 | 1 | 1.064444369 | 1.064444369 | 3.986811643 | 1.32859561 | 0.552519839 | 0.75355 | 1.32859561 | 12.38280355 | 26.28803785 | 0.088648725 | -1.052327504 | FALSE | FALSE | FALSE |
| ATMIN | 0.1177471 | 0.85702527 | 1 | 1.065345625 | 9.893102744 | 20.99561921 | 0.16024022 | 0.861976603 | 1 | 0.160240218 | 0.160240218 | 3.163535254 | 0.734815872 | -0.133821472 | FALSE | TRUE | FALSE |
| TMEM63A | 0.1176851 | 0.85703738 | 0.7555 | 0.11768511 | 1.028730041 | 2.314899095 | 0.19848145 | 0.852476608 | 0.5097 | 0.198481447 | 0.198481447 | 0.778543715 | 0.592927511 | -0.226998399 | FALSE | TRUE | FALSE |
| CD22 | 0.1176531 | 0.85704948 | 1 | 0.117653063 | 0.117653063 | 2.311083413 | 0.15120468 | 0.864323403 | 1 | 0.15120468 | 0.15120468 | 2.976524448 | 0.778104642 | -0.108961994 | FALSE | TRUE | FALSE |
| DGKI | 0.1175338 | 0.85706158 | 1 | 10.28163873 | 995.4212441 | 995.4212441 | 1.54046864 | 0.53177191 | 0.51325 | 1.540468644 | 1.540468644 | 30.28069095 | 0.076297446 | -1.117490002 | FALSE | FALSE | FALSE |
| KCTD18 | 0.1174333 | 0.85707369 | 0.5042 | 0.117433281 | 0.117433281 | 0.581326469 | 0.04476091 | 0.887933037 | 1 | 0.044760915 | 0.044760915 | 0.887044791 | 2.6235675 | 0.418892242 | FALSE | TRUE | FALSE |
| GK5 | 0.1174001 | 0.85708578 | 1 | 0.117400075 | 0.117400075 | 2.326703965 | 0.15115111 | 0.864334888 | 1 | 0.151151113 | 0.151151113 | 2.998870032 | 0.776706653 | -0.109742975 | FALSE | TRUE | FALSE |
| ABL1 | 0.1173294 | 0.85709788 | 1 | 0.469190469 | 0.469190469 | 9.123523097 | 0.15286224 | 0.863850935 | 1 | 0.15286224 | 0.15286224 | 2.982382472 | 0.767549996 | -0.114893326 | FALSE | TRUE | FALSE |
| PRPH2 | 0.1173012 | 0.85710998 | 0.0101 | 0.117301239 | 0.117301239 | 2.326584207 | 0.15092995 | 0.864415222 | 1 | 0.150929945 | 0.150929945 | 0.150929945 | 0.777189969 | -0.109472813 | FALSE | TRUE | FALSE |
| YARS2 | 0.1172759 | 0.85712207 | 0.9409 | 2.381409141 | 89.17122221 | 89.17122221 | 0.04733335 | 0.887258657 | 0.5046 | 0.424882943 | 0.424882943 | 0.936474649 | 2.477658276 | 0.394041407 | FALSE | TRUE | FALSE |

| | | | | | | | | | | | | | | | | |
|---|---|---|---|---|---|---|---|---|---|---|---|---|---|---|---|---|
| SLC7A11 | 0.1172197 | 0.85713416 | 0.6635 | 0.468632824 | 7.495870394 | 29.99020572 | 0.21358681 | 0.848718972 | 0.68765 | 0.213586813 | 3.415530732 | 3.415530732 | 0.548815457 | -0.260573666 | FALSE | TRUE | FALSE |
| TNR | 0.1171516 | 0.85714625 | 0.0425 | 0.117151582 | 0.117151582 | 2.272622229 | 0.17731074 | 0.857675106 | 0.51755 | 0.177310738 | 0.177310738 | 3.452112563 | 0.660713409 | -0.179986879 | FALSE | TRUE | FALSE |
| RUNX3 | 0.1171262 | 0.85715834 | 1 | 0.117126225 | 0.117126225 | 2.314237512 | 1.37132849 | 0.549004426 | 0.5103 | 1.37132849 | 1.37132849 | 2.866199749 | 0.085410772 | -1.068487351 | FALSE | FALSE | FALSE |
| NUB1 | 0.1170791 | 0.85717043 | 0.0176 | 0.117079051 | 0.117079051 | 2.314127798 | 0.1501704 | 0.864506927 | 1 | 0.150170405 | 0.150170405 | 2.972382361 | 0.779641311 | -0.108105157 | FALSE | TRUE | FALSE |
| EXOSC3 | 0.1170652 | 0.85718251 | 1 | 7.49293471 | 68.56979536 | 274.4256006 | 0.15855411 | 0.862437976 | 1 | 0.634100299 | 2.536570714 | 2.536570714 | 0.738329602 | -0.131749719 | FALSE | TRUE | FALSE |
| AHSG | 0.1170575 | 0.85719459 | 0.5029 | 1.054294729 | 1.054294729 | 2.322525154 | 0.15443257 | 0.863445101 | 1 | 0.154432568 | 0.154432568 | 0.154432568 | 0.757984303 | -0.120339788 | FALSE | TRUE | FALSE |
| SNAPIN | 0.1170024 | 0.85720667 | 1 | 0.117002395 | 0.117002395 | 0.117002395 | 0.12569146 | 0.869964413 | 1 | 0.125691463 | 0.125691463 | 0.125691463 | 0.930869868 | -0.031111027 | FALSE | TRUE | FALSE |
| NANS | 0.11697 | 0.85721875 | 1 | 0.116970039 | 0.116970039 | 2.319962516 | 0.14995183 | 0.864587078 | 1 | 0.14995183 | 0.14995183 | 0.14995183 | 0.780050763 | -0.107877134 | FALSE | TRUE | FALSE |
| BAX | 0.1169535 | 0.85723083 | 1 | 0.11695347 | 1.085321798 | 1.085321798 | 0.13530527 | 0.867689005 | 1 | 0.135305274 | 0.135305274 | 0.135305274 | 0.864367416 | -0.063301613 | FALSE | TRUE | FALSE |
| STIL | 0.1169268 | 0.8572429 | 0.7588 | 1.0684007 | 10.019637 | 35.99986275 | 0.15485961 | 0.863282117 | 1 | 1.422549345 | 1.422549345 | 27.90989827 | 0.755050097 | -0.122024232 | FALSE | TRUE | FALSE |
| HENMT1 | 0.1168858 | 0.85725497 | 1 | 1.052578735 | 1.052578735 | 2.320219369 | 0.15423135 | 0.863526451 | 1 | 0.154231353 | 0.154231353 | 0.154231353 | 0.757859865 | -0.120411092 | FALSE | TRUE | FALSE |
| RND2 | 0.1168648 | 0.85726704 | 0.50165 | 1.079119994 | 1.079119994 | 1.079119994 | 1.39284947 | 0.545703631 | 0.0062 | 1.392849473 | 1.392849473 | 1.392849473 | 0.083903407 | -1.076220402 | FALSE | FALSE | FALSE |
| VAX1 | 0.1168444 | 0.85727911 | 1 | 10.49465461 | 103.3461544 | 103.3461544 | 0.16541949 | 0.860852406 | 1 | 0.165419486 | 0.165419486 | 0.165419486 | 0.706352109 | -0.150978754 | FALSE | TRUE | FALSE |
| GAS2L3 | 0.1168388 | 0.85729118 | 0.01725 | 0.116838799 | 0.116838799 | 0.988290638 | 0.04454876 | 0.887981497 | 0.01465 | 0.044548757 | 0.044548757 | 0.88141899 | 2.622717377 | 0.418751494 | FALSE | TRUE | FALSE |
| ACBD5 | 0.1168369 | 0.85730324 | 1 | 0.116836902 | 0.116836902 | 2.314509942 | 12.5054805 | 0.175230268 | 0.25315 | 1.312596948 | 12.50548052 | 12.50548052 | 0.009342856 | -2.029520351 | FALSE | FALSE | FALSE |
| HPSE2 | 0.1168316 | 0.8573153 | 0.755 | 1.068394985 | 10.02767987 | 10.02767987 | 0.15956952 | 0.862154406 | 1 | 0.15956952 | 0.15956952 | 3.163063352 | 0.732167659 | -0.135389459 | FALSE | TRUE | FALSE |
| GRIK4 | 0.1168164 | 0.85732736 | 0.27165 | 0.116816423 | 0.467079908 | 2.286424131 | 0.04532623 | 0.88777122 | 0.02445 | 0.04532623 | 0.04532623 | 0.889443682 | 2.577236688 | 0.411154305 | FALSE | TRUE | FALSE |
| AFAP1L2 | 0.1167955 | 0.85733942 | 1 | 0.467119629 | 0.467119629 | 9.179963593 | 0.15182754 | 0.864162429 | 1 | 0.151827538 | 0.151827538 | 2.990002285 | 0.769264302 | -0.113924421 | FALSE | TRUE | FALSE |
| KCTD3 | 0.1167369 | 0.85735147 | 1 | 10.01926661 | 96.45174424 | 197.5314724 | 0.61172485 | 0.706944666 | 0.87715 | 5.623309195 | 53.05438958 | 53.05438958 | 0.190832358 | -0.719347983 | FALSE | TRUE | FALSE |
| CA6 | 0.1166404 | 0.85736353 | 1 | 1.077080995 | 1.077080995 | 1.077080995 | 0.15481628 | 0.86334037 | 1 | 0.154816279 | 0.154816279 | 0.154816279 | 0.753411416 | -0.122967804 | FALSE | TRUE | FALSE |
| EDC4 | 0.1166174 | 0.85737558 | 1 | 0.116617408 | 0.116617408 | 3.923096966 | 0.14541595 | 0.865553213 | 1 | 1.339341373 | 1.339341373 | 26.07177197 | 0.801957483 | -0.095848656 | FALSE | TRUE | FALSE |
| STX18 | 0.1165809 | 0.85738763 | 1 | 0.1165809 | 0.1165809 | 2.313369368 | 0.1495028 | 0.864701424 | 1 | 0.149502802 | 0.149502802 | 0.149502802 | 0.779790736 | -0.108021929 | FALSE | TRUE | FALSE |
| MVD | 0.1165145 | 0.85739968 | 0.26475 | 0.116514487 | 0.465990287 | 2.305088817 | 0.04491623 | 0.88787645 | 0.01305 | 0.044916232 | 0.044916232 | 0.889765793 | 2.594039665 | 0.413976612 | FALSE | TRUE | FALSE |
| PM20D2 | 0.1165143 | 0.85741172 | 0.0096 | 0.116514296 | 0.116514296 | 0.116514296 | 0.14517413 | 0.865620932 | 1 | 1.330553961 | 1.330553961 | 1.330553961 | 0.802583055 | -0.095510014 | FALSE | TRUE | FALSE |
| ATG4D | 0.1164868 | 0.85742377 | 1 | 0.11648683 | 0.11648683 | 2.303026117 | 0.14959403 | 0.864655708 | 1 | 0.149594028 | 0.149594028 | 2.961658138 | 0.778686368 | -0.108637428 | FALSE | TRUE | FALSE |
| NPFF | 0.1164813 | 0.85743581 | 1 | 0.11648127 | 0.11648127 | 0.11648127 | 0.14910593 | 0.864804175 | 1 | 0.149105928 | 0.149105928 | 0.149105928 | 0.78119812 | -0.107238811 | FALSE | TRUE | FALSE |
| IL1R2 | 0.1164781 | 0.85744785 | 0.5056 | 1.053088746 | 1.053088746 | 2.30878152 | 0.14487974 | 0.865654767 | 1 | 1.316075091 | 12.27019469 | 12.27019469 | 0.803964134 | -0.094763325 | FALSE | TRUE | FALSE |
| CFHR1 | 0.1161565 | 0.85745989 | 0.0099 | 0.116156524 | 0.116156524 | 0.116156524 | 0.14896925 | 0.864838393 | 1 | 0.148969255 | 0.148969255 | 0.148969255 | 0.779734877 | -0.10805304 | FALSE | TRUE | FALSE |
| EXO1 | 0.1161355 | 0.85747193 | 0.5165 | 0.116135505 | 0.116135505 | 2.287000052 | 0.71565559 | 0.658454243 | 0.69515 | 1.608973109 | 59.34007319 | 59.34007319 | 0.162278484 | -0.789739058 | FALSE | TRUE | FALSE |
| GAL3ST2 | 0.1160666 | 0.85748396 | 1 | 1.042121035 | 9.603589329 | 9.603589329 | 0.14860908 | 0.864929557 | 1 | 1.34284314 | 12.45388535 | 12.45388535 | 0.78101961 | -0.107338062 | FALSE | TRUE | FALSE |
| MYL10 | 0.1160476 | 0.857496 | 0.00545 | 0.116047556 | 0.116047556 | 0.116047556 | 0.14482081 | 0.865677316 | 1 | 0.144820815 | 1.337203182 | 1.337203182 | 0.801318209 | -0.096194988 | FALSE | TRUE | FALSE |
| DOK1 | 0.1160033 | 0.85750803 | 1 | 0.116003293 | 0.116003293 | 2.291995292 | 0.59006865 | 0.716502223 | 0.50145 | 0.590068653 | 0.590068653 | 2.919357814 | 0.196592876 | -0.706432224 | FALSE | TRUE | FALSE |
| NFATC4 | 0.1159306 | 0.85752006 | 1 | 0.11593062 | 0.11593062 | 2.266618106 | 0.14764868 | 0.86519101 | 1 | 0.590503169 | 0.590503169 | 11.57641499 | 0.785178834 | -0.105031416 | FALSE | TRUE | FALSE |
| APOBEC3G | 0.1159239 | 0.85753208 | 1 | 1.045637831 | 1.045637831 | 2.298396874 | 1.3072576 | 0.553810447 | 0.75095 | 1.307257596 | 12.16279776 | 12.16279776 | 0.088677178 | -1.052188137 | FALSE | FALSE | FALSE |
| COLEC11 | 0.1159161 | 0.85754411 | 1 | 0.463472019 | 4.31526984 | 4.31526984 | 0.15591036 | 0.863013535 | 1 | 0.155910359 | 0.155910359 | 0.155910359 | 0.743479294 | -0.128731122 | FALSE | TRUE | FALSE |
| GNG12 | 0.1158823 | 0.85755613 | 1 | 0.115882343 | 0.115882343 | 0.115882343 | 0.11803572 | 0.871703644 | 0.0014 | 0.118035716 | 0.118035716 | 0.118035716 | 0.981756603 | -0.007996169 | FALSE | TRUE | FALSE |
| GIPC1 | 0.1158598 | 0.85756815 | 0.01175 | 0.115859755 | 0.115859755 | 2.296484332 | 0.14946714 | 0.864712848 | 1 | 0.149467137 | 0.149467137 | 0.149467137 | 0.775152033 | -0.11061311 | FALSE | TRUE | FALSE |
| RABEPK | 0.1158176 | 0.85758017 | 0.62705 | 0.463275436 | 4.082606464 | 4.082606464 | 0.38337759 | 0.798044231 | 0.74925 | 0.383377586 | 3.493940096 | 3.493940096 | 0.302098074 | -0.519852044 | FALSE | TRUE | FALSE |
| CCNI | 0.1157824 | 0.85759219 | 1 | 0.115782433 | 0.115782433 | 0.115782433 | 0.12915633 | 0.869131897 | 1 | 0.129156332 | 0.129156332 | 0.129156332 | 0.896451849 | -0.047473033 | FALSE | TRUE | FALSE |
| IRF1 | 0.1157329 | 0.85760421 | 0.009 | 0.115732902 | 0.115732902 | 0.115732902 | 0.144794 | 0.865688588 | 1 | 1.368042745 | 1.368042745 | 1.368042745 | 0.799293509 | -0.097293714 | FALSE | TRUE | FALSE |
| CDC14B | 0.1156204 | 0.85761622 | 0.51155 | 1.072423561 | 1.072423561 | 2.288580875 | 0.60801901 | 0.708530341 | 0.5065 | 0.608019014 | 0.608019014 | 3.01340962 | 0.190159212 | -0.720882632 | FALSE | TRUE | FALSE |
| ABHD5 | 0.1155758 | 0.85762823 | 0.00935 | 0.1155758 | 0.1155758 | 0.1155758 | 0.14884643 | 0.864895384 | 0.00955 | 0.148846426 | 0.148846426 | 0.148846426 | 0.776476825 | -0.109871502 | FALSE | TRUE | FALSE |
| RHCG | 0.1154418 | 0.85764024 | 1 | 0.11544178 | 0.11544178 | 2.282117219 | 0.13803695 | 0.867172024 | 1 | 5.035735429 | 47.14188998 | 47.14188998 | 0.836310724 | -0.077632334 | FALSE | TRUE | FALSE |
| CCNJL | 0.1154173 | 0.85765225 | 0.0129 | 0.11541728 | 0.11541728 | 2.286397872 | 0.1481018 | 0.865043353 | 1 | 0.148101804 | 0.148101804 | 2.937228363 | 0.779310426 | -0.108289514 | FALSE | TRUE | FALSE |
| CASP9 | 0.1153778 | 0.85766426 | 1 | 0.11537779 | 0.11537779 | 2.288217206 | 0.14815019 | 0.865020608 | 0.0104 | 0.148150188 | 0.148150188 | 2.941133507 | 0.77878936 | -0.10857999 | FALSE | TRUE | FALSE |
| UGT1A8 | 0.1153775 | 0.85767626 | 1 | 1.053717588 | 1.053717588 | 3.905682547 | 0.15301632 | 0.86380467 | 0.0132 | 0.153016319 | 0.153016319 | 3.030824708 | 0.75402116 | -0.122616467 | FALSE | TRUE | FALSE |
| GTPBP8 | 0.1153651 | 0.85768827 | 1 | 0.115365062 | 0.115365062 | 0.115365062 | 0.14796338 | 0.865100181 | 1 | 0.147963377 | 0.147963377 | 0.147963377 | 0.7796866 | -0.10807993 | FALSE | TRUE | FALSE |
| TARS2 | 0.115345 | 0.85770027 | 0.5095 | 0.115345018 | 0.115345018 | 2.269413087 | 1.73521001 | 0.514208174 | 0.0159 | 0.187127212 | 1.735210008 | 3.68892977 | 0.066473232 | -1.177353202 | FALSE | FALSE | FALSE |
| SEC14L1 | 0.1153015 | 0.85771227 | 1 | 1.064533908 | 1.064533908 | 20.93894874 | 0.15326587 | 0.863735215 | 1 | 0.153265874 | 0.153265874 | 3.020672887 | 0.752296962 | -0.123610692 | FALSE | TRUE | FALSE |
| TMEM237 | 0.1152945 | 0.85772426 | 1 | 0.115294496 | 0.115294496 | 2.290258618 | 0.14338154 | 0.865958569 | 1 | 1.295912114 | 1.295912114 | 1.295912114 | 0.804109769 | -0.094684662 | FALSE | TRUE | FALSE |
| CARD14 | 0.1152382 | 0.85773626 | 0.52005 | 0.115238204 | 0.115238204 | 2.251123097 | 0.178834 | 0.857334476 | 0.5135 | 1.588000829 | 14.46369606 | 31.1078201 | 0.644386424 | -0.190853618 | FALSE | TRUE | FALSE |
| DENND4A | 0.1152348 | 0.85774825 | 0.8841 | 10.02449155 | 97.15335596 | 336.1349862 | 0.16285937 | 0.861392737 | 1 | 0.162859372 | 0.162859372 | 5.469956722 | 0.707572634 | -0.150228972 | FALSE | TRUE | FALSE |
| GIT1 | 0.1151892 | 0.85776024 | 1 | 102.131226 | 10677.00704 | 113087.8547 | 0.17564991 | 0.858176817 | 1 | 0.702370244 | 2.809265878 | 13.80361147 | 0.655788509 | -0.183236197 | FALSE | TRUE | FALSE |
| NR0B2 | 0.1151332 | 0.85777223 | 1 | 0.115133175 | 0.115133175 | 0.115133175 | 0.14696983 | 0.865315715 | 1 | 0.587630342 | 0.587630342 | 0.587630342 | 0.783379638 | -0.106027721 | FALSE | TRUE | FALSE |
| IFI16 | 0.11507 | 0.85778422 | 1 | 0.115069954 | 0.115069954 | 2.274938898 | 0.14769867 | 0.865168314 | 1 | 0.147698671 | 0.147698671 | 2.924049438 | 0.779085918 | -0.108414645 | FALSE | TRUE | FALSE |
| OR6B3 | 0.1149494 | 0.8577962 | 0.0095 | 0.114949382 | 0.114949382 | 0.114949382 | 1.19959402 | 0.56755794 | 0.0096 | 1.199594018 | 1.199594018 | 1.199594018 | 0.095823571 | -1.01852765 | FALSE | FALSE | FALSE |
| PTDSS2 | 0.1149416 | 0.85780818 | 1 | 0.11494162 | 0.11494162 | 2.273083523 | 0.14777186 | 0.865122899 | 1 | 0.147771858 | 0.147771858 | 2.926271712 | 0.77783159 | -0.109114423 | FALSE | TRUE | FALSE |
| DVL1 | 0.1149352 | 0.85782017 | 1 | 1.048876196 | 9.824115676 | 20.54194138 | 0.15723059 | 0.862755674 | 0.022 | 0.157230587 | 0.157230587 | 3.087017447 | 0.730997854 | -0.136083898 | FALSE | TRUE | FALSE |
| PRSS58 | 0.114907 | 0.85783214 | 1 | 0.114906979 | 0.114906979 | 0.114906979 | 0.14795828 | 0.86511154 | 1 | 0.147958284 | 0.147958284 | 0.147958284 | 0.776617411 | -0.109792877 | FALSE | TRUE | FALSE |
| DIABLO | 0.1147277 | 0.85784412 | 1 | 0.114727741 | 0.114727741 | 0.114727741 | 0.14770513 | 0.86514561 | 1 | 0.147705128 | 0.147705128 | 0.147705128 | 0.776734988 | -0.109727132 | FALSE | TRUE | FALSE |
| NDUFV2 | 0.1146514 | 0.8578561 | 1 | 1.046018957 | 1.046018957 | 1.046018957 | 0.58340505 | 0.719342932 | 0.5053 | 1.337049636 | 5.347199709 | 5.347199709 | 0.196521149 | -0.706590705 | FALSE | TRUE | FALSE |
| NEUROG3 | 0.1146498 | 0.85786807 | 1 | 0.114649833 | 0.458426675 | 2.274526908 | 0.14961687 | 0.864644274 | 1 | 0.149616867 | 0.149616867 | 0.149616867 | 0.766289491 | -0.11560713 | FALSE | TRUE | FALSE |
| KDM3B | 0.1145727 | 0.85788005 | 0.0484 | 0.114572651 | 0.114572651 | 3.8413475 | 0.14806741 | 0.86506609 | 1 | 0.148067411 | 0.148067411 | 4.972647484 | 0.773787088 | -0.111378522 | FALSE | TRUE | FALSE |
| KRTAP10-1 | 0.1145542 | 0.85789202 | 1 | 0.114554228 | 0.114554228 | 0.114554228 | 0.13830621 | 0.867050482 | 1 | 1.246824014 | 11.53636667 | 11.53636667 | 0.828265241 | -0.081830564 | FALSE | TRUE | FALSE |
| MTRF1L | 0.1145272 | 0.85790398 | 0.50485 | 1.031384489 | 1.031384489 | 1.031384489 | 0.58133167 | 0.720193519 | 0.74835 | 1.316790302 | 5.26697679 | 5.26697679 | 0.197008299 | -0.705515479 | FALSE | TRUE | FALSE |
| SOX11 | 0.1145088 | 0.85791595 | 1 | 0.114508793 | 0.114508793 | 2.263559409 | 99.4324076 | 0.029680783 | 0.000025 | 0.148077405 | 0.148077405 | 2.93122188 | 0.001151624 | -2.938689119 | FALSE | FALSE | FALSE |
| SLC51A | 0.114503 | 0.85792791 | 1 | 0.114503001 | 0.114503001 | 0.114503001 | 0.12660692 | 0.86974125 | 1 | 0.126606921 | 0.126606921 | 0.126606921 | 0.904397647 | -0.043640576 | FALSE | TRUE | FALSE |
| THEM5 | 0.1144515 | 0.85793988 | 0.5049 | 0.457792034 | 0.457792034 | 0.457792034 | 0.14845106 | 0.864963714 | 1 | 0.148451057 | 0.148451057 | 0.148451057 | 0.770971256 | -0.112961813 | FALSE | TRUE | FALSE |
| NLRP7 | 0.1144372 | 0.85795184 | 0.8214 | 9.33479064 | 87.52816668 | 840.7982933 | 0.02358429 | 0.892830731 | 1 | 0.207492114 | 1.873651892 | 7.00995821 | 4.852265986 | 0.685944599 | FALSE | TRUE | FALSE |
| RRP36 | 0.1144108 | 0.8579638 | 1 | 0.114410839 | 0.114410839 | 0.114410839 | 1.24855404 | 0.561410745 | 0.75 | 1.248554036 | 11.57484623 | 11.57484623 | 0.091634672 | -1.037940172 | FALSE | FALSE | FALSE |
| GALNT2 | 0.1143534 | 0.85797575 | 0.0177 | 0.114353402 | 0.114353402 | 2.256999492 | 0.14736132 | 0.865236382 | 1 | 0.147361318 | 0.147361318 | 2.91307997 | 0.776006918 | -0.110134407 | FALSE | TRUE | FALSE |
| SH3D21 | 0.1143445 | 0.85798771 | 0.5112 | 0.114344473 | 0.114344473 | 0.267195026 | 1.55924778 | 0.530364011 | 0.50075 | 1.559247783 | 14.09224851 | 14.09224851 | 0.0733331 | -1.134699958 | FALSE | FALSE | FALSE |
| ZNF440 | 0.1143342 | 0.85799966 | 0.75195 | 0.114334203 | 1.013401381 | 2.262706542 | 1.64214084 | 0.522335183 | 0.50495 | 1.642140837 | 15.07571311 | 15.07571311 | 0.069625089 | -1.157234234 | FALSE | FALSE | FALSE |

| | | | | | | | | | | | | | | | | | |
|---|---|---|---|---|---|---|---|---|---|---|---|---|---|---|---|---|---|
| ELANE | 0.114292 | 0.85801161 | 1 | 0.11429196 | 0.11429196 | 2.261997532 | 0.14303064 | 0.86610439 | 1 | 1.330894363 | 1.330894363 | 2.834327965 | 0.79907325 | -0.097413408 | FALSE | TRUE | FALSE |
| PTPRA | 0.114271 | 0.85802356 | 1 | 0.114270982 | 0.114270982 | 2.246361968 | 0.14314401 | 0.866037127 | 1 | 1.340976666 | 1.340976666 | 26.41542709 | 0.798293838 | -0.097837223 | FALSE | TRUE | FALSE |
| CD44 | 0.1142384 | 0.8580355 | 1 | 0.114238442 | 0.114238442 | 2.254176175 | 0.1468133 | 0.865349687 | 1 | 0.146813304 | 0.146813304 | 2.901617856 | 0.778120503 | -0.108953141 | FALSE | TRUE | FALSE |
| PIK3CB | 0.1141833 | 0.85804745 | 0.5077 | 1.065588977 | 1.065588977 | 20.90761649 | 0.15223915 | 0.864012623 | 1 | 0.152239153 | 0.152239153 | 5.130965839 | 0.750025524 | -0.124923957 | FALSE | TRUE | FALSE |
| GFPT1 | 0.1141291 | 0.85805939 | 0.0216 | 0.114129057 | 0.114129057 | 2.250296929 | 0.14738312 | 0.865225041 | 0.01635 | 0.147383117 | 0.147383117 | 2.910917908 | 0.774369948 | -0.11105151 | FALSE | TRUE | FALSE |
| FGD3 | 0.1141164 | 0.85807133 | 0.0244 | 0.114116439 | 0.114116439 | 2.242726386 | 5.70491386 | 0.276961688 | 0.5075 | 5.704913859 | 128.4001133 | 513.4856326 | 0.020003184 | -1.69890088 | FALSE | FALSE | FALSE |
| TSSK4 | 0.1140979 | 0.85808327 | 1 | 0.114097853 | 0.114097853 | 0.114097853 | 4.97017008 | 0.307378101 | 0.00245 | 0.146531626 | 0.146531626 | 0.146531626 | 0.022956529 | -1.639093777 | FALSE | FALSE | FALSE |
| ACTL6B | 0.1140365 | 0.85809521 | 0.7507 | 0.261642915 | 1.045870258 | 1.045870258 | 0.04519234 | 0.88779552 | 0.0128 | 0.045192344 | 0.045192344 | 0.895657572 | 2.523358057 | 0.40197888 | FALSE | TRUE | FALSE |
| CCAR1 | 0.1139654 | 0.85810714 | 1 | 0.113965357 | 0.113965357 | 3.83118437 | 0.14722686 | 0.865259057 | 0.02725 | 0.147226858 | 0.147226858 | 2.889217225 | 0.774079938 | -0.111214188 | FALSE | TRUE | FALSE |
| RNF17 | 0.1139246 | 0.85811908 | 1 | 0.113924636 | 0.113924636 | 3.827711026 | 0.14695986 | 0.86532704 | 1 | 0.146959858 | 0.146959858 | 2.870316396 | 0.775209216 | -0.110581073 | FALSE | TRUE | FALSE |
| CHGB | 0.1139072 | 0.85813101 | 0.5116 | 1.029058582 | 1.029058582 | 20.32183478 | 0.15091733 | 0.864426691 | 1 | 0.150917327 | 0.150917327 | 2.98483205 | 0.754765772 | -0.122187803 | FALSE | TRUE | FALSE |
| DDX60L | 0.1138717 | 0.85814294 | 0.34455 | 0.113871661 | 8.684180665 | 19.11161679 | 17.7727996 | 0.144773007 | 0.3532 | 1.981200694 | 163.3399961 | 1537.064996 | 0.006407075 | -2.193340189 | FALSE | FALSE | FALSE |
| CLEC14A | 0.1138715 | 0.85815487 | 1 | 0.113871533 | 0.113871533 | 2.251776599 | 0.14672679 | 0.865361008 | 0.01325 | 0.146726792 | 0.146726792 | 2.905411907 | 0.776078669 | -0.110094253 | FALSE | TRUE | FALSE |
| TGFBR3 | 0.1138443 | 0.85816679 | 1 | 0.113844334 | 0.113844334 | 2.24065711 | 0.14702127 | 0.865304387 | 1 | 0.147021266 | 0.147021266 | 2.899180019 | 0.774339236 | -0.111068735 | FALSE | TRUE | FALSE |
| PRSS16 | 0.1137203 | 0.85817871 | 1 | 1.028340933 | 1.028340933 | 3.849834816 | 0.14622946 | 0.865440208 | 1 | 1.329722575 | 1.329722575 | 2.891468027 | 0.777683603 | -0.109197058 | FALSE | TRUE | FALSE |
| UGT2A3 | 0.1135649 | 0.85819064 | 0.7556 | 0.113564927 | 1.022806763 | 2.251731441 | 143.219816 | 0.01976891 | 0.06605 | 1.613964848 | 143.2198156 | 143.2198156 | 0.000792941 | -3.100758884 | FALSE | FALSE | FALSE |
| USP19 | 0.1135374 | 0.85820256 | 1 | 1.044280881 | 9.858045867 | 191.1595141 | 1.39596619 | 0.545570621 | 0.51405 | 1.395966191 | 1.395966191 | 27.16884337 | 0.081332452 | -1.089736136 | FALSE | FALSE | FALSE |
| C6orf15 | 0.1134755 | 0.85821448 | 1 | 1.018132439 | 1.018132439 | 1.018132439 | 0.15009533 | 0.864541289 | 1 | 0.15009533 | 0.15009533 | 0.15009533 | 0.756022814 | -0.121465099 | FALSE | TRUE | FALSE |
| NT5DC2 | 0.113449 | 0.85822639 | 0.01915 | 0.113448975 | 0.113448975 | 2.234319918 | 1.29755342 | 0.5550948 | 0.50765 | 1.297553416 | 1.297553416 | 4.803918338 | 0.08743299 | -1.058324668 | FALSE | FALSE | FALSE |
| ZBTB25 | 0.1134408 | 0.85823831 | 1 | 0.113440814 | 0.113440814 | 2.251288285 | 1.28984668 | 0.555989445 | 0.51195 | 1.289846675 | 1.289846675 | 1.289846675 | 0.087949069 | -1.055768755 | FALSE | FALSE | FALSE |
| ZFYVE19 | 0.1134179 | 0.85825022 | 0.50635 | 0.988632482 | 8.839476948 | 8.839476948 | 0.78145715 | 0.627595692 | 0.25785 | 0.195371142 | 0.781457155 | 1.797008218 | 0.145136437 | -0.838223541 | FALSE | TRUE | FALSE |
| ZC3H14 | 0.1134145 | 0.85826213 | 1 | 0.113414508 | 0.113414508 | 2.236945336 | 0.14623528 | 0.865428899 | 1 | 0.146235277 | 0.146235277 | 2.88908648 | 0.775561894 | -0.110383537 | FALSE | TRUE | FALSE |
| SLC24A2 | 0.1133934 | 0.85827404 | 1 | 1.059296658 | 1.059296658 | 3.836798577 | 0.14963026 | 0.864632838 | 1 | 0.598310043 | 0.598310043 | 2.959119793 | 0.757824301 | -0.120431473 | FALSE | TRUE | FALSE |
| RB1 | 0.113301 | 0.85828595 | 0.51135 | 0.11330099 | 0.11330099 | 2.232672365 | 0.04374028 | 0.888150811 | 0.0194 | 0.043740283 | 0.043740283 | 0.863458421 | 2.59031223 | 0.413352116 | FALSE | TRUE | FALSE |
| PSIP1 | 0.1132789 | 0.85829785 | 0.8757 | 9.463667929 | 89.88412698 | 89.88412698 | 0.15950238 | 0.862201745 | 1 | 0.159502385 | 0.159502385 | 3.165099524 | 0.710202028 | -0.148618092 | FALSE | TRUE | FALSE |
| ZNF383 | 0.113208 | 0.85830975 | 1 | 1.021246696 | 1.021246696 | 2.246360944 | 0.15002183 | 0.864564188 | 1 | 0.150021834 | 0.150021834 | 0.150021834 | 0.754610442 | -0.122277189 | FALSE | TRUE | FALSE |
| PARVB | 0.113181 | 0.85832166 | 0.2544 | 0.050904015 | 0.452452977 | 0.452452977 | 2.99110347 | 0.412419714 | 0.1311 | 0.334008411 | 2.99110347 | 2.99110347 | 0.03783921 | -1.422057934 | FALSE | FALSE | FALSE |
| CD46 | 0.113151 | 0.85833355 | 0.01145 | 0.113150982 | 0.113150982 | 2.245826298 | 0.14554951 | 0.865519329 | 0.0097 | 0.145549506 | 0.145549506 | 0.145549506 | 0.777405466 | -0.10935241 | FALSE | TRUE | FALSE |
| UBAP2L | 0.1131038 | 0.85834545 | 1 | 1.064823136 | 10.28880887 | 35.89365047 | 0.14079037 | 0.866662406 | 1 | 0.14079037 | 0.14079037 | 2.761786134 | 0.803348726 | -0.09509589 | FALSE | TRUE | FALSE |
| NBR1 | 0.1130912 | 0.85835735 | 1 | 0.113091181 | 0.113091181 | 2.226668271 | 4.77622592 | 0.316183518 | 0.01245 | 1.292834956 | 1.292834956 | 4.776225922 | 0.023677938 | -1.625656123 | FALSE | FALSE | FALSE |
| ALX3 | 0.1130587 | 0.85836924 | 0.00925 | 0.113058653 | 0.113058653 | 0.113058653 | 0.13742876 | 0.867293354 | 1 | 1.294251505 | 12.50962156 | 12.50962156 | 0.822670984 | -0.08477382 | FALSE | TRUE | FALSE |
| PBX4 | 0.1130086 | 0.85838113 | 1 | 0.113008552 | 0.113008552 | 2.241903871 | 1.27162358 | 0.558274948 | 0.7469 | 1.271623579 | 12.09037975 | 12.09037975 | 0.0888695 | -1.051247262 | FALSE | FALSE | FALSE |
| SLC5A6 | 0.1129497 | 0.85839302 | 0.5049 | 0.112949681 | 0.112949681 | 0.956211714 | 0.1701289 | 0.859661755 | 0.7562 | 0.170128896 | 0.680238213 | 3.360923229 | 0.663906506 | -0.177893075 | FALSE | TRUE | FALSE |
| KRTAP4-1 | 0.112802 | 0.85840491 | 1 | 0.112802001 | 0.112802001 | 0.112802001 | 0.14473509 | 0.865711125 | 1 | 0.144735095 | 0.144735095 | 0.144735095 | 0.779368686 | -0.108257047 | FALSE | TRUE | FALSE |
| GALNT18 | 0.1127643 | 0.8584168 | 0.0209 | 0.11276434 | 0.11276434 | 2.220541691 | 0.14592507 | 0.865462822 | 1 | 0.14592507 | 0.14592507 | 2.878867421 | 0.772755084 | -0.111958129 | FALSE | TRUE | FALSE |
| AKR1D1 | 0.1127006 | 0.85842868 | 1 | 1.02698348 | 1.02698348 | 2.239876728 | 0.14979519 | 0.864609962 | 1 | 0.149795192 | 0.149795192 | 0.149795192 | 0.7523643 | -0.12357182 | FALSE | TRUE | FALSE |
| TMEM223 | 0.1126715 | 0.85844056 | 0.00455 | 0.112671475 | 0.112671475 | 0.112671475 | 1.10561477 | 0.577771506 | 0.75145 | 1.105614771 | 10.51097141 | 10.51097141 | 0.101908439 | -0.991789852 | FALSE | FALSE | FALSE |
| ITGA11 | 0.1126712 | 0.85845244 | 0.6986 | 0.255080069 | 4.077568393 | 8.775471029 | 0.20770788 | 0.850260603 | 0.5077 | 0.20770788 | 0.830638556 | 4.058432842 | 0.542450138 | -0.265640176 | FALSE | TRUE | FALSE |
| NRIP1 | 0.1126245 | 0.85846432 | 1 | 1.024666855 | 1.024666855 | 20.114028 | 0.1451938 | 0.86560965 | 0.52115 | 1.327484315 | 1.327484315 | 26.11635691 | 0.775684199 | -0.110315055 | FALSE | TRUE | FALSE |
| SPATA17 | 0.1126024 | 0.8584762 | 0.75825 | 1.019324684 | 4.076959248 | 4.076959248 | 0.15113247 | 0.86434637 | 0.00915 | 0.151132466 | 0.151132466 | 0.151132466 | 0.745057515 | -0.127810201 | FALSE | TRUE | FALSE |
| SQSTM1 | 0.112514 | 0.85848807 | 0.51225 | 0.112513996 | 0.449601939 | 0.449601939 | 0.18590537 | 0.855517018 | 0.7504 | 0.185905374 | 1.761052761 | 1.761052761 | 0.605221856 | -0.218085397 | FALSE | TRUE | FALSE |
| GINS2 | 0.1124943 | 0.85849995 | 1 | 0.112494271 | 1.020337199 | 1.020337199 | 0.14770065 | 0.865151998 | 1 | 0.590719634 | 0.590719634 | 0.590719634 | 0.761636944 | -0.118251998 | FALSE | TRUE | FALSE |
| CTSE | 0.1124926 | 0.85851182 | 0.7532 | 1.010949719 | 9.324798455 | 9.324798455 | 0.14902511 | 0.864826989 | 0.504 | 1.348876439 | 1.348876439 | 2.959513163 | 0.754856393 | -0.122135662 | FALSE | TRUE | FALSE |
| ULK1 | 0.1124157 | 0.85852369 | 1 | 0.112415739 | 0.112415739 | 2.17971549 | 0.14528465 | 0.865575793 | 1 | 0.145284649 | 0.145284649 | 2.827400766 | 0.773761992 | -0.111392607 | FALSE | TRUE | FALSE |
| CHL1 | 0.1124075 | 0.85853556 | 0.27315 | 0.028118161 | 0.112407531 | 0.948060553 | 0.04932537 | 0.88683241 | 0.7611 | 0.11192555 | 0.447392556 | 2.19610841 | 2.278898683 | 0.357725017 | FALSE | TRUE | FALSE |
| AAGAB | 0.1123797 | 0.85854742 | 0.0082 | 0.112379695 | 0.112379695 | 0.112379695 | 1.28295781 | 0.556753693 | 0.50505 | 1.282957814 | 1.282957814 | 1.282957814 | 0.087594224 | -1.057524528 | FALSE | FALSE | FALSE |
| TMEM50A | 0.1123365 | 0.85855928 | 0.0042 | 0.112336504 | 0.112336504 | 0.112336504 | 0.1218013 | 0.870839458 | 0.00435 | 0.121801298 | 0.121801298 | 0.121801298 | 0.922293156 | -0.035131014 | FALSE | TRUE | FALSE |
| PDK1 | 0.1123269 | 0.85857115 | 1 | 0.112326905 | 0.112326905 | 2.227636929 | 1.29589709 | 0.555478637 | 0.50095 | 1.295897093 | 1.295897093 | 2.793037117 | 0.086678877 | -1.062086722 | FALSE | FALSE | FALSE |
| HSPA4L | 0.1122309 | 0.85858301 | 0.02195 | 0.112230866 | 0.112230866 | 2.212549035 | 0.14462966 | 0.865733655 | 1 | 0.144629661 | 0.144629661 | 2.856169991 | 0.775987899 | -0.110145051 | FALSE | TRUE | FALSE |
| NLGN1 | 0.1122001 | 0.85859486 | 1 | 1.049279489 | 1.049279489 | 3.791950192 | 0.14083352 | 0.866629049 | 1 | 1.318232644 | 12.66382464 | 26.02043232 | 0.796686087 | -0.098712767 | FALSE | TRUE | FALSE |
| SSUH2 | 0.1121653 | 0.85860672 | 0.5099 | 1.011934463 | 1.011934463 | 1.011934463 | 1.31014963 | 0.553423785 | 0.50085 | 1.310149633 | 1.310149633 | 1.310149633 | 0.085612587 | -1.067462381 | FALSE | FALSE | FALSE |
| POLR2M | 0.1121292 | 0.85861857 | 0.01165 | 0.112129205 | 0.112129205 | 2.22473814 | 0.11624371 | 0.872136827 | 0.00955 | 0.116243711 | 0.116243711 | 0.116243711 | 0.964604489 | -0.015650721 | FALSE | TRUE | FALSE |
| SLC46A2 | 0.1119528 | 0.85863043 | 1 | 0.111952816 | 0.111952816 | 2.212463742 | 0.14295103 | 0.866149194 | 1 | 0.571710041 | 0.571710041 | 2.829101766 | 0.783155024 | -0.106152262 | FALSE | TRUE | FALSE |
| PTPRF | 0.1119171 | 0.85864228 | 0.064 | 0.027996741 | 0.234892499 | 2.124566639 | 0.01246433 | 0.895929819 | 1 | 0.115680054 | 0.238069056 | 2.209493033 | 8.978995969 | 0.953227777 | FALSE | TRUE | FALSE |
| VPS33A | 0.1118102 | 0.85865413 | 0.01945 | 0.111810187 | 0.111810187 | 2.205633177 | 0.14483145 | 0.865666043 | 1 | 0.144831454 | 0.144831454 | 2.86173031 | 0.772002101 | -0.112381518 | FALSE | TRUE | FALSE |
| ADAM9 | 0.1118009 | 0.85866597 | 1 | 0.111800855 | 0.111800855 | 2.202163107 | 0.14445394 | 0.865789949 | 0.01805 | 0.144453937 | 0.144453937 | 2.850522353 | 0.77395506 | -0.111284256 | FALSE | TRUE | FALSE |
| SLFN5 | 0.1117049 | 0.85867782 | 0.8748 | 0.993588592 | 9.06509893 | 19.54359936 | 0.02180615 | 0.893283736 | 1 | 0.191079587 | 1.718538555 | 3.765952168 | 5.122633682 | 0.709493301 | FALSE | TRUE | FALSE |
| SNX19 | 0.1117002 | 0.85868966 | 0.87665 | 0.98464442 | 8.903067211 | 19.29413916 | 0.02253433 | 0.893061383 | 0.51455 | 0.196043303 | 0.196043303 | 3.850842925 | 4.956890515 | 0.695209327 | FALSE | TRUE | FALSE |
| LHX4 | 0.1116953 | 0.8587015 | 0.0135 | 0.111695322 | 0.111695322 | 0.553815908 | 0.04268886 | 0.888407937 | 0.50415 | 0.170612087 | 0.170612087 | 0.846791433 | 2.616498357 | 0.417720466 | FALSE | TRUE | FALSE |
| MMADHC | 0.1116666 | 0.85871334 | 0.5097 | 0.446426743 | 1.785194325 | 1.785194325 | 0.68052443 | 0.674914414 | 0.5116 | 0.680524432 | 2.721568717 | 2.721568717 | 0.164089096 | -0.784920278 | FALSE | TRUE | FALSE |
| SENP6 | 0.111639 | 0.85872518 | 1 | 0.111639028 | 0.111639028 | 2.190256487 | 0.13609818 | 0.867590294 | 1 | 0.136098179 | 0.136098179 | 2.676246618 | 0.82028304 | -0.086036268 | FALSE | TRUE | FALSE |
| CCDC102A | 0.1116254 | 0.85873702 | 1 | 0.111625426 | 0.111625426 | 2.201139888 | 0.14465507 | 0.865722391 | 1 | 0.144655072 | 0.144655072 | 2.857274377 | 0.771666174 | -0.112570537 | FALSE | TRUE | FALSE |
| NAPA | 0.1115332 | 0.85874885 | 1 | 0.445935167 | 1.783396605 | 1.783396605 | 0.14775382 | 0.865134256 | 1 | 0.14775382 | 0.14775382 | 0.14775382 | 0.754858156 | -0.122134648 | FALSE | TRUE | FALSE |
| ZNF585A | 0.1114138 | 0.85876068 | 1 | 1.015942813 | 1.015942813 | 3.766181986 | 0.14834084 | 0.864975097 | 1 | 0.148340844 | 0.148340844 | 2.934275656 | 0.751065913 | -0.124321948 | FALSE | TRUE | FALSE |
| RBM26 | 0.1114006 | 0.85877251 | 0.0279 | 0.11140063 | 0.11140063 | 2.189100011 | 0.14443746 | 0.865801202 | 1 | 0.144437465 | 0.144437465 | 2.844265085 | 0.771272398 | -0.112792211 | FALSE | TRUE | FALSE |
| AFF4 | 0.1113837 | 0.85878434 | 0.876 | 4.172045995 | 40.1094911 | 82.22392074 | 0.15550615 | 0.863142115 | 1 | 0.155506154 | 0.155506154 | 3.050725753 | 0.716265273 | -0.144926104 | FALSE | TRUE | FALSE |
| XCL1 | 0.111256 | 0.85879617 | 1 | 0.111256039 | 0.111256039 | 0.111256039 | 0.11865899 | 0.871548266 | 1 | 0.118658986 | 0.118658986 | 0.118658986 | 0.937611575 | -0.02797704 | FALSE | TRUE | FALSE |
| TTC4 | 0.1112339 | 0.85880799 | 1 | 0.111233865 | 0.111233865 | 0.111233865 | 0.14337883 | 0.865969797 | 0.00925 | 0.143378825 | 0.143378825 | 0.143378825 | 0.77580399 | -0.110247991 | FALSE | TRUE | FALSE |
| PLCB1 | 0.1111758 | 0.85881982 | 0.02995 | 0.11117581 | 0.11117581 | 2.173416344 | 0.14383685 | 0.865902406 | 1 | 0.14383685 | 0.14383685 | 2.81955163 | 0.772929955 | -0.111859861 | FALSE | TRUE | FALSE |
| CYP4F22 | 0.1111729 | 0.85883164 | 0.50855 | 0.111172919 | 0.111172919 | 0.54831148 | 0.04190701 | 0.888552101 | 0.7569 | 0.167530852 | 0.66990224 | 3.308587444 | 2.652847491 | 0.423712284 | FALSE | TRUE | FALSE |
| ASB14 | 0.1111456 | 0.85884346 | 0.75165 | 0.444367848 | 4.006584822 | 4.006584822 | 0.18201557 | 0.856533638 | 0.87245 | 1.654129114 | 15.41881554 | 15.41881554 | 0.610637629 | -0.214216437 | FALSE | TRUE | FALSE |
| ADH1A | 0.1111369 | 0.85885527 | 1 | 0.111136897 | 0.111136897 | 0.111136897 | 0.14321198 | 0.86602591 | 0.00825 | 0.14321198 | 0.14321198 | 0.14321198 | 0.776030726 | -0.110121083 | FALSE | TRUE | FALSE |
| SMG9 | 0.1110461 | 0.85886709 | 1 | 0.111046094 | 0.111046094 | 2.194936792 | 0.14313202 | 0.866048342 | 0.0141 | 0.143132024 | 0.143132024 | 2.833129761 | 0.775829828 | -0.110233528 | FALSE | TRUE | FALSE |

| | | | | | | | | | | | | | | | | | |
|---|---|---|---|---|---|---|---|---|---|---|---|---|---|---|---|---|---|
| TNRC6A | 0.1110055 | 0.8588789 | 0.7611 | 1.026435847 | 9.741188571 | 187.8231184 | 0.13098958 | 0.868701079 | 1 | 112.0218569 | 2169.296183 | 11432.96005 | 0.847437413 | -0.071892366 | FALSE | TRUE | FALSE |
| ENTPD5 | 0.1109961 | 0.85889071 | 1 | 0.110996073 | 0.110996073 | 2.203710879 | 0.14310095 | 0.866081977 | 1 | 0.143100951 | 0.143100951 | 0.143100951 | 0.775648742 | -0.110334908 | FALSE | TRUE | FALSE |
| FBLN1 | 0.1109542 | 0.85890252 | 0.32915 | 0.110954191 | 1.773635657 | 3.751793427 | 0.05157917 | 0.88628713 | 0.02085 | 0.051579175 | 0.051579175 | 1.014868214 | 2.151143203 | 0.332669323 | FALSE | TRUE | FALSE |
| BOD1L1 | 0.1109476 | 0.85891433 | 0.7674 | 0.11094756 | 2.112525911 | 19.53515902 | 0.15640571 | 0.862861274 | 0.7111 | 1.452454233 | 135.0737849 | 264.9475361 | 0.709357467 | -0.149134855 | FALSE | TRUE | FALSE |
| B3GAT3 | 0.1109301 | 0.85892614 | 0.0112 | 0.110930095 | 0.110930095 | 2.198960677 | 0.56138872 | 0.728458119 | 0.7536 | 0.561388723 | 2.245521342 | 2.250924068 | 0.197599436 | -0.704214298 | FALSE | TRUE | FALSE |
| DDX24 | 0.1108772 | 0.85893794 | 1 | 0.110877172 | 0.110877172 | 2.180801505 | 0.14331005 | 0.865981023 | 1 | 0.143310051 | 0.143310051 | 2.824333873 | 0.773687342 | -0.111434509 | FALSE | TRUE | FALSE |
| IFNAR1 | 0.1108681 | 0.85894974 | 1 | 0.110868124 | 0.110868124 | 2.195177217 | 0.14309771 | 0.866093184 | 0.0127 | 0.143097707 | 0.143097707 | 2.836729544 | 0.774772191 | -0.110825976 | FALSE | TRUE | FALSE |
| GZMH | 0.110834 | 0.85896154 | 0.6851 | 0.443230832 | 7.093646518 | 7.093646518 | 0.17196119 | 0.859145443 | 0.74875 | 0.171961195 | 0.687726307 | 0.687726307 | 0.644529063 | -0.190757495 | FALSE | TRUE | FALSE |
| RINL | 0.1108212 | 0.85897334 | 0.50175 | 1.009600027 | 1.009600027 | 2.193867595 | 0.14013739 | 0.866773492 | 1 | 0.14013739 | 0.14013739 | 2.777621505 | 0.790803653 | -0.101931333 | FALSE | TRUE | FALSE |
| GABBR1 | 0.1107454 | 0.85898514 | 1 | 0.110745442 | 0.110745442 | 2.167362001 | 0.56833261 | 0.725377718 | 0.51805 | 0.568332607 | 0.568332607 | 11.15140803 | 0.194860265 | -0.710276712 | FALSE | TRUE | FALSE |
| HOXA6 | 0.1106806 | 0.85899693 | 0.2626 | 0.110680559 | 0.442501144 | 0.549578202 | 0.04270809 | 0.888375856 | 1 | 0.170720089 | 0.170720089 | 0.170720089 | 2.591559557 | 0.413561194 | FALSE | TRUE | FALSE |
| BLZF1 | 0.1106697 | 0.85900872 | 1 | 1.001191299 | 1.001191299 | 2.196516232 | 0.14713368 | 0.865281725 | 1 | 0.147133682 | 0.147133682 | 0.147133682 | 0.752171197 | -0.123683301 | FALSE | TRUE | FALSE |
| PDSS1 | 0.1106141 | 0.85902051 | 0.967 | 90.80621652 | 894.1268817 | 9019.140909 | 0.16658441 | 0.860574731 | 1 | 0.166584412 | 0.166584412 | 0.166584412 | 0.664012563 | -0.177823704 | FALSE | TRUE | FALSE |
| GORASP1 | 0.1106005 | 0.8590323 | 1 | 0.110600545 | 0.110600545 | 2.193563651 | 1.26677906 | 0.559659962 | 0.5016 | 1.266779058 | 1.266779058 | 2.751469495 | 0.087308473 | -1.058943609 | FALSE | FALSE | FALSE |
| LRRIQ4 | 0.1105865 | 0.85904409 | 1 | 0.11058652 | 0.11058652 | 2.186734146 | 0.14287518 | 0.866160391 | 1 | 0.142875178 | 0.142875178 | 2.829050376 | 0.774007923 | -0.111254594 | FALSE | TRUE | FALSE |
| ETFB | 0.1104731 | 0.85905587 | 0.7531 | 0.110473074 | 0.441695794 | 0.548377729 | 1.50098387 | 0.535672911 | 0.37555 | 0.653094028 | 6.001757144 | 6.001757144 | 0.07360044 | -1.133119588 | FALSE | FALSE | FALSE |
| ARRDC1 | 0.1104274 | 0.85906766 | 1 | 0.110427442 | 0.110427442 | 2.186576363 | 0.14223639 | 0.866383937 | 1 | 0.142236386 | 0.142236386 | 2.819796496 | 0.776365636 | -0.109933696 | FALSE | TRUE | FALSE |
| ZNF789 | 0.1104117 | 0.85907944 | 0.01115 | 0.110411682 | 0.110411682 | 2.190889324 | 0.13812427 | 0.867127852 | 1 | 1.251067311 | 1.251067311 | 2.743376622 | 0.799364811 | -0.097254974 | FALSE | TRUE | FALSE |
| BMP2K | 0.110395 | 0.85909122 | 1 | 0.110395019 | 0.110395019 | 2.165187158 | 0.1429534 | 0.866137996 | 1 | 0.142953404 | 0.142953404 | 2.810289473 | 0.772244776 | -0.112245021 | FALSE | TRUE | FALSE |
| TTLL3 | 0.1103637 | 0.859103 | 0.5056 | 1.000555884 | 1.000555884 | 19.65264463 | 1.21878753 | 0.565486093 | 0.87555 | 11.38088839 | 109.0023514 | 224.0224409 | 0.090552057 | -1.043101678 | FALSE | FALSE | FALSE |
| ACSBG1 | 0.1103473 | 0.85911477 | 0.02185 | 0.11034734 | 0.11034734 | 2.1673614 | 0.14273328 | 0.866249896 | 1 | 0.14273328 | 0.14273328 | 2.809516752 | 0.77310169 | -0.111763377 | FALSE | TRUE | FALSE |
| AMN1 | 0.1102821 | 0.85912654 | 1 | 0.11028211 | 0.11028211 | 0.11028211 | 0.11660001 | 0.87207511 | 1 | 0.116600007 | 0.116600007 | 0.116600007 | 0.945815636 | -0.024193511 | FALSE | TRUE | FALSE |
| THUMPD2 | 0.1102429 | 0.85913832 | 1 | 0.110242921 | 0.110242921 | 2.188820658 | 0.56238285 | 0.728035974 | 0.5068 | 0.562382853 | 0.562382853 | 2.794049869 | 0.19602853 | -0.707681359 | FALSE | TRUE | FALSE |
| NOP56 | 0.1101654 | 0.85915009 | 0.75475 | 0.110165432 | 0.440481869 | 2.174851746 | 10108.0573 | 0.000192287 | 0.0125 | 39.32490585 | 2523.510463 | 10108.05729 | 1.08988E-05 | -4.962622352 | FALSE | FALSE | FALSE |
| SLC18A3 | 0.1101486 | 0.85916185 | 1 | 0.110148605 | 0.110148605 | 2.160660174 | 0.14296461 | 0.866126796 | 0.0204 | 0.142964607 | 0.142964607 | 2.81086075 | 0.770460654 | -0.113249535 | FALSE | TRUE | FALSE |
| USPL1 | 0.1100815 | 0.85917362 | 0.02495 | 0.110081465 | 0.110081465 | 2.162600854 | 1.25647701 | 0.560536769 | 0.51175 | 1.256477005 | 1.256477005 | 24.73675153 | 0.087611205 | -1.057440345 | FALSE | FALSE | FALSE |
| SORCS3 | 0.1100182 | 0.85918539 | 1 | 0.440044827 | 1.760507884 | 14.83567604 | 0.14121428 | 0.866584546 | 1 | 1.295211321 | 1.295211321 | 25.42385678 | 0.779086719 | -0.108414199 | FALSE | TRUE | FALSE |
| ALPI | 0.1099691 | 0.85919715 | 0.38985 | 0.050029143 | 1.756226962 | 7.020988101 | 0.03418907 | 0.890318522 | 0.8581 | 0.136639809 | 2.184150325 | 8.735702193 | 3.216499777 | 0.507383526 | FALSE | TRUE | FALSE |
| STIM1 | 0.1099539 | 0.85920891 | 0.0193 | 0.109953874 | 0.109953874 | 2.166110466 | 0.14275681 | 0.866238714 | 0.01875 | 0.142756814 | 0.142756814 | 2.817408578 | 0.770218048 | -0.113386309 | FALSE | TRUE | FALSE |
| RBM45 | 0.1099353 | 0.85922067 | 1 | 1.003168753 | 1.003168753 | 3.720394566 | 5.15501005 | 0.299842615 | 0.2581 | 1.288937403 | 5.155010051 | 5.155010051 | 0.021325914 | -1.67109234 | FALSE | FALSE | FALSE |
| C15orf39 | 0.1099314 | 0.85923242 | 1 | 9107.751828 | 1000418.504 | 9261455.197 | 0.1149199 | 0.872393393 | 1 | 11159221.89 | 16502327107 | 2.32349E+12 | 0.95659196 | -0.019273274 | FALSE | TRUE | FALSE |
| SBSPON | 0.1098778 | 0.85924418 | 1 | 0.985478123 | 0.985478123 | 0.985478123 | 0.14155694 | 0.866450871 | 1 | 1.277878905 | 1.277878905 | 1.277878905 | 0.776209108 | -0.110021265 | FALSE | TRUE | FALSE |
| C1QTNF6 | 0.1098565 | 0.85925593 | 0.01045 | 0.109856546 | 0.109856546 | 2.179905363 | 0.14232006 | 0.866339286 | 1 | 0.142320065 | 0.142320065 | 0.142320065 | 0.771897809 | -0.112440192 | FALSE | TRUE | FALSE |
| CCDC61 | 0.1098255 | 0.85926768 | 1 | 0.109825542 | 0.109825542 | 2.169941952 | 0.1416359 | 0.866406255 | 1 | 0.141635896 | 0.141635896 | 2.802526313 | 0.775407541 | -0.11046998 | FALSE | TRUE | FALSE |
| TRIM40 | 0.1098007 | 0.85927943 | 0.0064 | 0.109800681 | 0.109800681 | 0.109800681 | 43.9669478 | 0.062553724 | 0.12885 | 4.750918019 | 43.96694778 | 43.96694778 | 0.002497346 | -2.602521284 | FALSE | FALSE | FALSE |
| TLR2 | 0.109773 | 0.85929118 | 0.81135 | 8.728701178 | 80.80863675 | 766.4273828 | 0.02352192 | 0.892860541 | 1 | 0.204764701 | 0.204764701 | 0.795831094 | 4.66683863 | 0.669022784 | FALSE | TRUE | FALSE |
| A2M | 0.1097186 | 0.85930293 | 0.04055 | 0.109718623 | 0.109718623 | 2.134980624 | 0.02122291 | 0.893483122 | 0.0326 | 0.021222908 | 0.021222908 | 0.414317243 | 5.169820437 | 0.713475459 | FALSE | TRUE | FALSE |
| TM9SF1 | 0.1096354 | 0.85931467 | 1 | 0.109635413 | 0.109635413 | 2.16183981 | 0.14227457 | 0.866350451 | 1 | 0.142274571 | 0.142274571 | 2.810186289 | 0.770590362 | -0.113176427 | FALSE | TRUE | FALSE |
| HMBOX1 | 0.1095824 | 0.85932641 | 0.01215 | 0.109582484 | 0.109582484 | 2.174045874 | 0.14238571 | 0.866316949 | 1 | 0.142385711 | 0.142385711 | 2.827571173 | 0.769616607 | -0.113725569 | FALSE | TRUE | FALSE |
| SLC30A9 | 0.1095467 | 0.85933815 | 0.01705 | 0.109546699 | 0.109546699 | 2.1674492 | 0.13759479 | 0.867249257 | 1 | 1.265262325 | 1.265262325 | 2.725908569 | 0.796154407 | -0.099002696 | FALSE | TRUE | FALSE |
| EOGT | 0.1095456 | 0.85934989 | 1 | 0.994891233 | 0.994891233 | 2.171853137 | 0.14161224 | 0.866428567 | 1 | 1.292703136 | 1.292703136 | 1.292703136 | 0.773560165 | -0.111505903 | FALSE | TRUE | FALSE |
| STXBP3 | 0.1093368 | 0.85936163 | 0.01385 | 0.109336836 | 0.109336836 | 2.165040242 | 0.13860681 | 0.86700623 | 1 | 0.554296401 | 2.217216545 | 4.693072859 | 0.788827303 | -0.103018066 | FALSE | TRUE | FALSE |
| MICU1 | 0.1091238 | 0.85937336 | 0.01315 | 0.109123769 | 0.109123769 | 2.16294157 | 1.17746606 | 0.569611916 | 0.87625 | 11.0391821 | 106.1537991 | 106.1537991 | 0.092676784 | -1.033029043 | FALSE | FALSE | FALSE |
| RXFP3 | 0.1091219 | 0.8593851 | 0.01835 | 0.10912189 | 0.10912189 | 2.149192539 | 0.14123814 | 0.866562286 | 1 | 0.141238143 | 0.141238143 | 2.7868334 | 0.772609207 | -0.112040121 | FALSE | TRUE | FALSE |
| TMEM104 | 0.1091122 | 0.85939683 | 1 | 0.109112162 | 0.109112162 | 2.15092044 | 0.14112609 | 0.866595674 | 1 | 0.141126091 | 0.141126091 | 2.786814643 | 0.773153715 | -0.111734153 | FALSE | TRUE | FALSE |
| HSPA1L | 0.1090705 | 0.85940856 | 0.02 | 0.109070513 | 0.109070513 | 2.151694788 | 0.136793 | 0.867436482 | 1 | 1.24479087 | 1.24479087 | 4.632821273 | 0.797339848 | -0.098356531 | FALSE | TRUE | FALSE |
| EPS15 | 0.1088751 | 0.85942029 | 0.02045 | 0.108875114 | 0.108875114 | 2.145877243 | 0.14126401 | 0.866540018 | 1 | 0.141264014 | 0.141264014 | 2.789113991 | 0.770720805 | -0.113102917 | FALSE | TRUE | FALSE |
| FMO4 | 0.1088686 | 0.85943201 | 0.7527 | 0.240994801 | 0.963711018 | 2.156225273 | 12.6560141 | 0.173889363 | 0.32225 | 3.164780216 | 118.6748476 | 118.6748476 | 0.008602128 | -2.065394121 | FALSE | FALSE | FALSE |
| CREB3L2 | 0.1088678 | 0.85944374 | 1 | 0.997866008 | 0.997866008 | 2.157347294 | 1.29853058 | 0.554838509 | 0.51165 | 1.29853058 | 1.29853058 | 2.799040647 | 0.083839262 | -1.076552552 | FALSE | FALSE | FALSE |
| ZNF527 | 0.1088666 | 0.85945546 | 1 | 0.108866601 | 0.108866601 | 2.155138036 | 0.13655378 | 0.867491452 | 0.50705 | 1.242271826 | 1.242271826 | 2.70655172 | 0.797243431 | -0.098409051 | FALSE | TRUE | FALSE |
| FA2H | 0.1088403 | 0.85946718 | 1 | 0.108840335 | 0.108840335 | 2.161170826 | 0.14144133 | 0.866462021 | 1 | 0.141441327 | 0.141441327 | 0.141441327 | 0.769508721 | -0.113786454 | FALSE | TRUE | FALSE |
| AGTPBP1 | 0.1087509 | 0.8594789 | 0.0332 | 0.108750926 | 0.108750926 | 3.661670715 | 1.24938392 | 0.561036577 | 0.52025 | 1.249383925 | 1.249383925 | 24.53158162 | 0.087043641 | -1.060262949 | FALSE | FALSE | FALSE |
| SNX25 | 0.1087112 | 0.85949062 | 0.0198 | 0.108711194 | 0.108711194 | 2.14228007 | 0.13637656 | 0.867524415 | 1 | 1.239697635 | 1.239697635 | 24.47287751 | 0.797139861 | -0.098465474 | FALSE | TRUE | FALSE |
| CDH6 | 0.1085979 | 0.85950234 | 1 | 0.108597947 | 0.108597947 | 2.13708349 | 2.78521708 | 0.428251528 | 0.0057 | 0.141260434 | 0.141260434 | 2.785217084 | 0.038990838 | -1.409037434 | FALSE | FALSE | FALSE |
| DNAJA3 | 0.1085007 | 0.85951405 | 1 | 0.108500732 | 0.108500732 | 2.145690398 | 1.26840836 | 0.559157378 | 0.5054 | 1.268408363 | 1.268408363 | 2.705441832 | 0.085540852 | -1.067826427 | FALSE | FALSE | FALSE |
| CHRNB4 | 0.1083906 | 0.85952576 | 0.93805 | 0.433320782 | 6.93059706 | 8.558975858 | 0.16783249 | 0.860210916 | 0.87405 | 0.671022019 | 2.683527457 | 3.319981506 | 0.645826271 | -0.189884293 | FALSE | TRUE | FALSE |
| PSMC4 | 0.1083718 | 0.85953747 | 1 | 0.108371791 | 0.108371791 | 2.144696775 | 1.28005902 | 0.557134935 | 0.5087 | 1.28005902 | 1.28005902 | 2.706577135 | 0.084661558 | -1.072313744 | FALSE | FALSE | FALSE |
| ZNF627 | 0.1082642 | 0.85954918 | 1 | 9.106693522 | 86.78903409 | 86.78903409 | 0.1534089 | 0.863700462 | 1 | 0.153408898 | 0.153408898 | 3.043733563 | 0.705723253 | -0.151365573 | FALSE | TRUE | FALSE |
| SMYD3 | 0.1082427 | 0.85956089 | 1 | 0.108242695 | 0.108242695 | 2.148663229 | 0.14018648 | 0.866751289 | 1 | 0.140186479 | 0.140186479 | 0.140186479 | 0.772133628 | -0.112307533 | FALSE | TRUE | FALSE |
| GALNT9 | 0.108205 | 0.85957259 | 0.0106 | 0.108205009 | 0.108205009 | 0.243674424 | 0.02078202 | 0.89363772 | 0.50475 | 0.083089128 | 0.083089128 | 0.41298841 | 5.206665384 | 0.716559668 | FALSE | TRUE | FALSE |
| GCC1 | 0.108143 | 0.8595843 | 1 | 0.108142979 | 0.108142979 | 2.127126582 | 0.13986897 | 0.86681788 | 1 | 0.139868972 | 0.139868972 | 2.756635143 | 0.773173477 | -0.111723053 | FALSE | TRUE | FALSE |
| PSMB8 | 0.1081213 | 0.859596 | 1 | 0.108121274 | 0.108121274 | 0.108121274 | 0.13985276 | 0.866828972 | 1 | 0.139852761 | 0.139852761 | 0.139852761 | 0.773107897 | -0.111759891 | FALSE | TRUE | FALSE |
| ITPRIP | 0.108084 | 0.8596077 | 1 | 0.976232569 | 0.976232569 | 3.660340189 | 1.26976476 | 0.558905642 | 0.50545 | 1.26976476 | 1.26976476 | 2.758923686 | 0.085121254 | -1.069961986 | FALSE | FALSE | FALSE |
| PFDN6 | 0.1080786 | 0.8596194 | 0.00365 | 0.108078559 | 0.108078559 | 0.108078559 | 0.10954372 | 0.873481043 | 0.00365 | 0.109543719 | 0.109543719 | 0.109543719 | 0.986624888 | -0.005847933 | FALSE | TRUE | FALSE |
| GPR84 | 0.1079651 | 0.85963109 | 1 | 0.431815505 | 0.431815505 | 2.141405288 | 0.1413104 | 0.866506602 | 1 | 0.141310401 | 0.141310401 | 2.805576786 | 0.764028231 | -0.116890594 | FALSE | TRUE | FALSE |
| ESRP1 | 0.1078815 | 0.85964279 | 1 | 0.107881524 | 0.107881524 | 2.129322421 | 0.13970682 | 0.866851151 | 1 | 0.139706825 | 0.139706825 | 2.761830951 | 0.772199383 | -0.11227055 | FALSE | TRUE | FALSE |
| PAIP1 | 0.1078525 | 0.85965448 | 1 | 0.107852498 | 0.107852498 | 2.14257144 | 0.11520251 | 0.872321647 | 1 | 0.115202507 | 0.115202507 | 0.115202507 | 0.936199227 | -0.028631722 | FALSE | TRUE | FALSE |
| SLC52A3 | 0.1078499 | 0.85966617 | 0.01375 | 0.107849897 | 0.107849897 | 2.133775555 | 0.14034629 | 0.866740184 | 0.0122 | 0.140346295 | 0.140346295 | 2.780302626 | 0.768455604 | -0.114381218 | FALSE | TRUE | FALSE |
| YES1 | 0.10779 | 0.85967786 | 1 | 0.431045271 | 0.431045271 | 3.646849722 | 0.139976 | 0.866784592 | 1 | 0.55977246 | 0.55977246 | 2.776359706 | 0.770060762 | -0.113475005 | FALSE | TRUE | FALSE |
| GLYCTK | 0.1077778 | 0.85968955 | 0.26165 | 0.107777831 | 0.431068376 | 2.128972462 | 0.04064883 | 0.888807592 | 0.5094 | 0.364451972 | 0.364451972 | 0.804141799 | 2.651437464 | 0.423481388 | FALSE | TRUE | FALSE |
| ANKRD44 | 0.1077612 | 0.85970123 | 0.7572 | 0.992674822 | 15.87373497 | 19.49238367 | 0.17203047 | 0.859083735 | 0.7561 | 0.172030473 | 0.687895687 | 3.385426788 | 0.626407417 | -0.203143109 | FALSE | TRUE | FALSE |
| JAK1 | 0.1076553 | 0.85971292 | 1 | 0.107655285 | 0.107655285 | 2.101154213 | 1.27339243 | 0.558022125 | 0.5158 | 1.273392429 | 1.273392429 | 24.9256017 | 0.084542112 | -1.072926908 | FALSE | FALSE | FALSE |
| ZSCAN31 | 0.1075765 | 0.8597246 | 1 | 9.089843413 | 86.82419509 | 86.82419509 | 1.27090692 | 0.558527463 | 0.8717 | 11.89714213 | 114.2305376 | 114.2305376 | 0.08464548 | -1.07239623 | FALSE | FALSE | FALSE |

| | | | | | | | | | | | | | | | | | |
|---|---|---|---|---|---|---|---|---|---|---|---|---|---|---|---|---|---|
| COG5 | 0.107538 | 0.85973628 | 0.5041 | 0.107537953 | 0.107537953 | 2.117084325 | 0.02117761 | 0.893505233 | 0.0205 | 0.021177607 | 0.021177607 | 0.417706272 | 5.077908725 | 0.70568489 | FALSE | TRUE | FALSE |
| KIRREL3 | 0.1074713 | 0.85974796 | 0.7533 | 1.00743354 | 9.692307019 | 19.75992879 | 0.57093072 | 0.724318429 | 0.75095 | 1.341726832 | 5.364975898 | 26.37794453 | 0.188238799 | -0.725290858 | FALSE | TRUE | FALSE |
| AADAC | 0.1074541 | 0.85975963 | 1 | 0.107454097 | 0.107454097 | 2.133716199 | 0.13488326 | 0.867809468 | 1 | 1.223126723 | 1.223126723 | 1.223126723 | 0.796645159 | -0.098735078 | FALSE | TRUE | FALSE |
| BTNL8 | 0.1074009 | 0.85977131 | 0.8751 | 0.966764828 | 8.931761523 | 8.931761523 | 1.33839396 | 0.551482875 | 0.5051 | 1.338393964 | 1.338393964 | 1.338393964 | 0.080246135 | -1.095575874 | FALSE | FALSE | FALSE |
| FAM162A | 0.1073462 | 0.85978298 | 1 | 0.107346236 | 0.107346236 | 0.107346236 | 0.11881685 | 0.871517148 | 0.00315 | 0.118816849 | 0.118816849 | 0.118816849 | 0.903459711 | -0.04409121 | FALSE | TRUE | FALSE |
| CLASP2 | 0.10734 | 0.85979465 | 1 | 0.107340042 | 0.107340042 | 3.604001265 | 1.1720607 | 0.570212848 | 0.8803 | 11.08532284 | 107.5359195 | 216.829368 | 0.091582324 | -1.038188342 | FALSE | FALSE | FALSE |
| ARHGEF11 | 0.1073346 | 0.85980632 | 1 | 118356012.8 | 2.5368E+12 | 3.13733E+13 | 134.17181 | 0.021213857 | 0.95535 | 216061.2873 | 313370305.8 | 6167318824 | 0.000799978 | -3.096921735 | FALSE | FALSE | FALSE |
| HOXD9 | 0.1072916 | 0.85981799 | 1 | 0.107291643 | 0.107291643 | 0.107291643 | 0.54599347 | 0.735296252 | 0.7475 | 0.545993467 | 2.183611235 | 2.183611235 | 0.196507192 | -0.706621549 | FALSE | TRUE | FALSE |
| OR6C2 | 0.1071873 | 0.85982965 | 1 | 0.107187292 | 0.107187292 | 0.107187292 | 0.13847573 | 0.867017295 | 1 | 0.138475727 | 0.138475727 | 0.138475727 | 0.774051125 | -0.111230354 | FALSE | TRUE | FALSE |
| NSUN4 | 0.1071827 | 0.85984132 | 0.87965 | 8.963740153 | 85.18037642 | 85.18037642 | 1.2993211 | 0.554581912 | 0.7559 | 1.299321099 | 12.1182578 | 12.1182578 | 0.082491283 | -1.083591943 | FALSE | FALSE | FALSE |
| AFAP1 | 0.1071231 | 0.85985298 | 1 | 0.977201408 | 0.977201408 | 19.19801331 | 0.14329787 | 0.865992248 | 1 | 0.143297868 | 0.143297868 | 2.82121833 | 0.747555577 | -0.126356514 | FALSE | TRUE | FALSE |
| SRA1 | 0.1071032 | 0.85986464 | 1 | 0.107103223 | 0.107103223 | 0.107103223 | 1.21048043 | 0.56646311 | 0.50015 | 1.210480433 | 1.210480433 | 1.210480433 | 0.088479929 | -1.053155235 | FALSE | FALSE | FALSE |
| PPP1R36 | 0.1070868 | 0.8598763 | 1 | 8.868798125 | 812.984668 | 812.984668 | 0.15619579 | 0.862931589 | 0.0097 | 0.156195794 | 0.156195794 | 0.156195794 | 0.685593235 | -0.163933477 | FALSE | TRUE | FALSE |
| ZFP90 | 0.1070649 | 0.85988795 | 0.5101 | 0.970141205 | 0.970141205 | 3.619015423 | 1.26380487 | 0.560161333 | 0.0132 | 1.263804869 | 1.263804869 | 2.748561703 | 0.084716291 | -1.072033065 | FALSE | FALSE | FALSE |
| PLLP | 0.1070503 | 0.85989961 | 1 | 0.107050345 | 0.107050345 | 0.107050345 | 0.11317543 | 0.872730628 | 1 | 0.113175428 | 0.113175428 | 0.113175428 | 0.945879746 | -0.024164074 | FALSE | TRUE | FALSE |
| HES4 | 0.1070164 | 0.85991126 | 0.75045 | 0.427884437 | 1.711241037 | 1.711241037 | 2.15738535 | 0.475176494 | 0.50135 | 2.157385354 | 20.0426462 | 20.0426462 | 0.049604677 | -1.30447737 | FALSE | FALSE | FALSE |
| GGTLC2 | 0.1069911 | 0.85992291 | 1 | 1.52434E+14 | 5.78331E+19 | 7.96658E+20 | 0.18684662 | 0.85529645 | 1 | 3005486381 | 5.91061E+13 | 7.35483E+14 | 0.572614611 | -0.242137574 | FALSE | TRUE | FALSE |
| HLA-C | 0.1069786 | 0.85993456 | 0.94025 | 8.87353023 | 814.6737351 | 814.6737351 | 0.1560915 | 0.862955012 | 0.013 | 0.156091504 | 0.156091504 | 3.090598009 | 0.685357944 | -0.164082549 | FALSE | TRUE | FALSE |
| ATXN7L1 | 0.1069603 | 0.8599462 | 1 | 0.106960346 | 0.106960346 | 0.106960346 | 0.12273383 | 0.870692639 | 1 | 0.122733828 | 0.122733828 | 0.122733828 | 0.871482196 | -0.059741481 | FALSE | TRUE | FALSE |
| TRIM2 | 0.1069461 | 0.85995785 | 0.0235 | 0.10694613 | 0.10694613 | 2.103026659 | 0.1393511 | 0.866884409 | 1 | 0.139351097 | 0.139351097 | 2.745785741 | 0.767458116 | -0.114945317 | FALSE | TRUE | FALSE |
| SMC6 | 0.1068714 | 0.85996949 | 0.02865 | 0.106871413 | 0.106871413 | 3.597950284 | 1.22845641 | 0.564012617 | 0.51045 | 1.228456406 | 1.228456406 | 24.15973317 | 0.086996504 | -1.060498199 | FALSE | FALSE | FALSE |
| MAGI2 | 0.106854 | 0.85998113 | 1 | 0.99559461 | 0.99559461 | 19.34400982 | 1.29917038 | 0.554710249 | 0.5191 | 1.299170377 | 1.299170377 | 25.3291572 | 0.082247861 | -1.084875387 | FALSE | FALSE | FALSE |
| ADARB1 | 0.1068412 | 0.85999277 | 1 | 0.427156749 | 0.427156749 | 8.39474116 | 0.53954215 | 0.738493865 | 0.75795 | 1.271505705 | 5.083837359 | 18.27206679 | 0.198022029 | -0.703286494 | FALSE | TRUE | FALSE |
| ATOH8 | 0.1068366 | 0.86000441 | 0.75485 | 0.427090381 | 1.70776542 | 2.115766296 | 0.12122916 | 0.870996432 | 1 | 109.3255701 | 1110.661098 | 11558.84601 | 0.881277805 | -0.054887168 | FALSE | TRUE | FALSE |
| SCN2B | 0.1068193 | 0.86001604 | 0.007 | 0.106819286 | 0.106819286 | 0.106819286 | 0.13905355 | 0.866961954 | 1 | 0.13905355 | 0.13905355 | 0.13905355 | 0.768188126 | -0.11453241 | FALSE | TRUE | FALSE |
| OR2Z1 | 0.1067888 | 0.86002767 | 1 | 0.106788796 | 0.106788796 | 0.106788796 | 1.17487236 | 0.569732238 | 0.75675 | 1.174872356 | 10.89115877 | 10.89115877 | 0.090893956 | -1.041464996 | FALSE | FALSE | FALSE |
| ATF5 | 0.1067619 | 0.8600393 | 1 | 0.959732489 | 8.854992714 | 8.854992714 | 0.14667679 | 0.865372327 | 1 | 0.146676787 | 0.146676787 | 0.146676787 | 0.72787168 | -0.137945178 | FALSE | TRUE | FALSE |
| NOP9 | 0.1067499 | 0.86005093 | 0.01995 | 0.106749869 | 0.106749869 | 2.102441224 | 0.13827638 | 0.867083651 | 1 | 0.138276379 | 0.138276379 | 2.728355111 | 0.772003645 | -0.112380649 | FALSE | TRUE | FALSE |
| PRR14 | 0.1067265 | 0.86006256 | 0.96775 | 90.50187328 | 900.7950021 | 9184.859773 | 0.13958664 | 0.866873324 | 1 | 1.295421532 | 12.33867307 | 25.55375574 | 0.764589843 | -0.116571475 | FALSE | TRUE | FALSE |
| OTUB2 | 0.1066682 | 0.86007419 | 0.00625 | 0.106668177 | 0.106668177 | 0.106668177 | 0.13821148 | 0.867105755 | 1 | 0.138211484 | 0.138211484 | 0.138211484 | 0.771775059 | -0.112509261 | FALSE | TRUE | FALSE |
| UGP2 | 0.1066205 | 0.86008581 | 0.01175 | 0.106620539 | 0.106620539 | 2.112957614 | 0.13797097 | 0.867183062 | 1 | 0.137970969 | 0.137970969 | 2.737246357 | 0.772775173 | -0.111946839 | FALSE | TRUE | FALSE |
| SNAPC3 | 0.106575 | 0.86009743 | 1 | 8.830572739 | 83.52690072 | 83.52690072 | 0.15093143 | 0.864403752 | 1 | 0.150931429 | 0.150931429 | 0.150931429 | 0.706115219 | -0.151124428 | FALSE | TRUE | FALSE |
| C14orf93 | 0.1065723 | 0.86010905 | 1 | 0.106572253 | 0.106572253 | 2.106755315 | 0.13791936 | 0.867194099 | 1 | 0.137919359 | 0.137919359 | 2.730234881 | 0.772714243 | -0.111981083 | FALSE | TRUE | FALSE |
| RPL10L | 0.1065611 | 0.86012067 | 1 | 0.426020053 | 0.426020053 | 0.426020053 | 0.14041251 | 0.866706862 | 1 | 0.140412513 | 0.140412513 | 0.140412513 | 0.75891423 | -0.119807304 | FALSE | TRUE | FALSE |
| KIF3C | 0.1065252 | 0.86013228 | 1 | 1.00218825 | 9.676794889 | 33.8712483 | 0.13888974 | 0.866984094 | 1 | 1.308136549 | 12.64504525 | 25.69457195 | 0.766976818 | -0.115217763 | FALSE | TRUE | FALSE |
| NSUN5 | 0.106521 | 0.8601439 | 1 | 0.106521007 | 0.106521007 | 2.10486729 | 0.13791739 | 0.867205134 | 1 | 0.13791739 | 0.13791739 | 2.729196301 | 0.772353707 | -0.112183765 | FALSE | TRUE | FALSE |
| SALL3 | 0.1064798 | 0.86015551 | 0.9853 | 89.62417487 | 9050.933399 | 94254.00486 | 0.16149756 | 0.86169121 | 0.51805 | 1.498123104 | 1.498123104 | 29.06424103 | 0.659327421 | -0.180898862 | FALSE | TRUE | FALSE |
| MMP2 | 0.1064775 | 0.86016712 | 1 | 0.425773554 | 0.425773554 | 8.376611636 | 0.55440764 | 0.731375194 | 0.511 | 0.554407636 | 0.554407636 | 4.693375326 | 0.192056286 | -0.716571473 | FALSE | TRUE | FALSE |
| TIMMDC1 | 0.1064447 | 0.86017873 | 1 | 0.95467234 | 0.95467234 | 0.95467234 | 10.4697518 | 0.192846091 | 0.68705 | 10.46975182 | 964.1396651 | 964.1396651 | 0.010166877 | -1.992812448 | FALSE | FALSE | FALSE |
| NEU4 | 0.1064329 | 0.86019034 | 0.75065 | 0.106432859 | 0.425526593 | 2.096264649 | 2.46582639 | 0.450386841 | 0.77585 | 9.86415327 | 157.9719053 | 632.4155364 | 0.04316316 | -1.364886766 | FALSE | FALSE | FALSE |
| SDHC | 0.1063818 | 0.86020194 | 0.0042 | 0.106381838 | 0.106381838 | 0.106381838 | 0.13702549 | 0.867381467 | 1 | 0.547860815 | 0.547860815 | 0.547860815 | 0.776365346 | -0.109933858 | FALSE | TRUE | FALSE |
| FSCN2 | 0.1063221 | 0.86021354 | 0.7544 | 0.956881365 | 3.827489249 | 8.402141635 | 0.12681251 | 0.869656044 | 1 | 10.61428287 | 983.3131012 | 983.3131012 | 0.838419791 | -0.076538479 | FALSE | TRUE | FALSE |
| GUSB | 0.1062262 | 0.86022514 | 0.6666 | 0.10622624 | 1.698019709 | 6.791424542 | 13.311108 | 0.167843578 | 0.21515 | 0.832091958 | 13.31110798 | 16.40649002 | 0.00798027 | -2.097982396 | FALSE | FALSE | FALSE |
| MATN1 | 0.1061808 | 0.86023674 | 0.50165 | 0.10618077 | 0.10618077 | 0.52482014 | 0.16105857 | 0.861786465 | 0.7536 | 0.161058568 | 0.643950152 | 3.186460991 | 0.659268063 | -0.180937962 | FALSE | TRUE | FALSE |
| AVIL | 0.1061325 | 0.86024834 | 0.0182 | 0.012480511 | 0.106132485 | 0.24556547 | 0.00304179 | 0.898653585 | 0.87855 | 0.220460155 | 1.950419838 | 4.346216598 | 34.89147304 | 1.542719305 | FALSE | TRUE | FALSE |
| ABI2 | 0.1061041 | 0.86025994 | 1 | 0.10610409 | 0.10610409 | 2.101041302 | 0.13835163 | 0.867039421 | 1 | 0.138351633 | 0.138351633 | 2.742864851 | 0.766916067 | -0.115252164 | FALSE | TRUE | FALSE |
| MEIS2 | 0.1060628 | 0.86027153 | 1 | 0.106062767 | 0.106062767 | 2.099567128 | 0.13835786 | 0.867028359 | 0.01245 | 0.138357861 | 0.138357861 | 2.742233668 | 0.766582878 | -0.115440885 | FALSE | TRUE | FALSE |
| ANXA4 | 0.1060282 | 0.86028312 | 1 | 0.106028212 | 0.106028212 | 2.105087548 | 0.13714456 | 0.867337425 | 1 | 0.137144564 | 0.137144564 | 0.137144564 | 0.773112759 | -0.111757159 | FALSE | TRUE | FALSE |
| NTNG1 | 0.1060279 | 0.86029471 | 1 | 0.106027907 | 0.106027907 | 2.095437691 | 1.22847202 | 0.563889379 | 0.50995 | 1.228472022 | 1.228472022 | 2.645394455 | 0.086308768 | -1.063945082 | FALSE | FALSE | FALSE |
| ZFYVE28 | 0.1059726 | 0.8603063 | 1 | 0.105972579 | 0.105972579 | 2.070102462 | 0.1337835 | 0.868027979 | 1 | 1.240773041 | 1.240773041 | 24.30565326 | 0.792119969 | -0.101209038 | FALSE | TRUE | FALSE |
| ZNF165 | 0.1059569 | 0.86031789 | 1 | 0.953063101 | 0.953063101 | 2.102590721 | 0.14140807 | 0.866473169 | 1 | 0.141408074 | 0.141408074 | 2.808712203 | 0.749298974 | -0.125344862 | FALSE | TRUE | FALSE |
| IL20RA | 0.1059501 | 0.86032947 | 0.51075 | 0.96248251 | 0.96248251 | 2.097243229 | 0.13752084 | 0.867271309 | 1 | 1.255639589 | 1.255639589 | 2.725537369 | 0.770429254 | -0.113267235 | FALSE | TRUE | FALSE |
| UGT2B15 | 0.1059234 | 0.86034106 | 0.50425 | 0.105923394 | 0.105923394 | 0.241391405 | 0.17195558 | 0.859157778 | 0.75785 | 0.17195558 | 1.549140825 | 1.549140825 | 0.615992769 | -0.210424386 | FALSE | TRUE | FALSE |
| CDCA3 | 0.1058618 | 0.86035264 | 1 | 0.105861798 | 0.105861798 | 0.105861798 | 0.12442377 | 0.870250285 | 1 | 0.124423775 | 0.124423775 | 0.124423775 | 0.850816475 | -0.070164109 | FALSE | TRUE | FALSE |
| CASP4 | 0.1058561 | 0.86036422 | 0.4997 | 0.105856091 | 0.954058909 | 0.954058909 | 1.24351296 | 0.562281242 | 0.5014 | 1.24351296 | 1.24351296 | 1.24351296 | 0.085126649 | -1.069934463 | FALSE | FALSE | FALSE |
| PTPN9 | 0.1058493 | 0.86037579 | 1 | 0.105849342 | 0.105849342 | 2.092565965 | 1.199717 | 0.567315146 | 0.7526 | 1.199716998 | 11.40095614 | 23.75046594 | 0.088228592 | -1.054390649 | FALSE | FALSE | FALSE |
| YIPF2 | 0.1057281 | 0.86038737 | 1 | 0.105728119 | 0.105728119 | 0.105728119 | 0.12879968 | 0.869249909 | 1 | 1.159689055 | 10.71690014 | 10.71690014 | 0.820872499 | -0.085724294 | FALSE | TRUE | FALSE |
| SLC6A20 | 0.1056901 | 0.86039894 | 0.14225 | 0.10569005 | 0.422584436 | 0.894903218 | 3.64721835 | 0.374714525 | 0.4994 | 2.636321215 | 138.7570882 | 554.8574842 | 0.028978262 | -1.537927659 | FALSE | FALSE | FALSE |
| TTC13 | 0.1056753 | 0.86041051 | 0.0221 | 0.105675255 | 0.105675255 | 2.082894555 | 0.1372915 | 0.867315392 | 1 | 0.137291496 | 0.137291496 | 2.710778206 | 0.769714503 | -0.113670331 | FALSE | TRUE | FALSE |
| HACL1 | 0.1056311 | 0.86042208 | 0.5054 | 0.105631054 | 0.422400692 | 0.422400692 | 0.02076313 | 0.893645071 | 1 | 0.083031084 | 0.083031084 | 0.411185637 | 5.087435102 | 0.706498882 | FALSE | TRUE | FALSE |
| SPAG4 | 0.1056013 | 0.86043365 | 1 | 0.105601265 | 0.105601265 | 2.087017414 | 0.13717083 | 0.867326409 | 1 | 0.137170833 | 0.137170833 | 2.71479471 | 0.769852181 | -0.113592656 | FALSE | TRUE | FALSE |
| TAX1BP1 | 0.1055397 | 0.86044522 | 1 | 0.105539682 | 0.105539682 | 2.082826886 | 0.13688055 | 0.867403479 | 1 | 0.136880545 | 0.136880545 | 2.705646155 | 0.771034935 | -0.112925944 | FALSE | TRUE | FALSE |
| CYB5D2 | 0.1055046 | 0.86045678 | 0.25535 | 0.105504554 | 0.421930524 | 0.523628121 | 1.32248413 | 0.553036593 | 0.7863 | 5.289122757 | 115.5024672 | 461.9387958 | 0.079777558 | -1.098119263 | FALSE | FALSE | FALSE |
| ACOT2 | 0.1054154 | 0.86046834 | 1 | 0.105415449 | 0.105415449 | 2.079614186 | 1.23347608 | 0.563395675 | 0.51 | 1.233476076 | 1.233476076 | 2.631114231 | 0.085462094 | -1.068226468 | FALSE | FALSE | FALSE |
| ZNF225 | 0.1054114 | 0.86047991 | 0.27095 | 0.105411387 | 0.934204667 | 2.081128168 | 0.0207529 | 0.89365977 | 1 | 0.18125968 | 0.18125968 | 0.410356415 | 5.079358032 | 0.705808826 | FALSE | TRUE | FALSE |
| UAP1L1 | 0.1053675 | 0.86049146 | 0.0101 | 0.105367492 | 0.105367492 | 2.088935476 | 0.13678759 | 0.867447479 | 1 | 0.136787593 | 0.136787593 | 2.714696426 | 0.770300065 | -0.113340065 | FALSE | TRUE | FALSE |
| BTNL2 | 0.1053184 | 0.86050302 | 0.50965 | 0.950782129 | 0.950782129 | 2.087017881 | 0.14080051 | 0.866651289 | 1 | 0.140800514 | 0.140800514 | 2.793223226 | 0.747997487 | -0.126099861 | FALSE | TRUE | FALSE |
| SEC22A | 0.1052634 | 0.86051458 | 0.00785 | 0.105263402 | 0.105263402 | 0.105263402 | 1.19013839 | 0.568526498 | 0.50015 | 1.190138392 | 1.190138392 | 1.190138392 | 0.088446354 | -1.053320065 | FALSE | FALSE | FALSE |
| SLC39A4 | 0.1052558 | 0.86052613 | 1 | 0.96474808 | 0.96474808 | 3.564936174 | 0.14124141 | 0.866551153 | 1 | 0.141241412 | 0.141241412 | 2.787902871 | 0.745219179 | -0.127715976 | FALSE | TRUE | FALSE |
| KCNJ10 | 0.1052483 | 0.86053768 | 1 | 0.105248319 | 0.105248319 | 2.087070211 | 0.13742176 | 0.867304374 | 0.0102 | 0.137421758 | 0.137421758 | 2.727854668 | 0.765878132 | -0.11584033 | FALSE | TRUE | FALSE |
| CCT2 | 0.1052008 | 0.86054923 | 1 | 6.726258113 | 26.90969464 | 107.684325 | 2.27148649 | 0.466907905 | 0.25525 | 0.568012137 | 2.27148649 | 2.818792328 | 0.046313628 | -1.334291202 | FALSE | FALSE | FALSE |
| USP30 | 0.105123 | 0.86056078 | 1 | 0.105123039 | 0.105123039 | 2.079669394 | 0.13687679 | 0.867425483 | 0.01395 | 0.136876788 | 0.136876788 | 2.711391289 | 0.768012168 | -0.114631899 | FALSE | TRUE | FALSE |
| SLC20A2 | 0.1051121 | 0.86057233 | 0.02065 | 0.105112054 | 0.105112054 | 2.067642868 | 0.15995716 | 0.862059633 | 0.75405 | 0.159957163 | 0.639463334 | 3.152726411 | 0.657126269 | -0.182351172 | FALSE | TRUE | FALSE |

| | | | | | | | | | | | | | | | | | |
|---|---|---|---|---|---|---|---|---|---|---|---|---|---|---|---|---|---|
| SLC9B1 | 0.1050332 | 0.86058387 | 1 | 0.987316488 | 0.987316488 | 2.082280408 | 1.06805766 | 0.581574297 | 0.50465 | 1.068057658 | 1.068057658 | 2.2521987 | 0.098340352 | -1.007268242 | FALSE | FALSE | FALSE |
| HOXA1 | 0.1050325 | 0.86059541 | 1 | 0.1050325 | 0.1050325 | 2.081983601 | 1.24680622 | 0.561784239 | 0.5003 | 1.246806219 | 1.246806219 | 2.636898309 | 0.084241238 | -1.074475257 | FALSE | FALSE | FALSE |
| BABAM1 | 0.1049696 | 0.86060696 | 0.0093 | 0.104969566 | 0.104969566 | 0.104969566 | 0.11147936 | 0.873045985 | 1 | 0.111479361 | 0.111479361 | 0.111479361 | 0.94160538 | -0.026131069 | FALSE | TRUE | FALSE |
| GPR132 | 0.1049637 | 0.8606185 | 0.75395 | 0.948294707 | 8.793255757 | 18.75978538 | 0.14311236 | 0.866059556 | 1 | 0.572408474 | 0.572408474 | 2.83484347 | 0.733435505 | -0.13463807 | FALSE | TRUE | FALSE |
| SCNN1G | 0.104939 | 0.86063003 | 0.02005 | 0.104938979 | 0.104938979 | 2.068039598 | 0.54105841 | 0.737713022 | 0.01555 | 0.541058414 | 0.541058414 | 2.671090128 | 0.193951293 | -0.71230732 | FALSE | TRUE | FALSE |
| EXTL2 | 0.1048434 | 0.86064157 | 1 | 0.104843428 | 0.104843428 | 0.104843428 | 0.53387138 | 0.740934402 | 0.74775 | 0.533871382 | 2.135443283 | 2.135443283 | 0.196383309 | -0.706895427 | FALSE | TRUE | FALSE |
| SLC35G2 | 0.104769 | 0.8606531 | 0.01045 | 0.104768991 | 0.104768991 | 2.080905595 | 1.15112443 | 0.572244598 | 0.75425 | 1.151124425 | 10.64025822 | 10.64025822 | 0.09101448 | -1.040889506 | FALSE | FALSE | FALSE |
| PTK2 | 0.1047051 | 0.86066463 | 1 | 0.10470513 | 0.10470513 | 2.050931535 | 1.19780922 | 0.567921603 | 0.7584 | 1.197809219 | 11.4662675 | 40.39823395 | 0.087413862 | -1.058419692 | FALSE | FALSE | FALSE |
| SPTLC3 | 0.1046436 | 0.86067616 | 0.0078 | 0.028033647 | 0.028033647 | 0.061118644 | 0.02408451 | 0.892718805 | 0.5091 | 0.054970631 | 0.219634857 | 0.219634857 | 4.344850874 | 0.637974875 | FALSE | TRUE | FALSE |
| ZWILCH | 0.1045856 | 0.86068769 | 1 | 0.94038405 | 8.678459987 | 18.61816277 | 0.14405267 | 0.865868687 | 1 | 0.144052672 | 0.144052672 | 2.855479107 | 0.726023074 | -0.139049577 | FALSE | TRUE | FALSE |
| PCDH17 | 0.1045655 | 0.86069922 | 1 | 0.974796701 | 9.326716759 | 180.0774684 | 0.14526343 | 0.865587081 | 1 | 0.145263429 | 0.145263429 | 2.81641589 | 0.719833934 | -0.142767684 | FALSE | TRUE | FALSE |
| SLC18B1 | 0.104442 | 0.86071075 | 1 | 0.104441985 | 0.104441985 | 2.072561488 | 0.13563331 | 0.867667079 | 0.00965 | 0.13563331 | 0.13563331 | 0.13563331 | 0.770031971 | -0.113491243 | FALSE | TRUE | FALSE |
| HSD17B1 | 0.1044057 | 0.86072227 | 0.01325 | 0.104405715 | 0.104405715 | 2.065253243 | 0.13630897 | 0.867546381 | 1 | 0.136308968 | 0.136308968 | 2.699883993 | 0.765948976 | -0.11580016 | FALSE | TRUE | FALSE |
| MOGAT1 | 0.1044042 | 0.86073379 | 0.5062 | 0.948622583 | 0.948622583 | 0.948622583 | 0.13993766 | 0.866795689 | 1 | 0.139937665 | 0.139937665 | 0.139937665 | 0.74607673 | -0.127216505 | FALSE | TRUE | FALSE |
| KNSTRN | 0.104302 | 0.86074531 | 1 | 0.933449054 | 0.933449054 | 0.933449054 | 0.13924647 | 0.866906573 | 0.0079 | 0.139246468 | 0.139246468 | 0.139246468 | 0.749045988 | -0.125491518 | FALSE | TRUE | FALSE |
| NCKIPSD | 0.104293 | 0.86075683 | 0.5109 | 0.949624724 | 0.949624724 | 18.71054669 | 0.13569656 | 0.867656115 | 1 | 1.241533714 | 1.241533714 | 4.591687609 | 0.768574825 | -0.114313845 | FALSE | TRUE | FALSE |
| OR2B6 | 0.1042426 | 0.86076835 | 1 | 0.104242599 | 0.970596455 | 0.970596455 | 9.36016912 | 0.207750921 | 0.68875 | 9.360169122 | 920.9394026 | 920.9394026 | 0.011136829 | -1.953238467 | FALSE | FALSE | FALSE |
| XKR6 | 0.1042143 | 0.86077986 | 1 | 0.104214317 | 0.104214317 | 2.050163267 | 1.24576483 | 0.562032871 | 0.02145 | 1.245764829 | 1.245764829 | 24.5556937 | 0.083654888 | -1.07750868 | FALSE | FALSE | FALSE |
| CPED1 | 0.1041827 | 0.86079137 | 0.9677 | 8.168926849 | 75.0990355 | 707.312044 | 1.4864977 | 0.537188926 | 0.8903 | 13.48134305 | 1193.673638 | 11640.30416 | 0.070085983 | -1.154368833 | FALSE | FALSE | FALSE |
| TAMM41 | 0.1041588 | 0.86080288 | 1 | 0.104158763 | 0.104158763 | 0.104158763 | 0.13518224 | 0.867754738 | 0.0071 | 0.135182239 | 0.135182239 | 0.135182239 | 0.770506267 | -0.113223824 | FALSE | TRUE | FALSE |
| PRIMPOL | 0.104143 | 0.86081439 | 0.50865 | 0.104143028 | 0.104143028 | 0.515883727 | 2.94438528 | 0.414976545 | 0.2591 | 0.328233663 | 2.944385283 | 2.944385283 | 0.035370041 | -1.451364438 | FALSE | FALSE | FALSE |
| ST6GALNAC: | 0.1041146 | 0.8608259 | 0.5042 | 0.416415022 | 0.416415022 | 0.416415022 | 0.52487008 | 0.744937674 | 0.75625 | 1.191900057 | 4.767162234 | 4.767162234 | 0.19836261 | -0.702540186 | FALSE | TRUE | FALSE |
| ZNF101 | 0.1040854 | 0.86083741 | 1 | 0.104085381 | 0.104085381 | 2.063704675 | 0.13526412 | 0.867710923 | 1 | 0.135264115 | 0.135264115 | 2.684676057 | 0.769497366 | -0.113792862 | FALSE | TRUE | FALSE |
| CLCC1 | 0.1040344 | 0.86084891 | 0.0166 | 0.104034439 | 0.104034439 | 2.059025677 | 0.13498126 | 0.867787581 | 1 | 0.134981257 | 0.134981257 | 2.674863907 | 0.770732486 | -0.113096335 | FALSE | TRUE | FALSE |
| ZNF354B | 0.1039996 | 0.86086041 | 0.0151 | 0.103999605 | 0.103999605 | 2.058711713 | 10.7662529 | 0.190128375 | 0.2527 | 1.155267612 | 10.7662529 | 22.89710663 | 0.009659777 | -2.015032888 | FALSE | FALSE | FALSE |
| ZNF212 | 0.1039517 | 0.86087191 | 0.50675 | 0.936983262 | 0.936983262 | 2.056760412 | 0.13099027 | 0.868690274 | 1 | 1.186698251 | 11.0341844 | 11.0341844 | 0.793583038 | -0.100407623 | FALSE | TRUE | FALSE |
| DZIP3 | 0.1039006 | 0.86088341 | 0.0275 | 0.103900608 | 0.103900608 | 2.037952546 | 0.08066594 | 0.880154864 | 0.5139 | 0.080665939 | 0.080665939 | 1.585890587 | 1.288035683 | 0.109927895 | FALSE | TRUE | FALSE |
| TBPL2 | 0.103896 | 0.86089491 | 1 | 0.103896004 | 0.103896004 | 2.062499966 | 0.13101518 | 0.868679467 | 1 | 0.131015181 | 1.191051969 | 1.191051969 | 0.793007364 | -0.10072278 | FALSE | TRUE | FALSE |
| YIF1B | 0.1037717 | 0.8609064 | 0.0101 | 0.103771706 | 0.103771706 | 2.060557091 | 0.13521976 | 0.867732834 | 0.0074 | 0.135219761 | 0.135219761 | 0.135219761 | 0.767430035 | -0.114961208 | FALSE | TRUE | FALSE |
| NCAPG2 | 0.1037716 | 0.86091789 | 0.033 | 0.103771574 | 0.103771574 | 3.499021447 | 0.13513109 | 0.867765687 | 1 | 0.135131087 | 0.135131087 | 2.649455262 | 0.767932652 | -0.114676866 | FALSE | TRUE | FALSE |
| STEAP1 | 0.1037443 | 0.86092938 | 1 | 0.103744323 | 0.103744323 | 0.103744323 | 0.13526645 | 0.867699965 | 1 | 0.135266454 | 0.135266454 | 0.135266454 | 0.766962689 | -0.115225763 | FALSE | TRUE | FALSE |
| NOL11 | 0.103647 | 0.86094087 | 0.01825 | 0.103646958 | 0.103646958 | 2.046468852 | 0.13065823 | 0.868809034 | 1 | 1.183859637 | 1.183859637 | 4.421560674 | 0.79326771 | -0.100580223 | FALSE | TRUE | FALSE |
| IGDCC4 | 0.1036248 | 0.86095236 | 0.5203 | 0.103624826 | 0.103624826 | 2.017680097 | 0.04011202 | 0.888934938 | 1 | 0.04011202 | 0.04011202 | 0.783511388 | 2.583385855 | 0.412189277 | FALSE | TRUE | FALSE |
| ATRIP | 0.1034925 | 0.86096385 | 1 | 0.947225598 | 0.947225598 | 18.63936156 | 0.13491379 | 0.867798526 | 1 | 1.240057611 | 1.240057611 | 4.564262745 | 0.767101193 | -0.115147342 | FALSE | TRUE | FALSE |
| PREX1 | 0.1034851 | 0.86097533 | 1 | 0.413815979 | 1.989973237 | 7.957504012 | 0.13687686 | 0.867414482 | 1 | 0.136876865 | 0.136876865 | 4.607410682 | 0.756044879 | -0.121452424 | FALSE | TRUE | FALSE |
| KLK1 | 0.1034213 | 0.86098681 | 1 | 0.103421255 | 0.103421255 | 0.103421255 | 1.1954607 | 0.568163709 | 0.5039 | 1.195460698 | 1.195460698 | 1.195460698 | 0.086511631 | -1.062925499 | FALSE | FALSE | FALSE |
| POC1A | 0.1034112 | 0.86099829 | 0.01095 | 0.103411193 | 0.103411193 | 2.049987639 | 0.13509134 | 0.867776635 | 0.00985 | 0.135091341 | 0.135091341 | 0.135091341 | 0.765490901 | -0.116059967 | FALSE | TRUE | FALSE |
| SNCAIP | 0.1033748 | 0.86100977 | 1 | 0.103374756 | 0.103374756 | 2.032169848 | 0.13470186 | 0.86782041 | 1 | 0.134701865 | 0.134701865 | 2.653453906 | 0.767433739 | -0.114959112 | FALSE | TRUE | FALSE |
| TUBGCP3 | 0.1032648 | 0.86102125 | 1 | 0.103264777 | 0.103264777 | 2.025520722 | 0.5323006 | 0.741737265 | 0.50915 | 0.532300602 | 0.532300602 | 10.46519879 | 0.193997107 | -0.712204747 | FALSE | TRUE | FALSE |
| MFSD12 | 0.1032237 | 0.86103273 | 0.99995 | 557003.6875 | 1169379444 | 18777209571 | 3.80870931 | 0.366009561 | 0.9577 | 553.1054437 | 82469.40124 | 787110.1445 | 0.027102014 | -1.566998437 | FALSE | FALSE | FALSE |
| NELFCD | 0.1032193 | 0.8610442 | 0.0177 | 0.103219346 | 0.103219346 | 2.03883979 | 1.20255715 | 0.56682866 | 0.50885 | 1.202557151 | 1.202557151 | 2.583517736 | 0.085833215 | -1.066344622 | FALSE | FALSE | FALSE |
| ZNF394 | 0.1031264 | 0.86105567 | 1 | 0.103126377 | 0.103126377 | 2.034621923 | 0.53057474 | 0.742702106 | 0.504 | 0.530574744 | 0.530574744 | 2.621354325 | 0.194367293 | -0.711376814 | FALSE | TRUE | FALSE |
| VANGL1 | 0.1030782 | 0.86106714 | 0.8947 | 0.411963305 | 6.585173773 | 26.33809061 | 0.0495473 | 0.886783039 | 0.00805 | 0.049547304 | 0.049547304 | 0.049547304 | 2.080400265 | 0.3181469 | FALSE | TRUE | FALSE |
| CLDN22 | 0.1030143 | 0.86107861 | 1 | 0.103014331 | 0.103014331 | 0.103014331 | 1.21414336 | 0.566097201 | 0.50065 | 1.214143365 | 1.214143365 | 1.214143365 | 0.084845278 | -1.071372324 | FALSE | FALSE | FALSE |
| TRHR | 0.1029935 | 0.86109008 | 1 | 0.927667046 | 0.927667046 | 0.927667046 | 97.5502646 | 0.030202676 | 0.0653 | 10.25320151 | 951.3486874 | 951.3486874 | 0.0010558 | -2.976418447 | FALSE | FALSE | FALSE |
| SLC25A48 | 0.102898 | 0.86110154 | 0.5084 | 0.944569005 | 0.944569005 | 0.944569005 | 1.2373047 | 0.563024787 | 0.5067 | 1.2373047 | 1.2373047 | 1.2373047 | 0.083162984 | -1.080069938 | FALSE | FALSE | FALSE |
| WWC1 | 0.1028534 | 0.86111301 | 1 | 0.102853382 | 0.102853382 | 2.008418319 | 0.13426294 | 0.867918806 | 0.02875 | 0.134262938 | 0.134262938 | 2.629223487 | 0.766059374 | -0.115737569 | FALSE | TRUE | FALSE |
| SORCS2 | 0.1028403 | 0.86112447 | 0.03355 | 0.102840301 | 0.102840301 | 2.008463434 | 0.1341579 | 0.867940654 | 1 | 0.134157897 | 0.134157897 | 2.627513457 | 0.766561661 | -0.115452905 | FALSE | TRUE | FALSE |
| CEACAM4 | 0.1027656 | 0.86113593 | 0.50855 | 0.102765597 | 0.102765597 | 0.102765597 | 0.01597844 | 0.894867183 | 1 | 0.015978439 | 0.015978439 | 0.015978439 | 6.431516604 | 0.808313395 | FALSE | TRUE | FALSE |
| ATAD3A | 0.1027527 | 0.86114738 | 0.9589 | 19.54461133 | 2915.88562 | 8976.660011 | 8.34459819 | 0.22055773 | 0.81415 | 177.6162338 | 67155.39711 | 569287.9283 | 0.012313674 | -1.90961235 | FALSE | FALSE | FALSE |
| WDR46 | 0.1027455 | 0.86115884 | 1 | 0.927850773 | 0.927850773 | 18.28391152 | 0.13368183 | 0.868060697 | 1 | 1.214170148 | 1.214170148 | 4.521771701 | 0.768582023 | -0.114309778 | FALSE | TRUE | FALSE |
| KDM3A | 0.1027403 | 0.8611703 | 0.5185 | 0.410872457 | 0.410872457 | 8.021123751 | 0.13435257 | 0.867907879 | 0.5107 | 0.537308105 | 0.537308105 | 10.51961811 | 0.764706311 | -0.116505325 | FALSE | TRUE | FALSE |
| CBR1 | 0.1025336 | 0.86118175 | 0.9368 | 8.675713626 | 812.4862808 | 812.4862808 | 0.12431523 | 0.870281976 | 1 | 406.1783965 | 40211.94002 | 103670.5043 | 0.824787278 | -0.083658046 | FALSE | TRUE | FALSE |
| UNC45A | 0.1024604 | 0.8611932 | 0.27475 | 0.102460394 | 0.922970472 | 3.459264435 | 0.02067949 | 0.893674466 | 1 | 0.020679488 | 0.020679488 | 0.405969007 | 4.954687287 | 0.695016249 | FALSE | TRUE | FALSE |
| CHKB | 0.1024518 | 0.86120465 | 1 | 0.102451753 | 0.102451753 | 2.029534757 | 0.13360863 | 0.8680825 | 0.01105 | 0.133608628 | 0.133608628 | 2.649771716 | 0.766804913 | -0.115315113 | FALSE | TRUE | FALSE |
| ZNF37A | 0.1024446 | 0.8612161 | 0.7494 | 0.102444613 | 0.893619559 | 2.031103749 | 0.01980254 | 0.893894467 | 1 | 0.170377865 | 1.504598893 | 1.504598893 | 5.173306279 | 0.713768191 | FALSE | TRUE | FALSE |
| ANTXR1 | 0.1024367 | 0.86122754 | 1 | 0.102436692 | 0.102436692 | 2.025748984 | 0.13261015 | 0.8682893 | 1 | 0.530226572 | 0.530226572 | 2.625987457 | 0.77246492 | -0.112121234 | FALSE | TRUE | FALSE |
| SCRN3 | 0.1023915 | 0.86123899 | 1 | 8.487151623 | 80.2937209 | 80.2937209 | 0.13309745 | 0.868191416 | 1 | 11.17160929 | 106.3535791 | 106.3535791 | 0.769297263 | -0.113905812 | FALSE | TRUE | FALSE |
| ZNF644 | 0.1023604 | 0.86125043 | 1 | 0.102360444 | 0.102360444 | 3.443692626 | 0.13398744 | 0.867962495 | 0.02945 | 0.133987436 | 0.133987436 | 2.627087489 | 0.763955545 | -0.116931913 | FALSE | TRUE | FALSE |
| NISCH | 0.1022808 | 0.86126187 | 1 | 0.939338322 | 0.939338322 | 18.0660387 | 0.13616308 | 0.867579319 | 1 | 0.54464278 | 0.54464278 | 10.52347112 | 0.751163631 | -0.124265447 | FALSE | TRUE | FALSE |
| ZXDC | 0.1022504 | 0.86127331 | 1 | 0.920386941 | 0.920386941 | 18.15888944 | 0.13707215 | 0.867359449 | 1 | 0.137072146 | 0.137072146 | 2.708781687 | 0.745960559 | -0.127284134 | FALSE | TRUE | FALSE |
| USP6NL | 0.1022328 | 0.86128474 | 0.02295 | 0.102232762 | 0.102232762 | 2.011756515 | 0.12727986 | 0.869592068 | 1 | 0.127279862 | 0.127279862 | 2.5094895 | 0.803212387 | -0.095169603 | FALSE | TRUE | FALSE |
| FKTN | 0.1022324 | 0.86129618 | 0.01275 | 0.102232446 | 0.102232446 | 0.224185251 | 0.16148585 | 0.861703123 | 0.68615 | 1.504994108 | 57.50261217 | 57.50261217 | 0.633073704 | -0.198545726 | FALSE | TRUE | FALSE |
| CEP170B | 0.1021535 | 0.86130761 | 1 | 0.94250666 | 0.94250666 | 18.26228907 | 0.13773879 | 0.867238229 | 1 | 0.137738787 | 0.137738787 | 2.678904772 | 0.741646426 | -0.129803092 | FALSE | TRUE | FALSE |
| EMILIN1 | 0.1021272 | 0.86131904 | 1 | 0.93822817 | 3.752318928 | 18.35554583 | 0.13759323 | 0.867260284 | 1 | 0.550296092 | 0.550296092 | 10.79429045 | 0.742239713 | -0.129455813 | FALSE | TRUE | FALSE |
| TMEM8B | 0.1020587 | 0.86133047 | 0.01565 | 0.10205868 | 0.10205868 | 2.014457872 | 4.50772146 | 0.32943808 | 0.00365 | 0.132998236 | 0.132998236 | 2.629284557 | 0.022640858 | -1.645107127 | FALSE | FALSE | FALSE |
| TDRD7 | 0.1019908 | 0.8613419 | 1 | 0.101990806 | 0.101990806 | 2.000779391 | 0.13296434 | 0.868213179 | 1 | 0.13296434 | 0.13296434 | 2.614406205 | 0.767053828 | -0.115174159 | FALSE | TRUE | FALSE |
| MAST1 | 0.1019824 | 0.86135333 | 0.88555 | 9.012890306 | 88.04094067 | 303.0736263 | 0.14640148 | 0.865406276 | 0.0542 | 0.146401481 | 2.808921268 | 4.929499368 | 0.696594341 | -0.157020058 | FALSE | TRUE | FALSE |
| FFAR2 | 0.1018644 | 0.86136475 | 1 | 0.101864446 | 0.101864446 | 0.101864446 | 0.12901282 | 0.869174834 | 1 | 1.184791931 | 1.184791931 | 2.560473858 | 0.789568421 | -0.10261023 | FALSE | TRUE | FALSE |
| SLC37A1 | 0.1017869 | 0.86137617 | 0.0158 | 0.101786864 | 0.101786864 | 2.00906814 | 0.13316306 | 0.868180532 | 0.0154 | 0.133163063 | 0.133163063 | 2.632514713 | 0.764377608 | -0.116692044 | FALSE | TRUE | FALSE |
| MPP3 | 0.1017017 | 0.8613876 | 1 | 0.920113372 | 8.543900654 | 18.15399276 | 1.24391563 | 0.562157094 | 0.50445 | 1.24391563 | 1.24391563 | 4.624318237 | 0.081759287 | -1.087462903 | FALSE | FALSE | FALSE |
| KLC1 | 0.1016939 | 0.86139902 | 1 | 0.101693947 | 0.101693947 | 2.009161272 | 4.72560391 | 0.318982795 | 0.2579 | 1.181739847 | 4.725603908 | 10.09372991 | 0.021519778 | -1.6671622 12 | FALSE | FALSE | FALSE |
| CLDN17 | 0.1015514 | 0.86141043 | 1 | 0.947461778 | 9.07249284 | 9.07249284 | 5.02094016 | 0.305649276 | 0.3794 | 2.145803855 | 20.07978378 | 20.07978378 | 0.020225565 | -1.694099338 | FALSE | FALSE | FALSE |

| | | | | | | | | | | | | | | | | | |
|---|---|---|---|---|---|---|---|---|---|---|---|---|---|---|---|---|---|
| CCDC50 | 0.1015334 | 0.86142185 | 1 | 0.101533436 | 0.101533436 | 2.011571281 | 0.13216985 | 0.868408748 | 1 | 0.132169846 | 0.132169846 | 2.621499097 | 0.768204242 | -0.114523299 | FALSE | TRUE | FALSE |
| USP17L2 | 0.1015161 | 0.86143326 | 1 | 0.932479011 | 8.791026398 | 18.44779844 | 91.9382453 | 0.032007222 | 0.5072 | 91.93824531 | 912.1171067 | 9270.085343 | 0.001104177 | -2.956961117 | FALSE | FALSE | FALSE |
| CLNS1A | 0.101516 | 0.86144468 | 1 | 0.10151604 | 0.10151604 | 0.10151604 | 0.13264874 | 0.868267559 | 1 | 0.132648738 | 0.132648738 | 0.132648738 | 0.765299705 | -0.116168454 | FALSE | TRUE | FALSE |
| PPP1R21 | 0.10149 | 0.86145609 | 1 | 0.10149003 | 0.10149003 | 2.002964618 | 0.13232252 | 0.868376192 | 1 | 0.13232252 | 0.13232252 | 2.615616422 | 0.76698985 | -0.115210383 | FALSE | TRUE | FALSE |
| SLC24A4 | 0.1014653 | 0.8614675 | 0.51275 | 0.101465263 | 0.101465263 | 0.858704673 | 0.154156 | 0.863549676 | 0.75915 | 0.154155996 | 0.616494554 | 3.043880239 | 0.658198613 | -0.181643037 | FALSE | TRUE | FALSE |
| TNFRSF18 | 0.1014454 | 0.8614789 | 1 | 0.907033648 | 8.323783577 | 8.323783577 | 1.22411852 | 0.564750519 | 0.50535 | 1.224118523 | 1.224118523 | 1.224118523 | 0.08287218 | -1.081591236 | FALSE | FALSE | FALSE |
| KRTAP10-7 | 0.1014296 | 0.86149031 | 1 | 3.55535E+17 | 1.23861E+22 | 2.56064E+24 | 0.12052699 | 0.87111132 | 1 | 3.77437E+18 | 2.06734E+24 | 4.58601E+26 | 0.841550702 | -0.074919713 | FALSE | TRUE | FALSE |
| SARM1 | 0.1013795 | 0.86150171 | 0.01265 | 0.101379497 | 0.101379497 | 2.006599561 | 0.13226869 | 0.868397897 | 1 | 0.132268687 | 0.132268687 | 2.621248105 | 0.766466339 | -0.115506913 | FALSE | TRUE | FALSE |
| PCDHB16 | 0.1013262 | 0.86151311 | 0.69835 | 0.912923291 | 79.9225804 | 79.9225804 | 0.01988138 | 0.893879826 | 0.8793 | 1.60320666 | 14.96009697 | 31.49393602 | 5.096536459 | 0.707275135 | FALSE | TRUE | FALSE |
| TNFRSF10B | 0.1013009 | 0.86152451 | 1 | 0.912068715 | 0.912068715 | 2.006676979 | 1.12309565 | 0.575670804 | 0.87795 | 10.42324318 | 99.22079276 | 99.22079276 | 0.090197938 | -1.044803393 | FALSE | FALSE | FALSE |
| TBRG4 | 0.1012849 | 0.86153591 | 0.01835 | 0.101284905 | 0.101284905 | 1.997743361 | 0.11850359 | 0.871589735 | 1 | 0.118503589 | 0.118503589 | 2.341246466 | 0.854699046 | -0.068186781 | FALSE | TRUE | FALSE |
| ADAMTS6 | 0.1012816 | 0.86154731 | 1 | 1.619743471 | 25.92960254 | 54.56767791 | 0.13132204 | 0.868582126 | 1 | 8.402803229 | 41.22426281 | 164.926444 | 0.771246349 | -0.112806879 | FALSE | TRUE | FALSE |
| EIF5B | 0.101277 | 0.8615587 | 1 | 0.101276999 | 0.101276999 | 1.984743993 | 0.12846665 | 0.869367711 | 1 | 1.184434401 | 1.184434401 | 23.26788282 | 0.788352456 | -0.103279575 | FALSE | TRUE | FALSE |
| ASB6 | 0.1012667 | 0.86157009 | 1 | 0.101266662 | 0.101266662 | 2.002211047 | 0.13215176 | 0.868419596 | 0.01225 | 0.132151759 | 0.132151759 | 2.616449795 | 0.766290686 | -0.115606453 | FALSE | TRUE | FALSE |
| RNF25 | 0.1012613 | 0.86158148 | 1 | 0.101261284 | 0.101261284 | 2.004735197 | 0.1319538 | 0.868484652 | 1 | 0.1319538 | 0.1319538 | 2.61555422 | 0.767399528 | -0.114978472 | FALSE | TRUE | FALSE |
| BPI | 0.1012121 | 0.86159287 | 0.01435 | 0.101212108 | 0.101212108 | 2.00308815 | 0.13171025 | 0.868506325 | 1 | 0.131710248 | 0.131710248 | 2.609953552 | 0.768445205 | -0.114387095 | FALSE | TRUE | FALSE |
| SDR16C5 | 0.1011599 | 0.86160426 | 0.50175 | 0.404457002 | 1.617502484 | 1.617502484 | 0.03863673 | 0.889291692 | 1 | 0.35385182 | 1.414542659 | 1.414542659 | 2.618230985 | 0.418007958 | FALSE | TRUE | FALSE |
| SLC4A7 | 0.1011465 | 0.86161564 | 0.8184 | 0.927589273 | 21.08866632 | 84.28840664 | 0.01247332 | 0.895915687 | 0.8816 | 0.199257149 | 0.796695888 | 3.90781565 | 8.109029062 | 0.908968857 | FALSE | TRUE | FALSE |
| TCP1 | 0.1011392 | 0.86162703 | 0.0142 | 0.101139242 | 0.101139242 | 2.001479351 | 0.13115699 | 0.868614588 | 1 | 0.131156987 | 0.524390285 | 2.598799524 | 0.771131178 | -0.112871738 | FALSE | TRUE | FALSE |
| OR5B17 | 0.1011155 | 0.86163841 | 0.50205 | 0.933038088 | 0.933038088 | 0.933038088 | 0.11963205 | 0.871288513 | 1 | 0.119632047 | 0.119632047 | 0.119632047 | 0.845220638 | -0.073029907 | FALSE | TRUE | FALSE |
| FBXO36 | 0.1011075 | 0.86164979 | 0.50475 | 0.90268414 | 0.90268414 | 0.90268414 | 0.13537332 | 0.867678043 | 1 | 0.135373323 | 0.135373323 | 0.135373323 | 0.746879328 | -0.126749561 | FALSE | TRUE | FALSE |
| AKAP5 | 0.1010845 | 0.86166116 | 0.5025 | 0.952886217 | 0.952886217 | 0.952886217 | 0.10716131 | 0.874043465 | 1 | 1.011072013 | 1.011072013 | 1.011072013 | 0.943292496 | -0.025353621 | FALSE | TRUE | FALSE |
| PSPH | 0.1010382 | 0.86167254 | 1 | 0.921920388 | 3.686756908 | 3.686756908 | 120.983541 | 0.024096253 | 0.8066 | 1108.983275 | 54295.00516 | 217549.608 | 0.00083514 | -3.07824064 | FALSE | FALSE | FALSE |
| MTFR2 | 0.1010323 | 0.86168391 | 1 | 0.101032274 | 0.101032274 | 2.006080204 | 1.0895993 | 0.579852793 | 0.877 | 10.13000305 | 96.5973982 | 96.5973982 | 0.092724246 | -1.032806687 | FALSE | FALSE | FALSE |
| PRL | 0.1008773 | 0.86169529 | 0.0087 | 0.10087726 | 0.10087726 | 0.10087726 | 0.13204763 | 0.868452132 | 0.00665 | 0.132047633 | 0.132047633 | 0.132047633 | 0.763945991 | -0.116937344 | FALSE | TRUE | FALSE |
| CBR3 | 0.1008527 | 0.86170666 | 1 | 0.10085265 | 0.10085265 | 0.10085265 | 0.13177528 | 0.868495489 | 0.0083 | 0.131775275 | 0.131775275 | 0.131775275 | 0.765338189 | -0.116146616 | FALSE | TRUE | FALSE |
| GTPBP3 | 0.1007633 | 0.86171802 | 0.02085 | 0.100763274 | 0.100763274 | 1.983994978 | 0.13162245 | 0.868549649 | 1 | 0.131622448 | 0.131622448 | 2.596442171 | 0.765547792 | -0.116027692 | FALSE | TRUE | FALSE |
| PRLHR | 0.1007551 | 0.86172939 | 0.50785 | 0.100755138 | 0.100755138 | 0.497051004 | 0.03911859 | 0.889157165 | 1 | 0.039118591 | 0.039118591 | 0.772847215 | 2.575633092 | 0.410883996 | FALSE | TRUE | FALSE |
| MRGPRX1 | 0.1007248 | 0.86174076 | 1 | 0.100724794 | 0.100724794 | 0.100724794 | 0.1310928 | 0.868636222 | 1 | 0.131092798 | 0.131092798 | 0.131092798 | 0.768347274 | -0.114442445 | FALSE | TRUE | FALSE |
| HTRA2 | 0.1006886 | 0.86175212 | 1 | 0.100688619 | 0.100688619 | 1.98638652 | 0.13035774 | 0.868862949 | 1 | 0.52130022 | 0.52130022 | 2.575908092 | 0.772402305 | -0.112156439 | FALSE | TRUE | FALSE |
| TAS2R42 | 0.100638 | 0.86176348 | 0.8737 | 3.414456658 | 77.40453395 | 77.40453395 | 0.13892948 | 0.866973025 | 0.504 | 1.251354488 | 1.251354488 | 1.251354488 | 0.724381834 | -0.140032449 | FALSE | TRUE | FALSE |
| ETNPPL | 0.1005857 | 0.86177484 | 0.0124 | 0.100585746 | 0.100585746 | 1.993216617 | 0.52149106 | 0.746648612 | 0.50685 | 0.521491059 | 0.521491059 | 2.587323204 | 0.192881075 | -0.714710426 | FALSE | TRUE | FALSE |
| SOD1 | 0.1005588 | 0.8617862 | 1 | 0.100558752 | 0.100558752 | 0.100558752 | 0.5213606 | 0.746769877 | 0.5026 | 0.521360598 | 0.521360598 | 0.521360598 | 0.192877545 | -0.714718331 | FALSE | TRUE | FALSE |
| LZTS2 | 0.1004139 | 0.86179756 | 1 | 0.100413887 | 0.100413887 | 1.97426552 | 1.0491693 | 0.583396885 | 0.51375 | 1.049169298 | 1.049169298 | 20.67006983 | 0.095707992 | -1.019051793 | FALSE | FALSE | FALSE |
| OR4D9 | 0.1003712 | 0.86180891 | 1 | 0.900623138 | 0.900623138 | 0.900623138 | 1.01184977 | 0.58677815 | 0.9857 | 88.38961698 | 8524.635677 | 86755.14088 | 0.099195777 | -1.003506815 | FALSE | FALSE | FALSE |
| ENO2 | 0.1003384 | 0.86182027 | 1 | 1.604584922 | 6.419086026 | 7.945807555 | 0.1304018 | 0.868841388 | 1 | 1.21083499 | 4.841737246 | 4.841737246 | 0.769455987 | -0.113816217 | FALSE | TRUE | FALSE |
| FCGR2A | 0.1003051 | 0.86183162 | 1 | 0.100305136 | 0.100305136 | 0.100305136 | 0.11122399 | 0.873096713 | 1 | 0.111223985 | 0.111223985 | 0.111223985 | 0.901830082 | -0.044875282 | FALSE | TRUE | FALSE |
| TMEM98 | 0.1002592 | 0.86184297 | 0.0072 | 0.10025921 | 0.10025921 | 0.10025921 | 0.10388832 | 0.874947368 | 0.5013 | 0.975586803 | 0.975586803 | 0.975586803 | 0.965067204 | -0.015442443 | FALSE | TRUE | FALSE |
| ISM2 | 0.1002342 | 0.86185432 | 1 | 8.419305632 | 80.1812905 | 80.1812905 | 0.14329522 | 0.86600347 | 1 | 0.143295224 | 0.143295224 | 2.836428908 | 0.699494565 | -0.155215656 | FALSE | TRUE | FALSE |
| LMAN2L | 0.1000551 | 0.86186566 | 0.01105 | 0.100055068 | 0.100055068 | 1.984565321 | 0.13103983 | 0.868657848 | 1 | 0.131039834 | 0.131039834 | 0.131039834 | 0.763546965 | -0.117164245 | FALSE | TRUE | FALSE |
| PLEKHM1 | 0.0999241 | 0.86187701 | 1 | 0.399616656 | 0.399616656 | 7.796451558 | 0.13246761 | 0.86834362 | 1 | 0.132467614 | 0.132467614 | 2.592052654 | 0.754328533 | -0.122439465 | FALSE | TRUE | FALSE |
| HSPD1 | 0.0999156 | 0.86188835 | 1 | 0.099915576 | 0.099915576 | 1.977850345 | 0.12981699 | 0.869002931 | 1 | 0.518998351 | 0.518998351 | 2.572920491 | 0.769664852 | -0.113698346 | FALSE | TRUE | FALSE |
| RNF170 | 0.0998325 | 0.8618997 | 1 | 0.923415897 | 0.923415897 | 0.923415897 | 2.08733924 | 0.481688251 | 0.5096 | 2.087339243 | 8.350476039 | 8.350476039 | 0.047827658 | -1.320320883 | FALSE | FALSE | FALSE |
| RNF152 | 0.0998162 | 0.86191104 | 1 | 0.099816204 | 0.099816204 | 0.099816204 | 0.51826542 | 0.748015805 | 0.5136 | 0.518265418 | 0.518265418 | 0.518265418 | 0.19259669 | -0.715351182 | FALSE | TRUE | FALSE |
| OR4X1 | 0.0997832 | 0.86192237 | 1 | 0.896272028 | 0.896272028 | 0.896272028 | 0.13007905 | 0.868949126 | 1 | 1.175818122 | 1.175818122 | 1.175818122 | 0.767096313 | -0.115150105 | FALSE | TRUE | FALSE |
| SERPINB9 | 0.0996688 | 0.86193371 | 1 | 0.09966882 | 0.09966882 | 1.977702958 | 0.13091062 | 0.868733484 | 1 | 0.130910615 | 0.130910615 | 0.130910615 | 0.761350176 | -0.118415547 | FALSE | TRUE | FALSE |
| GIT2 | 0.0996418 | 0.86194505 | 0.0219 | 0.099641757 | 0.099641757 | 1.960053279 | 0.13094294 | 0.868711882 | 1 | 0.13094294 | 0.13094294 | 2.580879672 | 0.760955555 | -0.118640708 | FALSE | TRUE | FALSE |
| NDUFA13 | 0.0996104 | 0.86195638 | 1 | 0.915903904 | 0.915903904 | 0.915903904 | 0.53221328 | 0.741863548 | 0.5024 | 0.53221328 | 0.53221328 | 0.53221328 | 0.187162634 | -0.727780852 | FALSE | TRUE | FALSE |
| GRXCR2 | 0.099567 | 0.86196771 | 1 | 0.099566951 | 0.398233554 | 0.398233554 | 0.12745443 | 0.869549388 | 1 | 1.152740139 | 1.152740139 | 1.152740139 | 0.781196482 | -0.107239721 | FALSE | TRUE | FALSE |
| HMGCLL1 | 0.0995294 | 0.86197904 | 0.5043 | 0.099529423 | 0.397811458 | 0.397811458 | 0.17922362 | 0.857233253 | 0.691 | 0.179223624 | 2.864719374 | 2.864719374 | 0.555336517 | -0.255443768 | FALSE | TRUE | FALSE |
| ALKBH2 | 0.0995183 | 0.86199037 | 1 | 0.099518347 | 0.099518347 | 0.099518347 | 0.12587177 | 0.869932577 | 0.50405 | 1.134699529 | 1.134699529 | 1.134699529 | 0.790632793 | -0.102025177 | FALSE | TRUE | FALSE |
| NAV1 | 0.0994659 | 0.8620017 | 1 | 0.099465858 | 1.9072813 | 3.337351451 | 0.13070911 | 0.868787457 | 0.05005 | 0.130709106 | 2.518857742 | 4.392590264 | 0.760971146 | -0.11863181 | FALSE | TRUE | FALSE |
| SMPD3 | 0.099465 | 0.86201302 | 1 | 0.099465049 | 0.099465049 | 1.95520174 | 0.13052777 | 0.868830605 | 1 | 0.130527768 | 0.130527768 | 2.571102025 | 0.762022138 | -0.118032412 | FALSE | TRUE | FALSE |
| SNX4 | 0.0994602 | 0.86202434 | 1 | 0.099460214 | 0.898485668 | 1.939401498 | 0.13389602 | 0.86798433 | 0.01025 | 0.13389602 | 0.13389602 | 2.65991237 | 0.742816809 | -0.129118277 | FALSE | TRUE | FALSE |
| DENND5A | 0.0994461 | 0.86203566 | 1 | 0.099446136 | 0.099446136 | 1.939238774 | 0.13035633 | 0.868873727 | 1 | 0.130356331 | 0.130356331 | 2.549651886 | 0.762879217 | -0.117544216 | FALSE | TRUE | FALSE |
| RTKN | 0.099424 | 0.86204698 | 0.87785 | 8.397193701 | 80.19010682 | 80.19010682 | 0.14242208 | 0.866294605 | 1 | 0.142422085 | 0.142422085 | 2.815916684 | 0.698093841 | -0.156086193 | FALSE | TRUE | FALSE |
| PLXNC1 | 0.0993225 | 0.8620583 | 0.51775 | 0.397260451 | 0.397260451 | 7.702557716 | 0.13165628 | 0.868538821 | 1 | 0.131656283 | 0.131656283 | 2.562109575 | 0.754407936 | -0.122393752 | FALSE | TRUE | FALSE |
| MTTP | 0.0992652 | 0.86206962 | 1 | 0.0992652 | 0.0992652 | 1.952931168 | 0.12850728 | 0.86935701 | 1 | 0.513967427 | 0.513967427 | 10.1315799 | 0.772448039 | -0.112130725 | FALSE | TRUE | FALSE |
| PCDH18 | 0.099262 | 0.86208093 | 0.5132 | 0.924342498 | 0.924342498 | 18.10256887 | 0.1345222 | 0.867886022 | 1 | 0.134522199 | 0.134522199 | 2.641114401 | 0.737885885 | -0.132010797 | FALSE | TRUE | FALSE |
| B3GNT4 | 0.0992493 | 0.86209224 | 0.9848 | 7.840220568 | 683.9123718 | 6619.868174 | 0.17622692 | 0.858039185 | 0.6932 | 1.5673001 | 133.6973602 | 133.6973602 | 0.563190423 | -0.249344739 | FALSE | TRUE | FALSE |
| CXADR | 0.0992143 | 0.86210355 | 0.00985 | 0.099214342 | 0.099214342 | 0.099214342 | 0.13032627 | 0.868884503 | 1 | 0.130326273 | 0.130326273 | 2.590207544 | 0.761276601 | -0.118457519 | FALSE | TRUE | FALSE |
| ADAM22 | 0.0991835 | 0.86211486 | 1 | 0.904567934 | 0.904567934 | 17.7565277 | 0.13389057 | 0.867995245 | 0.02205 | 0.133890574 | 0.133890574 | 2.634108347 | 0.740780403 | -0.130310515 | FALSE | TRUE | FALSE |
| ST3GAL6 | 0.0991375 | 0.86212617 | 0.0088 | 0.099137456 | 0.099137456 | 0.099137456 | 0.12547569 | 0.870028042 | 1 | 1.131733456 | 1.131733456 | 1.131733456 | 0.790092915 | -0.102321833 | FALSE | TRUE | FALSE |
| TCL1A | 0.0990285 | 0.86213747 | 1 | 0.099028524 | 0.099028524 | 0.099028524 | 0.10575772 | 0.874462208 | 0.0039 | 0.105757719 | 0.105757719 | 0.105757719 | 0.936371596 | -0.028551769 | FALSE | TRUE | FALSE |
| EEPD1 | 0.098967 | 0.86214878 | 1 | 2.08532E+15 | 1.01564E+20 | 6.5818E+21 | 0.03679764 | 0.88971705 | 1 | 1.27006E+30 | 5.12285E+36 | 4.41496E+39 | 2.689492423 | 0.429670325 | FALSE | TRUE | FALSE |
| ABCA3 | 0.0989262 | 0.86216008 | 0.41555 | 0.098926233 | 6.302095886 | 100.6489281 | 0.05133305 | 0.886320312 | 0.5116 | 0.051333055 | 1.865637585 | 29.7895544 | 1.927144875 | 0.284914364 | FALSE | TRUE | FALSE |
| SH3TC2 | 0.0989182 | 0.86217138 | 0.54715 | 0.098918174 | 25.72168409 | 410.1582193 | 0.41829084 | 0.785265301 | 0.6843 | 2.285215952 | 1422.635139 | 17283.29308 | 0.236481808 | -0.626202264 | FALSE | TRUE | FALSE |
| LCA5 | 0.0988656 | 0.86218268 | 1 | 0.905656518 | 8.514881231 | 17.90474329 | 0.13779797 | 0.867227199 | 0.01525 | 0.137797971 | 0.137797971 | 2.72802356 | 0.717467406 | -0.144197824 | FALSE | TRUE | FALSE |
| GH1 | 0.098838 | 0.86219397 | 1 | 0.098837963 | 0.098837963 | 0.098837963 | 0.51363434 | 0.749998427 | 0.50475 | 0.513634336 | 0.513634336 | 0.513634336 | 0.192428652 | -0.715730263 | FALSE | TRUE | FALSE |
| USP47 | 0.0988042 | 0.86220527 | 1 | 0.098804237 | 0.098804237 | 3.325444031 | 1.14475491 | 0.572839011 | 0.63805 | 1.968492294 | 18.31206577 | 66.48384172 | 0.086310384 | -1.06393695 | FALSE | FALSE | FALSE |
| NMRK2 | 0.0987878 | 0.86221656 | 1 | 0.098787813 | 0.098787813 | 0.098787813 | 0.11787529 | 0.871745019 | 0.87535 | 9.897194589 | 94.23669666 | 94.23669666 | 0.838070574 | -0.076719408 | FALSE | TRUE | FALSE |
| SREBF1 | 0.0987796 | 0.86222785 | 0.5156 | 0.22155694 | 0.885935831 | 4.319221134 | 0.15191825 | 0.864127887 | 0.51465 | 0.342951662 | 1.371408033 | 6.706143819 | 0.650215464 | -0.186942706 | FALSE | TRUE | FALSE |
| PVR | 0.0987773 | 0.86223914 | 1 | 8.282258332 | 78.8064677 | 78.8064677 | 10.9500772 | 0.188430694 | 0.5138 | 10.95007723 | 104.7782667 | 104.7782667 | 0.00902069 | -2.044760224 | FALSE | FALSE | FALSE |

| | | | | | | | | | | | | | | | | | |
|---|---|---|---|---|---|---|---|---|---|---|---|---|---|---|---|---|---|
| FCRL2 | 0.0987511 | 0.86225043 | 1 | 0.098751103 | 0.098751103 | 1.957417526 | 0.12885139 | 0.86920702 | 1 | 0.128851393 | 0.128851393 | 2.556795413 | 0.766395308 | -0.115547163 | FALSE | TRUE | FALSE |
| NDST1 | 0.0986991 | 0.86226171 | 1 | 0.09869911 | 0.09869911 | 1.928342734 | 0.51332041 | 0.750194887 | 0.51125 | 0.513320408 | 0.513320408 | 10.05697992 | 0.192275835 | -0.716075295 | FALSE | TRUE | FALSE |
| DDHD2 | 0.0986493 | 0.862273 | 1 | 0.098649302 | 0.098649302 | 1.948645272 | 0.1251819 | 0.8701022 | 1 | 1.13807027 | 1.13807027 | 4.233238336 | 0.788047645 | -0.103447524 | FALSE | TRUE | FALSE |
| ADAM18 | 0.0986342 | 0.86228428 | 0.5039 | 0.881238631 | 8.075873058 | 8.075873058 | 0.01928126 | 0.894011466 | 0.87325 | 0.678295593 | 6.124757801 | 6.124757801 | 5.115549691 | 0.708892308 | FALSE | TRUE | FALSE |
| GALNT15 | 0.0986328 | 0.86229556 | 0.88065 | 0.394515715 | 1.578394726 | 3.336148008 | 0.13582346 | 0.867634182 | 0.9854 | 11.03746162 | 992.7827266 | 3971.123829 | 0.726184117 | -0.138953254 | FALSE | TRUE | FALSE |
| GNB5 | 0.0985698 | 0.86230684 | 1 | 0.098569843 | 0.098569843 | 1.952648948 | 0.12905579 | 0.869164103 | 1 | 0.129055788 | 0.129055788 | 2.559493835 | 0.763777006 | -0.11703342 | FALSE | TRUE | FALSE |
| BCAS3 | 0.098569 | 0.86231811 | 1 | 0.892859576 | 0.892859576 | 17.50433874 | 1.17459619 | 0.569852485 | 0.51365 | 1.174596188 | 1.174596188 | 23.08252874 | 0.083917384 | -1.076148061 | FALSE | FALSE | FALSE |
| CD5 | 0.09855 | 0.86232939 | 1 | 0.098550022 | 0.098550022 | 1.949800507 | 0.12868704 | 0.869324898 | 1 | 0.128687044 | 0.128687044 | 2.549352354 | 0.765811531 | -0.115878099 | FALSE | TRUE | FALSE |
| RUFY4 | 0.0985196 | 0.86234066 | 1 | 0.098519611 | 0.098519611 | 0.098519611 | 9.67780372 | 0.202684598 | 0.31325 | 9.67780372 | 907.8140462 | 907.8140462 | 0.010179955 | -1.992254123 | FALSE | FALSE | FALSE |
| PPARG | 0.0984448 | 0.86235193 | 0.013 | 0.098444829 | 0.098444829 | 1.950203006 | 0.12955179 | 0.869056696 | 0.0111 | 0.129551795 | 0.129551795 | 2.569366342 | 0.759887807 | -0.119250524 | FALSE | TRUE | FALSE |
| TSPO2 | 0.0984087 | 0.8623632 | 1 | 0.098408651 | 0.098408651 | 0.098408651 | 0.98804826 | 0.589779642 | 0.502 | 0.988048257 | 0.988048257 | 0.988048257 | 0.099599033 | -1.001744879 | FALSE | TRUE | FALSE |
| IFT74 | 0.0984066 | 0.86237447 | 0.5013 | 0.884211032 | 0.884211032 | 3.327466543 | 0.13246525 | 0.868354479 | 1 | 0.132465251 | 0.132465251 | 2.625986094 | 0.742886336 | -0.129077629 | FALSE | TRUE | FALSE |
| DHX37 | 0.0983962 | 0.86238573 | 1 | 0.098396165 | 0.098396165 | 1.906404252 | 0.11776595 | 0.871786367 | 0.8828 | 10.28070928 | 99.81081454 | 199.9374317 | 0.835523066 | -0.078041556 | FALSE | TRUE | FALSE |
| TOP1MT | 0.0983916 | 0.862397 | 0.26265 | 0.011732605 | 0.098391639 | 0.230783744 | 0.02207616 | 0.893202306 | 0.89265 | 1.635057913 | 133.7306802 | 1253.4274 | 4.456917385 | 0.649034584 | FALSE | TRUE | FALSE |
| ERC1 | 0.0983867 | 0.86240826 | 0.66445 | 8.614472377 | 83.68158849 | 832.7582054 | 0.02155325 | 0.893350275 | 1 | 0.021553248 | 0.021553248 | 0.422722033 | 4.564821242 | 0.659423775 | FALSE | TRUE | FALSE |
| DYRK3 | 0.0983685 | 0.86241952 | 1 | 0.898157448 | 0.898157448 | 3.330792662 | 0.12130838 | 0.870965065 | 1 | 10.43643186 | 100.5864478 | 100.5864478 | 0.810896446 | -0.091034603 | FALSE | TRUE | FALSE |
| POGK | 0.0983488 | 0.86243078 | 0.018 | 0.098348841 | 0.098348841 | 1.939376906 | 0.10594827 | 0.8743727 | 1 | 0.105948265 | 0.105948265 | 2.092760769 | 0.928272312 | -0.032324603 | FALSE | TRUE | FALSE |
| PIP5K1A | 0.0983349 | 0.86244203 | 0.01755 | 0.098334901 | 0.098334901 | 1.942830244 | 0.12858573 | 0.869335603 | 1 | 0.128585731 | 0.128585731 | 2.544214031 | 0.764741936 | -0.116485094 | FALSE | TRUE | FALSE |
| ACVR1 | 0.0982759 | 0.86245329 | 1 | 0.09827593 | 0.09827593 | 1.945076179 | 0.51117973 | 0.751133346 | 0.50605 | 0.511179733 | 0.511179733 | 2.533556083 | 0.192253181 | -0.716126466 | FALSE | TRUE | FALSE |
| UGT1A7 | 0.0982637 | 0.86246454 | 1 | 0.098263661 | 0.098263661 | 1.943358295 | 0.12857583 | 0.869346308 | 0.01315 | 0.128575834 | 0.128575834 | 2.546252814 | 0.76424673 | -0.116766411 | FALSE | TRUE | FALSE |
| ZFAT | 0.0982344 | 0.86247579 | 1 | 0.098234434 | 0.098234434 | 1.914395397 | 0.12873097 | 0.869303481 | 0.0298 | 0.128730972 | 0.128730972 | 2.516452195 | 0.763098679 | -0.117419298 | FALSE | TRUE | FALSE |
| KCNJ13 | 0.0982031 | 0.86248704 | 1 | 0.098203059 | 0.098203059 | 0.098203059 | 0.49923642 | 0.75534193 | 0.87615 | 1.996588989 | 7.986925164 | 7.986925164 | 0.19670652 | -0.706181245 | FALSE | TRUE | FALSE |
| MMAA | 0.0982015 | 0.86249829 | 0.50525 | 0.910883409 | 8.666096818 | 8.666096818 | 0.01901871 | 0.894091767 | 1 | 0.173006359 | 1.615251959 | 1.615251959 | 5.163417229 | 0.71293722 | FALSE | TRUE | FALSE |
| MICU3 | 0.0981936 | 0.86250953 | 1 | 0.392693707 | 0.392693707 | 1.946352612 | 0.13016664 | 0.868927592 | 0.01235 | 0.130166637 | 0.130166637 | 2.582876172 | 0.754368734 | -0.12241632 | FALSE | TRUE | FALSE |
| CCKAR | 0.0981851 | 0.86252078 | 1 | 0.098185081 | 0.098185081 | 1.942725004 | 0.12884631 | 0.869217745 | 0.0119 | 0.128846311 | 0.128846311 | 2.552675153 | 0.762032533 | -0.118026487 | FALSE | TRUE | FALSE |
| RPL38 | 0.0981576 | 0.86253202 | 1 | 0.09815759 | 0.09815759 | 0.09815759 | 0.09915041 | 0.876188647 | 0.00245 | 0.09915041 | 0.09915041 | 0.09915041 | 0.98998673 | -0.004370627 | FALSE | TRUE | FALSE |
| NCF4 | 0.0981239 | 0.86254326 | 1 | 0.098123858 | 0.098123858 | 1.946741801 | 0.12874213 | 0.869282058 | 1 | 0.128742127 | 0.128742127 | 0.128742127 | 0.762173655 | -0.117946067 | FALSE | TRUE | FALSE |
| NDUFB10 | 0.0980188 | 0.8625545 | 1 | 3.54186E+14 | 1.35715E+20 | 1.87293E+21 | 0.16867712 | 0.85995514 | 1 | 1701216213 | 3.44082E+13 | 5.50901E+15 | 0.581103318 | -0.235746645 | FALSE | TRUE | FALSE |
| RASSF1 | 0.0980174 | 0.86256574 | 1 | 0.098017398 | 0.098017398 | 0.116479767 | 0.12103029 | 0.871038231 | 0.75625 | 1.11600078 | 10.56154155 | 10.56154155 | 0.809858407 | -0.091590905 | FALSE | TRUE | FALSE |
| OR8H3 | 0.0979998 | 0.86257697 | 1 | 0.097999837 | 0.097999837 | 0.097999837 | 1.01797062 | 0.58655419 | 0.93725 | 9.371847439 | 856.6093416 | 856.6093416 | 0.096269809 | -1.016509891 | FALSE | FALSE | FALSE |
| HDAC9 | 0.0979709 | 0.8625882 | 1 | 0.097970853 | 0.097970853 | 1.921318779 | 0.12882696 | 0.869239189 | 0.02645 | 0.128826965 | 0.128826965 | 2.532357609 | 0.760484057 | -0.118909886 | FALSE | TRUE | FALSE |
| PRMT2 | 0.0979652 | 0.86259944 | 1 | 0.883583782 | 0.883583782 | 0.883583782 | 0.13204707 | 0.868462974 | 1 | 0.132047073 | 0.132047073 | 0.132047073 | 0.741896216 | -0.129656844 | FALSE | TRUE | FALSE |
| MBIP | 0.0979499 | 0.86261066 | 0.75415 | 0.879091412 | 8.097835787 | 8.097835787 | 0.13589995 | 0.867601269 | 1 | 0.135899948 | 0.135899948 | 0.135899948 | 0.720749742 | -0.142215504 | FALSE | TRUE | FALSE |
| XRN2 | 0.0978496 | 0.86262189 | 1 | 0.391335996 | 0.391335996 | 7.693695537 | 10.5683332 | 0.191826127 | 0.26025 | 1.121573421 | 10.5683332 | 22.09533767 | 0.009258756 | -2.033447362 | FALSE | FALSE | FALSE |
| NDRG4 | 0.0977783 | 0.86263312 | 1 | 0.097778305 | 0.097778305 | 1.939148377 | 0.12766086 | 0.869517362 | 1 | 0.127660859 | 0.127660859 | 0.127660859 | 0.765922349 | -0.115815258 | FALSE | TRUE | FALSE |
| AQP9 | 0.0977772 | 0.86264434 | 1 | 0.097777177 | 0.097777177 | 0.097777177 | 0.50834768 | 0.752450242 | 0.5046 | 0.508347681 | 0.508347681 | 0.508347681 | 0.192343117 | -0.715923351 | FALSE | TRUE | FALSE |
| CDK9 | 0.097777 | 0.86265557 | 1 | 0.097777042 | 0.097777042 | 1.936257784 | 0.12050075 | 0.871121756 | 1 | 1.09469106 | 10.20683586 | 10.20683586 | 0.811422719 | -0.090752837 | FALSE | TRUE | FALSE |
| CCKBR | 0.0977646 | 0.86266679 | 1 | 0.898656899 | 0.898656899 | 3.309654327 | 0.12840133 | 0.86937841 | 1 | 1.184712261 | 1.184712261 | 4.34906209 | 0.761398884 | -0.118387764 | FALSE | TRUE | FALSE |
| HEATR3 | 0.0977123 | 0.86267801 | 1 | 0.09771234 | 0.09771234 | 1.93142973 | 1.11950456 | 0.576022322 | 0.50915 | 1.119504558 | 1.119504558 | 2.453347314 | 0.08728177 | -1.059076454 | FALSE | FALSE | FALSE |
| GOLM1 | 0.0976347 | 0.86268922 | 1 | 0.097634672 | 0.097634672 | 1.935541469 | 0.12808366 | 0.869431884 | 0.0109 | 0.128083657 | 0.128083657 | 2.541854519 | 0.762272674 | -0.117889649 | FALSE | TRUE | FALSE |
| MAP3K4 | 0.0976174 | 0.86270044 | 1 | 0.390420563 | 0.390420563 | 7.568790665 | 1.16672299 | 0.57081212 | 0.04185 | 1.166722994 | 1.166722994 | 22.70208482 | 0.083668009 | -1.077440568 | FALSE | FALSE | FALSE |
| ARHGAP20 | 0.0975353 | 0.86271165 | 1 | 0.097535306 | 0.097535306 | 1.910327119 | 0.12780314 | 0.869453263 | 1 | 0.127803137 | 0.127803137 | 2.509393797 | 0.763168324 | -0.117379663 | FALSE | TRUE | FALSE |
| GTSE1 | 0.0975082 | 0.86272287 | 1 | 0.097508163 | 0.097508163 | 1.916834325 | 0.1275886 | 0.869538714 | 1 | 0.1275886 | 0.1275886 | 2.513321995 | 0.764238837 | -0.116770896 | FALSE | TRUE | FALSE |
| MOB3C | 0.0974661 | 0.86273408 | 0.0098 | 0.097466056 | 0.097466056 | 0.097466056 | 0.12818099 | 0.869421192 | 0.0079 | 0.128180987 | 0.128180987 | 0.128180987 | 0.76037842 | -0.118970217 | FALSE | TRUE | FALSE |
| ETAA1 | 0.0973383 | 0.86274529 | 0.5102 | 0.876690718 | 0.876690718 | 17.28837401 | 0.13128999 | 0.868592948 | 1 | 0.131289992 | 0.131289992 | 2.593405164 | 0.74139941 | -0.129947764 | FALSE | TRUE | FALSE |
| GOLIM4 | 0.0973004 | 0.86275649 | 1 | 0.097300391 | 0.097300391 | 1.920174103 | 0.12361247 | 0.870461283 | 0.5046 | 1.121300829 | 1.121300829 | 4.176113257 | 0.78714058 | -0.103947697 | FALSE | TRUE | FALSE |
| CTNNA2 | 0.0972955 | 0.8627677 | 1 | 0.097295462 | 0.097295462 | 1.905245307 | 0.12772298 | 0.86946395 | 0.02555 | 0.127722983 | 0.127722983 | 2.507373966 | 0.761769416 | -0.118176468 | FALSE | TRUE | FALSE |
| CEBPZ | 0.0972477 | 0.8627789 | 0.5084 | 0.388987244 | 0.388987244 | 7.63862902 | 0.50390334 | 0.753980848 | 0.7572 | 0.503903335 | 2.01609371 | 9.916873593 | 0.192988776 | -0.714467948 | FALSE | TRUE | FALSE |
| ATRAID | 0.0972147 | 0.86279011 | 0.00615 | 0.097214692 | 0.097214692 | 0.097214692 | 1.11846113 | 0.576256365 | 0.50425 | 1.118461129 | 1.118461129 | 1.118461129 | 0.086918257 | -1.060888991 | FALSE | FALSE | FALSE |
| PIP5K1B | 0.0971462 | 0.86280131 | 0.01445 | 0.097146196 | 0.097146196 | 1.923580631 | 1.99091202 | 0.489698317 | 0.01375 | 0.4976463 | 1.990912023 | 9.865637631 | 0.048794821 | -1.311626271 | FALSE | FALSE | FALSE |
| EARS2 | 0.0971398 | 0.8628125 | 1 | 0.097139837 | 0.097139837 | 1.918515293 | 0.12738867 | 0.86956006 | 0.0153 | 0.127388668 | 0.127388668 | 2.519714368 | 0.762546927 | -0.117733425 | FALSE | TRUE | FALSE |
| TRIM62 | 0.0971268 | 0.8628237 | 0.9852 | 78.71349507 | 7751.352737 | 79711.60807 | 0.0835101 | 0.879466965 | 1 | 672455782.4 | 1.31713E+13 | 1.63741E+14 | 1.163054184 | 0.065599948 | FALSE | TRUE | FALSE |
| MSANTD3 | 0.0970405 | 0.8628349 | 1 | 0.097040504 | 0.097040504 | 0.097040504 | 0.09951072 | 0.876052949 | 1 | 0.099510725 | 0.099510725 | 0.099510725 | 0.975176336 | -0.010916846 | FALSE | TRUE | FALSE |
| IRF2 | 0.0970201 | 0.86284609 | 0.75035 | 0.387834454 | 1.550741652 | 1.924370283 | 0.12651198 | 0.869751893 | 1 | 2.022418123 | 8.089159818 | 8.089159818 | 0.766885001 | -0.115269756 | FALSE | TRUE | FALSE |
| ZNF766 | 0.0969999 | 0.86285728 | 0.01105 | 0.096999865 | 0.096999865 | 1.924545104 | 0.12699073 | 0.869602735 | 1 | 0.126990727 | 0.126990727 | 2.521998193 | 0.763834242 | -0.117000876 | FALSE | TRUE | FALSE |
| TRIM69 | 0.0969938 | 0.86286847 | 0.01105 | 0.09699382 | 0.09699382 | 1.925073964 | 0.12736882 | 0.8695814 | 1 | 0.127368818 | 0.127368818 | 2.530257999 | 0.761519358 | -0.118319053 | FALSE | TRUE | FALSE |
| OSBPL5 | 0.096926 | 0.86287966 | 0.02665 | 0.096926013 | 0.096926013 | 1.897735854 | 0.49898946 | 0.75552974 | 0.26975 | 0.498989458 | 1.995920542 | 9.794582927 | 0.19424461 | -0.711651023 | FALSE | TRUE | FALSE |
| KLK4 | 0.0969115 | 0.86289085 | 1 | 0.096911482 | 0.096911482 | 0.096911482 | 0.12768025 | 0.869496003 | 0.0066 | 0.127680251 | 0.127680251 | 0.127680251 | 0.759017005 | -0.119748494 | FALSE | TRUE | FALSE |
| IFT80 | 0.0968923 | 0.86290203 | 0.5122 | 0.09689226 | 0.387558217 | 0.479108732 | 0.01114304 | 0.896415212 | 1 | 0.044551745 | 0.044551745 | 0.220637458 | 8.695316549 | 0.939285397 | FALSE | TRUE | FALSE |
| ARL15 | 0.0968838 | 0.86291322 | 1 | 0.387457658 | 1.549908478 | 1.549908478 | 0.12989776 | 0.868992173 | 0.00495 | 0.129897762 | 0.129897762 | 0.129897762 | 0.745846294 | -0.127350664 | FALSE | TRUE | FALSE |
| BMP1 | 0.0967466 | 0.8629244 | 0.9412 | 0.880854101 | 14.09055501 | 30.10071694 | 0.59188827 | 0.715369438 | 0.81755 | 2.367714277 | 26.42269996 | 105.6715841 | 0.16345423 | -0.786603837 | FALSE | TRUE | FALSE |
| ZNF793 | 0.0966847 | 0.86293558 | 1 | 0.09668468 | 0.09668468 | 1.918445358 | 0.11569428 | 0.872249828 | 0.8782 | 9.710083037 | 92.43613684 | 92.43613684 | 0.835691063 | -0.077954242 | FALSE | TRUE | FALSE |
| TSR3 | 0.0966684 | 0.86294675 | 1 | 0.096668373 | 0.096668373 | 0.096668373 | 0.12696828 | 0.8696134 | 1 | 0.126968283 | 0.126968283 | 0.126968283 | 0.761358436 | -0.118410836 | FALSE | TRUE | FALSE |
| XPNPEP1 | 0.0966164 | 0.86295793 | 0.7532 | 0.386308467 | 1.544992484 | 7.621521779 | 0.12873489 | 0.86929277 | 0.50025 | 0.514756999 | 0.514756999 | 2.562122864 | 0.750506361 | -0.124645622 | FALSE | TRUE | FALSE |
| ZNF20 | 0.0965083 | 0.86296911 | 0.0121 | 0.096508325 | 0.096508325 | 1.912489702 | 0.12665894 | 0.869730605 | 1 | 0.126658938 | 0.126658938 | 2.512744255 | 0.76195432 | -0.118071064 | FALSE | TRUE | FALSE |
| OR9G4 | 0.0964949 | 0.86298028 | 0.50555 | 0.865851297 | 0.865851297 | 0.865851297 | 0.11890451 | 0.871496395 | 0.87435 | 1.071951458 | 94.13350426 | 94.13350426 | 0.811532984 | -0.090693824 | FALSE | TRUE | FALSE |
| ARIH2 | 0.0964227 | 0.86299145 | 1 | 0.096422699 | 0.096422699 | 1.908547114 | 0.12679163 | 0.869688009 | 1 | 0.126791629 | 0.126791629 | 2.512772305 | 0.760481585 | -0.118911298 | FALSE | TRUE | FALSE |
| LTA4H | 0.0963196 | 0.86300262 | 1 | 0.096319554 | 0.096319554 | 1.904808699 | 0.12616445 | 0.869836981 | 0.01395 | 0.126164447 | 0.126164447 | 2.498382128 | 0.763444506 | -0.117222526 | FALSE | TRUE | FALSE |
| ETV3L | 0.0963104 | 0.86301379 | 0.4992 | 0.096310426 | 0.864674632 | 1.912019211 | 0.12992268 | 0.868981414 | 1 | 0.12992268 | 0.12992268 | 0.12992268 | 0.741290329 | -0.130011666 | FALSE | TRUE | FALSE |
| TERT | 0.0962965 | 0.86302496 | 0.51915 | 0.096296454 | 0.384812747 | 6.818062963 | 0.01858879 | 0.894179243 | 1 | 0.018588791 | 0.018588791 | 0.362720956 | 5.180350722 | 0.714359164 | FALSE | TRUE | FALSE |
| DCSTAMP | 0.0962641 | 0.86303612 | 0.0109 | 0.096264087 | 0.096264087 | 1.908774803 | 1.10292766 | 0.578235754 | 0.50375 | 1.102927662 | 1.102927662 | 2.427037948 | 0.087280508 | -1.059082733 | FALSE | FALSE | FALSE |
| AKNAD1 | 0.096262 | 0.86304728 | 0.87535 | 0.835104461 | 7.431328761 | 7.431328761 | 0.17054358 | 0.859465499 | 0.0185 | 0.170543575 | 0.170543575 | 0.672471511 | 0.564442435 | -0.248380343 | FALSE | TRUE | FALSE |
| FIP1L1 | 0.0962254 | 0.86305844 | 0.01835 | 0.096225355 | 0.096225355 | 1.900078194 | 0.12675757 | 0.86969866 | 1 | 0.126757572 | 0.126757572 | 2.506794294 | 0.759129047 | -0.119684391 | FALSE | TRUE | FALSE |

| | | | | | | | | | | | | | | | | | |
|---|---|---|---|---|---|---|---|---|---|---|---|---|---|---|---|---|---|
| MUC15 | 0.0961188 | 0.8630696 | 0.9847 | 73.8712423 | 7022.246595 | 70952.32165 | 0.12947568 | 0.869088935 | 1 | 10.75724927 | 989.2136447 | 9838.794465 | 0.742369265 | -0.129380017 | FALSE | TRUE | FALSE |
| GAS8 | 0.0961016 | 0.86308076 | 1 | 0.86768804 | 8.04079693 | 17.14544413 | 0.13388744 | 0.868017069 | 1 | 0.133887445 | 0.133887445 | 2.64942595 | 0.717778802 | -0.144009372 | FALSE | TRUE | FALSE |
| TBX4 | 0.0960808 | 0.86309192 | 1 | 0.09608082 | 0.09608082 | 1.900049554 | 0.12680125 | 0.869677356 | 0.01235 | 0.126801251 | 0.126801251 | 2.510948283 | 0.757727702 | -0.120486835 | FALSE | TRUE | FALSE |
| PBK | 0.0960461 | 0.86310307 | 1 | 0.867508839 | 8.042116085 | 8.042116085 | 1.18027361 | 0.569250656 | 0.50475 | 1.180273609 | 1.180273609 | 1.180273609 | 0.081376116 | -1.089503042 | FALSE | FALSE | FALSE |
| PDLIM3 | 0.0960331 | 0.86311423 | 1 | 0.096033138 | 0.096033138 | 1.90332775 | 0.12640873 | 0.869773175 | 1 | 0.126408732 | 0.126408732 | 0.126408732 | 0.759703354 | -0.119355956 | FALSE | TRUE | FALSE |
| RBAK | 0.0960231 | 0.86312538 | 1 | 0.096023078 | 0.096023078 | 1.896866781 | 0.12607573 | 0.869890107 | 1 | 0.126075728 | 0.126075728 | 2.494216957 | 0.761630166 | -0.118255863 | FALSE | TRUE | FALSE |
| N4BP2L1 | 0.0959984 | 0.86313653 | 0.0064 | 0.095998414 | 0.095998414 | 0.095998414 | 0.1075606 | 0.873893268 | 1 | 0.107560596 | 0.107560596 | 0.107560596 | 0.892505413 | -0.049389141 | FALSE | TRUE | FALSE |
| SYT8 | 0.095966 | 0.86314767 | 1 | 7.949402843 | 728.8530556 | 728.8530556 | 11.2393301 | 0.185040281 | 0.0108 | 1.209148346 | 11.23933013 | 23.94966605 | 0.008538413 | -2.068622842 | FALSE | FALSE | FALSE |
| LRRC23 | 0.0959323 | 0.86315882 | 1 | 0.095932266 | 0.095932266 | 1.905619879 | 92.0137465 | 0.031826365 | 0.00435 | 9.658715731 | 92.01374653 | 92.01374653 | 0.001042586 | -2.981888009 | FALSE | FALSE | FALSE |
| CTRL | 0.0959294 | 0.86316996 | 1 | 0.095929412 | 0.095929412 | 0.095929412 | 1.11848433 | 0.57613938 | 0.50955 | 1.11848433 | 1.11848433 | 1.11848433 | 0.085767327 | -1.066678123 | FALSE | FALSE | FALSE |
| EFR3A | 0.0958976 | 0.8631811 | 1 | 0.868877374 | 0.868877374 | 3.238611103 | 0.12977541 | 0.869013687 | 0.0185 | 0.129775413 | 0.129775413 | 2.560027699 | 0.738950199 | -0.13138483 | FALSE | TRUE | FALSE |
| ECHDC1 | 0.0958846 | 0.86319224 | 1 | 0.095884606 | 0.095884606 | 0.095884606 | 9.93465599 | 0.198958314 | 0.005 | 1.070161786 | 9.934655988 | 9.934655988 | 0.009651528 | -2.015403944 | FALSE | FALSE | FALSE |
| CCDC15 | 0.0958562 | 0.86320338 | 0.941 | 0.836734806 | 68.76468345 | 68.76468345 | 1.46403938 | 0.538969843 | 0.13735 | 1.46403938 | 13.28083746 | 28.898871 | 0.06547376 | -1.183932718 | FALSE | FALSE | FALSE |
| SLC7A5 | 0.0957225 | 0.86321452 | 0.75565 | 0.095722467 | 0.382625329 | 1.885435305 | 0.03643018 | 0.889803338 | 1 | 0.582052793 | 9.308882316 | 11.48554547 | 2.62755932 | 0.41955253 | FALSE | TRUE | FALSE |
| PCDHB13 | 0.0956984 | 0.86322565 | 0.2686 | 0.095698352 | 0.850085807 | 3.237335506 | 0.01959682 | 0.893931054 | 0.02055 | 0.019596823 | 0.019596823 | 0.384921831 | 4.883360651 | 0.688718799 | FALSE | TRUE | FALSE |
| WBP11 | 0.0956229 | 0.86323679 | 1 | 8801.33839 | 11151166.69 | 126174491.1 | 0.12620958 | 0.869815719 | 0.99895 | 11705.8123 | 14941283.63 | 169372156.8 | 0.757651459 | -0.120530536 | FALSE | TRUE | FALSE |
| COLEC12 | 0.0955884 | 0.86324792 | 1 | 0.095588448 | 0.095588448 | 1.883655161 | 0.12611901 | 0.869858237 | 1 | 0.126119012 | 0.126119012 | 2.4896858 | 0.757922586 | -0.120375151 | FALSE | TRUE | FALSE |
| MAPKAPK3 | 0.095399 | 0.86325905 | 1 | 0.864823733 | 8.046594013 | 8.046594013 | 37.539433 | 0.073189182 | 0.13095 | 1.106952121 | 99.16220739 | 99.16220739 | 0.002541301 | -2.594943854 | FALSE | FALSE | FALSE |
| C2CD2 | 0.0952933 | 0.86327018 | 1 | 0.866127352 | 0.866127352 | 3.225063476 | 0.12914243 | 0.869142634 | 0.01555 | 0.129142427 | 0.129142427 | 2.550117972 | 0.737893358 | -0.132006399 | FALSE | TRUE | FALSE |
| MAP7D1 | 0.0952702 | 0.86328131 | 1 | 0.878759926 | 8.319112303 | 17.26004475 | 0.12159569 | 0.870912756 | 0.02065 | 0.121595694 | 0.121595694 | 2.393461925 | 0.783500143 | -0.10596092 | FALSE | TRUE | FALSE |
| CTSG | 0.0952553 | 0.86329243 | 0.00995 | 0.095255258 | 0.095255258 | 0.095255258 | 0.49547956 | 0.756650372 | 0.509 | 0.495479556 | 0.495479556 | 0.495479556 | 0.192248615 | -0.716136781 | FALSE | TRUE | FALSE |
| DGKE | 0.0951653 | 0.86330356 | 1 | 0.095165338 | 0.095165338 | 1.879405146 | 1.116998 | 0.576723592 | 0.5066 | 1.116997996 | 1.116997996 | 4.11459842 | 0.085197411 | -1.0695736 | FALSE | FALSE | FALSE |
| PAN2 | 0.0951489 | 0.86331468 | 1 | 0.095148918 | 0.095148918 | 1.856947335 | 0.12540673 | 0.870038641 | 0.0285 | 0.125406729 | 0.125406729 | 2.45459896 | 0.758722584 | -0.119916989 | FALSE | TRUE | FALSE |
| KATNA1 | 0.0950568 | 0.8633258 | 0.96755 | 8.024735127 | 306.296315 | 1224.820503 | 0.01203581 | 0.896056851 | 0.9402 | 0.192266942 | 3.074453397 | 3.074453397 | 7.897832678 | 0.897507929 | FALSE | TRUE | FALSE |
| TIAM2 | 0.0950469 | 0.86333692 | 0.5198 | 0.095046924 | 0.828265012 | 3.189209371 | 0.00291996 | 0.898707409 | 0.75795 | 0.02455511 | 0.211948062 | 4.106136259 | 32.5508139 | 1.512561852 | FALSE | TRUE | FALSE |
| HYAL3 | 0.0950382 | 0.86334804 | 1 | 8.033179213 | 751.535096 | 751.535096 | 4.79810877 | 0.315339519 | 0.49885 | 2.603352436 | 45.00224312 | 45.00224312 | 0.019807438 | -1.703171692 | FALSE | FALSE | FALSE |
| KDM8 | 0.0950036 | 0.86335915 | 1 | 0.095003593 | 0.095003593 | 1.881245302 | 0.12460034 | 0.870197433 | 1 | 0.124600343 | 0.124600343 | 2.470255895 | 0.762466541 | -0.11777921 | FALSE | TRUE | FALSE |
| INHBE | 0.0949783 | 0.86337027 | 1 | 0.094978348 | 0.094978348 | 1.885812824 | 1.10874494 | 0.577422681 | 0.5069 | 1.108744938 | 1.108744938 | 1.108744938 | 0.085662938 | -1.067207037 | FALSE | FALSE | FALSE |
| METTL17 | 0.0949242 | 0.86338138 | 0.01445 | 0.094924156 | 0.094924156 | 1.877828673 | 0.12456464 | 0.870208007 | 1 | 0.124564637 | 0.124564637 | 2.467414031 | 0.762047383 | -0.118018024 | FALSE | TRUE | FALSE |
| OR6T1 | 0.0948658 | 0.86339249 | 0.5026 | 0.84782153 | 0.84782153 | 0.84782153 | 1.1181346 | 0.576606882 | 0.50275 | 1.118134604 | 1.118134604 | 1.118134604 | 0.084842957 | -1.071384204 | FALSE | FALSE | FALSE |
| XRCC1 | 0.094821 | 0.8634036 | 0.5066 | 0.094821033 | 0.094821033 | 1.867907456 | 0.14589539 | 0.865485429 | 0.0191 | 0.145895391 | 0.145895391 | 1.234220698 | 0.649924799 | -0.187136891 | FALSE | TRUE | FALSE |
| TATDN2 | 0.0947751 | 0.86341471 | 0.02165 | 0.094775064 | 0.094775064 | 1.864623141 | 0.12445843 | 0.870239718 | 1 | 0.124458426 | 0.124458426 | 2.453421197 | 0.76149978 | -0.118330218 | FALSE | TRUE | FALSE |
| PLEKHA8 | 0.0947285 | 0.86342581 | 1 | 0.094728456 | 0.094728456 | 1.878826419 | 0.12456379 | 0.870218579 | 1 | 0.124563792 | 0.124563792 | 0.124563792 | 0.760481472 | -0.118911362 | FALSE | TRUE | FALSE |
| NSUN6 | 0.0947221 | 0.86343692 | 0.75455 | 0.094722068 | 0.378843712 | 0.470105325 | 2.87408669 | 0.421273412 | 0.26 | 0.31382839 | 2.874086686 | 2.874086686 | 0.032957276 | -1.482048693 | FALSE | FALSE | FALSE |
| ABHD6 | 0.0946386 | 0.86344802 | 1 | 0.094638603 | 0.094638603 | 0.094638603 | 0.12493605 | 0.870144541 | 1 | 0.124936047 | 0.124936047 | 0.124936047 | 0.757496376 | -0.120619441 | FALSE | TRUE | FALSE |
| STAMBPL1 | 0.0946233 | 0.86345912 | 1 | 0.094623331 | 0.094623331 | 1.877546183 | 1.05925835 | 0.582373415 | 0.75285 | 1.059258348 | 9.843823819 | 9.843823819 | 0.089329795 | -1.049003662 | FALSE | FALSE | FALSE |
| PTGFRN | 0.0945985 | 0.86347022 | 1 | 0.857017001 | 0.857017001 | 16.75326868 | 0.12821259 | 0.869399805 | 1 | 0.128212586 | 0.128212586 | 2.513163178 | 0.737825713 | -0.132046214 | FALSE | TRUE | FALSE |
| MGAT4B | 0.0945457 | 0.86348132 | 1 | 0.094545745 | 0.094545745 | 1.860275204 | 0.12295176 | 0.870629621 | 1 | 0.491761261 | 0.491761261 | 4.164460656 | 0.768966179 | -0.114092761 | FALSE | TRUE | FALSE |
| LRRC45 | 0.09445 | 0.86349241 | 1 | 0.847912098 | 0.847912098 | 1.866057246 | 0.12393281 | 0.870398051 | 1 | 1.119723809 | 1.119723809 | 4.197785743 | 0.762106475 | -0.117984348 | FALSE | TRUE | FALSE |
| DIP2B | 0.0943772 | 0.8635035 | 1 | 0.094377153 | 0.094377153 | 3.16716594 | 1.10795585 | 0.577539008 | 0.0383 | 1.10795585 | 1.10795585 | 21.55192277 | 0.08518133 | -1.069655584 | FALSE | FALSE | FALSE |
| BTBD1 | 0.0943299 | 0.8635146 | 1 | 0.377217198 | 0.377217198 | 1.869761593 | 0.12574453 | 0.869953803 | 1 | 0.125744533 | 0.125744533 | 2.49512166 | 0.750171268 | -0.124839574 | FALSE | TRUE | FALSE |
| KLK15 | 0.0942537 | 0.86352569 | 0.5028 | 0.871428273 | 0.871428273 | 0.871428273 | 10.2888617 | 0.19555941 | 0.508 | 10.28886173 | 100.2684822 | 100.2684822 | 0.00916075 | -2.038068962 | FALSE | FALSE | FALSE |
| FCGRT | 0.0942323 | 0.86353678 | 1 | 0.094232324 | 0.094232324 | 1.868353995 | 0.92265345 | 0.595889261 | 0.5044 | 0.922653446 | 0.922653446 | 0.922653446 | 0.102131872 | -0.990838708 | FALSE | TRUE | FALSE |
| CDC42EP1 | 0.0942161 | 0.86354786 | 1 | 0.094216065 | 0.094216065 | 1.864853701 | 1.11269836 | 0.577189848 | 0.01235 | 1.112698356 | 1.112698356 | 2.388597489 | 0.084673501 | -1.072252482 | FALSE | FALSE | FALSE |
| CWF19L1 | 0.0942156 | 0.86355895 | 0.0152 | 0.094215629 | 0.094215629 | 1.866359793 | 0.12385646 | 0.870408593 | 1 | 0.12385646 | 0.12385646 | 2.45634008 | 0.760684009 | -0.118795713 | FALSE | TRUE | FALSE |
| TDO2 | 0.0942001 | 0.86357003 | 0.81705 | 1.506907613 | 6.029301171 | 24.12991591 | 169.024515 | 0.015982242 | 0.44575 | 169.0245152 | 25579.39699 | 102402.4084 | 0.000557316 | -3.253898227 | FALSE | FALSE | FALSE |
| FAM3B | 0.0941994 | 0.86358112 | 0.74985 | 0.846624247 | 7.809754832 | 7.809754832 | 1.11838585 | 0.576373276 | 0.7504 | 1.118385854 | 10.38138133 | 10.38138133 | 0.084227987 | -1.074543579 | FALSE | FALSE | FALSE |
| PSD3 | 0.0941047 | 0.8635922 | 0.51355 | 0.376347972 | 0.376347972 | 7.378069292 | 1.12154037 | 0.57578804 | 0.5103 | 1.121540374 | 1.121540374 | 22.03948394 | 0.083906662 | -1.076203555 | FALSE | FALSE | FALSE |
| CXCL12 | 0.0941008 | 0.86360327 | 0.0026 | 0.094100797 | 0.094100797 | 0.094100797 | 0.09997548 | 0.875916963 | 1 | 0.099975478 | 0.099975478 | 0.099975478 | 0.941238782 | -0.026300187 | FALSE | TRUE | FALSE |
| PDE2A | 0.0940502 | 0.86361435 | 0.0275 | 0.094050169 | 0.094050169 | 1.842492829 | 0.12409375 | 0.870334762 | 0.02175 | 0.124093747 | 0.124093747 | 2.437057102 | 0.757896115 | -0.120390319 | FALSE | TRUE | FALSE |
| NCOA3 | 0.0940364 | 0.86362543 | 1 | 0.094036382 | 0.094036382 | 3.157856421 | 1.0984292 | 0.578930227 | 0.5151 | 1.098429197 | 1.098429197 | 21.48155969 | 0.085609872 | -1.067476155 | FALSE | FALSE | FALSE |
| ITCH | 0.093981 | 0.8636365 | 1 | 8.175092186 | 787.0362658 | 787.0362658 | 0.13454877 | 0.867875091 | 1 | 1.246471631 | 4.984804765 | 24.58547154 | 0.69849045 | -0.155839528 | FALSE | TRUE | FALSE |
| SERPINA9 | 0.0939538 | 0.86364757 | 0.26035 | 0.093953845 | 0.859095653 | 0.859095653 | 0.01900781 | 0.894099062 | 1 | 0.019007813 | 0.019007813 | 0.019007813 | 4.942906659 | 0.693982409 | FALSE | TRUE | FALSE |
| GNA12 | 0.0939324 | 0.86365864 | 1 | 0.093932411 | 0.093932411 | 1.861050145 | 0.12429273 | 0.870303095 | 0.0111 | 0.124292734 | 0.124292734 | 2.465343256 | 0.755735335 | -0.121630272 | FALSE | TRUE | FALSE |
| PTS | 0.0939311 | 0.86366971 | 1 | 0.375542841 | 0.375542841 | 0.375542841 | 0.12547726 | 0.870017441 | 1 | 0.125477263 | 0.125477263 | 0.125477263 | 0.74859024 | -0.12575584 | FALSE | TRUE | FALSE |
| OR2T3 | 0.0938976 | 0.86368078 | 0.74745 | 0.884354291 | 8.548362084 | 8.548362084 | 0.10596058 | 0.874362747 | 0.00715 | 0.105960579 | 0.105960579 | 0.105960579 | 0.886156055 | -0.052489791 | FALSE | TRUE | FALSE |
| COX10 | 0.0938564 | 0.86369184 | 1 | 0.375224866 | 1.500471705 | 1.8711516 | 0.42459905 | 0.782887407 | 0.99225 | 152.2625588 | 15110.00092 | 39036.72477 | 0.221047168 | -0.655515045 | FALSE | TRUE | FALSE |
| POLM | 0.0938445 | 0.8637029 | 0.87875 | 0.202937691 | 7.206091339 | 7.206091339 | 0.03802665 | 0.889441693 | 0.5091 | 0.15209814 | 0.15209814 | 0.752716955 | 2.467861792 | 0.392320834 | FALSE | TRUE | FALSE |
| RSPH4A | 0.0937649 | 0.86371396 | 0.02145 | 0.09376487 | 0.09376487 | 1.850246737 | 0.01834474 | 0.894237489 | 0.5088 | 0.157355611 | 0.157355611 | 0.620277964 | 5.11126856 | 0.708528701 | FALSE | TRUE | FALSE |
| PPFIA2 | 0.0936643 | 0.86372502 | 0.03225 | 0.093664319 | 0.093664319 | 3.148434828 | 0.12363786 | 0.870450748 | 0.0318 | 0.123637859 | 0.123637859 | 4.161642224 | 0.757569893 | -0.120577293 | FALSE | TRUE | FALSE |
| LRRC37A | 0.0935553 | 0.86373608 | 1 | 0.093555338 | 0.093555338 | 3.142402185 | 0.08483495 | 0.879171245 | 1 | 7.666176655 | 766.6810066 | 1474.272757 | 1.10279241 | 0.042493769 | FALSE | TRUE | FALSE |
| APMAP | 0.0935133 | 0.86374714 | 1 | 0.093513334 | 0.093513334 | 1.852597675 | 1.05308344 | 0.583169811 | 0.7555 | 1.05308344 | 9.822627923 | 9.822627923 | 0.088799548 | -1.051589244 | FALSE | FALSE | FALSE |
| ZNF112 | 0.0935015 | 0.86375819 | 1 | 0.093501514 | 0.093501514 | 1.841850404 | 0.10268053 | 0.875291778 | 1 | 82.59286774 | 8092.71094 | 83013.32649 | 0.910606057 | -0.040669465 | FALSE | TRUE | FALSE |
| TMPRSS11E | 0.093471 | 0.86376925 | 1 | 0.093470994 | 0.093470994 | 1.855042283 | 93.5265493 | 0.031283294 | 0.0035 | 9.69212567 | 93.52654925 | 93.52654925 | 0.000999406 | -3.00025805 | FALSE | FALSE | FALSE |
| PRRT3 | 0.0933935 | 0.8637803 | 1 | 0.093393516 | 0.093393516 | 1.838397595 | 0.11914684 | 0.871434097 | 1 | 1.077861579 | 1.077861579 | 4.029186876 | 0.783852217 | -0.105765809 | FALSE | TRUE | FALSE |
| VPS4A | 0.0933935 | 0.86379135 | 0.5043 | 0.373374784 | 0.373374784 | 1.850812653 | 0.48877275 | 0.75922398 | 0.25955 | 0.488772752 | 1.954652422 | 2.425356134 | 0.191077567 | -0.718790297 | FALSE | TRUE | FALSE |
| CYFIP1 | 0.0933453 | 0.8638024 | 1 | 0.851033264 | 0.851033264 | 16.4825243 | 0.12686939 | 0.869634725 | 1 | 0.126869394 | 0.126869394 | 2.466532887 | 0.735758841 | -0.133264511 | FALSE | TRUE | FALSE |
| SLC39A11 | 0.0933365 | 0.86381344 | 0.00965 | 0.093336506 | 0.093336506 | 0.093336506 | 0.12316799 | 0.870534982 | 1 | 0.123167995 | 0.123167995 | 0.123167995 | 0.757798376 | -0.12044633 | FALSE | TRUE | FALSE |
| TRAPPC2L | 0.0933174 | 0.86382449 | 0.50105 | 0.854917993 | 0.854917993 | 0.854917993 | 0.12313635 | 0.870545505 | 1 | 1.132771326 | 1.132771326 | 1.132771326 | 0.757838315 | -0.120423441 | FALSE | TRUE | FALSE |
| URI1 | 0.0932727 | 0.86383553 | 1 | 0.842479777 | 0.842479777 | 3.155766094 | 1.11515416 | 0.576956835 | 0.50475 | 1.115154161 | 1.115154161 | 2.432902223 | 0.083641086 | -1.077580339 | FALSE | FALSE | FALSE |
| DDX50 | 0.0932501 | 0.86384657 | 1 | 0.093250061 | 0.093250061 | 1.839978752 | 0.12275722 | 0.87067164 | 1 | 0.122757225 | 0.122757225 | 2.42611495 | 0.759629926 | -0.119397935 | FALSE | TRUE | FALSE |
| CLRN2 | 0.0931677 | 0.86385761 | 1 | 0.093167716 | 0.093167716 | 0.093167716 | 0.12311559 | 0.870566544 | 1 | 0.123115589 | 0.123115589 | 0.123115589 | 0.75674995 | -0.121047599 | FALSE | TRUE | FALSE |
| ZNF569 | 0.0930874 | 0.86386865 | 1 | 0.093087417 | 0.093087417 | 1.842081586 | 0.12274571 | 0.87068214 | 1 | 0.122745711 | 0.122745711 | 2.432068186 | 0.758376126 | -0.120115347 | FALSE | TRUE | FALSE |

| | | | | | | | | | | | | | | | | | |
|---|---|---|---|---|---|---|---|---|---|---|---|---|---|---|---|---|---|
| OSGEP | 0.0930809 | 0.86387969 | 1 | 0.093080928 | 0.093080928 | 1.846099815 | 1.09704057 | 0.579276595 | 0.502 | 1.097040573 | 1.097040573 | 1.097040573 | 0.084847297 | -1.071361987 | FALSE | FALSE | FALSE |
| GCLC | 0.0930149 | 0.86389072 | 1 | 0.093014895 | 0.093014895 | 1.837727254 | 1.10593426 | 0.577655291 | 0.5093 | 1.105934257 | 1.105934257 | 2.359716959 | 0.084105266 | -1.075176809 | FALSE | FALSE | FALSE |
| PNPO | 0.0929635 | 0.86390176 | 1 | 0.09296348 | 0.09296348 | 0.09296348 | 0.12297407 | 0.870608602 | 0.0056 | 0.122974066 | 0.122974066 | 0.122974066 | 0.755960038 | -0.121501162 | FALSE | TRUE | FALSE |
| NCSTN | 0.0929629 | 0.86391279 | 1 | 0.092962879 | 0.092962879 | 1.832171811 | 0.12313444 | 0.870556025 | 1 | 0.123134444 | 0.123134444 | 2.431067337 | 0.754970545 | -0.122069992 | FALSE | TRUE | FALSE |
| ZNF419 | 0.092905 | 0.86392382 | 0.01355 | 0.092904962 | 0.092904962 | 0.214909078 | 0.01809976 | 0.894288408 | 1 | 0.156900892 | 0.156900892 | 0.35919834 | 5.132939032 | 0.710366106 | FALSE | TRUE | FALSE |
| FGFR2 | 0.0927435 | 0.86393485 | 0.81635 | 0.370557418 | 14.16946327 | 14.16946327 | 0.07382289 | 0.881634015 | 0.51105 | 0.073822891 | 0.073822891 | 1.453248411 | 1.256297307 | 0.099092429 | FALSE | TRUE | FALSE |
| LRCH4 | 0.0926726 | 0.86394587 | 0.0223 | 0.092672555 | 0.092672555 | 1.819288977 | 0.4793738 | 0.763224923 | 0.75245 | 0.479373798 | 1.917451966 | 9.431649163 | 0.193320026 | -0.713723155 | FALSE | TRUE | FALSE |
| GGT6 | 0.0926458 | 0.8639569 | 0.5003 | 0.83023241 | 0.83023241 | 0.83023241 | 1.09839516 | 0.579045743 | 0.50455 | 1.098395163 | 1.098395163 | 2.418800056 | 0.084346499 | -1.073932937 | FALSE | FALSE | FALSE |
| PIK3R4 | 0.0925743 | 0.86396792 | 0.03615 | 0.092574295 | 0.092574295 | 3.113812485 | 0.12236228 | 0.87077657 | 1 | 0.122362277 | 0.122362277 | 2.392320121 | 0.756559106 | -0.121157137 | FALSE | TRUE | FALSE |
| SLC2A6 | 0.0925538 | 0.86397894 | 0.50995 | 0.092553779 | 0.092553779 | 0.784620596 | 1.26644797 | 0.55978541 | 0.25835 | 0.31676941 | 1.266447966 | 2.757235464 | 0.073081391 | -1.136193197 | FALSE | FALSE | FALSE |
| RNF31 | 0.0925217 | 0.86398996 | 1 | 0.092521732 | 0.092521732 | 3.115540139 | 0.48439103 | 0.761313477 | 0.51665 | 0.48439103 | 0.48439103 | 9.473739502 | 0.191006286 | -0.71895234 | FALSE | TRUE | FALSE |
| PDE7A | 0.0925009 | 0.86400098 | 1 | 0.850108634 | 0.850108634 | 1.832095244 | 69.8032799 | 0.041258376 | 0.00515 | 4.361887146 | 69.8032799 | 69.8032799 | 0.001325166 | -2.877729675 | FALSE | FALSE | FALSE |
| GABBR2 | 0.0924655 | 0.864012 | 0.02885 | 0.092465509 | 0.092465509 | 1.808780565 | 0.47933878 | 0.763260021 | 0.76035 | 0.47933878 | 1.916994285 | 9.40142597 | 0.192902209 | -0.7146628 | FALSE | TRUE | FALSE |
| PARP12 | 0.0924612 | 0.86402301 | 0.0167 | 0.092461167 | 0.092461167 | 1.825960357 | 0.11817475 | 0.871662244 | 0.51095 | 1.07174643 | 1.07174643 | 3.998765655 | 0.782410507 | -0.106565326 | FALSE | TRUE | FALSE |
| SAP130 | 0.0924507 | 0.86403403 | 1 | 0.092450713 | 0.092450713 | 1.809460704 | 0.10514792 | 0.874690379 | 1 | 0.105147916 | 0.105147916 | 2.063275285 | 0.879244369 | -0.055890405 | FALSE | TRUE | FALSE |
| MRPL39 | 0.0924368 | 0.86404504 | 1 | 0.828064947 | 0.828064947 | 1.835586933 | 1.09587066 | 0.579391931 | 0.5055 | 1.095870662 | 1.095870662 | 1.095870662 | 0.084350067 | -1.073914567 | FALSE | FALSE | FALSE |
| CHST2 | 0.0924274 | 0.86405605 | 1 | 0.092427419 | 0.092427419 | 1.822024232 | 0.12253423 | 0.870745108 | 1 | 0.122534232 | 0.122534232 | 2.419692005 | 0.754298753 | -0.12245661 | FALSE | TRUE | FALSE |
| GSTM2 | 0.0924173 | 0.86406706 | 0.50325 | 0.843999127 | 0.843999127 | 0.843999127 | 0.10982855 | 0.873400315 | 0.50455 | 1.007516686 | 1.007516686 | 1.007516686 | 0.841469055 | -0.07496185 | FALSE | TRUE | FALSE |
| C14orf28 | 0.0924086 | 0.86407806 | 1 | 0.823894906 | 0.823894906 | 0.823894906 | 1.0895442 | 0.57996786 | 0.4934 | 0.121333156 | 1.089544195 | 1.089544195 | 0.084813964 | -1.071532638 | FALSE | FALSE | FALSE |
| SLC25A39 | 0.0923708 | 0.86408907 | 0.7512 | 0.835674139 | 13.36462838 | 13.36462838 | 0.59145481 | 0.715679343 | 0.5103 | 0.591454806 | 2.365738738 | 2.365738738 | 0.156175575 | -0.806386885 | FALSE | TRUE | FALSE |
| SERPINH1 | 0.092313 | 0.86410007 | 0.50485 | 0.092312995 | 0.092312995 | 0.45930834 | 0.03647269 | 0.889787659 | 1 | 0.036472693 | 0.036472693 | 0.721170746 | 2.531016707 | 0.403295012 | FALSE | TRUE | FALSE |
| DMC1 | 0.092254 | 0.86411107 | 1 | 7.842856376 | 75.14184313 | 75.14184313 | 0.12942731 | 0.869099678 | 1 | 0.129427313 | 1.185791885 | 1.185791885 | 0.712786551 | -0.147040503 | FALSE | TRUE | FALSE |
| IYD | 0.0922355 | 0.86412207 | 1 | 0.092235532 | 0.092235532 | 0.092235532 | 0.12207907 | 0.87080802 | 0.0063 | 0.122079072 | 0.122079072 | 0.122079072 | 0.755539258 | -0.121742965 | FALSE | TRUE | FALSE |
| RNF175 | 0.0921197 | 0.86413307 | 0.74465 | 0.092119685 | 0.809465115 | 0.809465115 | 0.15564865 | 0.863095386 | 0.74925 | 0.155648648 | 1.383557385 | 1.383557385 | 0.591843784 | -0.227792909 | FALSE | TRUE | FALSE |
| GRIN2A | 0.0920622 | 0.86414407 | 1 | 0.092062234 | 0.092062234 | 3.099660518 | 4.39971439 | 0.335879884 | 0.2743 | 1.100274018 | 4.399714392 | 85.40564732 | 0.020924593 | -1.679342976 | FALSE | FALSE | FALSE |
| ZNF280D | 0.0920023 | 0.86415506 | 0.0262 | 0.092002316 | 0.092002316 | 1.810383485 | 0.10984915 | 0.873370017 | 0.02095 | 0.109849149 | 0.109849149 | 2.165760761 | 0.837533257 | -0.076997939 | FALSE | TRUE | FALSE |
| TMEM39B | 0.0919889 | 0.86416605 | 1 | 8.09255E+18 | 4.39966E+24 | 9.72386E+26 | 0.17438905 | 0.858476218 | 1 | 3.06775E+12 | 7.76753E+16 | 1.38223E+19 | 0.527492276 | -0.277783895 | FALSE | TRUE | FALSE |
| IFI6 | 0.0919775 | 0.86417704 | 1 | 0.091977542 | 0.091977542 | 0.091977542 | 0.09707902 | 0.87669024 | 1 | 0.09707902 | 0.09707902 | 0.09707902 | 0.947450253 | -0.023443584 | FALSE | TRUE | FALSE |
| KRTAP5-10 | 0.0918689 | 0.86418803 | 0.996 | 62.49335015 | 5461.304889 | 52868.05895 | 0.16732807 | 0.860344527 | 0.9706 | 1.458936726 | 119.6190084 | 1123.482289 | 0.549034312 | -0.260400513 | FALSE | TRUE | FALSE |
| ARHGAP29 | 0.0918086 | 0.86419902 | 1 | 0.091808599 | 0.091808599 | 1.796836256 | 0.12124741 | 0.870985978 | 0.0276 | 0.121247411 | 0.121247411 | 2.379122742 | 0.757200496 | -0.12078911 | FALSE | TRUE | FALSE |
| FCGR3B | 0.0916758 | 0.86421001 | 0.5033 | 0.854475259 | 0.854475259 | 0.854475259 | 0.10064718 | 0.875761209 | 1 | 0.940144837 | 0.940144837 | 0.940144837 | 0.910863342 | -0.040546776 | FALSE | TRUE | FALSE |
| CREB3L4 | 0.0916755 | 0.864221 | 1 | 0.366598042 | 0.366598042 | 1.819064882 | 1.0914777 | 0.579737654 | 0.5056 | 1.091477699 | 1.091477699 | 1.091477699 | 0.083992083 | -1.075761647 | FALSE | FALSE | FALSE |
| PSME4 | 0.0916726 | 0.86423198 | 0.04485 | 0.091672617 | 0.091672617 | 3.068097093 | 0.12010718 | 0.871215605 | 0.518 | 0.480364963 | 0.480364963 | 9.359780544 | 0.763256758 | -0.117329342 | FALSE | TRUE | FALSE |
| NLRC3 | 0.0916511 | 0.86424296 | 0.78725 | 0.790960075 | 28.02254967 | 112.1071563 | 0.02935134 | 0.89142004 | 0.5078 | 0.029351336 | 0.029351336 | 0.574368693 | 3.122551663 | 0.494509633 | FALSE | TRUE | FALSE |
| SLK | 0.0915999 | 0.86425394 | 1 | 0.091599857 | 0.091599857 | 3.081438701 | 0.12133003 | 0.870954607 | 0.0278 | 0.121330031 | 0.121330031 | 2.380926178 | 0.75496443 | -0.12207351 | FALSE | TRUE | FALSE |
| TPM4 | 0.0915326 | 0.86426492 | 0.00575 | 0.091532596 | 0.091532596 | 0.091532596 | 0.12142713 | 0.870933685 | 1 | 0.121427128 | 0.121427128 | 0.121427128 | 0.753806806 | -0.122739945 | FALSE | TRUE | FALSE |
| TMED9 | 0.0914578 | 0.8642759 | 1 | 0.091457841 | 0.091457841 | 0.091457841 | 0.09877121 | 0.876266065 | 1 | 0.09877121 | 0.09877121 | 0.09877121 | 0.925956467 | -0.033409431 | FALSE | TRUE | FALSE |
| HOXC9 | 0.0914329 | 0.86428687 | 0.5008 | 0.365495179 | 0.365495179 | 1.814074432 | 0.47990986 | 0.763049269 | 0.7566 | 0.479909864 | 1.919018052 | 1.919018052 | 0.190520901 | -0.720057373 | FALSE | TRUE | FALSE |
| BDH2 | 0.0914203 | 0.86429784 | 1 | 0.091420254 | 0.091420254 | 0.091420254 | 1.0722598 | 0.580772299 | 0.5035 | 1.072259802 | 1.072259802 | 1.072259802 | 0.085259425 | -1.069257602 | FALSE | FALSE | FALSE |
| MAMDC2 | 0.0913599 | 0.86430882 | 1 | 0.091359861 | 0.091359861 | 1.806052395 | 0.12048432 | 0.87113219 | 1 | 0.120484322 | 0.120484322 | 2.385117042 | 0.758271772 | -0.120175111 | FALSE | TRUE | FALSE |
| ZNF227 | 0.0913454 | 0.86431979 | 0.8753 | 0.80388205 | 7.256602525 | 15.84768448 | 0.15012009 | 0.864518383 | 0.93765 | 1.335099691 | 113.8870868 | 113.8870868 | 0.60848211 | -0.215752186 | FALSE | TRUE | FALSE |
| FBXO6 | 0.0911563 | 0.86433076 | 1 | 0.091156313 | 0.091156313 | 0.091156313 | 1.06133871 | 0.582145356 | 0.50745 | 1.061338708 | 1.061338708 | 1.061338708 | 0.085888051 | -1.06606725 | FALSE | FALSE | FALSE |
| STK3 | 0.0910999 | 0.86434172 | 0.7524 | 0.821661159 | 7.606246416 | 16.28964752 | 0.12767422 | 0.869506683 | 0.01065 | 0.127674216 | 0.127674216 | 2.533834494 | 0.713533979 | -0.14658534 | FALSE | TRUE | FALSE |
| KPNA5 | 0.0909933 | 0.86435269 | 1 | 0.835325859 | 0.835325859 | 1.803910243 | 0.12426448 | 0.870313652 | 1 | 0.124264481 | 0.124264481 | 2.46609841 | 0.732254777 | -0.135337786 | FALSE | TRUE | FALSE |
| ADAM7 | 0.0909684 | 0.86436365 | 0.9682 | 7.194627437 | 66.42776922 | 628.3432687 | 431.645428 | 0.005443642 | 0.378 | 107.9279498 | 10084.38792 | 40331.35018 | 0.000210748 | -3.676236658 | FALSE | FALSE | FALSE |
| DNAJA4 | 0.0909454 | 0.86437462 | 1 | 0.83273785 | 0.83273785 | 1.802893846 | 0.12038376 | 0.871153053 | 0.50635 | 1.106919683 | 1.106919683 | 2.389005225 | 0.75546208 | -0.12178733 | FALSE | TRUE | FALSE |
| SSRP1 | 0.0909265 | 0.86438558 | 1 | 0.854324972 | 0.854324972 | 16.81790811 | 0.12078747 | 0.871080005 | 1 | 1.136331363 | 1.136331363 | 4.087018298 | 0.752781154 | -0.123331262 | FALSE | TRUE | FALSE |
| GATA4 | 0.0909048 | 0.86439654 | 0.01105 | 0.090904757 | 0.090904757 | 0.451646912 | 0.0356203 | 0.88997555 | 1 | 0.142342145 | 0.142342145 | 0.142342145 | 2.552049035 | 0.406889015 | FALSE | TRUE | FALSE |
| ZGPAT | 0.0908898 | 0.86440749 | 0.5109 | 0.363518841 | 0.363518841 | 3.077406564 | 0.12141191 | 0.870944146 | 0.0159 | 0.12141191 | 0.12141191 | 2.397549019 | 0.748606892 | -0.125746179 | FALSE | TRUE | FALSE |
| OR51D1 | 0.0908881 | 0.86441845 | 1 | 0.81476586 | 7.4965928 | 7.4965928 | 0.12339537 | 0.870503406 | 1 | 1.11442448 | 1.11442448 | 1.11442448 | 0.73655968 | -0.132792059 | FALSE | TRUE | FALSE |
| TRAM1L1 | 0.0908843 | 0.8644294 | 0.50365 | 0.829337823 | 0.829337823 | 0.829337823 | 0.12396278 | 0.870376961 | 1 | 0.123962776 | 0.123962776 | 0.123962776 | 0.73315775 | -0.13480257 | FALSE | TRUE | FALSE |
| CREB3L3 | 0.0908411 | 0.86444036 | 0.878 | 0.821270176 | 30.44676882 | 30.44676882 | 1.38920144 | 0.546102229 | 0.3197 | 1.389201444 | 124.3260753 | 124.3260753 | 0.06539089 | -1.184482749 | FALSE | FALSE | FALSE |
| UFSP2 | 0.0905448 | 0.86445131 | 1 | 0.090544831 | 0.090544831 | 1.795203945 | 0.11841148 | 0.87162082 | 1 | 0.473560243 | 0.473560243 | 0.492326182 | 0.764662584 | -0.11653016 | FALSE | TRUE | FALSE |
| ANKFN1 | 0.0905418 | 0.86446226 | 1 | 7.581109343 | 72.0845554 | 149.442791 | 0.1160213 | 0.872208757 | 0.93745 | 9.808816324 | 917.8238806 | 917.8238806 | 0.780389578 | -0.107688539 | FALSE | TRUE | FALSE |
| IRF4 | 0.090508 | 0.86447321 | 1 | 0.090508005 | 0.090508005 | 1.790839506 | 0.11636621 | 0.872095688 | 1 | 1.860264071 | 7.440656687 | 9.216656767 | 0.777785964 | -0.109139898 | FALSE | TRUE | FALSE |
| CC2D1A | 0.0904411 | 0.86448415 | 0.031 | 0.09044115 | 0.09044115 | 1.765248684 | 0.11965371 | 0.871278102 | 0.0286 | 0.119653711 | 0.119653711 | 2.342204324 | 0.755857461 | -0.121560095 | FALSE | TRUE | FALSE |
| TRPV4 | 0.0904186 | 0.8644951 | 0.5162 | 0.115252513 | 7.350244552 | 29.38118114 | 0.00074754 | 0.899620938 | 0.8143 | 0.007036887 | 0.06790085 | 0.671208264 | 120.9556499 | 2.082626159 | FALSE | TRUE | FALSE |
| KBTBD7 | 0.0903377 | 0.86450604 | 1 | 0.090337685 | 0.090337685 | 1.77779198 | 1.06843727 | 0.581459921 | 0.0174 | 1.068437266 | 1.068437266 | 3.92779787 | 0.08455123 | -1.072880072 | FALSE | FALSE | FALSE |
| FAM222A | 0.0903239 | 0.86451698 | 1 | 0.090323885 | 0.090323885 | 1.784585885 | 0.11908454 | 0.871454869 | 1 | 0.119084541 | 0.119084541 | 2.356258937 | 0.758485391 | -0.12005278 | FALSE | TRUE | FALSE |
| CHRNA3 | 0.0903167 | 0.86452792 | 0.9383 | 5.769788373 | 214.1059053 | 1816.308468 | 0.16806 | 0.860089232 | 0.2595 | 0.168060002 | 1.569426349 | 1.569426349 | 0.53740725 | -0.269696479 | FALSE | TRUE | FALSE |
| LYRM4 | 0.0902987 | 0.86453886 | 0.0011 | 0.09029875 | 0.09029875 | 0.09029875 | 0.09674983 | 0.876767072 | 1 | 0.096749831 | 0.096749831 | 0.096749831 | 0.93332204 | -0.029968478 | FALSE | TRUE | FALSE |
| OCLN | 0.090275 | 0.8645498 | 1 | 0.360968542 | 1.443708579 | 3.057202757 | 0.12249246 | 0.870755597 | 0.01105 | 0.122492464 | 0.122492464 | 2.429604252 | 0.736984514 | -0.132541638 | FALSE | TRUE | FALSE |
| SLC6A11 | 0.0902154 | 0.86456073 | 1 | 0.360693734 | 0.360693734 | 3.053592904 | 0.47401964 | 0.765483803 | 0.75725 | 0.474019643 | 1.895767037 | 9.362559382 | 0.190319995 | -0.720515583 | FALSE | TRUE | FALSE |
| U2AF1L4 | 0.0901598 | 0.86457166 | 0.00675 | 0.090159772 | 0.090159772 | 0.090159772 | 0.11891916 | 0.871486016 | 1 | 0.118919161 | 0.118919161 | 0.118919161 | 0.758160178 | -0.120239031 | FALSE | TRUE | FALSE |
| TFAP2A | 0.0900757 | 0.86458259 | 0.5044 | 0.360062681 | 0.360062681 | 1.780699504 | 0.47911076 | 0.763365249 | 0.50965 | 0.479110757 | 0.479110757 | 2.372562161 | 0.188006046 | -0.725828185 | FALSE | TRUE | FALSE |
| NEXN | 0.09007 | 0.86459352 | 0.01855 | 0.090070027 | 0.090070027 | 0.760566012 | 0.03410815 | 0.890334065 | 1 | 0.300218844 | 0.300218844 | 0.675255824 | 2.640718922 | 0.421722177 | FALSE | TRUE | FALSE |
| PGBD2 | 0.090067 | 0.86460445 | 1 | 7.448691877 | 70.38958789 | 70.38958789 | 0.13007505 | 0.86895989 | 0.0135 | 0.130075048 | 0.130075048 | 2.573633512 | 0.692423063 | -0.159628475 | FALSE | TRUE | FALSE |
| HPX | 0.090002 | 0.86461538 | 1 | 0.805422747 | 0.805422747 | 3.049536829 | 1.06919441 | 0.581345478 | 0.50065 | 1.069194413 | 1.069194413 | 2.351961439 | 0.084177372 | -1.074804637 | FALSE | FALSE | FALSE |
| MGME1 | 0.0899032 | 0.8646263 | 0.50385 | 0.813461426 | 0.813461426 | 0.813461426 | 90.5695088 | 0.032550207 | 0.12535 | 9.472819901 | 90.56950875 | 90.56950875 | 0.000992644 | -3.003206663 | FALSE | FALSE | FALSE |
| ACTG2 | 0.0898103 | 0.86463723 | 1 | 0.35903345 | 0.35903345 | 1.781397191 | 0.11944665 | 0.871350945 | 1 | 0.477542956 | 0.477542956 | 0.477542956 | 0.751886121 | -0.123847932 | FALSE | TRUE | FALSE |
| PIK3R2 | 0.0897327 | 0.86464815 | 1 | 0.358807012 | 0.358807012 | 7.066562039 | 0.47658384 | 0.764446901 | 0.51005 | 0.476583843 | 0.476583843 | 4.033936933 | 0.188283097 | -0.725188667 | FALSE | TRUE | FALSE |
| LIM2 | 0.0896493 | 0.86465907 | 1 | 0.089649259 | 0.089649259 | 0.089649259 | 0.11924285 | 0.871413319 | 1 | 0.11924285 | 0.11924285 | 0.11924285 | 0.751820838 | -0.123885641 | FALSE | TRUE | FALSE |
| PRR22 | 0.0896411 | 0.86466999 | 0.00905 | 0.089641095 | 0.089641095 | 0.089641095 | 0.11460627 | 0.872475295 | 1 | 0.114606268 | 1.028014262 | 1.028014262 | 0.782165731 | -0.106701215 | FALSE | TRUE | FALSE |

| | | | | | | | | | | | | | | | | | |
|---|---|---|---|---|---|---|---|---|---|---|---|---|---|---|---|---|---|
| PITPNM3 | 0.0895953 | 0.86468091 | 0.8802 | 7.947289896 | 77.77429572 | 154.7049397 | 0.12284273 | 0.870661137 | 0.76155 | 1.145880965 | 10.97027793 | 38.72663146 | 0.729349354 | -0.137064397 | FALSE | TRUE | FALSE |
| UNKL | 0.0895136 | 0.86469182 | 0.0091 | 0.089513603 | 0.089513603 | 0.089513603 | 1.02766143 | 0.585656693 | 0.7536 | 1.027661429 | 9.704954476 | 9.704954476 | 0.087104177 | -1.059961016 | FALSE | FALSE | FALSE |
| SPARCL1 | 0.089501 | 0.86470274 | 1 | 0.089500974 | 0.089500974 | 1.770607842 | 0.11827664 | 0.871641535 | 1 | 0.11827664 | 0.11827664 | 2.342932662 | 0.7567088 | -0.121071216 | FALSE | TRUE | FALSE |
| RCC1 | 0.0894905 | 0.86471365 | 1 | 0.830062078 | 0.830062078 | 1.772962406 | 0.12271659 | 0.870724125 | 1 | 0.122716589 | 0.122716589 | 2.433982834 | 0.729244935 | -0.137126578 | FALSE | TRUE | FALSE |
| ACOT11 | 0.089479 | 0.86472456 | 0.6913 | 0.196390018 | 7.06548352 | 7.06548352 | 0.0413807 | 0.888640043 | 0.89445 | 3.327894533 | 295.517739 | 2885.974298 | 2.162335829 | 0.334923144 | FALSE | TRUE | FALSE |
| RNF14 | 0.089476 | 0.86473547 | 1 | 0.089476003 | 0.089476003 | 1.774887545 | 0.11897343 | 0.871465253 | 1 | 0.118973434 | 0.118973434 | 0.118973434 | 0.752067075 | -0.123743424 | FALSE | TRUE | FALSE |
| MRPL50 | 0.0894662 | 0.86474638 | 1 | 0.838957461 | 0.838957461 | 0.838957461 | 0.09874146 | 0.876304738 | 1 | 0.098741456 | 0.098741456 | 0.098741456 | 0.906064757 | -0.042840762 | FALSE | TRUE | FALSE |
| RICTOR | 0.0894588 | 0.86475728 | 1 | 0.829319898 | 3.316757334 | 64.35258953 | 0.12402209 | 0.870366414 | 1 | 0.12402209 | 0.12402209 | 4.15893765 | 0.721313695 | -0.141875822 | FALSE | TRUE | FALSE |
| WFDC9 | 0.0894104 | 0.86476818 | 1 | 0.089410427 | 0.089410427 | 0.089410427 | 0.09242818 | 0.877653884 | 1 | 0.092428184 | 0.092428184 | 0.092428184 | 0.967350258 | -0.014416248 | FALSE | TRUE | FALSE |
| REN | 0.0893984 | 0.86477909 | 1 | 0.089398439 | 0.089398439 | 1.771389376 | 1.04403334 | 0.583510406 | 0.50405 | 1.044033343 | 1.044033343 | 2.2767532 | 0.085627954 | -1.067384431 | FALSE | FALSE | FALSE |
| DCK | 0.0893507 | 0.86478999 | 1 | 0.089350747 | 0.089350747 | 0.089350747 | 9.87889467 | 0.199976665 | 0.2534 | 1.036455841 | 9.878894672 | 9.878894672 | 0.00904461 | -2.043610169 | FALSE | FALSE | FALSE |
| GDPD3 | 0.0893485 | 0.86480088 | 1 | 0.797246193 | 0.797246193 | 1.772862039 | 1.05860022 | 0.582487359 | 0.505 | 1.058600222 | 1.058600222 | 1.058600222 | 0.084402502 | -1.073644681 | FALSE | FALSE | FALSE |
| EFNB3 | 0.0893202 | 0.86481178 | 1 | 0.089320215 | 0.089320215 | 1.771841511 | 4.18664496 | 0.344577537 | 0.3733 | 1.04709277 | 16.74386319 | 16.74386319 | 0.021334557 | -1.670916374 | FALSE | FALSE | FALSE |
| CDC7 | 0.0892937 | 0.86482268 | 1 | 0.089293677 | 0.089293677 | 1.768390197 | 0.11437119 | 0.87251621 | 0.5081 | 1.029590493 | 1.029590493 | 2.267697611 | 0.780735757 | -0.10749593 | FALSE | TRUE | FALSE |
| RETNLB | 0.0892817 | 0.86483357 | 1 | 0.089281684 | 0.089281684 | 0.089281684 | 0.09748697 | 0.876594074 | 1 | 0.097486969 | 0.097486969 | 0.097486969 | 0.915831977 | -0.038184197 | FALSE | TRUE | FALSE |
| FCRL4 | 0.0892801 | 0.86484446 | 1 | 0.089280075 | 0.089280075 | 1.76829547 | 1.03140226 | 0.585206604 | 0.50475 | 1.031402261 | 1.031402261 | 2.268642255 | 0.086561837 | -1.062673534 | FALSE | FALSE | FALSE |
| ACTL9 | 0.0892626 | 0.86485535 | 0.7475 | 0.089262638 | 0.356838979 | 1.761551316 | 0.03577277 | 0.889944274 | 1 | 0.035772768 | 0.035772768 | 0.707084656 | 2.495267836 | 0.397117169 | FALSE | TRUE | FALSE |
| TBC1D9 | 0.0892464 | 0.86486624 | 1 | 0.089246433 | 0.089246433 | 3.005653124 | 0.1186417 | 0.871558636 | 0.0342 | 0.118641697 | 0.118641697 | 2.316162219 | 0.752234967 | -0.123646483 | FALSE | TRUE | FALSE |
| MROH1 | 0.0892099 | 0.86487712 | 0.8803 | 1.427132544 | 13.21004495 | 64.10395188 | 0.12506178 | 0.870123374 | 1 | 0.125061781 | 0.125061781 | 2.436256414 | 0.713326411 | -0.146711696 | FALSE | TRUE | FALSE |
| ALG9 | 0.0891858 | 0.86488801 | 1 | 0.089185824 | 0.089185824 | 1.769694601 | 0.11485278 | 0.872424117 | 1 | 1.060524186 | 1.060524186 | 2.281152825 | 0.77652298 | -0.109845688 | FALSE | TRUE | FALSE |
| LOX | 0.0891421 | 0.86489889 | 1 | 0.089142099 | 0.089142099 | 1.763908867 | 0.09621953 | 0.876872575 | 0.0123 | 0.096219528 | 0.096219528 | 1.906386585 | 0.926444988 | -0.033180364 | FALSE | TRUE | FALSE |
| HACE1 | 0.0890992 | 0.86490977 | 0.0221 | 0.089099187 | 0.089099187 | 1.75476292 | 0.11800259 | 0.87171399 | 0.01895 | 0.118002594 | 0.118002594 | 2.328273779 | 0.755061252 | -0.122017816 | FALSE | TRUE | FALSE |
| ARHGAP33 | 0.0890668 | 0.86492065 | 1 | 0.089066795 | 0.089066795 | 1.739664734 | 0.11794982 | 0.871724334 | 1 | 0.11794982 | 0.11794982 | 2.310305178 | 0.755124463 | -0.12198146 | FALSE | TRUE | FALSE |
| NFXL1 | 0.0890485 | 0.86493153 | 1 | 0.08904847 | 0.08904847 | 1.753687682 | 0.46732495 | 0.768001195 | 0.5131 | 0.467324945 | 0.467324945 | 3.945251883 | 0.190549361 | -0.719992503 | FALSE | TRUE | FALSE |
| UIMC1 | 0.0890317 | 0.86494241 | 0.0182 | 0.089031742 | 0.089031742 | 1.758432524 | 0.11760251 | 0.87182769 | 1 | 0.117602508 | 0.117602508 | 2.326207785 | 0.757056491 | -0.120871712 | FALSE | TRUE | FALSE |
| ZNF329 | 0.0888751 | 0.86495328 | 1 | 0.088875141 | 0.088875141 | 1.760499347 | 0.11774062 | 0.8717967 | 1 | 0.117740615 | 0.117740615 | 2.334969893 | 0.75483843 | -0.122145998 | FALSE | TRUE | FALSE |
| EFCAB3 | 0.0888744 | 0.86496415 | 1 | 0.355445556 | 0.355445556 | 1.765456256 | 0.11896405 | 0.871475635 | 1 | 0.118964046 | 0.118964046 | 0.141425395 | 0.747069465 | -0.126639014 | FALSE | TRUE | FALSE |
| FAM114A1 | 0.0888176 | 0.86497503 | 1 | 0.088817609 | 0.088817609 | 1.757641351 | 1.02694748 | 0.585769063 | 0.51075 | 1.02694748 | 1.02694748 | 2.256562885 | 0.086487002 | -1.063049157 | FALSE | FALSE | FALSE |
| MDH1B | 0.0887598 | 0.8649859 | 1 | 0.7981876 | 0.7981876 | 1.75738993 | 0.11737786 | 0.871879309 | 1 | 1.062109387 | 1.062109387 | 2.326814332 | 0.756188529 | -0.121369915 | FALSE | TRUE | FALSE |
| ZNF526 | 0.0887423 | 0.86499676 | 1 | 7.486632494 | 699.0635002 | 699.0635002 | 0.12883804 | 0.869228468 | 0.51075 | 1.177414056 | 1.177414056 | 23.17934841 | 0.688789893 | -0.161913234 | FALSE | TRUE | FALSE |
| PLK3 | 0.0887105 | 0.86500763 | 0.02005 | 0.088710495 | 0.088710495 | 1.748761403 | 0.11753835 | 0.871838017 | 0.01655 | 0.117538353 | 0.117538353 | 2.321048542 | 0.754736585 | -0.122204598 | FALSE | TRUE | FALSE |
| SHD | 0.0886474 | 0.8650185 | 1 | 0.803894338 | 0.803894338 | 1.756728714 | 0.12109452 | 0.871027783 | 0.00955 | 0.121094519 | 0.121094519 | 0.121094519 | 0.732051628 | -0.135458289 | FALSE | TRUE | FALSE |
| AUP1 | 0.0885847 | 0.86502936 | 1 | 0.354161724 | 0.354161724 | 1.755608017 | 0.11926452 | 0.871392534 | 0.0119 | 0.119264521 | 0.119264521 | 2.366215028 | 0.742758004 | -0.129152659 | FALSE | TRUE | FALSE |
| CTSV | 0.0885764 | 0.86504022 | 1 | 0.354197752 | 0.354197752 | 0.354197752 | 0.470242 | 0.766919521 | 0.0082 | 0.470241999 | 0.470241999 | 0.470241999 | 0.188363372 | -0.725003543 | FALSE | TRUE | FALSE |
| ZEB1 | 0.0885226 | 0.86505108 | 1 | 0.817249784 | 7.743705163 | 27.56048869 | 0.99137878 | 0.589226708 | 0.96945 | 91.3964982 | 910.9869223 | 9301.882385 | 0.089292419 | -1.04918541 | FALSE | TRUE | FALSE |
| APTX | 0.0884626 | 0.86506194 | 0.879 | 0.809358874 | 7.595286311 | 7.595286311 | 0.16205718 | 0.86154809 | 0.5062 | 0.162057179 | 0.162057179 | 0.162057179 | 0.545872687 | -0.262908635 | FALSE | TRUE | FALSE |
| GMFG | 0.0884537 | 0.8650728 | 0.0053 | 0.08845367 | 0.08845367 | 0.08845367 | 1.04022422 | 0.583737335 | 0.49865 | 0.113800816 | 1.040224218 | 1.040224218 | 0.085033273 | -1.070411104 | FALSE | FALSE | FALSE |
| ZNF200 | 0.0884212 | 0.86508365 | 0.7496 | 0.796967125 | 7.372714917 | 7.372714917 | 0.12432012 | 0.870271414 | 0.01 | 0.124320123 | 0.124320123 | 0.147782858 | 0.711237925 | -0.147985094 | FALSE | TRUE | FALSE |
| ART3 | 0.0884035 | 0.86509451 | 1 | 0.78895382 | 0.78895382 | 0.78895382 | 0.11671933 | 0.872054526 | 1 | 1.049021555 | 1.049021555 | 1.049021555 | 0.757402155 | -0.120673464 | FALSE | TRUE | FALSE |
| CELA3A | 0.0883741 | 0.86510536 | 0.98345 | 1.413136332 | 22.61919666 | 90.5287297 | 0.13587398 | 0.867612242 | 0.93735 | 1.237302917 | 46.24118812 | 46.24118812 | 0.650412214 | -0.186811312 | FALSE | TRUE | FALSE |
| TPR | 0.0883596 | 0.86511621 | 1 | 0.088359596 | 1.694668147 | 2.944372866 | 0.11749243 | 0.871858666 | 1 | 0.117492429 | 2.264578911 | 3.924549034 | 0.752045023 | -0.123756159 | FALSE | TRUE | FALSE |
| SYNDIG1 | 0.0883355 | 0.86512706 | 1 | 0.088335495 | 0.088335495 | 0.088335495 | 0.46540301 | 0.768872377 | 0.00595 | 0.465403014 | 0.465403014 | 0.465403014 | 0.189804303 | -0.721693945 | FALSE | TRUE | FALSE |
| WDR25 | 0.0882914 | 0.8651379 | 1 | 0.088291443 | 0.088291443 | 1.743279989 | 1.03282861 | 0.584868459 | 0.51065 | 1.032828611 | 1.032828611 | 2.245032433 | 0.085485087 | -1.068109644 | FALSE | FALSE | FALSE |
| ERBB4 | 0.0882331 | 0.86514875 | 0.88025 | 0.352757073 | 3.282021884 | 15.99764638 | 0.14286943 | 0.866171586 | 0.7553 | 0.142869427 | 0.571244194 | 4.812969886 | 0.617578483 | -0.209307844 | FALSE | TRUE | FALSE |
| MMS19 | 0.0881979 | 0.86515959 | 1 | 0.088197922 | 0.088197922 | 1.728943047 | 0.45781014 | 0.771933236 | 0.76075 | 0.45781014 | 1.83150389 | 8.99590246 | 0.192651745 | -0.715227053 | FALSE | TRUE | FALSE |
| CLSPN | 0.0880752 | 0.86517043 | 1 | 0.088075154 | 0.088075154 | 2.962180004 | 9.39528637 | 0.2067399 | 0.26755 | 1.003398734 | 9.395286373 | 33.79061267 | 0.009374398 | -2.02805661 | FALSE | FALSE | FALSE |
| CAMTA1 | 0.0880145 | 0.86518127 | 1 | 0.088014505 | 1.690379117 | 2.953707116 | 0.11717185 | 0.871899947 | 1 | 0.117171854 | 0.117171854 | 3.938340764 | 0.751157404 | -0.124269047 | FALSE | TRUE | FALSE |
| HYLS1 | 0.0879047 | 0.86519211 | 1 | 0.087904651 | 0.087904651 | 0.087904651 | 0.1168276 | 0.872023639 | 0.0087 | 0.116827596 | 0.116827596 | 0.116827596 | 0.752430541 | -0.123533585 | FALSE | TRUE | FALSE |
| SLC10A5 | 0.0878008 | 0.86520295 | 1 | 0.08780085 | 0.08780085 | 0.08780085 | 0.11609458 | 0.872177939 | 1 | 0.116094578 | 0.116094578 | 0.116094578 | 0.756287256 | -0.121313218 | FALSE | TRUE | FALSE |
| METTL22 | 0.0877574 | 0.86521379 | 1 | 0.789160314 | 3.15639564 | 6.951519655 | 3.832327 | 0.364784549 | 0.8169 | 35.49720272 | 821.105051 | 3284.194774 | 0.022899246 | -1.640178819 | FALSE | FALSE | FALSE |
| SYNPO2 | 0.0877462 | 0.86522462 | 1 | 0.087746191 | 0.087746191 | 1.718123884 | 0.11639509 | 0.8720854 | 1 | 0.116395093 | 0.116395093 | 2.284843217 | 0.753865036 | -0.122706398 | FALSE | TRUE | FALSE |
| BARHL1 | 0.0877044 | 0.86523546 | 0.0129 | 0.087704449 | 0.087704449 | 1.735140874 | 0.11700825 | 0.871961822 | 1 | 0.117008246 | 0.117008246 | 2.317894115 | 0.749557846 | -0.125194846 | FALSE | TRUE | FALSE |
| PAPOLB | 0.0877043 | 0.86524629 | 1 | 0.350722314 | 0.350722314 | 2.968357616 | 38.0144151 | 0.07179942 | 0.1291 | 4.030926833 | 38.01441509 | 38.01441509 | 0.002307132 | -2.636927473 | FALSE | FALSE | FALSE |
| ASPRV1 | 0.0876835 | 0.86525712 | 1 | 0.087683515 | 0.087683515 | 1.741304695 | 0.10111368 | 0.87566368 | 1 | 0.928637213 | 0.928637213 | 0.928637213 | 0.867177586 | -0.061891956 | FALSE | TRUE | FALSE |
| TMEM123 | 0.0875264 | 0.86526795 | 1 | 0.087526392 | 0.087526392 | 0.087526392 | 0.09178761 | 0.877776917 | 1 | 0.091787613 | 0.091787613 | 0.091787613 | 0.95357521 | -0.020645048 | FALSE | TRUE | FALSE |
| SEZ6 | 0.0874543 | 0.86527877 | 1 | 0.087454318 | 0.087454318 | 1.712639127 | 0.11622387 | 0.872147107 | 0.0255 | 0.116223873 | 0.116223873 | 2.281752802 | 0.752464322 | -0.123514087 | FALSE | TRUE | FALSE |
| DNAJB12 | 0.0872394 | 0.8652896 | 1 | 0.797230176 | 0.797230176 | 1.727528318 | 0.1196083 | 0.871298923 | 0.01245 | 0.119608304 | 0.119608304 | 2.37132109 | 0.729375653 | -0.137048738 | FALSE | TRUE | FALSE |
| MSX1 | 0.087237 | 0.86530042 | 1 | 0.348723912 | 0.348723912 | 0.348723912 | 0.46554111 | 0.768772209 | 0.50735 | 0.46554111 | 0.46554111 | 0.46554111 | 0.187388467 | -0.727257142 | FALSE | TRUE | FALSE |
| TF | 0.0872189 | 0.86531125 | 0.01935 | 0.08721892 | 0.08721892 | 1.719494685 | 0.11606725 | 0.872198486 | 0.01785 | 0.116067253 | 0.116067253 | 2.292159317 | 0.75145158 | -0.124098998 | FALSE | TRUE | FALSE |
| CCDC137 | 0.0871517 | 0.86532207 | 1 | 0.087151709 | 0.087151709 | 0.087151709 | 1.0329593 | 0.584755618 | 0.009 | 1.032959298 | 1.032959298 | 1.032959298 | 0.084370904 | -1.0738073 | FALSE | FALSE | FALSE |
| THPO | 0.0871179 | 0.86533289 | 0.0099 | 0.087117867 | 0.087117867 | 0.087117867 | 1.00211004 | 0.588339262 | 0.51035 | 1.002110044 | 1.002110044 | 1.002110044 | 0.086934431 | -1.060808184 | FALSE | FALSE | FALSE |
| SIDT2 | 0.0871109 | 0.8653437 | 1 | 0.785546222 | 7.270661895 | 15.37953813 | 0.12273136 | 0.870703136 | 0.0253 | 0.12273136 | 0.12273136 | 2.408957127 | 0.709768602 | -0.148883216 | FALSE | TRUE | FALSE |
| RXFP1 | 0.0870961 | 0.86535452 | 0.26095 | 0.087096111 | 0.348216313 | 0.787345533 | 0.01761941 | 0.894397368 | 0.0123 | 0.017619414 | 0.017619414 | 0.349184472 | 4.943189921 | 0.694007297 | FALSE | TRUE | FALSE |
| SNX29 | 0.087062 | 0.86536533 | 1 | 0.087061993 | 0.087061993 | 1.706542008 | 0.11552297 | 0.872270355 | 1 | 0.115522968 | 0.115522968 | 2.269852926 | 0.75363363 | -0.12283973 | FALSE | TRUE | FALSE |
| DCBLD2 | 0.087027 | 0.86537615 | 0.752 | 0.786276072 | 7.291186636 | 7.291186636 | 0.13205941 | 0.868441289 | 0.9403 | 2.794047228 | 106.757193 | 106.757193 | 0.658998857 | -0.181115339 | FALSE | TRUE | FALSE |
| ALG6 | 0.0870135 | 0.86538696 | 0.50985 | 0.087013539 | 0.087013539 | 0.431612928 | 0.53243234 | 0.741653002 | 0.4986 | 0.133149293 | 2.129602358 | 2.129602358 | 0.163426472 | -0.786677594 | FALSE | TRUE | FALSE |
| MICALL2 | 0.0870114 | 0.86539777 | 0.6323 | 0.087011392 | 3.015241506 | 3.015241506 | 0.04258459 | 0.888431987 | 0.8785 | 0.373518182 | 13.43668797 | 13.43668797 | 2.043260046 | 0.310323643 | FALSE | TRUE | FALSE |
| EPM2A | 0.0869248 | 0.86540857 | 0.50405 | 0.347558307 | 0.347558307 | 0.347558307 | 0.46300963 | 0.769769832 | 0.00535 | 0.463009632 | 0.463009632 | 0.463009632 | 0.187738635 | -0.726446343 | FALSE | TRUE | FALSE |
| SERPINB8 | 0.0868848 | 0.86541938 | 0.81265 | 0.347233871 | 12.85502632 | 12.85502632 | 0.27832009 | 0.83146248 | 0.5015 | 0.278320092 | 1.112970189 | 1.112970189 | 0.312175636 | -0.505600994 | FALSE | TRUE | FALSE |
| HHLA2 | 0.0868532 | 0.86543018 | 1 | 0.775838578 | 7.113170656 | 7.113170656 | 0.11843935 | 0.87161046 | 1 | 1.066291874 | 1.066291874 | 1.066291874 | 0.73331392 | -0.134710071 | FALSE | TRUE | FALSE |
| KLK11 | 0.0867791 | 0.86544099 | 0.0082 | 0.086779108 | 0.086779108 | 0.086779108 | 0.11564021 | 0.872260092 | 0.00855 | 0.115640205 | 0.115640205 | 0.115640205 | 0.75042333 | -0.124693673 | FALSE | TRUE | FALSE |
| ZNF502 | 0.0867109 | 0.86545179 | 1 | 0.086710948 | 0.086710948 | 1.718482712 | 0.11499328 | 0.872362654 | 1 | 0.11499328 | 0.11499328 | 2.281483954 | 0.754052301 | -0.122598531 | FALSE | TRUE | FALSE |
| JUP | 0.0867103 | 0.86546259 | 0.7534 | 0.346692797 | 1.38652385 | 6.794268125 | 0.46806522 | 0.767866559 | 0.5144 | 0.468065218 | 0.468065218 | 3.963015424 | 0.185252673 | -0.732235516 | FALSE | TRUE | FALSE |

| | | | | | | | | | | | | | | | | | |
|---|---|---|---|---|---|---|---|---|---|---|---|---|---|---|---|---|---|
| BACH1 | 0.0865959 | 0.86547339 | 1 | 0.796743204 | 0.796743204 | 15.71877253 | 0.11540282 | 0.872290876 | 1 | 1.065670096 | 1.065670096 | 3.903956017 | 0.750379248 | -0.124719185 | FALSE | TRUE | FALSE |
| WDR43 | 0.086574 | 0.86548418 | 1 | 0.086573988 | 0.086573988 | 1.711215943 | 0.11512103 | 0.872342154 | 1 | 0.11512103 | 0.11512103 | 2.278678942 | 0.752025831 | -0.123767242 | FALSE | TRUE | FALSE |
| C1QTNF8 | 0.0865572 | 0.86549498 | 0.878 | 3.162356671 | 29.65394643 | 29.65394643 | 0.120194 | 0.871184336 | 0.50615 | 1.104019021 | 1.104019021 | 1.104019021 | 0.720146167 | -0.142579346 | FALSE | TRUE | FALSE |
| FLVCR1 | 0.0865453 | 0.86550577 | 1 | 0.787010382 | 0.787010382 | 1.713925977 | 0.11863741 | 0.871569003 | 1 | 0.118637413 | 0.118637413 | 2.352244713 | 0.729493978 | -0.136978289 | FALSE | TRUE | FALSE |
| TCF7 | 0.0865259 | 0.86551656 | 0.5001 | 0.10776552 | 1.382974564 | 5.531092403 | 0.16517859 | 0.860876503 | 0.0066 | 0.041325593 | 0.16517859 | 0.16517859 | 0.523832531 | -0.280807534 | FALSE | TRUE | FALSE |
| AQP10 | 0.0863472 | 0.86552735 | 0.00735 | 0.086347155 | 0.086347155 | 0.086347155 | 0.11491597 | 0.872403636 | 0.00825 | 0.114915971 | 0.114915971 | 0.114915971 | 0.751393866 | -0.124132355 | FALSE | TRUE | FALSE |
| SLC25A12 | 0.0862511 | 0.86553814 | 0.0197 | 0.086251053 | 0.086251053 | 1.701964554 | 0.11523838 | 0.872311392 | 0.01685 | 0.115238378 | 0.115238378 | 2.277619694 | 0.748457718 | -0.125832729 | FALSE | TRUE | FALSE |
| PUS3 | 0.0862264 | 0.86554893 | 1 | 0.086226367 | 0.086226367 | 1.708462878 | 0.11496381 | 0.872372902 | 1 | 0.114963814 | 0.114963814 | 2.280410063 | 0.750030499 | -0.124921076 | FALSE | TRUE | FALSE |
| COL18A1 | 0.0862099 | 0.86555971 | 0.8128 | 0.775690179 | 647.8443064 | 6374.97552 | 3241.2465 | 0.000659545 | 0.13785 | 34.45385592 | 25923.09316 | 332472.7386 | 2.65978E-05 | -4.575154819 | FALSE | FALSE | FALSE |
| PLSCR2 | 0.086125 | 0.8655705 | 0.49775 | 0.086124987 | 0.787383195 | 0.787383195 | 8.01660742 | 0.227565739 | 0.3163 | 8.016607419 | 763.6999261 | 763.6999261 | 0.010743321 | -1.968861449 | FALSE | FALSE | FALSE |
| ARHGEF7 | 0.0860719 | 0.86558128 | 1 | 0.086071924 | 0.086071924 | 1.68967017 | 0.11494487 | 0.872383149 | 0.02085 | 0.11494487 | 0.11494487 | 2.261486544 | 0.74881049 | -0.12562808 | FALSE | TRUE | FALSE |
| MFSD6 | 0.0860674 | 0.86559206 | 1 | 0.086067367 | 0.086067367 | 1.693955806 | 0.45346919 | 0.773320201 | 0.5089 | 0.453469186 | 0.453469186 | 3.837179475 | 0.189797609 | -0.721709262 | FALSE | TRUE | FALSE |
| GJB5 | 0.0860606 | 0.86560284 | 0.9645 | 1.37562193 | 22.01040171 | 88.07548866 | 0.64962235 | 0.689900634 | 0.65555 | 0.649622352 | 10.3918628 | 41.57883535 | 0.132477821 | -0.877856824 | FALSE | TRUE | FALSE |
| SLC37A4 | 0.085992 | 0.86561362 | 0.01235 | 0.085992047 | 0.085992047 | 1.705577014 | 0.11052928 | 0.873198061 | 1 | 0.991570675 | 0.991570675 | 0.991570675 | 0.778002439 | -0.109019041 | FALSE | TRUE | FALSE |
| BORA | 0.0859376 | 0.86562439 | 1 | 0.76830794 | 0.76830794 | 1.702308758 | 1.02537302 | 0.586105808 | 0.51115 | 1.025373022 | 1.025373022 | 2.258967615 | 0.083811056 | -1.076698686 | FALSE | FALSE | FALSE |
| C2orf78 | 0.0858813 | 0.86563517 | 1 | 0.085881255 | 0.085881255 | 1.690350803 | 0.11071916 | 0.873177802 | 0.51145 | 1.003638887 | 1.003638887 | 19.79179901 | 0.775667517 | -0.110324395 | FALSE | TRUE | FALSE |
| ITGAM | 0.0858524 | 0.86564594 | 0.88265 | 0.779820762 | 7.265429104 | 15.18906152 | 1.23439652 | 0.563272108 | 0.89685 | 11.56220811 | 1091.936346 | 10996.71568 | 0.069550119 | -1.15770212 | FALSE | FALSE | FALSE |
| FGL2 | 0.0858201 | 0.86565671 | 1 | 0.785356327 | 7.376345596 | 7.376345596 | 7.1935518 | 0.240792461 | 0.7751 | 28.78745134 | 461.3688934 | 1847.708716 | 0.011930148 | -1.923354164 | FALSE | FALSE | FALSE |
| ZNF652 | 0.0857576 | 0.86566748 | 1 | 0.085757627 | 0.085757627 | 1.693887641 | 0.4533909 | 0.773384286 | 0.50875 | 0.453390895 | 0.453390895 | 2.243247746 | 0.189147219 | -0.723200039 | FALSE | TRUE | FALSE |
| ZBTB49 | 0.085745 | 0.86567825 | 1 | 0.085745024 | 0.085745024 | 1.687803892 | 0.11404717 | 0.872577537 | 0.0206 | 0.114047172 | 0.114047172 | 2.249172737 | 0.751838232 | -0.123875594 | FALSE | TRUE | FALSE |
| POLR2L | 0.0857373 | 0.86568902 | 1 | 0.085737319 | 0.085737319 | 0.085737319 | 0.08823049 | 0.878538519 | 1 | 0.088230487 | 0.088230487 | 0.088230487 | 0.971742551 | -0.01244878 | FALSE | TRUE | FALSE |
| TAS2R60 | 0.0856831 | 0.86569978 | 0.8815 | 6.955830256 | 65.12567273 | 65.12567273 | 0.1204049 | 0.871142622 | 0.5044 | 1.08068505 | 1.08068505 | 1.08068505 | 0.711625102 | -0.147748741 | FALSE | TRUE | FALSE |
| SLC1A5 | 0.085659 | 0.86571054 | 1 | 0.342504768 | 0.342504768 | 1.69120182 | 0.11436961 | 0.872526435 | 0.5066 | 0.457323891 | 0.457323891 | 2.26153334 | 0.748966419 | -0.125537654 | FALSE | TRUE | FALSE |
| KIAA0513 | 0.0856305 | 0.86572131 | 1 | 0.085630492 | 0.085630492 | 1.695499059 | 0.09157921 | 0.877814731 | 0.50355 | 0.857326299 | 0.857326299 | 1.81546394 | 0.935042935 | -0.029168447 | FALSE | TRUE | FALSE |
| HMGB4 | 0.0856018 | 0.86573207 | 1 | 0.784993485 | 0.784993485 | 0.784993485 | 833.695846 | 0.00278 | 0.18865 | 84.21530341 | 833.6958458 | 8454.791227 | 0.000102678 | -3.98852455 | FALSE | FALSE | FALSE |
| ANKRD1 | 0.0856017 | 0.86574282 | 1 | 0.085601729 | 0.085601729 | 0.085601729 | 0.96305369 | 0.591650495 | 0.7511 | 0.963053693 | 8.89925148 | 8.89925148 | 0.088885729 | -1.051167963 | FALSE | TRUE | FALSE |
| CD1E | 0.0855935 | 0.86575358 | 1 | 0.76457257 | 0.76457257 | 1.698669774 | 0.11012903 | 0.873299265 | 1 | 0.989727475 | 9.129162124 | 9.129162124 | 0.777210837 | -0.109461153 | FALSE | TRUE | FALSE |
| TCAIM | 0.0855919 | 0.86576433 | 1 | 0.085591879 | 0.085591879 | 1.697547207 | 0.11369933 | 0.872638812 | 1 | 0.113699335 | 0.113699335 | 2.257267474 | 0.752791379 | -0.123325363 | FALSE | TRUE | FALSE |
| DLX3 | 0.0855138 | 0.86577509 | 0.01095 | 0.085513836 | 0.085513836 | 1.696416265 | 0.45233984 | 0.773672211 | 0.50425 | 0.452339841 | 0.452339841 | 0.452339841 | 0.189047765 | -0.723428453 | FALSE | TRUE | FALSE |
| TMCO3 | 0.0855059 | 0.86578584 | 0.507 | 0.085505933 | 0.085505933 | 1.681685459 | 0.12494145 | 0.870133958 | 0.75855 | 0.12494145 | 1.125535802 | 2.462200768 | 0.684368027 | -0.164710289 | FALSE | TRUE | FALSE |
| DCLRE1C | 0.0854617 | 0.86579659 | 0.93905 | 0.765927277 | 28.17477984 | 28.17477984 | 1.2297814 | 0.563642664 | 0.00365 | 0.036312065 | 0.036312065 | 0.7200244 | 0.069493434 | -1.158056228 | FALSE | FALSE | FALSE |
| RHOBTB1 | 0.085406 | 0.86580734 | 1 | 0.085405999 | 0.085405999 | 1.680987739 | 1.02584504 | 0.585993624 | 0.5066 | 1.025845041 | 1.025845041 | 3.741613651 | 0.083254289 | -1.079593385 | FALSE | FALSE | FALSE |
| CRYGB | 0.0853458 | 0.86581808 | 1 | 1.364711084 | 5.459246844 | 5.459246844 | 0.1178347 | 0.871776032 | 1 | 0.117834704 | 0.117834704 | 0.117834704 | 0.724284361 | -0.140090892 | FALSE | TRUE | FALSE |
| ANXA13 | 0.0853328 | 0.86582883 | 1 | 0.341329583 | 0.341329583 | 1.692530166 | 0.07486646 | 0.881419608 | 1 | 0.074866462 | 0.074866462 | 0.074866462 | 1.139800436 | 0.056828819 | FALSE | TRUE | FALSE |
| RAD51AP1 | 0.085321 | 0.86583957 | 1 | 0.793365531 | 0.793365531 | 0.793365531 | 0.09516596 | 0.877111745 | 1 | 0.887082836 | 0.887082836 | 0.887082836 | 0.896549979 | -0.047425495 | FALSE | TRUE | FALSE |
| ATP1A4 | 0.0853196 | 0.86585031 | 0.96015 | 627.2635722 | 1758903.963 | 53064757.26 | 8.4152E-05 | 0.900238712 | 0.9909 | 8.445847239 | 20531.23803 | 797256.6764 | 1013.876269 | 3.005984958 | FALSE | TRUE | FALSE |
| GNG5 | 0.0853122 | 0.86586105 | 1 | 0.085312229 | 0.085312229 | 0.085312229 | 0.08623181 | 0.8789486 | 1 | 0.086231815 | 0.086231815 | 0.086231815 | 0.989335887 | -0.004656237 | FALSE | TRUE | FALSE |
| GPR83 | 0.085211 | 0.86587179 | 1 | 0.085211042 | 0.085211042 | 1.687921218 | 1.00888828 | 0.587448629 | 0.5045 | 1.008888279 | 1.008888279 | 2.184986107 | 0.084460335 | -1.073347199 | FALSE | FALSE | FALSE |
| MGAT1 | 0.0851932 | 0.86588252 | 1 | 0.085193186 | 0.085193186 | 1.681645825 | 0.11408133 | 0.872567319 | 1 | 0.114081327 | 0.114081327 | 2.25540822 | 0.746775903 | -0.126809704 | FALSE | TRUE | FALSE |
| ITGA9 | 0.0851679 | 0.86589326 | 1 | 0.340596871 | 0.340596871 | 6.66514173 | 0.45471739 | 0.772870542 | 0.5135 | 0.454717385 | 0.454717385 | 8.921465043 | 0.187298607 | -0.727465452 | FALSE | TRUE | FALSE |
| PLA1A | 0.0851585 | 0.86590399 | 0.51225 | 0.085158464 | 0.340296344 | 0.340296344 | 0.26341042 | 0.835678068 | 0.25755 | 0.06589399 | 0.263410425 | 0.263410425 | 0.323291928 | -0.490405139 | FALSE | TRUE | FALSE |
| PDHX | 0.0851559 | 0.86591472 | 0.0142 | 0.085155877 | 0.085155877 | 1.685437257 | 0.97450297 | 0.590991679 | 0.7542 | 0.974502965 | 9.128543459 | 9.128543459 | 0.087383907 | -1.058568539 | FALSE | TRUE | FALSE |
| MRPL15 | 0.085155 | 0.86592545 | 0.74845 | 0.777690059 | 7.289554413 | 7.289554413 | 1.02993782 | 0.585319208 | 0.7557 | 4.118618935 | 38.77839697 | 38.77839697 | 0.082679706 | -1.082601079 | FALSE | FALSE | FALSE |
| NECAP2 | 0.0850909 | 0.86593618 | 0.00975 | 0.085090946 | 0.085090946 | 0.085090946 | 0.1131436 | 0.872740822 | 0.00685 | 0.113143598 | 0.113143598 | 0.113143598 | 0.752061522 | -0.123746631 | FALSE | TRUE | FALSE |
| FOXF1 | 0.0850809 | 0.86594691 | 1 | 77.2222304 | 8199.590849 | 87553.46868 | 0.13115957 | 0.868603769 | 1 | 0.131159567 | 0.131159567 | 2.599220487 | 0.648682354 | -0.187967916 | FALSE | TRUE | FALSE |
| CCDC80 | 0.0850547 | 0.86595763 | 0.0296 | 0.085054697 | 0.085054697 | 1.665883572 | 0.11323116 | 0.872710235 | 1 | 0.113231165 | 0.113231165 | 2.223277541 | 0.751159781 | -0.124267673 | FALSE | TRUE | FALSE |
| LAMB3 | 0.085045 | 0.86596836 | 0.4277 | 0.085044992 | 1.629815861 | 6.515782741 | 0.02452004 | 0.892599192 | 0.70055 | 0.052416403 | 0.838125205 | 3.306460489 | 3.468387545 | 0.540127618 | FALSE | TRUE | FALSE |
| RASAL2 | 0.085034 | 0.86597908 | 0.52445 | 0.085034023 | 0.339799639 | 2.864195108 | 0.06591324 | 0.883376453 | 0.5067 | 0.065913244 | 0.065913244 | 1.289277231 | 1.290090085 | 0.110620038 | FALSE | TRUE | FALSE |
| RUNX2 | 0.0849759 | 0.8659898 | 1 | 0.084975921 | 0.084975921 | 1.678701963 | 0.11379116 | 0.872618393 | 1 | 0.113791161 | 0.113791161 | 2.251260831 | 0.746770846 | -0.126812645 | FALSE | TRUE | FALSE |
| PDLIM7 | 0.0849644 | 0.86600051 | 1 | 0.084964391 | 0.084964391 | 1.679634457 | 0.11013634 | 0.873289152 | 1 | 1.023376545 | 1.023376545 | 2.180280383 | 0.771447403 | -0.112693678 | FALSE | TRUE | FALSE |
| NPFFR1 | 0.0849624 | 0.86601123 | 0.0084 | 0.084962428 | 0.084962428 | 0.084962428 | 0.11307594 | 0.872761205 | 1 | 0.113075943 | 0.113075943 | 0.113075943 | 0.751374924 | -0.124143303 | FALSE | TRUE | FALSE |
| GHR | 0.0849617 | 0.86602194 | 0.5027 | 0.084961679 | 0.084961679 | 0.719227803 | 0.03134051 | 0.890906426 | 1 | 2.003406861 | 18.38490634 | 73.54720536 | 2.710922128 | 0.433117042 | FALSE | TRUE | FALSE |
| RABGGTA | 0.0849479 | 0.86603266 | 0.0193 | 0.084947933 | 0.084947933 | 1.676654387 | 0.11312077 | 0.872751014 | 1 | 0.113120769 | 0.113120769 | 2.236241125 | 0.750949038 | -0.124389535 | FALSE | TRUE | FALSE |
| WIZ | 0.0849207 | 0.86604337 | 1 | 0.790731868 | 7.556745794 | 26.71732355 | 0.10575814 | 0.874452269 | 0.0263 | 0.105758142 | 0.105758142 | 2.073200463 | 0.802970602 | -0.095300355 | FALSE | TRUE | FALSE |
| CSRP2 | 0.0849197 | 0.86605408 | 1 | 0.0849197 | 0.0849197 | 0.0849197 | 0.11323077 | 0.872720432 | 1 | 0.113230771 | 0.113230771 | 0.113230771 | 0.749970165 | -0.124956013 | FALSE | TRUE | FALSE |
| CLSTN2 | 0.0848837 | 0.86606479 | 1 | 0.084883698 | 0.084883698 | 1.663517384 | 0.11101192 | 0.873147404 | 1 | 0.443907198 | 1.775510794 | 8.720598949 | 0.764635882 | -0.116545326 | FALSE | TRUE | FALSE |
| KNG1 | 0.0848377 | 0.86607549 | 0.0171 | 0.084837663 | 0.084837663 | 1.675793078 | 0.98584685 | 0.590000411 | 0.5103 | 0.985846848 | 0.985846848 | 3.701563853 | 0.086055621 | -1.065220755 | FALSE | TRUE | FALSE |
| TLE6 | 0.0847979 | 0.8660862 | 1 | 0.757113723 | 0.757113723 | 1.677915134 | 0.98113718 | 0.590441445 | 0.26145 | 0.981137181 | 9.046188615 | 9.046188615 | 0.086428134 | -1.063344865 | FALSE | TRUE | FALSE |
| GRSF1 | 0.0847674 | 0.8660969 | 1 | 0.759993038 | 6.993512267 | 6.993512267 | 4.05283869 | 0.353817838 | 0.0035 | 0.119585838 | 0.119585838 | 0.119585838 | 0.020915563 | -1.679530433 | FALSE | FALSE | FALSE |
| ENDOD1 | 0.0847433 | 0.8661076 | 1 | 0.760855405 | 0.760855405 | 1.680319641 | 0.11607541 | 0.872188213 | 0.0106 | 0.116075406 | 0.116075406 | 2.303958865 | 0.730071253 | -0.136634752 | FALSE | TRUE | FALSE |
| PRDM10 | 0.0846102 | 0.8661183 | 0.0373 | 0.084610189 | 0.084610189 | 2.848256225 | 0.11283504 | 0.87279177 | 1 | 0.112835043 | 0.112835043 | 3.80292229 | 0.749857369 | -0.125021336 | FALSE | TRUE | FALSE |
| TFR2 | 0.0845132 | 0.866129 | 0.02325 | 0.084513201 | 0.084513201 | 1.659242651 | 8.67486054 | 0.215512055 | 0.5039 | 8.674860536 | 82.80226882 | 170.6893924 | 0.009742312 | -2.011337949 | FALSE | FALSE | FALSE |
| EXOSC1 | 0.0844739 | 0.8661397 | 1 | 0.084473931 | 0.084473931 | 0.084473931 | 0.00010593 | 0.900186425 | 0.0063 | 0.000105935 | 0.000105935 | 0.000105935 | 797.4134481 | 2.901683556 | FALSE | TRUE | FALSE |
| SIRPB1 | 0.0844352 | 0.86615039 | 1 | 6.911356416 | 626.1818197 | 626.1818197 | 1.00819966 | 0.587560166 | 0.936 | 9.305553004 | 854.8868853 | 854.8868853 | 0.08374852 | -1.077022857 | FALSE | FALSE | FALSE |
| NOL10 | 0.084402 | 0.86616108 | 0.50465 | 0.33746779 | 0.33746779 | 2.853647587 | 0.11422281 | 0.8725571 | 1 | 0.114222805 | 0.114222805 | 2.262933837 | 0.738924452 | -0.131399962 | FALSE | TRUE | FALSE |
| ZSCAN9 | 0.0843735 | 0.86617178 | 1 | 0.084373497 | 0.084373497 | 1.67559058 | 0.11221641 | 0.872863043 | 1 | 0.112216408 | 0.112216408 | 2.230415122 | 0.751881995 | -0.123850315 | FALSE | TRUE | FALSE |
| NARS2 | 0.0842706 | 0.86618247 | 0.8732 | 0.336910094 | 3.08696195 | 3.08696195 | 0.12932248 | 0.869121159 | 0.99295 | 8.271903834 | 132.4653292 | 530.289769 | 0.65163179 | -0.185997737 | FALSE | TRUE | FALSE |
| ADAM30 | 0.0842697 | 0.86619316 | 0.9383 | 0.738364333 | 61.13742878 | 61.13742878 | 0.15245955 | 0.863954918 | 0.75065 | 0.15245955 | 1.354012819 | 3.008709416 | 0.55273512 | -0.25748294 | FALSE | TRUE | FALSE |
| THAP3 | 0.0841807 | 0.86620384 | 0.75775 | 0.190849038 | 0.762859545 | 0.762859545 | 0.0344821 | 0.890256308 | 1 | 0.034482102 | 0.034482102 | 0.034482102 | 2.441285945 | 0.387618651 | FALSE | TRUE | FALSE |
| RET | 0.0841712 | 0.86621453 | 0.9228 | 7.586233975 | 756.6735017 | 7837.659617 | 0.05467744 | 0.885753928 | 0.1547 | 0.054677441 | 0.218479692 | 1.065190768 | 1.539414346 | 0.187355529 | FALSE | TRUE | FALSE |
| SGCD | 0.0840884 | 0.86622521 | 1 | 0.336157609 | 1.344183623 | 1.344183623 | 1.80136621 | 0.507059474 | 0.25125 | 0.450460327 | 1.801366211 | 1.801366211 | 0.046680322 | -1.330866159 | FALSE | FALSE | FALSE |
| CYB5R1 | 0.0840564 | 0.8662359 | 0.8117 | 1.344336279 | 5.378229264 | 21.52182938 | 0.30603138 | 0.823509749 | 0.00685 | 0.306031381 | 0.306031381 | 0.306031381 | 0.274665884 | -0.561195281 | FALSE | TRUE | FALSE |
| TPM2 | 0.0840547 | 0.86624658 | 1 | 0.084054692 | 0.084054692 | 0.084054692 | 0.11249572 | 0.872822324 | 0.0078 | 0.112495718 | 0.112495718 | 0.112495718 | 0.747181258 | -0.12657403 | FALSE | TRUE | FALSE |

| | | | | | | | | | | | | | | | | | |
|---|---|---|---|---|---|---|---|---|---|---|---|---|---|---|---|---|---|
| TAS2R20 | 0.0840526 | 0.86625726 | 0.50265 | 0.79733179 | 0.79733179 | 0.79733179 | 0.08014455 | 0.88029157 | 1 | 0.080144549 | 0.080144549 | 0.080144549 | 1.048761937 | 0.020676917 | FALSE | TRUE | FALSE |
| RILP | 0.0840429 | 0.86626794 | 0.008 | 0.084042946 | 0.084042946 | 0.084042946 | 0.11152704 | 0.873005374 | 1 | 0.111527039 | 0.111527039 | 0.111527039 | 0.753565649 | -0.122878907 | FALSE | TRUE | FALSE |
| MSI2 | 0.0840177 | 0.86627861 | 1 | 0.084017707 | 0.084017707 | 0.084017707 | 0.11254813 | 0.872812141 | 0.0069 | 0.112548131 | 0.112548131 | 0.112548131 | 0.74650469 | -0.12696746 | FALSE | TRUE | FALSE |
| CUX1 | 0.0839914 | 0.86628929 | 0.76295 | 0.192729167 | 0.770508981 | 14.87976173 | 0.30819529 | 0.822641845 | 0.5164 | 0.30819529 | 0.30819529 | 5.976459241 | 0.272526606 | -0.564591092 | FALSE | TRUE | FALSE |
| DPH6 | 0.0839535 | 0.86629996 | 0.50705 | 0.754399084 | 0.754399084 | 0.754399084 | 0.10839523 | 0.873752778 | 1 | 0.979311657 | 9.080951577 | 9.080951577 | 0.774513214 | -0.110971169 | FALSE | TRUE | FALSE |
| IL5RA | 0.0839148 | 0.86631063 | 0.7556 | 0.335417745 | 3.063216136 | 3.063216136 | 0.01689917 | 0.894629141 | 1 | 0.151610394 | 0.151610394 | 0.151610394 | 4.965616074 | 0.695973138 | FALSE | TRUE | FALSE |
| MIF4GD | 0.0838064 | 0.8663213 | 0.87325 | 6.800635892 | 63.65935431 | 63.65935431 | 0.99616977 | 0.588783394 | 0.8757 | 9.159106352 | 86.37532764 | 86.37532764 | 0.084128616 | -1.075056255 | FALSE | TRUE | FALSE |
| CCNG1 | 0.0837232 | 0.86633197 | 0.00635 | 0.083723223 | 0.083723223 | 0.083723223 | 0.09636112 | 0.876843818 | 1 | 0.096361124 | 0.096361124 | 0.096361124 | 0.868848559 | -0.061055915 | FALSE | TRUE | FALSE |
| SLC43A1 | 0.0837061 | 0.86634264 | 1 | 0.083706139 | 0.083706139 | 1.652802944 | 0.1114092 | 0.87306628 | 0.0158 | 0.111409202 | 0.111409202 | 2.203179204 | 0.751339546 | -0.124163752 | FALSE | TRUE | FALSE |
| TAF6 | 0.0836617 | 0.8663533 | 0.02305 | 0.083661695 | 0.083661695 | 1.644974859 | 0.11151633 | 0.873015529 | 1 | 0.111516333 | 0.111516333 | 2.197117115 | 0.750219214 | -0.124811817 | FALSE | TRUE | FALSE |
| IMP4 | 0.0836401 | 0.86636396 | 1 | 0.083640133 | 0.083640133 | 1.659124694 | 0.11148504 | 0.873035835 | 0.0094 | 0.11148504 | 0.11148504 | 0.11148504 | 0.750236384 | -0.124801878 | FALSE | TRUE | FALSE |
| DOCK3 | 0.0835766 | 0.86637463 | 1 | 0.083576571 | 1.600811729 | 2.796180234 | 0.4160556 | 0.786121796 | 0.87805 | 3.872639897 | 36.99574471 | 178.079317 | 0.200878371 | -0.697066823 | FALSE | TRUE | FALSE |
| KCNAB3 | 0.0835292 | 0.86638529 | 0.25455 | 0.083529212 | 0.737721688 | 0.737721688 | 10.6553947 | 0.191146841 | 0.19615 | 10.65539468 | 99.78770896 | 957.3737781 | 0.007839148 | -2.105731156 | FALSE | FALSE | FALSE |
| RSPO4 | 0.0835137 | 0.86639595 | 1 | 0.083513711 | 0.083513711 | 0.083513711 | 0.11180855 | 0.872934244 | 1 | 0.111808551 | 0.111808551 | 0.111808551 | 0.746934925 | -0.126717234 | FALSE | TRUE | FALSE |
| CRYBA1 | 0.0834987 | 0.8664066 | 1 | 0.083498652 | 0.083498652 | 0.083498652 | 0.11141909 | 0.873056133 | 1 | 0.111419094 | 0.111419094 | 0.111419094 | 0.749410618 | -0.125280158 | FALSE | TRUE | FALSE |
| MAX | 0.0834253 | 0.86641726 | 0.0034 | 0.083425349 | 0.083425349 | 0.083425349 | 0.11182829 | 0.872924077 | 1 | 0.111828288 | 0.111828288 | 0.111828288 | 0.746012933 | -0.127253643 | FALSE | TRUE | FALSE |
| DEXI | 0.0834242 | 0.86642791 | 1 | 0.083424249 | 0.083424249 | 0.083424249 | 0.08500045 | 0.879134187 | 1 | 0.085000452 | 0.085000452 | 0.085000452 | 0.98145653 | -0.008128931 | FALSE | TRUE | FALSE |
| WIPF3 | 0.0833564 | 0.86643857 | 0.5078 | 0.751248794 | 0.751248794 | 1.655441397 | 0.11110987 | 0.873106854 | 1 | 1.007362183 | 1.007362183 | 1.007362183 | 0.750216087 | -0.124813627 | FALSE | TRUE | FALSE |
| TIMELESS | 0.0833013 | 0.86644922 | 0.78455 | 0.333255208 | 6.137904622 | 24.55529247 | 0.00531473 | 0.898037624 | 0.5188 | 0.044635752 | 0.044635752 | 0.872604808 | 15.67367953 | 1.195170963 | FALSE | TRUE | FALSE |
| ZDHHC5 | 0.0832392 | 0.86645987 | 1 | 0.332776858 | 0.332776858 | 6.545134657 | 0.11301238 | 0.872771395 | 1 | 0.113012379 | 0.113012379 | 2.227193161 | 0.736549191 | -0.132798243 | FALSE | TRUE | FALSE |
| ATP8A1 | 0.0832013 | 0.86647051 | 0.27115 | 0.020815978 | 0.083201324 | 0.700781237 | 0.15270531 | 0.863908722 | 0.52025 | 0.152705314 | 0.610584509 | 2.997539315 | 0.544848908 | -0.263723915 | FALSE | TRUE | FALSE |
| C17orf58 | 0.0831166 | 0.86648116 | 1 | 0.770027003 | 0.770027003 | 0.770027003 | 0.09269739 | 0.877606502 | 1 | 0.861256275 | 0.861256275 | 0.861256275 | 0.896644572 | -0.047379676 | FALSE | TRUE | FALSE |
| SERPINE2 | 0.0831153 | 0.8664918 | 1 | 0.083115271 | 0.083115271 | 1.648300369 | 0.11153322 | 0.872995218 | 1 | 0.111533217 | 0.111533217 | 2.214111997 | 0.745206438 | -0.127723402 | FALSE | TRUE | FALSE |
| SLC15A1 | 0.0831034 | 0.86650245 | 0.937 | 0.733709249 | 26.59348364 | 26.59348364 | 0.13934608 | 0.866895492 | 0.5066 | 0.139346077 | 0.139346077 | 0.688287798 | 0.596381192 | -0.224476061 | FALSE | TRUE | FALSE |
| RAF1 | 0.0830389 | 0.86651309 | 1 | 0.083038886 | 0.083038886 | 1.638096922 | 0.11148888 | 0.873025683 | 1 | 0.111488879 | 0.111488879 | 2.202940638 | 0.744817664 | -0.127950032 | FALSE | TRUE | FALSE |
| LYSMD4 | 0.0830045 | 0.86652373 | 0.99975 | 59576.04637 | 6453694.656 | 69445283.99 | 9.97023254 | 0.198278805 | 0.96595 | 912.4011651 | 91965.19081 | 956020.5347 | 0.008325235 | -2.079603476 | FALSE | FALSE | FALSE |
| CRYZ | 0.0829858 | 0.86653437 | 1 | 0.082985802 | 0.082985802 | 0.082985802 | 0.11044494 | 0.873208188 | 0.00685 | 0.110444941 | 0.110444941 | 0.110444941 | 0.751377119 | -0.124142034 | FALSE | TRUE | FALSE |
| FMNL3 | 0.0829856 | 0.866545 | 0.27035 | 0.020764732 | 0.082985592 | 0.700833914 | 0.00977233 | 0.896792153 | 1 | 0.039050406 | 0.039050406 | 0.764815567 | 8.491894042 | 0.929004567 | FALSE | TRUE | FALSE |
| PROCA1 | 0.0828981 | 0.86655564 | 1 | 0.082898092 | 0.082898092 | 1.645707321 | 0.90233737 | 0.598565464 | 0.8761 | 8.289327251 | 78.10642898 | 78.10642898 | 0.091870397 | -1.036824408 | FALSE | TRUE | FALSE |
| TNFRSF10D | 0.082846 | 0.86656627 | 1 | 0.082846047 | 0.082846047 | 1.641459287 | 0.11034786 | 0.873248682 | 1 | 0.110347855 | 0.110347855 | 2.188816718 | 0.750771701 | -0.124492106 | FALSE | TRUE | FALSE |
| CCDC174 | 0.0827575 | 0.86657691 | 1 | 0.082757499 | 0.082757499 | 1.638937353 | 0.10716881 | 0.874033463 | 1 | 0.972022099 | 0.972022099 | 2.124881874 | 0.772216293 | -0.112261039 | FALSE | TRUE | FALSE |
| EPB42 | 0.0826499 | 0.86658754 | 0.51605 | 0.082649894 | 0.082649894 | 1.627659508 | 0.12949908 | 0.86907819 | 0.5058 | 0.129499077 | 0.129499077 | 1.095542813 | 0.638227669 | -0.195024372 | FALSE | TRUE | FALSE |
| WDR88 | 0.0826269 | 0.86659817 | 1 | 0.082626867 | 0.082626867 | 1.635104653 | 0.42277072 | 0.783530706 | 0.7514 | 0.954108554 | 3.816158426 | 3.816158426 | 0.195441322 | -0.708983609 | FALSE | TRUE | FALSE |
| BCR | 0.0826218 | 0.86660879 | 1 | 0.082621802 | 0.082621802 | 1.609547231 | 0.10741887 | 0.873973416 | 1 | 0.997130579 | 0.997130579 | 19.35883966 | 0.769155351 | -0.113985934 | FALSE | TRUE | FALSE |
| WDR6 | 0.0825951 | 0.86661942 | 0.27255 | 0.009214792 | 0.082595123 | 0.310430774 | 0.02124908 | 0.89347575 | 0.5152 | 0.021249082 | 0.021249082 | 0.415163708 | 3.886997258 | 0.589614234 | FALSE | TRUE | FALSE |
| NOX5 | 0.0825836 | 0.86663004 | 0.50875 | 0.082583597 | 1.321205891 | 5.28653983 | 0.20436962 | 0.850951383 | 0.50825 | 0.204369618 | 6.994629138 | 27.98459223 | 0.404089403 | -0.393522539 | FALSE | TRUE | FALSE |
| IL17RE | 0.0825768 | 0.86664067 | 1 | 0.082576763 | 0.082576763 | 1.634909182 | 0.10666903 | 0.874163378 | 1 | 0.957234466 | 0.957234466 | 2.114461125 | 0.774140031 | -0.111180475 | FALSE | TRUE | FALSE |
| CRYGA | 0.0825609 | 0.86665129 | 0.00735 | 0.082560855 | 0.082560855 | 0.082560855 | 0.11074894 | 0.873167671 | 1 | 0.11074894 | 0.11074894 | 0.11074894 | 0.745477607 | -0.127565397 | FALSE | TRUE | FALSE |
| OR4D1 | 0.0825575 | 0.86666191 | 1 | 2.49953E+19 | 1.41611E+25 | 3.18027E+27 | 15353.5459 | 0.000132542 | 0.99685 | 2726207197 | 5.37559E+13 | 8.49535E+15 | 5.3771E-06 | -5.269452039 | FALSE | FALSE | FALSE |
| DDX56 | 0.0824973 | 0.86667252 | 1 | 0.749241276 | 0.749241276 | 2.793518131 | 1.00686211 | 0.587894403 | 0.4975 | 0.110298771 | 1.006862106 | 2.182157728 | 0.081935011 | -1.086530484 | FALSE | FALSE | FALSE |
| GMIP | 0.0824507 | 0.86668314 | 1 | 0.082450666 | 0.082450666 | 1.608876401 | 0.97650574 | 0.590551609 | 0.51435 | 0.976505745 | 0.976505745 | 19.11063817 | 0.084434389 | -1.073480635 | FALSE | TRUE | FALSE |
| ZNF829 | 0.0823787 | 0.86669375 | 1 | 619.4472702 | 62246.54518 | 248966.3864 | 0.13705648 | 0.867370459 | 1 | 0.137056484 | 0.137056484 | 2.723719578 | 0.601056374 | -0.221084793 | FALSE | TRUE | FALSE |
| INA | 0.0823777 | 0.86670437 | 1 | 0.768428251 | 3.072156223 | 6.507258801 | 0.45634719 | 0.772289705 | 0.5081 | 0.456347186 | 0.456347186 | 2.258441724 | 0.180515499 | -0.743485504 | FALSE | TRUE | FALSE |
| HTATIP2 | 0.0823409 | 0.86671498 | 1 | 0.082340932 | 0.082340932 | 0.082340932 | 0.10966416 | 0.873470958 | 1 | 0.109664163 | 0.109664163 | 0.109664163 | 0.750846312 | -0.124448948 | FALSE | TRUE | FALSE |
| COQ5 | 0.0823323 | 0.86672559 | 1 | 1.31667229 | 12.0963196 | 12.0963196 | 0.11635381 | 0.872105975 | 1 | 0.116353812 | 0.116353812 | 0.116353812 | 0.707603256 | -0.150210177 | FALSE | TRUE | FALSE |
| KLK6 | 0.0822943 | 0.8667362 | 1 | 0.082294269 | 0.082294269 | 0.082294269 | 0.11018442 | 0.873279037 | 1 | 0.110184422 | 0.110184422 | 0.110184422 | 0.746877534 | -0.126750604 | FALSE | TRUE | FALSE |
| CEP78 | 0.0822367 | 0.8667468 | 0.0172 | 0.082236669 | 0.082236669 | 1.623660676 | 0.10968222 | 0.873440692 | 1 | 0.109682222 | 0.109682222 | 2.168877558 | 0.749772091 | -0.12507073 | FALSE | TRUE | FALSE |
| ESRRA | 0.0822213 | 0.86675741 | 1 | 0.774793555 | 3.0972005 | 3.0972005 | 0.11539681 | 0.872301135 | 1 | 0.115396807 | 0.115396807 | 2.287180077 | 0.712509436 | -0.14720938 | FALSE | TRUE | FALSE |
| ZNF672 | 0.0820803 | 0.86676801 | 1 | 0.082080266 | 0.082080266 | 1.618505828 | 0.10968416 | 0.8734306 | 1 | 0.109684159 | 0.109684159 | 2.166477871 | 0.748332909 | -0.125905155 | FALSE | TRUE | FALSE |
| KSR1 | 0.0820685 | 0.86677861 | 1 | 0.082068492 | 0.082068492 | 1.60874516 | 0.1103181 | 0.873258802 | 1 | 0.110318095 | 0.110318095 | 2.167683713 | 0.74392593 | -0.128470303 | FALSE | TRUE | FALSE |
| DLL4 | 0.08205 | 0.86678921 | 1 | 0.082049982 | 0.082049982 | 1.610284271 | 0.10362462 | 0.875006539 | 0.7564 | 0.978522042 | 9.483347794 | 19.24741519 | 0.791800103 | -0.101384446 | FALSE | TRUE | FALSE |
| MBD2 | 0.0820289 | 0.86679981 | 1 | 0.32796132 | 1.311556504 | 1.311556504 | 0.1126264 | 0.872801956 | 1 | 0.112626398 | 0.112626398 | 0.112626398 | 0.728327335 | -0.13767339 | FALSE | TRUE | FALSE |
| BEND3 | 0.0820039 | 0.86681041 | 1 | 0.767773952 | 0.767773952 | 14.99020648 | 0.85355416 | 0.606805659 | 0.75445 | 0.853554161 | 8.208204929 | 28.87296562 | 0.096073483 | -1.017396464 | FALSE | TRUE | FALSE |
| RBBP8NL | 0.0820014 | 0.866821 | 0.5063 | 0.082001351 | 0.082001351 | 0.178225505 | 0.01121783 | 0.896359152 | 0.4998 | 0.011217832 | 0.102492035 | 0.221972692 | 7.309910748 | 0.863912074 | FALSE | TRUE | FALSE |
| SUN2 | 0.0819801 | 0.8668316 | 1 | 0.081980141 | 0.081980141 | 1.608407221 | 0.10944726 | 0.87352137 | 0.02115 | 0.109447265 | 0.109447265 | 2.152217514 | 0.74903782 | -0.125496254 | FALSE | TRUE | FALSE |
| ANGPTL5 | 0.0819749 | 0.86684219 | 0.50545 | 0.081974914 | 0.081974914 | 0.081974914 | 1.93442799 | 0.495398908 | 0.50785 | 1.934427986 | 17.448025 | 69.80928555 | 0.042376824 | -1.372871594 | FALSE | FALSE | FALSE |
| CASP7 | 0.0819166 | 0.86685278 | 1 | 0.73454057 | 6.760277445 | 6.760277445 | 81.3261399 | 0.0361488 | 0.06995 | 8.574322989 | 790.6856635 | 790.6856635 | 0.001007261 | -2.99685812 | FALSE | FALSE | FALSE |
| TACC3 | 0.0818613 | 0.86686337 | 0.0224 | 0.081861274 | 0.081861274 | 1.608541333 | 0.10940262 | 0.873541524 | 1 | 0.109402616 | 0.109402616 | 2.154252259 | 0.748257004 | -0.12594921 | FALSE | TRUE | FALSE |
| AP4E1 | 0.0818536 | 0.86687396 | 1 | 0.327411437 | 0.327411437 | 6.442229014 | 0.10929685 | 0.873591882 | 1 | 0.437183978 | 0.437183978 | 8.618903309 | 0.748910744 | -0.125569939 | FALSE | TRUE | FALSE |
| TRAIP | 0.0818461 | 0.86688454 | 1 | 0.081846101 | 0.081846101 | 1.620461582 | 0.10628576 | 0.874293037 | 1 | 0.970949461 | 0.970949461 | 2.106884962 | 0.770057087 | -0.113477078 | FALSE | TRUE | FALSE |
| BZW2 | 0.0817481 | 0.86689513 | 1 | 0.081748147 | 0.081748147 | 1.622401582 | 0.96701892 | 0.591321296 | 0.50105 | 0.96701892 | 0.96701892 | 0.96701892 | 0.084536243 | -1.072957055 | FALSE | TRUE | FALSE |
| ATP6V1A | 0.0816341 | 0.86690571 | 1 | 0.081634139 | 0.081634139 | 1.612899561 | 0.10983211 | 0.873390218 | 0.01515 | 0.109832113 | 0.109832113 | 2.173189253 | 0.743262938 | -0.128857522 | FALSE | TRUE | FALSE |
| ABCG2 | 0.0816311 | 0.86691629 | 0.51135 | 0.175687929 | 6.197673162 | 24.78085189 | 0.19462239 | 0.853294784 | 0.4119 | 0.194622393 | 3.799110978 | 15.18827797 | 0.419433287 | -0.377337106 | FALSE | TRUE | FALSE |
| GAB4 | 0.0816194 | 0.86692687 | 0.87525 | 6.698405537 | 63.05707335 | 63.05707335 | 0.11216621 | 0.872883393 | 0.75505 | 1.010748703 | 9.348133266 | 9.454614739 | 0.727664927 | -0.138068557 | FALSE | TRUE | FALSE |
| ZNF839 | 0.081591 | 0.86693745 | 0.50815 | 0.081590969 | 0.081590969 | 1.603427062 | 0.01335949 | 0.895703278 | 0.0191 | 0.013359487 | 0.013359487 | 0.263090822 | 6.107343078 | 0.785852317 | FALSE | TRUE | FALSE |
| MAN1C1 | 0.0815242 | 0.86694803 | 0.51125 | 0.081524201 | 0.081524201 | 0.689941742 | 1.15328202 | 0.571887229 | 0.0129 | 0.28851652 | 1.153282019 | 4.232300845 | 0.070688869 | -1.150648968 | FALSE | FALSE | FALSE |
| COL8A1 | 0.0814806 | 0.8669586 | 0.51295 | 0.081480644 | 0.081480644 | 0.295856566 | 0.01580311 | 0.894917551 | 1 | 0.01580311 | 0.01580311 | 0.312220269 | 5.155987905 | 0.71231189 | FALSE | TRUE | FALSE |
| ABCA8 | 0.0814365 | 0.86696917 | 0.3726 | 0.009466598 | 0.717350131 | 6.466877037 | 0.0007119 | 0.899640769 | 0.15025 | 0.000711902 | 0.005979549 | 0.013893974 | 114.3929595 | 2.058399296 | FALSE | TRUE | FALSE |
| LZIC | 0.0814323 | 0.86697975 | 1 | 0.081432306 | 0.081432306 | 0.081432306 | 0.09518481 | 0.877102195 | 0.0033 | 0.095184809 | 0.095184809 | 0.095184809 | 0.855517885 | -0.067770907 | FALSE | TRUE | FALSE |
| COX15 | 0.0812643 | 0.86699032 | 0.939 | 0.738722388 | 11.81202247 | 11.81202247 | 0.12974323 | 0.869024442 | 0.87655 | 0.518786994 | 4.73961824 | 4.73961824 | 0.626347042 | -0.20318497 | FALSE | TRUE | FALSE |
| IFT52 | 0.0812538 | 0.86700089 | 1 | 0.324949374 | 0.324949374 | 1.612246492 | 0.10998508 | 0.873349809 | 1 | 0.109985081 | 0.109985081 | 0.109985081 | 0.738770778 | -0.131490291 | FALSE | TRUE | FALSE |
| RMI1 | 0.0812278 | 0.86701145 | 0.01405 | 0.081227777 | 0.081227777 | 1.608775562 | 0.42882499 | 0.781559279 | 0.50945 | 0.428824992 | 0.428824992 | 2.12585537 | 0.18941941 | -0.72257552 | FALSE | TRUE | FALSE |
| APOA4 | 0.0812051 | 0.86702202 | 1 | 0.081205085 | 0.081205085 | 1.607713481 | 3.85135044 | 0.364293188 | 0.2602 | 0.963137553 | 3.85135044 | 3.85135044 | 0.021084834 | -1.676029813 | FALSE | FALSE | FALSE |

| | | | | | | | | | | | | | | | | | |
|---|---|---|---|---|---|---|---|---|---|---|---|---|---|---|---|---|---|
| CHML | 0.081167 | 0.86703258 | 0.75875 | 0.081167044 | 0.324575172 | 1.605291473 | 0.27796193 | 0.831567426 | 0.25625 | 0.277961932 | 2.575510207 | 2.575510207 | 0.29200777 | -0.534605592 | FALSE | TRUE | FALSE |
| DPEP3 | 0.0810597 | 0.86704315 | 0.0146 | 0.081059657 | 0.081059657 | 1.603561069 | 0.10835833 | 0.873772864 | 1 | 0.108358332 | 0.108358332 | 2.146401254 | 0.748070365 | -0.12605755 | FALSE | TRUE | FALSE |
| TRPC7 | 0.0809716 | 0.86705371 | 0.7526 | 0.080971565 | 0.323714106 | 1.585466927 | 1.1296531 | 0.57496619 | 0.50895 | 1.129653104 | 10.73833349 | 22.17509389 | 0.071678257 | -1.144612566 | FALSE | FALSE | FALSE |
| CYB5R3 | 0.0809333 | 0.86706427 | 0.96975 | 1.293875582 | 11.82555392 | 47.30063707 | 0.03451416 | 0.890240744 | 0.00745 | 0.034514161 | 0.034514161 | 0.034514161 | 2.344929286 | 0.370129751 | FALSE | TRUE | FALSE |
| PLK4 | 0.0809271 | 0.86707482 | 1 | 0.080927117 | 0.080927117 | 1.592437022 | 0.42843614 | 0.78167801 | 0.50895 | 0.428436136 | 0.428436136 | 8.446886934 | 0.18888957 | -0.723792023 | FALSE | TRUE | FALSE |
| RALGDS | 0.080926 | 0.86708538 | 0.5092 | 0.728537763 | 0.728537763 | 14.31710001 | 0.11155096 | 0.872985059 | 1 | 0.111550956 | 0.111550956 | 2.196775021 | 0.725461953 | -0.139385359 | FALSE | TRUE | FALSE |
| LCN12 | 0.0809229 | 0.86709594 | 1 | 0.080922867 | 0.080922867 | 0.080922867 | 0.08732489 | 0.878715906 | 1 | 0.087324893 | 0.087324893 | 0.087324893 | 0.926687275 | -0.033066801 | FALSE | TRUE | FALSE |
| NFRKB | 0.0809226 | 0.86710649 | 1 | 0.080922586 | 0.080922586 | 2.724952834 | 0.1084711 | 0.873732686 | 0.03305 | 0.108471098 | 0.108471098 | 2.114151353 | 0.746029008 | -0.127244285 | FALSE | TRUE | FALSE |
| USP24 | 0.080902 | 0.86711704 | 0.06195 | 0.080901987 | 1.551751146 | 2.69231089 | 0.10871688 | 0.873712588 | 0.05805 | 0.10871688 | 2.095573044 | 3.627199117 | 0.744152947 | -0.128337794 | FALSE | TRUE | FALSE |
| KRT18 | 0.0809004 | 0.86712759 | 0.99915 | 7470.435662 | 9495717.534 | 107549221.2 | 0.45844487 | 0.771575629 | 0.99565 | 4152.725874 | 457661.0881 | 4975712.817 | 0.176467005 | -0.753336485 | FALSE | TRUE | FALSE |
| SEC22C | 0.0808056 | 0.86713814 | 1 | 0.080805628 | 0.080805628 | 0.080805628 | 0.09468224 | 0.877188097 | 1 | 0.094682243 | 0.094682243 | 0.094682243 | 0.853440156 | -0.068826926 | FALSE | TRUE | FALSE |
| NEUROD1 | 0.0807823 | 0.86714869 | 1 | 0.080782306 | 0.080782306 | 1.599761217 | 0.10883763 | 0.873672376 | 1 | 0.108837632 | 0.108837632 | 2.157900015 | 0.742227703 | -0.12946284 | FALSE | TRUE | FALSE |
| TMEM68 | 0.0807683 | 0.86715923 | 0.006 | 0.080768341 | 0.080768341 | 0.080768341 | 0.10854869 | 0.873722637 | 0.00595 | 0.108548687 | 0.108548687 | 0.108548687 | 0.744074785 | -0.128383413 | FALSE | TRUE | FALSE |
| RNF20 | 0.0807506 | 0.86716977 | 0.03165 | 0.080750643 | 0.080750643 | 2.71779668 | 0.10704748 | 0.874063466 | 1 | 0.428158378 | 0.428158378 | 8.410370744 | 0.754344207 | -0.122430441 | FALSE | TRUE | FALSE |
| NOLC1 | 0.0806932 | 0.86718032 | 0.01855 | 0.080693156 | 0.080693156 | 1.593457361 | 0.92639534 | 0.594917753 | 0.74825 | 0.926395342 | 8.653388643 | 18.32147159 | 0.087104449 | -1.059959663 | FALSE | TRUE | FALSE |
| TPK1 | 0.0806902 | 0.86719086 | 1 | 0.080690183 | 0.080690183 | 0.080690183 | 0.10704147 | 0.874073464 | 1 | 0.107041468 | 0.428040858 | 0.428040858 | 0.75382172 | -0.122731353 | FALSE | TRUE | FALSE |
| COL3A1 | 0.0806818 | 0.86720139 | 0.33335 | 0.020195574 | 0.32240623 | 1.568911087 | 0.04451155 | 0.887997642 | 0.3388 | 0.044511546 | 0.710883553 | 3.47018567 | 1.812603795 | 0.258302885 | FALSE | TRUE | FALSE |
| STAM2 | 0.0806696 | 0.86721193 | 0.5093 | 0.720492043 | 0.720492043 | 1.599209193 | 0.9686253 | 0.591211465 | 0.515 | 0.968625301 | 0.968625301 | 2.137316252 | 0.083282558 | -1.079445946 | FALSE | TRUE | FALSE |
| PPM1D | 0.0806354 | 0.86722247 | 1 | 0.080635414 | 0.080635414 | 1.593841116 | 3.62201605 | 0.37709219 | 0.00585 | 0.107069796 | 0.107069796 | 2.119322494 | 0.022262578 | -1.652424553 | FALSE | FALSE | FALSE |
| SLC5A12 | 0.0806337 | 0.867233 | 0.7529 | 0.080633714 | 0.32236912 | 1.595227107 | 0.12763965 | 0.869528039 | 0.7493 | 0.127639646 | 0.510342184 | 2.528461346 | 0.631729377 | -0.199468927 | FALSE | TRUE | FALSE |
| KDM4A | 0.0805785 | 0.86724353 | 1 | 773235.2698 | 1148267519 | 13377344784 | 0.07274358 | 0.881847736 | 1 | 1.61713E+13 | 4.4027E+17 | 8.63701E+18 | 1.107705534 | 0.044424325 | FALSE | TRUE | FALSE |
| TOM1 | 0.0805677 | 0.86725406 | 1 | 0.080567693 | 0.080567693 | 1.591729587 | 0.94416281 | 0.59350761 | 0.50325 | 0.944162812 | 0.944162812 | 2.068530709 | 0.085332415 | -1.068885964 | FALSE | TRUE | FALSE |
| TSPAN17 | 0.0804888 | 0.86726459 | 1 | 0.738409306 | 0.738409306 | 1.597769074 | 1.74387613 | 0.513456751 | 0.0084 | 0.436000375 | 1.743876131 | 1.743876131 | 0.046155093 | -1.335780373 | FALSE | FALSE | FALSE |
| DENND6A | 0.0804517 | 0.86727512 | 1 | 0.080451656 | 0.080451656 | 1.590645711 | 0.10791219 | 0.87381302 | 1 | 0.107912188 | 0.107912188 | 2.136513541 | 0.745528912 | -0.12753551 | FALSE | TRUE | FALSE |
| SRI | 0.0803868 | 0.86728565 | 1 | 0.080386848 | 0.080386848 | 0.080386848 | 0.10652073 | 0.874223252 | 1 | 0.425988023 | 0.425988023 | 0.425988023 | 0.754659224 | -0.122249115 | FALSE | TRUE | FALSE |
| EXOSC8 | 0.0803576 | 0.86729617 | 1 | 0.080357575 | 0.080357575 | 0.080357575 | 0.10730084 | 0.873983428 | 0.00735 | 0.107300843 | 0.107300843 | 0.107300843 | 0.748899754 | -0.125576312 | FALSE | TRUE | FALSE |
| HOXD10 | 0.0802818 | 0.86730669 | 0.01045 | 0.080281762 | 0.080281762 | 0.398400123 | 0.12666417 | 0.869719958 | 0.0112 | 0.126664173 | 0.126664173 | 0.62914102 | 0.63381586 | -0.198036897 | FALSE | TRUE | FALSE |
| ADCY1 | 0.080224 | 0.86731721 | 0.52015 | 0.320794631 | 0.320794631 | 6.241323436 | 0.10924004 | 0.873622077 | 1 | 0.109240041 | 0.109240041 | 2.132325426 | 0.734383167 | -0.134077286 | FALSE | TRUE | FALSE |
| ITGA2 | 0.0802009 | 0.86732773 | 0.2646 | 0.020059 | 0.080200923 | 0.393062952 | 2.5239642 | 0.445234957 | 0.07685 | 0.157753525 | 2.523964198 | 5.32236434 | 0.031775777 | -1.497903825 | FALSE | FALSE | FALSE |
| CAPRIN2 | 0.0801788 | 0.86733825 | 0.7577 | 0.080178787 | 0.720644766 | 1.578555484 | 0.01572791 | 0.894931934 | 0.75195 | 0.138988058 | 1.260639832 | 2.741528392 | 5.097866747 | 0.707388479 | FALSE | TRUE | FALSE |
| KRT38 | 0.0801282 | 0.86734877 | 0.87645 | 0.746427332 | 7.131886133 | 7.131886133 | 0.59039642 | 0.716348323 | 0.2563 | 0.147722199 | 0.590396422 | 0.597290888 | 0.135719294 | -0.867358409 | FALSE | TRUE | FALSE |
| SPICE1 | 0.0801222 | 0.86735928 | 1 | 0.08012218 | 0.08012218 | 1.576628825 | 0.10724107 | 0.874003446 | 1 | 0.107241071 | 0.107241071 | 2.114361443 | 0.747122157 | -0.126608384 | FALSE | TRUE | FALSE |
| RMDN3 | 0.0800958 | 0.8673698 | 1 | 0.080095827 | 0.080095827 | 1.584940069 | 0.10718575 | 0.874023459 | 1 | 0.107185749 | 0.107185749 | 2.123700749 | 0.747261899 | -0.126527161 | FALSE | TRUE | FALSE |
| EIF4ENIF1 | 0.0800378 | 0.86738031 | 0.0268 | 0.080037799 | 0.080037799 | 1.569325549 | 0.09309958 | 0.877540111 | 1 | 0.093099584 | 0.093099584 | 1.829748352 | 0.859700926 | -0.065652606 | FALSE | TRUE | FALSE |
| AKAP6 | 0.0799957 | 0.86739082 | 0.05855 | 0.079995749 | 1.538002297 | 2.671781922 | 0.09510124 | 0.877121294 | 1 | 8.284879696 | 798.7827224 | 1552.042499 | 0.841164062 | -0.07511929 | FALSE | TRUE | FALSE |
| LLGL1 | 0.0799373 | 0.86740133 | 0.0359 | 0.07993729 | 0.07993729 | 1.55438649 | 0.10754794 | 0.873903292 | 1 | 0.107547944 | 0.107547944 | 2.098272426 | 0.743271205 | -0.128852692 | FALSE | TRUE | FALSE |
| BCAS4 | 0.0799356 | 0.86741184 | 0.0044 | 0.079935625 | 0.079935625 | 0.079935625 | 0.084734 | 0.879208282 | 1 | 0.084733995 | 0.790813883 | 0.790813883 | 0.943371366 | -0.02531731 | FALSE | TRUE | FALSE |
| NOX3 | 0.0799162 | 0.86742234 | 0.50755 | 0.079916214 | 0.731515825 | 0.731515825 | 1.45079352 | 0.540467294 | 0.22735 | 0.629544389 | 13.70628579 | 54.78207383 | 0.055084485 | -1.258970704 | FALSE | FALSE | FALSE |
| SYT16 | 0.0798611 | 0.86743285 | 1 | 0.079861139 | 0.079861139 | 1.575332439 | 0.10751766 | 0.873923335 | 1 | 0.10751766 | 0.10751766 | 2.124380096 | 0.742772293 | -0.129144304 | FALSE | TRUE | FALSE |
| RRN3 | 0.0797446 | 0.86744335 | 1 | 0.079744644 | 0.079744644 | 1.575245323 | 0.10665599 | 0.874173361 | 0.01525 | 0.106655994 | 0.106655994 | 2.109964772 | 0.747680846 | -0.126283745 | FALSE | TRUE | FALSE |
| MMP7 | 0.0797255 | 0.86745385 | 1 | 0.079725454 | 0.079725454 | 0.079725454 | 0.94005339 | 0.594159442 | 0.5016 | 0.940053386 | 0.940053386 | 0.940053386 | 0.084809496 | -1.071555519 | FALSE | TRUE | FALSE |
| GH2 | 0.0797162 | 0.86746435 | 1 | 0.731697884 | 0.731697884 | 0.731697884 | 0.10721479 | 0.874013453 | 1 | 0.987965786 | 0.987965786 | 0.987965786 | 0.743518214 | -0.128708388 | FALSE | TRUE | FALSE |
| ETHE1 | 0.0796763 | 0.86747485 | 1 | 0.318509373 | 0.318509373 | 0.318509373 | 0.10744481 | 0.873953388 | 0.50755 | 0.429546468 | 0.429546468 | 0.429546468 | 0.741555238 | -0.129856493 | FALSE | TRUE | FALSE |
| ALAD | 0.0796682 | 0.86748534 | 0.75815 | 0.079668187 | 0.318466154 | 0.395288717 | 0.49422157 | 0.757169695 | 0.6954 | 0.494221566 | 7.904774466 | 7.904774466 | 0.161199334 | -0.792636756 | FALSE | TRUE | FALSE |
| VPS53 | 0.0796638 | 0.86749584 | 1 | 0.714913947 | 0.714913947 | 14.08619741 | 77.7897476 | 0.037325791 | 0.2265 | 14.17298317 | 311.1499626 | 1244.874882 | 0.001024092 | -2.989661195 | FALSE | FALSE | FALSE |
| MPI | 0.0796408 | 0.86750633 | 0.8181 | 0.318371481 | 1.273037787 | 5.091630895 | 0.00949758 | 0.896875581 | 1 | 0.151706411 | 0.606543807 | 0.753331017 | 8.385374021 | 0.923522439 | FALSE | TRUE | FALSE |
| MORN3 | 0.0796251 | 0.86751682 | 1 | 1.273550108 | 5.09520828 | 5.09520828 | 0.9817037 | 0.590221055 | 0.50795 | 0.981703702 | 0.981703702 | 0.981703702 | 0.081109071 | -1.090930573 | FALSE | TRUE | FALSE |
| FBXO43 | 0.079508 | 0.86752731 | 1 | 0.079507952 | 0.079507952 | 1.571731856 | 0.10661085 | 0.8742033 | 1 | 0.106610853 | 0.106610853 | 2.11044667 | 0.74577728 | -0.127390852 | FALSE | TRUE | FALSE |
| CIR1 | 0.0795023 | 0.8675378 | 1 | 0.079502334 | 0.079502334 | 1.577929797 | 8.29781467 | 0.221899674 | 0.25675 | 0.899898888 | 8.297814673 | 8.297814673 | 0.009581117 | -2.018583851 | FALSE | FALSE | FALSE |
| SLC5A1 | 0.0794992 | 0.86754829 | 0.4989 | 0.019892076 | 0.317800073 | 0.673107083 | 0.00949709 | 0.896882528 | 1 | 0.037951177 | 0.037951177 | 0.187505821 | 8.370899391 | 0.922772122 | FALSE | TRUE | FALSE |
| NTN4 | 0.0794634 | 0.86755877 | 0.51045 | 0.317727825 | 0.317727825 | 2.687102824 | 35.0525434 | 0.080174172 | 0.19555 | 3.686812952 | 140.1957941 | 140.1957941 | 0.002266979 | -2.644552544 | FALSE | FALSE | FALSE |
| HMCES | 0.0793997 | 0.86756926 | 1 | 0.079399714 | 0.079399714 | 1.57655218 | 0.10682451 | 0.87412343 | 0.01055 | 0.106824513 | 0.106824513 | 2.122933834 | 0.743272417 | -0.128851984 | FALSE | TRUE | FALSE |
| PDE12 | 0.0793046 | 0.86757974 | 1 | 0.079304558 | 0.079304558 | 1.560209213 | 0.10046054 | 0.875809922 | 1 | 0.930443488 | 8.844540563 | 18.34116724 | 0.789410035 | -0.102697357 | FALSE | TRUE | FALSE |
| SMG6 | 0.0792355 | 0.86759022 | 1 | 0.721243516 | 6.73818486 | 130.5582834 | 0.44770462 | 0.775069132 | 0.5203 | 0.447704624 | 0.447704624 | 8.707318107 | 0.176981753 | -0.752071507 | FALSE | TRUE | FALSE |
| CPB1 | 0.0791807 | 0.8676007 | 1 | 0.079180664 | 0.079180664 | 1.569433411 | 0.41993173 | 0.784574933 | 0.01005 | 0.419931726 | 0.419931726 | 2.083368904 | 0.188556041 | -0.724559549 | FALSE | TRUE | FALSE |
| ACTL6A | 0.0791644 | 0.86761118 | 0.8789 | 1.265126698 | 5.059398748 | 5.059398748 | 0.10574946 | 0.874492015 | 1 | 1.69025689 | 27.04340614 | 27.04340614 | 0.748603234 | -0.125748301 | FALSE | TRUE | FALSE |
| TESK2 | 0.0791401 | 0.86762166 | 0.74825 | 0.07914011 | 0.316445524 | 1.562688857 | 0.125237 | 0.87008102 | 0.75835 | 0.125237002 | 0.500802349 | 2.476692469 | 0.631922746 | -0.199336012 | FALSE | TRUE | FALSE |
| CXCR1 | 0.0791377 | 0.86763213 | 0.7501 | 0.316453952 | 1.265745301 | 1.265745301 | 0.10882859 | 0.873692485 | 1 | 0.108828592 | 0.108828592 | 0.108828592 | 0.727177234 | -0.138359726 | FALSE | TRUE | FALSE |
| ITFG2 | 0.0791323 | 0.8676426 | 0.51115 | 0.725836936 | 0.725836936 | 1.565722056 | 0.10984677 | 0.873380118 | 1 | 0.109846769 | 0.109846769 | 2.176239522 | 0.720387671 | -0.142433729 | FALSE | TRUE | FALSE |
| TIGD3 | 0.0791104 | 0.86765307 | 0.01655 | 0.079110412 | 0.079110412 | 1.562510658 | 0.1058319 | 0.874422442 | 1 | 0.105831903 | 0.105831903 | 2.09341834 | 0.747510052 | -0.126382963 | FALSE | TRUE | FALSE |
| RUSC1 | 0.0791016 | 0.86766354 | 1 | 1.03575E+51 | 2.17206E+61 | 1.93625E+61 | 0.27363618 | 0.832696136 | 1 | 9.64997E+37 | 5.66467E+46 | 3.92205E+50 | 0.289075764 | -0.538988319 | FALSE | TRUE | FALSE |
| SLC25A46 | 0.0790779 | 0.86767401 | 0.0102 | 0.07907788 | 0.07907788 | 1.570119905 | 0.10536207 | 0.874621019 | 0.50315 | 0.421290221 | 0.421290221 | 0.421290221 | 0.750534618 | -0.124629272 | FALSE | TRUE | FALSE |
| STAT6 | 0.079052 | 0.86768448 | 1 | 0.079052001 | 0.079052001 | 1.553751147 | 0.10674602 | 0.874133419 | 0.0217 | 0.106746023 | 0.106746023 | 2.102431389 | 0.740561557 | -0.130438836 | FALSE | TRUE | FALSE |
| PNPLA1 | 0.0790034 | 0.86769494 | 1 | 0.718975941 | 0.718975941 | 1.564722991 | 0.1095001 | 0.87350121 | 0.0138 | 0.109500104 | 0.109500104 | 2.171267682 | 0.721491316 | -0.141768892 | FALSE | TRUE | FALSE |
| CFHR2 | 0.0789724 | 0.86770541 | 1 | 0.710540681 | 6.561551188 | 6.561551188 | 82.1437428 | 0.035181095 | 0.12915 | 2.046536565 | 82.14374279 | 82.14374279 | 0.000961392 | -3.017099408 | FALSE | FALSE | FALSE |
| SSX2IP | 0.0789553 | 0.86771587 | 1 | 0.078955338 | 0.078955338 | 1.56287039 | 0.10564927 | 0.874531736 | 0.0119 | 0.10564927 | 0.10564927 | 2.093846267 | 0.747334442 | -0.126485002 | FALSE | TRUE | FALSE |
| FABP2 | 0.0789118 | 0.86772633 | 1 | 0.078911817 | 0.078911817 | 0.078911817 | 0.08451335 | 0.879226792 | 1 | 0.084513346 | 0.084513346 | 0.084513346 | 0.933720186 | -0.029783252 | FALSE | TRUE | FALSE |
| MFSD2A | 0.0788869 | 0.86773679 | 0.7558 | 0.735465257 | 7.037339602 | 14.53529077 | 0.1064836 | 0.874233226 | 1 | 0.994891461 | 9.540170792 | 19.69042017 | 0.740836416 | -0.130277678 | FALSE | TRUE | FALSE |
| MRS2 | 0.078839 | 0.86774724 | 1 | 0.078839005 | 0.078839005 | 1.561925299 | 0.10556804 | 0.874551587 | 0.01335 | 0.105568041 | 0.105568041 | 2.093839719 | 0.746807503 | -0.126791327 | FALSE | TRUE | FALSE |
| MCM3 | 0.0788041 | 0.8677577 | 1 | 0.078804112 | 0.078804112 | 1.547325888 | 0.1057561 | 0.874472145 | 1 | 0.105756101 | 0.105756101 | 2.081092571 | 0.745149565 | -0.127756548 | FALSE | TRUE | FALSE |
| NNT | 0.0787444 | 0.86776815 | 0.24615 | 0.03445341 | 1.256361848 | 5.020247186 | 7.6476E-06 | 0.900777988 | 0.94005 | 0.006517384 | 0.10399006 | 0.128076309 | 10296.65588 | 4.012696199 | FALSE | TRUE | FALSE |
| DYNC1I1 | 0.0786971 | 0.86777861 | 0.51205 | 0.722146881 | 0.722146881 | 2.66049737 | 0.1093322 | 0.873581813 | 1 | 0.109332196 | 0.109332196 | 2.159417075 | 0.719797814 | -0.142789476 | FALSE | TRUE | FALSE |
| B4GALNT2 | 0.0786561 | 0.86778906 | 1 | 0.078656122 | 0.078656122 | 1.553748711 | 0.10557049 | 0.874541662 | 1 | 0.105570495 | 0.105570495 | 2.088497127 | 0.745057811 | -0.127810028 | FALSE | TRUE | FALSE |

| | | | | | | | | | | | | | | | | | |
|---|---|---|---|---|---|---|---|---|---|---|---|---|---|---|---|---|---|
| RPUSD3 | 0.0786051 | 0.86779951 | 1 | 0.078605133 | 0.078605133 | 1.559072869 | 0.10520562 | 0.874650754 | 1 | 0.105205618 | 0.105205618 | 0.105205618 | 0.747157186 | -0.126588022 | FALSE | TRUE | FALSE |
| TTC24 | 0.0785979 | 0.86780996 | 1 | 0.707313354 | 6.533052694 | 6.533052694 | 81.8585664 | 0.035568856 | 0.004 | 8.595522704 | 81.85856638 | 81.85856638 | 0.000960168 | -3.01765299 | FALSE | FALSE | FALSE |
| PDZRN3 | 0.0785512 | 0.8678204 | 1 | 0.314085646 | 0.314085646 | 6.120657162 | 0.10725957 | 0.873993438 | 1 | 0.107259572 | 0.107259572 | 2.096649691 | 0.732346325 | -0.135283494 | FALSE | TRUE | FALSE |
| DHRS13 | 0.0785104 | 0.86783085 | 1 | 0.722826804 | 0.722826804 | 1.555894086 | 0.43161019 | 0.780544649 | 0.504 | 0.431610186 | 0.431610186 | 2.141483586 | 0.181901248 | -0.740164322 | FALSE | TRUE | FALSE |
| WRAP73 | 0.0785102 | 0.86784129 | 0.01445 | 0.078510186 | 0.078510186 | 1.551107678 | 0.10567238 | 0.874521808 | 0.01305 | 0.105672377 | 0.105672377 | 2.090799796 | 0.742958451 | -0.129035473 | FALSE | TRUE | FALSE |
| BNIP1 | 0.0783942 | 0.86785173 | 0.00835 | 0.078394239 | 0.078394239 | 0.078394239 | 0.1048741 | 0.874769556 | 0.00705 | 0.104874102 | 0.104874102 | 0.104874102 | 0.747508089 | -0.126384104 | FALSE | TRUE | FALSE |
| PFKFB4 | 0.0783439 | 0.86786217 | 0.0156 | 0.078343898 | 0.078343898 | 0.664079083 | 0.03142705 | 0.890875621 | 0.0136 | 0.031427045 | 0.031427045 | 0.621671279 | 2.492881462 | 0.396701628 | FALSE | TRUE | FALSE |
| PLCB3 | 0.0782873 | 0.86787261 | 1 | 0.078287321 | 0.078287321 | 1.517909921 | 0.10439908 | 0.874888147 | 1 | 0.417523344 | 0.417523344 | 8.125168983 | 0.749885198 | -0.125005219 | FALSE | TRUE | FALSE |
| PMEL | 0.0782724 | 0.86788305 | 0.50755 | 0.700970453 | 0.700970453 | 2.648983317 | 0.1049571 | 0.874749771 | 1 | 0.946228189 | 0.946228189 | 3.554047418 | 0.745756087 | -0.127403193 | FALSE | TRUE | FALSE |
| TM2D3 | 0.0782656 | 0.86789348 | 1 | 0.078265642 | 0.078265642 | 0.078265642 | 0.10539961 | 0.874611105 | 1 | 0.10539961 | 0.10539961 | 0.10539961 | 0.742561024 | -0.12926785 | FALSE | TRUE | FALSE |
| CLN6 | 0.0782402 | 0.86790392 | 0.6901 | 0.177385636 | 2.833749147 | 2.833749147 | 0.03853454 | 0.8893391 | 0.00815 | 0.03853454 | 0.03853454 | 0.03853454 | 2.030392323 | 0.307579963 | FALSE | TRUE | FALSE |
| PRKCQ | 0.0782256 | 0.86791435 | 1 | 0.312761933 | 0.312761933 | 6.159975104 | 0.95470024 | 0.59230758 | 0.26315 | 0.954700235 | 3.817290221 | 13.8574603 | 0.081937304 | -1.086518328 | FALSE | TRUE | FALSE |
| MMP26 | 0.0781772 | 0.86792478 | 1 | 0.078177157 | 0.078177157 | 0.078177157 | 0.10478842 | 0.874779447 | 1 | 0.104788421 | 0.104788421 | 0.104788421 | 0.746047667 | -0.127233423 | FALSE | TRUE | FALSE |
| MCOLN1 | 0.078172 | 0.86793521 | 0.0228 | 0.078171966 | 0.078171966 | 1.537620917 | 0.10545643 | 0.874581353 | 1 | 0.105456429 | 0.105456429 | 2.07842245 | 0.74127264 | -0.130022029 | FALSE | TRUE | FALSE |
| FOXB1 | 0.0781318 | 0.86794564 | 1 | 28.5518183 | 1134.146242 | 4535.793342 | 0.11752808 | 0.871848343 | 0.7564 | 0.469834469 | 1.878696735 | 2.32690481 | 0.664792857 | -0.177313656 | FALSE | TRUE | FALSE |
| RASGRP3 | 0.0781309 | 0.86795606 | 0.01825 | 0.07813093 | 0.07813093 | 1.543241265 | 0.10540821 | 0.874601189 | 1 | 0.105408209 | 0.105408209 | 2.085128326 | 0.74122244 | -0.130051441 | FALSE | TRUE | FALSE |
| CDH2 | 0.078115 | 0.86796649 | 1 | 0.312312666 | 0.312312666 | 6.138166503 | 0.10350683 | 0.875055807 | 1 | 0.968220121 | 0.968220121 | 19.06896032 | 0.754684507 | -0.122234566 | FALSE | TRUE | FALSE |
| SC5D | 0.078113 | 0.86797691 | 1 | 0.078113032 | 0.078113032 | 0.078113032 | 0.10544555 | 0.874591272 | 1 | 0.105445546 | 0.105445546 | 0.105445546 | 0.740790253 | -0.130304741 | FALSE | TRUE | FALSE |
| EIF2A | 0.0780023 | 0.86798733 | 1 | 0.078002342 | 0.078002342 | 1.54688279 | 0.10164711 | 0.875556233 | 1 | 0.101647114 | 0.919565146 | 2.017833525 | 0.767383734 | -0.114987411 | FALSE | TRUE | FALSE |
| PRSS54 | 0.077952 | 0.86799775 | 0.0121 | 0.077951981 | 0.077951981 | 1.546490151 | 8.69734637 | 0.21517464 | 0.25095 | 0.915612966 | 8.697346367 | 8.697346367 | 0.008962732 | -2.047559608 | FALSE | FALSE | FALSE |
| JMJD1C | 0.077937 | 0.86800817 | 0.06265 | 0.077937048 | 1.492737981 | 2.599142208 | 0.10534335 | 0.874630932 | 0.05435 | 0.105343348 | 2.027971298 | 3.521218922 | 0.739838345 | -0.130863163 | FALSE | TRUE | FALSE |
| GRB7 | 0.0779333 | 0.86801859 | 0.51295 | 0.0779333 | 0.0779333 | 0.660067878 | 0.0308288 | 0.891067883 | 0.51415 | 0.123259994 | 0.123259994 | 1.044418181 | 2.527937897 | 0.402766401 | FALSE | TRUE | FALSE |
| PPIAL4G | 0.0779207 | 0.868029 | 1 | 0.077920673 | 0.077920673 | 0.077920673 | 0.07915739 | 0.880573157 | 0.5078 | 0.745673957 | 0.745673957 | 0.745673957 | 0.984376495 | -0.006838765 | FALSE | TRUE | FALSE |
| PHLDB2 | 0.0778669 | 0.86803942 | 0.75485 | 0.173956784 | 0.695587017 | 5.855803374 | 0.26543919 | 0.835127062 | 0.81865 | 1.061429599 | 39.16158495 | 83.16228541 | 0.293351129 | -0.532612237 | FALSE | TRUE | FALSE |
| KIAA0319L | 0.0778459 | 0.86804983 | 1 | 0.077845939 | 0.077845939 | 1.528454229 | 0.1016309 | 0.875566008 | 0.509 | 0.925255001 | 0.925255001 | 18.20677056 | 0.76596722 | -0.115789816 | FALSE | TRUE | FALSE |
| ZNF461 | 0.0778208 | 0.86806024 | 1 | 0.077820796 | 0.077820796 | 1.542433561 | 0.10141357 | 0.875585554 | 0.50365 | 0.916118667 | 0.916118667 | 2.012221979 | 0.767360753 | -0.115000417 | FALSE | TRUE | FALSE |
| THSD1 | 0.0777841 | 0.86807065 | 1 | 0.077784121 | 0.077784121 | 1.528228865 | 0.10446766 | 0.87484864 | 1 | 0.104467657 | 0.104467657 | 2.056842708 | 0.744576107 | -0.128090904 | FALSE | TRUE | FALSE |
| PAX4 | 0.0777658 | 0.86808106 | 0.5003 | 0.077765837 | 0.077765837 | 0.077765837 | 0.12871319 | 0.86931419 | 0.8793 | 1.228006562 | 12.0166127 | 12.0166127 | 0.604179217 | -0.218834218 | FALSE | TRUE | FALSE |
| PLEKHH2 | 0.0777654 | 0.86809146 | 0.0573 | 0.009286789 | 0.181049114 | 0.653069239 | 0.00043099 | 0.899818904 | 1 | 0.000430992 | 0.000430992 | 0.014489822 | 180.4334796 | 2.256317124 | FALSE | TRUE | FALSE |
| TC2N | 0.0777274 | 0.86810187 | 0.01285 | 0.077727438 | 0.077727438 | 1.541734337 | 0.91682658 | 0.596534304 | 0.4997 | 0.101337303 | 0.916826578 | 0.927778631 | 0.084778779 | -1.07171284 | FALSE | TRUE | FALSE |
| CFB | 0.0777229 | 0.86811227 | 0.50545 | 0.310770884 | 0.310770884 | 6.114997971 | 0.10502216 | 0.874710182 | 0.5138 | 0.419944909 | 0.419944909 | 3.551706815 | 0.740061591 | -0.130732135 | FALSE | TRUE | FALSE |
| PTPN4 | 0.0777062 | 0.86812267 | 1 | 0.722275293 | 0.722275293 | 14.20929411 | 0.10837476 | 0.873762822 | 0.02035 | 0.108374765 | 0.108374765 | 2.136249445 | 0.7170135 | -0.144472667 | FALSE | TRUE | FALSE |
| SMAD3 | 0.0776761 | 0.86813307 | 1 | 0.310519048 | 0.310519048 | 1.536585286 | 0.10628689 | 0.874283072 | 0.01205 | 0.106286885 | 0.106286885 | 2.105319114 | 0.73081591 | -0.136192006 | FALSE | TRUE | FALSE |
| CPO | 0.0776551 | 0.86814347 | 1 | 0.691617162 | 0.691617162 | 1.542687307 | 0.1010015 | 0.875692954 | 1 | 0.905320028 | 8.328729403 | 8.328729403 | 0.768850456 | -0.114158124 | FALSE | TRUE | FALSE |
| DPYSL3 | 0.0776423 | 0.86815386 | 0.8755 | 0.310301433 | 1.240445823 | 2.629533437 | 0.03170262 | 0.890829385 | 0.51185 | 0.126687044 | 0.126687044 | 0.627222677 | 2.449081163 | 0.389003178 | FALSE | TRUE | FALSE |
| LIMD1 | 0.0776246 | 0.86816426 | 1 | 0.077624638 | 0.077624638 | 1.531158339 | 0.10174927 | 0.875526901 | 1 | 0.948089689 | 0.948089689 | 3.444232455 | 0.762901146 | -0.117531733 | FALSE | TRUE | FALSE |
| MAN1A1 | 0.0776016 | 0.86817465 | 0.01785 | 0.077601621 | 0.077601621 | 1.53050929 | 0.40528745 | 0.790381868 | 0.87885 | 1.621005324 | 6.485063895 | 13.72102 | 0.191473042 | -0.717892363 | FALSE | TRUE | FALSE |
| COPA | 0.0775439 | 0.86818504 | 1 | 0.077543876 | 0.077543876 | 2.610260511 | 0.10496648 | 0.874739876 | 1 | 0.104966476 | 0.104966476 | 2.052270799 | 0.738748975 | -0.131503109 | FALSE | TRUE | FALSE |
| ZNF790 | 0.0775118 | 0.86819543 | 0.75885 | 0.077511804 | 0.675707334 | 1.533426705 | 0.12565202 | 0.869985629 | 0.6928 | 1.107152412 | 92.70485128 | 92.70485128 | 0.616876688 | -0.209801641 | FALSE | TRUE | FALSE |
| IKBKB | 0.0775106 | 0.86820582 | 1 | 0.077510611 | 0.077510611 | 1.525141276 | 0.10134825 | 0.875614862 | 1 | 0.927369397 | 0.927369397 | 3.423361309 | 0.764794791 | -0.116455079 | FALSE | TRUE | FALSE |
| OSBPL7 | 0.0774727 | 0.8682162 | 0.0289 | 0.077472696 | 0.077472696 | 1.518460127 | 0.41230309 | 0.787505574 | 0.51095 | 0.41230309 | 0.41230309 | 8.100583288 | 0.187902292 | -0.726067923 | FALSE | TRUE | FALSE |
| CTAGE1 | 0.0772925 | 0.86822659 | 1 | 0.07729252 | 0.07729252 | 1.526185355 | 0.9044274 | 0.597819498 | 0.50065 | 0.904427398 | 0.904427398 | 3.405449182 | 0.08546017 | -1.068236245 | FALSE | TRUE | FALSE |
| ANKS1B | 0.0772888 | 0.86823697 | 1 | 0.70429064 | 0.70429064 | 13.77823681 | 0.1074532 | 0.873943372 | 0.0267 | 0.107453198 | 0.107453198 | 2.107666653 | 0.719278505 | -0.143102918 | FALSE | TRUE | FALSE |
| OR4C15 | 0.0772162 | 0.86824736 | 1 | 0.686681037 | 0.686681037 | 0.686681037 | 0.106627 | 0.874183342 | 1 | 0.106627001 | 0.106627001 | 0.106627001 | 0.724171089 | -0.140158818 | FALSE | TRUE | FALSE |
| TOPBP1 | 0.0771977 | 0.86825774 | 0.0373 | 0.077197652 | 0.077197652 | 2.592949817 | 0.10453423 | 0.87483876 | 1 | 0.104534229 | 0.104534229 | 3.516256718 | 0.738491626 | -0.131654425 | FALSE | TRUE | FALSE |
| P2RY6 | 0.0771753 | 0.86826812 | 1 | 0.308631785 | 1.234558514 | 1.528560584 | 0.10629493 | 0.874273106 | 1 | 0.106294932 | 0.106294932 | 2.107766684 | 0.72604849 | -0.139034373 | FALSE | TRUE | FALSE |
| FCRLA | 0.0771622 | 0.86827849 | 1 | 0.686038062 | 6.260372784 | 6.260372784 | 0.10978499 | 0.873420507 | 0.00945 | 0.109784992 | 0.109784992 | 0.109784992 | 0.702848305 | -0.153138398 | FALSE | TRUE | FALSE |
| PCDHB10 | 0.0769988 | 0.86828887 | 0.89315 | 6.265607377 | 234.6320229 | 2251.143652 | 0.15236951 | 0.863966463 | 0.5142 | 0.152369512 | 0.152369512 | 2.993392606 | 0.505342381 | -0.296414278 | FALSE | TRUE | FALSE |
| KCNS2 | 0.0769924 | 0.86829925 | 0.5105 | 0.076992442 | 0.076992442 | 0.653095115 | 0.12015015 | 0.871194761 | 0.8737 | 0.480277598 | 1.920299063 | 2.377057432 | 0.640801868 | -0.193276231 | FALSE | TRUE | FALSE |
| GSS | 0.0769703 | 0.86830962 | 0.5059 | 0.076970267 | 0.076970267 | 0.168813562 | 0.13526226 | 0.86772188 | 0.01285 | 0.135262261 | 0.135262261 | 0.295309285 | 0.569044655 | -0.244853651 | FALSE | TRUE | FALSE |
| IST1 | 0.0768125 | 0.86831999 | 1 | 0.076812488 | 0.076812488 | 0.076812488 | 0.1033614 | 0.875105037 | 1 | 0.103361398 | 0.103361398 | 0.103361398 | 0.743141481 | -0.128926544 | FALSE | TRUE | FALSE |
| CKAP5 | 0.0767812 | 0.86833036 | 1 | 0.076781192 | 1.482510006 | 2.569922562 | 0.10385594 | 0.874957234 | 0.045 | 0.103855941 | 0.103855941 | 3.482647642 | 0.739304764 | -0.131176495 | FALSE | TRUE | FALSE |
| METTL3 | 0.0767593 | 0.86834073 | 1 | 0.076759347 | 0.076759347 | 1.516607022 | 0.10013983 | 0.875887786 | 0.5125 | 0.902676499 | 0.902676499 | 1.981438534 | 0.766521628 | -0.115475586 | FALSE | TRUE | FALSE |
| ARAP3 | 0.0766838 | 0.8683511 | 0.5242 | 0.076683809 | 0.076683809 | 1.479576939 | 0.01548792 | 0.895003795 | 0.0393 | 0.015487917 | 0.015487917 | 0.300103685 | 4.951202384 | 0.694710679 | FALSE | TRUE | FALSE |
| SPATA6 | 0.0766636 | 0.86836147 | 1 | 0.076663635 | 0.076663635 | 1.518890017 | 0.8715402 | 0.602779513 | 0.75365 | 0.871540196 | 8.036344787 | 17.28677853 | 0.087963396 | -1.055698014 | FALSE | TRUE | FALSE |
| AK2 | 0.0766511 | 0.86837183 | 0.0074 | 0.076651114 | 0.076651114 | 0.076651114 | 0.1035134 | 0.875045956 | 0.0067 | 0.103513402 | 0.103513402 | 0.103513402 | 0.740494592 | -0.130478109 | FALSE | TRUE | FALSE |
| HOXD13 | 0.0766511 | 0.86838219 | 0.5049 | 0.306472053 | 0.306472053 | 1.520275029 | 0.41450454 | 0.786717129 | 0.50545 | 0.41450454 | 0.41450454 | 2.05812877 | 0.184922154 | -0.733011057 | FALSE | TRUE | FALSE |
| MKKS | 0.0766389 | 0.86839255 | 0.75365 | 0.076638945 | 0.306393518 | 0.379853102 | 0.11700498 | 0.871972129 | 0.93965 | 1.074244895 | 17.17704799 | 17.17704799 | 0.655005842 | -0.183754827 | FALSE | TRUE | FALSE |
| SIRT4 | 0.0765508 | 0.86840291 | 1 | 0.076550778 | 0.076550778 | 1.519154044 | 0.10303187 | 0.875193558 | 1 | 0.103031867 | 0.103031867 | 0.103031867 | 0.742981566 | -0.129021961 | FALSE | TRUE | FALSE |
| ZBTB11 | 0.0764974 | 0.86841327 | 1 | 0.69644055 | 0.69644055 | 13.63475665 | 0.1032616 | 0.875144394 | 1 | 0.944660197 | 0.944660197 | 18.54137225 | 0.740812025 | -0.130291976 | FALSE | TRUE | FALSE |
| CDH18 | 0.0764655 | 0.86842363 | 1 | 0.710783677 | 0.710783677 | 14.00073648 | 0.10127457 | 0.875624628 | 0.87305 | 1.619483553 | 15.09143599 | 18.62333766 | 0.755031215 | -0.122035093 | FALSE | TRUE | FALSE |
| DHRS4L2 | 0.0764611 | 0.86843399 | 0.74895 | 0.305669314 | 1.222283331 | 1.222283331 | 0.10587689 | 0.874382652 | 0.00285 | 0.105876888 | 0.105876888 | 0.105876888 | 0.722169577 | -0.141360811 | FALSE | TRUE | FALSE |
| LRRK1 | 0.0764532 | 0.86844434 | 0.15695 | 0.008843995 | 0.07645324 | 0.641053221 | 0.00133348 | 0.899282669 | 0.76405 | 0.011649194 | 0.046557665 | 0.893008804 | 57.33376845 | 1.758410488 | FALSE | TRUE | FALSE |
| CYP2A6 | 0.076452 | 0.86845469 | 1 | 0.076451978 | 0.076451978 | 1.509318204 | 75.344459 | 0.037934071 | 0.31885 | 7.832331478 | 742.9336796 | 742.9336796 | 0.001014699 | -2.993662595 | FALSE | FALSE | FALSE |
| KY | 0.0764181 | 0.86846504 | 0.01915 | 0.076418115 | 0.076418115 | 1.50720543 | 0.10312371 | 0.87515423 | 1 | 0.103123709 | 0.103123709 | 2.037312026 | 0.741033422 | -0.130162204 | FALSE | TRUE | FALSE |
| PARVG | 0.0763983 | 0.86847539 | 1 | 0.305475579 | 0.305475579 | 1.516998705 | 0.10443431 | 0.874858519 | 1 | 0.104434312 | 0.104434312 | 0.104434312 | 0.731543749 | -0.135759696 | FALSE | TRUE | FALSE |
| SLC9A5 | 0.0763498 | 0.86848574 | 1 | 0.076349788 | 0.076349788 | 1.496575101 | 0.09978974 | 0.875965562 | 0.51055 | 0.903651743 | 0.903651743 | 17.75548347 | 0.76510662 | -0.11627804 | FALSE | TRUE | FALSE |
| ANXA11 | 0.0763391 | 0.86849608 | 1 | 0.682335491 | 6.259724805 | 13.4852448 | 0.10893952 | 0.873642201 | 1 | 0.108939523 | 0.108939523 | 2.156076384 | 0.700747197 | -0.154438631 | FALSE | TRUE | FALSE |
| AEN | 0.0762314 | 0.86850643 | 1 | 0.076231374 | 0.076231374 | 1.512118158 | 0.10262616 | 0.875311404 | 0.0088 | 0.102626162 | 0.102626162 | 0.102626162 | 0.742806437 | -0.129124341 | FALSE | TRUE | FALSE |
| THAP2 | 0.0761824 | 0.86851677 | 0.00595 | 0.076182372 | 0.076182372 | 0.076182372 | 0.10304667 | 0.875183728 | 1 | 0.103046666 | 0.103046666 | 0.103046666 | 0.739299725 | -0.131179456 | FALSE | TRUE | FALSE |
| MOB1B | 0.0761787 | 0.86852711 | 1 | 0.076178702 | 0.076178702 | 0.076178702 | 0.1024783 | 0.875360443 | 1 | 0.102478299 | 0.102478299 | 0.102478299 | 0.743364233 | -0.128798339 | FALSE | TRUE | FALSE |
| CYP2E1 | 0.0761471 | 0.86853745 | 1 | 0.076147106 | 0.076147106 | 1.505233335 | 0.10294449 | 0.875213214 | 1 | 0.102944492 | 0.102944492 | 2.037795938 | 0.73969092 | -0.130949713 | FALSE | TRUE | FALSE |
| CDCA7L | 0.0761189 | 0.86854779 | 1 | 0.076118881 | 0.076118881 | 1.507027462 | 0.09940599 | 0.87609175 | 1 | 0.89611783 | 0.89611783 | 1.970460698 | 0.765737333 | -0.115920179 | FALSE | TRUE | FALSE |

| | | | | | | | | | | | | | | | | | |
|---|---|---|---|---|---|---|---|---|---|---|---|---|---|---|---|---|---|
| LSP1 | 0.0760651 | 0.86855813 | 1 | 0.689680499 | 6.418171983 | 6.418171983 | 0.09977183 | 0.875984992 | 0.0073 | 0.099771829 | 0.099771829 | 0.099771829 | 0.762390212 | -0.117822688 | FALSE | TRUE | FALSE |
| UBA6 | 0.0760528 | 0.86856846 | 1 | 0.076052806 | 0.076052806 | 1.49452613 | 0.1028875 | 0.875223039 | 1 | 0.102887503 | 0.102887503 | 2.026104925 | 0.7391841 | -0.131247384 | FALSE | TRUE | FALSE |
| PUS7 | 0.0760343 | 0.8685788 | 1 | 0.076034287 | 0.076034287 | 1.50236354 | 0.10235736 | 0.875389848 | 1 | 0.10235736 | 0.10235736 | 2.025413801 | 0.742831654 | -0.129109598 | FALSE | TRUE | FALSE |
| HSD17B12 | 0.0759701 | 0.86858913 | 0.00965 | 0.075970149 | 0.075970149 | 0.075970149 | 0.10272422 | 0.875262328 | 0.008 | 0.102724218 | 0.102724218 | 0.102724218 | 0.739554421 | -0.131029862 | FALSE | TRUE | FALSE |
| ZNF578 | 0.0758906 | 0.86859946 | 1 | 0.07589061 | 0.07589061 | 1.501714514 | 0.10197064 | 0.875497555 | 0.0142 | 0.101970643 | 0.101970643 | 2.020357571 | 0.744239789 | -0.128287115 | FALSE | TRUE | FALSE |
| SLC16A11 | 0.0758818 | 0.86860979 | 0.0107 | 0.075881757 | 0.075881757 | 1.504878318 | 7.98384293 | 0.228560598 | 0.0073 | 0.864772095 | 7.983842931 | 7.983842931 | 0.009504415 | -2.022074605 | FALSE | FALSE | FALSE |
| ZNF341 | 0.0758037 | 0.86862012 | 1 | 0.075803722 | 0.075803722 | 1.48520781 | 0.09633217 | 0.876853405 | 1 | 0.877397169 | 8.201965731 | 17.23282778 | 0.786899364 | -0.104080806 | FALSE | TRUE | FALSE |
| DHRS7C | 0.0757896 | 0.86863044 | 1 | 0.075789587 | 0.075789587 | 0.075789587 | 0.10258455 | 0.875321214 | 0.00855 | 0.102584545 | 0.102584545 | 0.102584545 | 0.738801215 | -0.131472399 | FALSE | TRUE | FALSE |
| CRNN | 0.0757846 | 0.86864077 | 1 | 0.075784617 | 0.075784617 | 1.501986158 | 0.0988108 | 0.876246719 | 0.50445 | 0.884926632 | 0.884926632 | 1.960475251 | 0.766966971 | -0.115223338 | FALSE | TRUE | FALSE |
| ZNF222 | 0.0757809 | 0.86865109 | 1 | 0.680190718 | 0.680190718 | 1.504582469 | 0.92193487 | 0.59610449 | 0.5065 | 0.92193487 | 0.92193487 | 0.92193487 | 0.082197687 | -1.085140401 | FALSE | TRUE | FALSE |
| RTCA | 0.0757285 | 0.86866141 | 1 | 0.075728506 | 0.075728506 | 1.503269665 | 0.10108637 | 0.875673439 | 0.50165 | 0.404258787 | 0.404258787 | 0.404258787 | 0.749146541 | -0.125433221 | FALSE | TRUE | FALSE |
| LGALS2 | 0.0755904 | 0.86867173 | 1 | 0.075590388 | 0.075590388 | 0.075590388 | 0.39276838 | 0.794548564 | 1 | 0.392768381 | 0.392768381 | 0.392768381 | 0.192455379 | -0.715669945 | FALSE | TRUE | FALSE |
| PIK3CD | 0.0754967 | 0.86868205 | 1 | 0.301877292 | 1.207373674 | 10.1878693 | 0.41411343 | 0.786886587 | 0.518 | 0.41411343 | 0.41411343 | 8.081743686 | 0.182309137 | -0.739191566 | FALSE | TRUE | FALSE |
| KRT34 | 0.0754948 | 0.86869237 | 0.69285 | 0.30183131 | 4.828183791 | 5.97520381 | 0.03147882 | 0.890860213 | 0.01115 | 0.031478817 | 0.031478817 | 0.623933163 | 2.398274688 | 0.379898924 | FALSE | TRUE | FALSE |
| CASKIN1 | 0.0754708 | 0.86870269 | 1 | 0.697601566 | 0.697601566 | 13.53867216 | 0.10550318 | 0.874561511 | 1 | 0.105503184 | 0.105503184 | 2.054862838 | 0.715340995 | -0.145486886 | FALSE | TRUE | FALSE |
| EFTUD2 | 0.075418 | 0.868713 | 1 | 0.075417977 | 0.075417977 | 1.477493987 | 0.10134905 | 0.875605094 | 1 | 0.405172493 | 0.405172493 | 7.957191868 | 0.744140913 | -0.128344817 | FALSE | TRUE | FALSE |
| ZKSCAN5 | 0.0753782 | 0.86872332 | 1 | 0.075378191 | 0.075378191 | 1.482699381 | 0.89328114 | 0.599732308 | 0.5109 | 0.893281136 | 0.893281136 | 3.335174819 | 0.084383503 | -1.07374245 | FALSE | TRUE | FALSE |
| CAPSL | 0.075356 | 0.86873363 | 1 | 0.075356032 | 0.075356032 | 0.075356032 | 0.1021381 | 0.875458409 | 0.0053 | 0.102138101 | 0.102138101 | 0.102138101 | 0.737785717 | -0.132069757 | FALSE | TRUE | FALSE |
| TRAF5 | 0.0753218 | 0.86874394 | 0.51105 | 0.075321823 | 0.075321823 | 0.37280326 | 0.25199021 | 0.838861112 | 0.5138 | 0.251990206 | 1.007801212 | 2.279382318 | 0.298907741 | -0.524462837 | FALSE | TRUE | FALSE |
| NET1 | 0.0752865 | 0.86875425 | 1 | 0.07528649 | 0.07528649 | 1.488150303 | 0.09874193 | 0.876295072 | 1 | 0.901584375 | 0.901584375 | 1.954523584 | 0.762457134 | -0.117784567 | FALSE | TRUE | FALSE |
| ZNF473 | 0.0752602 | 0.86876455 | 0.0256 | 0.075260207 | 0.075260207 | 1.478209084 | 0.10170791 | 0.875546457 | 1 | 0.101707915 | 0.101707915 | 2.00199048 | 0.739964109 | -0.130789344 | FALSE | TRUE | FALSE |
| RAB19 | 0.0751848 | 0.86877486 | 1 | 0.300603883 | 1.202174592 | 1.202174592 | 0.93267741 | 0.594592925 | 0.0051 | 0.932677409 | 0.932677409 | 0.932677409 | 0.080611821 | -1.093601269 | FALSE | TRUE | FALSE |
| FGR | 0.075084 | 0.86878516 | 0.5094 | 0.300208345 | 0.300208345 | 2.542773742 | 0.10298837 | 0.875203387 | 1 | 0.102988374 | 0.102988374 | 2.036562509 | 0.729052816 | -0.137241008 | FALSE | TRUE | FALSE |
| SMG1 | 0.0750772 | 0.86879546 | 0.0917 | 0.075077153 | 0.620337239 | 2.480371823 | 0.27443374 | 0.832419681 | 0.54435 | 0.274433745 | 0.973824723 | 9.097580572 | 0.273571143 | -0.562929716 | FALSE | TRUE | FALSE |
| DCXR | 0.0750482 | 0.86880577 | 1 | 3048553410 | 4.94369E+12 | 7.53378E+14 | 0.12829173 | 0.869389108 | 1 | 1066131.454 | 124866797 | 15544061084 | 0.584980946 | -0.23285828 | FALSE | TRUE | FALSE |
| CCDC65 | 0.0750454 | 0.86881607 | 0.25795 | 0.07504535 | 0.681879399 | 1.487962523 | 0.0122907 | 0.895986313 | 0.011 | 0.012290697 | 0.012290697 | 0.243946712 | 6.105865922 | 0.785747263 | FALSE | TRUE | FALSE |
| LRRC37B | 0.0749796 | 0.86882636 | 1 | 0.074979617 | 0.074979617 | 1.473451507 | 0.08818836 | 0.878557214 | 1 | 0.088188365 | 0.088188365 | 1.736658549 | 0.850221199 | -0.070468071 | FALSE | TRUE | FALSE |
| VENTX | 0.0749394 | 0.86883666 | 1 | 0.679654281 | 0.679654281 | 0.679654281 | 0.8405765 | 0.610048597 | 0.93505 | 7.844755617 | 736.7968224 | 736.7968224 | 0.089152374 | -1.049867089 | FALSE | TRUE | FALSE |
| FCN2 | 0.0749383 | 0.86884695 | 1 | 0.684862691 | 0.684862691 | 1.487577403 | 76.7578154 | 0.037527155 | 0.0675 | 7.964558147 | 758.2655181 | 758.2655181 | 0.000976295 | -3.010418945 | FALSE | FALSE | FALSE |
| FHL5 | 0.074844 | 0.86885725 | 1 | 0.074843961 | 0.074843961 | 0.074843961 | 0.40108791 | 0.791843553 | 0.5012 | 0.401087915 | 0.401087915 | 0.401087915 | 0.186602384 | -0.729082812 | FALSE | TRUE | FALSE |
| IAPP | 0.0747849 | 0.86886754 | 1 | 0.70157852 | 0.70157852 | 0.70157852 | 0.74388936 | 0.644314809 | 0.50295 | 0.743889359 | 0.743889359 | 0.743889359 | 0.100532267 | -0.997694524 | FALSE | TRUE | FALSE |
| TMEM184A | 0.074783 | 0.86887783 | 0.0149 | 0.074782995 | 0.074782995 | 1.477847392 | 0.10123919 | 0.875644157 | 1 | 0.10123919 | 0.10123919 | 2.00353649 | 0.738676344 | -0.131545809 | FALSE | TRUE | FALSE |
| ZNF276 | 0.0747798 | 0.86888812 | 1 | 0.679082734 | 0.679082734 | 2.53110968 | 0.92328554 | 0.595565953 | 0.50855 | 0.923285539 | 0.923285539 | 3.425597257 | 0.08099312 | -1.09155187 | FALSE | TRUE | FALSE |
| SPATA16 | 0.0747416 | 0.86889841 | 0.0152 | 0.074741601 | 0.074741601 | 1.480075973 | 0.10069391 | 0.875751463 | 1 | 0.100693911 | 0.100693911 | 1.996366143 | 0.742265351 | -0.129440812 | FALSE | TRUE | FALSE |
| HSD11B2 | 0.074716 | 0.86890869 | 1 | 0.298668636 | 0.298668636 | 1.481394684 | 0.10271618 | 0.875272146 | 1 | 0.102716183 | 0.102716183 | 2.038672532 | 0.727402658 | -0.138225116 | FALSE | TRUE | FALSE |
| IMPDH2 | 0.0746846 | 0.86891898 | 0.50775 | 0.074684594 | 0.074684594 | 0.369487562 | 4.11744499 | 0.348974945 | 0.1359 | 0.455812337 | 4.11744499 | 4.11744499 | 0.018138577 | -1.741396781 | FALSE | FALSE | FALSE |
| INHA | 0.0746696 | 0.86892926 | 1 | 0.074669584 | 0.074669584 | 1.481945304 | 0.10146834 | 0.875575781 | 0.00945 | 0.101468341 | 0.101468341 | 0.101468341 | 0.735890455 | -0.13318683 | FALSE | TRUE | FALSE |
| ABCB9 | 0.0745993 | 0.86893954 | 0.82335 | 0.695308878 | 26.55341469 | 106.0996488 | 0.0204993 | 0.89371119 | 0.89305 | 0.438825175 | 16.84363549 | 67.30036734 | 3.639113965 | 0.560995656 | FALSE | TRUE | FALSE |
| ARHGEF15 | 0.0745924 | 0.86894982 | 0.75635 | 0.1660287 | 0.663870059 | 3.248259273 | 0.0308395 | 0.891060204 | 0.0246 | 0.030839504 | 0.030839504 | 0.604940574 | 2.418727671 | 0.383586973 | FALSE | TRUE | FALSE |
| RNF183 | 0.0745151 | 0.8689601 | 1 | 6.220303899 | 574.7269353 | 574.7269353 | 0.94198567 | 0.593833765 | 0.87665 | 8.77837187 | 83.90656446 | 83.90656446 | 0.079104288 | -1.101799975 | FALSE | TRUE | FALSE |
| SUSD1 | 0.0744944 | 0.86897038 | 0.89255 | 6.041612453 | 541.0481463 | 5306.199978 | 0.14622375 | 0.865451515 | 0.7564 | 0.14622375 | 1.318110198 | 2.88758785 | 0.509454762 | -0.292894374 | FALSE | TRUE | FALSE |
| ZNF33B | 0.0744842 | 0.86898065 | 0.51265 | 0.074484231 | 0.074484231 | 0.296330259 | 0.01495472 | 0.895118585 | 0.49965 | 0.014954723 | 0.128656306 | 0.505919263 | 4.980649446 | 0.697285976 | FALSE | TRUE | FALSE |
| CORT | 0.0744768 | 0.86899093 | 0.0043 | 0.074476828 | 0.074476828 | 0.074476828 | 0.07737938 | 0.880952702 | 1 | 0.077379382 | 0.077379382 | 0.077379382 | 0.962489314 | -0.016604083 | FALSE | TRUE | FALSE |
| DYNLRB2 | 0.0744251 | 0.8690012 | 1 | 0.074425057 | 0.074425057 | 0.074425057 | 0.07732935 | 0.880970722 | 1 | 0.077329348 | 0.077329348 | 0.077329348 | 0.962442573 | -0.016625174 | FALSE | TRUE | FALSE |
| ASCC1 | 0.0742955 | 0.86901147 | 0.49485 | 0.000593493 | 0.074295481 | 0.074295481 | 0.10343354 | 0.875085349 | 0.00855 | 0.103433536 | 0.103433536 | 0.103433536 | 0.718291993 | -0.143698975 | FALSE | TRUE | FALSE |
| NAE1 | 0.0742798 | 0.86902174 | 1 | 0.666765074 | 0.666765074 | 1.471959033 | 0.90596138 | 0.597605834 | 0.50395 | 0.90596138 | 0.90596138 | 1.989438211 | 0.081989975 | -1.086239248 | FALSE | TRUE | FALSE |
| PCDHGA5 | 0.0741322 | 0.86903201 | 0.94215 | 6.207442461 | 572.5981609 | 5694.705707 | 0.02128394 | 0.893468377 | 0.51595 | 0.190299897 | 1.745223381 | 3.724050194 | 3.483011352 | 0.54195489 | FALSE | TRUE | FALSE |
| NMD3 | 0.0741177 | 0.86904227 | 0.0134 | 0.074117665 | 0.074117665 | 1.46843546 | 0.10037623 | 0.875848865 | 1 | 0.100376234 | 0.100376234 | 1.990921834 | 0.738398544 | -0.131709169 | FALSE | TRUE | FALSE |
| NFE2L1 | 0.0741052 | 0.86905254 | 1 | 0.074105189 | 0.074105189 | 1.457205403 | 0.10087837 | 0.875722214 | 0.0213 | 0.100878371 | 0.100878371 | 1.987685458 | 0.734599378 | -0.133949444 | FALSE | TRUE | FALSE |
| NUCB2 | 0.0741052 | 0.8690628 | 0.0093 | 0.074105184 | 0.074105184 | 0.074105184 | 3.43889153 | 0.386901849 | 0.1317 | 1.520141795 | 13.75676518 | 13.75676518 | 0.021549148 | -1.666569887 | FALSE | FALSE | FALSE |
| DHRS11 | 0.0740735 | 0.86907306 | 1 | 0.074073482 | 0.074073482 | 0.074073482 | 0.10052832 | 0.875790441 | 1 | 0.10052832 | 0.10052832 | 0.10052832 | 0.736841938 | -0.132625664 | FALSE | TRUE | FALSE |
| CTNNAL1 | 0.0740641 | 0.86908332 | 0.016 | 0.074064119 | 0.074064119 | 1.46470932 | 0.09990495 | 0.875936407 | 1 | 0.099904946 | 0.099904946 | 1.978402963 | 0.741345864 | -0.129979131 | FALSE | TRUE | FALSE |
| RCCD1 | 0.0740235 | 0.86909358 | 1 | 5304584.097 | 9839749672 | 2.88588E+11 | 3.94243393 | 0.358344212 | 0.9999 | 51705929.34 | 2.55832E+11 | 1.36179E+13 | 0.018776085 | -1.726394951 | FALSE | FALSE | FALSE |
| AMBN | 0.0739646 | 0.86910384 | 1 | 0.65615586 | 5.974469944 | 5.974469944 | 0.10257918 | 0.875331024 | 1 | 0.917835177 | 0.917835177 | 2.037306221 | 0.721048441 | -0.142035558 | FALSE | TRUE | FALSE |
| SLC19A2 | 0.0739592 | 0.8691141 | 0.5031 | 0.073959167 | 0.073959167 | 0.367101494 | 0.1181048 | 0.871693296 | 0.01045 | 0.118104799 | 0.118104799 | 0.586735677 | 0.626216444 | -0.203275532 | FALSE | TRUE | FALSE |
| STXBP5L | 0.0739523 | 0.86912435 | 1 | 0.073952333 | 0.073952333 | 2.489139146 | 0.10053903 | 0.875780699 | 1 | 0.10053903 | 0.10053903 | 1.972260373 | 0.735558449 | -0.133382812 | FALSE | TRUE | FALSE |
| ALG3 | 0.0739445 | 0.8691346 | 0.74895 | 0.073944524 | 0.295614306 | 0.366927807 | 0.11952633 | 0.871330141 | 0.50285 | 0.119526334 | 0.119526334 | 0.593651676 | 0.618646297 | -0.208557582 | FALSE | TRUE | FALSE |
| IL22 | 0.0738692 | 0.86914485 | 1 | 0.0738692 | 0.0738692 | 0.0738692 | 0.0790564 | 0.8806094 | 1 | 0.079056396 | 0.079056396 | 0.079056396 | 0.934386129 | -0.029473617 | FALSE | TRUE | FALSE |
| SLC2A12 | 0.0738673 | 0.8691551 | 0.0158 | 0.073867264 | 0.073867264 | 1.460585037 | 0.39596127 | 0.793469996 | 0.50835 | 0.395961267 | 0.395961267 | 3.353709223 | 0.186551741 | -0.729200694 | FALSE | TRUE | FALSE |
| GPATCH2 | 0.0738245 | 0.86916535 | 0.014 | 0.073824455 | 0.073824455 | 1.461401331 | 0.0997816 | 0.875975278 | 1 | 0.0997816 | 0.0997816 | 1.9776673 | 0.739860403 | -0.130850215 | FALSE | TRUE | FALSE |
| BRAT1 | 0.073823 | 0.8691756 | 0.87835 | 0.659494602 | 6.04311675 | 12.92300898 | 0.01555214 | 0.894982246 | 1 | 0.136924937 | 0.136924937 | 2.68962668 | 4.746806385 | 0.676401518 | FALSE | TRUE | FALSE |
| GRIK5 | 0.0737925 | 0.86918584 | 0.0291 | 0.07379246 | 0.07379246 | 1.440733672 | 0.10045662 | 0.87581966 | 1 | 0.100456622 | 0.100456622 | 1.966951095 | 0.734570391 | -0.133966581 | FALSE | TRUE | FALSE |
| OXCT1 | 0.073728 | 0.86919609 | 1 | 0.073727992 | 0.073727992 | 1.459184022 | 0.39644536 | 0.793205154 | 0.50845 | 0.396445357 | 0.396445357 | 1.964870739 | 0.185972645 | -0.730550931 | FALSE | TRUE | FALSE |
| MKNK2 | 0.0737013 | 0.86920633 | 0.51165 | 0.66398298 | 0.66398298 | 1.459941834 | 0.87584302 | 0.60183847 | 0.75215 | 0.875843022 | 8.140137009 | 8.140137009 | 0.084149019 | -1.074950942 | FALSE | TRUE | FALSE |
| CNPPD1 | 0.0737011 | 0.86921657 | 1 | 0.655049544 | 0.655049544 | 1.460962026 | 0.10232062 | 0.87541924 | 1 | 0.102320619 | 0.102320619 | 2.030443048 | 0.720295405 | -0.142489356 | FALSE | TRUE | FALSE |
| LSS | 0.0736571 | 0.86922681 | 0.7566 | 0.16367438 | 0.654438859 | 3.218370559 | 0.03048124 | 0.891144612 | 0.0185 | 0.030481245 | 0.030481245 | 0.600575314 | 2.4164733 | 0.383182001 | FALSE | TRUE | FALSE |
| SLAMF9 | 0.0736554 | 0.86923705 | 1 | 0.073655387 | 0.073655387 | 0.073655387 | 0.08868068 | 0.878444969 | 1 | 0.088680684 | 0.088680684 | 0.088680684 | 0.83056855 | -0.080624518 | FALSE | TRUE | FALSE |
| WFDC13 | 0.073652 | 0.86924728 | 0.50765 | 0.692292287 | 0.692292287 | 0.692292287 | 0.0798963 | 0.88037345 | 1 | 0.079896298 | 0.079896298 | 0.079896298 | 0.921844598 | -0.035342285 | FALSE | TRUE | FALSE |
| MARCKSL1 | 0.0735326 | 0.86925752 | 1 | 0.073532579 | 0.073532579 | 0.073532579 | 0.07666987 | 0.881132671 | 1 | 0.07666987 | 0.07666987 | 0.07666987 | 0.959080526 | -0.018144927 | FALSE | TRUE | FALSE |
| PPP2R2B | 0.0735247 | 0.86926775 | 1 | 0.66176074 | 0.66176074 | 1.455419404 | 0.10249223 | 0.875350638 | 0.0123 | 0.102492229 | 0.102492229 | 2.031333566 | 0.717368197 | -0.144257881 | FALSE | TRUE | FALSE |
| HNRNPUL1 | 0.0734867 | 0.86927798 | 1 | 0.073486676 | 0.073486676 | 1.43885794 | 0.09400953 | 0.877359575 | 1 | 0.88141594 | 8.481566599 | 29.74675448 | 0.781693881 | -0.106963288 | FALSE | TRUE | FALSE |
| ZNF132 | 0.0734378 | 0.86928821 | 1 | 6.120451211 | 564.5869746 | 564.5869746 | 0.10242592 | 0.873370246 | 1 | 8.654158629 | 82.6597258 | 82.6597258 | 0.716984937 | -0.144489968 | FALSE | TRUE | FALSE |
| ABCG5 | 0.0734173 | 0.86929844 | 0.98035 | 4.69997037 | 163.9479068 | 656.341933 | 0.1438727 | 0.865891168 | 0.65625 | 0.575447674 | 9.21268442 | 36.8755974 | 0.510293443 | -0.292180012 | FALSE | TRUE | FALSE |

| | | | | | | | | | | | | | | | | | |
|---|---|---|---|---|---|---|---|---|---|---|---|---|---|---|---|---|---|
| GINS4 | 0.0733434 | 0.86930867 | 1 | 0.655951526 | 0.655951526 | 0.655951526 | 0.09794947 | 0.876497769 | 1 | 0.391752641 | 3.527600947 | 3.527600947 | 0.748788469 | -0.125640852 | FALSE | TRUE | FALSE |
| KLF11 | 0.073289 | 0.86931889 | 0.01425 | 0.073288996 | 0.073288996 | 1.44996639 | 0.0994405 | 0.876072352 | 1 | 0.099440497 | 0.099440497 | 1.969907045 | 0.73701358 | -0.13252451 | FALSE | TRUE | FALSE |
| KIF27 | 0.0732742 | 0.86932912 | 0.99945 | 441756.9732 | 934842105.1 | 59547501675 | 1.37145271 | 0.548873343 | 1 | 3215509044 | 5.60878E+13 | 5.43002E+15 | 0.0534282 | -1.272229455 | FALSE | FALSE | FALSE |
| HAAO | 0.0732586 | 0.86933934 | 0.50125 | 0.651717681 | 0.651717681 | 0.651717681 | 0.88556684 | 0.600366401 | 0.49865 | 0.098811893 | 0.885566837 | 0.885566837 | 0.082725054 | -1.082362942 | FALSE | TRUE | FALSE |
| ESPNL | 0.0731562 | 0.86934956 | 0.63605 | 0.132341304 | 1.169450143 | 2.594879673 | 1.13911051 | 0.573668648 | 0.3914 | 0.498499535 | 18.21190165 | 18.21190165 | 0.064222187 | -1.192314911 | FALSE | FALSE | FALSE |
| PLEKHG4B | 0.073149 | 0.86935978 | 0.69695 | 0.073149 | 5.401424926 | 12.07652921 | 0.02018255 | 0.893777237 | 0.75255 | 0.020182547 | 0.170255769 | 0.680546456 | 3.624369177 | 0.559232428 | FALSE | TRUE | FALSE |
| SUPT20H | 0.0731098 | 0.86937 | 1 | 0.07310983 | 0.07310983 | 1.443128358 | 0.09875936 | 0.876285404 | 0.01625 | 0.098759364 | 0.098759364 | 1.952488352 | 0.740282509 | -0.130602512 | FALSE | TRUE | FALSE |
| CCDC59 | 0.0730817 | 0.86938021 | 1 | 0.073081674 | 0.073081674 | 0.073081674 | 0.09852688 | 0.876333729 | 1 | 0.098526879 | 0.098526879 | 0.098526879 | 0.741743521 | -0.129746239 | FALSE | TRUE | FALSE |
| SETMAR | 0.0729851 | 0.86939043 | 0.9405 | 5.764477644 | 503.9754291 | 2015.339535 | 75.9945358 | 0.037730007 | 0.11645 | 4.970046433 | 180.7634473 | 897.8066556 | 0.0009604 | -3.017548045 | FALSE | FALSE | FALSE |
| MSR1 | 0.0729782 | 0.86940064 | 0.68885 | 0.072978211 | 1.166763552 | 1.166763552 | 0.03313892 | 0.890574486 | 0.99095 | 2.118359967 | 33.90911792 | 135.7180188 | 2.202190483 | 0.342854881 | FALSE | TRUE | FALSE |
| MTG2 | 0.0729251 | 0.86941085 | 0.01705 | 0.072925119 | 0.072925119 | 1.441105369 | 0.09778013 | 0.876526676 | 1 | 0.391045378 | 0.391045378 | 1.935049198 | 0.745807165 | -0.127373449 | FALSE | TRUE | FALSE |
| NDST3 | 0.0728894 | 0.86942107 | 1 | 0.0728894 | 0.0728894 | 1.43626195 | 0.09919558 | 0.876178964 | 0.01865 | 0.099195584 | 0.099195584 | 1.958090583 | 0.734804885 | -0.133827965 | FALSE | TRUE | FALSE |
| PRR16 | 0.0728477 | 0.86943127 | 1 | 0.072847739 | 0.072847739 | 0.072847739 | 8.05091797 | 0.226568829 | 0.251 | 2.012241688 | 8.050917966 | 8.050917966 | 0.009048377 | -2.043429327 | FALSE | FALSE | FALSE |
| CRTAC1 | 0.0728307 | 0.86944148 | 1 | 0.07283065 | 0.07283065 | 1.432031057 | 1.95112005 | 0.493620825 | 0.004 | 0.099029432 | 0.099029432 | 1.951120053 | 0.037327611 | -1.427969804 | FALSE | FALSE | FALSE |
| ABCE1 | 0.0728141 | 0.86945169 | 1 | 0.291130499 | 1.164309549 | 5.759244465 | 0.10136979 | 0.875595325 | 1 | 0.101369794 | 0.101369794 | 2.007957796 | 0.718301788 | -0.143693052 | FALSE | TRUE | FALSE |
| ABHD3 | 0.0727925 | 0.86946189 | 1 | 0.072792537 | 0.072792537 | 1.444029947 | 0.38540499 | 0.797487125 | 0.75445 | 0.385404991 | 1.541780933 | 1.545498357 | 0.188872843 | -0.723830481 | FALSE | TRUE | FALSE |
| KRT35 | 0.0727865 | 0.8694721 | 0.7527 | 0.290821251 | 4.646250658 | 4.646250658 | 0.01477732 | 0.895175897 | 0.50245 | 0.059051582 | 0.059051582 | 0.292720651 | 4.925552323 | 0.692454937 | FALSE | TRUE | FALSE |
| TMOD1 | 0.0727761 | 0.8694823 | 0.50705 | 0.290926536 | 0.290926536 | 1.444131812 | 0.10032758 | 0.875868328 | 1 | 0.100327576 | 0.100327576 | 0.100327576 | 0.725384823 | -0.139431536 | FALSE | TRUE | FALSE |
| CD248 | 0.0727398 | 0.8694925 | 0.02485 | 0.072739787 | 0.072739787 | 1.427939282 | 0.09904118 | 0.87620801 | 1 | 0.099041181 | 0.099041181 | 1.948578189 | 0.734439819 | -0.134043785 | FALSE | TRUE | FALSE |
| DNASE1L2 | 0.0725692 | 0.8695027 | 0.87645 | 1.410233904 | 51.66231093 | 51.66231093 | 0.37243282 | 0.801687898 | 0.998 | 10149.44032 | 3884927.977 | 39341345.48 | 0.19485175 | -0.71029569 | FALSE | TRUE | FALSE |
| ALX1 | 0.0724938 | 0.86951289 | 1 | 6.579959055 | 660.9352614 | 660.9352614 | 0.11219559 | 0.872873219 | 1 | 0.112195588 | 0.112195588 | 2.22924395 | 0.646137981 | -0.18967473 | FALSE | TRUE | FALSE |
| CSF3R | 0.0724647 | 0.86952309 | 0.5112 | 0.65189313 | 0.65189313 | 12.81126587 | 0.10118076 | 0.875653919 | 1 | 0.101180763 | 0.101180763 | 1.992605799 | 0.716190928 | -0.144971184 | FALSE | TRUE | FALSE |
| RXRG | 0.0724331 | 0.86953329 | 0.9689 | 6.489400537 | 103.7319452 | 414.8865038 | 0.08432623 | 0.879273048 | 0.9998 | 4727.024567 | 761672.4732 | 7852544.491 | 0.85896235 | -0.066025872 | FALSE | TRUE | FALSE |
| PCYT2 | 0.0724262 | 0.86954348 | 1 | 0.671978802 | 0.671978802 | 0.671978802 | 1.5256995 | 0.533590241 | 0.69505 | 3.549291815 | 56.77696041 | 56.77696041 | 0.047470833 | -1.323573149 | FALSE | FALSE | FALSE |
| DHRS1 | 0.0724122 | 0.86955367 | 0.74945 | 0.16394777 | 0.6553313 | 0.6553313 | 0.11934384 | 0.871371743 | 0.00785 | 0.119343843 | 0.119343843 | 0.119343843 | 0.606752919 | -0.216988125 | FALSE | TRUE | FALSE |
| SERINC2 | 0.072376 | 0.86956386 | 1 | 2.59879201 | 23.94683623 | 23.94683623 | 0.09911267 | 0.876198329 | 0.7477 | 0.895518699 | 8.304620208 | 8.304620208 | 0.730239199 | -0.136534858 | FALSE | TRUE | FALSE |
| CDK11A | 0.0723752 | 0.86957405 | 0.51275 | 0.658910011 | 0.658910011 | 12.91592835 | 0.09833218 | 0.876372364 | 1 | 0.899562644 | 0.899562644 | 17.67545584 | 0.736027522 | -0.133105946 | FALSE | TRUE | FALSE |
| TPM3 | 0.0723234 | 0.86958424 | 0.0071 | 0.072323386 | 0.072323386 | 0.072323386 | 0.09846969 | 0.876343389 | 1 | 0.098469689 | 0.098469689 | 0.098469689 | 0.734473587 | -0.134023818 | FALSE | TRUE | FALSE |
| HOOK1 | 0.0722634 | 0.86959442 | 1 | 0.072263363 | 0.072263363 | 1.429137794 | 0.0948209 | 0.877178557 | 1 | 0.850662159 | 0.850662159 | 1.877771325 | 0.762103771 | -0.117985889 | FALSE | TRUE | FALSE |
| NR3C1 | 0.0722436 | 0.86960461 | 1 | 2.662503552 | 25.18710283 | 52.55948137 | 0.10585525 | 0.87440255 | 1 | 0.105855248 | 0.105855248 | 2.092909674 | 0.682475788 | -0.165912751 | FALSE | TRUE | FALSE |
| LRIT2 | 0.0722318 | 0.86961479 | 1 | 24.00171543 | 551.885856 | 2207.22032 | 749.552551 | 0.003068399 | 0.1889 | 17.22472721 | 749.552551 | 7494.389254 | 9.63666E-05 | -4.016073498 | FALSE | FALSE | FALSE |
| VCP | 0.0722046 | 0.86962497 | 1 | 0.288632194 | 0.288632194 | 5.669076456 | 0.09326414 | 0.877492649 | 0.9845 | 5.962854465 | 95.46037655 | 382.0940741 | 0.774195182 | -0.111149536 | FALSE | TRUE | FALSE |
| PRMT8 | 0.0721574 | 0.86963515 | 1 | 54.78376712 | 525.5865836 | 5165.699342 | 0.09463191 | 0.877216707 | 1 | 73.28054833 | 7000.989908 | 70913.56117 | 0.76250601 | -0.117756729 | FALSE | TRUE | FALSE |
| MCMDC2 | 0.0721483 | 0.86964533 | 1 | 5.845270439 | 54.67273545 | 54.67273545 | 7.76749824 | 0.231867611 | 0.68905 | 7.767498239 | 705.8750504 | 705.8750504 | 0.009288491 | -2.032054856 | FALSE | FALSE | FALSE |
| ACSS1 | 0.0721065 | 0.8696555 | 1 | 0.072106547 | 0.072106547 | 1.418287761 | 0.0981782 | 0.876410977 | 0.0194 | 0.098178199 | 0.098178199 | 1.934943341 | 0.734445606 | -0.134040363 | FALSE | TRUE | FALSE |
| LAMB4 | 0.0720863 | 0.86966568 | 0.74365 | 0.559458522 | 71.26067214 | 580.9157848 | 0.00130197 | 0.899295976 | 0.33095 | 0.000668361 | 0.020806744 | 0.051876495 | 55.3672431 | 1.743252899 | FALSE | TRUE | FALSE |
| ITPRIPL1 | 0.0720506 | 0.86967585 | 1 | 0.641111603 | 0.641111603 | 2.438776737 | 0.74906343 | 0.641807874 | 0.98585 | 64.47005188 | 6126.280344 | 61887.38896 | 0.096187643 | -1.016880719 | FALSE | TRUE | FALSE |
| COQ4 | 0.0719629 | 0.86968602 | 1 | 0.071962925 | 0.071962925 | 1.427205328 | 0.80427008 | 0.619843287 | 0.8757 | 7.471637447 | 119.5319376 | 478.2784989 | 0.08947607 | -1.048293101 | FALSE | TRUE | FALSE |
| CYP11A1 | 0.071875 | 0.86969619 | 0.505 | 0.65530672 | 0.65530672 | 1.424872369 | 0.09964942 | 0.876023833 | 1 | 0.099649415 | 0.398486097 | 1.977567341 | 0.721279029 | -0.141896695 | FALSE | TRUE | FALSE |
| WDR86 | 0.0718606 | 0.86970636 | 1 | 0.647425467 | 0.647425467 | 0.647425467 | 7.46731191 | 0.235507613 | 0.3201 | 7.467311909 | 693.1778396 | 693.1778396 | 0.00962335 | -2.016673728 | FALSE | FALSE | FALSE |
| SLC9B2 | 0.0718283 | 0.86971653 | 1 | 0.071828289 | 0.071828289 | 1.42373952 | 0.09717485 | 0.876661405 | 0.0105 | 0.097174855 | 0.097174855 | 1.928213142 | 0.739165383 | -0.131258381 | FALSE | TRUE | FALSE |
| CNTN5 | 0.0718272 | 0.8697267 | 0.98655 | 6.132903251 | 235.4542593 | 941.6839708 | 30.8998688 | 0.089908848 | 0.08135 | 7.721731378 | 123.6821523 | 495.1831502 | 0.002324516 | -2.633667482 | FALSE | FALSE | FALSE |
| SERPINB1 | 0.071813 | 0.86973686 | 1 | 0.07181299 | 0.07181299 | 1.425239957 | 0.38671293 | 0.796926849 | 0.50825 | 0.386712929 | 0.386712929 | 0.386712929 | 0.185701033 | -0.731185681 | FALSE | TRUE | FALSE |
| MFAP3L | 0.0718022 | 0.86974702 | 1 | 0.071802227 | 0.071802227 | 1.422836904 | 0.09808514 | 0.876468852 | 1 | 0.098085142 | 0.098085142 | 0.098085142 | 0.73203979 | -0.135465313 | FALSE | TRUE | FALSE |
| QDPR | 0.0717962 | 0.86975719 | 1 | 0.287014756 | 0.287014756 | 0.287014756 | 0.09688086 | 0.876747872 | 1 | 0.387319665 | 1.548851161 | 1.548851161 | 0.741077045 | -0.130136639 | FALSE | TRUE | FALSE |
| WDFY2 | 0.0717898 | 0.86976734 | 0.0131 | 0.07178975 | 0.07178975 | 1.422602086 | 0.38535426 | 0.797512515 | 0.5046 | 0.385354255 | 0.385354255 | 1.911524316 | 0.186295466 | -0.729797716 | FALSE | TRUE | FALSE |
| GPR135 | 0.0717497 | 0.8697775 | 1 | 0.071749709 | 0.071749709 | 1.41788412 | 0.09454192 | 0.877235774 | 1 | 0.859673069 | 0.859673069 | 1.870973443 | 0.758919486 | -0.119804296 | FALSE | TRUE | FALSE |
| SUCLA2 | 0.0717459 | 0.86978766 | 0.0108 | 0.071745908 | 0.071745908 | 1.422806201 | 0.09775812 | 0.876536308 | 1 | 0.097758119 | 0.097758119 | 1.940627146 | 0.733912524 | -0.134355701 | FALSE | TRUE | FALSE |
| COG1 | 0.0717336 | 0.86979781 | 1 | 0.64455308 | 0.64455308 | 12.6189498 | 0.39652207 | 0.793152099 | 0.5139 | 0.396522073 | 0.396522073 | 7.782778979 | 0.180906929 | -0.742544798 | FALSE | TRUE | FALSE |
| GBP6 | 0.0717324 | 0.86980797 | 0.7538 | 0.286838576 | 2.529451811 | 2.529451811 | 0.01541935 | 0.895032513 | 1 | 0.015419348 | 0.015419348 | 0.305262206 | 4.652105959 | 0.667649598 | FALSE | TRUE | FALSE |
| MOCS2 | 0.0717115 | 0.86981812 | 1 | 0.636376612 | 0.636376612 | 0.636376612 | 0.8664146 | 0.604027162 | 0.5002 | 0.866414597 | 0.866414597 | 0.866414597 | 0.082768138 | -1.082136814 | FALSE | TRUE | FALSE |
| CYP27C1 | 0.0716999 | 0.86982827 | 1 | 0.071699929 | 0.071699929 | 1.421298798 | 0.09754817 | 0.87658445 | 1 | 0.097548173 | 0.097548173 | 0.097548173 | 0.735020724 | -0.133700415 | FALSE | TRUE | FALSE |
| NCF2 | 0.0716983 | 0.86983842 | 0.01515 | 0.071698276 | 0.071698276 | 1.419746752 | 0.09744306 | 0.876603697 | 1 | 0.097443056 | 0.097443056 | 1.931838243 | 0.735796665 | -0.133242185 | FALSE | TRUE | FALSE |
| CRYBA4 | 0.0716755 | 0.86984857 | 1 | 0.071675522 | 0.071675522 | 0.071675522 | 0.38509299 | 0.797614011 | 0.50255 | 0.385092993 | 0.385092993 | 0.385092993 | 0.18612523 | -0.730194751 | FALSE | TRUE | FALSE |
| XPC | 0.0716547 | 0.86985871 | 0.756 | 0.071654709 | 0.638676877 | 2.41828436 | 0.01456335 | 0.895268903 | 1 | 0.127850516 | 0.511238221 | 2.511890651 | 4.920208362 | 0.691983495 | FALSE | TRUE | FALSE |
| TAPBPL | 0.0715875 | 0.86986886 | 1 | 0.636237339 | 0.636237339 | 1.418179068 | 0.09973131 | 0.876004415 | 1 | 0.099731311 | 0.099731311 | 1.977972972 | 0.717803481 | -0.14399444 | FALSE | TRUE | FALSE |
| TRIM41 | 0.0715755 | 0.869879 | 1 | 0.07157547 | 0.07157547 | 1.40682162 | 0.09719065 | 0.87665179 | 1 | 0.097190647 | 0.097190647 | 1.914256345 | 0.736444011 | -0.132860265 | FALSE | TRUE | FALSE |
| MDH2 | 0.0715176 | 0.86988914 | 1 | 1.143179969 | 10.67555236 | 10.67555236 | 0.10191218 | 0.875507338 | 1 | 0.101912182 | 0.40743211 | 0.40743211 | 0.701757402 | -0.153812998 | FALSE | TRUE | FALSE |
| KLHL12 | 0.071455 | 0.86989928 | 1 | 0.071454954 | 0.071454954 | 1.413233517 | 0.09743008 | 0.876613318 | 0.01415 | 0.097430075 | 0.097430075 | 1.929542199 | 0.733397297 | -0.134660695 | FALSE | TRUE | FALSE |
| TAF15 | 0.0714295 | 0.86990942 | 1 | 0.071429489 | 0.071429489 | 1.410450962 | 0.09740349 | 0.876622938 | 1 | 0.097403488 | 0.097403488 | 1.926273593 | 0.733336045 | -0.134696968 | FALSE | TRUE | FALSE |
| OXER1 | 0.0713137 | 0.86991956 | 1 | 0.63862251 | 0.63862251 | 1.41264606 | 0.0996204 | 0.876033539 | 1 | 0.099620405 | 0.099620405 | 1.975639376 | 0.715854098 | -0.145175485 | FALSE | TRUE | FALSE |
| RSPH3 | 0.0712618 | 0.86992969 | 1 | 0.071261803 | 0.071261803 | 1.405370504 | 0.09367825 | 0.877397623 | 1 | 0.840738589 | 0.840738589 | 3.170220735 | 0.760708134 | -0.11878194 | FALSE | TRUE | FALSE |
| ARHGDIA | 0.0712306 | 0.86993983 | 1 | 0.071230631 | 0.071230631 | 0.071230631 | 0.09632349 | 0.876862991 | 0.00675 | 0.096323493 | 0.096323493 | 0.096323493 | 0.739493856 | -0.13106543 | FALSE | TRUE | FALSE |
| SLC22A18 | 0.0712305 | 0.86994996 | 0.0117 | 0.071230537 | 0.071230537 | 1.412267773 | 0.09700547 | 0.876719062 | 0.0108 | 0.097005474 | 0.097005474 | 1.925304318 | 0.734293994 | -0.134130024 | FALSE | TRUE | FALSE |
| WFIKKN1 | 0.0712087 | 0.86996009 | 1 | 0.648498292 | 0.648498292 | 12.75002444 | 0.09692685 | 0.876728667 | 1 | 0.886957055 | 0.886957055 | 3.284368184 | 0.734664239 | -0.1339111 | FALSE | TRUE | FALSE |
| REEP4 | 0.0711859 | 0.86997022 | 1 | 59.8260818 | 593.6972517 | 6035.577814 | 0.09412335 | 0.877331027 | 0.9832 | 79.76953474 | 8092.470072 | 84464.05308 | 0.756304728 | -0.121303184 | FALSE | TRUE | FALSE |
| TNP1 | 0.0711775 | 0.86998035 | 0.00165 | 0.07117751 | 0.07117751 | 0.07117751 | 0.074142 | 0.881589405 | 1 | 0.074142001 | 0.074142001 | 0.074142001 | 0.960016036 | -0.017721513 | FALSE | TRUE | FALSE |
| HOXD4 | 0.0711466 | 0.86999047 | 1 | 0.284446767 | 0.284446767 | 1.411307056 | 0.07339898 | 0.881696422 | 0.99985 | 9835.603199 | 4173132.04 | 44500213.21 | 0.96931333 | -0.013535815 | FALSE | TRUE | FALSE |
| HOMER2 | 0.0711278 | 0.8700006 | 0.50515 | 0.284367672 | 0.284367672 | 1.410991765 | 0.38398106 | 0.797892593 | 0.7531 | 0.38398106 | 1.535617961 | 1.535617961 | 0.185237906 | -0.732270137 | FALSE | TRUE | FALSE |
| LAMP5 | 0.0711042 | 0.87001072 | 1 | 0.071104167 | 0.071104167 | 0.071104167 | 0.08086605 | 0.880081836 | 1 | 0.080866053 | 0.080866053 | 0.080866053 | 0.879283265 | -0.055871193 | FALSE | TRUE | FALSE |
| ZNF408 | 0.0710909 | 0.87002085 | 1 | 0.639188013 | 0.639188013 | 12.55540072 | 0.87325833 | 0.602152673 | 0.5068 | 0.873258325 | 0.873258325 | 17.1900747 | 0.081408746 | -1.089328937 | FALSE | TRUE | FALSE |
| CEP97 | 0.0710905 | 0.87003097 | 1 | 0.28434545 | 0.28434545 | 5.600455215 | 0.0975774 | 0.876565197 | 1 | 0.097577402 | 0.097577402 | 1.925396804 | 0.728555319 | -0.137537467 | FALSE | TRUE | FALSE |
| OR10A7 | 0.0710377 | 0.87004108 | 1 | 1.03915E+25 | 8.51884E+30 | 3.61403E+34 | 0.0781046 | 0.880844485 | 1 | 8.92285E+28 | 1.45794E+36 | 7.14021E+39 | 0.909520069 | -0.041187713 | FALSE | TRUE | FALSE |

| | | | | | | | | | | | | | | | | | |
|---|---|---|---|---|---|---|---|---|---|---|---|---|---|---|---|---|---|
| ACIN1 | 0.071005 | 0.8700512 | 1 | 0.651679902 | 0.651679902 | 12.63420058 | 7.7480611 | 0.23252818 | 0.0299 | 0.820043141 | 7.748061099 | 150.7677165 | 0.009164223 | -2.037904348 | FALSE | FALSE | FALSE |
| EDEM2 | 0.0709763 | 0.87006132 | 0.87755 | 0.283841813 | 1.135398161 | 1.401535424 | 0.10796194 | 0.873802983 | 0.9363 | 0.967376259 | 35.58048397 | 35.58048397 | 0.657419245 | -0.182157587 | FALSE | TRUE | FALSE |
| MLLT10 | 0.0709295 | 0.87007143 | 0.0252 | 0.070929523 | 0.070929523 | 1.39284694 | 0.85764641 | 0.606396484 | 0.5077 | 0.857646406 | 0.857646406 | 16.87844952 | 0.082702525 | -1.082481232 | FALSE | TRUE | FALSE |
| FRMD1 | 0.0707965 | 0.87008155 | 1 | 0.647554937 | 0.647554937 | 2.397588662 | 0.88667215 | 0.600260842 | 0.504 | 0.886672147 | 0.886672147 | 1.905466113 | 0.079845134 | -1.097751544 | FALSE | TRUE | FALSE |
| FOXJ1 | 0.070779 | 0.87009166 | 0.01335 | 0.070778988 | 0.070778988 | 1.401710101 | 0.09536312 | 0.877063981 | 1 | 0.381193626 | 0.381193626 | 1.890801081 | 0.742205035 | -0.129476104 | FALSE | TRUE | FALSE |
| SERPINF2 | 0.0707742 | 0.87010177 | 0.93635 | 6.203391613 | 601.764973 | 601.764973 | 0.1094393 | 0.873531448 | 1 | 0.109439304 | 0.109439304 | 2.166245871 | 0.646698214 | -0.189298338 | FALSE | TRUE | FALSE |
| GALNTL6 | 0.0706919 | 0.87011188 | 0.0169 | 0.070691891 | 0.070691891 | 1.396423706 | 0.38118377 | 0.798874121 | 0.51105 | 0.381183773 | 0.381183773 | 1.885748971 | 0.18545357 | -0.731764803 | FALSE | TRUE | FALSE |
| UNC5CL | 0.0706451 | 0.87012199 | 1 | 0.632912502 | 5.819837752 | 12.49936178 | 0.86482638 | 0.604544658 | 0.75985 | 0.864826382 | 8.005068348 | 8.096069814 | 0.081686988 | -1.087847117 | FALSE | TRUE | FALSE |
| AADACL2 | 0.0706439 | 0.87013209 | 0.49845 | 0.070643935 | 0.632200811 | 1.403150401 | 0.09877751 | 0.876256392 | 1 | 0.098777506 | 0.098777506 | 0.098777506 | 0.715182409 | -0.145583176 | FALSE | TRUE | FALSE |
| SLC25A16 | 0.0706133 | 0.8701422 | 0.49845 | 0.070613317 | 0.647886877 | 0.647886877 | 0.09934982 | 0.876130526 | 0.00735 | 0.099349823 | 0.099349823 | 0.099349823 | 0.71075433 | -0.148280486 | FALSE | TRUE | FALSE |
| GNB3 | 0.0706097 | 0.8701523 | 1 | 0.070609736 | 0.070609736 | 1.399852904 | 0.09609822 | 0.876882158 | 1 | 0.096098219 | 0.096098219 | 0.096098219 | 0.734766333 | -0.133850751 | FALSE | TRUE | FALSE |
| PANX2 | 0.0706078 | 0.8701624 | 0.9916 | 581.2853295 | 61111.2492 | 115884.5853 | 0.09418744 | 0.877302466 | 1 | 8.233770054 | 796.6077526 | 796.6077526 | 0.749652037 | -0.125140274 | FALSE | TRUE | FALSE |
| ZNF160 | 0.0705969 | 0.8701725 | 1 | 0.070596922 | 0.070596922 | 1.390867528 | 0.0959319 | 0.876920477 | 1 | 0.095931896 | 0.095931896 | 1.893400124 | 0.735906671 | -0.13317726 | FALSE | TRUE | FALSE |
| CCT5 | 0.0705945 | 0.8701826 | 1 | 0.07059448 | 0.07059448 | 1.395377906 | 0.38202471 | 0.79842233 | 0.50925 | 0.382024705 | 0.382024705 | 1.891514059 | 0.184790352 | -0.733320708 | FALSE | TRUE | FALSE |
| CRYBB1 | 0.0705753 | 0.87019269 | 1 | 0.07057529 | 0.07057529 | 0.07057529 | 0.09640966 | 0.87682464 | 1 | 0.096409662 | 0.096409662 | 0.096409662 | 0.732035445 | -0.13546789 | FALSE | TRUE | FALSE |
| SCO2 | 0.0705435 | 0.87020279 | 0.50405 | 0.070543544 | 0.070543544 | 0.070543544 | 0.24040943 | 0.841940461 | 0.6874 | 0.960986278 | 35.95882237 | 35.95882237 | 0.293430847 | -0.532494232 | FALSE | TRUE | FALSE |
| SLC27A2 | 0.0704646 | 0.87021288 | 0.0169 | 0.070464614 | 0.070464614 | 0.596641303 | 0.02760296 | 0.891800493 | 1 | 0.246323897 | 0.246323897 | 0.545218441 | 2.552792122 | 0.407015451 | FALSE | TRUE | FALSE |
| GPR108 | 0.0704483 | 0.87022298 | 1 | 0.070448338 | 0.070448338 | 1.38957704 | 0.09549136 | 0.877006617 | 0.01325 | 0.095491356 | 0.095491356 | 1.886664126 | 0.737745705 | -0.13209331 | FALSE | TRUE | FALSE |
| MMP8 | 0.0703508 | 0.87023307 | 0.5014 | 0.070350799 | 0.070350799 | 0.070350799 | 0.12231652 | 0.870787055 | 0.5061 | 0.122316522 | 0.122316522 | 0.282332853 | 0.575153687 | -0.240216092 | FALSE | TRUE | FALSE |
| APOH | 0.0702594 | 0.87024316 | 1 | 0.280956767 | 0.280956767 | 1.395522415 | 0.37131432 | 0.80202793 | 0.7524 | 0.371314325 | 3.384415707 | 3.384415707 | 0.189218178 | -0.723037144 | FALSE | TRUE | FALSE |
| AQP5 | 0.0702143 | 0.87025325 | 1 | 0.070214255 | 0.070214255 | 0.070214255 | 0.37134983 | 0.802003682 | 0.87955 | 1.485000985 | 5.939895286 | 5.939895286 | 0.189078463 | -0.723357937 | FALSE | TRUE | FALSE |
| ANAPC4 | 0.0702111 | 0.87026333 | 1 | 0.637487982 | 5.940706791 | 12.56789225 | 0.09464424 | 0.877197635 | 1 | 0.094644238 | 0.094644238 | 1.869087148 | 0.741842776 | -0.129688128 | FALSE | TRUE | FALSE |
| DCT | 0.0700854 | 0.87027342 | 0.3898 | 0.032167385 | 1.119168432 | 4.473959138 | 83851.9182 | 2.38314E-05 | 0.01315 | 44.16012205 | 8657.462998 | 83851.91823 | 8.35824E-07 | -6.077885282 | FALSE | FALSE | FALSE |
| MPDU1 | 0.0700483 | 0.8702835 | 1 | 0.640955105 | 0.640955105 | 0.640955105 | 0.09858352 | 0.876324066 | 1 | 0.098583523 | 0.098583523 | 0.098583523 | 0.710547619 | -0.148406811 | FALSE | TRUE | FALSE |
| FBXO10 | 0.070043 | 0.87029358 | 0.0305 | 0.070043016 | 0.070043016 | 1.369168234 | 0.250087 | 0.839419592 | 0.5112 | 0.250086997 | 0.250086997 | 4.901907756 | 0.280074603 | -0.552726271 | FALSE | TRUE | FALSE |
| LRRC25 | 0.0700107 | 0.87030367 | 1 | 0.070010694 | 0.070010694 | 1.389205293 | 0.07293904 | 0.881794371 | 0.01085 | 0.072939038 | 0.072939038 | 1.448680119 | 0.959852171 | -0.017795649 | FALSE | TRUE | FALSE |
| ENAH | 0.0699742 | 0.87031375 | 0.01685 | 0.069974194 | 0.069974194 | 1.384119482 | 0.09288416 | 0.877559086 | 1 | 0.869000883 | 0.869000883 | 1.839715101 | 0.753349023 | -0.12300377 | FALSE | TRUE | FALSE |
| ZW10 | 0.0699565 | 0.87032382 | 0.0183 | 0.069956547 | 0.069956547 | 1.379503239 | 0.09228497 | 0.877710698 | 1 | 0.831987042 | 0.831987042 | 1.822879446 | 0.758049166 | -0.120302626 | FALSE | TRUE | FALSE |
| RBBP8 | 0.0699293 | 0.8703339 | 0.75255 | 0.069929256 | 0.279757958 | 1.376716676 | 0.02833024 | 0.891640998 | 0.0206 | 0.028330239 | 0.028330239 | 0.558795031 | 2.468360954 | 0.392408668 | FALSE | TRUE | FALSE |
| AFP | 0.0699208 | 0.87034398 | 1 | 0.279628946 | 0.279628946 | 1.384547146 | 0.8481023 | 0.607926571 | 0.51235 | 0.848102299 | 0.848102299 | 1.852211064 | 0.082443843 | -1.083841774 | FALSE | TRUE | FALSE |
| STRADA | 0.0699074 | 0.87035405 | 0.01115 | 0.069907423 | 0.069907423 | 1.387807782 | 0.09532711 | 0.877073536 | 1 | 0.095327106 | 0.095327106 | 0.095327106 | 0.733342547 | -0.134693117 | FALSE | TRUE | FALSE |
| STOM | 0.0699014 | 0.87036412 | 1 | 0.069901405 | 0.279439667 | 0.279439667 | 0.09676063 | 0.876757473 | 1 | 0.096760633 | 0.096760633 | 0.096760633 | 0.722415747 | -0.141212796 | FALSE | TRUE | FALSE |
| MOS | 0.0698811 | 0.87037419 | 1 | 0.069881083 | 0.069881083 | 1.382156224 | 0.09456316 | 0.877226241 | 1 | 0.378081617 | 0.378081617 | 1.872825166 | 0.738988466 | -0.13136234 | FALSE | TRUE | FALSE |
| FAHD2B | 0.0698661 | 0.87038426 | 0.0088 | 0.069866099 | 0.069866099 | 0.069866099 | 0.09259137 | 0.877625459 | 1 | 0.092591374 | 0.853951249 | 0.853951249 | 0.754563798 | -0.122304035 | FALSE | TRUE | FALSE |
| TMEM91 | 0.0698498 | 0.87039433 | 1 | 0.63418785 | 0.63418785 | 0.63418785 | 0.09816173 | 0.876439921 | 0.0046 | 0.098161732 | 0.098161732 | 0.098161732 | 0.711578863 | -0.147776961 | FALSE | TRUE | FALSE |
| PSRC1 | 0.0697468 | 0.87040439 | 1 | 0.069746815 | 0.069746815 | 1.383694354 | 0.77037546 | 0.63210754 | 0.8776 | 7.042961171 | 66.04301571 | 66.04301571 | 0.090536133 | -1.043178057 | FALSE | TRUE | FALSE |
| DSC2 | 0.0697449 | 0.87041446 | 1 | 1.10645E+21 | 8.75492E+26 | 2.29761E+30 | 0.06201388 | 0.884315821 | 1 | 3.22393E+33 | 7.52533E+40 | 4.37606E+44 | 1.124666211 | 0.051023648 | FALSE | TRUE | FALSE |
| LRRC15 | 0.069687 | 0.87042452 | 1 | 0.069686974 | 0.069686974 | 1.37496226 | 0.08946617 | 0.878248067 | 1 | 0.816300247 | 7.644291129 | 16.13216453 | 0.778919813 | -0.108507249 | FALSE | TRUE | FALSE |
| DOK4 | 0.0696866 | 0.87043458 | 1 | 0.069686599 | 0.069686599 | 1.381305373 | 0.09538453 | 0.877044865 | 0.00965 | 0.095384529 | 0.095384529 | 0.095384529 | 0.730585971 | -0.136328671 | FALSE | TRUE | FALSE |
| RAB37 | 0.0696207 | 0.87044464 | 0.87725 | 2.554221864 | 24.05482546 | 24.05482546 | 0.10231375 | 0.875429035 | 0.00655 | 0.102313747 | 0.102313747 | 0.102313747 | 0.680462807 | -0.167195608 | FALSE | TRUE | FALSE |
| SEC14L2 | 0.0696034 | 0.8704547 | 1 | 0.069603435 | 0.069603435 | 1.381374343 | 0.09488543 | 0.877169016 | 1 | 0.094885426 | 0.094885426 | 0.094885426 | 0.733552432 | -0.134568838 | FALSE | TRUE | FALSE |
| EIF2B2 | 0.0695959 | 0.87046476 | 0.7401 | 0.619410446 | 9.903578911 | 39.615188 | 0.03535975 | 0.890022432 | 0.50505 | 0.035359749 | 0.035359749 | 0.035359749 | 1.968223211 | 0.294074349 | FALSE | TRUE | FALSE |
| HSD17B11 | 0.0695634 | 0.87047482 | 0.0076 | 0.069563378 | 0.069563378 | 0.069563378 | 0.09497991 | 0.877149931 | 1 | 0.094979914 | 0.094979914 | 0.094979914 | 0.732400941 | -0.135251106 | FALSE | TRUE | FALSE |
| OLFM2 | 0.0695274 | 0.87048487 | 0.01795 | 0.069527366 | 0.069527366 | 1.371800167 | 0.09425173 | 0.877283418 | 0.50635 | 0.376802961 | 0.376802961 | 1.862637845 | 0.737677363 | -0.132133544 | FALSE | TRUE | FALSE |
| FZD4 | 0.0695048 | 0.87049492 | 1 | 0.069504846 | 0.069504846 | 1.374231701 | 0.09315248 | 0.877530621 | 1 | 0.372410125 | 1.489214007 | 7.373306278 | 0.746140612 | -0.127179321 | FALSE | TRUE | FALSE |
| TMEM181 | 0.0693881 | 0.87050497 | 0.98355 | 53.21692803 | 5051.842371 | 51008.23181 | 0.10948073 | 0.873511291 | 0.50565 | 0.987852742 | 0.987852742 | 2.165612566 | 0.633792858 | -0.198052659 | FALSE | TRUE | FALSE |
| CWC22 | 0.0693776 | 0.87051502 | 0.0227 | 0.0693776 | 0.0693776 | 1.367361694 | 0.08814368 | 0.878575903 | 1 | 0.088143675 | 0.088143675 | 1.740264266 | 0.787096744 | -0.103971884 | FALSE | TRUE | FALSE |
| AQP8 | 0.0693752 | 0.87052507 | 1 | 0.27732633 | 0.27732633 | 0.27732633 | 0.09508426 | 0.877130841 | 0.50435 | 0.380126248 | 0.380126248 | 0.380126248 | 0.729617793 | -0.136904584 | FALSE | TRUE | FALSE |
| PDK4 | 0.0693398 | 0.87053512 | 0.01085 | 0.069339825 | 0.069339825 | 1.375319521 | 0.83285895 | 0.611749527 | 0.5063 | 0.832858948 | 0.832858948 | 0.832858948 | 0.083255185 | -1.079588712 | FALSE | TRUE | FALSE |
| CNPY1 | 0.0691649 | 0.87054517 | 1 | 0.069164928 | 0.069164928 | 0.069164928 | 0.07662084 | 0.881141656 | 1 | 0.076620841 | 0.076620841 | 0.076620841 | 0.9026908 | -0.044460984 | FALSE | TRUE | FALSE |
| DPP4 | 0.0691564 | 0.87055521 | 0.3231 | 0.069156435 | 1.105139779 | 1.105139779 | 44.6033059 | 0.061498467 | 0.05305 | 1.202004224 | 106.1690303 | 424.4125788 | 0.001550478 | -2.80953445 | FALSE | FALSE | FALSE |
| SERPINI1 | 0.0691198 | 0.87056525 | 0.00935 | 0.069119834 | 0.069119834 | 0.069119834 | 0.85864515 | 0.606089126 | 0.5016 | 0.858645147 | 0.858645147 | 0.858645147 | 0.080498718 | -1.094211036 | FALSE | TRUE | FALSE |
| CPD | 0.0690946 | 0.8705753 | 0.5207 | 0.624802611 | 0.624802611 | 12.19320472 | 0.09425756 | 0.877273892 | 1 | 0.85712046 | 0.85712046 | 16.77580221 | 0.733040178 | -0.134872221 | FALSE | TRUE | FALSE |
| METTL21C | 0.0690207 | 0.87058534 | 0.00625 | 0.069020662 | 0.069020662 | 0.069020662 | 0.80905498 | 0.617828492 | 0.50055 | 0.809054978 | 0.809054978 | 0.809054978 | 0.085310225 | -1.068998912 | FALSE | TRUE | FALSE |
| PNLDC1 | 0.068744 | 0.87059538 | 1 | 0.612386446 | 52.51018062 | 52.51018062 | 0.09072688 | 0.877994062 | 1 | 7.484362909 | 683.3674054 | 683.3674054 | 0.757702922 | -0.120501038 | FALSE | TRUE | FALSE |
| CSRP1 | 0.0687338 | 0.87060541 | 0.00525 | 0.068733839 | 0.068733839 | 0.068733839 | 0.09418346 | 0.877311987 | 0.005 | 0.094183459 | 0.094183459 | 0.094183459 | 0.729786731 | -0.136804037 | FALSE | TRUE | FALSE |
| ATP9A | 0.0687113 | 0.87061545 | 0.7065 | 0.274543389 | 4.386337742 | 9.276758335 | 0.21047912 | 0.849549705 | 0.89785 | 0.84151181 | 13.46133369 | 65.72418695 | 0.32645194 | -0.486180746 | FALSE | TRUE | FALSE |
| CDC42BPB | 0.0686734 | 0.87062549 | 1 | 0.068673385 | 1.320895678 | 2.301813302 | 0.09421509 | 0.877292942 | 1 | 0.094215092 | 0.094215092 | 3.163311991 | 0.72890005 | -0.13733202 | FALSE | TRUE | FALSE |
| IBSP | 0.0685847 | 0.87063552 | 0.51205 | 0.606666045 | 0.606666045 | 1.363119686 | 0.09587693 | 0.876930054 | 1 | 0.095876935 | 0.095876935 | 0.095876935 | 0.715341339 | -0.145486677 | FALSE | TRUE | FALSE |
| NASP | 0.0685773 | 0.87064555 | 1 | 0.068577292 | 0.068577292 | 1.353881725 | 0.09417049 | 0.877321508 | 1 | 0.09417049 | 0.09417049 | 1.862034605 | 0.728224857 | -0.137734501 | FALSE | TRUE | FALSE |
| ANGPT4 | 0.0685079 | 0.87065558 | 0.01595 | 0.068507905 | 0.068507905 | 1.354222976 | 0.09383428 | 0.877369089 | 1 | 0.093834276 | 0.093834276 | 1.857454831 | 0.730094672 | -0.136620821 | FALSE | TRUE | FALSE |
| CLGN | 0.0684899 | 0.87066561 | 1 | 0.613922609 | 2.455415144 | 5.42505717 | 0.09725683 | 0.876642174 | 1 | 0.097256832 | 0.097256832 | 1.928396981 | 0.704217203 | -0.15229337 | FALSE | TRUE | FALSE |
| GTF3C5 | 0.0684678 | 0.87067564 | 1 | 0.06846781 | 0.06846781 | 1.350004052 | 0.09339045 | 0.877464156 | 1 | 0.093390445 | 0.093390445 | 1.844544366 | 0.733135061 | -0.13481601 | FALSE | TRUE | FALSE |
| COX7B2 | 0.0683374 | 0.87068567 | 1 | 0.068337356 | 0.068337356 | 0.068337356 | 0.0702118 | 0.882440669 | 1 | 0.070211803 | 0.070211803 | 0.070211803 | 0.973302961 | -0.011751956 | FALSE | TRUE | FALSE |
| BAAT | 0.068282 | 0.87069569 | 0.50425 | 0.61483011 | 0.61483011 | 1.355295012 | 0.09602756 | 0.876910899 | 1 | 0.096027558 | 0.096027558 | 1.907738307 | 0.711066955 | -0.148089504 | FALSE | TRUE | FALSE |
| TP53BP2 | 0.0682772 | 0.87070572 | 0.7565 | 0.623706721 | 5.847673533 | 12.31988064 | 0.09938308 | 0.876101446 | 0.02385 | 0.099383078 | 0.099383078 | 1.953600577 | 0.687009932 | -0.163036984 | FALSE | TRUE | FALSE |
| DNAJC27 | 0.0682676 | 0.87071574 | 1 | 0.0682676 | 0.0682676 | 0.0682676 | 0.36853561 | 0.802680454 | 0.5022 | 0.368535605 | 0.368535605 | 0.368535605 | 0.185240175 | -0.732264817 | FALSE | TRUE | FALSE |
| CCDC93 | 0.0682481 | 0.87072576 | 1 | 0.068248122 | 0.068248122 | 1.34754362 | 0.09315763 | 0.877521131 | 1 | 0.09315763 | 0.09315763 | 1.842200663 | 0.732609041 | -0.135127726 | FALSE | TRUE | FALSE |
| DPYSL2 | 0.0682478 | 0.87073578 | 0.75705 | 0.272795715 | 1.090674305 | 5.377920616 | 1.40789799 | 0.544637927 | 0.69465 | 3.329198098 | 129.1752686 | 129.1752686 | 0.048474951 | -1.314482619 | FALSE | FALSE | FALSE |
| GJA8 | 0.0682228 | 0.8707458 | 0.5022 | 0.068222791 | 0.27271352 | 0.337636538 | 0.03275849 | 0.89062091 | 0.7493 | 0.032758489 | 0.130934184 | 0.162356873 | 2.082598813 | 0.318605617 | FALSE | TRUE | FALSE |
| DSG3 | 0.0681855 | 0.87075582 | 0.51075 | 0.068185481 | 0.068185481 | 1.33791984 | 0.01178569 | 0.896190628 | 0.9844 | 8.660748963 | 799.0750851 | 7954.202255 | 5.785449055 | 0.762337074 | FALSE | TRUE | FALSE |
| ABCG1 | 0.0681471 | 0.87076583 | 0.75765 | 0.0681471 | 0.27243815 | 1.339369782 | 0.25143018 | 0.838941133 | 0.5057 | 0.25143018 | 0.25143018 | 0.924964887 | 0.271037867 | -0.56697003 | FALSE | TRUE | FALSE |
| CELA3B | 0.0681231 | 0.87077585 | 1 | 0.068123111 | 0.068123111 | 0.068123111 | 0.34719884 | 0.809868426 | 0.87475 | 3.204601705 | 30.35721376 | 30.35721376 | 0.196207773 | -0.707283792 | FALSE | TRUE | FALSE |

| | | | | | | | | | | | | | | | | | |
|---|---|---|---|---|---|---|---|---|---|---|---|---|---|---|---|---|---|
| SLC24A1 | 0.0681013 | 0.87078586 | 0.5143 | 0.068101335 | 0.068101335 | 1.33560578 | 0.01237228 | 0.895972195 | 0.94155 | 1.008543101 | 90.99718005 | 90.99718005 | 5.504349466 | 0.740705999 | FALSE | TRUE | FALSE |
| GRID1 | 0.0679535 | 0.87079587 | 1 | 0.067953496 | 0.067953496 | 1.326802059 | 0.36527515 | 0.804019808 | 0.75315 | 0.365275151 | 1.460756429 | 12.34667908 | 0.186033723 | -0.730408322 | FALSE | TRUE | FALSE |
| OR10J5 | 0.0678687 | 0.87080588 | 1 | 0.629493588 | 5.992449042 | 5.992449042 | 0.07865314 | 0.880718004 | 0.50375 | 0.732181224 | 0.732181224 | 0.732181224 | 0.862886425 | -0.064046363 | FALSE | TRUE | FALSE |
| MCHR1 | 0.0677982 | 0.87081589 | 1 | 0.067798162 | 0.067798162 | 1.340773269 | 0.0928068 | 0.87758754 | 1 | 0.092806802 | 0.092806802 | 1.837816474 | 0.730530098 | -0.136361886 | FALSE | TRUE | FALSE |
| TMPRSS11F | 0.067746 | 0.87082589 | 1 | 0.067745967 | 0.067745967 | 1.344688691 | 0.36763698 | 0.803041079 | 0.50405 | 0.367636983 | 0.367636983 | 0.367636983 | 0.18427408 | -0.734535747 | FALSE | TRUE | FALSE |
| EPOR | 0.0677227 | 0.8708359 | 1 | 0.067722687 | 0.067722687 | 1.339942499 | 0.09252732 | 0.87764441 | 1 | 0.09252732 | 0.09252732 | 1.833079703 | 0.731920978 | -0.135535805 | FALSE | TRUE | FALSE |
| ZNF628 | 0.0677184 | 0.8708459 | 1 | 51289.60849 | 61408962.67 | 681245486.5 | 0.11764548 | 0.871817362 | 0.94 | 9.971572689 | 935.4361208 | 935.4361208 | 0.575614395 | -0.239868353 | FALSE | TRUE | FALSE |
| SBNO2 | 0.0676959 | 0.87085591 | 1 | 888993.6652 | 555010363.5 | 6338087550 | 1.17951981 | 0.56937114 | 0.9984 | 4838.49036 | 735682.6003 | 3458831.142 | 0.057392782 | -1.241142721 | FALSE | FALSE | FALSE |
| MRPL37 | 0.0676581 | 0.87086591 | 0.01455 | 0.067658053 | 0.067658053 | 0.15131278 | 0.01082099 | 0.896534151 | 1 | 0.010820992 | 0.010820992 | 0.214354508 | 6.252481357 | 0.796052405 | FALSE | TRUE | FALSE |
| AFMID | 0.0676418 | 0.87087591 | 0.0092 | 0.067641813 | 0.067641813 | 0.067641813 | 0.8023364 | 0.620224555 | 0.5059 | 0.8023364 | 0.8023364 | 0.8023364 | 0.084306051 | -1.074141255 | FALSE | TRUE | FALSE |
| ZDHHC11 | 0.0676327 | 0.87088591 | 0.75765 | 0.604175202 | 5.539560241 | 11.95728909 | 0.77793368 | 0.629233239 | 0.87645 | 7.176026176 | 67.89553203 | 67.89553203 | 0.086938932 | -1.060785701 | FALSE | TRUE | FALSE |
| SLC16A13 | 0.0676169 | 0.8708959 | 1 | 0.067616883 | 0.067616883 | 1.339426629 | 0.81050244 | 0.617345493 | 0.50135 | 0.810502441 | 0.810502441 | 1.777287561 | 0.083425884 | -1.07869918 | FALSE | TRUE | FALSE |
| ANO8 | 0.0676126 | 0.8709059 | 1 | 0.067612608 | 0.067612608 | 1.314967172 | 0.08728675 | 0.87872523 | 0.7544 | 0.808784376 | 7.691442187 | 27.29168482 | 0.774603313 | -0.11092065 | FALSE | TRUE | FALSE |
| AKR1E2 | 0.0675522 | 0.87091589 | 0.0094 | 0.067552248 | 0.067552248 | 0.067552248 | 0.80521492 | 0.619269928 | 0.50125 | 0.805214923 | 0.805214923 | 0.805214923 | 0.083893437 | -1.076272011 | FALSE | TRUE | FALSE |
| AMTN | 0.0675503 | 0.87092588 | 0.0048 | 0.06755028 | 0.06755028 | 0.06755028 | 0.07520781 | 0.881392758 | 0.0047 | 0.07520781 | 0.07520781 | 0.07520781 | 0.89818172 | -0.046635788 | FALSE | TRUE | FALSE |
| DENND6B | 0.0674906 | 0.87093587 | 1 | 0.067490601 | 0.067490601 | 1.33468758 | 0.09224938 | 0.877729624 | 0.01305 | 0.092249378 | 0.092249378 | 1.826773944 | 0.73161037 | -0.135720147 | FALSE | TRUE | FALSE |
| ACR | 0.0674849 | 0.87094586 | 0.99555 | 232.0108186 | 23766.4138 | 249263.8468 | 0.0997705 | 0.875994704 | 0.9694 | 35.48953824 | 873.9553658 | 3494.276797 | 0.676401374 | -0.169795519 | FALSE | TRUE | FALSE |
| CTSO | 0.0674752 | 0.87095585 | 1 | 0.067475225 | 0.067475225 | 0.067475225 | 0.09129778 | 0.87786197 | 1 | 0.091297779 | 0.365094584 | 0.365094584 | 0.739067543 | -0.13131587 | FALSE | TRUE | FALSE |
| ILVBL | 0.0674703 | 0.87096584 | 0.01915 | 0.067470349 | 0.067470349 | 1.327290286 | 0.08955656 | 0.87821989 | 0.50515 | 0.813895352 | 0.813895352 | 16.04270382 | 0.753382559 | -0.122984438 | FALSE | TRUE | FALSE |
| RGS7BP | 0.0674332 | 0.87097582 | 1 | 0.067433167 | 0.067433167 | 0.067433167 | 0.06982084 | 0.882511104 | 1 | 0.069820837 | 0.069820837 | 0.069820837 | 0.965802912 | -0.015111489 | FALSE | TRUE | FALSE |
| GLRA3 | 0.0673761 | 0.87098581 | 1 | 2.482879551 | 23.48711857 | 23.48711857 | 0.09953782 | 0.876043245 | 1 | 0.09953782 | 0.09953782 | 1.975254664 | 0.676889065 | -0.169482502 | FALSE | TRUE | FALSE |
| ZNRF3 | 0.0673175 | 0.87099579 | 0.75405 | 0.630806078 | 6.066675906 | 21.30706298 | 0.09264444 | 0.877615981 | 1 | 0.869502724 | 8.375453073 | 17.05135879 | 0.726622205 | -0.138691335 | FALSE | TRUE | FALSE |
| SLC22A7 | 0.0673034 | 0.87100577 | 0.0181 | 0.067303411 | 0.067303411 | 1.328293373 | 0.09221515 | 0.877748545 | 0.01595 | 0.092215146 | 0.092215146 | 1.822840839 | 0.729852022 | -0.136765184 | FALSE | TRUE | FALSE |
| GALNT6 | 0.0672869 | 0.87101575 | 0.51015 | 0.067286862 | 0.067286862 | 0.255443468 | 0.01374637 | 0.895539895 | 1 | 0.01374637 | 0.01374637 | 0.271537841 | 4.894882064 | 0.689742232 | FALSE | TRUE | FALSE |
| CHPF2 | 0.0672403 | 0.87102573 | 0.9692 | 5.180254569 | 47.20637612 | 440.7176109 | 0.998637 | 0.588561413 | 0.8925 | 8.98587176 | 783.2256912 | 7578.029106 | 0.06733207 | -1.171778031 | FALSE | TRUE | FALSE |
| MOAP1 | 0.0672112 | 0.8710357 | 1 | 0.067211216 | 0.067211216 | 0.067211216 | 0.0887258 | 0.878416877 | 1 | 0.789854982 | 0.789854982 | 0.789854982 | 0.75751601 | -0.120608184 | FALSE | TRUE | FALSE |
| CRISPLD2 | 0.0671166 | 0.87104568 | 0.9851 | 2.448654545 | 39.16653952 | 156.7010033 | 0.45223347 | 0.773736091 | 0.6864 | 0.452233467 | 7.234910742 | 7.234910742 | 0.14841139 | -0.828532768 | FALSE | TRUE | FALSE |
| LCN10 | 0.0670891 | 0.87105565 | 0.00555 | 0.067089082 | 0.067089082 | 0.067089082 | 0.6498682 | 0.68971458 | 0.2572 | 0.649868203 | 6.20714359 | 6.20714359 | 0.103234905 | -0.986173439 | FALSE | TRUE | FALSE |
| MYOT | 0.0668509 | 0.87106562 | 0.01285 | 0.066850943 | 0.066850943 | 1.324991506 | 0.09147127 | 0.87783363 | 1 | 0.091471267 | 0.091471267 | 1.814930002 | 0.730840901 | -0.136177156 | FALSE | TRUE | FALSE |
| DUS3L | 0.0668362 | 0.87107559 | 0.5103 | 0.267312758 | 0.267312758 | 5.233859609 | 0.08976448 | 0.878182303 | 0.50935 | 0.814136066 | 0.814136066 | 15.98077057 | 0.744573192 | -0.128092605 | FALSE | TRUE | FALSE |
| ZNF692 | 0.0668236 | 0.87108556 | 0.01585 | 0.066823639 | 0.066823639 | 1.321003852 | 0.08841219 | 0.878519819 | 0.50855 | 0.790348802 | 0.790348802 | 1.750211198 | 0.755819303 | -0.121582021 | FALSE | TRUE | FALSE |
| LYG2 | 0.0668179 | 0.87109553 | 1 | 0.066817949 | 0.066817949 | 0.066817949 | 0.0911248 | 0.877899737 | 1 | 0.091124804 | 0.091124804 | 0.091124804 | 0.733257531 | -0.134743468 | FALSE | TRUE | FALSE |
| SH2D3C | 0.0668124 | 0.8711055 | 1 | 0.600461769 | 5.538827187 | 11.74666712 | 0.8428254 | 0.609646245 | 0.5137 | 0.842825395 | 3.371117269 | 16.53138457 | 0.079271927 | -1.100880585 | FALSE | TRUE | FALSE |
| DHX32 | 0.0667952 | 0.87111546 | 1 | 0.066795188 | 0.066795188 | 1.317841717 | 0.36060007 | 0.805457724 | 0.0162 | 0.360600069 | 0.360600069 | 3.050959364 | 0.185233432 | -0.732280628 | FALSE | TRUE | FALSE |
| SLC16A5 | 0.0667116 | 0.87112543 | 1 | 0.266804925 | 0.266804925 | 1.317218003 | 0.36092379 | 0.805364093 | 0.7535 | 0.360923795 | 1.443851894 | 7.137357433 | 0.184835814 | -0.733213875 | FALSE | TRUE | FALSE |
| TRIM52 | 0.066661 | 0.87113539 | 1 | 0.066661027 | 0.066661027 | 0.066661027 | 0.0908855 | 0.877946916 | 1 | 0.0908855 | 0.0908855 | 0.0908855 | 0.733461628 | -0.134622602 | FALSE | TRUE | FALSE |
| LRRC34 | 0.0664955 | 0.87114535 | 1 | 0.591707743 | 5.40418252 | 5.40418252 | 0.83864585 | 0.610450156 | 0.49885 | 0.093473155 | 0.838645854 | 0.838645854 | 0.079289148 | -1.10078625 | FALSE | TRUE | FALSE |
| RDH13 | 0.0664922 | 0.87115531 | 0.50835 | 0.066492171 | 0.57763646 | 0.57763646 | 0.27439515 | 0.832454288 | 0.38165 | 0.125039367 | 1.097145159 | 1.097145159 | 0.242322692 | -0.615605914 | FALSE | TRUE | FALSE |
| PSTK | 0.0663796 | 0.87116527 | 1 | 0.066379569 | 0.066379569 | 1.317364642 | 0.78674586 | 0.625388221 | 0.01035 | 0.786745862 | 0.786745862 | 1.746501522 | 0.084372314 | -1.073800041 | FALSE | TRUE | FALSE |
| ASGR1 | 0.0663583 | 0.87117523 | 1 | 0.066358302 | 0.066358302 | 1.316759976 | 0.78396001 | 0.6264954 | 0.50635 | 0.783960009 | 0.783960009 | 0.783960009 | 0.084645009 | -1.072398642 | FALSE | TRUE | FALSE |
| CAB39L | 0.0663526 | 0.87118518 | 1 | 0.066352567 | 0.066352567 | 0.066352567 | 0.09045687 | 0.878088248 | 1 | 0.090456867 | 0.090456867 | 0.090456867 | 0.733527138 | -0.134583814 | FALSE | TRUE | FALSE |
| NEDD4L | 0.0663397 | 0.87119514 | 1 | 0.608477786 | 0.608477786 | 11.92180895 | 0.09404339 | 0.877350061 | 0.0246 | 0.094043386 | 0.094043386 | 1.847094981 | 0.705415922 | -0.151554742 | FALSE | TRUE | FALSE |
| GCOM1 | 0.0663325 | 0.87120509 | 0.0144 | 0.066332549 | 0.066332549 | 1.313442134 | 0.06977248 | 0.882519903 | 0.014 | 0.069772476 | 0.069772476 | 1.383210317 | 0.95069793 | -0.021957452 | FALSE | TRUE | FALSE |
| DONSON | 0.0662706 | 0.87121504 | 1 | 0.066270588 | 0.066270588 | 1.314953346 | 0.09069994 | 0.878003487 | 1 | 0.090699943 | 0.090699943 | 1.801392917 | 0.730657439 | -0.13628619 | FALSE | TRUE | FALSE |
| NCKAP5L | 0.0662599 | 0.87122499 | 1 | 0.066259852 | 0.066259852 | 2.230643748 | 0.0907945 | 0.877965779 | 0.03415 | 0.0907945 | 0.0907945 | 1.770902488 | 0.729778252 | -0.136809083 | FALSE | TRUE | FALSE |
| SLC25A47 | 0.0662592 | 0.87123494 | 0.012 | 0.066259153 | 0.066259153 | 0.328068554 | 0.41877961 | 0.784949463 | 0.5085 | 0.418779611 | 1.674863643 | 2.075428911 | 0.158219626 | -0.800739647 | FALSE | TRUE | FALSE |
| HOXA5 | 0.0662487 | 0.87124489 | 1 | 0.066248732 | 0.066248732 | 1.31271687 | 0.09137601 | 0.877843078 | 0.00925 | 0.091376013 | 0.091376013 | 0.091376013 | 0.725012294 | -0.139654629 | FALSE | TRUE | FALSE |
| IL16 | 0.0662319 | 0.87125483 | 0.5216 | 0.066231902 | 0.066231902 | 1.289239179 | 0.0138637 | 0.895490074 | 1 | 0.013863704 | 0.013863704 | 0.270733609 | 4.777359717 | 0.679187943 | FALSE | TRUE | FALSE |
| ZNF79 | 0.0661736 | 0.87126478 | 1 | 0.066173639 | 0.066173639 | 1.309657971 | 0.08794447 | 0.878613264 | 0.5047 | 0.795997816 | 0.795997816 | 1.742715054 | 0.752448015 | -0.123523499 | FALSE | TRUE | FALSE |
| TOP2B | 0.0661734 | 0.87127472 | 0.93915 | 5.673226929 | 538.3174184 | 1066.57818 | 1.6080639 | 0.524944566 | 0.0297 | 0.401984676 | 1.608063898 | 13.53125503 | 0.041150971 | -1.385619908 | FALSE | FALSE | FALSE |
| L3HYPDH | 0.066155 | 0.87128466 | 1 | 0.588760435 | 0.588760435 | 1.309642792 | 0.09202638 | 0.877767461 | 1 | 0.368083843 | 0.368083843 | 1.824037007 | 0.718869779 | -0.143349774 | FALSE | TRUE | FALSE |
| FASTK | 0.0661539 | 0.8712946 | 0.01785 | 0.066153852 | 0.066153852 | 1.305975387 | 0.0743539 | 0.881544764 | 1 | 0.074353904 | 0.074353904 | 1.47013964 | 0.889715928 | -0.050748634 | FALSE | TRUE | FALSE |
| CACNA2D2 | 0.0661483 | 0.87130454 | 0.52065 | 0.26449636 | 0.26449636 | 5.158980856 | 0.36430382 | 0.80432804 | 0.51315 | 0.36430382 | 0.36430382 | 7.126913333 | 0.181574592 | -0.740944924 | FALSE | TRUE | FALSE |
| LZTFL1 | 0.0661458 | 0.87131447 | 1 | 0.066145828 | 0.066145828 | 0.066145828 | 0.07132444 | 0.882158151 | 1 | 0.071324439 | 0.071324439 | 0.071324439 | 0.927393595 | -0.032735908 | FALSE | TRUE | FALSE |
| ENTHD1 | 0.0660819 | 0.87132441 | 1 | 0.066081894 | 0.066081894 | 1.308298132 | 0.09050819 | 0.878069422 | 1 | 0.090508189 | 0.090508189 | 1.794068774 | 0.730120605 | -0.136605395 | FALSE | TRUE | FALSE |
| OPN1SW | 0.0660287 | 0.87133434 | 1 | 0.066028706 | 0.264037296 | 1.309726659 | 0.09060701 | 0.878041172 | 0.502 | 0.362334194 | 0.362334194 | 1.799030364 | 0.728737279 | -0.137429013 | FALSE | TRUE | FALSE |
| CSN1S1 | 0.0660025 | 0.87134427 | 1 | 0.066002497 | 0.066002497 | 0.066002497 | 0.07611525 | 0.881213496 | 0.00375 | 0.076115251 | 0.076115251 | 0.076115251 | 0.867138922 | -0.06191132 | FALSE | TRUE | FALSE |
| DGAT2 | 0.0659629 | 0.8713542 | 1 | 51.07919376 | 4880.050472 | 49431.42876 | 72.8329182 | 0.039372143 | 0.31875 | 7.665740095 | 709.3231162 | 709.3231162 | 0.000905675 | -3.043027772 | FALSE | FALSE | FALSE |
| LAP3 | 0.0659169 | 0.87136413 | 1 | 0.065916851 | 0.065916851 | 1.304021375 | 0.0896146 | 0.878201099 | 1 | 0.358344235 | 0.358344235 | 1.775145633 | 0.735559294 | -0.133382313 | FALSE | TRUE | FALSE |
| PCID2 | 0.0658807 | 0.87137406 | 0.51045 | 0.26348878 | 0.26348878 | 1.306811104 | 0.0911468 | 0.877890297 | 1 | 0.091146803 | 0.091146803 | 1.809773347 | 0.722797466 | -0.140983379 | FALSE | TRUE | FALSE |
| STK36 | 0.0658701 | 0.87138399 | 0.50675 | 0.065870086 | 0.577495141 | 1.284204659 | 0.01826211 | 0.894259316 | 0.51885 | 0.018262109 | 0.018262109 | 0.357119569 | 3.606926629 | 0.557137308 | FALSE | TRUE | FALSE |
| VEPH1 | 0.0658361 | 0.87139391 | 0.0185 | 0.065836144 | 0.065836144 | 0.259284991 | 0.01374476 | 0.895547009 | 0.0174 | 0.013744764 | 0.013744764 | 0.271518563 | 4.789907252 | 0.680327104 | FALSE | TRUE | FALSE |
| GPAT2 | 0.065807 | 0.87140384 | 0.5042 | 0.065806952 | 0.065806952 | 1.293280913 | 0.10267259 | 0.875301592 | 0.5115 | 0.228482107 | 0.913639653 | 4.499667669 | 0.640939826 | -0.193182742 | FALSE | TRUE | FALSE |
| HSD11B1L | 0.065757 | 0.87141376 | 0.7527 | 0.583371058 | 5.311981085 | 5.311981085 | 0.75485728 | 0.639433966 | 0.9371 | 6.914464586 | 625.6463044 | 625.6463044 | 0.087111885 | -1.059922587 | FALSE | TRUE | FALSE |
| MLN | 0.0657037 | 0.87142368 | 0.0029 | 0.065703744 | 0.065703744 | 0.065703744 | 0.06824617 | 0.882853393 | 1 | 0.068246169 | 0.068246169 | 0.068246169 | 0.962746259 | -0.01648816 | FALSE | TRUE | FALSE |
| ANAPC5 | 0.0657026 | 0.8714336 | 1 | 0.065702589 | 0.065702589 | 1.292971113 | 0.09007986 | 0.878144695 | 1 | 0.090079861 | 0.090079861 | 1.776118428 | 0.729381555 | -0.137045224 | FALSE | TRUE | FALSE |
| PDE8B | 0.0656969 | 0.87144352 | 0.02405 | 0.065696947 | 0.065696947 | 1.289560788 | 0.35844926 | 0.806203481 | 0.5143 | 0.358449263 | 0.358449263 | 7.051694474 | 0.183281022 | -0.736882502 | FALSE | TRUE | FALSE |
| MINK1 | 0.0656907 | 0.87145343 | 1 | 0.611473771 | 2.444925739 | 20.58953847 | 0.09453342 | 0.877245305 | 1 | 0.094533416 | 0.094533416 | 1.847142975 | 0.69489409 | -0.158081382 | FALSE | TRUE | FALSE |
| TFAP4 | 0.0656311 | 0.87146335 | 1 | 0.065631091 | 0.065631091 | 1.299589129 | 0.34628042 | 0.810158235 | 0.75335 | 0.81905058 | 3.273992463 | 3.273992463 | 0.189531628 | -0.722318308 | FALSE | TRUE | FALSE |
| TMEM209 | 0.0655863 | 0.87147326 | 1 | 0.065586258 | 0.065586258 | 1.29842311 | 0.08960757 | 0.878210495 | 1 | 0.089607565 | 0.089607565 | 1.776141307 | 0.731927689 | -0.135531823 | FALSE | TRUE | FALSE |
| NDUFS2 | 0.065583 | 0.87148317 | 0.01435 | 0.06558304 | 0.06558304 | 1.298387672 | 0.09052001 | 0.878060007 | 1 | 0.090520013 | 0.090520013 | 1.79426166 | 0.724514255 | -0.139953065 | FALSE | TRUE | FALSE |
| ZSCAN20 | 0.0655605 | 0.87149308 | 1 | 5.575880276 | 53.43330949 | 109.2369268 | 0.09876056 | 0.876275735 | 0.0255 | 0.098760561 | 0.098760561 | 1.939581367 | 0.663832624 | -0.177941408 | FALSE | TRUE | FALSE |
| CERS5 | 0.0655485 | 0.87150299 | 1 | 0.262089427 | 0.262089427 | 1.300328657 | 0.35273992 | 0.808133473 | 0.8803 | 1.410806151 | 5.644020937 | 7.004368033 | 0.185826604 | -0.73089211 | FALSE | TRUE | FALSE |

| | | | | | | | | | | | | | | | | | |
|---|---|---|---|---|---|---|---|---|---|---|---|---|---|---|---|---|---|
| GPBAR1 | 0.0655373 | 0.8715129 | 1 | 1.048722559 | 4.203307027 | 4.203307027 | 0.79509644 | 0.622213456 | 0.5102 | 0.795096439 | 3.180226341 | 3.180226341 | 0.082426897 | -1.08393105 | FALSE | TRUE | FALSE |
| GSDMC | 0.0655237 | 0.8715228 | 0.5137 | 0.600033028 | 5.636028604 | 5.636028604 | 0.78114942 | 0.62777841 | 0.69315 | 0.781149424 | 71.52178994 | 71.52178994 | 0.083881125 | -1.076335756 | FALSE | TRUE | FALSE |
| SLC29A3 | 0.0655229 | 0.87153271 | 1 | 0.065522897 | 0.065522897 | 0.065522897 | 0.35690157 | 0.806782023 | 0.00725 | 0.356901574 | 0.356901574 | 0.356901574 | 0.183588144 | -0.736155369 | FALSE | TRUE | FALSE |
| RNPEP | 0.0654819 | 0.87154261 | 0.0166 | 0.065481893 | 0.065481893 | 1.295776826 | 0.75284905 | 0.64028559 | 0.75825 | 0.752849052 | 6.900406899 | 6.900406899 | 0.086978781 | -1.060586681 | FALSE | TRUE | FALSE |
| RMI2 | 0.0654246 | 0.87155251 | 1 | 0.065424602 | 0.065424602 | 0.065424602 | 0.06667888 | 0.88319387 | 1 | 0.066678885 | 0.066678885 | 0.066678885 | 0.981189207 | -0.008247238 | FALSE | TRUE | FALSE |
| ENPEP | 0.0653628 | 0.87156241 | 0.8745 | 0.567346206 | 5.051328847 | 11.13497768 | 1.03646157 | 0.584190515 | 0.26205 | 0.117788343 | 1.036461573 | 2.316963677 | 0.06306337 | -1.200222827 | FALSE | FALSE | FALSE |
| SOX9 | 0.0651879 | 0.87157231 | 0.0209 | 0.065187878 | 0.065187878 | 1.282410439 | 0.08622166 | 0.878957891 | 1 | 0.816806428 | 3.265103834 | 6.793751013 | 0.756049882 | -0.12144955 | FALSE | TRUE | FALSE |
| TTLL9 | 0.0651873 | 0.87158221 | 1 | 0.065187309 | 0.065187309 | 1.289771271 | 0.08910191 | 0.878313765 | 1 | 0.089101907 | 0.089101907 | 1.765209685 | 0.731603976 | -0.135723943 | FALSE | TRUE | FALSE |
| PREB | 0.0651045 | 0.87159211 | 1 | 0.592595135 | 0.592595135 | 1.289343101 | 0.09233377 | 0.877672828 | 0.01075 | 0.092333771 | 0.092333771 | 1.830751212 | 0.705099246 | -0.151749749 | FALSE | TRUE | FALSE |
| GJC1 | 0.0650956 | 0.871602 | 1 | 0.06509556 | 0.06509556 | 1.291131938 | 0.08978987 | 0.878172903 | 0.01065 | 0.08978987 | 0.08978987 | 1.7827023 | 0.724976663 | -0.139675973 | FALSE | TRUE | FALSE |
| SCN4B | 0.0650884 | 0.87161189 | 1 | 2.415456826 | 23.01351984 | 23.01351984 | 0.09369838 | 0.877388113 | 0.49745 | 0.093698377 | 0.872899723 | 0.872899723 | 0.694659273 | -0.158228162 | FALSE | TRUE | FALSE |
| ZKSCAN4 | 0.0650407 | 0.87162179 | 0.0155 | 0.065040661 | 0.065040661 | 1.286477547 | 6.90233979 | 0.246691403 | 0.25585 | 0.750996646 | 6.902339789 | 14.87409928 | 0.009422987 | -2.02581139 | FALSE | FALSE | FALSE |
| LCAT | 0.0649483 | 0.87163168 | 0.7499 | 0.064948285 | 0.259573344 | 0.551079166 | 0.10364994 | 0.874996681 | 0.75535 | 0.242901275 | 0.970669459 | 0.970669459 | 0.626611885 | -0.203001372 | FALSE | TRUE | FALSE |
| TAS2R14 | 0.0649451 | 0.87164157 | 0.75785 | 0.064945144 | 0.60334859 | 0.60334859 | 0.75541312 | 0.639006563 | 0.2529 | 0.080869972 | 0.755413115 | 0.755413115 | 0.085973016 | -1.065637837 | FALSE | TRUE | FALSE |
| LGSN | 0.0648744 | 0.87165146 | 0.93905 | 0.580714337 | 9.287473852 | 9.287473852 | 0.02793901 | 0.891747374 | 1 | 0.027939008 | 0.027939008 | 0.5544652 | 2.322000995 | 0.365862402 | FALSE | TRUE | FALSE |
| CCDC97 | 0.0648201 | 0.87166134 | 1 | 0.064820078 | 0.064820078 | 1.283311466 | 0.08872512 | 0.878426242 | 0.01045 | 0.088725123 | 0.088725123 | 1.758718416 | 0.730571858 | -0.136337061 | FALSE | TRUE | FALSE |
| GPRC6A | 0.0647614 | 0.87167123 | 0.85355 | 1.034506534 | 145.8699445 | 1232.033274 | 0.75551538 | 0.638920952 | 0.212 | 0.086531539 | 3.020437988 | 12.07827871 | 0.085718179 | -1.066927065 | FALSE | TRUE | FALSE |
| ZNF680 | 0.0646371 | 0.87168111 | 1 | 0.064637068 | 0.064637068 | 1.282056187 | 0.75239169 | 0.640455399 | 0.7482 | 0.752391691 | 6.958062017 | 6.958062017 | 0.085908801 | -1.065962342 | FALSE | TRUE | FALSE |
| SAR1B | 0.0646054 | 0.87169099 | 0.0057 | 0.064605406 | 0.064605406 | 0.064605406 | 0.08864103 | 0.87845433 | 0.0046 | 0.088641028 | 0.088641028 | 0.088641028 | 0.728843142 | -0.137365928 | FALSE | TRUE | FALSE |
| PAX5 | 0.0644179 | 0.87170088 | 0.5054 | 0.257498033 | 0.257498033 | 1.275235512 | 0.09011428 | 0.878135291 | 1 | 0.090114276 | 0.090114276 | 1.786115063 | 0.714846373 | -0.145787282 | FALSE | TRUE | FALSE |
| EIF2AK3 | 0.0643789 | 0.87171076 | 1 | 0.583905769 | 5.435540199 | 19.67987416 | 0.08340942 | 0.879485403 | 0.9362 | 7.091512925 | 667.302158 | 667.302158 | 0.771841793 | -0.112471709 | FALSE | TRUE | FALSE |
| SASH1 | 0.0643364 | 0.87172064 | 1 | 0.064336411 | 0.064336411 | 1.249313823 | 7.01912113 | 0.244402682 | 0.15375 | 7.019121133 | 67.91013603 | 136.7783315 | 0.009165878 | -2.037825909 | FALSE | FALSE | FALSE |
| LOXL3 | 0.0643153 | 0.87173051 | 1 | 0.064315332 | 0.064315332 | 1.262617121 | 0.08859338 | 0.878473048 | 1 | 0.08859338 | 0.08859338 | 1.743092549 | 0.725960917 | -0.13908676 | FALSE | TRUE | FALSE |
| ZNF709 | 0.0642821 | 0.87174039 | 1 | 0.064282067 | 0.064282067 | 1.271433417 | 0.08820146 | 0.878547867 | 0.0165 | 0.088201459 | 0.088201459 | 1.746853874 | 0.728809558 | -0.13738594 | FALSE | TRUE | FALSE |
| SOX10 | 0.0642774 | 0.87175027 | 1 | 0.064277358 | 0.064277358 | 1.269159541 | 0.0890074 | 0.878323145 | 1 | 0.089007398 | 0.089007398 | 1.76015113 | 0.722157482 | -0.141368085 | FALSE | TRUE | FALSE |
| CST3 | 0.064183 | 0.87176014 | 1 | 3.75612E+38 | 2.06404E+47 | 1.38929E+51 | 939.529309 | 0.002521623 | 1 | 1.86955E+31 | 3.95011E+38 | 2.19377E+42 | 6.8314E-05 | -4.165490503 | FALSE | FALSE | FALSE |
| TTPAL | 0.0641777 | 0.87177001 | 1 | 0.2565639 | 0.2565639 | 1.274086107 | 0.08698783 | 0.878799771 | 1 | 0.08698783 | 0.809349489 | 0.809349489 | 0.737777609 | -0.13207453 | FALSE | TRUE | FALSE |
| OR2T34 | 0.0641703 | 0.87177988 | 0.87625 | 5.805530864 | 57.3772291 | 57.3772291 | 0.07106046 | 0.882202376 | 0.7491 | 0.668687756 | 6.458081379 | 6.458081379 | 0.903038255 | -0.044293851 | FALSE | TRUE | FALSE |
| ZNF283 | 0.0641609 | 0.87178975 | 1 | 0.064160931 | 0.064160931 | 1.269395695 | 0.08807202 | 0.878603926 | 1 | 0.088072018 | 0.088072018 | 1.744724032 | 0.728505288 | -0.137567292 | FALSE | TRUE | FALSE |
| MAP3K11 | 0.0640766 | 0.87179962 | 0.02965 | 0.064076596 | 0.064076596 | 1.252945535 | 0.10428155 | 0.87489802 | 0.51165 | 0.104281549 | 0.104281549 | 2.044587507 | 0.614457655 | -0.211508041 | FALSE | TRUE | FALSE |
| RBM12B | 0.0640715 | 0.87180949 | 1 | 5.472204569 | 517.2785404 | 1028.571555 | 0.09238247 | 0.877663357 | 0.0264 | 0.092382471 | 0.092382471 | 1.812867875 | 0.693545816 | -0.158924844 | FALSE | TRUE | FALSE |
| KIF2C | 0.0640638 | 0.87181935 | 1 | 210.0121378 | 20839.56948 | 83355.3814 | 0.10647561 | 0.874243198 | 1 | 0.106475612 | 0.106475612 | 2.101075453 | 0.601675851 | -0.220637419 | FALSE | TRUE | FALSE |
| PRAME | 0.0640603 | 0.87182921 | 0.0157 | 0.064060288 | 0.064060288 | 1.268375823 | 0.08767769 | 0.878659938 | 1 | 0.087677689 | 0.087677689 | 1.738084891 | 0.73063386 | -0.136300205 | FALSE | TRUE | FALSE |
| DUOX2 | 0.0640099 | 0.87183908 | 0.7334 | 1.024482033 | 2284.531407 | 46034.22125 | 0.00011395 | 0.900160262 | 0.6154 | 0.000242125 | 0.134789963 | 1.201304188 | 561.7544635 | 2.749546532 | FALSE | TRUE | FALSE |
| MPP2 | 0.0639773 | 0.87184894 | 1 | 0.579261091 | 0.579261091 | 2.165960433 | 0.09081249 | 0.877956348 | 1 | 0.090812491 | 0.090812491 | 1.791242825 | 0.704498518 | -0.152119916 | FALSE | TRUE | FALSE |
| CDH24 | 0.0639651 | 0.8718588 | 0.5207 | 0.063965106 | 0.063965106 | 1.255737353 | 0.10356025 | 0.875026251 | 0.50555 | 0.103560252 | 0.103560252 | 2.037562669 | 0.617660779 | -0.209249975 | FALSE | TRUE | FALSE |
| CUX2 | 0.0639396 | 0.87186865 | 1 | 4.91318E+11 | 1.19093E+16 | 5.19475E+17 | 0.09208554 | 0.877758003 | 1 | 1.88332E+11 | 5.51306E+14 | 6.09604E+16 | 0.694350216 | -0.158421425 | FALSE | TRUE | FALSE |
| PIGB | 0.063902 | 0.87187851 | 1 | 0.063901973 | 0.063901973 | 1.267681403 | 0.08785524 | 0.878631938 | 1 | 0.087855238 | 0.087855238 | 1.74456279 | 0.727355295 | -0.138253395 | FALSE | TRUE | FALSE |
| OR6C75 | 0.0638831 | 0.87188836 | 1 | 5.097375618 | 47.31601933 | 47.31601933 | 0.09544258 | 0.877025744 | 0.00595 | 0.095442579 | 0.095442579 | 0.095442579 | 0.669335373 | -0.174356223 | FALSE | TRUE | FALSE |
| PBRM1 | 0.0638633 | 0.87189822 | 0.9431 | 5.733529423 | 569.6607988 | 1095.946343 | 0.10044794 | 0.875829397 | 0.03935 | 0.100447941 | 0.100447941 | 3.376331456 | 0.635785015 | -0.196689712 | FALSE | TRUE | FALSE |
| HECTD4 | 0.0638594 | 0.87190807 | 0.8868 | 0.603647134 | 11.12375206 | 107.8495374 | 0.10099092 | 0.875702709 | 0.9407 | 91.96849763 | 9758.461799 | 104079.1998 | 0.632328211 | -0.199057442 | FALSE | TRUE | FALSE |
| TNKS2 | 0.0638371 | 0.87191792 | 1 | 0.063837118 | 0.063837118 | 1.253301263 | 0.08809173 | 0.878594586 | 0.0225 | 0.088091733 | 0.088091733 | 1.73331068 | 0.724666387 | -0.139861883 | FALSE | TRUE | FALSE |
| AKAP12 | 0.0638329 | 0.87192777 | 0.04495 | 0.063832872 | 0.063832872 | 2.143966932 | 0.08790895 | 0.878622602 | 1 | 0.087908954 | 0.087908954 | 2.956928285 | 0.726124799 | -0.138988731 | FALSE | TRUE | FALSE |
| KLK14 | 0.063794 | 0.87193762 | 1 | 0.063793988 | 0.063793988 | 0.063793988 | 0.08539329 | 0.879069287 | 1 | 0.783954838 | 0.783954838 | 0.783954838 | 0.747060906 | -0.12664399 | FALSE | TRUE | FALSE |
| PARP9 | 0.0636805 | 0.87194746 | 0.01675 | 0.063680487 | 0.063680487 | 1.258850149 | 0.08473547 | 0.879199024 | 0.50885 | 0.757241553 | 0.757241553 | 1.677405244 | 0.751521036 | -0.124058859 | FALSE | TRUE | FALSE |
| CDHR4 | 0.0636047 | 0.87195731 | 1 | 0.570529121 | 5.252566989 | 5.252566989 | 0.71960166 | 0.656078106 | 0.9702 | 63.04312653 | 612.5783811 | 6097.765152 | 0.088388695 | -1.05360328 | FALSE | TRUE | FALSE |
| TSTD2 | 0.0635688 | 0.87196715 | 1 | 0.254260681 | 2.261763764 | 5.037529105 | 0.08559338 | 0.879050733 | 0.7527 | 0.766380016 | 7.042889899 | 7.042889899 | 0.742683305 | -0.129196338 | FALSE | TRUE | FALSE |
| CLASP1 | 0.0635527 | 0.87197699 | 1 | 0.581334519 | 0.581334519 | 11.26964831 | 0.09048501 | 0.878078835 | 1 | 0.090485008 | 0.090485008 | 3.041710133 | 0.702355955 | -0.153442731 | FALSE | TRUE | FALSE |
| COASY | 0.0634832 | 0.87198683 | 0.9693 | 2.284709215 | 50.01445856 | 199.984759 | 2.2171085 | 0.470455355 | 0.50035 | 2.217108495 | 21.13594287 | 21.13594287 | 0.028633346 | -1.543127895 | FALSE | FALSE | FALSE |
| KRTAP10-6 | 0.0634713 | 0.87199667 | 1 | 100.7791464 | 43972.8536 | 87218.15411 | 0.08950522 | 0.878238676 | 0.98355 | 68.46853019 | 6488.280923 | 65453.89561 | 0.709135412 | -0.149270826 | FALSE | TRUE | FALSE |
| SDHB | 0.0634614 | 0.87200651 | 0.5087 | 0.253660472 | 1.014155965 | 1.014155965 | 0.41351561 | 0.787027596 | 0.50475 | 0.413515609 | 1.653461054 | 1.653461054 | 0.153467872 | -0.81398253 | FALSE | TRUE | FALSE |
| ZNF567 | 0.063458 | 0.87201635 | 0.01355 | 0.063458015 | 0.063458015 | 1.256179085 | 0.08722057 | 0.878734552 | 1 | 0.087220574 | 0.087220574 | 1.728693632 | 0.727557868 | -0.138132458 | FALSE | TRUE | FALSE |
| FAM167B | 0.0634243 | 0.87202618 | 1 | 0.063424274 | 0.063424274 | 0.063424274 | 0.06965303 | 0.882537497 | 0.00435 | 0.069653026 | 0.069653026 | 0.069653026 | 0.910574557 | -0.040684489 | FALSE | TRUE | FALSE |
| FBP1 | 0.0634084 | 0.87203601 | 0.4975 | 0.063408416 | 0.592174537 | 0.598824416 | 0.08784139 | 0.878641272 | 0.5 | 0.454902847 | 0.821964303 | 0.821964303 | 0.721851238 | -0.141552294 | FALSE | TRUE | FALSE |
| BEND4 | 0.0633288 | 0.87204585 | 1 | 0.595890204 | 5.754619971 | 5.754619971 | 0.07261001 | 0.881883287 | 1 | 0.072610007 | 0.072610007 | 1.442663044 | 0.872177017 | -0.059395362 | FALSE | TRUE | FALSE |
| COL4A2 | 0.0633078 | 0.87205568 | 0.1547 | 0.001102031 | 0.252693516 | 2.250289445 | 0.00019254 | 0.900042358 | 0.35565 | 4.82425E-05 | 0.000768653 | 0.003725966 | 328.7994132 | 2.516931034 | FALSE | TRUE | FALSE |
| INTS5 | 0.063299 | 0.87206551 | 0.02805 | 0.063299028 | 0.063299028 | 1.237237388 | 0.00815349 | 0.89725639 | 1 | 0.008153491 | 0.008153491 | 0.159803352 | 7.763426314 | 0.890053435 | FALSE | TRUE | FALSE |
| GTF3C1 | 0.0632394 | 0.87207533 | 1 | 0.580458433 | 1.201136947 | 11.10882969 | 0.09005424 | 0.878154099 | 1 | 0.090054237 | 1.720849497 | 3.017921893 | 0.702236636 | -0.153516517 | FALSE | TRUE | FALSE |
| EXOC1 | 0.0632291 | 0.87208516 | 1 | 0.567434955 | 0.567434955 | 11.17397928 | 0.78591835 | 0.625850373 | 0.51185 | 0.785918354 | 0.785918354 | 15.5050533 | 0.080452497 | -1.094460472 | FALSE | TRUE | FALSE |
| SYCP2 | 0.0631489 | 0.87209499 | 1 | 0.063148915 | 0.063148915 | 2.122398576 | 0.08696127 | 0.878809083 | 1 | 0.08696127 | 0.08696127 | 2.926757496 | 0.72617287 | -0.13895998 | FALSE | TRUE | FALSE |
| CCDC27 | 0.0630518 | 0.87210481 | 0.9959 | 42.72276341 | 3723.816502 | 36001.37112 | 6753.08438 | 0.000255798 | 0.2583 | 705.3727903 | 66162.04418 | 663001.454 | 9.33675E-06 | -5.029804494 | FALSE | FALSE | FALSE |
| SH3YL1 | 0.0629754 | 0.87211463 | 1 | 0.06297538 | 0.558638146 | 0.558638146 | 0.74914024 | 0.64172366 | 0.7516 | 0.749140241 | 6.867459232 | 6.867459232 | 0.084063539 | -1.07539233 | FALSE | TRUE | FALSE |
| ANKS3 | 0.0629649 | 0.87212445 | 1 | 0.062964886 | 0.062964886 | 1.239055676 | 0.08703969 | 0.878753193 | 0.018 | 0.087039688 | 0.087039688 | 1.716126316 | 0.723404317 | -0.140618904 | FALSE | TRUE | FALSE |
| CWF19L2 | 0.0629229 | 0.87213427 | 1 | 5.104012458 | 47.76831094 | 47.76831094 | 0.7506345 | 0.641048431 | 0.9396 | 6.894384922 | 627.2051852 | 627.2051852 | 0.083826289 | -1.076619758 | FALSE | TRUE | FALSE |
| GAN | 0.0629225 | 0.87214409 | 0.0186 | 0.062922539 | 0.062922539 | 1.241132561 | 0.08438295 | 0.87925455 | 1 | 0.774934423 | 0.774934423 | 2.855826896 | 0.745678332 | -0.127448476 | FALSE | TRUE | FALSE |
| METTL4 | 0.0629147 | 0.87215391 | 0.01065 | 0.062914714 | 0.062914714 | 1.248408531 | 0.08657524 | 0.878892828 | 1 | 0.086575241 | 0.086575241 | 0.086575241 | 0.726705615 | -0.138641484 | FALSE | TRUE | FALSE |
| ZFP28 | 0.0628981 | 0.87216372 | 0.0182 | 0.062898088 | 0.062898088 | 1.239510119 | 0.08662137 | 0.878874227 | 1 | 0.086621373 | 0.086621373 | 1.71002992 | 0.726126665 | -0.138987614 | FALSE | TRUE | FALSE |
| SERPINB6 | 0.0628201 | 0.87217354 | 1 | 0.062820127 | 0.062820127 | 0.062820127 | 0.32324887 | 0.817636738 | 0.87445 | 2.975961777 | 28.11969752 | 28.11969752 | 0.194339818 | -0.711438208 | FALSE | TRUE | FALSE |
| BTNL3 | 0.0627956 | 0.87218335 | 1 | 0.558162959 | 0.558162959 | 1.244407868 | 0.08372054 | 0.879439298 | 1 | 0.749096352 | 6.879331853 | 6.879331853 | 0.750062165 | -0.124902741 | FALSE | TRUE | FALSE |
| VARS2 | 0.0627656 | 0.87219316 | 0.5142 | 0.062765614 | 0.062765614 | 1.225805762 | 0.01243924 | 0.895943948 | 0.7597 | 0.107900663 | 0.960654388 | 3.635071629 | 5.045775159 | 0.702927894 | FALSE | TRUE | FALSE |
| SCN1B | 0.0627473 | 0.87220297 | 1 | 0.062747307 | 0.062747307 | 0.062747307 | 0.08686119 | 0.878827702 | 0.0078 | 0.086861188 | 0.086861188 | 0.086861188 | 0.722386008 | -0.141230674 | FALSE | TRUE | FALSE |
| AXIN1 | 0.0627217 | 0.87221278 | 0.0272 | 0.062721651 | 0.062721651 | 1.227351664 | 0.08703406 | 0.878762512 | 0.0263 | 0.087034062 | 0.087034062 | 1.707531388 | 0.720656374 | -0.142271768 | FALSE | TRUE | FALSE |
| KRTAP10-10 | 0.062681 | 0.87222259 | 1 | 4.991770905 | 440.0388136 | 440.0388136 | 0.09662919 | 0.876786267 | 1 | 0.096629192 | 0.096629192 | 0.096629192 | 0.648675746 | -0.18797234 | FALSE | TRUE | FALSE |

| | | | | | | | | | | | | | | | | | |
|---|---|---|---|---|---|---|---|---|---|---|---|---|---|---|---|---|---|
| CMA1 | 0.0626582 | 0.87223239 | 0.0066 | 0.062658151 | 0.062658151 | 0.062658151 | 638.07248 | 0.003303298 | 0.0014 | 6.649616746 | 638.0724799 | 638.0724799 | 9.81991E-05 | -4.007892438 | FALSE | FALSE | FALSE |
| CDHR5 | 0.0626482 | 0.8722422 | 0.5101 | 0.062648204 | 0.062648204 | 1.226408309 | 0.11037788 | 0.873228439 | 0.5036 | 0.110377884 | 0.110377884 | 2.166286098 | 0.567579321 | -0.245973436 | FALSE | TRUE | FALSE |
| EHD4 | 0.0625466 | 0.872252 | 0.75935 | 0.062546581 | 0.250033362 | 1.232586831 | 0.02532534 | 0.892389266 | 0.5077 | 0.233478218 | 0.233478218 | 0.857821593 | 2.469722909 | 0.39264823 | FALSE | TRUE | FALSE |
| TPGS2 | 0.0625083 | 0.8722618 | 1 | 0.062508274 | 0.062508274 | 0.062508274 | 0.07071609 | 0.882264242 | 1 | 0.65165281 | 0.65165281 | 0.65165281 | 0.883932821 | -0.05358074 | FALSE | TRUE | FALSE |
| VKORC1L1 | 0.0624743 | 0.8722716 | 1 | 1.51367E+22 | 1.06071E+28 | 4.11846E+31 | 0.13251702 | 0.868321898 | 1 | 5.0504E+14 | 2.16013E+20 | 3.04743E+21 | 0.471443486 | -0.326570361 | FALSE | TRUE | FALSE |
| VAV2 | 0.0624189 | 0.8722814 | 1 | 0.249563659 | 0.249563659 | 4.883246778 | 0.34620881 | 0.81018049 | 0.5096 | 0.346208806 | 0.346208806 | 6.791976922 | 0.180292757 | -0.74402172 | FALSE | TRUE | FALSE |
| TYW5 | 0.0623884 | 0.8722912 | 1 | 0.062388371 | 0.062388371 | 0.062388371 | 0.3402576 | 0.812031174 | 0.00595 | 0.340257601 | 0.340257601 | 0.340257601 | 0.183356288 | -0.736704193 | FALSE | TRUE | FALSE |
| SERPINA11 | 0.0623598 | 0.87230099 | 0.503 | 0.249325483 | 0.249325483 | 1.237603098 | 0.34149249 | 0.811703371 | 0.25435 | 0.341492487 | 1.365756256 | 1.696585128 | 0.182609679 | -0.738476207 | FALSE | TRUE | FALSE |
| PER1 | 0.0623535 | 0.87231079 | 0.51605 | 0.563695172 | 0.563695172 | 10.93108761 | 26.3922481 | 0.10405688 | 0.0038 | 0.782595388 | 0.782595388 | 15.23161228 | 0.002362567 | -2.626615816 | FALSE | FALSE | FALSE |
| IRF6 | 0.0623298 | 0.87232058 | 1 | 0.062329785 | 0.062329785 | 1.235231861 | 0.08662818 | 0.878864925 | 1 | 0.086628176 | 0.086628176 | 1.718651965 | 0.719509375 | -0.142963543 | FALSE | TRUE | FALSE |
| KLHDC7A | 0.06222 | 0.87233037 | 0.51155 | 0.138875941 | 0.555248224 | 2.714552987 | 0.02553232 | 0.892284021 | 0.5144 | 0.229461267 | 0.229461267 | 4.497437908 | 2.436910335 | 0.38683955 | FALSE | TRUE | FALSE |
| HTRA1 | 0.0622189 | 0.87234016 | 1 | 0.24870873 | 0.24870873 | 1.233378533 | 0.08739455 | 0.878697255 | 1 | 0.087394548 | 0.087394548 | 1.734281373 | 0.7119317 | -0.147561669 | FALSE | TRUE | FALSE |
| COL28A1 | 0.0622009 | 0.87234995 | 0.90665 | 3.974085437 | 537.1792318 | 4654.580168 | 245.880104 | 0.010011519 | 0.5101 | 245.8801038 | 63203.11666 | 485830.9858 | 0.000252973 | -3.59692641 | FALSE | FALSE | FALSE |
| MPRIP | 0.0621974 | 0.87235974 | 1 | 0.062197398 | 0.062197398 | 1.209455007 | 0.08111126 | 0.88005443 | 1 | 0.753647963 | 7.186964426 | 25.44906608 | 0.766815833 | -0.115308928 | FALSE | TRUE | FALSE |
| ECI2 | 0.0621638 | 0.87236952 | 1 | 0.062163824 | 0.062163824 | 0.062163824 | 0.74760255 | 0.642480068 | 0.50005 | 0.747602555 | 0.747602555 | 0.747602555 | 0.083150898 | -1.080133055 | FALSE | TRUE | FALSE |
| DCDC2 | 0.0620801 | 0.87237931 | 1 | 0.062080066 | 0.062080066 | 1.228725395 | 0.08230458 | 0.879752181 | 0.75225 | 0.748841548 | 2.994630744 | 2.994630744 | 0.754272343 | -0.122471817 | FALSE | TRUE | FALSE |
| OR1L1 | 0.0619213 | 0.87238909 | 1 | 5.335975344 | 51.47066348 | 51.47066348 | 0.08005162 | 0.880309775 | 1 | 0.080051621 | 0.080051621 | 0.080051621 | 0.773516952 | -0.111530164 | FALSE | TRUE | FALSE |
| RELN | 0.0619031 | 0.87239888 | 0.9433 | 0.56197393 | 8.995821707 | 74.33082352 | 0.0260209 | 0.892148422 | 0.5398 | 0.237445102 | 0.863191263 | 7.876766821 | 2.37897663 | 0.376390176 | FALSE | TRUE | FALSE |
| PRB1 | 0.0618658 | 0.87240866 | 1 | 0.544422726 | 4.91738093 | 4.91738093 | 6.44284797 | 0.25642783 | 0.6903 | 6.44284797 | 574.6384277 | 574.6384277 | 0.009602248 | -2.01762709 | FALSE | FALSE | FALSE |
| AUH | 0.0618034 | 0.87241844 | 1 | 0.247107362 | 0.247107362 | 0.247107362 | 0.08657905 | 0.878883528 | 0.0062 | 0.086579046 | 0.086579046 | 0.086579046 | 0.713837532 | -0.146400622 | FALSE | TRUE | FALSE |
| MVK | 0.0617267 | 0.87242822 | 0.5089 | 0.246750257 | 0.246750257 | 1.223347386 | 0.08388338 | 0.879374697 | 1 | 1.340950258 | 5.363445393 | 5.363445393 | 0.735862729 | -0.133203193 | FALSE | TRUE | FALSE |
| SYT10 | 0.0617009 | 0.87243799 | 1 | 0.558708697 | 5.192549464 | 11.05942837 | 0.35064475 | 0.8089486 | 0.50645 | 1.402591645 | 5.611818662 | 6.949454735 | 0.175964114 | -0.754575892 | FALSE | TRUE | FALSE |
| OSBPL3 | 0.061681 | 0.87244777 | 0.50795 | 0.552943673 | 0.552943673 | 10.86712858 | 0.08510722 | 0.879124919 | 0.5096 | 0.767963351 | 0.767963351 | 15.1244658 | 0.724744633 | -0.139814992 | FALSE | TRUE | FALSE |
| GNAS | 0.0616622 | 0.87245754 | 0.27255 | 0.061662165 | 0.246510215 | 1.204363232 | 0.10516691 | 0.874680475 | 0.51385 | 0.957747795 | 8.946295637 | 18.75916524 | 0.586326669 | -0.231860352 | FALSE | TRUE | FALSE |
| SPHK2 | 0.0616529 | 0.87246731 | 0.02275 | 0.061652862 | 0.061652862 | 1.209578358 | 0.08183927 | 0.879816415 | 0.50595 | 0.73408822 | 0.73408822 | 14.43522949 | 0.753340852 | -0.123008481 | FALSE | TRUE | FALSE |
| ANKH | 0.061585 | 0.87247709 | 0.0139 | 0.061584976 | 0.061584976 | 0.304482868 | 0.10084857 | 0.875731965 | 0.5121 | 0.100848569 | 0.100848569 | 0.499277432 | 0.610667823 | -0.214194963 | FALSE | TRUE | FALSE |
| SGSM1 | 0.0614948 | 0.87248686 | 1 | 0.55931244 | 0.55931244 | 10.88084436 | 0.08288787 | 0.879586722 | 0.03065 | 0.082887874 | 0.082887874 | 1.617804406 | 0.741903359 | -0.129652663 | FALSE | TRUE | FALSE |
| XYLB | 0.0614544 | 0.87249663 | 0.50785 | 0.061454414 | 0.061454414 | 0.304114107 | 0.0251688 | 0.892404285 | 1 | 0.025168801 | 0.025168801 | 0.498629867 | 2.441690168 | 0.387690554 | FALSE | TRUE | FALSE |
| CNTN4 | 0.0614448 | 0.87250639 | 0.0297 | 0.061444773 | 0.061444773 | 1.204843042 | 0.02462097 | 0.892569249 | 1 | 0.225243926 | 0.225243926 | 4.427117744 | 2.495627718 | 0.397179801 | FALSE | TRUE | FALSE |
| LIN52 | 0.0613865 | 0.87251616 | 1 | 0.061386477 | 0.061386477 | 0.061386477 | 0.06236704 | 0.884213089 | 0.0034 | 0.06236704 | 0.06236704 | 0.06236704 | 0.984277536 | -0.006882427 | FALSE | TRUE | FALSE |
| KIAA0753 | 0.0613817 | 0.87252592 | 0.513 | 0.061381695 | 0.061381695 | 1.206247826 | 0.10549843 | 0.874571433 | 0.26445 | 0.105498433 | 0.933467589 | 2.092399929 | 0.58182566 | -0.235207129 | FALSE | TRUE | FALSE |
| PCDHGC3 | 0.0613584 | 0.87253569 | 1 | 0.061358423 | 0.061358423 | 1.19556599 | 0.08523228 | 0.879097109 | 0.0277 | 0.085232277 | 0.085232277 | 1.665901221 | 0.719896559 | -0.142729902 | FALSE | TRUE | FALSE |
| HSPB7 | 0.061348 | 0.87254545 | 1 | 0.061348032 | 0.061348032 | 0.061348032 | 0.08521835 | 0.87911565 | 1 | 0.085218352 | 0.085218352 | 0.085218352 | 0.719892262 | -0.142732495 | FALSE | TRUE | FALSE |
| RGS11 | 0.0613392 | 0.87255521 | 1 | 0.061339206 | 0.061339206 | 1.214685429 | 0.66761294 | 0.681746973 | 0.9373 | 6.10143336 | 549.5804178 | 549.5804178 | 0.091878397 | -1.036786592 | FALSE | TRUE | FALSE |
| HPS1 | 0.0613188 | 0.87256497 | 0.8734 | 4.975986784 | 46.57989493 | 46.57989493 | 0.75673449 | 0.63857808 | 0.87705 | 6.957757286 | 65.61631817 | 65.61631817 | 0.081030779 | -1.091349985 | FALSE | TRUE | FALSE |
| EXOC2 | 0.0612674 | 0.87257472 | 0.02435 | 0.061267373 | 0.061267373 | 1.203634137 | 0.08466346 | 0.879217537 | 0.02305 | 0.084663459 | 0.084663459 | 1.666812826 | 0.723657806 | -0.140466749 | FALSE | TRUE | FALSE |
| HEPACAM2 | 0.0612156 | 0.87258448 | 1 | 0.061215632 | 0.061215632 | 1.214982925 | 0.72985106 | 0.651775124 | 0.00935 | 0.729851063 | 0.729851063 | 0.729851063 | 0.083874143 | -1.076371904 | FALSE | TRUE | FALSE |
| CYP11B1 | 0.0611635 | 0.87259423 | 1 | 0.061163515 | 0.061163515 | 1.207906894 | 0.75772264 | 0.638320497 | 0.5046 | 0.757722635 | 0.757722635 | 1.628320752 | 0.08072019 | -1.093017827 | FALSE | TRUE | FALSE |
| ZNF350 | 0.061114 | 0.87260399 | 1 | 0.544230014 | 4.974301551 | 10.77912599 | 0.08944112 | 0.878257456 | 0.01265 | 0.089441118 | 0.089441118 | 1.773553083 | 0.683287133 | -0.165396757 | FALSE | TRUE | FALSE |
| LIPA | 0.061113 | 0.87261374 | 1 | 0.562436398 | 0.562436398 | 1.213699989 | 0.08752626 | 0.878678599 | 1 | 0.087526257 | 0.087526257 | 0.087526257 | 0.698225179 | -0.156004494 | FALSE | TRUE | FALSE |
| IPO8 | 0.0610649 | 0.87262349 | 0.02725 | 0.06106488 | 0.06106488 | 1.198369617 | 0.74435335 | 0.644065789 | 0.50765 | 0.744353345 | 2.976741071 | 14.64060742 | 0.08203749 | -1.085987636 | FALSE | TRUE | FALSE |
| PRDM5 | 0.0610119 | 0.87263324 | 0.7587 | 0.554487315 | 2.217313837 | 8.253128869 | 0.3280063 | 0.815959747 | 0.88095 | 2.993356617 | 28.03695731 | 28.03695731 | 0.186008192 | -0.730467928 | FALSE | TRUE | FALSE |
| AMPD3 | 0.0610058 | 0.87264299 | 0.74925 | 0.526881949 | 42.46856929 | 169.7543323 | 0.14529054 | 0.865564504 | 0.999 | 61217.0191 | 97543805.05 | 2376900935 | 0.419888307 | -0.37686622 | FALSE | TRUE | FALSE |
| CHST1 | 0.0608477 | 0.87265273 | 0.9385 | 5.334429478 | 517.5767404 | 517.5767404 | 0.0957667 | 0.876958775 | 1 | 0.095766697 | 0.095766697 | 1.89001443 | 0.635374691 | -0.196970089 | FALSE | TRUE | FALSE |
| MAP3K19 | 0.0608081 | 0.87266248 | 0.94165 | 4.608930736 | 384.8854299 | 3642.275027 | 0.00943498 | 0.896903363 | 0.51805 | 0.009434984 | 0.009434984 | 0.185010967 | 6.444961107 | 0.809220301 | FALSE | TRUE | FALSE |
| KRT25 | 0.0607996 | 0.87267222 | 0.7535 | 0.550099952 | 8.795305358 | 8.795305358 | 0.10462778 | 0.874818994 | 0.5056 | 0.104627781 | 0.104627781 | 0.519239374 | 0.581103574 | -0.235746454 | FALSE | TRUE | FALSE |
| RFC1 | 0.0607428 | 0.87268197 | 1 | 0.060742777 | 0.060742777 | 1.190443675 | 0.32667152 | 0.816522258 | 0.8813 | 1.306556785 | 5.227016056 | 25.66801338 | 0.185944512 | -0.730616635 | FALSE | TRUE | FALSE |
| SH2D7 | 0.0606829 | 0.87269171 | 0.8757 | 2.163940974 | 19.80254044 | 19.80254044 | 3.00822969 | 0.410919262 | 0.51235 | 3.008229691 | 27.72591481 | 27.72591481 | 0.020172295 | -1.695244692 | FALSE | FALSE | FALSE |
| GLP2R | 0.0606714 | 0.87270145 | 1 | 0.539514231 | 0.539514231 | 2.05322486 | 0.08368668 | 0.879448521 | 1 | 0.083686684 | 0.749732556 | 1.65660617 | 0.724982688 | -0.139672364 | FALSE | TRUE | FALSE |
| NFATC2 | 0.0606624 | 0.87271119 | 1 | 2.25454847 | 21.50902595 | 76.08389653 | 0.82170701 | 0.614420609 | 0.51565 | 0.821707008 | 0.821707008 | 16.07271228 | 0.073824832 | -1.131797531 | FALSE | TRUE | FALSE |
| GOSR2 | 0.0606234 | 0.87272092 | 0.51065 | 0.543711507 | 0.543711507 | 0.543711507 | 0.08636242 | 0.878930014 | 1 | 0.086362422 | 0.086362422 | 0.086362422 | 0.701964543 | -0.153684824 | FALSE | TRUE | FALSE |
| FGF22 | 0.0605793 | 0.87273066 | 0.0054 | 0.060579345 | 0.060579345 | 0.060579345 | 2.72431831 | 0.433480068 | 0.78395 | 10.89629881 | 243.9680157 | 975.7847995 | 0.022236515 | -1.65293328 | FALSE | FALSE | FALSE |
| TOR2A | 0.0605637 | 0.87274039 | 1 | 0.060563725 | 0.060563725 | 0.060563725 | 0.08262868 | 0.879641922 | 1 | 0.082628682 | 0.330473907 | 0.330473907 | 0.732962502 | -0.134918243 | FALSE | TRUE | FALSE |
| TSEN15 | 0.0605015 | 0.87275013 | 1 | 0.060501481 | 0.060501481 | 0.060501481 | 0.06329666 | 0.883946904 | 1 | 0.063296663 | 0.063296663 | 0.063296663 | 0.955839975 | -0.01961481 | FALSE | TRUE | FALSE |
| MTIF3 | 0.0604967 | 0.87275986 | 1 | 0.060496691 | 0.060496691 | 0.060496691 | 0.71665992 | 0.657997093 | 0.50605 | 0.716659922 | 0.716659922 | 0.716659922 | 0.084414782 | -1.073581499 | FALSE | TRUE | FALSE |
| SLC1A1 | 0.0604575 | 0.87276959 | 1 | 0.060457474 | 0.060457474 | 1.195591731 | 0.08148536 | 0.879944688 | 1 | 0.749857099 | 0.749857099 | 1.613607681 | 0.741942755 | -0.129629602 | FALSE | TRUE | FALSE |
| ZNF614 | 0.060441 | 0.87277932 | 0.51225 | 0.539813005 | 0.539813005 | 1.195918099 | 0.08604411 | 0.878985755 | 1 | 0.086044108 | 0.086044108 | 1.704700957 | 0.702442038 | -0.153389506 | FALSE | TRUE | FALSE |
| MMRN1 | 0.0603596 | 0.87278904 | 0.7244 | 0.291856216 | 4.665349521 | 18.65969841 | 0.02935644 | 0.891412406 | 0.8786 | 0.257334509 | 2.315273215 | 5.05243625 | 2.056092271 | 0.313042601 | FALSE | TRUE | FALSE |
| PROC | 0.0603264 | 0.87279877 | 0.75135 | 0.060326427 | 0.241103728 | 1.192442505 | 0.02507443 | 0.892434313 | 1 | 0.100201951 | 0.100201951 | 0.496330075 | 2.405893812 | 0.381276455 | FALSE | TRUE | FALSE |
| PGBD4 | 0.0602779 | 0.87280849 | 0.01655 | 0.060277906 | 0.060277906 | 1.190474394 | 0.0831048 | 0.879540686 | 0.0151 | 0.083104799 | 0.083104799 | 1.643771084 | 0.725324015 | -0.139467943 | FALSE | TRUE | FALSE |
| DLX5 | 0.0602689 | 0.87281822 | 1 | 0.060268864 | 0.060268864 | 0.060268864 | 0.08404533 | 0.87931003 | 1 | 0.084045334 | 0.084045334 | 0.084045334 | 0.717099455 | -0.144420608 | FALSE | TRUE | FALSE |
| INSC | 0.0602501 | 0.87282794 | 0.01795 | 0.060250135 | 0.060250135 | 0.230261194 | 0.00965728 | 0.896840835 | 1 | 0.009657281 | 0.009657281 | 0.190747944 | 6.238830334 | 0.795103175 | FALSE | TRUE | FALSE |
| DAG1 | 0.0601598 | 0.87283766 | 0.02585 | 0.060159838 | 0.060159838 | 1.178099298 | 0.0811759 | 0.880045292 | 1 | 1.297594598 | 5.189885731 | 25.47440293 | 0.741104648 | -0.130120463 | FALSE | TRUE | FALSE |
| OAS3 | 0.0601545 | 0.87284738 | 0.27035 | 0.060154495 | 0.524090729 | 2.028526881 | 0.10967346 | 0.873460871 | 0.5117 | 0.109673455 | 0.109673455 | 2.146861707 | 0.54848728 | -0.260833439 | FALSE | TRUE | FALSE |
| PEF1 | 0.0601504 | 0.8728571 | 1 | 0.06015045 | 0.06015045 | 0.06015045 | 0.08375156 | 0.879420848 | 0.00735 | 0.083751558 | 0.083751558 | 0.083751558 | 0.718200959 | -0.143754019 | FALSE | TRUE | FALSE |
| SLC11A2 | 0.060121 | 0.87286681 | 0.0167 | 0.060121016 | 0.060121016 | 1.189281408 | 0.08271755 | 0.879623527 | 1 | 0.330737947 | 0.330737947 | 1.638424723 | 0.726823005 | -0.138571335 | FALSE | TRUE | FALSE |
| C2 | 0.0600565 | 0.87287653 | 0.5162 | 0.060056494 | 0.060056494 | 1.181163976 | 0.02440559 | 0.892651553 | 1 | 0.097587473 | 0.097587473 | 1.923147051 | 2.460767624 | 0.391070604 | FALSE | TRUE | FALSE |
| TOP3B | 0.0599108 | 0.87288624 | 0.0297 | 0.059910842 | 0.059910842 | 1.171283845 | 0.73798886 | 0.647274312 | 0.51215 | 0.73798886 | 0.73798886 | 14.46708246 | 0.081181229 | -1.09054438 | FALSE | TRUE | FALSE |
| KLK5 | 0.0599065 | 0.87289595 | 1 | 0.239506479 | 0.957787534 | 0.957787534 | 0.08526371 | 0.879078562 | 0.0073 | 0.08526371 | 0.08526371 | 0.08526371 | 0.702602495 | -0.153290312 | FALSE | TRUE | FALSE |
| SH2D4A | 0.0598653 | 0.87290567 | 0.50775 | 0.527264837 | 4.763406999 | 4.763406999 | 0.94928675 | 0.593071964 | 0.5025 | 0.949286751 | 8.673158754 | 8.673158754 | 0.063063417 | -1.200222502 | FALSE | TRUE | FALSE |
| CDH20 | 0.0597235 | 0.87291538 | 1 | 0.059723496 | 0.059723496 | 1.171483057 | 0.08324662 | 0.879531475 | 0.0232 | 0.083246616 | 0.083246616 | 1.63667586 | 0.717428515 | -0.144221366 | FALSE | TRUE | FALSE |
| CCT6B | 0.0596935 | 0.87292508 | 0.63775 | 0.238670331 | 2.150236673 | 2.150236673 | 0.02572178 | 0.892208727 | 1 | 0.025721777 | 0.025721777 | 0.510011866 | 2.32073811 | 0.365626134 | FALSE | TRUE | FALSE |
| TRIM23 | 0.0596412 | 0.87293479 | 1 | 0.059641228 | 0.059641228 | 1.179536508 | 0.0828954 | 0.879577518 | 0.01415 | 0.082895403 | 0.082895403 | 1.641636178 | 0.719475692 | -0.142983874 | FALSE | TRUE | FALSE |

| | | | | | | | | | | | | | | | | | |
|---|---|---|---|---|---|---|---|---|---|---|---|---|---|---|---|---|---|
| IGDCC3 | 0.0595056 | 0.8729445 | 1 | 0.059505556 | 0.059505556 | 1.167464448 | 0.08268256 | 0.879632725 | 1 | 0.082682562 | 0.082682562 | 1.625901039 | 0.719686889 | -0.142856409 | FALSE | TRUE | FALSE |
| FTSJ3 | 0.0594761 | 0.8729542 | 1 | 0.059476082 | 0.059476082 | 1.166778288 | 0.08243635 | 0.879715449 | 1 | 0.082436355 | 0.082436355 | 1.620927335 | 0.721478798 | -0.141776427 | FALSE | TRUE | FALSE |
| APOBR | 0.059405 | 0.87296391 | 0.51135 | 0.510683452 | 4.503225098 | 10.00592099 | 7.91028619 | 0.229884811 | 0.3193 | 0.885478831 | 72.43831236 | 72.43831236 | 0.007509838 | -2.12436946 | FALSE | FALSE | FALSE |
| PAQR6 | 0.0594026 | 0.87297361 | 0.01105 | 0.059402633 | 0.059402633 | 1.178720268 | 0.67277237 | 0.678436351 | 0.5021 | 0.672772369 | 0.672772369 | 0.672772369 | 0.088295293 | -1.054062449 | FALSE | TRUE | FALSE |
| ARFGEF1 | 0.0593844 | 0.87298331 | 1 | 0.547217941 | 0.547217941 | 10.60509005 | 0.33755225 | 0.812834891 | 0.52225 | 0.337552255 | 0.337552255 | 6.566221922 | 0.175926409 | -0.754668963 | FALSE | TRUE | FALSE |
| SNX21 | 0.0592706 | 0.87299301 | 0.00665 | 0.059270604 | 0.059270604 | 0.059270604 | 0.08190978 | 0.879798069 | 0.0052 | 0.081909777 | 0.081909777 | 0.081909777 | 0.723608412 | -0.140496393 | FALSE | TRUE | FALSE |
| NR2C2 | 0.0592457 | 0.87300271 | 0.0179 | 0.059245662 | 0.059245662 | 1.16881762 | 0.08283237 | 0.879605127 | 1 | 0.082832371 | 0.082832371 | 1.636815187 | 0.715247683 | -0.14554354 | FALSE | TRUE | FALSE |
| SHC4 | 0.0592324 | 0.87301241 | 0.0169 | 0.059232407 | 0.059232407 | 1.169244316 | 0.08121798 | 0.880008727 | 1 | 0.324829655 | 0.324829655 | 2.748658656 | 0.729301688 | -0.137092781 | FALSE | TRUE | FALSE |
| FAM24B | 0.0592288 | 0.8730221 | 0.0022 | 0.059228846 | 0.059228846 | 0.059228846 | 0.06328926 | 0.883955508 | 1 | 0.063289262 | 0.063289262 | 0.063289262 | 0.935843522 | -0.028796762 | FALSE | TRUE | FALSE |
| DPPA4 | 0.0592003 | 0.8730318 | 0.0085 | 0.059200334 | 0.059200334 | 0.059200334 | 0.07269685 | 0.881874401 | 0.0073 | 0.072696852 | 0.072696852 | 0.072696852 | 0.814345216 | -0.08919145 | FALSE | TRUE | FALSE |
| TTLL12 | 0.0591081 | 0.87304149 | 1 | 0.526601061 | 0.526601061 | 10.35775697 | 6.55324743 | 0.253519751 | 0.01505 | 0.712125235 | 6.553247426 | 14.03474636 | 0.009019664 | -2.04480966 | FALSE | FALSE | FALSE |
| DYRK4 | 0.0591055 | 0.87305118 | 1 | 0.059105465 | 0.059105465 | 1.170002123 | 0.31447842 | 0.820483116 | 0.75135 | 0.708406521 | 2.833210928 | 2.833210928 | 0.187947604 | -0.725963207 | FALSE | TRUE | FALSE |
| MRPS26 | 0.0591011 | 0.87306087 | 0.0076 | 0.05910114 | 0.05910114 | 0.05910114 | 0.06379535 | 0.88383494 | 0.00615 | 0.063795352 | 0.063795352 | 0.063795352 | 0.926417642 | -0.033193183 | FALSE | TRUE | FALSE |
| ARMC3 | 0.0590541 | 0.87307056 | 0.02185 | 0.059054105 | 0.059054105 | 1.162962867 | 0.01251118 | 0.895887411 | 1 | 0.012511184 | 0.012511184 | 0.246839916 | 4.720105143 | 0.673951673 | FALSE | TRUE | FALSE |
| C14orf39 | 0.0590251 | 0.87308025 | 1 | 0.521320153 | 0.521320153 | 1.169836173 | 0.70402685 | 0.664866666 | 0.7561 | 0.704026852 | 6.428930237 | 6.428930237 | 0.083839314 | -1.076552282 | FALSE | TRUE | FALSE |
| JMJD8 | 0.0589834 | 0.87308993 | 0.00735 | 0.058983395 | 0.058983395 | 0.058983395 | 0.71663746 | 0.658073373 | 0.5004 | 0.716637455 | 0.716637455 | 0.716637455 | 0.082305767 | -1.084569733 | FALSE | TRUE | FALSE |
| R3HDM4 | 0.0589592 | 0.87309962 | 1 | 0.058959231 | 0.058959231 | 0.058959231 | 0.06700329 | 0.883115459 | 0.00845 | 0.067003289 | 0.067003289 | 0.067003289 | 0.87994533 | -0.055544309 | FALSE | TRUE | FALSE |
| MAP1A | 0.0589411 | 0.8731093 | 1 | 4.91962E+25 | 6.32545E+32 | 2.76266E+36 | 0.0119114 | 0.896099132 | 1 | 7.29393E+24 | 9.06774E+31 | 2.3665E+34 | 4.948291908 | 0.694455311 | FALSE | TRUE | FALSE |
| ATAD1 | 0.0589384 | 0.87311898 | 1 | 0.058938418 | 0.058938418 | 1.170777813 | 0.0823183 | 0.879743 | 1 | 0.0823183 | 0.0823183 | 0.0823183 | 0.715981966 | -0.145097916 | FALSE | TRUE | FALSE |
| LPCAT4 | 0.0588762 | 0.87312866 | 0.0147 | 0.058876151 | 0.058876151 | 1.165621536 | 0.32549533 | 0.816977979 | 0.5043 | 0.325495325 | 0.325495325 | 1.614000148 | 0.18088171 | -0.742605345 | FALSE | TRUE | FALSE |
| WWP1 | 0.0588567 | 0.87313834 | 1 | 0.941338346 | 3.766028495 | 18.50989009 | 0.08483936 | 0.879161982 | 1 | 0.084839362 | 0.084839362 | 1.671608585 | 0.693742467 | -0.15880172 | FALSE | TRUE | FALSE |
| PRAM1 | 0.0588201 | 0.87314802 | 1 | 44.36213156 | 424.6634734 | 4164.575418 | 0.08897048 | 0.878341902 | 0.75615 | 0.797399858 | 7.335145855 | 15.70658098 | 0.661119789 | -0.179719843 | FALSE | TRUE | FALSE |
| NPR2 | 0.0588173 | 0.87315769 | 0.0331 | 0.05881731 | 0.05881731 | 1.146818596 | 0.0244641 | 0.892644076 | 1 | 0.024464097 | 0.024464097 | 0.47844329 | 2.404229743 | 0.380975966 | FALSE | TRUE | FALSE |
| EIF2D | 0.0588118 | 0.87316737 | 1 | 0.05881178 | 0.05881178 | 1.163524148 | 0.08134974 | 0.879981289 | 1 | 0.081349743 | 0.081349743 | 1.611504476 | 0.722949787 | -0.140891866 | FALSE | TRUE | FALSE |
| PRRC2A | 0.0588084 | 0.87317704 | 0.97115 | 49.32379885 | 1642.095577 | 9282.742645 | 0.08712816 | 0.878743873 | 0.8843 | 7.637729416 | 74.30514131 | 254.9140872 | 0.674964523 | -0.170719053 | FALSE | TRUE | FALSE |
| BPGM | 0.0587924 | 0.87318671 | 1 | 0.939812586 | 8.698555995 | 8.698555995 | 0.08394124 | 0.879356228 | 1 | 0.779246232 | 0.779246232 | 0.779246232 | 0.700399836 | -0.154653965 | FALSE | TRUE | FALSE |
| LAD1 | 0.0587864 | 0.87319638 | 0.0144 | 0.058786431 | 0.058786431 | 1.162413763 | 0.70278439 | 0.665450096 | 0.5048 | 0.702784394 | 0.702784394 | 1.562013487 | 0.083647889 | -1.077545017 | FALSE | TRUE | FALSE |
| ULK2 | 0.0587837 | 0.87320605 | 1 | 0.058783697 | 0.058783697 | 1.15150368 | 0.08181399 | 0.879834756 | 0.02415 | 0.081813987 | 0.081813987 | 1.606608386 | 0.718504246 | -0.143570661 | FALSE | TRUE | FALSE |
| NOL4 | 0.0586681 | 0.87321572 | 1 | 5416.971734 | 621443.0109 | 6885263.252 | 0.06394572 | 0.883800451 | 1 | 79071819.81 | 1.28375E+11 | 1.57197E+12 | 0.917466596 | -0.037409739 | FALSE | TRUE | FALSE |
| ARHGAP32 | 0.0586513 | 0.87322539 | 1 | 0.058651304 | 1.126015111 | 1.964041807 | 0.08162101 | 0.879898906 | 1 | 0.081621006 | 1.574579747 | 2.73818863 | 0.718581005 | -0.143524267 | FALSE | TRUE | FALSE |
| BCAM | 0.058638 | 0.87323505 | 1 | 0.058638012 | 0.058638012 | 1.152373791 | 0.0812975 | 0.879999582 | 1 | 0.081297499 | 0.081297499 | 1.601029341 | 0.721276958 | -0.141897942 | FALSE | TRUE | FALSE |
| DGKA | 0.0586365 | 0.87324471 | 1 | 0.538935171 | 0.538935171 | 10.61973736 | 0.08182515 | 0.879825586 | 1 | 0.754903528 | 0.754903528 | 2.766920088 | 0.716607259 | -0.144718797 | FALSE | TRUE | FALSE |
| RRM2B | 0.0586215 | 0.87325438 | 1 | 0.058621451 | 0.058621451 | 0.058621451 | 0.08196418 | 0.879788893 | 1 | 0.081964182 | 0.081964182 | 0.081964182 | 0.715208148 | -0.145567546 | FALSE | TRUE | FALSE |
| PSG2 | 0.0584227 | 0.87326404 | 0.00955 | 0.058422696 | 0.058422696 | 0.058422696 | 0.6973349 | 0.667331613 | 0.4997 | 0.078407758 | 0.697334896 | 0.697334896 | 0.083779969 | -1.076859806 | FALSE | TRUE | FALSE |
| EPS8L3 | 0.0583468 | 0.8732737 | 0.7559 | 0.520178987 | 4.759878263 | 10.28072927 | 0.08336887 | 0.87949462 | 1 | 0.749250955 | 0.749250955 | 1.650029122 | 0.69986301 | -0.154986959 | FALSE | TRUE | FALSE |
| PNPT1 | 0.0583231 | 0.87328335 | 1 | 0.05832309 | 0.05832309 | 1.149472015 | 0.08143699 | 0.87995384 | 0.01905 | 0.081436992 | 0.081436992 | 1.607830449 | 0.716174421 | -0.144981195 | FALSE | TRUE | FALSE |
| CST1 | 0.0582423 | 0.87329301 | 1 | 0.058242281 | 0.058242281 | 0.058242281 | 0.06727095 | 0.883028223 | 0.0037 | 0.067270951 | 0.067270951 | 0.067270951 | 0.865786498 | -0.062589191 | FALSE | TRUE | FALSE |
| ARHGEF1 | 0.0582388 | 0.87330267 | 1 | 0.058238761 | 0.058238761 | 1.135942703 | 0.31202244 | 0.821352977 | 0.75665 | 0.722429756 | 2.888719513 | 14.13297049 | 0.18664927 | -0.728973705 | FALSE | TRUE | FALSE |
| HOXA3 | 0.0581856 | 0.87331232 | 1 | 0.058185572 | 0.058185572 | 1.149156328 | 0.08151596 | 0.879935534 | 1 | 0.081515959 | 0.081515959 | 1.612350865 | 0.713793626 | -0.146427334 | FALSE | TRUE | FALSE |
| CLIP2 | 0.0581641 | 0.87332198 | 1 | 0.058164099 | 0.058164099 | 1.128597164 | 0.08141627 | 0.879962991 | 1 | 0.081416272 | 0.081416272 | 1.585454255 | 0.71440386 | -0.146056207 | FALSE | TRUE | FALSE |
| HAUS5 | 0.0581595 | 0.87333163 | 0.7482 | 0.515951134 | 4.697921866 | 10.16534205 | 0.08550178 | 0.87906001 | 0.01845 | 0.085501778 | 0.085501778 | 1.687587708 | 0.680213488 | -0.16735476 | FALSE | TRUE | FALSE |
| FSHR | 0.0581058 | 0.87334128 | 0.00815 | 0.058105767 | 0.058105767 | 0.058105767 | 0.20585319 | 0.850648175 | 0.75445 | 0.822813209 | 7.73149279 | 7.73149279 | 0.282268001 | -0.549338352 | FALSE | TRUE | FALSE |
| RGCC | 0.0580486 | 0.87335093 | 0.00365 | 0.05804861 | 0.05804861 | 0.05804861 | 0.06126403 | 0.884486667 | 0.00355 | 0.061264028 | 0.061264028 | 0.061264028 | 0.947515411 | -0.023413718 | FALSE | TRUE | FALSE |
| HTR3C | 0.0579959 | 0.87336057 | 1 | 0.525287149 | 2.100497255 | 2.100497255 | 2.94221716 | 0.41540073 | 0.2541 | 0.735755483 | 2.942217164 | 2.942217164 | 0.01971164 | -1.705277234 | FALSE | FALSE | FALSE |
| SKI | 0.0579559 | 0.87337022 | 0.01995 | 0.057955853 | 0.057955853 | 0.490891915 | 0.02393362 | 0.892748673 | 1 | 0.09564538 | 0.09564538 | 0.810470125 | 2.42152496 | 0.38408895 | FALSE | TRUE | FALSE |
| IRF9 | 0.0579544 | 0.87337986 | 1 | 0.057954371 | 0.057954371 | 1.149577215 | 0.08120609 | 0.880027012 | 1 | 0.081206088 | 0.081206088 | 1.612385455 | 0.713670266 | -0.146502397 | FALSE | TRUE | FALSE |
| IL1RL2 | 0.0579487 | 0.87338951 | 1 | 0.513356437 | 2.053380956 | 4.582965169 | 0.08118403 | 0.880036153 | 0.4997 | 0.08118403 | 0.72486519 | 1.607378395 | 0.71379405 | -0.146427076 | FALSE | TRUE | FALSE |
| B4GALNT3 | 0.0579155 | 0.87339915 | 0.75995 | 0.05791548 | 0.505369769 | 1.95275373 | 0.10615544 | 0.87433288 | 0.51645 | 0.106155443 | 0.106155443 | 2.080530819 | 0.545572215 | -0.263147756 | FALSE | TRUE | FALSE |
| ANO3 | 0.0578767 | 0.87340879 | 0.27185 | 0.057876733 | 0.231475053 | 1.950990327 | 0.20921925 | 0.84995474 | 0.02655 | 0.209219251 | 0.209219251 | 4.115451221 | 0.276631967 | -0.558097635 | FALSE | TRUE | FALSE |
| FAM161B | 0.0577948 | 0.87341843 | 1 | 0.513575108 | 0.513575108 | 1.954619287 | 0.08016516 | 0.880282465 | 1 | 0.717736558 | 0.717736558 | 1.584265872 | 0.72094696 | -0.142096685 | FALSE | TRUE | FALSE |
| RUNDC3B | 0.0577728 | 0.87342807 | 1 | 0.057772802 | 0.057772802 | 1.145810953 | 0.06664082 | 0.883202576 | 1 | 0.066640817 | 0.066640817 | 1.323017598 | 0.86692818 | -0.06201688 | FALSE | TRUE | FALSE |
| ZNF267 | 0.0577397 | 0.8734377 | 1 | 0.51657352 | 4.74346637 | 10.20225147 | 59.3709516 | 0.047529036 | 0.3217 | 6.27217676 | 576.0680745 | 576.0680745 | 0.000972524 | -3.012099701 | FALSE | FALSE | FALSE |
| C1RL | 0.0576595 | 0.87344734 | 0.01435 | 0.057659464 | 0.057659464 | 1.140709636 | 0.07810997 | 0.880835459 | 0.5082 | 0.716422664 | 0.716422664 | 1.54730532 | 0.738183142 | -0.131835877 | FALSE | TRUE | FALSE |
| HSDL2 | 0.0576488 | 0.87345697 | 0.00895 | 0.057648847 | 0.057648847 | 0.057648847 | 0.08065101 | 0.880173108 | 0.00915 | 0.080651012 | 0.080651012 | 0.080651012 | 0.71479384 | -0.145819199 | FALSE | TRUE | FALSE |
| TCTN2 | 0.0576201 | 0.87346661 | 0.5098 | 0.057620053 | 0.057620053 | 1.135998133 | 0.01119782 | 0.896380182 | 0.8798 | 0.399923229 | 3.667326809 | 3.667326809 | 5.145649084 | 0.711440165 | FALSE | TRUE | FALSE |
| KLB | 0.0575972 | 0.87347624 | 1 | 4.85615986 | 46.33450309 | 94.83364398 | 0.75518536 | 0.639092132 | 0.27555 | 0.755185358 | 7.068801842 | 25.46455298 | 0.076268909 | -1.117652466 | FALSE | TRUE | FALSE |
| GGT5 | 0.0575722 | 0.87348587 | 1 | 0.057572192 | 0.057572192 | 1.136591328 | 0.07984048 | 0.880382542 | 1 | 0.079840482 | 0.079840482 | 1.578658864 | 0.721090236 | -0.142010385 | FALSE | TRUE | FALSE |
| SPEF1 | 0.0575558 | 0.8734955 | 1 | 0.057555781 | 0.057555781 | 0.057555781 | 0.06290948 | 0.884032895 | 1 | 0.062909478 | 0.062909478 | 0.062909478 | 0.914898403 | -0.03862713 | FALSE | TRUE | FALSE |
| LPP | 0.0575253 | 0.87350512 | 1 | 0.057525294 | 0.057525294 | 1.135817588 | 0.70087489 | 0.666176376 | 0.7551 | 0.700874885 | 6.659903712 | 13.85979802 | 0.082076409 | -1.085781652 | FALSE | TRUE | FALSE |
| GPX4 | 0.0575167 | 0.87351475 | 1 | 0.057516652 | 0.057516652 | 0.057516652 | 0.55243686 | 0.732332841 | 0.50665 | 0.55243686 | 0.55243686 | 0.55243686 | 0.104114435 | -0.982489052 | FALSE | TRUE | FALSE |
| GRAMD1A | 0.0575137 | 0.87352437 | 1 | 0.520498349 | 4.834691109 | 10.211783 | 0.75291041 | 0.640200623 | 0.50885 | 0.752910412 | 0.752910412 | 14.80539588 | 0.076388521 | -1.116971897 | FALSE | TRUE | FALSE |
| HIVEP1 | 0.0575111 | 0.873534 | 1 | 475.9738801 | 91889.70884 | 954600.6017 | 0.86749211 | 0.603612151 | 0.88435 | 8.241400028 | 80.30485667 | 275.2638853 | 0.066295767 | -1.178514197 | FALSE | TRUE | FALSE |
| NCKAP5 | 0.057498 | 0.87354362 | 1 | 0.05749804 | 1.110226877 | 1.926627981 | 0.08024881 | 0.880236922 | 1 | 0.08024881 | 0.08024881 | 2.693640054 | 0.716497112 | -0.144785556 | FALSE | TRUE | FALSE |
| L3MBTL4 | 0.0574942 | 0.87355324 | 0.0173 | 0.057494208 | 0.057494208 | 1.136369539 | 0.70866026 | 0.661990888 | 0.50345 | 0.708660263 | 0.708660263 | 1.540081378 | 0.081130848 | -1.090813983 | FALSE | TRUE | FALSE |
| TRIM38 | 0.0574796 | 0.87356285 | 1 | 0.057479645 | 0.057479645 | 1.139518261 | 0.07975748 | 0.880409808 | 1 | 0.079757479 | 0.079757479 | 1.582836822 | 0.720680309 | -0.142257344 | FALSE | TRUE | FALSE |
| TCOF1 | 0.0574795 | 0.87357247 | 1 | 0.057479546 | 0.057479546 | 1.11863766 | 0.08045598 | 0.880200464 | 1 | 0.080455975 | 0.080455975 | 1.570874933 | 0.714422334 | -0.146044977 | FALSE | TRUE | FALSE |
| PRKD2 | 0.0574068 | 0.87358209 | 1 | 0.057406755 | 0.057406755 | 1.120225867 | 0.08046912 | 0.880191346 | 1 | 0.080469117 | 0.080469117 | 1.57486036 | 0.713401081 | -0.146666237 | FALSE | TRUE | FALSE |
| TRIM60 | 0.0574043 | 0.8735917 | 1 | 4.664153001 | 43.68810022 | 43.68810022 | 0.73670338 | 0.648168167 | 0.75725 | 0.736703375 | 6.783246623 | 6.783246623 | 0.077920498 | -1.10834828 | FALSE | TRUE | FALSE |
| TM4SF18 | 0.0573335 | 0.87360131 | 1 | 0.523187843 | 0.523187843 | 0.523187843 | 0.07036631 | 0.88237016 | 0.00385 | 0.070366308 | 0.070366308 | 0.070366308 | 0.814786469 | -0.088956192 | FALSE | TRUE | FALSE |
| ITGA5 | 0.0573237 | 0.87361093 | 0.03235 | 0.057323685 | 0.057323685 | 1.120656979 | 0.31044612 | 0.821862967 | 0.88245 | 1.24164354 | 4.967251615 | 24.33952441 | 0.184649384 | -0.733652136 | FALSE | TRUE | FALSE |
| EXOC4 | 0.0572752 | 0.87362054 | 0.0244 | 0.057275226 | 0.057275226 | 1.123798607 | 0.07962454 | 0.880473382 | 1 | 0.079624543 | 0.079624543 | 1.56587998 | 0.719316225 | -0.143080144 | FALSE | TRUE | FALSE |
| ACTA1 | 0.0572364 | 0.87363014 | 1 | 0.05723639 | 0.05723639 | 1.129740199 | 0.07991856 | 0.880364358 | 1 | 0.079918565 | 0.079918565 | 1.579929521 | 0.71618391 | -0.14497544 | FALSE | TRUE | FALSE |
| NPL | 0.0571763 | 0.87363975 | 0.69295 | 0.057176289 | 0.914593554 | 0.914593554 | 0.02732519 | 0.891883873 | 0.93915 | 0.109255863 | 1.747968481 | 1.747968481 | 2.092439209 | 0.32065285 | FALSE | TRUE | FALSE |

| | | | | | | | | | | | | | | | | | |
|---|---|---|---|---|---|---|---|---|---|---|---|---|---|---|---|---|---|
| MCM10 | 0.0571722 | 0.87364936 | 1 | 0.057172244 | 0.057172244 | 1.122937701 | 0.07960199 | 0.880482459 | 1 | 0.079601992 | 0.079601992 | 1.566863481 | 0.718226302 | -0.143738694 | FALSE | TRUE | FALSE |
| HOXB5 | 0.0571266 | 0.87365896 | 1 | 0.057126571 | 0.057126571 | 1.132564765 | 0.08018866 | 0.880273359 | 0.00955 | 0.080188656 | 0.080188656 | 0.080188656 | 0.712402154 | -0.147274776 | FALSE | TRUE | FALSE |
| ENAM | 0.0571179 | 0.87366857 | 1 | 0.511851898 | 0.511851898 | 10.04064175 | 0.65398349 | 0.687840952 | 0.93855 | 6.038957243 | 555.2991407 | 555.2991407 | 0.087338434 | -1.058794598 | FALSE | TRUE | FALSE |
| ENPP5 | 0.057098 | 0.87367817 | 1 | 0.228312487 | 0.228312487 | 1.132238209 | 18.938913 | 0.138213918 | 0.3514 | 10.7937834 | 172.8049743 | 691.688002 | 0.003014853 | -2.520733909 | FALSE | FALSE | FALSE |
| TIMM44 | 0.0570904 | 0.87368777 | 1 | 0.057090409 | 0.057090409 | 1.127782751 | 0.08000811 | 0.880346169 | 1 | 0.080008108 | 0.080008108 | 1.582842478 | 0.713557797 | -0.146570844 | FALSE | TRUE | FALSE |
| SOAT1 | 0.057082 | 0.87369737 | 1 | 0.516890773 | 0.516890773 | 1.933396809 | 0.08212912 | 0.879770539 | 0.01345 | 0.082129122 | 0.082129122 | 1.626664819 | 0.695027646 | -0.15799792 | FALSE | TRUE | FALSE |
| IRS1 | 0.0570614 | 0.87370696 | 1 | 0.228152891 | 0.228152891 | 4.409742858 | 0.73126811 | 0.650821007 | 0.03865 | 0.731268113 | 0.731268113 | 14.19124884 | 0.078030687 | -1.10773457 | FALSE | TRUE | FALSE |
| THYN1 | 0.0569798 | 0.87371656 | 0.4987 | 0.01425003 | 0.056979765 | 0.056979765 | 1.72414912 | 0.515703939 | 0.2511 | 0.76612641 | 6.894424222 | 6.894424222 | 0.03304805 | -1.480854167 | FALSE | FALSE | FALSE |
| CCDC88B | 0.0569486 | 0.87372616 | 0.93685 | 0.512592861 | 44.78807275 | 91.84125525 | 0.01084007 | 0.896527162 | 0.52195 | 0.096165707 | 0.096165707 | 1.87292139 | 5.253527776 | 0.720451033 | FALSE | TRUE | FALSE |
| RAD52 | 0.0569178 | 0.87373575 | 0.0132 | 0.056917819 | 0.056917819 | 1.127898267 | 0.07908371 | 0.880591281 | 0.0113 | 0.079083707 | 0.079083707 | 1.568874935 | 0.719716122 | -0.142838769 | FALSE | TRUE | FALSE |
| MON2 | 0.0569111 | 0.87374534 | 0.04165 | 0.056911054 | 0.056911054 | 1.908352102 | 0.07965308 | 0.880455225 | 1 | 0.079653075 | 0.079653075 | 2.675364463 | 0.714486581 | -0.146005923 | FALSE | TRUE | FALSE |
| CDADC1 | 0.0568814 | 0.87375493 | 0.01285 | 0.056881443 | 0.056881443 | 1.128218452 | 0.07895187 | 0.880663725 | 1 | 0.078951874 | 0.078951874 | 0.078951874 | 0.72045716 | -0.142391838 | FALSE | TRUE | FALSE |
| GPSM2 | 0.0568555 | 0.87376452 | 0.25805 | 0.014228719 | 0.056855531 | 0.219755037 | 0.11957545 | 0.871319736 | 0.38205 | 0.054147749 | 0.477979552 | 0.591821154 | 0.475478272 | -0.322869324 | FALSE | TRUE | FALSE |
| LHX1 | 0.0567696 | 0.87377411 | 0.50415 | 0.531241629 | 0.531241629 | 1.120302634 | 0.07970244 | 0.880427979 | 1 | 0.747135068 | 0.747135068 | 1.575378904 | 0.7122695 | -0.147355652 | FALSE | TRUE | FALSE |
| EXOG | 0.0567668 | 0.8737837 | 1 | 0.056766834 | 0.056766834 | 1.126593674 | 6.13930364 | 0.265055326 | 0.2542 | 1.4631694 | 24.55252308 | 24.55252308 | 0.009246461 | -2.03402444 | FALSE | FALSE | FALSE |
| ASZ1 | 0.0567634 | 0.87379328 | 0.50565 | 0.509886691 | 0.509886691 | 1.125773336 | 0.08154913 | 0.879926379 | 1 | 0.081549129 | 0.081549129 | 1.618969793 | 0.696063373 | -0.157351218 | FALSE | TRUE | FALSE |
| FBXO25 | 0.0567575 | 0.87380286 | 1 | 0.525482061 | 4.993265663 | 4.993265663 | 0.06281418 | 0.88405867 | 1 | 5.548718253 | 54.18882918 | 54.18882918 | 0.903578023 | -0.044034341 | FALSE | TRUE | FALSE |
| CLDN25 | 0.0567103 | 0.87381245 | 1 | 0.519633188 | 4.886840956 | 4.886840956 | 0.0844123 | 0.879236046 | 0.00505 | 0.084412302 | 0.084412302 | 0.084412302 | 0.671825182 | -0.172743721 | FALSE | TRUE | FALSE |
| FOXJ3 | 0.0566421 | 0.87382203 | 1 | 0.056642092 | 0.056642092 | 1.120040246 | 0.07873792 | 0.880708961 | 1 | 0.314734992 | 0.314734992 | 1.559079599 | 0.719375057 | -0.143044625 | FALSE | TRUE | FALSE |
| ZNF326 | 0.0566385 | 0.87383161 | 0.0161 | 0.056638453 | 0.056638453 | 1.120182733 | 0.06310866 | 0.883989914 | 1 | 0.063108664 | 0.063108664 | 1.249817562 | 0.897475074 | -0.046977605 | FALSE | TRUE | FALSE |
| TMEM26 | 0.0564793 | 0.87384119 | 1 | 0.056479345 | 0.056479345 | 1.120417934 | 0.07645879 | 0.881177585 | 1 | 0.691425653 | 0.691425653 | 0.691425653 | 0.738690045 | -0.131537754 | FALSE | TRUE | FALSE |
| CFH | 0.0564361 | 0.87385077 | 1 | 0.05643607 | 0.05643607 | 1.103822296 | 0.07833007 | 0.880763206 | 1 | 0.313201777 | 0.313201777 | 6.142132309 | 0.720490467 | -0.142371761 | FALSE | TRUE | FALSE |
| SRL | 0.0564117 | 0.87386034 | 0.6868 | 0.225483843 | 3.605232974 | 3.605232974 | 0.37274262 | 0.801419885 | 0.69005 | 0.372742622 | 5.961101752 | 5.961101752 | 0.151342257 | -0.820039794 | FALSE | TRUE | FALSE |
| SBNO1 | 0.0564105 | 0.87386992 | 1 | 0.05641046 | 0.05641046 | 1.895644369 | 0.07930465 | 0.880527823 | 1 | 0.079304645 | 0.079304645 | 2.6687215 | 0.711313441 | -0.147938985 | FALSE | TRUE | FALSE |
| SERTAD2 | 0.056395 | 0.87387949 | 0.75005 | 0.056395006 | 0.548504855 | 0.548504855 | 0.00636313 | 0.897764749 | 1 | 0.060373879 | 0.060373879 | 0.060373879 | 8.862781137 | 0.947570025 | FALSE | TRUE | FALSE |
| CLEC1A | 0.0563348 | 0.87388906 | 1 | 0.498459459 | 0.498459459 | 0.498459459 | 0.08061672 | 0.880182227 | 1 | 0.080616723 | 0.080616723 | 0.080616723 | 0.69879807 | -0.155648303 | FALSE | TRUE | FALSE |
| MATK | 0.056229 | 0.87389863 | 0.01905 | 0.056228963 | 0.056228963 | 1.107385025 | 0.07328217 | 0.881740962 | 0.87375 | 2.68759882 | 25.29958837 | 25.29958837 | 0.767293921 | -0.115038242 | FALSE | TRUE | FALSE |
| CIDEB | 0.0561762 | 0.8739082 | 0.00815 | 0.056176178 | 0.056176178 | 0.056176178 | 0.30565293 | 0.823624813 | 0.749 | 0.305652935 | 1.222762604 | 1.222762604 | 0.183790736 | -0.735676382 | FALSE | TRUE | FALSE |
| ECEL1 | 0.0561735 | 0.87391777 | 0.62815 | 0.0561735 | 0.0561735 | 2.06287755 | 0.12429576 | 0.870292536 | 0.38735 | 0.055058996 | 0.496745164 | 1.083725729 | 0.451934157 | -0.344924834 | FALSE | TRUE | FALSE |
| FBXO34 | 0.0561418 | 0.87392734 | 0.8787 | 4.908907084 | 47.6967455 | 47.6967455 | 0.07274671 | 0.881838845 | 1 | 0.072746709 | 0.072746709 | 1.438684993 | 0.771743028 | -0.112527285 | FALSE | TRUE | FALSE |
| SEMA4A | 0.056133 | 0.8739369 | 0.0232 | 0.056132955 | 0.056132955 | 1.104230535 | 0.07821085 | 0.880817404 | 0.0178 | 0.078210852 | 0.078210852 | 1.541580547 | 0.71771312 | -0.144049115 | FALSE | TRUE | FALSE |
| MKLN1 | 0.0560718 | 0.87394647 | 1 | 0.512395425 | 4.805776177 | 10.12619027 | 0.07309516 | 0.881767672 | 1 | 25.02359916 | 600.5549479 | 2401.423904 | 0.76710694 | -0.115144088 | FALSE | TRUE | FALSE |
| CLIP1 | 0.0560603 | 0.87395603 | 0.7586 | 0.056060276 | 0.512482835 | 1.885621218 | 0.01206004 | 0.89604275 | 1 | 0.012060043 | 0.012060043 | 0.235615787 | 4.64843095 | 0.667306384 | FALSE | TRUE | FALSE |
| MAP1S | 0.0558588 | 0.87396559 | 0.03165 | 0.055858792 | 0.055858792 | 1.08815653 | 2.40411922 | 0.455261249 | 0.00585 | 0.071128334 | 0.071128334 | 1.38995245 | 0.023234618 | -1.633864457 | FALSE | FALSE | FALSE |
| PPP1R1B | 0.0558117 | 0.87397515 | 1 | 0.055811724 | 0.055811724 | 0.055811724 | 0.0609849 | 0.884546355 | 1 | 0.060984896 | 0.060984896 | 0.060984896 | 0.915172895 | -0.038496851 | FALSE | TRUE | FALSE |
| TRMT10C | 0.0557669 | 0.87398471 | 1 | 0.055766867 | 0.055766867 | 0.055766867 | 0.06331899 | 0.883938298 | 0.51045 | 0.583965275 | 0.583965275 | 0.583965275 | 0.880728902 | -0.055157751 | FALSE | TRUE | FALSE |
| ZNF57 | 0.0557406 | 0.87399427 | 1 | 0.055740631 | 0.055740631 | 1.103352076 | 5.838495 | 0.274162978 | 0.49815 | 0.635781547 | 54.99347618 | 54.99347618 | 0.009547089 | -2.020129035 | FALSE | FALSE | FALSE |
| ANPEP | 0.055611 | 0.87400383 | 1 | 0.889835411 | 7.993092343 | 39.03751871 | 0.07684998 | 0.881069739 | 1 | 2.778562795 | 44.48286435 | 54.44043819 | 0.723630102 | -0.140483375 | FALSE | TRUE | FALSE |
| UBL7 | 0.0555811 | 0.87401338 | 1 | 0.055581097 | 0.055581097 | 0.055581097 | 0.07738207 | 0.88094369 | 1 | 0.077382066 | 0.077382066 | 0.077382066 | 0.718268458 | -0.143713205 | FALSE | TRUE | FALSE |
| GALNT16 | 0.0555668 | 0.87402294 | 0.49925 | 0.055566837 | 0.222158912 | 1.881086892 | 59.1564137 | 0.047759937 | 0.31735 | 6.093136024 | 588.7053116 | 588.7053116 | 0.000939321 | -3.027186158 | FALSE | FALSE | FALSE |
| QRSL1 | 0.0554846 | 0.87403249 | 0.01345 | 0.055484586 | 0.055484586 | 1.099665674 | 166.655035 | 0.01623906 | 0.0042 | 4.606469206 | 41.64151408 | 166.6550347 | 0.000332931 | -3.47764609 | FALSE | FALSE | FALSE |
| WTIP | 0.0554563 | 0.87404204 | 0.7539 | 0.512463223 | 4.860347722 | 4.860347722 | 0.08292645 | 0.879568312 | 1 | 0.082926451 | 0.082926451 | 0.082926451 | 0.668741027 | -0.174742032 | FALSE | TRUE | FALSE |
| SSMEM1 | 0.0553932 | 0.87405159 | 0.00635 | 0.055393191 | 0.055393191 | 0.055393191 | 0.07691294 | 0.881060743 | 1 | 0.076912942 | 0.076912942 | 0.076912942 | 0.720206378 | -0.142543037 | FALSE | TRUE | FALSE |
| KCNC1 | 0.0553882 | 0.87406114 | 1 | 0.055388238 | 0.055388238 | 1.088120832 | 0.07805174 | 0.880862533 | 1 | 0.07805174 | 0.07805174 | 1.536626864 | 0.709634894 | -0.148965038 | FALSE | TRUE | FALSE |
| PRC1 | 0.0553614 | 0.87407069 | 1 | 0.055361361 | 0.055361361 | 1.09360679 | 0.07496265 | 0.881410658 | 1 | 0.67172435 | 0.67172435 | 2.533946779 | 0.738519305 | -0.131638148 | FALSE | TRUE | FALSE |
| PARD3 | 0.0553595 | 0.87408024 | 1 | 0.501478239 | 0.501478239 | 9.743640207 | 0.07994421 | 0.880355264 | 1 | 0.07994421 | 0.07994421 | 1.558641443 | 0.692476221 | -0.159595136 | FALSE | TRUE | FALSE |
| C5orf34 | 0.0552946 | 0.87408978 | 1 | 0.48986981 | 0.48986981 | 1.870588263 | 0.68778805 | 0.671799833 | 0.5063 | 0.687788053 | 0.687788053 | 1.526920835 | 0.080394853 | -1.094771756 | FALSE | TRUE | FALSE |
| A1CF | 0.0552642 | 0.87409933 | 0.0162 | 0.055264158 | 0.055264158 | 1.093386227 | 0.67166437 | 0.679304209 | 0.5069 | 0.671664374 | 0.671664374 | 1.483059992 | 0.082279424 | -1.084708757 | FALSE | TRUE | FALSE |
| SIK3 | 0.0552518 | 0.87410887 | 1 | 48.49753301 | 488.2860106 | 5036.190722 | 0.09095805 | 0.877918613 | 1 | 0.090958045 | 0.090958045 | 1.77574666 | 0.607442381 | -0.216494911 | FALSE | TRUE | FALSE |
| PCED1B | 0.0552467 | 0.87411841 | 0.0138 | 0.055246707 | 0.055246707 | 1.091708933 | 0.06825668 | 0.882844639 | 1 | 0.068256683 | 0.068256683 | 1.350736489 | 0.809396296 | -0.091838787 | FALSE | TRUE | FALSE |
| BROX | 0.0552321 | 0.87412795 | 0.011 | 0.05523211 | 0.05523211 | 1.097454328 | 0.06891595 | 0.882660529 | 0.01005 | 0.068915951 | 0.068915951 | 1.370421698 | 0.801441604 | -0.096128117 | FALSE | TRUE | FALSE |
| PPARD | 0.0552087 | 0.87413749 | 1 | 0.518179876 | 2.071237839 | 4.361583256 | 0.08027049 | 0.880218696 | 0.5083 | 0.320879479 | 0.320879479 | 1.588757969 | 0.68778337 | -0.162548329 | FALSE | TRUE | FALSE |
| SEL1L2 | 0.0552018 | 0.87414702 | 0.75615 | 0.491239205 | 4.486839026 | 9.70918353 | 0.08174834 | 0.879871421 | 0.01385 | 0.081748335 | 0.081748335 | 1.618016632 | 0.675265345 | -0.170525538 | FALSE | TRUE | FALSE |
| SLC16A6 | 0.0551797 | 0.87415656 | 0.0137 | 0.0551797 | 0.0551797 | 1.091317567 | 0.07713163 | 0.881051746 | 1 | 0.077131633 | 0.077131633 | 1.527513862 | 0.715396491 | -0.145453195 | FALSE | TRUE | FALSE |
| TNIK | 0.0551429 | 0.87416609 | 1 | 0.515741452 | 0.515741452 | 10.05245891 | 0.31698495 | 0.819704827 | 0.5193 | 0.316984946 | 0.316984946 | 6.197380345 | 0.173960701 | -0.759548852 | FALSE | TRUE | FALSE |
| PCDHA13 | 0.0551386 | 0.87417563 | 0.89685 | 4.586257716 | 173.6055626 | 1683.959189 | 0.0523076 | 0.886179174 | 0.0296 | 0.052307597 | 0.052307597 | 1.020346454 | 1.054122674 | 0.022891155 | FALSE | TRUE | FALSE |
| SERINC5 | 0.055114 | 0.87418516 | 0.01115 | 0.055114039 | 0.055114039 | 1.093184956 | 0.06443599 | 0.88370551 | 1 | 0.064435993 | 0.064435993 | 0.064435993 | 0.855330007 | -0.067866292 | FALSE | TRUE | FALSE |
| RPS6KL1 | 0.0550846 | 0.87419469 | 0.5132 | 0.489986101 | 0.489986101 | 1.864912839 | 0.07918899 | 0.880564093 | 1 | 0.079188991 | 0.079188991 | 1.566549195 | 0.695609872 | -0.157634263 | FALSE | TRUE | FALSE |
| TTK | 0.0549604 | 0.87420422 | 1 | 0.219761093 | 0.878942737 | 4.333599014 | 0.30877951 | 0.822408906 | 0.75325 | 0.308779508 | 1.235028455 | 6.099279893 | 0.177992262 | -0.749598878 | FALSE | TRUE | FALSE |
| GUCY1A2 | 0.0549599 | 0.87421374 | 0.021 | 0.054959937 | 0.054959937 | 1.084000838 | 0.07746276 | 0.880934677 | 1 | 0.077462759 | 0.077462759 | 1.530376848 | 0.70950141 | -0.149046737 | FALSE | TRUE | FALSE |
| OR51E2 | 0.0549539 | 0.87422327 | 1 | 0.054953907 | 0.054953907 | 1.091220657 | 0.06417951 | 0.883765943 | 0.98535 | 49.6146466 | 4734.673827 | 47930.83248 | 0.856253099 | -0.067397844 | FALSE | TRUE | FALSE |
| UBE2Q2 | 0.0549487 | 0.87423279 | 0.00995 | 0.054948733 | 0.054948733 | 0.054948733 | 0.07226802 | 0.881963206 | 1 | 0.647059904 | 5.946282695 | 5.946282695 | 0.760346475 | -0.118988463 | FALSE | TRUE | FALSE |
| STAP2 | 0.0549069 | 0.87424232 | 1 | 0.054906931 | 0.054906931 | 1.088136467 | 0.07638242 | 0.881195542 | 1 | 0.07638242 | 0.07638242 | 1.51539247 | 0.718842512 | -0.143366247 | FALSE | TRUE | FALSE |
| STEAP4 | 0.0548985 | 0.87425184 | 1 | 0.878164168 | 7.904413601 | 7.904413601 | 6.13949672 | 0.26473811 | 0.3159 | 2.646187797 | 233.6651628 | 233.6651628 | 0.00894185 | -2.048572603 | FALSE | FALSE | FALSE |
| MFHAS1 | 0.0548891 | 0.87426136 | 0.5138 | 0.500310826 | 0.500310826 | 9.757900887 | 0.07946375 | 0.880509681 | 1 | 0.079463753 | 0.079463753 | 1.554470091 | 0.690743437 | -0.160683233 | FALSE | TRUE | FALSE |
| FAM20C | 0.0548801 | 0.87427088 | 1 | 0.054880145 | 0.054880145 | 0.054880145 | 0.07565162 | 0.881339027 | 0.74605 | 0.302394658 | 1.209034191 | 1.209034191 | 0.725432552 | -0.139402961 | FALSE | TRUE | FALSE |
| UTP18 | 0.0548635 | 0.87428039 | 1 | 0.054863462 | 0.054863462 | 1.083919158 | 0.07681214 | 0.881087726 | 0.0141 | 0.076812136 | 0.076812136 | 1.519773342 | 0.714255128 | -0.146146633 | FALSE | TRUE | FALSE |
| CACNB3 | 0.0548506 | 0.87428991 | 1 | 0.219272692 | 0.219272692 | 1.857832598 | 0.30891975 | 0.822311663 | 0.01505 | 0.308919747 | 0.308919747 | 1.528229421 | 0.177556112 | -0.750664373 | FALSE | TRUE | FALSE |
| B3GALNT2 | 0.0548397 | 0.87429942 | 1 | 0.486731635 | 0.486731635 | 1.086545546 | 0.07210156 | 0.882007562 | 1 | 5.898927765 | 55.44432147 | 55.44432147 | 0.760589447 | -0.118849705 | FALSE | TRUE | FALSE |
| ACTL8 | 0.0547655 | 0.87430894 | 1 | 0.054765515 | 0.054765515 | 1.084181675 | 0.07713503 | 0.881042749 | 0.0114 | 0.07713503 | 0.07713503 | 1.52889271 | 0.709995373 | -0.148744481 | FALSE | TRUE | FALSE |
| GPR161 | 0.0547554 | 0.87431845 | 0.506 | 0.497434585 | 0.497434585 | 1.854451805 | 0.07923397 | 0.880555027 | 1 | 0.079233965 | 0.079233965 | 1.568177496 | 0.691059194 | -0.16048475 | FALSE | TRUE | FALSE |
| TMOD4 | 0.054702 | 0.87432796 | 0.8739 | 0.874825828 | 3.499804469 | 3.499804469 | 2.6596064 | 0.436654561 | 0.63015 | 2.659606402 | 99.40464353 | 99.40464353 | 0.020567688 | -1.686814515 | FALSE | FALSE | FALSE |
| ENTPD6 | 0.0545646 | 0.87433747 | 1 | 0.054564572 | 0.054564572 | 1.078338116 | 0.07626224 | 0.881204519 | 1 | 0.07626224 | 0.07626224 | 1.509292927 | 0.715486101 | -0.145398798 | FALSE | TRUE | FALSE |

| | | | | | | | | | | | | | | | | | |
|---|---|---|---|---|---|---|---|---|---|---|---|---|---|---|---|---|---|
| PARP14 | 0.0545336 | 0.87434698 | 1 | 0.054533584 | 0.054533584 | 1.830829722 | 0.07653162 | 0.881168605 | 1 | 0.076531617 | 0.076531617 | 2.573246184 | 0.712562804 | -0.147176852 | FALSE | TRUE | FALSE |
| KRT71 | 0.0544919 | 0.87435649 | 0.32605 | 0.054491943 | 0.87061685 | 1.076088232 | 0.10886398 | 0.873662319 | 0.2546 | 0.027241283 | 0.108863978 | 0.538768239 | 0.500550722 | -0.300551908 | FALSE | TRUE | FALSE |
| MOB3A | 0.0544858 | 0.87436599 | 1 | 0.49924021 | 0.49924021 | 0.49924021 | 0.07903089 | 0.880627514 | 1 | 0.07903089 | 0.07903089 | 0.07903089 | 0.689424065 | -0.161513561 | FALSE | TRUE | FALSE |
| AXIN2 | 0.0544848 | 0.8743755 | 1 | 0.054484804 | 0.054484804 | 1.06681591 | 0.07683508 | 0.881078733 | 1 | 0.076835082 | 0.076835082 | 1.508240543 | 0.709113629 | -0.149284168 | FALSE | TRUE | FALSE |
| MALSU1 | 0.0544681 | 0.874385 | 1 | 0.054468139 | 0.054468139 | 0.054468139 | 0.07595414 | 0.881258357 | 1 | 0.075954137 | 0.075954137 | 0.075954137 | 0.717118786 | -0.144408901 | FALSE | TRUE | FALSE |
| L2HGDH | 0.054421 | 0.8743945 | 0.74475 | 0.054421003 | 0.217558469 | 1.078937021 | 0.09111709 | 0.877909175 | 0.4966 | 0.02279581 | 0.091117092 | 0.452417528 | 0.597264491 | -0.223833304 | FALSE | TRUE | FALSE |
| FEZ1 | 0.0544071 | 0.874404 | 1 | 0.054407138 | 0.054407138 | 0.054407138 | 0.05629927 | 0.885418398 | 1 | 0.056299266 | 0.056299266 | 0.056299266 | 0.966391614 | -0.014846847 | FALSE | TRUE | FALSE |
| CAPG | 0.0543959 | 0.8744135 | 1 | 0.054395856 | 0.054395856 | 0.054395856 | 0.07585877 | 0.881294223 | 1 | 0.075858772 | 0.075858772 | 1.506952399 | 0.717067446 | -0.144439994 | FALSE | TRUE | FALSE |
| SLC16A7 | 0.0542987 | 0.874423 | 0.01155 | 0.054298706 | 0.054298706 | 1.076754264 | 0.07397778 | 0.881625095 | 1 | 0.673717301 | 0.673717301 | 1.468492096 | 0.733986721 | -0.134311797 | FALSE | TRUE | FALSE |
| RNF207 | 0.0542558 | 0.87443249 | 0.75445 | 0.48570424 | 4.462761179 | 9.59302989 | 0.08067717 | 0.88014574 | 1 | 0.080677169 | 0.080677169 | 1.595825041 | 0.672504388 | -0.172304878 | FALSE | TRUE | FALSE |
| TNKS1BP1 | 0.0542016 | 0.87444199 | 1 | 0.054201583 | 1.044501054 | 1.819601475 | 0.0710039 | 0.882220058 | 1 | 0.071003904 | 0.071003904 | 2.387291208 | 0.763360602 | -0.117270258 | FALSE | TRUE | FALSE |
| SNAP47 | 0.0541763 | 0.87445148 | 1 | 0.495696412 | 4.654991573 | 4.654991573 | 5.66317877 | 0.279435301 | 0.2551 | 0.60048742 | 5.663178774 | 11.89692617 | 0.00956642 | -2.019250558 | FALSE | FALSE | FALSE |
| PRPH | 0.0541692 | 0.87446097 | 0.14395 | 0.05416918 | 0.21655729 | 0.459123934 | 0.23492068 | 0.843452251 | 0.0118 | 0.058784221 | 0.234920677 | 0.516596895 | 0.230584981 | -0.637168984 | FALSE | TRUE | FALSE |
| KSR2 | 0.0541106 | 0.87447046 | 0.5172 | 0.216320961 | 0.216320961 | 4.220756176 | 0.07726595 | 0.880988737 | 1 | 0.077265946 | 0.077265946 | 1.512014628 | 0.700316046 | -0.154705923 | FALSE | TRUE | FALSE |
| PKD1L1 | 0.0540525 | 0.87447995 | 0.0808 | 0.000879207 | 0.054052493 | 0.22662202 | 0.02468495 | 0.892546782 | 0.1495 | 0.00310131 | 0.02468495 | 0.470249542 | 2.189694234 | 0.340383475 | FALSE | TRUE | FALSE |
| SSTR5 | 0.0540373 | 0.87448944 | 1 | 0.054037335 | 0.054037335 | 1.06617307 | 5.98113285 | 0.270101437 | 0.2526 | 0.644816714 | 5.981132845 | 5.981132845 | 0.009034632 | -2.044089525 | FALSE | FALSE | FALSE |
| EIF4EBP1 | 0.0540115 | 0.87449893 | 1 | 3.90459E+39 | 7.08142E+47 | 9.04595E+50 | 0.06857324 | 0.882748267 | 1 | 1.49882E+42 | 1.12158E+50 | 4.97424E+53 | 0.787647559 | -0.103668069 | FALSE | TRUE | FALSE |
| REV3L | 0.0539957 | 0.87450841 | 0.76555 | 0.497677458 | 9.439422644 | 89.29519985 | 0.07838386 | 0.880745129 | 1 | 0.72561027 | 1.495986294 | 24.12973809 | 0.688862789 | -0.161867274 | FALSE | TRUE | FALSE |
| SNX14 | 0.0539318 | 0.8745179 | 1 | 0.053931818 | 0.053931818 | 1.061566722 | 0.07318976 | 0.881749867 | 0.51055 | 0.65395359 | 0.65395359 | 12.89661342 | 0.736876519 | -0.132605282 | FALSE | TRUE | FALSE |
| IFRD2 | 0.0539198 | 0.87452738 | 1 | 0.215610436 | 0.86238275 | 1.827746836 | 93.5276854 | 0.031103705 | 0.22885 | 23.38290286 | 1497.439492 | 5993.997573 | 0.000576511 | -3.239192132 | FALSE | FALSE | FALSE |
| NEO1 | 0.0538988 | 0.87453686 | 0.88 | 4.641390303 | 44.75503717 | 90.94740939 | 0.07828199 | 0.880790311 | 0.75525 | 0.72067274 | 6.809376141 | 24.25198349 | 0.688520777 | -0.16208295 | FALSE | TRUE | FALSE |
| TDP1 | 0.0538816 | 0.87454634 | 0.5082 | 0.053881635 | 0.215454855 | 0.49923214 | 0.01071777 | 0.8965621 | 0.874 | 0.817379417 | 7.417709169 | 7.417709169 | 5.027318355 | 0.701336388 | FALSE | TRUE | FALSE |
| MYRIP | 0.0538713 | 0.87455582 | 0.51185 | 0.49083685 | 0.49083685 | 9.647046094 | 0.07581505 | 0.881312148 | 1 | 0.694059044 | 0.694059044 | 13.66962163 | 0.710561876 | -0.148398098 | FALSE | TRUE | FALSE |
| AEBP2 | 0.0538499 | 0.8745653 | 1 | 0.47621787 | 0.47621787 | 0.47621787 | 0.07907702 | 0.880600342 | 1 | 0.079077019 | 0.079077019 | 0.079077019 | 0.680980386 | -0.166865396 | FALSE | TRUE | FALSE |
| FZD3 | 0.0538199 | 0.87457478 | 1 | 4.731253335 | 184.2805467 | 184.2805467 | 0.07680517 | 0.881096718 | 0.8786 | 6.792287631 | 66.3709637 | 66.3709637 | 0.700733087 | -0.154447375 | FALSE | TRUE | FALSE |
| RALGAPB | 0.0537979 | 0.87458425 | 1 | 0.215075206 | 0.215075206 | 4.183828479 | 0.30403394 | 0.824064485 | 0.5143 | 0.304033939 | 0.304033939 | 5.933833696 | 0.17694716 | -0.752156402 | FALSE | TRUE | FALSE |
| ZNF420 | 0.0537903 | 0.87459372 | 1 | 0.053790261 | 0.053790261 | 1.062662216 | 0.07312887 | 0.88175877 | 0.51155 | 0.656942969 | 0.656942969 | 2.473234096 | 0.735554416 | -0.133385193 | FALSE | TRUE | FALSE |
| NGFR | 0.0537826 | 0.87460319 | 1 | 0.053782613 | 0.053782613 | 1.061994211 | 0.07602368 | 0.881240416 | 0.01405 | 0.076023683 | 0.076023683 | 1.503461089 | 0.707445505 | -0.150307008 | FALSE | TRUE | FALSE |
| AMHR2 | 0.053756 | 0.87461267 | 0.5064 | 0.053755969 | 0.053755969 | 0.201430658 | 0.30059599 | 0.825370314 | 0.99355 | 1025.902044 | 272325.5091 | 2907632.66 | 0.178831287 | -0.747556497 | FALSE | TRUE | FALSE |
| ZNF638 | 0.0537359 | 0.87462213 | 0.04695 | 0.053735903 | 0.053735903 | 1.799517678 | 0.07584427 | 0.881303186 | 0.0449 | 0.075844273 | 0.075844273 | 2.544488724 | 0.708503109 | -0.14965824 | FALSE | TRUE | FALSE |
| PRKAG1 | 0.0537344 | 0.8746316 | 1 | 0.493922138 | 0.493922138 | 0.493922138 | 0.29614163 | 0.826378795 | 0.872 | 1.184302544 | 10.92699241 | 10.92699241 | 0.181448348 | -0.741246981 | FALSE | TRUE | FALSE |
| NR4A2 | 0.0537288 | 0.87464107 | 1 | 4966.965874 | 570224.0512 | 6319427.046 | 0.09068703 | 0.87801291 | 1 | 8.26246913 | 833.0768777 | 833.0768777 | 0.592464172 | -0.227337907 | FALSE | TRUE | FALSE |
| PPP1R1A | 0.0537232 | 0.87465053 | 1 | 0.053723214 | 0.053723214 | 0.053723214 | 0.05681282 | 0.885325828 | 1 | 0.056812817 | 0.056812817 | 0.056812817 | 0.94561784 | -0.024284343 | FALSE | TRUE | FALSE |
| KDM1B | 0.0537198 | 0.87466 | 0.0173 | 0.053719757 | 0.053719757 | 1.061577088 | 0.66419025 | 0.683310271 | 0.25945 | 0.664190253 | 6.349228879 | 13.14416342 | 0.080880075 | -1.092158457 | FALSE | TRUE | FALSE |
| CIT | 0.0536826 | 0.87466946 | 0.06855 | 0.053682638 | 0.113405045 | 1.792769017 | 0.01091611 | 0.896492202 | 1 | 0.043639886 | 0.209445353 | 3.348192358 | 4.917742577 | 0.691765791 | FALSE | TRUE | FALSE |
| SMC5 | 0.0536508 | 0.87467892 | 0.0317 | 0.05365077 | 0.05365077 | 1.051116292 | 0.67034076 | 0.68003464 | 0.51755 | 0.67034076 | 0.67034076 | 13.1654784 | 0.08003507 | -1.09671967 | FALSE | TRUE | FALSE |
| NAPSA | 0.0536017 | 0.87468838 | 0.0128 | 0.053601745 | 0.053601745 | 1.060827742 | 0.0752353 | 0.881374853 | 1 | 0.075235304 | 0.075235304 | 1.490848254 | 0.712454683 | -0.147242755 | FALSE | TRUE | FALSE |
| VCL | 0.0535957 | 0.87469784 | 0.5171 | 0.485645625 | 0.485645625 | 9.482939385 | 0.07538404 | 0.881347985 | 1 | 0.686746372 | 0.686746372 | 13.4479799 | 0.710969378 | -0.148149104 | FALSE | TRUE | FALSE |
| FANCD2 | 0.0535914 | 0.87470729 | 0.5191 | 0.053591387 | 0.053591387 | 1.045814466 | 0.01121365 | 0.896366163 | 0.51175 | 0.096007968 | 0.096007968 | 1.879036176 | 4.779120483 | 0.679347979 | FALSE | TRUE | FALSE |
| RABGAP1L | 0.0533862 | 0.87471675 | 1 | 0.4814497 | 4.456313245 | 4.456313245 | 0.07968911 | 0.880446144 | 1 | 0.079689108 | 0.079689108 | 0.079689108 | 0.669930789 | -0.173970062 | FALSE | TRUE | FALSE |
| ANKRD34B | 0.0533844 | 0.8747262 | 1 | 0.05338441 | 0.05338441 | 1.05838521 | 0.07464751 | 0.881455392 | 0.01065 | 0.074647508 | 0.074647508 | 1.481492679 | 0.715153271 | -0.145600871 | FALSE | TRUE | FALSE |
| HSDL1 | 0.0533277 | 0.87473566 | 0.4984 | 0.053327743 | 0.213169091 | 1.058882654 | 0.2979717 | 0.825987962 | 0.74975 | 0.297971702 | 1.191480819 | 1.191480819 | 0.178969151 | -0.747221821 | FALSE | TRUE | FALSE |
| HPD | 0.0533101 | 0.87474511 | 1 | 0.053310088 | 0.053310088 | 1.056873204 | 0.07522629 | 0.881383806 | 1 | 0.075226286 | 0.075226286 | 1.492931473 | 0.70866303 | -0.149560223 | FALSE | TRUE | FALSE |
| HAUS8 | 0.0532537 | 0.87475456 | 1 | 0.053253729 | 0.053253729 | 1.055135066 | 0.07445282 | 0.8815269 | 1 | 0.07445282 | 0.07445282 | 1.476816032 | 0.71526812 | -0.145531131 | FALSE | TRUE | FALSE |
| LIMCH1 | 0.0532141 | 0.87476401 | 0.51645 | 0.053214126 | 0.053214126 | 1.04047737 | 0.83089327 | 0.612346273 | 0.2616 | 0.093299333 | 0.830893266 | 3.147596739 | 0.064044478 | -1.193518309 | FALSE | TRUE | FALSE |
| TMPRSS3 | 0.0532046 | 0.87477345 | 0.01395 | 0.053204611 | 0.053204611 | 0.117912331 | 0.01115993 | 0.896401203 | 0.0122 | 0.011159934 | 0.011159934 | 0.221170086 | 4.767466367 | 0.678287638 | FALSE | TRUE | FALSE |
| XDH | 0.0531614 | 0.8747829 | 0.6307 | 0.212517435 | 7.309039876 | 35.57503484 | 0.0171132 | 0.894535097 | 0.89215 | 0.273606181 | 11.38708767 | 81.56594538 | 3.10645885 | 0.492265605 | FALSE | TRUE | FALSE |
| NUP133 | 0.053116 | 0.87479235 | 1 | 0.053115975 | 0.053115975 | 1.039619451 | 0.65441069 | 0.687463328 | 0.51635 | 0.654410687 | 0.654410687 | 12.84150771 | 0.081166118 | -1.090625225 | FALSE | TRUE | FALSE |
| FOXP2 | 0.0530986 | 0.87480179 | 1 | 0.053098573 | 0.053098573 | 1.048291994 | 0.07518912 | 0.881401708 | 0.0188 | 0.075189124 | 0.075189124 | 1.486712809 | 0.706200178 | -0.151072177 | FALSE | TRUE | FALSE |
| BSND | 0.0530978 | 0.87481123 | 0.00915 | 0.053097822 | 0.053097822 | 0.053097822 | 0.07246561 | 0.881927699 | 1 | 0.65726497 | 0.65726497 | 0.65726497 | 0.732731302 | -0.135055255 | FALSE | TRUE | FALSE |
| RAD51B | 0.053054 | 0.87482067 | 1 | 0.053054016 | 0.053054016 | 0.053054016 | 2.5453978 | 0.444271714 | 0.24865 | 0.285173458 | 2.545397799 | 2.545397799 | 0.020843114 | -1.681037399 | FALSE | FALSE | FALSE |
| SLC35A5 | 0.0530325 | 0.87483011 | 0.01085 | 0.053032509 | 0.053032509 | 1.05289731 | 2.52040485 | 0.445811453 | 0.00235 | 0.074315916 | 0.074315916 | 0.074315916 | 0.021041266 | -1.676928128 | FALSE | FALSE | FALSE |
| ACVR2B | 0.0530296 | 0.87483955 | 1 | 0.211959522 | 0.211959522 | 1.047161437 | 0.07594832 | 0.881276293 | 1 | 0.075948322 | 0.075948322 | 1.502017465 | 0.698232197 | -0.156000129 | FALSE | TRUE | FALSE |
| P2RX7 | 0.053025 | 0.87484899 | 0.01775 | 0.053024986 | 0.053024986 | 1.046817435 | 0.0741858 | 0.881580479 | 1 | 0.074185797 | 0.074185797 | 1.466847146 | 0.714759272 | -0.145840202 | FALSE | TRUE | FALSE |
| ZNF292 | 0.0529827 | 0.87485842 | 1 | 0.052982743 | 1.014537423 | 1.764163069 | 0.07467556 | 0.881446448 | 1 | 0.074675558 | 1.437273507 | 2.492660083 | 0.709505826 | -0.149044034 | FALSE | TRUE | FALSE |
| IL34 | 0.0529812 | 0.87486786 | 0.0072 | 0.052981212 | 0.052981212 | 0.052981212 | 0.05748245 | 0.885165617 | 0.0076 | 0.057482452 | 0.057482452 | 0.057482452 | 0.921693659 | -0.0354134 | FALSE | TRUE | FALSE |
| GPLD1 | 0.0528898 | 0.87487729 | 0.50985 | 0.052889792 | 0.052889792 | 1.039279809 | 67.5549806 | 0.042123011 | 0.0667 | 0.783590241 | 67.55498055 | 67.55498055 | 0.000782915 | -3.106285512 | FALSE | FALSE | FALSE |
| PRLR | 0.0528835 | 0.87488672 | 1 | 1.83131E+36 | 9.17587E+44 | 5.95071E+48 | 0.01382827 | 0.895504314 | 1 | 1.10637E+32 | 2.34541E+39 | 1.28315E+45 | 3.824300112 | 0.582551966 | FALSE | TRUE | FALSE |
| ZNF862 | 0.0528668 | 0.87489615 | 0.6952 | 0.052866755 | 4.388570343 | 9.257869307 | 0.7874759 | 0.625017628 | 0.5029 | 0.787475896 | 7.343249518 | 70.15160791 | 0.067134442 | -1.173054615 | FALSE | TRUE | FALSE |
| YTHDC2 | 0.05286 | 0.87490558 | 0.0403 | 0.052859979 | 0.052859979 | 1.773422101 | 0.07235257 | 0.881945455 | 0.5176 | 0.663633921 | 0.663633921 | 12.96944388 | 0.73058882 | -0.136326978 | FALSE | TRUE | FALSE |
| RD3 | 0.052852 | 0.87491501 | 0.00695 | 0.052851968 | 0.052851968 | 0.052851968 | 0.05635167 | 0.885409988 | 1 | 0.056351668 | 0.056351668 | 0.056351668 | 0.93789536 | -0.027845613 | FALSE | TRUE | FALSE |
| ZNF700 | 0.0527943 | 0.87492444 | 0.5101 | 0.052794325 | 0.052794325 | 0.210435523 | 0.01137078 | 0.896303034 | 1 | 0.011370775 | 0.011370775 | 0.22495239 | 4.642983878 | 0.666797176 | FALSE | TRUE | FALSE |
| IGSF11 | 0.0527529 | 0.87493386 | 0.0102 | 0.052752901 | 0.052752901 | 1.046776186 | 0.07421569 | 0.881562624 | 1 | 0.074215688 | 0.074215688 | 0.074215688 | 0.710805261 | -0.148249366 | FALSE | TRUE | FALSE |
| CYP17A1 | 0.0527408 | 0.87494328 | 1 | 0.052740797 | 0.052740797 | 1.043433438 | 0.07441717 | 0.881535833 | 1 | 0.074417167 | 0.074417167 | 1.474193927 | 0.708718142 | -0.14952645 | FALSE | TRUE | FALSE |
| TAF1L | 0.0526253 | 0.87495271 | 1 | 0.05262525 | 1.015213667 | 1.763788863 | 0.0740982 | 0.881598329 | 1 | 0.074098201 | 0.074098201 | 2.48773127 | 0.710209552 | -0.148613491 | FALSE | TRUE | FALSE |
| VSTM1 | 0.0524647 | 0.87496213 | 0.0072 | 0.052464663 | 0.052464663 | 0.052464663 | 0.0609905 | 0.884537832 | 1 | 0.0609905 | 0.0609905 | 0.0609905 | 0.860210404 | -0.065395309 | FALSE | TRUE | FALSE |
| LMLN | 0.0524416 | 0.87497155 | 0.50885 | 0.464697464 | 0.464697464 | 9.162847422 | 0.29081341 | 0.827851945 | 0.75695 | 0.649318987 | 2.597207893 | 5.743957772 | 0.180327389 | -0.743938304 | FALSE | TRUE | FALSE |
| MCM2 | 0.0524163 | 0.87498097 | 0.03195 | 0.052416295 | 0.052416295 | 1.02176768 | 0.07400156 | 0.881616174 | 1 | 0.074001557 | 0.074001557 | 1.44694022 | 0.708313412 | -0.149774535 | FALSE | TRUE | FALSE |
| NEU3 | 0.0523902 | 0.87499039 | 0.99785 | 17.36513875 | 1111.492648 | 4448.357147 | 0.38169534 | 0.798548042 | 0.69055 | 0.381695336 | 6.107214977 | 6.107214977 | 0.137256514 | -0.862467034 | FALSE | TRUE | FALSE |
| SLC13A2 | 0.0523653 | 0.8749998 | 1 | 0.465320196 | 4.243931875 | 9.178024807 | 0.67838449 | 0.675938724 | 0.5055 | 0.678384486 | 0.678384486 | 1.495693055 | 0.077191214 | -1.112432128 | FALSE | TRUE | FALSE |
| RARRES2 | 0.0523575 | 0.87500922 | 1 | 0.492051156 | 0.492051156 | 0.492051156 | 0.05701762 | 0.885283706 | 0.0028 | 0.057017622 | 0.057017622 | 0.057017622 | 0.918268541 | -0.037030294 | FALSE | TRUE | FALSE |
| TJP3 | 0.0522992 | 0.87501863 | 0.517 | 0.113271475 | 0.453037399 | 3.819333033 | 5.31403181 | 0.293646748 | 0.21635 | 1.327652447 | 25.86053583 | 56.56038205 | 0.009841723 | -2.006928851 | FALSE | FALSE | FALSE |
| WWP2 | 0.0522436 | 0.87502804 | 1 | 0.487133068 | 0.487133068 | 9.526281309 | 0.28378992 | 0.829944058 | 0.8797 | 2.649396366 | 25.38544081 | 51.94971 | 0.184092626 | -0.734963608 | FALSE | TRUE | FALSE |

| | | | | | | | | | | | | | | | | | |
|---|---|---|---|---|---|---|---|---|---|---|---|---|---|---|---|---|---|
| GNL2 | 0.0521624 | 0.87503745 | 1 | 0.052162398 | 0.052162398 | 1.02772073 | 0.07336766 | 0.881705333 | 0.01845 | 0.073367663 | 0.073367663 | 1.448103323 | 0.710972589 | -0.148147143 | FALSE | TRUE | FALSE |
| MAPKBP1 | 0.0520883 | 0.87504686 | 0.04455 | 0.052088287 | 0.052088287 | 1.748872472 | 0.29093892 | 0.827778881 | 0.5166 | 0.290938916 | 0.290938916 | 5.650399456 | 0.179035133 | -0.747061738 | FALSE | TRUE | FALSE |
| NMRAL1 | 0.052055 | 0.87505627 | 1 | 0.052055049 | 0.052055049 | 1.033220133 | 0.07298099 | 0.881776573 | 1 | 0.072980988 | 0.072980988 | 0.072980988 | 0.713268624 | -0.14674688 | FALSE | TRUE | FALSE |
| SP140 | 0.0519885 | 0.87506568 | 1 | 0.051988528 | 0.051988528 | 1.023919368 | 0.07329294 | 0.881732057 | 0.0174 | 0.073292944 | 0.073292944 | 1.446160781 | 0.709325149 | -0.149154642 | FALSE | TRUE | FALSE |
| ZNF230 | 0.0519538 | 0.87507509 | 1 | 0.051953802 | 0.051953802 | 1.030234607 | 0.07285343 | 0.881821059 | 0.0101 | 0.072853425 | 0.072853425 | 1.446148811 | 0.713127783 | -0.146832643 | FALSE | TRUE | FALSE |
| MAP4K2 | 0.0519075 | 0.87508449 | 1 | 0.051907499 | 0.051907499 | 1.018949401 | 0.07095489 | 0.882228897 | 1 | 0.640262868 | 0.640262868 | 12.59641431 | 0.73155636 | -0.13575221 | FALSE | TRUE | FALSE |
| TAP2 | 0.0518947 | 0.8750939 | 0.8777 | 0.207450334 | 1.900559487 | 4.088153154 | 0.09024054 | 0.878116477 | 0.50405 | 0.09024054 | 0.09024054 | 1.781478873 | 0.575070753 | -0.240278719 | FALSE | TRUE | FALSE |
| GPS2 | 0.0518652 | 0.8751033 | 0.00825 | 0.05186523 | 0.05186523 | 0.05186523 | 0.05616556 | 0.885443622 | 1 | 0.056165561 | 0.056165561 | 0.056165561 | 0.923434735 | -0.034593793 | FALSE | TRUE | FALSE |
| SEC24D | 0.0518636 | 0.8751127 | 0.5095 | 0.051863623 | 0.454210752 | 1.749570633 | 0.10376193 | 0.87497696 | 0.0231 | 0.025951624 | 0.103761933 | 0.510680315 | 0.499832852 | -0.301175203 | FALSE | TRUE | FALSE |
| GUCY2C | 0.05185 | 0.8751221 | 0.1391 | 0.006156967 | 0.051849997 | 0.254207107 | 4.12467641 | 0.347945339 | 0.19935 | 1.031590181 | 36.99727251 | 147.9656475 | 0.012570682 | -1.900641146 | FALSE | FALSE | FALSE |
| ARG2 | 0.0518359 | 0.8751315 | 0.75245 | 0.051835939 | 0.207281687 | 0.257601913 | 0.0219237 | 0.893261539 | 1 | 0.021923701 | 0.021923701 | 0.021923701 | 2.364379046 | 0.373717102 | FALSE | TRUE | FALSE |
| POU1F1 | 0.0516749 | 0.87514089 | 1 | 0.051674895 | 0.051674895 | 0.051674895 | 0.07333024 | 0.88172315 | 1 | 0.073330241 | 0.073330241 | 0.073330241 | 0.704687368 | -0.152003513 | FALSE | TRUE | FALSE |
| DECR1 | 0.0516651 | 0.87515029 | 0.50675 | 0.051665077 | 0.051665077 | 0.051665077 | 0.02173603 | 0.893320711 | 0.0076 | 0.021736029 | 0.021736029 | 0.021736029 | 2.376932654 | 0.376016877 | FALSE | TRUE | FALSE |
| FHOD1 | 0.0516507 | 0.87515968 | 0.9412 | 0.452118699 | 37.36321842 | 78.742237 | 0.01144704 | 0.896274953 | 0.75675 | 0.098962212 | 0.878131118 | 3.340435556 | 4.512144778 | 0.654383026 | FALSE | TRUE | FALSE |
| PRICKLE2 | 0.0516286 | 0.87516908 | 1 | 0.051628641 | 0.051628641 | 1.011556127 | 0.07335986 | 0.881714242 | 0.02525 | 0.07335986 | 0.07335986 | 1.440857752 | 0.703772344 | -0.152567804 | FALSE | TRUE | FALSE |
| ABHD12B | 0.0516233 | 0.87517847 | 1 | 1.81618825 | 16.39648795 | 16.39648795 | 0.64891691 | 0.690395602 | 0.75385 | 0.648916912 | 5.909106595 | 5.909106595 | 0.079552975 | -1.099343575 | FALSE | TRUE | FALSE |
| SPON2 | 0.051543 | 0.87518786 | 1 | 0.051542951 | 0.051542951 | 1.022047664 | 0.07255004 | 0.881892172 | 1 | 0.072550038 | 0.072550038 | 0.086225308 | 0.710446922 | -0.148468363 | FALSE | TRUE | FALSE |
| DOK3 | 0.0515389 | 0.87519725 | 0.00775 | 0.051538897 | 0.051538897 | 0.051538897 | 0.07042863 | 0.882352519 | 1 | 0.633347534 | 0.633347534 | 0.633347534 | 0.731788963 | -0.135614145 | FALSE | TRUE | FALSE |
| SLC6A17 | 0.0515312 | 0.87520664 | 1 | 0.480954672 | 4.607093362 | 9.449367723 | 0.07788822 | 0.880889596 | 1 | 0.077888218 | 0.077888218 | 1.533529078 | 0.661604264 | -0.179401704 | FALSE | TRUE | FALSE |
| C5orf46 | 0.0514738 | 0.87521602 | 1 | 0.051473767 | 0.051473767 | 0.051473767 | 0.05318534 | 0.886037731 | 0.00195 | 0.053185337 | 0.053185337 | 0.053185337 | 0.967818772 | -0.014205959 | FALSE | TRUE | FALSE |
| HERC6 | 0.0514527 | 0.87522541 | 0.7484 | 0.051452681 | 0.461233623 | 1.011960216 | 0.17145996 | 0.859244065 | 0.5182 | 0.171459956 | 1.512922088 | 3.378980837 | 0.300085697 | -0.522754704 | FALSE | TRUE | FALSE |
| YEATS2 | 0.0513501 | 0.87523479 | 1 | 0.051350072 | 0.051350072 | 1.726716073 | 0.07249918 | 0.881918819 | 0.0349 | 0.072499183 | 0.072499183 | 2.441106766 | 0.70828483 | -0.14979206 | FALSE | TRUE | FALSE |
| ZNF234 | 0.0512759 | 0.87524418 | 0.75175 | 0.051275942 | 0.445019995 | 1.013084611 | 0.01048178 | 0.896638887 | 1 | 0.787910953 | 7.098847957 | 7.098847957 | 4.891911275 | 0.689478572 | FALSE | TRUE | FALSE |
| PLXNB2 | 0.0512478 | 0.87525356 | 0.05795 | 0.051247841 | 0.974133033 | 1.720314297 | 0.06619584 | 0.88330696 | 1 | 5.752862263 | 55.72687452 | 193.4099777 | 0.774185272 | -0.111155095 | FALSE | TRUE | FALSE |
| PCK1 | 0.0512021 | 0.87526294 | 1 | 38.74063595 | 3644.613389 | 36634.2155 | 0.66454771 | 0.683115745 | 0.81325 | 57.58703597 | 556.5163463 | 5509.574859 | 0.077048056 | -1.113238316 | FALSE | TRUE | FALSE |
| NOP14 | 0.0511972 | 0.87527232 | 0.0277 | 0.051197176 | 0.051197176 | 1.003716606 | 0.07003864 | 0.882458285 | 1 | 0.629829255 | 0.629829255 | 12.37712788 | 0.730984735 | -0.136091692 | FALSE | TRUE | FALSE |
| NCAPD2 | 0.0511965 | 0.8752817 | 0.51645 | 0.051196535 | 0.442994893 | 1.720537802 | 0.02826171 | 0.891671411 | 0.03495 | 0.028261715 | 0.028261715 | 0.550867459 | 1.81151553 | 0.258042062 | FALSE | TRUE | FALSE |
| CHRD | 0.0511536 | 0.87529107 | 1 | 0.4630574 | 0.4630574 | 9.045962962 | 0.07455938 | 0.881500094 | 0.02795 | 0.074559381 | 0.074559381 | 1.460616943 | 0.686078923 | -0.163625922 | FALSE | TRUE | FALSE |
| USP36 | 0.0511306 | 0.87530045 | 0.5171 | 0.457914553 | 0.457914553 | 8.91256568 | 0.07210876 | 0.881998694 | 1 | 0.650104659 | 0.650104659 | 12.69415578 | 0.709076033 | -0.149307193 | FALSE | TRUE | FALSE |
| SH3D19 | 0.0511098 | 0.87530982 | 0.50825 | 0.465277722 | 0.465277722 | 1.726519175 | 0.66197431 | 0.684857263 | 0.50485 | 0.66197431 | 0.66197431 | 2.446439662 | 0.077208085 | -1.112337217 | FALSE | TRUE | FALSE |
| DNTTIP2 | 0.0510859 | 0.87531919 | 1 | 0.051085921 | 0.051085921 | 1.009539812 | 5.30729002 | 0.294833152 | 0.5047 | 5.307290024 | 49.5861085 | 49.5861085 | 0.009625613 | -2.016571592 | FALSE | FALSE | FALSE |
| BTBD6 | 0.0510789 | 0.87532856 | 0.99335 | 89.82145047 | 8728.306244 | 89093.09006 | 0.0247803 | 0.89250182 | 0.7518 | 0.221162811 | 2.025915614 | 2.025915614 | 2.061271449 | 0.314135188 | FALSE | TRUE | FALSE |
| DNAJB7 | 0.051067 | 0.87533793 | 0.75395 | 0.451538842 | 4.097892062 | 4.097892062 | 0.0717574 | 0.882078469 | 0.7525 | 0.639620789 | 5.851707951 | 5.851707951 | 0.71166111 | -0.147726767 | FALSE | TRUE | FALSE |
| A4GALT | 0.0510626 | 0.8753473 | 1 | 0.467397435 | 0.467397435 | 1.009885383 | 0.07462183 | 0.881491156 | 0.01195 | 0.074621825 | 0.074621825 | 1.477801647 | 0.684285091 | -0.164762922 | FALSE | TRUE | FALSE |
| WDR7 | 0.0510358 | 0.87535667 | 1 | 0.051035779 | 0.051035779 | 1.713733537 | 0.07232648 | 0.881954331 | 1 | 0.072326483 | 0.072326483 | 2.432273258 | 0.705630601 | -0.151422594 | FALSE | TRUE | FALSE |
| CCDC150 | 0.0509684 | 0.87536603 | 0.9136 | 35.64483582 | 3164.096752 | 30846.58353 | 0.01352619 | 0.895618095 | 0.87695 | 0.116580148 | 1.030647993 | 2.294006313 | 3.768123648 | 0.576125145 | FALSE | TRUE | FALSE |
| ARHGEF12 | 0.0509133 | 0.8753754 | 1 | 43.23149025 | 4391.40107 | 45864.33351 | 0.08179622 | 0.879843924 | 0.7579 | 0.760074193 | 7.248835712 | 141.2368879 | 0.622440381 | -0.20590224 | FALSE | TRUE | FALSE |
| DIP2A | 0.0508528 | 0.87538476 | 0.8784 | 4.236982674 | 40.18803666 | 142.5783064 | 0.0676471 | 0.882958348 | 1 | 54.14198096 | 1048.836316 | 5287.399901 | 0.751736075 | -0.123934608 | FALSE | TRUE | FALSE |
| ZNF785 | 0.0508241 | 0.87539412 | 0.50925 | 0.450582006 | 0.450582006 | 1.004266926 | 0.60120011 | 0.711807083 | 0.8773 | 5.50362867 | 51.67702607 | 51.67702607 | 0.084537735 | -1.072949394 | FALSE | TRUE | FALSE |
| NDUFA10 | 0.0508158 | 0.87540348 | 0.7555 | 0.050815811 | 0.464067993 | 0.464067993 | 0.85958204 | 0.605472855 | 0.00685 | 0.093434527 | 0.859582042 | 0.859582042 | 0.059116883 | -1.228288469 | FALSE | TRUE | FALSE |
| ETV7 | 0.0508113 | 0.87541284 | 0.0107 | 0.050811257 | 0.050811257 | 1.009794851 | 0.0713439 | 0.882149302 | 1 | 0.071343903 | 0.071343903 | 0.071343903 | 0.712201811 | -0.147396926 | FALSE | TRUE | FALSE |
| CLU | 0.0507662 | 0.8754222 | 1 | 0.050766206 | 0.050766206 | 1.003353788 | 0.05766686 | 0.885114938 | 1 | 0.057666856 | 0.057666856 | 1.141356346 | 0.880335945 | -0.055351565 | FALSE | TRUE | FALSE |
| GLDC | 0.0507443 | 0.87543156 | 0.92495 | 10.72148631 | 3290.615816 | 29960.35793 | 0.2473647 | 0.840084691 | 0.6039 | 0.542044107 | 34.5842629 | 552.9036282 | 0.205139663 | -0.687950363 | FALSE | TRUE | FALSE |
| PNMT | 0.0507383 | 0.87544091 | 1 | 0.050738345 | 0.050738345 | 0.050738345 | 0.05231622 | 0.886170863 | 1 | 0.488704377 | 4.685367713 | 4.685367713 | 0.969839664 | -0.013300058 | FALSE | TRUE | FALSE |
| ANKRD17 | 0.0506579 | 0.87545027 | 1 | 44.87958754 | 4689.323947 | 49662.54253 | 0.08698962 | 0.878790459 | 1 | 0.086989619 | 1.671970453 | 2.911084954 | 0.58234434 | -0.234820141 | FALSE | TRUE | FALSE |
| FAM124A | 0.0506375 | 0.87545962 | 1 | 0.050637472 | 0.050637472 | 0.999399444 | 0.07122599 | 0.882166998 | 1 | 0.071225991 | 0.071225991 | 1.407971738 | 0.710940933 | -0.14816648 | FALSE | TRUE | FALSE |
| MAGI1 | 0.0506312 | 0.87546897 | 0.52545 | 0.202465595 | 0.202465595 | 3.931666523 | 0.5902215 | 0.716450942 | 0.69365 | 5.562306781 | 532.7066521 | 1047.724697 | 0.085783332 | -1.066597087 | FALSE | TRUE | FALSE |
| PCYOX1 | 0.0506186 | 0.87547832 | 1 | 0.450837935 | 0.450837935 | 1.001470677 | 0.64025926 | 0.694115171 | 0.50765 | 0.640259265 | 0.640259265 | 1.413776789 | 0.0790595 | -1.102045935 | FALSE | TRUE | FALSE |
| BBS1 | 0.0506123 | 0.87548767 | 0.25125 | 9.93997E-05 | 0.202220287 | 0.202220287 | 0.922734 | 0.595781555 | 0.11525 | 0.057737377 | 0.922734004 | 3.690191957 | 0.054850413 | -1.260820095 | FALSE | TRUE | FALSE |
| ERCC1 | 0.0505709 | 0.87549701 | 1 | 1.13464E+18 | 1.53537E+23 | 3.12569E+25 | 0.53047497 | 0.742743881 | 1 | 8.47663E+21 | 5.27095E+27 | 4.88977E+30 | 0.095331341 | -1.0207643 | FALSE | TRUE | FALSE |
| EVL | 0.0505621 | 0.87550636 | 1 | 8398777903 | 1.88741E+14 | 2.42598E+15 | 0.08648435 | 0.878911424 | 1 | 9424393.772 | 14304404822 | 2.0962E+12 | 0.584638542 | -0.233112557 | FALSE | TRUE | FALSE |
| CD163 | 0.0505433 | 0.8755157 | 1 | 0.050543285 | 0.050543285 | 0.989062712 | 0.07070189 | 0.882273075 | 1 | 0.282768286 | 0.282768286 | 5.547763476 | 0.714878857 | -0.145767547 | FALSE | TRUE | FALSE |
| GZMM | 0.05051 | 0.87552505 | 0.7552 | 0.459028635 | 1.835331866 | 1.835331866 | 0.64070353 | 0.693934432 | 0.75105 | 0.64070353 | 6.000899285 | 6.000899285 | 0.078835164 | -1.103280027 | FALSE | TRUE | FALSE |
| ANKZF1 | 0.0504818 | 0.87553439 | 0.2599 | 0.012629461 | 0.050481848 | 0.248262274 | 0.01272357 | 0.89584497 | 0.75135 | 0.012723566 | 0.050874802 | 0.250627964 | 3.967586341 | 0.598526387 | FALSE | TRUE | FALSE |
| FGD6 | 0.0504577 | 0.87554373 | 0.7581 | 0.05045767 | 0.20176187 | 1.697506597 | 0.02152837 | 0.893357664 | 1 | 0.021528374 | 0.021528374 | 0.421390746 | 2.34377522 | 0.369915958 | FALSE | TRUE | FALSE |
| TUBA3E | 0.0504334 | 0.87555307 | 0.894 | 0.201587909 | 3.223163751 | 12.8930008 | 5.59744564 | 0.283417428 | 0.83655 | 109.1496525 | 7550.28233 | 72015.94226 | 0.009010075 | -2.045271601 | FALSE | FALSE | FALSE |
| KRT31 | 0.0504151 | 0.87556241 | 0.26215 | 0.050415074 | 0.201488515 | 0.99727545 | 0.18915493 | 0.854578334 | 0.7484 | 0.189154935 | 1.806487521 | 1.806487521 | 0.266527933 | -0.574257269 | FALSE | TRUE | FALSE |
| WDR70 | 0.0503739 | 0.87557174 | 1 | 0.458510937 | 0.458510937 | 1.703810205 | 0.07365222 | 0.881660768 | 1 | 0.073652221 | 0.073652221 | 1.455065277 | 0.683942335 | -0.164980513 | FALSE | TRUE | FALSE |
| KRT85 | 0.0503708 | 0.87558108 | 0.51175 | 0.050370778 | 0.050370778 | 0.426775094 | 0.31965419 | 0.818879257 | 0.69135 | 0.735767539 | 11.76198217 | 11.76198217 | 0.157578971 | -0.802501739 | FALSE | TRUE | FALSE |
| ANKRD30A | 0.0503674 | 0.87559041 | 0.89455 | 3.844952528 | 324.1350653 | 3084.356681 | 4905.19442 | 0.000351822 | 0.50915 | 4905.194416 | 484639.3161 | 4980605.217 | 1.02682E-05 | -4.988506775 | FALSE | FALSE | FALSE |
| COL1A2 | 0.0502976 | 0.87559974 | 0.2658 | 0.012598325 | 0.200858656 | 0.977223112 | 4.9577E-06 | 0.900861927 | 0.15545 | 4.95766E-06 | 1.97781E-05 | 9.6653E-05 | 10145.44297 | 4.006271014 | FALSE | TRUE | FALSE |
| VPS16 | 0.0502614 | 0.87560907 | 1 | 0.050261412 | 0.050261412 | 0.985666979 | 5.70176126 | 0.277271842 | 0.2621 | 0.610362735 | 5.701761258 | 11.99775753 | 0.008815068 | -2.054774342 | FALSE | FALSE | FALSE |
| UPK1B | 0.0502224 | 0.8756184 | 1 | 0.200798712 | 0.200798712 | 0.200798712 | 0.25997953 | 0.836736344 | 0.93675 | 5.555914929 | 213.3796021 | 213.3796021 | 0.193178115 | -0.714042077 | FALSE | TRUE | FALSE |
| TRIM42 | 0.050215 | 0.87562773 | 1 | 0.050214972 | 0.050214972 | 0.987732479 | 0.07078007 | 0.882255407 | 1 | 0.070780068 | 0.070780068 | 1.395014327 | 0.709450751 | -0.149077747 | FALSE | TRUE | FALSE |
| IL27 | 0.050197 | 0.87563706 | 0.00665 | 0.050197041 | 0.050197041 | 0.050197041 | 0.05260152 | 0.886104331 | 0.006 | 0.052601519 | 0.052601519 | 0.052601519 | 0.954288825 | -0.020320162 | FALSE | TRUE | FALSE |
| MMP13 | 0.0501457 | 0.87564639 | 1 | 0.444391849 | 0.444391849 | 0.99378387 | 0.28794005 | 0.828723871 | 0.50585 | 0.287940046 | 0.287940046 | 1.428288934 | 0.174153417 | -0.759067999 | FALSE | TRUE | FALSE |
| EXOSC9 | 0.0501072 | 0.87565571 | 0.50315 | 0.050107194 | 0.050107194 | 0.118504314 | 0.01090268 | 0.896499196 | 1 | 0.01090268 | 0.01090268 | 0.216437214 | 4.595860331 | 0.662366822 | FALSE | TRUE | FALSE |
| GZMB | 0.0501072 | 0.87566503 | 0.00795 | 0.050107162 | 0.050107162 | 0.050107162 | 0.07034996 | 0.882387795 | 1 | 0.281280409 | 0.281280409 | 0.281280409 | 0.712255687 | -0.147364074 | FALSE | TRUE | FALSE |
| MRPS17 | 0.0500245 | 0.87567436 | 0.004 | 0.05002451 | 0.05002451 | 0.05002451 | 0.05344737 | 0.885971065 | 1 | 0.053447366 | 0.053447366 | 0.053447366 | 0.935958378 | -0.028743464 | FALSE | TRUE | FALSE |
| FDXR | 0.050016 | 0.87568368 | 1 | 0.050016044 | 0.050016044 | 0.98754856 | 210.527433 | 0.01276666 | 0.06715 | 5.479078274 | 210.5274333 | 210.5274333 | 0.000237575 | -3.624199353 | FALSE | FALSE | FALSE |
| ARFGAP3 | 0.0499902 | 0.875693 | 1 | 0.445885618 | 0.445885618 | 0.990313843 | 5.60311944 | 0.282502909 | 0.2563 | 2.432631894 | 22.40906309 | 22.40906309 | 0.008921845 | -2.049545333 | FALSE | FALSE | FALSE |
| LIG1 | 0.049989 | 0.87570231 | 0.75495 | 0.049989016 | 0.19988474 | 0.977567208 | 0.0213303 | 0.893446251 | 1 | 0.021330296 | 0.021330296 | 0.418244051 | 2.343568731 | 0.369877695 | FALSE | TRUE | FALSE |
| SLC52A2 | 0.0499737 | 0.87571163 | 1 | 0.049973725 | 0.049973725 | 0.9889432 | 0.07118143 | 0.88218469 | 1 | 0.071181431 | 0.071181431 | 1.410413584 | 0.702061256 | -0.153624993 | FALSE | TRUE | FALSE |

| | | | | | | | | | | | | | | | | | |
|---|---|---|---|---|---|---|---|---|---|---|---|---|---|---|---|---|---|
| SRPK1 | 0.0499508 | 0.87572095 | 1 | 0.049950803 | 0.049950803 | 0.987633493 | 0.07033027 | 0.882405425 | 1 | 0.070330273 | 0.070330273 | 1.392485085 | 0.710231893 | -0.14859983 | FALSE | TRUE | FALSE |
| FABP4 | 0.0498818 | 0.87573026 | 0.00385 | 0.049881806 | 0.049881806 | 0.049881806 | 0.0548213 | 0.885686967 | 1 | 0.054821295 | 0.054821295 | 0.054821295 | 0.909898338 | -0.041007128 | FALSE | TRUE | FALSE |
| NXPE1 | 0.0498589 | 0.87573957 | 1 | 3.999619513 | 37.2251618 | 37.2251618 | 0.07680382 | 0.881105708 | 1 | 0.076803819 | 0.076803819 | 0.076803819 | 0.649171751 | -0.187640387 | FALSE | TRUE | FALSE |
| SLC26A6 | 0.0498039 | 0.87574888 | 0.1979 | 0.006910103 | 0.110225712 | 0.440322921 | 0.00248247 | 0.898868563 | 0.50525 | 0.002482469 | 0.002482469 | 0.009643517 | 20.06224155 | 1.302379455 | FALSE | TRUE | FALSE |
| PEX5L | 0.0497505 | 0.87575819 | 1 | 0.049750459 | 0.049750459 | 0.983136876 | 0.07088175 | 0.882237735 | 1 | 0.070881754 | 0.070881754 | 1.402730941 | 0.701879629 | -0.153737362 | FALSE | TRUE | FALSE |
| CCHCR1 | 0.0496816 | 0.8757675 | 0.88425 | 0.42480032 | 3.725789379 | 8.330133296 | 0.01147017 | 0.896260908 | 0.02365 | 0.011470169 | 0.011470169 | 0.225453424 | 4.331376624 | 0.636625948 | FALSE | TRUE | FALSE |
| NAGPA | 0.0496677 | 0.87577681 | 1 | 0.049667661 | 0.049667661 | 0.979786987 | 0.07023999 | 0.88243186 | 1 | 0.070239988 | 0.070239988 | 1.387891679 | 0.707113742 | -0.150510723 | FALSE | TRUE | FALSE |
| PRKRA | 0.0496419 | 0.87578612 | 1 | 4.9164E+167 | 5.8761E+187 | 1.4844E+197 | 0.16300698 | 0.861356839 | 1 | 4.7453E+163 | 2.1601E+185 | 1.5317E+193 | 0.304538457 | -0.516357857 | FALSE | TRUE | FALSE |
| SBSN | 0.0495936 | 0.87579542 | 1 | 38.21914426 | 3639.749808 | 36809.20448 | 0.06870905 | 0.882721958 | 0.93675 | 5.727837813 | 528.4949707 | 528.4949707 | 0.721791906 | -0.141587992 | FALSE | TRUE | FALSE |
| ZNF264 | 0.0495926 | 0.87580473 | 1 | 0.049592616 | 0.049592616 | 0.979975682 | 0.60964784 | 0.707767113 | 0.5131 | 0.609647837 | 0.609647837 | 1.346252795 | 0.081346333 | -1.089662022 | FALSE | TRUE | FALSE |
| SERPINA5 | 0.0495129 | 0.87581403 | 0.7602 | 0.19792532 | 0.791394145 | 0.793298224 | 0.28531835 | 0.829479565 | 0.50645 | 0.285318351 | 0.285318351 | 0.285318351 | 0.173535661 | -0.760611265 | FALSE | TRUE | FALSE |
| HIBCH | 0.0494804 | 0.87582333 | 1 | 0.049480401 | 0.049480401 | 0.049480401 | 0.07025178 | 0.88242305 | 1 | 0.070251783 | 0.070251783 | 0.070251783 | 0.704329476 | -0.152224136 | FALSE | TRUE | FALSE |
| ARAP1 | 0.0494443 | 0.87583263 | 1 | 0.049444276 | 0.049444276 | 1.663565462 | 0.07027978 | 0.882414238 | 1 | 0.070279779 | 0.070279779 | 1.361675163 | 0.703534883 | -0.152714364 | FALSE | TRUE | FALSE |
| SLTM | 0.0494342 | 0.87584193 | 1 | 0.049434164 | 0.049434164 | 1.661121394 | 0.62492392 | 0.701105676 | 0.51945 | 0.62492392 | 0.62492392 | 12.25068107 | 0.079104292 | -1.101799953 | FALSE | TRUE | FALSE |
| NT5C3A | 0.0494287 | 0.87585123 | 1 | 0.049428741 | 0.049428741 | 0.049428741 | 0.07037451 | 0.88236134 | 0.00595 | 0.070374513 | 0.070374513 | 0.070374513 | 0.702367065 | -0.153435861 | FALSE | TRUE | FALSE |
| UBAP2 | 0.0493719 | 0.87586052 | 0.0295 | 0.049371889 | 0.049371889 | 0.966351267 | 0.06975912 | 0.8825287 | 1 | 0.069759121 | 0.069759121 | 1.368900744 | 0.707748153 | -0.150121255 | FALSE | TRUE | FALSE |
| LGR4 | 0.0493299 | 0.87586982 | 1 | 16716.08097 | 7409889.121 | 145947950 | 4.2148086 | 0.342487569 | 0.99885 | 701244.2252 | 646642483.8 | 14407161466 | 0.011703939 | -1.93166794 | FALSE | FALSE | FALSE |
| RIN2 | 0.0493192 | 0.87587911 | 1 | 7.77126E+16 | 3.48395E+22 | 7.15429E+24 | 0.08335757 | 0.879503836 | 1 | 1.06725E+14 | 4.47866E+18 | 5.73271E+20 | 0.591658141 | -0.227929155 | FALSE | TRUE | FALSE |
| SLC25A40 | 0.0491645 | 0.8758884 | 0.0078 | 0.04916454 | 0.04916454 | 0.04916454 | 0.27291957 | 0.832971665 | 0.75215 | 0.272919569 | 1.091600401 | 1.091600401 | 0.180142965 | -0.744382694 | FALSE | TRUE | FALSE |
| LEPR | 0.0491406 | 0.8758977 | 0.5036 | 0.441521446 | 0.441521446 | 1.660006129 | 0.07182781 | 0.88206075 | 0.01775 | 0.071827812 | 0.071827812 | 1.418619513 | 0.684143877 | -0.164852556 | FALSE | TRUE | FALSE |
| TLR9 | 0.0491392 | 0.87590699 | 1 | 0.443881474 | 0.443881474 | 8.640232278 | 0.07191733 | 0.882043026 | 0.02985 | 0.071917328 | 0.071917328 | 1.404395677 | 0.683273753 | -0.165405262 | FALSE | TRUE | FALSE |
| PRRC2B | 0.0491359 | 0.87591627 | 1 | 0.04913587 | 0.931540593 | 1.638125534 | 0.06983965 | 0.882502304 | 1 | 0.06983965 | 1.332388871 | 2.333816213 | 0.703552636 | -0.152703405 | FALSE | TRUE | FALSE |
| RASSF7 | 0.0491078 | 0.87592556 | 1 | 3.981710185 | 37.25685372 | 37.25685372 | 0.07595074 | 0.881267326 | 0.0063 | 0.075950742 | 0.075950742 | 0.075950742 | 0.646574259 | -0.189381589 | FALSE | TRUE | FALSE |
| F11 | 0.0491042 | 0.87593485 | 0.8149 | 0.436299483 | 9.296248777 | 37.14852423 | 0.0504594 | 0.886519045 | 0.8963 | 0.442484051 | 7.076125585 | 28.30787185 | 0.973142336 | -0.011823633 | FALSE | TRUE | FALSE |
| HOXB2 | 0.049101 | 0.87594413 | 0.8779 | 0.785131742 | 7.163846572 | 7.163846572 | 0.27402167 | 0.832558025 | 0.87715 | 2.509535085 | 23.58829341 | 23.58829341 | 0.179186507 | -0.746694696 | FALSE | TRUE | FALSE |
| MUS81 | 0.0490148 | 0.87595342 | 1 | 0.049014784 | 0.049014784 | 0.967227911 | 0.60162463 | 0.711647761 | 0.0153 | 0.601624633 | 0.601624633 | 2.277455224 | 0.081470706 | -1.08899852 | FALSE | TRUE | FALSE |
| XRN1 | 0.048977 | 0.8759627 | 1 | 0.04897703 | 0.04897703 | 1.643088517 | 0.27564933 | 0.832159685 | 0.52045 | 0.275649328 | 0.275649328 | 5.375768413 | 0.177678757 | -0.750364493 | FALSE | TRUE | FALSE |
| TLR4 | 0.0489409 | 0.87597198 | 1 | 0.431926782 | 0.431926782 | 1.653904632 | 0.07117063 | 0.882193533 | 1 | 0.071170634 | 0.071170634 | 1.407674665 | 0.687655569 | -0.162629036 | FALSE | TRUE | FALSE |
| DENND1B | 0.0489326 | 0.87598126 | 1 | 0.432543893 | 0.432543893 | 1.655525675 | 0.07119464 | 0.882175845 | 1 | 0.071194639 | 0.071194639 | 1.408834159 | 0.687307282 | -0.162849055 | FALSE | TRUE | FALSE |
| CCP110 | 0.0489157 | 0.87599054 | 0.02455 | 0.048915702 | 0.048915702 | 0.961932121 | 0.06285693 | 0.88405008 | 0.0225 | 0.062856934 | 0.062856934 | 1.23857963 | 0.778206939 | -0.108904901 | FALSE | TRUE | FALSE |
| CLYBL | 0.0488968 | 0.87599981 | 1 | 0.048896801 | 0.432912675 | 0.972141597 | 0.06511517 | 0.883558511 | 0.87385 | 5.303921408 | 49.74225843 | 49.74225843 | 0.750928006 | -0.124401698 | FALSE | TRUE | FALSE |
| ITGA8 | 0.048889 | 0.87600909 | 0.50985 | 0.048889021 | 0.048889021 | 0.957769291 | 0.08171263 | 0.879880584 | 0.02645 | 0.081712634 | 0.081712634 | 1.604752143 | 0.598304307 | -0.223077871 | FALSE | TRUE | FALSE |
| ZNF497 | 0.0488883 | 0.87601836 | 1 | 4.034901827 | 38.09068609 | 38.50336104 | 0.64554229 | 0.692175783 | 0.75655 | 0.645542295 | 5.985457785 | 12.72186047 | 0.075732162 | -1.120719642 | FALSE | TRUE | FALSE |
| MBD3 | 0.0488882 | 0.87602764 | 1 | 0.048888175 | 0.048888175 | 0.968218491 | 0.06990745 | 0.882467091 | 1 | 0.069907446 | 0.069907446 | 0.069907446 | 0.699327157 | -0.155319607 | FALSE | TRUE | FALSE |
| BCL9L | 0.0486807 | 0.87603691 | 1 | 0.0486807 | 0.0486807 | 1.638492496 | 0.06957621 | 0.882555086 | 1 | 0.069576206 | 0.069576206 | 2.344626288 | 0.699674544 | -0.155103927 | FALSE | TRUE | FALSE |
| RAPGEF2 | 0.0486294 | 0.87604618 | 1 | 435360.7082 | 91526154.06 | 5368374079 | 0.07366896 | 0.881651851 | 1 | 64731.96089 | 6239897.765 | 69515673.37 | 0.660107342 | -0.180385437 | FALSE | TRUE | FALSE |
| KYNU | 0.0485903 | 0.87605545 | 0.97025 | 0.777255299 | 3.109806405 | 12.44547554 | 0.02138375 | 0.893424118 | 0.01105 | 0.021383749 | 0.021383749 | 0.424558252 | 2.272302125 | 0.356466075 | FALSE | TRUE | FALSE |
| MYOD1 | 0.0484845 | 0.87606472 | 0.51115 | 0.193776111 | 0.193776111 | 0.956791247 | 0.06863328 | 0.882730729 | 1 | 0.274330065 | 1.096782805 | 5.422287206 | 0.706428054 | -0.150932062 | FALSE | TRUE | FALSE |
| CAMSAP1 | 0.0484429 | 0.87607398 | 1 | 0.048442864 | 0.048442864 | 1.628810625 | 0.06504355 | 0.883593129 | 0.7542 | 0.601209384 | 5.703465139 | 25.76777431 | 0.744775875 | -0.1279744 | FALSE | TRUE | FALSE |
| KIFC2 | 0.0484395 | 0.87608325 | 0.02605 | 0.048439456 | 0.048439456 | 0.948797042 | 0.06872649 | 0.882713186 | 0.0241 | 0.068726494 | 0.068726494 | 1.349510735 | 0.704814891 | -0.151924929 | FALSE | TRUE | FALSE |
| ATP1A1 | 0.0484359 | 0.87609251 | 1 | 0.453407424 | 0.453407424 | 8.877146099 | 0.07151323 | 0.882113896 | 1 | 0.071513225 | 0.071513225 | 1.40369104 | 0.677299831 | -0.169219033 | FALSE | TRUE | FALSE |
| IGSF9B | 0.0483784 | 0.87610178 | 1 | 0.048378425 | 0.048378425 | 0.933249999 | 0.06915618 | 0.882607828 | 1 | 0.069156183 | 0.069156183 | 2.329673983 | 0.699553139 | -0.15517929 | FALSE | TRUE | FALSE |
| TICRR | 0.0482955 | 0.87611104 | 0.0523 | 0.048295507 | 0.109877242 | 0.930224989 | 0.01051328 | 0.896617955 | 0.04685 | 0.010513278 | 0.010513278 | 0.352690897 | 4.593762971 | 0.662168583 | FALSE | TRUE | FALSE |
| ZFP69B | 0.0482606 | 0.8761203 | 1 | 0.048260642 | 0.048260642 | 0.048260642 | 0.06837143 | 0.882800852 | 1 | 0.068371429 | 0.068371429 | 1.359068716 | 0.705859772 | -0.151281568 | FALSE | TRUE | FALSE |
| BRD8 | 0.0482342 | 0.87612956 | 0.5209 | 0.429249192 | 0.429249192 | 8.398879414 | 0.07045728 | 0.882326049 | 1 | 0.070457277 | 0.070457277 | 1.382198125 | 0.684588375 | -0.16457048 | FALSE | TRUE | FALSE |
| FBXO7 | 0.0482206 | 0.87613882 | 1 | 0.43445049 | 0.43445049 | 0.955087171 | 5.60140095 | 0.282808087 | 0.2526 | 0.602653878 | 5.601400947 | 5.601400947 | 0.00860866 | -2.06506442 | FALSE | FALSE | FALSE |
| CKAP2L | 0.0480215 | 0.87614808 | 1 | 0.048021488 | 0.048021488 | 0.947855373 | 0.58586361 | 0.718284659 | 0.5131 | 0.585863612 | 0.585863612 | 2.231474156 | 0.081967008 | -1.086360916 | FALSE | TRUE | FALSE |
| POU2F1 | 0.0480091 | 0.87615733 | 1 | 0.191903142 | 0.76727114 | 3.783658697 | 0.0703311 | 0.88239661 | 1 | 0.070331099 | 0.070331099 | 1.389048857 | 0.682616004 | -0.165823534 | FALSE | TRUE | FALSE |
| MTFR1 | 0.0480022 | 0.87616659 | 1 | 0.426281307 | 0.426281307 | 0.426281307 | 0.06803901 | 0.882862146 | 0.5099 | 0.608819156 | 0.608819156 | 0.608819156 | 0.70550959 | -0.151497079 | FALSE | TRUE | FALSE |
| RIN1 | 0.0479976 | 0.87617584 | 1 | 0.42276987 | 0.42276987 | 1.62418444 | 0.06990257 | 0.8824847 | 1 | 0.069902573 | 0.069902573 | 1.381848449 | 0.68663607 | -0.163273386 | FALSE | TRUE | FALSE |
| CD101 | 0.047993 | 0.87618509 | 0.7543 | 0.047993038 | 0.418727688 | 1.619475945 | 0.74859292 | 0.642060258 | 0.5054 | 0.748592922 | 6.776417625 | 14.71812734 | 0.064110996 | -1.193067477 | FALSE | TRUE | FALSE |
| CDKN2AIP | 0.047965 | 0.87619434 | 1 | 0.047964959 | 0.047964959 | 0.948589267 | 0.05181427 | 0.886253931 | 1 | 0.051814266 | 0.051814266 | 1.02609224 | 0.925709514 | -0.033525273 | FALSE | TRUE | FALSE |
| MRPL55 | 0.0479445 | 0.87620359 | 0.006 | 0.047944452 | 0.047944452 | 0.047944452 | 0.05247043 | 0.886154236 | 1 | 0.052470427 | 0.052470427 | 0.052470427 | 0.913742362 | -0.03917624 | FALSE | TRUE | FALSE |
| FAP | 0.0479399 | 0.87621284 | 0.7562 | 0.047939918 | 0.435986736 | 0.946061812 | 0.03997452 | 0.888958786 | 0.8785 | 0.159826472 | 1.428542927 | 1.76679759 | 1.199261952 | 0.078914055 | FALSE | TRUE | FALSE |
| GALNT10 | 0.047915 | 0.87622209 | 0.02145 | 0.047914968 | 0.047914968 | 0.944869758 | 0.06849977 | 0.8827658 | 0.0161 | 0.068499768 | 0.068499768 | 1.353074423 | 0.699490957 | -0.155217895 | FALSE | TRUE | FALSE |
| AFF3 | 0.0479031 | 0.87623133 | 0.03595 | 0.047903131 | 0.047903131 | 1.614017281 | 0.06861563 | 0.882739499 | 1 | 0.068615628 | 0.068615628 | 1.338657996 | 0.698137322 | -0.156059144 | FALSE | TRUE | FALSE |
| FUT11 | 0.0478846 | 0.87624058 | 1 | 3.85508102 | 143.7660305 | 143.7660305 | 0.63140368 | 0.69861891 | 0.7578 | 0.63140368 | 5.789944706 | 12.46806665 | 0.075838364 | -1.120111042 | FALSE | TRUE | FALSE |
| PGS1 | 0.0478415 | 0.87624982 | 0.5069 | 0.191324107 | 0.191324107 | 1.61969092 | 0.06676068 | 0.883167744 | 1 | 0.60193636 | 0.60193636 | 1.320390378 | 0.716612446 | -0.144715653 | FALSE | TRUE | FALSE |
| RBM19 | 0.0477354 | 0.87625906 | 0.513 | 0.047735385 | 0.047735385 | 0.933744045 | 0.01027932 | 0.896694667 | 1 | 0.041108767 | 0.041108767 | 0.806248865 | 4.643824986 | 0.666875844 | FALSE | TRUE | FALSE |
| TMEM126B | 0.0477352 | 0.8762683 | 0.0048 | 0.047735217 | 0.047735217 | 0.047735217 | 0.06536216 | 0.883497883 | 1 | 0.575107369 | 0.575107369 | 0.575107369 | 0.730318794 | -0.136487523 | FALSE | TRUE | FALSE |
| CCDC125 | 0.0477293 | 0.87627754 | 0.01285 | 0.047729307 | 0.047729307 | 0.946343367 | 0.58347462 | 0.719242505 | 0.5018 | 0.583474616 | 0.583474616 | 1.302299968 | 0.081801857 | -1.087236837 | FALSE | TRUE | FALSE |
| OLFM4 | 0.0477261 | 0.87628678 | 0.7557 | 0.107194514 | 0.42847007 | 0.42847007 | 0.08002604 | 0.880337072 | 0.7578 | 0.180878008 | 0.723053581 | 0.723053581 | 0.596381565 | -0.22447579 | FALSE | TRUE | FALSE |
| STT3A | 0.0476976 | 0.87629602 | 1 | 0.04769762 | 0.04769762 | 0.939934869 | 0.06829466 | 0.882827128 | 0.0195 | 0.068294656 | 0.068294656 | 1.348202623 | 0.698409253 | -0.155890016 | FALSE | TRUE | FALSE |
| PARP10 | 0.0476818 | 0.87630525 | 1 | 0.047681773 | 0.047681773 | 0.928525138 | 0.58843194 | 0.717472773 | 0.51575 | 0.588431943 | 0.588431943 | 11.49512238 | 0.081031925 | -1.091343846 | FALSE | TRUE | FALSE |
| NEK8 | 0.0476508 | 0.87631448 | 0.89445 | 1.715957935 | 63.39850858 | 253.5519415 | 0.01084448 | 0.896520171 | 1 | 0.173351259 | 0.693344355 | 3.419524358 | 4.394012673 | 0.642861305 | FALSE | TRUE | FALSE |
| FLVCR2 | 0.0476496 | 0.87632372 | 1 | 0.190528474 | 0.190528474 | 0.942238828 | 0.06873047 | 0.882704413 | 1 | 0.068730475 | 0.068730475 | 1.360943346 | 0.693282331 | -0.159089868 | FALSE | TRUE | FALSE |
| PRMT3 | 0.0475762 | 0.87633295 | 1 | 0.047576183 | 0.047576183 | 0.942240015 | 0.0676188 | 0.882975824 | 1 | 0.067618795 | 0.067618795 | 1.340752936 | 0.703594065 | -0.152677833 | FALSE | TRUE | FALSE |
| CTNND2 | 0.0475527 | 0.87634218 | 0.8816 | 0.190048015 | 0.759731859 | 3.696123172 | 0.02080948 | 0.893623014 | 1 | 0.020809479 | 0.020809479 | 0.701102481 | 2.285144493 | 0.358913666 | FALSE | TRUE | FALSE |
| FKBP10 | 0.0475218 | 0.87635141 | 0.98405 | 16.05183938 | 1027.162657 | 4110.496696 | 0.08881269 | 0.878370028 | 0.8796 | 0.35509414 | 1.420105262 | 3.008951133 | 0.535079368 | -0.271581795 | FALSE | TRUE | FALSE |
| PIWIL1 | 0.0474973 | 0.87636063 | 0.0256 | 0.047497255 | 0.047497255 | 0.933123786 | 0.58404808 | 0.718991098 | 0.5116 | 0.584048082 | 0.584048082 | 11.498582 | 0.081324221 | -1.089780089 | FALSE | TRUE | FALSE |
| SLCO2B1 | 0.0474934 | 0.87636986 | 0.02235 | 0.047493395 | 0.047493395 | 0.934602892 | 0.06791211 | 0.882888397 | 1 | 0.067912109 | 0.067912109 | 1.338999674 | 0.69933618 | -0.155314003 | FALSE | TRUE | FALSE |
| MAPK8IP3 | 0.0474923 | 0.87637908 | 1 | 0.047492291 | 0.912005491 | 1.599311393 | 0.06793306 | 0.882879648 | 1 | 0.067933065 | 0.067933065 | 1.310808427 | 0.699104199 | -0.155458089 | FALSE | TRUE | FALSE |
| ERICH1 | 0.0474919 | 0.87638831 | 1 | 3.913244493 | 36.91202799 | 36.91202799 | 0.06787681 | 0.882923382 | 1 | 0.067876805 | 0.067876805 | 1.345105273 | 0.699677893 | -0.155101848 | FALSE | TRUE | FALSE |

| | | | | | | | | | | | | | | | | | |
|---|---|---|---|---|---|---|---|---|---|---|---|---|---|---|---|---|---|
| TMEM132B | 0.0474897 | 0.87639753 | 0.02905 | 0.047489662 | 0.047489662 | 0.929042645 | 0.05312138 | 0.886046059 | 1 | 0.05312138 | 0.05312138 | 1.041951192 | 0.893983973 | -0.048670267 | FALSE | TRUE | FALSE |
| EME1 | 0.0474059 | 0.87640675 | 1 | 0.047405905 | 0.047405905 | 0.938380771 | 0.06713734 | 0.883071856 | 1 | 0.06713734 | 0.06713734 | 1.330598386 | 0.706103423 | -0.151131683 | FALSE | TRUE | FALSE |
| PHF3 | 0.0473652 | 0.87641597 | 1 | 0.430724469 | 4.020125461 | 77.71884858 | 0.07189289 | 0.882051888 | 1 | 0.071892894 | 0.071892894 | 2.410270177 | 0.658829518 | -0.181226951 | FALSE | TRUE | FALSE |
| MATR3 | 0.047352 | 0.87642519 | 1 | 0.04735202 | 0.04735202 | 0.932934462 | 0.06760324 | 0.88298456 | 1 | 0.067603238 | 0.067603238 | 1.334313864 | 0.70044012 | -0.154628986 | FALSE | TRUE | FALSE |
| BUB1 | 0.0472909 | 0.8764344 | 1 | 0.424774709 | 3.916015058 | 14.32859828 | 0.62816378 | 0.699896606 | 0.02265 | 0.628163775 | 0.628163775 | 12.36358997 | 0.075284295 | -1.12329561 | FALSE | TRUE | FALSE |
| VIPR2 | 0.0472098 | 0.87644362 | 1 | 0.047209756 | 0.047209756 | 0.935287899 | 0.06706412 | 0.883098022 | 1 | 0.067064117 | 0.067064117 | 1.330136721 | 0.70394956 | -0.152458458 | FALSE | TRUE | FALSE |
| MPP7 | 0.0470794 | 0.87645284 | 1 | 0.047079437 | 0.047079437 | 0.931571852 | 0.067081 | 0.883089301 | 1 | 0.067081004 | 0.067081004 | 1.329046228 | 0.701829649 | -0.153768289 | FALSE | TRUE | FALSE |
| UBTF | 0.0470657 | 0.87646205 | 1 | 0.047065704 | 0.047065704 | 0.924769148 | 0.06762508 | 0.882967087 | 1 | 0.067625077 | 0.067625077 | 1.331541249 | 0.695980035 | -0.157403218 | FALSE | TRUE | FALSE |
| POC1B | 0.047025 | 0.87647126 | 0.01215 | 0.04702498 | 0.04702498 | 0.932437627 | 0.58362283 | 0.719142 | 0.509 | 0.583622828 | 0.583622828 | 0.583622828 | 0.080574265 | -1.093803649 | FALSE | TRUE | FALSE |
| KRT222 | 0.0470134 | 0.87648047 | 1 | 0.431892824 | 0.431892824 | 0.431892824 | 0.61395708 | 0.705951233 | 0.5016 | 0.613957084 | 2.454588068 | 2.454588068 | 0.076574365 | -1.115916597 | FALSE | TRUE | FALSE |
| NDUFAF1 | 0.0469876 | 0.87648968 | 0.01 | 0.046987578 | 0.046987578 | 0.055853423 | 0.06726153 | 0.883036952 | 0.0069 | 0.067261525 | 0.067261525 | 0.067261525 | 0.698580325 | -0.15578365 | FALSE | TRUE | FALSE |
| SELE | 0.0469814 | 0.87649889 | 1 | 0.046981394 | 0.046981394 | 0.929566658 | 0.06691097 | 0.883132892 | 1 | 0.066910972 | 0.066910972 | 1.325595566 | 0.702147836 | -0.153571439 | FALSE | TRUE | FALSE |
| WDR91 | 0.0469706 | 0.8765081 | 0.0224 | 0.04697064 | 0.04697064 | 0.923091264 | 0.06716014 | 0.883063132 | 1 | 0.067160141 | 0.067160141 | 1.322626631 | 0.699382687 | -0.155285122 | FALSE | TRUE | FALSE |
| RUFY1 | 0.0469419 | 0.8765173 | 0.01875 | 0.046941884 | 0.046941884 | 0.927823566 | 0.06492795 | 0.88361908 | 1 | 0.585607091 | 0.585607091 | 1.28513605 | 0.72298429 | -0.14087114 | FALSE | TRUE | FALSE |
| FCHSD1 | 0.0469412 | 0.87652651 | 0.7544 | 0.10007496 | 0.400268762 | 1.970533158 | 0.16417121 | 0.861081029 | 0.7558 | 0.164171213 | 1.452925833 | 3.238645038 | 0.285928237 | -0.543742954 | FALSE | TRUE | FALSE |
| GPC2 | 0.0469207 | 0.87653571 | 1 | 4.7813E+22 | 4.73004E+29 | 1.14885E+32 | 0.09820249 | 0.876401326 | 1 | 3.34242E+16 | 1.57005E+22 | 3.28627E+24 | 0.477795611 | -0.320757844 | FALSE | TRUE | FALSE |
| SPATA20 | 0.0469174 | 0.87654491 | 0.8839 | 0.931275067 | 34.47225926 | 34.47225926 | 0.01959833 | 0.893923738 | 1 | 1.582600054 | 59.09240647 | 59.09240647 | 2.393950836 | 0.379115227 | FALSE | TRUE | FALSE |
| PARM1 | 0.0469169 | 0.87655411 | 0.00895 | 0.046916936 | 0.046916936 | 0.046916936 | 0.05072042 | 0.886469422 | 0.0074 | 0.050720416 | 0.050720416 | 0.050720416 | 0.925010863 | -0.033853167 | FALSE | TRUE | FALSE |
| NUP188 | 0.0469083 | 0.87656331 | 0.51895 | 0.046908268 | 0.046908268 | 0.905596726 | 0.0828812 | 0.879595926 | 0.2755 | 0.082881195 | 0.727466288 | 14.10306404 | 0.565969982 | -0.247206602 | FALSE | TRUE | FALSE |
| DHX36 | 0.0468687 | 0.87657251 | 1 | 0.432129121 | 0.432129121 | 8.483030655 | 0.62172764 | 0.703041446 | 0.51415 | 0.621727643 | 0.621727643 | 12.23212795 | 0.075384649 | -1.122717084 | FALSE | TRUE | FALSE |
| IFT57 | 0.0468678 | 0.8765817 | 0.01045 | 0.046867755 | 0.046867755 | 0.928959404 | 0.06607407 | 0.883341716 | 0.5061 | 0.264231499 | 0.264231499 | 1.31105608 | 0.709321407 | -0.149156933 | FALSE | TRUE | FALSE |
| RNF32 | 0.0468177 | 0.8765909 | 0.01025 | 0.046817719 | 0.046817719 | 0.929481985 | 0.56134648 | 0.728504937 | 0.75585 | 0.561346482 | 5.154723695 | 5.154723695 | 0.083402534 | -1.078820755 | FALSE | TRUE | FALSE |
| AFAP1L1 | 0.0468164 | 0.87660009 | 1 | 0.046816363 | 0.046816363 | 0.919529812 | 0.06297831 | 0.884024301 | 1 | 0.57294915 | 5.349810893 | 11.27771965 | 0.743372847 | -0.128793306 | FALSE | TRUE | FALSE |
| HTR1D | 0.046808 | 0.87660929 | 1 | 0.187126375 | 0.187126375 | 0.928165215 | 5.56110989 | 0.28523892 | 0.2611 | 0.587914508 | 5.561109892 | 5.561109892 | 0.008417025 | -2.074841375 | FALSE | FALSE | FALSE |
| ESRP2 | 0.0467979 | 0.87661848 | 1 | 0.046797905 | 0.046797905 | 0.91850384 | 0.57443143 | 0.722713171 | 0.50805 | 0.574431434 | 0.574431434 | 11.29969962 | 0.081468218 | -1.089011784 | FALSE | TRUE | FALSE |
| PCDH10 | 0.0467906 | 0.87662767 | 1 | 6.47614E+19 | 3.83297E+25 | 8.74766E+27 | 0.4453084 | 0.775666946 | 1 | 1.65264E+25 | 1.48951E+31 | 6.67138E+34 | 0.105074529 | -0.97850255 | FALSE | TRUE | FALSE |
| BOD1 | 0.0467756 | 0.87663685 | 1 | 0.046775599 | 0.046775599 | 0.046775599 | 0.04968773 | 0.886717149 | 1 | 0.049687732 | 0.049687732 | 0.049687732 | 0.94139132 | -0.02622981 | FALSE | TRUE | FALSE |
| NIPAL1 | 0.0467386 | 0.87664604 | 0.2538 | 0.046738585 | 0.186848833 | 0.186848833 | 0.07954136 | 0.880491534 | 0.0076 | 0.079541358 | 0.079541358 | 0.079541358 | 0.587601047 | -0.230917439 | FALSE | TRUE | FALSE |
| PCDHGA11 | 0.0467384 | 0.87665523 | 0.76 | 0.046738443 | 0.418874855 | 1.578200636 | 0.0103784 | 0.896659811 | 1 | 0.010378397 | 0.010378397 | 0.203120537 | 4.503435524 | 0.653543949 | FALSE | TRUE | FALSE |
| NEU1 | 0.0467306 | 0.87666441 | 1 | 0.186825467 | 0.747100376 | 1.582988788 | 0.27078978 | 0.833588295 | 0.01205 | 0.270789783 | 0.270789783 | 1.341900107 | 0.172571664 | -0.763030513 | FALSE | TRUE | FALSE |
| SUMF1 | 0.0466675 | 0.87667359 | 0.75555 | 0.04666751 | 0.18653933 | 0.231751166 | 0.07964492 | 0.880464304 | 0.00875 | 0.079644921 | 0.079644921 | 0.079644921 | 0.585944578 | -0.23214346 | FALSE | TRUE | FALSE |
| ZNF300 | 0.0466112 | 0.87668278 | 1 | 0.04661117 | 0.04661117 | 0.923274613 | 5.1451888 | 0.301884585 | 0.009 | 0.559789895 | 5.145188799 | 5.145188799 | 0.009059176 | -2.042911308 | FALSE | FALSE | FALSE |
| PMPCB | 0.0465784 | 0.87669196 | 1 | 0.046578355 | 0.046578355 | 0.922258297 | 0.06640259 | 0.883237389 | 0.01285 | 0.066402593 | 0.066402593 | 1.316361509 | 0.701453854 | -0.154000894 | FALSE | TRUE | FALSE |
| TP53 | 0.0465548 | 0.87670114 | 0.01085 | 0.046554802 | 0.046554802 | 0.231128423 | 0.0199615 | 0.893872504 | 0.01115 | 0.019961499 | 0.019961499 | 0.396308553 | 2.332229821 | 0.367771344 | FALSE | TRUE | FALSE |
| VAC14 | 0.0465374 | 0.87671031 | 1 | 0.426384195 | 0.426384195 | 8.373154828 | 0.06878486 | 0.882695639 | 1 | 0.068784859 | 0.068784859 | 1.353715546 | 0.676564041 | -0.169691088 | FALSE | TRUE | FALSE |
| LRIG3 | 0.0465325 | 0.87671949 | 1 | 0.046532502 | 0.046532502 | 0.912799769 | 0.06442505 | 0.883714147 | 1 | 0.580758749 | 0.580758749 | 11.41870438 | 0.722273428 | -0.141298362 | FALSE | TRUE | FALSE |
| SLC6A4 | 0.0465009 | 0.87672867 | 0.0189 | 0.046500914 | 0.046500914 | 0.91688688 | 0.06681802 | 0.88315032 | 1 | 0.066818018 | 0.066818018 | 1.31973037 | 0.695933756 | -0.157432098 | FALSE | TRUE | FALSE |
| PDHB | 0.0464692 | 0.87673784 | 0.0092 | 0.046469228 | 0.046469228 | 0.046469228 | 0.26407906 | 0.835494801 | 0.50805 | 0.264079063 | 0.264079063 | 0.264079063 | 0.175967104 | -0.754568512 | FALSE | TRUE | FALSE |
| PLEKHA7 | 0.0464555 | 0.87674701 | 1 | 0.046455477 | 0.046455477 | 1.562369608 | 0.0663154 | 0.883263487 | 1 | 0.066315399 | 0.066315399 | 1.296129885 | 0.70052322 | -0.154577465 | FALSE | TRUE | FALSE |
| ZNF470 | 0.0464369 | 0.87675618 | 0.8741 | 0.185742678 | 0.743137119 | 0.917570094 | 1.42827431 | 0.543165702 | 0.50435 | 1.428274308 | 13.11051247 | 13.11051247 | 0.032512575 | -1.48794864 | FALSE | FALSE | FALSE |
| UPK1A | 0.0463827 | 0.87676535 | 1 | 0.04638273 | 0.04638273 | 0.04638273 | 0.06565373 | 0.883445868 | 1 | 0.262511446 | 0.262511446 | 0.262511446 | 0.706475187 | -0.150903087 | FALSE | TRUE | FALSE |
| SLC35B3 | 0.0463773 | 0.87677452 | 1 | 0.405939446 | 0.405939446 | 0.405939446 | 0.06765791 | 0.882940867 | 1 | 0.067657907 | 0.067657907 | 0.067657907 | 0.685467369 | -0.164013215 | FALSE | TRUE | FALSE |
| MYLK4 | 0.0463639 | 0.87678369 | 0.01075 | 0.046363886 | 0.046363886 | 0.919584843 | 0.54107811 | 0.737669497 | 0.8778 | 4.973436862 | 46.88891681 | 46.88891681 | 0.085687972 | -1.067080135 | FALSE | TRUE | FALSE |
| SLC4A2 | 0.0463455 | 0.87679286 | 0.94075 | 2.95734154 | 137.0480597 | 1108.063177 | 0.04088774 | 0.888759768 | 0.93925 | 0.163413471 | 2.612178402 | 12.70642212 | 1.133480809 | 0.054414172 | FALSE | TRUE | FALSE |
| SPTBN1 | 0.0461993 | 0.87680202 | 0.53445 | 0.046199311 | 0.218738849 | 1.542187455 | 0.17457725 | 0.85843886 | 0.64685 | 0.299594937 | 2.790532522 | 23.39922537 | 0.264635345 | -0.577352152 | FALSE | TRUE | FALSE |
| DEPDC7 | 0.046188 | 0.87681119 | 0.5114 | 0.412421566 | 0.412421566 | 0.916040758 | 0.06787958 | 0.882914638 | 1 | 0.067879583 | 0.067879583 | 1.347599417 | 0.680440649 | -0.16720975 | FALSE | TRUE | FALSE |
| CELA2A | 0.0461391 | 0.87682035 | 0.50975 | 0.18448094 | 0.737806394 | 0.737806394 | 0.0783383 | 0.880754168 | 0.25575 | 0.078338296 | 0.313250551 | 0.313250551 | 0.588972484 | -0.229904994 | FALSE | TRUE | FALSE |
| MTUS1 | 0.0461186 | 0.87682951 | 0.941 | 3.442656617 | 283.1477133 | 2659.204092 | 0.90570287 | 0.597712689 | 0.6469 | 8.053753331 | 684.403004 | 6533.284612 | 0.050920195 | -1.29310994 | FALSE | TRUE | FALSE |
| LRRC17 | 0.0461062 | 0.87683867 | 0.0087 | 0.046106213 | 0.046106213 | 0.046106213 | 0.06587633 | 0.883402492 | 1 | 0.06587633 | 0.06587633 | 0.06587633 | 0.699890434 | -0.154969942 | FALSE | TRUE | FALSE |
| KNOP1 | 0.0461041 | 0.87684783 | 1 | 0.046104147 | 0.046104147 | 0.913609621 | 0.0653796 | 0.883489217 | 0.0107 | 0.065379596 | 0.065379596 | 1.297007998 | 0.705176379 | -0.151702244 | FALSE | TRUE | FALSE |
| TUBGCP5 | 0.0460392 | 0.87685699 | 0.02965 | 0.046039206 | 0.046039206 | 1.551900321 | 0.06176978 | 0.884384215 | 1 | 0.55042609 | 5.034151385 | 10.81659152 | 0.745335442 | -0.127648227 | FALSE | TRUE | FALSE |
| TLL1 | 0.0460097 | 0.87686614 | 1 | 0.046009743 | 0.046009743 | 0.902392377 | 0.06028391 | 0.884691085 | 1 | 5.280026272 | 51.34585026 | 103.7987998 | 0.763217598 | -0.117351624 | FALSE | TRUE | FALSE |
| KLHL8 | 0.045929 | 0.8768753 | 1 | 0.419057985 | 3.924270906 | 8.289353331 | 22.2107148 | 0.122188489 | 0.1951 | 2.360956423 | 88.8307474 | 88.8307474 | 0.002067878 | -2.684475099 | FALSE | FALSE | FALSE |
| PML | 0.0459066 | 0.87688445 | 0.5228 | 0.408345868 | 0.408345868 | 7.968543345 | 0.56884483 | 0.725185755 | 0.7555 | 0.568844834 | 5.227432694 | 11.13304203 | 0.080701381 | -1.093119036 | FALSE | TRUE | FALSE |
| MTRR | 0.0458652 | 0.87689361 | 0.9375 | 0.406568197 | 14.79193103 | 14.79193103 | 0.02090248 | 0.893600949 | 1 | 0.02090248 | 0.02090248 | 0.413227278 | 2.194245157 | 0.341285148 | FALSE | TRUE | FALSE |
| MAK | 0.045853 | 0.87690276 | 0.8718 | 0.878522481 | 31.95575002 | 31.95575002 | 0.02081682 | 0.89361566 | 0.509 | 0.083247172 | 0.083247172 | 0.412128255 | 2.202687634 | 0.342952914 | FALSE | TRUE | FALSE |
| UNC80 | 0.0458483 | 0.87691191 | 1 | 0.045848253 | 0.045848253 | 1.535289564 | 0.06534134 | 0.883506547 | 1 | 0.065341336 | 0.065341336 | 2.192020146 | 0.701673027 | -0.153865218 | FALSE | TRUE | FALSE |
| DSC1 | 0.0458385 | 0.87692106 | 0.5102 | 0.04583855 | 0.04583855 | 0.902628648 | 0.66306657 | 0.684150259 | 0.3871 | 0.295838806 | 2.651580855 | 5.605711875 | 0.069131143 | -1.160326264 | FALSE | TRUE | FALSE |
| TAL1 | 0.0457857 | 0.8769302 | 1 | 1.80269E+16 | 7.64363E+21 | 1.53241E+24 | 0.73406955 | 0.649459715 | 1 | 4.15461E+17 | 2.01265E+23 | 2.90055E+24 | 0.062372364 | -1.205007797 | FALSE | TRUE | FALSE |
| MGAT2 | 0.0457853 | 0.87693935 | 1 | 0.045785342 | 0.045785342 | 0.906192788 | 0.06590375 | 0.883385134 | 1 | 0.065903747 | 0.065903747 | 1.306010628 | 0.694730477 | -0.158183649 | FALSE | TRUE | FALSE |
| CBL | 0.0456789 | 0.8769485 | 0.87655 | 0.413514703 | 3.842087894 | 8.129206672 | 0.02073912 | 0.893667119 | 1 | 0.020739122 | 0.020739122 | 0.408543807 | 2.20254872 | 0.342925524 | FALSE | TRUE | FALSE |
| ATP6V0A2 | 0.0455508 | 0.87695764 | 0.24115 | 0.045550842 | 0.72763135 | 2.909253421 | 0.00208022 | 0.899055969 | 0.265 | 0.002080224 | 0.008308927 | 0.040949073 | 21.8970865 | 1.340386334 | FALSE | TRUE | FALSE |
| PYCARD | 0.0455461 | 0.87696678 | 1 | 0.419829618 | 3.97181803 | 3.97181803 | 0.46924841 | 0.767292538 | 0.5036 | 4.450634635 | 43.29592567 | 43.29592567 | 0.09706173 | -1.012951971 | FALSE | TRUE | FALSE |
| MIIP | 0.0455428 | 0.87697592 | 0.01335 | 0.045542751 | 0.045542751 | 0.901718851 | 0.06281201 | 0.884067259 | 1 | 0.554767538 | 0.554767538 | 1.245140169 | 0.725064404 | -0.139623416 | FALSE | TRUE | FALSE |
| RFX5 | 0.045531 | 0.87698506 | 1 | 0.41426883 | 0.41426883 | 8.182608502 | 0.06737054 | 0.883019492 | 1 | 0.067370539 | 0.067370539 | 1.332684085 | 0.675829619 | -0.170162779 | FALSE | TRUE | FALSE |
| DLL1 | 0.0454477 | 0.8769942 | 1 | 0.429336363 | 0.429336363 | 8.4374415 | 0.0621263 | 0.884273037 | 0.93905 | 2.345289572 | 90.8885926 | 90.8885926 | 0.731537829 | -0.135763211 | FALSE | TRUE | FALSE |
| PFKFB3 | 0.0454238 | 0.87700334 | 0.01845 | 0.045423845 | 0.045423845 | 0.894860987 | 0.06540093 | 0.883480549 | 0.01565 | 0.065400929 | 0.065400929 | 1.290738686 | 0.694544344 | -0.158300021 | FALSE | TRUE | FALSE |
| PLXND1 | 0.0454128 | 0.87701248 | 0.06085 | 0.045412837 | 0.866905497 | 1.525078415 | 0.05773437 | 0.885098036 | 1 | 5.056900482 | 490.1994206 | 943.9229568 | 0.786582407 | -0.104255771 | FALSE | TRUE | FALSE |
| ZAP70 | 0.0453889 | 0.87702161 | 1 | 0.423143222 | 1.691482635 | 6.139407647 | 1.06695738 | 0.581688618 | 0.26135 | 0.26682528 | 1.06695738 | 5.253623632 | 0.042540513 | -1.371197281 | FALSE | FALSE | FALSE |
| PIKFYVE | 0.0452886 | 0.87703075 | 0.5365 | 0.045288586 | 0.181097417 | 3.48586218 | 0.7421957 | 0.645142229 | 0.2745 | 0.082250318 | 0.742195702 | 14.3509714 | 0.061019736 | -1.214529675 | FALSE | TRUE | FALSE |
| RPL37 | 0.0452331 | 0.87703988 | 1 | 0.045233067 | 0.045233067 | 0.045233067 | 0.04683269 | 0.887389234 | 0.00375 | 0.046832685 | 0.046832685 | 0.046832685 | 0.965843984 | -0.015093021 | FALSE | TRUE | FALSE |
| SCUBE1 | 0.0451927 | 0.87704901 | 0.89105 | 0.180617335 | 2.88713122 | 14.0806763 | 0.0233813 | 0.892882889 | 0.51285 | 0.093438287 | 0.373499121 | 1.827647834 | 1.932858 | 0.286199949 | FALSE | TRUE | FALSE |
| AMDHD1 | 0.0451879 | 0.87705814 | 1 | 0.180680254 | 0.722615818 | 0.89597706 | 0.06631978 | 0.883254789 | 0.01085 | 0.066319778 | 0.066319778 | 1.316336654 | 0.681364365 | -0.166620584 | FALSE | TRUE | FALSE |

| | | | | | | | | | | | | | | | | |
|---|---|---|---|---|---|---|---|---|---|---|---|---|---|---|---|---|
| IGF2R | 0.0450437 | 0.87706727 | 0.5335 | 0.411256556 | 0.854006777 | 13.73591017 | 0.06669175 | 0.883185162 | 1 | 0.066691755 | 1.272397533 | 2.232228972 | 0.675400678 | -0.170438508 | FALSE | TRUE | FALSE |
| CDC25C | 0.0450381 | 0.8770764 | 0.5022 | 0.399105756 | 0.399105756 | 0.892794978 | 0.57414349 | 0.72281102 | 0.009 | 0.574143494 | 0.574143494 | 0.574143494 | 0.078443923 | -1.105440697 | FALSE | TRUE | FALSE |
| SYNE4 | 0.0450215 | 0.87708553 | 0.8784 | 0.412717261 | 3.880668252 | 3.880668252 | 0.00683532 | 0.897586656 | 1 | 0.061529927 | 0.568479893 | 0.568479893 | 6.586595138 | 0.818660969 | FALSE | TRUE | FALSE |
| FAM13A | 0.0449747 | 0.87709465 | 1 | 0.402918261 | 0.402918261 | 7.896990407 | 0.06637567 | 0.88324609 | 0.02515 | 0.066375667 | 0.066375667 | 1.304066452 | 0.677578082 | -0.169040651 | FALSE | TRUE | FALSE |
| LCN1 | 0.0449493 | 0.87710378 | 1 | 0.415141386 | 0.415141386 | 0.415141386 | 0.05274649 | 0.886079363 | 1 | 0.052746492 | 0.052746492 | 0.052746492 | 0.852177006 | -0.069470188 | FALSE | TRUE | FALSE |
| ELMO3 | 0.0448458 | 0.8771129 | 0.7616 | 0.095472423 | 0.381871448 | 1.867896729 | 5.50242056 | 0.286450146 | 0.13525 | 0.622508023 | 5.502420563 | 12.21115692 | 0.008150188 | -2.088832386 | FALSE | FALSE | FALSE |
| TRMT11 | 0.0448163 | 0.87712203 | 1 | 0.398274762 | 0.398274762 | 0.889082635 | 0.25335092 | 0.838508134 | 0.50955 | 0.567205652 | 2.268452352 | 2.268452352 | 0.176894301 | -0.752286158 | FALSE | TRUE | FALSE |
| PCDHB6 | 0.0447539 | 0.87713115 | 0.2696 | 0.044753855 | 0.40361427 | 0.875912164 | 0.00995832 | 0.896764316 | 0.02335 | 0.009958324 | 0.009958324 | 0.195404705 | 4.494115115 | 0.652644193 | FALSE | TRUE | FALSE |
| PDLIM1 | 0.0447453 | 0.87714027 | 1 | 0.044745349 | 0.044745349 | 0.044745349 | 0.06389935 | 0.883817698 | 0.0085 | 0.063899348 | 0.063899348 | 0.063899348 | 0.700247355 | -0.154748523 | FALSE | TRUE | FALSE |
| SHBG | 0.0447204 | 0.87714939 | 1 | 0.404260216 | 0.404260216 | 0.887122711 | 5.27558513 | 0.295129535 | 0.0073 | 0.565923452 | 5.275585128 | 5.275585128 | 0.008476858 | -2.071765112 | FALSE | FALSE | FALSE |
| HEATR4 | 0.0447094 | 0.8771585 | 0.1459 | 0.044709357 | 0.391173868 | 0.876477153 | 0.00122674 | 0.899322578 | 1 | 0.00122674 | 0.00122674 | 0.024106225 | 36.4456794 | 1.56164605 | FALSE | TRUE | FALSE |
| FBXL18 | 0.0446942 | 0.87716762 | 1 | 0.044694177 | 0.044694177 | 0.874780096 | 0.05919736 | 0.884869429 | 1 | 0.059197363 | 0.059197363 | 1.161627433 | 0.755002832 | -0.12205142 | FALSE | TRUE | FALSE |
| FABP9 | 0.044668 | 0.87717673 | 0.0032 | 0.044668003 | 0.044668003 | 0.044668003 | 0.048441 | 0.887021297 | 0.0023 | 0.048441 | 0.048441 | 0.048441 | 0.922111503 | -0.03521656 | FALSE | TRUE | FALSE |
| NUDCD1 | 0.0446489 | 0.87718585 | 1 | 0.04464886 | 0.04464886 | 0.883439994 | 0.55118509 | 0.732876806 | 0.50445 | 0.551185086 | 0.551185086 | 1.226237702 | 0.081005203 | -1.091487083 | FALSE | TRUE | FALSE |
| DNAJC13 | 0.0446087 | 0.87719496 | 1 | 0.044608719 | 0.858085195 | 1.490238414 | 0.06224137 | 0.884247352 | 0.5284 | 0.56950866 | 0.56950866 | 11.00548683 | 0.716705251 | -0.144659413 | FALSE | TRUE | FALSE |
| LIPG | 0.0446028 | 0.87720407 | 0.01435 | 0.044602775 | 0.044602775 | 0.220517927 | 0.01894709 | 0.894113648 | 1 | 0.075724009 | 0.075724009 | 0.374887043 | 2.354069394 | 0.371819261 | FALSE | TRUE | FALSE |
| SKA3 | 0.0445774 | 0.87721318 | 1 | 0.407014697 | 0.407014697 | 0.884622274 | 0.05544014 | 0.885536036 | 0.01045 | 0.055440138 | 0.055440138 | 1.101217205 | 0.804063591 | -0.094709603 | FALSE | TRUE | FALSE |
| HLA-DQB1 | 0.0445439 | 0.87722229 | 1 | 0.044543927 | 0.044543927 | 0.884143555 | 0.05267976 | 0.886087687 | 0.00895 | 0.052679762 | 0.052679762 | 0.052679762 | 0.845560519 | -0.072855304 | FALSE | TRUE | FALSE |
| AKR1C4 | 0.0445002 | 0.87723139 | 0.009 | 0.044500249 | 0.044500249 | 0.044500249 | 0.54860069 | 0.733778352 | 0.4971 | 0.06168259 | 0.54860069 | 0.54860069 | 0.081115919 | -1.090893906 | FALSE | TRUE | FALSE |
| DDOST | 0.0444758 | 0.8772405 | 1 | 0.044475838 | 0.044475838 | 0.880880005 | 0.06421325 | 0.883748682 | 1 | 0.064213253 | 0.064213253 | 1.273280817 | 0.692627092 | -0.159500525 | FALSE | TRUE | FALSE |
| CTNND1 | 0.0444476 | 0.8772496 | 1 | 0.044447609 | 0.044447609 | 0.868826812 | 0.06033815 | 0.884665568 | 1 | 0.563690503 | 5.404767347 | 11.04862984 | 0.73664191 | -0.132743576 | FALSE | TRUE | FALSE |
| TMEM175 | 0.0443565 | 0.87725871 | 1 | 0.395024476 | 0.395024476 | 1.502775817 | 0.56983482 | 0.724849138 | 0.5114 | 0.569834823 | 0.569834823 | 1.254127704 | 0.077840978 | -1.108791718 | FALSE | TRUE | FALSE |
| ACOT12 | 0.0443472 | 0.87726781 | 0.50265 | 0.177363904 | 0.177363904 | 1.501139219 | 41.7518846 | 0.06548861 | 0.3474 | 41.75188463 | 1604.66045 | 15796.9906 | 0.001062159 | -2.9738103 | FALSE | FALSE | FALSE |
| EML2 | 0.0443259 | 0.87727691 | 0.94115 | 0.388230142 | 32.12117112 | 32.12117112 | 0.00918025 | 0.896965823 | 1 | 6.371433342 | 59.32613358 | 565.9182796 | 4.828401522 | 0.683803378 | FALSE | TRUE | FALSE |
| TMEM255B | 0.0443169 | 0.87728601 | 1 | 0.044316892 | 0.044316892 | 0.87895373 | 4.79039674 | 0.315621 | 0.2606 | 0.527388532 | 4.790396744 | 4.790396744 | 0.009251194 | -2.033802191 | FALSE | FALSE | FALSE |
| GRB10 | 0.0442559 | 0.87729511 | 0.01535 | 0.04425591 | 0.04425591 | 0.874945442 | 0.06302693 | 0.88400711 | 0.5142 | 0.252011204 | 0.252011204 | 1.247774113 | 0.702174582 | -0.153554895 | FALSE | TRUE | FALSE |
| RSPH6A | 0.0442338 | 0.87730421 | 1 | 0.044233804 | 0.044233804 | 0.868562871 | 0.54928668 | 0.733418487 | 0.514 | 0.549286681 | 0.549286681 | 10.80929673 | 0.08052954 | -1.094044782 | FALSE | TRUE | FALSE |
| HGF | 0.0442104 | 0.8773133 | 1 | 0.176738126 | 0.176738126 | 3.484905524 | 0.06263657 | 0.884127349 | 1 | 0.582194741 | 0.582194741 | 2.114711715 | 0.705824456 | -0.151303298 | FALSE | TRUE | FALSE |
| CPA3 | 0.0441839 | 0.8773224 | 0.01125 | 0.044183915 | 0.044183915 | 0.876502861 | 2.04133255 | 0.485723042 | 0.68885 | 4.674022104 | 175.5797142 | 175.5797142 | 0.021644643 | -1.664649566 | FALSE | FALSE | FALSE |
| WDR38 | 0.0441204 | 0.87733149 | 1 | 0.044120377 | 0.044120377 | 0.044120377 | 0.06131862 | 0.884469603 | 1 | 0.548321529 | 0.548321529 | 0.548321529 | 0.719526638 | -0.142953123 | FALSE | TRUE | FALSE |
| BTAF1 | 0.0440439 | 0.87734058 | 0.98505 | 62.39874832 | 9764.672751 | 62913.52102 | 0.00527525 | 0.898058037 | 1 | 183.241349 | 79108.44712 | 834199.7585 | 8.349162221 | 0.921642899 | FALSE | TRUE | FALSE |
| MARS2 | 0.0440073 | 0.87734967 | 1 | 0.044007256 | 0.044007256 | 0.866358947 | 0.2508115 | 0.839100938 | 0.01795 | 0.250811499 | 0.250811499 | 2.12275756 | 0.175459483 | -0.755823155 | FALSE | TRUE | FALSE |
| FRRS1 | 0.0439747 | 0.87735876 | 1 | 0.39574165 | 0.39574165 | 1.488297251 | 0.06515051 | 0.883549853 | 0.0141 | 0.065150512 | 0.065150512 | 1.290572536 | 0.674971125 | -0.170714806 | FALSE | TRUE | FALSE |
| FARSB | 0.0439066 | 0.87736785 | 0.50075 | 0.402253958 | 0.402253958 | 0.868215998 | 0.25834411 | 0.837162402 | 0.50955 | 0.258344107 | 0.258344107 | 1.279441784 | 0.169953851 | -0.769668991 | FALSE | TRUE | FALSE |
| AREL1 | 0.0438996 | 0.87737694 | 1 | 0.043899574 | 0.043899574 | 0.862776364 | 0.0632051 | 0.883972713 | 1 | 0.063205103 | 0.063205103 | 1.244787556 | 0.694557431 | -0.158291838 | FALSE | TRUE | FALSE |
| TOM1L1 | 0.0438895 | 0.87738603 | 1 | 0.043889497 | 0.043889497 | 0.870535062 | 0.06260002 | 0.884135928 | 0.0105 | 0.062600025 | 0.062600025 | 1.242886948 | 0.701109907 | -0.154213896 | FALSE | TRUE | FALSE |
| ECSIT | 0.0438451 | 0.87739511 | 1 | 0.394348652 | 1.576909681 | 1.576909681 | 0.06569853 | 0.883428521 | 0.01335 | 0.065698527 | 0.065698527 | 1.301276505 | 0.667367356 | -0.175635041 | FALSE | TRUE | FALSE |
| GPR155 | 0.0437315 | 0.87740419 | 1 | 0.386926456 | 3.513759829 | 7.625810539 | 5.11221595 | 0.303335464 | 0.2609 | 0.558471963 | 5.112215949 | 11.02604213 | 0.008554322 | -2.067814398 | FALSE | FALSE | FALSE |
| CYP2U1 | 0.0437241 | 0.87741328 | 1 | 0.399575582 | 3.747787354 | 7.92783989 | 0.06708986 | 0.883080579 | 1 | 0.067089863 | 0.067089863 | 0.067089863 | 0.651723818 | -0.185936407 | FALSE | TRUE | FALSE |
| PKD2 | 0.0437095 | 0.87742236 | 0.7567 | 0.043709464 | 0.174715262 | 1.473397487 | 0.01806684 | 0.894310216 | 1 | 0.288656748 | 2.666185977 | 5.630958084 | 2.419320123 | 0.383693338 | FALSE | TRUE | FALSE |
| CARD16 | 0.0436725 | 0.87743144 | 0.7502 | 0.043672485 | 0.417310334 | 0.417310334 | 0.00529195 | 0.89804443 | 1 | 0.005291953 | 0.049428118 | 0.049428118 | 8.252622214 | 0.916591965 | FALSE | TRUE | FALSE |
| ACSL5 | 0.0436574 | 0.87744052 | 1 | 0.043657355 | 0.043657355 | 0.862078393 | 0.06273041 | 0.884101603 | 1 | 0.06273041 | 0.06273041 | 1.240593266 | 0.695952014 | -0.157420704 | FALSE | TRUE | FALSE |
| MIOX | 0.043635 | 0.8774496 | 1 | 0.174474625 | 0.174474625 | 0.866110913 | 0.55881727 | 0.729251831 | 0.50095 | 0.558817274 | 0.558817274 | 0.558817274 | 0.078084581 | -1.107434718 | FALSE | TRUE | FALSE |
| DEPTOR | 0.0436062 | 0.87745867 | 0.0123 | 0.043606193 | 0.043606193 | 0.863897312 | 0.53406983 | 0.740764698 | 0.50175 | 0.534069832 | 0.534069832 | 0.534069832 | 0.081648861 | -1.088049869 | FALSE | TRUE | FALSE |
| KIF25 | 0.0435962 | 0.87746775 | 0.01085 | 0.04359617 | 0.04359617 | 0.864697528 | 0.05725817 | 0.885233123 | 1 | 4.747038435 | 44.91401777 | 44.91401777 | 0.761396479 | -0.118389136 | FALSE | TRUE | FALSE |
| RAD9A | 0.043582 | 0.87747682 | 1 | 0.400403438 | 0.400403438 | 0.862872108 | 4.20726249 | 0.34301142 | 0.137 | 4.207262486 | 40.77272251 | 40.77272251 | 0.010358756 | -1.984692394 | FALSE | FALSE | FALSE |
| IQSEC3 | 0.043524 | 0.87748589 | 1 | 25.72387629 | 976.1774924 | 9496.172409 | 0.06439272 | 0.883722782 | 1 | 9.545513491 | 221.6028812 | 886.2694012 | 0.67591419 | -0.170108436 | FALSE | TRUE | FALSE |
| KL | 0.0434954 | 0.87749496 | 1 | 0.173923868 | 0.173923868 | 3.40164065 | 0.06348237 | 0.883903864 | 0.02575 | 0.063482367 | 0.063482367 | 1.244905714 | 0.685156535 | -0.164210195 | FALSE | TRUE | FALSE |
| TMEM214 | 0.0434941 | 0.87750403 | 1 | 0.39356175 | 0.39356175 | 1.470935378 | 0.06461545 | 0.883662309 | 0.01415 | 0.06461545 | 0.06461545 | 1.278300963 | 0.673122507 | -0.171905888 | FALSE | TRUE | FALSE |
| MFSD8 | 0.043457 | 0.8775131 | 1 | 0.04345701 | 0.04345701 | 0.861014032 | 0.0625578 | 0.884161659 | 1 | 0.062557798 | 0.062557798 | 1.240851943 | 0.694669747 | -0.158221614 | FALSE | TRUE | FALSE |
| GRIP1 | 0.0433246 | 0.87752217 | 1 | 0.393672696 | 0.393672696 | 7.70676314 | 0.057092 | 0.88525842 | 1 | 4.861327623 | 458.134733 | 458.134733 | 0.75885621 | -0.119840508 | FALSE | TRUE | FALSE |
| DHH | 0.0432554 | 0.87753124 | 1 | 0.043255355 | 0.043255355 | 0.855365894 | 0.06273509 | 0.884093019 | 1 | 0.062735093 | 0.062735093 | 1.24225433 | 0.689492172 | -0.16147066 | FALSE | TRUE | FALSE |
| FARSA | 0.0432314 | 0.8775403 | 0.0155 | 0.043231423 | 0.043231423 | 0.853457881 | 0.06254177 | 0.884170234 | 1 | 0.062541765 | 0.062541765 | 1.236594574 | 0.69124085 | -0.160370604 | FALSE | TRUE | FALSE |
| DTNB | 0.0432229 | 0.87754937 | 0.02005 | 0.04322294 | 0.04322294 | 0.852445955 | 0.24383883 | 0.840900874 | 0.7562 | 0.243838826 | 0.975289263 | 4.817049375 | 0.177260288 | -0.75138855 | FALSE | TRUE | FALSE |
| RUNDC3A | 0.0432135 | 0.87755843 | 0.00935 | 0.043213529 | 0.043213529 | 0.043213529 | 0.42555913 | 0.782681897 | 0.5091 | 0.425559132 | 0.425559132 | 0.425559132 | 0.101545299 | -0.993340176 | FALSE | TRUE | FALSE |
| C1orf122 | 0.0432056 | 0.87756749 | 1 | 0.043205612 | 0.043205612 | 0.043205612 | 0.04662822 | 0.887405536 | 0.00085 | 0.046628223 | 0.046628223 | 0.046628223 | 0.926597866 | -0.033108704 | FALSE | TRUE | FALSE |
| MRC2 | 0.0431718 | 0.87757655 | 0.76105 | 0.389517407 | 3.607082324 | 69.62822392 | 0.06609125 | 0.88332434 | 1 | 0.066091249 | 0.066091249 | 2.222973618 | 0.653215221 | -0.184943704 | FALSE | TRUE | FALSE |
| TIAM1 | 0.0431661 | 0.87758561 | 0.05075 | 0.043166091 | 0.831945033 | 1.449406516 | 0.55166511 | 0.732831562 | 0.0429 | 0.551665109 | 0.551665109 | 10.67893525 | 0.078246912 | -1.106532793 | FALSE | TRUE | FALSE |
| DDX49 | 0.0431205 | 0.87759467 | 0.0155 | 0.043120484 | 0.043120484 | 0.851079738 | 0.06197066 | 0.884324374 | 1 | 0.061970663 | 0.061970663 | 1.225064066 | 0.695820923 | -0.157502516 | FALSE | TRUE | FALSE |
| GALK1 | 0.0430582 | 0.87760372 | 1 | 0.043058223 | 0.043058223 | 0.852431963 | 0.06231221 | 0.884230223 | 0.01095 | 0.062312213 | 0.062312213 | 1.235111675 | 0.69100776 | -0.160517075 | FALSE | TRUE | FALSE |
| VPS4B | 0.0430063 | 0.87761278 | 1 | 0.043006283 | 0.043006283 | 0.853018821 | 0.24622304 | 0.840258022 | 0.50755 | 0.246223039 | 0.246223039 | 1.222810954 | 0.174663926 | -0.757796781 | FALSE | TRUE | FALSE |
| CDKL4 | 0.0429859 | 0.87762183 | 1 | 0.042985905 | 0.042985905 | 0.042985905 | 0.06158585 | 0.884418384 | 1 | 0.061585845 | 0.061585845 | 0.061585845 | 0.697983526 | -0.156154828 | FALSE | TRUE | FALSE |
| PIGO | 0.0429391 | 0.87763089 | 1 | 3632122577 | 5.5522E+13 | 8.81859E+15 | 0.07475424 | 0.881437502 | 1 | 59924143261 | 1.26324E+15 | 2.05413E+17 | 0.57440323 | -0.240783126 | FALSE | TRUE | FALSE |
| KLRB1 | 0.0428709 | 0.87763994 | 1 | 0.042870868 | 0.042870868 | 0.042870868 | 0.05182993 | 0.886245629 | 1 | 0.051829928 | 0.051829928 | 0.051829928 | 0.827145049 | -0.082418325 | FALSE | TRUE | FALSE |
| TMEM44 | 0.0428358 | 0.87764899 | 0.01 | 0.042835791 | 0.042835791 | 0.043378555 | 2.05555001 | 0.48438439 | 0.0012 | 0.06061304 | 0.06061304 | 0.06061304 | 0.02083909 | -1.681121259 | FALSE | FALSE | FALSE |
| PLCB4 | 0.0427734 | 0.87765804 | 0.5104 | 0.390425006 | 0.390425006 | 7.639993502 | 0.56747742 | 0.725760807 | 0.0291 | 0.567477421 | 0.567477421 | 11.13349222 | 0.075374565 | -1.122775183 | FALSE | TRUE | FALSE |
| MYO1B | 0.0427296 | 0.87766709 | 0.50895 | 0.097562001 | 1.558248618 | 3.283773093 | 0.76972337 | 0.632552413 | 0.36605 | 0.769723369 | 12.30353069 | 49.20848942 | 0.055512931 | -1.255605842 | FALSE | TRUE | FALSE |
| TBC1D5 | 0.0427289 | 0.87767613 | 1 | 0.375655601 | 3.389771303 | 7.410492401 | 0.06493987 | 0.883610431 | 1 | 0.064939874 | 0.064939874 | 1.283163257 | 0.657976244 | -0.181789786 | FALSE | TRUE | FALSE |
| RORA | 0.0427184 | 0.87768518 | 1 | 0.399133388 | 0.399133388 | 0.844827848 | 0.06064525 | 0.884614503 | 1 | 0.969313845 | 9.073937624 | 11.24254497 | 0.704397758 | -0.152182035 | FALSE | TRUE | FALSE |
| SPEG | 0.0426973 | 0.87769422 | 0.8847 | 0.394636241 | 3.741202171 | 69.83003708 | 0.31344468 | 0.820859793 | 0.79475 | 1.253559414 | 97.08133873 | 388.2574348 | 0.136219575 | -0.86576048 | FALSE | TRUE | FALSE |
| RAD51D | 0.0426905 | 0.87770327 | 0.00995 | 0.042690482 | 0.042690482 | 0.042690482 | 0.06152233 | 0.884426923 | 1 | 0.061522233 | 0.061522233 | 1.222290344 | 0.693903312 | -0.158701039 | FALSE | TRUE | FALSE |
| SLC18A1 | 0.04269 | 0.87771231 | 0.3804 | 0.019552196 | 0.170606088 | 0.170606088 | 0.01096853 | 0.896485207 | 0.752 | 0.010968532 | 0.043840577 | 0.093048422 | 3.892038799 | 0.590177161 | FALSE | TRUE | FALSE |
| KIF18B | 0.0426798 | 0.87772135 | 0.02165 | 0.042679772 | 0.042679772 | 0.838093454 | 0.06193073 | 0.884332927 | 1 | 0.061930728 | 0.061930728 | 1.218779594 | 0.689153405 | -0.161684094 | FALSE | TRUE | FALSE |

| | | | | | | | | | | | | | | | | | |
|---|---|---|---|---|---|---|---|---|---|---|---|---|---|---|---|---|---|
| KNDC1 | 0.0426779 | 0.87773039 | 0.9428 | 3.612709094 | 338.4813481 | 661.040423 | 0.06963335 | 0.882546292 | 1 | 0.069633355 | 0.069633355 | 1.339631635 | 0.612894155 | -0.212614521 | FALSE | TRUE | FALSE |
| ASB4 | 0.042651 | 0.87773942 | 0.5022 | 0.379923456 | 0.379923456 | 0.845594026 | 0.06136445 | 0.88446107 | 1 | 0.550594966 | 0.550594966 | 1.217878142 | 0.695044425 | -0.157987436 | FALSE | TRUE | FALSE |
| IQUB | 0.0426453 | 0.87774846 | 0.0177 | 0.04264528 | 0.04264528 | 0.171635205 | 0.07824447 | 0.880808374 | 0.5139 | 0.078244469 | 0.078244469 | 0.310885142 | 0.545026121 | -0.263582684 | FALSE | TRUE | FALSE |
| ASCC3 | 0.0426408 | 0.8777575 | 0.03725 | 0.005021853 | 0.042640807 | 0.355710424 | 0.00330605 | 0.898552521 | 0.0477 | 0.003306045 | 0.003306045 | 0.06391359 | 12.89782926 | 1.110516624 | FALSE | TRUE | FALSE |
| CLDN8 | 0.0426257 | 0.87776653 | 0.00615 | 0.042625704 | 0.042625704 | 0.042625704 | 0.06164197 | 0.884409843 | 0.0057 | 0.061641968 | 0.061641968 | 0.061641968 | 0.691504586 | -0.160204935 | FALSE | TRUE | FALSE |
| PDGFRA | 0.0426241 | 0.87777556 | 0.02745 | 0.0426241 | 0.0426241 | 0.834178139 | 0.06190104 | 0.884341478 | 1 | 0.061901037 | 0.061901037 | 1.214572054 | 0.688584587 | -0.162042703 | FALSE | TRUE | FALSE |
| ARMC6 | 0.0426177 | 0.87778459 | 1 | 0.37586808 | 3.40245455 | 7.419701551 | 0.06486539 | 0.883636375 | 1 | 0.064865389 | 0.064865389 | 1.282455797 | 0.657017059 | -0.182423354 | FALSE | TRUE | FALSE |
| ZNF804A | 0.0425998 | 0.87779362 | 1 | 0.042599833 | 0.042599833 | 0.835583953 | 0.05335115 | 0.885987738 | 1 | 0.053351154 | 0.053351154 | 1.04889576 | 0.798480061 | -0.097735924 | FALSE | TRUE | FALSE |
| CYP4F12 | 0.0425441 | 0.87780265 | 0.0175 | 0.04254414 | 0.04254414 | 0.840020628 | 0.92443896 | 0.595458061 | 0.69445 | 2.131080464 | 34.08928581 | 34.08928581 | 0.046021579 | -1.337038488 | FALSE | TRUE | FALSE |
| BCAT1 | 0.0425432 | 0.87781168 | 0.00925 | 0.042543173 | 0.042543173 | 0.042543173 | 0.06115431 | 0.884512255 | 1 | 0.061154312 | 0.061154312 | 0.061154312 | 0.695669228 | -0.157597207 | FALSE | TRUE | FALSE |
| ZNF215 | 0.0425281 | 0.87782071 | 1 | 3.386372935 | 125.5955464 | 125.5955464 | 0.06741644 | 0.883002028 | 0.0124 | 0.067416443 | 0.067416443 | 1.337738368 | 0.630827037 | -0.200089702 | FALSE | TRUE | FALSE |
| ANKRD53 | 0.0424825 | 0.87782973 | 1 | 352753069.8 | 7.68917E+11 | 1.08108E+14 | 0.0795128 | 0.880500608 | 1 | 5851.97365 | 6493157.232 | 70296078.63 | 0.534285596 | -0.272226534 | FALSE | TRUE | FALSE |
| DNAJC4 | 0.0424818 | 0.87783875 | 0.50235 | 0.372244494 | 0.372244494 | 0.372244494 | 0.53788612 | 0.739398569 | 0.5034 | 0.537886124 | 0.537886124 | 0.537886124 | 0.078979169 | -1.102487441 | FALSE | TRUE | FALSE |
| ASNSD1 | 0.0424675 | 0.87784778 | 1 | 0.04246748 | 0.04246748 | 0.839538684 | 40.9930004 | 0.067377529 | 0.1965 | 40.99300041 | 396.4619286 | 3928.077052 | 0.001035969 | -2.984653215 | FALSE | FALSE | FALSE |
| FAM83D | 0.0424614 | 0.8778568 | 1 | 0.383384979 | 3.552864557 | 7.581265598 | 0.0575645 | 0.885148728 | 0.0138 | 0.057564502 | 0.057564502 | 1.139855885 | 0.73763092 | -0.132160887 | FALSE | TRUE | FALSE |
| DAPK1 | 0.0424417 | 0.87786582 | 1 | 0.042441735 | 0.042441735 | 1.427974356 | 0.06090545 | 0.884563399 | 0.52575 | 0.243530343 | 0.243530343 | 4.723723219 | 0.696846295 | -0.156863005 | FALSE | TRUE | FALSE |
| GBGT1 | 0.0423927 | 0.87787483 | 0.01035 | 0.042392664 | 0.042392664 | 0.840664211 | 0.04946831 | 0.8867995 | 1 | 0.049468311 | 0.049468311 | 0.981977606 | 0.856966068 | -0.067036374 | FALSE | TRUE | FALSE |
| CCDC62 | 0.0423112 | 0.87788385 | 1 | 0.042311156 | 0.042311156 | 0.835444212 | 0.06074958 | 0.884588956 | 0.0163 | 0.06074958 | 0.06074958 | 1.20135396 | 0.696484742 | -0.157088394 | FALSE | TRUE | FALSE |
| GRAMD1C | 0.0423091 | 0.87789287 | 1 | 0.376372497 | 3.436445595 | 7.44476712 | 0.56292281 | 0.727565291 | 0.50885 | 0.562922812 | 0.562922812 | 1.243027827 | 0.075159764 | -1.12401459 | FALSE | TRUE | FALSE |
| SPTLC2 | 0.0422605 | 0.87790188 | 1 | 0.395671336 | 1.581465831 | 3.342998487 | 0.0634098 | 0.883912475 | 1 | 0.253466786 | 0.253466786 | 1.256542077 | 0.666466823 | -0.176221465 | FALSE | TRUE | FALSE |
| CSF1R | 0.0422175 | 0.8779109 | 1 | 0.0422175 | 0.0422175 | 0.826609 | 0.06127477 | 0.884478136 | 1 | 0.061274773 | 0.061274773 | 1.202782361 | 0.688986642 | -0.161789198 | FALSE | TRUE | FALSE |
| ARHGEF10L | 0.0421896 | 0.87791991 | 1 | 0.383001602 | 0.383001602 | 7.429909985 | 0.05923546 | 0.88484398 | 1 | 0.540045855 | 5.05331088 | 18.22125103 | 0.712235743 | -0.147376235 | FALSE | TRUE | FALSE |
| LGR6 | 0.0421812 | 0.87792892 | 1 | 0.042181183 | 0.042181183 | 0.825877371 | 0.06114624 | 0.884520781 | 0.02375 | 0.061146244 | 0.061146244 | 1.200233079 | 0.68984095 | -0.161251029 | FALSE | TRUE | FALSE |
| THAP7 | 0.0421322 | 0.87793793 | 1 | 0.04213224 | 0.04213224 | 0.04213224 | 0.06126177 | 0.884495198 | 1 | 0.06126177 | 0.06126177 | 0.06126177 | 0.687741142 | -0.162574994 | FALSE | TRUE | FALSE |
| RIOK2 | 0.0421101 | 0.87794694 | 0.0148 | 0.042110066 | 0.042110066 | 0.833454605 | 0.05863135 | 0.884920299 | 1 | 0.518198857 | 0.518198857 | 1.161898508 | 0.718217516 | -0.143744007 | FALSE | TRUE | FALSE |
| EGFLAM | 0.0420484 | 0.87795595 | 0.33105 | 0.010519877 | 0.168110949 | 0.354842348 | 0.11707887 | 0.871920579 | 0.5067 | 0.117078871 | 1.869648263 | 7.474177695 | 0.359145601 | -0.444729449 | FALSE | TRUE | FALSE |
| KIR3DL2 | 0.0420458 | 0.87796495 | 1 | 0.04204584 | 0.04204584 | 0.832230858 | 0.41704823 | 0.78572298 | 0.4994 | 0.044409855 | 0.417048225 | 0.88011501 | 0.100817693 | -0.996463246 | FALSE | TRUE | FALSE |
| DGUOK | 0.0420077 | 0.87797396 | 0.7539 | 0.042007663 | 0.167900362 | 0.167900362 | 0.28705125 | 0.829012548 | 0.25385 | 0.071809817 | 0.287051245 | 0.287051245 | 0.146342032 | -0.834630919 | FALSE | TRUE | FALSE |
| OR2T5 | 0.0419025 | 0.87798296 | 1 | 0.041902482 | 0.041902482 | 0.041902482 | 0.06320081 | 0.883981314 | 0.5012 | 0.599748617 | 0.599748617 | 0.599748617 | 0.663005504 | -0.178482866 | FALSE | TRUE | FALSE |
| BRDT | 0.0418637 | 0.87799196 | 0.5169 | 0.369906067 | 0.369906067 | 7.289068042 | 0.06204059 | 0.884307267 | 1 | 0.062040595 | 0.062040595 | 1.224690428 | 0.674779636 | -0.170838033 | FALSE | TRUE | FALSE |
| RPS6KC1 | 0.0418589 | 0.87800096 | 1 | 314.3400701 | 31566.22272 | 104999.4506 | 0.0746392 | 0.881464335 | 0.0244 | 0.074639195 | 0.074639195 | 1.469102296 | 0.560817051 | -0.25117879 | FALSE | TRUE | FALSE |
| SLFN11 | 0.0417365 | 0.87800997 | 0.96585 | 29.1698159 | 2595.476026 | 25352.46478 | 0.73796331 | 0.647355774 | 0.8117 | 6.653522754 | 61.493326 | 582.2443719 | 0.056556381 | -1.247518387 | FALSE | TRUE | FALSE |
| STAR | 0.0417169 | 0.87801896 | 0.50765 | 0.379454582 | 0.379454582 | 0.828477719 | 0.0580154 | 0.885055762 | 1 | 2.118460343 | 19.85739181 | 19.85739181 | 0.719065954 | -0.143231273 | FALSE | TRUE | FALSE |
| ECT2 | 0.0417162 | 0.87802796 | 1 | 0.373254319 | 0.373254319 | 7.351621369 | 0.062098 | 0.884290154 | 1 | 0.062097998 | 0.062097998 | 1.225320466 | 0.67178072 | -0.172772465 | FALSE | TRUE | FALSE |
| PAPPA2 | 0.0416988 | 0.87803696 | 0.5302 | 0.041698829 | 0.041698829 | 0.803785139 | 0.00877167 | 0.897097456 | 1 | 0.078405487 | 0.078405487 | 1.517943593 | 4.75380614 | 0.677041467 | FALSE | TRUE | FALSE |
| CYP19A1 | 0.0416777 | 0.87804595 | 1 | 0.166606619 | 0.166606619 | 0.824823563 | 0.24228687 | 0.841336981 | 0.50435 | 0.242286871 | 0.242286871 | 1.200918486 | 0.172018127 | -0.764425785 | FALSE | TRUE | FALSE |
| LIPN | 0.0416411 | 0.87805495 | 1 | 0.374900143 | 0.374900143 | 0.374900143 | 0.23113699 | 0.844563717 | 0.8768 | 2.091795433 | 19.42978765 | 19.42978765 | 0.180157514 | -0.744347618 | FALSE | TRUE | FALSE |
| MAP2 | 0.0415822 | 0.87806394 | 1 | 0.041582224 | 0.041582224 | 1.396368676 | 0.06038292 | 0.884648551 | 1 | 0.060382924 | 0.060382924 | 1.174240103 | 0.688642098 | -0.162006432 | FALSE | TRUE | FALSE |
| F13B | 0.0415604 | 0.87807293 | 0.75165 | 0.041560434 | 0.166186357 | 0.822071708 | 0.01805064 | 0.89432475 | 1 | 0.018050642 | 0.018050642 | 0.357516008 | 2.302435228 | 0.362187422 | FALSE | TRUE | FALSE |
| EPHX2 | 0.0415581 | 0.87808192 | 1 | 0.368183714 | 0.368183714 | 1.407528592 | 0.06168867 | 0.884401302 | 0.0136 | 0.061688666 | 0.061688666 | 1.220528638 | 0.673674748 | -0.171549731 | FALSE | TRUE | FALSE |
| ALG12 | 0.0415398 | 0.87809091 | 0.5097 | 0.04153976 | 0.04153976 | 0.205236899 | 21.4252598 | 0.126759724 | 0.13035 | 1.340325486 | 21.42525984 | 85.6932912 | 0.001938822 | -2.71246211 | FALSE | FALSE | FALSE |
| ZNF25 | 0.0415238 | 0.8780999 | 1 | 0.371356444 | 0.371356444 | 0.824622147 | 0.06182172 | 0.884358576 | 0.0092 | 0.061821719 | 0.061821719 | 0.061821719 | 0.671669957 | -0.172844077 | FALSE | TRUE | FALSE |
| ZMYM6 | 0.0414592 | 0.87810889 | 0.02495 | 0.041459173 | 0.041459173 | 0.81661711 | 0.05976242 | 0.884776068 | 1 | 0.059762422 | 0.059762422 | 1.179281581 | 0.693733145 | -0.158807555 | FALSE | TRUE | FALSE |
| F2 | 0.0414347 | 0.87811787 | 0.50755 | 0.041434747 | 0.041434747 | 0.815690609 | 0.01734906 | 0.894491639 | 0.50585 | 0.15675814 | 0.15675814 | 0.586849599 | 2.3882989 | 0.378088679 | FALSE | TRUE | FALSE |
| THAP6 | 0.041425 | 0.87812686 | 1 | 0.165584267 | 0.165584267 | 0.165584267 | 0.53040245 | 0.742785641 | 0.75485 | 0.530402455 | 5.039343527 | 5.039343527 | 0.078101087 | -1.107342919 | FALSE | TRUE | FALSE |
| ZMYM1 | 0.0414196 | 0.87813584 | 0.51825 | 0.369992153 | 0.369992153 | 7.267505685 | 0.05202702 | 0.886229021 | 0.9394 | 4.297919652 | 392.9771333 | 392.9771333 | 0.796117222 | -0.099022981 | FALSE | TRUE | FALSE |
| KIF23 | 0.0414176 | 0.87814482 | 1 | 0.041417646 | 0.041417646 | 1.394290534 | 0.060359 | 0.88465706 | 1 | 0.060359005 | 0.060359005 | 1.186762034 | 0.686188357 | -0.163556655 | FALSE | TRUE | FALSE |
| EOMES | 0.0413859 | 0.8781538 | 1 | 0.38659845 | 0.38659845 | 0.818577283 | 0.05846378 | 0.884962662 | 1 | 0.546688637 | 5.246629139 | 10.82738573 | 0.707890157 | -0.150034126 | FALSE | TRUE | FALSE |
| WFIKKN2 | 0.0413445 | 0.87816278 | 0.9918 | 350.335343 | 37364.76643 | 70339.3217 | 0.05259718 | 0.886112652 | 1 | 47436.04283 | 23459527.37 | 262420675.6 | 0.786059646 | -0.104544498 | FALSE | TRUE | FALSE |
| AFG3L2 | 0.0413165 | 0.87817176 | 0.6954 | 0.165178129 | 2.642026422 | 3.253766273 | 0.01827061 | 0.894252041 | 1 | 0.073032963 | 0.073032963 | 1.441252088 | 2.261360919 | 0.354369882 | FALSE | TRUE | FALSE |
| MCM4 | 0.0413024 | 0.87818074 | 1 | 0.041302377 | 0.041302377 | 0.810255752 | 0.05955618 | 0.884793053 | 1 | 0.059556184 | 0.059556184 | 1.171042076 | 0.693502753 | -0.15895181 | FALSE | TRUE | FALSE |
| LOXL4 | 0.0413016 | 0.87818971 | 0.02305 | 0.041301568 | 0.041301568 | 0.810491594 | 0.273317 | 0.832834017 | 0.50915 | 0.273317005 | 1.093436605 | 5.375654452 | 0.151112324 | -0.820700115 | FALSE | TRUE | FALSE |
| PROX2 | 0.0412635 | 0.87819869 | 1 | 0.041263518 | 0.041263518 | 0.816409238 | 0.05702115 | 0.885275279 | 1 | 0.505725208 | 2.022778049 | 2.022778049 | 0.723652904 | -0.140469691 | FALSE | TRUE | FALSE |
| CRIP3 | 0.0412582 | 0.87820766 | 0.0045 | 0.04125817 | 0.04125817 | 0.04125817 | 0.22249111 | 0.84672645 | 0.8762 | 2.014109292 | 18.71338642 | 18.71338642 | 0.185437384 | -0.731802709 | FALSE | TRUE | FALSE |
| CAST | 0.0412512 | 0.87821663 | 1 | 0.041251248 | 0.041251248 | 0.814722868 | 0.05948745 | 0.884810033 | 0.01445 | 0.05948745 | 0.05948745 | 1.176657867 | 0.693444554 | -0.158988258 | FALSE | TRUE | FALSE |
| CTSF | 0.0411984 | 0.8782256 | 1 | 0.041198376 | 0.041198376 | 0.816587294 | 0.05956415 | 0.884784561 | 1 | 0.059564154 | 0.059564154 | 0.070787359 | 0.691663912 | -0.160104883 | FALSE | TRUE | FALSE |
| VWCE | 0.0411247 | 0.87823457 | 0.7545 | 0.164412825 | 1.463372419 | 3.223399339 | 0.00930187 | 0.89694501 | 1 | 0.037190366 | 0.037190366 | 0.730868508 | 4.421116401 | 0.645531949 | FALSE | TRUE | FALSE |
| SLC2A7 | 0.0411193 | 0.87824354 | 0.2622 | 0.041119287 | 0.164403392 | 0.812638099 | 0.01720616 | 0.894513372 | 1 | 0.154619493 | 0.618099309 | 1.361283551 | 2.389799669 | 0.378361497 | FALSE | TRUE | FALSE |
| PRKD1 | 0.0410527 | 0.87825251 | 0.9718 | 3.470934753 | 221.9328519 | 887.8991467 | 0.01953242 | 0.893945683 | 1 | 0.078075008 | 0.078075008 | 0.660299502 | 2.101774177 | 0.322586052 | FALSE | TRUE | FALSE |
| EFNA4 | 0.0410049 | 0.87826147 | 0.5022 | 0.369632252 | 0.369632252 | 0.369632252 | 0.2395682 | 0.842140604 | 0.2513 | 0.239568197 | 0.958153128 | 0.958153128 | 0.171161769 | -0.766593235 | FALSE | TRUE | FALSE |
| TAS2R46 | 0.0409854 | 0.87827044 | 1 | 0.388398615 | 3.777537313 | 3.777537313 | 0.03988731 | 0.88897468 | 1 | 0.039887308 | 0.039887308 | 0.039887308 | 1.027528729 | 0.011793973 | FALSE | TRUE | FALSE |
| DGKZ | 0.0409468 | 0.8782794 | 1 | 0.372580939 | 0.372580939 | 7.266456577 | 0.54415262 | 0.736444589 | 0.5032 | 0.544152618 | 0.544152618 | 10.64440192 | 0.075248737 | -1.123500788 | FALSE | TRUE | FALSE |
| ZNF780B | 0.0409179 | 0.87828836 | 0.6912 | 0.355950626 | 29.05275129 | 29.05275129 | 0.6676401 | 0.681615955 | 0.69095 | 0.667640105 | 55.8795997 | 55.8795997 | 0.061287427 | -1.212628613 | FALSE | TRUE | FALSE |
| ZNF223 | 0.0409038 | 0.87829732 | 1 | 0.040903789 | 0.040903789 | 0.810974199 | 0.05898775 | 0.88487791 | 1 | 0.05898775 | 0.05898775 | 0.05898775 | 0.693428538 | -0.158998289 | FALSE | TRUE | FALSE |
| SPATA7 | 0.0408812 | 0.87830628 | 1 | 0.357626253 | 0.357626253 | 1.383337434 | 0.5035886 | 0.754094861 | 0.26005 | 0.503588595 | 4.55956297 | 9.976859179 | 0.081179724 | -1.090552429 | FALSE | TRUE | FALSE |
| CCL23 | 0.0407529 | 0.87831524 | 1 | 0.040752882 | 0.040752882 | 0.040752882 | 0.04993658 | 0.886642937 | 1 | 0.04993658 | 0.04993658 | 0.04993658 | 0.816092781 | -0.088260464 | FALSE | TRUE | FALSE |
| ACOT4 | 0.0407449 | 0.8783242 | 1 | 0.364470436 | 0.364470436 | 0.807876462 | 31.8843635 | 0.08760358 | 0.132 | 4.601257441 | 73.59956745 | 294.4675481 | 0.001277895 | -2.893504779 | FALSE | FALSE | FALSE |
| CHN2 | 0.0407384 | 0.87833315 | 0.5057 | 0.040738417 | 0.040738417 | 0.20179339 | 0.01741989 | 0.894448149 | 0.0124 | 0.017419893 | 0.017419893 | 0.345415914 | 2.33861459 | 0.368958655 | FALSE | TRUE | FALSE |
| BCS1L | 0.0407044 | 0.87834211 | 0.87785 | 0.371186521 | 3.471892547 | 3.471892547 | 0.03667399 | 0.889748441 | 0.7521 | 0.036673988 | 0.146614503 | 0.310661088 | 1.109897281 | 0.045282787 | FALSE | TRUE | FALSE |
| ALDH16A1 | 0.040693 | 0.87835106 | 0.997 | 2743.90717 | 2870811.497 | 30449905.9 | 0.02750559 | 0.891845988 | 1 | 184775473.5 | 48239011812 | 2.25216E+12 | 1.479446024 | 0.170099125 | FALSE | TRUE | FALSE |
| ABCG8 | 0.0406929 | 0.87836001 | 0.9431 | 1.413268663 | 115.0305675 | 459.9996201 | 1.67760299 | 0.519703017 | 0.7775 | 15.49390373 | 2343.031453 | 9370.508883 | 0.024256547 | -1.615171014 | FALSE | FALSE | FALSE |
| SFTPD | 0.0405224 | 0.87836897 | 0.0099 | 0.040522376 | 0.040522376 | 0.040522376 | 2.08843066 | 0.481518826 | 0.2544 | 0.522393028 | 2.088430662 | 2.088430662 | 0.019403266 | -1.712125159 | FALSE | FALSE | FALSE |
| NR1D2 | 0.0404487 | 0.87837792 | 0.5029 | 0.040448742 | 0.040448742 | 0.200353644 | 0.0170888 | 0.894556814 | 1 | 0.159975146 | 0.159975146 | 0.338595483 | 2.36697444 | 0.374193568 | FALSE | TRUE | FALSE |

| | | | | | | | | | | | | | | | | | |
|---|---|---|---|---|---|---|---|---|---|---|---|---|---|---|---|---|---|
| SEMA5A | 0.0404445 | 0.87838687 | 1 | 0.04044447 | 0.04044447 | 1.363191054 | 0.05886107 | 0.884911822 | 1 | 0.058861072 | 0.058861072 | 1.149439353 | 0.687117451 | -0.162969021 | FALSE | TRUE | FALSE |
| BMPER | 0.0404194 | 0.87839581 | 0.02165 | 0.040419351 | 0.040419351 | 0.795919843 | 0.05897957 | 0.88488639 | 1 | 0.058979569 | 0.058979569 | 1.16355535 | 0.685311054 | -0.164112263 | FALSE | TRUE | FALSE |
| EZH1 | 0.0404179 | 0.87840476 | 1 | 0.161579569 | 0.161579569 | 1.365574453 | 0.05832274 | 0.884996531 | 0.75 | 0.233172778 | 0.932451615 | 4.603311585 | 0.693004683 | -0.159263831 | FALSE | TRUE | FALSE |
| CDT1 | 0.040408 | 0.8784137 | 1 | 0.040408015 | 0.040408015 | 0.795114209 | 0.49566358 | 0.756538784 | 0.7536 | 0.495663575 | 4.571339148 | 9.772195142 | 0.081523067 | -1.08871949 | FALSE | TRUE | FALSE |
| PIGW | 0.0404054 | 0.87842265 | 1 | 0.359900843 | 0.359900843 | 0.801333545 | 0.05679026 | 0.885334249 | 1 | 0.509085288 | 4.683920547 | 4.683920547 | 0.711484036 | -0.14783484 | FALSE | TRUE | FALSE |
| GSTCD | 0.040389 | 0.87843159 | 0.51115 | 0.364909744 | 0.364909744 | 0.800649031 | 0.53339484 | 0.741273093 | 0.50555 | 0.53339484 | 0.53339484 | 1.164923971 | 0.075720727 | -1.120785223 | FALSE | TRUE | FALSE |
| RSC1A1 | 0.0403626 | 0.87844053 | 1 | 0.353009358 | 0.353009358 | 1.366512701 | 4.5064219 | 0.329708924 | 0.2542 | 0.497820318 | 4.506421897 | 9.864282361 | 0.008956695 | -2.0478522 | FALSE | FALSE | FALSE |
| VILL | 0.0402808 | 0.87844947 | 0.97025 | 3.073830169 | 27.87800401 | 259.0330603 | 0.59503732 | 0.71433121 | 0.9636 | 48.80560408 | 4409.815582 | 43405.64576 | 0.067694641 | -1.16944571 | FALSE | TRUE | FALSE |
| HSD17B6 | 0.040253 | 0.87845841 | 0.98385 | 13.68398504 | 524.3393148 | 2097.350938 | 78.7980066 | 0.037126572 | 0.34635 | 19.70324334 | 758.0238049 | 7474.616747 | 0.000510837 | -3.291717397 | FALSE | FALSE | FALSE |
| PRB4 | 0.0402242 | 0.87846735 | 0.75225 | 0.349769503 | 3.121711517 | 3.121711517 | 0.05952271 | 0.884801543 | 1 | 0.059522707 | 0.523163003 | 0.523163003 | 0.675778783 | -0.170195447 | FALSE | TRUE | FALSE |
| PBXIP1 | 0.0402162 | 0.87847629 | 1 | 0.040216183 | 0.040216183 | 0.792395949 | 0.05801966 | 0.885047304 | 0.01895 | 0.058019657 | 0.058019657 | 1.145225525 | 0.69314755 | -0.159174308 | FALSE | TRUE | FALSE |
| ZNF16 | 0.0402144 | 0.87848522 | 1 | 0.040214433 | 0.040214433 | 0.793702777 | 0.05814586 | 0.885021923 | 1 | 0.05814586 | 0.05814586 | 1.149428734 | 0.691613005 | -0.160136849 | FALSE | TRUE | FALSE |
| SLC5A2 | 0.0401703 | 0.87849416 | 0.51105 | 0.040170332 | 0.641023807 | 3.143317882 | 0.00094903 | 0.899495173 | 0.26775 | 0.000949031 | 0.003789753 | 0.018638114 | 42.32773358 | 1.626625015 | FALSE | TRUE | FALSE |
| NLRP3 | 0.0401547 | 0.87850309 | 1 | 0.370319899 | 0.370319899 | 7.247860527 | 0.06047624 | 0.884631529 | 0.02595 | 0.060476243 | 0.060476243 | 1.186686268 | 0.66397538 | -0.177848024 | FALSE | TRUE | FALSE |
| SRRM4 | 0.0401293 | 0.87851202 | 0.01975 | 0.040129315 | 0.040129315 | 0.790172929 | 0.04441911 | 0.888021852 | 1 | 0.044419113 | 0.044419113 | 0.876270235 | 0.903424495 | -0.044108138 | FALSE | TRUE | FALSE |
| KLK9 | 0.040106 | 0.87852095 | 1 | 3.439777038 | 326.522013 | 326.522013 | 0.05500177 | 0.885645077 | 1 | 45.74994071 | 4583.831261 | 47546.64 | 0.729176533 | -0.137167317 | FALSE | TRUE | FALSE |
| CHAT | 0.0400806 | 0.87852988 | 0.89535 | 0.63984696 | 10.22473398 | 40.88864382 | 4.87452857 | 0.312233549 | 0.2731 | 1.219368251 | 19.49121286 | 77.95672562 | 0.008222459 | -2.084998275 | FALSE | FALSE | FALSE |
| WDHD1 | 0.0400109 | 0.87853881 | 0.815 | 0.349826809 | 12.53270626 | 115.0821839 | 0.0124734 | 0.895908619 | 0.2703 | 0.0124734 | 0.049861561 | 0.245205787 | 3.207701694 | 0.506193973 | FALSE | TRUE | FALSE |
| TM6SF2 | 0.04 | 0.87854774 | 1 | 0.040000028 | 0.040000028 | 0.040000028 | 0.42177003 | 0.783821816 | 0.5088 | 0.421770029 | 0.421770029 | 0.421770029 | 0.094838478 | -1.023015424 | FALSE | TRUE | FALSE |
| RASEF | 0.0399902 | 0.87855666 | 1 | 0.159896304 | 0.159896304 | 3.154387271 | 0.05545611 | 0.88552764 | 1 | 0.505432824 | 4.727962859 | 9.9873668 | 0.72111423 | -0.141995934 | FALSE | TRUE | FALSE |
| AARS2 | 0.0399241 | 0.87856559 | 0.5165 | 0.086345719 | 1.38014479 | 5.519875799 | 0.00632106 | 0.897778424 | 0.02545 | 0.006321063 | 0.006321063 | 0.123736174 | 6.31604318 | 0.80044509 | FALSE | TRUE | FALSE |
| SIX4 | 0.0398733 | 0.87857451 | 0.5083 | 0.091459352 | 0.365487392 | 1.348669022 | 0.01790052 | 0.894361069 | 1 | 0.017900522 | 0.017900522 | 0.352868523 | 2.227491641 | 0.347816083 | FALSE | TRUE | FALSE |
| SETD4 | 0.0398428 | 0.87858343 | 1 | 0.350668838 | 3.167793602 | 3.167793602 | 0.05930083 | 0.884835495 | 0.50615 | 0.526869233 | 0.526869233 | 0.526869233 | 0.671875078 | -0.172711468 | FALSE | TRUE | FALSE |
| ICOS | 0.0398384 | 0.87859235 | 1 | 0.03983837 | 0.03983837 | 0.03983837 | 0.0423474 | 0.888472051 | 0.00435 | 0.042347403 | 0.042347403 | 0.042347403 | 0.940751197 | -0.02652522 | FALSE | TRUE | FALSE |
| BCL2 | 0.0398316 | 0.87860127 | 1 | 0.039831557 | 0.039831557 | 0.039831557 | 0.03268192 | 0.890644109 | 1 | 293.2646751 | 32223.46763 | 349506.1145 | 1.218764288 | 0.08591972 | FALSE | TRUE | FALSE |
| STARD3 | 0.0398305 | 0.87861019 | 1 | 0.039830503 | 0.039830503 | 0.78729192 | 0.23033758 | 0.84477225 | 0.50415 | 0.230337576 | 0.230337576 | 1.140511553 | 0.1729223 | -0.762148997 | FALSE | TRUE | FALSE |
| ZNF610 | 0.0398206 | 0.87861911 | 1 | 0.039820601 | 0.039820601 | 0.789305495 | 0.05760979 | 0.885140283 | 1 | 0.057609792 | 0.057609792 | 1.14314012 | 0.691212372 | -0.160388497 | FALSE | TRUE | FALSE |
| HEATR1 | 0.0397324 | 0.87862802 | 1 | 0.039732365 | 0.765799606 | 1.330099597 | 0.52250632 | 0.745796223 | 0.5248 | 0.522506317 | 0.522506317 | 10.11487709 | 0.076041884 | -1.118947132 | FALSE | TRUE | FALSE |
| WDR89 | 0.0397255 | 0.87863694 | 1 | 2656.792741 | 2784095.315 | 29573535.88 | 0.0645356 | 0.883679593 | 1 | 48.81588041 | 4591.474139 | 46146.21252 | 0.61555887 | -0.210730406 | FALSE | TRUE | FALSE |
| ANKRD12 | 0.0396527 | 0.87864585 | 0.7659 | 0.357055786 | 3.299914653 | 15.98354034 | 0.16074972 | 0.861893477 | 0.5199 | 0.160749722 | 0.160749722 | 3.12472282 | 0.246673543 | -0.607877428 | FALSE | TRUE | FALSE |
| SLC38A2 | 0.0396427 | 0.87865476 | 1 | 0.039642719 | 0.039642719 | 0.785723382 | 0.04296436 | 0.888335729 | 1 | 0.042964357 | 0.042964357 | 0.852479825 | 0.922688516 | -0.034944885 | FALSE | TRUE | FALSE |
| STAT2 | 0.0394964 | 0.87866367 | 1 | 0.039496386 | 0.039496386 | 0.777447089 | 0.05781428 | 0.885089583 | 1 | 0.057814281 | 0.057814281 | 1.140180991 | 0.683159682 | -0.165477772 | FALSE | TRUE | FALSE |
| WDR83 | 0.0394874 | 0.87867258 | 1 | 0.157896429 | 0.631530991 | 1.338976321 | 0.05745266 | 0.8851825 | 1 | 0.229742403 | 0.918926449 | 1.139748222 | 0.687303227 | -0.162851617 | FALSE | TRUE | FALSE |
| VTCN1 | 0.0394711 | 0.87868149 | 0.00485 | 0.039471077 | 0.039471077 | 0.039471077 | 0.22488317 | 0.846143773 | 0.50255 | 0.22488317 | 0.22488317 | 0.22488317 | 0.175518145 | -0.75567798 | FALSE | TRUE | FALSE |
| PTPRM | 0.039468 | 0.8786904 | 0.0434 | 0.039467998 | 0.039467998 | 1.325396217 | 0.52331758 | 0.74534726 | 0.52145 | 0.523317576 | 0.523317576 | 10.18154689 | 0.075418827 | -1.122520225 | FALSE | TRUE | FALSE |
| RASIP1 | 0.0394064 | 0.87869931 | 0.0238 | 0.039406377 | 0.039406377 | 0.77626809 | 0.04427745 | 0.888062179 | 1 | 0.044277451 | 0.044277451 | 0.873803356 | 0.889987471 | -0.050616107 | FALSE | TRUE | FALSE |
| DCST1 | 0.0393835 | 0.87870821 | 1 | 0.039383452 | 0.039383452 | 0.773191733 | 0.4921898 | 0.75798107 | 0.5082 | 0.492189798 | 0.492189798 | 9.684252924 | 0.080016798 | -1.096818831 | FALSE | TRUE | FALSE |
| GEMIN4 | 0.0393489 | 0.87871712 | 0.99625 | 26.17064219 | 2253.024322 | 21647.64669 | 0.01075277 | 0.896555114 | 0.0276 | 0.010752773 | 0.010752773 | 0.210679446 | 3.65941979 | 0.563412232 | FALSE | TRUE | FALSE |
| GPATCH1 | 0.0392655 | 0.87872602 | 1 | 0.039265471 | 0.039265471 | 0.770806876 | 0.05693307 | 0.885292133 | 0.0225 | 0.056933068 | 0.056933068 | 1.120118612 | 0.689677768 | -0.161353774 | FALSE | TRUE | FALSE |
| CCT3 | 0.0392625 | 0.87873492 | 0.0174 | 0.039262502 | 0.039262502 | 0.775566229 | 0.05762978 | 0.885123388 | 1 | 0.057629784 | 0.057629784 | 1.140070881 | 0.681288379 | -0.166669019 | FALSE | TRUE | FALSE |
| USP50 | 0.039261 | 0.87874382 | 1 | 0.34205667 | 3.058791015 | 3.058791015 | 0.05663437 | 0.885367921 | 0.7486 | 0.498212144 | 4.498388807 | 4.498388807 | 0.693236976 | -0.159118281 | FALSE | TRUE | FALSE |
| ATP1A2 | 0.0392026 | 0.87875272 | 0.94185 | 0.156695932 | 2.505349856 | 12.2180951 | 0.06844359 | 0.882774565 | 0.87755 | 0.273606964 | 1.094032049 | 5.352062856 | 0.572772814 | -0.242017603 | FALSE | TRUE | FALSE |
| ACSF2 | 0.0391868 | 0.87876162 | 1 | 0.03918682 | 0.03918682 | 0.773321191 | 0.05705049 | 0.88526685 | 0.0145 | 0.057050487 | 0.057050487 | 1.127647093 | 0.686879673 | -0.163119335 | FALSE | TRUE | FALSE |
| PLA2G4D | 0.0391613 | 0.87877052 | 0.7581 | 0.345148217 | 3.122233005 | 6.787543072 | 0.06019966 | 0.88470809 | 1 | 0.060199658 | 0.060199658 | 1.186244778 | 0.650524058 | -0.186736637 | FALSE | TRUE | FALSE |
| KCNB2 | 0.0391486 | 0.87877941 | 1 | 0.039148554 | 0.039148554 | 0.76820228 | 0.05747414 | 0.885174059 | 0.02215 | 0.057474136 | 0.057474136 | 1.130362245 | 0.681150807 | -0.166756724 | FALSE | TRUE | FALSE |
| INF2 | 0.0391342 | 0.8787883 | 1 | 0.039134224 | 0.039134224 | 0.760982696 | 0.05557716 | 0.885494045 | 0.5167 | 0.514003975 | 0.514003975 | 10.0284476 | 0.704142174 | -0.152339643 | FALSE | TRUE | FALSE |
| F7 | 0.0390525 | 0.8787972 | 0.50575 | 0.039052548 | 0.039052548 | 0.193184528 | 1.20545228 | 0.566585014 | 0.85035 | 19.2621422 | 1797.973041 | 8903.387999 | 0.032396594 | -1.489500647 | FALSE | FALSE | FALSE |
| USP13 | 0.039036 | 0.87880609 | 0.02385 | 0.039035952 | 0.039035952 | 0.767746772 | 0.48694232 | 0.760273548 | 0.6325 | 0.863346424 | 7.789552004 | 9.596184589 | 0.080165453 | -1.096012747 | FALSE | TRUE | FALSE |
| CRNKL1 | 0.0390293 | 0.87881498 | 0.7531 | 0.350480285 | 3.230286078 | 11.81886429 | 0.06041321 | 0.884640041 | 0.0233 | 0.06041321 | 0.06041321 | 1.188535842 | 0.646038996 | -0.189741266 | FALSE | TRUE | FALSE |
| GLYAT | 0.0389913 | 0.87882387 | 1 | 0.03899128 | 0.03899128 | 0.03899128 | 0.04315051 | 0.88827951 | 1 | 0.04315051 | 0.04315051 | 0.04315051 | 0.903611112 | -0.044018437 | FALSE | TRUE | FALSE |
| C1orf210 | 0.0389726 | 0.87883276 | 1 | 0.354294429 | 0.354294429 | 0.354294429 | 0.04831756 | 0.887045894 | 0.0026 | 0.048317557 | 0.048317557 | 0.048317557 | 0.806593802 | -0.093345119 | FALSE | TRUE | FALSE |
| CTTNBP2 | 0.0389284 | 0.87884165 | 0.27525 | 0.004437807 | 0.038928411 | 0.755421894 | 0.01917592 | 0.894047981 | 0.8195 | 0.092118053 | 1.793996714 | 7.169692552 | 2.030067553 | 0.30751049 | FALSE | TRUE | FALSE |
| CHRNA4 | 0.0389029 | 0.87885053 | 0.87385 | 0.155502682 | 0.621731353 | 3.052480579 | 0.015974 | 0.894874381 | 1 | 0.586026937 | 22.07110731 | 22.07110731 | 2.435386451 | 0.386567886 | FALSE | TRUE | FALSE |
| JAKMIP1 | 0.0389001 | 0.87885942 | 0.0268 | 0.038900133 | 0.038900133 | 0.762569793 | 0.04331208 | 0.888247361 | 1 | 0.043312076 | 0.043312076 | 0.851086657 | 0.898135957 | -0.046657916 | FALSE | TRUE | FALSE |
| ZNF571 | 0.0388912 | 0.8788683 | 0.8774 | 0.33204263 | 2.907908278 | 2.907908278 | 0.07602202 | 0.881249387 | 0.01295 | 0.076022025 | 0.076022025 | 0.178188566 | 0.511578714 | -0.291087534 | FALSE | TRUE | FALSE |
| RBP2 | 0.0388851 | 0.87887718 | 0.0037 | 0.038885083 | 0.038885083 | 0.038885083 | 0.04121935 | 0.888664006 | 1 | 0.041219354 | 0.041219354 | 0.041219354 | 0.943369546 | -0.025318148 | FALSE | TRUE | FALSE |
| ASAH1 | 0.0388552 | 0.87888606 | 1 | 0.352843414 | 3.288629021 | 3.288629021 | 0.51838099 | 0.747975813 | 0.7535 | 0.518380992 | 4.856246565 | 4.856246565 | 0.074954919 | -1.125199863 | FALSE | TRUE | FALSE |
| LIMK2 | 0.0388324 | 0.87889494 | 0.01875 | 0.038832432 | 0.038832432 | 0.76547838 | 0.01673024 | 0.894665271 | 1 | 0.066870659 | 0.066870659 | 0.330363235 | 2.321091852 | 0.365692327 | FALSE | TRUE | FALSE |
| NCOA6 | 0.0388013 | 0.87890382 | 0.05015 | 0.03880132 | 0.747605394 | 1.298539069 | 0.05001158 | 0.886634685 | 1 | 0.050011577 | 0.050011577 | 1.676867627 | 0.775846776 | -0.11022404 | FALSE | TRUE | FALSE |
| UGT1A3 | 0.0387757 | 0.8789127 | 0.75665 | 0.343701682 | 3.126891143 | 6.79911421 | 4.472066 | 0.331329387 | 0.1345 | 4.472065996 | 42.02023049 | 42.02023049 | 0.008670651 | -2.061948301 | FALSE | FALSE | FALSE |
| ACP2 | 0.0387746 | 0.87892157 | 0.01505 | 0.038774553 | 0.038774553 | 0.766587086 | 0.0545727 | 0.885770667 | 1 | 0.483659927 | 0.483659927 | 1.08041253 | 0.710511963 | -0.148428605 | FALSE | TRUE | FALSE |
| TBC1D17 | 0.0387629 | 0.87893045 | 0.75515 | 0.341957821 | 3.096275284 | 6.732609815 | 0.5003478 | 0.755003117 | 0.75625 | 0.5003478 | 4.568173815 | 9.869531534 | 0.077471966 | -1.110855422 | FALSE | TRUE | FALSE |
| SIKE1 | 0.0387521 | 0.87893932 | 1 | 0.349082163 | 0.349082163 | 0.349082163 | 0.05013295 | 0.886576893 | 0.00585 | 0.050132952 | 0.050132952 | 0.050132952 | 0.77298689 | -0.111827872 | FALSE | TRUE | FALSE |
| RBM34 | 0.0387464 | 0.87894819 | 1 | 0.038746398 | 0.038746398 | 0.767784001 | 0.48834925 | 0.759550753 | 0.50615 | 0.488349252 | 0.488349252 | 1.084591004 | 0.079341574 | -1.100499186 | FALSE | TRUE | FALSE |
| RPL3L | 0.0386919 | 0.87895706 | 0.2629 | 0.038691936 | 0.154748937 | 0.764238735 | 0.01633494 | 0.894780735 | 1 | 0.14280568 | 0.14280568 | 0.323127395 | 2.368661791 | 0.374503054 | FALSE | TRUE | FALSE |
| GSDMD | 0.038683 | 0.87896593 | 1 | 0.350670076 | 3.26270473 | 6.937595648 | 0.44098513 | 0.777101016 | 0.75145 | 0.440985129 | 4.124825378 | 4.124825378 | 0.087719568 | -1.056903516 | FALSE | TRUE | FALSE |
| SLC52A1 | 0.0386824 | 0.8789748 | 0.0112 | 0.038682392 | 0.038682392 | 0.766776446 | 0.05639663 | 0.885401577 | 0.00995 | 0.056396632 | 0.056396632 | 0.056396632 | 0.685898976 | -0.163739846 | FALSE | TRUE | FALSE |
| MARCO | 0.0386087 | 0.87898367 | 0.75375 | 0.038608702 | 0.344016294 | 0.764093013 | 0.00844415 | 0.897194251 | 1 | 0.074108769 | 0.296262127 | 0.668913274 | 4.572243518 | 0.660129353 | FALSE | TRUE | FALSE |
| UPF1 | 0.0385333 | 0.87899253 | 1 | 0.154026283 | 0.154026283 | 2.98818695 | 0.05730261 | 0.885207817 | 1 | 0.057302613 | 0.057302613 | 1.115717433 | 0.672451977 | -0.172338725 | FALSE | TRUE | FALSE |
| DHX34 | 0.038518 | 0.8790014 | 0.2796 | 0.038517971 | 0.154041017 | 1.297846635 | 0.07057093 | 0.882308396 | 0.28145 | 0.070570929 | 0.621444704 | 12.08983791 | 0.545805066 | -0.262962438 | FALSE | TRUE | FALSE |
| TTYH1 | 0.0385153 | 0.87901026 | 1 | 0.03851533 | 0.03851533 | 0.763874179 | 0.04229653 | 0.88848006 | 1 | 0.393364664 | 0.393364664 | 0.839709074 | 0.91060266 | -0.040671085 | FALSE | TRUE | FALSE |
| PTPN7 | 0.0385083 | 0.87901912 | 1 | 0.03850827 | 0.03850827 | 0.763056421 | 4.13212708 | 0.347170734 | 0.50535 | 4.132127084 | 38.579772 | 38.579772 | 0.009319237 | -2.030619663 | FALSE | FALSE | FALSE |
| ME3 | 0.0384784 | 0.87902799 | 1 | 0.153862895 | 0.61540258 | 3.033463185 | 0.05684331 | 0.885308983 | 1 | 0.227307974 | 0.227307974 | 1.924620789 | 0.676920575 | -0.169462285 | FALSE | TRUE | FALSE |

| | | | | | | | | | | | | | | | | | |
|---|---|---|---|---|---|---|---|---|---|---|---|---|---|---|---|---|---|
| TGM7 | 0.0384562 | 0.87903685 | 0.90805 | 0.614349039 | 22.05119625 | 88.19166389 | 0.35684549 | 0.806805091 | 0.35955 | 0.356845486 | 5.705139504 | 22.82033592 | 0.107767231 | -0.967513275 | FALSE | TRUE | FALSE |
| EXOC3L2 | 0.0384494 | 0.8790457 | 1 | 0.038449358 | 0.038449358 | 0.760123987 | 0.0412974 | 0.888648032 | 1 | 0.041297399 | 0.041297399 | 0.817560417 | 0.931035821 | -0.031033609 | FALSE | TRUE | FALSE |
| ANTXR2 | 0.0384282 | 0.87905456 | 1 | 0.344433425 | 0.344433425 | 0.761761269 | 0.05380952 | 0.885912676 | 0.8795 | 1.939650411 | 17.94533764 | 17.94533764 | 0.714151792 | -0.146209469 | FALSE | TRUE | FALSE |
| TAS2R1 | 0.0383841 | 0.87906342 | 1 | 0.038384125 | 0.038384125 | 0.038384125 | 0.04307755 | 0.888319672 | 1 | 0.043077546 | 0.043077546 | 0.043077546 | 0.891047176 | -0.050099302 | FALSE | TRUE | FALSE |
| RGS9 | 0.0383832 | 0.87907227 | 0.0217 | 0.038383214 | 0.038383214 | 0.756277431 | 0.05575271 | 0.885468837 | 0.01955 | 0.05575271 | 0.05575271 | 1.100475659 | 0.688454675 | -0.162124646 | FALSE | TRUE | FALSE |
| NUDT16L1 | 0.0382761 | 0.87908113 | 1 | 0.038276081 | 0.038276081 | 0.038276081 | 0.04212853 | 0.888520093 | 1 | 0.042128525 | 0.042128525 | 0.042128525 | 0.908554976 | -0.041648789 | FALSE | TRUE | FALSE |
| TUBG2 | 0.0382659 | 0.87908998 | 0.0137 | 0.038265852 | 0.038265852 | 0.756745216 | 17.5080369 | 0.145720608 | 0.19585 | 1.875095255 | 70.022147 | 70.022147 | 0.002185616 | -2.66042607 | FALSE | FALSE | FALSE |
| IRX6 | 0.0382222 | 0.87909883 | 0.9101 | 0.61046336 | 13.26498922 | 53.01918926 | 0.07925064 | 0.880545961 | 0.5104 | 0.079250641 | 0.079250641 | 0.296877354 | 0.482295291 | -0.316686979 | FALSE | TRUE | FALSE |
| PALMD | 0.0381576 | 0.87910768 | 1 | 0.038157603 | 0.038157603 | 0.756324989 | 0.40193557 | 0.791601384 | 0.0111 | 0.401935566 | 0.401935566 | 0.864266671 | 0.094934627 | -1.022575354 | FALSE | TRUE | FALSE |
| SHROOM3 | 0.0381006 | 0.87911653 | 1 | 0.038100558 | 0.732216492 | 1.277729838 | 1.9506754 | 0.493782956 | 0.2703 | 0.487738475 | 1.950675398 | 37.66501521 | 0.019531983 | -1.709253667 | FALSE | FALSE | FALSE |
| FAM131C | 0.0380462 | 0.87912538 | 1 | 0.038046167 | 0.038046167 | 0.038046167 | 0.04043378 | 0.88886334 | 1 | 0.04043378 | 0.04043378 | 0.04043378 | 0.940950041 | -0.026433435 | FALSE | TRUE | FALSE |
| RECQL | 0.038006 | 0.87913423 | 0.8166 | 3.124348576 | 29.40935851 | 283.6031175 | 0.71281724 | 0.659591547 | 0.2571 | 0.078665415 | 0.712817245 | 1.557317434 | 0.053318058 | -1.273125679 | FALSE | TRUE | FALSE |
| RABL3 | 0.0379909 | 0.87914307 | 1 | 0.037990895 | 0.151833828 | 0.151833828 | 0.05530043 | 0.885577997 | 1 | 0.221034287 | 0.883688766 | 0.883688766 | 0.686990939 | -0.163048991 | FALSE | TRUE | FALSE |
| EIF2AK4 | 0.0379768 | 0.87915192 | 0.51945 | 0.037976778 | 0.322985916 | 1.560502354 | 0.33639164 | 0.813373953 | 0.1471 | 0.039962089 | 0.336391642 | 2.825236147 | 0.112894535 | -0.94732708 | FALSE | TRUE | FALSE |
| SEPSECS | 0.0379572 | 0.87916076 | 0.75445 | 0.037957158 | 0.151740329 | 0.188484455 | 0.14248991 | 0.86628343 | 0.75275 | 0.142489912 | 1.321973155 | 1.321973155 | 0.266384882 | -0.574490427 | FALSE | TRUE | FALSE |
| UBXN11 | 0.0379458 | 0.8791696 | 0.99195 | 24.88051544 | 2121.749224 | 4485.013129 | 0.00932331 | 0.896938071 | 1 | 0.676594873 | 5.989702946 | 5.989702946 | 4.069994868 | 0.609593862 | FALSE | TRUE | FALSE |
| COG4 | 0.0379057 | 0.87917844 | 1 | 0.037905715 | 0.037905715 | 0.745284665 | 0.05528275 | 0.885586386 | 1 | 0.055282745 | 0.055282745 | 1.089158308 | 0.685669908 | -0.16388491 | FALSE | TRUE | FALSE |
| PTPRO | 0.0378344 | 0.87918728 | 1 | 0.15131417 | 0.15131417 | 2.957670945 | 0.22110149 | 0.846972818 | 0.51765 | 0.221101493 | 0.221101493 | 4.333577181 | 0.171117794 | -0.766704828 | FALSE | TRUE | FALSE |
| MICALL1 | 0.0378227 | 0.87919612 | 0.75595 | 0.341053304 | 1.363757928 | 6.713607459 | 0.05373201 | 0.885937706 | 1 | 1.947224413 | 18.11195791 | 18.11195791 | 0.703913016 | -0.152481004 | FALSE | TRUE | FALSE |
| ESYT1 | 0.0377863 | 0.87920496 | 0.9425 | 0.604359694 | 21.1686753 | 84.71209948 | 0.00519463 | 0.898085243 | 0.51 | 0.020764102 | 0.020764102 | 0.406507913 | 7.274110076 | 0.861779869 | FALSE | TRUE | FALSE |
| RNF10 | 0.0377256 | 0.87921379 | 0.754 | 0.037725604 | 0.150790748 | 0.741562532 | 0.0165397 | 0.894730247 | 1 | 0.06610127 | 0.06610127 | 1.302020514 | 2.280912504 | 0.358108626 | FALSE | TRUE | FALSE |
| KRIT1 | 0.0377022 | 0.87922263 | 1 | 0.03770222 | 0.03770222 | 0.744265269 | 0.0555163 | 0.885510845 | 1 | 0.0555163 | 0.0555163 | 1.097636582 | 0.679119813 | -0.168053599 | FALSE | TRUE | FALSE |
| SLC27A4 | 0.0376848 | 0.87923146 | 1 | 0.150662052 | 0.150662052 | 2.958662221 | 0.05538574 | 0.885561216 | 0.5072 | 0.221443001 | 0.221443001 | 1.875811461 | 0.680405972 | -0.167231883 | FALSE | TRUE | FALSE |
| PLEKHG3 | 0.0376824 | 0.8792403 | 0.0387 | 0.03768245 | 0.03768245 | 0.731974262 | 0.07224252 | 0.88197208 | 0.5157 | 0.072242523 | 0.072242523 | 1.408162624 | 0.521610379 | -0.282653776 | FALSE | TRUE | FALSE |
| USH1C | 0.037641 | 0.87924913 | 1 | 0.333055382 | 1.332134749 | 6.521910153 | 152.746725 | 0.018254041 | 0.0743 | 16.30995247 | 366.6095037 | 1466.353752 | 0.000246427 | -3.608310949 | FALSE | FALSE | FALSE |
| GBP4 | 0.0375792 | 0.87925796 | 0.75485 | 0.037579238 | 0.327988755 | 0.742975796 | 0.07250819 | 0.881909938 | 0.0146 | 0.072508187 | 0.072508187 | 0.166622464 | 0.518275787 | -0.285439081 | FALSE | TRUE | FALSE |
| PSAT1 | 0.037478 | 0.87926679 | 0.75665 | 0.149817363 | 0.599041469 | 0.599041469 | 0.21137165 | 0.849212845 | 0.9381 | 1.947518227 | 31.14876853 | 31.14876853 | 0.177308368 | -0.751270766 | FALSE | TRUE | FALSE |
| XKR3 | 0.0374541 | 0.87927562 | 1 | 0.037454131 | 0.037454131 | 0.743142554 | 0.04459178 | 0.887973423 | 1 | 0.044591779 | 0.044591779 | 0.044591779 | 0.839933548 | -0.075755072 | FALSE | TRUE | FALSE |
| BEND7 | 0.0374031 | 0.87928444 | 0.01285 | 0.03740307 | 0.03740307 | 0.741115873 | 0.04477321 | 0.887916875 | 0.01265 | 0.044773212 | 0.044773212 | 0.888140843 | 0.835389478 | -0.078110999 | FALSE | TRUE | FALSE |
| PCDHGC5 | 0.0374023 | 0.87929327 | 1 | 0.037402264 | 0.037402264 | 0.731576356 | 0.05491279 | 0.885653457 | 1 | 0.054912794 | 0.054912794 | 1.076925317 | 0.681121119 | -0.166775654 | FALSE | TRUE | FALSE |
| TP53BP1 | 0.0373985 | 0.87930209 | 1 | 0.037398508 | 0.037398508 | 1.251986003 | 0.05432917 | 0.885820818 | 1 | 0.217273614 | 0.217273614 | 4.2147897 | 0.688368831 | -0.162178803 | FALSE | TRUE | FALSE |
| PZP | 0.037396 | 0.87931092 | 0.65505 | 0.304727676 | 21.75002692 | 190.3001336 | 0.01514389 | 0.895089908 | 0.6926 | 0.015143893 | 1.035829863 | 2.41731134 | 2.469375565 | 0.392587146 | FALSE | TRUE | FALSE |
| TRAF3IP3 | 0.0372848 | 0.87931974 | 1 | 0.037284767 | 0.037284767 | 0.738620366 | 0.05303211 | 0.886062713 | 0.50875 | 0.479612416 | 0.479612416 | 1.051778048 | 0.703060196 | -0.153007489 | FALSE | TRUE | FALSE |
| NDST4 | 0.0372836 | 0.87932856 | 1 | 0.149085633 | 0.149085633 | 2.938354091 | 0.05470303 | 0.885737195 | 1 | 0.218748795 | 0.218748795 | 1.8470589 | 0.681564437 | -0.166493079 | FALSE | TRUE | FALSE |
| WWC2 | 0.0372571 | 0.87933738 | 1 | 3.05481841 | 28.74401472 | 59.85616414 | 0.056189 | 0.885435215 | 1 | 0.506120459 | 4.679044562 | 17.08204204 | 0.663066983 | -0.178442597 | FALSE | TRUE | FALSE |
| PMM1 | 0.0372048 | 0.8793462 | 0.9998 | 342.8261624 | 72913.5759 | 292467.7824 | 0.25993013 | 0.836752771 | 0.88445 | 21.89130099 | 32640.52136 | 326852.4369 | 0.143133771 | -0.844257886 | FALSE | TRUE | FALSE |
| SCGB1D1 | 0.0372001 | 0.87935502 | 1 | 0.03720011 | 0.03720011 | 0.03720011 | 0.03957351 | 0.889054089 | 1 | 0.039573508 | 0.039573508 | 0.039573508 | 0.940025594 | -0.026860322 | FALSE | TRUE | FALSE |
| SLC38A1 | 0.0371887 | 0.87936383 | 1 | 0.037188748 | 0.037188748 | 0.736759253 | 0.36606728 | 0.80371057 | 0.50195 | 0.366067281 | 0.366067281 | 0.772148184 | 0.101589927 | -0.99314935 | FALSE | TRUE | FALSE |
| AOC2 | 0.0371379 | 0.87937265 | 0.50875 | 0.037137925 | 0.037137925 | 0.730868075 | 0.00751609 | 0.897449274 | 1 | 0.545624137 | 43.84840121 | 43.84840121 | 4.941121343 | 0.693825519 | FALSE | TRUE | FALSE |
| SULT6B1 | 0.0371231 | 0.87938146 | 1 | 0.037123078 | 0.037123078 | 0.037123078 | 0.05426474 | 0.885837529 | 1 | 0.054264744 | 0.054264744 | 0.054264744 | 0.684110443 | -0.16487378 | FALSE | TRUE | FALSE |
| RNASEH2B | 0.0371088 | 0.87939027 | 0.87325 | 0.14833093 | 2.371745044 | 2.371745044 | 0.22353472 | 0.846479331 | 0.0039 | 0.055917857 | 0.223534717 | 0.223534717 | 0.166008919 | -0.779868578 | FALSE | TRUE | FALSE |
| HOXD3 | 0.03707 | 0.87939908 | 1 | 0.037069966 | 0.037069966 | 0.73236541 | 0.05477927 | 0.885703714 | 1 | 0.054779273 | 0.054779273 | 1.083822473 | 0.676715192 | -0.169594074 | FALSE | TRUE | FALSE |
| RB1CC1 | 0.0369918 | 0.87940789 | 1 | 0.036991786 | 0.036991786 | 1.241721611 | 0.04543366 | 0.887730698 | 1 | 0.045433661 | 0.045433661 | 1.527428593 | 0.814193395 | -0.089272425 | FALSE | TRUE | FALSE |
| SV2C | 0.0369497 | 0.8794167 | 0.5128 | 0.147735541 | 0.147735541 | 2.909980347 | 0.81529796 | 0.615596056 | 0.8137 | 3.261555741 | 29.76343727 | 119.0967863 | 0.045320464 | -1.343705652 | FALSE | TRUE | FALSE |
| SDS | 0.0368786 | 0.87942551 | 0.7507 | 0.083274434 | 0.332801514 | 0.332801514 | 0.06578814 | 0.88341117 | 0.0103 | 0.065788142 | 0.065788142 | 0.326811581 | 0.560566349 | -0.251372977 | FALSE | TRUE | FALSE |
| SLC37A2 | 0.0368535 | 0.87943432 | 1 | 0.03685352 | 0.03685352 | 0.72831106 | 0.05415954 | 0.885854236 | 1 | 0.054159544 | 0.054159544 | 1.071848626 | 0.680462157 | -0.167196023 | FALSE | TRUE | FALSE |
| AHCTF1 | 0.0368531 | 0.87944312 | 1 | 0.036853124 | 0.708995715 | 1.230049279 | 0.05255949 | 0.886120971 | 1 | 0.479261584 | 0.479261584 | 9.262610141 | 0.701169752 | -0.154176827 | FALSE | TRUE | FALSE |
| TMCO6 | 0.0367907 | 0.87945193 | 0.012 | 0.036790656 | 0.036790656 | 0.72908177 | 0.05345953 | 0.885962727 | 0.01075 | 0.053459526 | 0.053459526 | 1.060575195 | 0.688196451 | -0.162287571 | FALSE | TRUE | FALSE |
| IER3 | 0.0367735 | 0.87946073 | 0.5031 | 0.346782572 | 0.346782572 | 0.346782572 | 0.03971267 | 0.888998514 | 0.0055 | 0.039712675 | 0.039712675 | 0.039712675 | 0.925988674 | -0.033394325 | FALSE | TRUE | FALSE |
| XPA | 0.0367192 | 0.87946953 | 1 | 0.322957053 | 1.291631132 | 1.291631132 | 0.79894777 | 0.621174322 | 0.77915 | 7.075742425 | 149.9008676 | 599.6704299 | 0.045959467 | -1.337625013 | FALSE | TRUE | FALSE |
| MMP20 | 0.03671 | 0.87947834 | 0.01225 | 0.036710024 | 0.036710024 | 0.727073628 | 0.21226498 | 0.849043863 | 0.01165 | 0.212264983 | 0.212264983 | 1.052437125 | 0.172944325 | -0.762093683 | FALSE | TRUE | FALSE |
| HSD3B7 | 0.036684 | 0.87948714 | 1 | 0.586379079 | 9.382401156 | 9.382401156 | 0.21851025 | 0.847520871 | 0.86945 | 0.218510246 | 3.495068195 | 3.495068195 | 0.167882438 | -0.774994733 | FALSE | TRUE | FALSE |
| GLB1L2 | 0.036675 | 0.87949593 | 0.81225 | 0.585331192 | 69.46446907 | 598.2434128 | 0.12912706 | 0.869153369 | 0.49665 | 0.111171626 | 4.616386437 | 18.44921209 | 0.284022717 | -0.546646923 | FALSE | TRUE | FALSE |
| KLHL41 | 0.0366178 | 0.87950473 | 1 | 0.036617819 | 0.036617819 | 0.724164398 | 0.05389754 | 0.88590433 | 1 | 0.053897541 | 0.053897541 | 1.067327481 | 0.679396843 | -0.167876475 | FALSE | TRUE | FALSE |
| ZNF45 | 0.036597 | 0.87951353 | 1 | 0.323533225 | 0.323533225 | 1.236549907 | 0.47156057 | 0.766613409 | 0.509 | 0.471560573 | 1.886015905 | 4.188056663 | 0.077608362 | -1.110091482 | FALSE | TRUE | FALSE |
| MMP1 | 0.0365287 | 0.87952232 | 1 | 27.5560866 | 2587.260645 | 25980.35784 | 0.06393827 | 0.883809075 | 1 | 0.06393827 | 0.06393827 | 1.268904925 | 0.571312435 | -0.243126323 | FALSE | TRUE | FALSE |
| ZNF514 | 0.0364839 | 0.87953112 | 1 | 0.320888709 | 2.896807499 | 2.896807499 | 0.45842384 | 0.771608189 | 0.8774 | 4.170643398 | 38.91878849 | 38.91878849 | 0.079585547 | -1.099165792 | FALSE | TRUE | FALSE |
| MTBP | 0.0364625 | 0.87953991 | 1 | 0.036462528 | 0.036462528 | 0.71859955 | 0.0532404 | 0.88601274 | 1 | 0.053240398 | 0.053240398 | 1.051100416 | 0.68486581 | -0.164394514 | FALSE | TRUE | FALSE |
| MUC21 | 0.0363585 | 0.8795487 | 1 | 20984881710 | 3.43656E+14 | 6.55502E+16 | 0.00742467 | 0.897456151 | 1 | 5.30531E+12 | 1.7779E+17 | 2.86566E+19 | 4.896989433 | 0.689929167 | FALSE | TRUE | FALSE |
| DCLK2 | 0.0363407 | 0.87955749 | 1 | 3.199382141 | 311.7299552 | 311.7299552 | 0.05542515 | 0.885544431 | 0.87855 | 4.916793804 | 48.11915291 | 48.11915291 | 0.655671929 | -0.183313409 | FALSE | TRUE | FALSE |
| CRELD2 | 0.0363118 | 0.87956628 | 1 | 0.145171602 | 0.145171602 | 0.719431885 | 0.21401943 | 0.848563102 | 0.50355 | 0.21401943 | 0.21401943 | 1.061753476 | 0.169665772 | -0.770405761 | FALSE | TRUE | FALSE |
| ZNF226 | 0.0362681 | 0.87957507 | 0.68915 | 0.036268144 | 2.664920194 | 2.664920194 | 6.20372527 | 0.262829819 | 0.11355 | 0.703175004 | 56.07318704 | 518.9495583 | 0.005846188 | -2.233127222 | FALSE | FALSE | FALSE |
| HOXC5 | 0.0362644 | 0.87958386 | 1 | 0.03626443 | 0.03626443 | 0.03626443 | 0.05348945 | 0.885954387 | 1 | 0.053489446 | 0.053489446 | 0.053489446 | 0.677973558 | -0.168787244 | FALSE | TRUE | FALSE |
| SERPINB2 | 0.0361955 | 0.87959264 | 1 | 0.329874836 | 0.329874836 | 0.329874836 | 0.05504765 | 0.88561993 | 1 | 0.055047652 | 0.055047652 | 0.055047652 | 0.657530005 | -0.182084424 | FALSE | TRUE | FALSE |
| AURKAIP1 | 0.0361904 | 0.87960143 | 1 | 0.036190399 | 0.036190399 | 0.036190399 | 0.04056803 | 0.888823525 | 0.00605 | 0.040568033 | 0.040568033 | 0.040568033 | 0.892091549 | -0.049590575 | FALSE | TRUE | FALSE |
| TRIM55 | 0.0361545 | 0.87961021 | 1 | 0.319440099 | 0.319440099 | 0.715816049 | 0.05081162 | 0.886444596 | 1 | 1.808617681 | 16.5211216 | 16.5211216 | 0.711540944 | -0.147800104 | FALSE | TRUE | FALSE |
| PLA2G6 | 0.0360858 | 0.87961899 | 0.7544 | 0.079696922 | 0.318610938 | 1.566347908 | 1.20319341 | 0.566706872 | 0.5078 | 1.203193412 | 11.25648286 | 23.69661184 | 0.029991707 | -1.522998813 | FALSE | FALSE | FALSE |
| BCHE | 0.0360288 | 0.87962778 | 0.9976 | 1381.534871 | 1092978.119 | 23579777.78 | 4.28596951 | 0.338799218 | 0.115 | 0.268267959 | 9.718189669 | 38.84899377 | 0.008406231 | -2.075398688 | FALSE | FALSE | FALSE |
| IBTK | 0.0360258 | 0.87963656 | 1 | 0.324670185 | 0.324670185 | 6.352395645 | 0.20977678 | 0.849745508 | 0.75395 | 0.475571265 | 1.901850666 | 9.329219168 | 0.171733948 | -0.765143846 | FALSE | TRUE | FALSE |
| ZNF671 | 0.0359328 | 0.87964533 | 0.0162 | 0.035932792 | 0.035932792 | 0.710845967 | 0.05255517 | 0.886129861 | 1 | 0.052555165 | 0.052555165 | 1.041039861 | 0.683715699 | -0.165124448 | FALSE | TRUE | FALSE |
| CYP2W1 | 0.0359309 | 0.87965411 | 1 | 0.325053952 | 1.29962399 | 1.29962399 | 0.21586827 | 0.8482077 | 0.7484 | 0.21586827 | 0.86333895 | 1.07081079 | 0.166448111 | -0.77872113 | FALSE | TRUE | FALSE |
| PLA2G4F | 0.0359079 | 0.87966289 | 0.9687 | 1.212757303 | 23.75704203 | 217.4576703 | 0.00887484 | 0.897062846 | 0.5131 | 0.074825774 | 0.074825774 | 1.473552279 | 4.046033479 | 0.607029472 | FALSE | TRUE | FALSE |
| GPBP1 | 0.035906 | 0.87967166 | 1 | 0.035906014 | 0.035906014 | 0.711404969 | 0.03896562 | 0.889188847 | 0.0123 | 0.038965622 | 0.038965622 | 0.772893254 | 0.921479297 | -0.035514418 | FALSE | TRUE | FALSE |

| | | | | | | | | | | | | | | | | |
|---|---|---|---|---|---|---|---|---|---|---|---|---|---|---|---|---|
| SLC25A21 | 0.035852 | 0.87968044 | 0.74615 | 0.143346219 | 1.290673575 | 1.290673575 | 0.05503031 | 0.885628314 | 0.00595 | 0.055030305 | 0.055030305 | 0.055030305 | 0.651495898 | -0.186088314 | FALSE | TRUE | FALSE |
| SEMA6C | 0.0358415 | 0.87968921 | 0.93875 | 2.856593524 | 252.0160752 | 252.0160752 | 4.25363863 | 0.340648582 | 0.694 | 4.253638625 | 381.6220628 | 381.6220628 | 0.008426074 | -2.074374723 | FALSE | FALSE | FALSE |
| SLC2A4 | 0.0358396 | 0.87969798 | 1 | 0.143259853 | 0.143259853 | 0.708269077 | 0.20977669 | 0.849759475 | 0.75425 | 0.209776688 | 0.838803944 | 4.151575731 | 0.170846549 | -0.76739379 | FALSE | TRUE | FALSE |
| DNER | 0.035777 | 0.87970675 | 0.87305 | 0.142977462 | 0.571530465 | 2.819136909 | 0.01532771 | 0.895054043 | 1 | 0.244803425 | 2.27840222 | 2.817582957 | 2.334141209 | 0.368127126 | FALSE | TRUE | FALSE |
| AP3M2 | 0.0357666 | 0.87971552 | 0.50885 | 0.321097015 | 0.321097015 | 0.709188058 | 0.05306291 | 0.886054387 | 1 | 0.212181192 | 0.84865516 | 0.84865516 | 0.674042247 | -0.171312883 | FALSE | TRUE | FALSE |
| RHPN1 | 0.0357247 | 0.87972429 | 1 | 0.313022482 | 2.815117115 | 6.183408449 | 4.19039665 | 0.344317054 | 0.2606 | 0.4617541 | 4.190396648 | 9.134940953 | 0.008525366 | -2.069286953 | FALSE | FALSE | FALSE |
| ST7L | 0.0357066 | 0.87973306 | 1 | 0.311764551 | 1.247020392 | 2.828063841 | 0.05429165 | 0.885829174 | 0.0143 | 0.054291647 | 0.054291647 | 1.076329432 | 0.657681911 | -0.181984103 | FALSE | TRUE | FALSE |
| FMNL1 | 0.0357024 | 0.87974183 | 0.0297 | 0.03570237 | 0.03570237 | 0.698581109 | 1.89325173 | 0.499711857 | 0.0197 | 0.473595829 | 1.893251734 | 9.290932599 | 0.018857698 | -1.724511314 | FALSE | FALSE | FALSE |
| FRMD8 | 0.0356598 | 0.87975059 | 1 | 0.142549613 | 0.142549613 | 1.207811154 | 0.21106328 | 0.849325299 | 0.01405 | 0.211063282 | 0.211063282 | 1.044370458 | 0.16895329 | -0.772233348 | FALSE | TRUE | FALSE |
| DNAAF2 | 0.0356418 | 0.87975935 | 1 | 0.317133229 | 0.317133229 | 6.236621171 | 0.04888678 | 0.886963869 | 0.5084 | 0.438203094 | 0.438203094 | 1.6544581 | 0.729068584 | -0.137231615 | FALSE | TRUE | FALSE |
| GLS2 | 0.0356402 | 0.87976812 | 1 | 0.035640158 | 0.035640158 | 0.703785607 | 0.05068283 | 0.886485967 | 1 | 0.449600943 | 0.449600943 | 1.002356575 | 0.703199797 | -0.152921263 | FALSE | TRUE | FALSE |
| LCN6 | 0.0355672 | 0.87977688 | 0.00555 | 0.035567227 | 0.035567227 | 0.035567227 | 0.03964862 | 0.889022338 | 1 | 0.366027168 | 0.366027168 | 0.366027168 | 0.897060897 | -0.047178074 | FALSE | TRUE | FALSE |
| KCNU1 | 0.0355234 | 0.87978564 | 0.89135 | 2.736932961 | 232.8637323 | 2226.07451 | 432.099283 | 0.005377721 | 0.1553 | 45.76342906 | 4177.380427 | 41328.05891 | 8.22112E-05 | -4.085069097 | FALSE | FALSE | FALSE |
| SNX31 | 0.0354631 | 0.8797944 | 1 | 839307259.1 | 8.16837E+12 | 1.19336E+15 | 0.54537243 | 0.735606473 | 1 | 4600209.057 | 6016096549 | 73674618628 | 0.065025509 | -1.186916236 | FALSE | TRUE | FALSE |
| SLC14A1 | 0.0354617 | 0.87980315 | 1 | 0.035461711 | 0.035461711 | 0.035461711 | 0.0496509 | 0.886733628 | 0.87675 | 0.793966571 | 7.205465939 | 7.205465939 | 0.714220844 | -0.146167479 | FALSE | TRUE | FALSE |
| SGSM2 | 0.0354546 | 0.87981191 | 0.0358 | 0.035454612 | 0.035454612 | 0.689623374 | 0.44891202 | 0.774753193 | 0.516 | 0.448912018 | 0.448912018 | 8.760631947 | 0.078978978 | -1.102488489 | FALSE | TRUE | FALSE |
| LCK | 0.0353722 | 0.87982067 | 0.50365 | 0.035372219 | 0.141441002 | 0.331427503 | 0.03201016 | 0.89079083 | 0.50595 | 0.032010157 | 0.032010157 | 0.158383078 | 1.105031094 | 0.043374499 | FALSE | TRUE | FALSE |
| USP16 | 0.0353375 | 0.87982942 | 1 | 0.035337513 | 0.31024827 | 6.12018619 | 0.05331093 | 0.886004407 | 0.0173 | 0.053310931 | 0.053310931 | 1.05337992 | 0.66285679 | -0.178580291 | FALSE | TRUE | FALSE |
| OASL | 0.0352773 | 0.87983817 | 1 | 0.308476013 | 2.768608739 | 6.105006661 | 0.46944417 | 0.767258679 | 0.50335 | 0.469444168 | 0.469444168 | 1.053100526 | 0.075146966 | -1.124088552 | FALSE | TRUE | FALSE |
| EMILIN3 | 0.0352065 | 0.87984693 | 1 | 0.035206493 | 0.035206493 | 0.691939592 | 0.04610375 | 0.887543941 | 0.93785 | 3.902420279 | 365.5904155 | 365.5904155 | 0.763636138 | -0.117113527 | FALSE | TRUE | FALSE |
| GPR176 | 0.0351968 | 0.87985568 | 1 | 63538593807 | 1.13397E+14 | 1.83636E+16 | 0.08198039 | 0.879779717 | 0.9959 | 679.1041735 | 71614.1551 | 761453.2062 | 0.429332248 | -0.36720649 | FALSE | TRUE | FALSE |
| GZMA | 0.0351432 | 0.87986443 | 0.75445 | 0.312446407 | 2.851139583 | 2.851139583 | 0.47756475 | 0.763959709 | 0.49955 | 0.053269496 | 0.47756475 | 0.47756475 | 0.073588258 | -1.133191476 | FALSE | TRUE | FALSE |
| PDZD7 | 0.0351361 | 0.87987317 | 0.66195 | 0.2893953 | 21.13872263 | 187.2506126 | 5.4654E-06 | 0.900849022 | 0.507 | 4.20905E-05 | 4.20905E-05 | 0.000184843 | 6428.792431 | 3.808129404 | FALSE | TRUE | FALSE |
| PPWD1 | 0.0350941 | 0.87988192 | 1 | 0.035094136 | 0.035094136 | 0.692760269 | 0.05177618 | 0.886262232 | 1 | 0.051776183 | 0.051776183 | 1.02366333 | 0.677804613 | -0.16889548 | FALSE | TRUE | FALSE |
| MS4A14 | 0.0350494 | 0.87989067 | 1 | 0.305476466 | 0.305476466 | 1.185580933 | 0.45098334 | 0.774054949 | 0.50645 | 0.450983342 | 0.450983342 | 1.01530074 | 0.077717827 | -1.10947935 | FALSE | TRUE | FALSE |
| CCDC146 | 0.0350447 | 0.87989941 | 0.27225 | 0.035044654 | 0.298528987 | 1.181522907 | 0.06356569 | 0.883878026 | 0.8775 | 0.549105207 | 4.865463969 | 10.82037068 | 0.551313938 | -0.258601028 | FALSE | TRUE | FALSE |
| ACTRT2 | 0.0350446 | 0.87990816 | 0.01205 | 0.035044608 | 0.035044608 | 0.173728956 | 0.01535205 | 0.895046867 | 0.49835 | 0.015352046 | 0.06134164 | 0.304405316 | 2.282732054 | 0.358454937 | FALSE | TRUE | FALSE |
| SMOX | 0.0350147 | 0.8799169 | 0.019 | 0.035014704 | 0.035014704 | 0.689964425 | 0.20548024 | 0.85070339 | 0.50165 | 0.205480243 | 0.205480243 | 1.014631619 | 0.170404239 | -0.768519607 | FALSE | TRUE | FALSE |
| CAPN10 | 0.0349484 | 0.87992564 | 0.513 | 0.03494837 | 0.03494837 | 0.687215892 | 0.0645022 | 0.883696872 | 0.5009 | 0.14138995 | 2.378271188 | 2.378271188 | 0.541816732 | -0.266147588 | FALSE | TRUE | FALSE |
| OSGIN1 | 0.0349346 | 0.87993438 | 1 | 0.139687051 | 0.139687051 | 0.689780917 | 1.72960261 | 0.515106673 | 0.0117 | 0.204017177 | 0.816004822 | 4.034483846 | 0.020198057 | -1.694690398 | FALSE | FALSE | FALSE |
| JAK2 | 0.0349093 | 0.87994312 | 0.88095 | 0.139542082 | 0.557927363 | 2.736073647 | 0.06102537 | 0.884529307 | 0.5155 | 0.243961927 | 0.975533549 | 4.794792321 | 0.572045846 | -0.242569164 | FALSE | TRUE | FALSE |
| SCO1 | 0.0348866 | 0.87995186 | 1 | 0.034886585 | 0.034886585 | 0.034886585 | 0.05165251 | 0.886278832 | 0.00835 | 0.051652505 | 0.051652505 | 0.051652505 | 0.675409352 | -0.170432931 | FALSE | TRUE | FALSE |
| C7 | 0.0348815 | 0.8799606 | 0.7551 | 0.034881531 | 0.315418754 | 0.687017093 | 0.00752619 | 0.897435519 | 1 | 0.066869459 | 0.609797622 | 1.319454312 | 4.63468617 | 0.666020332 | FALSE | TRUE | FALSE |
| CLUL1 | 0.0348743 | 0.87996933 | 1 | 0.034874296 | 0.034874296 | 0.692146827 | 0.04475953 | 0.887941116 | 1 | 0.401587054 | 0.401587054 | 0.401587054 | 0.779147857 | -0.10838012 | FALSE | TRUE | FALSE |
| CKAP2 | 0.0348493 | 0.87997807 | 0.0172 | 0.034849267 | 0.034849267 | 0.688616448 | 0.05103777 | 0.886378342 | 1 | 0.051037771 | 0.051037771 | 1.009955915 | 0.682813255 | -0.165698057 | FALSE | TRUE | FALSE |
| SSPN | 0.0348256 | 0.8799868 | 1 | 0.32564016 | 0.32564016 | 0.32564016 | 0.03890508 | 0.889204681 | 1 | 0.038905078 | 0.038905078 | 0.038905078 | 0.895143112 | -0.048107526 | FALSE | TRUE | FALSE |
| MORN1 | 0.0348255 | 0.87999553 | 1 | 0.034825534 | 0.034825534 | 0.689124999 | 0.19716322 | 0.852799552 | 0.7583 | 0.443381682 | 1.773033067 | 3.906427339 | 0.17663302 | -0.752928105 | FALSE | TRUE | FALSE |
| SLAMF1 | 0.0348154 | 0.88000426 | 1 | 0.034815409 | 0.034815409 | 0.034815409 | 0.03749921 | 0.889567724 | 1 | 0.349617575 | 0.349617575 | 0.349617575 | 0.928430565 | -0.03225057 | FALSE | TRUE | FALSE |
| MON1A | 0.0348034 | 0.88001299 | 1 | 0.034803447 | 0.034803447 | 0.684984088 | 0.2031837 | 0.851362908 | 0.0181 | 0.203183705 | 0.203183705 | 1.720330777 | 0.171290544 | -0.766266612 | FALSE | TRUE | FALSE |
| DNAI2 | 0.0347761 | 0.88002172 | 0.8737 | 0.139076919 | 0.556336834 | 2.738188052 | 0.0146418 | 0.895247454 | 1 | 0.128714082 | 1.16135902 | 2.539430341 | 2.375125063 | 0.375686482 | FALSE | TRUE | FALSE |
| HTR2B | 0.0347492 | 0.88003044 | 1 | 0.034749215 | 0.034749215 | 0.688712918 | 0.04816791 | 0.887086867 | 0.7504 | 0.428380938 | 3.910276359 | 3.910276359 | 0.721418371 | -0.141812803 | FALSE | TRUE | FALSE |
| ATG2A | 0.0347488 | 0.88003917 | 1 | 0.312995319 | 0.662878866 | 5.970788982 | 3.9320664 | 0.360089759 | 0.3394 | 3.932066397 | 364.862335 | 717.6525903 | 0.008837297 | -2.05368055 | FALSE | FALSE | FALSE |
| ZNF682 | 0.0346959 | 0.88004789 | 1 | 0.034695889 | 0.034695889 | 0.688154131 | 0.4451392 | 0.775729691 | 0.5059 | 0.445139203 | 0.445139203 | 0.985788479 | 0.077943907 | -1.108217828 | FALSE | TRUE | FALSE |
| NOSTRIN | 0.0346881 | 0.88005662 | 1 | 0.034688085 | 0.034688085 | 0.688109146 | 0.050891 | 0.886403195 | 0.0097 | 0.050890996 | 0.050890996 | 0.050890996 | 0.681615364 | -0.166460629 | FALSE | TRUE | FALSE |
| CYP51A1 | 0.0345916 | 0.88006534 | 1 | 0.314233042 | 0.314233042 | 0.684968112 | 0.46809823 | 0.767832876 | 0.50665 | 0.468098231 | 0.468098231 | 1.016406396 | 0.073898081 | -1.131366838 | FALSE | TRUE | FALSE |
| NCR1 | 0.034587 | 0.88007406 | 1 | 0.034586959 | 0.034586959 | 0.034586959 | 0.04145731 | 0.888616071 | 1 | 0.041457313 | 0.041457313 | 0.041457313 | 0.834278829 | -0.078688777 | FALSE | TRUE | FALSE |
| GC | 0.0345528 | 0.88008278 | 1 | 0.034552847 | 0.034552847 | 0.685712418 | 0.45215079 | 0.773768018 | 0.51125 | 0.452150789 | 0.452150789 | 0.98643353 | 0.076418859 | -1.116799453 | FALSE | TRUE | FALSE |
| TNNT1 | 0.0345516 | 0.8800915 | 1 | 0.034551572 | 0.034551572 | 0.034551572 | 0.05132786 | 0.886328605 | 0.00945 | 0.051327857 | 0.051327857 | 0.051327857 | 0.673154382 | -0.171885323 | FALSE | TRUE | FALSE |
| MMAB | 0.0344995 | 0.88010022 | 1 | 0.137928187 | 0.137928187 | 0.137928187 | 4.12112424 | 0.348460633 | 0.0046 | 0.441950836 | 4.121124236 | 4.121124236 | 0.008371388 | -2.077202505 | FALSE | FALSE | FALSE |
| RTN3 | 0.0344796 | 0.88010893 | 1 | 0.302237753 | 2.71924308 | 5.939401902 | 0.0506275 | 0.886494238 | 1 | 0.447799263 | 4.065247721 | 8.818358175 | 0.681045745 | -0.166823716 | FALSE | TRUE | FALSE |
| FAM13B | 0.0343563 | 0.88011765 | 1 | 0.318126003 | 0.318126003 | 6.265098759 | 0.04957527 | 0.886766573 | 1 | 0.459979337 | 4.380277888 | 9.075420333 | 0.693013088 | -0.159258564 | FALSE | TRUE | FALSE |
| SERPINE1 | 0.0342514 | 0.88012636 | 1 | 3.88052E+20 | 1.23608E+26 | 2.23436E+28 | 0.0807943 | 0.880109231 | 1 | 8.30181E+11 | 5.44968E+15 | 2.58391E+17 | 0.423933779 | -0.372701977 | FALSE | TRUE | FALSE |
| NIF3L1 | 0.0342209 | 0.88013508 | 1 | 0.034220907 | 0.034220907 | 0.679607711 | 34.2979171 | 0.08130572 | 0.00425 | 3.688507444 | 34.29791707 | 34.29791707 | 0.000997755 | -3.000976228 | FALSE | FALSE | FALSE |
| DEPDC5 | 0.0341765 | 0.88014379 | 1 | 0.034176486 | 0.034176486 | 1.145640618 | 0.44444997 | 0.775917711 | 0.5215 | 0.444449972 | 0.444449972 | 8.61501822 | 0.076896138 | -1.114095472 | FALSE | TRUE | FALSE |
| ZNF136 | 0.0341658 | 0.8801525 | 1 | 0.034165841 | 0.034165841 | 0.676864201 | 0.05036061 | 0.886527312 | 1 | 0.050360613 | 0.050360613 | 0.998834288 | 0.678423846 | -0.168498895 | FALSE | TRUE | FALSE |
| IL12RB1 | 0.0341541 | 0.88016121 | 0.96815 | 25.92714311 | 248.7296962 | 2444.51719 | 0.05682659 | 0.885317406 | 1 | 0.51074419 | 0.51074419 | 1.921989003 | 0.601022684 | -0.221109137 | FALSE | TRUE | FALSE |
| NUDT12 | 0.0341249 | 0.88016992 | 0.01075 | 0.03412494 | 0.03412494 | 0.67756173 | 0.05020058 | 0.886552107 | 1 | 0.05020058 | 0.05020058 | 0.05020058 | 0.679771823 | -0.167636841 | FALSE | TRUE | FALSE |
| PCDHA12 | 0.0340947 | 0.88017863 | 0.76095 | 0.034094675 | 0.301854161 | 1.151905664 | 0.00790157 | 0.897339136 | 0.03015 | 0.007901572 | 0.007901572 | 0.154165974 | 4.314922926 | 0.634973043 | FALSE | TRUE | FALSE |
| CDK10 | 0.0340191 | 0.88018733 | 1 | 0.304126532 | 0.304126532 | 0.673939839 | 4.05471094 | 0.353564812 | 0.25605 | 0.439268619 | 4.054710944 | 4.054710944 | 0.008390028 | -2.076236587 | FALSE | FALSE | FALSE |
| SLC34A3 | 0.0340024 | 0.88019604 | 0.26075 | 0.034002363 | 0.304836505 | 0.669895987 | 0.00783826 | 0.897373577 | 1 | 0.00783826 | 0.00783826 | 0.15470257 | 4.337998772 | 0.637289425 | FALSE | TRUE | FALSE |
| TMF1 | 0.0339255 | 0.88020474 | 1 | 0.135633842 | 0.135633842 | 2.662929726 | 0.05106706 | 0.886361767 | 1 | 0.05106706 | 0.05106706 | 1.004825003 | 0.664331795 | -0.177614962 | FALSE | TRUE | FALSE |
| LRRC40 | 0.0338864 | 0.88021345 | 1 | 0.033886444 | 0.033886444 | 0.670447443 | 0.04988426 | 0.886659436 | 1 | 0.049884265 | 0.049884265 | 0.988241927 | 0.679301261 | -0.167937579 | FALSE | TRUE | FALSE |
| KIF1C | 0.033801 | 0.88022215 | 0.0382 | 0.033800986 | 0.033800986 | 0.656911504 | 0.1146773 | 0.872434356 | 0.27495 | 0.028692854 | 0.114677303 | 0.967552899 | 0.294748701 | -0.530548101 | FALSE | TRUE | FALSE |
| IL9 | 0.0337683 | 0.88023085 | 0.4979 | 0.033768266 | 0.315124897 | 0.315124897 | 0.03780254 | 0.889481106 | 0.00305 | 0.03780254 | 0.03780254 | 0.03780254 | 0.893280352 | -0.049012218 | FALSE | TRUE | FALSE |
| TRAPPC11 | 0.0337441 | 0.88023955 | 1 | 0.033744068 | 0.033744068 | 0.661466775 | 0.04736226 | 0.887250487 | 1 | 0.757788639 | 6.781503554 | 25.55643917 | 0.712467381 | -0.147235014 | FALSE | TRUE | FALSE |
| ACTN1 | 0.0336659 | 0.88024825 | 0.51195 | 0.134561194 | 0.134561194 | 2.62902947 | 0.05087161 | 0.886411478 | 1 | 0.050871614 | 0.050871614 | 0.996686683 | 0.661781839 | -0.179285155 | FALSE | TRUE | FALSE |
| TBCCD1 | 0.0336404 | 0.88025695 | 1 | 0.033640356 | 0.033640356 | 0.666030067 | 0.05008398 | 0.886585152 | 1 | 0.050083975 | 0.050083975 | 0.99279215 | 0.671679034 | -0.172838208 | FALSE | TRUE | FALSE |
| SEMA4G | 0.0336207 | 0.88026564 | 1 | 9.30304E+17 | 6.24288E+23 | 3.82147E+26 | 0.11008906 | 0.873319488 | 1 | 7.82466E+20 | 4.35633E+26 | 9.68618E+28 | 0.305395689 | -0.515137098 | FALSE | TRUE | FALSE |
| PLA2G4E | 0.0335773 | 0.88027434 | 0.51065 | 0.294693746 | 0.294693746 | 5.805904885 | 0.42506512 | 0.782770019 | 0.75185 | 0.425065119 | 3.859804316 | 8.38945903 | 0.078993356 | -1.102409435 | FALSE | TRUE | FALSE |
| SLC5A5 | 0.0335575 | 0.88028303 | 0.0192 | 0.033557522 | 0.033557522 | 0.28429339 | 0.22755942 | 0.845453566 | 0.32265 | 0.227559416 | 3.639235613 | 4.485916736 | 0.147467077 | -0.831304929 | FALSE | TRUE | FALSE |
| LIG4 | 0.0335387 | 0.88029172 | 0.021 | 0.033538658 | 0.033538658 | 0.660086337 | 0.48276301 | 0.761989683 | 0.3887 | 0.482763008 | 17.58828971 | 17.58828971 | 0.069472304 | -1.158188299 | FALSE | TRUE | FALSE |
| EGFR | 0.0335198 | 0.88030042 | 0.9375 | 0.309978425 | 28.58731795 | 57.13975311 | 0.06837217 | 0.882792091 | 0.75935 | 0.273254783 | 2.546596097 | 9.214441751 | 0.490254529 | -0.309578385 | FALSE | TRUE | FALSE |
| ALX4 | 0.0335064 | 0.88030911 | 1 | 0.033506359 | 0.033506359 | 0.663702243 | 0.05007685 | 0.886601668 | 1 | 0.050076852 | 0.050076852 | 0.993078592 | 0.669098749 | -0.174509782 | FALSE | TRUE | FALSE |

| | | | | | | | | | | | | | | | | | |
|---|---|---|---|---|---|---|---|---|---|---|---|---|---|---|---|---|---|
| PCDHGA1 | 0.0335037 | 0.8803178 | 0.8785 | 0.295261739 | 2.669043836 | 5.772886301 | 0.00757471 | 0.897414879 | 0.75465 | 0.065827224 | 0.587161221 | 1.290512948 | 4.423106954 | 0.645727441 | FALSE | TRUE | FALSE |
| FRMD6 | 0.0334704 | 0.88032648 | 0.50385 | 0.133791965 | 0.133791965 | 1.132847016 | 0.193093 | 0.853601087 | 0.93745 | 0.772099569 | 12.35413742 | 12.35413742 | 0.173338401 | -0.761105213 | FALSE | TRUE | FALSE |
| MZT2B | 0.0334588 | 0.88033517 | 1 | 0.033458812 | 0.033458812 | 0.033458812 | 0.03881112 | 0.889260067 | 1 | 0.038811122 | 0.038811122 | 0.038811122 | 0.862093399 | -0.06444568 | FALSE | TRUE | FALSE |
| ARHGEF28 | 0.033414 | 0.88034386 | 0.7593 | 0.033414034 | 0.28899203 | 5.616703481 | 0.0632093 | 0.883964111 | 0.7583 | 0.063209304 | 0.553953639 | 10.80299788 | 0.528625245 | -0.276852101 | FALSE | TRUE | FALSE |
| ZSWIM2 | 0.033402 | 0.88035254 | 1 | 0.295028509 | 0.295028509 | 1.129437077 | 0.05083616 | 0.886419759 | 0.0126 | 0.050836161 | 0.050836161 | 1.007458528 | 0.657051446 | -0.182400624 | FALSE | TRUE | FALSE |
| SCRN2 | 0.0333928 | 0.88036122 | 1 | 0.290290857 | 0.290290857 | 0.6612477 | 0.41807581 | 0.78535127 | 0.2578 | 0.418075808 | 3.764215938 | 3.764215938 | 0.079872708 | -1.097601589 | FALSE | TRUE | FALSE |
| HIRIP3 | 0.0333517 | 0.88036991 | 1 | 0.033351727 | 0.033351727 | 0.658333638 | 0.0491163 | 0.886898175 | 1 | 0.049116297 | 0.049116297 | 0.971034122 | 0.679035856 | -0.168107293 | FALSE | TRUE | FALSE |
| FHL2 | 0.0333444 | 0.88037859 | 1 | 0.033344425 | 0.033344425 | 0.033344425 | 0.04979066 | 0.886708907 | 1 | 0.04979066 | 0.04979066 | 0.04979066 | 0.669692365 | -0.174124652 | FALSE | TRUE | FALSE |
| KIT | 0.0333378 | 0.88038727 | 0.0268 | 0.033337844 | 0.033337844 | 0.653483182 | 0.04987565 | 0.886675931 | 0.02325 | 0.04987565 | 0.04987565 | 0.979998643 | 0.668419235 | -0.174951061 | FALSE | TRUE | FALSE |
| SEC23A | 0.0333182 | 0.88039595 | 0.96255 | 0.532467918 | 8.518026398 | 41.99135954 | 1.45501444 | 0.539651705 | 0.00935 | 0.157508773 | 1.45501444 | 1.45501444 | 0.022898895 | -1.64018547 | FALSE | FALSE | FALSE |
| IL36G | 0.0332867 | 0.88040462 | 1 | 0.306505357 | 0.306505357 | 0.306505357 | 0.0393984 | 0.889101684 | 1 | 0.039398396 | 0.039398396 | 0.039398396 | 0.84487385 | -0.073208132 | FALSE | TRUE | FALSE |
| ELAC2 | 0.0332719 | 0.8804133 | 0.51475 | 0.033271942 | 0.033271942 | 0.653237986 | 0.02993752 | 0.89126718 | 0.02085 | 0.029937521 | 0.029937521 | 0.589069947 | 1.111379311 | 0.045862308 | FALSE | TRUE | FALSE |
| ATP13A3 | 0.0331999 | 0.88042197 | 1 | 0.033199908 | 0.033199908 | 0.649198695 | 0.04962454 | 0.886741866 | 1 | 0.049624536 | 0.049624536 | 0.972975139 | 0.669022035 | -0.174559578 | FALSE | TRUE | FALSE |
| PTPRZ1 | 0.0331902 | 0.88043065 | 0.52845 | 0.033190212 | 0.159791735 | 0.63871986 | 0.01425607 | 0.895347481 | 0.8834 | 0.227869329 | 0.911363578 | 7.639782854 | 2.328145471 | 0.367010113 | FALSE | TRUE | FALSE |
| CCDC157 | 0.0331646 | 0.88043932 | 0.98305 | 23.78995121 | 2160.084054 | 21331.03332 | 0.05655027 | 0.885376337 | 1 | 0.500179482 | 0.500179482 | 1.914312872 | 0.586461735 | -0.231760319 | FALSE | TRUE | FALSE |
| EFHC1 | 0.0331583 | 0.88044799 | 0.8787 | 0.132591671 | 0.530333208 | 1.12121181 | 0.01476444 | 0.895190216 | 0.5047 | 0.05902889 | 0.05902889 | 0.49957214 | 2.245820914 | 0.351375122 | FALSE | TRUE | FALSE |
| FOXP4 | 0.0330873 | 0.88045666 | 0.02095 | 0.033087252 | 0.033087252 | 0.651302898 | 0.04954827 | 0.886774807 | 1 | 0.049548265 | 0.049548265 | 0.977180739 | 0.667778219 | -0.17536775 | FALSE | TRUE | FALSE |
| ABCB10 | 0.033055 | 0.88046533 | 0.87325 | 0.132145792 | 0.528418768 | 0.653095277 | 0.12196309 | 0.87082898 | 0.6959 | 1.127624539 | 42.7433099 | 42.7433099 | 0.271024512 | -0.566991429 | FALSE | TRUE | FALSE |
| DDX28 | 0.0330328 | 0.880474 | 1 | 0.03303285 | 0.03303285 | 0.651412446 | 0.19368555 | 0.853428104 | 0.0156 | 0.193685545 | 0.193685545 | 1.639632498 | 0.170548865 | -0.768151165 | FALSE | TRUE | FALSE |
| NEMF | 0.0330172 | 0.88048267 | 0.02775 | 0.03301721 | 0.03301721 | 0.648253052 | 0.0484118 | 0.887029497 | 1 | 0.193618662 | 0.193618662 | 3.809849921 | 0.682007451 | -0.166210881 | FALSE | TRUE | FALSE |
| PIGV | 0.0330012 | 0.88049133 | 1 | 0.033001221 | 0.033001221 | 0.654064173 | 0.04919616 | 0.886865302 | 1 | 0.049196157 | 0.049196157 | 0.976096997 | 0.670808916 | -0.173401174 | FALSE | TRUE | FALSE |
| RINT1 | 0.0329857 | 0.8805 | 1 | 0.032985668 | 0.032985668 | 0.650582221 | 0.04586653 | 0.887592716 | 1 | 0.403734266 | 3.647569745 | 7.976186907 | 0.719166352 | -0.143170641 | FALSE | TRUE | FALSE |
| ZFC3H1 | 0.032977 | 0.88050866 | 1 | 0.032976967 | 0.032976967 | 1.101798033 | 0.04102589 | 0.888695945 | 1 | 0.380887694 | 0.380887694 | 7.380779989 | 0.803808611 | -0.094847346 | FALSE | TRUE | FALSE |
| PCDHA10 | 0.0329346 | 0.88051732 | 0.5101 | 0.059999339 | 0.526281591 | 1.178585264 | 0.00755542 | 0.89742176 | 1 | 0.007555418 | 0.007555418 | 0.148732079 | 4.359072315 | 0.639394074 | FALSE | TRUE | FALSE |
| TPD52L2 | 0.0329137 | 0.88052598 | 1 | 0.032913674 | 0.032913674 | 0.032913674 | 0.03749847 | 0.889575592 | 0.0052 | 0.037498466 | 0.037498466 | 0.037498466 | 0.877733867 | -0.056637144 | FALSE | TRUE | FALSE |
| UTS2B | 0.0329081 | 0.88053464 | 1 | 0.303915983 | 0.303915983 | 0.303915983 | 0.34489224 | 0.810713001 | 0.50155 | 0.34489224 | 0.34489224 | 0.34489224 | 0.09541569 | -1.020380205 | FALSE | TRUE | FALSE |
| ANO4 | 0.0328959 | 0.8805433 | 1 | 4.57069E+11 | 6.41236E+15 | 4.11459E+17 | 0.01521624 | 0.895082737 | 1 | 15068621617 | 2.75932E+14 | 1.35042E+16 | 2.161894857 | 0.334834568 | FALSE | TRUE | FALSE |
| RAB11FIP1 | 0.0328223 | 0.88055196 | 0.0337 | 0.032822258 | 0.032822258 | 0.639811667 | 0.42213347 | 0.78379274 | 0.0303 | 0.422133466 | 0.422133466 | 8.253870073 | 0.077753272 | -1.109281329 | FALSE | TRUE | FALSE |
| NEUROD4 | 0.0328028 | 0.88056061 | 1 | 0.032802817 | 0.032802817 | 0.032802817 | 0.44080793 | 0.777162937 | 0.00725 | 0.440807932 | 0.440807932 | 0.440807932 | 0.074415215 | -1.128338259 | FALSE | TRUE | FALSE |
| SLC5A7 | 0.0327819 | 0.88056927 | 0.0163 | 0.032781933 | 0.032781933 | 0.648608741 | 0.04762225 | 0.887225971 | 1 | 0.448239043 | 0.448239043 | 0.943447956 | 0.688374344 | -0.162175324 | FALSE | TRUE | FALSE |
| DBF4B | 0.0327117 | 0.88057792 | 1 | 0.032711654 | 0.032711654 | 0.646179635 | 0.04834588 | 0.887037696 | 0.0145 | 0.048345879 | 0.048345879 | 0.956429579 | 0.676617214 | -0.169656957 | FALSE | TRUE | FALSE |
| LIPF | 0.0327013 | 0.88058658 | 1 | 0.291882422 | 2.673973098 | 2.673973098 | 0.05157616 | 0.886295427 | 1 | 0.051576156 | 0.051576156 | 0.051576156 | 0.634039932 | -0.197883389 | FALSE | TRUE | FALSE |
| STAT5A | 0.0326673 | 0.88059523 | 1 | 0.032667278 | 0.032667278 | 0.640912983 | 0.04898163 | 0.886939242 | 1 | 0.048981628 | 0.048981628 | 0.963193134 | 0.666929196 | -0.17592027 | FALSE | TRUE | FALSE |
| HYI | 0.0326325 | 0.88060388 | 0.50735 | 0.287550146 | 0.287550146 | 0.287550146 | 0.04689461 | 0.887372927 | 1 | 0.416364633 | 3.794287509 | 3.794287509 | 0.695869377 | -0.157472275 | FALSE | TRUE | FALSE |
| MSANTD1 | 0.0325942 | 0.88061253 | 1 | 0.032594205 | 0.032594205 | 0.032594205 | 0.03563396 | 0.889967732 | 1 | 0.035633958 | 0.035633958 | 0.035633958 | 0.914695067 | -0.038723663 | FALSE | TRUE | FALSE |
| ANKMY1 | 0.0325881 | 0.88062118 | 0.70225 | 0.067772222 | 1.08445365 | 2.28898549 | 0.0043487 | 0.898234646 | 1 | 0.004348703 | 0.004348703 | 0.085240607 | 7.493758996 | 0.874699722 | FALSE | TRUE | FALSE |
| AGT | 0.0325045 | 0.88062982 | 1 | 0.129953559 | 0.129953559 | 0.643545183 | 0.41984667 | 0.78460379 | 0.755 | 0.419846668 | 3.918357587 | 3.918357587 | 0.077419897 | -1.111147411 | FALSE | TRUE | FALSE |
| GSDMA | 0.0324709 | 0.88063847 | 0.0116 | 0.032470929 | 0.032470929 | 0.64378172 | 0.04782352 | 0.887176911 | 0.01155 | 0.047823524 | 0.047823524 | 0.949158277 | 0.678973987 | -0.168146864 | FALSE | TRUE | FALSE |
| CEP135 | 0.0324405 | 0.88064711 | 0.5157 | 0.032440521 | 0.032440521 | 0.636042446 | 0.05931823 | 0.884827009 | 0.7564 | 0.059318226 | 0.511478232 | 1.996558536 | 0.546889609 | -0.262100328 | FALSE | TRUE | FALSE |
| NAGS | 0.0324401 | 0.88065576 | 1 | 0.032440149 | 0.032440149 | 0.640881936 | 0.04860819 | 0.886996691 | 1 | 0.048608188 | 0.048608188 | 0.961705522 | 0.667380352 | -0.175626584 | FALSE | TRUE | FALSE |
| TMPRSS7 | 0.0323461 | 0.8806644 | 0.9522 | 1.158162216 | 100.2353363 | 400.7367079 | 0.55288978 | 0.732014425 | 0.8127 | 5.124066184 | 194.5862733 | 1893.790725 | 0.058503715 | -1.232816554 | FALSE | TRUE | FALSE |
| ANKEF1 | 0.0323344 | 0.88067304 | 0.74845 | 0.070752662 | 0.282870039 | 1.092983868 | 0.01467534 | 0.895225997 | 0.0176 | 0.014675338 | 0.014675338 | 0.289571615 | 2.203313582 | 0.343076312 | FALSE | TRUE | FALSE |
| PAX9 | 0.0323336 | 0.88068168 | 1 | 0.129205401 | 0.129205401 | 0.639873482 | 0.04856708 | 0.887004894 | 1 | 0.194097132 | 0.194097132 | 0.962344086 | 0.665752341 | -0.176687298 | FALSE | TRUE | FALSE |
| THSD4 | 0.032332 | 0.88069032 | 1 | 0.516921254 | 2.067678137 | 10.11124531 | 0.43229823 | 0.780214278 | 0.7608 | 0.432298231 | 4.074920521 | 8.539606338 | 0.074791022 | -1.126150533 | FALSE | TRUE | FALSE |
| EPB41L1 | 0.0323124 | 0.88069896 | 0.7572 | 0.075595124 | 0.302014928 | 1.479748494 | 0.0143032 | 0.895333202 | 1 | 0.133937605 | 0.535179014 | 2.628498644 | 2.259103971 | 0.353936219 | FALSE | TRUE | FALSE |
| GPR39 | 0.0322737 | 0.8807076 | 0.50665 | 0.129080943 | 0.129080943 | 0.637224039 | 32.4375898 | 0.084728086 | 0.1281 | 3.492957844 | 129.7382167 | 129.7382167 | 0.000994946 | -3.002200363 | FALSE | FALSE | FALSE |
| AIFM3 | 0.0322603 | 0.88071623 | 1 | 0.288181794 | 0.288181794 | 1.091653112 | 0.04801986 | 0.887135998 | 1 | 0.431949377 | 0.431949377 | 1.625865979 | 0.671811098 | -0.172752826 | FALSE | TRUE | FALSE |
| SPATA21 | 0.0322519 | 0.88072487 | 1 | 0.281426347 | 2.520517833 | 2.551016359 | 3.79548636 | 0.366498646 | 0.25675 | 0.419782277 | 3.795486357 | 8.319225339 | 0.008497423 | -2.070712761 | FALSE | FALSE | FALSE |
| NDUFAF7 | 0.0322029 | 0.8807335 | 0.5035 | 0.032202928 | 0.032202928 | 0.159719269 | 0.01421043 | 0.895361757 | 1 | 0.014210427 | 0.014210427 | 0.282048384 | 2.266147898 | 0.35528825 | FALSE | TRUE | FALSE |
| ARHGAP19 | 0.0321593 | 0.88074213 | 0.0136 | 0.03215931 | 0.03215931 | 0.636945204 | 0.04483665 | 0.887900708 | 0.75115 | 0.394213902 | 3.557454607 | 7.816887731 | 0.71725504 | -0.144326391 | FALSE | TRUE | FALSE |
| ZNF251 | 0.0321514 | 0.88075076 | 0.0212 | 0.032151405 | 0.032151405 | 0.634290375 | 0.04622503 | 0.887486989 | 0.5046 | 0.408521141 | 0.408521141 | 1.563985363 | 0.695540984 | -0.157677275 | FALSE | TRUE | FALSE |
| SLC9A4 | 0.0321227 | 0.88075939 | 1 | 0.032122685 | 0.032122685 | 0.631929928 | 0.18818877 | 0.854996997 | 0.50315 | 0.188188773 | 0.188188773 | 1.592035759 | 0.170693952 | -0.767781866 | FALSE | TRUE | FALSE |
| POLR2B | 0.0321156 | 0.88076802 | 0.99945 | 176.0499919 | 11286.04584 | 108566.9285 | 0.12340204 | 0.870492877 | 1 | 6134846904 | 3.26297E+13 | 1.08965E+15 | 0.260252181 | -0.584605622 | FALSE | TRUE | FALSE |
| INTS8 | 0.0320614 | 0.88077665 | 0.02525 | 0.032061444 | 0.032061444 | 0.630899121 | 0.03835158 | 0.889362791 | 1 | 0.038351585 | 0.038351585 | 0.756138813 | 0.835987451 | -0.077800241 | FALSE | TRUE | FALSE |
| CIITA | 0.03203 | 0.88078527 | 1 | 0.128050468 | 0.128050468 | 2.489556062 | 0.43576879 | 0.778943375 | 0.51345 | 0.435768793 | 0.435768793 | 8.500708134 | 0.073502372 | -1.133698645 | FALSE | TRUE | FALSE |
| ZNF648 | 0.0320296 | 0.8807939 | 0.75305 | 0.281136437 | 2.53276974 | 5.533559883 | 0.04887827 | 0.886972076 | 1 | 0.433215223 | 0.433215223 | 1.655065258 | 0.655293027 | -0.183564453 | FALSE | TRUE | FALSE |
| AQR | 0.0319999 | 0.88080252 | 0.7468 | 0.127796871 | 4.807811924 | 46.44137076 | 0.0026801 | 0.898814898 | 0.5131 | 0.002680096 | 0.002680096 | 0.052328016 | 11.93983831 | 1.076998446 | FALSE | TRUE | FALSE |
| ETV4 | 0.0319982 | 0.88081114 | 1 | 0.031998203 | 0.031998203 | 0.632906289 | 0.40622642 | 0.789862451 | 0.50715 | 0.406226417 | 0.406226417 | 0.911261834 | 0.07876938 | -1.103642571 | FALSE | TRUE | FALSE |
| NADK | 0.0319858 | 0.88081977 | 0.759 | 0.281040718 | 2.534507751 | 5.551034726 | 0.05029811 | 0.886535578 | 1 | 0.050298113 | 0.050298113 | 0.994957741 | 0.635924696 | -0.196594309 | FALSE | TRUE | FALSE |
| RUNX1 | 0.0319785 | 0.88082839 | 0.5118 | 0.300351489 | 0.300351489 | 0.632808975 | 0.04514597 | 0.887819811 | 1 | 4.08324321 | 40.34979019 | 40.34979019 | 0.70833517 | -0.149761194 | FALSE | TRUE | FALSE |
| ZIM3 | 0.0318955 | 0.880837 | 1 | 0.278935127 | 0.278935127 | 0.632539822 | 0.04722365 | 0.887283161 | 1 | 0.416824955 | 0.416824955 | 0.937469453 | 0.675414327 | -0.170429731 | FALSE | TRUE | FALSE |
| CDH3 | 0.0318694 | 0.88084562 | 0.26845 | 0.031869424 | 0.127408305 | 0.625883878 | 0.05671079 | 0.885342669 | 0.5103 | 0.056710792 | 0.056710792 | 1.11616418 | 0.561964006 | -0.250291501 | FALSE | TRUE | FALSE |
| MTMR6 | 0.0318667 | 0.88085424 | 0.93815 | 0.293665833 | 11.10052572 | 11.10052572 | 0.23230163 | 0.844354661 | 0.5031 | 0.232301633 | 2.150130354 | 2.150130354 | 0.137178102 | -0.862715212 | FALSE | TRUE | FALSE |
| RAPSN | 0.0318433 | 0.88086285 | 1 | 0.29730338 | 0.29730338 | 0.6316339 | 0.04587591 | 0.887576462 | 0.87375 | 1.556252405 | 16.44014828 | 16.44014828 | 0.69411876 | -0.158566218 | FALSE | TRUE | FALSE |
| FILIP1 | 0.0317919 | 0.88087147 | 1 | 0.281095153 | 2.550941001 | 9.453453904 | 0.40864516 | 0.789009186 | 0.87985 | 3.735199202 | 35.01870153 | 73.21382399 | 0.077798317 | -1.109029797 | FALSE | TRUE | FALSE |
| UNC5B | 0.0317555 | 0.88088008 | 1 | 2.84206E+21 | 2.15227E+28 | 4.77011E+30 | 0.00091058 | 0.899521675 | 1 | 1.24535E+26 | 1.48823E+33 | 3.81364E+35 | 34.87408503 | 1.542502822 | FALSE | TRUE | FALSE |
| IL27RA | 0.0317048 | 0.88088869 | 0.879 | 2.552653553 | 23.80146465 | 23.80146465 | 0.0480648 | 0.887103248 | 1 | 0.048064797 | 0.048064797 | 0.949656645 | 0.659627187 | -0.180701453 | FALSE | TRUE | FALSE |
| SRR | 0.0317019 | 0.88089731 | 1 | 0.126724545 | 0.506693129 | 0.506693129 | 0.74311016 | 0.644646274 | 0.688 | 0.743110156 | 11.89071355 | 11.89071355 | 0.042661079 | -1.369968168 | FALSE | TRUE | FALSE |
| CYBA | 0.0316727 | 0.88090592 | 1 | 0.126600699 | 1.161013493 | 1.161013493 | 0.04804582 | 0.887119626 | 1 | 0.442386297 | 0.442386297 | 0.442386297 | 0.659217732 | -0.180971119 | FALSE | TRUE | FALSE |
| NEK3 | 0.0316657 | 0.88091452 | 1 | 0.278574351 | 0.278574351 | 1.072272595 | 0.0470195 | 0.887332141 | 1 | 0.417225286 | 0.417225286 | 0.933320221 | 0.673459373 | -0.171688598 | FALSE | TRUE | FALSE |
| EPHB3 | 0.0316545 | 0.88092313 | 1 | 0.031654501 | 0.031654501 | 0.6161437 | 0.18829312 | 0.854905624 | 0.5224 | 0.188293115 | 0.188293115 | 3.676883856 | 0.168112899 | -0.774398964 | FALSE | TRUE | FALSE |
| NIPAL3 | 0.0316406 | 0.88093174 | 0.7541 | 0.071786489 | 0.286865192 | 0.286865192 | 0.01456205 | 0.895276051 | 1 | 0.014562045 | 0.014562045 | 0.288771922 | 2.172815688 | 0.337022888 | FALSE | TRUE | FALSE |
| SLITRK6 | 0.0316136 | 0.88094034 | 1 | 0.031613639 | 0.031613639 | 0.623337432 | 0.04564295 | 0.887657689 | 0.50725 | 0.406158914 | 0.406158914 | 1.543274038 | 0.692629207 | -0.159499199 | FALSE | TRUE | FALSE |

| | | | | | | | | | | | | | | | | | |
|---|---|---|---|---|---|---|---|---|---|---|---|---|---|---|---|---|---|
| TRAK2 | 0.0315207 | 0.88094895 | 0.7561 | 0.275704713 | 2.475175674 | 9.296958897 | 3.3038516 | 0.394644036 | 0.8913 | 30.60017488 | 2825.698348 | 28139.70416 | 0.009540605 | -2.020424083 | FALSE | FALSE | FALSE |
| FGFR4 | 0.0314862 | 0.88095755 | 0.27165 | 0.007879196 | 0.031486155 | 0.266153837 | 0.01636157 | 0.894759103 | 0.7586 | 0.016361568 | 0.065392092 | 0.3206745 | 1.924397152 | 0.284294706 | FALSE | TRUE | FALSE |
| NYNRIN | 0.0313778 | 0.88096615 | 1 | 0.279479984 | 0.604003334 | 5.379811689 | 0.04822033 | 0.887062286 | 0.0453 | 0.048220332 | 0.048220332 | 1.61887974 | 0.650717936 | -0.186607222 | FALSE | TRUE | FALSE |
| RFTN1 | 0.0313693 | 0.88097475 | 1 | 0.272435863 | 0.272435863 | 1.062226035 | 0.04778799 | 0.887185091 | 1 | 0.04778799 | 0.04778799 | 0.945637897 | 0.656426742 | -0.182813734 | FALSE | TRUE | FALSE |
| DNAJC16 | 0.031368 | 0.88098335 | 1 | 0.031368024 | 0.031368024 | 0.617785551 | 0.04689714 | 0.887364772 | 0.01875 | 0.046897141 | 0.046897141 | 0.925324437 | 0.668868589 | -0.174659199 | FALSE | TRUE | FALSE |
| VRK3 | 0.0313654 | 0.88099195 | 0.74355 | 0.273902727 | 2.455028428 | 2.455028428 | 0.42252865 | 0.783618125 | 0.5066 | 0.422528645 | 0.422528645 | 0.949727222 | 0.074232607 | -1.129405286 | FALSE | TRUE | FALSE |
| MAT1A | 0.0313647 | 0.88100055 | 0.01135 | 0.031364658 | 0.031364658 | 0.62136222 | 2.91731391 | 0.417932052 | 0.1366 | 0.729487118 | 2.917313907 | 2.917313907 | 0.010751211 | -1.968542607 | FALSE | FALSE | FALSE |
| ZNF560 | 0.0313604 | 0.88100914 | 0.87555 | 2.485459861 | 22.99335968 | 22.99335968 | 0.43899639 | 0.777687845 | 0.01765 | 0.438996389 | 0.438996389 | 1.670620943 | 0.071436556 | -1.14607949 | FALSE | TRUE | FALSE |
| GPX5 | 0.0313346 | 0.88101774 | 1 | 2.76020168 | 26.91851587 | 26.91851587 | 0.03747764 | 0.889583459 | 0.50595 | 0.348831558 | 0.348831558 | 0.348831558 | 0.836088969 | -0.077747507 | FALSE | TRUE | FALSE |
| RIMS2 | 0.0312497 | 0.88102633 | 0.9726 | 2.66156453 | 51.76394664 | 250.292408 | 0.55860272 | 0.729391393 | 0.52105 | 5.260846495 | 102.6565607 | 502.2729659 | 0.05594258 | -1.252257507 | FALSE | TRUE | FALSE |
| SORBS1 | 0.0312379 | 0.88103493 | 1 | 0.031237905 | 0.031237905 | 1.05094225 | 0.04654746 | 0.88743813 | 1 | 0.046547461 | 0.046547461 | 1.567981405 | 0.671097921 | -0.173214107 | FALSE | TRUE | FALSE |
| CDS1 | 0.0312101 | 0.88104352 | 1 | 0.270755053 | 0.270755053 | 0.270755053 | 0.04755649 | 0.887234144 | 1 | 0.047556488 | 0.047556488 | 0.047556488 | 0.656275024 | -0.182914124 | FALSE | TRUE | FALSE |
| DYNC1H1 | 0.0311542 | 0.88105211 | 0.5656 | 0.031154207 | 0.560878517 | 1.153416185 | 0.02679379 | 0.891974703 | 0.1213 | 0.026793794 | 0.488232871 | 0.882376521 | 1.16273966 | 0.065482486 | FALSE | TRUE | FALSE |
| PALLD | 0.0311392 | 0.8810607 | 0.33405 | 0.031139173 | 2.33121148 | 5.207881745 | 0.00976529 | 0.896813022 | 0.51595 | 0.009765293 | 0.009765293 | 0.191607975 | 3.188759848 | 0.503621813 | FALSE | TRUE | FALSE |
| AGBL5 | 0.0311234 | 0.88106928 | 1 | 2.16461E+27 | 2.65718E+35 | 6.7803E+37 | 4.64727565 | 0.321483345 | 1 | 4.58746E+27 | 5.48214E+34 | 2.29541E+38 | 0.006697137 | -2.174110846 | FALSE | FALSE | FALSE |
| BRI3 | 0.0311188 | 0.88107787 | 1 | 0.293066704 | 0.293066704 | 0.293066704 | 0.30660413 | 0.823298399 | 0.5057 | 0.30660413 | 0.30660413 | 0.30660413 | 0.101494972 | -0.993555472 | FALSE | TRUE | FALSE |
| RASA2 | 0.0310776 | 0.88108646 | 0.9406 | 0.124279165 | 1.988955917 | 2.450664295 | 0.05627397 | 0.885426807 | 0.51355 | 0.056273975 | 0.056273975 | 0.475549335 | 0.552255575 | -0.257859891 | FALSE | TRUE | FALSE |
| SULF2 | 0.0309602 | 0.88109504 | 0.2622 | 0.003264841 | 0.030960243 | 0.063818762 | 0.00096153 | 0.899481917 | 1 | 0.000961532 | 0.000961532 | 0.018846591 | 32.19886741 | 1.507840596 | FALSE | TRUE | FALSE |
| KIF5A | 0.030953 | 0.88110363 | 1 | 0.030953017 | 0.030953017 | 0.604230654 | 0.04618424 | 0.88751954 | 1 | 0.184614314 | 0.184614314 | 3.614239052 | 0.670207329 | -0.173790827 | FALSE | TRUE | FALSE |
| IL2 | 0.0309512 | 0.88111221 | 1 | 0.030951231 | 0.030951231 | 0.030951231 | 0.03129931 | 0.890914124 | 0.00285 | 0.031299308 | 0.031299308 | 0.031299308 | 0.988879084 | -0.004856809 | FALSE | TRUE | FALSE |
| PLEKHA4 | 0.0309259 | 0.88112079 | 0.0289 | 0.030925877 | 0.030925877 | 0.605488557 | 0.39599806 | 0.793417085 | 0.52145 | 0.39599806 | 0.39599806 | 7.772800065 | 0.07809603 | -1.10737104 | FALSE | TRUE | FALSE |
| ARHGEF17 | 0.0308713 | 0.88112937 | 0.06395 | 0.030871329 | 0.586903194 | 1.033178732 | 0.04501078 | 0.887860273 | 0.5247 | 0.414378181 | 0.860824965 | 7.92492601 | 0.68586527 | -0.163761188 | FALSE | TRUE | FALSE |
| PWP1 | 0.0308705 | 0.88113795 | 0.0146 | 0.030870493 | 0.030870493 | 0.61128892 | 0.1818436 | 0.85657197 | 0.5075 | 0.1818436 | 0.1818436 | 0.901414286 | 0.169763976 | -0.770154461 | FALSE | TRUE | FALSE |
| ENPP6 | 0.0308639 | 0.88114653 | 0.0127 | 0.030863923 | 0.030863923 | 0.611013149 | 0.18361867 | 0.856033484 | 0.50965 | 0.183618666 | 0.183618666 | 0.910406438 | 0.168087067 | -0.7744657 | FALSE | TRUE | FALSE |
| ISYNA1 | 0.0308471 | 0.8811551 | 1 | 0.030847137 | 0.030847137 | 0.606599334 | 0.04620281 | 0.887511404 | 0.01775 | 0.046202814 | 0.046202814 | 0.910397752 | 0.667646276 | -0.175453569 | FALSE | TRUE | FALSE |
| MS4A8 | 0.0308228 | 0.88116368 | 0.0071 | 0.030822802 | 0.030822802 | 0.030822802 | 0.03487582 | 0.890162865 | 1 | 0.034875816 | 0.034875816 | 0.034875816 | 0.883787254 | -0.053652266 | FALSE | TRUE | FALSE |
| RGMB | 0.0307829 | 0.88117225 | 1 | 0.030782918 | 0.030782918 | 0.610088206 | 0.71826134 | 0.65715558 | 0.50915 | 0.718261339 | 2.872081409 | 2.872081409 | 0.042857545 | -1.367972709 | FALSE | TRUE | FALSE |
| SMARCA2 | 0.0307797 | 0.88118083 | 0.52555 | 0.030779713 | 0.030779713 | 0.59530019 | 0.00629812 | 0.897805764 | 0.87805 | 0.231598976 | 2.187417542 | 7.783708988 | 4.887130418 | 0.689053929 | FALSE | TRUE | FALSE |
| TTC8 | 0.0307655 | 0.8811894 | 0.0136 | 0.03076546 | 0.03076546 | 0.609309581 | 0.04484446 | 0.887892623 | 1 | 0.408899225 | 0.408899225 | 0.889185709 | 0.686048151 | -0.163645402 | FALSE | TRUE | FALSE |
| MORC1 | 0.0307434 | 0.88119797 | 1 | 0.272336516 | 0.272336516 | 5.358413165 | 0.04729878 | 0.887266827 | 1 | 0.047298781 | 0.047298781 | 0.93245343 | 0.649983522 | -0.187097653 | FALSE | TRUE | FALSE |
| MAP3K13 | 0.0307291 | 0.88120654 | 0.02895 | 0.030729076 | 0.030729076 | 0.602061287 | 0.04630273 | 0.887470707 | 1 | 0.046302728 | 0.046302728 | 0.90941487 | 0.663655847 | -0.178057075 | FALSE | TRUE | FALSE |
| GABRP | 0.0307066 | 0.88121511 | 0.50795 | 0.122748763 | 0.490806679 | 0.608437398 | 0.86748185 | 0.603715985 | 0.13465 | 0.216931102 | 0.867481847 | 0.867481847 | 0.035397446 | -1.451028077 | FALSE | TRUE | FALSE |
| SULT1A2 | 0.0306455 | 0.88122367 | 0.51335 | 0.122541719 | 0.122541719 | 0.127403324 | 0.1812033 | 0.856776068 | 0.75155 | 0.181203303 | 0.724782238 | 0.724782238 | 0.169122016 | -0.771799852 | FALSE | TRUE | FALSE |
| FRMPD1 | 0.0305933 | 0.88123224 | 0.75975 | 0.12233441 | 1.071313686 | 5.189988043 | 2.02228313 | 0.488214197 | 0.33325 | 0.505711535 | 18.3669666 | 39.32387507 | 0.015128077 | -1.820216267 | FALSE | FALSE | FALSE |
| NDUFAF6 | 0.0305835 | 0.8812408 | 1 | 0.122280331 | 0.122280331 | 0.122280331 | 0.40583619 | 0.790026774 | 0.50315 | 0.40583619 | 0.40583619 | 0.40583619 | 0.075359298 | -1.122863158 | FALSE | TRUE | FALSE |
| C3 | 0.0305668 | 0.88124937 | 1 | 0.030566848 | 0.586387716 | 1.025611907 | 0.36180845 | 0.805059144 | 0.96935 | 32.80454423 | 632.416057 | 3283.618904 | 0.084483512 | -1.07322804 | FALSE | TRUE | FALSE |
| DZIP1L | 0.030539 | 0.88125793 | 1 | 0.266371523 | 2.384707853 | 5.238892747 | 0.41209438 | 0.787561663 | 0.51005 | 0.412094377 | 0.412094377 | 8.121129347 | 0.074106919 | -1.130141243 | FALSE | TRUE | FALSE |
| CPNE7 | 0.0305327 | 0.88126649 | 1 | 0.269361525 | 1.077244979 | 2.409372014 | 0.04218435 | 0.888496077 | 0.9389 | 3.408477085 | 304.8342709 | 304.8342709 | 0.723792959 | -0.140385646 | FALSE | TRUE | FALSE |
| FGD4 | 0.0305103 | 0.88127505 | 1 | 0.030510349 | 0.030510349 | 0.60243334 | 0.045308 | 0.887779321 | 0.5156 | 0.181167086 | 0.181167086 | 1.531630638 | 0.673398744 | -0.171727698 | FALSE | TRUE | FALSE |
| HNRNPL | 0.030509 | 0.88128361 | 1 | 0.030509011 | 0.030509011 | 0.603023796 | 1.5354367 | 0.532472815 | 0.00345 | 0.04531247 | 0.04531247 | 0.896884375 | 0.019869924 | -1.701803788 | FALSE | FALSE | FALSE |
| SLC9A9 | 0.0304996 | 0.88129217 | 1 | 0.030499645 | 0.030499645 | 0.6028263 | 0.04537462 | 0.88774691 | 0.0158 | 0.045374618 | 0.045374618 | 0.898098235 | 0.672174148 | -0.172518195 | FALSE | TRUE | FALSE |
| AGXT | 0.0304918 | 0.88130073 | 0.88635 | 0.974658226 | 15.58654962 | 15.58654962 | 28.6058959 | 0.0963585 | 0.42985 | 16.45841341 | 2487.125717 | 2487.125717 | 0.001065926 | -2.972272906 | FALSE | FALSE | FALSE |
| BMPR1B | 0.0304654 | 0.88130928 | 1 | 0.486646857 | 1.945719919 | 2.41240158 | 0.04765113 | 0.887217797 | 0.0111 | 0.047651126 | 0.047651126 | 0.945256543 | 0.639342515 | -0.194266415 | FALSE | TRUE | FALSE |
| KLC2 | 0.0304613 | 0.88131784 | 1 | 0.030461258 | 0.030461258 | 0.599137977 | 0.71914381 | 0.656463765 | 0.2632 | 0.179866036 | 0.719143808 | 3.544810071 | 0.042357673 | -1.37306791 | FALSE | TRUE | FALSE |
| AP4B1 | 0.0304164 | 0.88132639 | 1 | 0.030416369 | 0.030416369 | 0.599316697 | 0.37476488 | 0.800930677 | 0.7543 | 0.374764883 | 3.37798257 | 7.397440263 | 0.081161202 | -1.090651528 | FALSE | TRUE | FALSE |
| LIG3 | 0.0304136 | 0.88133494 | 0.2711 | 0.030413643 | 0.121625 | 1.025527042 | 0.11621124 | 0.872157386 | 0.50785 | 0.116211238 | 0.464644533 | 2.280121791 | 0.261709998 | -0.582179685 | FALSE | TRUE | FALSE |
| C2CD5 | 0.0303738 | 0.88134349 | 1 | 0.268051614 | 0.268051614 | 5.264835856 | 0.40377728 | 0.791088268 | 0.50435 | 0.403777278 | 0.403777278 | 7.947774945 | 0.07522419 | -1.123642481 | FALSE | TRUE | FALSE |
| PDE4D | 0.0303609 | 0.88135204 | 1 | 0.030360942 | 0.030360942 | 0.599530117 | 0.04578791 | 0.887608966 | 0.01565 | 0.045787913 | 0.045787913 | 0.905540351 | 0.663077649 | -0.178435611 | FALSE | TRUE | FALSE |
| PARPBP | 0.0303429 | 0.88136059 | 1 | 0.030342891 | 0.030342891 | 0.601899294 | 0.04508097 | 0.887852182 | 0.00995 | 0.045080969 | 0.045080969 | 0.045080969 | 0.673075386 | -0.171936291 | FALSE | TRUE | FALSE |
| ACCSL | 0.0302941 | 0.88136914 | 0.75405 | 0.0664364 | 0.265589634 | 0.59895765 | 0.05420986 | 0.885845883 | 0.75245 | 0.054209857 | 0.21678054 | 0.45941405 | 0.558830053 | -0.252720246 | FALSE | TRUE | FALSE |
| ITGA3 | 0.0302873 | 0.88137768 | 1 | 0.030287318 | 0.030287318 | 0.591532604 | 0.0455643 | 0.887682035 | 1 | 0.045564304 | 0.045564304 | 0.892418749 | 0.664715924 | -0.177363917 | FALSE | TRUE | FALSE |
| VIPR1 | 0.0302647 | 0.88138623 | 1 | 0.030264678 | 0.030264678 | 0.599904807 | 3.44718038 | 0.386210101 | 0.25025 | 0.378346427 | 3.447180378 | 3.447180378 | 0.008779546 | -2.056527956 | FALSE | FALSE | FALSE |
| PCDHGA4 | 0.0302347 | 0.88139477 | 1 | 0.03023472 | 0.03023472 | 0.591509722 | 0.04126127 | 0.88865602 | 1 | 3.28135967 | 30.40788784 | 64.36487869 | 0.732762725 | -0.135036631 | FALSE | TRUE | FALSE |
| PI4KA | 0.0301618 | 0.88140331 | 0.70625 | 0.260070439 | 10.98358222 | 43.91395887 | 0.00892623 | 0.897042069 | 0.52365 | 0.008926229 | 0.04285477 | 0.17134897 | 3.379010614 | 0.528789556 | FALSE | TRUE | FALSE |
| MRPS22 | 0.030075 | 0.88141186 | 1 | 0.030074989 | 0.030074989 | 0.596566184 | 3.49376337 | 0.383195712 | 0.5023 | 3.493763367 | 33.37079069 | 33.37079069 | 0.008608193 | -2.065088006 | FALSE | FALSE | FALSE |
| ZDHHC13 | 0.0300705 | 0.8814204 | 0.50035 | 0.263474443 | 0.263474443 | 0.596324061 | 0.04620406 | 0.887503267 | 0.01075 | 0.04620406 | 0.04620406 | 0.917198017 | 0.650819761 | -0.186539269 | FALSE | TRUE | FALSE |
| TAOK2 | 0.0300695 | 0.88142894 | 0.0436 | 0.030069525 | 0.030069525 | 1.012305129 | 0.04540226 | 0.887738805 | 0.0388 | 0.045402257 | 0.045402257 | 1.530297872 | 0.662291424 | -0.178950868 | FALSE | TRUE | FALSE |
| CA9 | 0.0300362 | 0.88143748 | 1 | 0.264901356 | 0.264901356 | 0.594154289 | 0.38748421 | 0.796568612 | 0.2579 | 0.387484209 | 3.533825107 | 3.533825107 | 0.077515967 | -1.11060883 | FALSE | TRUE | FALSE |
| TMEM154 | 0.0300249 | 0.88144601 | 1 | 0.030024895 | 0.030024895 | 0.030024895 | 0.03500914 | 0.890116091 | 0.0046 | 0.035009135 | 0.035009135 | 0.035009135 | 0.857630293 | -0.066699887 | FALSE | TRUE | FALSE |
| NTN5 | 0.0299046 | 0.88145455 | 0.9927 | 208.3641174 | 20156.90678 | 20156.90678 | 0.05276329 | 0.886071039 | 0.752 | 0.470369977 | 1.881149973 | 1.881149973 | 0.566769279 | -0.246593698 | FALSE | TRUE | FALSE |
| FAM81B | 0.0298941 | 0.88146308 | 0.012 | 0.029894101 | 0.029894101 | 0.066552404 | 3.15713224 | 0.403318385 | 0.3147 | 0.334926663 | 30.50572098 | 30.50572098 | 0.009468752 | -2.023707273 | FALSE | FALSE | FALSE |
| PRKCSH | 0.0298569 | 0.88147162 | 1 | 0.029856943 | 0.029856943 | 0.58927429 | 0.04512756 | 0.887827905 | 0.0146 | 0.045127555 | 0.045127555 | 0.892075717 | 0.661612233 | -0.179396473 | FALSE | TRUE | FALSE |
| EHBP1 | 0.0298477 | 0.88148015 | 1 | 0.276073861 | 0.276073861 | 5.401833612 | 0.04659057 | 0.887421835 | 1 | 0.046590566 | 0.046590566 | 0.913999617 | 0.640638012 | -0.193387296 | FALSE | TRUE | FALSE |
| GIMAP8 | 0.0298389 | 0.88148868 | 1 | 0.26037124 | 0.26037124 | 1.009480949 | 13.5905971 | 0.165501728 | 0.13725 | 1.504412428 | 13.59059707 | 13.59059707 | 0.002195554 | -2.658455784 | FALSE | FALSE | FALSE |
| IQCG | 0.0298357 | 0.88149721 | 0.0111 | 0.029835716 | 0.029835716 | 0.591748899 | 0.04312763 | 0.888287544 | 1 | 0.377688029 | 0.377688029 | 0.856231014 | 0.691800559 | -0.160019091 | FALSE | TRUE | FALSE |
| GALNT11 | 0.0298314 | 0.88150574 | 0.0192 | 0.029831415 | 0.029831415 | 0.588310145 | 0.04191554 | 0.888544101 | 1 | 1.537133085 | 14.47132213 | 14.47132213 | 0.711702935 | -0.147701243 | FALSE | TRUE | FALSE |
| DIAPH1 | 0.0297901 | 0.88151427 | 0.5169 | 0.02979015 | 0.02979015 | 0.582790726 | 0.00673002 | 0.89762095 | 0.0272 | 0.006730022 | 0.006730022 | 0.132007167 | 4.426456329 | 0.646056184 | FALSE | TRUE | FALSE |
| ME2 | 0.0297822 | 0.8815228 | 1 | 0.029782243 | 0.029782243 | 0.589412042 | 0.04335808 | 0.888215195 | 1 | 0.387346493 | 0.387346493 | 1.467315633 | 0.686890192 | -0.163112685 | FALSE | TRUE | FALSE |
| SIX5 | 0.0297453 | 0.88153132 | 1 | 0.275175418 | 0.275175418 | 1.006951652 | 0.18357721 | 0.856072077 | 0.50755 | 0.183577207 | 0.183577207 | 0.908336475 | 0.162031288 | -0.790401115 | FALSE | TRUE | FALSE |
| PHLPP2 | 0.0297168 | 0.88153985 | 1 | 0.029716754 | 0.029716754 | 0.999537809 | 0.04461981 | 0.887965348 | 0.03055 | 0.044619809 | 0.044619809 | 0.873031027 | 0.665999121 | -0.176526344 | FALSE | TRUE | FALSE |
| ZNF823 | 0.0296798 | 0.88154837 | 1 | 0.029679766 | 0.029679766 | 0.587002095 | 0.0444986 | 0.888005713 | 1 | 0.044498597 | 0.044498597 | 0.881262808 | 0.666982058 | -0.175885848 | FALSE | TRUE | FALSE |
| ARHGAP39 | 0.0296597 | 0.88155689 | 1 | 2500.118227 | 3094758.73 | 34473459.92 | 1.66891075 | 0.520436654 | 0.9946 | 6117.384072 | 1742229.785 | 20114836.82 | 0.017771879 | -1.750266647 | FALSE | FALSE | FALSE |
| STXBP4 | 0.0296004 | 0.88156541 | 0.5073 | 0.118380664 | 0.118380664 | 0.58642833 | 0.04479647 | 0.887908792 | 0.01255 | 0.04479647 | 0.04479647 | 0.888491279 | 0.660775353 | -0.179946165 | FALSE | TRUE | FALSE |

| | | | | | | | | | | | | | | | | | |
|---|---|---|---|---|---|---|---|---|---|---|---|---|---|---|---|---|---|
| CYP2C19 | 0.0295883 | 0.88157393 | 0.7496 | 0.118311422 | 0.473197461 | 2.343534085 | 0.04547749 | 0.88772259 | 0.01175 | 0.045477493 | 0.045477493 | 0.901865642 | 0.650613374 | -0.186677014 | FALSE | TRUE | FALSE |
| MEIS3 | 0.0295451 | 0.88158245 | 0.0153 | 0.029545141 | 0.029545141 | 0.584915369 | 0.39299985 | 0.794417649 | 0.50495 | 0.392999846 | 0.392999846 | 0.857179437 | 0.075178505 | -1.123906316 | FALSE | TRUE | FALSE |
| ADAMTS2 | 0.0295145 | 0.88159097 | 0.8785 | 1.087585369 | 10.28853007 | 36.66096708 | 0.18583648 | 0.855529973 | 0.88 | 0.743182653 | 2.972820577 | 14.44641763 | 0.158819579 | -0.79909596 | FALSE | TRUE | FALSE |
| KTN1 | 0.029506 | 0.88159949 | 0.51995 | 0.029505988 | 0.029505988 | 0.578200399 | 0.00668127 | 0.897634662 | 0.02615 | 0.006681268 | 0.006681268 | 0.131244862 | 4.416225719 | 0.645051263 | FALSE | TRUE | FALSE |
| SLC4A5 | 0.0294812 | 0.881608 | 1 | 3.01728E+58 | 4.15539E+70 | 1.93337E+75 | 0.32644476 | 0.81658454 | 1 | 3.22652E+42 | 4.53819E+52 | 3.67307E+56 | 0.09030985 | -1.044264881 | FALSE | TRUE | FALSE |
| MYCBPAP | 0.0294803 | 0.88161652 | 0.0288 | 0.0294803 | 0.0294803 | 0.577773791 | 0.36729845 | 0.803232848 | 0.75345 | 0.367298452 | 3.329275454 | 12.40616102 | 0.080262523 | -1.095487193 | FALSE | TRUE | FALSE |
| EML4 | 0.0294622 | 0.88162503 | 0.51345 | 0.029462201 | 0.029462201 | 0.578837964 | 0.0132767 | 0.895724554 | 0.02365 | 0.013276699 | 0.013276699 | 0.26139927 | 2.219090844 | 0.346175081 | FALSE | TRUE | FALSE |
| NUAK2 | 0.029457 | 0.88163354 | 1 | 0.263075256 | 0.263075256 | 5.176452889 | 0.04431892 | 0.888046051 | 1 | 0.398571415 | 0.398571415 | 1.500354832 | 0.664659306 | -0.17740091 | FALSE | TRUE | FALSE |
| CYP2C8 | 0.0294358 | 0.88164205 | 0.5069 | 0.262636356 | 0.262636356 | 0.584384677 | 0.04561882 | 0.887673921 | 1 | 0.045618822 | 0.045618822 | 0.045618822 | 0.645255939 | -0.19026799 | FALSE | TRUE | FALSE |
| SAMD9 | 0.0294249 | 0.88165056 | 0.85325 | 0.87891874 | 242.847918 | 975.7336406 | 1.24169501 | 0.562529333 | 0.5176 | 2.550849051 | 199.5721455 | 1377.877866 | 0.02369739 | -1.625299476 | FALSE | FALSE | FALSE |
| KRT24 | 0.0294053 | 0.88165907 | 0.90695 | 0.469685604 | 9.952969258 | 39.78300792 | 0.91016401 | 0.597070737 | 0.6461 | 2.085414018 | 78.31944708 | 313.1637065 | 0.032307715 | -1.490693756 | FALSE | TRUE | FALSE |
| ROBO2 | 0.0294045 | 0.88166758 | 0.5179 | 0.271509255 | 0.271509255 | 5.277227996 | 0.04595856 | 0.887560204 | 0.0377 | 0.045958562 | 0.045958562 | 1.546147821 | 0.639803698 | -0.193953254 | FALSE | TRUE | FALSE |
| ADAMDEC1 | 0.0294033 | 0.88167609 | 1 | 0.25851241 | 0.25851241 | 0.25851241 | 0.37886896 | 0.799696954 | 0.7512 | 0.378868963 | 3.445687079 | 3.445687079 | 0.077608187 | -1.110092461 | FALSE | TRUE | FALSE |
| NCAN | 0.0293946 | 0.88168459 | 0.0385 | 0.029394594 | 0.029394594 | 0.989388331 | 0.04269017 | 0.888399919 | 1 | 0.393736789 | 1.57424474 | 13.26872446 | 0.688556528 | -0.1620604 | FALSE | TRUE | FALSE |
| TRIM50 | 0.029387 | 0.88169309 | 1 | 2.351220404 | 21.85459161 | 21.85459161 | 0.04816324 | 0.887095058 | 1 | 0.048163236 | 0.048163236 | 0.953318955 | 0.610154388 | -0.214560261 | FALSE | TRUE | FALSE |
| ZNF620 | 0.0293657 | 0.8817016 | 0.0094 | 0.029365686 | 0.029365686 | 0.029365686 | 0.04390136 | 0.888126652 | 0.00985 | 0.043901355 | 0.043901355 | 0.043901355 | 0.668901573 | -0.174637783 | FALSE | TRUE | FALSE |
| SEMA4D | 0.0293566 | 0.8817101 | 1 | 0.029356564 | 0.029356564 | 0.575119635 | 0.04399499 | 0.888094424 | 1 | 0.043994988 | 0.043994988 | 0.864022314 | 0.667270646 | -0.17569798 | FALSE | TRUE | FALSE |
| FZD1 | 0.029324 | 0.8817186 | 0.9917 | 41.32105011 | 4032.29865 | 41312.99761 | 0.21001609 | 0.849703592 | 0.51825 | 0.210016092 | 0.210016092 | 4.131380451 | 0.13962733 | -0.855029567 | FALSE | TRUE | FALSE |
| SLC29A4 | 0.0292965 | 0.8817271 | 0.0206 | 0.029296469 | 0.029296469 | 0.576163931 | 0.04420475 | 0.888070242 | 1 | 0.044204747 | 0.044204747 | 0.871104392 | 0.662744858 | -0.178653633 | FALSE | TRUE | FALSE |
| CES4A | 0.0292926 | 0.88173559 | 0.0104 | 0.029292579 | 0.029292579 | 0.580960923 | 0.37889288 | 0.799672122 | 0.50665 | 0.378892883 | 0.378892883 | 0.846745172 | 0.077310976 | -1.111758844 | FALSE | TRUE | FALSE |
| KRT7 | 0.0292842 | 0.88174409 | 0.76045 | 0.029284199 | 0.117041049 | 0.577487269 | 1.81608615 | 0.505977107 | 0.38415 | 0.791572428 | 7.261080352 | 29.03852455 | 0.016124895 | -1.792503101 | FALSE | FALSE | FALSE |
| APEH | 0.0292443 | 0.88175259 | 0.0235 | 0.029244311 | 0.029244311 | 0.575192124 | 0.04378025 | 0.888142759 | 1 | 0.043780255 | 0.043780255 | 0.862810653 | 0.667979464 | -0.175236889 | FALSE | TRUE | FALSE |
| FOXO1 | 0.0291935 | 0.88176108 | 0.5068 | 0.116652636 | 0.116652636 | 0.988116292 | 0.17528173 | 0.858251779 | 0.7483 | 0.175281735 | 0.700657561 | 1.4853651 | 0.16655161 | -0.778451164 | FALSE | TRUE | FALSE |
| TULP4 | 0.0291787 | 0.88176957 | 0.94175 | 1.084283876 | 41.36369344 | 145.8156099 | 0.04819338 | 0.887078674 | 1 | 0.048193383 | 0.048193383 | 1.622893212 | 0.605450126 | -0.217921626 | FALSE | TRUE | FALSE |
| HAUS1 | 0.0291646 | 0.88177807 | 0.0067 | 0.029164561 | 0.029164561 | 0.029164561 | 0.03266523 | 0.890675027 | 1 | 0.032665232 | 0.032665232 | 0.032665232 | 0.892831887 | -0.049230308 | FALSE | TRUE | FALSE |
| LILRB2 | 0.0291341 | 0.88178656 | 1 | 3.75596E+21 | 2.22332E+27 | 7.7995E+30 | 0.0599509 | 0.884733589 | 1 | 2.51745E+15 | 9.3939E+20 | 1.79682E+23 | 0.48596593 | -0.313394177 | FALSE | TRUE | FALSE |
| ACTL7B | 0.029106 | 0.88179505 | 1 | 0.11632724 | 0.11632724 | 0.574724402 | 0.40403781 | 0.79095281 | 0.50825 | 0.404037809 | 0.404037809 | 0.856918716 | 0.072037872 | -1.142439125 | FALSE | TRUE | FALSE |
| NOM1 | 0.0290889 | 0.88180354 | 0.0232 | 0.029088877 | 0.029088877 | 0.571682723 | 0.0422551 | 0.888488069 | 0.51115 | 0.371780837 | 0.371780837 | 7.321860164 | 0.68841097 | -0.162152218 | FALSE | TRUE | FALSE |
| SHC2 | 0.0290818 | 0.88181202 | 1 | 0.029081787 | 0.029081787 | 0.5746833 | 0.04388361 | 0.888134706 | 1 | 0.043883611 | 0.043883611 | 0.868428341 | 0.662702692 | -0.178681265 | FALSE | TRUE | FALSE |
| CC2D2A | 0.0290643 | 0.88182051 | 0.49095 | 0.015607873 | 0.565405602 | 1.213282808 | 0.00230791 | 0.898942265 | 0.8203 | 0.019558875 | 0.680093838 | 1.525992058 | 12.59334103 | 1.100140964 | FALSE | TRUE | FALSE |
| DXO | 0.0290576 | 0.88182899 | 0.01445 | 0.029057555 | 0.029057555 | 0.57544774 | 0.0434124 | 0.88820715 | 1 | 0.043412398 | 0.043412398 | 0.860741942 | 0.669337714 | -0.174354704 | FALSE | TRUE | FALSE |
| ITGAL | 0.0290555 | 0.88183748 | 1 | 0.029055494 | 0.029055494 | 0.566738415 | 0.04393189 | 0.88811054 | 1 | 0.04393189 | 0.04393189 | 0.85946188 | 0.661375911 | -0.179551627 | FALSE | TRUE | FALSE |
| PDXDC1 | 0.0290481 | 0.88184596 | 0.02065 | 0.029048067 | 0.029048067 | 0.571710969 | 0.04236077 | 0.888456028 | 1 | 0.376779629 | 0.376779629 | 1.43314173 | 0.685730334 | -0.163846639 | FALSE | TRUE | FALSE |
| KIFC3 | 0.0290465 | 0.88185444 | 0.75835 | 0.264294365 | 2.468207716 | 8.926140189 | 0.04621124 | 0.887495129 | 1 | 0.184767268 | 0.184767268 | 3.626580835 | 0.628558656 | -0.201654189 | FALSE | TRUE | FALSE |
| TK2 | 0.0289535 | 0.88186292 | 1 | 0.028953511 | 0.028953511 | 0.028953511 | 0.16971242 | 0.859771909 | 0.87275 | 0.678580039 | 2.713920238 | 2.713920238 | 0.170603376 | -0.768012378 | FALSE | TRUE | FALSE |
| ABRA | 0.0289018 | 0.8818714 | 1 | 0.115533054 | 0.115533054 | 0.573106735 | 309.573584 | 0.007828786 | 0.19265 | 31.38751553 | 309.5735835 | 3127.884751 | 9.33602E-05 | -4.029838407 | FALSE | FALSE | FALSE |
| TANC2 | 0.0288906 | 0.88187988 | 1 | 2.54418074 | 247.9585079 | 476.6319056 | 0.03222234 | 0.890736815 | 1 | 305166.7941 | 394633924 | 7617991219 | 0.896601315 | -0.047400628 | FALSE | TRUE | FALSE |
| TRPC1 | 0.028839 | 0.88188836 | 0.51035 | 0.11528349 | 0.11528349 | 0.974479738 | 0.0416297 | 0.888592089 | 1 | 0.38320583 | 3.620378685 | 7.56441263 | 0.692751167 | -0.159422734 | FALSE | TRUE | FALSE |
| IL21 | 0.0287906 | 0.88189683 | 0.00365 | 0.028790573 | 0.028790573 | 0.028790573 | 0.02948573 | 0.891358943 | 1 | 0.029485731 | 0.029485731 | 0.029485731 | 0.976423973 | -0.010361594 | FALSE | TRUE | FALSE |
| CNTN6 | 0.0287689 | 0.88190531 | 0.5064 | 0.06168147 | 2.164436829 | 19.47451781 | 0.01060756 | 0.896603996 | 0.3859 | 0.01060756 | 0.360993895 | 0.360993895 | 2.71211324 | 0.433307819 | FALSE | TRUE | FALSE |
| TMEM37 | 0.0287439 | 0.88191378 | 1 | 0.028743928 | 0.028743928 | 0.028743928 | 0.03009734 | 0.891213586 | 0.00545 | 0.030097344 | 0.030097344 | 0.030097344 | 0.955032027 | -0.019982064 | FALSE | TRUE | FALSE |
| ATP2A2 | 0.0287101 | 0.88192226 | 1 | 0.269026236 | 0.269026236 | 5.263961402 | 0.40564301 | 0.790190823 | 0.5106 | 0.405643013 | 1.62142921 | 7.957816439 | 0.070776884 | -1.150108559 | FALSE | TRUE | FALSE |
| DICER1 | 0.0286876 | 0.88193073 | 1 | 0.265901429 | 0.265901429 | 5.135647513 | 0.17788219 | 0.857498678 | 0.52315 | 0.17788219 | 0.17788219 | 3.449849431 | 0.161273178 | -0.792437857 | FALSE | TRUE | FALSE |
| APLF | 0.0286128 | 0.8819392 | 1 | 0.249913753 | 0.249913753 | 0.567856048 | 0.37817688 | 0.799895387 | 0.01005 | 0.378176883 | 0.378176883 | 0.852049805 | 0.075659943 | -1.121133988 | FALSE | TRUE | FALSE |
| DDX55 | 0.0285849 | 0.88194767 | 0.01645 | 0.028584919 | 0.028584919 | 0.065152611 | 0.1993676 | 0.852247039 | 0.76055 | 1.787058404 | 154.8497866 | 619.1551206 | 0.143377955 | -0.843517619 | FALSE | TRUE | FALSE |
| CHST3 | 0.0285694 | 0.88195614 | 0.9931 | 49.82987182 | 9860.674039 | 26307.70002 | 0.05242345 | 0.88616255 | 0.50635 | 0.209546212 | 0.209546212 | 1.03964519 | 0.544973418 | -0.263624681 | FALSE | TRUE | FALSE |
| GNAT2 | 0.0285191 | 0.8819646 | 1 | 0.028519117 | 0.028519117 | 0.028519117 | 0.04335545 | 0.888223238 | 1 | 0.043355447 | 0.043355447 | 0.043355447 | 0.65779779 | -0.18190759 | FALSE | TRUE | FALSE |
| DHX33 | 0.0285113 | 0.88197307 | 1 | 2.296125564 | 204.6842335 | 204.6842335 | 0.04049224 | 0.88883149 | 0.9853 | 30.97373904 | 2935.068642 | 29608.54499 | 0.704116584 | -0.152355427 | FALSE | TRUE | FALSE |
| FAM83C | 0.028505 | 0.88198153 | 1 | 0.028505006 | 0.028505006 | 0.55996712 | 0.33242365 | 0.814718265 | 0.9367 | 2.985371208 | 259.6648905 | 259.6648905 | 0.085749034 | -1.066770765 | FALSE | TRUE | FALSE |
| TBX3 | 0.0284924 | 0.88199 | 1 | 0.028492437 | 0.028492437 | 0.560955324 | 0.04342513 | 0.888199105 | 1 | 0.043425125 | 0.043425125 | 0.856554182 | 0.656127928 | -0.183011476 | FALSE | TRUE | FALSE |
| SPN | 0.0284803 | 0.88199846 | 0.01195 | 0.028480316 | 0.028480316 | 0.564280631 | 0.03023975 | 0.891198265 | 1 | 0.283275105 | 0.283275105 | 0.283275105 | 0.941817292 | -0.02603334 | FALSE | TRUE | FALSE |
| NAA16 | 0.0284101 | 0.88200692 | 0.5157 | 0.028410144 | 0.028410144 | 0.55991586 | 36.8123185 | 0.075138227 | 0.06745 | 0.442225154 | 36.81231847 | 36.81231847 | 0.000771756 | -3.112519729 | FALSE | FALSE | FALSE |
| PHF10 | 0.0283983 | 0.88201538 | 0.50705 | 0.113542756 | 0.113542756 | 0.563590043 | 0.17181872 | 0.859207097 | 0.49955 | 0.042971472 | 0.171818719 | 0.853598833 | 0.165280414 | -0.781778608 | FALSE | TRUE | FALSE |
| AK7 | 0.0283405 | 0.88202384 | 0.02075 | 0.028340475 | 0.028340475 | 0.558542975 | 0.00655042 | 0.897703167 | 0.01785 | 0.006550417 | 0.006550417 | 0.129324271 | 4.326514895 | 0.636138203 | FALSE | TRUE | FALSE |
| CRISPLD1 | 0.0282968 | 0.8820323 | 1 | 0.028296762 | 0.028296762 | 0.56074451 | 0.04309616 | 0.88830361 | 0.01105 | 0.043096162 | 0.043096162 | 0.854959845 | 0.656595877 | -0.182701849 | FALSE | TRUE | FALSE |
| ORM2 | 0.0282824 | 0.88204076 | 1 | 0.028282441 | 0.028282441 | 0.028282441 | 0.03277046 | 0.890613175 | 1 | 0.032770458 | 0.032770458 | 0.032770458 | 0.863046865 | -0.06396562 | FALSE | TRUE | FALSE |
| COPB2 | 0.0282589 | 0.88204921 | 1 | 0.11293885 | 0.11293885 | 2.223920192 | 0.68119046 | 0.674502751 | 0.2706 | 0.17038446 | 0.681190461 | 3.361334722 | 0.041484614 | -1.382112951 | FALSE | TRUE | FALSE |
| PIK3AP1 | 0.0281588 | 0.88205767 | 1 | 0.266254021 | 2.583812563 | 5.265685523 | 0.03264653 | 0.890690479 | 1 | 0.032646527 | 0.032646527 | 0.642430751 | 0.862536222 | -0.064222658 | FALSE | TRUE | FALSE |
| MIS18A | 0.0281554 | 0.88206612 | 0.0061 | 0.02815542 | 0.02815542 | 0.02815542 | 2.40028551 | 0.45563269 | 0.50305 | 2.400285509 | 2.400285509 | 2.400285509 | 0.01173003 | -1.930700892 | FALSE | FALSE | FALSE |
| CYB5R2 | 0.0280645 | 0.88207458 | 0.50515 | 0.028064517 | 0.028064517 | 0.028064517 | 0.00640175 | 0.897730545 | 0.4953 | 0.006401749 | 0.02559092 | 0.02559092 | 4.383882906 | 0.641858946 | FALSE | TRUE | FALSE |
| TMPRSS13 | 0.0280393 | 0.88208303 | 1 | 0.028039319 | 0.028039319 | 0.553710496 | 1.49639165 | 0.536225282 | 0.26225 | 0.374293747 | 1.496391651 | 3.233662983 | 0.018737955 | -1.727277817 | FALSE | FALSE | FALSE |
| ZNF107 | 0.0280255 | 0.88209148 | 1 | 0.028025489 | 0.028025489 | 0.553757754 | 3.20875333 | 0.40000854 | 0.01185 | 0.352806379 | 3.208753331 | 6.981231743 | 0.008734074 | -2.058783133 | FALSE | FALSE | FALSE |
| ZFP30 | 0.028018 | 0.88209993 | 0.0129 | 0.028018023 | 0.028018023 | 0.555456386 | 0.36187857 | 0.804988634 | 0.5035 | 0.361878571 | 0.361878571 | 0.81257501 | 0.077423825 | -1.111125377 | FALSE | TRUE | FALSE |
| NT5E | 0.0280087 | 0.88210838 | 1 | 0.028008714 | 0.028008714 | 0.552895418 | 0.3470379 | 0.809935383 | 0.877 | 3.22019496 | 122.5423401 | 122.5423401 | 0.080707938 | -1.093083746 | FALSE | TRUE | FALSE |
| ADAMTS19 | 0.0280046 | 0.88211683 | 1 | 0.111968566 | 0.111968566 | 2.1944095 | 0.04305168 | 0.888327701 | 1 | 0.043051684 | 0.043051684 | 0.845786189 | 0.650486823 | -0.186761496 | FALSE | TRUE | FALSE |
| ELN | 0.0280008 | 0.88212527 | 1 | 0.028000815 | 0.028000815 | 0.551003113 | 0.04235917 | 0.88846404 | 0.01935 | 0.042359168 | 0.042359168 | 0.835163293 | 0.661033163 | -0.179776752 | FALSE | TRUE | FALSE |
| LAMC1 | 0.0279851 | 0.88213372 | 0.8843 | 0.111856421 | 0.447202126 | 2.162140533 | 0.01300927 | 0.895781252 | 1 | 0.013009267 | 0.013009267 | 0.437716623 | 2.15116484 | 0.332673691 | FALSE | TRUE | FALSE |
| PRODH2 | 0.0278375 | 0.88214216 | 1 | 0.243756925 | 0.243756925 | 0.942238844 | 195468.277 | 1.00591E-05 | 0.50425 | 195468.2771 | 40073646.16 | 2567778375 | 1.42415E-07 | -6.84644537 | FALSE | FALSE | FALSE |
| ZNF646 | 0.0278096 | 0.8821506 | 1 | 0.027809645 | 0.531873264 | 0.932907249 | 0.04217797 | 0.888504083 | 1 | 0.042177965 | 0.810934637 | 1.41718213 | 0.659340602 | -0.18089018 | FALSE | TRUE | FALSE |
| HSBP1 | 0.0277952 | 0.88215905 | 1 | 0.027795174 | 0.027795174 | 0.027795174 | 0.02820485 | 0.891694211 | 1 | 0.028204853 | 0.028204853 | 0.028204853 | 0.985474852 | -0.006354454 | FALSE | TRUE | FALSE |
| CDH12 | 0.0277928 | 0.88216749 | 1 | 2.438541431 | 236.7940929 | 236.7940929 | 1.30917924 | 0.553552747 | 0.9971 | 12304.60565 | 5510569.595 | 40590884.87 | 0.021229167 | -1.673067056 | FALSE | FALSE | FALSE |
| TMPRSS2 | 0.0277926 | 0.88217593 | 1 | 0.249663512 | 0.249663512 | 0.941144767 | 0.04345333 | 0.888183011 | 1 | 0.04345333 | 0.04345333 | 0.859972304 | 0.639595626 | -0.194094515 | FALSE | TRUE | FALSE |
| GPAA1 | 0.0277912 | 0.88218436 | 0.51485 | 0.24601379 | 0.24601379 | 4.843903596 | 2.67740295 | 0.435467045 | 0.0156 | 0.669163807 | 2.677402954 | 5.666723405 | 0.010379914 | -1.983806239 | FALSE | FALSE | FALSE |

| | | | | | | | | | | | | | | | | | |
|---|---|---|---|---|---|---|---|---|---|---|---|---|---|---|---|---|---|
| HBB | 0.0277694 | 0.8821928 | 1 | 0.243673677 | 0.243673677 | 0.243673677 | 0.04183482 | 0.888568099 | 1 | 0.041834824 | 0.370374953 | 0.370374953 | 0.663786741 | -0.177971427 | FALSE | TRUE | FALSE |
| PAPPA | 0.0277622 | 0.88220124 | 0.5156 | 0.254965749 | 0.254965749 | 4.936333816 | 0.04363836 | 0.888174962 | 0.04185 | 0.043638355 | 0.043638355 | 1.468267542 | 0.636187573 | -0.196414819 | FALSE | TRUE | FALSE |
| DIDO1 | 0.0277545 | 0.88220967 | 1 | 0.027754509 | 0.523570078 | 0.926959218 | 0.04213765 | 0.888512089 | 1 | 0.042137651 | 0.800358596 | 1.410332331 | 0.658662938 | -0.181336773 | FALSE | TRUE | FALSE |
| TATDN1 | 0.0277331 | 0.88221811 | 0.74885 | 0.244606763 | 2.214366039 | 2.214366039 | 0.04441503 | 0.888029919 | 0.00545 | 0.044415025 | 0.044415025 | 0.044415025 | 0.624408426 | -0.204531245 | FALSE | TRUE | FALSE |
| WDR81 | 0.0276577 | 0.88222654 | 0.9959 | 20.74514011 | 3866.415452 | 19447.72059 | 0.05077972 | 0.886452872 | 0.9929 | 16.62933785 | 1507.286248 | 7373.808032 | 0.544660259 | -0.263874312 | FALSE | TRUE | FALSE |
| SULT2A1 | 0.0276293 | 0.88223497 | 1 | 0.027629328 | 0.027629328 | 0.027629328 | 0.04036942 | 0.888895171 | 1 | 0.355120809 | 0.355120809 | 0.355120809 | 0.684412344 | -0.164682166 | FALSE | TRUE | FALSE |
| MSLN | 0.0275987 | 0.8822434 | 0.5117 | 0.239532413 | 0.239532413 | 4.704825573 | 0.35270413 | 0.808201674 | 0.26535 | 0.352704134 | 3.171032643 | 6.942654371 | 0.078248979 | -1.106521321 | FALSE | TRUE | FALSE |
| MET | 0.0275959 | 0.88225183 | 0.0351 | 0.027595934 | 0.027595934 | 0.927828821 | 0.65874571 | 0.686134132 | 0.2721 | 0.164730265 | 0.658745713 | 5.545960031 | 0.041891634 | -1.377872701 | FALSE | TRUE | FALSE |
| PUS7L | 0.0275886 | 0.88226026 | 0.01645 | 0.027588638 | 0.027588638 | 0.545322468 | 0.0404449 | 0.888839454 | 1 | 0.358793318 | 0.358793318 | 0.800566278 | 0.682128974 | -0.166133503 | FALSE | TRUE | FALSE |
| CCDC77 | 0.0275863 | 0.88226868 | 0.8775 | 2.11365702 | 19.22636445 | 19.22636445 | 0.38341329 | 0.798018974 | 0.5077 | 0.383413294 | 0.383413294 | 0.869319651 | 0.071949164 | -1.14297425 | FALSE | TRUE | FALSE |
| FAM217B | 0.0275855 | 0.88227711 | 1 | 0.027585496 | 0.027585496 | 0.027585496 | 0.03365646 | 0.890427244 | 1 | 0.033656458 | 0.033656458 | 0.033656458 | 0.819619682 | -0.086387621 | FALSE | TRUE | FALSE |
| EPHB1 | 0.0275619 | 0.88228553 | 0.5179 | 0.110155193 | 0.110155193 | 2.151109444 | 0.16856609 | 0.85996734 | 0.0277 | 0.168566088 | 0.168566088 | 3.301117009 | 0.163507933 | -0.786461172 | FALSE | TRUE | FALSE |
| SDCCAG8 | 0.0275361 | 0.88229396 | 0.0157 | 0.027536094 | 0.027536094 | 0.544049495 | 0.04146143 | 0.888608078 | 1 | 0.041461427 | 0.041461427 | 0.820376003 | 0.664137647 | -0.177741901 | FALSE | TRUE | FALSE |
| CDH15 | 0.0274388 | 0.88230238 | 0.76185 | 0.061088868 | 0.244180521 | 1.196813716 | 0.94931189 | 0.59296294 | 0.507 | 0.949311889 | 35.75165107 | 35.75165107 | 0.028903848 | -1.539044334 | FALSE | TRUE | FALSE |
| SMPDL3B | 0.0274096 | 0.8823108 | 0.68965 | 0.027409624 | 0.43802953 | 0.43802953 | 157.98255 | 0.017038485 | 0.12425 | 7.352544171 | 631.7174906 | 2526.650458 | 0.000173498 | -3.760706037 | FALSE | FALSE | FALSE |
| ADAT1 | 0.0272717 | 0.88231922 | 0.50525 | 0.027271699 | 0.027271699 | 0.060926344 | 0.00621638 | 0.897819429 | 1 | 0.006216375 | 0.006216375 | 0.123310184 | 4.387074023 | 0.642174962 | FALSE | TRUE | FALSE |
| LRRFIP1 | 0.0272205 | 0.88232764 | 0.87635 | 2.19675885 | 20.50707551 | 43.25585174 | 0.04510566 | 0.887835999 | 0.01725 | 0.045105656 | 0.045105656 | 0.889817644 | 0.603482754 | -0.219335136 | FALSE | TRUE | FALSE |
| ERP27 | 0.0272094 | 0.88233606 | 0.0063 | 0.027209391 | 0.027209391 | 0.027209391 | 24.2831064 | 0.113053263 | 0.1249 | 0.265556429 | 24.28310639 | 24.28310639 | 0.001120507 | -2.950585416 | FALSE | FALSE | FALSE |
| LGALS12 | 0.0271752 | 0.88234448 | 1 | 0.027175243 | 0.246922111 | 0.246922111 | 0.29312932 | 0.827136889 | 0.26045 | 0.293129316 | 2.74505656 | 2.74505656 | 0.092707352 | -1.032885823 | FALSE | TRUE | FALSE |
| CGA | 0.0271748 | 0.88235289 | 1 | 0.027174835 | 0.027174835 | 0.027174835 | 0.34644308 | 0.810069159 | 0.49975 | 0.036509181 | 0.346443078 | 0.346443078 | 0.078439537 | -1.10546498 | FALSE | TRUE | FALSE |
| PLXNA4 | 0.0271326 | 0.88236131 | 1 | 0.027132598 | 0.518417574 | 0.91022713 | 0.16375732 | 0.861189078 | 0.5274 | 0.16375732 | 0.786670861 | 3.145759874 | 0.165687846 | -0.780709348 | FALSE | TRUE | FALSE |
| MCTP1 | 0.0270332 | 0.88236972 | 0.0233 | 0.027033202 | 0.027033202 | 0.531748877 | 0.04076996 | 0.888775714 | 1 | 0.163013444 | 0.163013444 | 3.212874413 | 0.66306663 | -0.178442828 | FALSE | TRUE | FALSE |
| EFCC1 | 0.0269864 | 0.88237814 | 1 | 0.247377367 | 2.327400612 | 2.327400612 | 0.03271976 | 0.890628644 | 1 | 0.301424843 | 0.301424843 | 0.301424843 | 0.824774165 | -0.083664951 | FALSE | TRUE | FALSE |
| DZIP1 | 0.0269554 | 0.88238655 | 0.02325 | 0.026955426 | 0.026955426 | 0.529537409 | 0.0395147 | 0.889062024 | 1 | 0.347805051 | 0.347805051 | 6.84721042 | 0.682161902 | -0.166112539 | FALSE | TRUE | FALSE |
| AKNA | 0.0269009 | 0.88239496 | 0.52225 | 0.238947942 | 0.238947942 | 4.638952029 | 0.03900269 | 0.889180928 | 1 | 0.349071961 | 0.349071961 | 6.800845129 | 0.68971882 | -0.161327923 | FALSE | TRUE | FALSE |
| SLC47A1 | 0.0269003 | 0.88240337 | 1 | 0.026900298 | 0.026900298 | 0.531905554 | 0.04075364 | 0.888783685 | 0.0133 | 0.040753642 | 0.040753642 | 0.806930084 | 0.660071019 | -0.180409335 | FALSE | TRUE | FALSE |
| PEX12 | 0.0268529 | 0.88241178 | 1 | 0.026852867 | 0.238741817 | 0.238741817 | 1.44470598 | 0.54141518 | 0.2577 | 0.361295787 | 1.444705981 | 1.444705981 | 0.018587081 | -1.730788817 | FALSE | FALSE | FALSE |
| POLR1E | 0.0268278 | 0.88242019 | 1 | 0.026827848 | 0.026827848 | 0.531871342 | 0.03176031 | 0.890821676 | 0.0109 | 0.03176031 | 0.03176031 | 0.630320423 | 0.844697305 | -0.073298891 | FALSE | TRUE | FALSE |
| SEC24C | 0.0267649 | 0.88242859 | 0.7617 | 0.250273322 | 2.401881757 | 8.440960631 | 0.63482065 | 0.697506286 | 0.8221 | 3.577660359 | 57.17591028 | 228.6561855 | 0.042161341 | -1.375085583 | FALSE | TRUE | FALSE |
| PKP2 | 0.0267303 | 0.882437 | 0.50985 | 0.05841217 | 0.23353031 | 1.145117921 | 0.04903414 | 0.886931031 | 0.5093 | 0.04903414 | 0.04903414 | 0.9635617 | 0.545136099 | -0.263495058 | FALSE | TRUE | FALSE |
| MANEAL | 0.026706 | 0.8824454 | 1 | 0.026706027 | 0.026706027 | 0.529015484 | 0.03965964 | 0.889014398 | 1 | 0.365445248 | 0.365445248 | 0.786514691 | 0.673380511 | -0.171739457 | FALSE | TRUE | FALSE |
| CDC20B | 0.0266728 | 0.88245381 | 1 | 0.232266888 | 2.075969618 | 2.075969618 | 0.04269794 | 0.888383878 | 0.0124 | 0.042697938 | 0.042697938 | 0.846149548 | 0.624686823 | -0.204337655 | FALSE | TRUE | FALSE |
| DHX35 | 0.0266598 | 0.88246221 | 1 | 0.23507802 | 2.127541218 | 4.630526793 | 0.04043698 | 0.888855379 | 0.7559 | 0.359554964 | 3.281382316 | 7.095331939 | 0.659293094 | -0.180921474 | FALSE | TRUE | FALSE |
| TSEN2 | 0.0266399 | 0.88247061 | 1 | 2.133714162 | 189.1704523 | 189.1704523 | 122.032955 | 0.023632257 | 0.06765 | 13.01553604 | 293.2861096 | 1172.841857 | 0.000218301 | -3.660944301 | FALSE | FALSE | FALSE |
| LMAN1 | 0.0266344 | 0.88247901 | 0.0157 | 0.026634409 | 0.026634409 | 0.526963289 | 0.03939983 | 0.889093754 | 1 | 0.355484587 | 0.355484587 | 0.780535739 | 0.676003116 | -0.170051302 | FALSE | TRUE | FALSE |
| JARID2 | 0.0266326 | 0.88248741 | 1 | 0.02663263 | 0.02663263 | 0.897214315 | 0.04089507 | 0.888743819 | 0.0358 | 0.040895072 | 0.040895072 | 0.795802177 | 0.651243018 | -0.18625692 | FALSE | TRUE | FALSE |
| ACSS2 | 0.0266102 | 0.88249581 | 0.50885 | 0.23820474 | 0.23820474 | 0.89991558 | 1.44485212 | 0.541280016 | 0.25715 | 0.361336091 | 1.444852119 | 3.169773646 | 0.018417222 | -1.734775868 | FALSE | FALSE | FALSE |
| TMC6 | 0.026608 | 0.8825042 | 0.513 | 0.239391093 | 0.239391093 | 4.703171097 | 0.04183926 | 0.888560101 | 1 | 0.04183926 | 0.04183926 | 0.823740262 | 0.635957804 | -0.196571699 | FALSE | TRUE | FALSE |
| RAD9B | 0.026587 | 0.8825126 | 0.0088 | 0.026587036 | 0.026587036 | 0.026587036 | 0.29283589 | 0.827173704 | 0.50395 | 0.292835895 | 0.292835895 | 0.292835895 | 0.090791589 | -1.041954382 | FALSE | TRUE | FALSE |
| HERC1 | 0.0265538 | 0.88252099 | 0.988 | 89.94395823 | 94298.90603 | 1729673.17 | 0.31107016 | 0.821706364 | 0.98915 | 2712.703554 | 3124846.575 | 49951295.23 | 0.085362727 | -1.06873172 | FALSE | TRUE | FALSE |
| APLP1 | 0.026452 | 0.88252939 | 1 | 0.979014867 | 9.304183556 | 19.25826271 | 0.04369583 | 0.888166913 | 0.0183 | 0.043695827 | 0.043695827 | 0.861245166 | 0.605365852 | -0.217982081 | FALSE | TRUE | FALSE |
| PTPRE | 0.0264514 | 0.88253778 | 1 | 0.026451381 | 0.026451381 | 0.520115894 | 0.15990643 | 0.862095187 | 0.50495 | 0.159906431 | 0.159906431 | 3.150631172 | 0.165417866 | -0.781417585 | FALSE | TRUE | FALSE |
| FAM174A | 0.026445 | 0.88254617 | 1 | 0.026444953 | 0.026444953 | 0.026444953 | 0.02825889 | 0.891679012 | 1 | 0.02825889 | 0.02825889 | 0.02825889 | 0.935810029 | -0.028812305 | FALSE | TRUE | FALSE |
| FASTKD5 | 0.0264344 | 0.88255456 | 0.88085 | 2.184716291 | 20.63854956 | 20.63854956 | 0.03831977 | 0.889378579 | 1 | 0.038319773 | 0.038319773 | 0.757155853 | 0.689837166 | -0.161253411 | FALSE | TRUE | FALSE |
| ANKRD50 | 0.0263963 | 0.88256295 | 1 | 0.239158734 | 2.223977296 | 8.03393193 | 0.37528013 | 0.800734323 | 0.51825 | 0.375280132 | 1.500622051 | 12.62476622 | 0.070337606 | -1.152812418 | FALSE | TRUE | FALSE |
| NADSYN1 | 0.0263815 | 0.88257133 | 0.14475 | 0.026381549 | 0.105482217 | 0.518573985 | 0.00358121 | 0.898471537 | 0.02115 | 0.003581207 | 0.003581207 | 0.070540171 | 7.366662998 | 0.867270803 | FALSE | TRUE | FALSE |
| NUBP1 | 0.0263613 | 0.88257972 | 1 | 0.23186506 | 0.927221943 | 0.927221943 | 0.34812864 | 0.809622518 | 0.7533 | 0.34812864 | 3.167304747 | 3.167304747 | 0.075722769 | -1.120773516 | FALSE | TRUE | FALSE |
| CSF3 | 0.0263597 | 0.88258811 | 0.4994 | 0.026359728 | 0.248480099 | 0.248480099 | 0.02851165 | 0.891595349 | 1 | 0.028511646 | 0.028511646 | 0.028511646 | 0.924524933 | -0.034081372 | FALSE | TRUE | FALSE |
| THADA | 0.0263588 | 0.88259649 | 0.8952 | 0.223691058 | 17.34538126 | 158.3036689 | 0.00892474 | 0.897048996 | 0.0406 | 0.008924739 | 0.008924739 | 0.173434674 | 2.953451602 | 0.470329859 | FALSE | TRUE | FALSE |
| SLC25A24 | 0.0263519 | 0.88260487 | 1 | 0.237388091 | 0.237388091 | 0.52225682 | 0.04149373 | 0.888600084 | 1 | 0.041493726 | 0.041493726 | 0.823242788 | 0.635082042 | -0.197170168 | FALSE | TRUE | FALSE |
| FCAR | 0.0263276 | 0.88261325 | 0.00855 | 0.026327625 | 0.026327625 | 0.026327625 | 0.03317114 | 0.890551262 | 0.00715 | 0.03317114 | 0.03317114 | 0.03317114 | 0.793690691 | -0.100348713 | FALSE | TRUE | FALSE |
| TMEM132E | 0.026277 | 0.88262164 | 0.0341 | 0.026276979 | 0.026276979 | 0.511094447 | 0.03399976 | 0.890365141 | 0.03115 | 0.033999755 | 0.033999755 | 0.663495647 | 0.772857882 | -0.11190036 | FALSE | TRUE | FALSE |
| DNAJC2 | 0.0262646 | 0.88263002 | 1 | 5.4025E+12 | 1.24263E+17 | 2.10261E+19 | 0.03240664 | 0.890713651 | 1 | 7.77897E+16 | 2.48048E+21 | 4.90517E+23 | 0.810468218 | -0.091264011 | FALSE | TRUE | FALSE |
| PTPN22 | 0.0262341 | 0.88263839 | 0.74875 | 0.056399266 | 0.225510776 | 1.113353483 | 0.01211771 | 0.896035698 | 0.01525 | 0.012117713 | 0.012117713 | 0.239583908 | 2.164938434 | 0.335445551 | FALSE | TRUE | FALSE |
| GAL3ST4 | 0.0261943 | 0.88264677 | 1 | 2.11114553 | 19.69482992 | 19.69482992 | 0.0375811 | 0.889551985 | 1 | 28.77634911 | 276.854512 | 2728.700821 | 0.697007228 | -0.156762718 | FALSE | TRUE | FALSE |
| CNNM4 | 0.0261528 | 0.88265515 | 1 | 0.240184707 | 0.240184707 | 4.71212426 | 0.0414082 | 0.888632053 | 0.0213 | 0.041408204 | 0.041408204 | 0.814242105 | 0.631585205 | -0.199568052 | FALSE | TRUE | FALSE |
| ZNF600 | 0.0260888 | 0.88266352 | 0.6959 | 0.222203882 | 17.38647075 | 17.38647075 | 3.86448045 | 0.363555568 | 0.31205 | 0.43583949 | 35.12528053 | 35.12528053 | 0.006750924 | -2.170636812 | FALSE | FALSE | FALSE |
| RTTN | 0.0260775 | 0.8826719 | 0.5158 | 0.026077518 | 0.224624044 | 0.871446946 | 0.00100006 | 0.899442128 | 1 | 0.001000061 | 0.019333942 | 0.033488505 | 26.07593899 | 1.416239956 | FALSE | TRUE | FALSE |
| DHRS9 | 0.0260376 | 0.88268027 | 1 | 0.239995737 | 0.239995737 | 0.239995737 | 0.03879597 | 0.889275882 | 0.75565 | 0.358506235 | 3.400152835 | 3.400152835 | 0.671141707 | -0.173185772 | FALSE | TRUE | FALSE |
| GCK | 0.0260314 | 0.88268864 | 1 | 0.026031395 | 0.026031395 | 0.513074387 | 3.41158917 | 0.388968086 | 0.25885 | 0.353806873 | 3.411589175 | 6.98565023 | 0.007630284 | -2.117459279 | FALSE | FALSE | FALSE |
| SOWAHB | 0.0259759 | 0.88269701 | 1 | 0.025975926 | 0.025975926 | 0.511510544 | 0.03932263 | 0.889109613 | 0.0184 | 0.039322626 | 0.039322626 | 0.77576664 | 0.660584716 | -0.180071479 | FALSE | TRUE | FALSE |
| HELLS | 0.0259728 | 0.88270538 | 1 | 0.10380954 | 0.415014906 | 2.045980966 | 0.04088896 | 0.888751794 | 0.01825 | 0.040888961 | 0.040888961 | 0.807291333 | 0.635204138 | -0.197086682 | FALSE | TRUE | FALSE |
| CEP72 | 0.0259667 | 0.88271375 | 0.51125 | 0.225241808 | 0.225241808 | 0.878718931 | 0.04043991 | 0.888847417 | 0.0173 | 0.040439906 | 0.040439906 | 0.798314681 | 0.64210472 | -0.192394138 | FALSE | TRUE | FALSE |
| HOOK2 | 0.0259591 | 0.88272212 | 1 | 0.025959125 | 0.025959125 | 0.510042691 | 0.03644311 | 0.889795499 | 0.02015 | 0.036443109 | 0.036443109 | 0.717548778 | 0.712319162 | -0.147325373 | FALSE | TRUE | FALSE |
| CEP128 | 0.0259487 | 0.88273048 | 0.26835 | 0.025948682 | 0.220807702 | 0.872590322 | 0.05015954 | 0.88656037 | 0.2711 | 0.050159541 | 0.433236329 | 1.688964842 | 0.517322964 | -0.286238243 | FALSE | TRUE | FALSE |
| PTPRN2 | 0.0259252 | 0.88273885 | 0.98425 | 19.95521983 | 769.0809719 | 7593.458741 | 0.04251984 | 0.888440002 | 1 | 3.54210226 | 33.59395449 | 69.36785163 | 0.609720438 | -0.214869247 | FALSE | TRUE | FALSE |
| LTN1 | 0.0258342 | 0.88274721 | 0.7597 | 0.229226826 | 0.91681882 | 17.8040189 | 0.04105014 | 0.888679978 | 1 | 0.041050143 | 0.041050143 | 1.378855149 | 0.629333167 | -0.201119379 | FALSE | TRUE | FALSE |
| TECPR2 | 0.0258135 | 0.88275558 | 0.76235 | 0.233945417 | 2.176120883 | 42.12509813 | 0.04206305 | 0.888528096 | 1 | 0.042063055 | 0.042063055 | 0.817404782 | 0.613685866 | -0.212053879 | FALSE | TRUE | FALSE |
| NUP210L | 0.0257739 | 0.88276394 | 1 | 0.227535869 | 2.061726487 | 39.86201771 | 0.04043123 | 0.888871299 | 0.52165 | 0.36019616 | 0.36019616 | 6.992098898 | 0.637474051 | -0.195537489 | FALSE | TRUE | FALSE |
| GJC3 | 0.0257664 | 0.8827723 | 0.0076 | 0.025766381 | 0.025766381 | 0.025766381 | 0.01173881 | 0.896204691 | 0.00695 | 0.01173881 | 0.01173881 | 0.01173881 | 2.194973867 | 0.341429354 | FALSE | TRUE | FALSE |
| TTC31 | 0.0257472 | 0.88278066 | 0.50865 | 0.226070221 | 0.226070221 | 0.87125974 | 27.5880424 | 0.101081463 | 0.1305 | 2.963431358 | 27.58804237 | 27.58804237 | 0.000933273 | -3.02999119 | FALSE | FALSE | FALSE |
| MPHOSPH9 | 0.0257298 | 0.88278902 | 1 | 0.227252928 | 0.227252928 | 4.459465031 | 0.040391 | 0.888887215 | 1 | 0.040390997 | 0.040390997 | 0.794404313 | 0.637017757 | -0.195848462 | FALSE | TRUE | FALSE |
| CCDC38 | 0.0257266 | 0.88279738 | 0.4984 | 0.025726555 | 0.22375476 | 0.509752937 | 2.99446025 | 0.412205803 | 0.2536 | 0.332438092 | 2.994460246 | 2.994460246 | 0.008591383 | -2.065936922 | FALSE | FALSE | FALSE |

| | | | | | | | | | | | | | | | | | |
|---|---|---|---|---|---|---|---|---|---|---|---|---|---|---|---|---|---|
| CSAD | 0.0257197 | 0.88280573 | 1 | 0.025719703 | 0.025719703 | 0.50836002 | 0.15486763 | 0.86327046 | 0.5068 | 0.154867629 | 0.154867629 | 0.766497657 | 0.166075398 | -0.779694697 | FALSE | TRUE | FALSE |
| DBNDD2 | 0.0256763 | 0.88281409 | 1 | 0.025676335 | 0.025676335 | 0.025676335 | 0.02792328 | 0.891754966 | 0.00455 | 0.027923281 | 0.027923281 | 0.027923281 | 0.919531457 | -0.036433409 | FALSE | TRUE | FALSE |
| OGDH | 0.025664 | 0.88282244 | 1 | 0.025663976 | 0.025663976 | 0.500272561 | 0.03909698 | 0.889165087 | 0.5127 | 0.156294418 | 0.156294418 | 3.05597971 | 0.656418452 | -0.18281922 | FALSE | TRUE | FALSE |
| GPAM | 0.0255972 | 0.88283079 | 1 | 0.025597222 | 0.025597222 | 0.504344791 | 0.03880335 | 0.889267975 | 1 | 0.155155331 | 0.155155331 | 1.310693941 | 0.659665209 | -0.18067642 | FALSE | TRUE | FALSE |
| ACSS3 | 0.0255759 | 0.88283915 | 0.0199 | 0.025575915 | 0.025575915 | 0.504433429 | 0.03878634 | 0.889283787 | 1 | 0.155080497 | 0.155080497 | 1.312570689 | 0.659405234 | -0.18084761 | FALSE | TRUE | FALSE |
| ST14 | 0.0255277 | 0.8828475 | 0.0307 | 0.025527688 | 0.025527688 | 0.498695302 | 0.35359939 | 0.807860162 | 0.51835 | 0.353599387 | 1.413357016 | 6.927059382 | 0.072193814 | -1.141500016 | FALSE | TRUE | FALSE |
| OTOGL | 0.0255199 | 0.88285585 | 0.8304 | 1.452797388 | 372.9377736 | 3137.943378 | 1.5403604 | 0.531912251 | 0.5185 | 1.967076899 | 506.2318681 | 7697.356827 | 0.01656751 | -1.780742762 | FALSE | FALSE | FALSE |
| TNFAIP2 | 0.0255027 | 0.8828642 | 0.01665 | 0.025502733 | 0.025502733 | 0.50431047 | 0.0275992 | 0.891808078 | 1 | 0.258079405 | 0.258079405 | 0.546507016 | 0.924038927 | -0.034309733 | FALSE | TRUE | FALSE |
| ART4 | 0.0254921 | 0.88287254 | 1 | 0.025492129 | 0.025492129 | 0.025492129 | 0.02938305 | 0.891404771 | 1 | 0.029383051 | 0.029383051 | 0.029383051 | 0.867579362 | -0.061690788 | FALSE | TRUE | FALSE |
| ZNF665 | 0.0254601 | 0.88288089 | 0.5104 | 0.224051839 | 0.224051839 | 0.860787931 | 0.039965 | 0.888966733 | 1 | 0.039965002 | 0.039965002 | 0.79088433 | 0.637061046 | -0.19581895 | FALSE | TRUE | FALSE |
| KEAP1 | 0.0254259 | 0.88288923 | 1 | 1.80356E+22 | 4.91645E+28 | 1.18306E+31 | 0.04429031 | 0.888054116 | 1 | 2.27028E+20 | 1.45453E+26 | 3.42354E+28 | 0.574074701 | -0.241031592 | FALSE | TRUE | FALSE |
| ADM2 | 0.0254056 | 0.88289758 | 1 | 0.025405564 | 0.025405564 | 0.025405564 | 0.02744655 | 0.891861144 | 0.0037 | 0.027446548 | 0.027446548 | 0.027446548 | 0.925637863 | -0.033558889 | FALSE | TRUE | FALSE |
| ZDHHC8 | 0.0254026 | 0.88290592 | 0.0273 | 0.025402615 | 0.025402615 | 0.496358813 | 0.03921523 | 0.889141319 | 0.0268 | 0.039215234 | 0.039215234 | 0.76837766 | 0.647774158 | -0.188576381 | FALSE | TRUE | FALSE |
| TRPC4 | 0.0253914 | 0.88291426 | 0.5144 | 0.234787562 | 0.234787562 | 4.611876404 | 0.04039574 | 0.888879257 | 1 | 0.040395738 | 0.040395738 | 0.795190375 | 0.62856518 | -0.201649681 | FALSE | TRUE | FALSE |
| SLC12A8 | 0.0253797 | 0.8829226 | 0.9794 | 17.56705173 | 4485.757261 | 41731.50563 | 0.48305656 | 0.761847652 | 0.35735 | 0.213660058 | 13.64468069 | 115.5205247 | 0.052539752 | -1.279511985 | FALSE | TRUE | FALSE |
| CCDC51 | 0.0253692 | 0.88293094 | 0.01235 | 0.025369218 | 0.025369218 | 0.50294759 | 0.03467297 | 0.890186239 | 1 | 0.034672968 | 0.034672968 | 0.68811841 | 0.731671369 | -0.135683939 | FALSE | TRUE | FALSE |
| CHRNA5 | 0.0253534 | 0.88293928 | 0.81465 | 0.101326312 | 3.622344631 | 3.622344631 | 0.20473688 | 0.850841261 | 0.5089 | 0.204736883 | 3.273241593 | 3.273241593 | 0.123834167 | -0.907159513 | FALSE | TRUE | FALSE |
| C10orf90 | 0.0253381 | 0.88294762 | 1 | 0.21936306 | 1.949263148 | 4.320559715 | 0.04070877 | 0.888791655 | 0.01775 | 0.040708775 | 0.040708775 | 0.803264473 | 0.622423733 | -0.205913856 | FALSE | TRUE | FALSE |
| MTMR12 | 0.0253239 | 0.88295596 | 1 | 0.101210075 | 0.101210075 | 1.995472028 | 0.15633818 | 0.862884721 | 0.01825 | 0.156338184 | 0.156338184 | 1.322260873 | 0.161981496 | -0.790534595 | FALSE | TRUE | FALSE |
| GALNTL5 | 0.0253055 | 0.88296429 | 1 | 0.025305515 | 0.025305515 | 0.502015812 | 0.03763122 | 0.889536241 | 1 | 0.338448999 | 0.338448999 | 0.747261836 | 0.672460726 | -0.172333075 | FALSE | TRUE | FALSE |
| SLC26A8 | 0.0252353 | 0.88297262 | 0.2678 | 0.025235335 | 0.100835854 | 0.495421016 | 6.6198145 | 0.252545843 | 0.47885 | 3.913492082 | 249.9531898 | 2112.169715 | 0.003812091 | -2.418836738 | FALSE | FALSE | FALSE |
| CPNE3 | 0.0252305 | 0.88298096 | 1 | 0.225288864 | 2.064712964 | 2.064712964 | 0.04104117 | 0.888687962 | 1 | 0.041041169 | 0.041041169 | 0.814123667 | 0.614761413 | -0.2112934 | FALSE | TRUE | FALSE |
| CNDP1 | 0.0252204 | 0.88298929 | 1 | 0.22302831 | 0.891860957 | 1.993336511 | 1.35859282 | 0.550050804 | 0.37975 | 0.60959983 | 5.434353774 | 5.434353774 | 0.018563622 | -1.731337288 | FALSE | FALSE | FALSE |
| PPARA | 0.0252124 | 0.88299762 | 1 | 0.025212437 | 0.025212437 | 0.498392517 | 0.03854091 | 0.889331202 | 0.50245 | 0.15403545 | 0.15403545 | 0.762930587 | 0.65417342 | -0.184307106 | FALSE | TRUE | FALSE |
| IPO4 | 0.0251647 | 0.88300595 | 1 | 0.025164721 | 0.025164721 | 0.490762824 | 0.32822519 | 0.815897025 | 0.51165 | 0.328225194 | 0.328225194 | 6.420259586 | 0.076669072 | -1.115379794 | FALSE | TRUE | FALSE |
| PIGQ | 0.0251616 | 0.88301428 | 1 | 0.219892878 | 0.219892878 | 4.330516551 | 2.87554958 | 0.420649539 | 0.1365 | 2.875549584 | 26.6582982 | 26.6582982 | 0.008750173 | -2.057983373 | FALSE | FALSE | FALSE |
| UMOD | 0.0251557 | 0.8830226 | 0.26055 | 0.02515572 | 0.100527898 | 0.386221231 | 0.05068733 | 0.886477695 | 0.5107 | 0.050687334 | 0.050687334 | 0.193201396 | 0.496292041 | -0.30426269 | FALSE | TRUE | FALSE |
| GAPDHS | 0.0251494 | 0.88303093 | 1 | 0.22461071 | 2.058925151 | 4.442442567 | 0.03972159 | 0.88899057 | 1 | 0.357547158 | 0.357547158 | 0.786677031 | 0.633140679 | -0.198499783 | FALSE | TRUE | FALSE |
| SLC37A3 | 0.0251 | 0.88303926 | 1 | 0.025100019 | 0.025100019 | 0.497062963 | 0.03682997 | 0.8897092 | 0.7529 | 0.327680908 | 1.310391326 | 1.310391326 | 0.681510657 | -0.166527348 | FALSE | TRUE | FALSE |
| DGKD | 0.0250633 | 0.88304758 | 0.03705 | 0.025063284 | 0.025063284 | 0.487163409 | 0.09937592 | 0.876120834 | 0.75895 | 0.397156422 | 6.348163745 | 13.38923605 | 0.252206817 | -0.59824318 | FALSE | TRUE | FALSE |
| USP34 | 0.0250211 | 0.8830559 | 0.08345 | 0.025021104 | 0.471187587 | 0.826198233 | 1.19851851 | 0.567679235 | 0.30185 | 0.329527911 | 6.249403317 | 58.4341961 | 0.020876694 | -1.680338275 | FALSE | FALSE | FALSE |
| FAM184A | 0.025018 | 0.88306422 | 0.02765 | 0.025017977 | 0.025017977 | 0.842347555 | 2.91045295 | 0.418561401 | 0.26175 | 0.319672026 | 2.91045295 | 10.77583573 | 0.008595905 | -2.065708387 | FALSE | FALSE | FALSE |
| AMIGO3 | 0.0250122 | 0.88307254 | 1 | 0.100024575 | 0.100024575 | 1.965164079 | 0.33393888 | 0.814378733 | 0.01615 | 0.333938881 | 0.333938881 | 0.739154331 | 0.074900548 | -1.125515006 | FALSE | TRUE | FALSE |
| PREPL | 0.0249792 | 0.88308086 | 0.0195 | 0.024979188 | 0.024979188 | 0.492698616 | 0.32430347 | 0.817390259 | 0.50865 | 0.324303471 | 0.324303471 | 1.249585377 | 0.077024115 | -1.113373281 | FALSE | TRUE | FALSE |
| SLC22A11 | 0.0249781 | 0.88308918 | 1 | 0.024978092 | 0.024978092 | 0.493131862 | 0.03833317 | 0.889370685 | 0.01485 | 0.038333172 | 0.038333172 | 0.757967643 | 0.651605161 | -0.186015485 | FALSE | TRUE | FALSE |
| BOC | 0.0249499 | 0.8830975 | 0.52325 | 0.223222828 | 0.223222828 | 4.34400051 | 3.08999951 | 0.406379247 | 0.0234 | 0.334523969 | 3.089999506 | 11.29333219 | 0.008074389 | -2.092890323 | FALSE | FALSE | FALSE |
| CEP95 | 0.0249464 | 0.88310581 | 0.8771 | 0.212928884 | 1.864257148 | 4.200260371 | 0.00608081 | 0.89784675 | 1 | 0.051394463 | 0.051394463 | 0.213586636 | 4.102486681 | 0.61304718 | FALSE | TRUE | FALSE |
| STAC2 | 0.0249247 | 0.88311413 | 0.5001 | 0.227076206 | 0.227076206 | 0.493384975 | 0.03961842 | 0.889038216 | 0.01305 | 0.039618423 | 0.039618423 | 0.785214128 | 0.629119193 | -0.201267066 | FALSE | TRUE | FALSE |
| TAS2R43 | 0.0249112 | 0.88312244 | 0.00675 | 0.024911178 | 0.024911178 | 0.024911178 | 0.02543403 | 0.892336668 | 0.00675 | 0.025434025 | 0.025434025 | 0.025434025 | 0.979442992 | -0.009020837 | FALSE | TRUE | FALSE |
| GLT8D2 | 0.0249092 | 0.88313076 | 1 | 0.214450539 | 0.214450539 | 0.494792393 | 0.32859437 | 0.815729556 | 0.00745 | 0.32859437 | 0.32859437 | 0.32859437 | 0.075805235 | -1.120300802 | FALSE | TRUE | FALSE |
| DOCK4 | 0.0248674 | 0.88313907 | 1 | 0.223184881 | 0.478445718 | 4.294056597 | 0.33457959 | 0.814101934 | 0.76145 | 0.334579593 | 3.09914431 | 59.90076128 | 0.074324198 | -1.12886977 | FALSE | TRUE | FALSE |
| CDR2 | 0.0248618 | 0.88314738 | 1 | 0.024861774 | 0.024861774 | 0.492624209 | 0.0297373 | 0.891320725 | 0.01145 | 0.029737303 | 0.029737303 | 0.589887736 | 0.836046714 | -0.077769456 | FALSE | TRUE | FALSE |
| STXBP2 | 0.024834 | 0.88315569 | 0.2627 | 0.024834023 | 0.099264062 | 0.4898029 | 0.01090148 | 0.896506188 | 1 | 0.696395231 | 2.785241729 | 11.14239315 | 2.278040589 | 0.357561458 | FALSE | TRUE | FALSE |
| RC3H1 | 0.0248182 | 0.88316399 | 0.0321 | 0.024818202 | 0.024818202 | 0.834093283 | 0.14702859 | 0.865293057 | 0.7562 | 0.338867178 | 1.354769808 | 6.698298528 | 0.168798474 | -0.772631483 | FALSE | TRUE | FALSE |
| RELT | 0.0247814 | 0.8831723 | 0.0163 | 0.024781372 | 0.024781372 | 0.490184378 | 0.038085 | 0.889418033 | 0.01225 | 0.038085003 | 0.038085003 | 0.754330252 | 0.650685828 | -0.186628652 | FALSE | TRUE | FALSE |
| IFFO1 | 0.0247812 | 0.88318061 | 1 | 0.024781242 | 0.024781242 | 0.487708821 | 0.0381847 | 0.889410144 | 0.0192 | 0.038184701 | 0.038184701 | 0.752941286 | 0.648983535 | -0.187766321 | FALSE | TRUE | FALSE |
| RAD51C | 0.0247554 | 0.88318891 | 1 | 0.024755382 | 0.024755382 | 0.491373153 | 0.03777898 | 0.889504742 | 0.01 | 0.037778982 | 0.037778982 | 0.044906798 | 0.655268649 | -0.18358061 | FALSE | TRUE | FALSE |
| EIF2B1 | 0.0247291 | 0.88319722 | 0.0078 | 0.024729056 | 0.024729056 | 0.024729056 | 0.0377792 | 0.889496864 | 0.5009 | 0.151013907 | 0.151013907 | 0.151013907 | 0.654568083 | -0.184045175 | FALSE | TRUE | FALSE |
| BLM | 0.0246783 | 0.88320552 | 0.88075 | 0.216361189 | 1.945720646 | 4.223860137 | 0.19832661 | 0.852530525 | 0.2601 | 0.198326606 | 1.76084764 | 6.686159779 | 0.124432485 | -0.905066227 | FALSE | TRUE | FALSE |
| RPAP1 | 0.0246282 | 0.88321382 | 0.5182 | 0.024628191 | 0.024628191 | 0.479914189 | 0.00541997 | 0.898017204 | 1 | 0.047502021 | 0.427303861 | 1.600132238 | 4.543970773 | 0.657435529 | FALSE | TRUE | FALSE |
| ZNF311 | 0.0246007 | 0.88322212 | 1 | 1.908103224 | 17.46299245 | 17.46299245 | 0.34965699 | 0.80919626 | 0.5067 | 0.349656992 | 0.349656992 | 1.344673591 | 0.070356539 | -1.152695532 | FALSE | TRUE | FALSE |
| KIAA1549L | 0.0245644 | 0.88323042 | 1 | 0.024564415 | 0.472893348 | 0.824743396 | 0.03775707 | 0.889512618 | 1 | 0.03775707 | 0.03775707 | 1.26959105 | 0.650591131 | -0.186691862 | FALSE | TRUE | FALSE |
| FUCA2 | 0.0245211 | 0.88323872 | 1 | 0.024521092 | 0.024521092 | 0.486290403 | 0.03771171 | 0.889528368 | 1 | 0.037711715 | 0.037711715 | 0.748638155 | 0.650224793 | -0.186936475 | FALSE | TRUE | FALSE |
| ZNF33A | 0.0244924 | 0.88324702 | 1 | 0.21431475 | 0.21431475 | 0.827864108 | 0.03854344 | 0.889323302 | 0.01645 | 0.038543442 | 0.038543442 | 0.762310066 | 0.63544894 | -0.196919341 | FALSE | TRUE | FALSE |
| PCDH1 | 0.0244701 | 0.88325531 | 0.2762 | 0.024470085 | 0.20998329 | 0.825013084 | 0.04923477 | 0.886848858 | 0.5185 | 0.049234771 | 0.049234771 | 0.959444163 | 0.49700821 | -0.303636437 | FALSE | TRUE | FALSE |
| CPSF1 | 0.0244645 | 0.88326361 | 1 | 0.02446445 | 0.02446445 | 0.823515275 | 0.3080346 | 0.822758072 | 0.88175 | 2.833077811 | 26.72602138 | 54.81875337 | 0.079421111 | -1.100064041 | FALSE | TRUE | FALSE |
| DOCK8 | 0.0244451 | 0.8832719 | 0.0577 | 0.024445118 | 0.11711511 | 0.468346461 | 0.01097765 | 0.896478212 | 1 | 0.04390119 | 0.211389791 | 0.845377689 | 2.226807226 | 0.347682622 | FALSE | TRUE | FALSE |
| CCDC13 | 0.0244184 | 0.88328019 | 1 | 1.883514957 | 17.19038136 | 37.09239287 | 0.33571172 | 0.813652999 | 0.2696 | 0.335711724 | 3.020271334 | 6.623438333 | 0.072736116 | -1.138249896 | FALSE | TRUE | FALSE |
| SRMS | 0.0244144 | 0.88328848 | 1 | 0.221974386 | 0.221974386 | 0.827378198 | 0.03888565 | 0.889220511 | 1 | 0.038885647 | 0.038885647 | 0.768263224 | 0.627850855 | -0.20214351 | FALSE | TRUE | FALSE |
| NR1D1 | 0.0244079 | 0.88329678 | 1 | 0.097535915 | 0.097535915 | 0.82598776 | 2.36722315 | 0.458586575 | 0.13505 | 0.592026886 | 2.367223148 | 5.014631571 | 0.010310785 | -1.986708286 | FALSE | FALSE | FALSE |
| TMEM106C | 0.0243619 | 0.88330506 | 0.0073 | 0.024361936 | 0.024361936 | 0.024361936 | 0.0281379 | 0.891717001 | 1 | 0.0281379 | 0.0281379 | 0.0281379 | 0.86580503 | -0.062579895 | FALSE | TRUE | FALSE |
| GYPC | 0.0243437 | 0.88331335 | 0.003 | 0.024343744 | 0.024343744 | 0.024343744 | 0.0260537 | 0.892133336 | 1 | 0.026053696 | 0.026053696 | 0.026053696 | 0.934368171 | -0.029481964 | FALSE | TRUE | FALSE |
| TRAPPC9 | 0.0243407 | 0.88332164 | 0.0388 | 0.02434072 | 0.02434072 | 0.472562929 | 0.03723719 | 0.889630642 | 1 | 0.037237194 | 0.037237194 | 0.725493248 | 0.653666878 | -0.184643521 | FALSE | TRUE | FALSE |
| PTPRU | 0.0243342 | 0.88332993 | 0.02495 | 0.024334191 | 0.024334191 | 0.467686779 | 0.04332295 | 0.888239321 | 0.5259 | 0.173209164 | 0.69267936 | 3.344648914 | 0.561692851 | -0.250501103 | FALSE | TRUE | FALSE |
| GDAP1 | 0.0243116 | 0.88333821 | 0.75305 | 0.22330479 | 0.892536203 | 0.892536203 | 0.35104104 | 0.808813243 | 0.4978 | 0.038071923 | 0.351041037 | 0.351041037 | 0.069255647 | -1.159544809 | FALSE | TRUE | FALSE |
| ILDR1 | 0.0242444 | 0.8833465 | 0.87815 | 0.217525513 | 2.001878333 | 2.001878333 | 0.04986966 | 0.886684177 | 0.7459 | 0.049869665 | 0.452003987 | 0.986780197 | 0.48615515 | -0.313225109 | FALSE | TRUE | FALSE |
| FASN | 0.0241934 | 0.88335478 | 0.539 | 0.221781337 | 0.80851245 | 7.411636903 | 0.03855562 | 0.889307499 | 1 | 0.038555625 | 0.728427472 | 1.291129902 | 0.627494366 | -0.202390169 | FALSE | TRUE | FALSE |
| SON | 0.024156 | 0.88336306 | 1 | 0.217988914 | 2.019043479 | 38.49517045 | 0.03726962 | 0.889614918 | 1 | 0.33829019 | 3.151532977 | 60.42628533 | 0.648142797 | -0.188329301 | FALSE | TRUE | FALSE |
| SLC28A2 | 0.0240916 | 0.88337134 | 1 | 0.209183356 | 1.864257476 | 4.135246037 | 0.32260413 | 0.817882545 | 0.25685 | 0.322604133 | 2.904013345 | 2.904013345 | 0.074678412 | -1.126804926 | FALSE | TRUE | FALSE |
| TOMM5 | 0.0240871 | 0.88337962 | 1 | 0.024087119 | 0.024087119 | 0.024087119 | 0.02553024 | 0.892291545 | 1 | 0.025530236 | 0.025530236 | 0.025530236 | 0.943474179 | -0.025269981 | FALSE | TRUE | FALSE |
| ETFA | 0.0240703 | 0.8833879 | 1 | 0.024070346 | 0.024070346 | 0.024070346 | 0.0360745 | 0.889881675 | 1 | 0.036074496 | 0.32703181 | 0.32703181 | 0.667239995 | -0.17571793 | FALSE | TRUE | FALSE |
| FYCO1 | 0.0240673 | 0.88339618 | 0.9415 | 0.206425237 | 16.37915534 | 35.23445984 | 0.04881859 | 0.886980282 | 0.81825 | 0.19524556 | 6.805384634 | 15.20818188 | 0.492994998 | -0.307157487 | FALSE | TRUE | FALSE |
| RBP5 | 0.0240313 | 0.88340445 | 1 | 0.024031301 | 0.024031301 | 0.024031301 | 0.02569764 | 0.892223794 | 1 | 0.025697643 | 0.23938579 | 0.23938579 | 0.935155838 | -0.029116011 | FALSE | TRUE | FALSE |

| | | | | | | | | | | | | | | | | | |
|---|---|---|---|---|---|---|---|---|---|---|---|---|---|---|---|---|---|
| PPP1R9A | 0.0240309 | 0.88341273 | 0.75865 | 0.219811392 | 2.063609221 | 7.392139289 | 3.1232912 | 0.405069879 | 0.14815 | 3.1232912 | 30.17723413 | 61.25269023 | 0.007694096 | -2.113842385 | FALSE | FALSE | FALSE |
| MAP3K6 | 0.0240078 | 0.883421 | 0.2748 | 0.006003566 | 0.024007765 | 0.465116944 | 0.00171667 | 0.899162767 | 0.03585 | 0.001716675 | 0.001716675 | 0.0333838 | 13.98503878 | 1.145663675 | FALSE | TRUE | FALSE |
| IL4I1 | 0.0239795 | 0.88342928 | 1 | 0.217190678 | 0.217190678 | 0.47266533 | 2.86811867 | 0.421895803 | 0.3226 | 2.868118667 | 268.7561504 | 268.7561504 | 0.008360713 | -2.077756704 | FALSE | FALSE | FALSE |
| SPAG16 | 0.0239712 | 0.88343755 | 0.50175 | 0.056627916 | 1.812595391 | 1.812595391 | 0.00586016 | 0.89792863 | 0.50165 | 0.049792237 | 0.049792237 | 0.11596604 | 4.090536407 | 0.611780262 | FALSE | TRUE | FALSE |
| FLT3 | 0.0239639 | 0.88344582 | 1 | 0.023963861 | 0.023963861 | 0.470741709 | 0.03676723 | 0.889732748 | 0.5101 | 0.146981917 | 0.146981917 | 2.893476998 | 0.651772228 | -0.185904149 | FALSE | TRUE | FALSE |
| ALDH3B2 | 0.0239022 | 0.88345409 | 0.5045 | 0.023902249 | 0.023902249 | 0.053358858 | 0.04515943 | 0.887811715 | 0.5112 | 0.413367197 | 3.880984568 | 3.880984568 | 0.529285845 | -0.276309721 | FALSE | TRUE | FALSE |
| IL11RA | 0.0239007 | 0.88346236 | 1 | 0.210353918 | 0.210353918 | 0.47357211 | 0.03779538 | 0.889488986 | 1 | 0.037795381 | 0.037795381 | 0.749721613 | 0.632369806 | -0.199028875 | FALSE | TRUE | FALSE |
| NFATC2IP | 0.0238482 | 0.88347062 | 1 | 0.023848201 | 0.023848201 | 0.023848201 | 0.02807408 | 0.891739782 | 1 | 0.028074084 | 0.028074084 | 0.028074084 | 0.849473893 | -0.070849964 | FALSE | TRUE | FALSE |
| IQCH | 0.0238385 | 0.88347889 | 0.998 | 142.7242228 | 12757.32761 | 124801.5383 | 0.04915756 | 0.886881741 | 0.941 | 33.92940519 | 2993.769084 | 6238.8479 | 0.484940184 | -0.314311827 | FALSE | TRUE | FALSE |
| ESRRB | 0.0238307 | 0.88348716 | 1 | 0.023830718 | 0.023830718 | 0.47108236 | 0.03698323 | 0.889669933 | 0.01375 | 0.036983229 | 0.036983229 | 0.732100295 | 0.644365541 | -0.190867692 | FALSE | TRUE | FALSE |
| DPP7 | 0.0238115 | 0.88349542 | 0.13825 | 0.023811465 | 0.095224797 | 0.469401885 | 0.00316652 | 0.898619917 | 1 | 0.012658074 | 0.050612941 | 0.249962607 | 7.519757651 | 0.876203844 | FALSE | TRUE | FALSE |
| PTPRG | 0.0237628 | 0.88350368 | 0.96865 | 0.379903506 | 7.371332527 | 16.59058964 | 0.04563763 | 0.887665806 | 0.27845 | 0.04563763 | 0.182473472 | 1.536741509 | 0.520683334 | -0.283426323 | FALSE | TRUE | FALSE |
| IARS2 | 0.0237077 | 0.88351195 | 1 | 0.023707708 | 0.023707708 | 0.464429112 | 0.03558393 | 0.889983366 | 1 | 0.321924442 | 0.321924442 | 6.322012164 | 0.666247545 | -0.176364378 | FALSE | TRUE | FALSE |
| PLEKHM3 | 0.0236707 | 0.88352021 | 1 | 0.094618351 | 0.094618351 | 1.863516275 | 0.03458811 | 0.890209605 | 1 | 1.272636358 | 12.02196956 | 12.02196956 | 0.684359225 | -0.164715874 | FALSE | TRUE | FALSE |
| FMO1 | 0.0235482 | 0.88352847 | 0.50675 | 0.210064234 | 0.210064234 | 0.466568283 | 0.32674117 | 0.816480713 | 0.01265 | 0.326741172 | 0.326741172 | 0.721396801 | 0.072069783 | -1.142246783 | FALSE | TRUE | FALSE |
| SPAG17 | 0.0235383 | 0.88353673 | 0.5257 | 0.023538305 | 0.054520793 | 0.787167944 | 0.04609374 | 0.887552073 | 0.51935 | 0.04609374 | 0.04609374 | 0.892089214 | 0.510661653 | -0.291866753 | FALSE | TRUE | FALSE |
| BUB1B | 0.0234949 | 0.88354499 | 1 | 0.023494942 | 0.023494942 | 0.461332777 | 0.03314855 | 0.890566746 | 1 | 2.700549783 | 25.328936 | 53.14278975 | 0.708777447 | -0.14949011 | FALSE | TRUE | FALSE |
| TACC2 | 0.0234866 | 0.88355324 | 0.0183 | 1.37491E-05 | 0.023486576 | 0.443367146 | 9824.21592 | 0.00020172 | 0.0074 | 3.625748949 | 613.2678307 | 5588.450419 | 2.39068E-06 | -5.621478188 | FALSE | FALSE | FALSE |
| PARD3B | 0.0234608 | 0.8835615 | 0.75745 | 0.023460842 | 0.202381309 | 0.790355948 | 0.0220694 | 0.893209714 | 0.75685 | 0.047052028 | 0.188154107 | 0.92331431 | 1.063048535 | 0.026553093 | FALSE | TRUE | FALSE |
| DLGAP5 | 0.0234013 | 0.88356975 | 0.5075 | 0.202407878 | 0.202407878 | 0.78985369 | 0.03580355 | 0.889928631 | 1 | 0.312932368 | 0.312932368 | 1.20951833 | 0.65360315 | -0.184685863 | FALSE | TRUE | FALSE |
| WDPCP | 0.0233574 | 0.88357801 | 1 | 0.20622741 | 1.868869567 | 4.073623706 | 0.0382021 | 0.889402254 | 0.01605 | 0.038202102 | 0.038202102 | 0.755729694 | 0.611416761 | -0.21366266 | FALSE | TRUE | FALSE |
| PDE8A | 0.0233398 | 0.88358626 | 0.01995 | 0.023339755 | 0.023339755 | 0.460060123 | 0.03521725 | 0.89006928 | 1 | 0.324061539 | 0.324061539 | 1.190494389 | 0.66273646 | -0.178659136 | FALSE | TRUE | FALSE |
| SAMD15 | 0.0233037 | 0.88359451 | 0.016 | 0.023303702 | 0.023303702 | 0.460592641 | 0.30305565 | 0.824350069 | 0.5105 | 0.303055653 | 0.303055653 | 0.686113642 | 0.076895785 | -1.114097466 | FALSE | TRUE | FALSE |
| HLTF | 0.0232998 | 0.88360276 | 0.0816 | 0.005830693 | 0.093130795 | 0.11459129 | 0.02983101 | 0.891290134 | 0.6727 | 0.029831005 | 0.477080639 | 1.908606674 | 0.781060349 | -0.107315409 | FALSE | TRUE | FALSE |
| PTPRD | 0.023267 | 0.88361101 | 0.52845 | 0.023267015 | 0.112069433 | 0.44778132 | 0.15050197 | 0.86447255 | 0.9451 | 5.616262459 | 451.6154531 | 2113.958969 | 0.154596083 | -0.810801514 | FALSE | TRUE | FALSE |
| CCPG1 | 0.0232618 | 0.88361926 | 0.01795 | 0.02326176 | 0.02326176 | 0.459358034 | 0.02828862 | 0.891648603 | 1 | 0.028288624 | 0.028288624 | 0.559474412 | 0.822300845 | -0.084969263 | FALSE | TRUE | FALSE |
| NID1 | 0.0232578 | 0.88362751 | 0.3923 | 0.010492276 | 0.09293129 | 0.450865716 | 0.0248924 | 0.892471827 | 0.33325 | 0.024892399 | 0.39783935 | 1.93605351 | 0.934333823 | -0.02949793 | FALSE | TRUE | FALSE |
| TXNDC16 | 0.0232293 | 0.88363576 | 1 | 0.207584521 | 0.207584521 | 0.785570866 | 0.03016226 | 0.891205926 | 1 | 0.271168368 | 2.502162486 | 5.362034152 | 0.770144023 | -0.113428051 | FALSE | TRUE | FALSE |
| FOXP1 | 0.0232066 | 0.883644 | 1 | 0.212246032 | 0.212246032 | 4.184823193 | 0.03611356 | 0.889873846 | 1 | 0.331722438 | 0.331722438 | 1.22136001 | 0.642601136 | -0.192058511 | FALSE | TRUE | FALSE |
| PRICKLE1 | 0.0231984 | 0.88365225 | 1 | 0.092706421 | 0.092706421 | 1.822894323 | 5.21481244 | 0.297787622 | 0.0129 | 0.554727418 | 5.214812441 | 10.92977384 | 0.004448568 | -2.351779739 | FALSE | FALSE | FALSE |
| PDE1C | 0.023145 | 0.88366049 | 0.5084 | 0.210922937 | 0.210922937 | 4.161086766 | 0.32964131 | 0.81543574 | 0.51 | 0.329641311 | 0.329641311 | 1.218677279 | 0.070212556 | -1.153585219 | FALSE | TRUE | FALSE |
| FBLN7 | 0.0231069 | 0.88366873 | 1 | 0.092353571 | 0.369209958 | 1.826343726 | 0.03683505 | 0.889701349 | 1 | 0.036835052 | 0.036835052 | 0.729417266 | 0.627308626 | -0.20251874 | FALSE | TRUE | FALSE |
| LRRCC1 | 0.023075 | 0.88367697 | 0.9691 | 16.22461107 | 151.7302893 | 1453.690165 | 3.05261381 | 0.409412065 | 0.26395 | 0.338853922 | 3.052613814 | 11.43863661 | 0.007559086 | -2.121530721 | FALSE | FALSE | FALSE |
| EXOC3 | 0.023032 | 0.88368521 | 1 | 0.023032025 | 0.023032025 | 0.451975412 | 0.02613669 | 0.892088054 | 1 | 0.026136693 | 0.026136693 | 0.514062519 | 0.881214229 | -0.054918499 | FALSE | TRUE | FALSE |
| CLN8 | 0.023019 | 0.88369345 | 1 | 0.091983737 | 0.091983737 | 0.091983737 | 0.14371078 | 0.865947339 | 0.5042 | 0.143710781 | 0.143710781 | 0.143710781 | 0.160175808 | -0.795403075 | FALSE | TRUE | FALSE |
| NFATC1 | 0.0229812 | 0.88370169 | 0.0316 | 0.022981237 | 0.022981237 | 0.448351722 | 0.14137615 | 0.866495459 | 0.02615 | 0.141376154 | 0.141376154 | 2.766325251 | 0.162553843 | -0.78900276 | FALSE | TRUE | FALSE |
| PLXNA2 | 0.022953 | 0.88370992 | 1 | 0.022953026 | 0.438236036 | 0.769563027 | 0.0349335 | 0.890155072 | 1 | 0.139689505 | 0.670631123 | 10.72594549 | 0.6570491 | -0.182402175 | FALSE | TRUE | FALSE |
| KIF18A | 0.0229395 | 0.88371816 | 1 | 0.091709851 | 0.091709851 | 1.804747349 | 0.03500261 | 0.890123889 | 1 | 0.318411946 | 0.318411946 | 6.278101783 | 0.655364512 | -0.183517079 | FALSE | TRUE | FALSE |
| BAZ1A | 0.0228741 | 0.88372639 | 0.04275 | 0.022874084 | 0.022874084 | 0.766382479 | 0.03574371 | 0.889952095 | 0.03655 | 0.035743706 | 0.035743706 | 1.199672404 | 0.639947191 | -0.193855863 | FALSE | TRUE | FALSE |
| PIGZ | 0.0227956 | 0.88373463 | 0.50715 | 0.091152327 | 0.091152327 | 0.450163192 | 0.03462531 | 0.890201817 | 1 | 0.308824121 | 0.308824121 | 0.684811874 | 0.658350403 | -0.181542895 | FALSE | TRUE | FALSE |
| SLC7A4 | 0.0227764 | 0.88374286 | 0.4672 | 0.022776418 | 5.834412263 | 40.84211904 | 0.00025705 | 0.899937324 | 0.41445 | 0.000257051 | 0.016464657 | 0.148264591 | 88.60648836 | 1.947465525 | FALSE | TRUE | FALSE |
| SEC16A | 0.0227469 | 0.88375109 | 0.8835 | 1.904487516 | 18.10812307 | 130.4652162 | 0.0353438 | 0.890030243 | 1 | 2.993185884 | 28.62232885 | 100.2775091 | 0.643590591 | -0.191390314 | FALSE | TRUE | FALSE |
| TBL3 | 0.0226754 | 0.88375932 | 0.02715 | 0.022675408 | 0.022675408 | 0.443773552 | 0.30900278 | 0.822272735 | 0.51055 | 0.30900278 | 0.30900278 | 6.063036073 | 0.073382536 | -1.134407281 | FALSE | TRUE | FALSE |
| ZNF773 | 0.0226651 | 0.88376755 | 1 | 0.197245689 | 0.197245689 | 0.449812003 | 0.28873787 | 0.828470477 | 0.87565 | 2.599992014 | 24.0139675 | 24.0139675 | 0.078497179 | -1.10514595 | FALSE | TRUE | FALSE |
| TMEM132A | 0.0226608 | 0.88377578 | 0.7639 | 0.022660781 | 0.20519021 | 0.764823853 | 0.00375596 | 0.898410719 | 1 | 0.033411811 | 0.30505907 | 1.128674919 | 6.033286384 | 0.780553941 | FALSE | TRUE | FALSE |
| TES | 0.0226163 | 0.88378401 | 1 | 0.090409742 | 0.090409742 | 0.448927 | 0.13659873 | 0.867480462 | 0.7551 | 0.310601469 | 1.241731612 | 1.241731612 | 0.165567786 | -0.781024158 | FALSE | TRUE | FALSE |
| PCDHA3 | 0.0225915 | 0.88379224 | 0.51365 | 0.022591536 | 0.090314916 | 0.76294993 | 0.04432277 | 0.888037986 | 0.27445 | 0.044322768 | 0.394603576 | 1.498055769 | 0.509704976 | -0.292681127 | FALSE | TRUE | FALSE |
| TBX10 | 0.022588 | 0.88380046 | 0.75295 | 0.205024698 | 0.819562168 | 0.819562168 | 0.3078317 | 0.822835468 | 0.81345 | 1.230611485 | 46.03866739 | 46.03866739 | 0.073377733 | -1.134435712 | FALSE | TRUE | FALSE |
| VWA2 | 0.0225798 | 0.88380868 | 0.60125 | 0.022579783 | 0.756733452 | 3.026202983 | 0.01703599 | 0.894600221 | 0.00815 | 0.002010969 | 0.002010969 | 0.017035985 | 1.325416915 | 0.122352509 | FALSE | TRUE | FALSE |
| COG6 | 0.0225533 | 0.88381691 | 0.01645 | 0.022553275 | 0.022553275 | 0.44566314 | 0.03382139 | 0.890388437 | 1 | 0.033821393 | 0.299057219 | 0.669289447 | 0.66683459 | -0.17598188 | FALSE | TRUE | FALSE |
| OR4C46 | 0.0225402 | 0.88382513 | 0.0065 | 0.022540221 | 0.022540221 | 0.022540221 | 0.261962 | 0.83614274 | 0.50305 | 0.261961997 | 0.261961997 | 0.261961997 | 0.08604386 | -1.065280116 | FALSE | TRUE | FALSE |
| BCAS1 | 0.02253 | 0.88383335 | 0.5081 | 0.022530044 | 0.022530044 | 0.050055548 | 0.02876162 | 0.891534428 | 0.87895 | 0.264156441 | 2.488435077 | 2.488435077 | 0.783337178 | -0.106051261 | FALSE | TRUE | FALSE |
| SIGLEC12 | 0.0225292 | 0.88384157 | 1 | 0.022529155 | 0.022529155 | 0.44527963 | 19.7707995 | 0.133544091 | 0.78105 | 184.5857366 | 17289.93625 | 34943.98781 | 0.001139517 | -2.943279322 | FALSE | FALSE | FALSE |
| TBC1D9B | 0.022468 | 0.88384979 | 0.76445 | 0.049735786 | 0.198819882 | 3.860749107 | 0.04102525 | 0.888703926 | 0.7547 | 0.091535122 | 0.365940369 | 3.086790492 | 0.547662306 | -0.261487149 | FALSE | TRUE | FALSE |
| QRFP | 0.0224581 | 0.88385801 | 0.25025 | 0.022458132 | 0.207027856 | 0.207027856 | 0.02947068 | 0.891381862 | 0.50165 | 0.029470679 | 0.029470679 | 0.029470679 | 0.762050028 | -0.118016517 | FALSE | TRUE | FALSE |
| PCDHGA8 | 0.0224368 | 0.88386622 | 0.7603 | 0.089682192 | 0.79930352 | 1.762326801 | 218.88107 | 0.011782921 | 0.000025 | 0.045590717 | 0.410420439 | 1.541217167 | 0.000102507 | -3.989247948 | FALSE | FALSE | FALSE |
| TEKT2 | 0.0224277 | 0.88387444 | 1 | 0.192923102 | 1.703350661 | 3.816921379 | 0.30826762 | 0.82256427 | 0.0135 | 0.308267624 | 0.308267624 | 0.701549484 | 0.072753911 | -1.138143657 | FALSE | TRUE | FALSE |
| ASXL3 | 0.0224137 | 0.88388265 | 1 | 1.896445495 | 177.5985386 | 348.7719738 | 0.03316133 | 0.890559005 | 1 | 0.303176309 | 2.844814891 | 54.98729741 | 0.675898212 | -0.170118703 | FALSE | TRUE | FALSE |
| NLRP10 | 0.0224084 | 0.88389086 | 0.0172 | 0.022408403 | 0.022408403 | 0.442697702 | 0.03465433 | 0.890194029 | 0.0143 | 0.034654331 | 0.034654331 | 0.685632242 | 0.646626339 | -0.189346609 | FALSE | TRUE | FALSE |
| NES | 0.0223921 | 0.88389908 | 0.9999 | 6388.856889 | 6915465.666 | 74184994.06 | 0.00583405 | 0.897935448 | 0.9965 | 40.53409981 | 3915.470674 | 39847.49091 | 3.838170703 | 0.584124286 | FALSE | TRUE | FALSE |
| SYNGAP1 | 0.0223752 | 0.88390729 | 1 | 1585.325969 | 163605.9072 | 1719620.884 | 1.16886001 | 0.570692373 | 0.9816 | 2555.168414 | 267743.9523 | 2835663.627 | 0.019142732 | -1.71799608 | FALSE | FALSE | FALSE |
| DNAAF3 | 0.0223522 | 0.8839155 | 0.87265 | 1.689170176 | 15.25967879 | 15.25967879 | 0.3155656 | 0.820044914 | 0.4989 | 0.0363019 | 0.315565603 | 0.720322026 | 0.070832195 | -1.149769302 | FALSE | TRUE | FALSE |
| KLHL36 | 0.0223484 | 0.8839237 | 1 | 2.18856E+18 | 1.19035E+24 | 2.63129E+26 | 0.03378191 | 0.890411724 | 1 | 3.46132E+18 | 2.87473E+25 | 6.63589E+27 | 0.661548104 | -0.179438571 | FALSE | TRUE | FALSE |
| XPNPEP3 | 0.0223258 | 0.88393191 | 1 | 0.022325796 | 0.022325796 | 0.441827956 | 0.13654368 | 0.867502441 | 0.75545 | 0.136543676 | 0.545936575 | 2.705615007 | 0.163506625 | -0.786464645 | FALSE | TRUE | FALSE |
| FASTKD3 | 0.0222875 | 0.88394012 | 1 | 0.022287513 | 0.022287513 | 0.440612503 | 2.38135214 | 0.457113592 | 0.5116 | 2.381352139 | 21.91554028 | 21.91554028 | 0.009359184 | -2.028762007 | FALSE | FALSE | FALSE |
| ABCC1 | 0.0222872 | 0.88394832 | 0.99995 | 7899191012 | 3.86332E+14 | 3.77543E+16 | 0.00357988 | 0.89847829 | 0.9959 | 733.790576 | 1841532.493 | 47110838.56 | 6.225675834 | 0.794186504 | FALSE | TRUE | FALSE |
| FGA | 0.0222678 | 0.88395653 | 0.51385 | 0.201845698 | 1.876667909 | 3.972491359 | 0.37489545 | 0.800881625 | 0.8925 | 3.505091366 | 329.809955 | 3315.38697 | 0.059397254 | -1.22623363 | FALSE | TRUE | FALSE |
| FBRS | 0.022145 | 0.88396473 | 1 | 4.07692E+53 | 3.15583E+65 | 3.51437E+69 | 0.27706984 | 0.831794366 | 1 | 1.07591E+55 | 9.99624E+66 | 1.18059E+71 | 0.079925757 | -1.097313243 | FALSE | TRUE | FALSE |
| WNT10B | 0.0221013 | 0.88397293 | 0.5141 | 0.199371147 | 0.199371147 | 0.437476172 | 0.03554662 | 0.890006808 | 0.0122 | 0.035546616 | 0.035546616 | 0.704485611 | 0.621755596 | -0.206380298 | FALSE | TRUE | FALSE |
| KLHL40 | 0.022099 | 0.88398113 | 1 | 0.022098969 | 0.022098969 | 0.433360614 | 0.13367523 | 0.868071599 | 0.87895 | 0.534520528 | 2.137895032 | 10.50655903 | 0.165318355 | -0.781678925 | FALSE | TRUE | FALSE |
| BTN2A2 | 0.0220857 | 0.88398933 | 1 | 0.193065996 | 1.732265221 | 3.820322151 | 0.03580649 | 0.889920807 | 0.5065 | 0.143199375 | 0.143199375 | 0.709431293 | 0.61680756 | -0.209850312 | FALSE | TRUE | FALSE |
| SLC5A4 | 0.0220734 | 0.88399753 | 0.93025 | 1.798191061 | 162.1453288 | 570.3727762 | 0.21170847 | 0.849184706 | 0.3552 | 0.211708469 | 3.385385887 | 13.54271838 | 0.104262955 | -0.981869973 | FALSE | TRUE | FALSE |
| BBOX1 | 0.0220533 | 0.88400573 | 0.9398 | 0.35258986 | 12.61261278 | 12.61261278 | 25.9588697 | 0.106152863 | 0.0684 | 2.743415775 | 103.7905792 | 103.7905792 | 0.000849549 | -3.070811312 | FALSE | FALSE | FALSE |

| | | | | | | | | | | | | | | | | | |
|---|---|---|---|---|---|---|---|---|---|---|---|---|---|---|---|---|---|
| MARVELD2 | 0.0220248 | 0.88401393 | 0.5064 | 0.191041393 | 0.191041393 | 0.745291202 | 0.03397424 | 0.890372907 | 0.5095 | 0.297693985 | 0.297693985 | 0.67259677 | 0.648279607 | -0.18823764 | FALSE | TRUE | FALSE |
| LARP1B | 0.0220203 | 0.88402212 | 0.5092 | 0.191836848 | 0.191836848 | 3.775137358 | 0.03210379 | 0.890783117 | 1 | 2.540989829 | 23.49113645 | 50.10051539 | 0.685909369 | -0.163733265 | FALSE | TRUE | FALSE |
| EVI2B | 0.0220118 | 0.88403032 | 0.01015 | 0.022011777 | 0.022011777 | 0.437167854 | 0.02636196 | 0.892050293 | 0.0083 | 0.026361962 | 0.026361962 | 0.026361962 | 0.834982519 | -0.078322617 | FALSE | TRUE | FALSE |
| FRMD5 | 0.0219942 | 0.88403851 | 1 | 0.0219942 | 0.0219942 | 0.434970242 | 0.03444066 | 0.890264088 | 0.0139 | 0.034440658 | 0.034440658 | 0.682034075 | 0.638611484 | -0.194763276 | FALSE | TRUE | FALSE |
| SCP2 | 0.0219903 | 0.8840467 | 1 | 0.021990316 | 0.021990316 | 0.435678459 | 0.13465078 | 0.867853224 | 0.7513 | 0.134650775 | 0.538369724 | 0.66814431 | 0.163313695 | -0.786977396 | FALSE | TRUE | FALSE |
| OVCH1 | 0.0219645 | 0.88405489 | 0.93845 | 1.528869313 | 117.2618677 | 446.3613175 | 0.0576288 | 0.885131836 | 0.8123 | 0.485564282 | 4.19402572 | 37.11369208 | 0.381138006 | -0.418917743 | FALSE | TRUE | FALSE |
| FAM20A | 0.0219634 | 0.88406308 | 1 | 16.35407885 | 1522.249299 | 15219.89239 | 0.03410137 | 0.890341835 | 1 | 26.02161196 | 2461.806446 | 24814.22183 | 0.644061554 | -0.191072625 | FALSE | TRUE | FALSE |
| ZNF142 | 0.0218837 | 0.88407127 | 1 | 5292559.767 | 17581695737 | 7.94207E+11 | 0.02473909 | 0.892524305 | 1 | 1.42919E+17 | 5.35505E+22 | 1.01979E+25 | 0.884580942 | -0.053262421 | FALSE | TRUE | FALSE |
| CNTD1 | 0.0218809 | 0.88407946 | 0.00895 | 0.021880906 | 0.021880906 | 0.021880906 | 0.02567688 | 0.892246387 | 0.0068 | 0.025676882 | 0.025676882 | 0.025676882 | 0.852163681 | -0.069476979 | FALSE | TRUE | FALSE |
| PPP2R3A | 0.0218798 | 0.88408765 | 1 | 0.194061991 | 0.194061991 | 3.807561698 | 0.03506277 | 0.89009269 | 1 | 0.035062768 | 0.035062768 | 0.68951593 | 0.624017841 | -0.204802993 | FALSE | TRUE | FALSE |
| MTMR10 | 0.0218711 | 0.88409583 | 1 | 0.021871095 | 0.021871095 | 0.431965721 | 0.30456366 | 0.823969085 | 0.5086 | 0.304563659 | 0.304563659 | 1.124351554 | 0.071811243 | -1.143807557 | FALSE | TRUE | FALSE |
| DSPP | 0.021848 | 0.88410402 | 1 | 16.73694908 | 161.05136 | 1587.610356 | 0.03837757 | 0.889354895 | 1 | 0.345865851 | 0.345865851 | 1.297115155 | 0.569290969 | -0.244665706 | FALSE | TRUE | FALSE |
| REG1B | 0.0218203 | 0.8841122 | 1 | 16.38953362 | 15385.86003 | 15385.86003 | 18042.1936 | 0.000105201 | 0.384 | 18042.19357 | 21063494.89 | 231712011 | 1.2094E-06 | -5.917428828 | FALSE | FALSE | FALSE |
| SPAG6 | 0.021816 | 0.88412038 | 1 | 0.021816027 | 0.021816027 | 0.432680146 | 0.0339227 | 0.890380672 | 0.0108 | 0.033922703 | 0.033922703 | 0.673468544 | 0.643109926 | -0.191714787 | FALSE | TRUE | FALSE |
| P2RX2 | 0.0218124 | 0.88412856 | 1 | 0.021812402 | 0.021812402 | 0.430360452 | 0.03356097 | 0.890458272 | 1 | 0.033560967 | 0.033560967 | 0.663236005 | 0.649933653 | -0.187130975 | FALSE | TRUE | FALSE |
| EIF4G1 | 0.0218039 | 0.88413674 | 1 | 2.43988E+11 | 5.90361E+15 | 7.71602E+16 | 0.0395112 | 0.889069958 | 1 | 527260227.9 | 1.03614E+13 | 1.28908E+14 | 0.551841072 | -0.25818598 | FALSE | TRUE | FALSE |
| CDC42BPA | 0.021795 | 0.88414492 | 1 | 0.200286572 | 0.200286572 | 3.885852439 | 0.03526448 | 0.890061474 | 1 | 0.035264476 | 0.035264476 | 1.183418743 | 0.618043214 | -0.208981158 | FALSE | TRUE | FALSE |
| CATSPER4 | 0.0217307 | 0.8841531 | 0.017 | 0.021730658 | 0.021730658 | 0.081595888 | 0.01931308 | 0.893996854 | 0.49535 | 0.019313084 | 0.175221201 | 0.175221201 | 1.125178064 | 0.051221257 | FALSE | TRUE | FALSE |
| RDH8 | 0.0217066 | 0.88416128 | 1 | 0.021706558 | 0.021706558 | 0.021706558 | 0.03401133 | 0.890357373 | 1 | 0.034011331 | 0.034011331 | 0.034011331 | 0.638215478 | -0.195032668 | FALSE | TRUE | FALSE |
| PTGIS | 0.0217005 | 0.88416945 | 1 | 16.08154171 | 1029.053173 | 2453.631582 | 0.03925573 | 0.889117541 | 0.0138 | 0.039255732 | 0.039255732 | 0.776976979 | 0.552797197 | -0.257434168 | FALSE | TRUE | FALSE |
| GRIN3A | 0.0216923 | 0.88417763 | 1 | 0.086728418 | 0.086728418 | 1.698957789 | 2.72626596 | 0.433280739 | 0.2613 | 0.292964123 | 2.726265962 | 5.753182365 | 0.007956797 | -2.099261738 | FALSE | FALSE | FALSE |
| LRBA | 0.0216165 | 0.8841858 | 0.52975 | 0.021616536 | 0.195906643 | 3.722862487 | 0.00655399 | 0.89769632 | 0.3005 | 0.006553992 | 0.058594275 | 1.120188904 | 3.298224467 | 0.518280209 | FALSE | TRUE | FALSE |
| TMEM260 | 0.0216119 | 0.88419398 | 1 | 0.021611866 | 0.021611866 | 0.427162867 | 0.28229463 | 0.830512227 | 0.5076 | 0.282294628 | 0.282294628 | 0.641193103 | 0.076557837 | -1.116010344 | FALSE | TRUE | FALSE |
| CDAN1 | 0.0216007 | 0.88420215 | 0.5125 | 0.021600745 | 0.74200726 | 2.966016819 | 0.00504253 | 0.898112436 | 0.51475 | 0.005042534 | 0.024704164 | 0.098677457 | 4.283707963 | 0.631819856 | FALSE | TRUE | FALSE |
| FOCAD | 0.0215841 | 0.88421032 | 0.28005 | 0.021584068 | 0.184257785 | 1.031559465 | 0.00492219 | 0.898159991 | 1 | 0.359224578 | 3.18908152 | 12.06665129 | 4.385051213 | 0.64197467 | FALSE | TRUE | FALSE |
| MTHFD1 | 0.0215511 | 0.88421849 | 1 | 0.193410427 | 0.193410427 | 3.794103885 | 0.03471928 | 0.890178449 | 1 | 0.034719285 | 0.034719285 | 0.682654327 | 0.620724801 | -0.207100902 | FALSE | TRUE | FALSE |
| PDZK1IP1 | 0.0215464 | 0.88422665 | 1 | 0.021546361 | 0.021546361 | 0.021546361 | 0.02272218 | 0.893009378 | 1 | 0.022722184 | 0.022722184 | 0.022722184 | 0.94825222 | -0.023076132 | FALSE | TRUE | FALSE |
| DAO | 0.0215413 | 0.88423482 | 0.8776 | 0.086141879 | 0.740933998 | 0.740933998 | 0.01000957 | 0.896750393 | 0.5009 | 0.087051674 | 0.087051674 | 0.087051674 | 2.152068191 | 0.332856028 | FALSE | TRUE | FALSE |
| ADAMTS7 | 0.0215405 | 0.88424299 | 0.8806 | 0.193208502 | 1.777563801 | 6.487653833 | 0.00523698 | 0.898071642 | 0.52525 | 0.046273999 | 0.046273999 | 0.890608673 | 4.113158626 | 0.614175459 | FALSE | TRUE | FALSE |
| TOR3A | 0.0214679 | 0.88425115 | 1 | 0.18811065 | 0.18811065 | 0.425677939 | 0.03296411 | 0.890597702 | 1 | 0.291469588 | 1.165597864 | 1.165597864 | 0.651251786 | -0.186251073 | FALSE | TRUE | FALSE |
| CDH4 | 0.0214581 | 0.88425932 | 1 | 0.021458079 | 0.021458079 | 0.4175996 | 0.3065376 | 0.823336864 | 0.51905 | 0.3065376 | 0.3065376 | 5.984956364 | 0.070001458 | -1.154892912 | FALSE | TRUE | FALSE |
| CMTM2 | 0.0214166 | 0.88426748 | 1 | 0.02141665 | 0.02141665 | 0.02141665 | 0.02734891 | 0.891876297 | 1 | 0.027348909 | 0.027348909 | 0.027348909 | 0.783089714 | -0.10618848 | FALSE | TRUE | FALSE |
| FAM167A | 0.02141 | 0.88427564 | 1 | 0.02141004 | 0.02141004 | 0.02141004 | 0.02460401 | 0.892576736 | 1 | 0.024604005 | 0.225780632 | 0.225780632 | 0.870185162 | -0.060388326 | FALSE | TRUE | FALSE |
| FASTKD2 | 0.0213924 | 0.8842838 | 1 | 0.021392401 | 0.021392401 | 0.42284339 | 0.02947672 | 0.891374223 | 0.0145 | 0.02947672 | 0.02947672 | 0.583458451 | 0.725738864 | -0.13921962 | FALSE | TRUE | FALSE |
| OGFOD1 | 0.0213848 | 0.88429196 | 1 | 0.021384756 | 0.021384756 | 0.423396448 | 0.03319432 | 0.890528028 | 0.0121 | 0.033194316 | 0.033194316 | 0.658008317 | 0.644229451 | -0.190959425 | FALSE | TRUE | FALSE |
| GPT | 0.0213515 | 0.88430012 | 0.81555 | 0.085328861 | 3.055315212 | 3.055315212 | 0.00496268 | 0.898139616 | 0.8757 | 0.174751136 | 1.580211904 | 1.580211904 | 4.302402895 | 0.633711077 | FALSE | TRUE | FALSE |
| ELP2 | 0.0213433 | 0.88430828 | 0.506 | 0.085350154 | 0.341394561 | 1.683021441 | 0.08559501 | 0.879041454 | 0.50715 | 0.085595009 | 0.085595009 | 1.690703487 | 0.249351574 | -0.603187887 | FALSE | TRUE | FALSE |
| KLHL5 | 0.0213407 | 0.88431643 | 1 | 0.194527831 | 0.194527831 | 0.721401718 | 1.1999533 | 0.567193631 | 0.38505 | 0.523922964 | 4.798215891 | 10.35232888 | 0.017784626 | -1.749955275 | FALSE | FALSE | FALSE |
| FNDC1 | 0.0213027 | 0.88432459 | 1 | 0.021302652 | 0.409102655 | 0.71447046 | 0.03318044 | 0.890543518 | 0.04885 | 0.033180443 | 0.033180443 | 1.114655804 | 0.642024331 | -0.192448513 | FALSE | TRUE | FALSE |
| MAGI3 | 0.0212468 | 0.88433274 | 1 | 0.021246818 | 0.021246818 | 0.416018979 | 0.03309884 | 0.890589964 | 1 | 0.033098839 | 0.033098839 | 0.649723191 | 0.641920323 | -0.192518874 | FALSE | TRUE | FALSE |
| SELP | 0.0210158 | 0.8843409 | 0.8774 | 0.182973585 | 1.634069657 | 3.602430817 | 0.37150109 | 0.8019309 | 0.5045 | 0.371501092 | 3.359423079 | 3.359423079 | 0.056569831 | -1.247415121 | FALSE | TRUE | FALSE |
| MAP10 | 0.0209716 | 0.88434905 | 0.76155 | 0.020971595 | 0.175122121 | 0.707691476 | 0.33923318 | 0.812314349 | 0.50655 | 0.339233178 | 2.952664266 | 6.664492182 | 0.061820588 | -1.208866868 | FALSE | TRUE | FALSE |
| WDR3 | 0.020948 | 0.8843572 | 0.7602 | 0.02094797 | 0.083774812 | 0.411963392 | 0.14271378 | 0.866261075 | 0.825 | 0.693731682 | 11.09351712 | 44.37832804 | 0.146783086 | -0.833323985 | FALSE | TRUE | FALSE |
| MASTL | 0.0209014 | 0.88436535 | 1 | 0.020901388 | 0.020901388 | 0.411696481 | 0.03263845 | 0.890698204 | 1 | 0.032638446 | 0.032638446 | 0.644054306 | 0.640391642 | -0.193554345 | FALSE | TRUE | FALSE |
| DAGLB | 0.0208644 | 0.8843735 | 0.0224 | 0.02086441 | 0.02086441 | 0.410423322 | 0.03218957 | 0.890744535 | 1 | 0.1287296 | 0.1287296 | 1.089646251 | 0.648173019 | -0.188309051 | FALSE | TRUE | FALSE |
| MS4A7 | 0.0208582 | 0.88438165 | 1 | 0.020858239 | 0.020858239 | 0.020858239 | 0.02458927 | 0.892584222 | 1 | 0.024589269 | 0.024589269 | 0.024589269 | 0.848265917 | -0.071467982 | FALSE | TRUE | FALSE |
| FRY | 0.0207919 | 0.8843898 | 1 | 2.33376E+22 | 1.64763E+28 | 6.4099E+31 | 0.17501566 | 0.858314192 | 1 | 6.6614E+37 | 2.98732E+46 | 1.83855E+50 | 0.118800473 | -0.925181829 | FALSE | TRUE | FALSE |
| ASS1 | 0.0207503 | 0.88439795 | 0.96355 | 0.331336623 | 5.296006533 | 21.18120688 | 2.8677656 | 0.422309848 | 0.26515 | 2.867765597 | 45.87788807 | 183.5675511 | 0.007235702 | -2.140519352 | FALSE | FALSE | FALSE |
| OR2T33 | 0.0207158 | 0.88440609 | 1 | 7.54337E+24 | 5.66181E+30 | 2.27117E+34 | 0.01837266 | 0.894230211 | 1 | 1.61064E+33 | 6.50683E+41 | 2.33135E+44 | 1.127535899 | 0.052130378 | FALSE | TRUE | FALSE |
| SPINT1 | 0.0207158 | 0.88441424 | 1 | 0.020715799 | 0.020715799 | 0.408108639 | 0.0326718 | 0.89065184 | 1 | 0.032671801 | 0.032671801 | 0.644806823 | 0.634057446 | -0.197871393 | FALSE | TRUE | FALSE |
| TTF1 | 0.0206898 | 0.88442238 | 0.87595 | 1.590183387 | 14.48716355 | 31.27019036 | 0.03505206 | 0.890108292 | 0.0208 | 0.035052063 | 0.035052063 | 0.690673865 | 0.590260334 | -0.228956401 | FALSE | TRUE | FALSE |
| ABCF2 | 0.0206879 | 0.88443052 | 1 | 0.02068787 | 0.02068787 | 0.408342992 | 0.03271975 | 0.890636377 | 0.01725 | 0.032719748 | 0.032719748 | 0.646848558 | 0.632274738 | -0.19909417 | FALSE | TRUE | FALSE |
| BSN | 0.020683 | 0.88443866 | 1 | 0.020683035 | 0.377963044 | 0.68338675 | 0.03149884 | 0.890852508 | 1 | 0.291950761 | 5.39102067 | 9.679127928 | 0.65662855 | -0.182680238 | FALSE | TRUE | FALSE |
| PPP1R2 | 0.0206691 | 0.8844468 | 1 | 0.176524297 | 1.54742597 | 1.54742597 | 0.03379578 | 0.890403963 | 0.00555 | 0.033795777 | 0.033795777 | 0.033795777 | 0.61158788 | -0.21354113 | FALSE | TRUE | FALSE |
| LCTL | 0.0205627 | 0.88445494 | 0.0155 | 0.020562655 | 0.020562655 | 0.406293295 | 0.03211991 | 0.890775403 | 0.0141 | 0.032119914 | 0.032119914 | 0.635571732 | 0.640184003 | -0.193695182 | FALSE | TRUE | FALSE |
| IDH3B | 0.0205516 | 0.88446308 | 1 | 0.020551608 | 0.020551608 | 0.020551608 | 0.26054459 | 0.836588349 | 0.6894 | 2.390304123 | 89.93166308 | 89.93166308 | 0.078879429 | -1.103036241 | FALSE | TRUE | FALSE |
| C5orf15 | 0.0205462 | 0.88447122 | 1 | 0.020546155 | 0.020546155 | 0.020546155 | 0.02178425 | 0.893305924 | 1 | 0.02178425 | 0.02178425 | 0.02178425 | 0.943165583 | -0.025412055 | FALSE | TRUE | FALSE |
| CABIN1 | 0.0205266 | 0.88447935 | 0.2837 | 0.002360576 | 0.020526568 | 0.391351139 | 0.00298538 | 0.898687231 | 0.5291 | 0.002985377 | 0.014321722 | 0.057239275 | 6.87570336 | 0.837317132 | FALSE | TRUE | FALSE |
| TRIM32 | 0.0205231 | 0.88448749 | 1 | 0.186860751 | 1.746189768 | 3.681086587 | 0.03441045 | 0.890271867 | 1 | 0.034410449 | 0.034410449 | 0.679098298 | 0.596421674 | -0.224446582 | FALSE | TRUE | FALSE |
| EPHA5 | 0.0205117 | 0.88449562 | 1 | 0.190869908 | 0.190869908 | 3.741140376 | 0.03350314 | 0.89047378 | 1 | 0.033503143 | 0.033503143 | 0.658257294 | 0.61223092 | -0.21308474 | FALSE | TRUE | FALSE |
| CCDC115 | 0.020489 | 0.88450375 | 1 | 0.183794287 | 0.183794287 | 0.183794287 | 0.03319403 | 0.890535774 | 1 | 0.033194029 | 0.033194029 | 0.033194029 | 0.617248626 | -0.209539868 | FALSE | TRUE | FALSE |
| NAT2 | 0.0204546 | 0.88451189 | 1 | 0.02045463 | 0.02045463 | 0.02045463 | 0.02477364 | 0.892509316 | 1 | 0.024773636 | 0.024773636 | 0.024773636 | 0.825661226 | -0.08319811 | FALSE | TRUE | FALSE |
| LRSAM1 | 0.0204334 | 0.88452002 | 0.0229 | 0.0204334 | 0.0204334 | 0.401625794 | 0.03216316 | 0.890759971 | 0.02035 | 0.032163156 | 0.032163156 | 0.633489886 | 0.635304575 | -0.197018017 | FALSE | TRUE | FALSE |
| SELPLG | 0.0204133 | 0.88452815 | 0.0121 | 0.020413344 | 0.020413344 | 0.404754314 | 0.02393773 | 0.892741207 | 0.0098 | 0.023937728 | 0.023937728 | 0.023937728 | 0.852768624 | -0.069168787 | FALSE | TRUE | FALSE |
| QKI | 0.0203706 | 0.88453628 | 1 | 0.020370624 | 0.020370624 | 0.020370624 | 0.03044057 | 0.891175276 | 1 | 0.030440573 | 0.030440573 | 0.030440573 | 0.669193188 | -0.174448489 | FALSE | TRUE | FALSE |
| ADCY2 | 0.0203556 | 0.8845444 | 1 | 0.183968144 | 0.183968144 | 3.585119564 | 1.15319297 | 0.57200641 | 0.26275 | 0.288419585 | 1.153192969 | 5.637990204 | 0.017651522 | -1.753217846 | FALSE | FALSE | FALSE |
| SCAND1 | 0.0203543 | 0.88455253 | 0.0039 | 0.020354259 | 0.020354259 | 0.020354259 | 0.0224868 | 0.893076233 | 0.0032 | 0.0224868 | 0.0224868 | 0.0224868 | 0.90516477 | -0.043272357 | FALSE | TRUE | FALSE |
| BAG5 | 0.0202867 | 0.88456065 | 1 | 0.185613475 | 0.741889546 | 0.741889546 | 0.03344669 | 0.890489284 | 1 | 0.033446694 | 0.033446694 | 0.663245924 | 0.606538788 | -0.217141421 | FALSE | TRUE | FALSE |
| APOC1 | 0.0202831 | 0.88456878 | 0.747 | 0.020283145 | 0.190967155 | 0.190967155 | 0.02432852 | 0.892681453 | 0.0015 | 0.024328519 | 0.024328519 | 0.024328519 | 0.833718852 | -0.078980378 | FALSE | TRUE | FALSE |
| SLC9C2 | 0.020281 | 0.8845769 | 1 | 0.177431724 | 0.177431724 | 3.481877195 | 0.27984065 | 0.831111744 | 0.5184 | 0.27984065 | 0.27984065 | 5.503965071 | 0.072473397 | -1.139821383 | FALSE | TRUE | FALSE |
| P4HA3 | 0.0202071 | 0.88458502 | 1 | 0.020207148 | 0.020207148 | 0.400239263 | 0.03072366 | 0.891090912 | 0.87825 | 0.491296023 | 1.965355865 | 2.435974787 | 0.657706371 | -0.181967951 | FALSE | TRUE | FALSE |
| ACAA2 | 0.0200852 | 0.88459315 | 0.9359 | 0.08027256 | 1.283139145 | 1.283139145 | 0.1510281 | 0.864392279 | 0.50065 | 0.151028102 | 0.603830129 | 0.603830129 | 0.13298995 | -0.876181178 | FALSE | TRUE | FALSE |
| AXL | 0.0200646 | 0.88460127 | 1 | 16067.4953 | 7216182.259 | 79072972.33 | 0.11928911 | 0.871382139 | 1 | 257019.1381 | 216904921.8 | 5489988148 | 0.16820174 | -0.774169516 | FALSE | TRUE | FALSE |

| | | | | | | | | | | | | | | | | | |
|---|---|---|---|---|---|---|---|---|---|---|---|---|---|---|---|---|---|
| NOL8 | 0.0200641 | 0.88460939 | 1 | 0.174041737 | 1.549541178 | 5.871856842 | 20.699348 | 0.129216265 | 0.1979 | 20.69934797 | 195.241669 | 1886.614354 | 0.000969313 | -3.013535977 | FALSE | FALSE | FALSE |
| THSD7A | 0.0200129 | 0.8846175 | 0.0454 | 0.020012855 | 0.020012855 | 0.669601427 | 0.03136002 | 0.890898726 | 0.52075 | 0.12538091 | 0.12538091 | 2.437276618 | 0.638164568 | -0.195067313 | FALSE | TRUE | FALSE |
| PTCD3 | 0.0199651 | 0.88462562 | 1 | 0.01996512 | 0.01996512 | 0.394225471 | 0.03102824 | 0.890998741 | 1 | 0.031028238 | 0.031028238 | 0.613611922 | 0.64345002 | -0.191485181 | FALSE | TRUE | FALSE |
| EVX1 | 0.019963 | 0.88463374 | 0.0125 | 0.019963015 | 0.019963015 | 0.394957426 | 0.03100858 | 0.891014113 | 1 | 0.12392384 | 0.495377702 | 1.051718255 | 0.643790106 | -0.191255702 | FALSE | TRUE | FALSE |
| CHRNA1 | 0.0199509 | 0.88464185 | 0.75795 | 0.180529305 | 1.676617879 | 1.676617879 | 0.00977076 | 0.89679911 | 0.75495 | 0.039045229 | 0.156068811 | 0.331249316 | 2.041899597 | 0.310034383 | FALSE | TRUE | FALSE |
| DNMT1 | 0.0199496 | 0.88464997 | 1 | 0.019949593 | 0.382823429 | 0.670024388 | 0.03158824 | 0.890844801 | 0.04615 | 0.031588241 | 0.031588241 | 1.0624733 | 0.631551252 | -0.199591399 | FALSE | TRUE | FALSE |
| SLC51B | 0.0199151 | 0.88465808 | 0.0025 | 0.019915087 | 0.019915087 | 0.019915087 | 0.02092225 | 0.893586234 | 0.00185 | 0.020922255 | 0.020922255 | 0.020922255 | 0.951861426 | -0.021426273 | FALSE | TRUE | FALSE |
| SPAG1 | 0.0198866 | 0.88466619 | 1 | 0.019886577 | 0.019886577 | 0.392251488 | 2.38280766 | 0.45692887 | 0.2581 | 0.2614387 | 2.382807664 | 5.165281032 | 0.008345859 | -2.078528948 | FALSE | FALSE | FALSE |
| RNF123 | 0.0197831 | 0.8846743 | 0.9407 | 0.401461842 | 15.01848899 | 31.10211103 | 0.00986018 | 0.896771277 | 0.51695 | 0.088467558 | 0.088467558 | 1.721044001 | 2.006364957 | 0.302409934 | FALSE | TRUE | FALSE |
| SLC6A5 | 0.0197781 | 0.88468241 | 0.8754 | 0.079068217 | 0.316175015 | 1.552499722 | 0.03630071 | 0.889834685 | 0.93805 | 0.14513493 | 2.321727781 | 2.855978824 | 0.544841052 | -0.263730177 | FALSE | TRUE | FALSE |
| HELZ | 0.0197655 | 0.88469052 | 0.79995 | 0.44685162 | 44.65576001 | 178.3680692 | 7.7197E-05 | 0.900251775 | 0.5226 | 0.012536108 | 0.012536108 | 0.242997074 | 256.0378573 | 2.408304184 | FALSE | TRUE | FALSE |
| CPAMD8 | 0.0197281 | 0.88469863 | 0.7009 | 0.019728105 | 1.471767888 | 3.227980683 | 0.00634788 | 0.897771587 | 0.28835 | 0.00634788 | 0.053797168 | 1.036959532 | 3.1078256 | 0.49245664 | FALSE | TRUE | FALSE |
| DDX60 | 0.0197229 | 0.88470674 | 0.87955 | 0.167134434 | 1.45280674 | 5.605455235 | 0.00495302 | 0.898146408 | 1 | 0.041592306 | 0.041592306 | 0.810747666 | 3.981995034 | 0.600100714 | FALSE | TRUE | FALSE |
| ZSCAN5B | 0.0197207 | 0.88471484 | 1 | 0.171106978 | 0.171106978 | 0.390641634 | 0.03082361 | 0.89107556 | 1 | 0.270159064 | 0.270159064 | 0.611277803 | 0.639792721 | -0.193960705 | FALSE | TRUE | FALSE |
| SEMA4F | 0.0197048 | 0.88472295 | 0.5106 | 0.171150169 | 0.171150169 | 3.359266369 | 0.12407763 | 0.870345314 | 0.26425 | 0.124077634 | 0.496389223 | 2.440806034 | 0.158810204 | -0.799121597 | FALSE | TRUE | FALSE |
| SOX13 | 0.0196793 | 0.88473105 | 1 | 0.019679268 | 0.019679268 | 0.388689217 | 0.48943131 | 0.759078445 | 0.25815 | 0.122439055 | 0.489431311 | 2.421938351 | 0.040208438 | -1.395682799 | FALSE | TRUE | FALSE |
| LRRC43 | 0.0196775 | 0.88473915 | 1 | 1.516736378 | 13.83790347 | 29.8349642 | 0.03355127 | 0.890466027 | 1 | 0.033551266 | 0.033551266 | 0.661279128 | 0.586491069 | -0.231738597 | FALSE | TRUE | FALSE |
| CDH1 | 0.019632 | 0.88474725 | 1 | 6623.508449 | 2930441.849 | 33138197.32 | 0.5639093 | 0.727282047 | 0.9646 | 120.8559183 | 12351.15288 | 129388.0124 | 0.034814149 | -1.458244214 | FALSE | TRUE | FALSE |
| TLE2 | 0.0196145 | 0.88475535 | 1 | 0.173663475 | 0.173663475 | 3.407559492 | 0.03089095 | 0.891037163 | 1 | 0.275676757 | 0.275676757 | 5.421548924 | 0.634959618 | -0.197253894 | FALSE | TRUE | FALSE |
| CTSH | 0.0195897 | 0.88476345 | 1 | 14.52008142 | 1347.435582 | 13451.61004 | 0.56908867 | 0.725089669 | 0.49935 | 0.142287062 | 2.276685278 | 2.276685278 | 0.0344229 | -1.463152547 | FALSE | TRUE | FALSE |
| CCDC158 | 0.0195868 | 0.88477155 | 0.99195 | 12.42703565 | 1036.788013 | 3778.981216 | 2158.89698 | 0.001096853 | 0.0443 | 25.08154552 | 2158.896979 | 20741.07104 | 9.07261E-06 | -5.042267576 | FALSE | FALSE | FALSE |
| SLC25A35 | 0.0195455 | 0.88477965 | 1 | 0.019545531 | 0.019545531 | 0.019545531 | 0.03086909 | 0.891052525 | 0.0081 | 0.030869093 | 0.030869093 | 0.030869093 | 0.633174784 | -0.198476389 | FALSE | TRUE | FALSE |
| DSTYK | 0.019542 | 0.88478774 | 1 | 16.74688171 | 668.5668442 | 6840.906293 | 0.0216072 | 0.893342885 | 1 | 52104912.89 | 1.88492E+11 | 3.01685E+12 | 0.904418992 | -0.043630326 | FALSE | TRUE | FALSE |
| PRMT7 | 0.0194907 | 0.88479584 | 1 | 0.019490717 | 0.019490717 | 0.383615291 | 0.03067644 | 0.891098586 | 0.01745 | 0.030676445 | 0.030676445 | 0.604927996 | 0.635364265 | -0.196977215 | FALSE | TRUE | FALSE |
| SLC2A10 | 0.0194475 | 0.88480393 | 1 | 1019.476587 | 261288.0026 | 2760621.972 | 0.1234641 | 0.870482347 | 0.998 | 1642.016918 | 426621.8429 | 3255747.007 | 0.157515384 | -0.802677023 | FALSE | TRUE | FALSE |
| PITPNM1 | 0.0194396 | 0.88481202 | 1 | 0.01943964 | 0.01943964 | 0.655130744 | 0.02985578 | 0.891282484 | 0.519 | 0.269792313 | 0.269792313 | 5.237186497 | 0.651118202 | -0.186340164 | FALSE | TRUE | FALSE |
| MCOLN3 | 0.0193693 | 0.88482012 | 1 | 0.172240805 | 0.172240805 | 0.383692232 | 2.41465698 | 0.453769999 | 0.1336 | 1.050911783 | 9.654828221 | 9.654828221 | 0.00802155 | -2.095741709 | FALSE | FALSE | FALSE |
| NR6A1 | 0.0193662 | 0.88482821 | 0.51125 | 0.077384674 | 0.077384674 | 0.383649098 | 0.03088639 | 0.891044845 | 0.5025 | 0.123432978 | 0.123432978 | 0.612565875 | 0.627013156 | -0.202723347 | FALSE | TRUE | FALSE |
| ZNF91 | 0.019328 | 0.8848363 | 1 | 0.019327987 | 0.019327987 | 0.651154413 | 0.02874709 | 0.891542047 | 1 | 0.256361207 | 2.346469257 | 8.646216601 | 0.672345967 | -0.172407196 | FALSE | TRUE | FALSE |
| WDSUB1 | 0.0193188 | 0.88484439 | 1 | 0.019318787 | 0.019318787 | 0.383199666 | 0.11706026 | 0.871930892 | 0.75115 | 0.26153385 | 1.045725564 | 1.045725564 | 0.165032844 | -0.782429617 | FALSE | TRUE | FALSE |
| TREX1 | 0.0192817 | 0.88485247 | 1 | 0.019281682 | 0.019281682 | 0.019281682 | 0.03052221 | 0.891136944 | 1 | 0.030522207 | 0.030522207 | 0.030522207 | 0.631726343 | -0.199471013 | FALSE | TRUE | FALSE |
| CCDC88C | 0.0192743 | 0.88486056 | 1 | 0.171025766 | 1.557588307 | 29.78837337 | 0.2804138 | 0.831006244 | 0.5192 | 0.280413796 | 0.602406198 | 5.391171483 | 0.068735326 | -1.162820005 | FALSE | TRUE | FALSE |
| GRIK1 | 0.0192654 | 0.88486865 | 0.75345 | 0.042379901 | 0.169400209 | 0.832755271 | 2.79871918 | 0.427235332 | 0.13395 | 0.307700035 | 2.798719184 | 6.05898224 | 0.006883663 | -2.162180369 | FALSE | FALSE | FALSE |
| UVSSA | 0.0192442 | 0.88487673 | 1 | 0.16708674 | 0.16708674 | 3.275196131 | 0.03106506 | 0.890991054 | 1 | 0.031065059 | 0.031065059 | 0.610390406 | 0.619481547 | -0.207971626 | FALSE | TRUE | FALSE |
| POPDC3 | 0.019232 | 0.88488481 | 1 | 0.019232048 | 0.019232048 | 0.019232048 | 0.02131732 | 0.893453628 | 1 | 0.195887134 | 1.847450576 | 1.847450576 | 0.902179306 | -0.044707139 | FALSE | TRUE | FALSE |
| LCP1 | 0.0191915 | 0.8848929 | 1 | 0.019191538 | 0.019191538 | 0.379639854 | 0.1211081 | 0.871017334 | 0.5094 | 0.121108097 | 0.121108097 | 0.600262898 | 0.158466186 | -0.800063394 | FALSE | TRUE | FALSE |
| SIPA1L1 | 0.0191659 | 0.88490098 | 0.7597 | 0.019165864 | 0.092418613 | 1.475898699 | 0.08282168 | 0.879614328 | 0.04345 | 0.082821681 | 0.082821681 | 1.602819428 | 0.231411195 | -0.635615634 | FALSE | TRUE | FALSE |
| EWSR1 | 0.0191446 | 0.88490906 | 1 | 18188107.73 | 1.13162E+11 | 1.36645E+12 | 0.0521723 | 0.886195794 | 1 | 0.0521723 | 0.0521723 | 1.029835889 | 0.366949558 | -0.435393631 | FALSE | TRUE | FALSE |
| AFF1 | 0.0190424 | 0.88491714 | 1 | 0.174974489 | 1.650150337 | 5.896892165 | 0.03228588 | 0.890729095 | 0.02925 | 0.032285876 | 0.032285876 | 0.632065748 | 0.589805328 | -0.229291309 | FALSE | TRUE | FALSE |
| TET1 | 0.0190368 | 0.88492522 | 1 | 1.683546406 | 164.775001 | 316.8405577 | 0.18203135 | 0.856508072 | 1 | 0.728122597 | 0.728122597 | 14.08606573 | 0.104579997 | -0.980551375 | FALSE | TRUE | FALSE |
| MEN1 | 0.0190111 | 0.88493329 | 1 | 0.019011137 | 0.019011137 | 0.373823486 | 0.02946617 | 0.891389499 | 0.50495 | 0.278789865 | 0.278789865 | 5.493062341 | 0.645185267 | -0.190315558 | FALSE | TRUE | FALSE |
| NPR1 | 0.0190004 | 0.88494137 | 1 | 0.303670951 | 2.779778943 | 10.24775363 | 0.03193339 | 0.890806255 | 0.02465 | 0.031933392 | 0.031933392 | 0.627229448 | 0.595001734 | -0.225481769 | FALSE | TRUE | FALSE |
| NAT16 | 0.0189979 | 0.88494944 | 1 | 0.01899786 | 0.01899786 | 0.375378665 | 0.02204821 | 0.89321712 | 1 | 0.2028075 | 0.2028075 | 0.436282161 | 0.861650888 | -0.06466866 | FALSE | TRUE | FALSE |
| KCNC2 | 0.0189921 | 0.88495752 | 1 | 0.177459636 | 1.701820221 | 3.497035337 | 0.12489465 | 0.870155123 | 0.8743 | 0.499263707 | 1.996294923 | 2.465538142 | 0.15206525 | -0.81797002 | FALSE | TRUE | FALSE |
| LRP12 | 0.0189686 | 0.88496559 | 0.5125 | 0.075807785 | 0.075807785 | 1.490944513 | 0.03064657 | 0.89110626 | 1 | 0.030646566 | 0.030646566 | 0.60394584 | 0.618945456 | -0.208347621 | FALSE | TRUE | FALSE |
| CCDC148 | 0.0189165 | 0.88497366 | 1 | 0.018916523 | 0.018916523 | 0.374794926 | 0.02869578 | 0.891549664 | 1 | 0.248010776 | 0.248010776 | 0.569191336 | 0.659209268 | -0.180976695 | FALSE | TRUE | FALSE |
| FAHD2A | 0.0188893 | 0.88498173 | 1 | 0.075490337 | 0.075490337 | 0.075490337 | 0.47103134 | 0.766715536 | 0.5016 | 0.471031342 | 1.883579445 | 1.883579445 | 0.04010206 | -1.396833319 | FALSE | TRUE | FALSE |
| GCC2 | 0.0188862 | 0.8849898 | 0.7623 | 0.16469036 | 1.474028429 | 28.70083971 | 0.03060884 | 0.891121604 | 0.5198 | 0.269710356 | 0.269710356 | 5.268277189 | 0.617019348 | -0.209701218 | FALSE | TRUE | FALSE |
| TMPO | 0.018841 | 0.88499787 | 0.7508 | 0.165310956 | 1.488719485 | 1.488719485 | 0.27122031 | 0.833485841 | 0.50545 | 0.271220307 | 0.271220307 | 0.607568457 | 0.069467422 | -1.158218817 | FALSE | TRUE | FALSE |
| SLC7A7 | 0.0188275 | 0.88500594 | 0.2601 | 0.018827538 | 0.17144892 | 0.372867869 | 0.00461798 | 0.898193936 | 1 | 0.004617977 | 0.004617977 | 0.091563801 | 4.077010029 | 0.61034178 | FALSE | TRUE | FALSE |
| ESRRG | 0.018819 | 0.88501401 | 0.50975 | 0.176765998 | 0.176765998 | 0.372805308 | 0.03005278 | 0.891221245 | 1 | 0.480029243 | 1.919208041 | 1.919208041 | 0.626199939 | -0.203286979 | FALSE | TRUE | FALSE |
| GALNT3 | 0.0188011 | 0.88502207 | 1 | 0.018801102 | 0.018801102 | 0.37175518 | 0.11790921 | 0.871734677 | 0.5068 | 0.117909207 | 0.117909207 | 0.998363064 | 0.15945406 | -0.797364418 | FALSE | TRUE | FALSE |
| IL12RB2 | 0.0187061 | 0.88503014 | 1 | 0.1621743 | 1.443105598 | 3.19473727 | 0.03117861 | 0.890960294 | 0.02125 | 0.031178614 | 0.031178614 | 0.615311245 | 0.599965658 | -0.221873608 | FALSE | TRUE | FALSE |
| MFSD11 | 0.0186992 | 0.8850382 | 1 | 0.166363459 | 1.519149696 | 1.519149696 | 0.03147667 | 0.890867918 | 1 | 0.031476668 | 0.031476668 | 0.031476668 | 0.594065698 | -0.226165523 | FALSE | TRUE | FALSE |
| PCDHA7 | 0.0186306 | 0.88504626 | 1 | 0.018630627 | 0.018630627 | 0.362489925 | 1.00775806 | 0.587671641 | 0.3909 | 0.449027096 | 4.030742652 | 19.67182127 | 0.018487202 | -1.73312881 | FALSE | FALSE | FALSE |
| EFEMP2 | 0.0186291 | 0.88505432 | 1 | 0.018629114 | 0.018629114 | 0.368236953 | 0.45173266 | 0.773863744 | 0.505 | 0.451732658 | 4.206209119 | 4.206209119 | 0.041239245 | -1.384689294 | FALSE | TRUE | FALSE |
| ZNF335 | 0.0185812 | 0.88506238 | 1 | 0.162504972 | 1.458727558 | 28.23489627 | 0.03110037 | 0.890983365 | 0.03955 | 0.031100374 | 0.031100374 | 0.60431231 | 0.597458417 | -0.223692316 | FALSE | TRUE | FALSE |
| LAMC2 | 0.0184975 | 0.88507044 | 0.9999 | 487100.9455 | 573271480.6 | 6312284623 | 0.01542571 | 0.895025335 | 1 | 102968947.7 | 1.15846E+11 | 1.34666E+12 | 1.199132506 | 0.078867176 | FALSE | TRUE | FALSE |
| DNMT3L | 0.0184609 | 0.8850785 | 0.0119 | 0.018460923 | 0.018460923 | 0.365686115 | 0.02926226 | 0.891442934 | 0.0112 | 0.029262261 | 0.029262261 | 0.580313138 | 0.630878242 | -0.200054451 | FALSE | TRUE | FALSE |
| ITGAV | 0.0184469 | 0.88508656 | 1 | 0.073745741 | 0.073745741 | 1.445305058 | 0.02888721 | 0.891511567 | 1 | 0.263062879 | 0.263062879 | 5.168100142 | 0.638583886 | -0.194782045 | FALSE | TRUE | FALSE |
| AGXT2 | 0.018415 | 0.88509462 | 0.6305 | 0.073627967 | 0.641006043 | 0.641006043 | 0.00901915 | 0.897014355 | 1 | 0.009019151 | 0.009019151 | 0.178840837 | 2.041771121 | 0.310007057 | FALSE | TRUE | FALSE |
| VWA5A | 0.0184115 | 0.88510267 | 1 | 0.018411522 | 0.018411522 | 0.362942924 | 0.22274567 | 0.846668372 | 0.69535 | 1.97733841 | 168.0494554 | 168.0494554 | 0.08265715 | -1.082719574 | FALSE | TRUE | FALSE |
| DPF3 | 0.0183951 | 0.88511073 | 1 | 0.018395145 | 0.018395145 | 0.364313849 | 0.02911073 | 0.891473447 | 0.51035 | 0.116356611 | 0.116356611 | 0.577211397 | 0.63190254 | -0.199349899 | FALSE | TRUE | FALSE |
| SLC16A4 | 0.0183755 | 0.88511878 | 1 | 0.161142523 | 0.161142523 | 0.364565479 | 0.25723858 | 0.837390926 | 0.5074 | 0.257238579 | 0.257238579 | 0.257238579 | 0.07143363 | -1.14609728 | FALSE | TRUE | FALSE |
| INTU | 0.0183214 | 0.88512683 | 1 | 0.018321392 | 0.018321392 | 0.360233846 | 0.0280834 | 0.891732189 | 1 | 0.246422035 | 0.246422035 | 4.854991588 | 0.652392259 | -0.185491201 | FALSE | TRUE | FALSE |
| LILRA5 | 0.0182817 | 0.88513488 | 1 | 0.018281748 | 0.018281748 | 0.018281748 | 0.02239116 | 0.893105922 | 0.0065 | 0.022391156 | 0.022391156 | 0.022391156 | 0.816471831 | -0.088058794 | FALSE | TRUE | FALSE |
| FRK | 0.0182717 | 0.88514293 | 1 | 0.018271692 | 0.018271692 | 0.361981919 | 0.25134255 | 0.839021073 | 0.50615 | 0.251342549 | 0.251342549 | 0.558817506 | 0.072696374 | -1.138487251 | FALSE | TRUE | FALSE |
| WDR76 | 0.0182532 | 0.88515098 | 1 | 0.018253225 | 0.018253225 | 0.36075933 | 2.21473924 | 0.471159487 | 0.25565 | 0.242740434 | 2.214739237 | 4.804363482 | 0.008241704 | -2.083982994 | FALSE | FALSE | FALSE |
| ZDBF2 | 0.0182526 | 0.88515903 | 0.5228 | 0.160371884 | 0.350667388 | 3.081054851 | 2.25882875 | 0.467620188 | 0.27225 | 0.248511859 | 2.258828746 | 43.60289883 | 0.008080549 | -2.092559127 | FALSE | FALSE | FALSE |
| ATOH1 | 0.0182514 | 0.88516708 | 0.01485 | 0.018251354 | 0.018251354 | 0.360871931 | 0.02867985 | 0.891557281 | 1 | 0.11460187 | 0.458052343 | 0.567844238 | 0.636382514 | -0.196281762 | FALSE | TRUE | FALSE |
| ORC1 | 0.0182443 | 0.88517512 | 1 | 0.163059519 | 0.163059519 | 3.201106774 | 0.02960779 | 0.891336015 | 1 | 0.118386008 | 0.118386008 | 2.329253548 | 0.616199691 | -0.210278524 | FALSE | TRUE | FALSE |
| DSCAM | 0.018206 | 0.88518317 | 1 | 0.018205987 | 0.347116488 | 0.609209224 | 0.02918442 | 0.891458193 | 1 | 0.029184422 | 0.559571429 | 0.978429933 | 0.623825504 | -0.204936874 | FALSE | TRUE | FALSE |
| CEACAM21 | 0.0181737 | 0.88519121 | 0.0085 | 0.018173655 | 0.018173655 | 0.018173655 | 0.02688154 | 0.891959574 | 1 | 0.231308475 | 2.042886258 | 2.042886258 | 0.676064444 | -0.170011904 | FALSE | TRUE | FALSE |

| | | | | | | | | | | | | | | | | | |
|---|---|---|---|---|---|---|---|---|---|---|---|---|---|---|---|---|---|
| CDK5RAP1 | 0.0181679 | 0.88519926 | 1 | 0.018167948 | 0.018167948 | 0.358928834 | 0.25129547 | 0.839037051 | 0.5062 | 0.251295472 | 0.251295472 | 0.555280569 | 0.072297155 | -1.140878791 | FALSE | TRUE | FALSE |
| WDR59 | 0.0181667 | 0.8852073 | 0.88035 | 0.290496814 | 1.162081478 | 5.679408002 | 0.02995079 | 0.891259527 | 1 | 0.029950789 | 0.029950789 | 0.587142876 | 0.606552406 | -0.21713167 | FALSE | TRUE | FALSE |
| ADAM28 | 0.0181621 | 0.88521534 | 0.5141 | 0.038782977 | 0.155064669 | 0.614460264 | 0.63101179 | 0.698852059 | 0.25615 | 0.071219993 | 0.631011791 | 1.407399061 | 0.028782522 | -1.540871156 | FALSE | TRUE | FALSE |
| SNX32 | 0.0181257 | 0.88522338 | 1 | 0.154712015 | 0.154712015 | 0.358534331 | 0.23151861 | 0.844518963 | 0.87765 | 2.049056836 | 18.60163013 | 18.60163013 | 0.078290366 | -1.106291678 | FALSE | TRUE | FALSE |
| KIAA1217 | 0.0180858 | 0.88523142 | 1 | 0.01808578 | 0.347073561 | 0.606651794 | 0.02883778 | 0.891519189 | 1 | 0.028837782 | 0.028837782 | 0.968871016 | 0.627155713 | -0.202624617 | FALSE | TRUE | FALSE |
| SCLT1 | 0.0180025 | 0.88523945 | 1 | 0.153980973 | 1.351833665 | 3.044919372 | 0.02995469 | 0.891251873 | 1 | 0.029954694 | 0.029954694 | 0.59314646 | 0.600990754 | -0.221132209 | FALSE | TRUE | FALSE |
| UROD | 0.0179543 | 0.88524749 | 1 | 0.071768803 | 0.071768803 | 0.356041282 | 0.02905172 | 0.891488698 | 1 | 0.029051722 | 0.029051722 | 0.029051722 | 0.618011775 | -0.20900325 | FALSE | TRUE | FALSE |
| LRPPRC | 0.0179353 | 0.88525553 | 0.75575 | 0.158937907 | 1.445626564 | 5.349064249 | 0.0253611 | 0.892374243 | 1 | 19.32040563 | 1825.832779 | 18393.76499 | 0.707196758 | -0.150459739 | FALSE | TRUE | FALSE |
| INPP5E | 0.0179207 | 0.88526356 | 0.2576 | 0.017920659 | 0.071604504 | 0.353914739 | 2.11911832 | 0.47913524 | 0.0719 | 0.13259469 | 2.119118317 | 2.119118317 | 0.008456658 | -2.072801236 | FALSE | FALSE | FALSE |
| RCBTB1 | 0.017895 | 0.8852716 | 1 | 0.017894997 | 0.017894997 | 0.354124657 | 0.02865163 | 0.891564896 | 0.013 | 0.028651627 | 0.028651627 | 0.567707433 | 0.624571742 | -0.204417669 | FALSE | TRUE | FALSE |
| UCP2 | 0.0178692 | 0.88527963 | 0.0113 | 0.017869213 | 0.017869213 | 0.35465806 | 0.02863229 | 0.891572511 | 1 | 0.028632287 | 0.028632287 | 0.028632287 | 0.624093122 | -0.204750603 | FALSE | TRUE | FALSE |
| SLC22A6 | 0.0178122 | 0.88528766 | 0.7518 | 0.159075782 | 0.635982742 | 1.406504698 | 0.24326801 | 0.841041313 | 0.87535 | 2.241518769 | 21.18438636 | 21.18438636 | 0.073220309 | -1.135368442 | FALSE | TRUE | FALSE |
| DNAI1 | 0.0178083 | 0.88529569 | 0.51295 | 0.071187977 | 0.071187977 | 0.602641889 | 0.99414783 | 0.589005157 | 0.37885 | 0.43750746 | 3.975337341 | 3.975337341 | 0.01791316 | -1.746827793 | FALSE | TRUE | FALSE |
| LMAN1L | 0.0177794 | 0.88530372 | 1 | 0.153666463 | 0.153666463 | 0.352034667 | 0.24535412 | 0.840587911 | 0.50215 | 0.245354121 | 0.245354121 | 0.556863966 | 0.072464334 | -1.139875692 | FALSE | TRUE | FALSE |
| MEI1 | 0.0177601 | 0.88531175 | 1 | 0.017760074 | 0.017760074 | 0.347009406 | 0.02809032 | 0.891724596 | 1 | 0.028090321 | 0.028090321 | 0.55037442 | 0.63224888 | -0.199111931 | FALSE | TRUE | FALSE |
| DHX29 | 0.0177436 | 0.88531977 | 1 | 121.262246 | 11618.1365 | 117756.5044 | 0.23418288 | 0.843558074 | 0.9884 | 1822.967076 | 1939734.611 | 20709198.25 | 0.075768255 | -1.120512717 | FALSE | TRUE | FALSE |
| OGFOD2 | 0.0177354 | 0.8853278 | 0.25385 | 0.017735369 | 0.070888 | 0.088003277 | 1.1028689 | 0.578351642 | 0.5937 | 1.102868899 | 40.61064428 | 162.4192755 | 0.016081122 | -1.793683649 | FALSE | FALSE | FALSE |
| TCTN1 | 0.0177012 | 0.88533583 | 0.01625 | 0.01770115 | 0.01770115 | 0.350070347 | 0.02822205 | 0.891686612 | 0.01265 | 0.028222049 | 0.028222049 | 0.558888713 | 0.627209958 | -0.202587055 | FALSE | TRUE | FALSE |
| TANGO6 | 0.0176988 | 0.88534385 | 1 | 0.017698781 | 0.017698781 | 0.346844314 | 2.15284508 | 0.475696311 | 0.26025 | 0.236092812 | 2.152845079 | 7.969668211 | 0.008221112 | -2.085069424 | FALSE | FALSE | FALSE |
| ANXA3 | 0.0176983 | 0.88535187 | 0.00765 | 0.01769832 | 0.01769832 | 0.01769832 | 0.18914295 | 0.854604571 | 0.25295 | 0.189142954 | 1.759754795 | 1.759754795 | 0.093571131 | -1.028858121 | FALSE | TRUE | FALSE |
| CSPG4 | 0.0176907 | 0.8853599 | 0.07235 | 0.017690682 | 0.083725215 | 0.591979691 | 0.00808515 | 0.89727019 | 1 | 0.07055618 | 0.154021517 | 1.34408992 | 2.188045965 | 0.340056441 | FALSE | TRUE | FALSE |
| PAX2 | 0.0176455 | 0.88536792 | 1 | 0.070512935 | 0.070512935 | 0.349189655 | 0.11361262 | 0.87264902 | 0.4985 | 0.028427658 | 0.113612617 | 0.563273644 | 0.155312636 | -0.808793208 | FALSE | TRUE | FALSE |
| TAS2R38 | 0.0176128 | 0.88537594 | 1 | 0.017612817 | 0.017612817 | 0.017612817 | 0.02087234 | 0.893608305 | 0.00575 | 0.020872343 | 0.020872343 | 0.020872343 | 0.843835204 | -0.07374236 | FALSE | TRUE | FALSE |
| STRN3 | 0.017599 | 0.88538395 | 1 | 2.62622E+25 | 3.08629E+32 | 3.23314E+35 | 0.01564035 | 0.894953502 | 1 | 1.82823E+14 | 7.78986E+18 | 9.98889E+20 | 1.125230045 | 0.05124132 | FALSE | TRUE | FALSE |
| ABI3BP | 0.0175813 | 0.88539197 | 1 | 0.017581311 | 0.017581311 | 0.345193916 | 0.02783796 | 0.891777733 | 1 | 0.027837955 | 0.027837955 | 0.547778208 | 0.631558998 | -0.199586073 | FALSE | TRUE | FALSE |
| SKIL | 0.0175747 | 0.88539999 | 0.51 | 0.163672875 | 0.163672875 | 0.594271087 | 0.02923735 | 0.891450564 | 0.0139 | 0.029237355 | 0.029237355 | 0.579062431 | 0.60110538 | -0.221049385 | FALSE | TRUE | FALSE |
| ADAMTS18 | 0.0175581 | 0.885408 | 0.6947 | 0.03792631 | 1.34228044 | 2.957999291 | 0.01119164 | 0.89638719 | 0.2756 | 0.011191639 | 0.044733785 | 0.376346305 | 1.568861319 | 0.195584555 | FALSE | TRUE | FALSE |
| CCDC33 | 0.0175432 | 0.88541602 | 1 | 0.150751381 | 0.150751381 | 2.966806175 | 0.22687285 | 0.845556749 | 0.88 | 2.020021057 | 18.44830158 | 18.44830158 | 0.077325966 | -1.111674647 | FALSE | TRUE | FALSE |
| DNAAF1 | 0.0175183 | 0.88542403 | 1 | 0.150216146 | 0.150216146 | 2.962585878 | 0.0260998 | 0.8921107 | 1 | 2.008245838 | 18.30589969 | 18.30589969 | 0.671202956 | -0.17314614 | FALSE | TRUE | FALSE |
| CCDC172 | 0.0175047 | 0.88543205 | 0.00655 | 0.017504721 | 0.017504721 | 0.017504721 | 0.02150778 | 0.893372438 | 1 | 0.021507784 | 0.021507784 | 0.021507784 | 0.813878408 | -0.089440473 | FALSE | TRUE | FALSE |
| STYK1 | 0.0174902 | 0.88544006 | 1 | 0.279656628 | 1.118672395 | 1.386857471 | 0.10762653 | 0.873873215 | 0.8779 | 0.959658948 | 8.782508367 | 8.782508367 | 0.162508391 | -0.789124209 | FALSE | TRUE | FALSE |
| KRT39 | 0.0174885 | 0.88544807 | 0.0141 | 0.017488527 | 0.017488527 | 0.086628911 | 0.12568892 | 0.869975022 | 0.50995 | 0.50261638 | 2.010410711 | 8.043432486 | 0.139141359 | -0.856543761 | FALSE | TRUE | FALSE |
| MSANTD2 | 0.0174627 | 0.88545608 | 0.99585 | 155.2841618 | 16994.78704 | 183915.7484 | 0.02497443 | 0.892456824 | 1 | 0.024974425 | 0.024974425 | 0.49490378 | 0.699224825 | -0.155383161 | FALSE | TRUE | FALSE |
| TMEM252 | 0.017398 | 0.88546408 | 1 | 0.01739796 | 0.162507812 | 0.162507812 | 0.01947688 | 0.893952996 | 1 | 0.019476877 | 0.019476877 | 0.019476877 | 0.893262273 | -0.049021008 | FALSE | TRUE | FALSE |
| FKBP9 | 0.0172958 | 0.88547209 | 0.015 | 0.017295751 | 0.017295751 | 0.341430147 | 0.11006028 | 0.873329596 | 0.50645 | 0.110060278 | 0.110060278 | 0.544372648 | 0.157147989 | -0.803691172 | FALSE | TRUE | FALSE |
| FUS | 0.0172736 | 0.8854801 | 0.0179 | 0.017273572 | 0.017273572 | 0.341038314 | 0.02792119 | 0.891762556 | 1 | 0.027921189 | 0.027921189 | 0.552109212 | 0.618654606 | -0.208551749 | FALSE | TRUE | FALSE |
| PLEKHG4 | 0.0172465 | 0.8854881 | 0.5125 | 0.017246549 | 0.017246549 | 0.335884668 | 0.03496712 | 0.890139483 | 0.5183 | 0.034967121 | 0.034967121 | 0.683153744 | 0.493221879 | -0.306957667 | FALSE | TRUE | FALSE |
| TRPC3 | 0.0172121 | 0.88549611 | 0.7539 | 0.154241091 | 0.616663895 | 3.025986779 | 0.4501212 | 0.774309374 | 0.2679 | 0.112550943 | 0.450121198 | 2.213157804 | 0.038238833 | -1.417495367 | FALSE | TRUE | FALSE |
| ARFGEF2 | 0.0171859 | 0.88550411 | 0.05225 | 0.017185868 | 0.331726777 | 0.576099188 | 0.10967363 | 0.873450782 | 0.51825 | 0.10967363 | 0.10967363 | 2.125863258 | 0.156700096 | -0.804930737 | FALSE | TRUE | FALSE |
| ZNF302 | 0.0171823 | 0.88551211 | 0.50645 | 0.147436463 | 0.147436463 | 0.147436463 | 0.22231837 | 0.846798984 | 0.8741 | 0.222318367 | 18.03124738 | 18.03124738 | 0.077287077 | -1.111893116 | FALSE | TRUE | FALSE |
| CILP | 0.0171116 | 0.88552011 | 0.52745 | 0.143611151 | 1.236334694 | 2.797187788 | 0.03506041 | 0.890100491 | 0.75395 | 0.140230713 | 1.197252019 | 4.731573752 | 0.488060111 | -0.311526686 | FALSE | TRUE | FALSE |
| POLRMT | 0.0170972 | 0.88552812 | 0.04175 | 0.017097204 | 0.017097204 | 0.33085288 | 0.00418066 | 0.89826855 | 0.03705 | 0.004180659 | 0.004180659 | 0.081217632 | 4.089595335 | 0.611680337 | FALSE | TRUE | FALSE |
| GALNT8 | 0.0170706 | 0.88553611 | 1 | 0.148071275 | 0.592181857 | 2.310015271 | 0.23341413 | 0.843874698 | 0.7545 | 0.233414129 | 2.097452528 | 4.612324539 | 0.073134512 | -1.135877631 | FALSE | TRUE | FALSE |
| SCGB3A2 | 0.0170426 | 0.88554411 | 1 | 0.017042574 | 0.017042574 | 0.017042574 | 0.01908621 | 0.894069878 | 1 | 0.01908621 | 0.01908621 | 0.01908621 | 0.89292604 | -0.049184512 | FALSE | TRUE | FALSE |
| STRIP2 | 0.0170347 | 0.88555211 | 1 | 0.149644553 | 1.349270733 | 2.946994904 | 0.24857541 | 0.839863597 | 0.50715 | 0.248575412 | 0.248575412 | 0.947666275 | 0.068529423 | -1.164122924 | FALSE | TRUE | FALSE |
| KIF7 | 0.0170275 | 0.88556011 | 0.8157 | 0.598659118 | 12.49541946 | 49.94877833 | 0.03886521 | 0.889236337 | 0.5147 | 0.038865212 | 0.038865212 | 0.763338462 | 0.43811623 | -0.358410658 | FALSE | TRUE | FALSE |
| METTL25 | 0.0170204 | 0.8855681 | 0.0168 | 0.017020416 | 0.017020416 | 0.336661359 | 0.02671708 | 0.89200495 | 0.50665 | 0.106854889 | 0.106854889 | 0.529159716 | 0.637061289 | -0.195818784 | FALSE | TRUE | FALSE |
| PTK2B | 0.0170181 | 0.88557609 | 1 | 0.15629023 | 1.47315672 | 5.272517198 | 0.02607265 | 0.892118247 | 0.96845 | 3.838508416 | 88.00047813 | 351.8628448 | 0.652717171 | -0.185274962 | FALSE | TRUE | FALSE |
| CACHD1 | 0.0170074 | 0.88558409 | 0.0344 | 0.017007369 | 0.017007369 | 0.331506767 | 0.22910121 | 0.845128443 | 0.51605 | 0.229101205 | 0.229101205 | 4.479299748 | 0.074235178 | -1.129390244 | FALSE | TRUE | FALSE |
| GREB1 | 0.0170033 | 0.88559208 | 0.06045 | 0.017003303 | 0.32397681 | 0.570097385 | 0.02617255 | 0.892072952 | 1 | 0.235091214 | 0.940209383 | 18.01693497 | 0.64966159 | -0.187312809 | FALSE | TRUE | FALSE |
| HECTD1 | 0.0169754 | 0.88560007 | 1 | 0.154510908 | 0.323439311 | 5.136852774 | 0.02705049 | 0.891929305 | 1 | 0.247413831 | 0.989379951 | 18.95884158 | 0.627546251 | -0.20235426 | FALSE | TRUE | FALSE |
| CLN5 | 0.0169075 | 0.88560806 | 1 | 0.016907519 | 0.016907519 | 0.33510171 | 2.14749818 | 0.476560264 | 0.2588 | 0.231561926 | 2.147498182 | 2.147498182 | 0.007873124 | -2.103852932 | FALSE | FALSE | FALSE |
| MRPS30 | 0.0168788 | 0.88561605 | 1 | 0.016878849 | 0.016878849 | 0.333416994 | 0.0222361 | 0.893157841 | 1 | 0.022236096 | 0.022236096 | 0.439892868 | 0.75907429 | -0.119715718 | FALSE | TRUE | FALSE |
| GNAT1 | 0.0168633 | 0.88562404 | 0.0137 | 0.016863265 | 0.016863265 | 0.333334706 | 0.10786197 | 0.873823056 | 0.0132 | 0.107861968 | 0.107861968 | 0.534208604 | 0.156341162 | -0.805926663 | FALSE | TRUE | FALSE |
| LOXL2 | 0.0168514 | 0.88563203 | 0.51495 | 0.149826429 | 0.149826429 | 2.936408247 | 0.02787235 | 0.891770145 | 1 | 0.027872352 | 0.027872352 | 0.547582751 | 0.604593342 | -0.21853664 | FALSE | TRUE | FALSE |
| HGD | 0.016843 | 0.88564001 | 0.0153 | 0.000263541 | 0.01684298 | 0.067374036 | 9.3984E-05 | 0.900219111 | 0.50375 | 9.3984E-05 | 0.001501922 | 0.006006298 | 179.2111151 | 2.253364942 | FALSE | TRUE | FALSE |
| TUT1 | 0.0168034 | 0.88564799 | 1 | 0.016803426 | 0.016803426 | 0.329376254 | 0.02677676 | 0.891982266 | 1 | 0.02677676 | 0.02677676 | 0.526129633 | 0.627537684 | -0.20236019 | FALSE | TRUE | FALSE |
| PSEN1 | 0.0167938 | 0.88565598 | 1 | 17.15318357 | 1099.420901 | 4402.038679 | 0.02661175 | 0.892020067 | 1 | 108.823896 | 6982.545838 | 27967.93251 | 0.63106674 | -0.199924709 | FALSE | TRUE | FALSE |
| OPTN | 0.0167851 | 0.88566396 | 0.50915 | 0.016785085 | 0.016785085 | 0.038995733 | 0.00401592 | 0.898336295 | 1 | 0.034633553 | 0.034633553 | 0.079596033 | 4.179638609 | 0.621138732 | FALSE | TRUE | FALSE |
| STOML1 | 0.0167479 | 0.88567194 | 1 | 0.01674789 | 0.01674789 | 0.332160426 | 0.02705455 | 0.891921735 | 1 | 0.027054547 | 0.027054547 | 0.027054547 | 0.619041595 | -0.208280168 | FALSE | TRUE | FALSE |
| NR2C1 | 0.016723 | 0.88567992 | 0.7539 | 0.066822757 | 0.267081187 | 1.322689219 | 0.02774966 | 0.891785321 | 0.0139 | 0.02774966 | 0.02774966 | 0.549957187 | 0.602637799 | -0.219943632 | FALSE | TRUE | FALSE |
| C19orf53 | 0.0166409 | 0.8856879 | 1 | 0.016640878 | 0.016640878 | 0.016640878 | 0.017102 | 0.894549576 | 0.00295 | 0.017101999 | 0.017101999 | 0.017101999 | 0.973037024 | -0.011870635 | FALSE | TRUE | FALSE |
| PHGDH | 0.0166396 | 0.88569588 | 1 | 0.0166396 | 0.0166396 | 0.328838573 | 0.02698764 | 0.891944441 | 1 | 0.026987643 | 0.026987643 | 0.534103154 | 0.616563637 | -0.210022092 | FALSE | TRUE | FALSE |
| TTI1 | 0.0166363 | 0.88570386 | 0.5157 | 0.147209902 | 0.147209902 | 2.884696592 | 0.02751794 | 0.891823245 | 1 | 0.027517945 | 0.027517945 | 0.540548794 | 0.604560321 | -0.21856036 | FALSE | TRUE | FALSE |
| DLC1 | 0.0166342 | 0.88571183 | 1 | 0.016634239 | 0.016634239 | 0.558741743 | 0.10327869 | 0.875134557 | 0.75895 | 0.237877651 | 0.950983011 | 18.4724576 | 0.161061681 | -0.793007772 | FALSE | TRUE | FALSE |
| NAP1L5 | 0.0166162 | 0.88571981 | 1 | 0.016616226 | 0.016616226 | 0.016616226 | 0.01737431 | 0.894477146 | 1 | 0.017374307 | 0.017374307 | 0.017374307 | 0.956367669 | -0.019375114 | FALSE | TRUE | FALSE |
| MSH4 | 0.0165875 | 0.88572778 | 0.75785 | 0.016587496 | 0.1505521 | 0.326577336 | 176.459408 | 0.015221752 | 0.01325 | 2.058670826 | 75.26114326 | 705.6822308 | 9.40018E-05 | -4.026863981 | FALSE | FALSE | FALSE |
| ZBTB40 | 0.0165777 | 0.88573575 | 1 | 0.148515797 | 0.148515797 | 2.903061726 | 0.02753857 | 0.891815662 | 1 | 0.027538574 | 0.027538574 | 0.539769207 | 0.601981607 | -0.220416778 | FALSE | TRUE | FALSE |
| DPP3 | 0.0165282 | 0.88574373 | 1 | 0.016528178 | 0.016528178 | 0.324304196 | 0.02638309 | 0.892042738 | 1 | 0.02638309 | 0.02638309 | 0.518850673 | 0.626468624 | -0.203100676 | FALSE | TRUE | FALSE |
| KRR1 | 0.0164298 | 0.8857517 | 0.0096 | 0.016429787 | 0.016429787 | 0.016429787 | 0.02602218 | 0.89214088 | 1 | 0.02602218 | 0.02602218 | 0.02602218 | 0.631376256 | -0.199711755 | FALSE | TRUE | FALSE |
| FOXR1 | 0.0163977 | 0.88575967 | 0.4964 | 0.016397695 | 0.146063965 | 0.146063965 | 0.02722063 | 0.89189902 | 1 | 0.027220626 | 0.027220626 | 0.027220626 | 0.602399638 | -0.220115298 | FALSE | TRUE | FALSE |
| PPL | 0.0163269 | 0.88576764 | 0.3752 | 0.016326938 | 0.564188215 | 4.999013007 | 0.01200483 | 0.8960639 | 0.8529 | 0.402061848 | 30.43138464 | 274.6226386 | 1.360030891 | 0.133548773 | FALSE | TRUE | FALSE |
| KRT10 | 0.0163264 | 0.88577561 | 0.96905 | 13.09810203 | 127.9878971 | 1281.19403 | 0.02564502 | 0.892261443 | 1 | 20.85646426 | 2058.611676 | 21194.21744 | 0.63663093 | -0.196112266 | FALSE | TRUE | FALSE |

| | | | | | | | | | | | | | | | | | |
|---|---|---|---|---|---|---|---|---|---|---|---|---|---|---|---|---|---|
| HTR3A | 0.0162427 | 0.88578357 | 1 | 0.064913705 | 0.064913705 | 0.3209918 | 2.04142125 | 0.485556071 | 0.01005 | 0.412386691 | 1.649077614 | 2.041421246 | 0.007956577 | -2.099273733 | FALSE | FALSE | FALSE |
| FKBP11 | 0.0162317 | 0.88579154 | 1 | 0.016231723 | 0.016231723 | 0.016231723 | 0.02585191 | 0.892193656 | 1 | 0.025851911 | 0.025851911 | 0.025851911 | 0.627873236 | -0.202128029 | FALSE | TRUE | FALSE |
| SYCE1 | 0.0162295 | 0.88579951 | 0.50095 | 0.016229529 | 0.016229529 | 0.016229529 | 0.00211886 | 0.899029237 | 1 | 0.019323074 | 0.18086155 | 0.18086155 | 7.659552235 | 0.884203382 | FALSE | TRUE | FALSE |
| SUCLG2 | 0.0162271 | 0.88580747 | 0.5058 | 0.064871209 | 0.2594011 | 0.2594011 | 0.25831912 | 0.837195084 | 0.63025 | 0.258319122 | 9.354766871 | 9.354766871 | 0.062818006 | -1.201915854 | FALSE | TRUE | FALSE |
| TRIM37 | 0.0161956 | 0.88581543 | 1 | 0.016195619 | 0.016195619 | 0.317925376 | 0.02598007 | 0.892171043 | 1 | 0.025980065 | 0.025980065 | 0.511132353 | 0.623386401 | -0.205242676 | FALSE | TRUE | FALSE |
| TMTC2 | 0.0161697 | 0.8858234 | 0.7517 | 0.146943226 | 0.587366056 | 2.89260883 | 3.62901809 | 0.376143504 | 0.29345 | 2.126395865 | 33.98878662 | 135.9390549 | 0.004455668 | -2.351087223 | FALSE | FALSE | FALSE |
| MYADM | 0.0161509 | 0.88583136 | 1 | 0.016150896 | 0.016150896 | 0.319826504 | 0.01771716 | 0.89439011 | 1 | 0.017717162 | 0.017717162 | 0.351259286 | 0.911596109 | -0.040197537 | FALSE | TRUE | FALSE |
| C19orf44 | 0.0161331 | 0.88583932 | 0.93775 | 1.216445796 | 101.536901 | 101.536901 | 1.92317678 | 0.496844768 | 0.49815 | 1.923176776 | 163.9461514 | 1570.065476 | 0.008388763 | -2.076302076 | FALSE | FALSE | FALSE |
| STOX1 | 0.0161304 | 0.88584728 | 0.9389 | 0.296424056 | 18.95495266 | 75.83612259 | 0.03734221 | 0.889599191 | 0.6954 | 0.080100641 | 1.280816984 | 1.280816984 | 0.431962821 | -0.364553631 | FALSE | TRUE | FALSE |
| TMEM79 | 0.01606 | 0.88585523 | 1 | 5.57638E+19 | 3.05328E+25 | 6.75995E+27 | 0.0495785 | 0.886758339 | 1 | 409273164.3 | 1.23506E+13 | 1.49284E+14 | 0.323931494 | -0.489546826 | FALSE | TRUE | FALSE |
| ZNF443 | 0.016055 | 0.88586319 | 0.50705 | 0.139673946 | 0.139673946 | 0.542923071 | 0.21945746 | 0.84727616 | 0.25505 | 0.219457456 | 1.977241932 | 4.343544667 | 0.073157547 | -1.135740863 | FALSE | TRUE | FALSE |
| COMMD6 | 0.0160269 | 0.88587115 | 0.0028 | 0.016026857 | 0.016026857 | 0.016026857 | 0.01672533 | 0.894672494 | 1 | 0.016725326 | 0.016725326 | 0.016725326 | 0.958238874 | -0.018526215 | FALSE | TRUE | FALSE |
| C8B | 0.0160212 | 0.8858791 | 0.81365 | 0.137887916 | 4.870731171 | 4.870731171 | 0.00785052 | 0.897366691 | 1 | 0.068369986 | 0.273341374 | 0.621378445 | 2.040783155 | 0.309796861 | FALSE | TRUE | FALSE |
| UGGT1 | 0.0160031 | 0.88588706 | 1 | 0.016003129 | 0.016003129 | 0.537638786 | 0.02577281 | 0.892201192 | 1 | 0.025772813 | 0.025772813 | 0.506683191 | 0.620930625 | -0.20695692 | FALSE | TRUE | FALSE |
| LDHD | 0.0159675 | 0.88589501 | 0.93695 | 0.142321714 | 2.274395067 | 2.274395067 | 9.44607469 | 0.206064744 | 0.3057 | 2.36296956 | 89.11834983 | 356.3435633 | 0.001690389 | -2.772013374 | FALSE | FALSE | FALSE |
| SDAD1 | 0.0159448 | 0.88590296 | 0.0182 | 0.015944831 | 0.015944831 | 0.315027738 | 0.025687 | 0.892238857 | 1 | 0.025687003 | 0.025687003 | 0.508248163 | 0.620735335 | -0.207093532 | FALSE | TRUE | FALSE |
| YY1AP1 | 0.0159154 | 0.88591091 | 1 | 0.015915365 | 0.015915365 | 0.313597415 | 0.02538011 | 0.89236673 | 0.01765 | 0.025380111 | 0.025380111 | 0.500982017 | 0.627080196 | -0.202676915 | FALSE | TRUE | FALSE |
| HELQ | 0.015913 | 0.88591886 | 0.87975 | 0.139277383 | 1.25119104 | 2.731812878 | 0.00785808 | 0.897359804 | 0.514 | 0.069470631 | 0.069470631 | 1.36577546 | 2.025045432 | 0.306434771 | FALSE | TRUE | FALSE |
| EEA1 | 0.0159038 | 0.88592681 | 1 | 9.89612E+14 | 1.29308E+20 | 8.29942E+21 | 0.20341467 | 0.851239686 | 1 | 1.95114E+17 | 6.23924E+22 | 1.23411E+25 | 0.07818391 | -1.106882617 | FALSE | TRUE | FALSE |
| DISC1 | 0.0158985 | 0.88593476 | 1 | 0.137450108 | 0.137450108 | 2.703824145 | 0.02545967 | 0.892321631 | 0.5078 | 0.222433945 | 0.222433945 | 0.860753223 | 0.624456643 | -0.20449771 | FALSE | TRUE | FALSE |
| DMP1 | 0.0158932 | 0.8859427 | 1 | 0.015893171 | 0.015893171 | 0.315054275 | 0.01924525 | 0.894018771 | 1 | 0.175145029 | 0.175145029 | 0.381908971 | 0.825822839 | -0.083113111 | FALSE | TRUE | FALSE |
| TYK2 | 0.0158773 | 0.88595065 | 1 | 0.015877314 | 0.015877314 | 0.308074407 | 0.10226321 | 0.875438827 | 0.0372 | 0.102263207 | 0.102263207 | 1.991393881 | 0.155259304 | -0.808942365 | FALSE | TRUE | FALSE |
| AGRP | 0.0158654 | 0.88595859 | 1 | 0.147600036 | 0.147600036 | 0.147600036 | 0.01802589 | 0.894332016 | 1 | 0.018025889 | 0.018025889 | 0.018025889 | 0.880145579 | -0.055445488 | FALSE | TRUE | FALSE |
| TCL1B | 0.0158286 | 0.88596654 | 1 | 0.015828579 | 0.015828579 | 0.015828579 | 0.15653598 | 0.862849548 | 0.5042 | 0.156535981 | 0.156535981 | 0.156535981 | 0.101117831 | -0.995172253 | FALSE | TRUE | FALSE |
| KLHDC4 | 0.0158211 | 0.88597448 | 0.9703 | 1.223008882 | 44.62682934 | 417.3400436 | 0.00445714 | 0.898221079 | 1 | 0.004457138 | 0.004457138 | 0.08812725 | 3.549610898 | 0.550180749 | FALSE | TRUE | FALSE |
| FASTKD1 | 0.0157921 | 0.88598242 | 0.0226 | 0.015792075 | 0.015792075 | 0.311196205 | 0.02449777 | 0.892621638 | 0.5057 | 0.212020549 | 0.212020549 | 4.185440295 | 0.644633179 | -0.190687346 | FALSE | TRUE | FALSE |
| TTC12 | 0.015791 | 0.88599036 | 0.01785 | 0.015790973 | 0.015790973 | 0.311749464 | 1.84447003 | 0.504579438 | 0.1372 | 1.844470027 | 17.06553867 | 17.06553867 | 0.008561252 | -2.067462706 | FALSE | FALSE | FALSE |
| SLC1A7 | 0.0157003 | 0.8859983 | 1 | 0.015700284 | 0.015700284 | 0.309246072 | 0.02538702 | 0.892359216 | 0.0157 | 0.025387017 | 0.025387017 | 0.500956458 | 0.618437526 | -0.208704166 | FALSE | TRUE | FALSE |
| CEP76 | 0.0156946 | 0.88600624 | 0.01995 | 0.01569458 | 0.01569458 | 0.309849036 | 0.02470657 | 0.892539291 | 1 | 0.222387011 | 0.222387011 | 0.830065399 | 0.635239173 | -0.197062729 | FALSE | TRUE | FALSE |
| PCCA | 0.0156935 | 0.88601418 | 0.99025 | 16.01848837 | 2245.124529 | 16452.59249 | 0.0014398 | 0.89924939 | 0.9943 | 1.469289822 | 23.5173626 | 94.12213303 | 10.89974108 | 1.037416182 | FALSE | TRUE | FALSE |
| TTC14 | 0.01568 | 0.88602211 | 1 | 0.01567998 | 0.01567998 | 0.31092808 | 0.02513134 | 0.892419301 | 0.01055 | 0.02513134 | 0.02513134 | 0.498855565 | 0.623921354 | -0.20487015 | FALSE | TRUE | FALSE |
| LMF1 | 0.0156606 | 0.88603005 | 1 | 0.135794887 | 1.208579435 | 2.680431241 | 0.22031272 | 0.847102954 | 0.75435 | 0.220312719 | 1.980815226 | 4.355547423 | 0.071083577 | -1.148230727 | FALSE | TRUE | FALSE |
| AACS | 0.0156323 | 0.88603798 | 0.5111 | 0.137674556 | 0.137674556 | 0.529124218 | 0.02600465 | 0.892163504 | 1 | 0.026004654 | 0.026004654 | 0.513371927 | 0.601134325 | -0.221028473 | FALSE | TRUE | FALSE |
| NBEA | 0.0155773 | 0.88604591 | 1 | 0.015577347 | 0.296033044 | 0.517538957 | 0.02541147 | 0.892344185 | 0.06825 | 0.025411466 | 0.485827954 | 0.846587938 | 0.613004655 | -0.212536227 | FALSE | TRUE | FALSE |
| BRIP1 | 0.0155721 | 0.88605385 | 0.59145 | 0.015572107 | 1.114261543 | 4.453937978 | 0.01146581 | 0.896267931 | 0.7812 | 0.389715198 | 30.346009 | 149.0437831 | 1.358133637 | 0.132942506 | FALSE | TRUE | FALSE |
| UGT2B10 | 0.0155599 | 0.88606178 | 0.01265 | 0.015559872 | 0.015559872 | 0.037782991 | 0.26218256 | 0.83607652 | 0.25405 | 0.030794261 | 0.26218256 | 0.26218256 | 0.05934747 | -1.226597788 | FALSE | TRUE | FALSE |
| SMARCAL1 | 0.0155409 | 0.88606971 | 1 | 4307171152 | 2.1843E+13 | 9.27995E+14 | 0.01259412 | 0.895866194 | 0.99955 | 457.8338887 | 66886.91189 | 625297.5803 | 1.233978794 | 0.091307697 | FALSE | TRUE | FALSE |
| HPS5 | 0.0155344 | 0.88607764 | 0.997 | 23.96163053 | 2181.176049 | 17716.0784 | 5.07826484 | 0.303914912 | 0.77825 | 47.297234 | 4408.857838 | 35471.9169 | 0.003059004 | -2.514420013 | FALSE | FALSE | FALSE |
| LMNB1 | 0.0155147 | 0.88608556 | 0.0128 | 0.015514669 | 0.015514669 | 0.307473165 | 0.02540227 | 0.892351701 | 1 | 0.025402266 | 0.025402266 | 0.503978089 | 0.610759263 | -0.214129938 | FALSE | TRUE | FALSE |
| LRRC36 | 0.0155012 | 0.88609349 | 0.0196 | 0.015501211 | 0.015501211 | 0.306094953 | 0.0233784 | 0.892890337 | 1 | 0.201360639 | 1.780120662 | 3.982235434 | 0.663057058 | -0.178449098 | FALSE | TRUE | FALSE |
| IL7 | 0.0154982 | 0.88610142 | 0.004 | 0.01549822 | 0.01549822 | 0.01549822 | 0.01583284 | 0.89489597 | 0.00265 | 0.015832843 | 0.015832843 | 0.015832843 | 0.978865254 | -0.009277087 | FALSE | TRUE | FALSE |
| PRCD | 0.0154879 | 0.88610934 | 0.00195 | 0.015487872 | 0.015487872 | 0.015487872 | 0.01668696 | 0.894679716 | 0.00165 | 0.016686961 | 0.016686961 | 0.016686961 | 0.928142125 | -0.032385516 | FALSE | TRUE | FALSE |
| UCMA | 0.0154855 | 0.88611726 | 1 | 0.015485455 | 0.015485455 | 0.015485455 | 0.01811579 | 0.894273864 | 1 | 0.018115788 | 0.018115788 | 0.018115788 | 0.854804418 | -0.068133242 | FALSE | TRUE | FALSE |
| CRAT | 0.0154815 | 0.88612519 | 1 | 1.20079E+14 | 1.64421E+19 | 2.98931E+21 | 0.0270663 | 0.891914164 | 1 | 2.28233E+19 | 3.47283E+25 | 1.41325E+28 | 0.571985507 | -0.242614975 | FALSE | TRUE | FALSE |
| PRDM15 | 0.0154364 | 0.88613311 | 0.52245 | 0.134947901 | 0.134947901 | 2.615448273 | 0.02564375 | 0.89226897 | 0.03715 | 0.025643754 | 0.025643754 | 0.498860643 | 0.60195436 | -0.220436436 | FALSE | TRUE | FALSE |
| IL13 | 0.0154227 | 0.88614103 | 1 | 0.015422714 | 0.015422714 | 0.015422714 | 0.01663417 | 0.894701378 | 0.0036 | 0.016634172 | 0.016634172 | 0.016634172 | 0.927170527 | -0.032840382 | FALSE | TRUE | FALSE |
| BCL2L10 | 0.0153662 | 0.88614895 | 0.007 | 0.015366214 | 0.015366214 | 0.015366214 | 0.01738179 | 0.894469899 | 1 | 0.017381787 | 0.017381787 | 0.017381787 | 0.884041104 | -0.053527542 | FALSE | TRUE | FALSE |
| SLC12A2 | 0.0153168 | 0.88615687 | 0.6953 | 0.035592932 | 0.567863738 | 1.199642653 | 0.17655428 | 0.857964007 | 0.0236 | 0.035948208 | 0.14363268 | 0.705288866 | 0.086754255 | -1.061709215 | FALSE | TRUE | FALSE |
| TELO2 | 0.0153114 | 0.88616478 | 1 | 0.015311448 | 0.015311448 | 0.299264647 | 0.02480194 | 0.892494323 | 1 | 0.024801939 | 0.024801939 | 0.486085927 | 0.617348842 | -0.209469362 | FALSE | TRUE | FALSE |
| SERINC4 | 0.0152835 | 0.8861727 | 1 | 0.015283528 | 0.015283528 | 0.015283528 | 15.0955458 | 0.157901238 | 0.128 | 1.6774281 | 15.09554584 | 15.09554584 | 0.001012453 | -2.994625205 | FALSE | FALSE | FALSE |
| CEP192 | 0.0152523 | 0.88618062 | 0.5257 | 0.015252344 | 0.035370141 | 0.294350697 | 0.00372499 | 0.898430999 | 1 | 0.031453114 | 0.031453114 | 0.609572763 | 4.094597655 | 0.612211233 | FALSE | TRUE | FALSE |
| HAUS6 | 0.015248 | 0.88618853 | 1 | 0.060981586 | 0.060981586 | 1.19931792 | 15.9466955 | 0.154029149 | 0.07895 | 1.733844248 | 150.3435122 | 150.3435122 | 0.000956188 | -3.01945687 | FALSE | FALSE | FALSE |
| SCGB1C1 | 0.0152472 | 0.88619644 | 1 | 0.015247239 | 0.015247239 | 0.015247239 | 0.17763771 | 0.857612146 | 0.75105 | 0.17763771 | 1.728367414 | 1.728367414 | 0.085833346 | -1.066343958 | FALSE | TRUE | FALSE |
| TBATA | 0.015245 | 0.88620436 | 1 | 0.015245009 | 0.015245009 | 0.302574787 | 0.01825307 | 0.89426659 | 1 | 0.018253074 | 0.018253074 | 0.018253074 | 0.835202304 | -0.078208317 | FALSE | TRUE | FALSE |
| CILP2 | 0.0152021 | 0.88621227 | 1 | 0.015202096 | 0.015202096 | 0.292862298 | 0.09705136 | 0.876709456 | 0.03985 | 0.097051358 | 0.097051358 | 1.877951829 | 0.156639705 | -0.805098142 | FALSE | TRUE | FALSE |
| TBC1D26 | 0.0151393 | 0.88622018 | 1 | 0.015139259 | 0.015139259 | 0.015139259 | 0.15442409 | 0.863456728 | 0.0069 | 0.154424093 | 0.154424093 | 0.154424093 | 0.098036896 | -1.008610447 | FALSE | TRUE | FALSE |
| TUBGCP4 | 0.0150774 | 0.88622809 | 0.02005 | 0.015077357 | 0.015077357 | 0.29763798 | 0.0201554 | 0.893799236 | 1 | 0.020155403 | 0.020155403 | 0.398502991 | 0.748055309 | -0.126066291 | FALSE | TRUE | FALSE |
| TLN1 | 0.0150664 | 0.886236 | 0.8815 | 0.060216946 | 0.560627714 | 10.62326448 | 0.12007678 | 0.871226025 | 0.30095 | 0.030039774 | 0.251125451 | 2.289778346 | 0.125473378 | -0.901448408 | FALSE | TRUE | FALSE |
| FRYL | 0.0149101 | 0.8862439 | 1 | 113.0701595 | 11410.18785 | 118684.4589 | 0.02828046 | 0.891656207 | 0.76335 | 0.256715062 | 4.90066375 | 45.65810502 | 0.527224437 | -0.278004468 | FALSE | TRUE | FALSE |
| PTPDC1 | 0.0148919 | 0.88625181 | 1 | 0.014891882 | 0.014891882 | 0.293592412 | 0.02270894 | 0.893024241 | 0.7531 | 0.198746074 | 1.785296646 | 3.924986426 | 0.655771789 | -0.183247271 | FALSE | TRUE | FALSE |
| MTMR14 | 0.0148233 | 0.88625972 | 0.26415 | 0.003709398 | 0.014823308 | 0.073119966 | 0.00206667 | 0.89906933 | 0.8804 | 0.033003949 | 0.131940046 | 0.163060231 | 7.172574294 | 0.855675055 | FALSE | TRUE | FALSE |
| ZNF484 | 0.0148129 | 0.88626762 | 0.9918 | 96.71304836 | 9057.336418 | 18243.96168 | 0.2356612 | 0.843118744 | 0.8721 | 0.530862874 | 19.58798606 | 19.58798606 | 0.062856631 | -1.201648898 | FALSE | TRUE | FALSE |
| DMBX1 | 0.0147513 | 0.88627552 | 1 | 0.01475129 | 0.01475129 | 0.29209742 | 0.02430676 | 0.892696396 | 1 | 0.02430676 | 0.02430676 | 0.48188455 | 0.606880161 | -0.216897059 | FALSE | TRUE | FALSE |
| TUBA3D | 0.0147167 | 0.88628343 | 1 | 0.058823646 | 0.058823646 | 0.290783285 | 0.3769192 | 0.80026638 | 0.50675 | 0.376919199 | 1.507442219 | 1.865317631 | 0.039044758 | -1.408437261 | FALSE | TRUE | FALSE |
| CCDC122 | 0.0147158 | 0.88629133 | 1 | 0.014715847 | 0.014715847 | 0.014715847 | 0.15574082 | 0.863048626 | 0.5038 | 0.155740823 | 0.155740823 | 0.155740823 | 0.094489339 | -1.024617188 | FALSE | TRUE | FALSE |
| ZNF106 | 0.0146668 | 0.88629923 | 0.0513 | 0.014666808 | 0.283209882 | 0.490403962 | 0.20236364 | 0.851567832 | 0.2827 | 0.202363642 | 1.862600725 | 36.11582578 | 0.072477487 | -1.139796876 | FALSE | TRUE | FALSE |
| ACTR6 | 0.0146614 | 0.88630713 | 1 | 0.234229797 | 0.936566317 | 0.936566317 | 0.02463026 | 0.892561761 | 0.4995 | 0.024630259 | 0.098445617 | 0.098445617 | 0.595259052 | -0.225293992 | FALSE | TRUE | FALSE |
| SEC24A | 0.0146285 | 0.88631503 | 0.0282 | 0.014628506 | 0.014628506 | 0.28690604 | 0.10924825 | 0.873612013 | 0.3288 | 0.109248249 | 1.746786599 | 2.147660016 | 0.133901515 | -0.873214509 | FALSE | TRUE | FALSE |
| SLC24A5 | 0.0145942 | 0.88632293 | 0.7503 | 0.05834869 | 0.233339944 | 0.289303806 | 0.02432605 | 0.892688925 | 1 | 0.02432605 | 0.02432605 | 0.482731569 | 0.599943153 | -0.221889899 | FALSE | TRUE | FALSE |
| AASS | 0.0145277 | 0.88633082 | 0.8756 | 0.058076587 | 0.512312962 | 1.141614279 | 0.00706147 | 0.897511138 | 1 | 0.062811865 | 0.251061869 | 0.954612201 | 2.05732548 | 0.313303005 | FALSE | TRUE | FALSE |
| SLC8A3 | 0.0145203 | 0.88633872 | 1 | 0.058027859 | 0.058027859 | 1.138172642 | 0.36665783 | 0.803543644 | 0.5084 | 0.366657826 | 3.403154199 | 7.208420611 | 0.039601682 | -1.402286371 | FALSE | TRUE | FALSE |
| COL8A2 | 0.0144962 | 0.88634662 | 0.75585 | 0.134533819 | 0.537589845 | 1.961355083 | 0.0963639 | 0.87683423 | 0.5049 | 0.38520435 | 1.540197962 | 1.906915618 | 0.150431361 | -0.822661616 | FALSE | TRUE | FALSE |
| VPS41 | 0.0144913 | 0.88635451 | 0.5119 | 0.125285099 | 0.125285099 | 2.467477926 | 0.81067839 | 0.617151928 | 0.2518 | 0.20269819 | 0.810678393 | 3.135058477 | 0.017875471 | -1.747742502 | FALSE | TRUE | FALSE |

| | | | | | | | | | | | | | | | | | |
|---|---|---|---|---|---|---|---|---|---|---|---|---|---|---|---|---|---|
| SP110 | 0.0144863 | 0.8863624 | 1 | 0.014486339 | 0.014486339 | 0.285759698 | 0.02286576 | 0.892979639 | 0.5096 | 0.201416342 | 0.201416342 | 0.773185695 | 0.633538512 | -0.19822698 | FALSE | TRUE | FALSE |
| SURF6 | 0.0144719 | 0.88637029 | 1 | 0.014471938 | 0.014471938 | 0.285971934 | 0.02174857 | 0.893313318 | 1 | 0.021748568 | 0.021748568 | 0.430380708 | 0.665420299 | -0.176903955 | FALSE | TRUE | FALSE |
| LDLRAD3 | 0.0144597 | 0.88637819 | 1 | 0.014459737 | 0.014459737 | 0.286809444 | 0.02359112 | 0.892823276 | 0.5015 | 0.09427286 | 0.09427286 | 0.09427286 | 0.612931205 | -0.212588267 | FALSE | TRUE | FALSE |
| ZNF831 | 0.0144587 | 0.88638608 | 0.51825 | 0.129484923 | 0.278231811 | 2.491710889 | 0.02025687 | 0.893762566 | 0.7623 | 0.182463674 | 1.686868518 | 32.60907406 | 0.71376637 | -0.146443918 | FALSE | TRUE | FALSE |
| ADAM12 | 0.0143613 | 0.88639396 | 0.0267 | 0.014361336 | 0.014361336 | 0.281757211 | 0.02358092 | 0.892845638 | 0.02135 | 0.023580921 | 0.023580921 | 0.463698515 | 0.609023532 | -0.215365927 | FALSE | TRUE | FALSE |
| TRMT1 | 0.014351 | 0.88640185 | 1 | 0.12573688 | 0.502790924 | 2.468963653 | 0.02363783 | 0.892808362 | 1 | 0.208988096 | 0.208988096 | 4.112674765 | 0.607119831 | -0.216725581 | FALSE | TRUE | FALSE |
| BBS12 | 0.0143441 | 0.88640974 | 1 | 0.014344103 | 0.014344103 | 0.283376739 | 0.19661504 | 0.852933708 | 0.259 | 0.19661504 | 1.796147353 | 3.889963776 | 0.072955268 | -1.136943343 | FALSE | TRUE | FALSE |
| PRF1 | 0.0143114 | 0.88641762 | 0.0181 | 0.014311357 | 0.014311357 | 0.121323876 | 0.0070179 | 0.897531744 | 1 | 0.007017898 | 0.007017898 | 0.138536049 | 2.039265537 | 0.30947378 | FALSE | TRUE | FALSE |
| CAPRIN1 | 0.01431 | 0.88642551 | 1 | 0.01430997 | 0.01430997 | 0.281975793 | 0.01676359 | 0.894658047 | 1 | 0.016763586 | 0.016763586 | 0.330910637 | 0.853634196 | -0.068728196 | FALSE | TRUE | FALSE |
| FRMD3 | 0.0143016 | 0.88643339 | 0.87845 | 0.228468114 | 0.913500136 | 1.131008045 | 0.02438827 | 0.892659029 | 0.0134 | 0.024388271 | 0.024388271 | 0.483222922 | 0.586413792 | -0.231795824 | FALSE | TRUE | FALSE |
| MIB1 | 0.0142994 | 0.88644128 | 0.78395 | 0.113916641 | 7.771607887 | 31.31698452 | 2.92053606 | 0.417721962 | 0.3786 | 0.730009379 | 95.66314418 | 802.2587842 | 0.004896172 | -2.310143375 | FALSE | FALSE | FALSE |
| TMPRSS11A | 0.0142421 | 0.88644916 | 0.011 | 0.014242142 | 0.014242142 | 0.070713789 | 0.00679126 | 0.897593517 | 1 | 0.027150179 | 0.027150179 | 0.027150179 | 2.097129049 | 0.321625156 | FALSE | TRUE | FALSE |
| MAMDC4 | 0.0142178 | 0.88645704 | 0.99575 | 560.8024117 | 420492.1518 | 3984720.4 | 0.41720823 | 0.785694434 | 0.73575 | 29.84331205 | 2341.806378 | 21449.95667 | 0.034078366 | -1.467521242 | FALSE | TRUE | FALSE |
| KAT6A | 0.0142051 | 0.88646492 | 0.7639 | 0.133504805 | 1.287760894 | 24.73015293 | 0.02503151 | 0.892441817 | 1 | 0.025031514 | 0.48301473 | 0.837954422 | 0.56748872 | -0.246042767 | FALSE | TRUE | FALSE |
| INPP5K | 0.014203 | 0.8864728 | 1 | 0.014203007 | 0.014203007 | 0.281705088 | 0.02343898 | 0.892867991 | 0.00975 | 0.023438982 | 0.023438982 | 0.023438982 | 0.605956672 | -0.217558428 | FALSE | TRUE | FALSE |
| IGSF9 | 0.0141369 | 0.88648067 | 1 | 0.014136945 | 0.014136945 | 0.475963459 | 0.02305802 | 0.892927562 | 1 | 0.023058022 | 0.023058022 | 0.453347289 | 0.613103106 | -0.212466484 | FALSE | TRUE | FALSE |
| PILRB | 0.0141339 | 0.88648855 | 1 | 0.125322325 | 0.125322325 | 0.125322325 | 0.01940789 | 0.893989546 | 0.00545 | 0.019407895 | 0.019407895 | 0.019407895 | 0.728255795 | -0.137716051 | FALSE | TRUE | FALSE |
| IFT88 | 0.0141159 | 0.88649643 | 1 | 0.056428608 | 0.056428608 | 1.112900787 | 0.09280582 | 0.877597021 | 0.5135 | 0.092805818 | 0.092805818 | 0.784521772 | 0.152101367 | -0.817866884 | FALSE | TRUE | FALSE |
| HARS2 | 0.0141025 | 0.8865043 | 1 | 0.056370279 | 0.056370279 | 0.279450607 | 0.02269117 | 0.893031671 | 1 | 0.3626919 | 1.450578115 | 1.45406653 | 0.621498389 | -0.206559993 | FALSE | TRUE | FALSE |
| CCDC39 | 0.0141015 | 0.88651217 | 0.01985 | 0.014101533 | 0.014101533 | 0.278212212 | 0.19780835 | 0.852597872 | 0.5094 | 0.197808349 | 0.197808349 | 0.755713833 | 0.071288866 | -1.146978296 | FALSE | TRUE | FALSE |
| TBX19 | 0.0140908 | 0.88652005 | 1 | 0.126011361 | 1.156621031 | 1.156621031 | 0.0971338 | 0.876671018 | 0.5037 | 0.0971338 | 0.0971338 | 0.481873844 | 0.14506542 | -0.838436101 | FALSE | TRUE | FALSE |
| KRT80 | 0.0140306 | 0.88652792 | 1 | 0.014030618 | 0.014030618 | 0.277741222 | 0.08884463 | 0.878360655 | 0.74835 | 0.204220381 | 0.816233321 | 0.816233321 | 0.157923082 | -0.801554389 | FALSE | TRUE | FALSE |
| PLEKHG1 | 0.0140228 | 0.88653579 | 0.51975 | 0.014022787 | 0.014022787 | 0.272524795 | 0.00348201 | 0.898532285 | 1 | 0.003482012 | 0.003482012 | 0.067901601 | 4.027207934 | 0.605004054 | FALSE | TRUE | FALSE |
| MADD | 0.0139998 | 0.88654366 | 0.0523 | 0.013999764 | 0.269763975 | 0.469571596 | 0.0215141 | 0.893365051 | 0.76205 | 0.188718831 | 1.699091079 | 32.88446111 | 0.650725015 | -0.186602498 | FALSE | TRUE | FALSE |
| RBL2 | 0.0139786 | 0.88655152 | 0.73335 | 0.055843656 | 2.055590771 | 8.216047276 | 0.52311194 | 0.745469873 | 0.35335 | 0.130884429 | 2.091268355 | 8.362446494 | 0.02672206 | -1.573130072 | FALSE | TRUE | FALSE |
| COL11A1 | 0.0138676 | 0.88655939 | 0.219 | 0.001530154 | 0.055342714 | 0.268415869 | 0.00165568 | 0.899182769 | 0.24135 | 0.001094423 | 0.01612211 | 0.040795054 | 8.375792616 | 0.923025916 | FALSE | TRUE | FALSE |
| EML5 | 0.0138121 | 0.88656726 | 0.049 | 0.013812138 | 0.013812138 | 0.462222334 | 0.02198479 | 0.893224526 | 1 | 0.195351504 | 0.195351504 | 3.794261492 | 0.628258722 | -0.201861474 | FALSE | TRUE | FALSE |
| MCM7 | 0.013787 | 0.88657512 | 1 | 0.117183342 | 0.117183342 | 0.46660786 | 0.0229374 | 0.892964765 | 1 | 0.0229374 | 0.0229374 | 0.453207784 | 0.601069132 | -0.221075575 | FALSE | TRUE | FALSE |
| DDX20 | 0.0137709 | 0.88658299 | 0.0215 | 0.01377091 | 0.01377091 | 0.271258462 | 0.02245921 | 0.893083657 | 1 | 0.022459213 | 0.022459213 | 0.44320432 | 0.613151915 | -0.212431911 | FALSE | TRUE | FALSE |
| LAMA4 | 0.0137467 | 0.88659085 | 0.165 | 0.013746706 | 0.121156492 | 0.461224588 | 0.00055401 | 0.899739807 | 1 | 0.000554014 | 0.000554014 | 0.018617575 | 24.81292418 | 1.394677949 | FALSE | TRUE | FALSE |
| RASA3 | 0.0137445 | 0.88659872 | 1 | 0.013744469 | 0.013744469 | 0.269424555 | 0.02271227 | 0.89301681 | 1 | 0.022712275 | 0.022712275 | 0.446281005 | 0.605155962 | -0.218132684 | FALSE | TRUE | FALSE |
| THBS3 | 0.0137401 | 0.88660658 | 0.13065 | 0.0008608 | 0.013740051 | 0.067317541 | 0.01187702 | 0.896127301 | 0.669 | 0.025219206 | 0.877533349 | 3.507248937 | 1.156860249 | 0.063280899 | FALSE | TRUE | FALSE |
| EPB41L4B | 0.0137224 | 0.88661444 | 1 | 0.054833059 | 0.054833059 | 1.07871299 | 0.02304664 | 0.892935005 | 1 | 0.023046636 | 0.023046636 | 0.454282266 | 0.595419124 | -0.22517722 | FALSE | TRUE | FALSE |
| PDE9A | 0.0136998 | 0.8866223 | 1 | 0.013699762 | 0.013699762 | 0.270156133 | 0.02256563 | 0.893053956 | 0.01735 | 0.022565633 | 0.022565633 | 0.445739416 | 0.607107371 | -0.216734494 | FALSE | TRUE | FALSE |
| EDIL3 | 0.0136916 | 0.88663016 | 1 | 0.218625262 | 2.04385752 | 2.04385752 | 0.02400438 | 0.892726273 | 1 | 0.024004376 | 0.024004376 | 0.476303058 | 0.570378304 | -0.243837002 | FALSE | TRUE | FALSE |
| TTC16 | 0.0136321 | 0.88663801 | 0.5125 | 0.114287931 | 0.982852743 | 2.241471855 | 0.23763806 | 0.842677006 | 0.5061 | 0.237638058 | 8.30830612 | 8.30830612 | 0.057364934 | -1.241353499 | FALSE | TRUE | FALSE |
| LRWD1 | 0.0136228 | 0.88664587 | 1 | 0.013622755 | 0.013622755 | 0.268545463 | 0.0222207 | 0.893165254 | 1 | 0.022220698 | 0.022220698 | 0.438793274 | 0.613066014 | -0.212492759 | FALSE | TRUE | FALSE |
| CAPN2 | 0.0136159 | 0.88665372 | 0.15055 | 0.000848715 | 0.218657355 | 3.514914844 | 0.038878 | 0.889228424 | 0.3424 | 0.009704467 | 1.645285277 | 21.52064991 | 0.350221311 | -0.455657431 | FALSE | TRUE | FALSE |
| PRKAG2 | 0.0135876 | 0.88666158 | 0.0181 | 0.013587644 | 0.013587644 | 0.268141293 | 0.02234782 | 0.893120761 | 1 | 0.089301788 | 0.089301788 | 0.441721573 | 0.608007713 | -0.216090911 | FALSE | TRUE | FALSE |
| BFSP2 | 0.0135721 | 0.88666943 | 0.01405 | 0.013572054 | 0.013572054 | 0.268754959 | 0.08774982 | 0.878650606 | 0.7553 | 0.087749824 | 0.350851856 | 0.43521599 | 0.154667595 | -0.810600668 | FALSE | TRUE | FALSE |
| TMEM213 | 0.0135667 | 0.88667728 | 1 | 0.013566695 | 0.013566695 | 0.013566695 | 0.01407101 | 0.895411693 | 0.0032 | 0.01407101 | 0.01407101 | 0.01407101 | 0.964159284 | -0.015851212 | FALSE | TRUE | FALSE |
| DENND4C | 0.0135327 | 0.88668514 | 0.99875 | 952.4335899 | 1028994.151 | 11041850.26 | 0.02826426 | 0.891663809 | 1 | 0.254152419 | 2.345590351 | 8.548578762 | 0.478792247 | -0.31985289 | FALSE | TRUE | FALSE |
| INPP5F | 0.0135286 | 0.88669299 | 0.75745 | 0.116908913 | 1.036957499 | 3.933892661 | 0.02145127 | 0.893401975 | 0.87635 | 1.676389235 | 15.3998555 | 32.94465402 | 0.630666248 | -0.200200411 | FALSE | TRUE | FALSE |
| MIB2 | 0.0135049 | 0.88670083 | 0.75585 | 0.053979626 | 0.215813034 | 1.823194988 | 0.090555 | 0.87805059 | 0.5148 | 0.090555004 | 0.090555004 | 1.769393714 | 0.149134519 | -0.826421823 | FALSE | TRUE | FALSE |
| PCDHGB7 | 0.0134932 | 0.88670868 | 1 | 0.013493193 | 0.013493193 | 0.263188207 | 0.0221968 | 0.893172667 | 0.0273 | 0.022196803 | 0.022196803 | 0.434244941 | 0.607889046 | -0.216175683 | FALSE | TRUE | FALSE |
| KCTD19 | 0.0134725 | 0.88671653 | 1 | 0.013472546 | 0.013472546 | 0.264622451 | 0.02196006 | 0.893246737 | 1 | 0.021960058 | 0.021960058 | 0.432261467 | 0.613502286 | -0.212183815 | FALSE | TRUE | FALSE |
| HBS1L | 0.0134592 | 0.88672438 | 1 | 0.118185087 | 0.472558804 | 1.819063732 | 0.02235544 | 0.893113342 | 0.5018 | 0.198041319 | 0.198041319 | 0.756178025 | 0.602053933 | -0.220364602 | FALSE | TRUE | FALSE |
| GAMT | 0.0134436 | 0.88673222 | 1 | 0.053738113 | 0.479681564 | 0.479681564 | 0.72702463 | 0.653352974 | 0.807 | 1.67352254 | 152.8192206 | 610.9132995 | 0.018491248 | -1.733033773 | FALSE | TRUE | FALSE |
| GLB1L3 | 0.0134332 | 0.88674007 | 0.21125 | 0.000916583 | 0.33442443 | 3.306249567 | 3.0932E-06 | 0.900958603 | 0.95525 | 0.000590667 | 0.090459296 | 0.887531437 | 4342.785308 | 3.63776836 | FALSE | TRUE | FALSE |
| ANO2 | 0.0134073 | 0.88674791 | 0.8788 | 0.120345244 | 1.108013696 | 2.356240276 | 0.0035525 | 0.898505294 | 0.02545 | 0.003552495 | 0.003552495 | 0.069730674 | 3.774050989 | 0.576807763 | FALSE | TRUE | FALSE |
| RDX | 0.013402 | 0.88675575 | 1 | 0.013401966 | 0.013401966 | 0.265131705 | 0.02230019 | 0.893128179 | 1 | 0.022300187 | 0.022300187 | 0.441741335 | 0.600979985 | -0.221139991 | FALSE | TRUE | FALSE |
| ECI1 | 0.0132926 | 0.88676359 | 1 | 0.117938694 | 0.117938694 | 0.117938694 | 0.19018785 | 0.854315448 | 0.756 | 0.190187849 | 1.743789335 | 1.743789335 | 0.069891723 | -1.155574252 | FALSE | TRUE | FALSE |
| POPDC2 | 0.0132532 | 0.88677143 | 0.0098 | 0.01325315 | 0.01325315 | 0.01325315 | 0.00180645 | 0.899129415 | 0.00855 | 0.001806446 | 0.001806446 | 0.001806446 | 7.336588192 | 0.865494142 | FALSE | TRUE | FALSE |
| SLC17A3 | 0.0132448 | 0.88677927 | 0.01255 | 0.013244827 | 0.013244827 | 0.262629392 | 0.02098798 | 0.893564154 | 1 | 0.181877548 | 0.181877548 | 0.416595444 | 0.631067207 | -0.199924387 | FALSE | TRUE | FALSE |
| PCP2 | 0.0132436 | 0.88678711 | 0.5021 | 0.120109843 | 0.120109843 | 0.120109843 | 0.01664064 | 0.894686938 | 0.005 | 0.016640642 | 0.016640642 | 0.016640642 | 0.795859769 | -0.099163449 | FALSE | TRUE | FALSE |
| CROCC | 0.013236 | 0.88679495 | 0.2918 | 0.013235959 | 0.113294445 | 2.178045511 | 0.21432157 | 0.848506344 | 0.8964 | 1.902521757 | 161.3951161 | 1540.574076 | 0.061757476 | -1.209310461 | FALSE | TRUE | FALSE |
| AMDHD2 | 0.0132203 | 0.88680278 | 1 | 0.013220254 | 0.013220254 | 0.260211608 | 0.02178477 | 0.893298529 | 1 | 0.087062209 | 0.087062209 | 0.4296038 | 0.606857728 | -0.216913113 | FALSE | TRUE | FALSE |
| AMBRA1 | 0.0131128 | 0.88681062 | 1 | 0.120477599 | 0.120477599 | 2.341839029 | 0.0225309 | 0.893068808 | 1 | 0.022530897 | 0.022530897 | 0.439438405 | 0.581992299 | -0.235082762 | FALSE | TRUE | FALSE |
| CLDN18 | 0.0130735 | 0.88681845 | 0.0071 | 0.013073464 | 0.013073464 | 0.013073464 | 0.74923322 | 0.641639404 | 0.25675 | 0.083132718 | 0.749233221 | 0.749233221 | 0.017449125 | -1.758226358 | FALSE | TRUE | FALSE |
| TRPV2 | 0.013059 | 0.88682629 | 1 | 0.052200599 | 0.052200599 | 1.026220197 | 0.02194157 | 0.893254138 | 1 | 0.021941571 | 0.021941571 | 0.432237965 | 0.595170647 | -0.225358495 | FALSE | TRUE | FALSE |
| COMMD2 | 0.013042 | 0.88683412 | 1 | 0.013042049 | 0.013042049 | 0.013042049 | 0.01494376 | 0.895125752 | 1 | 0.014943762 | 0.014943762 | 0.014943762 | 0.872742001 | -0.059114123 | FALSE | TRUE | FALSE |
| TMC1 | 0.0130204 | 0.88684195 | 1 | 0.456852412 | 4.114744769 | 4.114744769 | 1.67403809 | 0.51999674 | 0.5107 | 1.674038092 | 62.33746831 | 62.33746831 | 0.007777853 | -2.109140264 | FALSE | FALSE | FALSE |
| PLEKHH1 | 0.0130192 | 0.88684978 | 0.51715 | 0.01301924 | 0.01301924 | 0.252989199 | 0.23152825 | 0.844504039 | 0.15185 | 0.231528246 | 2.104056022 | 4.514450097 | 0.056231757 | -1.25001835 | FALSE | TRUE | FALSE |
| EVI5 | 0.0130171 | 0.88685761 | 0.51955 | 0.112723143 | 0.112723143 | 2.222680876 | 0.01917786 | 0.89404068 | 1 | 0.167653371 | 1.504301063 | 3.311406614 | 0.678755895 | -0.168286385 | FALSE | TRUE | FALSE |
| IL17RD | 0.0130103 | 0.88686544 | 0.01965 | 0.013010291 | 0.013010291 | 0.255932909 | 0.01513412 | 0.895097078 | 1 | 0.139825516 | 0.139825516 | 2.756025433 | 0.85966647 | -0.065670012 | FALSE | TRUE | FALSE |
| WDR72 | 0.0129919 | 0.88687326 | 1 | 0.11141962 | 0.11141962 | 2.189899558 | 0.17887119 | 0.857309184 | 0.7577 | 0.178871191 | 1.59113499 | 3.522939425 | 0.072632952 | -1.138866305 | FALSE | TRUE | FALSE |
| LCA5L | 0.0129864 | 0.88688109 | 1 | 0.012986371 | 0.012986371 | 0.256967201 | 0.02136582 | 0.893438874 | 1 | 0.021365818 | 0.021365818 | 0.423315608 | 0.607810617 | -0.216231718 | FALSE | TRUE | FALSE |
| BIN2 | 0.0129357 | 0.88688891 | 1 | 0.116299107 | 0.116299107 | 0.256149838 | 0.02214971 | 0.893180078 | 1 | 0.022149706 | 0.022149706 | 0.439126715 | 0.584011827 | -0.233578358 | FALSE | TRUE | FALSE |
| DMKN | 0.0129332 | 0.88689674 | 0.8734 | 0.111680139 | 0.989203901 | 0.989203901 | 0.00275415 | 0.898754462 | 1 | 0.002754153 | 0.002754153 | 0.054615123 | 4.695906932 | 0.671719481 | FALSE | TRUE | FALSE |
| TNFRSF10A | 0.0129264 | 0.88690456 | 1 | 3.04389E+14 | 1.17167E+20 | 2.266E+22 | 1.31369946 | 0.553294751 | 1 | 5.59437E+18 | 2.6899E+24 | 5.66228E+26 | 0.009839669 | -2.007019528 | FALSE | FALSE | FALSE |
| TRABD2A | 0.0129249 | 0.88691238 | 0.8755 | 0.948927564 | 8.449646621 | 8.449646621 | 0.02223698 | 0.893150427 | 1 | 0.190817013 | 0.190817013 | 0.440195887 | 0.581234339 | -0.235648736 | FALSE | TRUE | FALSE |
| ALDH18A1 | 0.0128353 | 0.8869202 | 0.75875 | 0.051290444 | 0.205009622 | 1.008158486 | 0.02196013 | 0.893239334 | 1 | 0.021960131 | 0.021960131 | 0.432539723 | 0.584483568 | -0.233227694 | FALSE | TRUE | FALSE |
| SERAC1 | 0.0127894 | 0.88692802 | 1 | 0.012789384 | 0.012789384 | 0.252618817 | 0.1838445 | 0.85599487 | 0.5121 | 0.183844497 | 0.183844497 | 0.407142852 | 0.069566315 | -1.157601001 | FALSE | TRUE | FALSE |

| | | | | | | | | | | | | | | | | | |
|---|---|---|---|---|---|---|---|---|---|---|---|---|---|---|---|---|---|
| ERBB3 | 0.0127876 | 0.88693584 | 0.3982 | 0.012787643 | 0.442281388 | 0.994331598 | 0.0010129 | 0.899428858 | 0.03455 | 0.001012897 | 0.001012897 | 0.034097645 | 12.62481771 | 1.101225116 | FALSE | TRUE | FALSE |
| ASIP | 0.0127874 | 0.88694366 | 1 | 0.012787392 | 0.012787392 | 0.012787392 | 0.01313662 | 0.895738734 | 0.00255 | 0.013136621 | 0.013136621 | 0.013136621 | 0.973415623 | -0.011701688 | FALSE | TRUE | FALSE |
| DNAJC10 | 0.0127548 | 0.88695148 | 0.01805 | 0.012754763 | 0.012754763 | 0.251742186 | 0.08178846 | 0.879853091 | 0.75205 | 0.081788458 | 0.327191496 | 1.616821223 | 0.155948209 | -0.807019609 | FALSE | TRUE | FALSE |
| DMXL2 | 0.0127526 | 0.88695929 | 1 | 10.17753276 | 337.588302 | 1887.649044 | 0.02324252 | 0.892905229 | 1 | 0.211845607 | 1.981765563 | 37.90588489 | 0.54867421 | -0.260685453 | FALSE | TRUE | FALSE |
| MTOR | 0.0127475 | 0.88696711 | 0.94535 | 0.269503134 | 10.28899522 | 48.73488355 | 0.52222364 | 0.746040398 | 0.0356 | 0.027407907 | 0.130646905 | 0.913683249 | 0.024410109 | -1.612430275 | FALSE | TRUE | FALSE |
| PTPRJ | 0.012741 | 0.88697492 | 1 | 0.012740963 | 0.012740963 | 0.429194417 | 0.02104148 | 0.893542066 | 1 | 0.021041481 | 0.021041481 | 0.410812519 | 0.605516485 | -0.217874029 | FALSE | TRUE | FALSE |
| CNTROB | 0.0127359 | 0.88698274 | 0.51585 | 0.108885222 | 0.108885222 | 2.131723489 | 0.02144938 | 0.893409357 | 1 | 0.021449384 | 0.021449384 | 0.420997827 | 0.59376654 | -0.226384279 | FALSE | TRUE | FALSE |
| FOXN2 | 0.012664 | 0.88699055 | 1 | 0.050594102 | 0.050594102 | 0.251404317 | 0.02148118 | 0.893379824 | 0.00925 | 0.021481177 | 0.021481177 | 0.021481177 | 0.589537241 | -0.229488755 | FALSE | TRUE | FALSE |
| ZNF880 | 0.0125946 | 0.88699836 | 1 | 2.1825E+124 | 7.4005E+142 | 1.3073E+150 | 8.2546E-05 | 0.900245244 | 1 | 3.6973E+99 | 4.9444E+114 | 4.9462E+121 | 152.578088 | 2.183492168 | FALSE | TRUE | FALSE |
| SNRNP200 | 0.0125924 | 0.88700617 | 1 | 0.012592388 | 0.239843916 | 0.420555107 | 0.02111636 | 0.893512602 | 0.0584 | 0.021116357 | 0.404514156 | 0.706739428 | 0.596333353 | -0.2245109 | FALSE | TRUE | FALSE |
| GART | 0.0125752 | 0.88701398 | 1 | 0.050267034 | 0.050267034 | 0.987933754 | 0.02013122 | 0.893806567 | 0.873 | 0.321796355 | 2.917309665 | 6.337676422 | 0.624661411 | -0.204355322 | FALSE | TRUE | FALSE |
| BSCL2 | 0.0125686 | 0.88702179 | 1 | 0.112336684 | 0.112336684 | 0.249095973 | 0.02092119 | 0.893593592 | 0.5025 | 0.188281619 | 0.188281619 | 0.415086203 | 0.600760239 | -0.221298818 | FALSE | TRUE | FALSE |
| EVC | 0.0125445 | 0.88702959 | 0.0273 | 0.0125445 | 0.0125445 | 0.24579805 | 0.02001985 | 0.893843208 | 1 | 0.173672428 | 0.173672428 | 3.411271013 | 0.626603151 | -0.203007426 | FALSE | TRUE | FALSE |
| RECQL5 | 0.0124843 | 0.8870374 | 0.75685 | 0.012484306 | 0.106568313 | 0.421415031 | 0.00330318 | 0.898559265 | 1 | 0.003303178 | 0.003303178 | 0.064789125 | 3.779483118 | 0.57743241 | FALSE | TRUE | FALSE |
| ERCC4 | 0.0124775 | 0.8870452 | 0.7537 | 0.026854776 | 0.107356936 | 0.526523642 | 0.02489691 | 0.892464326 | 0.51035 | 0.024896907 | 0.024896907 | 0.48929413 | 0.501167269 | -0.3000173 | FALSE | TRUE | FALSE |
| VNN2 | 0.0124718 | 0.88705301 | 1 | 1.027521077 | 9.691567959 | 9.691567959 | 7.08321419 | 0.24309093 | 0.1286 | 0.76453846 | 7.083214193 | 7.083214193 | 0.001760752 | -2.754301721 | FALSE | FALSE | FALSE |
| KCNK10 | 0.0124666 | 0.88706081 | 1 | 0.012466556 | 0.012466556 | 0.245869853 | 0.02093236 | 0.893578875 | 1 | 0.020932363 | 0.020932363 | 0.413525224 | 0.595563711 | -0.225071773 | FALSE | TRUE | FALSE |
| BEND5 | 0.0124424 | 0.88706861 | 1 | 0.012442431 | 0.012442431 | 0.012442431 | 0.01401913 | 0.895433081 | 0.0078 | 0.014019129 | 0.014019129 | 0.014019129 | 0.887532386 | -0.051815791 | FALSE | TRUE | FALSE |
| ZNF786 | 0.0124342 | 0.88707641 | 0.02865 | 0.012434199 | 0.012434199 | 0.243481586 | 0.17292318 | 0.858848759 | 0.51265 | 0.172923175 | 0.172923175 | 3.394645521 | 0.071905915 | -1.143235384 | FALSE | TRUE | FALSE |
| SEC23IP | 0.0123309 | 0.88708421 | 0.02645 | 0.012330859 | 0.012330859 | 0.242083725 | 0.02065821 | 0.893689158 | 0.0222 | 0.020658211 | 0.020658211 | 0.406466924 | 0.596898668 | -0.22409939 | FALSE | TRUE | FALSE |
| MLYCD | 0.0122979 | 0.88709201 | 0.0819 | 0.000769149 | 0.049193301 | 0.19682848 | 6.3477E-06 | 0.900803831 | 0.1982 | 6.34771E-06 | 2.5366E-05 | 0.00010139 | 1937.381155 | 3.287215071 | FALSE | TRUE | FALSE |
| UGT3A2 | 0.0122508 | 0.88709981 | 1 | 0.012250825 | 0.10639685 | 0.242638394 | 0.17779059 | 0.857511295 | 0.5134 | 0.177790588 | 0.177790588 | 0.40188338 | 0.068905923 | -1.161743449 | FALSE | TRUE | FALSE |
| ZNF426 | 0.0122347 | 0.88710761 | 1 | 0.105115704 | 0.105115704 | 0.24238337 | 0.02015879 | 0.893791904 | 1 | 0.175140772 | 0.175140772 | 0.399821175 | 0.606917285 | -0.216870494 | FALSE | TRUE | FALSE |
| SLC6A12 | 0.0122109 | 0.8871154 | 1 | 0.012210897 | 0.012210897 | 0.241326888 | 0.02048238 | 0.893718532 | 1 | 0.02048238 | 0.02048238 | 0.405374752 | 0.596165911 | -0.224632861 | FALSE | TRUE | FALSE |
| TNC | 0.0121979 | 0.8871232 | 0.89895 | 1.035652207 | 97.08743609 | 974.1628078 | 0.01306015 | 0.895767082 | 0.9412 | 0.11707956 | 4.306569557 | 20.63385137 | 0.933980607 | -0.029662141 | FALSE | TRUE | FALSE |
| CADPS2 | 0.0121931 | 0.88713099 | 1 | 0.108445491 | 0.108445491 | 2.125908375 | 0.0209612 | 0.893571515 | 1 | 0.020961201 | 0.020961201 | 0.411893504 | 0.58169664 | -0.235303445 | FALSE | TRUE | FALSE |
| RAP1GAP2 | 0.0121926 | 0.88713879 | 1 | 0.453969582 | 4.341702995 | 4.341702995 | 0.0218561 | 0.893276338 | 1 | 0.087340457 | 0.087340457 | 0.739921425 | 0.557857336 | -0.253476851 | FALSE | TRUE | FALSE |
| PTPRB | 0.0121804 | 0.88714658 | 0.763 | 0.109271022 | 0.436926448 | 8.390794036 | 0.08390774 | 0.879365463 | 0.5236 | 0.083907743 | 0.083907743 | 1.61911644 | 0.145164388 | -0.838139913 | FALSE | TRUE | FALSE |
| ZZEF1 | 0.0121775 | 0.88715437 | 1 | 6.82581E+14 | 2.88592E+20 | 5.78032E+22 | 0.00292736 | 0.898700684 | 1 | 1.57575E+16 | 1.28103E+21 | 1.59762E+24 | 4.159899981 | 0.619082889 | FALSE | TRUE | FALSE |
| VIL1 | 0.0121513 | 0.88716216 | 1 | 0.106342097 | 0.106342097 | 2.088340116 | 0.0821711 | 0.879761361 | 0.5178 | 0.082171102 | 0.082171102 | 1.617188792 | 0.147877673 | -0.830097393 | FALSE | TRUE | FALSE |
| ERBB2 | 0.0121121 | 0.88716995 | 0.98525 | 0.193516242 | 3.09491083 | 15.03211073 | 0.21623027 | 0.848150692 | 0.63905 | 0.379617577 | 3.4556635 | 16.8484627 | 0.056014988 | -1.251695749 | FALSE | TRUE | FALSE |
| LAPTM4B | 0.0120985 | 0.88717774 | 0.00685 | 0.001579894 | 0.012098458 | 0.012098458 | 0.00355402 | 0.898498545 | 0.896 | 0.223584957 | 15.53561434 | 134.2204621 | 3.40415751 | 0.532009647 | FALSE | TRUE | FALSE |
| LBP | 0.0120966 | 0.88718552 | 0.75495 | 0.012096633 | 0.109280558 | 0.239403455 | 0.00203646 | 0.899076009 | 1 | 0.002036461 | 0.002036461 | 0.040354109 | 5.940027784 | 0.773788476 | FALSE | TRUE | FALSE |
| HUS1 | 0.0120928 | 0.88719331 | 1 | 0.012092834 | 0.012092834 | 0.012092834 | 0.01362297 | 0.895589671 | 1 | 0.013622965 | 0.013622965 | 0.013622965 | 0.887680019 | -0.051743556 | FALSE | TRUE | FALSE |
| CYP2C9 | 0.01206 | 0.88720109 | 0.0118 | 0.012060025 | 0.012060025 | 0.239066065 | 0.17418711 | 0.858575747 | 0.51005 | 0.174187114 | 0.174187114 | 0.387575063 | 0.069236034 | -1.159667819 | FALSE | TRUE | FALSE |
| BPTF | 0.0120367 | 0.88720888 | 0.0728 | 0.012036732 | 0.229027041 | 0.399566991 | 0.02017444 | 0.893784571 | 0.06565 | 0.020174436 | 0.386122055 | 0.671612255 | 0.59663285 | -0.224292839 | FALSE | TRUE | FALSE |
| TNNT3 | 0.0120363 | 0.88721666 | 1 | 0.048102438 | 0.048102438 | 0.048102438 | 0.020212 | 0.893769902 | 1 | 0.020211997 | 0.080784522 | 0.080784522 | 0.595503751 | -0.225115498 | FALSE | TRUE | FALSE |
| PTH2R | 0.0119884 | 0.88722444 | 1 | 0.415383073 | 14.77616575 | 14.77616575 | 0.0212958 | 0.893461003 | 1 | 0.085160949 | 0.085160949 | 0.421866622 | 0.562947505 | -0.249532101 | FALSE | TRUE | FALSE |
| PCDHB14 | 0.0119672 | 0.88723222 | 0.8774 | 0.223996755 | 3.581062184 | 14.32384933 | 0.00636517 | 0.89775791 | 0.02175 | 0.006365174 | 0.006365174 | 0.125086885 | 1.880108881 | 0.274183001 | FALSE | TRUE | FALSE |
| PLB1 | 0.0119601 | 0.88724 | 0.02105 | 0.00153358 | 0.011960142 | 0.029772248 | 0.0312512 | 0.890921822 | 0.9794 | 18.1928884 | 13045.14083 | 122007.2485 | 0.382709788 | -0.417130431 | FALSE | TRUE | FALSE |
| FBF1 | 0.0119594 | 0.88724778 | 0.0338 | 0.011959372 | 0.011959372 | 0.233035529 | 0.01973772 | 0.893916421 | 1 | 0.01973772 | 0.01973772 | 0.385793449 | 0.605914563 | -0.217588609 | FALSE | TRUE | FALSE |
| FIBIN | 0.011929 | 0.88725556 | 1 | 0.112351888 | 0.112351888 | 0.112351888 | 0.01297708 | 0.895802498 | 1 | 0.012977079 | 0.012977079 | 0.012977079 | 0.919232915 | -0.036574433 | FALSE | TRUE | FALSE |
| DAAM1 | 0.0119266 | 0.88726334 | 1 | 0.047663444 | 0.047663444 | 0.933809116 | 0.71262106 | 0.659893462 | 0.39035 | 0.308139876 | 2.848926413 | 10.39310387 | 0.016736212 | -1.776342844 | FALSE | TRUE | FALSE |
| ARG1 | 0.011906 | 0.88727111 | 1 | 0.047588813 | 0.047588813 | 0.047588813 | 0.30263644 | 0.824558894 | 0.50685 | 0.682122602 | 10.9077519 | 10.9077519 | 0.039340935 | -1.405155326 | FALSE | TRUE | FALSE |
| TMC4 | 0.0118964 | 0.88727889 | 1 | 0.103757639 | 0.103757639 | 2.040202956 | 0.17418336 | 0.858588178 | 0.0175 | 0.174183361 | 0.174183361 | 0.669499293 | 0.068298141 | -1.165591114 | FALSE | TRUE | FALSE |
| KRT8 | 0.0118519 | 0.88728666 | 1 | 0.011851948 | 0.011851948 | 0.234014835 | 0.01999744 | 0.893857858 | 0.0157 | 0.019997438 | 0.019997438 | 0.395452377 | 0.592673345 | -0.227184605 | FALSE | TRUE | FALSE |
| C11orf16 | 0.0117965 | 0.88729443 | 0.2629 | 0.011796544 | 0.106400341 | 0.106400341 | 0.00182489 | 0.899122742 | 1 | 0.146937544 | 1.37010678 | 1.37010678 | 6.464245575 | 0.810517847 | FALSE | TRUE | FALSE |
| ZDHHC3 | 0.0117801 | 0.88730221 | 0.0104 | 0.011780099 | 0.011780099 | 0.233946774 | 0.15530838 | 0.863200481 | 0.75545 | 0.155308383 | 1.356432842 | 1.356432842 | 0.075849727 | -1.120045976 | FALSE | TRUE | FALSE |
| MEP1B | 0.0117612 | 0.88730998 | 0.9428 | 0.903939196 | 32.93211428 | 32.93211428 | 0.17590898 | 0.858114289 | 0.7527 | 0.703472586 | 6.318699198 | 6.318699198 | 0.066859634 | -1.174836007 | FALSE | TRUE | FALSE |
| SUMO2 | 0.0117472 | 0.88731775 | 0.00305 | 0.011747163 | 0.011747163 | 0.011747163 | 0.01187573 | 0.896134341 | 1 | 0.011875727 | 0.011875727 | 0.011875727 | 0.989174233 | -0.004727205 | FALSE | TRUE | FALSE |
| EXD2 | 0.0117204 | 0.88732552 | 1 | 0.011720426 | 0.011720426 | 0.232042914 | 0.01946426 | 0.893967619 | 1 | 0.019464259 | 0.019464259 | 0.385824378 | 0.602151176 | -0.220294461 | FALSE | TRUE | FALSE |
| DDX54 | 0.0117172 | 0.88733328 | 1 | 0.04685624 | 0.187421046 | 1.581398233 | 0.07896338 | 0.880654675 | 0.02855 | 0.078963384 | 0.078963384 | 1.543865193 | 0.148388183 | -0.828600682 | FALSE | TRUE | FALSE |
| KRT19 | 0.0117171 | 0.88734105 | 0.0172 | 0.011717076 | 0.011717076 | 0.231338448 | 0.01979373 | 0.893901786 | 1 | 0.019793729 | 0.019793729 | 0.391404072 | 0.591959035 | -0.227708346 | FALSE | TRUE | FALSE |
| SCAF1 | 0.0116713 | 0.88734882 | 0.7606 | 0.107308102 | 1.012605318 | 3.617755581 | 0.01411052 | 0.895390297 | 1 | 1.23118016 | 11.94989025 | 24.08845387 | 0.827135924 | -0.082423116 | FALSE | TRUE | FALSE |
| ZSCAN29 | 0.0116537 | 0.88735658 | 0.7574 | 0.099830425 | 0.877778384 | 1.963334243 | 0.02043101 | 0.893733213 | 1 | 0.020431007 | 0.020431007 | 0.40261646 | 0.570390718 | -0.24382755 | FALSE | TRUE | FALSE |
| PIGN | 0.0116313 | 0.88736434 | 1 | 0.102340019 | 0.102340019 | 2.020013064 | 0.02003421 | 0.893835882 | 0.0176 | 0.020034215 | 0.020034215 | 0.39606314 | 0.580574215 | -0.236142257 | FALSE | TRUE | FALSE |
| FKRP | 0.0115644 | 0.88737211 | 0.49715 | 0.011564367 | 0.046206261 | 0.228761053 | 2.66615339 | 0.436456914 | 0.4704 | 1.590967247 | 25.41174967 | 101.5977403 | 0.004337473 | -2.362763253 | FALSE | FALSE | FALSE |
| AGMO | 0.0115603 | 0.88737987 | 1 | 0.899802767 | 77.52848279 | 77.52848279 | 0.17465418 | 0.858426403 | 0.87855 | 1.577651344 | 14.617252 | 14.617252 | 0.066189379 | -1.179211693 | FALSE | TRUE | FALSE |
| ZFYVE26 | 0.0115538 | 0.88738763 | 0.3033 | 0.0115538 | 0.220018832 | 1.887239331 | 0.00299211 | 0.898673775 | 0.5294 | 0.025380262 | 0.057308233 | 0.486110592 | 3.861417907 | 0.586746806 | FALSE | TRUE | FALSE |
| ARHGEF16 | 0.0115459 | 0.88739539 | 1 | 0.011545927 | 0.011545927 | 0.228242495 | 1.41059002 | 0.544370859 | 0.1361 | 1.410590018 | 13.09763543 | 13.09763543 | 0.008185176 | -2.086972003 | FALSE | FALSE | FALSE |
| P2RX3 | 0.0115082 | 0.88740315 | 0.01155 | 0.01150822 | 0.01150822 | 0.228024272 | 0.0142016 | 0.895368893 | 1 | 0.128633426 | 0.128633426 | 0.281705741 | 0.810346604 | -0.091329184 | FALSE | TRUE | FALSE |
| PLCG2 | 0.0114903 | 0.88741091 | 1 | 0.105123238 | 0.105123238 | 2.037459875 | 0.02003508 | 0.893828555 | 0.0373 | 0.020035081 | 0.020035081 | 0.389760403 | 0.573506591 | -0.241461586 | FALSE | TRUE | FALSE |
| GFPT2 | 0.0114531 | 0.88741866 | 0.51345 | 0.103729958 | 0.103729958 | 2.039597195 | 0.01876277 | 0.89414281 | 1 | 0.170704728 | 1.594010832 | 3.363310631 | 0.610416689 | -0.214373601 | FALSE | TRUE | FALSE |
| TJP2 | 0.011448 | 0.88742642 | 1 | 0.099670379 | 0.099670379 | 1.937759058 | 0.01910898 | 0.894055281 | 0.5162 | 0.167998442 | 0.167998442 | 3.27716258 | 0.59908864 | -0.222508916 | FALSE | TRUE | FALSE |
| MS4A6A | 0.0114211 | 0.88743417 | 1 | 0.011421147 | 0.011421147 | 0.011421147 | 0.01366284 | 0.895575454 | 0.00475 | 0.01366284 | 0.01366284 | 0.01366284 | 0.835927756 | -0.077831255 | FALSE | TRUE | FALSE |
| MCM8 | 0.0114135 | 0.88744193 | 1 | 7.524430098 | 68.87469365 | 645.8840218 | 0.18210899 | 0.856444116 | 0.50745 | 0.182108985 | 0.182108985 | 0.714278543 | 0.06267412 | -1.202911755 | FALSE | TRUE | FALSE |
| MARVELD3 | 0.0114036 | 0.88744968 | 1 | 0.401868672 | 14.53936338 | 14.53936338 | 0.02028629 | 0.893747892 | 1 | 0.0811056 | 0.324345339 | 0.324345339 | 0.562134443 | -0.250159804 | FALSE | TRUE | FALSE |
| PDZD2 | 0.0113938 | 0.88745743 | 0.0793 | 0.011393794 | 0.214628511 | 0.379553952 | 0.01910222 | 0.89406258 | 1 | 0.019102221 | 0.362365548 | 0.637889764 | 0.59646434 | -0.224415516 | FALSE | TRUE | FALSE |
| CCDC30 | 0.0113602 | 0.88746518 | 0.76005 | 0.096144628 | 0.835196439 | 1.900787129 | 858294.187 | 1.93674E-06 | 0.0041 | 892.3496536 | 84995.85397 | 858294.1874 | 1.32358E-08 | -7.878249591 | FALSE | FALSE | FALSE |
| IDH1 | 0.0113581 | 0.88747293 | 1 | 0.045388597 | 0.045388597 | 0.225277389 | 0.01945068 | 0.893974929 | 0.0087 | 0.019450679 | 0.019450679 | 0.019450679 | 0.583942781 | -0.233629706 | FALSE | TRUE | FALSE |
| NEURL4 | 0.0113064 | 0.88748068 | 1 | 4.13055E+43 | 3.83562E+52 | 3.18959E+56 | 0.00550988 | 0.897989965 | 1 | 2.47826E+30 | 9.51174E+37 | 2.5392E+41 | 2.052018064 | 0.31218118 | FALSE | TRUE | FALSE |
| PPP1R14D | 0.0112909 | 0.88748843 | 1 | 0.105466785 | 1.011095172 | 1.011095172 | 0.0118575 | 0.89614138 | 1 | 0.110873809 | 10.47313474 | 10.47313474 | 0.9522144 | -0.021265255 | FALSE | TRUE | FALSE |
| RRP12 | 0.011289 | 0.88749618 | 0.5178 | 0.101106846 | 0.101106846 | 1.961385764 | 0.01904147 | 0.894084472 | 1 | 0.171678428 | 0.171678428 | 3.3424784 | 0.59286256 | -0.227045975 | FALSE | TRUE | FALSE |

| | | | | | | | | | | | | | | | | | |
|---|---|---|---|---|---|---|---|---|---|---|---|---|---|---|---|---|---|
| ATAD2B | 0.011261 | 0.88750392 | 0.7605 | 0.105141091 | 0.420142729 | 7.393051947 | 0.01975625 | 0.893909104 | 0.5206 | 0.078955116 | 0.078955116 | 1.545981217 | 0.569997586 | -0.244126984 | FALSE | TRUE | FALSE |
| KLKB1 | 0.0112489 | 0.88751167 | 0.969 | 0.211079633 | 3.373919773 | 13.49403329 | 101.476939 | 0.028989378 | 0.22075 | 11.44025445 | 923.2669822 | 3694.692553 | 0.000110852 | -3.955257387 | FALSE | FALSE | FALSE |
| HERC2 | 0.0112487 | 0.88751941 | 0.13855 | 0.00281486 | 0.05051794 | 0.806946466 | 0.00225931 | 0.898975732 | 0.7257 | 0.005145061 | 0.164049933 | 1.492482204 | 4.978815131 | 0.697126001 | FALSE | TRUE | FALSE |
| CCDC42 | 0.0112315 | 0.88752715 | 1 | 0.011231522 | 0.011231522 | 0.222726412 | 0.013025 | 0.895774167 | 1 | 0.013024997 | 0.013024997 | 0.013024997 | 0.862305192 | -0.064338999 | FALSE | TRUE | FALSE |
| EXOSC10 | 0.0112313 | 0.8875349 | 0.02775 | 0.011231341 | 0.011231341 | 0.220427093 | 0.15855853 | 0.862426183 | 0.756 | 0.158558535 | 1.4490587 | 3.118886931 | 0.070834036 | -1.149758015 | FALSE | TRUE | FALSE |
| MERTK | 0.0112282 | 0.88754264 | 1 | 6.43593E+47 | 8.04245E+57 | 2.4014E+62 | 0.25860363 | 0.837096996 | 1 | 2.40195E+52 | 3.89101E+62 | 1.59225E+67 | 0.043418452 | -1.362325664 | FALSE | TRUE | FALSE |
| GLB1L | 0.0111952 | 0.88755038 | 0.2597 | 0.002803472 | 0.044717061 | 0.044717061 | 0.0037876 | 0.898397195 | 0.73495 | 0.033123047 | 0.528828035 | 2.113826317 | 2.955741035 | 0.470666381 | FALSE | TRUE | FALSE |
| PAG1 | 0.0111817 | 0.88755811 | 1 | 0.011181663 | 0.011181663 | 0.221659085 | 0.10828063 | 0.873782905 | 0.7517 | 0.108280629 | 1.038813397 | 1.038813397 | 0.103265592 | -0.986044362 | FALSE | TRUE | FALSE |
| HBP1 | 0.0111773 | 0.88756585 | 0.50575 | 0.103793142 | 0.103793142 | 0.221564127 | 0.1769648 | 0.857838541 | 0.0111 | 0.176964802 | 0.176964802 | 0.377180035 | 0.06316121 | -1.199549556 | FALSE | TRUE | FALSE |
| SLC27A1 | 0.0111681 | 0.88757359 | 0.76055 | 0.098172239 | 0.39252438 | 1.922656611 | 0.07725369 | 0.880997742 | 0.51575 | 0.077253686 | 0.077253686 | 1.51676233 | 0.144563797 | -0.839940454 | FALSE | TRUE | FALSE |
| SMTN | 0.0111562 | 0.88758132 | 0.94075 | 0.403586282 | 6.453401582 | 24.03763372 | 0.01953803 | 0.893938369 | 1 | 0.177797021 | 0.177797021 | 3.477862464 | 0.570999388 | -0.243364357 | FALSE | TRUE | FALSE |
| FBXL13 | 0.0111261 | 0.88758906 | 0.01735 | 0.011126075 | 0.011126075 | 0.219609937 | 11.6411103 | 0.180292115 | 0.13305 | 1.286218243 | 11.64111032 | 11.64111032 | 0.000955757 | -3.019652438 | FALSE | FALSE | FALSE |
| ADM | 0.0111192 | 0.88759679 | 0.0082 | 0.011119157 | 0.011119157 | 0.011119157 | 0.02454874 | 0.892591707 | 1 | 0.098194495 | 0.098194495 | 0.098194495 | 0.452942157 | -0.343957256 | FALSE | TRUE | FALSE |
| BANK1 | 0.0111145 | 0.88760452 | 0.50995 | 0.09400579 | 0.09400579 | 0.375447038 | 1.30527083 | 0.554067877 | 0.50995 | 1.305270825 | 11.75333844 | 11.75333844 | 0.008515097 | -2.069810392 | FALSE | FALSE | FALSE |
| NUP107 | 0.0110742 | 0.88761226 | 1 | 0.01107424 | 0.01107424 | 0.217745716 | 0.01853078 | 0.894201092 | 1 | 0.018530784 | 0.018530784 | 0.36509951 | 0.597613124 | -0.223579874 | FALSE | TRUE | FALSE |
| LDB3 | 0.0110645 | 0.88761999 | 0.7517 | 0.024744723 | 0.098871765 | 0.48616092 | 0.0056178 | 0.897962713 | 1 | 0.02244965 | 0.089734951 | 0.442006646 | 1.969552239 | 0.294367504 | FALSE | TRUE | FALSE |
| EMILIN2 | 0.0110576 | 0.88762772 | 0.51595 | 0.094806131 | 0.094806131 | 1.860015124 | 0.01896217 | 0.894106355 | 1 | 0.018962174 | 0.018962174 | 0.372874719 | 0.583137978 | -0.234228673 | FALSE | TRUE | FALSE |
| CDH17 | 0.0110509 | 0.88763544 | 0.02105 | 0.011050933 | 0.011050933 | 0.217568917 | 0.15778501 | 0.862579347 | 0.5091 | 0.157785008 | 0.157785008 | 0.608178834 | 0.070037915 | -1.154666792 | FALSE | TRUE | FALSE |
| ZNF343 | 0.011048 | 0.88764317 | 0.01585 | 0.011047952 | 0.011047952 | 0.218629087 | 1.17049946 | 0.570572582 | 0.89125 | 10.57800879 | 930.0090602 | 9037.179932 | 0.009438665 | -2.02508943 | FALSE | FALSE | FALSE |
| NPAT | 0.0110268 | 0.8876509 | 1 | 0.011026812 | 0.011026812 | 0.371154396 | 0.12738099 | 0.869570731 | 0.50685 | 0.127380993 | 0.127380993 | 2.494792857 | 0.086565596 | -1.062654678 | FALSE | TRUE | FALSE |
| KRBA2 | 0.0110237 | 0.88765862 | 0.87695 | 0.101090045 | 0.950848402 | 0.950848402 | 0.00172882 | 0.899156098 | 1 | 0.015580843 | 0.015580843 | 0.034292605 | 6.37639418 | 0.804575157 | FALSE | TRUE | FALSE |
| RBM28 | 0.010986 | 0.88766635 | 1 | 0.010986043 | 0.010986043 | 0.216475037 | 0.01844545 | 0.894222932 | 1 | 0.018445453 | 0.018445453 | 0.364105588 | 0.595596261 | -0.225048037 | FALSE | TRUE | FALSE |
| LCN2 | 0.010985 | 0.88767407 | 1 | 0.010985037 | 0.010985037 | 0.010985037 | 0.01300031 | 0.895788335 | 0.0054 | 0.013000314 | 0.013000314 | 0.013000314 | 0.844982409 | -0.073152332 | FALSE | TRUE | FALSE |
| BMF | 0.010974 | 0.88768179 | 1 | 0.010974003 | 0.010974003 | 0.010974003 | 0.01180983 | 0.896169528 | 1 | 0.011809826 | 0.011809826 | 0.011809826 | 0.929226442 | -0.031878441 | FALSE | TRUE | FALSE |
| FREM2 | 0.0109507 | 0.88768951 | 0.9999 | 791644.2671 | 11366244464 | 2.12792E+11 | 0.00063333 | 0.899680407 | 1 | 0.43792629 | 14.5942266 | 76.8352413 | 17.29075668 | 1.237813999 | FALSE | TRUE | FALSE |
| PANK4 | 0.0109358 | 0.88769723 | 0.75415 | 0.043695319 | 0.17463308 | 0.855333759 | 0.01849208 | 0.894215653 | 1 | 0.172730859 | 0.172730859 | 3.389853725 | 0.591379448 | -0.228133772 | FALSE | TRUE | FALSE |
| EHMT1 | 0.0109259 | 0.88770495 | 1 | 0.01092587 | 0.01092587 | 0.367669277 | 0.01865292 | 0.894164672 | 0.0405 | 0.018652925 | 0.018652925 | 0.361780021 | 0.585745695 | -0.232290895 | FALSE | TRUE | FALSE |
| NCOR1 | 0.0109084 | 0.88771267 | 1 | 0.010908434 | 0.207962372 | 0.363188112 | 0.01856279 | 0.894186527 | 0.0573 | 0.018562794 | 0.355888184 | 0.619579439 | 0.587650434 | -0.230880939 | FALSE | TRUE | FALSE |
| ZNF337 | 0.010893 | 0.88772039 | 0.81665 | 0.765780005 | 6.66640262 | 59.45697153 | 0.00311646 | 0.898633387 | 0.5105 | 0.025667327 | 0.025667327 | 0.105230459 | 3.495310284 | 0.543485735 | FALSE | TRUE | FALSE |
| FZD6 | 0.0108853 | 0.88772811 | 0.9672 | 0.695596629 | 6.272667836 | 25.09044516 | 0.30774398 | 0.82289347 | 0.50395 | 0.307743975 | 1.230771921 | 2.603441664 | 0.035371285 | -1.451349168 | FALSE | TRUE | FALSE |
| KRTAP3-1 | 0.0108732 | 0.88773582 | 1 | 0.01087316 | 0.01087316 | 0.01087316 | 0.01119797 | 0.896373173 | 0.00195 | 0.01119797 | 0.01119797 | 0.01119797 | 0.970993824 | -0.012783532 | FALSE | TRUE | FALSE |
| NBEAL2 | 0.0108648 | 0.88774353 | 0.7154 | 0.097403018 | 3.580382126 | 16.82313806 | 0.01151598 | 0.896246858 | 0.88455 | 0.046048516 | 0.217892583 | 3.48482297 | 0.943451691 | -0.025280333 | FALSE | TRUE | FALSE |
| HPS4 | 0.0108288 | 0.88775125 | 1 | 0.010828773 | 0.010828773 | 0.213406948 | 0.01707361 | 0.894564051 | 1 | 0.610632987 | 5.606756168 | 5.606756168 | 0.634240432 | -0.197746076 | FALSE | TRUE | FALSE |
| CCDC28A | 0.0108243 | 0.88775896 | 0.50315 | 0.097012749 | 0.097012749 | 0.097012749 | 0.01350047 | 0.895646504 | 0.7525 | 0.121689647 | 1.125793479 | 1.125793479 | 0.801771846 | -0.095949198 | FALSE | TRUE | FALSE |
| FMN2 | 0.0108227 | 0.88776667 | 1 | 0.095366432 | 0.862515599 | 16.67870915 | 0.01880532 | 0.894135521 | 1 | 0.16725528 | 0.16725528 | 3.247190192 | 0.575512579 | -0.239945179 | FALSE | TRUE | FALSE |
| ENPP1 | 0.0108155 | 0.88777438 | 1 | 0.010815454 | 0.010815454 | 0.213129707 | 0.01828261 | 0.894244765 | 1 | 0.018282606 | 0.018282606 | 0.360915 | 0.591570712 | -0.227993336 | FALSE | TRUE | FALSE |
| HPS6 | 0.0108009 | 0.88778209 | 1 | 0.097001242 | 0.894125399 | 1.911639557 | 0.01889011 | 0.89412094 | 0.5088 | 0.170879642 | 0.170879642 | 0.63864196 | 0.571776352 | -0.24277381 | FALSE | TRUE | FALSE |
| EPHA2 | 0.0107974 | 0.8877898 | 1 | 0.043156904 | 0.17254081 | 1.456887707 | 0.29037489 | 0.827924949 | 0.52015 | 0.290374893 | 1.16130439 | 5.671251472 | 0.037184182 | -1.429641766 | FALSE | TRUE | FALSE |
| KCNQ2 | 0.0107614 | 0.8877975 | 0.0148 | 0.010761352 | 0.010761352 | 0.210670224 | 0.00055685 | 0.899733211 | 0.26325 | 0.000556849 | 0.00537677 | 0.010928988 | 19.3254377 | 1.286129339 | FALSE | TRUE | FALSE |
| NSMAF | 0.010749 | 0.88780521 | 0.63515 | 0.022803417 | 0.793346509 | 0.793346509 | 0.01311315 | 0.895745822 | 0.87585 | 0.115478042 | 4.168865539 | 4.168865539 | 0.819713062 | -0.086338144 | FALSE | TRUE | FALSE |
| KIFAP3 | 0.0107394 | 0.88781292 | 1 | 0.010739379 | 0.010739379 | 0.212036042 | 0.1194726 | 0.871340544 | 0.01765 | 0.1194726 | 0.1194726 | 0.441962073 | 0.089889892 | -1.046289143 | FALSE | TRUE | FALSE |
| SRBD1 | 0.0107248 | 0.88782062 | 1 | 0.010724802 | 0.010724802 | 0.210696514 | 0.15121004 | 0.864311917 | 0.5131 | 0.151210043 | 0.151210043 | 2.976966465 | 0.070926522 | -1.149191336 | FALSE | TRUE | FALSE |
| PCDHB8 | 0.0106644 | 0.88782832 | 1 | 0.092726445 | 0.092726445 | 1.817780549 | 0.15291867 | 0.863827806 | 0.2747 | 0.152918668 | 1.377075247 | 3.004924151 | 0.06973911 | -1.1565236 | FALSE | TRUE | FALSE |
| UQCR11 | 0.0106427 | 0.88783603 | 0.00245 | 0.010642658 | 0.010642658 | 0.010642658 | 0.01117441 | 0.896394197 | 1 | 0.011174413 | 0.011174413 | 0.011174413 | 0.952413092 | -0.021174643 | FALSE | TRUE | FALSE |
| TAS2R5 | 0.0106419 | 0.88784373 | 1 | 0.010641866 | 0.010641866 | 0.010641866 | 0.10357264 | 0.875016396 | 0.50615 | 0.103572645 | 0.103572645 | 0.103572645 | 0.102747842 | -0.98822729 | FALSE | TRUE | FALSE |
| SLCO1B1 | 0.0106204 | 0.88785143 | 1 | 0.092014166 | 0.092014166 | 0.35930152 | 0.0174595 | 0.894433645 | 0.88265 | 0.279254616 | 2.444404244 | 2.444404244 | 0.608288091 | -0.215890686 | FALSE | TRUE | FALSE |
| IL31RA | 0.0106189 | 0.88785913 | 1 | 0.010618861 | 0.010618861 | 0.20983973 | 1.26483109 | 0.560036094 | 0.25905 | 0.143568147 | 1.264831089 | 2.841134253 | 0.008395477 | -2.075954601 | FALSE | FALSE | FALSE |
| ALDH4A1 | 0.0106148 | 0.88786682 | 0.50565 | 0.094768213 | 0.094768213 | 0.20939392 | 0.29025403 | 0.827997891 | 0.26385 | 0.072588706 | 0.29025403 | 1.434288697 | 0.036570589 | -1.436868043 | FALSE | TRUE | FALSE |
| ABCB5 | 0.010607 | 0.88787452 | 0.4745 | 0.006056551 | 24.62846872 | 916.7225082 | 3.9377E-06 | 0.900907066 | 0.5426 | 7.1935E-06 | 0.007332946 | 0.081877404 | 2693.734422 | 3.430354776 | FALSE | TRUE | FALSE |
| PSG4 | 0.0106004 | 0.88788222 | 1 | 0.089058419 | 0.089058419 | 0.210238033 | 0.01808837 | 0.894295678 | 1 | 0.01808837 | 0.01808837 | 0.01808837 | 0.586035368 | -0.232076173 | FALSE | TRUE | FALSE |
| N4BP2 | 0.0105572 | 0.88788991 | 0.0418 | 0.010557229 | 0.010557229 | 0.35469051 | 0.01720467 | 0.894520615 | 0.519 | 0.149460694 | 0.149460694 | 2.913932558 | 0.613625697 | -0.212096462 | FALSE | TRUE | FALSE |
| CLCN6 | 0.010539 | 0.88789761 | 0.5075 | 0.010539011 | 0.051652226 | 0.206247231 | 0.00022974 | 0.899957035 | 0.878 | 0.016833654 | 0.268475636 | 0.330286075 | 45.87441326 | 1.661570523 | FALSE | TRUE | FALSE |
| NAV2 | 0.0105376 | 0.8879053 | 0.07085 | 0.01053763 | 0.021920652 | 0.199225211 | 0.02343683 | 0.892875441 | 0.53495 | 0.023436828 | 0.048830343 | 0.446029455 | 0.449618451 | -0.347155875 | FALSE | TRUE | FALSE |
| ALDOA | 0.0105133 | 0.88791299 | 1 | 0.010513328 | 0.010513328 | 0.208350493 | 0.01801043 | 0.894339281 | 0.0107 | 0.018010428 | 0.018010428 | 0.35731734 | 0.583735598 | -0.233783821 | FALSE | TRUE | FALSE |
| TEX9 | 0.0105 | 0.88792068 | 0.8726 | 0.791123507 | 7.136216345 | 7.136216345 | 0.01917929 | 0.894033378 | 1 | 0.019179288 | 0.019179288 | 0.019179288 | 0.547465256 | -0.261643437 | FALSE | TRUE | FALSE |
| VDR | 0.0104968 | 0.88792837 | 1 | 0.010496771 | 0.010496771 | 0.207683517 | 2.4667965 | 0.449819311 | 0.50335 | 2.466796497 | 39.43093614 | 157.7072666 | 0.004255224 | -2.371077599 | FALSE | FALSE | FALSE |
| SDK2 | 0.0104416 | 0.88793606 | 1 | 807851.2054 | 1841367827 | 60698057950 | 0.0225808 | 0.893046529 | 1 | 178324.2006 | 86113876.37 | 1582396956 | 0.46241095 | -0.334971891 | FALSE | TRUE | FALSE |
| GRM3 | 0.0104396 | 0.88794375 | 1 | 0.097591918 | 0.097591918 | 1.912321279 | 0.01546382 | 0.895018156 | 1 | 22.07436982 | 2183.337712 | 22520.34008 | 0.675097302 | -0.170633628 | FALSE | TRUE | FALSE |
| IGFALS | 0.01042 | 0.88795144 | 0.876 | 0.362462422 | 3.236480612 | 7.136118492 | 0.01676975 | 0.894650822 | 1 | 1.323927145 | 115.69417 | 115.69417 | 0.621358788 | -0.206657555 | FALSE | TRUE | FALSE |
| KRT5 | 0.0104125 | 0.88795912 | 1 | 0.041596493 | 0.16621377 | 0.819801302 | 0.01808427 | 0.894302947 | 1 | 0.072255366 | 0.072255366 | 0.357037823 | 0.575776618 | -0.239745976 | FALSE | TRUE | FALSE |
| NLRP14 | 0.0103954 | 0.88796681 | 0.7552 | 0.01039537 | 0.088784581 | 0.350808093 | 0.02119084 | 0.893497864 | 0.939 | 0.183332226 | 14.79977391 | 14.79977391 | 0.490559641 | -0.309308184 | FALSE | TRUE | FALSE |
| DMPK | 0.0103891 | 0.88797449 | 1 | 0.088746014 | 0.088746014 | 0.351772291 | 0.07085443 | 0.882246572 | 0.5065 | 0.070854427 | 0.070854427 | 0.600163796 | 0.146626065 | -0.83378882 | FALSE | TRUE | FALSE |
| ATP9B | 0.0103876 | 0.88798217 | 0.1445 | 9.23611E-06 | 0.088788185 | 0.354764165 | 5.7615E-05 | 0.900310518 | 0.146 | 8.01968E-06 | 0.000127904 | 0.000510985 | 180.2945291 | 2.255982548 | FALSE | TRUE | FALSE |
| SCIN | 0.0103814 | 0.88798986 | 0.5112 | 0.090083488 | 0.090083488 | 0.206036152 | 0.14456759 | 0.865744918 | 0.87755 | 1.298422074 | 11.96149091 | 11.96149091 | 0.071810106 | -1.143814431 | FALSE | TRUE | FALSE |
| KIR2DL3 | 0.0103509 | 0.88799754 | 1 | 1.85484E+61 | 7.52448E+71 | 1.92612E+77 | 0.83213975 | 0.612048139 | 1 | 4.1412E+71 | 8.70535E+83 | 3.31391E+89 | 0.012438911 | -1.905217632 | FALSE | TRUE | FALSE |
| CEP350 | 0.0103433 | 0.88800522 | 1 | 0.010343337 | 0.196718928 | 0.342682965 | 0.01714558 | 0.894527856 | 0.5298 | 0.156808724 | 0.328023377 | 5.211203522 | 0.603265537 | -0.219491484 | FALSE | TRUE | FALSE |
| SCLY | 0.0103178 | 0.88801289 | 1 | 0.164804427 | 0.658904228 | 0.815258442 | 0.16087788 | 0.861845936 | 0.51395 | 0.160877877 | 0.642962752 | 1.387122115 | 0.064134445 | -1.192908662 | FALSE | TRUE | FALSE |
| GALE | 0.010304 | 0.88802057 | 1 | 0.010304041 | 0.010304041 | 0.20436017 | 0.01761571 | 0.894404625 | 1 | 0.017615711 | 0.017615711 | 0.017615711 | 0.584934752 | -0.232892576 | FALSE | TRUE | FALSE |
| POLR1C | 0.0102749 | 0.88802825 | 0.75205 | 0.092408154 | 0.852995039 | 0.852995039 | 0.01863227 | 0.894171958 | 1 | 0.018632274 | 0.018632274 | 0.018632274 | 0.551457483 | -0.258487965 | FALSE | TRUE | FALSE |
| NRDE2 | 0.0102657 | 0.88803593 | 0.51655 | 0.087357301 | 0.087357301 | 1.707894599 | 1.27107968 | 0.558401246 | 0.02105 | 0.143885987 | 1.271079685 | 4.850882608 | 0.008076363 | -2.092784193 | FALSE | FALSE | FALSE |
| DBX1 | 0.0102476 | 0.8880436 | 1 | 0.010247614 | 0.010247614 | 0.203185169 | 0.01741421 | 0.8944554 | 1 | 0.069582801 | 0.069582801 | 0.345639345 | 0.588462712 | -0.230281051 | FALSE | TRUE | FALSE |
| CCDC138 | 0.0102455 | 0.88805127 | 1 | 0.088768894 | 0.088768894 | 0.346655255 | 0.15141642 | 0.864265955 | 0.5086 | 0.15141642 | 0.15141642 | 0.342564523 | 0.067664341 | -1.169640144 | FALSE | TRUE | FALSE |
| EEF1D | 0.0101956 | 0.88805895 | 1 | 5.13067E+24 | 6.02318E+31 | 1.54219E+34 | 0.01583725 | 0.894888774 | 1 | 2.1692E+27 | 3.23014E+34 | 8.9219E+36 | 0.643771937 | -0.191267959 | FALSE | TRUE | FALSE |

| | | | | | | | | | | | | | | | | | |
|---|---|---|---|---|---|---|---|---|---|---|---|---|---|---|---|---|---|
| CBLC | 0.0101617 | 0.88806662 | 1 | 0.0101617 | 0.0101617 | 0.200969118 | 0.01741263 | 0.89446265 | 1 | 0.017412628 | 0.017412628 | 0.344833272 | 0.5835822 | -0.233897963 | FALSE | TRUE | FALSE |
| LPL | 0.0101492 | 0.88807429 | 1 | 0.010149151 | 0.010149151 | 0.200995355 | 1.42503509 | 0.543299867 | 0.2558 | 0.150869115 | 1.425035094 | 1.425035094 | 0.007122036 | -2.147395864 | FALSE | FALSE | FALSE |
| SRGN | 0.010104 | 0.88808196 | 1 | 0.010104027 | 0.010104027 | 0.010104027 | 0.01037849 | 0.896652837 | 0.00335 | 0.010378492 | 0.010378492 | 0.010378492 | 0.973554465 | -0.011639747 | FALSE | TRUE | FALSE |
| POLR1A | 0.0100705 | 0.88808963 | 0.92215 | 0.367581899 | 13.7740454 | 66.28140448 | 0.00554286 | 0.897976341 | 1 | 0.022149579 | 0.088533132 | 1.710887424 | 1.816851117 | 0.25931934 | FALSE | TRUE | FALSE |
| EYA2 | 0.0100473 | 0.8880973 | 0.96825 | 0.641182358 | 2.563552872 | 10.25206791 | 0.07155464 | 0.882096185 | 0.75605 | 0.071554645 | 0.285988255 | 1.415204077 | 0.140414756 | -0.852587251 | FALSE | TRUE | FALSE |
| RNF19A | 0.010031 | 0.88810497 | 1 | 0.16020988 | 0.640508215 | 3.152749165 | 1.06945652 | 0.581116403 | 0.2716 | 0.619199326 | 9.896847348 | 12.20869401 | 0.009379528 | -2.027819019 | FALSE | FALSE | FALSE |
| FRAT2 | 0.0100221 | 0.88811263 | 0.0052 | 0.010022083 | 0.010022083 | 0.010022083 | 0.0104999 | 0.896624933 | 1 | 0.010499903 | 0.010499903 | 0.010499903 | 0.954492915 | -0.020227291 | FALSE | TRUE | FALSE |
| MROH6 | 0.0099528 | 0.8881203 | 0.50575 | 0.083637949 | 0.083637949 | 0.33681835 | 0.01617148 | 0.894809565 | 1 | 0.137630451 | 1.202258461 | 2.722386298 | 0.615451018 | -0.210806505 | FALSE | TRUE | FALSE |
| SMPD2 | 0.0099445 | 0.88812796 | 1 | 0.084976279 | 0.084976279 | 0.197177078 | 10.6568122 | 0.190807305 | 0.0657 | 1.173716955 | 99.18560015 | 99.18560015 | 0.000933154 | -3.03004651 | FALSE | FALSE | FALSE |
| BFSP1 | 0.0099273 | 0.88813563 | 1 | 706.2952128 | 768029.0966 | 8259289.465 | 1.30869311 | 0.553681629 | 0.99725 | 11837.40326 | 14013830.19 | 154878664.2 | 0.007585667 | -2.120006209 | FALSE | FALSE | FALSE |
| TRIM48 | 0.0099242 | 0.88814329 | 1 | 0.009924194 | 0.009924194 | 0.009924194 | 0.01607253 | 0.894838381 | 0.5018 | 0.13604873 | 0.13604873 | 0.13604873 | 0.61746315 | -0.209388956 | FALSE | TRUE | FALSE |
| MCC | 0.0099236 | 0.88815095 | 1 | 0.086483146 | 0.086483146 | 1.69068591 | 0.0173551 | 0.894484393 | 0.02695 | 0.017355105 | 0.017355105 | 0.340201399 | 0.571795071 | -0.242759592 | FALSE | TRUE | FALSE |
| WTAP | 0.0099031 | 0.88815861 | 1 | 0.093007551 | 0.896503272 | 0.896503272 | 0.00944792 | 0.896896419 | 0.985 | 8.397113645 | 879.256144 | 9329.301149 | 1.048176425 | 0.020434388 | FALSE | TRUE | FALSE |
| LYVE1 | 0.0098977 | 0.88816627 | 1 | 0.090030364 | 0.090030364 | 0.090030364 | 0.10657855 | 0.874213277 | 0.75655 | 0.106578553 | 0.999025126 | 0.999025126 | 0.092867932 | -1.032134225 | FALSE | TRUE | FALSE |
| ADAMTS17 | 0.0098751 | 0.88817393 | 0.76335 | 0.009875062 | 0.039471506 | 0.332733727 | 0.02012764 | 0.893813897 | 0.5211 | 0.02012764 | 0.02012764 | 0.394146724 | 0.490621969 | -0.309253009 | FALSE | TRUE | FALSE |
| CYP4A22 | 0.0098291 | 0.88818158 | 1 | 0.083746627 | 0.083746627 | 0.194455082 | 0.01704085 | 0.894585756 | 0.01395 | 0.017040849 | 0.017040849 | 0.33756795 | 0.576797092 | -0.238976938 | FALSE | TRUE | FALSE |
| ARHGEF40 | 0.0098174 | 0.88818924 | 0.0457 | 0.009817402 | 0.009817402 | 0.329130627 | 0.01659965 | 0.894715814 | 0.04255 | 0.016599655 | 0.016599655 | 0.557442391 | 0.591422073 | -0.228102471 | FALSE | TRUE | FALSE |
| TYW1 | 0.0098151 | 0.8881969 | 0.5159 | 0.039229742 | 0.039229742 | 0.772904977 | 0.01705519 | 0.894578522 | 1 | 0.017055186 | 0.017055186 | 0.336624125 | 0.575491102 | -0.239961387 | FALSE | TRUE | FALSE |
| D2HGDH | 0.0097751 | 0.88820455 | 0.01725 | 0.009775088 | 0.009775088 | 0.192714857 | 0.26058754 | 0.836538959 | 0.1369 | 0.065192649 | 1.041878778 | 2.209871804 | 0.037511726 | -1.425832953 | FALSE | TRUE | FALSE |
| SPTLC1 | 0.0097511 | 0.8882122 | 0.75795 | 0.009751062 | 0.038973499 | 0.1932009 | 4.79998229 | 0.315057861 | 0.11915 | 0.300032748 | 4.799982291 | 19.20603943 | 0.002031479 | -2.692187736 | FALSE | FALSE | FALSE |
| BIVM | 0.0097491 | 0.88821986 | 1 | 0.009749081 | 0.009749081 | 0.193236771 | 0.01068398 | 0.896576069 | 0.0118 | 0.010683979 | 0.010683979 | 0.211995332 | 0.912495345 | -0.039769342 | FALSE | TRUE | FALSE |
| PMS1 | 0.009745 | 0.88822751 | 0.7253 | 0.03893216 | 0.621850991 | 3.063243597 | 0.01365674 | 0.895582563 | 0.80185 | 0.218222096 | 7.514565909 | 30.05735863 | 0.713567724 | -0.146564802 | FALSE | TRUE | FALSE |
| EIF2B4 | 0.0097428 | 0.88823516 | 1 | 0.009742804 | 0.009742804 | 0.192585999 | 0.06577606 | 0.883419846 | 0.75295 | 0.065776056 | 0.262913946 | 1.302017716 | 0.148120824 | -0.829383882 | FALSE | TRUE | FALSE |
| KRT2 | 0.0097225 | 0.88824281 | 0.01735 | 0.009722479 | 0.009722479 | 0.191792663 | 0.01679638 | 0.894643595 | 0.01655 | 0.016796381 | 0.016796381 | 0.331882407 | 0.578843676 | -0.237438707 | FALSE | TRUE | FALSE |
| DCHS1 | 0.0097127 | 0.88825046 | 0.0913 | 0.009712661 | 0.080657607 | 0.322335687 | 0.00498579 | 0.898126027 | 1 | 0.004985786 | 0.093772052 | 0.16594073 | 1.948070217 | 0.289604607 | FALSE | TRUE | FALSE |
| PROM1 | 0.0097026 | 0.8882581 | 1 | 0.082662241 | 0.082662241 | 1.628144418 | 0.01684858 | 0.894636368 | 0.0207 | 0.016848579 | 0.016848579 | 0.332461943 | 0.575871919 | -0.239674098 | FALSE | TRUE | FALSE |
| MS4A13 | 0.0096397 | 0.88826575 | 1 | 0.009639706 | 0.009639706 | 0.009639706 | 0.01028149 | 0.896687697 | 1 | 0.010281494 | 0.010281494 | 0.010281494 | 0.937578382 | -0.027992415 | FALSE | TRUE | FALSE |
| MTMR11 | 0.0096012 | 0.8882734 | 0.1389 | 0.009601172 | 0.038386188 | 0.081131238 | 0.00568117 | 0.897949082 | 0.7528 | 0.005681168 | 0.02271076 | 0.112348415 | 1.689999788 | 0.22788665 | FALSE | TRUE | FALSE |
| CDH16 | 0.0095803 | 0.88828104 | 1 | 0.009580275 | 0.009580275 | 0.188158827 | 0.01626669 | 0.894795151 | 1 | 0.016266689 | 0.016266689 | 0.320173493 | 0.588950561 | -0.22992116 | FALSE | TRUE | FALSE |
| ABCF1 | 0.0095691 | 0.88828868 | 1 | 0.342122266 | 3.140650498 | 6.715855754 | 0.01390893 | 0.895461585 | 0.99595 | 104.4290434 | 10485.41493 | 108791.1872 | 0.687984244 | -0.162421508 | FALSE | TRUE | FALSE |
| KRT17 | 0.0095522 | 0.88829633 | 0.7504 | 0.00955219 | 0.038179094 | 0.188846184 | 0.00478813 | 0.89818036 | 1 | 0.042478091 | 0.042478091 | 0.094791958 | 1.994974497 | 0.299937348 | FALSE | TRUE | FALSE |
| KDR | 0.0095191 | 0.88830397 | 1 | 0.009519052 | 0.009519052 | 0.185398323 | 0.01649868 | 0.894737462 | 0.0338 | 0.016498675 | 0.016498675 | 0.322350929 | 0.576958603 | -0.238855346 | FALSE | TRUE | FALSE |
| SHMT1 | 0.0095147 | 0.88831161 | 1 | 0.009514742 | 0.009514742 | 0.188299982 | 0.01601505 | 0.894852784 | 1 | 0.064039173 | 0.064039173 | 0.317342943 | 0.594112593 | -0.226131242 | FALSE | TRUE | FALSE |
| BBS9 | 0.009495 | 0.88831925 | 0.26885 | 0.009494963 | 0.037947219 | 0.186934749 | 0.08293155 | 0.879559105 | 0.3847 | 0.038369854 | 0.331556399 | 0.756836804 | 0.114491562 | -0.941226519 | FALSE | TRUE | FALSE |
| UNC93A | 0.0094319 | 0.88832689 | 1 | 0.009431894 | 0.009431894 | 0.18657134 | 0.01605427 | 0.894845853 | 1 | 0.016054271 | 0.016054271 | 0.317986425 | 0.587500638 | -0.230991658 | FALSE | TRUE | FALSE |
| PTTG2 | 0.0094149 | 0.88833453 | 0.4992 | 0.009414901 | 0.086443178 | 0.086443178 | 0.01128416 | 0.8963311 | 0.0047 | 0.011284156 | 0.011284156 | 0.011284156 | 0.834346998 | -0.078653292 | FALSE | TRUE | FALSE |
| IFNA5 | 0.009414 | 0.88834216 | 1 | 0.009414046 | 0.088038251 | 0.088038251 | 0.01047078 | 0.896645862 | 1 | 0.010470783 | 0.010470783 | 0.010470783 | 0.899077572 | -0.046202836 | FALSE | TRUE | FALSE |
| HIP1R | 0.0093951 | 0.8883498 | 0.0304 | 0.009395129 | 0.009395129 | 0.183407185 | 0.12648227 | 0.869762535 | 0.884 | 1.121829349 | 10.20592246 | 21.96294072 | 0.074280207 | -1.129126893 | FALSE | TRUE | FALSE |
| SCPEP1 | 0.0093521 | 0.88835743 | 1 | 0.082627876 | 0.749297215 | 0.749297215 | 0.06740106 | 0.883010761 | 0.50935 | 0.067401061 | 0.067401061 | 0.334772482 | 0.138753153 | -0.857757139 | FALSE | TRUE | FALSE |
| CRABP2 | 0.0093483 | 0.88836507 | 0.00425 | 0.009348331 | 0.009348331 | 0.009348331 | 0.01013439 | 0.896722537 | 0.00335 | 0.01013439 | 0.01013439 | 0.01013439 | 0.92243648 | -0.03506353 | FALSE | TRUE | FALSE |
| RBP7 | 0.0093257 | 0.8883727 | 0.49165 | 0.009325683 | 0.087293479 | 0.087293479 | 0.01031533 | 0.896680727 | 0.00325 | 0.010315334 | 0.010315334 | 0.010315334 | 0.904060234 | -0.043802633 | FALSE | TRUE | FALSE |
| CCDC141 | 0.0093206 | 0.88838033 | 0.98495 | 0.64736471 | 49.65970362 | 450.7891258 | 0.17186329 | 0.85919477 | 0.517 | 1.478646373 | 13.04120676 | 117.8379015 | 0.054232492 | -1.265740438 | FALSE | TRUE | FALSE |
| SAMD1 | 0.0093037 | 0.88838796 | 1 | 63807356961 | 5.73445E+15 | 6.97998E+16 | 0.01784007 | 0.894368331 | 1 | 114606763.8 | 2.89865E+11 | 3.90448E+13 | 0.521504633 | -0.282741829 | FALSE | TRUE | FALSE |
| ACAT1 | 0.0092964 | 0.88839559 | 1 | 0.009296384 | 0.009296384 | 0.009296384 | 0.06351779 | 0.883895253 | 0.51145 | 0.063517793 | 0.063517793 | 0.063517793 | 0.146358731 | -0.834581364 | FALSE | TRUE | FALSE |
| MAP9 | 0.0092913 | 0.88840322 | 1 | 0.077844597 | 0.669439007 | 0.669439007 | 0.01608113 | 0.894831178 | 1 | 0.136720842 | 0.136720842 | 0.318987362 | 0.57777352 | -0.238242366 | FALSE | TRUE | FALSE |
| ECM1 | 0.0092864 | 0.88841085 | 0.01645 | 0.00928645 | 0.00928645 | 0.183519523 | 0.01571832 | 0.894939124 | 1 | 0.062848826 | 0.062848826 | 0.532126352 | 0.59080407 | -0.228556521 | FALSE | TRUE | FALSE |
| HR | 0.009262 | 0.88841848 | 1 | 0.009262026 | 0.009262026 | 0.18078802 | 0.06338952 | 0.883921084 | 0.51145 | 0.063389522 | 0.063389522 | 1.240900932 | 0.146112887 | -0.835311479 | FALSE | TRUE | FALSE |
| SLC34A2 | 0.0092612 | 0.8884261 | 1 | 0.082548797 | 0.082548797 | 1.624236554 | 0.06505938 | 0.883584476 | 0.5054 | 0.065059383 | 0.065059383 | 0.551108612 | 0.142350579 | -0.846640762 | FALSE | TRUE | FALSE |
| FLT1 | 0.0092399 | 0.88843373 | 1 | 0.084776408 | 0.084776408 | 1.652360454 | 0.01654596 | 0.894723031 | 1 | 0.016545961 | 0.016545961 | 0.323483585 | 0.55843824 | -0.25302485 | FALSE | TRUE | FALSE |
| MFSD1 | 0.0092236 | 0.88844135 | 1 | 0.00922356 | 0.00922356 | 0.182862557 | 0.06227229 | 0.884238788 | 0.25865 | 0.062272289 | 0.248980511 | 0.309182083 | 0.148116605 | -0.82939625 | FALSE | TRUE | FALSE |
| IRF5 | 0.0092039 | 0.88844898 | 0.01625 | 0.00920392 | 0.00920392 | 0.18197893 | 0.01597375 | 0.894881578 | 1 | 0.015973748 | 0.015973748 | 0.316264639 | 0.576190354 | -0.239434017 | FALSE | TRUE | FALSE |
| TMIGD2 | 0.0091997 | 0.8884566 | 0.00825 | 0.009199685 | 0.009199685 | 0.009199685 | 0.01132224 | 0.896317069 | 0.0073 | 0.011322243 | 0.011322243 | 0.011322243 | 0.812532083 | -0.090159482 | FALSE | TRUE | FALSE |
| SLC12A5 | 0.0091632 | 0.88846422 | 1 | 0.036608684 | 0.036608684 | 0.712873275 | 0.01499622 | 0.895111417 | 1 | 0.563716117 | 5.442814442 | 19.03647958 | 0.611031021 | -0.213936741 | FALSE | TRUE | FALSE |
| LILRB4 | 0.0091387 | 0.88847184 | 1 | 0.009138724 | 0.009138724 | 0.181102747 | 0.01180554 | 0.896176562 | 1 | 0.105825252 | 0.105825252 | 0.234208594 | 0.774104716 | -0.111200286 | FALSE | TRUE | FALSE |
| CKMT2 | 0.0091122 | 0.88847946 | 0.50585 | 0.036439138 | 0.036439138 | 0.180317064 | 4.49634422 | 0.330519973 | 0.12915 | 0.506753608 | 4.496344218 | 4.496344218 | 0.002026572 | -2.693237944 | FALSE | FALSE | FALSE |
| TALDO1 | 0.0091 | 0.88848708 | 1 | 0.580899544 | 2.322757861 | 9.28999162 | 0.06545296 | 0.88345454 | 0.75215 | 0.065452959 | 0.26162358 | 0.26162358 | 0.139031817 | -0.8568858 | FALSE | TRUE | FALSE |
| HEATR5B | 0.009079 | 0.88849469 | 0.928 | 0.310449686 | 10.89868971 | 52.36156611 | 0.01759576 | 0.894411881 | 0.95465 | 1.346795826 | 114.1099712 | 456.4095648 | 0.515977274 | -0.287369426 | FALSE | TRUE | FALSE |
| SERPINA3 | 0.0090161 | 0.88850231 | 1 | 0.07947382 | 0.07947382 | 0.178858076 | 0.09689594 | 0.87673827 | 0.99995 | 29918.74604 | 33947426.61 | 369917826.2 | 0.093049725 | -1.031284905 | FALSE | TRUE | FALSE |
| ATP8A2 | 0.0089945 | 0.88850993 | 0.879 | 0.035951018 | 0.3191576 | 1.560533142 | 0.07462461 | 0.881482217 | 0.14955 | 0.074624606 | 0.298381881 | 1.462428965 | 0.120530545 | -0.918902881 | FALSE | TRUE | FALSE |
| PDZD8 | 0.0089679 | 0.88851754 | 1 | 3.220350963 | 126.7789236 | 506.7918645 | 0.01634874 | 0.894766314 | 1 | 0.612964408 | 23.59286294 | 47.90944405 | 0.548536424 | -0.260794529 | FALSE | TRUE | FALSE |
| PTPN14 | 0.0089646 | 0.88852515 | 1 | 0.082774015 | 0.784425678 | 5.864467295 | 0.0166367 | 0.894694159 | 0.0358 | 0.016636705 | 0.016636705 | 0.560399389 | 0.538843032 | -0.268537729 | FALSE | TRUE | FALSE |
| RGS22 | 0.00896 | 0.88853277 | 0.51625 | 0.008960041 | 0.008960041 | 0.175672593 | 0.01922373 | 0.894026075 | 0.51065 | 0.019223733 | 0.019223733 | 0.37779855 | 0.466092655 | -0.331527741 | FALSE | TRUE | FALSE |
| ETFDH | 0.0089588 | 0.88854038 | 0.8819 | 0.324452382 | 28.77241539 | 114.9404066 | 0.04293281 | 0.888343756 | 0.88335 | 1.529674393 | 131.3583555 | 525.1215449 | 0.208671185 | -0.680537517 | FALSE | TRUE | FALSE |
| ZCWPW1 | 0.0089371 | 0.88854799 | 1 | 0.008937078 | 0.008937078 | 0.176716847 | 0.01478105 | 0.895161575 | 1 | 0.126975737 | 0.126975737 | 0.292670624 | 0.604630813 | -0.218509724 | FALSE | TRUE | FALSE |
| ALOX12 | 0.0089346 | 0.8885556 | 1 | 0.008934608 | 0.008934608 | 0.176220245 | 0.01537398 | 0.895039691 | 0.0157 | 0.015373979 | 0.015373979 | 0.303730899 | 0.581151314 | -0.235710776 | FALSE | TRUE | FALSE |
| VPS33B | 0.0089121 | 0.8885632 | 0.0179 | 0.008912073 | 0.008912073 | 0.175945155 | 0.06081282 | 0.884580438 | 0.01645 | 0.060812821 | 0.060812821 | 0.514705477 | 0.146549247 | -0.834016409 | FALSE | TRUE | FALSE |
| ZMYND8 | 0.0088866 | 0.88857081 | 1 | 0.794161931 | 78.5437018 | 151.9905872 | 0.01553876 | 0.89498943 | 1 | 1.394355903 | 138.4685753 | 268.2370191 | 0.571896503 | -0.242682559 | FALSE | TRUE | FALSE |
| NFASC | 0.0088593 | 0.88857842 | 1 | 0.008859278 | 0.008859278 | 0.171767788 | 0.01547107 | 0.895010976 | 1 | 0.015471075 | 0.015471075 | 0.301066395 | 0.572634904 | -0.242122183 | FALSE | TRUE | FALSE |
| POLR3A | 0.0088589 | 0.88858602 | 0.0365 | 0.002216348 | 0.008858906 | 0.172168772 | 0.02055051 | 0.893703847 | 0.6959 | 0.020550507 | 0.32864459 | 1.602405029 | 0.431079671 | -0.365442458 | FALSE | TRUE | FALSE |
| ARVCF | 0.0088534 | 0.88859363 | 0.51705 | 0.078025148 | 0.078025148 | 1.515983088 | 0.01529733 | 0.895061218 | 1 | 0.135955178 | 0.135955178 | 2.650615076 | 0.578756349 | -0.237504232 | FALSE | TRUE | FALSE |
| MED25 | 0.0088345 | 0.88860123 | 0.0246 | 0.00883451 | 0.00883451 | 0.173430778 | 0.13105755 | 0.868647036 | 0.02035 | 0.131057554 | 0.131057554 | 2.578513328 | 0.06740939 | -1.171279606 | FALSE | TRUE | FALSE |
| PITRM1 | 0.0087988 | 0.88860883 | 1 | 0.075794099 | 0.075794099 | 1.485221433 | 1.13937197 | 0.573550321 | 0.25925 | 0.127580432 | 1.139371968 | 4.309071271 | 0.007722528 | -2.112240526 | FALSE | FALSE | FALSE |
| F13A1 | 0.0087574 | 0.88861644 | 1 | 0.139911691 | 0.559445093 | 2.753227426 | 0.0617043 | 0.884392759 | 0.26365 | 0.061704302 | 0.246692541 | 1.216588086 | 0.141924467 | -0.847942728 | FALSE | TRUE | FALSE |

| | | | | | | | | | | | | | | | | | |
|---|---|---|---|---|---|---|---|---|---|---|---|---|---|---|---|---|---|
| SPATC1 | 0.0087418 | 0.88862404 | 0.9398 | 0.074084389 | 5.739191331 | 5.739191331 | 0.01573075 | 0.894924743 | 0.694 | 0.135373114 | 10.81279296 | 10.81279296 | 0.555712529 | -0.255149811 | FALSE | TRUE | FALSE |
| OR2T12 | 0.008712 | 0.88863164 | 1 | 1.41226E+28 | 2.16932E+35 | 6.0442E+37 | 0.48665701 | 0.760453527 | 1 | 1.30748E+40 | 9.96065E+48 | 7.66353E+52 | 0.01790165 | -1.747106934 | FALSE | TRUE | FALSE |
| UFM1 | 0.0086881 | 0.88863924 | 1 | 0.008688118 | 0.008688118 | 0.008688118 | 0.00963446 | 0.896847786 | 1 | 0.009634456 | 0.009634456 | 0.009634456 | 0.901775659 | -0.044901492 | FALSE | TRUE | FALSE |
| DIS3L | 0.008687 | 0.88864683 | 0.51885 | 0.008687008 | 0.008687008 | 0.170342008 | 0.00410785 | 0.898295658 | 1 | 0.139373751 | 1.213828681 | 2.739392555 | 2.114731218 | 0.325255176 | FALSE | TRUE | FALSE |
| CGREF1 | 0.0086541 | 0.88865443 | 1 | 0.0730114 | 0.0730114 | 0.0730114 | 9.35130619 | 0.208087221 | 0.1246 | 0.277247053 | 9.35130619 | 9.35130619 | 0.00092544 | -3.033651871 | FALSE | FALSE | FALSE |
| USP28 | 0.0086444 | 0.88866203 | 1 | 0.008644421 | 0.008644421 | 0.291009914 | 0.01444938 | 0.895304634 | 1 | 0.125971361 | 0.125971361 | 2.477548583 | 0.598255436 | -0.223113346 | FALSE | TRUE | FALSE |
| SEMA3G | 0.0085724 | 0.88866962 | 1 | 0.073600504 | 0.073600504 | 1.441984232 | 0.91155713 | 0.596856338 | 0.264 | 0.494512939 | 7.91481379 | 9.712320618 | 0.009404179 | -2.026679098 | FALSE | TRUE | FALSE |
| WDR27 | 0.0085593 | 0.88867721 | 0.27305 | 0.008559271 | 0.034229972 | 0.288968171 | 0.30492196 | 0.823911798 | 0.26515 | 0.139643831 | 1.219200009 | 2.74893588 | 0.028070366 | -1.551751932 | FALSE | TRUE | FALSE |
| GOLGB1 | 0.0085528 | 0.88868481 | 0.89765 | 0.619709934 | 49.59001458 | 459.7585869 | 1.48538303 | 0.537326307 | 0.3379 | 0.169258354 | 13.36268631 | 49.3690051 | 0.005757963 | -2.239731131 | FALSE | FALSE | FALSE |
| ADAMTS15 | 0.0085455 | 0.8886924 | 1 | 0.008545531 | 0.008545531 | 0.166079916 | 0.05563234 | 0.885477241 | 0.87685 | 0.508922365 | 4.778273224 | 9.924460096 | 0.153607246 | -0.813588297 | FALSE | TRUE | FALSE |
| KIF20A | 0.00853 | 0.88869999 | 1 | 0.074774808 | 0.074774808 | 1.468900885 | 0.06018839 | 0.884716591 | 0.51405 | 0.060188387 | 0.060188387 | 1.184892092 | 0.141721133 | -0.848565384 | FALSE | TRUE | FALSE |
| INTS10 | 0.0085101 | 0.88870758 | 0.75305 | 0.079463402 | 0.76153129 | 1.566287666 | 1.63407883 | 0.522771536 | 0.51025 | 1.634078829 | 1.634078829 | 13.81694633 | 0.005207894 | -2.283337843 | FALSE | FALSE | FALSE |
| PCDH12 | 0.0084766 | 0.88871517 | 0.51625 | 0.074499669 | 0.074499669 | 1.45328103 | 0.01428408 | 0.895340342 | 1 | 0.126531111 | 1.150399239 | 4.269213526 | 0.593428863 | -0.226631334 | FALSE | TRUE | FALSE |
| RDH5 | 0.0084756 | 0.88872276 | 1 | 0.135466786 | 2.16735403 | 2.16735403 | 0.01488991 | 0.895140084 | 1 | 0.238026179 | 0.952037235 | 0.952037235 | 0.569217182 | -0.244721999 | FALSE | TRUE | FALSE |
| HECW2 | 0.0084614 | 0.88873034 | 1 | 0.008461415 | 0.008461415 | 0.283884026 | 0.01486092 | 0.895147248 | 1 | 0.014860922 | 0.014860922 | 0.499383493 | 0.56937353 | -0.244602727 | FALSE | TRUE | FALSE |
| ATG2B | 0.0084472 | 0.88873793 | 1 | 0.008447189 | 0.162693647 | 0.28254781 | 0.01459483 | 0.895261755 | 0.0441 | 0.014594831 | 0.014594831 | 0.489131498 | 0.578779491 | -0.237486866 | FALSE | TRUE | FALSE |
| PCDH15 | 0.0084171 | 0.88874552 | 0.6463 | 0.008417124 | 1.314138807 | 10.84980687 | 0.00065584 | 0.899667197 | 0.6996 | 0.00065584 | 0.04210457 | 0.100885344 | 12.83410217 | 1.108365492 | FALSE | TRUE | FALSE |
| NBN | 0.0083909 | 0.8887531 | 1 | 0.008390941 | 0.008390941 | 0.165745795 | 0.01452119 | 0.895290344 | 1 | 0.01452119 | 0.01452119 | 0.287262585 | 0.577841166 | -0.238191521 | FALSE | TRUE | FALSE |
| PEAK1 | 0.0083586 | 0.88876068 | 1 | 0.075768551 | 0.075768551 | 1.467740461 | 0.13352273 | 0.868093399 | 0.0393 | 0.133522727 | 0.133522727 | 2.596322411 | 0.062600848 | -1.203419786 | FALSE | TRUE | FALSE |
| NRCAM | 0.0083253 | 0.88876827 | 1 | 0.008325293 | 0.008325293 | 0.162738543 | 0.01415534 | 0.895376029 | 1 | 0.128430386 | 0.128430386 | 2.517334017 | 0.588137824 | -0.230520889 | FALSE | TRUE | FALSE |
| EHBP1L1 | 0.0083115 | 0.88877585 | 0.98465 | 0.607701014 | 21.59630791 | 86.39131506 | 2.11965898 | 0.478964296 | 0.9307 | 74.4394403 | 2898.161791 | 11608.06345 | 0.003921172 | -2.406584069 | FALSE | FALSE | FALSE |
| MX2 | 0.0082859 | 0.88878343 | 0.02185 | 0.008285908 | 0.008285908 | 0.163094409 | 0.00424045 | 0.89824821 | 0.50985 | 0.016948256 | 0.016948256 | 0.14340383 | 1.954017963 | 0.290928552 | FALSE | TRUE | FALSE |
| CFI | 0.0082387 | 0.88879101 | 0.7509 | 0.008238694 | 0.032925554 | 0.163038586 | 0.03717975 | 0.889646361 | 0.51035 | 0.037179748 | 0.148570447 | 0.329768278 | 0.221590897 | -0.654448084 | FALSE | TRUE | FALSE |
| NEDD4 | 0.0082208 | 0.88879859 | 0.27725 | 0.008220808 | 0.032860718 | 0.276556797 | 0.00850216 | 0.897166612 | 0.63195 | 0.033996472 | 0.290305777 | 1.424731489 | 0.966908095 | -0.014614804 | FALSE | TRUE | FALSE |
| AGER | 0.0082127 | 0.88880616 | 0.7541 | 0.06919846 | 0.276756996 | 0.650973343 | 0.01351633 | 0.895632302 | 1 | 0.461172069 | 16.15450543 | 16.15450543 | 0.607611085 | -0.216374312 | FALSE | TRUE | FALSE |
| GFAP | 0.0082039 | 0.88881374 | 1 | 0.075475288 | 0.075475288 | 0.162037192 | 0.01490442 | 0.895132918 | 1 | 0.014904419 | 0.014904419 | 0.29482142 | 0.550433849 | -0.259294867 | FALSE | TRUE | FALSE |
| ASXL1 | 0.0081342 | 0.88882132 | 1 | 0.008134175 | 0.008134175 | 0.27329283 | 0.01361278 | 0.895596779 | 1 | 0.117148694 | 0.117148694 | 2.278921404 | 0.597539742 | -0.223633205 | FALSE | TRUE | FALSE |
| YLPM1 | 0.0081218 | 0.88882889 | 1 | 0.008121809 | 0.155587437 | 0.2704156 | 0.01309495 | 0.895759997 | 0.52795 | 0.115294664 | 0.252133855 | 2.21991598 | 0.620224501 | -0.207451082 | FALSE | TRUE | FALSE |
| TGS1 | 0.0081046 | 0.88883646 | 0.0212 | 0.008104595 | 0.008104595 | 0.159758584 | 0.01392321 | 0.89545446 | 0.0168 | 0.013923215 | 0.013923215 | 0.274931271 | 0.582092254 | -0.23500818 | FALSE | TRUE | FALSE |
| MYBPC1 | 0.0080937 | 0.88884404 | 1 | 0.073124217 | 0.292264871 | 1.430608416 | 0.01483622 | 0.895154412 | 0.02425 | 0.014836217 | 0.014836217 | 0.291019085 | 0.545539833 | -0.263173534 | FALSE | TRUE | FALSE |
| RADIL | 0.0080895 | 0.88885161 | 0.76225 | 0.06824714 | 0.590983632 | 2.304441854 | 0.0146927 | 0.895211687 | 1 | 0.0146927 | 0.0146927 | 0.28723706 | 0.550576889 | -0.259182022 | FALSE | TRUE | FALSE |
| CTSA | 0.0080884 | 0.88885918 | 0.50765 | 0.073225396 | 0.073225396 | 0.159959671 | 0.01466746 | 0.89523315 | 0.01395 | 0.014667458 | 0.014667458 | 0.290459089 | 0.551454126 | -0.25849061 | FALSE | TRUE | FALSE |
| TTF2 | 0.0080661 | 0.88886675 | 0.51705 | 0.008066074 | 0.039484508 | 0.157813687 | 0.02147429 | 0.893387209 | 0.86085 | 0.182024525 | 2.910828561 | 14.27701964 | 0.375615458 | -0.425256544 | FALSE | TRUE | FALSE |
| TDRD3 | 0.0080394 | 0.88887432 | 1 | 0.073833178 | 0.073833178 | 0.271648582 | 0.05784731 | 0.88508113 | 0.51025 | 0.057847314 | 0.057847314 | 0.48948527 | 0.138976974 | -0.857057148 | FALSE | TRUE | FALSE |
| SLC39A12 | 0.0080079 | 0.88888189 | 1 | 0.00800786 | 0.00800786 | 0.158335957 | 0.1118597 | 0.872913908 | 0.75645 | 0.111859695 | 0.985043679 | 2.214779454 | 0.071588427 | -1.145157183 | FALSE | TRUE | FALSE |
| PPIP5K2 | 0.0079907 | 0.88888945 | 0.5159 | 0.06775121 | 0.06775121 | 1.324803021 | 0.11822753 | 0.87165189 | 0.5115 | 0.11822753 | 0.11822753 | 2.318025262 | 0.067587408 | -1.17013421 | FALSE | TRUE | FALSE |
| PXDNL | 0.0079387 | 0.88889702 | 0.1594 | 0.007938664 | 0.067698116 | 0.153864001 | 0.00033568 | 0.899878148 | 1 | 0.002768439 | 0.023435523 | 0.455911555 | 23.64966968 | 1.373825079 | FALSE | TRUE | FALSE |
| SVIL | 0.0079298 | 0.88890459 | 0.0686 | 0.007929811 | 0.150832268 | 0.264731809 | 0.12159916 | 0.87090229 | 0.5296 | 0.121599162 | 0.258615927 | 2.326611656 | 0.065212708 | -1.185667762 | FALSE | TRUE | FALSE |
| RHBDF1 | 0.0079108 | 0.88891215 | 1 | 0.007910754 | 0.007910754 | 0.15443851 | 0.01388157 | 0.895475831 | 1 | 0.013881571 | 0.013881571 | 0.271783835 | 0.56987454 | -0.244220746 | FALSE | TRUE | FALSE |
| SKA2 | 0.0079091 | 0.88891971 | 1 | 0.007909122 | 0.007909122 | 0.007909122 | 0.00900784 | 0.897021285 | 0.00315 | 0.009007837 | 0.009007837 | 0.009007837 | 0.878026718 | -0.056492268 | FALSE | TRUE | FALSE |
| GIGYF2 | 0.0078965 | 0.88892727 | 1 | 0.031552281 | 0.031552281 | 0.615971155 | 0.50018174 | 0.755116162 | 0.39375 | 0.214389175 | 1.999338878 | 9.792772644 | 0.015787228 | -1.801694126 | FALSE | TRUE | FALSE |
| PRG4 | 0.0078914 | 0.88893484 | 1 | 48.26105348 | 4374.659936 | 43129.56547 | 0.01340564 | 0.89568909 | 1 | 85.78140864 | 7952.841348 | 79292.42145 | 0.588661139 | -0.230134633 | FALSE | TRUE | FALSE |
| SPEF2 | 0.0078781 | 0.8889424 | 0.96905 | 0.563587605 | 10.92135986 | 44.5281498 | 1.28329455 | 0.556626477 | 0.5185 | 1.283294547 | 24.96126962 | 104.8506814 | 0.006138947 | -2.211906114 | FALSE | FALSE | FALSE |
| SPTAN1 | 0.007871 | 0.88894996 | 1 | 0.070821348 | 1.340497179 | 12.3794455 | 0.05644932 | 0.885393164 | 0.76055 | 0.127920682 | 0.511492859 | 9.744227682 | 0.139435652 | -0.855626169 | FALSE | TRUE | FALSE |
| ITGA6 | 0.007867 | 0.88895751 | 0.03075 | 0.007866964 | 0.007866964 | 0.265196357 | 0.0122761 | 0.896007484 | 0.9343 | 1.005062518 | 91.07758333 | 91.07758333 | 0.640835923 | -0.193253151 | FALSE | TRUE | FALSE |
| NLRC4 | 0.0078564 | 0.88896507 | 1 | 0.066799236 | 0.582967244 | 2.254576376 | 0.01437871 | 0.89531892 | 1 | 0.014378714 | 0.014378714 | 0.283032112 | 0.546389588 | -0.262497585 | FALSE | TRUE | FALSE |
| COL21A1 | 0.0078531 | 0.88897263 | 0.9796 | 0.501925623 | 8.033700637 | 32.15262209 | 48.3525413 | 0.056577052 | 0.28565 | 12.09473859 | 1805.920037 | 7223.344415 | 0.000162412 | -3.789380882 | FALSE | FALSE | FALSE |
| SHH | 0.0078407 | 0.88898018 | 1 | 0.031339964 | 0.031339964 | 0.15528536 | 0.01372242 | 0.895554122 | 1 | 0.05485419 | 0.05485419 | 0.272090564 | 0.571381553 | -0.243073785 | FALSE | TRUE | FALSE |
| MCF2L | 0.0078244 | 0.88898774 | 0.51985 | 0.069126703 | 0.069126703 | 1.341525721 | 0.12228951 | 0.870797538 | 0.5161 | 0.122289515 | 0.122289515 | 2.381731702 | 0.063982565 | -1.193938353 | FALSE | TRUE | FALSE |
| GP6 | 0.0078025 | 0.88899529 | 0.5127 | 0.065191729 | 0.065191729 | 0.263984254 | 0.01376734 | 0.895525665 | 1 | 0.013767345 | 0.013767345 | 0.272104043 | 0.566742837 | -0.24661396 | FALSE | TRUE | FALSE |
| ZNF254 | 0.0077998 | 0.88900284 | 0.01535 | 0.007799796 | 0.007799796 | 0.032915035 | 0.01703008 | 0.894607452 | 0.5052 | 0.017030081 | 0.017030081 | 0.041337198 | 0.458001121 | -0.339133459 | FALSE | TRUE | FALSE |
| POLR2A | 0.0077977 | 0.88901039 | 1 | 0.007797653 | 0.148653879 | 0.261318444 | 0.01382441 | 0.895511432 | 1 | 0.013824414 | 0.265037953 | 0.464089489 | 0.564049476 | -0.2486828 | FALSE | TRUE | FALSE |
| ACAN | 0.0077851 | 0.88901794 | 0.0659 | 0.007785062 | 0.148199675 | 0.260487124 | 0.01380125 | 0.895518549 | 0.05645 | 0.013801248 | 0.264255915 | 0.462670949 | 0.564083948 | -0.248656259 | FALSE | TRUE | FALSE |
| MMP3 | 0.0077259 | 0.88902549 | 1 | 0.065402754 | 0.065402754 | 0.153216768 | 0.11450259 | 0.872485527 | 0.5015 | 0.114502591 | 0.114502591 | 0.265114639 | 0.067473453 | -1.170867063 | FALSE | TRUE | FALSE |
| RSRC1 | 0.0077081 | 0.88903304 | 1 | 0.071263205 | 0.071263205 | 0.153032231 | 0.00914804 | 0.896972758 | 1 | 0.009148042 | 0.009148042 | 0.18178023 | 0.842594786 | -0.074381233 | FALSE | TRUE | FALSE |
| VRK2 | 0.0076999 | 0.88904059 | 0.00995 | 0.007699879 | 0.007699879 | 0.007699879 | 0.01293548 | 0.895809579 | 1 | 0.110697396 | 0.110697396 | 0.110697396 | 0.595252787 | -0.225298563 | FALSE | TRUE | FALSE |
| TAS2R40 | 0.0076923 | 0.88904813 | 0.0072 | 0.007692325 | 0.007692325 | 0.007692325 | 0.00845083 | 0.897187342 | 1 | 0.008450829 | 0.008450829 | 0.008450829 | 0.910245047 | -0.040841676 | FALSE | TRUE | FALSE |
| DHX16 | 0.0076832 | 0.88905568 | 1 | 0.574092222 | 5.157030542 | 11.17298625 | 0.01453422 | 0.895283198 | 0.0349 | 0.014534217 | 0.014534217 | 0.489295601 | 0.528628246 | -0.276849635 | FALSE | TRUE | FALSE |
| MAN2A1 | 0.0076744 | 0.88906322 | 1 | 0.007674377 | 0.007674377 | 0.258403262 | 0.0537885 | 0.88592102 | 0.5158 | 0.0537885 | 0.0537885 | 1.056068898 | 0.142676904 | -0.845646324 | FALSE | TRUE | FALSE |
| RARS2 | 0.0076295 | 0.88907077 | 0.0154 | 0.007629473 | 0.007629473 | 0.150945541 | 0.7416534 | 0.645472045 | 0.8673 | 11.8747152 | 549.8253321 | 2199.791955 | 0.010287114 | -1.987706436 | FALSE | TRUE | FALSE |
| TTC21A | 0.0076083 | 0.88907831 | 0.0352 | 0.007608281 | 0.007608281 | 0.148370695 | 0.00200782 | 0.899082687 | 1 | 0.017162071 | 0.017162071 | 0.335686999 | 3.789331044 | 0.578562548 | FALSE | TRUE | FALSE |
| KIF15 | 0.0075831 | 0.88908585 | 0.70225 | 0.015695772 | 0.250968576 | 0.594181918 | 0.00969942 | 0.896819976 | 0.512 | 0.009699418 | 0.038773973 | 0.326516696 | 0.781812461 | -0.106897412 | FALSE | TRUE | FALSE |
| ABLIM2 | 0.0075622 | 0.88909339 | 0.514 | 0.06818734 | 0.06818734 | 1.343837221 | 4.10294371 | 0.350256666 | 0.5016 | 4.102943711 | 156.3498587 | 625.226409 | 0.001843108 | -2.734449212 | FALSE | FALSE | FALSE |
| GARNL3 | 0.0075513 | 0.88910093 | 0.0275 | 0.007551254 | 0.007551254 | 0.148064402 | 0.01343163 | 0.895667801 | 1 | 0.013431635 | 0.013431635 | 0.26398821 | 0.562199205 | -0.250109773 | FALSE | TRUE | FALSE |
| TTC7A | 0.0075441 | 0.88910847 | 0.93765 | 0.255228665 | 4.085780004 | 5.009944639 | 0.05502412 | 0.885636696 | 0.0225 | 0.055024118 | 0.055024118 | 1.082492578 | 0.137105188 | -0.86294611 | FALSE | TRUE | FALSE |
| LSMEM1 | 0.0075306 | 0.88911601 | 1 | 0.007530623 | 0.007530623 | 0.007530623 | 0.00885194 | 0.89706977 | 1 | 0.008851937 | 0.008851937 | 0.008851937 | 0.850731671 | -0.070207399 | FALSE | TRUE | FALSE |
| PYDC1 | 0.0075104 | 0.88912355 | 0.0047 | 0.007510351 | 0.007510351 | 0.007510351 | 0.00812306 | 0.89726329 | 1 | 0.008123057 | 0.008123057 | 0.008123057 | 0.924572059 | -0.034059235 | FALSE | TRUE | FALSE |
| WRN | 0.007446 | 0.88913108 | 0.9991 | 340.7275143 | 280919.8415 | 2749979.813 | 5.3299E-05 | 0.900330083 | 0.9847 | 0.860150384 | 65.01996305 | 585.8908522 | 139.7028497 | 2.145205265 | FALSE | TRUE | FALSE |
| LLGL2 | 0.0074433 | 0.88913862 | 1 | 0.029759171 | 0.029759171 | 0.578249362 | 0.01273604 | 0.895837893 | 1 | 0.112829296 | 0.451128253 | 2.199901905 | 0.584429882 | -0.233267587 | FALSE | TRUE | FALSE |
| THNSL1 | 0.007434 | 0.88914615 | 1 | 0.064979275 | 0.582966861 | 1.28370707 | 10902.7004 | 0.000187635 | 0.428 | 4017.864977 | 1732217.248 | 17597052.09 | 6.81846E-07 | -6.166313887 | FALSE | FALSE | FALSE |
| APLP2 | 0.0074279 | 0.88915368 | 0.7517 | 0.02967393 | 0.118575571 | 0.583714364 | 0.01341699 | 0.895681995 | 1 | 0.053608423 | 0.053608423 | 1.056617154 | 0.553615544 | -0.256791725 | FALSE | TRUE | FALSE |
| OSBPL1A | 0.0074126 | 0.88916122 | 0.96745 | 0.573144574 | 9.165223342 | 36.66446061 | 0.11668833 | 0.872064819 | 0.5105 | 0.466591053 | 4.19865276 | 4.19865276 | 0.063524887 | -1.197056098 | FALSE | TRUE | FALSE |
| GYS1 | 0.0074067 | 0.88916875 | 1 | 0.029599288 | 0.029599288 | 0.58143924 | 0.18946122 | 0.854447006 | 0.93975 | 4.033658887 | 110.3906038 | 933.7355689 | 0.039093546 | -1.40789494 | FALSE | TRUE | FALSE |

| | | | | | | | | | | | | | | | | | |
|---|---|---|---|---|---|---|---|---|---|---|---|---|---|---|---|---|---|
| KIR3DL1 | 0.0074057 | 0.88917628 | 1 | 47632.08061 | 55920539.41 | 615771237.5 | 0.01292048 | 0.895816659 | 0.9957 | 92.12914143 | 9015.53599 | 92347.11988 | 0.573178573 | -0.241710053 | FALSE | TRUE | FALSE |
| IFNL1 | 0.0074023 | 0.8891838 | 0.00495 | 0.007402327 | 0.007402327 | 0.007402327 | 0.00858871 | 0.8971597 | 1 | 0.008588706 | 0.008588706 | 0.008588706 | 0.861867598 | -0.064559446 | FALSE | TRUE | FALSE |
| SAMHD1 | 0.0073959 | 0.88919133 | 0.5073 | 0.066982175 | 0.066982175 | 0.249872794 | 0.01355827 | 0.895603885 | 0.0172 | 0.013558272 | 0.013558272 | 0.26795777 | 0.545490932 | -0.263212464 | FALSE | TRUE | FALSE |
| NAV3 | 0.00738 | 0.88919886 | 0.0608 | 0.007380021 | 0.141426851 | 0.246642031 | 0.11619763 | 0.872167663 | 0.0533 | 0.11619763 | 0.245296659 | 2.237996775 | 0.063512664 | -1.197139669 | FALSE | TRUE | FALSE |
| RNF168 | 0.0073356 | 0.88920639 | 0.7599 | 0.061858449 | 0.535415088 | 1.222857533 | 0.01347746 | 0.895653604 | 0.01425 | 0.013477457 | 0.013477457 | 0.266801829 | 0.544284344 | -0.264174158 | FALSE | TRUE | FALSE |
| DNM2 | 0.0073194 | 0.88921391 | 0.50855 | 0.029239121 | 0.029239121 | 0.571297422 | 12.7901086 | 0.171877183 | 0.0436 | 0.799856281 | 3.920544727 | 15.67165277 | 0.000572268 | -3.242400563 | FALSE | FALSE | FALSE |
| NLRC5 | 0.0072877 | 0.88922144 | 0.0503 | 0.007287719 | 0.14047427 | 0.244784463 | 0.0128112 | 0.895823738 | 1 | 0.012811203 | 0.012811203 | 0.430937109 | 0.568855199 | -0.244998268 | FALSE | TRUE | FALSE |
| ATP11B | 0.0072378 | 0.88922896 | 1 | 0.0289266 | 0.115636519 | 0.974119459 | 0.05060793 | 0.886510777 | 0.5161 | 0.115174577 | 0.460353043 | 2.263239327 | 0.143017702 | -0.844610203 | FALSE | TRUE | FALSE |
| HAGH | 0.007224 | 0.88923648 | 0.50305 | 0.028864148 | 0.028864148 | 0.028864148 | 0.05154755 | 0.886303723 | 0.0082 | 0.05154755 | 0.05154755 | 0.05154755 | 0.140143388 | -0.853427388 | FALSE | TRUE | FALSE |
| RSF1 | 0.0072157 | 0.889244 | 1 | 0.007215706 | 0.007215706 | 0.24230108 | 0.40872899 | 0.788953892 | 0.2696 | 0.11490045 | 1.114841216 | 3.864059385 | 0.01765401 | -1.753156633 | FALSE | TRUE | FALSE |
| NFAM1 | 0.0071989 | 0.88925152 | 1 | 0.007198873 | 0.007198873 | 0.007198873 | 0.00808318 | 0.897277088 | 1 | 0.008083179 | 0.008083179 | 0.008083179 | 0.890599231 | -0.050317684 | FALSE | TRUE | FALSE |
| ALDH6A1 | 0.0071975 | 0.88925904 | 0.51085 | 0.0287636 | 0.0287636 | 0.143494591 | 0.01268497 | 0.895852045 | 0.7509 | 0.050698467 | 0.202678979 | 0.251421755 | 0.567406077 | -0.246106017 | FALSE | TRUE | FALSE |
| FAM186B | 0.0071912 | 0.88926656 | 0.51315 | 0.060339906 | 0.060339906 | 1.18514698 | 0.87377003 | 0.601943261 | 0.51425 | 0.873770026 | 7.812098683 | 17.19894728 | 0.008230136 | -2.084593009 | FALSE | TRUE | FALSE |
| LMNA | 0.0071762 | 0.88927408 | 0.75925 | 0.066689903 | 0.26643604 | 1.30907658 | 0.7696292 | 0.632641249 | 0.6815 | 1.792008859 | 68.43876321 | 273.664142 | 0.009324265 | -2.030385403 | FALSE | TRUE | FALSE |
| SLC22A10 | 0.0071744 | 0.88928159 | 0.8789 | 0.544705876 | 4.932238503 | 4.932238503 | 0.10516911 | 0.874670569 | 0.9387 | 3.750316038 | 80.44794893 | 321.6783844 | 0.068217898 | -1.166101667 | FALSE | TRUE | FALSE |
| USP53 | 0.007137 | 0.88928911 | 1 | 0.060516308 | 0.060516308 | 1.188055597 | 0.10703108 | 0.87408346 | 0.0226 | 0.107031083 | 0.107031083 | 2.105888742 | 0.0666811 | -1.175997244 | FALSE | TRUE | FALSE |
| MALL | 0.0070975 | 0.88929662 | 0.00425 | 0.007097451 | 0.007097451 | 0.007097451 | 0.00719666 | 0.897483651 | 1 | 0.007196662 | 0.007196662 | 0.007196662 | 0.986214349 | -0.006028683 | FALSE | TRUE | FALSE |
| FAH | 0.007071 | 0.88930414 | 1 | 0.007070973 | 0.007070973 | 0.140086535 | 0.01228538 | 0.895993371 | 0.50625 | 0.112840948 | 0.112840948 | 0.243667762 | 0.575559719 | -0.239909608 | FALSE | TRUE | FALSE |
| N4BP2L2 | 0.0070661 | 0.88931165 | 1 | 0.504621645 | 4.431667931 | 4.431667931 | 0.10673289 | 0.874143407 | 0.25245 | 0.106732892 | 0.927705083 | 0.927705083 | 0.066203795 | -1.179117114 | FALSE | TRUE | FALSE |
| UGT2B7 | 0.0070425 | 0.88931916 | 1 | 0.007042525 | 0.007042525 | 0.139579861 | 0.10640905 | 0.874253169 | 0.5081 | 0.106409048 | 0.106409048 | 0.240075353 | 0.066183515 | -1.179250172 | FALSE | TRUE | FALSE |
| AKAP13 | 0.0070068 | 0.88932667 | 0.07525 | 0.007006814 | 0.132879946 | 0.233345859 | 0.0124287 | 0.895951011 | 0.06645 | 0.012428697 | 0.237178156 | 0.414931607 | 0.563760972 | -0.248904993 | FALSE | TRUE | FALSE |
| PXDN | 0.0070043 | 0.88933418 | 0.87985 | 0.027990905 | 0.253563438 | 1.219967851 | 0.01476796 | 0.895183057 | 0.64235 | 0.059024394 | 0.537579048 | 2.598150627 | 0.474292782 | -0.323953485 | FALSE | TRUE | FALSE |
| FGFR3 | 0.0070021 | 0.88934169 | 1 | 2.10825E+29 | 3.76335E+37 | 1.93738E+41 | 0.00239822 | 0.898902076 | 1 | 3.54013E+40 | 1.20709E+50 | 8.81258E+53 | 2.919725481 | 0.46534202 | FALSE | TRUE | FALSE |
| KDM5B | 0.0069972 | 0.8893492 | 0.00005 | 2.77415E-06 | 2.42402E-05 | 0.000203041 | 3.2028E-06 | 0.900952163 | 0.8209 | 1.27992E-05 | 0.000462744 | 0.000995829 | 2184.666057 | 3.339385061 | FALSE | TRUE | FALSE |
| ARAP2 | 0.0069624 | 0.88935671 | 0.75535 | 0.059720349 | 0.525781232 | 2.555069346 | 0.03265202 | 0.890682753 | 0.03965 | 0.032652016 | 0.032652016 | 0.636506699 | 0.213231053 | -0.671149549 | FALSE | TRUE | FALSE |
| RNF111 | 0.0069622 | 0.88936421 | 1 | 2.49796E+64 | 2.69542E+76 | 3.29585E+80 | 0.0219687 | 0.89323193 | 1 | 6.97401E+51 | 2.39149E+62 | 2.56001E+66 | 0.31691237 | -0.499060809 | FALSE | TRUE | FALSE |
| BMPR2 | 0.0069588 | 0.88937172 | 1 | 0.027798825 | 0.027798825 | 0.545109384 | 0.05008235 | 0.886593411 | 0.0252 | 0.050082351 | 0.050082351 | 0.984380497 | 0.13894698 | -0.857150888 | FALSE | TRUE | FALSE |
| ZNF85 | 0.0069415 | 0.88937922 | 0.7528 | 0.059054336 | 0.515670283 | 1.170273294 | 0.10186907 | 0.87551712 | 0.8774 | 0.899694757 | 8.150374978 | 8.150374978 | 0.068141644 | -1.166587393 | FALSE | TRUE | FALSE |
| MROH9 | 0.0068634 | 0.88938673 | 0.01375 | 0.006863445 | 0.006863445 | 0.135949807 | 0.08621074 | 0.87896718 | 0.0122 | 0.086210742 | 0.086210742 | 0.195103126 | 0.079612413 | -1.099019212 | FALSE | TRUE | FALSE |
| SCUBE3 | 0.0068368 | 0.88939423 | 1 | 1.012451833 | 38.52421951 | 154.0363359 | 0.012496 | 0.895901551 | 0.87785 | 1.107232073 | 10.82981942 | 21.69532159 | 0.54711695 | -0.261919831 | FALSE | TRUE | FALSE |
| SECISBP2L | 0.0068359 | 0.88940173 | 0.02655 | 0.006835903 | 0.006835903 | 0.134363535 | 0.01229296 | 0.895979255 | 0.02295 | 0.012292965 | 0.012292965 | 0.242126157 | 0.556082518 | -0.254860758 | FALSE | TRUE | FALSE |
| NEK9 | 0.0068331 | 0.88940923 | 1 | 0.059134661 | 0.52527122 | 1.994713342 | 0.01245571 | 0.895936884 | 1 | 0.109011773 | 0.109011773 | 2.146687055 | 0.548592661 | -0.260750007 | FALSE | TRUE | FALSE |
| PCDHGB4 | 0.006786 | 0.88941673 | 1 | 512795.2807 | 674110422.7 | 57278010237 | 0.07292704 | 0.881803269 | 1 | 2.9544E+13 | 7.60291E+17 | 1.35809E+20 | 0.093052025 | -1.031274169 | FALSE | TRUE | FALSE |
| CAMSAP2 | 0.0067735 | 0.88942423 | 1 | 9.59584E+47 | 2.29374E+58 | 8.71573E+62 | 0.01710746 | 0.894542337 | 1 | 5.80045E+41 | 4.45035E+50 | 3.43207E+54 | 0.395939541 | -0.402371125 | FALSE | TRUE | FALSE |
| TRIO | 0.0067245 | 0.88943172 | 1 | 0.006724525 | 0.12588046 | 0.223366243 | 0.01171884 | 0.896211721 | 1 | 0.107346879 | 0.390341188 | 3.575601852 | 0.573821723 | -0.241223015 | FALSE | TRUE | FALSE |
| DAZAP1 | 0.0067244 | 0.88943922 | 1 | 4.87717E+41 | 2.55031E+50 | 1.66509E+54 | 0.01465235 | 0.895240303 | 1 | 1.411E+37 | 5.78115E+45 | 3.43106E+49 | 0.458931632 | -0.338252007 | FALSE | TRUE | FALSE |
| TRMU | 0.0067109 | 0.88944672 | 0.0126 | 0.0067109 | 0.0067109 | 0.133074336 | 0.01191375 | 0.896092088 | 1 | 0.011913745 | 0.011913745 | 0.236486697 | 0.563290494 | -0.249267578 | FALSE | TRUE | FALSE |
| KRTAP8-1 | 0.0066801 | 0.88945421 | 1 | 0.006680068 | 0.006680068 | 0.006680068 | 0.00709623 | 0.897497396 | 0.00155 | 0.007096234 | 0.007096234 | 0.007096234 | 0.941353936 | -0.026247057 | FALSE | TRUE | FALSE |
| KIF6 | 0.0066759 | 0.8894617 | 0.96985 | 0.106769754 | 0.899244564 | 3.597582842 | 0.4681037 | 0.767799183 | 0.1977 | 0.117055855 | 4.097262897 | 4.097262897 | 0.014261518 | -1.845834254 | FALSE | TRUE | FALSE |
| LIPI | 0.0066643 | 0.8894692 | 1 | 0.006664318 | 0.006664318 | 0.132263012 | 0.01190151 | 0.896106176 | 1 | 0.011901506 | 0.011901506 | 0.011901506 | 0.559955886 | -0.251846186 | FALSE | TRUE | FALSE |
| KRT77 | 0.0066076 | 0.88947669 | 0.7476 | 0.014670768 | 0.058612346 | 0.130363996 | 0.02849943 | 0.891610569 | 0.9367 | 1.042087438 | 23.55211308 | 94.10828799 | 0.231850127 | -0.634792663 | FALSE | TRUE | FALSE |
| PGM1 | 0.0066071 | 0.88948418 | 1 | 7.2149E+19 | 5.52366E+26 | 1.23813E+29 | 0.07655357 | 0.881159623 | 1 | 1.14845E+15 | 4.67478E+20 | 9.23982E+22 | 0.086306509 | -1.063956451 | FALSE | TRUE | FALSE |
| ALOX5 | 0.0065984 | 0.88949167 | 0.7585 | 0.026363195 | 0.105357704 | 0.518884538 | 0.04714325 | 0.887307656 | 0.51405 | 0.188427848 | 0.753319513 | 1.616541956 | 0.139964807 | -0.853981152 | FALSE | TRUE | FALSE |
| SMC1B | 0.0065922 | 0.88949916 | 1 | 0.060268682 | 0.060268682 | 1.17951379 | 0.01227804 | 0.896000428 | 0.02865 | 0.012278044 | 0.012278044 | 0.242638079 | 0.53690602 | -0.270101726 | FALSE | TRUE | FALSE |
| MLC1 | 0.0065783 | 0.88950665 | 0.00985 | 0.006578281 | 0.006578281 | 0.006578281 | 0.0453367 | 0.887763118 | 0.9402 | 0.181220306 | 2.89765332 | 2.89765332 | 0.145098372 | -0.838337459 | FALSE | TRUE | FALSE |
| RPE65 | 0.006575 | 0.88951413 | 0.0069 | 3.72069E-05 | 0.001325956 | 0.005298698 | 1.8907E-06 | 0.901074365 | 0.6956 | 4.15871E-06 | 6.63592E-05 | 6.63592E-05 | 3477.491043 | 3.54126602 | FALSE | TRUE | FALSE |
| PDCD11 | 0.0065611 | 0.88952162 | 0.5283 | 0.026237791 | 0.126310267 | 0.505110222 | 0.01181895 | 0.896162492 | 0.04195 | 0.011818945 | 0.011818945 | 0.396932546 | 0.555138103 | -0.255598963 | FALSE | TRUE | FALSE |
| SLCO6A1 | 0.006528 | 0.8895291 | 0.0188 | 0.006527959 | 0.006527959 | 0.128753626 | 0.0111383 | 0.896422216 | 0.50785 | 0.094534326 | 0.094534326 | 0.377120767 | 0.586082381 | -0.232041334 | FALSE | TRUE | FALSE |
| KCNC3 | 0.0065271 | 0.88953659 | 0.9384 | 0.241469135 | 3.856931014 | 4.754084637 | 0.11106902 | 0.873127132 | 0.62985 | 0.190985371 | 1.773719173 | 2.190849387 | 0.05876604 | -1.230873574 | FALSE | TRUE | FALSE |
| SCFD2 | 0.0065265 | 0.88954407 | 0.50975 | 0.056184205 | 0.056184205 | 1.107654682 | 0.01159473 | 0.896239832 | 0.5099 | 0.10091393 | 0.10091393 | 0.39247266 | 0.562885909 | -0.249579623 | FALSE | TRUE | FALSE |
| SCAP | 0.0064482 | 0.88955155 | 0.5225 | 0.025774789 | 0.025774789 | 0.500504054 | 0.01161686 | 0.896225778 | 1 | 0.046438779 | 0.046438779 | 0.90492666 | 0.555073026 | -0.255649877 | FALSE | TRUE | FALSE |
| SLC28A3 | 0.0064333 | 0.88955904 | 1 | 0.006433291 | 0.006433291 | 0.127014655 | 0.01115971 | 0.896408208 | 0.50695 | 0.044636124 | 0.044636124 | 0.377774242 | 0.576474595 | -0.239219827 | FALSE | TRUE | FALSE |
| PTRHD1 | 0.0064314 | 0.88956652 | 0.4991 | 0.006431363 | 0.058808868 | 0.058808868 | 0.06765402 | 0.882949608 | 0.24745 | 0.067654016 | 0.637129573 | 0.637129573 | 0.095062548 | -1.021990551 | FALSE | TRUE | FALSE |
| CATSPER3 | 0.0063991 | 0.889574 | 1 | 0.058729606 | 0.058729606 | 0.126865711 | 0.01183261 | 0.896148419 | 0.50915 | 0.047280422 | 0.047280422 | 0.234833432 | 0.54080394 | -0.266960153 | FALSE | TRUE | FALSE |
| MOGS | 0.0063696 | 0.88958147 | 0.02585 | 0.00636958 | 0.00636958 | 0.125116621 | 0.01133895 | 0.896310052 | 0.0209 | 0.011338952 | 0.011338952 | 0.223208106 | 0.561743231 | -0.250462152 | FALSE | TRUE | FALSE |
| CTBP2 | 0.0063652 | 0.88958895 | 0.2367 | 0.000254911 | 0.101472111 | 1.048254555 | 0.00082791 | 0.899574639 | 0.83125 | 0.032864751 | 36.97416775 | 1116.337207 | 7.688267659 | 0.885828494 | FALSE | TRUE | FALSE |
| SPHKAP | 0.0063565 | 0.88959643 | 1 | 0.006356547 | 0.006356547 | 0.213473196 | 0.00835763 | 0.897201158 | 0.0367 | 0.008357629 | 0.008357629 | 0.281089943 | 0.760568185 | -0.118861845 | FALSE | TRUE | FALSE |
| MYOM2 | 0.006349 | 0.8896039 | 0.5813 | 0.021234258 | 58.79178747 | 1281.834501 | 6.1403E-06 | 0.900816747 | 0.94685 | 0.008248207 | 11.10607777 | 254.6829055 | 1033.994613 | 3.014518276 | FALSE | TRUE | FALSE |
| PCDHA6 | 0.006329 | 0.88961138 | 1 | 0.055274554 | 0.495481196 | 1.867100855 | 0.894938 | 0.599520608 | 0.1421 | 0.395450553 | 3.578256556 | 7.71389651 | 0.007072046 | -2.150454951 | FALSE | TRUE | FALSE |
| RAD54B | 0.0063187 | 0.88961885 | 0.7321 | 0.025246989 | 0.403370771 | 1.612926512 | 0.00214474 | 0.899022551 | 0.25075 | 0.00214474 | 0.008571027 | 0.008571027 | 2.946119423 | 0.469250347 | FALSE | TRUE | FALSE |
| CCDC57 | 0.0062989 | 0.88962632 | 1 | 0.006298944 | 0.006298944 | 0.123634207 | 0.08855726 | 0.87849176 | 0.2661 | 0.088557262 | 0.768218227 | 1.742076667 | 0.071128491 | -1.147956405 | FALSE | TRUE | FALSE |
| ZNF615 | 0.0062283 | 0.8896338 | 1 | 0.006228268 | 0.006228268 | 0.122940749 | 0.0110716 | 0.896443221 | 1 | 0.011071605 | 0.011071605 | 0.218887115 | 0.562544277 | -0.249843289 | FALSE | TRUE | FALSE |
| PROZ | 0.0062238 | 0.88964127 | 0.5094 | 0.024873732 | 0.024873732 | 0.123378985 | 1.44002594 | 0.542089873 | 0.64145 | 1.614199133 | 124.824391 | 499.1599958 | 0.00432204 | -2.364311249 | FALSE | FALSE | FALSE |
| WDR62 | 0.0061821 | 0.88964874 | 0.04515 | 0.006182112 | 0.006182112 | 0.208083018 | 0.01101207 | 0.896464218 | 0.03955 | 0.011012072 | 0.011012072 | 0.213607906 | 0.561394106 | -0.250732151 | FALSE | TRUE | FALSE |
| ATP13A1 | 0.0061633 | 0.88965621 | 1 | 0.006163267 | 0.006163267 | 0.207742601 | 0.1703512 | 0.859526889 | 0.4025 | 0.170351198 | 1.593572583 | 7.771241656 | 0.036179769 | -1.441534215 | FALSE | TRUE | FALSE |
| SLC45A4 | 0.0061357 | 0.88966367 | 0.7502 | 0.006135652 | 0.024516428 | 0.119750144 | 1.74533524 | 0.513306236 | 0.3428 | 0.43662606 | 16.19486903 | 64.75229311 | 0.003515458 | -2.454018129 | FALSE | FALSE | FALSE |
| WDR54 | 0.0060859 | 0.88967114 | 0.50885 | 0.05462891 | 0.05462891 | 0.05462891 | 0.00807786 | 0.897283986 | 1 | 0.008077855 | 0.008077855 | 0.008077855 | 0.753409525 | -0.122968894 | FALSE | TRUE | FALSE |
| MAN1B1 | 0.0060675 | 0.88967861 | 1 | 0.862307041 | 18.43755755 | 73.71010024 | 0.04579062 | 0.887600842 | 0.75705 | 0.410665722 | 3.780093443 | 3.780093443 | 0.132505349 | -0.87776659 | FALSE | TRUE | FALSE |
| ANO7 | 0.0060535 | 0.88968607 | 0.98425 | 0.439767142 | 35.28259206 | 327.5379665 | 0.01421219 | 0.89535462 | 0.9685 | 1.067853749 | 9.609999591 | 88.60198031 | 0.425939171 | -0.370652419 | FALSE | TRUE | FALSE |
| TEX14 | 0.0059955 | 0.88969354 | 0.32945 | 0.001457271 | 0.104825718 | 0.237600884 | 0.00062342 | 0.899706815 | 0.8749 | 0.005193193 | 0.044374822 | 0.101407827 | 9.617234376 | 0.9830502 | FALSE | TRUE | FALSE |
| KRTAP2-4 | 0.0059837 | 0.889701 | 1 | 1.00145E+22 | 8.93651E+28 | 2.08894E+31 | 0.00401342 | 0.898343065 | 1 | 2.55947E+11 | 8.53781E+15 | 1.32869E+18 | 1.490927583 | 0.17345655 | FALSE | TRUE | FALSE |
| TTC3 | 0.005977 | 0.88970846 | 0.3438 | 0.005977013 | 0.388941129 | 0.918755704 | 0.00029447 | 0.899904457 | 1 | 0.002287477 | 0.002287477 | 0.044321903 | 20.29786524 | 1.307450365 | FALSE | TRUE | FALSE |
| TBXAS1 | 0.0059607 | 0.88971593 | 0.969 | 0.106831579 | 1.707760833 | 6.830516795 | 0.00327373 | 0.89857949 | 1 | 0.027763518 | 0.110998396 | 0.25878838 | 1.82076191 | 0.260253159 | FALSE | TRUE | FALSE |

| | | | | | | | | | | | | | | | | | |
|---|---|---|---|---|---|---|---|---|---|---|---|---|---|---|---|---|---|
| FGD5 | 0.0059424 | 0.88972339 | 0.036 | 0.005942389 | 0.005942389 | 0.200268572 | 0.01089517 | 0.89651318 | 1 | 0.010895165 | 0.010895165 | 0.212730575 | 0.545415217 | -0.26327275 | FALSE | TRUE | FALSE |
| UNC13D | 0.0059298 | 0.88973085 | 0.9847 | 0.201097758 | 3.217622887 | 12.87543267 | 0.01342322 | 0.895674899 | 0.0312 | 0.013423218 | 0.013423218 | 0.262323529 | 0.441755226 | -0.354818304 | FALSE | TRUE | FALSE |
| TMC2 | 0.005928 | 0.88973831 | 0.81995 | 0.023696281 | 0.805719605 | 0.805719605 | 0.05612978 | 0.885452027 | 0.14265 | 0.026665552 | 0.224444375 | 0.474389042 | 0.105612142 | -0.976286148 | FALSE | TRUE | FALSE |
| GANC | 0.0059143 | 0.88974576 | 0.70215 | 0.045711577 | 3.186836377 | 27.602774 | 0.05522209 | 0.885611546 | 0.12215 | 2.83523E-05 | 0.055222089 | 0.882191656 | 0.107100185 | -0.970209778 | FALSE | TRUE | FALSE |
| EPB41L4A | 0.0059065 | 0.88975322 | 0.9306 | 0.199563349 | 27.67479534 | 110.6651374 | 0.34159604 | 0.811615763 | 0.2116 | 0.085446608 | 2.928262476 | 11.70944832 | 0.017290845 | -1.762183782 | FALSE | TRUE | FALSE |
| NOS3 | 0.0058834 | 0.88976068 | 1 | 0.02352146 | 0.02352146 | 0.455641387 | 0.04093313 | 0.888719887 | 0.63725 | 0.163697977 | 1.450783746 | 7.054016657 | 0.14373232 | -0.842445565 | FALSE | TRUE | FALSE |
| GNPTG | 0.0058833 | 0.88976813 | 0.75125 | 0.051442202 | 0.205626924 | 0.205626924 | 0.09497589 | 0.877159475 | 0.5078 | 0.094975887 | 0.094975887 | 0.094975887 | 0.061945118 | -1.207992914 | FALSE | TRUE | FALSE |
| ABL2 | 0.0058475 | 0.88977558 | 1 | 1.28329E+11 | 2.73709E+15 | 3.46603E+16 | 0.04312244 | 0.888295578 | 1 | 50802746390 | 1.8657E+15 | 7.48842E+16 | 0.13560292 | -0.867730959 | FALSE | TRUE | FALSE |
| ITGA2B | 0.0058187 | 0.88978304 | 0.5166 | 0.023260924 | 0.023260924 | 0.454068124 | 0.01037532 | 0.896666784 | 1 | 0.092096525 | 0.092096525 | 1.802974975 | 0.560824944 | -0.251172678 | FALSE | TRUE | FALSE |
| WNK2 | 0.0057956 | 0.88979049 | 1 | 9.31529E+27 | 1.43088E+35 | 6.79247E+38 | 517023152 | 4.28976E-09 | 1 | 1.71174E+37 | 5.07429E+45 | 3.51284E+49 | 1.12096E-11 | -10.95040979 | FALSE | FALSE | FALSE |
| ASPH | 0.0057752 | 0.88979794 | 1 | 0.005775168 | 0.005775168 | 0.113962273 | 0.01035361 | 0.896673756 | 1 | 0.010353609 | 0.010353609 | 0.20463739 | 0.557792733 | -0.253527148 | FALSE | TRUE | FALSE |
| KIF14 | 0.0057652 | 0.88980539 | 0.9783 | 1.759012538 | 148.604823 | 1248.287558 | 0.01376041 | 0.895532781 | 0.87805 | 0.055018597 | 0.481186813 | 2.349475943 | 0.418968762 | -0.377818357 | FALSE | TRUE | FALSE |
| CDH26 | 0.0057597 | 0.88981284 | 1 | 3.702846816 | 33.61141866 | 312.571502 | 0.07579843 | 0.881321109 | 0.98875 | 507.4882265 | 482408.7177 | 4960597.319 | 0.075987534 | -1.11925765 | FALSE | TRUE | FALSE |
| CASR | 0.0057331 | 0.88982029 | 1 | 0.005733111 | 0.005733111 | 0.111926534 | 0.01055314 | 0.896610976 | 0.02975 | 0.010553141 | 0.010553141 | 0.206619438 | 0.543261045 | -0.264991435 | FALSE | TRUE | FALSE |
| NME8 | 0.0057199 | 0.88982774 | 1 | 0.415254644 | 3.676902071 | 3.676902071 | 0.09227458 | 0.877720162 | 0.51535 | 0.092274579 | 0.092274579 | 0.214269831 | 0.06198796 | -1.207692656 | FALSE | TRUE | FALSE |
| VN1R2 | 0.0057067 | 0.88983518 | 0.0099 | 0.005706692 | 0.005706692 | 0.005706692 | 0.05919895 | 0.884860947 | 0.75645 | 0.05919895 | 0.555512169 | 0.555512169 | 0.096398529 | -1.015929593 | FALSE | TRUE | FALSE |
| FARP1 | 0.0056691 | 0.88984263 | 0.1141 | 0.000146959 | 0.014025469 | 0.056055331 | 1.9288E-05 | 0.900551293 | 0.02055 | 2.13442E-06 | 1.92884E-05 | 9.42871E-05 | 293.9111636 | 2.468216082 | FALSE | TRUE | FALSE |
| SCHIP1 | 0.0056362 | 0.88985007 | 0.01245 | 0.005636224 | 0.005636224 | 0.111754242 | 0.00594831 | 0.89790135 | 1 | 0.005948315 | 0.005948315 | 0.118064427 | 0.947532886 | -0.023405708 | FALSE | TRUE | FALSE |
| MAN2A2 | 0.0056126 | 0.88985752 | 0.97285 | 5.289003675 | 6106.672333 | 118633.6947 | 0.02615038 | 0.892080504 | 0.127 | 0.002750095 | 0.25474193 | 2.540790279 | 0.214628711 | -0.668312183 | FALSE | TRUE | FALSE |
| ANO6 | 0.0055719 | 0.88986496 | 0.7523 | 0.022273529 | 0.188988693 | 0.43743634 | 0.00158065 | 0.899202764 | 1 | 0.013277559 | 0.053080309 | 0.261328123 | 3.525095261 | 0.547170858 | FALSE | TRUE | FALSE |
| IQGAP2 | 0.0055629 | 0.8898724 | 0.54785 | 0.01168255 | 0.432512099 | 0.90807993 | 0.51463251 | 0.749801655 | 0.1403 | 0.032205901 | 0.514632508 | 2.511918113 | 0.010809511 | -1.966193933 | FALSE | TRUE | FALSE |
| PALB2 | 0.0055255 | 0.88987984 | 0.8793 | 0.392452453 | 3.437194167 | 7.696093571 | 6.0266245 | 0.268530625 | 0.20375 | 6.026624505 | 54.80885469 | 510.6612385 | 0.000916852 | -3.037700703 | FALSE | FALSE | FALSE |
| TAS2R9 | 0.0055194 | 0.88988728 | 0.0074 | 0.005519447 | 0.005519447 | 0.005519447 | 0.00650575 | 0.897723701 | 1 | 0.006505751 | 0.006505751 | 0.006505751 | 0.848394988 | -0.071401906 | FALSE | TRUE | FALSE |
| PIK3C2B | 0.005483 | 0.88989472 | 1 | 0.005483034 | 0.005483034 | 0.184040827 | 0.01012347 | 0.896736466 | 1 | 0.010123467 | 0.010123467 | 0.340321859 | 0.541616185 | -0.266308366 | FALSE | TRUE | FALSE |
| BBS2 | 0.0054802 | 0.88990216 | 0.6784 | 0.02188781 | 0.781542206 | 3.123069323 | 172.482641 | 0.015472385 | 0.0714 | 2.705460296 | 418.1835908 | 3404.274797 | 3.17724E-05 | -4.497950543 | FALSE | FALSE | FALSE |
| ALPK1 | 0.0054749 | 0.8899096 | 0.5093 | 0.005474871 | 0.046492503 | 0.107023659 | 0.00017337 | 0.900075137 | 0.5138 | 0.001425625 | 0.001425625 | 0.02794419 | 31.57848553 | 1.499391298 | FALSE | TRUE | FALSE |
| R3HCC1L | 0.0054592 | 0.88991703 | 1 | 0.047037587 | 0.415990818 | 0.928003114 | 0.61453491 | 0.705618517 | 0.69275 | 0.614534905 | 52.79303541 | 52.79303541 | 0.008883397 | -2.051420906 | FALSE | TRUE | FALSE |
| TBC1D4 | 0.0054549 | 0.88992447 | 0.039 | 0.005454923 | 0.005454923 | 0.183566773 | 0.71018087 | 0.661319746 | 0.0276 | 0.080277819 | 0.710180872 | 2.704805689 | 0.007681033 | -2.114580371 | FALSE | TRUE | FALSE |
| NEK5 | 0.0054347 | 0.8899319 | 1 | 0.005434664 | 0.005434664 | 0.107384293 | 0.00947074 | 0.896889474 | 1 | 0.080277556 | 0.080277556 | 0.18740485 | 0.573837288 | -0.241211235 | FALSE | TRUE | FALSE |
| NLRP1 | 0.0054174 | 0.88993934 | 0.5162 | 0.045868583 | 0.045868583 | 0.892171398 | 0.00976834 | 0.896806067 | 1 | 0.083765697 | 0.083765697 | 1.634688049 | 0.554591166 | -0.256027052 | FALSE | TRUE | FALSE |
| KIF9 | 0.0053957 | 0.88994677 | 0.02055 | 0.005395671 | 0.005395671 | 0.106163828 | 0.00983191 | 0.896778236 | 1 | 0.009831907 | 0.009831907 | 0.193827615 | 0.548791908 | -0.260592301 | FALSE | TRUE | FALSE |
| RCBTB2 | 0.0053908 | 0.8899542 | 1 | 0.021544948 | 0.086128698 | 0.426062959 | 0.01012514 | 0.896729502 | 1 | 0.010125141 | 0.010125141 | 0.200500475 | 0.53241525 | -0.273749513 | FALSE | TRUE | FALSE |
| ROBO1 | 0.0053621 | 0.88996163 | 1 | 0.005362116 | 0.005362116 | 0.179603594 | 0.15386484 | 0.863596106 | 0.287 | 0.038475297 | 0.153864843 | 2.991074836 | 0.034849522 | -1.457803177 | FALSE | TRUE | FALSE |
| FOXN1 | 0.0053605 | 0.88996906 | 1 | 51714.20547 | 75747422.21 | 882967829.7 | 0.00777246 | 0.897387347 | 1 | 1009496158 | 2.26636E+13 | 2.91253E+14 | 0.689684858 | -0.161349309 | FALSE | TRUE | FALSE |
| ANKDD1A | 0.0053067 | 0.88997649 | 1 | 0.005306659 | 0.005306659 | 0.104894413 | 0.07904049 | 0.880618458 | 0.25975 | 0.079040491 | 0.703560706 | 0.703560706 | 0.06713849 | -1.17302843 | FALSE | TRUE | FALSE |
| CHST4 | 0.0052973 | 0.88998392 | 1 | 0.005297338 | 0.005297338 | 0.104940166 | 0.1482185 | 0.864997856 | 0.8127 | 0.592528509 | 2.369324821 | 9.476510283 | 0.03574006 | -1.446844721 | FALSE | TRUE | FALSE |
| ZNF347 | 0.0052879 | 0.88999134 | 0.9691 | 3.415749319 | 31.05463739 | 289.2530113 | 0.08631695 | 0.878939308 | 0.88005 | 0.341322068 | 6.819169782 | 6.819169782 | 0.061261226 | -1.212814319 | FALSE | TRUE | FALSE |
| KIAA0319 | 0.0052782 | 0.88999877 | 1 | 0.005278162 | 0.005278162 | 0.103386067 | 0.00954582 | 0.896868634 | 0.026 | 0.009545821 | 0.009545821 | 0.187443357 | 0.552929086 | -0.257330564 | FALSE | TRUE | FALSE |
| FAM151A | 0.0052751 | 0.89000619 | 0.75525 | 0.043487756 | 0.367992887 | 0.858752515 | 0.65770521 | 0.686641845 | 0.5014 | 0.657705214 | 5.793110603 | 5.793110603 | 0.008020408 | -2.095803552 | FALSE | TRUE | FALSE |
| AK9 | 0.0052651 | 0.89001362 | 0.63795 | 0.021058874 | 0.173062538 | 0.850751348 | 0.09232687 | 0.877701233 | 0.38245 | 0.044231253 | 0.369294514 | 1.819024646 | 0.057026779 | -1.243921158 | FALSE | TRUE | FALSE |
| DZANK1 | 0.0052464 | 0.89002104 | 0.7546 | 0.044670786 | 0.390399648 | 0.882181988 | 0.0097961 | 0.896785195 | 0.5043 | 0.084497943 | 0.084497943 | 0.193751278 | 0.535559326 | -0.271192414 | FALSE | TRUE | FALSE |
| PEX5 | 0.0052445 | 0.89002846 | 0.0198 | 0.005244541 | 0.005244541 | 0.103398612 | 0.00967451 | 0.896833883 | 0.01845 | 0.009674507 | 0.009674507 | 0.191064395 | 0.542099019 | -0.265921379 | FALSE | TRUE | FALSE |
| NUP155 | 0.0052277 | 0.89003588 | 1 | 0.020890546 | 0.083501725 | 0.702725822 | 2.34233686 | 0.461327103 | 0.24555 | 2.342336858 | 37.49589485 | 150.0767997 | 0.002231841 | -2.651336658 | FALSE | FALSE | FALSE |
| ANXA7 | 0.0051993 | 0.8900433 | 0.8745 | 0.048165147 | 0.457655697 | 0.457655697 | 0.0070912 | 0.897504267 | 0.0116 | 0.007091201 | 0.007091201 | 0.015463215 | 0.73320248 | -0.134776074 | FALSE | TRUE | FALSE |
| DARS2 | 0.0051825 | 0.89005072 | 0.01825 | 0.00518254 | 0.00518254 | 0.102433215 | 0.00957597 | 0.896854736 | 1 | 0.009575966 | 0.009575966 | 0.189537058 | 0.541202797 | -0.266639967 | FALSE | TRUE | FALSE |
| IFNA4 | 0.0051541 | 0.89005814 | 1 | 0.048298837 | 0.464526509 | 0.464526509 | 0.00551368 | 0.897983153 | 1 | 0.051750002 | 0.498499306 | 0.498499306 | 0.934776892 | -0.029292032 | FALSE | TRUE | FALSE |
| KEL | 0.0051522 | 0.89006556 | 0.75545 | 0.045121843 | 0.180370498 | 0.888151736 | 2.94353345 | 0.415188725 | 0.01095 | 0.32477691 | 2.943533455 | 6.404898138 | 0.001750334 | -2.75687896 | FALSE | FALSE | FALSE |
| XIRP1 | 0.0051394 | 0.89007297 | 0.87135 | 0.042008895 | 0.352234206 | 1.408872986 | 1.8662E-05 | 0.900564275 | 0.88155 | 0.097749431 | 0.8353132 | 3.283637371 | 275.3856393 | 2.439941289 | FALSE | TRUE | FALSE |
| KCNQ1 | 0.0051174 | 0.89008039 | 0.625 | 0.020438546 | 0.726899305 | 2.90472792 | 0.00022808 | 0.899970171 | 0.87775 | 0.009004826 | 0.143617137 | 0.143617137 | 22.43644958 | 1.350954134 | FALSE | TRUE | FALSE |
| MYZAP | 0.0051001 | 0.8900878 | 1 | 0.005100071 | 0.005100071 | 0.101186174 | 0.0488891 | 0.886955661 | 0.0104 | 0.048889102 | 0.048889102 | 0.102502192 | 0.104319179 | -0.981635838 | FALSE | TRUE | FALSE |
| CEP85 | 0.0050937 | 0.89009522 | 1 | 0.005093715 | 0.005093715 | 0.100375598 | 0.00607411 | 0.897853578 | 0.7534 | 0.055366077 | 0.517966275 | 1.092963268 | 0.838594582 | -0.076447948 | FALSE | TRUE | FALSE |
| NOS2 | 0.0050796 | 0.89010263 | 1 | 0.044219112 | 0.395095466 | 7.705050002 | 0.07876291 | 0.88069087 | 0.87835 | 0.710024007 | 26.2489434 | 26.2489434 | 0.064492915 | -1.190487996 | FALSE | TRUE | FALSE |
| FLT4 | 0.0050732 | 0.89011004 | 0.9391 | 0.18674298 | 7.056881583 | 25.11635753 | 0.04066439 | 0.888799624 | 0.5156 | 0.040664393 | 0.040664393 | 0.786507182 | 0.124756816 | -0.903935718 | FALSE | TRUE | FALSE |
| TAS2R4 | 0.0050184 | 0.89011745 | 0.0066 | 0.005018425 | 0.005018425 | 0.005018425 | 0.00594974 | 0.897894528 | 0.0056 | 0.005949743 | 0.005949743 | 0.005949743 | 0.843469162 | -0.073930791 | FALSE | TRUE | FALSE |
| KLHL38 | 0.0050087 | 0.89012486 | 1 | 0.044271967 | 0.044271967 | 0.098844987 | 0.08258554 | 0.879651118 | 0.01485 | 0.082585537 | 0.082585537 | 0.183191502 | 0.060648339 | -1.217181091 | FALSE | TRUE | FALSE |
| KIF13B | 0.0049985 | 0.89013227 | 0.52515 | 0.004998475 | 0.079786874 | 0.386251686 | 0.00024651 | 0.899950465 | 1 | 0.00098425 | 0.003930894 | 0.07633909 | 20.27725238 | 1.307009107 | FALSE | TRUE | FALSE |
| OTOA | 0.0049865 | 0.89013968 | 1 | 0.043632636 | 0.391867301 | 1.471731767 | 0.07884832 | 0.880681823 | 0.51575 | 0.714060729 | 6.632842923 | 13.99519505 | 0.063241975 | -1.198994579 | FALSE | TRUE | FALSE |
| KIDINS220 | 0.0049853 | 0.89014708 | 1 | 0.004985261 | 0.004985261 | 0.167217329 | 5.5690886 | 0.28463279 | 0.67105 | 22.26762757 | 2045.226866 | 9926.674887 | 0.000895166 | -3.048096206 | FALSE | FALSE | FALSE |
| PLEKHG2 | 0.0049457 | 0.89015449 | 1 | 0.043586541 | 0.043586541 | 0.845807382 | 0.00943112 | 0.896910307 | 1 | 0.00943112 | 0.00943112 | 0.183669398 | 0.524397404 | -0.280339467 | FALSE | TRUE | FALSE |
| C1orf87 | 0.0049375 | 0.8901619 | 1 | 0.383554867 | 3.512983634 | 3.512983634 | 0.06724043 | 0.88304568 | 0.5107 | 0.067240429 | 0.067240429 | 0.151529862 | 0.073430496 | -1.134123536 | FALSE | TRUE | FALSE |
| PDE6C | 0.0049261 | 0.8901693 | 0.9852 | 3.596446571 | 330.4025504 | 3281.91844 | 0.01075442 | 0.896548127 | 1 | 0.042991663 | 0.042991663 | 0.363454831 | 0.458049505 | -0.339087582 | FALSE | TRUE | FALSE |
| TRPV3 | 0.0048917 | 0.8901767 | 0.07795 | 0.000530309 | 0.046253273 | 0.046253273 | 0.01440854 | 0.895311777 | 0.75875 | 0.014408539 | 0.130976455 | 0.283222171 | 0.339498329 | -0.46916236 | FALSE | TRUE | FALSE |
| MAST4 | 0.0048598 | 0.8901841 | 0.7769 | 0.0048598 | 0.091994776 | 0.790189095 | 7.45898019 | 0.236169072 | 0.2052 | 0.816470968 | 15.55559908 | 142.1102647 | 0.000651537 | -3.186061076 | FALSE | FALSE | FALSE |
| ITGA1 | 0.0048584 | 0.89019151 | 1 | 0.004858448 | 0.004858448 | 0.163669979 | 0.03529452 | 0.890045861 | 0.2696 | 0.035294523 | 0.141121136 | 0.693171816 | 0.137654456 | -0.861209726 | FALSE | TRUE | FALSE |
| DSCAML1 | 0.004848 | 0.89019891 | 1 | 86845474602 | 2.1092E+15 | 3.67492E+17 | 0.00800932 | 0.897297779 | 1 | 1.32599E+13 | 5.18938E+18 | 9.86577E+19 | 0.605298513 | -0.218030393 | FALSE | TRUE | FALSE |
| INVS | 0.0048191 | 0.89020631 | 1 | 0.041521226 | 0.367193414 | 1.399523917 | 0.07733318 | 0.880961713 | 0.75685 | 0.077333184 | 0.691353878 | 1.51986567 | 0.0623159 | -1.20540113 | FALSE | TRUE | FALSE |
| MMS22L | 0.0048009 | 0.8902137 | 1 | 0.043190824 | 0.043190824 | 0.846938851 | 0.006679 | 0.897641516 | 0.0269 | 0.006679005 | 0.006679005 | 0.225053951 | 0.718806482 | -0.143388015 | FALSE | TRUE | FALSE |
| B3GNT3 | 0.0047876 | 0.8902211 | 1 | 3.76773E+46 | 5.34137E+56 | 4.13175E+60 | 0.01351303 | 0.895639403 | 1 | 1.32001E+37 | 5.40833E+45 | 3.20979E+49 | 0.354295335 | -0.450634566 | FALSE | TRUE | FALSE |
| ENOSF1 | 0.0047817 | 0.8902285 | 1 | 0.004781715 | 0.004781715 | 0.094875764 | 0.00868482 | 0.897125129 | 1 | 0.008684819 | 0.008684819 | 0.172483066 | 0.550583099 | -0.259177124 | FALSE | TRUE | FALSE |
| UACA | 0.004771 | 0.89023589 | 0.49715 | 0.000777683 | 0.004770973 | 0.093369083 | 0.01102085 | 0.896450221 | 0.028 | 0.01102085 | 0.01102085 | 0.216238934 | 0.432904257 | -0.363608144 | FALSE | TRUE | FALSE |
| MELK | 0.0047674 | 0.89024329 | 0.5059 | 0.040230501 | 0.040230501 | 0.161351531 | 0.07450158 | 0.881517966 | 0.50825 | 0.074501577 | 0.074501577 | 0.172487553 | 0.0639909 | -1.193881784 | FALSE | TRUE | FALSE |
| PCDHA2 | 0.0047667 | 0.89025068 | 1 | 0.004766704 | 0.004766704 | 0.09271791 | 0.28862482 | 0.82852484 | 0.5093 | 0.288624816 | 2.597752524 | 5.632658159 | 0.016515224 | -1.782115523 | FALSE | TRUE | FALSE |
| IRF7 | 0.004762 | 0.89025807 | 0.0178 | 0.004761992 | 0.004761992 | 0.018867031 | 0.01109175 | 0.89643622 | 0.2604 | 0.011091746 | 0.098411564 | 0.219018977 | 0.429327553 | -0.367211239 | FALSE | TRUE | FALSE |

| | | | | | | | | | | | | | | | | | |
|---|---|---|---|---|---|---|---|---|---|---|---|---|---|---|---|---|---|
| DSG2 | 0.0047468 | 0.89026547 | 0.5105 | 0.04111889 | 0.04111889 | 0.80613747 | 0.00903753 | 0.897007425 | 0.02435 | 0.009037528 | 0.009037528 | 0.177602479 | 0.525229048 | -0.279651263 | FALSE | TRUE | FALSE |
| MBD4 | 0.0046957 | 0.89027286 | 0.50795 | 0.040500263 | 0.040500263 | 0.092932516 | 0.07567757 | 0.881330069 | 0.51085 | 0.075677575 | 0.075677575 | 0.172010066 | 0.062049264 | -1.207263364 | FALSE | TRUE | FALSE |
| PIK3C2A | 0.0046937 | 0.89028025 | 1 | 0.042553846 | 0.042553846 | 0.827567757 | 0.00908323 | 0.896986628 | 0.03565 | 0.009083233 | 0.009083233 | 0.304899476 | 0.516741374 | -0.286726764 | FALSE | TRUE | FALSE |
| DSP | 0.0046899 | 0.89028764 | 0.56255 | 0.010519461 | 1.53916036 | 9.714429857 | 0.00029859 | 0.899897881 | 0.5245 | 0.000645406 | 0.010293607 | 0.049022007 | 15.70699942 | 1.196093228 | FALSE | TRUE | FALSE |
| NLRP4 | 0.0046896 | 0.89029502 | 1 | 0.039981133 | 0.039981133 | 0.782902311 | 0.008879 | 0.897055921 | 1 | 0.008878999 | 0.008878999 | 0.174304636 | 0.528170421 | -0.277225925 | FALSE | TRUE | FALSE |
| DAAM2 | 0.0046865 | 0.89030241 | 1 | 0.004686515 | 0.004686515 | 0.091441615 | 0.00871124 | 0.897118212 | 0.51055 | 0.034812043 | 0.034812043 | 0.681238742 | 0.537984863 | -0.269229943 | FALSE | TRUE | FALSE |
| ITSN2 | 0.0046525 | 0.8903098 | 0.93905 | 0.039901332 | 3.165511706 | 6.804205991 | 0.00921489 | 0.896958886 | 0.9987 | 240.1755744 | 55986.43788 | 571911.0256 | 0.504891879 | -0.296801615 | FALSE | TRUE | FALSE |
| CEP63 | 0.0046252 | 0.89031718 | 0.50705 | 0.004625161 | 0.004625161 | 0.018776267 | 0.00098514 | 0.899455394 | 0.01555 | 0.000985141 | 0.000985141 | 0.019497501 | 4.694922111 | 0.671628392 | FALSE | TRUE | FALSE |
| MEFV | 0.0046245 | 0.89032457 | 1 | 0.004624506 | 0.004624506 | 0.090806553 | 0.00823777 | 0.897235685 | 0.5118 | 0.07020737 | 0.07020737 | 0.278738815 | 0.561378669 | -0.250744093 | FALSE | TRUE | FALSE |
| DYM | 0.0045962 | 0.89033195 | 1 | 0.004596189 | 0.004596189 | 0.090860743 | 0.0086275 | 0.89713896 | 1 | 0.008627505 | 0.008627505 | 0.170791967 | 0.532736758 | -0.273487337 | FALSE | TRUE | FALSE |
| KIF21B | 0.004552 | 0.89033933 | 1 | 0.00455199 | 0.087224099 | 0.152694622 | 0.05851697 | 0.88494572 | 0.9982 | 509.8228495 | 219613.2692 | 2358814.509 | 0.077789226 | -1.109080549 | FALSE | TRUE | FALSE |
| CDON | 0.0045364 | 0.89034671 | 1 | 0.039754179 | 0.158921166 | 1.338552483 | 0.00882105 | 0.897083614 | 0.03155 | 0.008821054 | 0.008821054 | 0.172497277 | 0.514274589 | -0.288804934 | FALSE | TRUE | FALSE |
| DNM3 | 0.0045318 | 0.89035409 | 0.02295 | 0.004531755 | 0.004531755 | 0.0890134 | 0.07639309 | 0.881186564 | 0.51135 | 0.076393085 | 0.076393085 | 1.503756918 | 0.059321532 | -1.226787642 | FALSE | TRUE | FALSE |
| THRSP | 0.0045192 | 0.89036147 | 1 | 0.0045192 | 0.042479611 | 0.042479611 | 0.00492811 | 0.8981532 | 1 | 0.004928108 | 0.004928108 | 0.004928108 | 0.917025406 | -0.037618632 | FALSE | TRUE | FALSE |
| GBF1 | 0.0044982 | 0.89036885 | 0.05185 | 0.004498167 | 0.086529285 | 0.150618659 | 9.96855389 | 0.198618527 | 0.07795 | 0.274426672 | 9.968553891 | 48.17021939 | 0.000451236 | -3.345596587 | FALSE | FALSE | FALSE |
| SPTB | 0.0044914 | 0.89037623 | 0.9731 | 3.737930397 | 124.878763 | 699.540749 | 0.07102198 | 0.882211218 | 0.9659 | 64.04538458 | 12375.80826 | 128571.0156 | 0.063240099 | -1.199007457 | FALSE | TRUE | FALSE |
| CCDC136 | 0.0044908 | 0.8903836 | 1 | 0.037865279 | 0.327706236 | 1.276712647 | 0.00850102 | 0.897173523 | 0.5126 | 0.072688215 | 0.072688215 | 1.429127541 | 0.528266067 | -0.277147285 | FALSE | TRUE | FALSE |
| CSRNP3 | 0.0044908 | 0.89039098 | 1 | 0.004490789 | 0.004490789 | 0.088739533 | 0.00479024 | 0.898173572 | 1 | 0.044993177 | 0.044993177 | 0.162127715 | 0.937486405 | -0.028035021 | FALSE | TRUE | FALSE |
| CPT1A | 0.0044343 | 0.89039836 | 0.7495 | 0.004434285 | 0.01771231 | 0.086872787 | 0.15278144 | 0.863897168 | 0.5138 | 0.152781436 | 0.750859885 | 3.000535155 | 0.029023713 | -1.537247032 | FALSE | TRUE | FALSE |
| TRPM6 | 0.0044308 | 0.89040573 | 0.94165 | 0.155634465 | 5.612382853 | 27.07788428 | 0.03356345 | 0.890450517 | 0.76555 | 0.297473344 | 2.706048629 | 13.1138613 | 0.132013629 | -0.879381232 | FALSE | TRUE | FALSE |
| GP2 | 0.0044013 | 0.8904131 | 1 | 0.004401348 | 0.004401348 | 0.087092997 | 0.00829394 | 0.897214971 | 0.0128 | 0.008293944 | 0.008293944 | 0.1643284 | 0.530670031 | -0.275175438 | FALSE | TRUE | FALSE |
| HSF1 | 0.0043985 | 0.89042047 | 1 | 5.43983E+69 | 1.97914E+83 | 5.48846E+88 | 0.01226721 | 0.896014538 | 1 | 4.57475E+60 | 4.36505E+72 | 5.15547E+76 | 0.358559462 | -0.445438813 | FALSE | TRUE | FALSE |
| GBA2 | 0.004397 | 0.89042784 | 1 | 0.004396981 | 0.004396981 | 0.086158881 | 0.00820988 | 0.897249489 | 1 | 0.00820988 | 0.00820988 | 0.161265009 | 0.535571891 | -0.271182225 | FALSE | TRUE | FALSE |
| CYP4F2 | 0.0043915 | 0.89043521 | 0.01515 | 0.004391527 | 0.004391527 | 0.08669422 | 0.03102609 | 0.891006428 | 0.8757 | 0.124076732 | 1.076333244 | 1.076333244 | 0.141543019 | -0.849111547 | FALSE | TRUE | FALSE |
| NT5C1A | 0.004335 | 0.89044258 | 1 | 1.93863E+40 | 5.50848E+49 | 4.11497E+53 | 0.01345838 | 0.895660703 | 1 | 5.07428E+30 | 1.48512E+39 | 8.1864E+42 | 0.322103777 | -0.492004182 | FALSE | TRUE | FALSE |
| RPTN | 0.004333 | 0.89044995 | 0.88925 | 0.038292244 | 3.219671956 | 30.59734568 | 0.00084197 | 0.899554784 | 0.99255 | 0.60342662 | 54.59361202 | 191.468745 | 5.146216056 | 0.711488015 | FALSE | TRUE | FALSE |
| SPECC1 | 0.0043237 | 0.89045732 | 1 | 0.017279014 | 0.017279014 | 0.34004017 | 0.00801299 | 0.897290883 | 1 | 0.07193347 | 0.07193347 | 0.27100908 | 0.539589681 | -0.267936364 | FALSE | TRUE | FALSE |
| NIN | 0.0043018 | 0.89046469 | 1 | 0.037142889 | 0.082846686 | 1.240453209 | 5.08929999 | 0.303625253 | 0.5161 | 5.089299993 | 98.45054404 | 459.2802041 | 0.000845271 | -3.073003995 | FALSE | FALSE | FALSE |
| PREX2 | 0.0042821 | 0.89047205 | 1 | 0.004282127 | 0.004282127 | 0.143961409 | 0.00786877 | 0.897352915 | 1 | 0.031459221 | 0.12580496 | 1.059057211 | 0.544192799 | -0.264247209 | FALSE | TRUE | FALSE |
| LRIG2 | 0.0042649 | 0.89047942 | 1 | 0.004264947 | 0.004264947 | 0.083575049 | 0.56180355 | 0.728270671 | 0.2655 | 0.0639622 | 0.56180355 | 2.158377785 | 0.007591527 | -2.119670837 | FALSE | TRUE | FALSE |
| KRT78 | 0.0042637 | 0.89048678 | 0.5078 | 0.037975958 | 0.037975958 | 0.084306698 | 21.837382 | 0.124625239 | 0.3429 | 2.320154464 | 210.6833395 | 2082.31918 | 0.00019525 | -3.709410059 | FALSE | FALSE | FALSE |
| LRP10 | 0.0042479 | 0.89049414 | 0.881 | 0.06783865 | 0.271199972 | 1.333391557 | 0.00833816 | 0.897208065 | 0.01845 | 0.008338159 | 0.008338159 | 0.164229357 | 0.509457278 | -0.292892229 | FALSE | TRUE | FALSE |
| ENTPD2 | 0.0042219 | 0.8905015 | 1 | 2.55577E+66 | 1.75397E+78 | 4.06928E+83 | 0.00461595 | 0.898200723 | 1 | 1.1664E+87 | 7.8097E+100 | 3.3287E+106 | 0.914625698 | -0.038756601 | FALSE | TRUE | FALSE |
| CEP112 | 0.0041994 | 0.89050886 | 0.52135 | 0.034731054 | 0.034731054 | 0.682953123 | 0.06477019 | 0.883645021 | 0.02305 | 0.064770194 | 0.064770194 | 1.276219819 | 0.064835912 | -1.188184375 | FALSE | TRUE | FALSE |
| SLC2A9 | 0.0041871 | 0.89051622 | 0.0129 | 0.004187064 | 0.004187064 | 0.082856629 | 0.03117877 | 0.890952602 | 0.50575 | 0.031178772 | 0.031178772 | 0.154549177 | 0.13429215 | -0.871949374 | FALSE | TRUE | FALSE |
| F10 | 0.0041639 | 0.89052358 | 1 | 2.12396E+12 | 5.9096E+16 | 7.62652E+18 | 0.00604146 | 0.897874057 | 1 | 4.36654E+17 | 2.08338E+23 | 3.99057E+25 | 0.6892165 | -0.161644334 | FALSE | TRUE | FALSE |
| EVC2 | 0.0040814 | 0.89053094 | 0.52415 | 0.034749548 | 0.303481023 | 0.677394049 | 0.00126866 | 0.899309279 | 1 | 0.001268659 | 0.001268659 | 0.04275042 | 3.217122091 | 0.507467543 | FALSE | TRUE | FALSE |
| CEP152 | 0.0040461 | 0.8905383 | 0.5098 | 0.004046147 | 0.033029729 | 0.160790241 | 0.02290873 | 0.892972202 | 0.15355 | 0.022908729 | 0.195380183 | 0.447250199 | 0.17662031 | -0.752959356 | FALSE | TRUE | FALSE |
| FIG4 | 0.0040186 | 0.89054565 | 0.8901 | 0.064322335 | 1.0305815 | 5.061839276 | 2.04978138 | 0.484887109 | 0.7301 | 8.280634693 | 369.2273241 | 1479.229791 | 0.001960504 | -2.707632253 | FALSE | FALSE | FALSE |
| KRTAP5-5 | 0.0039885 | 0.89055301 | 1 | 0.269493568 | 20.16078849 | 20.16078849 | 0.00790815 | 0.897332246 | 1 | 0.007908153 | 0.065429515 | 0.065429515 | 0.504355931 | -0.297262868 | FALSE | TRUE | FALSE |
| SETBP1 | 0.0039679 | 0.89056036 | 1 | 333851.4334 | 430963607.8 | 59696895334 | 0.00908363 | 0.896979693 | 1 | 722.1410986 | 839094.9061 | 9116254.228 | 0.436819707 | -0.359697777 | FALSE | TRUE | FALSE |
| COX6C | 0.0039446 | 0.89056772 | 1 | 0.003944591 | 0.003944591 | 0.003944591 | 0.00403079 | 0.898322753 | 0.00275 | 0.004030789 | 0.004030789 | 0.004030789 | 0.978615147 | -0.009388066 | FALSE | TRUE | FALSE |
| BARD1 | 0.0039439 | 0.89057507 | 1 | 0.03456702 | 0.138163316 | 0.532849087 | 0.06696825 | 0.883124176 | 0.51155 | 0.066968247 | 0.066968247 | 0.256134098 | 0.058891819 | -1.22994503 | FALSE | TRUE | FALSE |
| MUC2 | 0.0039235 | 0.89058242 | 0.99595 | 24.65549531 | 4372.330039 | 42409.00866 | 0.05845384 | 0.884971131 | 0.9997 | 4118.209771 | 422386.8654 | 7968471.928 | 0.067121671 | -1.173137238 | FALSE | TRUE | FALSE |
| MED23 | 0.003838 | 0.89058977 | 1 | 0.003838032 | 0.003838032 | 0.129050562 | 0.02882001 | 0.891526809 | 0.517 | 0.028820011 | 0.028820011 | 0.564186333 | 0.133172465 | -0.875585561 | FALSE | TRUE | FALSE |
| STAG3 | 0.0038233 | 0.89059712 | 0.99805 | 213.2949575 | 20010.09377 | 200540.1487 | 0.06521147 | 0.883532534 | 0.992 | 48.04427197 | 4427.31155 | 15367.23656 | 0.058629995 | -1.231880142 | FALSE | TRUE | FALSE |
| FANCI | 0.0038187 | 0.89060447 | 0.2524 | 0.000307651 | 0.003818672 | 0.01871655 | 0.62090086 | 0.70332386 | 0.31165 | 0.1552214 | 20.73343562 | 82.97721728 | 0.006150211 | -2.211109955 | FALSE | TRUE | FALSE |
| TRPM5 | 0.0038169 | 0.89061182 | 1 | 0.003816934 | 0.003816934 | 0.074367423 | 0.51555733 | 0.749169773 | 0.5087 | 0.515557332 | 4.723074477 | 10.07614702 | 0.007403511 | -2.130562295 | FALSE | TRUE | FALSE |
| TMC3 | 0.0038022 | 0.89061917 | 1 | 0.032840091 | 0.032840091 | 0.642009636 | 0.0074231 | 0.897463027 | 1 | 0.007423102 | 0.007423102 | 0.14551217 | 0.512217101 | -0.290545926 | FALSE | TRUE | FALSE |
| IL17RC | 0.0037999 | 0.89062652 | 1 | 0.003799862 | 0.003799862 | 0.074399634 | 0.04716515 | 0.887291327 | 0.51305 | 0.047165152 | 0.047165152 | 0.925811304 | 0.080565033 | -1.093853411 | FALSE | TRUE | FALSE |
| HAP1 | 0.0037925 | 0.89063386 | 0.26605 | 0.003792529 | 0.03347258 | 0.074658402 | 4.01172651 | 0.355835315 | 0.3499 | 0.428992455 | 38.4335381 | 376.9992465 | 0.000945361 | -3.024402459 | FALSE | FALSE | FALSE |
| IFNA8 | 0.003785 | 0.89064121 | 1 | 0.003784986 | 0.003784986 | 0.003784986 | 0.0041527 | 0.898288882 | 1 | 0.004152695 | 0.004152695 | 0.004152695 | 0.911452775 | -0.040265829 | FALSE | TRUE | FALSE |
| FXR2 | 0.0037664 | 0.89064855 | 0.02335 | 0.003766412 | 0.003766412 | 0.074161963 | 0.00697548 | 0.897559207 | 1 | 0.006975479 | 0.006975479 | 0.137605578 | 0.539950288 | -0.267646223 | FALSE | TRUE | FALSE |
| HPS3 | 0.0037646 | 0.89065589 | 1 | 0.060194166 | 0.514857328 | 1.525758357 | 0.05939909 | 0.884818521 | 0.88185 | 0.526684201 | 4.79015168 | 10.35430021 | 0.063377865 | -1.198062396 | FALSE | TRUE | FALSE |
| HELB | 0.003703 | 0.89066323 | 1 | 2.260249401 | 20.16550565 | 184.3225553 | 0.00792829 | 0.897318461 | 1 | 0.007928288 | 0.007928288 | 0.155820552 | 0.46705698 | -0.330630133 | FALSE | TRUE | FALSE |
| ADAMTS14 | 0.0037026 | 0.89067057 | 0.6827 | 0.003702576 | 0.059155294 | 0.287284642 | 1.27403202 | 0.557769011 | 0.29745 | 0.31865228 | 5.095093258 | 42.94822952 | 0.002906188 | -2.536676345 | FALSE | FALSE | FALSE |
| FAT3 | 0.0036977 | 0.89067791 | 0.84435 | 0.014774089 | 1.079237922 | 7.785811872 | 0.00080869 | 0.899587871 | 0.9996 | 3.297238585 | 211.2502959 | 1747.284031 | 4.572400001 | 0.660144216 | FALSE | TRUE | FALSE |
| PLCE1 | 0.0036538 | 0.89068525 | 0.99655 | 4.996750808 | 666.1257835 | 4107.145127 | 0.00240124 | 0.898888673 | 0.5257 | 0.020272072 | 0.04625672 | 0.390514223 | 1.521616454 | 0.182305196 | FALSE | TRUE | FALSE |
| TRPM3 | 0.0036473 | 0.89069259 | 1 | 0.032258041 | 0.032258041 | 0.621745663 | 0.00700697 | 0.897552343 | 0.52535 | 0.062471319 | 0.062471319 | 1.209353627 | 0.520530795 | -0.283553573 | FALSE | TRUE | FALSE |
| MYO10 | 0.0036379 | 0.89069993 | 0.3763 | 0.001608774 | 0.231210464 | 0.923162584 | 1.9404E-06 | 0.901061516 | 0.7635 | 7.73713E-06 | 6.85966E-05 | 0.001316917 | 1874.813298 | 3.272958025 | FALSE | TRUE | FALSE |
| FBXW5 | 0.0036314 | 0.89070726 | 1 | 0.014515908 | 0.014515908 | 0.285798146 | 0.00701125 | 0.897545477 | 1 | 0.007011253 | 0.007011253 | 0.138300566 | 0.517945518 | -0.285715921 | FALSE | TRUE | FALSE |
| MYCBP2 | 0.0036253 | 0.8907146 | 1 | 0.003625278 | 0.066943864 | 0.118786929 | 0.06183054 | 0.884350027 | 0.10135 | 0.061830544 | 1.152374191 | 2.034828697 | 0.058632477 | -1.231861755 | FALSE | TRUE | FALSE |
| KCNA5 | 0.0036046 | 0.89072193 | 0.5074 | 0.014396172 | 0.057508493 | 0.282921938 | 0.03295173 | 0.890605438 | 0.50415 | 0.032951732 | 0.131662674 | 0.279452954 | 0.109390207 | -0.961021555 | FALSE | TRUE | FALSE |
| ALOX15 | 0.0035995 | 0.89072927 | 1 | 0.030203102 | 0.260126765 | 0.594143803 | 0.43427353 | 0.779550474 | 0.93835 | 35.40969382 | 3180.602724 | 31214.37758 | 0.008288627 | -2.08151739 | FALSE | TRUE | FALSE |
| DDX31 | 0.0035988 | 0.8907366 | 1 | 0.030955805 | 0.273301288 | 0.607013264 | 0.24024092 | 0.842017499 | 0.38595 | 0.110373312 | 0.960741481 | 2.169408604 | 0.014980123 | -1.824484611 | FALSE | TRUE | FALSE |
| ANKLE2 | 0.0035943 | 0.89074393 | 1 | 0.003594269 | 0.003594269 | 0.070585003 | 0.00656112 | 0.897682624 | 1 | 0.055346663 | 0.055346663 | 1.089276724 | 0.547813682 | -0.261367125 | FALSE | TRUE | FALSE |
| LPIN2 | 0.003587 | 0.89075126 | 1 | 0.032631208 | 0.032631208 | 0.640924776 | 0.00717173 | 0.897490523 | 1 | 0.007171727 | 0.007171727 | 0.141167281 | 0.500162418 | -0.300888944 | FALSE | TRUE | FALSE |
| SMARCA4 | 0.003583 | 0.89075859 | 1 | 0.014313935 | 0.068557957 | 0.273886785 | 0.00697288 | 0.89756607 | 0.5149 | 0.027860547 | 0.027860547 | 0.535878283 | 0.513847027 | -0.289166152 | FALSE | TRUE | FALSE |
| LARS2 | 0.003541 | 0.89076592 | 0.0257 | 0.003541028 | 0.003541028 | 0.069572237 | 0.00655845 | 0.897689473 | 1 | 0.056752649 | 0.056752649 | 1.117412184 | 0.539918689 | -0.26767164 | FALSE | TRUE | FALSE |
| INO80 | 0.0035118 | 0.89077324 | 0.28585 | 0.003511826 | 0.033728144 | 0.653752639 | 0.00101635 | 0.899422222 | 0.04165 | 0.001016347 | 0.001016347 | 0.034117033 | 3.455341477 | 0.538490973 | FALSE | TRUE | FALSE |
| PTPRCAP | 0.0035045 | 0.89078057 | 1 | 0.003504495 | 0.003504495 | 0.003504495 | 0.00386325 | 0.898383667 | 0.00445 | 0.003863247 | 0.003863247 | 0.003863247 | 0.907137093 | -0.042327074 | FALSE | TRUE | FALSE |
| CD163L1 | 0.0034919 | 0.8907879 | 0.9808 | 18.31425232 | 1532.355738 | 14503.99751 | 0.07825216 | 0.880799343 | 0.819 | 0.672473588 | 5.924157028 | 53.46773879 | 0.044623806 | -1.350433388 | FALSE | TRUE | FALSE |
| THAP9 | 0.00349 | 0.89079522 | 1 | 0.029829518 | 0.029829518 | 0.587380441 | 0.00665826 | 0.897655222 | 1 | 0.057580301 | 0.057580301 | 1.135934869 | 0.524156629 | -0.280538917 | FALSE | TRUE | FALSE |

| | | | | | | | | | | | | | | | | | |
|---|---|---|---|---|---|---|---|---|---|---|---|---|---|---|---|---|---|
| FHOD3 | 0.0034886 | 0.89080255 | 1 | 2008051.054 | 2510854890 | 7673524154 | 0.0070377 | 0.897524876 | 1 | 437633.6923 | 482829010.8 | 534842922.7 | 0.495704351 | -0.304777269 | FALSE | TRUE | FALSE |
| KBTBD12 | 0.0034744 | 0.89080987 | 1 | 0.120110852 | 1.066199792 | 1.066199792 | 0.06067357 | 0.884605988 | 0.0149 | 0.060673566 | 0.060673566 | 0.137389276 | 0.057263661 | -1.242120886 | FALSE | TRUE | FALSE |
| TRPV5 | 0.0034667 | 0.89081719 | 1 | 0.003466656 | 0.003466656 | 0.068203367 | 0.00673339 | 0.897607235 | 1 | 0.006733386 | 0.006733386 | 0.132733222 | 0.514845864 | -0.288322772 | FALSE | TRUE | FALSE |
| MICAL3 | 0.0034491 | 0.89082451 | 1 | 0.013788767 | 0.066094398 | 0.264228675 | 0.00653438 | 0.897710013 | 1 | 0.057872038 | 0.12584888 | 1.114585801 | 0.527843885 | -0.277494505 | FALSE | TRUE | FALSE |
| TBX6 | 0.0034475 | 0.89083183 | 0.5001 | 0.030054388 | 0.030054388 | 0.068155494 | 0.05841933 | 0.884988066 | 0.0133 | 0.058419332 | 0.058419332 | 0.131395981 | 0.059013363 | -1.229049639 | FALSE | TRUE | FALSE |
| SLC6A16 | 0.0034377 | 0.89083915 | 0.5046 | 0.029010072 | 0.029010072 | 0.571801154 | 0.0065321 | 0.897716857 | 1 | 0.055847487 | 0.055847487 | 0.221077549 | 0.526285516 | -0.278778582 | FALSE | TRUE | FALSE |
| RFC4 | 0.0034284 | 0.89084647 | 0.01075 | 0.003428379 | 0.003428379 | 0.06808876 | 0.00638374 | 0.897744229 | 1 | 0.025529328 | 0.025529328 | 0.025529328 | 0.537048337 | -0.269986624 | FALSE | TRUE | FALSE |
| TRPA1 | 0.003421 | 0.89085379 | 0.98525 | 0.250930266 | 20.32668121 | 189.6065074 | 17404.6499 | 0.000113696 | 0.0207 | 47.64124977 | 17404.64986 | 172161.9631 | 1.96558E-07 | -6.706508175 | FALSE | FALSE | FALSE |
| TNS3 | 0.0034198 | 0.8908611 | 0.7018 | 0.003419845 | 0.054564909 | 0.263904145 | 0.00936956 | 0.896931131 | 0.51735 | 0.009369558 | 0.037430304 | 0.181774688 | 0.364995381 | -0.437712632 | FALSE | TRUE | FALSE |
| ATP10B | 0.0034187 | 0.89086842 | 0.4057 | 0.001589963 | 0.05452171 | 0.217814723 | 0.00297326 | 0.898693958 | 0.0986 | 0.000744255 | 0.011881006 | 0.057919192 | 1.149810486 | 0.060626265 | FALSE | TRUE | FALSE |
| FLG2 | 0.0033958 | 0.89087573 | 0.99395 | 129.4272825 | 95643.694 | 1842516.387 | 40.0769787 | 0.068468786 | 0.9863 | 2670677.365 | 5096805159 | 2.9936E+11 | 8.47323E-05 | -4.071951214 | FALSE | FALSE | FALSE |
| SVEP1 | 0.0033702 | 0.89088304 | 0.6416 | 0.007491448 | 0.47758606 | 2.251024175 | 0.11393008 | 0.872587753 | 0.2062 | 0.012735855 | 0.455253138 | 2.160849498 | 0.029581041 | -1.52898655 | FALSE | TRUE | FALSE |
| ALS2 | 0.003366 | 0.89089036 | 1 | 0.013456085 | 0.013456085 | 0.261398899 | 0.00638514 | 0.897737387 | 0.51675 | 0.056190744 | 0.056190744 | 1.095341981 | 0.527157632 | -0.278059501 | FALSE | TRUE | FALSE |
| TARBP1 | 0.0033414 | 0.89089767 | 0.0354 | 0.003341401 | 0.003341401 | 0.112319305 | 0.00630904 | 0.89778526 | 0.0329 | 0.006309044 | 0.006309044 | 0.212364258 | 0.529620879 | -0.276034902 | FALSE | TRUE | FALSE |
| CLEC4M | 0.0033293 | 0.89090498 | 0.0115 | 0.003329279 | 0.003329279 | 0.00865545 | 0.00101778 | 0.899415585 | 1 | 0.001017785 | 0.001017785 | 0.020215349 | 3.271103551 | 0.514694292 | FALSE | TRUE | FALSE |
| MICAL1 | 0.0033214 | 0.89091229 | 0.75695 | 0.006836531 | 0.027333232 | 0.230437813 | 0.22137126 | 0.846929394 | 0.77095 | 1.040387383 | 66.55014313 | 598.966202 | 0.015003731 | -1.823800745 | FALSE | TRUE | FALSE |
| CDHR1 | 0.0033199 | 0.8909196 | 1 | 0.028253106 | 0.246791446 | 0.554672079 | 0.00673242 | 0.897614093 | 1 | 0.006732424 | 0.006732424 | 0.13246527 | 0.493121875 | -0.307045732 | FALSE | TRUE | FALSE |
| ADAMTS9 | 0.0033077 | 0.8909269 | 1 | 0.003307749 | 0.06341343 | 0.110519578 | 0.00636594 | 0.89775107 | 0.5268 | 0.02544831 | 0.122653171 | 0.490315174 | 0.519601265 | -0.2843298 | FALSE | TRUE | FALSE |
| LEF1 | 0.0032764 | 0.89093421 | 1 | 0.013082974 | 0.052253382 | 0.064961578 | 0.02605986 | 0.892125792 | 0.50565 | 0.026059856 | 0.026059856 | 0.129505634 | 0.125727276 | -0.900570494 | FALSE | TRUE | FALSE |
| SLC45A1 | 0.003263 | 0.89094152 | 1 | 5.59641E+27 | 7.87029E+34 | 8.5288E+37 | 0.03581091 | 0.889912983 | 1 | 9.76761E+20 | 2.10169E+27 | 6.24332E+29 | 0.091117611 | -1.040397677 | FALSE | TRUE | FALSE |
| ITIH5 | 0.0032502 | 0.89094882 | 1 | 0.051957234 | 0.44746151 | 2.189610272 | 36.9550463 | 0.07485927 | 0.27405 | 6.906753082 | 590.8333712 | 2898.525821 | 8.79507E-05 | -4.055760564 | FALSE | FALSE | FALSE |
| TTLL4 | 0.0032264 | 0.89095613 | 1 | 0.003226382 | 0.003226382 | 0.062973368 | 0.00593407 | 0.897914992 | 1 | 0.049607746 | 0.049607746 | 0.971067019 | 0.543704981 | -0.264636688 | FALSE | TRUE | FALSE |
| CGNL1 | 0.0032129 | 0.89096343 | 0.92475 | 0.051321131 | 1.775216736 | 7.099366465 | 0.00103966 | 0.899395669 | 1 | 0.001039659 | 0.001039659 | 0.020295587 | 3.090378654 | 0.490011695 | FALSE | TRUE | FALSE |
| TIE1 | 0.0031928 | 0.89097073 | 1 | 0.003192756 | 0.003192756 | 0.062029763 | 0.02449262 | 0.892636598 | 0.51195 | 0.024492617 | 0.024492617 | 0.477489329 | 0.130355867 | -0.884869417 | FALSE | TRUE | FALSE |
| TROAP | 0.0031053 | 0.89097803 | 1 | 0.0031053 | 0.0031053 | 0.061086894 | 0.00423994 | 0.898254991 | 0.0199 | 0.004239938 | 0.004239938 | 0.083572048 | 0.732392756 | -0.13525596 | FALSE | TRUE | FALSE |
| ADCY3 | 0.0031037 | 0.89098533 | 1 | 0.028012184 | 0.028012184 | 0.543654957 | 0.00629929 | 0.89779893 | 1 | 0.00629929 | 0.00629929 | 0.122691856 | 0.492708353 | -0.307410075 | FALSE | TRUE | FALSE |
| NAGLU | 0.0030801 | 0.89099263 | 0.6969 | 0.027934309 | 0.445780332 | 0.550588901 | 0.02982837 | 0.891297783 | 0.2647 | 0.007466283 | 0.029828366 | 0.147418126 | 0.103261312 | -0.986062361 | FALSE | TRUE | FALSE |
| C2CD3 | 0.0030594 | 0.89099993 | 0.97105 | 0.228701573 | 8.205824367 | 75.46514839 | 0.03194417 | 0.890798543 | 0.79235 | 0.127726077 | 4.284859847 | 11.97096827 | 0.095774594 | -1.018749683 | FALSE | TRUE | FALSE |
| ACSF3 | 0.0030144 | 0.89100723 | 0.019 | 0.003014388 | 0.003014388 | 0.059385482 | 0.04784035 | 0.887168731 | 0.5059 | 0.047840349 | 0.047840349 | 0.111243381 | 0.063009321 | -1.200595198 | FALSE | TRUE | FALSE |
| IFNL3 | 0.0030084 | 0.89101453 | 1 | 0.003008433 | 0.027536878 | 0.027536878 | 0.00365929 | 0.898451271 | 1 | 0.003659288 | 0.003659288 | 0.003659288 | 0.822136022 | -0.085056323 | FALSE | TRUE | FALSE |
| DNPEP | 0.0030036 | 0.89102182 | 0.7529 | 0.026824605 | 0.107179406 | 0.107179406 | 1.86127223 | 0.502863104 | 0.2913 | 0.465777271 | 7.439608188 | 29.74395839 | 0.001613724 | -2.792170628 | FALSE | FALSE | FALSE |
| KIF24 | 0.0030022 | 0.89102912 | 0.27975 | 0.000750912 | 0.003002158 | 0.025265672 | 0.48735187 | 0.760057193 | 0.35255 | 0.121887752 | 9.31597612 | 37.2486844 | 0.006160144 | -2.21040911 | FALSE | TRUE | FALSE |
| CDK5RAP2 | 0.0029978 | 0.89103641 | 0.76035 | 0.024927026 | 0.212746881 | 4.110165357 | 0.00528581 | 0.898051234 | 1 | 3.405739784 | 66.06729543 | 287.8998809 | 0.56714862 | -0.24630312 | FALSE | TRUE | FALSE |
| RHOT2 | 0.0029932 | 0.89104371 | 1 | 0.191365749 | 0.765580743 | 3.758580176 | 0.00615776 | 0.89782626 | 1 | 0.006157764 | 0.006157764 | 0.12120505 | 0.486084706 | -0.313288043 | FALSE | TRUE | FALSE |
| UGT2B28 | 0.0029581 | 0.891051 | 0.74845 | 0.024440475 | 0.207271959 | 0.207271959 | 0.00598853 | 0.897880881 | 1 | 0.005988529 | 0.005988529 | 0.118839857 | 0.493960729 | -0.306307577 | FALSE | TRUE | FALSE |
| ERN2 | 0.0029055 | 0.89105829 | 0.9381 | 0.215101345 | 17.6291745 | 37.56264638 | 0.00615284 | 0.897833091 | 1 | 0.05303963 | 0.05303963 | 1.038796468 | 0.472222139 | -0.325853656 | FALSE | TRUE | FALSE |
| FMN1 | 0.002901 | 0.89106558 | 0.01435 | 0.000349399 | 0.002900968 | 0.056797837 | 0.00079015 | 0.899601101 | 1 | 0.426470584 | 3.65208448 | 32.04153777 | 3.671409072 | 0.564832777 | FALSE | TRUE | FALSE |
| CDHR2 | 0.0028961 | 0.89107287 | 0.97085 | 0.052932056 | 0.845843243 | 4.10142625 | 0.0150642 | 0.895104248 | 0.03415 | 0.015064201 | 0.015064201 | 0.293113753 | 0.192248561 | -0.716136902 | FALSE | TRUE | FALSE |
| NOTCH2 | 0.0028877 | 0.89108016 | 0.6498 | 0.004938837 | 0.046034247 | 0.786060492 | 0.11124779 | 0.87308657 | 0.52845 | 0.266549601 | 10.4497621 | 41.75843297 | 0.025957064 | -1.585744438 | FALSE | TRUE | FALSE |
| MEGF6 | 0.0028825 | 0.89108745 | 0.8333 | 0.024111247 | 1.538789535 | 14.19819551 | 0.0492908 | 0.886840634 | 0.7039 | 0.438700603 | 149.1759426 | 1425.304377 | 0.058478922 | -1.233000644 | FALSE | TRUE | FALSE |
| CAND2 | 0.0028434 | 0.89109473 | 0.51635 | 0.011365324 | 0.011365324 | 0.221597266 | 0.02240414 | 0.893098501 | 0.0292 | 0.022404137 | 0.022404137 | 0.438150509 | 0.126915006 | -0.896487027 | FALSE | TRUE | FALSE |
| C1orf94 | 0.0028015 | 0.89110202 | 1 | 0.002801486 | 0.002801486 | 0.055581925 | 0.28495712 | 0.82958698 | 0.25625 | 0.030787997 | 0.284957117 | 0.284957117 | 0.009831253 | -2.007391113 | FALSE | TRUE | FALSE |
| P2RX4 | 0.0027824 | 0.8911093 | 0.50865 | 0.025227899 | 0.025227899 | 0.05507825 | 0.00356934 | 0.898485042 | 1 | 0.003569339 | 0.003569339 | 0.070743381 | 0.779522185 | -0.10817152 | FALSE | TRUE | FALSE |
| GLDN | 0.0027412 | 0.89111659 | 0.49995 | 0.002741196 | 0.010951684 | 0.054243027 | 0.02185995 | 0.893268939 | 0.01485 | 0.021859949 | 0.021859949 | 0.10839374 | 0.125398113 | -0.901709 | FALSE | TRUE | FALSE |
| KAT6B | 0.0027373 | 0.89112387 | 0.9395 | 0.061481391 | 2.416305807 | 8.213900594 | 0.00180291 | 0.899136087 | 1 | 0.001802906 | 0.001802906 | 0.06042963 | 1.51828808 | 0.181354182 | FALSE | TRUE | FALSE |
| ACAD10 | 0.0027364 | 0.89113115 | 0.419 | 0.000730794 | 26.09164576 | 231.0833932 | 51.0720099 | 0.053541331 | 0.3182 | 9.899606581 | 13134.20536 | 210942.2564 | 5.35801E-05 | -4.270996871 | FALSE | FALSE | FALSE |
| CCDC180 | 0.0027353 | 0.89113843 | 0.1564 | 0.002735299 | 0.021070815 | 0.091785018 | 0.00108801 | 0.899369102 | 0.2787 | 0.001088014 | 0.008402117 | 0.036566516 | 2.514030337 | 0.400370514 | FALSE | TRUE | FALSE |
| LIMA1 | 0.0026896 | 0.89114571 | 1 | 1.955692119 | 73.91369367 | 716.0995554 | 0.05075537 | 0.886461148 | 0.5086 | 0.461439512 | 4.30296852 | 4.351138545 | 0.052991204 | -1.275796214 | FALSE | TRUE | FALSE |
| ADCY9 | 0.0026701 | 0.89115299 | 1 | 0.097774859 | 0.919796771 | 4.444418006 | 0.00570765 | 0.897942265 | 1 | 0.022805852 | 0.022805852 | 0.44209428 | 0.467816406 | -0.329924552 | FALSE | TRUE | FALSE |
| HEXIM1 | 0.0026489 | 0.89116027 | 1 | 0.002648929 | 0.002648929 | 0.052384243 | 0.00266884 | 0.898821609 | 0.75645 | 0.025127348 | 0.242804 | 0.242804 | 0.992540701 | -0.003251675 | FALSE | TRUE | FALSE |
| NDUFC1 | 0.0026273 | 0.89116755 | 1 | 0.002627299 | 0.002627299 | 0.002627299 | 0.00271695 | 0.898788047 | 0.0017 | 0.002716949 | 0.002716949 | 0.002716949 | 0.967003369 | -0.014572013 | FALSE | TRUE | FALSE |
| AURKA | 0.0026272 | 0.89117483 | 0.0105 | 0.00262716 | 0.00262716 | 0.052178861 | 0.00503434 | 0.898119232 | 0.0084 | 0.005034344 | 0.005034344 | 0.005034344 | 0.521847488 | -0.282456403 | FALSE | TRUE | FALSE |
| CPT1C | 0.0026215 | 0.8911821 | 1 | 0.002621478 | 0.002621478 | 0.051324995 | 0.00511596 | 0.898092042 | 0.51195 | 0.04662308 | 0.04662308 | 0.915133189 | 0.51241224 | -0.290380505 | FALSE | TRUE | FALSE |
| ARHGEF10 | 0.0026111 | 0.89118938 | 0.51895 | 0.002611065 | 0.002611065 | 0.050609019 | 0.00080241 | 0.899594486 | 1 | 0.000802408 | 0.000802408 | 0.027034675 | 3.254038639 | 0.512422705 | FALSE | TRUE | FALSE |
| SLC26A9 | 0.0025955 | 0.89119665 | 0.514 | 0.005934455 | 0.023702316 | 0.11662235 | 0.13056677 | 0.86881982 | 0.2666 | 0.013880709 | 0.130566771 | 0.273351933 | 0.019879078 | -1.70160377 | FALSE | TRUE | FALSE |
| FAM8A1 | 0.0025745 | 0.89120393 | 1 | 8.05662E+92 | 1.0314E+107 | 5.7291E+112 | 0.00896895 | 0.897028214 | 1 | 3.67628E+79 | 5.69745E+92 | 1.49633E+98 | 0.287049332 | -0.542043459 | FALSE | TRUE | FALSE |
| CAPN13 | 0.0025386 | 0.8912112 | 1 | 0.002538557 | 0.002538557 | 0.050044853 | 0.03922643 | 0.889133394 | 0.753 | 0.156856998 | 1.358955392 | 1.358955392 | 0.064715479 | -1.188991827 | FALSE | TRUE | FALSE |
| NOD1 | 0.0025287 | 0.89121847 | 1 | 0.021766033 | 0.087007912 | 0.425449099 | 1.38249287 | 0.54689737 | 0.6639 | 3.157663218 | 118.1542886 | 1132.968494 | 0.001829079 | -2.737767619 | FALSE | FALSE | FALSE |
| TRPV1 | 0.0025282 | 0.89122574 | 0.51605 | 0.002528241 | 0.002528241 | 0.049587237 | 0.00073424 | 0.899627549 | 0.748 | 0.006350209 | 0.056370496 | 0.124838751 | 3.44334524 | 0.536980568 | FALSE | TRUE | FALSE |
| ACRV1 | 0.00252 | 0.89123301 | 0.00625 | 0.002519982 | 0.002519982 | 0.002519982 | 0.00300325 | 0.898660316 | 1 | 0.003003251 | 0.003003251 | 0.003003251 | 0.839084784 | -0.076194154 | FALSE | TRUE | FALSE |
| TTLL5 | 0.002512 | 0.89124028 | 1 | 0.020843462 | 0.177522443 | 0.700445433 | 0.04019963 | 0.888926986 | 0.52305 | 0.347201165 | 3.075929295 | 6.787297933 | 0.062487877 | -1.20420423 | FALSE | TRUE | FALSE |
| CUL7 | 0.0025033 | 0.89124755 | 0.34425 | 0.000626767 | 0.010000574 | 0.048162715 | 0.00053005 | 0.899752999 | 0.51785 | 0.00053005 | 0.00053005 | 0.010244818 | 4.722746555 | 0.67419464 | FALSE | TRUE | FALSE |
| CACNA1I | 0.0025023 | 0.89125481 | 0.61425 | 0.002502336 | 0.159075026 | 0.765558777 | 0.06622505 | 0.883289574 | 0.1364 | 0.013723837 | 0.264299688 | 3.490253716 | 0.037785337 | -1.422676703 | FALSE | TRUE | FALSE |
| PCDHA11 | 0.0024954 | 0.89126208 | 0.518 | 0.021558312 | 0.021558312 | 0.419311586 | 0.04092223 | 0.888735843 | 0.87975 | 0.366099274 | 3.359418787 | 7.14427148 | 0.060979106 | -1.214818944 | FALSE | TRUE | FALSE |
| PEAR1 | 0.0024163 | 0.89126934 | 0.9383 | 0.020482645 | 0.712372815 | 1.504805534 | 0.01278523 | 0.895830816 | 0.51025 | 0.012785231 | 0.012785231 | 0.250310987 | 0.188993627 | -0.72355284 | FALSE | TRUE | FALSE |
| PCDHGB2 | 0.0023318 | 0.89127661 | 0.031 | 0.002331846 | 0.002331846 | 0.045490625 | 0.00452114 | 0.898207509 | 0.51375 | 0.038824042 | 0.038824042 | 0.759638861 | 0.515765449 | -0.287547754 | FALSE | TRUE | FALSE |
| SPTBN2 | 0.0023314 | 0.89128387 | 0.54345 | 0.021070626 | 0.077588417 | 0.701214091 | 0.01930818 | 0.89400416 | 0.54275 | 0.019308183 | 0.161162863 | 0.64420211 | 0.120748653 | -0.918117704 | FALSE | TRUE | FALSE |
| DIS3 | 0.0023117 | 0.89129113 | 0.75435 | 0.019200152 | 0.076778095 | 0.377383415 | 0.3083626 | 0.822506042 | 0.6924 | 0.308362597 | 24.77195984 | 24.77195984 | 0.007496544 | -2.125138888 | FALSE | TRUE | FALSE |
| SMPDL3A | 0.0022969 | 0.89129839 | 0.01085 | 0.002296933 | 0.002296933 | 0.045639836 | 0.14568731 | 0.865496731 | 0.62725 | 0.145687307 | 5.136296347 | 5.136296347 | 0.015766182 | -1.802273466 | FALSE | TRUE | FALSE |
| GIPR | 0.0022964 | 0.89130565 | 1 | 0.019029683 | 0.019029683 | 0.045351173 | 0.0370764 | 0.889662076 | 0.74895 | 0.037076405 | 0.319852825 | 0.319852825 | 0.061936936 | -1.208050285 | FALSE | TRUE | FALSE |
| CEP290 | 0.0022713 | 0.89131291 | 0.52455 | 0.018041223 | 1.22361833 | 10.43636394 | 0.58949175 | 0.716911744 | 0.27605 | 0.589491753 | 44.05592264 | 394.1104961 | 0.003852945 | -2.41420715 | FALSE | TRUE | FALSE |
| ABCC4 | 0.0022256 | 0.89132017 | 0.6957 | 0.002225562 | 0.190315641 | 0.395902611 | 0.06455203 | 0.883670952 | 0.51545 | 0.608942432 | 58.23000646 | 198.3211307 | 0.034477022 | -1.462470253 | FALSE | TRUE | FALSE |
| COL9A1 | 0.0022167 | 0.89132743 | 0.26425 | 0.000555293 | 0.002216719 | 0.010904567 | 0.00020274 | 0.900009558 | 1 | 0.000809538 | 0.000809538 | 0.015931988 | 10.93362705 | 1.038764256 | FALSE | TRUE | FALSE |

| | | | | | | | | | | | | | | | | | |
|---|---|---|---|---|---|---|---|---|---|---|---|---|---|---|---|---|---|
| ADAMTS20 | 0.0022119 | 0.89133469 | 0.96315 | 0.159454131 | 21.30854809 | 117.3978611 | 0.00092383 | 0.899515051 | 0.64205 | 0.003692759 | 0.030785144 | 0.258027141 | 2.394289932 | 0.379176739 | FALSE | TRUE | FALSE |
| PLCB2 | 0.0022039 | 0.89134194 | 0.5209 | 0.008811401 | 0.008811401 | 0.171605755 | 0.01747849 | 0.894426391 | 0.75285 | 0.017478486 | 0.069902422 | 0.590133304 | 0.126090839 | -0.899316464 | FALSE | TRUE | FALSE |
| IFNA17 | 0.0021903 | 0.8913492 | 1 | 0.002190251 | 0.002190251 | 0.002190251 | 0.00230569 | 0.898955655 | 0.87635 | 0.200366552 | 1.94084927 | 1.94084927 | 0.949934426 | -0.022306373 | FALSE | TRUE | FALSE |
| UNC79 | 0.0021879 | 0.89135645 | 0.06765 | 0.002187893 | 0.04166332 | 0.073004971 | 0.00324185 | 0.898599707 | 1 | 0.00324185 | 0.062090549 | 0.10841533 | 0.674890272 | -0.170766832 | FALSE | TRUE | FALSE |
| OIT3 | 0.0021681 | 0.89136371 | 1 | 0.008662074 | 0.008662074 | 0.170962176 | 0.00444451 | 0.898227863 | 1 | 0.01775914 | 0.01775914 | 0.087859603 | 0.487822349 | -0.311738307 | FALSE | TRUE | FALSE |
| HTR3B | 0.002163 | 0.89137096 | 1 | 2.59858E+32 | 9.72173E+40 | 1.40737E+44 | 0.00191441 | 0.899102718 | 1 | 2.50368E+51 | 5.80881E+61 | 5.36085E+65 | 1.129843635 | 0.053018343 | FALSE | TRUE | FALSE |
| PIF1 | 0.0021625 | 0.89137821 | 0.5098 | 0.018369957 | 0.018369957 | 0.042726441 | 0.00447999 | 0.898214295 | 1 | 0.004479988 | 0.004479988 | 0.088643912 | 0.482702988 | -0.316320013 | FALSE | TRUE | FALSE |
| ABCC3 | 0.0021563 | 0.89138546 | 0.68445 | 0.008620721 | 4.304505951 | 68.9556311 | 1.20134651 | 0.566950387 | 0.21085 | 0.075047719 | 38.25417223 | 613.2799999 | 0.001794895 | -2.745960923 | FALSE | FALSE | FALSE |
| TRPC6 | 0.0021207 | 0.89139271 | 0.5056 | 0.00847243 | 0.00847243 | 0.166616294 | 0.00415765 | 0.898282106 | 1 | 0.037567523 | 0.348399336 | 0.740278396 | 0.510079841 | -0.29236184 | FALSE | TRUE | FALSE |
| UTRN | 0.0021165 | 0.89139996 | 0.54605 | 0.002116534 | 0.073419601 | 0.606895786 | 0.00157977 | 0.899209427 | 0.8129 | 0.013240742 | 0.455397202 | 4.009047605 | 1.339774456 | 0.127031693 | FALSE | TRUE | FALSE |
| MGA | 0.0021052 | 0.89140721 | 0.5335 | 0.002105209 | 0.004319159 | 0.069874095 | 0.00063153 | 0.89968701 | 1 | 0.000631526 | 0.012068949 | 0.021021117 | 3.333524378 | 0.522903635 | FALSE | TRUE | FALSE |
| CHD4 | 0.0020916 | 0.89141445 | 0.7641 | 0.002091567 | 0.010050968 | 0.160315124 | 0.00129376 | 0.899302627 | 0.5256 | 0.00516683 | 0.00516683 | 0.09964512 | 1.616654621 | 0.208617248 | FALSE | TRUE | FALSE |
| KCNH1 | 0.0020721 | 0.8914217 | 0.75835 | 0.002072089 | 0.008274825 | 0.069943248 | 0.0981611 | 0.876449566 | 0.0216 | 0.019985156 | 0.079847477 | 0.39208887 | 0.02110907 | -1.675530909 | FALSE | TRUE | FALSE |
| MDC1 | 0.0020557 | 0.89142894 | 1 | 0.017407063 | 0.151292793 | 2.9158808 | 0.28680102 | 0.829120546 | 0.52035 | 2.583029816 | 50.00132541 | 225.5524371 | 0.007167654 | -2.144622961 | FALSE | TRUE | FALSE |
| POMGNT1 | 0.0020469 | 0.89143619 | 0.986 | 0.032692863 | 0.522691602 | 2.090761536 | 0.00497784 | 0.898132822 | 0.96745 | 0.079529989 | 0.318008433 | 1.567476631 | 0.41120025 | -0.38594663 | FALSE | TRUE | FALSE |
| ENO3 | 0.0020462 | 0.89144343 | 0.50695 | 0.008176466 | 0.008176466 | 0.040503672 | 0.01627472 | 0.894787943 | 0.76195 | 0.035873455 | 0.143367691 | 0.143367691 | 0.125726022 | -0.900574824 | FALSE | TRUE | FALSE |
| SLC22A16 | 0.0020196 | 0.89145067 | 0.5046 | 0.017307291 | 0.017307291 | 0.068371387 | 0.13292637 | 0.868224059 | 0.6843 | 0.295364241 | 10.75621767 | 10.75621767 | 0.015193496 | -1.818342294 | FALSE | TRUE | FALSE |
| FBN2 | 0.0020172 | 0.89145791 | 0.23395 | 0.000506143 | 0.127882943 | 0.603374942 | 1.7892E-05 | 0.900577253 | 0.29785 | 4.49384E-06 | 3.76686E-05 | 0.000339096 | 112.7430438 | 2.052089756 | FALSE | TRUE | FALSE |
| CNKSR1 | 0.0020055 | 0.89146515 | 1 | 0.133884584 | 1.136854704 | 2.635688041 | 0.26331294 | 0.835694709 | 0.69835 | 1.053210718 | 20.13159952 | 80.52326072 | 0.007616524 | -2.118243171 | FALSE | TRUE | FALSE |
| POLN | 0.0020038 | 0.89147239 | 0.75515 | 0.008010047 | 0.065900404 | 0.157552557 | 0.32211449 | 0.818127662 | 0.73905 | 2.829253615 | 406.9801927 | 2005.904625 | 0.006220804 | -2.206153498 | FALSE | TRUE | FALSE |
| LMO7 | 0.0019995 | 0.89147963 | 0.5231 | 0.016925884 | 0.016925884 | 0.329922462 | 0.00417394 | 0.898275328 | 1 | 0.004173941 | 0.004173941 | 0.140527285 | 0.479042144 | -0.319626278 | FALSE | TRUE | FALSE |
| TRPS1 | 0.0019973 | 0.89148687 | 0.7563 | 0.001997256 | 0.007973608 | 0.067257817 | 0.00118175 | 0.899335875 | 1 | 0.0111368 | 0.107715391 | 0.37546403 | 1.690076835 | 0.227906449 | FALSE | TRUE | FALSE |
| PCDHGA6 | 0.0019897 | 0.8914941 | 0.76175 | 0.017047208 | 0.068143994 | 0.332590037 | 0.03455663 | 0.890217391 | 0.75625 | 0.034556629 | 0.307133478 | 0.676188453 | 0.057577201 | -1.239749453 | FALSE | TRUE | FALSE |
| ZNF747 | 0.0019866 | 0.89150134 | 0.0056 | 0.001986648 | 0.001986648 | 0.001986648 | 0.00265549 | 0.898828319 | 1 | 0.002655494 | 0.002655494 | 0.002655494 | 0.748127625 | -0.126024308 | FALSE | TRUE | FALSE |
| TBCD | 0.0019701 | 0.89150857 | 1 | 0.001970098 | 0.001970098 | 0.03833314 | 0.0038857 | 0.898370136 | 1 | 0.033171551 | 0.033171551 | 0.647545032 | 0.507012674 | -0.294981185 | FALSE | TRUE | FALSE |
| PHRF1 | 0.00196 | 0.89151581 | 1 | 0.017417583 | 0.158865495 | 3.041099164 | 0.01702036 | 0.894614683 | 0.5304 | 0.017020364 | 0.017020364 | 0.327500897 | 0.115155775 | -0.938714276 | FALSE | TRUE | FALSE |
| BRMS1 | 0.0019589 | 0.89152304 | 1 | 0.001958913 | 0.001958913 | 0.001958913 | 0.00227217 | 0.898962348 | 1 | 0.002272166 | 0.002272166 | 0.002272166 | 0.862134838 | -0.064424805 | FALSE | TRUE | FALSE |
| NLRP2 | 0.0019264 | 0.89153027 | 1 | 0.00192636 | 0.00192636 | 0.037621114 | 0.00386511 | 0.898376902 | 0.02845 | 0.003865109 | 0.003865109 | 0.075698088 | 0.498397207 | -0.3024244 | FALSE | TRUE | FALSE |
| PLOD1 | 0.0019122 | 0.8915375 | 0.8778 | 0.030526228 | 0.122013717 | 0.599192113 | 0.06150463 | 0.884435461 | 0.6606 | 0.245868194 | 3.932030444 | 15.73029351 | 0.031089941 | -1.507380097 | FALSE | TRUE | FALSE |
| MAST2 | 0.0018698 | 0.89154473 | 0.52295 | 0.003824348 | 0.015290563 | 0.294133387 | 0.00111912 | 0.899349168 | 0.5261 | 0.009302538 | 0.009302538 | 0.179770896 | 1.670773121 | 0.22291748 | FALSE | TRUE | FALSE |
| PGBD1 | 0.0018681 | 0.89155196 | 1 | 0.016582679 | 0.016582679 | 0.063117735 | 0.02103861 | 0.89354943 | 0.9354 | 0.192645642 | 17.41914266 | 17.41914266 | 0.088793572 | -1.051618472 | FALSE | TRUE | FALSE |
| SPAG5 | 0.0018225 | 0.89155919 | 0.0315 | 0.00182245 | 0.00182245 | 0.035660046 | 0.00034607 | 0.899858407 | 0.0281 | 0.000346066 | 0.000346066 | 0.006789151 | 5.266199526 | 0.72149731 | FALSE | TRUE | FALSE |
| TRRAP | 0.0018215 | 0.89156642 | 1 | 316934159.8 | 6.59681E+12 | 1.06862E+15 | 0.00403665 | 0.898315981 | 1 | 64210086.58 | 1.26357E+12 | 2.33462E+13 | 0.451237464 | -0.34559485 | FALSE | TRUE | FALSE |
| PLIN4 | 0.0018012 | 0.89157364 | 1 | 1.72027E+43 | 5.71099E+52 | 2.94814E+56 | 0.0801969 | 0.880264252 | 1 | 4.38396E+43 | 1.84361E+53 | 8.93324E+56 | 0.022459259 | -1.648604569 | FALSE | TRUE | FALSE |
| BTD | 0.0017928 | 0.89158087 | 1 | 0.015972989 | 0.146066361 | 0.316021988 | 0.00398621 | 0.898349834 | 1 | 0.003986206 | 0.003986206 | 0.078968295 | 0.44974857 | -0.347030209 | FALSE | TRUE | FALSE |
| DOCK6 | 0.0017761 | 0.89158809 | 0.5318 | 0.001776122 | 0.008424726 | 0.033687513 | 0.0086002 | 0.897152788 | 0.53175 | 0.008600204 | 0.019811655 | 0.164137781 | 0.206520933 | -0.685035922 | FALSE | TRUE | FALSE |
| TSC1 | 0.0017759 | 0.89159532 | 1 | 0.001775891 | 0.001775891 | 0.034682647 | 0.03422171 | 0.890302974 | 0.0284 | 0.034221714 | 0.034221714 | 0.670233794 | 0.051893693 | -1.284885422 | FALSE | TRUE | FALSE |
| PFKM | 0.0017184 | 0.89160254 | 1 | 0.006866775 | 0.006866775 | 0.134856164 | 0.01397561 | 0.895440208 | 0.76085 | 0.03093746 | 0.123638663 | 0.608899564 | 0.122960165 | -0.910235564 | FALSE | TRUE | FALSE |
| MANBA | 0.001706 | 0.89160976 | 1 | 0.01476065 | 0.01476065 | 0.290585196 | 0.03112749 | 0.890975676 | 0.5096 | 0.03112749 | 0.03112749 | 0.61394693 | 0.05480817 | -1.261154701 | FALSE | TRUE | FALSE |
| ADCY7 | 0.0017047 | 0.89161698 | 0.0345 | 0.001704709 | 0.001704709 | 0.033194377 | 0.00348433 | 0.898525539 | 1 | 0.031172388 | 0.031172388 | 0.608924634 | 0.489249977 | -0.310469186 | FALSE | TRUE | FALSE |
| SNED1 | 0.0016968 | 0.8916242 | 1 | 0.015004053 | 0.136173885 | 1.120823544 | 0.00298893 | 0.898680504 | 0.996 | 21.55231148 | 2120.818517 | 21784.0809 | 0.567698386 | -0.245882341 | FALSE | TRUE | FALSE |
| SPATA31D1 | 0.0016804 | 0.89163142 | 0.27995 | 0.001680375 | 0.014613903 | 0.284162014 | 0.0026883 | 0.898801474 | 0.97065 | 0.212823505 | 1.965918396 | 18.60426285 | 0.625069388 | -0.20407177 | FALSE | TRUE | FALSE |
| TRIP11 | 0.0016592 | 0.89163864 | 0.52375 | 0.01399261 | 0.01399261 | 0.467682438 | 0.20523159 | 0.850758566 | 0.69725 | 0.20523159 | 16.94088599 | 35.8274461 | 0.008084632 | -2.092339731 | FALSE | TRUE | FALSE |
| CCDC78 | 0.001659 | 0.89164586 | 0.0116 | 0.001658988 | 0.001658988 | 0.032858208 | 0.00218233 | 0.899015865 | 1 | 0.019345478 | 0.176012988 | 0.176012988 | 0.760191558 | -0.119076958 | FALSE | TRUE | FALSE |
| CENPE | 0.00165 | 0.89165307 | 1 | 0.014302585 | 0.031703841 | 0.47607993 | 0.26390199 | 0.83556149 | 0.28025 | 0.029386116 | 0.263901988 | 5.094706917 | 0.006252414 | -2.203952297 | FALSE | TRUE | FALSE |
| ITGAX | 0.0016443 | 0.89166029 | 0.51495 | 0.014095145 | 0.014095145 | 0.274531958 | 0.05522967 | 0.88560316 | 0.02955 | 0.013812577 | 0.055229667 | 0.466243562 | 0.029772718 | -1.526181518 | FALSE | TRUE | FALSE |
| LIAS | 0.0016235 | 0.8916675 | 1 | 0.015124625 | 0.015124625 | 0.015124625 | 0.00358149 | 0.898464782 | 1 | 0.003581495 | 0.003581495 | 0.003581495 | 0.453309269 | -0.343605401 | FALSE | TRUE | FALSE |
| SPTA1 | 0.0016127 | 0.89167472 | 1 | 0.013720703 | 0.03081657 | 0.456678267 | 0.00326721 | 0.89858623 | 1 | 0.028114539 | 0.248313529 | 4.770419888 | 0.493609684 | -0.306616329 | FALSE | TRUE | FALSE |
| PNPLA6 | 0.0016047 | 0.89168193 | 0.52075 | 0.01391728 | 0.01391728 | 0.267904359 | 0.2424887 | 0.841305909 | 0.3364 | 0.242488702 | 21.04062544 | 43.14053762 | 0.006617463 | -2.179308479 | FALSE | TRUE | FALSE |
| TYRO3 | 0.0016009 | 0.89168914 | 0.515 | 0.006402298 | 0.02560974 | 0.125567843 | 0.01525253 | 0.895068391 | 0.5108 | 0.01525253 | 0.061014864 | 0.299844036 | 0.104962291 | -0.978966699 | FALSE | TRUE | FALSE |
| NDOR1 | 0.001593 | 0.89169635 | 1 | 0.014181694 | 0.014181694 | 0.279168604 | 0.0034857 | 0.898518792 | 0.0175 | 0.003485698 | 0.003485698 | 0.06874648 | 0.457017533 | -0.340067138 | FALSE | TRUE | FALSE |
| CASKIN2 | 0.0015657 | 0.89170356 | 0.9982 | 86.0839184 | 8029.132216 | 80234.56029 | 0.02275109 | 0.893001944 | 1 | 145462.9502 | 169004839.4 | 20730982053 | 0.06881881 | -1.162292842 | FALSE | TRUE | FALSE |
| ACD | 0.0015472 | 0.89171077 | 1 | 0.001547171 | 0.001547171 | 0.03048489 | 0.0022005 | 0.899002491 | 0.50355 | 0.019363485 | 0.019363485 | 0.074534311 | 0.703100355 | -0.152982683 | FALSE | TRUE | FALSE |
| ANXA1 | 0.0015307 | 0.89171798 | 1 | 0.001530673 | 0.001530673 | 0.001530673 | 0.15010524 | 0.864529837 | 0.24625 | 0.016032949 | 0.150105243 | 0.150105243 | 0.010197332 | -1.991513439 | FALSE | TRUE | FALSE |
| HEXB | 0.0015275 | 0.89172519 | 1 | 0.01333489 | 0.053282633 | 0.120815559 | 0.98967273 | 0.58944806 | 0.19535 | 0.109504912 | 3.955937795 | 3.955937795 | 0.001543401 | -2.811521158 | FALSE | TRUE | FALSE |
| ALPK3 | 0.0015233 | 0.89173239 | 1 | 0.001523293 | 0.029325954 | 0.051095303 | 0.00308089 | 0.898646853 | 0.52305 | 0.026050091 | 0.026050091 | 0.50377278 | 0.494433139 | -0.305892429 | FALSE | TRUE | FALSE |
| UROC1 | 0.0015089 | 0.8917396 | 0.87855 | 0.024102085 | 0.20918804 | 0.473800446 | 0.00333015 | 0.898539032 | 0.5158 | 0.013309379 | 0.013309379 | 0.112722492 | 0.453101999 | -0.343804022 | FALSE | TRUE | FALSE |
| OPLAH | 0.001493 | 0.8917468 | 0.52255 | 0.001493008 | 0.001493008 | 0.028848907 | 0.29664261 | 0.826248705 | 0.3207 | 0.032915829 | 2.740450921 | 5.755345388 | 0.00503302 | -2.298171342 | FALSE | TRUE | FALSE |
| SH3TC1 | 0.0014898 | 0.89175401 | 1 | 1.04059E+25 | 6.17831E+31 | 4.6357E+34 | 2.4842E-08 | 0.901718936 | 1 | 2.99755E+25 | 2.03025E+32 | 1.60004E+35 | 59972.17016 | 4.777949765 | FALSE | TRUE | FALSE |
| BEST1 | 0.001489 | 0.89176121 | 0.87415 | 0.00594985 | 0.023780448 | 0.029512276 | 0.01462138 | 0.895254605 | 0.6919 | 0.014621379 | 0.23369764 | 0.23369764 | 0.101838552 | -0.992087786 | FALSE | TRUE | FALSE |
| CNGA3 | 0.0014859 | 0.89176841 | 0.891 | 0.005934138 | 0.094713846 | 0.378533295 | 4.15723343 | 0.346394485 | 0.2986 | 4.157233116 | 266.1585002 | 1065.292801 | 0.000357428 | -3.446811936 | FALSE | FALSE | FALSE |
| BAZ2B | 0.0014753 | 0.89177561 | 0.053 | 0.001475262 | 0.028453191 | 0.049275891 | 0.00309419 | 0.89864012 | 1 | 0.012365386 | 0.049428454 | 0.957421665 | 0.476784716 | -0.321677675 | FALSE | TRUE | FALSE |
| AHI1 | 0.0014624 | 0.89178281 | 1 | 0.011965449 | 0.100490425 | 0.402816158 | 0.02418851 | 0.892711336 | 0.879 | 0.20630677 | 7.218146658 | 7.218146658 | 0.060458813 | -1.218540381 | FALSE | TRUE | FALSE |
| GLB1 | 0.0014551 | 0.89179001 | 0.99445 | 2.544212966 | 394.5431393 | 3928.553201 | 0.00063923 | 0.899673803 | 1 | 6524862755 | 1.83756E+14 | 9.21617E+15 | 2.276293717 | 0.3572283 | FALSE | TRUE | FALSE |
| POSTN | 0.0014486 | 0.89179721 | 0.9889 | 4.467135189 | 4289.316318 | 44299.44041 | 0.00045815 | 0.899792552 | 0.99915 | 31.18111727 | 12646.51349 | 131858.4116 | 3.161736579 | 0.499925684 | FALSE | TRUE | FALSE |
| INTS1 | 0.001444 | 0.89180441 | 0.05685 | 0.001444032 | 0.027553644 | 0.048418268 | 0.01198925 | 0.896070948 | 0.528 | 0.011989249 | 0.057526137 | 0.230037388 | 0.12044387 | -0.919215299 | FALSE | TRUE | FALSE |
| CDC42BPG | 0.0014225 | 0.8918116 | 0.04905 | 0.001422506 | 0.001422506 | 0.047806548 | 0.01143548 | 0.896281975 | 0.88 | 0.045731281 | 0.393319426 | 1.90036842 | 0.124393993 | -0.90520059 | FALSE | TRUE | FALSE |
| WDR17 | 0.0014119 | 0.8918188 | 1 | 0.001411864 | 0.001411864 | 0.047496112 | 0.02377001 | 0.892785985 | 0.5143 | 0.023770007 | 0.023770007 | 0.466080422 | 0.059396876 | -1.226236395 | FALSE | TRUE | FALSE |
| CHD3 | 0.0014105 | 0.89182599 | 0.9701 | 1.917018177 | 180.7032991 | 1819.658664 | 0.00378193 | 0.898403957 | 0.97745 | 1.303740901 | 60.60488891 | 421.0109353 | 0.372950101 | -0.428349271 | FALSE | TRUE | FALSE |
| MACC1 | 0.0014036 | 0.89183318 | 1 | 0.011693111 | 0.011693111 | 0.047425779 | 0.02001896 | 0.893850534 | 0.7562 | 0.020018961 | 0.173516109 | 0.395557029 | 0.070115779 | -1.154184238 | FALSE | TRUE | FALSE |
| ATR | 0.0013977 | 0.89184038 | 1 | 0.101104353 | 8.096601039 | 29.73468881 | 0.00320467 | 0.898613182 | 0.5283 | 0.027339534 | 0.061593888 | 0.912049018 | 0.436150965 | -0.360363162 | FALSE | TRUE | FALSE |
| SLC26A3 | 0.0013612 | 0.89184757 | 0.87795 | 0.005436254 | 0.021716817 | 0.046043007 | 0.01387671 | 0.895482953 | 0.1447 | 0.013876709 | 0.055446593 | 0.117577822 | 0.098090147 | -1.008374614 | FALSE | TRUE | FALSE |
| LYST | 0.0013579 | 0.89185476 | 0.09635 | 0.001357866 | 0.005264885 | 0.044761616 | 0.00177654 | 0.899149429 | 0.5425 | 0.001776541 | 0.014706205 | 0.058778164 | 0.764331044 | -0.116718501 | FALSE | TRUE | FALSE |

| | | | | | | | | | | | | | | | | | |
|---|---|---|---|---|---|---|---|---|---|---|---|---|---|---|---|---|---|
| CC2D1B | 0.0013574 | 0.89186195 | 1 | 0.001357391 | 0.001357391 | 0.026648704 | 0.00273362 | 0.898774615 | 1 | 0.02242148 | 0.02242148 | 0.441159638 | 0.496554229 | -0.304033315 | FALSE | TRUE | FALSE |
| ACE | 0.0013552 | 0.89186914 | 0.2776 | 0.001355248 | 0.005416968 | 0.045614924 | 0.01250885 | 0.895894481 | 0.8936 | 0.106626984 | 3.730170206 | 14.91869066 | 0.108343185 | -0.965198402 | FALSE | TRUE | FALSE |
| MYOM1 | 0.0013476 | 0.89187632 | 0.5252 | 0.002710031 | 0.103752297 | 0.414756662 | 0.00907942 | 0.896993561 | 0.5182 | 0.009079419 | 0.175681195 | 0.619390366 | 0.148426089 | -0.828489755 | FALSE | TRUE | FALSE |
| CD93 | 0.001346 | 0.89188351 | 1 | 8.30795E+40 | 4.34431E+49 | 2.83641E+53 | 0.00247082 | 0.898875267 | 1 | 1.15218E+44 | 7.9999E+52 | 1.16386E+58 | 0.544767634 | -0.263788703 | FALSE | TRUE | FALSE |
| C8orf58 | 0.0013431 | 0.8918907 | 1 | 0.001343094 | 0.001343094 | 0.026638124 | 0.00151774 | 0.89921609 | 1 | 0.014020857 | 0.014020857 | 0.014020857 | 0.884929436 | -0.053091359 | FALSE | TRUE | FALSE |
| ATP8B1 | 0.001341 | 0.89189788 | 1 | 0.001341027 | 0.001341027 | 0.026122131 | 0.00290042 | 0.898714134 | 1 | 0.002900423 | 0.002900423 | 0.056675229 | 0.462355646 | -0.335023835 | FALSE | TRUE | FALSE |
| CRB1 | 0.0013322 | 0.89190507 | 0.5142 | 0.001332217 | 0.011234253 | 0.026030334 | 0.18344878 | 0.856136353 | 0.39595 | 0.032089573 | 16.55204648 | 58.03736054 | 0.007262066 | -2.138939792 | FALSE | TRUE | FALSE |
| NUP210 | 0.0013246 | 0.89191225 | 1 | 0.010872862 | 0.091606599 | 1.749833434 | 0.02381169 | 0.892778525 | 0.5197 | 0.023811692 | 0.023811692 | 0.457309666 | 0.055629503 | -1.254694823 | FALSE | TRUE | FALSE |
| HEATR6 | 0.0013052 | 0.89191943 | 0.7625 | 0.005219319 | 0.020876616 | 0.175774366 | 0.18269509 | 0.856303212 | 0.14405 | 0.182695092 | 1.597646505 | 3.583757157 | 0.007144128 | -2.146050765 | FALSE | TRUE | FALSE |
| UTP20 | 0.0013044 | 0.89192661 | 0.9968 | 7.920499014 | 1416.946549 | 13626.42253 | 0.00286623 | 0.89872758 | 0.9927 | 18.34621278 | 1701.14127 | 16962.19764 | 0.455103402 | -0.341889918 | FALSE | TRUE | FALSE |
| PFKP | 0.0012983 | 0.89193379 | 0.48035 | 0.001298325 | 0.020698934 | 0.082678585 | 0.00107903 | 0.899375745 | 0.51725 | 0.001079028 | 0.004310979 | 0.021149745 | 1.203235728 | 0.080350719 | FALSE | TRUE | FALSE |
| OPA1 | 0.0012964 | 0.89194097 | 0.9698 | 0.020674528 | 0.10161108 | 0.405826358 | 12.8869803 | 0.170532342 | 0.1828 | 0.806125677 | 51.54517088 | 206.2211366 | 0.000100599 | -3.997405385 | FALSE | FALSE | FALSE |
| VWA3B | 0.0012886 | 0.89194815 | 0.6937 | 0.016703898 | 1.131820844 | 2.648674552 | 0.00537032 | 0.898030818 | 0.52075 | 0.043545399 | 0.362190058 | 0.851854102 | 0.239942149 | -0.619893456 | FALSE | TRUE | FALSE |
| KLC4 | 0.0012785 | 0.89195533 | 1 | 3.26129E+35 | 1.19851E+44 | 6.79101E+47 | 0.22946734 | 0.845039544 | 1 | 3.03694E+33 | 1.04412E+42 | 5.78255E+45 | 0.005571717 | -2.254010935 | FALSE | TRUE | FALSE |
| MCCC1 | 0.0012785 | 0.8919625 | 0.5047 | 0.001278461 | 0.011302969 | 0.025210652 | 0.02855888 | 0.891587737 | 0.5008 | 0.02855888 | 0.45589831 | 0.45589831 | 0.044765797 | -1.349053679 | FALSE | TRUE | FALSE |
| TBC1D31 | 0.0012757 | 0.89196968 | 1 | 0.010731305 | 0.042899046 | 0.210697909 | 0.00275308 | 0.898761181 | 1 | 0.023471324 | 0.023471324 | 0.461850402 | 0.463379219 | -0.334063447 | FALSE | TRUE | FALSE |
| CELSR3 | 0.001274 | 0.89197685 | 0.55425 | 0.005090982 | 0.093141589 | 0.3722838 | 3082.24242 | 0.000723438 | 0.20985 | 192.9265472 | 125776.2472 | 1313510.877 | 4.13345E-07 | -6.383687213 | FALSE | FALSE | FALSE |
| NUP88 | 0.0012658 | 0.89198403 | 0.75775 | 0.011111464 | 0.100116745 | 0.218794433 | 0.00188034 | 0.899109393 | 0.5162 | 0.016667683 | 0.016667683 | 0.063595048 | 0.67315009 | -0.171888092 | FALSE | TRUE | FALSE |
| ZNF721 | 0.0012459 | 0.8919912 | 1 | 0.085457186 | 0.735532087 | 1.682301691 | 0.02121075 | 0.893490493 | 0.9428 | 0.180762736 | 14.15995457 | 14.15995457 | 0.058738408 | -1.231077827 | FALSE | TRUE | FALSE |
| RGS3 | 0.0012439 | 0.89199837 | 0.9405 | 0.088038649 | 6.898620082 | 14.97748365 | 0.00287508 | 0.898720857 | 1 | 0.024289079 | 0.024289079 | 0.474170138 | 0.432644114 | -0.3638692 | FALSE | TRUE | FALSE |
| TRPM8 | 0.0012414 | 0.89200554 | 0.8183 | 0.010538806 | 0.366990931 | 0.822318133 | 0.00691844 | 0.897572933 | 0.75835 | 0.006918438 | 0.061037631 | 0.233452615 | 0.179436845 | -0.746088374 | FALSE | TRUE | FALSE |
| SLC38A10 | 0.0012283 | 0.89201271 | 1 | 0.010113694 | 0.010113694 | 0.196254161 | 0.0194108 | 0.893982238 | 0.51435 | 0.019410803 | 0.019410803 | 0.378014268 | 0.063280158 | -1.198732446 | FALSE | TRUE | FALSE |
| COL5A1 | 0.0012252 | 0.89201988 | 0.29435 | 0.001225241 | 0.019506935 | 0.093834617 | 0.36959605 | 0.802294282 | 0.4126 | 0.092553326 | 5.898247793 | 94.22192919 | 0.003315081 | -2.4795058 | FALSE | TRUE | FALSE |
| AHNAK | 0.0011989 | 0.89202705 | 1 | 0.805619308 | 70.03769232 | 244.3288122 | 0.00280583 | 0.898734302 | 0.7751 | 0.024296077 | 0.445214518 | 7.136370639 | 0.427284379 | -0.369282984 | FALSE | TRUE | FALSE |
| IFT122 | 0.0011879 | 0.89203422 | 0.3389 | 0.000297351 | 0.004746671 | 0.023095967 | 0.00025441 | 0.899943895 | 0.7598 | 0.002136497 | 0.018415915 | 0.072030163 | 4.66919233 | 0.669241763 | FALSE | TRUE | FALSE |
| CSPP1 | 0.0011774 | 0.89204138 | 0.7589 | 0.009449331 | 0.077839582 | 0.317664672 | 0.02075342 | 0.893652421 | 0.03095 | 0.02075342 | 0.02075342 | 0.406679622 | 0.05673522 | -1.246147258 | FALSE | TRUE | FALSE |
| SH3BP2 | 0.0011534 | 0.89204855 | 0.8785 | 0.040046634 | 0.357177013 | 0.357177013 | 0.09125803 | 0.877871414 | 0.2586 | 0.022836558 | 0.091258034 | 0.205246234 | 0.012638905 | -1.898290544 | FALSE | TRUE | FALSE |
| GPR179 | 0.0011483 | 0.89205571 | 0.8947 | 0.009290352 | 0.654779234 | 5.697492394 | 0.01608309 | 0.894823975 | 0.7812 | 0.27148687 | 20.96380724 | 190.7703658 | 0.071399257 | -1.14630631 | FALSE | TRUE | FALSE |
| KRT32 | 0.001128 | 0.89206288 | 1 | 3.53193E+35 | 1.29657E+44 | 7.34214E+47 | 1.52395309 | 0.53372955 | 1 | 2.2991E+37 | 2.37707E+45 | 1.23088E+49 | 0.000740207 | -3.130647056 | FALSE | FALSE | FALSE |
| DSG4 | 0.001111 | 0.89207004 | 0.7555 | 0.009556014 | 0.084364336 | 0.322289576 | 0.02180473 | 0.893291133 | 0.5153 | 0.021804735 | 0.021804735 | 0.428682525 | 0.050952297 | -1.292836231 | FALSE | TRUE | FALSE |
| PKP4 | 0.0010785 | 0.8920772 | 1 | 0.009556745 | 0.009556745 | 0.186426582 | 0.00232564 | 0.898935569 | 1 | 0.020752577 | 0.190066108 | 0.698771016 | 0.463721164 | -0.333743083 | FALSE | TRUE | FALSE |
| GPSM1 | 0.0010737 | 0.89208436 | 0.5081 | 0.004289544 | 0.004289544 | 0.036358197 | 0.14080739 | 0.86664017 | 0.77385 | 0.562725381 | 20.87771208 | 83.47777811 | 0.007625642 | -2.117723583 | FALSE | TRUE | FALSE |
| SLC26A11 | 0.0010726 | 0.89209152 | 0.87995 | 0.068838041 | 17.7492347 | 71.1685313 | 4.24422638 | 0.341437751 | 0.2027 | 1.061007028 | 16.98193784 | 20.93612663 | 0.000252728 | -3.59734616 | FALSE | FALSE | FALSE |
| NF1 | 0.0010648 | 0.89209868 | 0.5338 | 0.004255217 | 0.020251386 | 0.14150935 | 1.26961685 | 0.559031542 | 0.0713 | 0.141995256 | 5.077999524 | 22.65276843 | 0.000838683 | -3.076402007 | FALSE | FALSE | FALSE |
| TRPM7 | 0.0010501 | 0.89210584 | 1 | 0.00895258 | 0.00895258 | 0.17357719 | 0.00230692 | 0.89894896 | 1 | 0.019904429 | 0.019904429 | 0.387340059 | 0.455204179 | -0.341793759 | FALSE | TRUE | FALSE |
| SOX30 | 0.0010231 | 0.892113 | 0.51535 | 0.004085554 | 0.004085554 | 0.080570311 | 0.00226226 | 0.898969041 | 0.51485 | 0.02092589 | 0.02092589 | 0.076521446 | 0.452225412 | -0.344645037 | FALSE | TRUE | FALSE |
| CR1 | 0.0010102 | 0.89212016 | 1 | 0.001010225 | 0.019234367 | 0.033690751 | 0.00211453 | 0.899035921 | 0.7671 | 0.017801536 | 0.071191015 | 1.363319782 | 0.477753798 | -0.320795852 | FALSE | TRUE | FALSE |
| IFT172 | 0.0010059 | 0.89212731 | 0.05285 | 0.000251713 | 0.001215202 | 0.019401605 | 0.00017674 | 0.900068583 | 1 | 0.012632032 | 0.44344544 | 0.977737183 | 5.691251986 | 0.755207815 | FALSE | TRUE | FALSE |
| CEP250 | 0.0009902 | 0.89213447 | 1 | 0.008073791 | 0.018939221 | 0.268523297 | 0.00219845 | 0.899009179 | 0.05785 | 0.00219845 | 0.042270719 | 0.073309512 | 0.450408958 | -0.346392981 | FALSE | TRUE | FALSE |
| PRSS36 | 0.0009787 | 0.89214162 | 0.993 | 0.066080394 | 4.22556738 | 16.90594678 | 0.00067245 | 0.899653985 | 1 | 0.01074725 | 0.042981156 | 0.211545131 | 1.455384916 | 0.162977869 | FALSE | TRUE | FALSE |
| GUCY2D | 0.0009724 | 0.89214877 | 1 | 0.015522282 | 0.062039969 | 0.302254779 | 0.0022381 | 0.898995803 | 1 | 0.008942275 | 0.008942275 | 0.174680529 | 0.434479628 | -0.362030582 | FALSE | TRUE | FALSE |
| ATAT1 | 0.0009624 | 0.89215592 | 1 | 9.61964E+36 | 2.30411E+45 | 1.07891E+49 | 0.00040488 | 0.89982549 | 1 | 2.45185E+33 | 3.07983E+40 | 1.29938E+44 | 2.377121316 | 0.376051346 | FALSE | TRUE | FALSE |
| NID2 | 0.0009582 | 0.89216307 | 0.98615 | 0.293587673 | 10.67873926 | 42.6975525 | 0.02028658 | 0.893740553 | 0.5201 | 0.020286584 | 0.081082009 | 0.683197834 | 0.04723142 | -1.325768997 | FALSE | TRUE | FALSE |
| TBC1D8 | 0.0009443 | 0.89217022 | 1 | 0.007664781 | 0.007664781 | 0.149528983 | 0.00210424 | 0.899049287 | 0.0308 | 0.002104237 | 0.002104237 | 0.041172224 | 0.44876388 | -0.347982106 | FALSE | TRUE | FALSE |
| KIF21A | 0.000935 | 0.89217737 | 1 | 0.008350919 | 0.076549718 | 1.487352678 | 0.13066005 | 0.868798246 | 0.54495 | 0.169934361 | 2.71341848 | 10.84669386 | 0.007156301 | -2.145311392 | FALSE | TRUE | FALSE |
| AEBP1 | 0.0009212 | 0.89218452 | 0.6372 | 0.003680969 | 0.031524144 | 0.153064442 | 0.08066532 | 0.880163987 | 0.4843 | 0.044883238 | 0.716658752 | 2.86476722 | 0.011420153 | -1.942328065 | FALSE | TRUE | FALSE |
| STRA6 | 0.0008918 | 0.89219167 | 0.2641 | 0.000891836 | 0.006848989 | 0.007544543 | 0.01099319 | 0.896471215 | 0.29275 | 0.005731224 | 0.086572088 | 0.34626756 | 0.081126229 | -1.090838712 | FALSE | TRUE | FALSE |
| MTMR3 | 0.0008907 | 0.89219882 | 0.87765 | 0.014206008 | 0.056750742 | 0.277383562 | 0.63843039 | 0.695314303 | 0.00705 | 0.03260363 | 0.130273792 | 0.638430386 | 0.001395164 | -2.855374613 | FALSE | TRUE | FALSE |
| CLTCL1 | 0.0008872 | 0.89220596 | 0.8773 | 0.006975858 | 0.056267003 | 0.234436848 | 0.00245996 | 0.89888197 | 0.7546 | 0.009837649 | 0.079202264 | 0.331072625 | 0.36064548 | -0.442919507 | FALSE | TRUE | FALSE |
| ITGAE | 0.0008808 | 0.89221311 | 0.99815 | 1847.146317 | 2920494.862 | 30029978.41 | 0.4929972 | 0.757797156 | 0.9996 | 1504255.111 | 4921117628 | 1.76493E+11 | 0.001786551 | -2.747984498 | FALSE | TRUE | FALSE |
| GEMIN5 | 0.0008666 | 0.89222025 | 1 | 0.029262331 | 0.253739629 | 1.235149577 | 0.00207595 | 0.89906265 | 1 | 0.002075948 | 0.002075948 | 0.069837517 | 0.41742874 | -0.379417653 | FALSE | TRUE | FALSE |
| USHBP1 | 0.00086 | 0.89222739 | 0.51055 | 0.007301142 | 0.007301142 | 0.143692456 | 0.1003961 | 0.875839132 | 0.5068 | 0.100396095 | 0.907011849 | 0.907011849 | 0.008566429 | -2.067200163 | FALSE | TRUE | FALSE |
| KRT75 | 0.0008504 | 0.89223453 | 0.8909 | 0.067629416 | 23.81386933 | 340.097596 | 0.46091099 | 0.770692835 | 0.46985 | 0.460910988 | 118.2964282 | 833.2185678 | 0.001845087 | -2.733983097 | FALSE | TRUE | FALSE |
| IGHMBP2 | 0.0008294 | 0.89224168 | 0.8759 | 0.00741316 | 0.271507763 | 0.271507763 | 0.01003771 | 0.89674343 | 0.3906 | 0.010037711 | 0.362191467 | 0.444386734 | 0.08262499 | -1.082888579 | FALSE | TRUE | FALSE |
| CTNS | 0.0008162 | 0.89224882 | 0.20765 | 8.84069E-05 | 0.003262912 | 0.01304734 | 5.9169E-06 | 0.900823204 | 0.8771 | 5.91689E-06 | 9.45614E-05 | 9.45614E-05 | 137.9442337 | 2.139703551 | FALSE | TRUE | FALSE |
| SCN8A | 0.0008026 | 0.89225595 | 0.8972 | 0.071791085 | 7.106347281 | 73.32856757 | 0.00063071 | 0.899693613 | 0.5285 | 0.005921169 | 0.005921169 | 0.114215105 | 1.272595069 | 0.104690236 | FALSE | TRUE | FALSE |
| SCN3A | 0.0007963 | 0.89226309 | 0.9446 | 0.20214685 | 12.88976885 | 51.52141798 | 14.4715094 | 0.160525634 | 0.1685 | 0.906179986 | 57.85301677 | 280.35049 | 5.50258E-05 | -4.259433655 | FALSE | FALSE | FALSE |
| KCNQ5 | 0.000774 | 0.89227023 | 0.5161 | 0.003089815 | 0.003089815 | 0.060669803 | 0.96193325 | 0.591869697 | 0.00015 | 0.007133718 | 0.028492453 | 0.140380996 | 0.000804602 | -3.094419058 | FALSE | TRUE | FALSE |
| SLX4 | 0.000756 | 0.89227737 | 0.05385 | 0.000756024 | 0.014494462 | 0.025364071 | 0.00158262 | 0.8991961 | 1 | 0.012912708 | 0.108140483 | 2.083630538 | 0.47770429 | -0.320840859 | FALSE | TRUE | FALSE |
| CEP104 | 0.0007488 | 0.8922845 | 1 | 0.000748778 | 0.000748778 | 0.014687013 | 0.41126669 | 0.788009252 | 0.32035 | 0.411266688 | 7.910256058 | 31.62985628 | 0.001820663 | -2.739770546 | FALSE | TRUE | FALSE |
| KNTC1 | 0.0007484 | 0.89229164 | 0.0519 | 0.000748399 | 0.001924483 | 0.014455068 | 0.00026633 | 0.899917606 | 1 | 0.002050621 | 0.002050621 | 0.039770766 | 2.810053898 | 0.44871465 | FALSE | TRUE | FALSE |
| ATP13A4 | 0.0007449 | 0.89229877 | 0.6939 | 0.000744886 | 0.01189048 | 0.058215128 | 0.13619397 | 0.867568341 | 0.6573 | 0.292221895 | 18.68294798 | 164.4821993 | 0.005469305 | -2.262067832 | FALSE | TRUE | FALSE |
| TNXB | 0.0007365 | 0.8923059 | 0.54785 | 0.000736471 | 0.013286829 | 0.111939737 | 4.3787E-05 | 0.900408269 | 1 | 4.37866E-05 | 0.000799369 | 0.001451183 | 16.81955692 | 1.225814551 | FALSE | TRUE | FALSE |
| SLIT2 | 0.0007105 | 0.89231304 | 1 | 0.026283572 | 0.419354866 | 2.03317473 | 0.00178547 | 0.899142758 | 0.0397 | 0.001785475 | 0.001785475 | 0.060078333 | 0.397948205 | -0.40017345 | FALSE | TRUE | FALSE |
| RALGAPA2 | 0.0006998 | 0.89232017 | 0.5249 | 0.000699849 | 0.005981209 | 0.050156294 | 0.00014374 | 0.900120991 | 1 | 0.001242916 | 0.001242916 | 0.024155511 | 4.869007986 | 0.687440487 | FALSE | TRUE | FALSE |
| CCDC68 | 0.0006979 | 0.8923273 | 1 | 0.006295422 | 0.006295422 | 0.006295422 | 0.00090235 | 0.899528298 | 1 | 0.000902348 | 0.000902348 | 0.000902348 | 0.773409999 | -0.111590217 | FALSE | TRUE | FALSE |
| ATP10A | 0.0006934 | 0.89233443 | 0.52355 | 0.000693417 | 0.000693417 | 0.013361654 | 0.00210534 | 0.899042604 | 0.51825 | 0.002105344 | 0.002105344 | 0.040747342 | 0.329360493 | -0.482328497 | FALSE | TRUE | FALSE |
| SLIT1 | 0.0006844 | 0.89234155 | 1 | 0.000684415 | 0.000684415 | 0.023015021 | 0.00161805 | 0.899189435 | 1 | 0.001618045 | 0.001618045 | 0.031335411 | 0.422988804 | -0.373671128 | FALSE | TRUE | FALSE |
| TCF7L2 | 0.0006824 | 0.89234868 | 1 | 0.000682408 | 0.000682408 | 0.013485682 | 0.00629792 | 0.897812597 | 0.25465 | 0.00629792 | 0.025156722 | 0.124637143 | 0.108354524 | -0.96515295 | FALSE | TRUE | FALSE |
| MYO3B | 0.0006741 | 0.89235581 | 0.70665 | 0.002693177 | 0.172003562 | 0.68787848 | 0.00087537 | 0.899541542 | 0.9158 | 0.027998491 | 3.672891331 | 30.92007996 | 0.770076387 | -0.113466193 | FALSE | TRUE | FALSE |
| CHRND | 0.0006667 | 0.89236293 | 0.40435 | 0.000666672 | 0.023660781 | 0.094706422 | 0.00169682 | 0.899169435 | 0.60865 | 0.001696816 | 0.108938049 | 0.436426631 | 0.392896116 | -0.405722264 | FALSE | TRUE | FALSE |
| CGN | 0.0006627 | 0.89237006 | 0.52055 | 0.005565124 | 0.005565124 | 0.108170412 | 0.8733028 | 0.602047998 | 0.36285 | 0.873302802 | 74.88312682 | 718.6262596 | 0.000758872 | -3.119831402 | FALSE | TRUE | FALSE |
| SPG7 | 0.00066 | 0.89237718 | 0.33005 | 0.000660041 | 0.051513036 | 0.051513036 | 0.00273315 | 0.898781331 | 0.01725 | 0.000316025 | 0.002733147 | 0.006207055 | 0.241495036 | -0.617091793 | FALSE | TRUE | FALSE |

| | | | | | | | | | | | | | | | | | |
|---|---|---|---|---|---|---|---|---|---|---|---|---|---|---|---|---|---|
| TPCN1 | 0.0006577 | 0.8923843 | 1 | 0.010486145 | 0.167344495 | 0.354244942 | 0.39367414 | 0.794207825 | 0.2001 | 0.098511802 | 0.393674139 | 1.93479848 | 0.001670773 | -2.777082478 | FALSE | TRUE | FALSE |
| WNK1 | 0.0006531 | 0.89239143 | 1 | 0.022129534 | 0.1925344 | 3.502113828 | 0.00138888 | 0.899276015 | 0.98565 | 0.942147602 | 72.02459561 | 329.5062313 | 0.470205849 | -0.327711973 | FALSE | TRUE | FALSE |
| ANK3 | 0.0006495 | 0.89239855 | 1 | 0.005965754 | 0.110229088 | 0.196728296 | 0.00149148 | 0.899222751 | 0.77315 | 0.013740313 | 0.256223666 | 4.299771949 | 0.435457685 | -0.361054041 | FALSE | TRUE | FALSE |
| CACNA1E | 0.0006477 | 0.89240567 | 0.5333 | 0.002584348 | 0.049047521 | 0.656579962 | 0.02717663 | 0.891906593 | 0.4097 | 0.011624601 | 0.433105688 | 2.071472941 | 0.023833751 | -1.622807602 | FALSE | TRUE | FALSE |
| MYH6 | 0.0006131 | 0.89241279 | 0.75145 | 0.009774739 | 9.962344292 | 159.3748068 | 2.1609E-08 | 0.901731614 | 0.95965 | 2.17808E-05 | 0.012169721 | 0.194116895 | 28374.2181 | 4.452923903 | FALSE | TRUE | FALSE |
| ZNF318 | 0.0006039 | 0.8924199 | 0.0583 | 0.000603936 | 0.011611352 | 0.020157729 | 0.00085867 | 0.899548163 | 0.0502 | 0.000858674 | 0.016586086 | 0.028723397 | 0.703335467 | -0.152837482 | FALSE | TRUE | FALSE |
| TANC1 | 0.0006009 | 0.89242702 | 1 | 0.000600867 | 0.011567127 | 0.020149536 | 0.00126632 | 0.899315929 | 0.94045 | 0.100283103 | 8.790629536 | 17.93304546 | 0.474499736 | -0.323764025 | FALSE | TRUE | FALSE |
| DNMT3B | 0.0005939 | 0.89243414 | 0.52235 | 0.002370264 | 0.002370264 | 0.046369181 | 0.00144613 | 0.899242731 | 0.02505 | 0.001446127 | 0.001446127 | 0.028364883 | 0.410701804 | -0.386473389 | FALSE | TRUE | FALSE |
| SOX6 | 0.0005762 | 0.89244125 | 1 | 0.005393694 | 0.051820441 | 0.106154412 | 0.0014834 | 0.899236072 | 1 | 0.001483401 | 0.001483401 | 0.029251133 | 0.388419082 | -0.410699443 | FALSE | TRUE | FALSE |
| BAIAP3 | 0.0005602 | 0.89244837 | 1 | 0.004651572 | 0.004651572 | 0.0903169 | 0.00133078 | 0.899289323 | 1 | 0.00531992 | 0.00531992 | 0.103657214 | 0.420936538 | -0.375783375 | FALSE | TRUE | FALSE |
| PHF12 | 0.0005558 | 0.89245548 | 1 | 0.005203303 | 0.005203303 | 0.101700001 | 0.00139267 | 0.89926936 | 1 | 0.001392666 | 0.001392666 | 0.027294598 | 0.399095659 | -0.398922996 | FALSE | TRUE | FALSE |
| SETD5 | 0.0005522 | 0.89246259 | 0.6389 | 0.005255653 | 0.079854079 | 0.17649988 | 0.00249044 | 0.898861857 | 0.8812 | 0.009944675 | 0.092555839 | 0.452389204 | 0.221737155 | -0.654161529 | FALSE | TRUE | FALSE |
| PKD1 | 0.0005405 | 0.89246971 | 0.9538 | 0.176790849 | 39.59753707 | 286.5090803 | 0.00485125 | 0.898166782 | 1 | 4121659.062 | 24538014042 | 1.14051E+12 | 0.11140902 | -0.953079646 | FALSE | TRUE | FALSE |
| RTEL1 | 0.0005364 | 0.89247682 | 0.03955 | 0.000536424 | 0.000536424 | 0.010401436 | 0.00509557 | 0.898098841 | 0.5191 | 0.005095571 | 0.005095571 | 0.099167904 | 0.105272691 | -0.977684274 | FALSE | TRUE | FALSE |
| TLN2 | 0.0005176 | 0.89248393 | 0.07465 | 0.000517599 | 0.009778175 | 0.017277371 | 0.0012193 | 0.899329227 | 1 | 0.010999893 | 0.023188747 | 0.210784608 | 0.424504571 | -0.372117629 | FALSE | TRUE | FALSE |
| TEP1 | 0.0005096 | 0.89249104 | 0.94395 | 0.032405434 | 2.271040473 | 19.68959254 | 13491.0515 | 0.000149291 | 0.000025 | 0.062289675 | 4.381390352 | 38.08513417 | 3.77698E-08 | -7.422855093 | FALSE | FALSE | FALSE |
| TYR | 0.000466 | 0.89249814 | 0.92675 | 0.014054972 | 0.473665228 | 1.893249511 | 56.0216957 | 0.049876983 | 0.17495 | 2.999189394 | 284.3499994 | 2012.127681 | 8.31896E-06 | -5.079931113 | FALSE | FALSE | FALSE |
| PAMR1 | 0.0004539 | 0.89250525 | 0.5109 | 0.000453921 | 0.000453921 | 0.001801457 | 0.00017152 | 0.90008169 | 0.01945 | 0.000171525 | 0.000171525 | 0.003386488 | 2.64638977 | 0.422653809 | FALSE | TRUE | FALSE |
| RPGRIP1 | 0.0004454 | 0.89251236 | 0.52645 | 0.000445378 | 0.000445378 | 0.008702123 | 0.00538552 | 0.898024011 | 0.15025 | 0.002601426 | 0.02152401 | 0.105518579 | 0.082699212 | -1.082498627 | FALSE | TRUE | FALSE |
| TECTA | 0.0004436 | 0.89251946 | 0.7183 | 0.003731652 | 0.237933331 | 1.991067088 | 0.31495348 | 0.820284203 | 0.1134 | 0.016467204 | 1.050681228 | 5.027730506 | 0.001408412 | -2.851270202 | FALSE | TRUE | FALSE |
| ZFHX4 | 0.0004348 | 0.89252657 | 0.942 | 0.041509115 | 4.364439605 | 46.33802442 | 0.0065628 | 0.897675775 | 1 | 0.573765777 | 55.51621718 | 185.2539475 | 0.066252747 | -1.178796108 | FALSE | TRUE | FALSE |
| ADCY6 | 0.0004336 | 0.89253367 | 0.13195 | 2.7111E-05 | 0.000433598 | 0.002100748 | 2.4266E-05 | 0.900525319 | 0.27955 | 2.42656E-05 | 9.69703E-05 | 0.000819504 | 17.86880744 | 1.252095569 | FALSE | TRUE | FALSE |
| CHD2 | 0.000431 | 0.89254077 | 0.0047 | 1.344E-06 | 1.28857E-05 | 0.000248794 | 0.00010123 | 0.900212575 | 0.00075 | 2.5647E-07 | 2.5647E-07 | 8.59495E-06 | 4.257498348 | 0.629154488 | FALSE | TRUE | FALSE |
| EPB41L3 | 0.0004293 | 0.89254788 | 0.7598 | 0.00388201 | 0.015500847 | 0.075728775 | 46.3434473 | 0.059401875 | 0.00135 | 0.148316913 | 2.368631662 | 9.469208787 | 9.26383E-06 | -5.033209192 | FALSE | FALSE | FALSE |
| FADS6 | 0.0004182 | 0.89255498 | 0.5042 | 0.003191316 | 0.003191316 | 0.008288013 | 0.00098053 | 0.899468657 | 1 | 0.000980532 | 0.000980532 | 0.000980532 | 0.426548694 | -0.370031384 | FALSE | TRUE | FALSE |
| POLE | 0.0004157 | 0.89256208 | 1 | 0.00041575 | 0.007864098 | 0.013887267 | 0.00358623 | 0.898458027 | 0.91005 | 0.118787638 | 4.80620636 | 19.22403092 | 0.115929577 | -0.93580575 | FALSE | TRUE | FALSE |
| VIT | 0.0004084 | 0.89256918 | 0.93915 | 0.001631213 | 0.026044881 | 0.032133622 | 0.07462898 | 0.881473277 | 0.04385 | 0.018670983 | 0.074628976 | 0.298373243 | 0.005472179 | -2.261839683 | FALSE | TRUE | FALSE |
| LZTR1 | 0.0003981 | 0.89257627 | 0.5204 | 0.000398108 | 0.000398108 | 0.007782187 | 0.00826441 | 0.897221877 | 0.9383 | 0.578920618 | 44.68183536 | 169.9852407 | 0.048171429 | -1.317210471 | FALSE | TRUE | FALSE |
| LTBP2 | 0.0003873 | 0.89258337 | 0.15025 | 0.000387342 | 0.001546607 | 0.013002733 | 5.3505E-06 | 0.900855475 | 0.8812 | 4.76888E-05 | 0.011785489 | 0.025394528 | 72.3943124 | 1.859704448 | FALSE | TRUE | FALSE |
| SLC19A3 | 0.0003819 | 0.89259047 | 0.50845 | 0.000381904 | 0.000381904 | 0.000817722 | 0.04003119 | 0.888950837 | 0.01185 | 0.040031188 | 0.040031188 | 0.793944613 | 0.00954016 | -2.020444341 | FALSE | TRUE | FALSE |
| ACAD9 | 0.0003798 | 0.89259756 | 0.6952 | 0.001516291 | 0.024190087 | 0.02986837 | 0.01732949 | 0.894498885 | 0.77425 | 0.069241943 | 1.106270747 | 4.423542198 | 0.021914522 | -1.659267994 | FALSE | TRUE | FALSE |
| MST1R | 0.0003732 | 0.89260466 | 0.51115 | 0.003141704 | 9.653415073 | 84.48726108 | 57.9322399 | 0.048226069 | 0.26265 | 6.506439962 | 528.163256 | 4927.787939 | 6.44116E-06 | -5.191035922 | FALSE | FALSE | FALSE |
| NOD2 | 0.0003643 | 0.89261175 | 0.51455 | 0.003031038 | 0.003031038 | 0.059238222 | 0.00089683 | 0.899534921 | 1 | 0.007570946 | 0.007570946 | 0.148372147 | 0.406221388 | -0.391237214 | FALSE | TRUE | FALSE |
| TET2 | 0.0003534 | 0.89261884 | 0.51835 | 0.002771718 | 0.022298115 | 0.054431063 | 0.00011352 | 0.900166804 | 0.75785 | 0.000888319 | 0.007134893 | 0.029879676 | 3.113198589 | 0.493206825 | FALSE | TRUE | FALSE |
| ABCB4 | 0.0003507 | 0.89262594 | 1 | 0.003154442 | 0.0291201 | 0.10618913 | 0.00094904 | 0.899488545 | 1 | 0.000949044 | 0.000949044 | 0.018580308 | 0.369547107 | -0.432330193 | FALSE | TRUE | FALSE |
| FAT2 | 0.0003506 | 0.89263303 | 0.84255 | 0.006413731 | 0.907697536 | 8.252071525 | 1.14475072 | 0.572957708 | 0.2636 | 0.135843187 | 34.65784135 | 312.6691917 | 0.000306273 | -3.513890583 | FALSE | FALSE | FALSE |
| VPS13D | 0.0003474 | 0.89264012 | 1 | 0.000347415 | 0.006389987 | 0.011400675 | 0.00081496 | 0.899581256 | 1 | 0.006970619 | 0.129460025 | 1.13654302 | 0.426295236 | -0.370289521 | FALSE | TRUE | FALSE |
| KRT76 | 0.0003453 | 0.89264721 | 0.18235 | 4.78418E-05 | 0.005527867 | 0.02212625 | 1.7391E-05 | 0.9006032 | 0.6456 | 6.9534E-05 | 0.001112375 | 0.00445083 | 19.85407538 | 1.297849666 | FALSE | TRUE | FALSE |
| NEK4 | 0.0003303 | 0.89265429 | 1 | 0.001320162 | 0.001320162 | 0.011143947 | 0.00630575 | 0.897792096 | 0.50685 | 0.025208046 | 0.21272915 | 0.21272915 | 0.052374816 | -1.28087749 | FALSE | TRUE | FALSE |
| NRXN2 | 0.0003293 | 0.89266138 | 0.29005 | 8.24505E-05 | 0.001315673 | 0.006333724 | 0.18338736 | 0.856149202 | 0.6408 | 1.710858377 | 46.89395399 | 230.2476029 | 0.001795755 | -2.745752875 | FALSE | TRUE | FALSE |
| PXN | 0.0003271 | 0.89266847 | 0.50815 | 0.003088783 | 0.02991788 | 0.02991788 | 0.03324381 | 0.890520281 | 1 | 0.307547892 | 0.307547892 | 0.657462041 | 0.009838804 | -2.007057692 | FALSE | TRUE | FALSE |
| MTMR7 | 0.0003231 | 0.89267555 | 1 | 0.001290661 | 0.00515456 | 0.025467364 | 0.00083934 | 0.899568021 | 1 | 0.003351999 | 0.003351999 | 0.028400857 | 0.384990963 | -0.414549464 | FALSE | TRUE | FALSE |
| PCSK5 | 0.0003211 | 0.89268264 | 0.528 | 0.002695534 | 0.006167149 | 0.05176338 | 0.00665977 | 0.89764837 | 0.29105 | 0.006659771 | 0.058113745 | 1.121346758 | 0.048222239 | -1.316752632 | FALSE | TRUE | FALSE |
| NEFH | 0.0003164 | 0.89268972 | 0.12255 | 4.36755E-05 | 0.000698521 | 0.003448041 | 3.8771E-05 | 0.900453821 | 0.23935 | 3.87706E-05 | 0.000620546 | 0.002483545 | 8.159833433 | 0.911681294 | FALSE | TRUE | FALSE |
| BRCA1 | 0.0003083 | 0.8926968 | 0.4024 | 0.000308285 | 0.010251377 | 0.041326985 | 0.00237245 | 0.898915476 | 0.5266 | 0.009475548 | 0.182960747 | 0.730743907 | 0.129943527 | -0.88624535 | FALSE | TRUE | FALSE |
| EDAR | 0.0003079 | 0.89270389 | 0.015 | 0.000307852 | 0.000307852 | 0.00066292 | 4.5613E-05 | 0.900375707 | 1 | 0.001546425 | 0.032121599 | 0.032121599 | 6.749232889 | 0.829254414 | FALSE | TRUE | FALSE |
| ACACB | 0.0003006 | 0.89271097 | 0.04625 | 3.17188E-07 | 0.00015638 | 0.002365932 | 3.3459E-08 | 0.901693568 | 0.97515 | 0.000117292 | 0.014721456 | 0.141558131 | 8984.450171 | 3.953491504 | FALSE | TRUE | FALSE |
| MYO1F | 0.0002987 | 0.89271805 | 0.04125 | 0.000298665 | 0.000298665 | 0.010057776 | 0.00668764 | 0.897627806 | 0.76255 | 0.006687636 | 0.062746349 | 1.219375816 | 0.044659223 | -1.350088839 | FALSE | TRUE | FALSE |
| EXTL3 | 0.0002944 | 0.89272512 | 0.75955 | 0.001175518 | 0.004692608 | 0.039690303 | 0.00690021 | 0.897579795 | 0.6379 | 0.011897216 | 0.110036399 | 0.402033863 | 0.042665704 | -1.369921084 | FALSE | TRUE | FALSE |
| TP63 | 0.0002868 | 0.8927322 | 0.02345 | 0.000286814 | 0.000286814 | 0.00564354 | 0.00075092 | 0.899614327 | 1 | 0.000750919 | 0.000750919 | 0.01480431 | 0.381951193 | -0.417992129 | FALSE | TRUE | FALSE |
| PAPLN | 0.0002776 | 0.89273928 | 0.87755 | 0.018804166 | 0.160846022 | 0.364456053 | 0.00559368 | 0.897969527 | 0.69795 | 0.04740565 | 3.672367499 | 8.016976396 | 0.049623197 | -1.30431526 | FALSE | TRUE | FALSE |
| DNMBP | 0.0002727 | 0.89274636 | 0.1456 | 3.24232E-05 | 0.001090638 | 0.002511678 | 0.00032862 | 0.899891304 | 0.51555 | 0.040004048 | 13.33910817 | 118.6062265 | 0.829915048 | -0.080966361 | FALSE | TRUE | FALSE |
| DYNC2H1 | 0.0002671 | 0.89275343 | 0.46705 | 0.000267071 | 0.016779345 | 0.078541092 | 0.00027528 | 0.899911032 | 0.9384 | 0.00880133 | 0.607353845 | 5.233651837 | 0.970182229 | -0.013146685 | FALSE | TRUE | FALSE |
| SECISBP2 | 0.0002661 | 0.89276051 | 0.75365 | 0.002146186 | 0.008578646 | 0.042290242 | 0.00274302 | 0.898767899 | 0.0171 | 0.00274302 | 0.00274302 | 0.023193127 | 0.09699626 | -1.01324501 | FALSE | TRUE | FALSE |
| NBEAL1 | 0.0002639 | 0.89276758 | 0.9842 | 0.017149034 | 1.230888653 | 10.80843253 | 0.0066497 | 0.897662074 | 0.93905 | 0.473758178 | 37.18850864 | 138.8460423 | 0.039686349 | -1.401358849 | FALSE | TRUE | FALSE |
| NUTM1 | 0.0002575 | 0.89277465 | 1 | 0.000257509 | 0.000257509 | 0.005043303 | 0.00037172 | 0.899851825 | 0.02505 | 0.000371724 | 0.000371724 | 0.007298396 | 0.692742297 | -0.159428295 | FALSE | TRUE | FALSE |
| SMC4 | 0.0002558 | 0.89278172 | 0.03285 | 0.00025577 | 0.00025577 | 0.00860834 | 0.04587175 | 0.88758459 | 0.14665 | 0.045871748 | 0.413823799 | 0.900020686 | 0.005575773 | -2.253694939 | FALSE | TRUE | FALSE |
| MFGE8 | 0.0002535 | 0.89278879 | 0.8366 | 0.016210526 | 1.039125525 | 4.160781132 | 2.4935E-07 | 0.901521992 | 0.9395 | 3.98093E-06 | 6.36192E-05 | 0.000254421 | 1016.453238 | 3.007087404 | FALSE | TRUE | FALSE |
| ATP12A | 0.0002534 | 0.89279586 | 0.3726 | 6.3375E-05 | 0.028730493 | 0.260216432 | 0.00397209 | 0.898356602 | 0.0074 | 5.80096E-06 | 0.000810418 | 0.003244965 | 0.063806419 | -1.195135628 | FALSE | TRUE | FALSE |
| COL20A1 | 0.0002503 | 0.89280293 | 0.9401 | 0.008199756 | 0.275741811 | 0.637774867 | 0.01049577 | 0.896631911 | 0.39555 | 0.010495768 | 0.087481582 | 0.427096339 | 0.023852214 | -1.622471306 | FALSE | TRUE | FALSE |
| EPHB2 | 0.0002489 | 0.89281 | 0.8463 | 0.229594958 | 53.75351718 | 536.47771 | 9.26366122 | 0.20876073 | 0.2352 | 3.886141976 | 90.55313365 | 362.0396663 | 2.68672E-05 | -4.570776968 | FALSE | FALSE | FALSE |
| HELZ2 | 0.0002486 | 0.89281707 | 0.8438 | 0.004365259 | 0.278396069 | 2.423062136 | 6.8594E-05 | 0.900258306 | 0.8824 | 0.002301498 | 0.077166632 | 0.373200006 | 3.624016123 | 0.559190121 | FALSE | TRUE | FALSE |
| SIPA1L2 | 0.0002451 | 0.89282413 | 1 | 3.88267E+43 | 7.90917E+53 | 7.0274E+57 | 0.00224514 | 0.898989113 | 1 | 1.42395E+20 | 9.73177E+25 | 2.34709E+28 | 0.109149266 | -0.961979182 | FALSE | TRUE | FALSE |
| GNE | 0.0002403 | 0.8928312 | 1 | 0.000240339 | 0.000240339 | 0.004737441 | 0.00062023 | 0.899713415 | 1 | 0.005571287 | 0.005571287 | 0.020989881 | 0.387502937 | -0.411725002 | FALSE | TRUE | FALSE |
| HADHB | 0.0002366 | 0.89283826 | 0.32 | 5.91965E-05 | 1.170734005 | 1.170734005 | 1.1742E-05 | 0.900700381 | 0.50905 | 4.69352E-05 | 0.276924031 | 0.276924031 | 20.14896464 | 1.304252735 | FALSE | TRUE | FALSE |
| ALDOB | 0.0002342 | 0.89284533 | 0.0105 | 0.000234212 | 0.000234212 | 0.000508169 | 0.003325 | 0.898545777 | 0.3132 | 0.003325002 | 0.317162035 | 0.317162035 | 0.070439606 | -1.15218308 | FALSE | TRUE | FALSE |
| PLEKHS1 | 0.0002281 | 0.89285239 | 1 | 0.000228088 | 0.000228088 | 0.000228088 | 0.02704856 | 0.891936873 | 0.00465 | 0.003158309 | 0.027048557 | 0.027048557 | 0.008432541 | -2.074041544 | FALSE | TRUE | FALSE |
| RALGAPA1 | 0.0002207 | 0.89285945 | 1 | 0.019175456 | 1.844131878 | 3.588399607 | 0.00605775 | 0.897867231 | 0.5219 | 0.006057749 | 0.006057749 | 0.117514235 | 0.03642828 | -1.438561335 | FALSE | TRUE | FALSE |
| TUBGCP6 | 0.0002158 | 0.89286651 | 1 | 0.013172308 | 0.890364964 | 2.039944395 | 2.3535972 | 0.460051834 | 0.2019 | 0.276759736 | 5.307994422 | 21.50410912 | 9.16851E-05 | -4.037701468 | FALSE | FALSE | FALSE |
| SEC31B | 0.0002016 | 0.89287357 | 0.93715 | 0.230571367 | 39.00638738 | 155.9365582 | 408.370516 | 0.006048372 | 0.1359 | 20.06166428 | 3818.552851 | 36540.36533 | 4.93688E-07 | -6.306547488 | FALSE | FALSE | FALSE |
| COL9A2 | 0.0001981 | 0.89288063 | 0.8818 | 0.003158922 | 0.027743807 | 0.05885759 | 0.00861571 | 0.897145874 | 0.32555 | 0.008615715 | 0.137587248 | 0.170152211 | 0.022989083 | -1.638478354 | FALSE | TRUE | FALSE |
| CLCN2 | 0.0001952 | 0.89288769 | 0.50025 | 0.000195193 | 0.026656462 | 0.11763564 | 0.00026168 | 0.89992418 | 0.3043 | 6.54176E-05 | 0.004190089 | 0.016773205 | 0.745937669 | -0.127297461 | FALSE | TRUE | FALSE |
| MYO7B | 0.0001944 | 0.89289474 | 0.47685 | 0.000194399 | 1.729389297 | 110.3121896 | 1.5491E-05 | 0.900642095 | 0.7307 | 0.000247104 | 2.599595472 | 41.48766566 | 12.54904069 | 1.098610527 | FALSE | TRUE | FALSE |

| | | | | | | | | | | | | | | | | | |
|---|---|---|---|---|---|---|---|---|---|---|---|---|---|---|---|---|---|
| MYO18A | 0.0001835 | 0.8929018 | 0.9374 | 0.015462055 | 0.981699191 | 3.918312545 | 0.20733587 | 0.850357676 | 0.43935 | 0.118482927 | 7.543527225 | 70.52719072 | 0.000885085 | -3.053014882 | FALSE | TRUE | FALSE |
| ACACA | 0.0001808 | 0.89290885 | 0.06705 | 0.000180832 | 0.003449963 | 0.006030564 | 0.00048426 | 0.899779371 | 1 | 0.004420657 | 0.009290343 | 0.084808006 | 0.373417016 | -0.427805896 | FALSE | TRUE | FALSE |
| MADCAM1 | 0.0001756 | 0.89291591 | 1 | 0.001407461 | 0.097771235 | 0.097771235 | 0.00034494 | 0.899864988 | 0.75335 | 0.002818917 | 0.023645603 | 0.023645603 | 0.508948968 | -0.293325762 | FALSE | TRUE | FALSE |
| NR3C2 | 0.0001752 | 0.89292296 | 0.26235 | 4.38563E-05 | 0.000175236 | 0.000861044 | 9.7259E-06 | 0.900745666 | 0.0227 | 9.7259E-06 | 9.7259E-06 | 0.000191373 | 18.01743732 | 1.25569302 | FALSE | TRUE | FALSE |
| WDR11 | 0.0001737 | 0.89293001 | 0.51555 | 0.001544396 | 0.001544396 | 0.030166296 | 0.0004689 | 0.899785962 | 0.87995 | 0.007479638 | 0.066986572 | 0.328487322 | 0.370524923 | -0.431182574 | FALSE | TRUE | FALSE |
| CHD5 | 0.0001711 | 0.89293706 | 1 | 0.010885659 | 0.173720449 | 0.82866461 | 0.00182921 | 0.899116068 | 0.97045 | 0.040691926 | 1.302419033 | 5.202358286 | 0.093519025 | -1.029100029 | FALSE | TRUE | FALSE |
| EFCAB6 | 0.0001681 | 0.89294411 | 0.5236 | 0.001297813 | 0.001297813 | 0.025185035 | 0.00043202 | 0.899812317 | 0.5148 | 0.003409854 | 0.003409854 | 0.066407829 | 0.389118356 | -0.409918281 | FALSE | TRUE | FALSE |
| GAA | 0.0001664 | 0.89295116 | 0.11415 | 2.5974E-06 | 0.000202469 | 0.002666795 | 0.00053387 | 0.899746403 | 0.54655 | 0.000533868 | 0.175171018 | 2.806335231 | 0.311637819 | -0.506349844 | FALSE | TRUE | FALSE |
| MUC5B | 0.0001633 | 0.89295821 | 1 | 4.56082E+43 | 6.46571E+53 | 5.00147E+57 | 0.95153641 | 0.5926355 | 1 | 5.60854E+52 | 7.55741E+63 | 9.48021E+67 | 0.000171667 | -3.765313205 | FALSE | TRUE | FALSE |
| CNTNAP5 | 0.0001466 | 0.89296526 | 0.5141 | 0.000146641 | 0.000146641 | 0.002855524 | 0.00040299 | 0.899832075 | 0.0333 | 0.000402992 | 0.000402992 | 0.007872351 | 0.363880484 | -0.439041237 | FALSE | TRUE | FALSE |
| TGM1 | 0.0001411 | 0.89297231 | 0.2344 | 2.02592E-05 | 0.001290894 | 0.005158584 | 4.1776E-07 | 0.901394513 | 0.75245 | 4.17761E-07 | 1.66857E-06 | 8.20013E-06 | 337.6589481 | 2.528478263 | FALSE | TRUE | FALSE |
| SPIDR | 0.000141 | 0.89297935 | 1 | 0.001146458 | 0.001146458 | 0.022541817 | 0.01779276 | 0.894375591 | 0.51185 | 0.017792762 | 0.154705037 | 0.154705037 | 0.00792245 | -2.101140483 | FALSE | TRUE | FALSE |
| NRAP | 0.0001405 | 0.8929864 | 0.5226 | 0.000561817 | 0.004321409 | 0.020869249 | 0.0037366 | 0.89841748 | 0.27235 | 0.000473805 | 0.003736605 | 0.072440742 | 0.037610384 | -1.424692238 | FALSE | TRUE | FALSE |
| COLQ | 0.0001276 | 0.89299344 | 0.50735 | 0.000127587 | 0.094107759 | 0.094107759 | 0.09707425 | 0.876699849 | 0.38045 | 0.097074251 | 0.87421203 | 3.494381026 | 0.001314321 | -2.881298427 | FALSE | TRUE | FALSE |
| KIF1B | 0.0001263 | 0.89300049 | 1 | 0.000126298 | 0.002433311 | 0.004229303 | 0.00312207 | 0.898626653 | 0.7585 | 0.003122073 | 0.029209488 | 0.565252971 | 0.040453279 | -1.39304627 | FALSE | TRUE | FALSE |
| GEN1 | 0.0001236 | 0.89300753 | 1 | 47480970467 | 3.07339E+14 | 6.49703E+15 | 0.00239829 | 0.898895375 | 1 | 975475.9509 | 3469778344 | 1.29349E+11 | 0.051534389 | -1.287902873 | FALSE | TRUE | FALSE |
| ABCA4 | 0.0001173 | 0.89301457 | 0.7142 | 0.007676703 | 1123.280806 | 144131.8588 | 0.57280004 | 0.723445269 | 0.27565 | 0.004249081 | 1990.553039 | 529057.9248 | 0.000204739 | -3.688799939 | FALSE | TRUE | FALSE |
| MYH7 | 0.0001115 | 0.89302161 | 0.12665 | 5.8465E-06 | 0.219774739 | 3.507809001 | 1.1203E-05 | 0.900719794 | 0.31685 | 2.80673E-06 | 0.000178609 | 0.103425769 | 9.95304461 | 0.997955951 | FALSE | TRUE | FALSE |
| TRANK1 | 0.0001108 | 0.89302865 | 1 | 5.63722E+28 | 1.03312E+37 | 4.34331E+40 | 0.15564952 | 0.863083699 | 1 | 1.16007E+26 | 2.35301E+33 | 3.81402E+36 | 0.000711691 | -3.147708577 | FALSE | TRUE | FALSE |
| IGSF22 | 0.0001071 | 0.89303569 | 0.27055 | 1.38975E-05 | 0.000107126 | 0.000468627 | 0.61402928 | 0.705895836 | 0.1302 | 0.000961022 | 0.614029281 | 1.430024465 | 0.000174464 | -3.758293504 | FALSE | TRUE | FALSE |
| LAMC3 | 0.0001051 | 0.89304273 | 0.99205 | 0.25202898 | 20.32684646 | 189.1955583 | 0.00542728 | 0.898010395 | 0.69695 | 0.005427285 | 0.086783286 | 0.419760759 | 0.019358464 | -1.713129099 | FALSE | TRUE | FALSE |
| TP73 | 0.000105 | 0.89304976 | 0.51185 | 0.001366167 | 0.001366167 | 0.026852477 | 0.03120629 | 0.890944908 | 0.3843 | 0.013436077 | 0.124612084 | 0.264638621 | 0.003363904 | -2.473156357 | FALSE | TRUE | FALSE |
| FLNB | 0.0001027 | 0.8930568 | 0.8653 | 0.01683384 | 22.41700811 | 356.5736236 | 0.00033101 | 0.899884726 | 0.042 | 2.46829E-06 | 0.00033101 | 0.005264308 | 0.310198665 | -0.508360076 | FALSE | TRUE | FALSE |
| GON4L | 0.0001015 | 0.89306383 | 1 | 0.000890327 | 0.001950612 | 0.017118401 | 0.00016079 | 0.900088242 | 0.52795 | 0.001423689 | 0.001423689 | 0.027501092 | 0.630943753 | -0.200009355 | FALSE | TRUE | FALSE |
| KCNN2 | 0.0001008 | 0.89307087 | 1 | 0.000945636 | 0.000945636 | 0.003412656 | 0.01477931 | 0.895168736 | 0.0162 | 0.014779314 | 0.014779314 | 0.291892012 | 0.006819487 | -2.166248316 | FALSE | TRUE | FALSE |
| FOXK1 | 9.902E-05 | 0.8930779 | 0.02075 | 9.90243E-05 | 9.90243E-05 | 0.001948553 | 0.01742399 | 0.894440897 | 1 | 0.017423985 | 0.017423985 | 0.343526451 | 0.005683217 | -2.245405745 | FALSE | TRUE | FALSE |
| HCN4 | 9.729E-05 | 0.89308493 | 0.51735 | 9.72905E-05 | 0.771366851 | 0.771366851 | 0.00021213 | 0.899989868 | 0.4493 | 0.000212127 | 3.945963863 | 3.945963863 | 0.458643538 | -0.338524721 | FALSE | TRUE | FALSE |
| KCNH2 | 9.483E-05 | 0.89309196 | 0.83315 | 0.000260394 | 0.009566543 | 0.038170274 | 0.04020588 | 0.888919034 | 0.00115 | 0.000298306 | 0.001190141 | 0.00582797 | 0.002358539 | -2.627356946 | FALSE | TRUE | FALSE |
| EIF2B5 | 9.366E-05 | 0.89309899 | 1 | 9.36574E-05 | 9.36574E-05 | 0.001844857 | 0.00350995 | 0.898512043 | 0.7535 | 0.032682155 | 0.312326748 | 0.644941262 | 0.026683393 | -1.573758943 | FALSE | TRUE | FALSE |
| LAMA2 | 9.181E-05 | 0.89310602 | 0.87185 | 0.001466106 | 0.194433921 | 2.841089507 | 0.03579105 | 0.889936453 | 0.5448 | 0.071818317 | 18.66396174 | 156.0840657 | 0.002565173 | -2.590883345 | FALSE | TRUE | FALSE |
| NMNAT2 | 8.983E-05 | 0.89311305 | 0.00995 | 8.98304E-05 | 8.98304E-05 | 8.98304E-05 | 0.07420026 | 0.881571552 | 1 | 0.07420026 | 0.07420026 | 0.07420026 | 0.001210649 | -2.916981896 | FALSE | TRUE | FALSE |
| AKAP3 | 8.717E-05 | 0.89312008 | 0.5081 | 0.002946826 | 0.857905242 | 16.882025 | 1.03160347 | 0.585093951 | 0.94065 | 9.565062392 | 886.7647743 | 886.7647743 | 8.45021E-05 | -4.073132316 | FALSE | FALSE | FALSE |
| TMTC4 | 8.379E-05 | 0.89312711 | 0.878 | 0.08508142 | 0.75588158 | 1.673662812 | 0.04079802 | 0.888767742 | 0.51495 | 0.040798022 | 0.040798022 | 0.804138254 | 0.002053718 | -2.687459173 | FALSE | TRUE | FALSE |
| CUBN | 8.334E-05 | 0.89313413 | 0.87295 | 0.008575618 | 5.056418948 | 44.39642671 | 0.12375251 | 0.870419134 | 0.15875 | 0.003555101 | 0.494403915 | 4.41094781 | 0.000673417 | -3.171716014 | FALSE | TRUE | FALSE |
| CELSR1 | 8.249E-05 | 0.89314116 | 1 | 0.000753075 | 0.013866354 | 0.129926028 | 0.0002273 | 0.899976737 | 1 | 0.187905956 | 6.277629507 | 34.47564199 | 0.362906195 | -0.440205618 | FALSE | TRUE | FALSE |
| MSH2 | 8.105E-05 | 0.89314818 | 0.09965 | 5.06933E-06 | 0.000324225 | 0.002260536 | 3.3691E-08 | 0.901687224 | 0.50675 | 1.34596E-07 | 5.37858E-07 | 2.14986E-06 | 2405.829271 | 3.381264805 | FALSE | TRUE | FALSE |
| MYO9B | 7.885E-05 | 0.8931552 | 1 | 1.37267E+12 | 4.91132E+17 | 9.23998E+19 | 0.0002119 | 0.899996433 | 1 | 1.11659E+15 | 4.96862E+20 | 1.01556E+23 | 0.37210353 | -0.42933621 | FALSE | TRUE | FALSE |
| CPS1 | 7.847E-05 | 0.89316223 | 0.8361 | 0.000313021 | 0.01990194 | 0.079472948 | 0.00014578 | 0.900114443 | 0.5971 | 0.000258058 | 0.004107398 | 0.016392826 | 0.538258541 | -0.26900907 | FALSE | TRUE | FALSE |
| COL24A1 | 7.689E-05 | 0.89316925 | 0.4102 | 4.37052E-05 | 0.180669276 | 2.905350236 | 1.0353E-10 | 0.902079009 | 0.8965 | 2.62291E-08 | 1.41172E-05 | 0.0002262 | 742671.9442 | 5.870797018 | FALSE | TRUE | FALSE |
| CPOX | 7.511E-05 | 0.89317627 | 1 | 7.51141E-05 | 7.51141E-05 | 0.0014896 | 0.0659715 | 0.883359087 | 1 | 0.065971502 | 0.065971502 | 1.309619994 | 0.001138584 | -2.943635018 | FALSE | TRUE | FALSE |
| MYO1C | 7.385E-05 | 0.89318329 | 0.27695 | 7.38536E-05 | 0.000672987 | 0.002490925 | 0.00020228 | 0.90001612 | 0.7575 | 0.000202279 | 0.001857317 | 0.006828997 | 0.365108153 | -0.437578469 | FALSE | TRUE | FALSE |
| LILRA4 | 7.051E-05 | 0.8931903 | 1 | 8.17685E+57 | 2.56208E+68 | 6.06906E+73 | 5.1273E-05 | 0.900343123 | 1 | 9.78736E+46 | 1.89492E+57 | 1.64063E+61 | 1.375198305 | 0.138365328 | FALSE | TRUE | FALSE |
| CATSPER1 | 6.949E-05 | 0.89319732 | 1 | 0.000613946 | 0.000613946 | 0.012035432 | 0.00021976 | 0.899983303 | 1 | 0.000219756 | 0.000219756 | 0.004318246 | 0.316236628 | -0.49998783 | FALSE | TRUE | FALSE |
| ABCC12 | 6.715E-05 | 0.89320434 | 0.646 | 0.000613604 | 1.332857952 | 54.01077254 | 0.03892202 | 0.889196764 | 0.12125 | 0.000202126 | 0.229323104 | 4.473707357 | 0.001725131 | -2.763177884 | FALSE | TRUE | FALSE |
| GNPTAB | 6.621E-05 | 0.89321135 | 0.83385 | 0.005350966 | 1.897818014 | 18.50943832 | 0.01982544 | 0.893887147 | 0.1437 | 0.000937337 | 0.388717797 | 2.651954412 | 0.00333957 | -2.476309423 | FALSE | TRUE | FALSE |
| DNMT3A | 6.481E-05 | 0.89321837 | 1 | 0.001034564 | 0.008440437 | 0.04127078 | 0.00373302 | 0.89842424 | 0.5131 | 0.006309462 | 0.053675426 | 0.12362323 | 0.017360187 | -1.760445601 | FALSE | TRUE | FALSE |
| CPT2 | 6.373E-05 | 0.89322538 | 0.94395 | 0.005118107 | 0.576621108 | 5.235226371 | 45424.846 | 4.09975E-05 | 0.0739 | 402.5776048 | 60551.19918 | 971999.7279 | 1.40293E-09 | -8.852963915 | FALSE | FALSE | FALSE |
| TPP1 | 6.324E-05 | 0.8932324 | 0.5024 | 6.32418E-05 | 0.00238591 | 0.009540486 | 0.00015681 | 0.900101344 | 0.07175 | 1.73478E-05 | 0.000627265 | 0.002509815 | 0.403305539 | -0.394365814 | FALSE | TRUE | FALSE |
| KATNAL1 | 6.168E-05 | 0.89323941 | 0.5052 | 0.002088913 | 0.143071445 | 0.143071445 | 0.4869774 | 0.760237518 | 0.01305 | 0.486977404 | 0.486977404 | 1.032125269 | 0.000126651 | -3.89739277 | FALSE | TRUE | FALSE |
| LHX9 | 6.108E-05 | 0.89324642 | 1 | 0.000572557 | 0.005508269 | 0.005508269 | 0.05892435 | 0.884903346 | 1 | 0.235525514 | 0.941650391 | 1.166984661 | 0.001036606 | -2.984386118 | FALSE | TRUE | FALSE |
| HCN2 | 5.836E-05 | 0.89325343 | 0.02425 | 5.83584E-05 | 5.83584E-05 | 0.001144528 | 0.20699591 | 0.85042694 | 0.6405 | 0.827586447 | 7.7979084 | 16.26858451 | 0.00028193 | -3.54985841 | FALSE | TRUE | FALSE |
| TAS2R30 | 5.574E-05 | 0.89326044 | 1 | 3.00801E+81 | 2.73107E+95 | 1.3277E+101 | 0.00011731 | 0.900153719 | 1 | 1.31633E+78 | 1.07336E+92 | 1.89839E+96 | 0.475200019 | -0.323123551 | FALSE | TRUE | FALSE |
| NOTCH1 | 5.475E-05 | 0.89326745 | 0.0841 | 5.47531E-05 | 0.000197541 | 0.001829562 | 3.134E-05 | 0.900486333 | 1 | 3.13396E-05 | 0.000589826 | 0.001049374 | 1.747092097 | 0.242315799 | FALSE | TRUE | FALSE |
| GJA3 | 5.473E-05 | 0.89327445 | 1 | 5.47255E-05 | 5.47255E-05 | 0.001086045 | 0.04774628 | 0.887201446 | 1 | 0.047746277 | 0.047746277 | 0.047746277 | 0.001146174 | -2.940749537 | FALSE | TRUE | FALSE |
| EXPH5 | 5.282E-05 | 0.89328146 | 1 | 17565.07361 | 18306500.91 | 193419652.1 | 0.07900736 | 0.880636569 | 0.88835 | 58.95817868 | 5449.560054 | 54170.87774 | 0.00066851 | -3.174891944 | FALSE | TRUE | FALSE |
| ATP2C1 | 5.244E-05 | 0.89328847 | 0.02305 | 5.24376E-05 | 5.24376E-05 | 0.001031744 | 0.040938 | 0.888711907 | 1 | 0.163632155 | 0.163632155 | 1.383621235 | 0.001280902 | -2.892484183 | FALSE | TRUE | FALSE |
| MCTP2 | 5.15E-05 | 0.89329547 | 0.9095 | 0.000822669 | 0.012220266 | 0.048846191 | 0.000102 | 0.900206039 | 0.96895 | 0.001630366 | 0.024255645 | 0.096988246 | 0.50495479 | -0.296747504 | FALSE | TRUE | FALSE |
| LRP6 | 5.116E-05 | 0.89330247 | 0.38225 | 5.11635E-05 | 0.08427921 | 0.706186844 | 2.5874E-06 | 0.901010086 | 0.99875 | 0.035980592 | 2.293876631 | 9.168270707 | 19.77395743 | 1.296093595 | FALSE | TRUE | FALSE |
| UNC13C | 5.108E-05 | 0.89330948 | 1 | 2.90193E+37 | 9.36936E+46 | 8.78541E+50 | 1.7538E-05 | 0.900596715 | 1 | 1.73334E+28 | 6.48006E+36 | 3.70672E+40 | 2.912254648 | 0.464229347 | FALSE | TRUE | FALSE |
| COL6A2 | 5.076E-05 | 0.89331648 | 0.7721 | 0.000202532 | 0.003226353 | 0.012882 | 10.0418861 | 0.19725829 | 0.17435 | 0.629900154 | 160.2479888 | 1500.91304 | 5.05514E-06 | -5.296266767 | FALSE | FALSE | FALSE |
| CHST6 | 4.958E-05 | 0.89332348 | 0.2069 | 5.47784E-06 | 0.000198162 | 0.000792293 | 8.3734E-08 | 0.901636442 | 0.5074 | 8.37336E-08 | 3.34574E-07 | 4.13544E-07 | 592.0597722 | 2.772365554 | FALSE | TRUE | FALSE |
| AGAP3 | 4.88E-05 | 0.89333048 | 1 | 0.005817097 | 0.178058847 | 0.712535587 | 0.00104271 | 0.899389028 | 0.98505 | 0.139781722 | 8.388517758 | 33.58591081 | 0.046798843 | -1.329764887 | FALSE | TRUE | FALSE |
| AMY2B | 4.64E-05 | 0.89333748 | 0.5008 | 4.64045E-05 | 4.64045E-05 | 0.000105502 | 1.103E-05 | 0.900726263 | 1 | 1.10302E-05 | 1.10302E-05 | 0.000218772 | 4.207061756 | 0.623978887 | FALSE | TRUE | FALSE |
| SLC45A2 | 4.497E-05 | 0.89334448 | 0.89065 | 0.155278821 | 5.640790718 | 22.54931186 | 3.30594991 | 0.394418801 | 0.00125 | 0.000254647 | 0.166691472 | 0.166691472 | 1.36014E-05 | -4.866417719 | FALSE | FALSE | FALSE |
| L1TD1 | 4.495E-05 | 0.89335147 | 0.26185 | 4.495E-05 | 0.098589319 | 0.098589319 | 0.00010291 | 0.900199502 | 0.01125 | 0.000102915 | 0.000102915 | 0.000244759 | 0.436768641 | -0.359748551 | FALSE | TRUE | FALSE |
| NRP1 | 4.494E-05 | 0.89335847 | 0.70665 | 0.000179412 | 0.011431977 | 0.040440651 | 1.4207E-05 | 0.900661531 | 0.50665 | 3.31743E-05 | 0.000529351 | 0.000652643 | 3.163339759 | 0.50014584 | FALSE | TRUE | FALSE |
| ARSB | 4.374E-05 | 0.89336547 | 0.50795 | 4.37396E-05 | 0.000398762 | 0.000398762 | 3.708E-06 | 0.900919955 | 0.8761 | 5.92298E-05 | 0.35512919 | 0.35512919 | 11.79607044 | 1.071737357 | FALSE | TRUE | FALSE |
| MYO5A | 4.346E-05 | 0.89337246 | 0.05465 | 2.2549E-06 | 4.34591E-05 | 0.000424912 | 7.6004E-07 | 0.901253903 | 0.5189 | 7.60043E-07 | 1.21278E-05 | 5.87638E-05 | 57.17982201 | 1.757242799 | FALSE | TRUE | FALSE |
| SLC12A3 | 4.322E-05 | 0.89337945 | 0.7564 | 4.32205E-05 | 0.000360634 | 0.001459346 | 2.1898E-05 | 0.900538308 | 0.5134 | 0.000181199 | 0.000181199 | 0.00354785 | 1.973721423 | 0.295285855 | FALSE | TRUE | FALSE |
| FUT1 | 4.247E-05 | 0.89338645 | 0.01295 | 4.24655E-05 | 4.24655E-05 | 0.000840097 | 0.06593655 | 0.88336777 | 0.7499 | 0.065936552 | 0.621481076 | 0.621481076 | 0.000644036 | -3.191089939 | FALSE | TRUE | FALSE |
| KCNH3 | 4.229E-05 | 0.89339344 | 1 | 0.000392263 | 0.000392263 | 0.007606183 | 0.00260828 | 0.898835028 | 0.5014 | 0.002608281 | 0.002608281 | 0.050761794 | 0.01621206 | -1.790161797 | FALSE | TRUE | FALSE |
| ALPP | 4.118E-05 | 0.89340043 | 0.82855 | 0.00036735 | 0.023459843 | 0.093818902 | 6.6017E-07 | 0.901285895 | 0.84435 | 1.05342E-05 | 9.46427E-05 | 0.0003782 | 62.37544908 | 1.795013685 | FALSE | TRUE | FALSE |
| MYH15 | 4.074E-05 | 0.89340742 | 0.51645 | 0.000398077 | 4.397651637 | 147.2594683 | 1.2815E-09 | 0.901997119 | 0.9736 | 9.10747E-06 | 0.090277132 | 1.753300808 | 31793.02451 | 4.502331845 | FALSE | TRUE | FALSE |

| | | | | | | | | | | | | | | | | | |
|---|---|---|---|---|---|---|---|---|---|---|---|---|---|---|---|---|---|
| TFAP2D | 4.063E-05 | 0.89341441 | 1 | 4.0628E-05 | 4.0628E-05 | 0.000804243 | 5.9171E-05 | 0.90029747 | 1 | 5.91714E-05 | 5.91714E-05 | 0.001172756 | 0.686616015 | -0.163286071 | FALSE | TRUE | FALSE |
| PCNT | 4.015E-05 | 0.8934214 | 0.83885 | 0.122331279 | 68.59916979 | 591.7398872 | 3.8466E-07 | 0.901426413 | 0.7731 | 2.95255E-06 | 5.51514E-05 | 0.000434467 | 104.3853805 | 2.018639679 | FALSE | TRUE | FALSE |
| SYTL3 | 3.95E-05 | 0.89342838 | 1 | 0.006800624 | 0.006800624 | 0.006800624 | 0.99047332 | 0.589337412 | 0.3193 | 0.115743553 | 8.688849183 | 8.688849183 | 3.98764E-05 | -4.399283678 | FALSE | TRUE | FALSE |
| NALCN | 3.862E-05 | 0.89343537 | 0.75875 | 0.000269699 | 0.009041887 | 0.068380702 | 0.00605847 | 0.897860405 | 0.00355 | 4.52196E-05 | 0.000719932 | 0.003489936 | 0.006374513 | -2.195552995 | FALSE | TRUE | FALSE |
| NLRP12 | 3.549E-05 | 0.89344236 | 0.36285 | 8.87694E-06 | 0.000141918 | 0.000692245 | 0.30623054 | 0.823432954 | 0.12855 | 0.000120722 | 1.2244706 | 4.897299895 | 0.000115891 | -3.935952008 | FALSE | TRUE | FALSE |
| KRT13 | 3.427E-05 | 0.89344934 | 0.7484 | 0.000137086 | 0.020714823 | 0.082913052 | 0.00045478 | 0.899799141 | 0.822 | 0.00986692 | 3.794827139 | 38.58312139 | 0.075366426 | -1.122822077 | FALSE | TRUE | FALSE |
| EYS | 3.396E-05 | 0.89345632 | 0.36545 | 8.49692E-06 | 0.001151004 | 0.009992508 | 0.05415319 | 0.885862588 | 0.23435 | 0.003388471 | 1.135581638 | 16.45090774 | 0.0006271 | -3.202663061 | FALSE | TRUE | FALSE |
| FARP2 | 3.357E-05 | 0.89346331 | 0.78815 | 0.000271723 | 0.008596727 | 0.0419447 | 9.0273E-07 | 0.901221891 | 0.76265 | 7.21126E-06 | 0.000115266 | 0.000243391 | 37.18279674 | 1.570342053 | FALSE | TRUE | FALSE |
| SZT2 | 3.202E-05 | 0.89347029 | 1 | 3.20198E-05 | 0.000594282 | 0.001058067 | 0.00059134 | 0.899720015 | 0.0881 | 0.000591343 | 0.002352776 | 0.019606672 | 0.054147536 | -1.266421299 | FALSE | TRUE | FALSE |
| OLFML2A | 3.075E-05 | 0.89347727 | 0.44355 | 3.07493E-05 | 0.007538314 | 0.030103075 | 0.00018227 | 0.900055472 | 0.99685 | 78.31682969 | 161716.2708 | 5519681.52 | 0.168703175 | -0.772876744 | FALSE | TRUE | FALSE |
| CYLD | 3.008E-05 | 0.89348425 | 1 | 66486.7583 | 721795757.5 | 1.12638E+11 | 0.08989604 | 0.878163501 | 1 | 1.60775E+12 | 4.2016E+16 | 7.59573E+18 | 0.000334557 | -3.475529414 | FALSE | TRUE | FALSE |
| PCDHA4 | 2.986E-05 | 0.89349123 | 0.8537 | 0.022859751 | 5.551593514 | 52.0721724 | 0.03267059 | 0.89065957 | 0.9923 | 25.57021589 | 2453.06479 | 8368.112016 | 0.000913901 | -3.039100719 | FALSE | TRUE | FALSE |
| ATP6V1B1 | 2.964E-05 | 0.89349821 | 0.8483 | 0.000474016 | 0.030375166 | 0.121612226 | 0.0003762 | 0.899845243 | 0.28605 | 0.000376196 | 0.006027238 | 0.029743235 | 0.078783994 | -1.103562004 | FALSE | TRUE | FALSE |
| NOTCH3 | 2.908E-05 | 0.89350518 | 0.00715 | 9.61376E-08 | 1.52914E-06 | 1.28342E-05 | 2.4635E-09 | 0.901946658 | 1 | 9.82311E-09 | 4.71345E-08 | 1.87945E-07 | 11806.11102 | 4.072106863 | FALSE | TRUE | FALSE |
| KLHL1 | 2.823E-05 | 0.89351216 | 0.01995 | 2.82339E-05 | 2.82339E-05 | 0.000557083 | 0.1006291 | 0.875770955 | 0.51085 | 0.231589765 | 0.925691435 | 3.40368008 | 0.000280574 | -3.55195301 | FALSE | TRUE | FALSE |
| SIPA1 | 2.792E-05 | 0.89351914 | 1 | 0.068093811 | 0.27222117 | 1.334829218 | 0.02816463 | 0.891709405 | 1 | 0.02816463 | 0.02816463 | 0.553426073 | 0.000991256 | -3.003813969 | FALSE | TRUE | FALSE |
| PNLIP | 2.73E-05 | 0.89352611 | 1 | 2.72964E-05 | 2.72964E-05 | 0.000541084 | 4.0759E-05 | 0.900427797 | 0.5047 | 0.000162916 | 0.000162916 | 0.000808812 | 0.669699324 | -0.17412014 | FALSE | TRUE | FALSE |
| KCNA10 | 2.69E-05 | 0.89353308 | 0.9137 | 0.000429425 | 0.006862668 | 0.027444686 | 1.17890394 | 0.569491557 | 0.78615 | 16.37964891 | 262.1841762 | 302.8769127 | 2.28159E-05 | -4.641762165 | FALSE | FALSE | FALSE |
| AKAP9 | 2.664E-05 | 0.89354006 | 1 | 2.66353E-05 | 0.000502262 | 0.000875181 | 0.03435775 | 0.890279645 | 0.15805 | 0.003923486 | 0.034357753 | 0.652364203 | 0.000775233 | -3.110567521 | FALSE | TRUE | FALSE |
| KCNC4 | 2.658E-05 | 0.89354703 | 0.32875 | 6.64812E-06 | 0.000106273 | 0.000130147 | 1.4899E-06 | 0.901119311 | 0.5085 | 5.95375E-06 | 5.95375E-06 | 0.000116851 | 17.83870341 | 1.251363285 | FALSE | TRUE | FALSE |
| KCNQ4 | 2.61E-05 | 0.893554 | 0.6611 | 0.000104329 | 4.285999089 | 4.285999089 | 9.1705E-06 | 0.900758598 | 0.5159 | 3.66471E-05 | 3.66471E-05 | 0.000720114 | 2.845815432 | 0.45420673 | FALSE | TRUE | FALSE |
| TAS2R19 | 2.561E-05 | 0.89356097 | 1 | 3.24838E+94 | 1.926E+110 | 1.3243E+116 | 0.40204213 | 0.791574441 | 1 | 1.57391E+86 | 8.04948E+98 | 1.2381E+106 | 6.37115E-05 | -4.195782115 | FALSE | TRUE | FALSE |
| TSHZ1 | 2.541E-05 | 0.89356794 | 1 | 1.07198E+19 | 4.48035E+25 | 8.14542E+27 | 0.00099794 | 0.899448761 | 1 | 9.27062E+20 | 4.96253E+27 | 9.77981E+29 | 0.025463267 | -1.594085875 | FALSE | TRUE | FALSE |
| ATAD3B | 2.475E-05 | 0.89357491 | 1 | 0.001764741 | 0.015486521 | 0.034654305 | 9.1165E-05 | 0.900225645 | 1 | 9.11652E-05 | 9.11652E-05 | 0.001794127 | 0.271471876 | -0.566275156 | FALSE | TRUE | FALSE |
| ABCB8 | 2.443E-05 | 0.89358187 | 1 | 0.006248486 | 0.099758791 | 0.122476367 | 0.06267649 | 0.884118768 | 0.3291 | 0.015684854 | 0.250517133 | 0.308177014 | 0.0003897 | -3.409269902 | FALSE | TRUE | FALSE |
| ZYX | 2.384E-05 | 0.89358884 | 1 | 0.000216903 | 0.000216903 | 0.000806717 | 0.08373044 | 0.879430074 | 0.50215 | 0.083730444 | 0.083730444 | 0.709315968 | 0.000284697 | -3.54561725 | FALSE | TRUE | FALSE |
| BCKDHA | 2.282E-05 | 0.89359581 | 0.70675 | 9.11605E-05 | 0.005821737 | 0.023281879 | 0.0059454 | 0.897908171 | 0.5979 | 0.010363116 | 1.522891113 | 8.654470999 | 0.003837905 | -2.41590579 | FALSE | TRUE | FALSE |
| PLA2G15 | 2.26E-05 | 0.89360277 | 0.50865 | 2.26049E-05 | 2.26049E-05 | 0.000111934 | 0.00019527 | 0.900029241 | 0.3206 | 0.000195268 | 5.035439073 | 5.035439073 | 0.115763554 | -0.936428149 | FALSE | TRUE | FALSE |
| KRT72 | 2.212E-05 | 0.89360973 | 0.35065 | 5.53219E-06 | 8.84787E-05 | 0.000353975 | 0.00057675 | 0.899726613 | 0.54415 | 0.000741352 | 0.047488861 | 0.331650646 | 0.038355455 | -1.416172863 | FALSE | TRUE | FALSE |
| SLC25A13 | 2.118E-05 | 0.8936167 | 0.7543 | 0.003206317 | 0.02701987 | 0.06331406 | 0.00199398 | 0.899089365 | 0.9853 | 0.148738749 | 5.334942442 | 21.32556462 | 0.010622553 | -1.973771077 | FALSE | TRUE | FALSE |
| TLL2 | 1.99E-05 | 0.89362366 | 0.6654 | 4.12117E-05 | 0.882260487 | 7.695223352 | 0.33471868 | 0.814016594 | 0.2771 | 0.000258276 | 5.350520273 | 26.17194844 | 5.9457E-05 | -4.225796837 | FALSE | TRUE | FALSE |
| THBS2 | 1.986E-05 | 0.89363062 | 0.03845 | 1.98552E-05 | 1.98552E-05 | 0.00038486 | 5.1075E-05 | 0.900349641 | 0.51825 | 0.000474533 | 0.000474533 | 0.009232239 | 0.388747009 | -0.41033294 | FALSE | TRUE | FALSE |
| BEST3 | 1.749E-05 | 0.89363758 | 0.1958 | 4.37834E-06 | 0.00015639 | 0.0006248 | 0.0004913 | 0.899772779 | 0.02 | 5.85657E-06 | 0.000462619 | 0.001962296 | 0.035594622 | -1.448615613 | FALSE | TRUE | FALSE |
| COL13A1 | 1.674E-05 | 0.89364454 | 0.01955 | 1.67422E-05 | 1.67422E-05 | 6.35823E-05 | 3.6058E-06 | 0.900932841 | 0.0175 | 3.60576E-06 | 3.60576E-06 | 7.11418E-05 | 4.643176652 | 0.666815207 | FALSE | TRUE | FALSE |
| ABCA10 | 1.653E-05 | 0.8936515 | 0.3254 | 2.0002E-06 | 0.088100383 | 0.088100383 | 0.00401941 | 0.898329525 | 0.9398 | 0.29428259 | 23.73836972 | 220.7835152 | 0.004111998 | -2.385947141 | FALSE | TRUE | FALSE |
| CLMN | 1.633E-05 | 0.89365845 | 0.75515 | 6.52558E-05 | 0.000596489 | 0.001279423 | 1.441E-05 | 0.900655053 | 0.51295 | 3.23771E-05 | 0.000129386 | 0.0006363 | 1.133328512 | 0.054355815 | FALSE | TRUE | FALSE |
| ITGB3 | 1.624E-05 | 0.89366541 | 1 | 1.62404E-05 | 1.62404E-05 | 0.000319362 | 4.488E-05 | 0.900395247 | 0.0194 | 4.48804E-05 | 4.48804E-05 | 0.000884341 | 0.361858676 | -0.441461011 | FALSE | TRUE | FALSE |
| NRXN1 | 1.617E-05 | 0.89367236 | 1 | 0.013677846 | 7.431229585 | 14.79310681 | 1.2259548 | 0.564504818 | 0.0887 | 0.076707903 | 1.225954805 | 5.94640332 | 1.31911E-05 | -4.879717709 | FALSE | FALSE | FALSE |
| FOXM1 | 1.616E-05 | 0.89367932 | 0.6926 | 0.000150414 | 0.003518171 | 0.014057495 | 9.5116E-06 | 0.900752132 | 0.88165 | 8.90916E-05 | 0.000855915 | 0.000855915 | 1.698677438 | 0.230110919 | FALSE | TRUE | FALSE |
| MTHFR | 1.603E-05 | 0.89368627 | 0.89295 | 6.41026E-05 | 1.930807987 | 7.722234993 | 0.00327455 | 0.898572749 | 0.0991 | 0.000818392 | 0.013105373 | 0.052463352 | 0.004896842 | -2.310083886 | FALSE | TRUE | FALSE |
| COL4A4 | 1.566E-05 | 0.89369323 | 0.19685 | 9.85492E-07 | 6.24445E-05 | 0.000584543 | 3.5348E-07 | 0.901439168 | 0.26605 | 8.86478E-08 | 3.53476E-07 | 2.98586E-06 | 44.30091462 | 1.646412693 | FALSE | TRUE | FALSE |
| NBAS | 1.558E-05 | 0.89370018 | 0.88325 | 0.020054611 | 0.661815579 | 2.674663759 | 0.00166515 | 0.899176103 | 0.94115 | 0.984790754 | 78.40089677 | 724.4101423 | 0.009357016 | -2.028862642 | FALSE | TRUE | FALSE |
| PER3 | 1.531E-05 | 0.89370713 | 0.03315 | 1.53053E-05 | 1.53053E-05 | 0.000298228 | 1.5754E-05 | 0.900635615 | 1 | 0.016287045 | 0.135476341 | 0.549549086 | 0.97152259 | -0.012547097 | FALSE | TRUE | FALSE |
| DNAH8 | 1.517E-05 | 0.89371408 | 0.9795 | 0.049031908 | 43.62995812 | 765.6340865 | 0.07796751 | 0.880871555 | 0.41345 | 0.023917997 | 11.94108526 | 147.5739249 | 0.000194613 | -3.710827624 | FALSE | TRUE | FALSE |
| MYO3A | 1.478E-05 | 0.89372103 | 0.5163 | 0.000116963 | 0.002272887 | 0.00788634 | 8.888E-05 | 0.900232179 | 0.5113 | 8.88797E-05 | 0.000721962 | 0.00173306 | 0.166304688 | -0.779095508 | FALSE | TRUE | FALSE |
| ABCC11 | 1.453E-05 | 0.89372797 | 0.1411 | 7.64477E-07 | 0.000131047 | 0.00120994 | 2.4038E-05 | 0.900531814 | 0.32855 | 1.0695E-05 | 0.002072633 | 0.019914043 | 0.604273062 | -0.218766766 | FALSE | TRUE | FALSE |
| PRUNE2 | 1.394E-05 | 0.89373492 | 0.5378 | 0.000113524 | 0.586272309 | 11.1141311 | 1.56117425 | 0.530222866 | 0.6496 | 1.56117425 | 264.1237757 | 2398.449077 | 8.93004E-06 | -5.049146482 | FALSE | FALSE | FALSE |
| ITGB6 | 1.382E-05 | 0.89374187 | 0.23935 | 1.38201E-05 | 0.113455527 | 0.453440377 | 2.8342E-06 | 0.900977915 | 0.65995 | 1.1323E-05 | 0.097889207 | 0.391239104 | 4.87613042 | 0.688075313 | FALSE | TRUE | FALSE |
| COL19A1 | 1.369E-05 | 0.89374881 | 0.51795 | 0.000117022 | 0.00102623 | 0.002292134 | 0.00011142 | 0.900173345 | 1 | 0.000942296 | 0.008179739 | 0.018502852 | 0.122850828 | -0.910621912 | FALSE | TRUE | FALSE |
| ACTR5 | 1.357E-05 | 0.89375576 | 0.879 | 0.007765893 | 0.031027945 | 0.031027945 | 0.00466255 | 0.898187149 | 0.7489 | 0.018627738 | 0.166657039 | 0.166657039 | 0.002911093 | -2.535943955 | FALSE | TRUE | FALSE |
| KCNH5 | 1.293E-05 | 0.8937627 | 0.14795 | 1.29276E-05 | 0.006673359 | 0.006673359 | 8.941E-06 | 0.900765062 | 0.64085 | 8.15511E-05 | 0.044060039 | 0.044060039 | 1.445876034 | 0.160131059 | FALSE | TRUE | FALSE |
| INSR | 1.28E-05 | 0.89376964 | 0.63805 | 0.000116665 | 0.211410217 | 0.448231935 | 7.6544E-06 | 0.900771526 | 1 | 0.01306491 | 0.01306491 | 0.253567053 | 1.671743422 | 0.223169623 | FALSE | TRUE | FALSE |
| ALOX12B | 1.278E-05 | 0.89377659 | 0.5168 | 0.000114885 | 0.000114885 | 0.002259193 | 0.01336295 | 0.895696185 | 1 | 0.120865246 | 0.120865246 | 2.381585029 | 0.000956489 | -3.019320232 | FALSE | TRUE | FALSE |
| IHH | 1.208E-05 | 0.89378353 | 0.63555 | 4.82567E-05 | 0.000770808 | 0.003805402 | 1.4499E-05 | 0.900648575 | 0.506 | 1.44992E-05 | 5.79398E-05 | 0.00023159 | 0.833073277 | -0.079316796 | FALSE | TRUE | FALSE |
| TXNDC2 | 1.207E-05 | 0.89379047 | 0.99785 | 36.4658582 | 2924.094016 | 27095.36558 | 3.7361E-05 | 0.900460325 | 1 | 82.75109057 | 65278.18893 | 630842.2318 | 0.322986614 | -0.490815476 | FALSE | TRUE | FALSE |
| PARP4 | 1.177E-05 | 0.89379741 | 1 | 0.002145262 | 0.017126757 | 0.331981582 | 0.00548641 | 0.897996776 | 0.50345 | 0.005486411 | 0.005486411 | 0.106742593 | 0.002145551 | -2.668461152 | FALSE | TRUE | FALSE |
| TMPRSS9 | 1.146E-05 | 0.89380434 | 0.52125 | 9.28984E-05 | 9.28984E-05 | 0.001805998 | 0.00033582 | 0.899871568 | 0.2676 | 0.000335817 | 0.002840972 | 0.006550494 | 0.034118944 | -1.467004414 | FALSE | TRUE | FALSE |
| ATP10D | 1.108E-05 | 0.89381128 | 0.92335 | 0.016182908 | 4.855845621 | 43.65107999 | 1.0839E-06 | 0.901189857 | 0.9078 | 3.67892E-05 | 0.064346888 | 0.56104321 | 10.22259635 | 1.009561213 | FALSE | TRUE | FALSE |
| HEPHL1 | 1.105E-05 | 0.89381822 | 0.1183 | 3.65824E-07 | 2.32322E-05 | 0.000114135 | 2.186E-06 | 0.901042236 | 0.2663 | 5.47539E-07 | 8.72954E-06 | 0.007701969 | 5.054332529 | 0.703663811 | FALSE | TRUE | FALSE |
| EPB41L5 | 1.094E-05 | 0.89382515 | 1 | 1.09406E-05 | 1.09406E-05 | 0.000215853 | 0.01871006 | 0.894157386 | 0.87715 | 0.074769845 | 0.298872689 | 1.477571814 | 0.000584747 | -3.233032146 | FALSE | TRUE | FALSE |
| ABCC10 | 1.088E-05 | 0.89383209 | 0.92275 | 0.003337756 | 3.416387933 | 54.74812787 | 4.8486E-06 | 0.900868378 | 0.6722 | 1.93865E-05 | 0.004967276 | 0.019885827 | 2.24430434 | 0.351081749 | FALSE | TRUE | FALSE |
| KMT2C | 1.066E-05 | 0.89383902 | 0.0151 | 1.50779E-08 | 1.43983E-06 | 1.29702E-05 | 1.8686E-09 | 0.901965587 | 0.98215 | 2.70644E-06 | 0.000708935 | 0.007537959 | 5703.040876 | 3.756106484 | FALSE | TRUE | FALSE |
| SCN7A | 1.065E-05 | 0.89384595 | 0.1456 | 1.06458E-05 | 9.34843E-05 | 0.000208168 | 9.4509E-07 | 0.90120908 | 1 | 8.00256E-06 | 6.95511E-05 | 0.000269372 | 11.26430067 | 1.051704234 | FALSE | TRUE | FALSE |
| NELL1 | 1.031E-05 | 0.89385289 | 1 | 1.03059E-05 | 1.03059E-05 | 0.000202854 | 5.7841E-06 | 0.90082966 | 1 | 0.237029333 | 0.947694096 | 3.790030771 | 1.781765718 | 0.250850599 | FALSE | TRUE | FALSE |
| KCNH4 | 1.03E-05 | 0.89385982 | 1 | 9.61949E-05 | 0.000752453 | 0.003678132 | 0.00235081 | 0.898928872 | 0.06765 | 4.78798E-05 | 0.009382048 | 0.009382048 | 0.004382972 | -2.358231313 | FALSE | TRUE | FALSE |
| BDP1 | 1.018E-05 | 0.89386675 | 0.06135 | 1.01825E-05 | 0.000195137 | 0.000339692 | 0.00823033 | 0.897242587 | 0.52485 | 0.06848012 | 0.584463621 | 2.289694967 | 0.001237193 | -2.90756249 | FALSE | TRUE | FALSE |
| MASP1 | 1.007E-05 | 0.89387368 | 0.9542 | 0.055808522 | 17.98740144 | 88.78506967 | 2.45678092 | 0.451330027 | 0.4946 | 2.093426749 | 157.1424955 | 628.7619808 | 4.09773E-06 | -5.387457116 | FALSE | FALSE | FALSE |
| TPH1 | 9.712E-06 | 0.8938806 | 1 | 9.71238E-06 | 9.71238E-06 | 0.000192681 | 0.000143 | 0.900127538 | 0.88235 | 0.001321169 | 0.012519737 | 0.012519737 | 0.067918573 | -1.168011451 | FALSE | TRUE | FALSE |
| MYO16 | 9.471E-06 | 0.89388753 | 1 | 1983.900816 | 127474.4961 | 320253.3901 | 1.7552E-05 | 0.900590228 | 1 | 316.106141 | 20293.54627 | 75558.2532 | 0.539557886 | -0.267961955 | FALSE | TRUE | FALSE |
| MYO6 | 9.44E-06 | 0.89389446 | 0.99 | 0.945560311 | 60.46136529 | 241.8935001 | 0.76275783 | 0.635110813 | 0.5131 | 0.762757835 | 12.19445194 | 102.673538 | 1.23755E-05 | -4.907436614 | FALSE | TRUE | FALSE |
| ARSI | 9.409E-06 | 0.89390138 | 0.99745 | 0.688961863 | 1311.943303 | 13261.91614 | 1.3841E-05 | 0.900668008 | 0.5078 | 3.13371E-05 | 0.000125132 | 0.000468565 | 0.679746101 | -0.167653275 | FALSE | TRUE | FALSE |
| CHD9 | 8.551E-06 | 0.89390831 | 0.96655 | 0.001786088 | 0.33678315 | 2.931722128 | 2.7251E-06 | 0.901003654 | 0.88305 | 0.000100299 | 0.000950201 | 0.00976667 | 3.137765592 | 0.496620496 | FALSE | TRUE | FALSE |
| JAG2 | 8.463E-06 | 0.89391523 | 0.5206 | 3.37864E-05 | 3.37864E-05 | 0.000653899 | 0.01297972 | 0.895795417 | 0.74895 | 0.012979716 | 0.051856454 | 0.438133975 | 0.000652035 | -3.185729258 | FALSE | TRUE | FALSE |

| | | | | | | | | | | | | | | | | | |
|---|---|---|---|---|---|---|---|---|---|---|---|---|---|---|---|---|---|
| HNF1B | 7.411E-06 | 0.89392215 | 0.01575 | 7.41112E-06 | 7.41112E-06 | 0.000146502 | 1.734E-05 | 0.900609685 | 0.01375 | 1.73399E-05 | 1.73399E-05 | 0.000343252 | 0.427401866 | -0.369163586 | FALSE | TRUE | FALSE |
| KIF26A | 7.099E-06 | 0.89392908 | 0.96905 | 0.021929983 | 1.968533558 | 19.36170608 | 1.9758E-05 | 0.900544801 | 1 | 1.97579E-05 | 1.97579E-05 | 0.000665182 | 0.359316177 | -0.444523229 | FALSE | TRUE | FALSE |
| PNLIPRP3 | 6.859E-06 | 0.893936 | 0.51225 | 0.017002157 | 0.017002157 | 0.017002157 | 0.00800662 | 0.897304674 | 1 | 0.032003343 | 0.032003343 | 0.032003343 | 0.000856702 | -3.067170439 | FALSE | TRUE | FALSE |
| KDM6B | 6.767E-06 | 0.89394292 | 1 | 0.23363075 | 17.68972484 | 159.5368207 | 0.00026059 | 0.899930752 | 0.98435 | 0.581694708 | 45.74348808 | 420.410096 | 0.025967945 | -1.585562423 | FALSE | TRUE | FALSE |
| FAM120B | 6.716E-06 | 0.89394984 | 0.87885 | 8.58511E-06 | 5.72813E-05 | 0.000168709 | 3.6104E-05 | 0.900466828 | 0.5141 | 3.61044E-05 | 0.00025067 | 0.000710991 | 0.186023868 | -0.73043133 | FALSE | TRUE | FALSE |
| FLNC | 6.666E-06 | 0.89395675 | 0.6008 | 2.65911E-05 | 0.026966948 | 0.430552791 | 0.00015411 | 0.900107894 | 0.86925 | 0.039250098 | 91.42400999 | 1464.101852 | 0.043251222 | -1.36400162 | FALSE | TRUE | FALSE |
| ANKRD27 | 6.57E-06 | 0.89396367 | 1 | 1.00114E+12 | 7.92643E+14 | 3.73475E+16 | 6.7417E-05 | 0.900264835 | 1 | 6.29165E+13 | 1.70397E+19 | 2.93089E+20 | 0.097446235 | -1.011234937 | FALSE | TRUE | FALSE |
| ACTR8 | 6.526E-06 | 0.89397059 | 0.51055 | 1.20864E-05 | 0.00041754 | 0.00167086 | 4.3952E-06 | 0.900887725 | 0.7937 | 7.03808E-05 | 0.004518376 | 0.018101248 | 1.484725707 | 0.171646228 | FALSE | TRUE | FALSE |
| ADCY10 | 6.44E-06 | 0.8939775 | 0.5238 | 6.43959E-06 | 6.43959E-06 | 0.000125145 | 0.02968279 | 0.89132837 | 0.1537 | 0.029682788 | 0.249866877 | 0.578815471 | 0.000216947 | -3.663646331 | FALSE | TRUE | FALSE |
| RNLS | 6.307E-06 | 0.89398442 | 1 | 6.30749E-06 | 6.30749E-06 | 0.000125313 | 0.00676612 | 0.897600376 | 1 | 0.006766117 | 0.006766117 | 0.006766117 | 0.000932216 | -3.030483224 | FALSE | TRUE | FALSE |
| CNTRL | 5.938E-06 | 0.89399133 | 0.28955 | 5.93764E-06 | 4.55799E-05 | 0.000877775 | 1.6592E-05 | 0.900622651 | 0.28535 | 1.65925E-05 | 0.002630897 | 0.050892521 | 0.35785131 | -0.446297389 | FALSE | TRUE | FALSE |
| CEP89 | 5.815E-06 | 0.89399824 | 0.8968 | 0.000369278 | 15.48431241 | 138.0744803 | 37.9669107 | 0.072077558 | 0.15685 | 4.279971753 | 344.858971 | 3205.639616 | 1.53165E-07 | -6.81483942 | FALSE | FALSE | FALSE |
| INPP5J | 5.7E-06 | 0.89400516 | 0.12035 | 6.26351E-07 | 2.27719E-05 | 0.000212556 | 1.443E-06 | 0.901132145 | 0.5036 | 1.85629E-06 | 0.000118453 | 0.000473586 | 3.95005652 | 0.59660331 | FALSE | TRUE | FALSE |
| ABCD3 | 5.402E-06 | 0.89401207 | 0.5097 | 2.15675E-05 | 2.15675E-05 | 0.000182418 | 0.01180344 | 0.896183596 | 0.4989 | 0.000485305 | 0.011803441 | 0.011803441 | 0.000457705 | -3.339413885 | FALSE | TRUE | FALSE |
| USH2A | 5.394E-06 | 0.89401898 | 0.82175 | 0.000629582 | 2.302004837 | 73.43295041 | 3.2503E-07 | 0.901458294 | 0.92395 | 0.000328493 | 1.711280756 | 49.42413859 | 16.595003 | 1.219977335 | FALSE | TRUE | FALSE |
| GANAB | 5.358E-06 | 0.89402589 | 0.51655 | 2.14214E-05 | 0.000104835 | 0.000419096 | 6.4672E-06 | 0.900797371 | 0.02865 | 6.46719E-06 | 6.46719E-06 | 0.000126858 | 0.828560198 | -0.081675933 | FALSE | TRUE | FALSE |
| SLC7A14 | 5.221E-06 | 0.8940328 | 0.8509 | 0.000333839 | 0.021393865 | 0.085655762 | 0.00068782 | 0.899647377 | 0.4003 | 0.000171928 | 0.011017058 | 0.054247879 | 0.007590735 | -2.119716143 | FALSE | TRUE | FALSE |
| GALNS | 5.145E-06 | 0.8940397 | 0.88595 | 0.000145796 | 0.026042489 | 0.116254936 | 2.8595E-06 | 0.900971478 | 0.9743 | 0.000730767 | 0.10922677 | 0.750895283 | 1.79931869 | 0.255108091 | FALSE | TRUE | FALSE |
| PDE6B | 4.725E-06 | 0.89404661 | 0.90015 | 0.000358002 | 0.121626726 | 1.144821126 | 6.0099E-07 | 0.90130508 | 0.8908 | 1.21242E-05 | 0.000774892 | 0.007224105 | 7.862536389 | 0.895562668 | FALSE | TRUE | FALSE |
| KCNN3 | 4.696E-06 | 0.89405351 | 0.7573 | 0.001194698 | 0.009764929 | 0.023501788 | 0.00077532 | 0.899607714 | 1 | 0.00077532 | 0.00077532 | 0.015282031 | 0.006056539 | -2.217775484 | FALSE | TRUE | FALSE |
| KCNG4 | 4.66E-06 | 0.89406042 | 0.95495 | 0.000297631 | 0.019050156 | 0.172935894 | 2.7466E-06 | 0.90099722 | 0.9824 | 0.0001754 | 0.01122619 | 0.04492798 | 1.696800601 | 0.229630809 | FALSE | TRUE | FALSE |
| SLC26A7 | 4.634E-06 | 0.89406732 | 0.5117 | 1.85167E-05 | 0.000164647 | 0.000164647 | 1.1606E-06 | 0.901177038 | 1 | 4.63784E-06 | 4.63784E-06 | 2.2978E-05 | 3.993033258 | 0.601302927 | FALSE | TRUE | FALSE |
| LAMA3 | 4.568E-06 | 0.89407423 | 0.54195 | 4.56813E-06 | 0.000143225 | 0.000877581 | 5.9505E-05 | 0.900290945 | 0.535 | 5.95051E-05 | 0.003764736 | 0.01554938 | 0.076768804 | -1.114815225 | FALSE | TRUE | FALSE |
| SCN11A | 4.493E-06 | 0.89408113 | 0.23415 | 2.81476E-07 | 7.53089E-05 | 0.001202635 | 3.527E-11 | 0.902135621 | 0.98035 | 8.92943E-09 | 1.17735E-06 | 9.98633E-06 | 127381.3216 | 5.10510575 | FALSE | TRUE | FALSE |
| PIEZO2 | 4.089E-06 | 0.89408803 | 0.5154 | 3.57557E-05 | 3.57557E-05 | 0.000707819 | 0.39263235 | 0.794600882 | 0.26305 | 0.043382838 | 0.392632348 | 1.457259712 | 1.04149E-05 | -4.98234442 | FALSE | TRUE | FALSE |
| COL10A1 | 4.016E-06 | 0.89409493 | 1 | 4.01608E-06 | 4.01608E-06 | 7.93843E-05 | 0.00010746 | 0.900179886 | 0.0147 | 0.000107459 | 0.000107459 | 0.000412234 | 0.037373146 | -1.427440338 | FALSE | TRUE | FALSE |
| EPHA10 | 3.942E-06 | 0.89410183 | 0.51565 | 3.94191E-06 | 3.94191E-06 | 7.6923E-05 | 0.01124009 | 0.896345128 | 0.7835 | 0.093770803 | 1.763994844 | 7.052611655 | 0.000350701 | -3.455063101 | FALSE | TRUE | FALSE |
| KDM5A | 3.937E-06 | 0.89410873 | 1 | 3.93675E-06 | 3.93675E-06 | 0.000131916 | 6.7666E-06 | 0.900790911 | 0.04145 | 6.76661E-06 | 6.76661E-06 | 0.000227135 | 0.581791332 | -0.235232754 | FALSE | TRUE | FALSE |
| CNTNAP1 | 3.642E-06 | 0.89411563 | 0.53005 | 3.64167E-06 | 1.4551E-05 | 0.000280141 | 3.3018E-06 | 0.900945723 | 0.8799 | 2.94373E-05 | 0.000269374 | 0.001303609 | 1.10295047 | 0.04255601 | FALSE | TRUE | FALSE |
| PANK1 | 3.409E-06 | 0.89412252 | 0.87715 | 0.000124085 | 0.001160167 | 0.001160167 | 0.00021174 | 0.900002996 | 0.25765 | 0.000211736 | 0.001989121 | 0.001989121 | 0.01610022 | -1.793168196 | FALSE | TRUE | FALSE |
| CAPN15 | 3.229E-06 | 0.89412942 | 0.8826 | 1.28926E-05 | 5.14948E-05 | 0.000251894 | 0.00044946 | 0.899805729 | 0.5103 | 0.015180433 | 11.35336272 | 45.39512621 | 0.007183438 | -2.143667676 | FALSE | TRUE | FALSE |
| GAK | 3.22E-06 | 0.89413631 | 0.6995 | 1.28688E-05 | 0.000205713 | 0.000994098 | 2.864E-05 | 0.900512327 | 0.16065 | 2.86398E-05 | 0.000114508 | 0.000555678 | 0.112426072 | -0.949132963 | FALSE | TRUE | FALSE |
| LTBP1 | 3.077E-06 | 0.89414321 | 0.5205 | 3.07719E-06 | 5.95585E-05 | 0.005482294 | 0.3384693 | 0.812531598 | 0.2146 | 0.084727511 | 5.405478283 | 26.27838382 | 9.09148E-06 | -5.04136551 | FALSE | TRUE | FALSE |
| MASP2 | 3.025E-06 | 0.8941501 | 0.7608 | 2.59456E-05 | 0.000228405 | 0.000512058 | 0.0555323 | 0.885502446 | 0.89405 | 0.482128017 | 7.7123679 | 30.85767184 | 5.4475E-05 | -4.263802414 | FALSE | TRUE | FALSE |
| DNAH2 | 2.981E-06 | 0.89415699 | 1 | 694.7311977 | 11360080.71 | 673812446.8 | 0.00017808 | 0.900062028 | 0.99995 | 6365275.626 | 80206374706 | 1.00349E+13 | 0.016736614 | -1.776332389 | FALSE | TRUE | FALSE |
| HSD17B4 | 2.9E-06 | 0.89416388 | 1 | 1.15868E-05 | 1.15868E-05 | 9.81243E-05 | 1.6683E-05 | 0.900616169 | 0.261 | 1.6683E-05 | 0.047780437 | 0.047780437 | 0.173819988 | -0.759900284 | FALSE | TRUE | FALSE |
| TCHH | 2.823E-06 | 0.89417077 | 1 | 10125740.83 | 1.55343E+11 | 1.48404E+13 | 1.7824E-06 | 0.901087211 | 1 | 141766.666 | 1071299336 | 1.04024E+11 | 1.583902743 | 0.199728511 | FALSE | TRUE | FALSE |
| COL12A1 | 2.786E-06 | 0.89417766 | 0.5391 | 2.78574E-06 | 4.44646E-05 | 0.000210352 | 0.00062756 | 0.899700215 | 0.76515 | 0.001422411 | 0.011941263 | 0.108066174 | 0.004439009 | -2.352713991 | FALSE | TRUE | FALSE |
| WDR35 | 2.698E-06 | 0.89418455 | 0.50795 | 2.69762E-06 | 5.26875E-05 | 0.000210523 | 1.6132E-07 | 0.901572892 | 0.81615 | 6.44652E-07 | 2.22849E-05 | 2.22849E-05 | 16.72259644 | 1.223303709 | FALSE | TRUE | FALSE |
| AGRN | 2.643E-06 | 0.89419144 | 0.04025 | 7.28759E-08 | 2.64304E-06 | 1.25896E-05 | 1.0304E-06 | 0.901196266 | 0.06405 | 1.15454E-07 | 4.11787E-06 | 1.97211E-05 | 2.564977011 | 0.409083477 | FALSE | TRUE | FALSE |
| RAD54L | 2.549E-06 | 0.89419833 | 0.18645 | 6.37917E-07 | 4.07389E-05 | 0.00016292 | 4.1313E-07 | 0.901400894 | 0.889 | 6.60203E-06 | 0.000417599 | 0.001752573 | 6.170499964 | 0.790320354 | FALSE | TRUE | FALSE |
| TAS1R2 | 2.486E-06 | 0.89420521 | 0.25975 | 6.22036E-07 | 9.93855E-06 | 1.21744E-05 | 2.7478E-07 | 0.901490153 | 0.75305 | 5.94975E-07 | 2.37764E-06 | 1.16748E-05 | 9.047590899 | 0.956532955 | FALSE | TRUE | FALSE |
| SCARA3 | 2.44E-06 | 0.8942121 | 0.5102 | 9.75076E-06 | 3.89804E-05 | 4.81742E-05 | 1.6641E-06 | 0.901106473 | 1 | 1.52738E-05 | 6.10371E-05 | 0.000131513 | 1.466073608 | 0.166155776 | FALSE | TRUE | FALSE |
| RAD50 | 2.432E-06 | 0.89421898 | 1 | 0.000168332 | 0.394412459 | 0.394412459 | 1.0465E-05 | 0.9007392 | 0.03405 | 1.04647E-05 | 1.04647E-05 | 0.00035217 | 0.232442532 | -0.633684402 | FALSE | TRUE | FALSE |
| COL9A3 | 2.215E-06 | 0.89422586 | 1 | 1.93476E-05 | 1.93476E-05 | 0.00038089 | 6.2945E-06 | 0.900810289 | 1 | 6.29446E-06 | 6.29446E-06 | 0.000124151 | 0.351847402 | -0.453645652 | FALSE | TRUE | FALSE |
| BRCA2 | 2.214E-06 | 0.89423275 | 0.93925 | 0.005236926 | 21.19318703 | 671.5772709 | 3.9036E-06 | 0.900913511 | 0.83755 | 0.000129247 | 0.152149805 | 2.904088745 | 0.567102302 | -0.24633859 | FALSE | TRUE | FALSE |
| MYH14 | 2.207E-06 | 0.89423963 | 1 | 0.000185349 | 0.001765156 | 0.00621071 | 1.2644E-05 | 0.900687434 | 1 | 0.000115223 | 0.000242783 | 0.002212427 | 0.174521583 | -0.758150857 | FALSE | TRUE | FALSE |
| SBF2 | 1.955E-06 | 0.89424651 | 0.5247 | 1.95516E-06 | 1.95516E-06 | 3.7852E-05 | 0.00011808 | 0.900147175 | 0.51375 | 0.000118075 | 0.004447735 | 0.009163157 | 0.016558637 | -1.780975421 | FALSE | TRUE | FALSE |
| SGCE | 1.933E-06 | 0.89425339 | 1 | 1.79604E-05 | 1.79604E-05 | 3.82998E-05 | 4.5018E-05 | 0.900388734 | 0.50205 | 4.5018E-05 | 4.5018E-05 | 9.58626E-05 | 0.042927935 | -1.367260006 | FALSE | TRUE | FALSE |
| NFE2L3 | 1.913E-06 | 0.89426027 | 0.87585 | 7.64881E-06 | 3.05913E-05 | 3.78882E-05 | 9.4713E-07 | 0.901202673 | 0.7571 | 7.71838E-06 | 3.08566E-05 | 3.20776E-05 | 2.019719196 | 0.305290993 | FALSE | TRUE | FALSE |
| KLHL25 | 1.889E-06 | 0.89426714 | 0.85 | 3.01787E-05 | 0.00048266 | 0.002372242 | 5.3143E-07 | 0.901337039 | 1 | 4.68746E-06 | 1.8732E-05 | 4.18337E-05 | 3.554257951 | 0.550748944 | FALSE | TRUE | FALSE |
| THBS1 | 1.884E-06 | 0.89427402 | 1 | 7.52203E-06 | 7.52203E-06 | 0.000146342 | 5.515E-06 | 0.900836115 | 0.93915 | 0.000206107 | 5.872009454 | 5.872009454 | 0.341654484 | -0.466412875 | FALSE | TRUE | FALSE |
| ABCB11 | 1.87E-06 | 0.8942809 | 0.0348 | 1.86951E-06 | 1.86951E-06 | 6.29409E-05 | 4.4224E-05 | 0.900401758 | 0.51875 | 4.42241E-05 | 4.42241E-05 | 0.000865616 | 0.042273596 | -1.373930804 | FALSE | TRUE | FALSE |
| SEC23B | 1.84E-06 | 0.89428777 | 0.51335 | 1.84007E-06 | 1.84007E-06 | 3.62404E-05 | 5.8294E-05 | 0.900303994 | 0.14195 | 5.82936E-05 | 0.000518239 | 0.000518365 | 0.031565475 | -1.500787664 | FALSE | TRUE | FALSE |
| FAT4 | 1.819E-06 | 0.89429465 | 0.46965 | 1.81894E-06 | 0.000155756 | 0.000709743 | 4.8867E-07 | 0.901362591 | 0.33065 | 4.88667E-07 | 8.98912E-06 | 6.44253E-05 | 3.722246905 | 0.570805178 | FALSE | TRUE | FALSE |
| ABCA9 | 1.814E-06 | 0.89430152 | 0.97955 | 0.000603052 | 0.046871081 | 0.18730681 | 64.2832674 | 0.043879806 | 0.07825 | 0.372342075 | 113.4866653 | 1025.728965 | 2.82252E-08 | -7.549362609 | FALSE | FALSE | FALSE |
| ALAS1 | 1.782E-06 | 0.89430839 | 0.1224 | 4.45927E-07 | 7.11974E-06 | 2.84595E-05 | 6.4821E-09 | 0.901883507 | 0.9406 | 5.63151E-07 | 2.17846E-05 | 2.17846E-05 | 274.8505967 | 2.439096684 | FALSE | TRUE | FALSE |
| TCF7L1 | 1.762E-06 | 0.89431526 | 0.99935 | 0.006161064 | 0.638773171 | 3.952381496 | 4.5471E-06 | 0.900874828 | 0.98905 | 0.004018305 | 0.256223781 | 2.585520682 | 0.387499067 | -0.411729339 | FALSE | TRUE | FALSE |
| HMBS | 1.701E-06 | 0.89432213 | 1 | 1.7014E-06 | 1.7014E-06 | 1.7014E-06 | 2.5677E-06 | 0.901016518 | 0.0075 | 2.56768E-06 | 2.56768E-06 | 2.56768E-06 | 0.662622086 | -0.178734093 | FALSE | TRUE | FALSE |
| TMPRSS6 | 1.694E-06 | 0.894329 | 0.0263 | 1.69406E-06 | 1.69406E-06 | 3.31822E-05 | 3.3959E-07 | 0.901445544 | 0.02415 | 3.39591E-07 | 3.39591E-07 | 6.66824E-06 | 4.988525442 | 0.697972192 | FALSE | TRUE | FALSE |
| CELSR2 | 1.667E-06 | 0.89433587 | 0.67105 | 6.66543E-06 | 0.000426804 | 0.001979842 | 5.4674E-06 | 0.900842569 | 0.8027 | 5.03186E-05 | 0.003767997 | 0.030424037 | 0.30488428 | -0.515864967 | FALSE | TRUE | FALSE |
| IGF1R | 1.552E-06 | 0.89434274 | 0.04295 | 1.55248E-06 | 1.55248E-06 | 5.22572E-05 | 2.7894E-06 | 0.900984351 | 0.0394 | 2.7894E-06 | 2.7894E-06 | 9.40052E-05 | 0.556564857 | -0.254484219 | FALSE | TRUE | FALSE |
| DISP1 | 1.509E-06 | 0.89434961 | 1 | 1.50854E-06 | 1.50854E-06 | 5.0732E-05 | 3.8949E-05 | 0.900447316 | 0.03495 | 3.89488E-05 | 3.89488E-05 | 0.000759532 | 0.038731299 | -1.411937935 | FALSE | TRUE | FALSE |
| MYH2 | 1.444E-06 | 0.89435647 | 0.80375 | 2.29978E-05 | 0.225027593 | 14.31659759 | 8.4664E-07 | 0.901247502 | 0.83755 | 1.35021E-05 | 0.001730192 | 0.013773635 | 1.70571997 | 0.231907734 | FALSE | TRUE | FALSE |
| KRT15 | 1.389E-06 | 0.89436334 | 0.68645 | 5.54529E-06 | 8.84868E-05 | 0.000109761 | 1.9746E-06 | 0.90105509 | 0.50855 | 1.9746E-06 | 1.9746E-06 | 9.80019E-06 | 0.70328344 | -0.152869609 | FALSE | TRUE | FALSE |
| MX1 | 1.346E-06 | 0.8943702 | 0.54605 | 2.98332E-06 | 0.000190238 | 0.000760409 | 0.00051806 | 0.899759593 | 0.0989 | 1.38106E-05 | 0.000880635 | 0.008270787 | 0.002598354 | -2.585301617 | FALSE | TRUE | FALSE |
| COL2A1 | 1.286E-06 | 0.89437707 | 0.79505 | 5.13396E-06 | 0.000327017 | 0.005222256 | 4.4638E-07 | 0.901381747 | 0.251 | 1.11834E-07 | 1.78211E-06 | 7.11669E-06 | 2.881343742 | 0.459595072 | FALSE | TRUE | FALSE |
| APC2 | 1.25E-06 | 0.89438393 | 0.0574 | 1.24986E-06 | 2.39518E-05 | 4.19082E-05 | 2.9759E-06 | 0.900965041 | 0.5265 | 2.74694E-05 | 5.73174E-05 | 0.000529068 | 0.41999028 | -0.376760761 | FALSE | TRUE | FALSE |
| DST | 1.245E-06 | 0.89439079 | 1 | 105466.542 | 1405325006 | 1.8049E+11 | 1.3824E-08 | 0.901788629 | 1 | 0.041592953 | 35.01452436 | 344.8378079 | 90.02721909 | 1.954373835 | FALSE | TRUE | FALSE |
| CAT | 1.244E-06 | 0.89439765 | 0.50345 | 1.13035E-05 | 1.13035E-05 | 2.45936E-05 | 3.5192E-06 | 0.900939282 | 0.0155 | 3.51921E-06 | 3.51921E-06 | 6.96462E-05 | 0.35362704 | -0.451454534 | FALSE | TRUE | FALSE |
| KIF1A | 1.162E-06 | 0.89440451 | 0.53225 | 4.64418E-06 | 2.22677E-05 | 8.89747E-05 | 2.5141E-06 | 0.901022949 | 0.04775 | 2.51407E-06 | 2.51407E-06 | 8.45866E-05 | 0.462318335 | -0.335058882 | FALSE | TRUE | FALSE |
| F5 | 1.156E-06 | 0.89441137 | 0.78835 | 9.79692E-06 | 0.000275364 | 0.001086408 | 1.3373E-05 | 0.900674484 | 0.3528 | 1.33734E-05 | 0.000212911 | 0.001031533 | 0.08645467 | -1.063211542 | FALSE | TRUE | FALSE |
| VPS13C | 1.12E-06 | 0.89441823 | 0.54055 | 1.23985E-05 | 3.70606E-05 | 0.000410275 | 0.00039887 | 0.899838659 | 0.53105 | 0.000398874 | 0.003171477 | 0.06021213 | 0.002807831 | -2.551629018 | FALSE | TRUE | FALSE |

| | | | | | | | | | | | | | | | | | |
|---|---|---|---|---|---|---|---|---|---|---|---|---|---|---|---|---|---|
| DMBT1 | 1.12E-06 | 0.89442509 | 0.70595 | 1.11975E-06 | 7.49539E-05 | 0.000300797 | 0.01997405 | 0.893865182 | 0.66395 | 0.019974052 | 1.464762891 | 13.00040428 | 5.606E-05 | -4.251347041 | FALSE | TRUE | FALSE |
| SYNJ1 | 1.101E-06 | 0.89443194 | 1 | 1.10132E-06 | 1.10132E-06 | 3.70203E-05 | 3.669E-06 | 0.900926398 | 1 | 3.66901E-06 | 3.66901E-06 | 0.0001235 | 0.300167668 | -0.522636089 | FALSE | TRUE | FALSE |
| NOTCH4 | 1.055E-06 | 0.8944388 | 0.847 | 8.84857E-06 | 0.000564726 | 0.002702404 | 0.10578113 | 0.874432386 | 0.15325 | 0.001652821 | 29.38582997 | 470.2924397 | 9.97448E-06 | -5.001109825 | FALSE | TRUE | FALSE |
| SULF1 | 1.036E-06 | 0.89444565 | 0.27275 | 2.5924E-07 | 4.14293E-06 | 1.65681E-05 | 1.4235E-06 | 0.901144976 | 0.51125 | 1.42353E-06 | 2.27716E-05 | 0.000111985 | 0.727920362 | -0.137916132 | FALSE | TRUE | FALSE |
| NDUFS1 | 1.023E-06 | 0.89445251 | 0.78545 | 9.04751E-06 | 0.000328246 | 0.001311714 | 1.3232E-07 | 0.901591966 | 0.69535 | 5.2848E-07 | 8.437E-06 | 8.437E-06 | 7.728390589 | 0.888089063 | FALSE | TRUE | FALSE |
| CLUH | 9.933E-07 | 0.89445936 | 0.0446 | 9.93347E-07 | 9.93347E-07 | 1.9159E-05 | 1.8325E-06 | 0.901080788 | 0.03805 | 1.83249E-06 | 1.83249E-06 | 3.54958E-05 | 0.542074559 | -0.265940975 | FALSE | TRUE | FALSE |
| FBLN2 | 9.859E-07 | 0.89446621 | 0.3346 | 9.85903E-07 | 1.57522E-05 | 4.28088E-05 | 3.3706E-07 | 0.901451919 | 0.03025 | 3.37055E-07 | 3.37055E-07 | 6.58033E-06 | 2.925050414 | 0.466133356 | FALSE | TRUE | FALSE |
| LPIN3 | 9.849E-07 | 0.89447306 | 0.85165 | 1.8874E-05 | 0.002741739 | 0.025525603 | 3.3335E-05 | 0.900473331 | 0.54215 | 7.67964E-05 | 0.011578405 | 0.111839898 | 0.029544077 | -1.529529578 | FALSE | TRUE | FALSE |
| PTPN21 | 8.965E-07 | 0.89447991 | 0.02615 | 1.04207E-07 | 8.96476E-07 | 4.35322E-06 | 2.3009E-07 | 0.901528358 | 0.52255 | 2.30088E-07 | 2.00134E-06 | 9.75183E-06 | 3.896235012 | 0.590645145 | FALSE | TRUE | FALSE |
| SNAPC4 | 8.674E-07 | 0.89448676 | 1 | 2.419622929 | 188.5766036 | 709.6169405 | 4.6076E-05 | 0.900369192 | 0.0388 | 4.6076E-05 | 4.6076E-05 | 0.000893967 | 0.018826394 | -1.725232848 | FALSE | TRUE | FALSE |
| LPIN1 | 7.644E-07 | 0.89449361 | 0.75415 | 6.68253E-06 | 5.9965E-05 | 0.000131123 | 2.3368E-06 | 0.901029379 | 1 | 0.000186126 | 0.0164091 | 0.0164091 | 0.32709002 | -0.485332707 | FALSE | TRUE | FALSE |
| CTC1 | 7.482E-07 | 0.89450046 | 0.35755 | 7.48248E-07 | 6.54811E-05 | 0.000634838 | 0.00013926 | 0.900134085 | 0.5069 | 0.000139261 | 0.013635485 | 0.139602039 | 0.005372975 | -2.269785148 | FALSE | TRUE | FALSE |
| KCNS3 | 7.147E-07 | 0.89450731 | 1 | 2.85579E-06 | 2.85579E-06 | 1.41416E-05 | 1.4471E-06 | 0.901125728 | 1 | 1.44706E-06 | 1.44706E-06 | 2.86686E-05 | 0.493910927 | -0.306351366 | FALSE | TRUE | FALSE |
| KCNH8 | 7.055E-07 | 0.89451415 | 1 | 7.0548E-07 | 7.0548E-07 | 1.38136E-05 | 0.0008404 | 0.899561403 | 0.13155 | 0.000210603 | 0.000840399 | 0.004134125 | 0.000839459 | -3.076000575 | FALSE | TRUE | FALSE |
| IFT140 | 6.767E-07 | 0.894521 | 1 | 0.013879087 | 0.478394079 | 1.865023141 | 0.00108944 | 0.899362458 | 1 | 0.001089444 | 0.001089444 | 0.036669887 | 0.000621147 | -3.206805688 | FALSE | TRUE | FALSE |
| UMODL1 | 6.715E-07 | 0.89452784 | 0.9924 | 0.12508993 | 9.822740655 | 90.14176856 | 0.02368578 | 0.892800904 | 0.7782 | 0.439246656 | 35.86237126 | 335.5634656 | 2.83496E-05 | -4.547452917 | FALSE | TRUE | FALSE |
| SLC6A19 | 6.157E-07 | 0.89453468 | 0.24315 | 1.53941E-07 | 2.46294E-06 | 9.85519E-06 | 4.1975E-05 | 0.900421288 | 0.0527 | 6.56805E-07 | 4.19747E-05 | 0.000167901 | 0.014667707 | -1.833637768 | FALSE | TRUE | FALSE |
| CHD8 | 5.604E-07 | 0.89454153 | 1 | 5.60442E-07 | 1.06813E-05 | 1.86327E-05 | 1.6414E-06 | 0.901112893 | 1 | 1.64136E-06 | 3.14602E-05 | 5.47151E-05 | 0.341449414 | -0.466673629 | FALSE | TRUE | FALSE |
| FOXJ2 | 5.595E-07 | 0.89454837 | 0.01595 | 1.39991E-07 | 5.59491E-07 | 2.76811E-06 | 2.7808E-07 | 0.901477412 | 0.0131 | 2.78081E-07 | 2.78081E-07 | 1.37785E-06 | 2.011975372 | 0.30362266 | FALSE | TRUE | FALSE |
| SETD2 | 5.372E-07 | 0.89455521 | 1 | 5.37202E-07 | 1.03634E-05 | 1.79212E-05 | 0.00100388 | 0.899435493 | 0.0007 | 1.54973E-06 | 1.54973E-06 | 5.18164E-05 | 0.000535125 | -3.27154447 | FALSE | TRUE | FALSE |
| WDR19 | 5.269E-07 | 0.89456205 | 0.8811 | 3.3743E-05 | 0.004735071 | 0.018963519 | 6.5374E-05 | 0.900271364 | 0.1535 | 9.01799E-06 | 0.000577638 | 0.002312358 | 0.008059344 | -2.093700317 | FALSE | TRUE | FALSE |
| ROR2 | 5.164E-07 | 0.89456889 | 0.75795 | 5.1635E-07 | 2.06186E-06 | 1.74181E-05 | 1.4395E-06 | 0.901138561 | 0.2768 | 1.43955E-06 | 5.74968E-06 | 4.86063E-05 | 0.358688865 | -0.445282105 | FALSE | TRUE | FALSE |
| NRXN3 | 4.92E-07 | 0.89457572 | 0.9691 | 7.86161E-06 | 3.83351E-05 | 0.000153251 | 5.8646E-07 | 0.901311474 | 0.51635 | 2.34363E-06 | 2.34363E-06 | 4.58252E-05 | 0.839002706 | -0.076236639 | FALSE | TRUE | FALSE |
| MAN2B1 | 4.825E-07 | 0.89458256 | 0.87325 | 7.69176E-06 | 0.026729267 | 0.026729267 | 0.01598316 | 0.894859984 | 0.68695 | 0.063873748 | 0.573222843 | 2.803448303 | 3.01886E-05 | -4.520156943 | FALSE | TRUE | FALSE |
| MYH3 | 4.526E-07 | 0.8945894 | 0.94065 | 3.43356E-05 | 0.002883781 | 0.005969637 | 2.4631E-05 | 0.900518824 | 0.2881 | 2.46311E-05 | 0.000220152 | 0.004248839 | 0.018373598 | -1.735805794 | FALSE | TRUE | FALSE |
| VPS13A | 4.484E-07 | 0.89459623 | 0.9835 | 1.048303077 | 85.1866141 | 794.6252957 | 2.1123E-07 | 0.901547449 | 1 | 1.47323E-05 | 0.057929417 | 0.057929417 | 2.122945429 | 0.326938831 | FALSE | TRUE | FALSE |
| COL14A1 | 4.073E-07 | 0.89460306 | 0.87885 | 1.62644E-06 | 1.39527E-05 | 0.000117164 | 3.8909E-07 | 0.901420035 | 0.7601 | 1.55367E-06 | 0.003840234 | 0.07456109 | 1.046821062 | 0.019872452 | FALSE | TRUE | FALSE |
| LRP4 | 3.928E-07 | 0.8946099 | 0.6416 | 3.92849E-07 | 2.49858E-05 | 0.000119526 | 1.4365E-07 | 0.901585609 | 0.80215 | 5.72944E-07 | 2.11316E-05 | 0.000101721 | 2.734723347 | 0.436913398 | FALSE | TRUE | FALSE |
| KRT4 | 3.885E-07 | 0.89461673 | 0.96955 | 3.17222E-05 | 0.000506587 | 0.002025174 | 0.00010506 | 0.900192964 | 0.0695 | 2.62835E-05 | 0.000420053 | 0.000420053 | 0.003698016 | -2.432031191 | FALSE | TRUE | FALSE |
| EGF | 3.726E-07 | 0.89462356 | 0.8898 | 5.95219E-06 | 9.51699E-05 | 0.000800638 | 2.9222E-05 | 0.900499332 | 0.30405 | 7.31078E-06 | 0.000467216 | 0.001868899 | 0.012752102 | -1.894418232 | FALSE | TRUE | FALSE |
| GPI | 3.587E-07 | 0.89463039 | 1 | 2.28385E-05 | 9.12429E-05 | 0.000364619 | 6.0162E-05 | 0.900284418 | 0.5027 | 0.000138057 | 0.002204028 | 0.00880967 | 0.005962124 | -2.224599005 | FALSE | TRUE | FALSE |
| DNAH3 | 3.53E-07 | 0.89463722 | 0.0003 | 6.34117E-12 | 2.3072E-09 | 1.04031E-08 | 3.0994E-12 | 0.902267463 | 0.967 | 1.52189E-10 | 3.65629E-08 | 2.71876E-07 | 113895.7299 | 5.056507442 | FALSE | TRUE | FALSE |
| MED13 | 3.434E-07 | 0.89464405 | 0.76725 | 1.37228E-06 | 6.61708E-06 | 0.000105671 | 2.7489E-06 | 0.900990786 | 0.7631 | 2.58966E-05 | 0.000250388 | 0.001212544 | 0.124938036 | -0.903305327 | FALSE | TRUE | FALSE |
| BEST2 | 2.459E-07 | 0.89465088 | 0.96705 | 1.56899E-05 | 0.000250876 | 0.001241105 | 4.8611E-05 | 0.900362676 | 0.9614 | 0.027982815 | 2.21795777 | 28.76443974 | 0.005057516 | -2.296062709 | FALSE | TRUE | FALSE |
| MTMR4 | 2.397E-07 | 0.89465771 | 0.03495 | 2.39739E-07 | 2.39739E-07 | 4.67765E-06 | 6.1703E-07 | 0.901298686 | 0.03005 | 6.17028E-07 | 6.17028E-07 | 1.20746E-05 | 0.388537603 | -0.410566944 | FALSE | TRUE | FALSE |
| ABCB1 | 2.395E-07 | 0.89466453 | 1 | 2.39508E-07 | 2.39508E-07 | 8.06304E-06 | 5.7217E-07 | 0.901324258 | 1 | 5.72169E-07 | 5.72169E-07 | 1.12055E-05 | 0.418596079 | -0.378204844 | FALSE | TRUE | FALSE |
| MATN2 | 2.306E-07 | 0.89467136 | 0.97055 | 3.19842E-05 | 0.002050863 | 0.010031139 | 4.0147E-07 | 0.901407275 | 0.90965 | 6.42249E-06 | 0.000102846 | 0.00041171 | 0.574460283 | -0.240739992 | FALSE | TRUE | FALSE |
| MYO9A | 2.166E-07 | 0.89467818 | 0.3553 | 5.42416E-08 | 8.65539E-07 | 7.20568E-06 | 6.2171E-08 | 0.901655492 | 0.2954 | 6.21706E-08 | 2.99042E-07 | 4.76933E-06 | 3.484739179 | 0.542170278 | FALSE | TRUE | FALSE |
| COL25A1 | 2.152E-07 | 0.89468501 | 1 | 2.15185E-07 | 2.15185E-07 | 4.2467E-06 | 6.2553E-07 | 0.901292291 | 0.01485 | 6.25528E-07 | 6.25528E-07 | 1.23645E-05 | 0.34400515 | -0.463435056 | FALSE | TRUE | FALSE |
| SLC4A1 | 2.13E-07 | 0.89469183 | 0.509 | 8.51259E-07 | 8.51259E-07 | 1.66664E-05 | 1.3187E-06 | 0.901164216 | 0.51265 | 1.31872E-06 | 1.31872E-06 | 2.58842E-05 | 0.161554418 | -0.791681161 | FALSE | TRUE | FALSE |
| MLH3 | 2.119E-07 | 0.89469865 | 1 | 0.019671899 | 1.534854533 | 6.132692631 | 0.00093218 | 0.899508425 | 0.26705 | 0.000932176 | 0.00372521 | 0.03140357 | 0.000227295 | -3.643409687 | FALSE | TRUE | FALSE |
| SDK1 | 2.11E-07 | 0.89470547 | 0.99855 | 0.081212571 | 1.536009293 | 10.8730557 | 2.5442E-07 | 0.901515626 | 1 | 4.05775E-06 | 0.016333272 | 0.016333272 | 0.829226854 | -0.081326642 | FALSE | TRUE | FALSE |
| MBD1 | 2.029E-07 | 0.89471229 | 0.51355 | 2.02907E-07 | 2.02907E-07 | 3.99214E-06 | 1.1368E-07 | 0.901604677 | 0.0201 | 1.13679E-07 | 1.13679E-07 | 2.24098E-06 | 1.784914719 | 0.251617471 | FALSE | TRUE | FALSE |
| CHD1 | 2.029E-07 | 0.89471911 | 0.88145 | 1.77354E-05 | 0.000172305 | 0.000594379 | 3.1844E-07 | 0.901464667 | 1 | 0.000270938 | 0.027557942 | 0.288007151 | 0.63711065 | -0.195785135 | FALSE | TRUE | FALSE |
| POLQ | 1.954E-07 | 0.89472593 | 0.8053 | 3.12612E-06 | 0.00187102 | 0.029956227 | 5.7183E-09 | 0.901889826 | 0.65805 | 1.12516E-08 | 1.45214E-06 | 6.98334E-06 | 34.17957255 | 1.533766627 | FALSE | TRUE | FALSE |
| ALK | 1.843E-07 | 0.89473275 | 0.66615 | 3.01406E-07 | 1.38758E-05 | 5.54491E-05 | 1.0993E-08 | 0.901813948 | 0.941 | 4.9586E-07 | 1.38262E-05 | 3.83873E-05 | 16.76731074 | 1.224463413 | FALSE | TRUE | FALSE |
| BBX | 1.733E-07 | 0.89473956 | 0.37355 | 1.73281E-07 | 3.40828E-06 | 1.36199E-05 | 4.6387E-09 | 0.901921407 | 0.87915 | 7.3981E-08 | 2.9556E-07 | 1.45804E-06 | 37.35558466 | 1.572355538 | FALSE | TRUE | FALSE |
| MFN2 | 1.606E-07 | 0.89474638 | 0.8963 | 6.41573E-07 | 1.0243E-05 | 4.09432E-05 | 1.7441E-06 | 0.901093633 | 0.66455 | 1.74407E-06 | 2.78577E-05 | 0.000111378 | 0.092099113 | -1.035744554 | FALSE | TRUE | FALSE |
| ZNFX1 | 1.499E-07 | 0.89475319 | 0.8839 | 5.98392E-07 | 5.46514E-06 | 4.57992E-05 | 3.0234E-05 | 0.900492833 | 0.14275 | 3.21385E-06 | 3.02339E-05 | 0.000146429 | 0.004956512 | -2.304823804 | FALSE | TRUE | FALSE |
| TNS1 | 1.446E-07 | 0.89476001 | 1 | 1.31578E-06 | 2.77783E-06 | 2.52728E-05 | 6.751E-07 | 0.901279498 | 0.04815 | 6.75097E-07 | 6.75097E-07 | 2.26956E-05 | 0.214224142 | -0.669131589 | FALSE | TRUE | FALSE |
| ACLY | 1.423E-07 | 0.89476682 | 1 | 5.68272E-07 | 5.68272E-07 | 1.10767E-05 | 2.0866E-06 | 0.901048663 | 0.2744 | 2.08664E-06 | 8.33421E-06 | 7.04182E-05 | 0.068218596 | -1.166097222 | FALSE | TRUE | FALSE |
| GRM6 | 1.349E-07 | 0.89477363 | 0.08045 | 8.86222E-10 | 2.25341E-07 | 2.64327E-06 | 1.265E-14 | 0.902373933 | 0.9823 | 3.209E-12 | 2.76328E-10 | 1.10349E-09 | 10662113.35 | 7.027843295 | FALSE | TRUE | FALSE |
| PDE6A | 1.278E-07 | 0.89478044 | 0.61305 | 5.10662E-07 | 3.99955E-05 | 0.000159855 | 0.00019992 | 0.900022681 | 0.5964 | 0.000370497 | 0.051208836 | 0.204992565 | 0.000639252 | -3.194327765 | FALSE | TRUE | FALSE |
| SPG11 | 1.073E-07 | 0.89478725 | 0.2342 | 1.33018E-08 | 6.82644E-06 | 6.50494E-05 | 3.4505E-12 | 0.902254922 | 0.98545 | 8.96932E-10 | 6.3018E-08 | 5.47356E-07 | 31110.2993 | 4.492904189 | FALSE | TRUE | FALSE |
| COL4A1 | 1.021E-07 | 0.89479406 | 0.0022 | 3.31724E-10 | 5.27895E-09 | 2.5582E-08 | 1.0344E-08 | 0.901839254 | 0.38875 | 1.0344E-08 | 3.48245E-07 | 1.38999E-06 | 9.866999969 | 0.994185127 | FALSE | TRUE | FALSE |
| HLCS | 1.018E-07 | 0.89480087 | 1 | 1.01833E-07 | 1.01833E-07 | 2.00518E-06 | 2.5529E-07 | 0.901509259 | 1 | 2.55294E-07 | 2.55294E-07 | 5.03629E-06 | 0.398886518 | -0.399150642 | FALSE | TRUE | FALSE |
| CNTNAP4 | 9.361E-08 | 0.89480768 | 1 | 9.36128E-08 | 9.36128E-08 | 3.15195E-06 | 1.7023E-07 | 0.901560172 | 1 | 1.52221E-06 | 1.39707E-05 | 5.13127E-05 | 0.5499257 | -0.259695984 | FALSE | TRUE | FALSE |
| ITGB2 | 8.674E-08 | 0.89481449 | 0.52105 | 7.82686E-07 | 7.82686E-07 | 1.53106E-05 | 1.4062E-06 | 0.90115139 | 0.0234 | 1.40625E-06 | 1.40625E-06 | 2.7581E-05 | 0.061681055 | -1.209848207 | FALSE | TRUE | FALSE |
| ATP13A2 | 8.462E-08 | 0.89482129 | 0.8809 | 2.92078E-06 | 2.59196E-05 | 9.8458E-05 | 1.0707E-05 | 0.900732732 | 0.26775 | 2.68145E-06 | 1.07072E-05 | 9.04832E-05 | 0.0079032 | -2.102197044 | FALSE | TRUE | FALSE |
| PYGB | 8.078E-08 | 0.8948281 | 0.9995 | 0.035020038 | 21.62094107 | 347.1690987 | 0.00072819 | 0.899634159 | 0.71425 | 0.002913045 | 0.747880743 | 12.00117147 | 0.000110936 | -3.954926748 | FALSE | TRUE | FALSE |
| EPB41 | 7.675E-08 | 0.8948349 | 0.8554 | 3.06792E-07 | 4.90552E-06 | 2.42178E-05 | 5.3169E-05 | 0.900336604 | 0.06405 | 3.32436E-06 | 0.000212712 | 0.000851208 | 0.001443551 | -2.840567942 | FALSE | TRUE | FALSE |
| PTCH2 | 7.19E-08 | 0.8948417 | 0.67725 | 2.8671E-07 | 9.74403E-06 | 4.76127E-05 | 1.6711E-06 | 0.901100053 | 0.0501 | 1.98198E-07 | 1.67807E-06 | 8.18941E-06 | 0.043023578 | -1.366293476 | FALSE | TRUE | FALSE |
| CBLB | 7.047E-08 | 0.89484851 | 1 | 7.04657E-08 | 7.04657E-08 | 1.38252E-06 | 1.124E-07 | 0.901611032 | 0.02345 | 1.12397E-07 | 1.12397E-07 | 2.21025E-06 | 0.626935306 | -0.202777272 | FALSE | TRUE | FALSE |
| ATIC | 6.986E-08 | 0.89485531 | 1 | 5.92887E-07 | 2.36899E-06 | 5.51778E-06 | 4.8357E-07 | 0.901368977 | 0.26285 | 4.83569E-07 | 1.93292E-06 | 4.09837E-06 | 0.144477844 | -0.840198746 | FALSE | TRUE | FALSE |
| COL22A1 | 6.953E-08 | 0.89486211 | 0.91895 | 0.00350992 | 1.098722059 | 10.0801243 | 5.4348E-05 | 0.900323562 | 0.14995 | 6.53249E-06 | 5.43478E-05 | 0.000861674 | 0.001279403 | -2.892992526 | FALSE | TRUE | FALSE |
| EPPK1 | 6.879E-08 | 0.89486891 | 0.83515 | 0.028985346 | 22.19565991 | 2054.598704 | 8.732E-07 | 0.901234698 | 0.98005 | 0.074654556 | 943.2873357 | 17821.20477 | 0.078782885 | -1.103568122 | FALSE | TRUE | FALSE |
| SLC12A6 | 6.423E-08 | 0.89487571 | 0.8191 | 3.23579E-07 | 5.16731E-06 | 2.06571E-05 | 2.8855E-07 | 0.90147104 | 0.513 | 2.88552E-07 | 4.61144E-06 | 1.01151E-05 | 0.22260378 | -0.652467466 | FALSE | TRUE | FALSE |
| SORL1 | 6.225E-08 | 0.89488251 | 0.0415 | 4.34807E-10 | 2.76131E-08 | 2.48422E-07 | 1.2734E-10 | 0.902072715 | 0.9908 | 1.79301E-08 | 2.59374E-06 | 1.03578E-05 | 488.8329148 | 2.689160441 | FALSE | TRUE | FALSE |
| COL15A1 | 5.994E-08 | 0.8948893 | 0.4003 | 2.75107E-08 | 2.39569E-07 | 1.16472E-06 | 1.9996E-08 | 0.90174429 | 0.51725 | 1.99956E-08 | 1.99956E-08 | 3.89862E-07 | 2.997777066 | 0.476799333 | FALSE | TRUE | FALSE |
| SCN9A | 5.924E-08 | 0.8948961 | 0.94245 | 4.53629E-06 | 0.000384534 | 0.000796766 | 3.6684E-07 | 0.901432791 | 0.043 | 3.66842E-07 | 3.66842E-07 | 1.23175E-05 | 0.161484779 | -0.791868407 | FALSE | TRUE | FALSE |
| MYO19 | 5.691E-08 | 0.89490289 | 0.8823 | 4.74167E-07 | 0.001796585 | 0.001796585 | 0.00726572 | 0.897469902 | 0.6543 | 0.007265721 | 0.561259067 | 5.112528201 | 7.83273E-06 | -5.10608672 | FALSE | TRUE | FALSE |
| LRP1 | 5.209E-08 | 0.89490969 | 0.92675 | 1.88611E-06 | 0.0002793 | 0.002302001 | 1.6197E-07 | 0.901566532 | 0.96405 | 8.55437E-05 | 0.035080968 | 0.38872646 | 0.321566245 | -0.492729545 | FALSE | TRUE | FALSE |
| MST1 | 5.203E-08 | 0.89491648 | 0.5127 | 9.5532E-08 | 3.32123E-06 | 2.9627E-05 | 2.7654E-07 | 0.901483783 | 0.04965 | 3.21742E-08 | 2.76535E-07 | 1.10555E-06 | 0.18813607 | -0.725527932 | FALSE | TRUE | FALSE |

| | | | | | | | | | | | | | | | | | |
|---|---|---|---|---|---|---|---|---|---|---|---|---|---|---|---|---|---|
| SACS | 4.964E-08 | 0.89492328 | 0.9524 | 5.28166E-05 | 0.110045364 | 2.264445693 | 2.2961E-06 | 0.901035808 | 0.9306 | 0.001258453 | 5.1147855 | 175.8124463 | 0.021616889 | -1.665206814 | FALSE | TRUE | FALSE |
| TPH2 | 4.569E-08 | 0.89493007 | 0.7583 | 1.82562E-07 | 7.2964E-07 | 3.61669E-06 | 1.1305E-06 | 0.901183448 | 0.0126 | 2.82822E-07 | 1.13049E-06 | 2.39784E-06 | 0.040415951 | -1.393447202 | FALSE | TRUE | FALSE |
| SRCAP | 4.547E-08 | 0.89493686 | 0.0023 | 1.59971E-10 | 1.33151E-09 | 5.307E-09 | 2.1239E-10 | 0.902060123 | 1 | 2.12394E-10 | 3.98776E-09 | 7.06672E-09 | 214.0916847 | 2.3305998 | FALSE | TRUE | FALSE |
| MEGF11 | 4.344E-08 | 0.89494365 | 0.5111 | 3.84475E-07 | 3.48821E-06 | 3.24227E-05 | 1.5421E-09 | 0.901990814 | 1 | 1.31832E-08 | 1.31832E-08 | 5.211E-08 | 28.1695076 | 1.449779256 | FALSE | TRUE | FALSE |
| ACADM | 4.152E-08 | 0.89495044 | 0.94145 | 3.17032E-06 | 0.000459682 | 0.001837551 | 4.7438E-07 | 0.901375362 | 0.23455 | 4.74385E-07 | 7.58205E-06 | 3.03234E-05 | 0.08752389 | -1.057873389 | FALSE | TRUE | FALSE |
| ATP6V0A4 | 4.007E-08 | 0.89495723 | 0.99625 | 0.000101601 | 0.018050539 | 0.135741734 | 8.9825E-07 | 0.901228295 | 0.8178 | 3.58857E-06 | 1.43402E-05 | 7.06718E-05 | 0.044606222 | -1.35060456 | FALSE | TRUE | FALSE |
| VCAN | 3.858E-08 | 0.89496401 | 1 | 1.41547E-06 | 2.25756E-05 | 0.000187963 | 4.014E-07 | 0.901413656 | 0.7643 | 4.01401E-07 | 7.62633E-06 | 3.04602E-05 | 0.096113037 | -1.017217699 | FALSE | TRUE | FALSE |
| TPCN2 | 3.739E-08 | 0.8949708 | 0.80805 | 3.2906E-07 | 1.18727E-05 | 4.7433E-05 | 4.3238E-05 | 0.900414779 | 0.04255 | 6.76572E-07 | 1.0812E-05 | 5.329E-05 | 0.000864707 | -3.063131016 | FALSE | TRUE | FALSE |
| FBN1 | 3.691E-08 | 0.89497759 | 0.02375 | 4.90872E-10 | 9.24869E-09 | 7.70353E-08 | 7.9762E-11 | 0.902091595 | 0.71115 | 7.97616E-11 | 1.51335E-09 | 1.06031E-08 | 462.7369191 | 2.665334151 | FALSE | TRUE | FALSE |
| HK2 | 3.533E-08 | 0.89498437 | 0.3845 | 3.53329E-08 | 6.9133E-07 | 2.76265E-06 | 3.1415E-08 | 0.901699911 | 0.3587 | 3.1415E-08 | 5.0178E-07 | 2.00616E-06 | 1.12471345 | 0.051041889 | FALSE | TRUE | FALSE |
| CSMD3 | 3.502E-08 | 0.89499115 | 1 | 3.30965E+29 | 5.34704E+36 | 1.85135E+40 | 1.535E-08 | 0.901769632 | 1 | 6.47325E+38 | 1.11502E+48 | 3.35386E+51 | 2.281153349 | 0.358154481 | FALSE | TRUE | FALSE |
| ALMS1 | 3.34E-08 | 0.89499794 | 0.7152 | 3.34016E-08 | 2.12227E-06 | 8.66713E-06 | 1.7445E-07 | 0.901553811 | 0.29915 | 2.21452E-08 | 4.17627E-07 | 3.28995E-06 | 0.191464175 | -0.717912474 | FALSE | TRUE | FALSE |
| H6PD | 3.079E-08 | 0.89500472 | 0.77395 | 4.90893E-07 | 0.000166383 | 0.002657034 | 1.786E-05 | 0.900583741 | 0.16655 | 9.08923E-07 | 0.000285455 | 0.002628944 | 0.001723738 | -2.763528715 | FALSE | TRUE | FALSE |
| GJD4 | 2.836E-08 | 0.8950115 | 1 | 1.13299E-07 | 1.13299E-07 | 5.61581E-07 | 3.3066E-05 | 0.900479832 | 0.13455 | 2.07012E-06 | 3.30664E-05 | 0.000132204 | 0.00085753 | -3.066750896 | FALSE | TRUE | FALSE |
| HSPG2 | 2.811E-08 | 0.89501828 | 0.7846 | 2.31544E-07 | 4.14987E-06 | 2.42532E-05 | 4.2294E-07 | 0.90138813 | 0.7801 | 1.34946E-06 | 3.93605E-05 | 0.000116928 | 0.06645202 | -1.177491812 | FALSE | TRUE | FALSE |
| NDUFV1 | 2.739E-08 | 0.89502506 | 0.14005 | 2.73903E-08 | 1.09409E-07 | 2.32383E-07 | 5.0998E-09 | 0.901902461 | 1 | 8.13418E-08 | 3.24981E-07 | 4.02426E-07 | 5.370888502 | 0.730046137 | FALSE | TRUE | FALSE |
| SCUBE2 | 2.515E-08 | 0.89503184 | 0.50985 | 2.51512E-08 | 2.1405E-07 | 4.93447E-07 | 1.1459E-07 | 0.901598322 | 0.5108 | 1.14592E-07 | 9.92433E-07 | 2.25336E-06 | 0.219485324 | -0.658594514 | FALSE | TRUE | FALSE |
| SLC26A4 | 2.329E-08 | 0.89503862 | 0.2639 | 2.61645E-09 | 2.3286E-08 | 5.16202E-08 | 8.5907E-10 | 0.902034931 | 0.01625 | 8.59066E-10 | 8.59066E-10 | 1.69762E-08 | 27.10622654 | 1.433069064 | FALSE | TRUE | FALSE |
| ABCC2 | 2.107E-08 | 0.89504539 | 0.93875 | 1.22472E-06 | 7.88196E-05 | 0.000188671 | 1.1386E-05 | 0.900713324 | 0.89795 | 9.23167E-05 | 0.006534111 | 0.056974967 | 0.001850809 | -2.732638464 | FALSE | TRUE | FALSE |
| EPB41L2 | 1.945E-08 | 0.89505217 | 1 | 7.19602E-07 | 2.73377E-05 | 5.63584E-05 | 2.2038E-07 | 0.901534722 | 0.27005 | 2.20376E-07 | 8.802E-07 | 4.33235E-06 | 0.088238789 | -1.054340461 | FALSE | TRUE | FALSE |
| MOV10L1 | 1.719E-08 | 0.89505895 | 0.76325 | 1.71902E-08 | 6.8643E-08 | 5.79115E-07 | 4.8323E-08 | 0.901674533 | 0.5156 | 4.83227E-08 | 4.83227E-08 | 9.44643E-07 | 0.355738172 | -0.44886953 | FALSE | TRUE | FALSE |
| PRB3 | 1.692E-08 | 0.89506572 | 1 | 1.6922E-08 | 1.6922E-08 | 1.6922E-08 | 2.1709E-08 | 0.901725275 | 0.752 | 1.66632E-07 | 1.31268E-06 | 1.31268E-06 | 0.779491209 | -0.108188778 | FALSE | TRUE | FALSE |
| COL27A1 | 1.674E-08 | 0.89507249 | 0.9997 | 1.69036E-05 | 1.097508215 | 4.386385463 | 7.5929E-07 | 0.901260303 | 0.52645 | 3.02989E-06 | 4.82822E-05 | 0.000232939 | 0.022050292 | -1.656585664 | FALSE | TRUE | FALSE |
| ASPM | 1.661E-08 | 0.89507927 | 0.5324 | 1.66119E-08 | 4.64221E-06 | 4.02013E-05 | 3.5759E-09 | 0.901934034 | 0.8725 | 2.50134E-07 | 0.000174703 | 0.001621417 | 4.645485831 | 0.66703114 | FALSE | TRUE | FALSE |
| ACAD8 | 1.654E-08 | 0.89508604 | 1 | 2.64118E-07 | 4.22203E-06 | 4.22203E-06 | 5.847E-07 | 0.901317866 | 0.85415 | 2.33829E-06 | 3.74237E-05 | 0.000149773 | 0.028286071 | -1.54842738 | FALSE | TRUE | FALSE |
| DCTN1 | 1.553E-08 | 0.89509281 | 0.75675 | 1.40756E-07 | 1.30924E-06 | 4.73767E-06 | 4.9502E-08 | 0.901668187 | 0.0316 | 4.95022E-08 | 4.95022E-08 | 9.65264E-07 | 0.313753562 | -0.503411335 | FALSE | TRUE | FALSE |
| SCN5A | 1.543E-08 | 0.89509958 | 0.9288 | 7.52766E-06 | 0.007596139 | 0.110657243 | 8.3853E-08 | 0.901630091 | 0.7663 | 7.63597E-07 | 0.000452786 | 0.003080461 | 0.183959896 | -0.735276845 | FALSE | TRUE | FALSE |
| COL6A6 | 1.466E-08 | 0.89510635 | 0.8291 | 9.34417E-07 | 0.000574024 | 0.009168933 | 2.1526E-07 | 0.901541086 | 0.8389 | 1.37362E-05 | 0.009145787 | 0.146417892 | 0.068125037 | -1.166693248 | FALSE | TRUE | FALSE |
| FOXK2 | 1.452E-08 | 0.89511312 | 1 | 4.95988E+17 | 5.30917E+24 | 1.32056E+27 | 1.7305E-08 | 0.901756963 | 1 | 6.32323E+26 | 2.55439E+35 | 1.52215E+39 | 0.839090999 | -0.076190938 | FALSE | TRUE | FALSE |
| CACNB2 | 1.414E-08 | 0.89511988 | 1 | 1.18367E-07 | 1.18367E-07 | 4.78494E-07 | 0.01388902 | 0.895468709 | 0.19755 | 0.013889016 | 0.05551968 | 0.221988718 | 1.01802E-06 | -5.992242434 | FALSE | TRUE | FALSE |
| MYH9 | 1.332E-08 | 0.89512665 | 0.34565 | 3.32883E-09 | 2.53198E-07 | 3.42627E-06 | 1.1737E-09 | 0.902003423 | 0.8708 | 1.75926E-07 | 4.53078E-05 | 0.000728188 | 11.35031494 | 1.055007912 | FALSE | TRUE | FALSE |
| ITPR1 | 1.32E-08 | 0.89513342 | 0.0752 | 1.31953E-08 | 2.70767E-08 | 4.39643E-07 | 8.3374E-09 | 0.901851902 | 1 | 8.33739E-09 | 1.58742E-07 | 2.78458E-07 | 1.582662484 | 0.199388308 | FALSE | TRUE | FALSE |
| KRT73 | 1.26E-08 | 0.89514018 | 0.51045 | 1.25987E-08 | 5.03247E-08 | 1.11062E-07 | 1.417E-08 | 0.901782298 | 0.8766 | 5.66051E-08 | 2.26185E-07 | 2.80059E-07 | 0.889141281 | -0.051029226 | FALSE | TRUE | FALSE |
| PFKL | 1.197E-08 | 0.89514694 | 0.8743 | 4.78292E-08 | 1.91206E-07 | 9.35036E-07 | 8.5841E-07 | 0.901241101 | 0.7606 | 7.31621E-06 | 0.001024796 | 0.004102236 | 0.013941169 | -1.855700808 | FALSE | TRUE | FALSE |
| RAD54L2 | 1.184E-08 | 0.89515371 | 1 | 1.11108E-07 | 1.11108E-07 | 2.15501E-06 | 1.9789E-08 | 0.901750627 | 0.0363 | 1.97889E-08 | 1.97889E-08 | 3.85219E-07 | 0.598426849 | -0.22298893 | FALSE | TRUE | FALSE |
| COL5A2 | 1.081E-08 | 0.89516047 | 0.1328 | 6.79083E-10 | 1.08067E-08 | 5.26092E-08 | 2.9131E-11 | 0.902154478 | 0.87695 | 4.6329E-10 | 1.84825E-09 | 9.02962E-09 | 370.964208 | 2.569332009 | FALSE | TRUE | FALSE |
| JAG1 | 1.039E-08 | 0.89516723 | 1 | 4.14654E-08 | 4.14654E-08 | 8.05756E-07 | 5.5337E-07 | 0.901330649 | 0.27895 | 1.38547E-07 | 5.53372E-07 | 4.67325E-06 | 0.018769515 | -1.72654696 | FALSE | TRUE | FALSE |
| SLC4A9 | 1.005E-08 | 0.89517399 | 0.9532 | 7.28381E-07 | 4.64117E-05 | 0.000185467 | 0.00380423 | 0.898390431 | 0.3502 | 0.00095199 | 0.139566268 | 0.972494323 | 2.64075E-06 | -5.578273461 | FALSE | TRUE | FALSE |
| PRPF8 | 9.784E-09 | 0.89518075 | 0.70955 | 3.90708E-08 | 7.40252E-07 | 5.20069E-06 | 4.8074E-09 | 0.901908777 | 1 | 1.91968E-08 | 3.66027E-07 | 1.46197E-06 | 2.035267203 | 0.308621434 | FALSE | TRUE | FALSE |
| MACF1 | 8.913E-09 | 0.89518751 | 1 | 8.08155E-08 | 1.4639E-06 | 3.07064E-06 | 2.6115E-07 | 0.901502891 | 0.3451 | 2.61149E-07 | 8.50383E-06 | 7.94892E-05 | 0.034128688 | -1.466880413 | FALSE | TRUE | FALSE |
| FER1L6 | 8.259E-09 | 0.89519427 | 0.03225 | 1.09469E-09 | 8.25932E-09 | 6.93521E-08 | 5.2536E-07 | 0.901349817 | 0.46205 | 5.25365E-07 | 6.81524E-05 | 0.000567427 | 0.015721123 | -1.803516425 | FALSE | TRUE | FALSE |
| MAP3K9 | 7.364E-09 | 0.89520102 | 1 | 7.36421E-09 | 7.36421E-09 | 1.43494E-07 | 2.0034E-08 | 0.901737953 | 1 | 1.84674E-07 | 1.84674E-07 | 3.60959E-06 | 0.367590825 | -0.434635337 | FALSE | TRUE | FALSE |
| FRAS1 | 7.029E-09 | 0.89520778 | 1 | 18186792.2 | 3.40703E+12 | 4.94238E+14 | 2.6659E-07 | 0.901496523 | 1 | 3.32087E+15 | 4.57378E+21 | 2.05194E+24 | 0.026365335 | -1.5789667 | FALSE | TRUE | FALSE |
| APC | 6.391E-09 | 0.89521453 | 0.07175 | 6.3906E-09 | 3.05462E-08 | 2.12148E-07 | 7.033E-08 | 0.901649143 | 0.94185 | 2.56713E-06 | 0.000232193 | 0.000926546 | 0.090866205 | -1.041597609 | FALSE | TRUE | FALSE |
| HRNR | 6.283E-09 | 0.89522129 | 1 | 3.2866E+172 | 6.1837E+193 | 1.0916E+203 | 3.998E-05 | 0.900440811 | 1 | 1.6999E+159 | 6.5144E+178 | 9.6383E+187 | 0.000157162 | -3.803651625 | FALSE | TRUE | FALSE |
| NPHP3 | 5.994E-09 | 0.89522804 | 0.84385 | 5.12114E-08 | 1.79201E-06 | 7.15754E-06 | 5.5828E-08 | 0.90166184 | 0.33235 | 2.56975E-08 | 8.90378E-07 | 2.00594E-06 | 0.107374942 | -0.969097058 | FALSE | TRUE | FALSE |
| LDLR | 5.808E-09 | 0.89523479 | 0.51795 | 5.02471E-08 | 5.02471E-08 | 9.82757E-07 | 1.6373E-08 | 0.901763298 | 1 | 1.4314E-07 | 1.4314E-07 | 2.80705E-06 | 0.3547091 | -0.45012767 | FALSE | TRUE | FALSE |
| PANK2 | 5.56E-09 | 0.89524155 | 0.016 | 5.55979E-09 | 5.55979E-09 | 1.09835E-07 | 1.4891E-08 | 0.901775965 | 0.97 | 1.12529E-05 | 0.000107788 | 0.001057717 | 0.373359639 | -0.427872632 | FALSE | TRUE | FALSE |
| EP300 | 5.55E-09 | 0.8952483 | 0.0217 | 2.90758E-10 | 5.54985E-09 | 9.68356E-09 | 6.5976E-09 | 0.901877188 | 0.29425 | 1.65184E-09 | 7.93956E-09 | 1.26626E-07 | 0.841194014 | -0.075103826 | FALSE | TRUE | FALSE |
| TGFBI | 5.251E-09 | 0.89525505 | 1 | 2.09812E-08 | 8.38473E-08 | 4.13731E-07 | 6.8386E-06 | 0.90078445 | 0.02735 | 4.27935E-07 | 6.83855E-06 | 2.73477E-05 | 0.00076792 | -3.114683926 | FALSE | TRUE | FALSE |
| CLN3 | 5.127E-09 | 0.8952618 | 1 | 5.12652E-09 | 5.12652E-09 | 1.01451E-07 | 9.1673E-09 | 0.901845578 | 1 | 9.16728E-09 | 9.16728E-09 | 1.81645E-07 | 0.559218674 | -0.252418335 | FALSE | TRUE | FALSE |
| PC | 5.024E-09 | 0.89526854 | 1 | 307.3107687 | 8157814.427 | 401696089 | 4.0423E-05 | 0.900434304 | 0.65495 | 0.000160834 | 0.02745527 | 0.221470475 | 0.00012429 | -3.905564531 | FALSE | TRUE | FALSE |
| ADSL | 4.611E-09 | 0.89527529 | 1 | 4.61069E-09 | 4.61069E-09 | 9.11292E-08 | 1.0413E-07 | 0.901623739 | 0.01445 | 1.04131E-07 | 1.04131E-07 | 2.23608E-07 | 0.044277802 | -1.353813945 | FALSE | TRUE | FALSE |
| PNPLA8 | 4.446E-09 | 0.89528204 | 1 | 4.0443E-08 | 3.77569E-07 | 7.97996E-07 | 7.6206E-09 | 0.901858225 | 1 | 6.5239E-07 | 6.27227E-06 | 6.27227E-06 | 0.583446398 | -0.233999037 | FALSE | TRUE | FALSE |
| KCNV2 | 4.308E-09 | 0.89528878 | 0.856 | 6.84869E-08 | 1.08988E-06 | 4.34936E-06 | 5.0061E-07 | 0.901356204 | 0.03935 | 3.14778E-08 | 5.00607E-07 | 1.99713E-06 | 0.008605474 | -2.065225208 | FALSE | TRUE | FALSE |
| GGCX | 3.939E-09 | 0.89529553 | 1 | 1.36427E-07 | 1.21498E-06 | 2.68631E-06 | 1.3352E-06 | 0.901157803 | 0.9855 | 0.001756531 | 0.553473663 | 2.554033162 | 0.002949828 | -2.530203296 | FALSE | TRUE | FALSE |
| SLC12A7 | 3.45E-09 | 0.89530227 | 0.99665 | 8.79349E-07 | 5.62415E-05 | 0.000272466 | 5.1072E-05 | 0.900356159 | 0.12525 | 3.19589E-06 | 0.000204242 | 0.001825731 | 6.75513E-05 | -4.170366562 | FALSE | TRUE | FALSE |
| ACAD11 | 3.16E-09 | 0.89530901 | 0.2626 | 7.90865E-10 | 3.16037E-09 | 1.55783E-08 | 4.0013E-09 | 0.901927721 | 0.8129 | 1.59869E-08 | 5.58474E-07 | 5.58474E-07 | 0.789833336 | -0.10246454 | FALSE | TRUE | FALSE |
| TRPM1 | 2.95E-09 | 0.89531576 | 0.52155 | 2.94996E-09 | 8.95476E-08 | 4.32547E-07 | 1.2435E-08 | 0.90180762 | 0.2229 | 1.5593E-09 | 4.97047E-08 | 2.09594E-07 | 0.237231015 | -0.624828534 | FALSE | TRUE | FALSE |
| ABCA7 | 2.891E-09 | 0.8953225 | 0.1434 | 2.87616E-12 | 2.45286E-06 | 0.000520007 | 3.6257E-16 | 0.902448948 | 0.8324 | 2.82416E-14 | 3.02535E-08 | 8.84364E-07 | 7974901.108 | 6.901725306 | FALSE | TRUE | FALSE |
| CAPN3 | 2.434E-09 | 0.89532924 | 0.90625 | 1.76898E-07 | 2.09502E-05 | 0.000180946 | 9.0508E-07 | 0.901215486 | 0.3266 | 1.93865E-07 | 1.4471E-05 | 0.000130138 | 0.00268931 | -2.570359056 | FALSE | TRUE | FALSE |
| GIGYF1 | 2.045E-09 | 0.89533598 | 1 | 2.04531E-09 | 2.04531E-09 | 3.98302E-08 | 4.4101E-06 | 0.900881277 | 0.1343 | 4.61762E-07 | 4.41011E-06 | 9.02083E-06 | 0.000463778 | -3.33369022 | FALSE | TRUE | FALSE |
| IL23R | 1.987E-09 | 0.89534272 | 1 | 9.95264E-09 | 9.95264E-09 | 3.93418E-08 | 1.3289E-08 | 0.90179496 | 0.5048 | 7.05718E-08 | 7.05718E-08 | 2.63436E-07 | 0.149521068 | -0.825297611 | FALSE | TRUE | FALSE |
| CACNA1S | 1.841E-09 | 0.89534945 | 0.95515 | 4.65161E-07 | 0.000471149 | 0.007518603 | 2.0514E-11 | 0.902167045 | 0.88255 | 7.20809E-10 | 9.6535E-08 | 7.2654E-07 | 89.75481907 | 1.953057776 | FALSE | TRUE | FALSE |
| KRT83 | 1.781E-09 | 0.89535619 | 1 | 1.78062E-09 | 1.78062E-09 | 3.51145E-08 | 2.9565E-08 | 0.901712595 | 0.01485 | 2.95647E-08 | 2.95647E-08 | 1.46254E-07 | 0.060228092 | -1.220200893 | FALSE | TRUE | FALSE |
| NABP2 | 1.778E-09 | 0.89536293 | 0.00725 | 1.77835E-09 | 1.77835E-09 | 1.77835E-09 | 1.0468E-08 | 0.901832929 | 1 | 1.0468E-08 | 1.0468E-08 | 1.0468E-08 | 0.169884111 | -0.769847238 | FALSE | TRUE | FALSE |
| RPGRIP1L | 1.687E-09 | 0.89536966 | 0.9684 | 1.48602E-05 | 0.000127415 | 0.001119296 | 4.5575E-10 | 0.902053827 | 0.99255 | 0.000603053 | 0.049358386 | 0.18151518 | 3.700627249 | 0.568275342 | FALSE | TRUE | FALSE |
| ANK2 | 1.682E-09 | 0.8953764 | 1 | 1.3045E-05 | 0.001331299 | 0.013925672 | 0.00097029 | 0.899475287 | 0.9946 | 575.0638679 | 8307885.696 | 415127527.2 | 1.73333E-06 | -5.761117972 | FALSE | TRUE | FALSE |
| STAB1 | 1.403E-09 | 0.89538313 | 0.77705 | 2.63286E-09 | 2.6325E-08 | 1.96901E-07 | 2.1976E-09 | 0.901952968 | 0.6545 | 9.54996E-09 | 4.31383E-08 | 7.20222E-07 | 0.638430888 | -0.194886109 | FALSE | TRUE | FALSE |
| ASH1L | 1.355E-09 | 0.89538986 | 1 | 1.26808E-08 | 4.49115E-08 | 4.20332E-07 | 7.5881E-09 | 0.901864546 | 0.07095 | 7.58813E-09 | 1.4464E-07 | 2.52285E-07 | 0.17855667 | -0.748223923 | FALSE | TRUE | FALSE |
| SYNE1 | 1.244E-09 | 0.89539659 | 1 | 6.09744E+34 | 4.5436E+44 | 4.51177E+48 | 3.3895E-09 | 0.901940346 | 1 | 4.9855E+27 | 3.40115E+36 | 5.52746E+40 | 0.367121228 | -0.435190502 | FALSE | TRUE | FALSE |
| ITPR2 | 1.065E-09 | 0.89540332 | 1 | 6.74879E-08 | 2.24667E-06 | 0.000139859 | 1.079E-07 | 0.901617386 | 0.1774 | 1.07896E-07 | 5.17825E-07 | 3.6009E-06 | 0.009871368 | -2.005622651 | FALSE | TRUE | FALSE |
| WDR33 | 8.657E-10 | 0.89541005 | 1 | 7.19764E-08 | 6.81968E-07 | 2.41307E-06 | 1.9338E-09 | 0.901959278 | 1 | 1.54327E-06 | 3.00923E-05 | 0.000150426 | 0.44769237 | -0.349020307 | FALSE | TRUE | FALSE |

| | | | | | | | | | | | | | | | | | |
|---|---|---|---|---|---|---|---|---|---|---|---|---|---|---|---|---|---|
| CADPS | 5.673E-10 | 0.89541678 | 1 | 4.24305E-09 | 3.25715E-08 | 6.33824E-07 | 1.304E-08 | 0.90180129 | 0.5247 | 1.30403E-08 | 1.30403E-08 | 2.54584E-07 | 0.043502225 | -1.361488526 | FALSE | TRUE | FALSE |
| INPPL1 | 4.251E-10 | 0.89542351 | 0.88275 | 6.75422E-09 | 6.09531E-08 | 2.97062E-07 | 1.7571E-09 | 0.901978202 | 1 | 1.75707E-09 | 1.75707E-09 | 3.42567E-08 | 0.241917092 | -0.616333446 | FALSE | TRUE | FALSE |
| SYNJ2 | 4.217E-10 | 0.89543024 | 0.7481 | 3.48601E-09 | 5.54475E-08 | 2.68839E-07 | 2.9718E-08 | 0.901706253 | 0.8715 | 1.01882E-06 | 0.000143036 | 0.000695804 | 0.014191018 | -1.847986444 | FALSE | TRUE | FALSE |
| ABCC8 | 3.425E-10 | 0.89543697 | 0.86705 | 6.6052E-09 | 4.18844E-07 | 1.67106E-06 | 8.3186E-08 | 0.901642793 | 0.1378 | 2.08518E-08 | 3.31947E-07 | 1.61191E-06 | 0.004117585 | -2.385357393 | FALSE | TRUE | FALSE |
| COL5A3 | 2.89E-10 | 0.89544369 | 0.51975 | 2.88995E-10 | 2.12269E-08 | 8.48254E-08 | 3.5366E-12 | 0.902248651 | 0.9584 | 4.73516E-10 | 3.02613E-08 | 1.21047E-07 | 81.71469274 | 1.912300152 | FALSE | TRUE | FALSE |
| SETX | 2.736E-10 | 0.89545042 | 1 | 2.73572E-10 | 5.21683E-09 | 9.10818E-09 | 9.0761E-10 | 0.902028631 | 0.06075 | 9.07612E-10 | 1.74048E-08 | 3.02932E-08 | 0.301419561 | -0.520828566 | FALSE | TRUE | FALSE |
| VPS13B | 2.704E-10 | 0.89545714 | 0.53315 | 2.70372E-10 | 3.87009E-08 | 5.25125E-07 | 6.6342E-12 | 0.902229831 | 0.9758 | 8.31623E-10 | 1.09976E-07 | 8.84998E-07 | 40.75410439 | 1.610171353 | FALSE | TRUE | FALSE |
| SCN1A | 2.09E-10 | 0.89546386 | 0.0524 | 2.08986E-10 | 4.02585E-09 | 6.98807E-09 | 1.0696E-09 | 0.902022331 | 0.049 | 1.06959E-09 | 1.06959E-09 | 3.58362E-08 | 0.195388373 | -0.709101284 | FALSE | TRUE | FALSE |
| NIPBL | 2.036E-10 | 0.89547059 | 1 | 2.03642E-10 | 3.89358E-09 | 6.76612E-09 | 1.0746E-09 | 0.902016029 | 0.05825 | 1.07464E-09 | 2.0656E-08 | 3.58034E-08 | 0.18949809 | -0.722395163 | FALSE | TRUE | FALSE |
| RNF213 | 1.818E-10 | 0.89547731 | 0.3961 | 1.81846E-10 | 8.39335E-09 | 1.52551E-07 | 1.3296E-11 | 0.902179608 | 1 | 8.83579E-11 | 1.62372E-09 | 2.91278E-09 | 13.67674651 | 1.135982798 | FALSE | TRUE | FALSE |
| CHD7 | 1.728E-10 | 0.89548403 | 0.9929 | 3.83215E-06 | 0.002569707 | 0.055996674 | 2.4041E-11 | 0.902160762 | 0.96765 | 2.1589E-09 | 1.37764E-07 | 1.14654E-06 | 7.189460701 | 0.856696314 | FALSE | TRUE | FALSE |
| TSC2 | 1.556E-10 | 0.89549075 | 1 | 40863932.7 | 4.43234E+13 | 9.66459E+15 | 3.1966E-11 | 0.902148193 | 0.9998 | 0.027916129 | 404.0379459 | 26237.88431 | 4.867658547 | 0.687320106 | FALSE | TRUE | FALSE |
| KCNH7 | 1.506E-10 | 0.89549747 | 0.1538 | 1.50623E-10 | 6.01609E-10 | 2.94667E-09 | 4.4627E-11 | 0.902123047 | 0.02885 | 4.46265E-11 | 4.46265E-11 | 1.50362E-09 | 3.375188464 | 0.528298028 | FALSE | TRUE | FALSE |
| ABCA2 | 1.074E-10 | 0.89550418 | 0.42465 | 1.0737E-10 | 3.92907E-09 | 1.84292E-08 | 1.8586E-09 | 0.901971895 | 0.31385 | 4.65101E-10 | 2.70789E-08 | 1.18778E-07 | 0.057769836 | -1.238298866 | FALSE | TRUE | FALSE |
| SETD1A | 1.068E-10 | 0.8955109 | 1 | 2502496017 | 1.30971E+15 | 2.85457E+17 | 1.1128E-09 | 0.902009727 | 1 | 27801.24091 | 691710372 | 36465697404 | 0.095968195 | -1.017872674 | FALSE | TRUE | FALSE |
| PRDM9 | 4.294E-11 | 0.89551762 | 1 | 8.31954E-08 | 4.25624E-06 | 1.15588E-05 | 5.5407E-09 | 0.901896144 | 0.81515 | 3.8841E-08 | 2.7928E-07 | 2.05873E-06 | 0.007750597 | -2.110664832 | FALSE | TRUE | FALSE |
| PRSS3 | 3.988E-11 | 0.89552433 | 1 | 1.227E+32 | 7.0124E+305 | 6.3744E+307 | 7.0259E-12 | 0.90221728 | 1 | 1.1264E+261 | 5.3833E+286 | 4.9745E+296 | 5.676587381 | 0.754087327 | FALSE | TRUE | FALSE |
| NPC1L1 | 3.598E-11 | 0.89553105 | 0.3688 | 9.02163E-12 | 1.43567E-10 | 6.98702E-10 | 1.2764E-11 | 0.902185889 | 0.8939 | 1.10048E-10 | 1.75189E-09 | 6.9925E-09 | 2.819230219 | 0.450130542 | FALSE | TRUE | FALSE |
| STOX2 | 3.279E-11 | 0.89553776 | 0.87945 | 2.50128E-10 | 7.82744E-09 | 7.82744E-09 | 1.3616E-10 | 0.902066419 | 0.75535 | 1.36156E-10 | 5.44355E-10 | 2.6761E-09 | 0.240856304 | -0.618241983 | FALSE | TRUE | FALSE |
| SLC4A11 | 1.742E-11 | 0.89554447 | 0.0287 | 1.74216E-11 | 1.74216E-11 | 3.40301E-10 | 1.0277E-10 | 0.902085302 | 0.0269 | 1.02769E-10 | 1.02769E-10 | 2.01305E-09 | 0.169522818 | -0.770771838 | FALSE | TRUE | FALSE |
| NSD1 | 1.206E-11 | 0.89555118 | 0.53895 | 4.80695E-11 | 2.29482E-10 | 1.5991E-09 | 5.3503E-11 | 0.902110468 | 0.53435 | 4.9895E-10 | 1.02402E-09 | 1.66414E-08 | 0.225416121 | -0.647015028 | FALSE | TRUE | FALSE |
| CACNA1H | 9.379E-12 | 0.89555789 | 0.0518 | 6.73837E-14 | 1.12429E-11 | 1.48845E-10 | 1.2657E-13 | 0.902336382 | 0.73305 | 6.13121E-13 | 1.27416E-10 | 6.15688E-10 | 74.09820917 | 1.869807712 | FALSE | TRUE | FALSE |
| MYOF | 8.492E-12 | 0.8955646 | 0.75895 | 2.67151E-10 | 3.3447E-07 | 8.84228E-06 | 3.9383E-08 | 0.901680879 | 0.2852 | 4.67985E-09 | 1.00709E-05 | 0.000161285 | 0.000215628 | -3.666293986 | FALSE | TRUE | FALSE |
| DNAH10 | 6.575E-12 | 0.89557131 | 0.5623 | 5.04732E-11 | 9.22973E-10 | 7.26681E-09 | 5.8773E-11 | 0.902097887 | 0.3311 | 5.87735E-11 | 1.08593E-09 | 8.55468E-09 | 0.111871666 | -0.951279893 | FALSE | TRUE | FALSE |
| MYO18B | 6.073E-12 | 0.89557802 | 0.07445 | 6.07349E-12 | 2.88137E-11 | 2.02397E-10 | 3.5221E-11 | 0.902141908 | 0.8858 | 1.40477E-10 | 1.15564E-09 | 9.67889E-09 | 0.172438335 | -0.76336618 | FALSE | TRUE | FALSE |
| NTRK3 | 6.027E-12 | 0.89558473 | 0.7555 | 5.65037E-11 | 5.43638E-10 | 1.11396E-09 | 0.05894402 | 0.884894868 | 0.998 | 1751.54704 | 611200.4622 | 6754112.508 | 1.02256E-10 | -9.990309123 | FALSE | TRUE | FALSE |
| ATP7B | 2.585E-12 | 0.89559144 | 0.9842 | 3.88018E-06 | 0.208074095 | 16.4190587 | 4.7875E-09 | 0.901915093 | 0.93285 | 0.000156969 | 20.05489196 | 2650.69913 | 0.000539957 | -3.267640607 | FALSE | TRUE | FALSE |
| SYNE2 | 2.542E-12 | 0.89559814 | 1 | 217002.386 | 4493268425 | 96420829128 | 4.1408E-11 | 0.902129334 | 1 | 279607.9754 | 836420281.2 | 43624546987 | 0.061393406 | -1.211878272 | FALSE | TRUE | FALSE |
| MYO5B | 2.065E-12 | 0.89560485 | 0.2244 | 2.06459E-12 | 3.29614E-11 | 1.58426E-10 | 3.3843E-12 | 0.902261193 | 0.8587 | 2.93413E-11 | 5.66257E-10 | 7.49318E-09 | 0.610058161 | -0.214628759 | FALSE | TRUE | FALSE |
| PKHD1 | 2.046E-12 | 0.89561155 | 0.9708 | 3.68812E-09 | 1.14624E-06 | 1.8036E-05 | 0.0001264 | 0.90014063 | 0.40915 | 3.48771E-05 | 0.042941334 | 1.070796451 | 1.61827E-08 | -7.790949012 | FALSE | TRUE | FALSE |
| ROS1 | 1.948E-12 | 0.89561825 | 0.6759 | 7.78262E-12 | 1.49729E-10 | 1.03918E-09 | 6.9859E-07 | 0.901273101 | 0.18055 | 5.88124E-08 | 6.4024E-06 | 4.46845E-05 | 2.789E-06 | -5.554551547 | FALSE | TRUE | FALSE |
| ATP2A1 | 1.811E-12 | 0.89562496 | 0.99895 | 3.51816E-07 | 0.000335601 | 0.004237677 | 3.9011E-12 | 0.902242378 | 0.98385 | 4.95383E-09 | 8.98902E-07 | 6.97004E-06 | 0.464154264 | -0.333337655 | FALSE | TRUE | FALSE |
| STAB2 | 1.604E-12 | 0.89563166 | 0.7674 | 1.74006E-11 | 1.105E-09 | 9.17807E-09 | 1.5902E-12 | 0.902280001 | 0.76845 | 1.08198E-11 | 5.40067E-10 | 2.52279E-09 | 1.008856545 | 0.003829416 | FALSE | TRUE | FALSE |
| POP4 | 1.287E-12 | 0.89563836 | 0.5016 | 6.44516E-12 | 6.44516E-12 | 6.44516E-12 | 5.6535E-11 | 0.902104178 | 0.74945 | 5.65354E-11 | 3.08131E-10 | 3.08131E-10 | 0.022760125 | -1.642825363 | FALSE | TRUE | FALSE |
| COL11A2 | 1.268E-12 | 0.89564506 | 0.9246 | 2.13945E-10 | 8.41288E-08 | 1.54187E-06 | 1.0655E-11 | 0.902192169 | 0.47205 | 1.0655E-11 | 6.77025E-10 | 2.98853E-09 | 0.119034264 | -0.924328009 | FALSE | TRUE | FALSE |
| ZFHX3 | 9.452E-13 | 0.89565176 | 0.90155 | 1.64411E-11 | 1.2589E-09 | 1.04513E-08 | 3.5574E-15 | 0.902411455 | 1 | 5.20196E-07 | 0.00046221 | 0.015359017 | 265.698736 | 2.424389488 | FALSE | TRUE | FALSE |
| ITGB4 | 7.101E-13 | 0.89565846 | 0.9849 | 3.72403E-10 | 5.0293E-08 | 5.04512E-07 | 5.0073E-10 | 0.902047529 | 0.67995 | 2.26723E-09 | 3.20911E-07 | 2.30542E-06 | 0.001418222 | -2.848255665 | FALSE | TRUE | FALSE |
| CCDC40 | 4.889E-13 | 0.89566515 | 0.9943 | 2.53698E-09 | 2.45991E-07 | 1.17736E-06 | 6.7941E-12 | 0.902223556 | 0.98425 | 1.17676E-08 | 1.34964E-06 | 6.83742E-06 | 0.071965573 | -1.142875214 | FALSE | TRUE | FALSE |
| MUC6 | 4.489E-13 | 0.89567185 | 1 | 2.9308E+268 | 3.1546E+296 | 1.4529E+303 | 7.183E-12 | 0.902204726 | 1 | 6.482E+277 | 1.5846E+301 | 1.7882E+306 | 0.062495395 | -1.204151985 | FALSE | TRUE | FALSE |
| IVL | 3.887E-13 | 0.89567854 | 0.9983 | 1.03623E-08 | 3.7268E-07 | 1.46078E-05 | 2.4431E-12 | 0.902273733 | 0.9974 | 1.50428E-08 | 3.73942E-06 | 2.45614E-05 | 0.159109774 | -0.798303142 | FALSE | TRUE | FALSE |
| KCNH6 | 3.544E-13 | 0.89568524 | 0.7358 | 2.08798E-11 | 2.30454E-08 | 1.1248E-07 | 1.0532E-08 | 0.901826603 | 0.64215 | 8.10133E-08 | 9.22214E-05 | 0.001479132 | 3.36464E-05 | -4.473061523 | FALSE | TRUE | FALSE |
| UNC13B | 2.954E-13 | 0.89569193 | 0.9797 | 2.78703E-08 | 0.000390668 | 0.004467899 | 1.579E-07 | 0.901579251 | 0.42675 | 1.57898E-07 | 0.002340762 | 0.027146096 | 1.87065E-06 | -5.728006607 | FALSE | TRUE | FALSE |
| NPC1 | 2.626E-13 | 0.89569862 | 0.9459 | 9.3171E-12 | 5.95012E-10 | 5.42294E-09 | 1.5178E-11 | 0.902173327 | 0.92355 | 3.13955E-09 | 8.01778E-07 | 7.90323E-06 | 0.017302964 | -1.761879487 | FALSE | TRUE | FALSE |
| VWF | 1.911E-13 | 0.89570532 | 0.2685 | 4.7833E-14 | 7.6404E-13 | 6.36851E-12 | 1.4759E-13 | 0.90233012 | 0.7074 | 5.89333E-13 | 2.07761E-11 | 9.84174E-11 | 1.295134811 | 0.112314977 | FALSE | TRUE | FALSE |
| ABCA13 | 1.639E-13 | 0.89571201 | 1 | 2.44258E+19 | 9.66525E+27 | 6.97398E+31 | 2.0729E-18 | 0.90248017 | 1 | 7.68256E+24 | 5.7444E+34 | 2.39144E+38 | 79087.55912 | 4.898108172 | FALSE | TRUE | FALSE |
| EP400 | 1.315E-13 | 0.8957187 | 1 | 7.11476E+44 | 3.98017E+57 | 3.21068E+62 | 1.6505E-20 | 0.902505134 | 1 | 7.23091E+22 | 1.68003E+32 | 1.15021E+36 | 7969908.07 | 6.901453312 | FALSE | TRUE | FALSE |
| PTCH1 | 1.301E-13 | 0.89572539 | 0.38505 | 3.25882E-14 | 1.09696E-12 | 4.37795E-12 | 3.73E-13 | 0.902311332 | 0.51645 | 6.49013E-13 | 2.37272E-11 | 9.47639E-11 | 0.348681803 | -0.457570717 | FALSE | TRUE | FALSE |
| SCN4A | 1.19E-13 | 0.89573208 | 0.9959 | 1.2529E-06 | 0.026195619 | 1.665565971 | 8.864E-15 | 0.902392697 | 0.99975 | 2.17437E-05 | 0.989925814 | 63.13398031 | 13.42124936 | 1.127792946 | FALSE | TRUE | FALSE |
| ATP2B4 | 1.049E-13 | 0.89573876 | 0.86295 | 2.21979E-12 | 1.4076E-10 | 5.61587E-10 | 1.5029E-14 | 0.902367676 | 1 | 5.99268E-14 | 5.99268E-14 | 1.17177E-12 | 6.978436188 | 0.843758112 | FALSE | TRUE | FALSE |
| DNAH1 | 8.877E-14 | 0.89574545 | 1 | 3.92501E+19 | 3.49112E+27 | 9.73046E+30 | 5.9673E-17 | 0.902473927 | 1 | 9.99667E+35 | 1.99589E+46 | 1.95022E+50 | 1487.644729 | 3.172499228 | FALSE | TRUE | FALSE |
| COL17A1 | 8.269E-14 | 0.89575214 | 0.04385 | 8.26942E-14 | 8.26942E-14 | 2.77741E-12 | 2.8309E-13 | 0.902323858 | 1 | 2.34163E-12 | 2.34163E-12 | 4.55326E-11 | 0.292114678 | -0.53444662 | FALSE | TRUE | FALSE |
| DNAH9 | 7.387E-14 | 0.89575882 | 0.9988 | 1.88322E-09 | 1.71656E-06 | 3.15132E-05 | 1.0632E-08 | 0.901820276 | 0.74755 | 8.35212E-08 | 7.15063E-05 | 0.00116371 | 6.94853E-06 | -5.158106785 | FALSE | TRUE | FALSE |
| CHD6 | 6.896E-14 | 0.89576551 | 0.2956 | 6.89585E-14 | 5.62048E-13 | 5.211E-12 | 3.5872E-13 | 0.902317596 | 0.85105 | 5.70781E-12 | 2.1176E-10 | 1.01246E-09 | 0.192232454 | -0.716173291 | FALSE | TRUE | FALSE |
| KIF26B | 6.57E-14 | 0.89577219 | 0.9406 | 6.08322E-13 | 2.30847E-11 | 1.10396E-10 | 4.9505E-14 | 0.902348902 | 1 | 1.83574E-12 | 1.74968E-11 | 8.41316E-11 | 1.32710031 | 0.122903751 | FALSE | TRUE | FALSE |
| LAMA1 | 4.795E-14 | 0.89577887 | 0.90895 | 5.76201E-12 | 5.91358E-09 | 1.0483E-07 | 7.2288E-12 | 0.902198448 | 0.3027 | 1.80808E-12 | 5.26281E-10 | 3.899E-09 | 0.006633185 | -2.178277915 | FALSE | TRUE | FALSE |
| PHKB | 3.106E-14 | 0.89578555 | 0.12995 | 7.77347E-15 | 1.24101E-13 | 6.08295E-13 | 3.0631E-15 | 0.902423956 | 0.8577 | 4.8892E-14 | 7.81184E-13 | 3.83701E-12 | 10.13875256 | 1.005984524 | FALSE | TRUE | FALSE |
| ATP2A3 | 2.457E-14 | 0.89579223 | 0.81475 | 4.73634E-13 | 3.67387E-11 | 2.74745E-10 | 6.1705E-13 | 0.902292536 | 0.11435 | 1.54526E-13 | 2.46457E-12 | 1.20321E-11 | 0.039824713 | -1.399847347 | FALSE | TRUE | FALSE |
| CFTR | 1.998E-14 | 0.89579891 | 0.9965 | 3.45365E-07 | 0.012013327 | 0.879768919 | 1.5837E-09 | 0.901984509 | 0.82155 | 7.07324E-07 | 0.012859163 | 0.836888333 | 1.26152E-05 | -4.899104368 | FALSE | TRUE | FALSE |
| CACNA1G | 1.311E-14 | 0.89580559 | 0.5334 | 1.31079E-14 | 2.47676E-13 | 1.74532E-12 | 4.1623E-14 | 0.902355161 | 0.2735 | 4.1623E-14 | 6.59959E-13 | 3.15219E-12 | 0.314920566 | -0.501798976 | FALSE | TRUE | FALSE |
| MYO5C | 6.637E-15 | 0.89581227 | 0.8747 | 1.059E-13 | 6.76302E-12 | 2.70462E-11 | 5.1903E-13 | 0.902298802 | 0.51255 | 5.19031E-13 | 1.00659E-11 | 1.32948E-10 | 0.012786396 | -1.893251852 | FALSE | TRUE | FALSE |
| CACNA1C | 6.286E-15 | 0.89581895 | 0.3886 | 6.2857E-15 | 1.19222E-13 | 8.38928E-13 | 2.9295E-15 | 0.902430205 | 0.763 | 1.16862E-14 | 1.86099E-13 | 8.89939E-13 | 2.145629227 | 0.331554676 | FALSE | TRUE | FALSE |
| MYH1 | 4.76E-15 | 0.89582563 | 0.95165 | 4.05089E-11 | 3.89811E-07 | 2.25464E-05 | 4.8419E-12 | 0.902236105 | 0.6326 | 2.23579E-11 | 1.1576E-07 | 4.30525E-06 | 0.000983018 | -3.007438745 | FALSE | TRUE | FALSE |
| COL1A1 | 4.505E-15 | 0.8958323 | 0.8941 | 4.23368E-14 | 6.74197E-13 | 3.25918E-12 | 3.9834E-15 | 0.902405203 | 0.28835 | 3.98344E-15 | 1.59005E-14 | 3.08083E-13 | 1.130969927 | 0.053451057 | FALSE | TRUE | FALSE |
| MYH10 | 4.204E-15 | 0.89583898 | 0.647 | 1.6759E-14 | 1.53009E-13 | 1.28379E-12 | 1.0871E-14 | 0.902380188 | 0.94325 | 1.7288E-13 | 2.75213E-12 | 1.32653E-11 | 0.386706134 | -0.412618939 | FALSE | TRUE | FALSE |
| PLEKHG6 | 4.081E-15 | 0.89584565 | 0.93925 | 1.6979E-13 | 7.81657E-12 | 7.81657E-12 | 2.4836E-14 | 0.902361419 | 0.7571 | 1.6465E-13 | 1.12026E-12 | 3.24456E-12 | 0.164328361 | -0.784287475 | FALSE | TRUE | FALSE |
| LILRA2 | 2.567E-15 | 0.89585232 | 1 | 0.254948502 | 0.254948502 | 5.048143535 | 1.2935E-05 | 0.90068096 | 1 | 8.9088E+282 | 2.7686E+304 | 1.2799E+306 | 1.98418E-10 | -9.702419572 | FALSE | TRUE | FALSE |
| NEB | 2.003E-15 | 0.895859 | 0.8423 | 3.02754E-14 | 4.32851E-12 | 3.57617E-11 | 4.6867E-15 | 0.902398951 | 0.9813 | 1.18727E-12 | 1.18982E-08 | 1.00826E-07 | 0.427468996 | -0.369095379 | FALSE | TRUE | FALSE |
| DGKH | 1.735E-15 | 0.89586567 | 0.62995 | 1.13574E-14 | 4.11652E-12 | 1.56424E-11 | 4.5592E-11 | 0.902116758 | 0.0883 | 8.36513E-12 | 1.82281E-10 | 3.84313E-10 | 3.80543E-05 | -4.419595836 | FALSE | TRUE | FALSE |
| KMT2A | 1.466E-15 | 0.89587234 | 0.7798 | 1.36835E-14 | 2.5456E-13 | 4.31507E-12 | 4.9955E-14 | 0.902342642 | 0.54665 | 4.99548E-14 | 4.14022E-13 | 1.65088E-12 | 0.029349477 | -1.532399636 | FALSE | TRUE | FALSE |
| MYO7A | 5.959E-16 | 0.89587901 | 0.2328 | 3.71504E-17 | 3.83582E-14 | 1.25384E-12 | 6.8661E-22 | 0.902511373 | 0.99685 | 1.12371E-17 | 1.15841E-14 | 1.86166E-13 | 867940.5912 | 5.93849 | FALSE | TRUE | FALSE |
| ATM | 5.849E-16 | 0.89588568 | 0.70485 | 5.8491E-16 | 3.70821E-14 | 1.5257E-13 | 1.1722E-15 | 0.902442701 | 0.06365 | 1.17216E-15 | 2.92207E-15 | 3.89948E-14 | 0.499001034 | -0.301898554 | FALSE | TRUE | FALSE |
| FAT1 | 2.497E-16 | 0.89589234 | 0.8948 | 4.59783E-15 | 7.45344E-13 | 5.97128E-12 | 1.0574E-12 | 0.902286269 | 0.36515 | 4.79084E-13 | 6.7373E-11 | 3.10161E-10 | 0.000236177 | -3.626762073 | FALSE | TRUE | FALSE |
| SBF1 | 1.449E-16 | 0.89589901 | 1 | 1.44926E-16 | 2.75024E-15 | 4.86394E-15 | 3.872E-13 | 0.902305067 | 0.3707 | 9.70298E-14 | 1.85271E-12 | 7.39325E-12 | 0.000374294 | -3.426787582 | FALSE | TRUE | FALSE |

| | | | | | | | | | | | | | | | | | |
|---|---|---|---|---|---|---|---|---|---|---|---|---|---|---|---|---|---|
| DNAH5 | 1.245E-16 | 0.89590568 | 0.15605 | 1.07807E-17 | 7.88097E-16 | 6.47642E-15 | 6.8036E-09 | 0.901870868 | 0.24675 | 4.26524E-10 | 4.23413E-07 | 7.22281E-06 | 1.82933E-08 | -7.737707588 | FALSE | TRUE | FALSE |
| NXNL1 | 7.031E-17 | 0.89591234 | 1 | 1.71375E-07 | 0.000159385 | 0.002749836 | 2.0311E-15 | 0.902436453 | 1 | 0.000784459 | 7.020077006 | 164.7971599 | 0.034615781 | -1.460725862 | FALSE | TRUE | FALSE |
| KMT2E | 5.404E-17 | 0.89591901 | 1 | 4.80724E-16 | 4.3894E-15 | 8.49598E-14 | 3.1655E-16 | 0.902461439 | 1 | 2.83947E-15 | 2.83947E-15 | 5.51734E-14 | 0.170705967 | -0.767751297 | FALSE | TRUE | FALSE |
| CDH23 | 4.093E-17 | 0.89592567 | 0.8624 | 7.51301E-16 | 5.05589E-14 | 4.68258E-13 | 5.746E-10 | 0.902041231 | 0.1561 | 1.6597E-11 | 4.2316E-09 | 4.25641E-08 | 7.12266E-08 | -7.147357973 | FALSE | TRUE | FALSE |
| FBN3 | 3.886E-17 | 0.89593233 | 0.2445 | 3.88595E-17 | 1.29396E-15 | 5.87509E-15 | 9.613E-15 | 0.902386443 | 0.9838 | 5.67398E-11 | 4.82924E-08 | 4.77345E-07 | 0.004042395 | -2.393361209 | FALSE | TRUE | FALSE |
| HMCN1 | 3.533E-17 | 0.895939 | 0.01385 | 6.80162E-20 | 4.91557E-18 | 3.7545E-17 | 8.427E-17 | 0.902467684 | 0.25025 | 8.91851E-18 | 2.46078E-15 | 2.13777E-14 | 0.41920913 | -0.377569267 | FALSE | TRUE | FALSE |
| RYR3 | 2.715E-17 | 0.89594566 | 1 | 2.71527E-17 | 4.93696E-16 | 8.90267E-16 | 3.0953E-15 | 0.902417706 | 1 | 3.09529E-15 | 5.69005E-14 | 1.01923E-13 | 0.008772261 | -2.056888456 | FALSE | TRUE | FALSE |
| CACNA1B | 5.42E-18 | 0.89595232 | 0.0493 | 3.40088E-19 | 5.41968E-18 | 4.54656E-17 | 4.0682E-19 | 0.902492654 | 0.8918 | 2.58818E-17 | 4.96645E-16 | 6.59634E-15 | 13.32207162 | 1.124571764 | FALSE | TRUE | FALSE |
| RYR2 | 1.453E-18 | 0.89595898 | 0.92655 | 5.01655E-17 | 1.26106E-14 | 1.12326E-13 | 3.5556E-16 | 0.902455194 | 0.8016 | 5.6377E-15 | 1.42158E-12 | 1.36031E-11 | 0.00408555 | -2.38874945 | FALSE | TRUE | FALSE |
| MYH4 | 1.25E-18 | 0.89596564 | 0.95195 | 1.26331E-15 | 2.50521E-12 | 4.00179E-11 | 2.3512E-20 | 0.902498894 | 0.9444 | 3.27881E-17 | 9.23328E-14 | 2.82402E-12 | 53.17928544 | 1.725742498 | FALSE | TRUE | FALSE |
| COL6A3 | 7.841E-19 | 0.89597229 | 0.6133 | 3.12024E-18 | 1.01757E-15 | 9.55402E-15 | 1.5828E-18 | 0.902486412 | 0.76455 | 1.39671E-17 | 3.31613E-15 | 3.76154E-14 | 0.495402145 | -0.305042117 | FALSE | TRUE | FALSE |
| DYSF | 3.865E-19 | 0.89597895 | 0.0321 | 1.53274E-21 | 1.86065E-19 | 1.84628E-18 | 2.604E-23 | 0.902523848 | 0.87925 | 4.12679E-22 | 5.25551E-20 | 2.19688E-19 | 14840.72353 | 4.171455075 | FALSE | TRUE | FALSE |
| PKHD1L1 | 3.407E-20 | 0.89598561 | 0.97835 | 3.50655E-15 | 5.64788E-11 | 2.08735E-09 | 7.1105E-12 | 0.902211003 | 0.03565 | 2.84916E-16 | 2.07323E-12 | 1.17791E-10 | 4.79101E-09 | -8.319572645 | FALSE | TRUE | FALSE |
| TENM4 | 3.313E-23 | 0.89599226 | 0.75155 | 3.05588E-22 | 4.6104E-20 | 4.47641E-19 | 5.1295E-24 | 0.902542555 | 0.78675 | 2.04686E-23 | 3.26173E-22 | 1.56669E-21 | 6.459148681 | 0.810175282 | FALSE | TRUE | FALSE |
| MYH13 | 1.679E-23 | 0.89599892 | 0.83905 | 5.63282E-22 | 5.65417E-19 | 4.89194E-18 | 2.4827E-23 | 0.902530084 | 0.8928 | 3.21898E-21 | 9.23086E-19 | 1.46953E-17 | 0.676300129 | -0.16986053 | FALSE | TRUE | FALSE |
| COL7A1 | 1.285E-23 | 0.89600557 | 0.051 | 1.611E-24 | 2.9965E-23 | 2.38962E-22 | 2.2741E-25 | 0.902555022 | 1 | 1.76672E-24 | 7.57496E-24 | 5.885E-23 | 56.49471679 | 1.752007836 | FALSE | TRUE | FALSE |
| TENM2 | 2.421E-24 | 0.89601222 | 0.5656 | 3.32923E-24 | 3.55884E-22 | 3.36889E-21 | 1.7205E-23 | 0.90253632 | 0.1526 | 2.50663E-24 | 1.44104E-22 | 6.36352E-22 | 0.140733877 | -0.851601346 | FALSE | TRUE | FALSE |
| LRP5 | 3.038E-25 | 0.89601887 | 0.41125 | 3.03778E-25 | 1.91622E-23 | 9.15725E-23 | 2.6037E-22 | 0.902517611 | 0.4186 | 2.60366E-22 | 1.64816E-20 | 2.62111E-19 | 0.001166734 | -2.933028293 | FALSE | TRUE | FALSE |
| ATXN3 | 3.077E-26 | 0.89602553 | 1 | 2.25767E-23 | 7.08416E-22 | 7.64953E-22 | 5.7941E-25 | 0.902548789 | 0.99705 | 1.01145E-22 | 3.56751E-21 | 2.19366E-20 | 0.053108159 | -1.274838752 | FALSE | TRUE | FALSE |
| ITPR3 | 1.79E-30 | 0.89603218 | 0.9997 | 7.08631E-25 | 6.14053E-22 | 1.14658E-20 | 5.0585E-29 | 0.902561255 | 1 | 2.01558E-21 | 2.08583E-18 | 2.20038E-17 | 0.035387599 | -1.451148905 | FALSE | TRUE | FALSE |
| PTPRS | 9.061E-34 | 0.89603883 | 0.99995 | 6.51378E-29 | 1.3598E-26 | 2.57169E-25 | 9.1767E-32 | 0.902567487 | 1 | 6.41799E-26 | 1.77507E-23 | 4.5417E-22 | 0.009873893 | -2.005511567 | FALSE | TRUE | FALSE |
| LRP1B | 6.486E-39 | 0.89604547 | 0.951 | 2.55098E-35 | 5.41512E-32 | 9.97904E-31 | 3.0994E-37 | 0.902573717 | 0.49885 | 3.09943E-37 | 3.0967E-34 | 4.91487E-33 | 0.020926444 | -1.679304563 | FALSE | TRUE | FALSE |
| PLEC | 1.926E-41 | 0.89605212 | 0.9989 | 9.28841E-35 | 1.00788E-30 | 4.29186E-29 | 5.798E-42 | 0.902579948 | 1 | 1.03896E-31 | 1.57032E-27 | 9.77845E-26 | 3.322169699 | 0.521421813 | FALSE | TRUE | FALSE |
| LRP2 | 1.063E-46 | 0.89605877 | 1 | 1.06291E-46 | 1.94147E-45 | 3.4822E-45 | 1.2839E-45 | 0.902586177 | 1 | 1.28389E-45 | 2.36979E-44 | 4.22464E-44 | 0.082788435 | -1.082030328 | FALSE | TRUE | FALSE |
| RYR1 | 5.202E-54 | 0.89606541 | 0.9853 | 2.69792E-48 | 4.45045E-44 | 2.76905E-42 | 9.8742E-51 | 0.902592405 | 0.7105 | 3.38359E-49 | 2.78001E-45 | 9.16047E-44 | 0.000526859 | -3.278305557 | FALSE | TRUE | FALSE |
| TTN | 6.684E-65 | 0.89607206 | 0.88465 | 3.14523E-62 | 1.38999E-58 | 9.59081E-57 | 3.8489E-63 | 0.902598633 | 0.79355 | 9.28174E-62 | 1.64272E-58 | 4.55142E-57 | 0.017364799 | -1.760330249 | FALSE | TRUE | FALSE |
