## Supplementary Table 2 for "Comparing the Phenotypic and Genetic Differences Between Autistic Regression and Autism Without Regression Using the SPARK Cohort"

| **Supplementary Table 2.** Autreg-specific candidates with an empirical Autreg P-value <0.05. For each gene, gene-specific mutation rates are provided for loss-of-function (LoF; mut.lof) and damaging missense (misD; mut.dmis) variants. Variant counts are shown separately for Autreg and Autnreg cohorts, including de novo (dn), transmitted (trans), and non-transmitted (ntrans) LoF and misD variants. Autreg and Autnreg Bayes factors (BFs), Bayesian false discovery rates (q-values), empirical P-values, and corresponding empirical null-distribution statistics (median, 95th percentile [q95], and 99th percentile [q99]) are also shown. The Autreg/Autnreg BF ratio and its log10 transformation quantify the relative strength of genetic association in Autreg compared with Autnreg. | | | | | | | | | | | | | | | | | | | | | | | | | | | | |
| --- | --- | --- | --- | --- | --- | --- | --- | --- | --- | --- | --- | --- | --- | --- | --- | --- | --- | --- | --- | --- | --- | --- | --- | --- | --- | --- | --- | --- |
| Gene | mut.lof | mut.dmis | Autreg_dn.lof | Autreg_trans.lof | Autreg_ntrans.lof | Autreg_dn.dmis | Autreg_trans.dmis | Autreg_ntrans.dmis | Autnreg_dn.lof | Autnreg_trans.lof | Autnreg_ntrans.lof | Autnreg_dn.dmis | Autnreg_trans.dmis | Autnreg_ntrans.dmis | Autreg_BF | Autreg_qval | Autreg_pval | Autreg_null_median | Autreg_null_q95 | Autreg_null_q99 | Autnreg_BF | Autnreg_qval | Autnreg_pval | Autnreg_null_median | Autnreg_null_q95 | Autnreg_null_q99 | Autnreg_Control_BF_ratio | Autreg_Autnreg_log10BF |
| ANGPTL3 | 1.68826830345713e-06 | 3.26027432246058e-06 | 0 | 6 | 0 | 1 | 1 | 0 | 0 | 1 | 4 | 0 | 0 | 2 | 2376300.86 | 7.95635661087037e-07 | 2.5e-05 | 31.9334659 | 11942.0718 | 29922.8135 | 0.5566129 | 0.73013318 | 0.9783 | 81.6338138 | 7397.29886 | 26010.6626 | 4269216.32 | 6.63034816 |
| AUTS2 | 4.15978707847664e-06 | 1.44068345906197e-05 | 2 | 0 | 0 | 0 | 0 | 0 | 0 | 0 | 0 | 0 | 0 | 0 | 131.289317 | 0.02047002 | 2.5e-05 | 0.11236869 | 0.11236869 | 2.17790573 | 0.13018902 | 0.86891682 | 1 | 0.13018902 | 0.13018902 | 2.53270136 | 1008.45154 | 3.00365503 |
| CCDC34 | 7.88060499063327e-07 | 2.66066283957106e-06 | 1 | 4 | 2 | 0 | 0 | 0 | 0 | 1 | 3 | 0 | 0 | 0 | 15763.2491 | 0.00013842 | 0.0175 | 48.739398 | 4533.80317 | 45315.8489 | 0.84544575 | 0.60863653 | 0.94005 | 7.73861963 | 699.201957 | 699.201957 | 18644.8971 | 4.27055999 |
| CTTNBP2NL | 2.15862376758603e-06 | 5.14356590224113e-06 | 2 | 0 | 0 | 0 | 0 | 0 | 0 | 0 | 0 | 0 | 0 | 0 | 501.946877 | 0.00529094 | 2.5e-05 | 0.4258455 | 0.4258455 | 8.42101271 | 0.47091435 | 0.76674956 | 1 | 0.47091435 | 0.47091435 | 9.32485502 | 1065.89846 | 3.02771583 |
| GTPBP2 | 2.33059082669932e-06 | 6.58526416498377e-06 | 1 | 0 | 0 | 0 | 1 | 1 | 0 | 0 | 0 | 0 | 0 | 0 | 34.0143368 | 0.07423069 | 0.00695 | 1.00558378 | 4.02301675 | 19.8226655 | 0.30251145 | 0.82461576 | 1 | 0.30251145 | 0.30251145 | 5.97359916 | 112.439832 | 2.05092019 |
| IREB2 | 3.64684076875944e-06 | 7.78515022026159e-06 | 1 | 0 | 0 | 0 | 1 | 0 | 0 | 0 | 0 | 0 | 0 | 0 | 30.1451948 | 0.08579153 | 0.00405 | 0.89378207 | 0.89378207 | 17.5727517 | 0.26982468 | 0.83382688 | 1 | 0.26982468 | 0.26982468 | 5.31587588 | 111.721413 | 2.04813642 |
| ITGB1 | 2.60530619168175e-06 | 6.56498954824448e-06 | 0 | 2 | 0 | 1 | 0 | 0 | 0 | 1 | 0 | 0 | 0 | 0 | 29.2791514 | 0.08770339 | 0.00495 | 0.15721026 | 1.48523603 | 3.0991593 | 0.26309165 | 0.83579448 | 0.50795 | 0.26309165 | 0.26309165 | 0.96253279 | 111.288791 | 2.04645142 |
| PSAP | 1.57428897948807e-06 | 5.03976580266397e-06 | 1 | 0 | 0 | 0 | 3 | 1 | 0 | 0 | 0 | 0 | 1 | 1 | 98.9535856 | 0.02703561 | 0.00205 | 0.73028264 | 11.6827463 | 11.6827463 | 0.2690167 | 0.83408176 | 0.7546 | 0.2690167 | 1.07567864 | 2.28104368 | 367.834358 | 2.56565229 |
| PTEN | 1.49213905988425e-06 | 3.24488035069535e-06 | 1 | 0 | 0 | 2 | 0 | 0 | 0 | 0 | 1 | 0 | 0 | 1 | 3340.45941 | 0.00071267 | 2.5e-05 | 0.23808286 | 0.23808286 | 4.72769276 | 0.27026392 | 0.83370768 | 0.7512 | 2.52799447 | 10.1101088 | 10.1101088 | 12359.9902 | 4.09201813 |
| RAI1 | 6.24749741881723e-06 | 2.15600641038189e-05 | 2 | 1 | 0 | 0 | 0 | 0 | 0 | 0 | 2 | 0 | 1 | 1 | 322.899157 | 0.00892117 | 5e-05 | 0.27890218 | 0.56463431 | 5.32391202 | 0.16990295 | 0.85971073 | 0.9421 | 1.60192575 | 61.9853631 | 297.529842 | 1900.49176 | 3.27886599 |
| SRSF11 | 2.07572554997359e-06 | 5.02169504490295e-06 | 3 | 0 | 0 | 0 | 0 | 0 | 0 | 0 | 0 | 0 | 0 | 0 | 10436.4494 | 0.00019779 | 2.5e-05 | 0.24698877 | 0.24698877 | 4.88545862 | 0.29151035 | 0.82759594 | 1 | 0.29151035 | 0.29151035 | 5.77372691 | 35801.2997 | 4.55389879 |
| ZNF143 | 2.02030779943832e-06 | 5.55301240392584e-06 | 1 | 1 | 0 | 0 | 0 | 0 | 0 | 0 | 0 | 0 | 0 | 0 | 61.4493566 | 0.03954328 | 5e-05 | 1.81541815 | 1.81541815 | 6.67956349 | 0.24595812 | 0.84032097 | 1 | 0.24595812 | 0.24595812 | 4.86652004 | 249.836661 | 2.39765617 |
| ACY1 | 1.16050536156581e-06 | 4.54596708658721e-06 | 0 | 2 | 0 | 0 | 0 | 0 | 0 | 0 | 0 | 0 | 0 | 0 | 58.9559297 | 0.04126888 | 0.01055 | 6.06709825 | 58.9559297 | 120.132855 | 0.71939264 | 0.65615531 | 1 | 0.71939264 | 0.71939264 | 14.2615589 | 81.9523672 | 1.9135615 |
| ALDH1L2 | 2.98693909878168e-06 | 7.5948498987517e-06 | 0 | 4 | 0 | 0 | 5 | 3 | 0 | 1 | 3 | 0 | 1 | 5 | 11129.6761 | 0.00017298 | 0.0211 | 32.3439325 | 4742.30097 | 44532.035 | 0.1230587 | 0.87058758 | 0.95235 | 17.7411595 | 2626.08574 | 10508.7427 | 90442.0104 | 4.95637021 |
| ALG10 | 1.55233059827745e-06 | 3.7126434291159e-06 | 0 | 6 | 0 | 0 | 0 | 0 | 0 | 0 | 0 | 0 | 0 | 0 | 67785.9089 | 3.05401995493725e-05 | 0.01795 | 68.3262584 | 6636.91338 | 67785.9089 | 0.13714087 | 0.86734844 | 1 | 0.13714087 | 0.13714087 | 2.72312308 | 494279.426 | 5.69397253 |
| ALG14 | 9.47057136779498e-07 | 2.31158711593866e-06 | 0 | 3 | 0 | 0 | 1 | 1 | 0 | 0 | 0 | 0 | 0 | 0 | 971.910796 | 0.00267033 | 0.03365 | 41.1822175 | 971.910796 | 3887.8871 | 0.36012565 | 0.80562135 | 1 | 0.36012565 | 0.36012565 | 0.36012565 | 2698.81023 | 3.43117235 |
| ANGEL2 | 2.1968333588414e-06 | 4.40476010079303e-06 | 0 | 4 | 1 | 0 | 0 | 0 | 0 | 0 | 1 | 0 | 0 | 0 | 212.322961 | 0.01322849 | 0.03605 | 21.6926866 | 212.322961 | 2128.94982 | 0.04638302 | 0.88745442 | 1 | 0.42442667 | 0.42442667 | 0.9197684 | 4577.60083 | 3.66063792 |
| C4orf17 | 1.01100515689972e-06 | 2.6381243087298e-06 | 0 | 3 | 0 | 0 | 0 | 0 | 0 | 0 | 0 | 0 | 0 | 0 | 135.096417 | 0.01982546 | 0.00345 | 1.52025377 | 135.096417 | 135.096417 | 0.2256408 | 0.84593881 | 1 | 0.2256408 | 0.2256408 | 0.2256408 | 598.723356 | 2.7772262 |
| CHRNA6 | 1.45702753172138e-06 | 4.13593865349136e-06 | 0 | 4 | 1 | 0 | 0 | 0 | 0 | 0 | 0 | 0 | 1 | 1 | 83.8455035 | 0.02967198 | 0.0361 | 8.56900485 | 83.8455035 | 840.454351 | 0.08247366 | 0.87968789 | 0.75345 | 0.08247366 | 0.32971855 | 0.4093933 | 1016.63368 | 3.00716449 |
| CNGA4 | 2.09550065992676e-06 | 6.72779739736111e-06 | 0 | 2 | 1 | 0 | 5 | 0 | 0 | 1 | 0 | 0 | 1 | 5 | 36.019097 | 0.07000341 | 0.02865 | 0.41613902 | 21.8142985 | 87.1435446 | 0.03987659 | 0.88898263 | 0.8259 | 0.27525263 | 10.1599234 | 21.7176366 | 903.264295 | 2.95581484 |
| DEFB134 | 2.03724991264681e-07 | 4.45189073238577e-07 | 0 | 2 | 0 | 0 | 0 | 0 | 0 | 0 | 0 | 0 | 0 | 0 | 39.8078697 | 0.06249693 | 0.00115 | 4.16844339 | 39.8078697 | 39.8078697 | 0.52357395 | 0.74522452 | 1 | 0.52357395 | 0.52357395 | 0.52357395 | 76.0310356 | 1.88099091 |
| ENTPD1 | 1.56402789994127e-06 | 3.89356817686991e-06 | 0 | 5 | 0 | 0 | 0 | 0 | 0 | 0 | 0 | 0 | 0 | 0 | 25123.2914 | 7.61887412269232e-05 | 0.0027 | 246.231957 | 2457.37812 | 25123.2914 | 0.38902143 | 0.7960288 | 1 | 0.38902143 | 0.38902143 | 7.72186074 | 64580.7397 | 4.81010301 |
| ENTPD4 | 3.15190034468799e-06 | 6.10017739543368e-06 | 0 | 5 | 0 | 0 | 0 | 0 | 0 | 0 | 5 | 0 | 0 | 0 | 2513.81399 | 0.00096845 | 0.00365 | 27.2568822 | 258.616599 | 2513.81399 | 0.05408136 | 0.88587929 | 0.96855 | 40.7613287 | 390.102991 | 3824.72652 | 46482.0786 | 4.66728554 |
| GOT1L1 | 1.36533109197872e-06 | 3.33267223860317e-06 | 0 | 5 | 1 | 0 | 1 | 1 | 0 | 0 | 1 | 0 | 1 | 0 | 8822.05664 | 0.00025319 | 0.0416 | 91.0381046 | 8822.05664 | 89996.9564 | 0.20469376 | 0.85086881 | 0.7535 | 0.46622693 | 1.86410049 | 1.86410049 | 43098.8057 | 4.63446524 |
| GPR25 | 1.23120517733288e-06 | 5.17192587066443e-06 | 0 | 2 | 0 | 0 | 0 | 0 | 0 | 0 | 0 | 0 | 0 | 0 | 43.0379179 | 0.05848822 | 0.0112 | 4.46068337 | 43.0379179 | 88.2036785 | 0.55405665 | 0.73155816 | 1 | 0.55405665 | 0.55405665 | 10.9706051 | 77.6778292 | 1.89029708 |
| GRAMD1B | 2.65423158556662e-06 | 7.96319394253855e-06 | 0 | 2 | 0 | 0 | 0 | 0 | 0 | 0 | 1 | 0 | 0 | 0 | 32.5978625 | 0.07904387 | 0.0181 | 3.3817421 | 32.5978625 | 66.4630643 | 0.41928013 | 0.78474796 | 1 | 3.94668815 | 3.94668815 | 77.7280225 | 77.747215 | 1.89068484 |
| GSTA5 | 6.43271049333658e-07 | 1.7442828202925e-06 | 0 | 7 | 2 | 0 | 0 | 0 | 0 | 0 | 1 | 0 | 0 | 0 | 100643.272 | 2.33508980146243e-05 | 0.02195 | 955.4055 | 100643.272 | 1068737.28 | 0.02078594 | 0.89363037 | 1 | 0.18886886 | 0.18886886 | 0.18886886 | 4841891.28 | 6.68501503 |
| IMPACT | 1.0586177578132e-06 | 2.65525026979969e-06 | 0 | 2 | 0 | 0 | 1 | 0 | 0 | 0 | 0 | 0 | 0 | 0 | 73.7868123 | 0.03319626 | 0.0034 | 1.94701193 | 73.7868123 | 73.7868123 | 0.27214585 | 0.83312625 | 1 | 0.27214585 | 0.27214585 | 0.27214585 | 271.129666 | 2.43317704 |
| L3MBTL1 | 2.6503007830862e-06 | 6.75722531289427e-06 | 0 | 3 | 0 | 0 | 3 | 0 | 0 | 0 | 4 | 0 | 1 | 1 | 1044.44813 | 0.00241252 | 0.0212 | 6.9633407 | 261.080243 | 1044.44813 | 0.13113868 | 0.86862541 | 0.9841 | 10.8106482 | 986.401275 | 3945.29001 | 7964.45514 | 3.90115607 |
| LAYN | 1.34007039320562e-06 | 3.01613620107524e-06 | 0 | 3 | 0 | 0 | 0 | 0 | 0 | 0 | 1 | 0 | 0 | 0 | 123.51271 | 0.02205681 | 0.00555 | 12.9269183 | 123.51271 | 123.51271 | 0.20188523 | 0.85167691 | 1 | 1.84512318 | 1.84512318 | 1.84512318 | 611.796664 | 2.7866071 |
| LCE3D | 3.7489646603341e-07 | 9.42121650441243e-07 | 0 | 2 | 0 | 0 | 0 | 0 | 0 | 0 | 0 | 0 | 0 | 0 | 43.5740256 | 0.05725693 | 0.0019 | 4.5538084 | 43.5740256 | 43.5740256 | 0.56519124 | 0.72642856 | 1 | 0.56519124 | 0.56519124 | 0.56519124 | 77.0960739 | 1.88703226 |
| MCL1 | 1.29097327499404e-06 | 4.73794856121348e-06 | 0 | 2 | 0 | 0 | 0 | 0 | 0 | 0 | 0 | 0 | 0 | 0 | 42.8462898 | 0.05898059 | 0.01045 | 4.45228249 | 42.8462898 | 88.1212736 | 0.55202318 | 0.73260512 | 1 | 0.55202318 | 0.55202318 | 10.9394973 | 77.6168306 | 1.8899559 |
| MED15 | 3.11550191613295e-06 | 8.33008738952378e-06 | 0 | 4 | 0 | 0 | 0 | 0 | 0 | 0 | 0 | 0 | 0 | 0 | 2214.51221 | 0.00118007 | 0.0074 | 22.3861844 | 2214.51221 | 2214.51221 | 0.33417413 | 0.81425109 | 1 | 0.33417413 | 0.33417413 | 6.57676142 | 6626.8212 | 3.82130525 |
| NMI | 1.0421959754251e-06 | 2.46665631154692e-06 | 0 | 3 | 0 | 0 | 0 | 0 | 0 | 0 | 1 | 0 | 0 | 0 | 90.9541821 | 0.0286482 | 0.00385 | 1.04411439 | 90.9541821 | 90.9541821 | 0.15809908 | 0.86253225 | 1 | 1.43114679 | 1.43114679 | 1.43114679 | 575.298612 | 2.75989333 |
| OR10H4 | 7.68869407503178e-07 | 2.72714101581512e-06 | 0 | 3 | 0 | 0 | 0 | 0 | 0 | 0 | 0 | 0 | 0 | 0 | 34.3028981 | 0.07369935 | 0.0037 | 3.53775347 | 34.3028981 | 34.3028981 | 0.05148458 | 0.88631202 | 1 | 0.05148458 | 0.05148458 | 0.05148458 | 666.275129 | 2.8236536 |
| OR2B11 | 9.99977881421091e-07 | 3.24698948316146e-06 | 0 | 3 | 0 | 0 | 0 | 0 | 0 | 0 | 1 | 0 | 0 | 0 | 223.332043 | 0.0125007 | 0.0049 | 22.8292311 | 223.332043 | 223.332043 | 0.32138176 | 0.81833142 | 1 | 2.99959975 | 2.99959975 | 2.99959975 | 694.91201 | 2.84192982 |
| OR4F5 | 7.95069846545341e-07 | 2.49267173937533e-06 | 0 | 2 | 0 | 0 | 0 | 0 | 0 | 0 | 0 | 0 | 0 | 0 | 71.1096591 | 0.03416093 | 0.0054 | 7.30951338 | 71.1096591 | 71.1096591 | 0.45152902 | 0.77392752 | 1 | 0.45152902 | 0.45152902 | 0.45152902 | 157.486353 | 2.19724293 |
| PDCL2 | 6.58433739164008e-07 | 1.41309079070735e-06 | 0 | 2 | 0 | 0 | 0 | 0 | 0 | 0 | 0 | 0 | 0 | 0 | 50.8821477 | 0.04712595 | 0.00315 | 5.28679367 | 50.8821477 | 50.8821477 | 0.64201389 | 0.69345135 | 1 | 0.64201389 | 0.64201389 | 0.64201389 | 79.2539674 | 1.89902101 |
| PPP1R12B | 3.97891763016181e-06 | 9.40154874779639e-06 | 0 | 10 | 3 | 0 | 0 | 0 | 0 | 1 | 7 | 0 | 0 | 0 | 61258323.6 | 2.93351337278609e-08 | 0.01495 | 52196.8407 | 18154181.4 | 674463881 | 0.14433006 | 0.86581245 | 0.96705 | 115.948812 | 11318.3098 | 115790.238 | 424432179 | 8.6278083 |
| PTPN13 | 8.83265169020113e-06 | 1.90928614323912e-05 | 0 | 4 | 1 | 0 | 6 | 9 | 0 | 1 | 3 | 0 | 7 | 11 | 8383.75173 | 0.00027271 | 0.0424 | 27.0044734 | 8383.75173 | 78149.1479 | 0.22333075 | 0.84655209 | 0.98175 | 70.6175761 | 18188.902 | 292349.995 | 37539.6211 | 4.57448988 |
| RARA | 1.52673390612171e-06 | 5.25487007072346e-06 | 0 | 1 | 0 | 0 | 5 | 1 | 0 | 1 | 0 | 0 | 0 | 1 | 415.239474 | 0.0064832 | 0.01335 | 11.1339542 | 415.239474 | 880.727065 | 0.62324508 | 0.70196303 | 0.5068 | 0.62324508 | 2.49146452 | 2.49146452 | 666.253911 | 2.82363977 |
| RNH1 | 1.54201033609932e-06 | 5.09613008499202e-06 | 0 | 3 | 0 | 0 | 0 | 0 | 0 | 0 | 0 | 0 | 0 | 0 | 103.330284 | 0.02564202 | 0.0074 | 10.8278194 | 103.330284 | 103.330284 | 0.1783719 | 0.85739767 | 1 | 0.1783719 | 0.1783719 | 3.53237132 | 579.29686 | 2.76290117 |
| SGCZ | 1.35802485230303e-06 | 2.39019838555116e-06 | 0 | 0 | 0 | 0 | 7 | 2 | 0 | 0 | 0 | 0 | 2 | 2 | 403.263413 | 0.00671352 | 0.0229 | 25.1807457 | 403.263413 | 1614.40149 | 0.64503318 | 0.69254137 | 0.6865 | 0.64503318 | 10.3171839 | 10.3171839 | 625.182434 | 2.79600677 |
| SLC14A2 | 2.94017224621376e-06 | 8.65766090004545e-06 | 0 | 5 | 0 | 0 | 0 | 0 | 0 | 0 | 3 | 0 | 1 | 0 | 539.504651 | 0.00485478 | 0.0366 | 6.01551415 | 56.2837806 | 539.504651 | 0.061779 | 0.88437567 | 0.8789 | 1.22057787 | 45.0811009 | 45.0811009 | 8732.81688 | 3.94115435 |
| SLC28A1 | 2.02548756569045e-06 | 6.33413177801397e-06 | 0 | 4 | 0 | 0 | 0 | 0 | 0 | 0 | 0 | 0 | 0 | 0 | 77.8822553 | 0.03167688 | 0.0055 | 0.93786541 | 77.8822553 | 77.8822553 | 0.02299621 | 0.89295733 | 1 | 0.02299621 | 0.02299621 | 0.45431838 | 3386.74337 | 3.52978229 |
| SLC34A1 | 1.82815270032656e-06 | 6.75901850240293e-06 | 0 | 1 | 0 | 0 | 7 | 2 | 0 | 1 | 0 | 0 | 1 | 3 | 397.616254 | 0.00683109 | 0.01755 | 6.20562286 | 397.616254 | 1591.86859 | 0.20495061 | 0.85081371 | 0.63135 | 0.37133124 | 3.27677955 | 13.1071721 | 1940.05888 | 3.28781491 |
| SLCO1B3 | 2.04724750375345e-06 | 5.16721991054701e-06 | 0 | 6 | 0 | 0 | 1 | 0 | 0 | 2 | 2 | 0 | 0 | 0 | 1788.4905 | 0.00146873 | 0.00845 | 2.45967195 | 194.204665 | 447.297518 | 0.17053068 | 0.85947778 | 0.31345 | 0.17053068 | 13.3017763 | 13.3017763 | 10487.7933 | 4.02068412 |
| SURF1 | 1.00483846130382e-06 | 2.48266938846846e-06 | 0 | 4 | 0 | 0 | 5 | 1 | 0 | 0 | 1 | 0 | 1 | 2 | 32560.9926 | 5.67662744757885e-05 | 0.00715 | 23.1531803 | 3430.42566 | 13727.2013 | 0.03849361 | 0.889347 | 0.93725 | 0.15389942 | 5.47426454 | 5.47426454 | 845880.356 | 5.92730894 |
| TFIP11 | 3.08187882431473e-06 | 8.66551875534587e-06 | 0 | 3 | 0 | 0 | 0 | 0 | 0 | 0 | 0 | 0 | 0 | 1 | 28.2923613 | 0.09180158 | 0.0132 | 3.04370926 | 28.2923613 | 59.7282708 | 0.05980058 | 0.88476757 | 1 | 0.23918988 | 0.23918988 | 4.70438548 | 473.111839 | 2.67496382 |
| TRAPPC12 | 2.2735235085009e-06 | 8.4994579616113e-06 | 0 | 1 | 0 | 0 | 6 | 0 | 0 | 0 | 1 | 0 | 0 | 4 | 1791.73162 | 0.00145107 | 0.01345 | 12.4327113 | 447.46355 | 1791.73162 | 0.07050127 | 0.88231722 | 0.9709 | 4.51316679 | 40.8293327 | 163.412079 | 25414.1768 | 4.40507605 |
| ZNF429 | 2.25606184076819e-06 | 5.06969009891304e-06 | 0 | 5 | 0 | 0 | 0 | 0 | 0 | 0 | 4 | 0 | 0 | 0 | 267.425859 | 0.01091394 | 0.03135 | 3.22640594 | 29.0204482 | 267.425859 | 0.0095459 | 0.89686169 | 1 | 0.70149106 | 57.0662109 | 57.0662109 | 28014.7251 | 4.44738636 |
| ZNF446 | 1.66630311277142e-06 | 5.08178182433256e-06 | 0 | 3 | 0 | 0 | 0 | 0 | 0 | 1 | 0 | 0 | 0 | 0 | 32.7845204 | 0.0777079 | 0.0069 | 3.54133593 | 32.7845204 | 32.7845204 | 0.59951895 | 0.71265365 | 0.5104 | 0.59951895 | 0.59951895 | 1.33622679 | 54.6847109 | 1.73786592 |
| ZSCAN32 | 1.78491095998775e-06 | 4.2434132696719e-06 | 0 | 4 | 0 | 0 | 0 | 0 | 0 | 1 | 2 | 0 | 0 | 0 | 356.394441 | 0.00794225 | 0.0037 | 3.9931588 | 356.394441 | 356.394441 | 0.64382696 | 0.69308802 | 0.87485 | 5.90792004 | 55.6054838 | 55.6054838 | 553.556257 | 2.74316176 |
