## Supplementary Table 3 for "Comparing the Phenotypic and Genetic Differences Between Autistic Regression and Autism Without Regression Using the SPARK Cohort"

| **Supplementary Table 3**. Function of the Autreg-specific candidate genes, their presence in Zhou et al., 2022, published ultra-rare de novo variants in the SPARK exomes datasets, and their score in the SFARI GENE database (release 2026 Q2). We selected only genes that harobored LoF or misD de novo variants in the Zhou et al., 2022 dataset. S: Genes with mutations associated with a substantial degree of increased ASD risk and consistently linked to additional characteristics not required for an ASD diagnosis; 1: Genes clearly implicated in ASD; 2: Strong candidate genes with two reported de novo likely-gene-disrupting mutations in autistic individuals or implicated by genome-wide association studies. | | | | |
| --- | --- | --- | --- | --- |
| Gene | Presence in the validation cohort | Score in the SFARI GENE database | Product’s Function | References |
| *ANGPTL3* | absent | absent | An angiopoietin, its dysregulation causes recessive familial hypobetalipoproteinemia. | (Musunuru et al., 2010) |
| *AUTS2* | Autrg:1; Autnreg:1 | 1 | A transcription regulator, its dysregulation causes a neurodevelopmental syndrome. | (Beunders et al., 2013) |
| *CCDC34* | absent | absent | A protein important for flagella formations and its knockout causes infertility. | (Cong et al., 2022) |
| *CTTNBP2NL* | absent | absent | A *CTTNBP2* homologue with low-brain expression. CTTNBP2 but not *CTTNBP2NL* is known to function in dendritic spinogenesis and maintenance. | (Chen et al., 2012) |
| *GTPBP2* | absent | absent | A GTP-binding protein with functions in ribosomal homeostasis and mRNA transcription. Bi-allelic mutations cause a neurodevelopmental syndrome accompanied in some cases with brain iron accumulation. | (Terrey et al., 2020, Jaberi et al., 2016) |
| *IREB2* | Autrg:1; Autnreg:0 | absent | A key RNA-binding protein involved cellular iron balance. Bi-allelic mutations cause an early-onset neurodegenerative syndrome. | (Costain et al., 2019) |
| *ITGB1* | Autrg:0; Autnreg:1 | absent | A cell membrane receptor protein involved in angiogenesis, neural growth, and other developmental processes with suggested links to autism. | (Liao et al., 2026, Su et al., 2024) |
| *PSAP* | absent | absent | A lysosomal protein and precursor of saposins A-D with regulatory roles in cellular and brain homeostasis implicated in multiple neurodegenerative diseases. For example, bi-allelic variants cause metachromatic leukodystrophy. | (Taha et al., 2026, Kretz et al., 1990) |
| *PTEN* | Autrg:6; Autnreg:0 | 1S | A lipid phosphatase that acts as a tumour suppressor gene and regulator of neuronal development. Its dysregulation causes a neurodevelopmental syndrome. | (Butler et al., 2005; Skelton et al., 2020) |
| *RAI1* | Autrg:0; Autnreg:1 | 1S | A transcriptional regulator, its heterozygous loss causes Smith-Magenis syndrome. | (Bi et al., 2005; Slager et al., 2003) |
| *SRSF11* | absent | 2 | A splicing factor involved in splicing neuronal microexons. | (Gehring & Roignant, 2021) |
| *ZNF143* | absent | absent | A transcription factor with key roles in mitochondrial homeostasis. | (Magnitov et al., 2025) |
| *ACY1* | Autrg:1; Autnreg:0 | S | Aminoacylase-1, a zinc-binding metalloenzyme. Bi-allelic mutations cause an inborn error of metabolism that present with neurological features that can include autism. | (Tylki-Szymanska et al., 2010, Sass et al., 2006) |
| *ALDH1L2* | absent | absent | The mitochondrial form of 10-formyltetrahydrofolate dehydrogenase. Bi-allelic mutations cause a neuro-ichthyotic syndrome. | (Sarret et al., 2019) |
| *ALG10* | absent | absent | An endoplasmic reticulum membrane enzyme called alpha-1,2-glucosyltransferase involved in the biosynthesis of the lipid-linked oligosaccharide substrate for N-linked protein glycosylation. | (Burda and Aebi, 1998) |
| *ALG14* | absent | absent | An endoplasmic reticulum membrane UDP-GlcNAc transferase enzyme involved in the biosynthesis of the lipid-linked oligosaccharide substrate for N-linked protein glycosylation. Bi-allelic mutations cause a congenital disorder of glycosylation that include NDD phenotypes. | (Marquez et al., 2026, Kvarnung et al., 2018) |
| *ANGEL2* | absent | absent | A mitochondrial protein involved in non-canonical mitochondrial RNA processing. | (Clemente et al., 2022) |
| *C4orf17* | absent | absent | A protein without an established function but maybe involved in embryonic Wnt/β-catenin signalling pathway. | (Wada et al., 2008) |
| *CHRNA6* | absent | absent | A subunit of neuronal nicotinic acetylcholine receptors that may play a role in nicotine response and addiction. | (Heydary et al., 2025) |
| *CNGA4* | absent | absent | A cyclic nucleotide-gated channel subunit primarily involved in olfactory signal transduction and sensory adaptation. | (Munger et al., 2001) |
| *DEFB134* | absent | absent | No sufficient data on its function. |  |
| *ENTPD1* | absent | absent | A member of the ecto‐nucleoside triphosphate diphosphohydrolases family involved in the breakdown of extracellular ATP to ADP and then to AMP. Bi-alleic mutations cause spastic paraplegia type 64 that can include NDD phenotypes. | (Nardi-Schreiber et al., 2017, Novarino et al., 2014) |
| *ENTPD4* | absent | absent | A member of the ecto‐nucleoside triphosphate diphosphohydrolases family, found in Golgi and lysosomes where it has unknown roles. | (Gorelik et al., 2020) |
| *GOT1L1* | absent | absent | No sufficient data on its function. |  |
| *GPR25* | absent | absent | A G protein-coupled receptor that acts as a key driver in the development and maintenance of tissue-resident memory T cells. | (Feng et al., 2025) |
| *GRAMD1B* | absent | absent | A regulator of lipid homeostasis, autophagic flux, and tau pathology. | (Acosta Ingram et al., 2025) |
| *GSTA5* | absent | absent | Glutathione S-transferase alpha 5. | (Pulford and Hayes, 1996) |
| *IMPACT* | absent | absent | A modulator of the eIF2α kinase general control non-derepressible-2 (Gcn2) important in coping with amino acids starvation. | (Sansom et al., 2025) |
| *L3MBTL1* | absent | absent | A polycomb group epigenetic regulator recently suggested as a risk factor for a subtype of frontotemporal lobar degeneration. | (Pottier et al., 2025) |
| *LAYN* | absent | absent | A C-type lectin transmembrane receptor with important roles in cell adhesion and migration. | (Jin and Zong, 2025) |
| *LCE3D* | absent | absent | A protein important for cutaneous homeostasis and involved in the pathogenesis of atopic dermatitis. | (Li et al., 2026) |
| *MCL1* | absent | absent | An antiapoptotic protein that regulates cell death, cell cycle progression, and mitochondrial homeostasis. | (Sancho et al., 2022) |
| *MED15* | absent | absent | A component of the mediator complex required for basal activity of the RNA polymerase II. | (Cooper and Fassler, 2019) |
| *NMI* | absent | absent | A transcriptional regulator with roles in carcinogenesis. | (Pruitt et al., 2016) |
| *OR10H4* | absent | absent | Olfactory receptor. |  |
| *OR2B11* | absent | absent | Olfactory receptor. |  |
| *OR4F5* | absent | absent | Olfactory receptor. |  |
| *PDCL2* | absent | absent | A member of the phosducin family essential for male fertility. | (Li et al., 2022) |
| *PPP1R12B* | absent | absent | A regulatory subunit of myosin light chain phosphatase. It is also expressed in brain regions and implicated in substance abuse disorder. | (Lee et al., 2024; Liu et al., 2020; Moorhead et al., 1998) |
| *PTPN13* | absent | absent | A non-receptor protein tyrosine phosphatase involved in cell signalling and has roles in cancer and neurodegeneration. | (He et al., 2023) |
| *RARA* | absent | absent | A retinoic acid nuclear receptor that function as a ligand-activated transcription factor. | (Song et al., 2025) |
| *RNH1* | absent | absent | Ribonuclease inhibitor 1 an inflammasome modulator. Bi-allelic mutations are associated with a distinctive subtype of acute, necrotizing encephalopathy that includes psychomotor regression. | (Hedberg-Oldfors et al., 2023, Song et al., 2025) |
| *SGCZ* | absent | absent | Zeta-sarcoglycan, a transmembrane protein that forms part of the dystrophin-glycoprotein complex. | (Ozawa et al., 2005) |
| *SLC14A2* | absent | absent | The UT-A family of urea transporter proteins with roles in hypertension and metabolic syndrome. | (Huang et al., 2025) |
| *SLC28A1* | absent | absent | A concentrative nucleoside transporter-1 that preferentially transports pyrimidine nucleosides. | (Wevers et al., 2019) |
| *SLC34A1* | absent | absent | Renal sodium-phosphate cotransporter 2A involved in phosphate transport. Bi-allelic mutations cause infantile hypercalcemia. | (Schlingmann et al., 2016) |
| *SLCO1B3* | absent | 2 | A liver-specific membrane protein involved in transporting bilirubin, bile acids, and other biomolecules. It has been suggested to be associated with autism. | (De Rubeis et al., 2014, van de Steeg et al., 2012) |
| *SURF1* | absent | absent | An assembly factor of mitochondrial complex IV. Bi-allelic mutations cause Leigh syndrome. | (Zhu et al., 1998) |
| *TFIP11* | absent | absent | A core nuclear factor with roles in pre-mRNA splicing and genomic stability. | (Duchemin et al., 2021) |
| *TRAPPC12* | absent | absent | A subunit of the multi-protein Trafficking Protein Particle (TRAPP) complex essential for intracellular vesicle-mediated transport with roles in cell division. Bi-allelic mutations cause progressive childhood-onset encephalopathy. | (Milev et al., 2017) |
| *ZNF429* | absent | absent | A transcription factor with possible roles on the expression of inflammatory and adhesion molecules. | (Wang et al., 2024) |
| *ZNF446* | absent | absent | A transcription repressor. | (Liu et al., 2005) |
| *ZSCAN32* | absent | absent | No sufficient data on its function. |  |

POTTIER, C., KUCUKALI, F., BAKER, M., BATZLER, A., JENKINS, G. D., VAN BLITTERSWIJK, M., VICENTE, C. T., DE COSTER, W., WYNANTS, S., VAN DE WALLE, P., ROSS, O. A., MURRAY, M. E., FAURA, J., HAGGARTY, S. J., VAN ROOIJ, J. G., MOL, M. O., HSIUNG, G. R., GRAFF, C., OIJERSTEDT, L., NEUMANN, M., ASMANN, Y., MCDONNELL, S. K., BAHETI, S., JOSEPHS, K. A., WHITWELL, J. L., BIENIEK, K. F., FORSBERG, L., HEUER, H., LAGO, A. L., GEIER, E. G., YOKOYAMA, J. S., ODDI, A. P., FLANAGAN, M., MAO, Q., HODGES, J. R., KWOK, J. B., DOMOTO-REILLY, K., SYNOFZIK, M., WILKE, C., ONYIKE, C., DICKERSON, B. C., EVERS, B. M., DUGGER, B. N., MUNOZ, D. G., KEITH, J., ZINMAN, L., ROGAEVA, E., SUH, E., GEFEN, T., GEULA, C., WEINTRAUB, S., DIEHL-SCHMID, J., FARLOW, M. R., EDBAUER, D., WOODRUFF, B. K., CASELLI, R. J., DONKER KAAT, L. L., HUEY, E. D., REIMAN, E. M., MEAD, S., KING, A., ROEBER, S., NANA, A. L., ERTEKIN-TANER, N., KNOPMAN, D. S., PETERSEN, R. C., PETRUCELLI, L., UITTI, R. J., WSZOLEK, Z. K., RAMOS, E. M., GRINBERG, L. T., TEMPINI, M. L. G., ROSEN, H. J., SPINA, S., PIGUET, O., GROSSMAN, M., TROJANOWSKI, J. Q., KEENE, C. D., JIN, L. W., PRUDLO, J., GESCHWIND, D. H., RISSMAN, R. A., CRUCHAGA, C., GHETTI, B., HALLIDAY, G. M., BEACH, T. G., SERRANO, G. E., ARZBERGER, T., HERMS, J., BOXER, A. L., HONIG, L. S., VONSATTEL, J. P., LOPEZ, O. L., KOFLER, J., WHITE, C. L., 3RD, GEARING, M., GLASS, J., ROHRER, J. D., IRWIN, D. J., LEE, E. B., et al. 2025. Deciphering distinct genetic risk factors for FTLD-TDP pathological subtypes via whole-genome sequencing. *Nat Commun,* 16**,** 3914.

PRUITT, H. C., DEVINE, D. J. & SAMANT, R. S. 2016. Roles of N-Myc and STAT interactor in cancer: From initiation to dissemination. *Int J Cancer,* 139**,** 491-500.

PULFORD, D. J. & HAYES, J. D. 1996. Characterization of the rat glutathione S-transferase Yc2 subunit gene, GSTA5: identification of a putative antioxidant-responsive element in the 5'-flanking region of rat GSTA5 that may mediate chemoprotection against aflatoxin B1. *Biochem J,* 318 ( Pt 1)**,** 75-84.

SANCHO, M., LEIVA, D., LUCENDO, E. & ORZAEZ, M. 2022. Understanding MCL1: from cellular function and regulation to pharmacological inhibition. *FEBS J,* 289**,** 6209-6234.

SANSOM, B. J. M., GIBBS, V. J., SCHIEMANN, A. H. & SATTLEGGER, E. 2025. Mutational analysis of Yih1 and IMPACT reveals amino acids required for Gcn2 inhibition. *FEBS Lett,* 599**,** 3488-3500.

SARRET, C., ASHKAVAND, Z., PAULES, E., DORBOZ, I., PEDIADITAKIS, P., SUMNER, S., EYMARD-PIERRE, E., FRANCANNET, C., KRUPENKO, N. I., BOESPFLUG-TANGUY, O. & KRUPENKO, S. A. 2019. Deleterious mutations in ALDH1L2 suggest a novel cause for neuro-ichthyotic syndrome. *NPJ Genom Med,* 4**,** 17.

SASS, J. O., MOHR, V., OLBRICH, H., ENGELKE, U., HORVATH, J., FLIEGAUF, M., LOGES, N. T., SCHWEITZER-KRANTZ, S., MOEBUS, R., WEILER, P., KISPERT, A., SUPERTI-FURGA, A., WEVERS, R. A. & OMRAN, H. 2006. Mutations in ACY1, the gene encoding aminoacylase 1, cause a novel inborn error of metabolism. *Am J Hum Genet,* 78**,** 401-9.

SCHLINGMANN, K. P., RUMINSKA, J., KAUFMANN, M., DURSUN, I., PATTI, M., KRANZ, B., PRONICKA, E., CIARA, E., AKCAY, T., BULUS, D., CORNELISSEN, E. A., GAWLIK, A., SIKORA, P., PATZER, L., GALIANO, M., BOYADZHIEV, V., DUMIC, M., VIVANTE, A., KLETA, R., DEKEL, B., LEVTCHENKO, E., BINDELS, R. J., RUST, S., FORSTER, I. C., HERNANDO, N., JONES, G., WAGNER, C. A. & KONRAD, M. 2016. Autosomal-Recessive Mutations in SLC34A1 Encoding Sodium-Phosphate Cotransporter 2A Cause Idiopathic Infantile Hypercalcemia. *J Am Soc Nephrol,* 27**,** 604-14.

SKELTON, P. D., STAN, R. V. & LUIKART, B. W. 2020. The Role of PTEN in Neurodevelopment. *Mol Neuropsychiatry,* 5**,** 60-71.

SLAGER, R. E., NEWTON, T. L., VLANGOS, C. N., FINUCANE, B. & ELSEA, S. H. 2003. Mutations in RAI1 associated with Smith-Magenis syndrome. *Nat Genet,* 33**,** 466-8.

SONG, Y. N., ZHAO, W. R., HUANG, X., WEI, L., HAN, J. Y., HOU, J. Y., LI, M. & CAO, X. 2025. Structural insights into retinoic acid receptor activation and selective modulators. *Clinical and Translational Discovery,* 5.

SU, C., MO, J., DONG, S., LIAO, Z., ZHANG, B. & ZHU, P. 2024. Integrinbeta-1 in disorders and cancers: molecular mechanisms and therapeutic targets. *Cell Commun Signal,* 22**,** 71.

TAHA, H. B., ZHU, S. & WANG, E. 2026. Prosaposin in CNS health and disease, metabolic stress and exercise adaptation. *J Mol Med (Berl),* 104**,** 38.

TERREY, M., ADAMSON, S. I., GIBSON, A. L., DENG, T., ISHIMURA, R., CHUANG, J. H. & ACKERMAN, S. L. 2020. GTPBP1 resolves paused ribosomes to maintain neuronal homeostasis. *Elife,* 9.

TYLKI-SZYMANSKA, A., GRADOWSKA, W., SOMMER, A., HEER, A., WALTER, M., REINHARD, C., OMRAN, H., SASS, J. O. & JURECKA, A. 2010. Aminoacylase 1 deficiency associated with autistic behavior. *J Inherit Metab Dis,* 33 Suppl 3**,** S211-4.

VAN DE STEEG, E., STRANECKY, V., HARTMANNOVA, H., NOSKOVA, L., HREBICEK, M., WAGENAAR, E., VAN ESCH, A., DE WAART, D. R., OUDE ELFERINK, R. P., KENWORTHY, K. E., STICOVA, E., AL-EDREESI, M., KNISELY, A. S., KMOCH, S., JIRSA, M. & SCHINKEL, A. H. 2012. Complete OATP1B1 and OATP1B3 deficiency causes human Rotor syndrome by interrupting conjugated bilirubin reuptake into the liver. *J Clin Invest,* 122**,** 519-28.

WADA, S., HAMADA, M., KOBAYASHI, K. & SATOH, N. 2008. Novel genes involved in canonical Wnt/beta-catenin signaling pathway in early Ciona intestinalis embryos. *Dev Growth Differ,* 50**,** 215-27.

WANG, H., WU, B., HE, X., LI, W. & GUAN, W. 2024. ZNF429 Participates in the Progression of Coronary Heart Disease through Regulating Inflammatory and Adhesive Factors. *Front Biosci (Landmark Ed),* 29**,** 335.

WEVERS, R. A., CHRISTENSEN, M., ENGELKE, U. F. H., GEUER, S., COENE, K. L. M., KWAST, J. T., LUND, A. M. & VISSERS, L. 2019. Functional disruption of pyrimidine nucleoside transporter CNT1 results in a novel inborn error of metabolism with high excretion of uridine and cytidine. *J Inherit Metab Dis,* 42**,** 494-500.

ZHOU, X., FELICIANO, P., SHU, C., WANG, T., ASTROVSKAYA, I., HALL, J. B., OBIAJULU, J. U., WRIGHT, J. R., MURALI, S. C., XU, S. X., BRUEGGEMAN, L., THOMAS, T. R., MARCHENKO, O., FLEISCH, C., BARNS, S. D., SNYDER, L. G., HAN, B., CHANG, T. S., TURNER, T. N., HARVEY, W. T., NISHIDA, A., O'ROAK, B. J., GESCHWIND, D. H., CONSORTIUM, S., MICHAELSON, J. J., VOLFOVSKY, N., EICHLER, E. E., SHEN, Y. & CHUNG, W. K. 2022. Integrating de novo and inherited variants in 42,607 autism cases identifies mutations in new moderate-risk genes. *Nat Genet,* 54**,** 1305-1319.

ZHU, Z., YAO, J., JOHNS, T., FU, K., DE BIE, I., MACMILLAN, C., CUTHBERT, A. P., NEWBOLD, R. F., WANG, J., CHEVRETTE, M., BROWN, G. K., BROWN, R. M. & SHOUBRIDGE, E. A. 1998. SURF1, encoding a factor involved in the biogenesis of cytochrome c oxidase, is mutated in Leigh syndrome. *Nat Genet,* 20**,** 337-43.
