## Supplementary Table 4 for "Comparing the Phenotypic and Genetic Differences Between Autistic Regression and Autism Without Regression Using the SPARK Cohort"

| **Supplementary Table 4**. Age group and regression status of the individuals who received a clinical genetic diagnosis of *PTEN* -related disorder in the SPARK collection version 14. *Individuals present also in our validation cohort as *PTEN* carriers (see Supplementary Table 3). Toddler/preschooler: 1-6 years; Child: 6-12 years; Teenager: 12-18 years; Adult > 18 years. | | |
| --- | --- | --- |
| Age group at evaluation | Language regression | Other regression |
| Adult* | Yes | No |
| Teenager | Yes | No |
| Toddler/preschooler | No | No |
| Adult* | Yes | No |
| Adult | Yes | No |
| Child | Yes | Yes |
| Toddler/preschooler | No | No |
| Child | Yes | No |
| Teenager | No | No |
| Toddler/preschooler | No | Yes |
| Child | Yes | No |
| Toddler/preschooler | Yes | Yes |
| Child | Yes | No |
| Toddler/preschooler | No | No |
| Adult | No | No |
