## Supplementary Table 5 for "Comparing the Phenotypic and Genetic Differences Between Autistic Regression and Autism Without Regression Using the SPARK Cohort"

| **Supplementary Table 5**. Enrichr-KG enrichment analysis | | | | | | |
| --- | --- | --- | --- | --- | --- | --- |
| Term | Library | p-value | q-value | z-score | combined score | overlaps |
| Phosphate Bond Hydrolysis By NTPDase Proteins R-HSA-8850843 | Reactome_2022 | 0.000191 | 0.0397 | 130.3 | 1116 | ENTPD1; ENTPD4 |
| nucleoside diphosphate catabolic process (GO:0009134) | GO_Biological_Process_2021 | 0.000245 | 0.0883 | 111.7 | 928.7 | ENTPD1; ENTPD4 |
| nucleoside phosphate catabolic process (GO:1901292) | GO_Biological_Process_2021 | 0.000614 | 0.0883 | 65.15 | 481.8 | ENTPD1; ENTPD4 |
| nucleobase-containing small molecule catabolic process (GO:0034656) | GO_Biological_Process_2021 | 0.000708 | 0.0883 | 60.13 | 436.2 | ENTPD1; ENTPD4 |
| nucleoside diphosphate metabolic process (GO:0009132) | GO_Biological_Process_2021 | 0.000807 | 0.0883 | 55.83 | 397.6 | ENTPD1; ENTPD4 |
| RNA Polymerase II Transcription R-HSA-73857 | Reactome_2022 | 0.002084 | 0.1779 | 3.33 | 20.56 | L3MBTL1; ZNF143; SURF1; ZNF429; ZSCAN32; RARA; PTEN; ZNF446; MED15; SRSF11 |
| Nucleotide Catabolism R-HSA-8956319 | Reactome_2022 | 0.003876 | 0.1779 | 23.66 | 131.4 | ENTPD1; ENTPD4 |
| Gene Expression (Transcription) R-HSA-74160 | Reactome_2022 | 0.004277 | 0.1779 | 2.991 | 16.32 | L3MBTL1; ZNF143; SURF1; ZNF429; ZSCAN32; RARA; PTEN; ZNF446; MED15; SRSF11 |
| Pyrimidine metabolism | KEGG_2021_Human | 0.009684 | 0.2679 | 14.45 | 66.99 | ENTPD1; ENTPD4 |
| Focal adhesion | KEGG_2021_Human | 0.01621 | 0.2691 | 5.985 | 24.67 | ITGB1; PTEN; PPP1R12B |
