## Supplementary Table 6 for "Comparing the Phenotypic and Genetic Differences Between Autistic Regression and Autism Without Regression Using the SPARK Cohort"

| **Supplementary Table 6.** Multilinominal logistic regression results. We tested the association between Autreg and ASD, ADHD, epilepsy, and schizophrenia PGS strata using exploratory data. The included PGS strata were 0–10%, 10–90%, and 90–100%. After adjusting for sex and age at evaluation a nominal significant association between schizophrenia PGS strata and regression status was noticed | | | |
| --- | --- | --- | --- |
| **Phenotype** | **p_value** | **adjusted_p_value** | **significance** |
| ADHD | 0.594 | 0.594 | ns |
| ASD | 0.424 | 0.566 | ns |
| Epilepsy | 0.385 | 0.566 | ns |
| Schizophrenia | 0.0313 | 0.125 | ns |
