## Supplementary Methods and Figures for "Comparing the Phenotypic and Genetic Differences Between Autistic Regression and Autism Without Regression Using the SPARK Cohort"

**Genome sequencing and variants calling, annotation, and filtering**

In SPARK, genome sequencing (GS) was done on the Illumina NovaSeq 6000. Variants were called using GATK (version 3.5)’s HaplotypeCaller and jointly using GLnexus (version 1.4.1).

Variant annotation and filtering

Variants were annotated using Ensembl Variant Effect Predictor (VEP, version111.0) on GRCh38 genomic build. Coding variants were selected using the “--coding_only” flag. These resulted in 2 046 DNV (1 123 in Autreg and 923 in Autnreg), 945,968 transmitted (509 989 and 435 979) and 1 521 792 non-transmitted (830 112 and 691 680) variants in 2 081 individuals (1 107 with Autreg and 974 with Autnreg). To proceed only with rare variants, we filtered out variants with a maximum observed allele frequency in 1000 Genomes (max_af) ≥ 0.01. We selected only variants predicted to be loss-of-function variants (LoF), missense variants, and synonymous variants. Under LoF category we included nonsense, frameshift, splice-donor, and splice-acceptor variants. Based on these criteria, we analyzed 1 935 DNV (955 and 980), 36 453 transmitted (18 287 and 18 166), and 36 717 non-transmitted (18 420 and 18 297) variants.

De novo calling

We included 5 712 trios from 3 364 families and identified de novo variants (DNV) as previously described^1^. In brief, DNV was called if it was called so in Slivar^2^ (version 0.2.8), with allele balance in children higher than 0.25 and identified as high confidence DNV in GATK^3^ (version 4.1.4.1). We separately considered the variant genotype as 1/0 in children when assessing the pseudo-autosomal regions on the sex chromosome and did not find DNVs on chrY’s pseudo-autosomal regions. We removed variants with GQ < 20, DP < 10, gnomAD population allele frequencies > 0.001, variants of either 10 Adenosines or Thymidines in a row, and those on genomic centromeres and low complex regions. We then removed DNV found in other familys’ parents, found in children from > 3 families, and on positions with > 3 alleles.

Transmitted and non-transmitted variant calling

Transmitted and non-transmitted variants were identified using Slivar (version 0.2.7) and bcftools (version 1.17). Variants were first normalized to split multiallelic sites and indexed per chromosome. For both categories, we required depth (DP) ≥ 10 and genotype quality (GQ) ≥ 20 in child and both parents, and considered only variants that passed VCF quality filters (PASS). For transmitted variants, the child was required to carry the alternate allele (heterozygous or homozygous alternate) with an allele balance ≥ 0.25, and at least one parent also carried the alternate allele in a genotype consistent with Mendelian transmission (e.g., heterozygous in one parent and homozygous reference in the other, or both parents heterozygous). For non-transmitted variants, the child was required to be homozygous for the reference allele, while at least one parent carried the alternate allele (heterozygous or homozygous for the alternate allele). On chromosome X, we applied sex-aware rules. For transmitted variants, female children were required to carry the alternate allele if at least one parent also carried it; male children were required to carry the alternate allele if the mother carried it. For non-transmitted variants, female children were required to be homozygous reference with at least one parent carrying the alternate allele, while male children were required to be reference with the mother carrying the alternate allele. Following per-chromosome filtering, variants from autosomes and chromosome X were merged and re-indexed to generate the final callsets of transmitted and non-transmitted variants.

**Statistical association analysis using TADA**

Risk-gene association was evaluated using the Transmission and De Novo Association (TADA) framework, which integrates evidence from de novo and inherited damaging variants within a hierarchical Bayesian model. TADA calculates a gene-level Bayes factor (BF) comparing a disease-risk model (H1) with a neutral model (H0), thereby quantifying the evidence that the observed variant burden in a gene is associated with phenotype^4-6^.

The analysis was performed separately in 1 107 Autreg trios (cases) and 974 Autnreg trios (controls). Variants were divided into two functional classes: loss-of-function (LoF) variants and damaging missense (misD) variants. For each gene and variant class, we incorporated counts of de novo variants (DN), transmitted variants (Trans), and non-transmitted variants (NonTrans). Gene-specific mutation rates were incorporated for both LoF and misD variants. Genes were retained only when corresponding mutation-rate information and prior parameters were available, and gene identifiers were explicitly matched and aligned across all input matrices before analysis. We used the TADA-R^6^ TADA version without including recessive variants. Gene priors were obtained from the TADA-R^6^ github page (<https://github.com/limo936/TADA-R>; accessed in August 2026). Refer to the main manuscript for the mutation rates source.

Transmission analysis

Transmitted and non-transmitted variants were modelled using trio-based transmission framework. Under the neutral hypothesis, the number of transmitted variants was modelled as a binomial outcome conditional on the total number of transmission opportunities (N_trans_= N_Tran_s + N_NonTrans_) with a transmission probability of 0.5. Thus, the case and control analyses retained the within-family 50:50 transmission null rather than comparing transmission counts directly between populations.

Case and control trios were analyzed as independent datasets. Separate sample-count matrices and variant-count matrices were constructed for each cohort, preventing control-family transmission information from entering the case analysis and vice versa. This design allowed the resulting case and control BFs to be compared directly to identify genes showing disease-associated enrichment specifically in the case cohort.

TADA model specification

TADA was applied separately to LoF and misD variants and the resulting class-specific evidence was combined multiplicatively to obtain an overall gene-level Bayes factor (BF_total_ = BF_LoF_× BF_misD_).

The model parameters for the de novo and case-control components were specified according to the TADA parameterization. For LoF variants, the hyperparameters were set to gamma.mean.dn = 34, beta.dn = 1, gamma.mean.CC = 9.5, and beta.CC = 4. For damaging missense variants, the corresponding values were gamma.mean.dn = 20, beta.dn = 1, gamma.mean.CC = 4, and beta.CC = 1000. Gene-specific prior parameters were incorporated from the LoF and damaging-missense prior files.

Empirical null distributions

To obtain empirical significance measures, null distributions were generated independently for the case and control cohorts using 20 000 null replicates. In each replicate, de novo counts were simulated according to the gene-specific neutral mutation model, while transmitted and non-transmitted counts were generated under the neutral 50:50 transmission model.

Because transmission counts are constrained by the observed number of transmission opportunities, the possible combinations were precomputed for each gene according to its observed transmission total. Their corresponding case-control Bayes factors were calculated once and stored as lookup tables. During the 20,000 null simulations, these precomputed values were retrieved rather than repeatedly evaluating the numerical integration required for each possible transmission configuration. This substantially reduced computational burden while preserving the underlying 50:50 transmission null model.

The null calculations also incorporated numerical safeguards for vectorized mutation-rate calculations and numerical integration. These safeguards prevented zero-likelihood and numerical underflow conditions from generating undefined Bayes factors and ensured that the null calculations remained stable when applied genome-wide.

Multiple-testing correction and case specificity

Bayesian false discovery rates (FDRs) were calculated from the gene-level total Bayes factors using a prior null probability of π_0_ = 0.95. Empirical p-values were obtained by comparing the observed gene-level BF with the corresponding independently generated null distribution.

Finally, case and control results were integrated into a side-by-side table containing the gene-level BF, Bayesian FDR, and empirical p-value for each cohort. Genes were prioritized based on strong evidence of association in cases (Autreg) together with comparatively weak evidence in controls (Autnreg).

**Polygenic scores exploratory analysis**

Quality control

We performed quality control (QC) for 12 519 SPARK participants using PLINK1.9^7^ as described previously^1^. In brief, we excluded variants with genotype missingness > 0.01, variants not in Hardy-Weinberg equilibrium (p < 1 × 10−^6^), individuals with heterozygosity rate > ±3 standard deviations from the samples’ heterozygosity rate mean. After QC we kept 11 933 individuals and 9 558 997 variants with a total genotyping rate of 99.9%.

The genomic coordinates were lifted over from hg38 to hg19 using liftOver^8^.

Population stratification and selection of individuals of European ancestry

We performed ancestry prediction to identify individuals of European ancestry using a support vector machine (SVM) approach implemented in R (package e1071). First, we used 1000 Genomes Project reference samples representing African (AFR), Admixed American (AMR), East Asian (ASN), and European (EUR) superpopulations. Reference samples were randomly split into training (80%) and testing (20%) sets. Labels were assigned as 1 for EUR and 0 for non-EUR. An SVM classifier with a linear kernel was trained using the first two multidimensional scaling (MDS) components (C1 and C2) as input features. Model performance was evaluated on the held-out test set using accuracy and confusion matrices. Once accuracy was confirmed, the trained model was applied to our samples to predict ancestry. Individuals predicted as EUR were retained for downstream analysis. For visualization, we generated multiple quality-control plots, including MDS plots of the 1000 Genomes superpopulations, training vs. testing set separation, the SVM decision boundary, prediction accuracy on the test set, and predicted ancestry assignments for our samples (supplementary figures S2-6).

Base data and polygenic score calculations

We selected four genome-wide association studies (GWAS) to serve as base data to calculate polygenic scores (PGS) for autism (46 350 participants)^9^, ADHD (70 953 participants)^10^, epilepsy (69 995 participants)^11^, and schizophrenia (130,644 participants)^12^. We only used summary statistics of European ancestry individuals as base data. We filtered SNPs with MAF < 1% and information imputation score < 0.8 when those parameters were provided. We checked for duplicated and ambiguous SNPs and arranged the base data files per the requirements of PRS-CS^13^. PGS was calculated using the default parameters of PRS-CS. We calculated the per-individual PGS using the “--score” PLINK command.

**Phasing and imputation**

Phasing was performed using SHAPEIT^14^ (v5.1.1) with pedigree information. Imputation was performed on version 2 of the TOPMed Imputation Server^15^ using TOPMed reference panel (version r3). Post-imputation filtering retained SNPs with imputation quality R² ≥0.5, genotype missingness <0.01, MAF >0.01, and HWE P ≥1e-6, while removing duplicated variants.

**Supplementary Figures**

**Methods Figures**

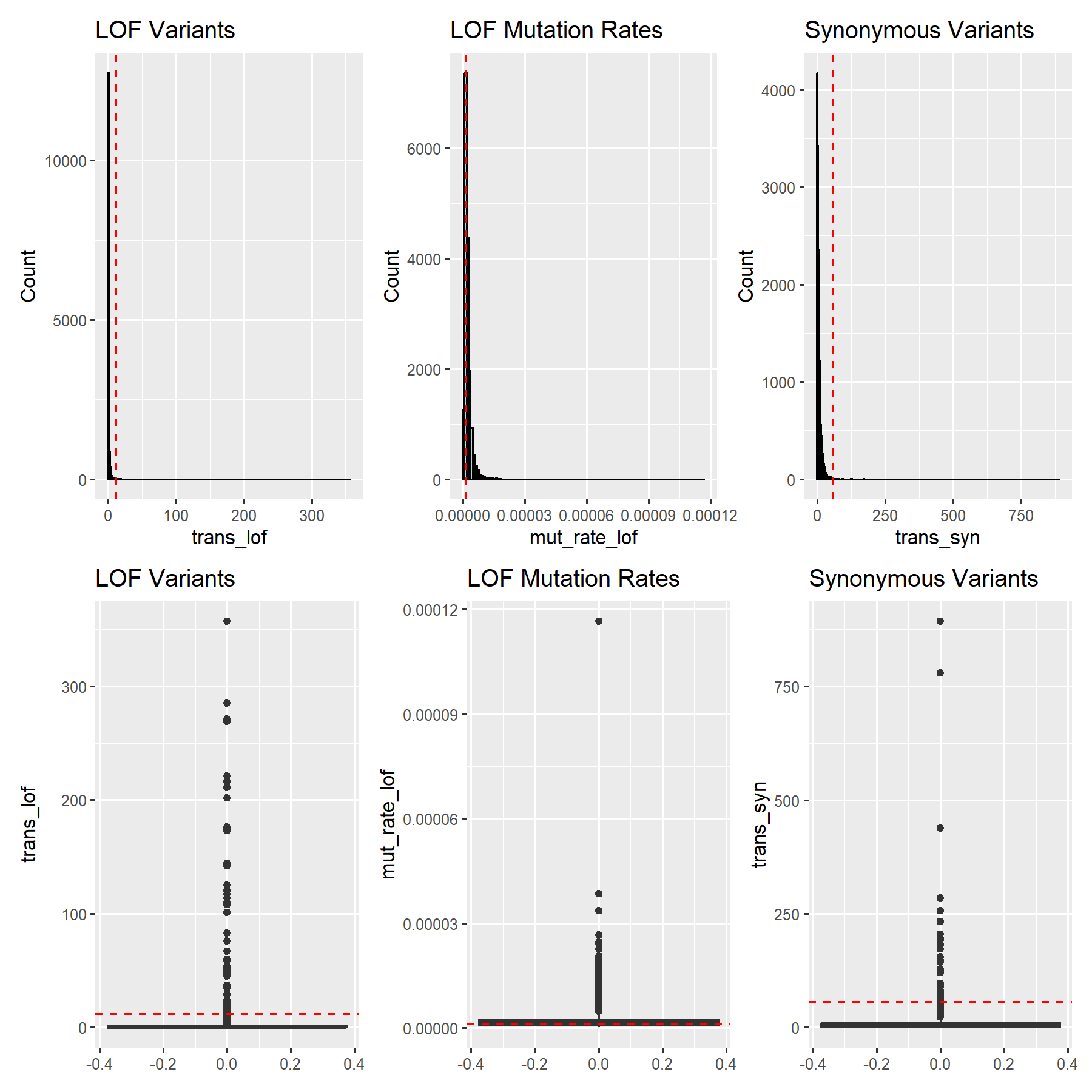

*Supplementary Figure 1. Cutoffs used to reduce bias from potential technical or annotation artifacts. We applied cutoffs that corresponded to 99.5% (interrupted red lines) of transmitted loss-of-function (LOF) variants (trans_lof), transmitted synonymous (syn) variants (trans_syn), LOF mutation rates (mut_rate_lof) and syn mutation rates (mut_rate_syn). We excluded genes with extreme variant counts (transmitted LoF > 12, transmitted synonymous > 57, or transmitted LoF > 12 with LoF mutation rate < 1e-06). The upper panel shows histograms and the lower panel shows scatter plot.*

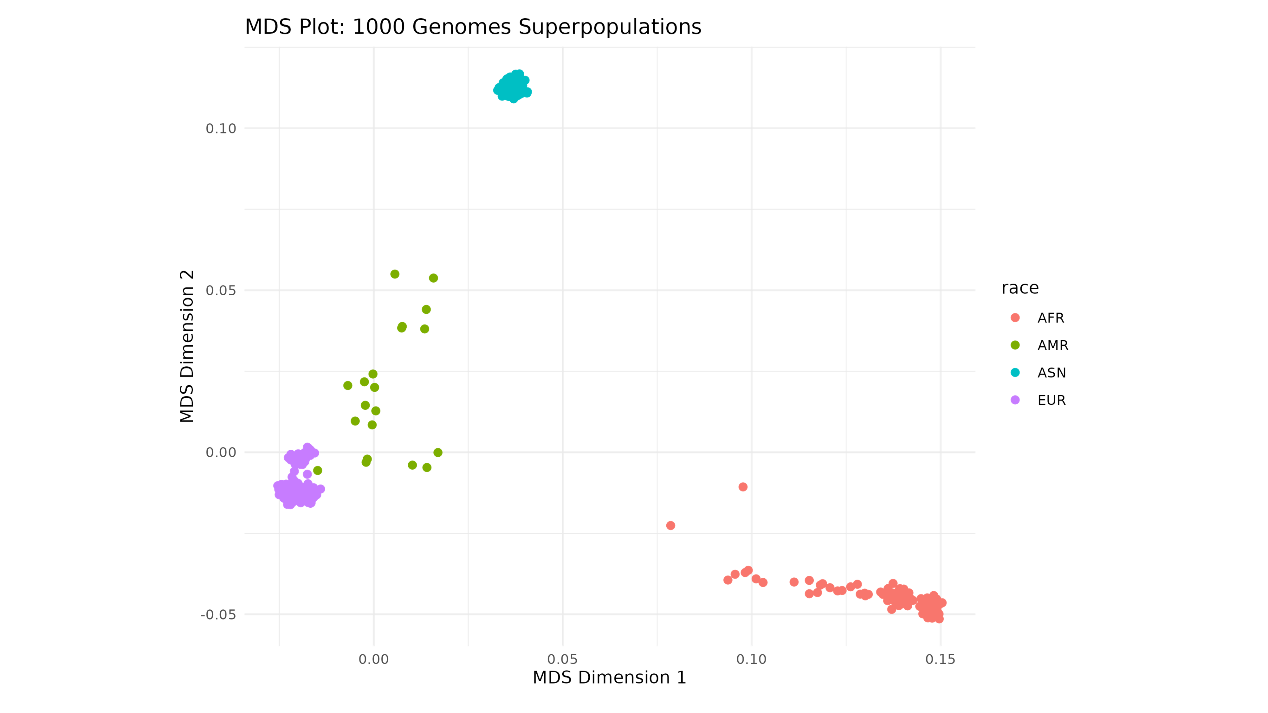

*Supplementary Figure 2: 1000 Genomes MDS plot. This plot shows the clustering of individuals from different ancestry groups (AFR, AMR, ASN, EUR) using the first two MDS dimensions (C1 and C2).*

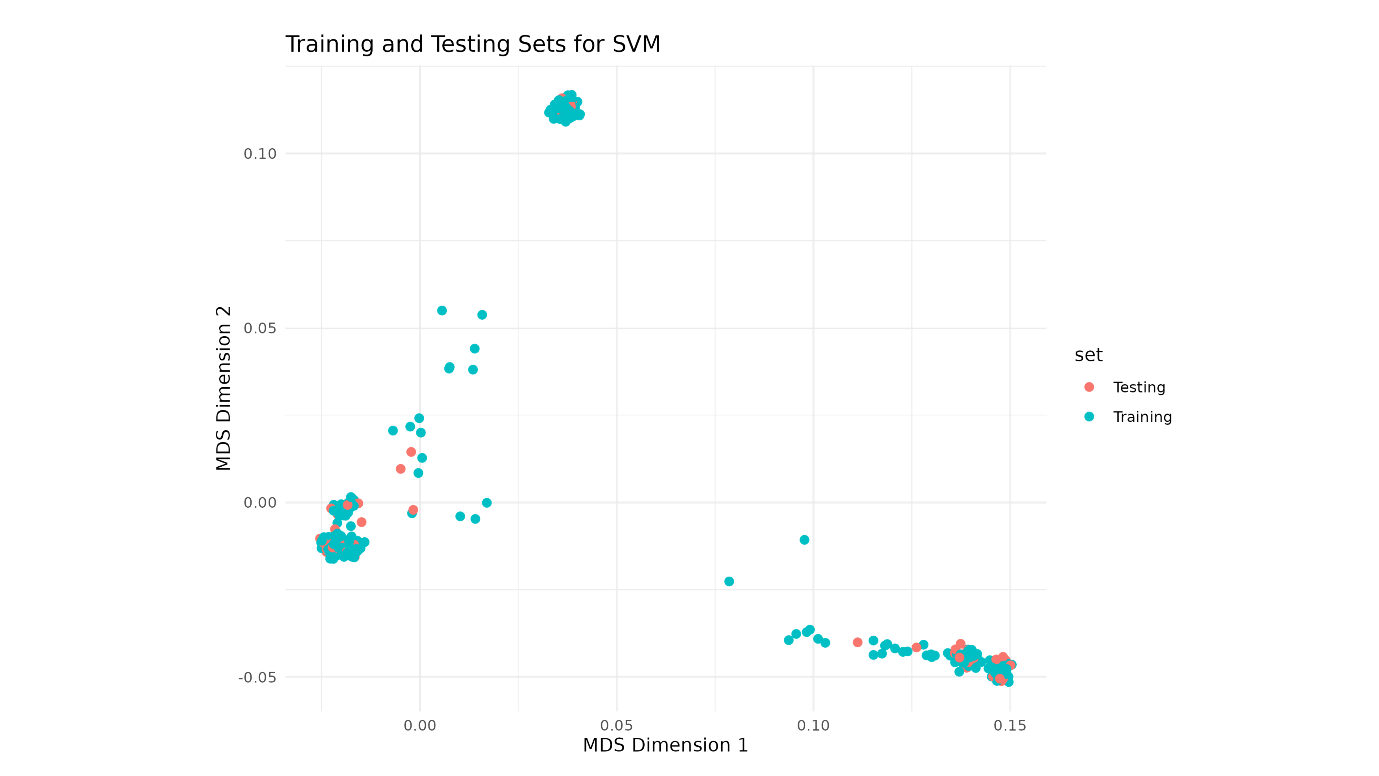

*Supplementary Figure 3: Training and Test Set MDS Plot. Data separation after splitting the 1000 Genomes samples into 80% training and 20% test sets.*

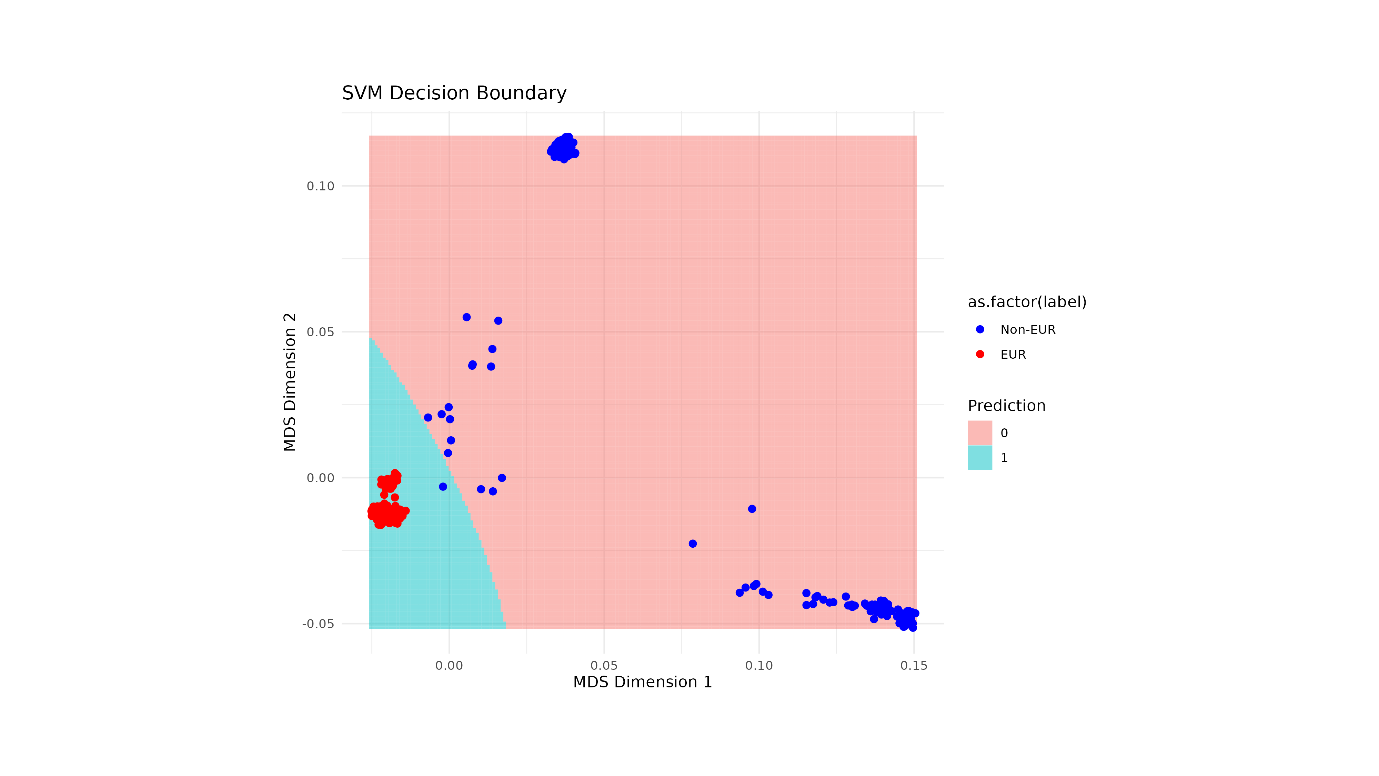

*Supplementary Figure 4: SVM Decision Boundary Plot. SVM decision boundary after training.*

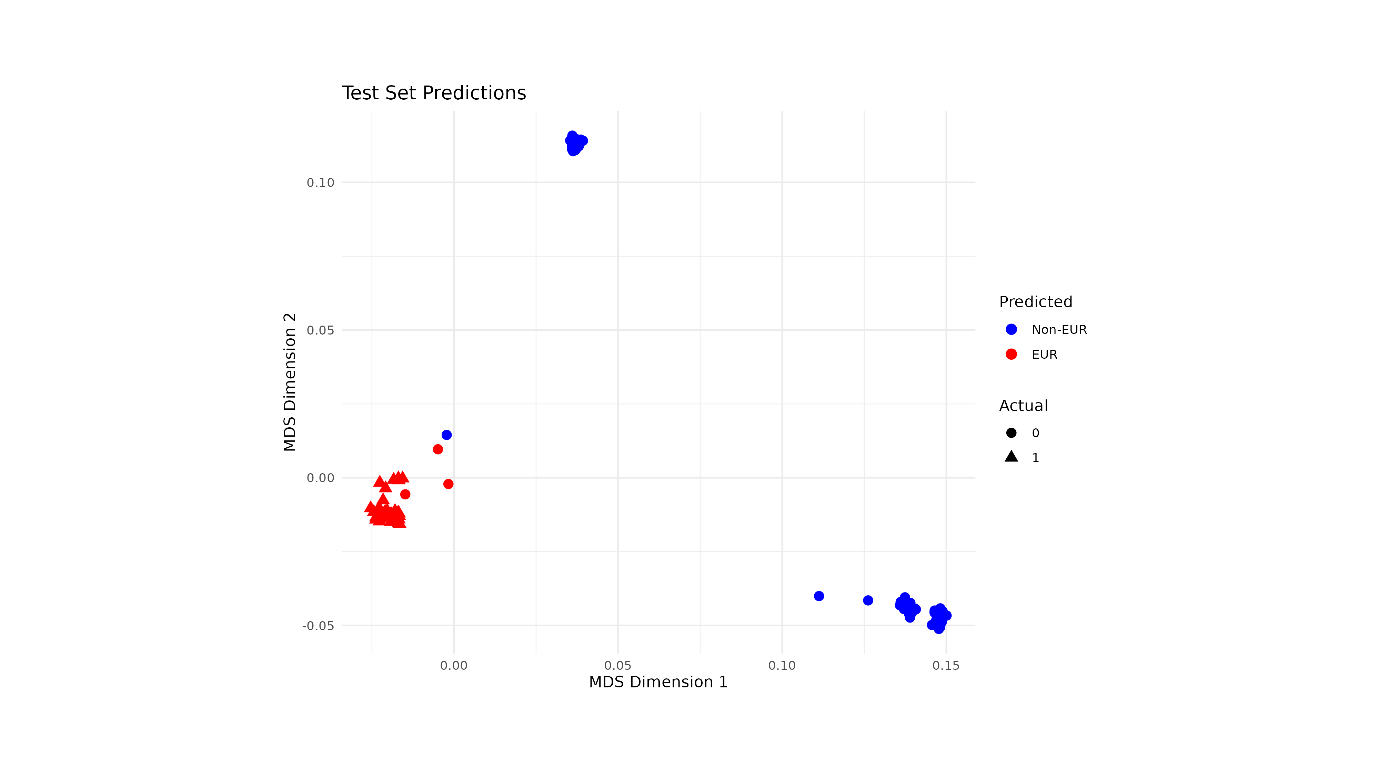

*Supplementary Figure 5: Test Set Predictions Plot. The predictions of the SVM model on the test set to visualize how well it classified European vs non-European individuals. An accuracy of 0.97 was obtained. Note: I have tried a linear Kernel but there was no difference in accuracy.*

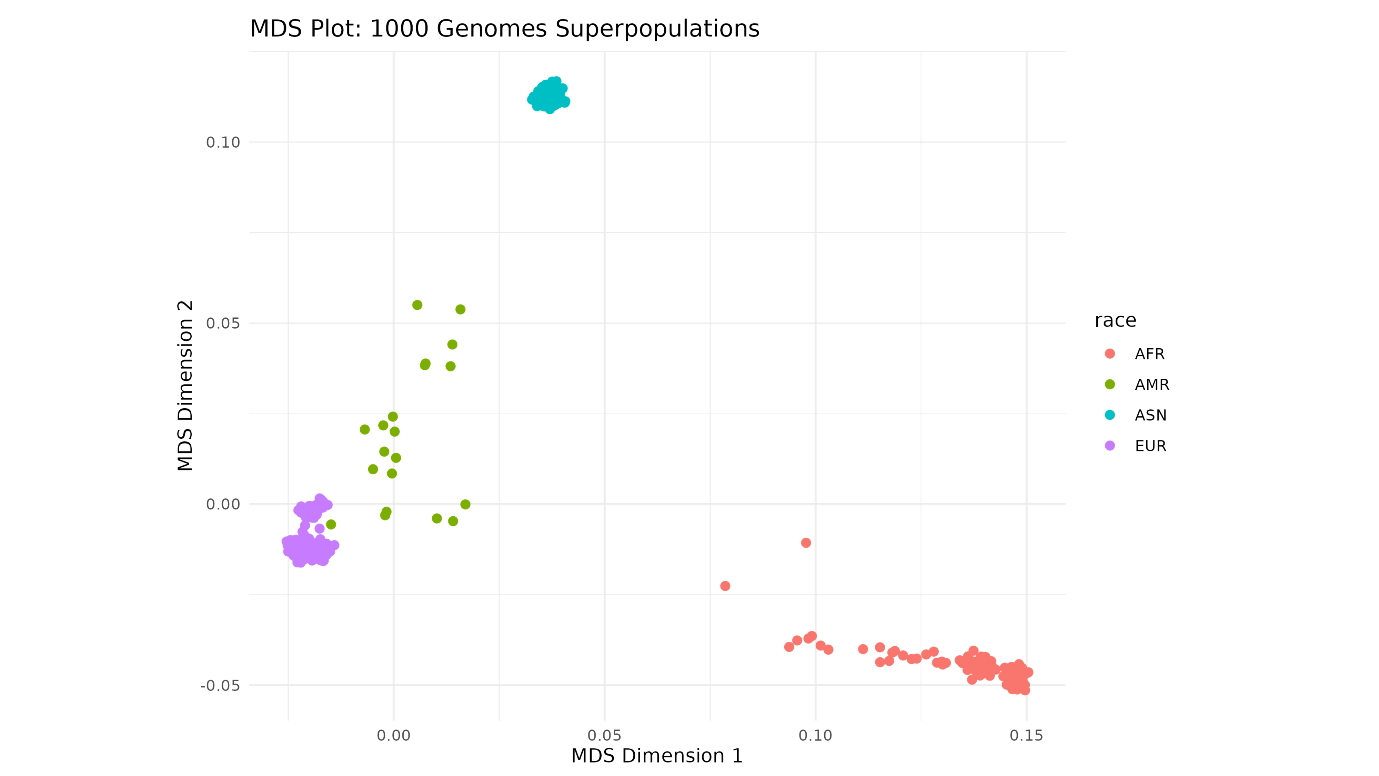

*Supplementary Figure 6: SPARK MDS Plot. This plot shows the clustering of individuals from different ancestry groups (AFR, AMR, ASN, EUR) using the first two MDS dimensions (C1 and C2). 9871 individuals were predicted as of European ancestry.*

**Results Figures**

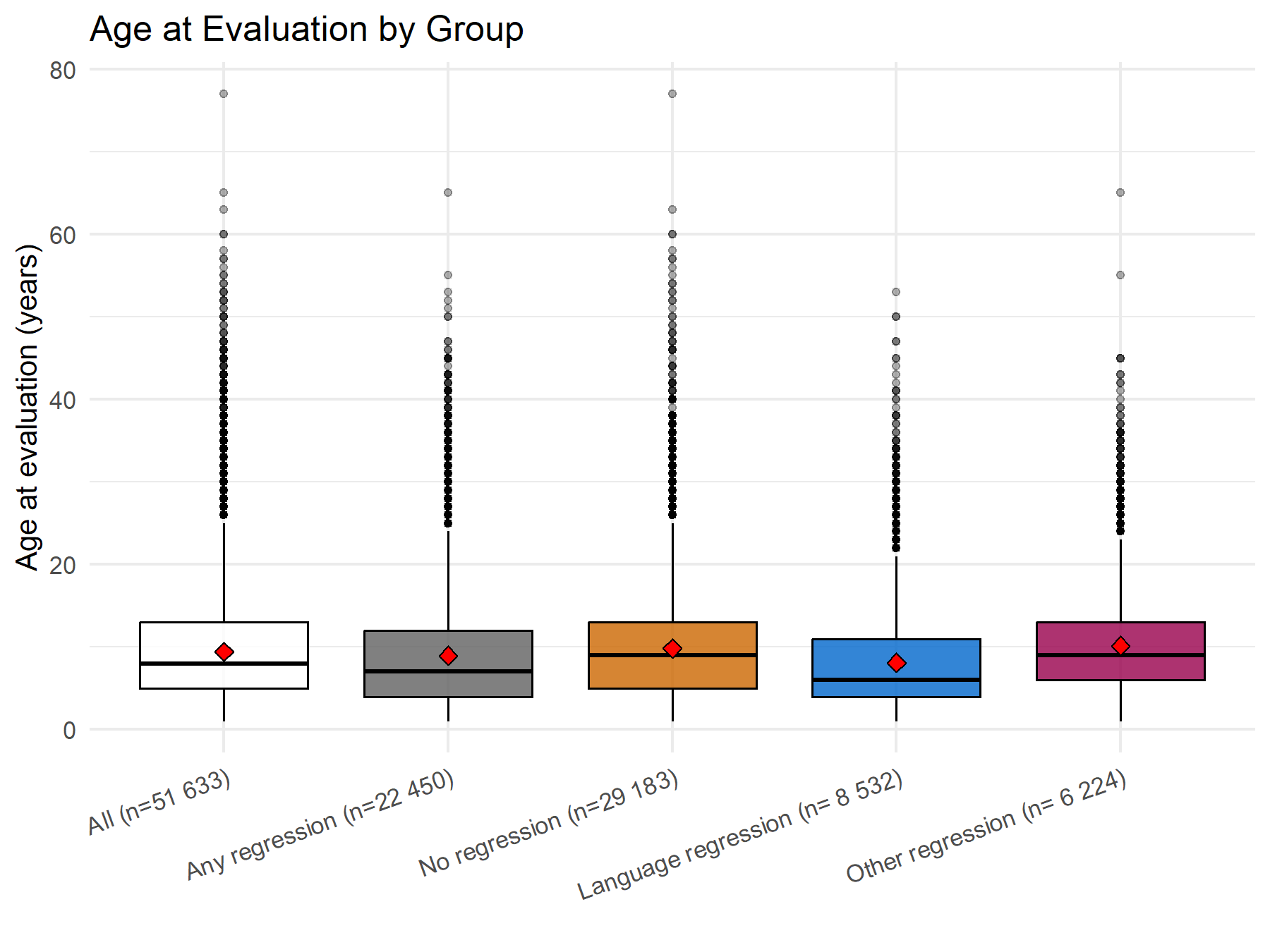

*Supplementary Figure 7. Age at evaluation in the different groups. The mean age at evaluation in all the studied individuals was 9.4 years (standard deviation 6.1, median 8, range 1-77, 95^th^ centile 21), in the those with any type of autistic regression 8.9 (standard deviation 6.2, median 7, range 1-65, 95^th^ centile 21), in those without regression 9.8 (standard deviation 6, median 9, range 1-77, 95^th^ centile 21), in those with only language regression 8.1 (standard deviation 6.1, median 6, range 1-53, 95^th^ centile 20), and in those with only other regression was 10.1 (standard deviation 6, median 9, range 1-65, 95^th^ centile 21). Red diamonds indicate the mean values.*

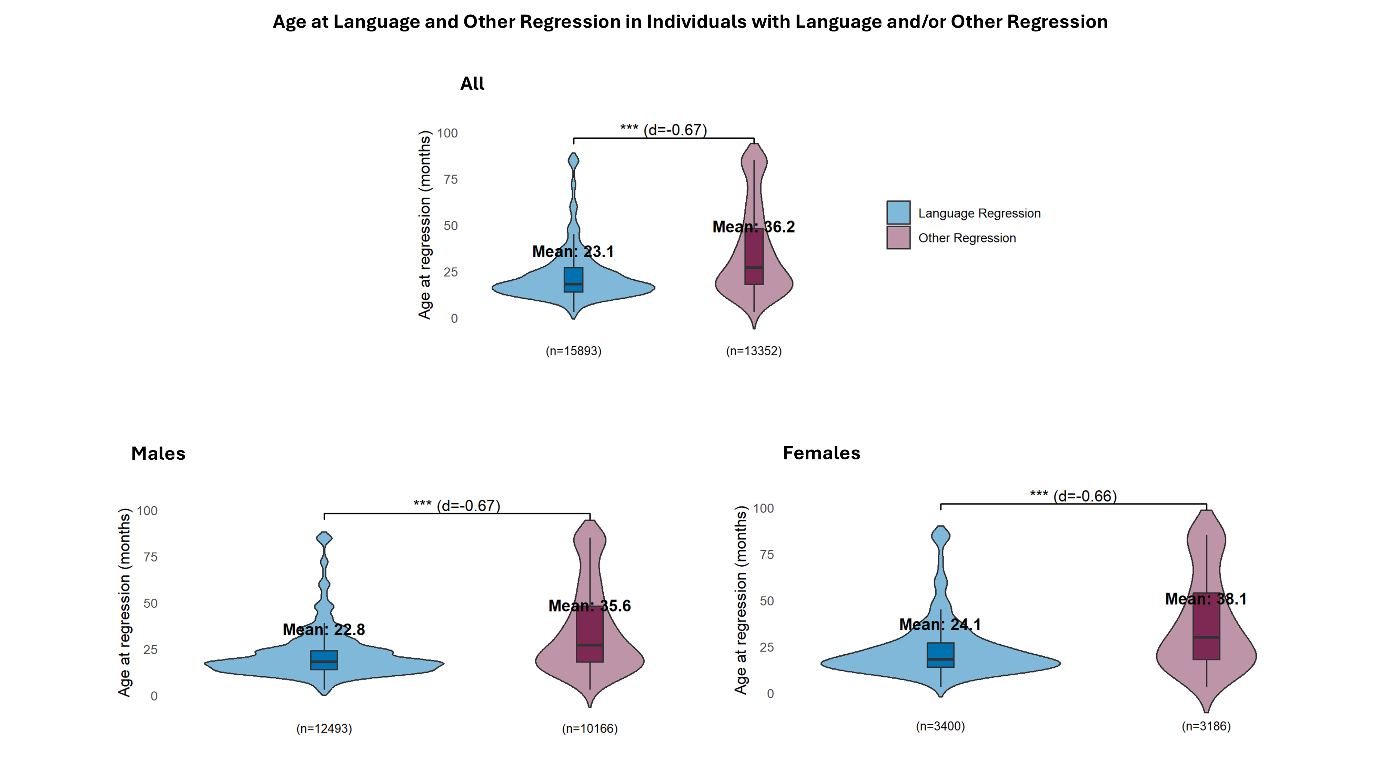

*Supplementary Figure 8. Age at language and other regression in individuals with regression. Both males and females: The mean age at language regression was 23.1 months (range = 3 – 85 months, median = 18, 95^th^ centile = 54) while the mean age at other regression was 36.2 months (range = 3 – 85 months, median = 27, 95 ^th^ centile = 85). The difference between the two means was statistically significant using two sample t-test (t = -54.999, df = 21297, p-value < 2.2e-16). Cohen’s d indicates a moderate effect size (0.67). Males: The mean age at language regression was 22.8 months (range = 3 – 85 months, median = 18, 95 ^th^ centile = 51) while the mean age at other regression was 35.6 months (range = 3 – 85 months, median = 27, 95 ^th^ centile = 85). The difference between the two means was statistically significant using two sample t-test (t = -48.037, df = 15959, p-value < 2.2e-16). Cohen’s d indicates a moderate effect size (0.67). Females: The mean age at language regression was 24.1 months (range = 3 – 85 months, median = 18, 95 ^th^ centile = 60) while the mean age at other regression was 38.1 months (range = 3 – 85 months, median = 30, 95 ^th^ centile = 85). The difference between the two means was statistically significant using two sample t-test (t = -26.5, df = 5357.5, p-value < 2.2e-16). Cohen’s d indicates a moderate effect size (0.66).*

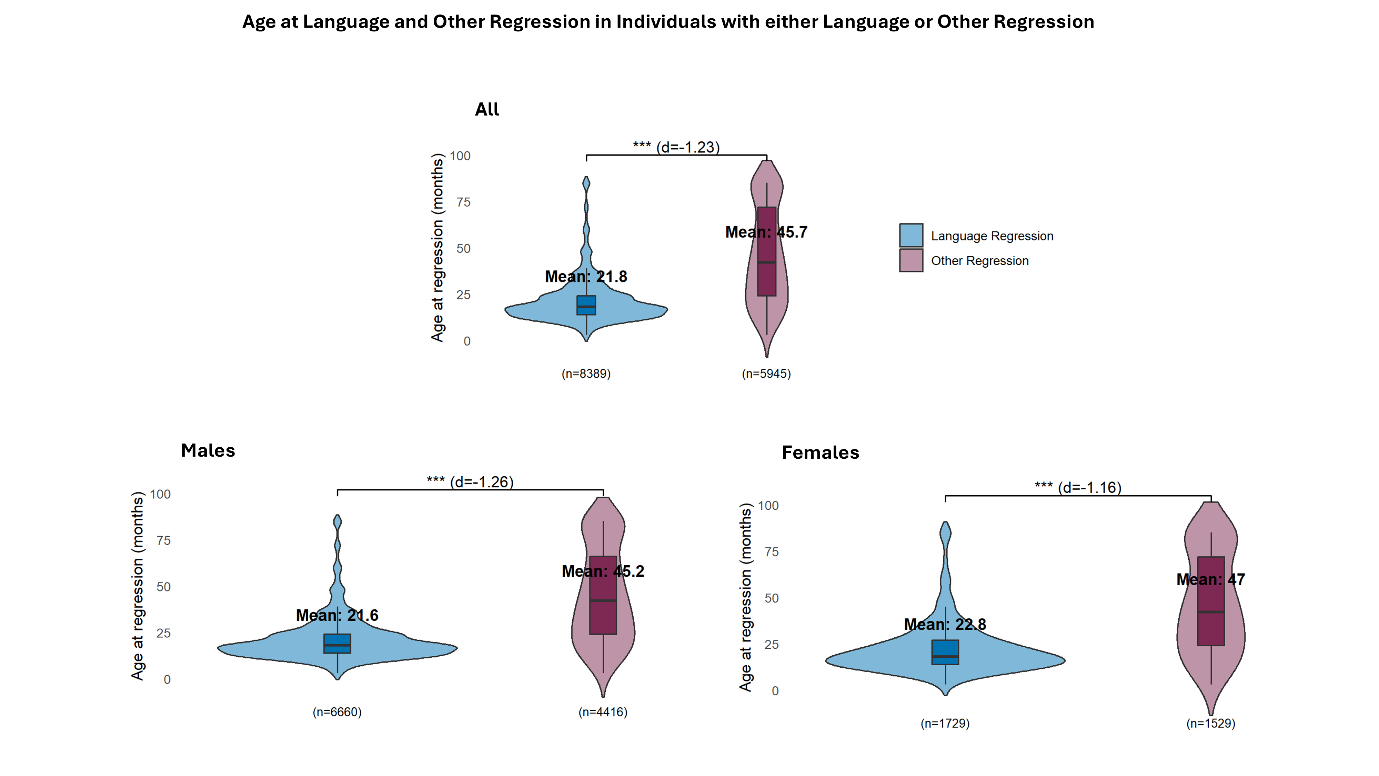

*Supplementary Figure 9. Age at language and other regression in individuals with one regression type. Both males and females: The mean age at language regression was 21.8 months (range = 3 – 85 months, median = 18, 95 ^th^ centile = 48) while the mean age at other regression was 45.7 months (range = 3 – 85 months, median = 42, 95 ^th^ centile = 85). The difference between the two means was statistically significant using two sample t-test (t = -65.815, df = 8147.4, p-value < 2.2e-16). Cohen’s d indicates a moderate effect size (-1.23). Males: The mean age at language regression was 21.6 months (range = 3 – 85 months, median = 18, 95 ^th^ centile = 48) while the mean age at other regression was 45.2 months (range = 3 – 85 months, median = 42, 95 ^th^ centile = 85). The difference between the two means was statistically significant using two sample t-test (t = -57.279, df = 5900.6, p-value < 2.2e-16). Cohen’s d indicates a moderate effect size (-1.26). Females: The mean age at language regression was 22.8 months (range = 3 – 85 months, median = 18, 95 ^th^ centile = 49.8) while the mean age at other regression was 47 months (range = 3 – 85 months, median = 42, 95 ^th^ centile = 85). The difference between the two means was statistically significant using two sample t-test (t = -31.879, df = 2300.6, p-value < 2.2e-16). Cohen’s d indicates a moderate effect size (-1.16).*

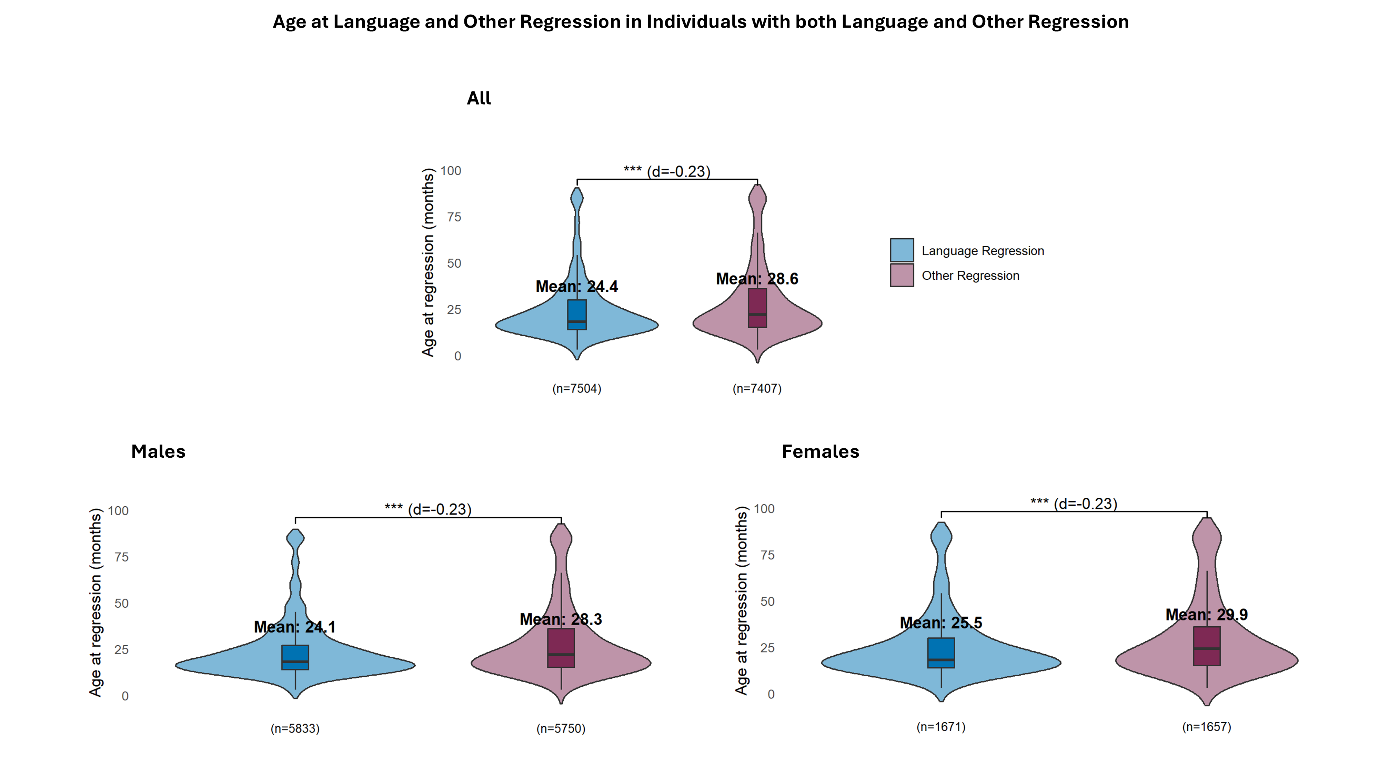

*Supplementary Figure 10. Age at language and other regression in individuals with both regression types. Both males and females: The mean age at language regression was 24.4 months (range = 3 – 85 months, median = 18, 95 ^th^ centile = 60) while the mean age at other regression was 28.6 months (range = 3 – 85 months, median = 22, 95 ^th^ centile = 84). The difference between the two means was statistically significant using two sample t-test (t = -14.17, df = 14319, p-value < 2.2e-16). Cohen’s d indicates a moderate effect size (-0.23). Males: The mean age at language regression was 24.1 months (range = 3 – 85 months, median = 18, 95 ^th^ centile = 60) while the mean age at other regression was 28.3 months (range = 3 – 85 months, median = 22, 95 ^th^ centile = 84). The difference between the two means was statistically significant using two sample t-test (t = -12.623, df = 11094, p-value < 2.2e-16). Cohen’s d indicates a moderate effect size (-0.23). Females: The mean age at language regression was 25.5 months (range = 3 – 85 months, median = 18, 95 ^th^ centile = 72) while the mean age at other regression was 29.9 months (range = 3 – 85 months, median = 24, 95 ^th^ centile = 85). The difference between the two means was statistically significant using two sample t-test (t = -6.4979, df = 3217.8, p-value = 9.407e-11). Cohen’s d indicates a moderate effect size (-0.23).*

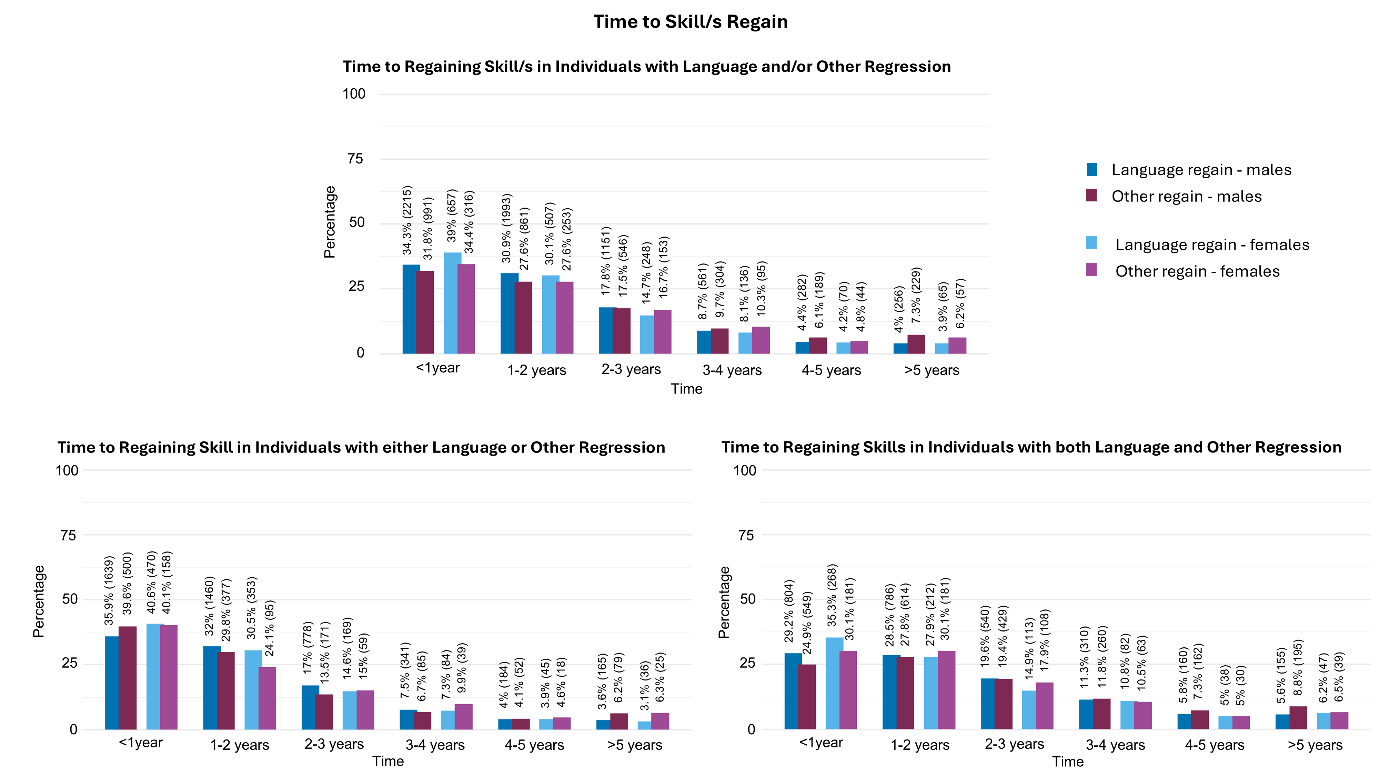

*Supplementary Figure 11. Time to skill/s regain. Upper panel: Times to skill/s regain according to sex and the type of regression is provided. Lower panel: Times to skill/s regain in those with single and double regression is provided according to sex and regression type.*

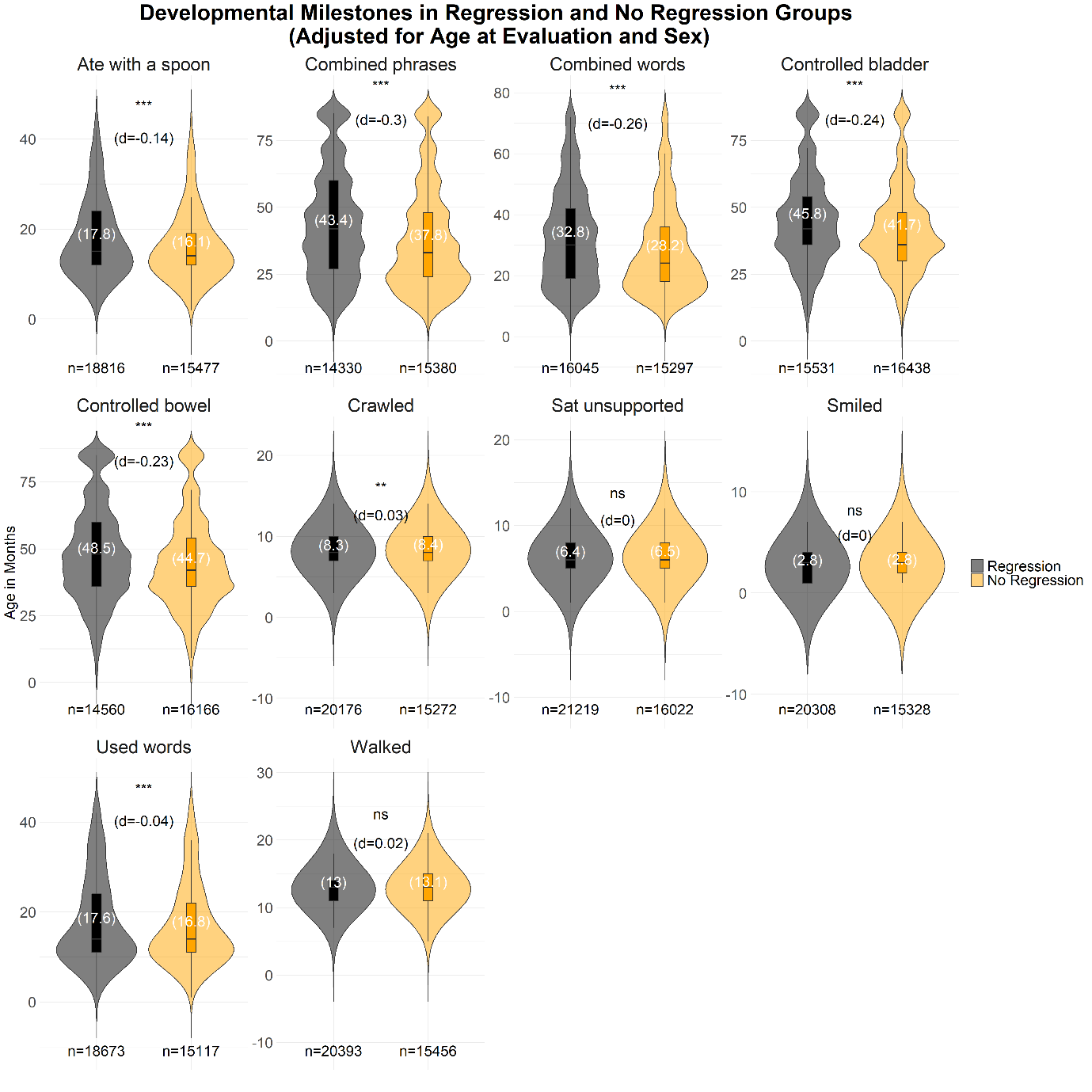

*Supplementary Figure 12. Comparing ages at reaching developmental milestones in the regression and no regression groups. In the no regression category, we included individuals without regression who were >7.08 years old at evaluation, which was the age at which 95% of individuals in SPARK v14 phenotype data with autistic regression developed regression. In the regression category we included all individuals with autistic regression (language regression, other regression, or both). Age at evaluation and sex were adjusted for in a regression model.*

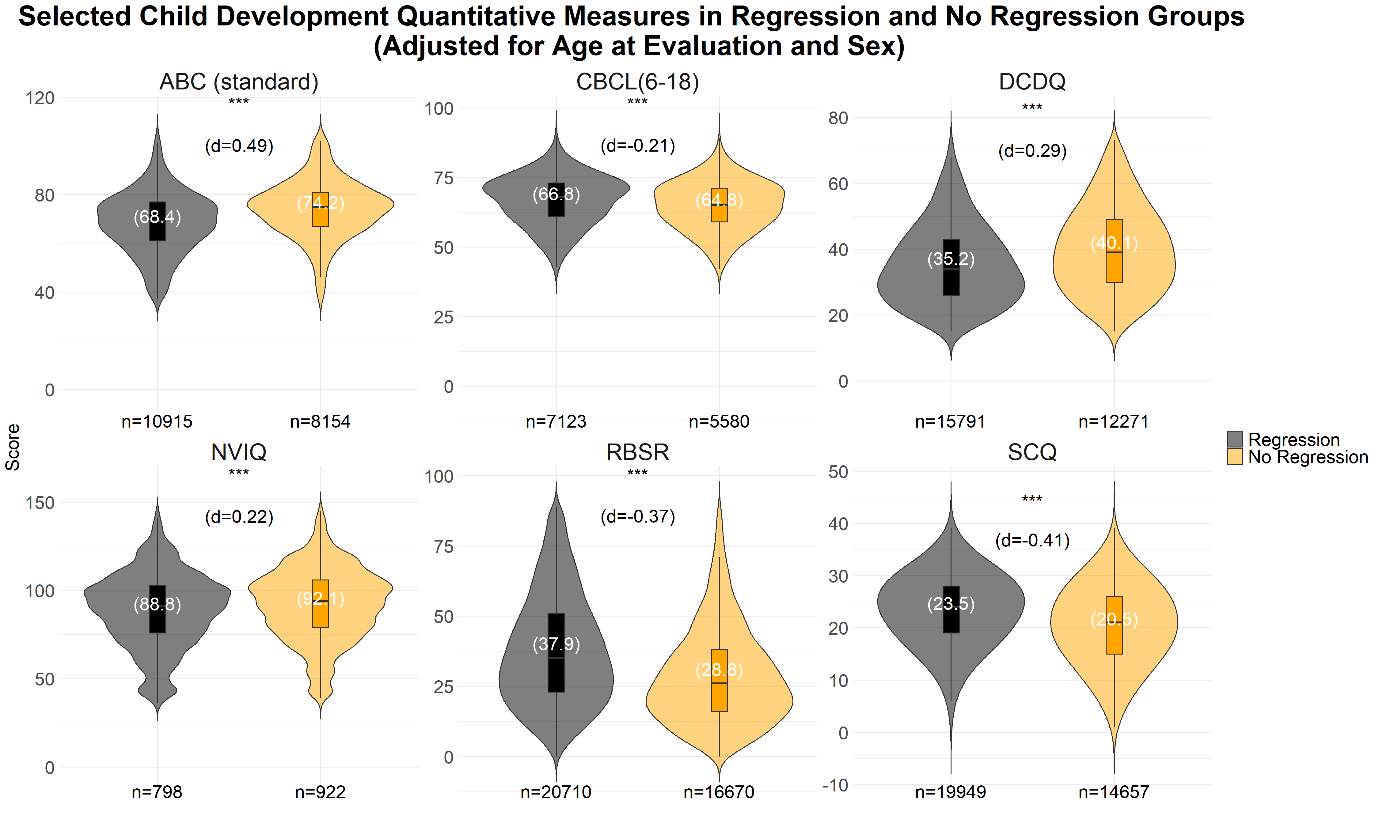

*Supplementary Figure 13. Comparing the scores of selected quantitative measures in the regression and no regression groups. In the no regression category, we included individuals without regression who were >7.08 years old at evaluation, which was the age at which 95% of individuals in SPARK v14 phenotype data with autistic regression developed regression. In the regression category we included all individuals with autistic regression (language regression, other regression, or both). Age at evaluation and sex were adjusted for in a regression model. Abc_standard, Vineland-3 adaptive behavior scales standard Adaptive Behavior Composite (ABC) score; cbcl_6_18, Child Behavior Checklist 6-18 years old total problems T score; dcdq, Developmental Coordination Disorder Questionnaire final score; nviq, Non-Verbal Intelligence Quotient overall full scale IQ score; rbsr, Repetitive Behavior Scale-Revised; scq, Social-Communication Questionnaire – Lifetime total final score.*

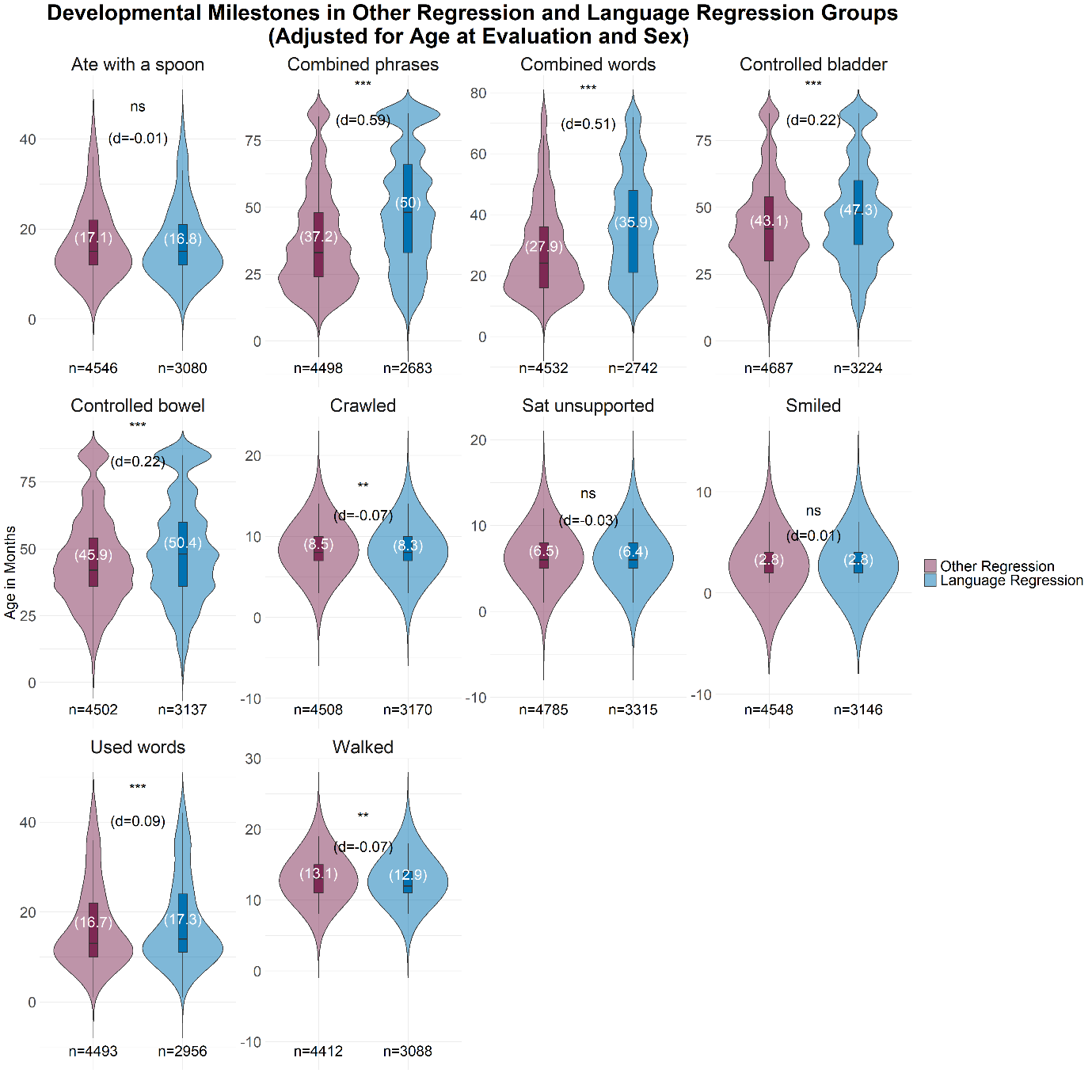

*Supplementary Figure 14. Comparing ages at reaching developmental milestones in the language and other regression groups. Here we compare individuals with only one type of regression. In the language regression category, we included individuals with only language regression and were > 7.08 years old at evaluation which was the age at which 95% of individuals with regression in SPARK v14 phenotype data manifested other regression. In the other regression category, we included individuals with only other regression and were > 4.5 years old at evaluation which was the age at which 95% of individuals with regression in SPARK v14 phenotype data manifested language regression. Age at evaluation and sex were adjusted for in a regression model.*

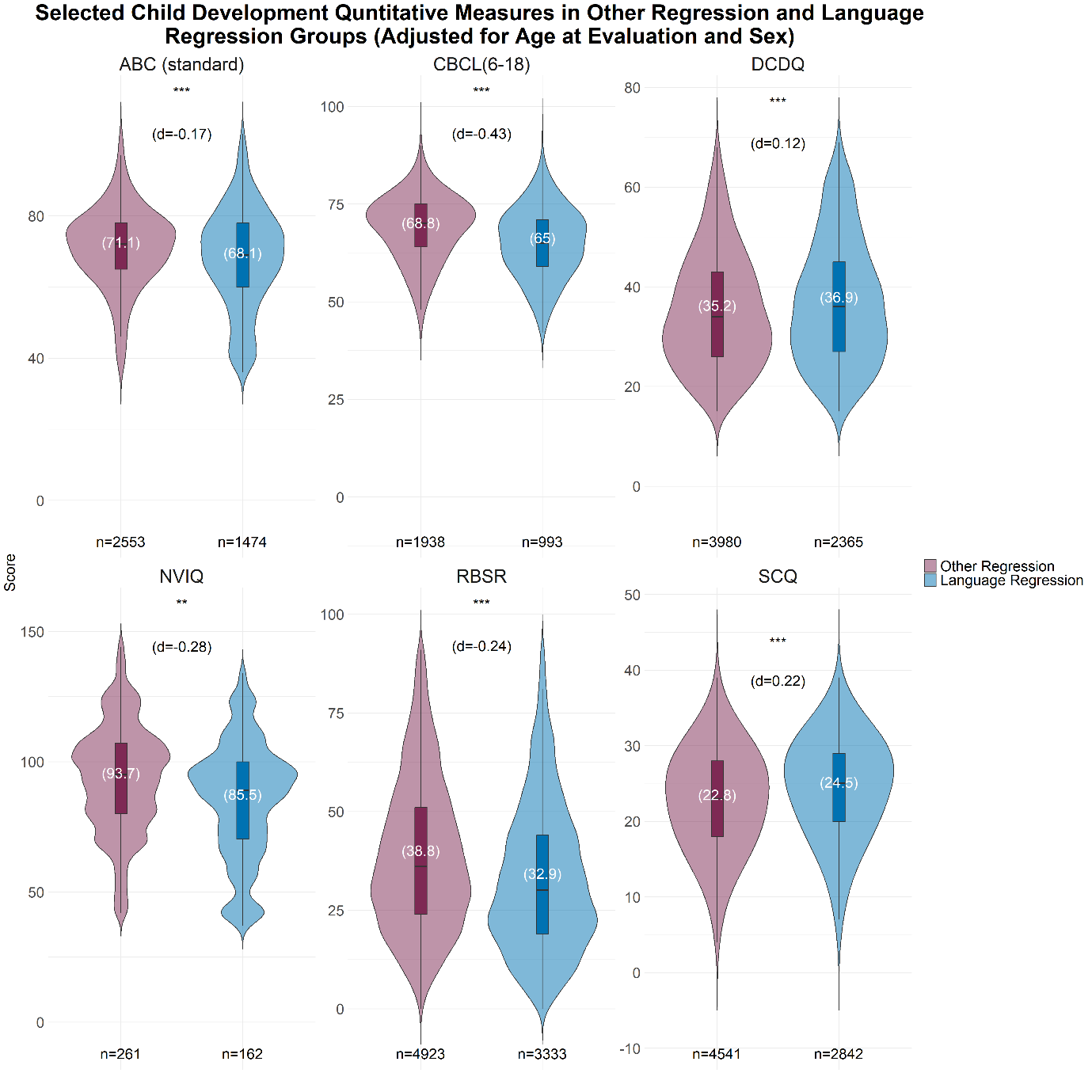

*Supplementary Figure 15. Comparing the scores of selected quantitative measures in the language and other regression groups. Here we compare individuals with only one type of regression. In the language regression category, we included individuals with only language regression and were > 7.08 years old at evaluation which was the age at which 95% of individuals with regression in SPARK v14 phenotype data manifested other regression. In the other regression category, we included individuals with only other regression and were > 4.5 years old at evaluation which was the age at which 95% of individuals with regression in SPARK v14 phenotype data manifested language regression. Age at evaluation and sex were adjusted for in a regression model. Abc_standard, Vineland-3 adaptive behavior scales standard Adaptive Behavior Composite (ABC) score; cbcl_6_18, Child Behavior Checklist 6-18 years old total problems T score; dcdq, Developmental Coordination Disorder Questionnaire final score; nviq, Non-Verbal Intelligence Quotient overall full scale IQ score; rbsr, Repetitive Behavior Scale-Revised; scq, Social-Communication Questionnaire – Lifetime total final score.*

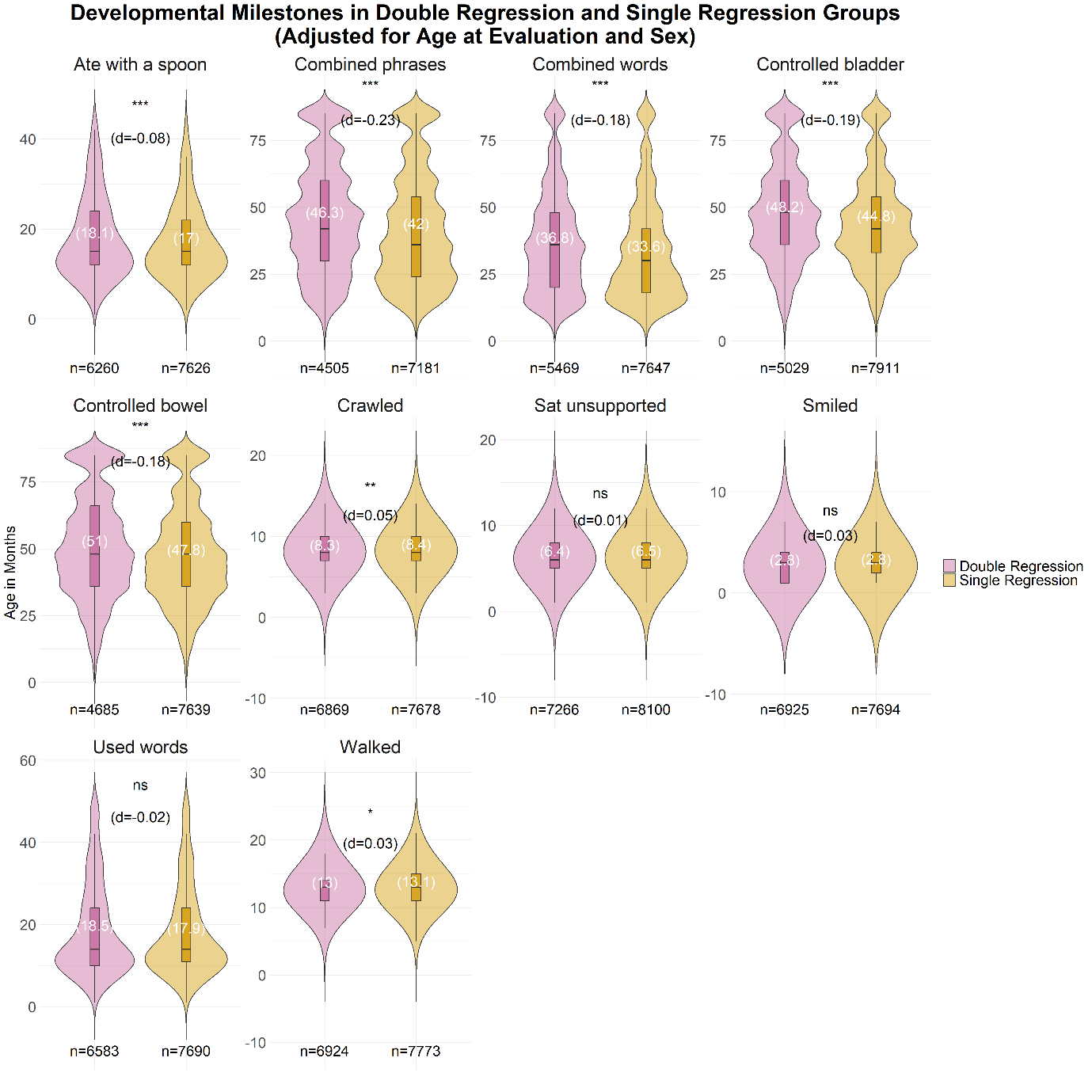

*Supplementary Figure 16. Comparing ages at reaching developmental milestones in the single regression and double regression groups. Double regression indicates both language and other regression while single regression indicates only one of them. In the single regression category we included only individuals with language regression and were > 7.08 years old at evaluation or individuals with other regression and were > 4.5 years old at evaluation which were the ages at which 95% of individuals with regression in SPARK v14 phenotype data manifested other and language regression, respectively. Age at evaluation and sex were adjusted for in a regression model.*

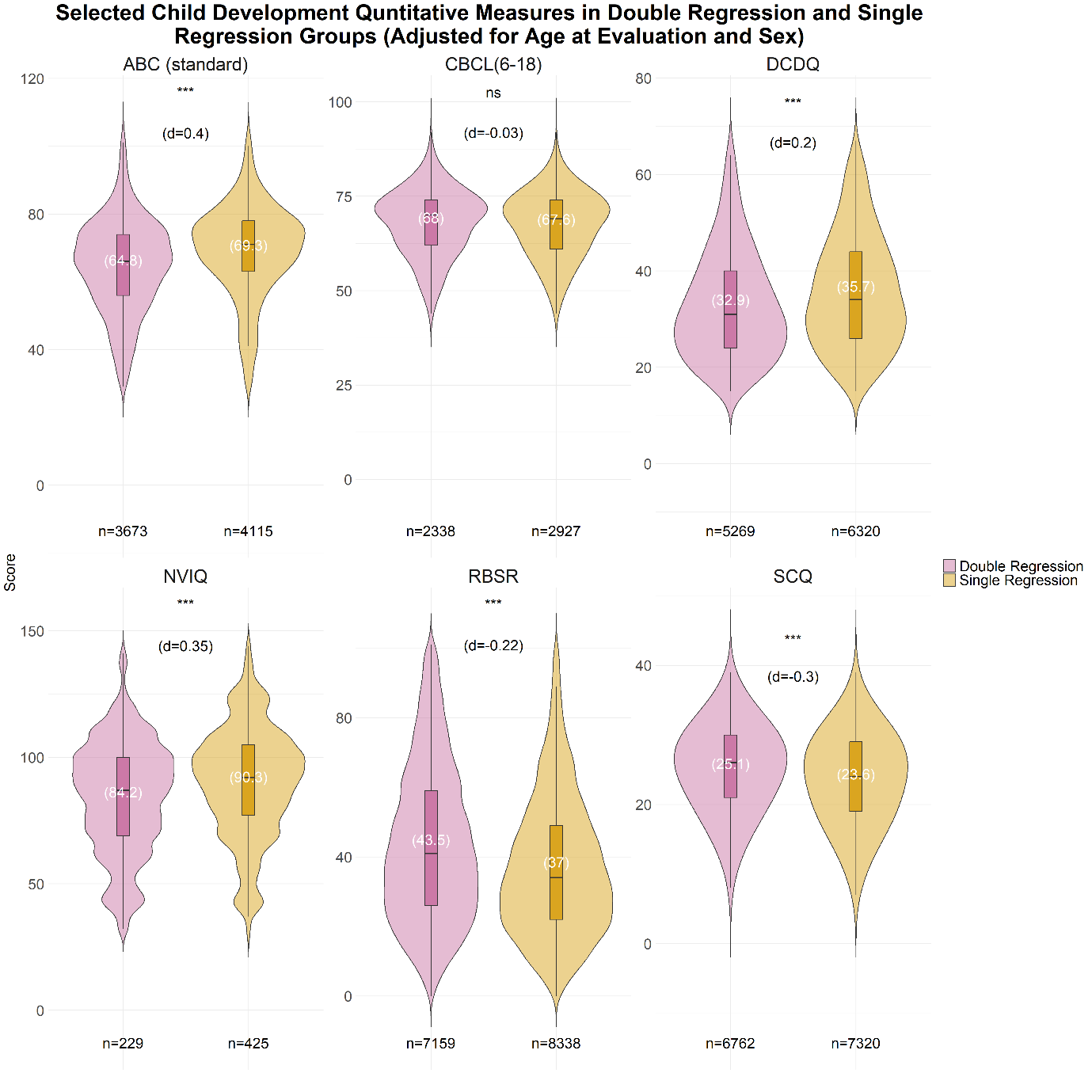

*Supplementary Figure 17. Comparing the scores of selected quantitative measures in the single and double regression groups. Double regression indicates both language and other regression while single regression indicates only one of them. In the single regression category we included only individuals with language regression and were > 7.08 years old at evaluation or individuals with other regression and were > 4.5 years old at evaluation which were the ages at which 95% of individuals with regression in SPARK v14 phenotype data manifested other and language regression, respectively. Age at evaluation and sex were adjusted for in a regression model. Abc_standard, Vineland-3 adaptive behavior scales standard Adaptive Behavior Composite (ABC) score; cbcl_6_18, Child Behavior Checklist 6-18 years old total problems T score; dcdq, Developmental Coordination Disorder Questionnaire final score; nviq, Non-Verbal Intelligence Quotient overall full scale IQ score; rbsr, Repetitive Behavior Scale-Revised; scq, Social-Communication Questionnaire – Lifetime total final score.*

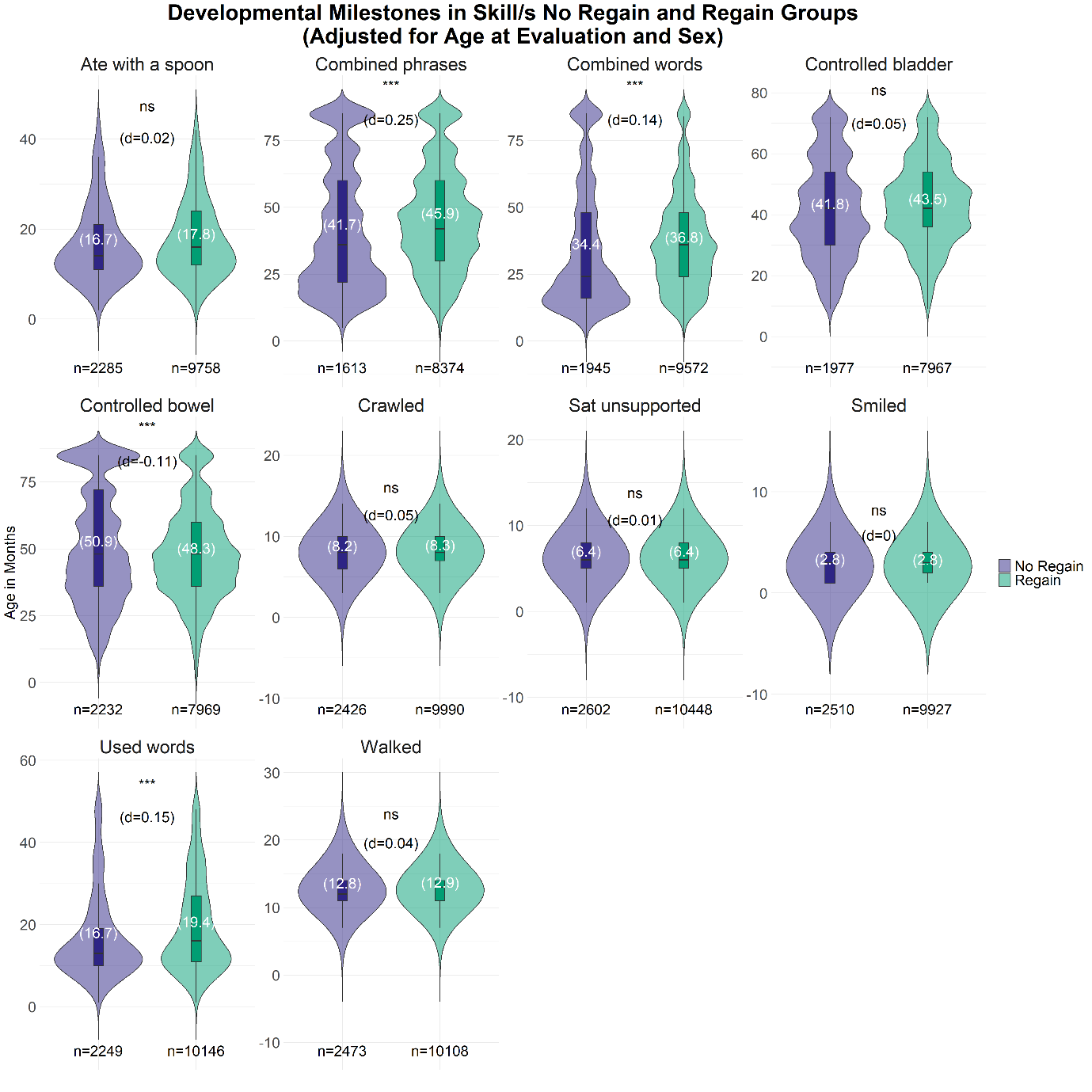

*Supplementary Figure 18. Comparing ages at reaching developmental milestones in the skill/s regain and no regain groups. Individuals in the no regain group were >5 years old at evaluation, which was the age at which ~95% of individuals with regression and regain in SPARK v14 phenotype data regained their lost skills. Age at evaluation and sex were adjusted for in a regression model.*

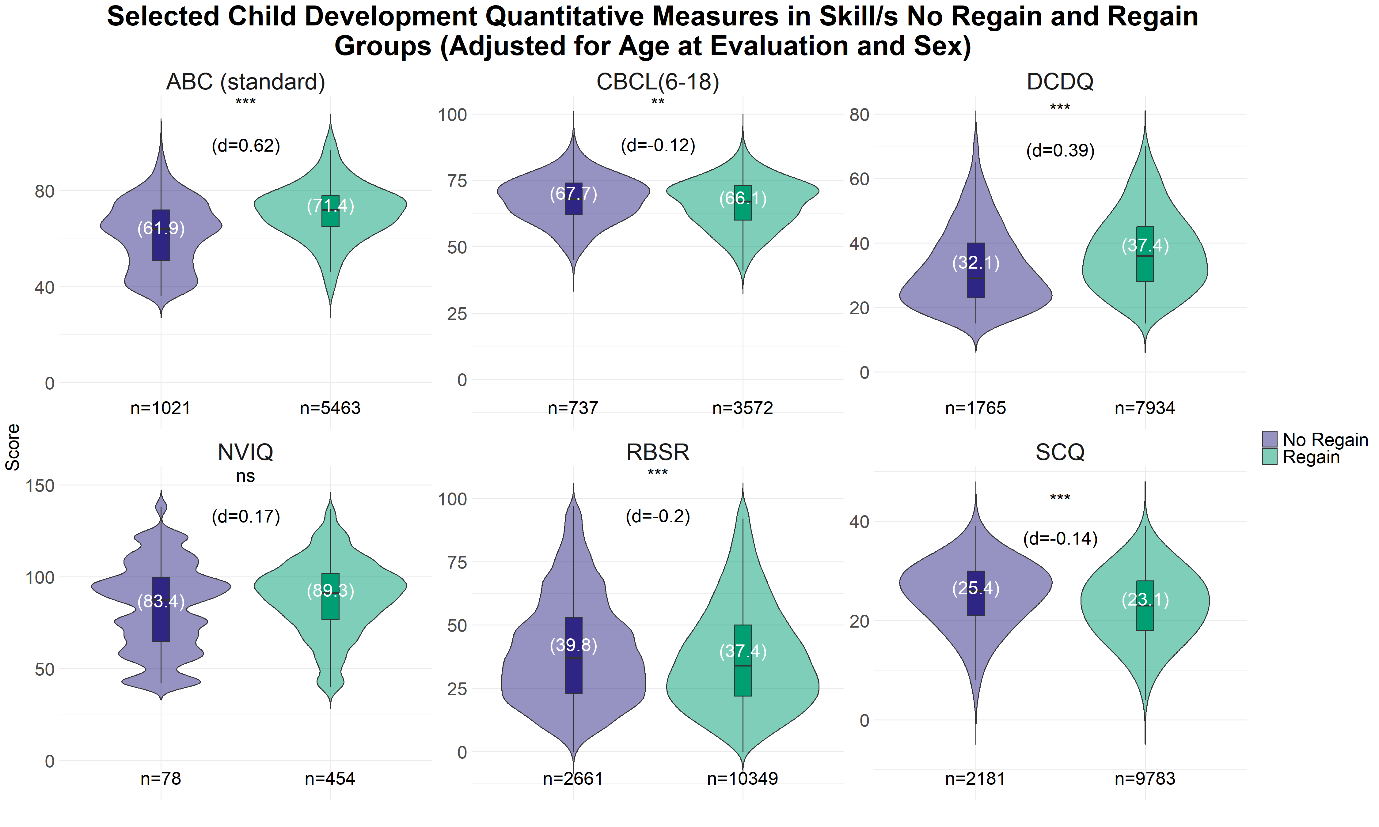

*Supplementary Figure 19. Comparing the scores of selected quantitative measures in the skill/s regain and no regain groups. Individuals in the no regain group were >5 years old at evaluation, which was the age at which ~95% of individuals with regression and regain in SPARK v14 phenotype data regained their lost skills. Age at evaluation and sex were adjusted for in a regression model. Abc_standard, Vineland-3 adaptive behavior scales standard Adaptive Behavior Composite (ABC) score; cbcl_6_18, Child Behavior Checklist 6-18 years old total problems T score; dcdq, Developmental Coordination Disorder Questionnaire final score; nviq, Non-Verbal Intelligence Quotient overall full scale IQ score; rbsr, Repetitive Behavior Scale-Revised; scq, Social-Communication Questionnaire – Lifetime total final score.*

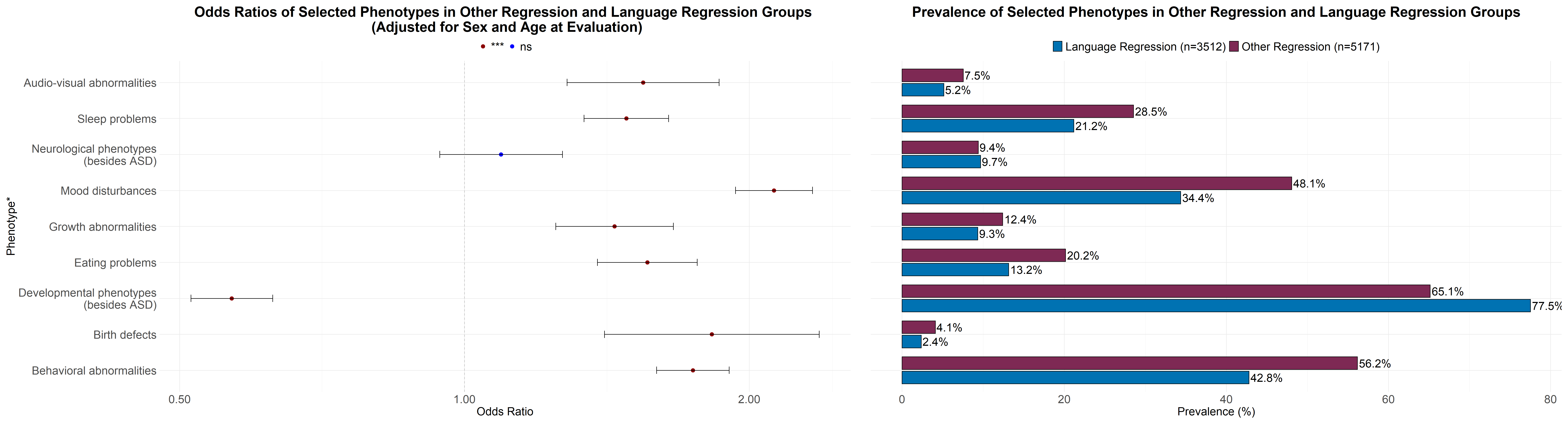

*Supplementary Figure 20. Comparing the occurrence of selected phenotypes in other regression and language regression groups. Here we compare individuals with only one type of regression. In the language regression category, we included individuals with only language regression and were > 7.08 years old at evaluation which was the age at which 95% of individuals with regression in SPARK v14 phenotype data manifested other regression. In the other regression category, we included individuals with only other regression and were > 4.5 years old at evaluation which was the age at which 95% of individuals with regression in SPARK v14 phenotype data manifested language regression. Age at evaluation and sex were adjusted for in a regression model.*

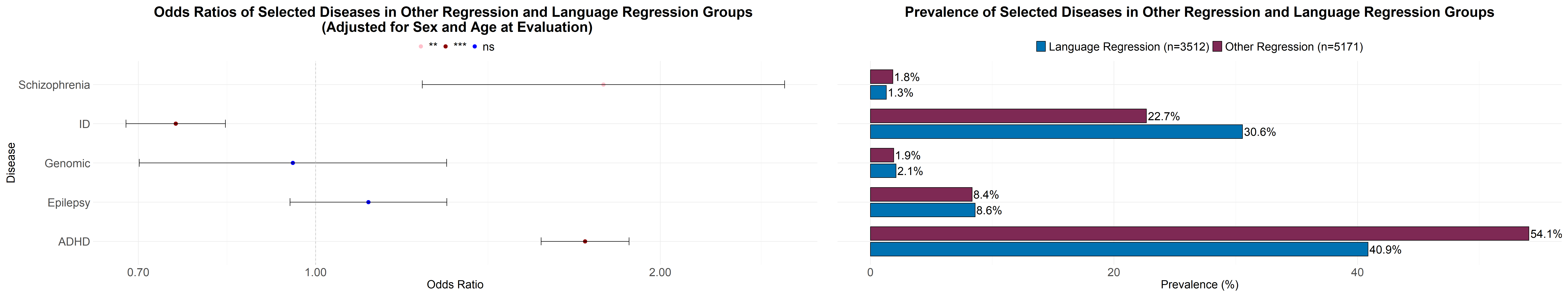

*Supplementary Figure 21. Comparing the occurrence of selected diseases in other regression and language regression groups. Here we compare individuals with only one type of regression. In the language regression category, we included individuals with only language regression and were > 7.08 years old at evaluation which was the age at which 95% of individuals with regression in SPARK v14 phenotype data manifested other regression. In the other regression category, we included individuals with only other regression and were > 4.5 years old at evaluation which was the age at which 95% of individuals with regression in SPARK v14 phenotype data manifested language regression. Age at evaluation and sex were adjusted for in a regression model.*

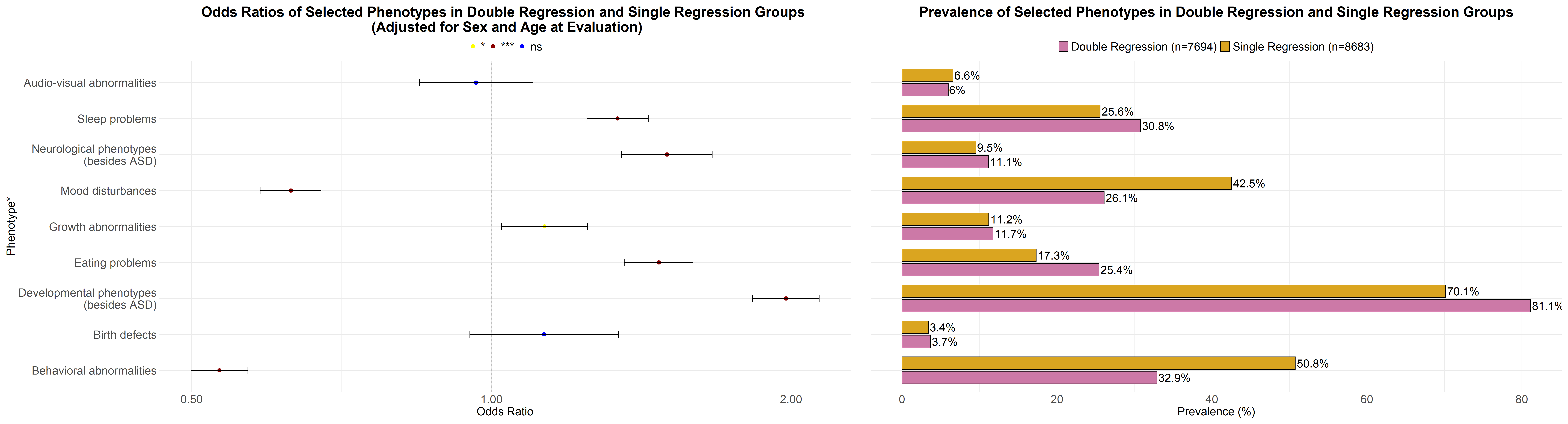

*Supplementary Figure 22. Comparing the occurrence of selected phenotypes in double regression and single regression groups. Double regression indicates both language and other regression while single regression indicates only one of them. In the single regression category we included only individuals with language regression and were > 7.08 years old at evaluation or individuals with other regression and were > 4.5 years old at evaluation which were the ages at which 95% of individuals with regression in SPARK v14 phenotype data manifested other and language regression, respectively. Age at evaluation and sex were adjusted for in a regression model.*

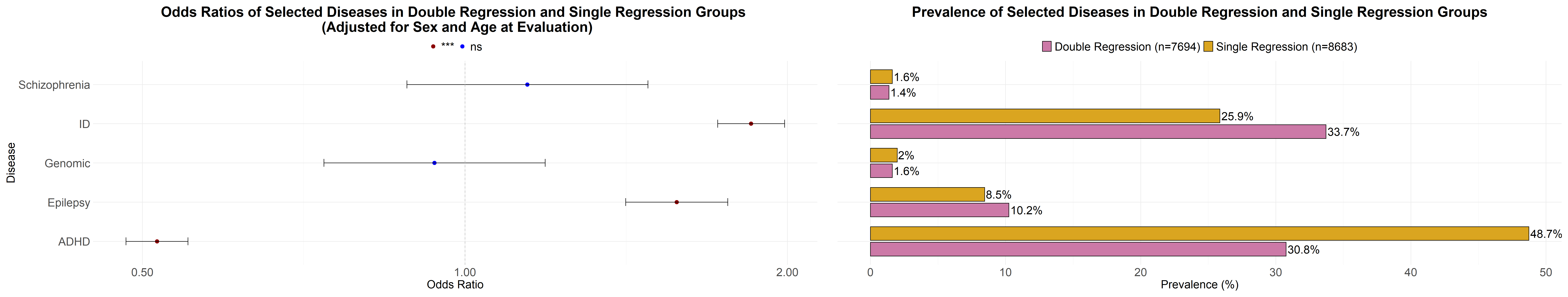

*Supplementary Figure 23. Comparing the occurrence of selected diseases in double regression and single regression groups. Double regression indicates both language and other regression while single regression indicates only one of them. In the single regression category we included only individuals with language regression and were > 7.08 years old at evaluation or individuals with other regression and were > 4.5 years old at evaluation which were the ages at which 95% of individuals with regression in SPARK v14 phenotype data manifested other and language regression, respectively. Age at evaluation and sex were adjusted for in a regression model.*

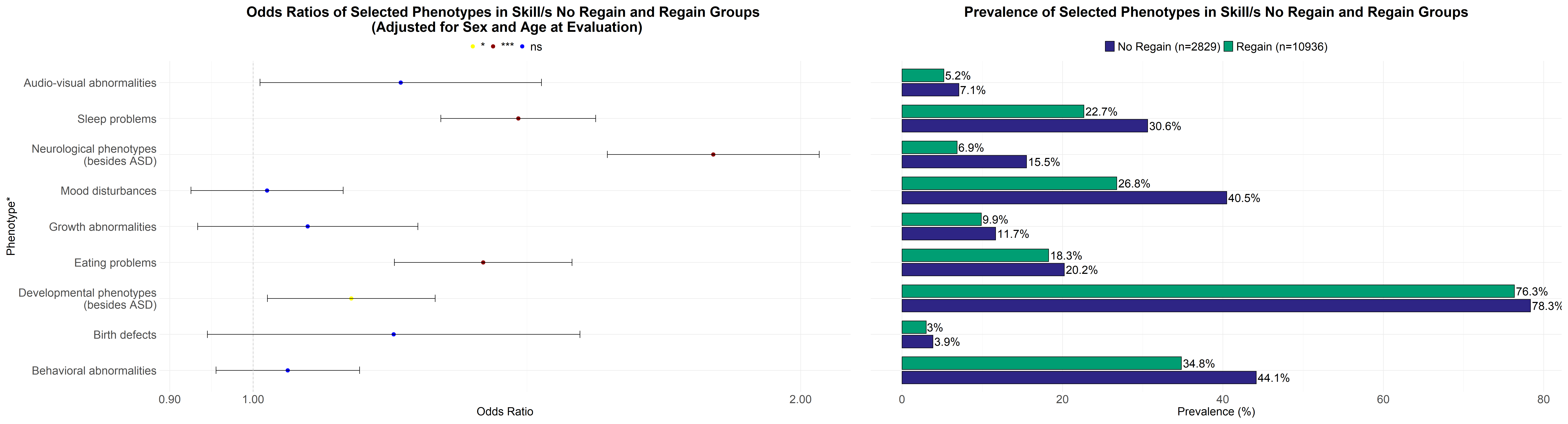

*Supplementary Figure 24. Comparing the occurrence of selected phenotypes in the skill/s regain and no regain groups. In the skill/s regain category we included individuals who regained all of their previously lost skills (other regression, language regression, or both). In the no regain category we included individuals who did not regain any of their lost skills. Individuals in the no regain group were >5 years old at evaluation, which was the age at which ~95% of individuals with regression and regain in SPARK v14 phenotype data regained their lost skills. Age at evaluation and sex were adjusted for in a regression model.*

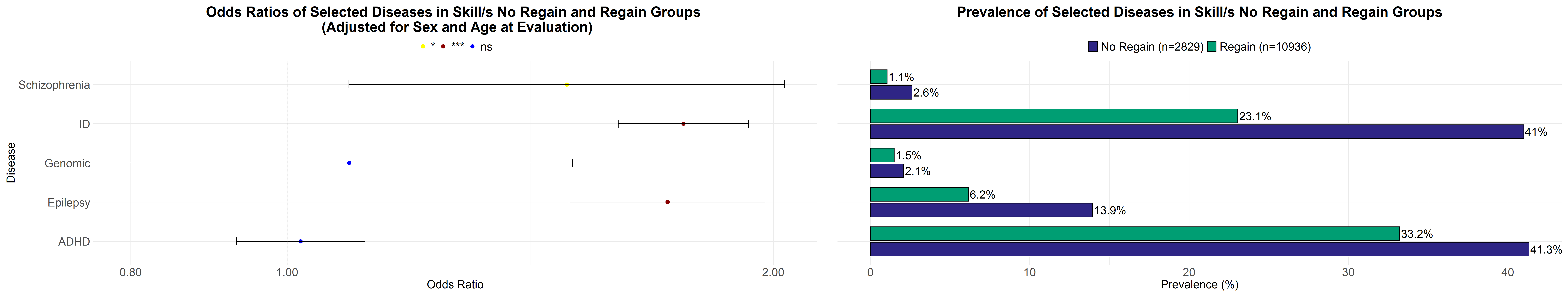

*Supplementary Figure 25. Comparing the occurrence of selected diseases in the skill/s regain and no regain groups. In the skill/s regain category we included individuals who regained all of their previously lost skills (other regression, language regression, or both). In the no regain category we included individuals who did not regain any of their lost skills. Individuals in the no regain group were >5 years old at evaluation, which was the age at which ~95% of individuals with regression and regain in SPARK v14 phenotype data regained their lost skills. Age at evaluation and sex were adjusted for in a regression model.*

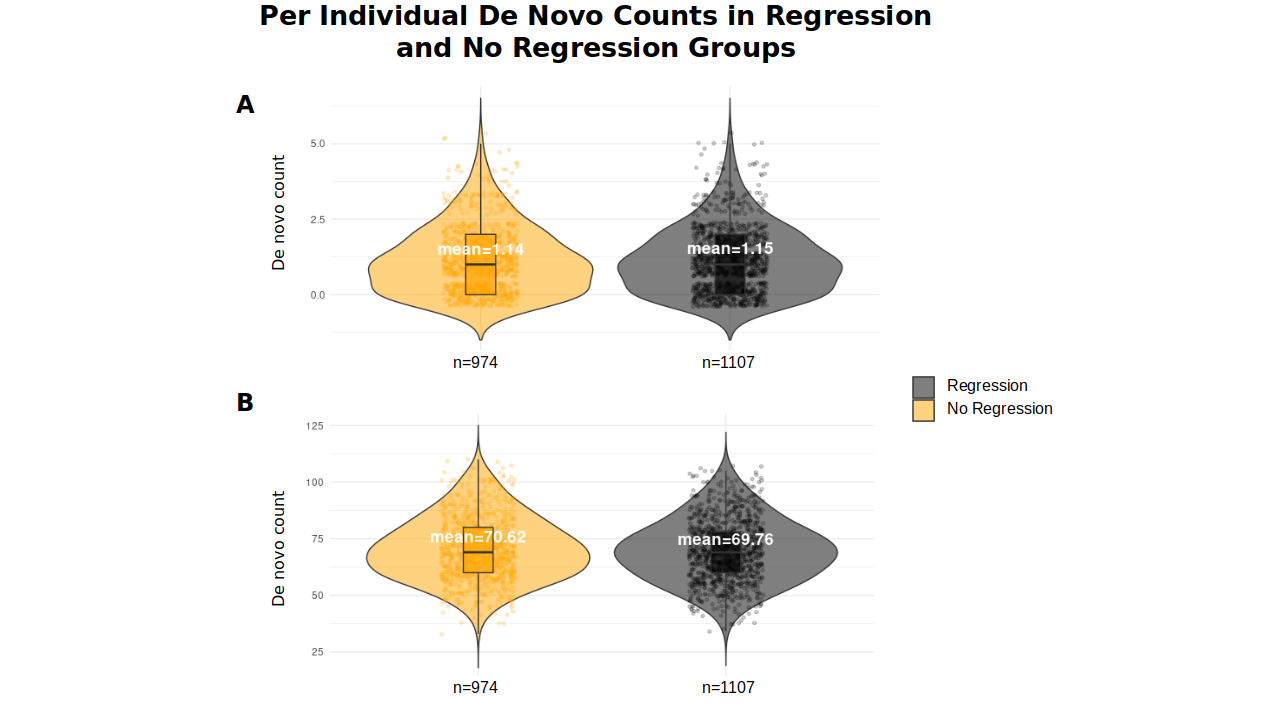
*Supplementary Figure 26. Per individual de novo counts in the regression and no regression groups. A. De novo counts in the coding DNA sequence obtained by selecting only coding sequences. B. De novo counts in the coding and non-coding DNA sequence obtained from genome sequencing.*
